# Multi-omic and functional screening reveal targetable vulnerabilities in *TP53* mutated multiple myeloma

**DOI:** 10.1101/2024.08.23.24312359

**Authors:** Dimitrios Tsallos, Nemo Ikonen, Juho J. Miettinen, Muntasir Mamun Majumder, Samuli Eldfors, Imre Västrik, Alun Parsons, Minna Suvela, Katie Dunphy, Paul Dowling, Despina Bazou, Peter O’Gorman, Juha Lievonen, Raija Silvennoinen, Pekka Anttila, Caroline A. Heckman

## Abstract

Despite development of several effective therapies for multiple myeloma (MM), the prognosis of patients with partial deletion of chromosome 17 (del(17p)) and *TP53* aberrations remains poor. By applying comprehensive multi-omics profiling analyses (whole exome and transcriptome sequencing plus proteomics) and functional *ex vivo* drug screening to samples from 167 patients with MM, we uncovered novel therapeutic vulnerabilities specific to *TP53* mutated MM. Our findings revealed a distinct sensitivity profile to a range of inhibitors (mitotic, topoisomerase, HDAC, HSP90, IGF1R and PI3K/AKT/mTOR inhibitors) irrespective of 17p deletion status. Conversely, no increase in sensitivity was observed for monoallelic *TP53* (del(17p) with WT *TP53*) when compared to WT *TP53* samples, highlighting the remaining unmet clinical need. Notably, plicamycin, an RNA synthesis inhibitor linked to modulation of chromatin structure and increased transcription, emerged as particularly efficacious for *TP53* mutated MM. The increased sensitivity correlated with higher protein expression of the drug targets: HDAC2, HSP90AA1 and multiple ribosomal subunits. Additionally, we observed increased RNA expression of G2M checkpoint, E2F targets and mTORC1 signaling in our cohort and the MMRF-CoMMpass (NCT01454297) study in *TP53* mutated MM. Harmonization of multi-omics data with *ex vivo* drug screening results revealed that *TP53* mutated MM is functionally distinct from MM with monoallelic *TP53*, and demonstrates that MM with mutated *TP53*, with and without del(17p), may be targetable by approved drugs. These results further indicate the need for regular monitoring by sequencing to identify these patients.

**KEY POINTS:** *TP53* mutation in myeloma confers sensitivity to multiple compounds, including approved drugs, irrespective of del(17p) status.

*TP53* mutated myeloma links to higher expression of drug targets involved in cell proliferation, mRNA processing, and chromatin modulation.

## INTRODUCTION

Multiple myeloma (MM) is a complex and incurable disease characterized by a clonal proliferation of plasma cells in the bone marrow (BM). Significant advancements in MM treatment modalities have been made over the past decades^1^, but despite the availability of novel treatments, the clinical and genetic heterogeneity of the disease continues to complicate prognosis and therapeutic strategies^1^. Genetic aberrations traditionally detected by fluorescence *in situ* hybridization (FISH) are known to play a pivotal role in MM progression and treatment outcomes^2^. Transitioning from FISH to next generation sequencing (NGS) could enhance the detection of recurrent aberrations to driver genes such as *TP53*, offering a more comprehensive genomic landscape and potentially refining therapeutic approaches and management of patients with MM^2^.

Among the genetic aberrations influencing MM prognosis, deletion of the p arm of chromosome 17, del(17p), stands out as a high-risk factor with profound implications for patient prognosis and response to treatment^2,3^. Del(17p) is identified in 10% of patients with newly diagnosed MM and is primarily monoallelic. Importantly, the *TP53* gene is located within the minimally deleted region on 17p13. The co-occurrence of del(17p) together with *TP53* mutation, often referred to as ‘double hit’ MM, is associated with worse prognosis^2,3^. *TP53* mutations alone are detected in 1-7% of newly diagnosed patients with MM^4–6^. However, both *TP53* mutation and del(17p) aberrations are more prevalent after relapse occurring in approximately 23-45% of cases^5,7^. The emergence of *TP53* mutations as a key risk factor for stratification is evident given the significant adverse prognosis, even in the absence of del(17p)^6,8^. Moreover, *TP53* mutations in various cancers can lead to gain-of-function or dominant negative phenotypes, altering treatment response^9^.

Functional *ex vivo* drug sensitivity testing of patients’ tumor cells to hundreds of inhibitors can reveal cell dependencies and molecular vulnerabilities while accounting for patient variability^10–13^. Here we show that integration of drug sensitivity data with genomic, transcriptomic, and proteomic data from the same samples revealed a deeper functional view of the impact of *TP53* aberrations in CD138+ MM cells. The presence of *TP53* mutation associated with increased sensitivity to approved drugs, including conventional chemotherapeutics, irrespective of del(17p) status. In contrast, monoallelic *TP53* samples had a distinct molecular profile without a significant change in *ex vivo* drug responses. Overall, we explored novel therapeutic options for *TP53* aberrations in MM and identified vulnerabilities associated with *TP53* mutation to multiple compounds, providing a foundation for targeted treatment strategies to improve patient outcome.

## MATERIALS AND METHODS

### Patients and samples

Bone marrow (BM) aspirates and skin biopsies were collected from patients under approved protocols (239/13/03/00/2010 and 303/13/03/01/2011) in line with the Declaration of Helsinki as previously described^10^. Following Ficoll (GE Healthcare) gradient separation of the mononuclear cell fraction, CD138+ cells were enriched by immunomagnetic bead separation (StemCell Technologies) and used for downstream assays. Interphase FISH was conducted according to the European Myeloma Network 2012 guidelines^29^. Patient characteristics and assays performed are detailed in **Table 1**.

**Table 1.**
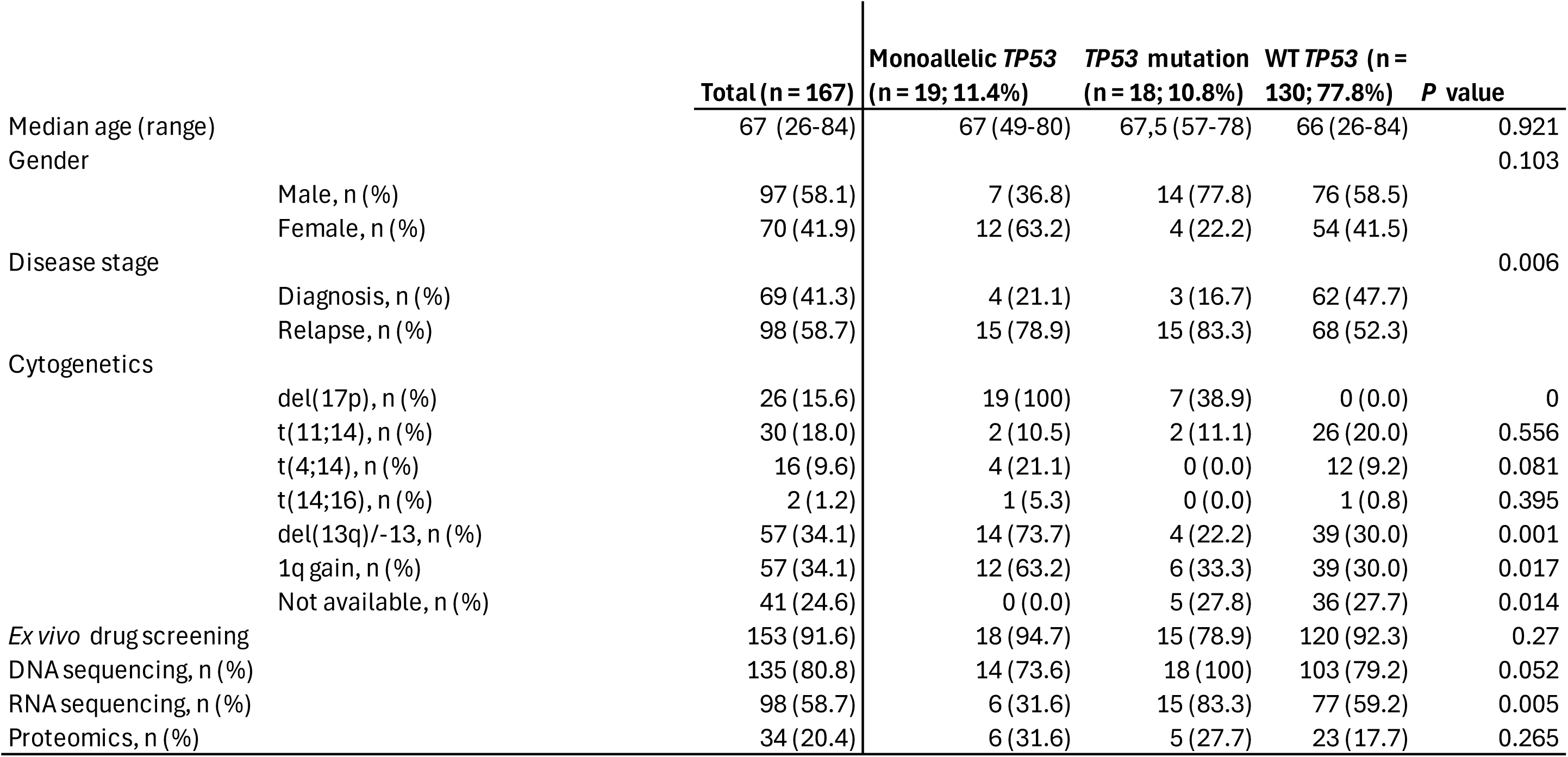
Summarized characteristics of MM patients providing samples to the FIMM cohort and the analyses performed.

### Ex vivo drug screening and analysis

Drug sensitivity testing was conducted using an established *ex vivo* drug screening method on CD138+ cells, with viability measured by CellTiter-Glo reagent (Promega) after 72 hours^10–12^. BM CD138+ cells were added to pre-drugged plates, which contained up to 348 compounds (**supplemental Table 1**). Quality control analysis, dose response curve fitting, and drug sensitivity scores (DSS) were calculated as previously described^15^.

### Whole Exome Sequencing

Genomic DNA was extracted from skin biopsies and CD138+ BM cells using the Qiagen DNeasy Blood & Tissue kit or the Qiagen AllPrep DNA/RNA/miRNA Universal kit. The isolated DNA was processed, exome libraries prepared and sequenced as described previously^11,31^. Somatic mutations were identified and annotated using established methods^32^. The presence of a mutation is considered if the variant frequency exceeded 5% (somatic p-value ≤ 0.05).

### RNA Sequencing

RNA from CD138+ cells isolated using the Qiagen AllPrep DNA/RNA/miRNA Universal kit, RNAseq libraries were prepared using ribosome depletion, data pre-processing and the analysis pipeline were carried out as described earlier^33^. Differential gene expression (DGE) analysis was performed using DESeq2 R package (1.36.0)^34^ from log_2_CPM raw counts calculated with edgeR after TMM normalized. Within DESeq2, raw count data were filtered to remove genes with less than 10 reads in at least 95% of the samples. For the gene set enrichment analysis, we used clusterProfiler (4.4.4)^35^ package and MSigDB (7.5.1) and from that H (Hallmark) and C5 (GO) genesets, excluding human phenotype ontology sets (HPO) ^36,37^.

### Label-Free LC-MS/MS Analysis and Data Processing of CD138+ cells

500 ng of each digested sample was analysed using Q-Exactive (ThermoFisher Scientific, Hemel Hempstead, UK) high-resolution accurate mass spectrometer connected to a Dionex Ultimate 3000 (RSLCnano) chromatography system (ThermoFisher Scientific, Hemel Hempstead, UK). Peptides were separated and data acquired with MS/MS scans. Protein identification and label-free quantification (LFQ) normalization were performed using MaxQuant v1.5.2.8 (http://www.maxquant.org), with subsequent data analysis using Perseus. In our figures, uniprot accession IDs are presented as gene symbol IDs. More details are provided in the **supplemental Methods**.

### Validation using public data

We utilized the IA21 version of the MMRF-CoMMpass study (NCT01454297), available from MMRF research gateway (https://research.themmrf.org/). Only CD138+ BM samples at diagnosis were selected. The del(17p) status was determined using available seqFISH data^38^. DGE analysis was performed as described above. Survival analysis was performed in R using survival (3.5-7)^39^ and survminer (0.4.9)^40^ packages.

### Statistical Analyses

Statistical analysis was done in R. Normality of the data was assessed using Shapiro-Wilk test and based on the result either the Welch Two Sample t-test or Wilcoxon rank-sum test were applied to subsequent analysis. Categorical data was analyzed using Fisher’s exact test or chi-square test according to the sample size. Kruskal-Wallis Test was applied to estimate the difference between the age distribution of the groups. Resulted p-values were adjusted with Benjamini-Hochberg. Survival analysis employed the Kaplan-Meier method and Cox proportional hazards models.

## RESULTS

### Study sample characteristics

Our study utilized data from 167 individuals receiving treatment throughout Finland. Collected BM samples were processed for mononuclear cell selection and immunomagnetic separation of CD138+ cells. DNA from the selected cells and a matching skin biopsy underwent whole exome sequencing to identify somatic mutations. Subsets of CD138+ cells were also used for bulk RNA sequencing and proteomic evaluation (**Figure 1A, B**). Additionally, the *ex vivo* sensitivity of the same CD138+ MM patient cells was assessed against an oncology drug collection of 348 small molecule inhibitors, with 153 total screened samples (**supplemental Table S1**). Other clinical data, such as karyotyping, was available and summarized information is provided in **Table 1**.

**Figure 1.**
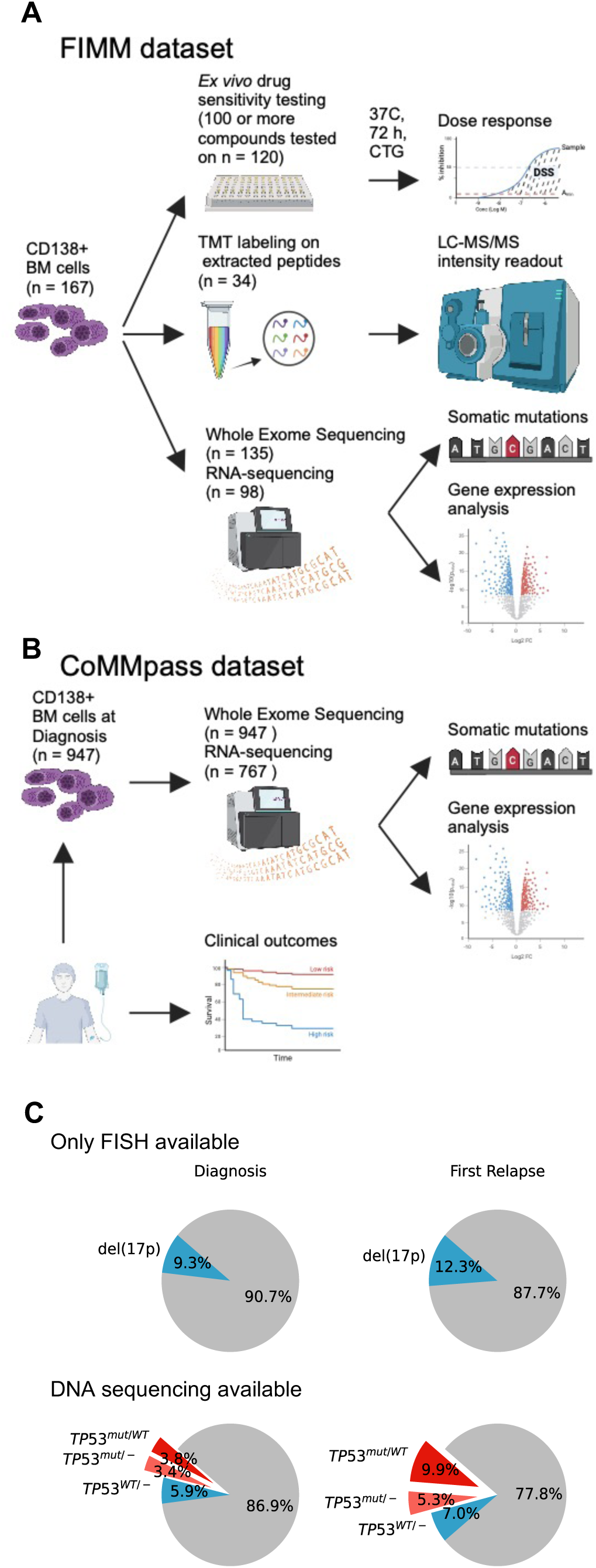
Workflow and patient stratification according to *TP53* status. (A) CD138+ cells from 167 bone marrow samples were molecularly and functionally profiled depending on viable cell numbers. (B) Dataset used from the MMRF-CoMMpass study for validation purposes. (C) Comparison of patient *TP53* status with and without DNA sequencing in the MMRF-CoMMpass patient population. Remaining patient proportion in grey represents wild-type (WT) *TP53* status.

When focusing on *TP53* aberrations, the use of next-generation sequencing provides a more detailed view of the patient subpopulations as presented in the MMRF-CoMMpass cohort (**Figure 1C**). Characteristically, *TP53* mutation becomes more prevalent at relapse (15.2%) compared to diagnosis (7.2%). Given that mutated *TP53* leads to functional impairment of wild-type (WT) p53 protein^9,14^, we grouped all samples with *TP53* mutation (*TP53*^mut/-^ and *TP53*^mut/WT^) in one category. This includes samples with additional detection of del(17p), also known as double-hit MM. Samples containing only wild-type copies of *TP53* but presented del(17p) from karyotyping are referred to as monoallelic *TP53* from here on. Five samples lacking sequencing results but with a detected del(17p) karyotype are also included in this category, for a total of 19 monoallelic *TP53* patient samples. The remaining samples (n = 130) were *TP53* WT.

### Presence of TP53 mutation is associated with increased sensitivity to multiple drugs

In search of novel therapeutic vulnerabilities in high-risk patients harboring *TP53* aberrations, we analyzed data from our *ex vivo* drug screening platform. To measure the drug sensitivity of the samples, we used the drug sensitivity score (DSS), which is a modified version of the area under the curve. The DSS allows for comparison of dose-response curves across a range of concentrations, providing a single measure of the sensitivity of the cancer cells to the drug^15^. Confirming the difficulty to treat del(17p) patients, CD138+ MM cells with monoallelic *TP53* showed no significant increase in *ex vivo* sensitivity to the tested compounds. Conversely, samples with *TP53* mutation, regardless of the presence of del(17p), displayed higher sensitivity to certain mitotic inhibitors and inhibitors targeting RNA synthesis, HDAC, topoisomerase, HSP90, and the PI3K/AKT/mTOR pathway (**Figure 2A,B; supplemental Table S2**). The mutant p53 targeting compound APR-246 demonstrated enhanced activity in *TP53* mutated MM, suggesting that it effectively reactivates the mutant p53 protein leading to cell death. The biological significance of the specific grouping is also observed for monoallelic *TP53* MM where increased resistance to the MDM2 antagonists idasanutlin (median difference = - 6.8; p-value = 0.074) and nutlin-3 (median difference = -2.6; p-value = 0.014) was observed. Resistance to MDM2 antagonists suggests the reduced availability of p53 in samples with monoallelic *TP53*, while the activity of APR-246 highlights the dominant effect of the mutant p53 protein (**Figure 2C**). The impact of *TP53* mutation on *ex vivo* drug response is highlighted by the comparison of DSS between *TP53* mutated and monoallelic *TP53* samples (**supplemental Figure S1**).

**Figure 2.**
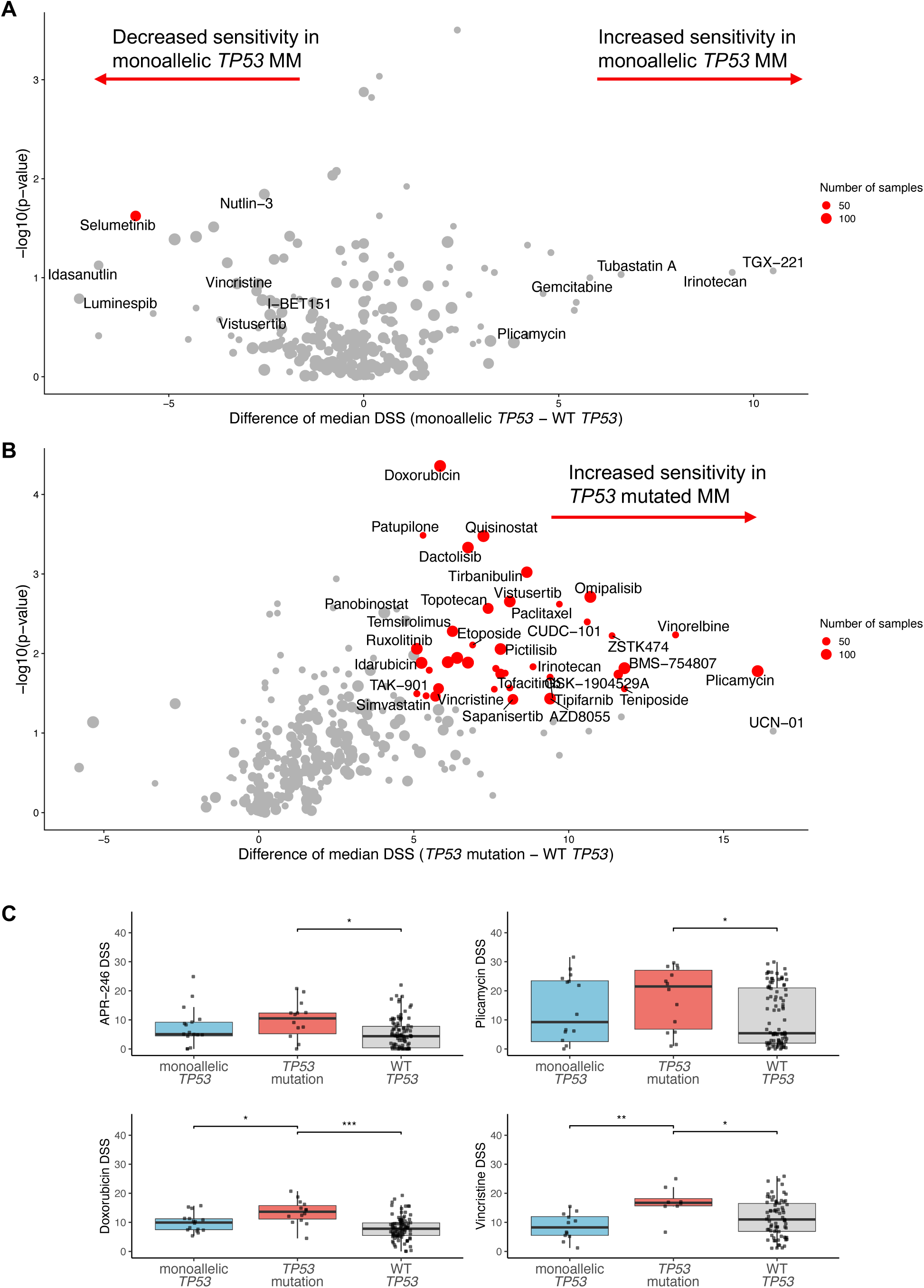
*Ex vivo* drug screening identifies compounds with enhanced activity in MM patient samples with *TP53* mutation. (A) Volcano plot illustrating the comparison of drug sensitivity between MM samples with monoallelic *TP53* and WT *TP53* samples. No compounds presented significantly increased sensitivity for samples with monoallelic *TP53*. (B) Samples harboring *TP53* mutations, irrespective of del(17p) status, exhibit increased sensitivity to a range of therapeutic agents. The volcano plot’s x-axis shows the median difference in DSS between the two groups, while the y-axis represents the negative logarithm (base10) of the p-value obtained from the Wilcoxon rank-sum test. Data points represent individual compounds, with the datapoint size reflecting the number of samples tested per compound. Significant hits with median difference above 5 are highlighted in red, and a few additional selected compounds are also labeled but remain in grey. (C) Box plot showing the distribution of *ex vivo* sensitivity to selected compounds. DSS below 10 indicates overall resistance to the compounds. The center line represents the median value. The upper and lower limits of the box represent the upper (75th percentile) and lower (25th percentile) quartiles, respectively. The whiskers extend to the largest and smallest values within 1.5 times the interquartile range from the quartiles. Points beyond the whiskers are outliers and are plotted individually. Statistical significance indicated as * p-value < 0.05, ** p-value < 0.01, *** p-value < 0.001 by Wilcoxon rank-sum test.

The most remarkable increase in sensitivity of *TP53* mutated MM samples was observed for plicamycin, an RNA synthesis inhibitor. The response to plicamycin was greater than that to another RNA and DNA synthesis inhibitor, dactinomycin, which also showed a significant increase in activity. Multiple conventional chemotherapeutics showed increased activity in samples with *TP53* mutation, specifically mitotic inhibitors (vinorelbine, paclitaxel, docetaxel, vincristine, ABT-751, patupilone) and topoisomerase inhibitors (teniposide, irinotecan, daunarubicin, camptothecin, topotecan, etoposide, doxorubicin, idarubicin). Many chemotherapeutics were tested on fewer samples, however, the consistency of the increased sensitivity on similar compounds supports the observation that *TP53* mutated cells are more sensitive to these drug classes (**supplemental Table S2**).

This increased activity was also evident with HDAC inhibitors quisinostat and panobinostat, both approved treatments. In a smaller set of samples, CUDC-101, targeting class I/II HDAC and EGFR also showed significantly increased activity in *TP53* mutated MM samples, while belinostat showed increased, but not a significant difference in activity (median difference = 9.5; p-value = 0.072).

The heightened activity of PI3K/AKT/mTOR pathway inhibitors in *TP53* mutated MM CD138+ cells suggested a potential dependency on growth signals involving this pathway. We showed increased *ex vivo* sensitivity to vistusertib, omipalisib, pictilisib, temsirolimus, and dactolisib. Additionally, kinase inhibitors tirbanibulin, GSK-1904529A, BMS-754807, and ruxolitinib all demonstrated increased activity in *TP53* mutated MM samples. GSK-1904529A and BMS-754807 target IGF1-R and ruxolitinib specifically targets JAK1 and JAK2 kinases, which are upstream of PI3K, AKT and mTOR. The HSP90 inhibitor tanespimycin, which indirectly suppresses the PI3K/AKT/mTOR pathway^16^, also showed significant increase in activity towards *TP53* mutated CD138+ cells.

To assess the impact of *TP53* mutation frequency on the response to these compounds, we correlated the response of each sample with the mutation frequency (**supplemental Figure S2; supplemental Table S3**). To establish correlation, we selected compounds tested in at least 5 samples and highlighted compounds with significant difference observed in *TP53* mutated MM (**supplemental Figure S2A)**. Except for JQ1, most compounds showed weak and non-significant correlation to mutation frequency, probably owing to overall limited selective toxicity of the compounds.

### TP53 mutations frequently co-occur with KRAS mutations in MM

To understand the broader genetic landscape of *TP53* aberrations in MM and search for possible confounding factors in drug response, we explored the frequency of co-occurrence of common mutations. In our cohort, almost half of the samples with *TP53* mutation also had a mutation to *KRAS*, while *KRAS* mutations were rare in samples with monoallelic *TP53* (Fisher’s exact test: p = 0.029; OR = 6.44, 95% CI = [1.01, 73.93]; **Figure 3A**). However, *KRAS* mutations were not more likely to be present in *TP53* mutated MM compared to MM with WT *TP53* (Fisher’s exact test: p = 0.080; OR = 0.378, 95% CI = [0.12, 1.21]). To assess whether *TP53* and *KRAS* mutation co-occurrence is common in MM, we analyzed diagnostic samples from the MMRF-CoMMpass study (**Figure 3B**). In this cohort, *KRAS* mutations were present in 14 out of 85 samples with *TP53* mutation (16.5% frequency), which was more common than with WT *TP53* (χ² = 5.089, df = 2, p-value = 0.024). To assess the impact of co-occurring *KRAS* mutations on *ex vivo* drug response, we compared the drug sensitivity profiles of *TP53* mutated MM with and without *KRAS* mutation. We found that samples with co-occurring *TP53* and *KRAS* mutations were significantly more resistant to 5 compounds compared to samples with mutated *TP53* and WT *KRAS* (**supplemental Figure S3**, **supplemental Table S4**). *TP53* is often characterized by hotspot mutations, with the location of the mutation having different functional impact on the resulting protein. Using the MMRF-CoMMpass dataset, we found that most mutations in *TP53* were spread across the protein but occurred more frequently in the DNA binding domain (**Figure 2C**). Similarly, in the FIMM cohort, most mutations occurred in the DNA binding domain, with some samples containing more than one mutation (**Table 2**). No other hotspot mutation was detected, in concordance with previous observations in MM^17^.

**Figure 3.**
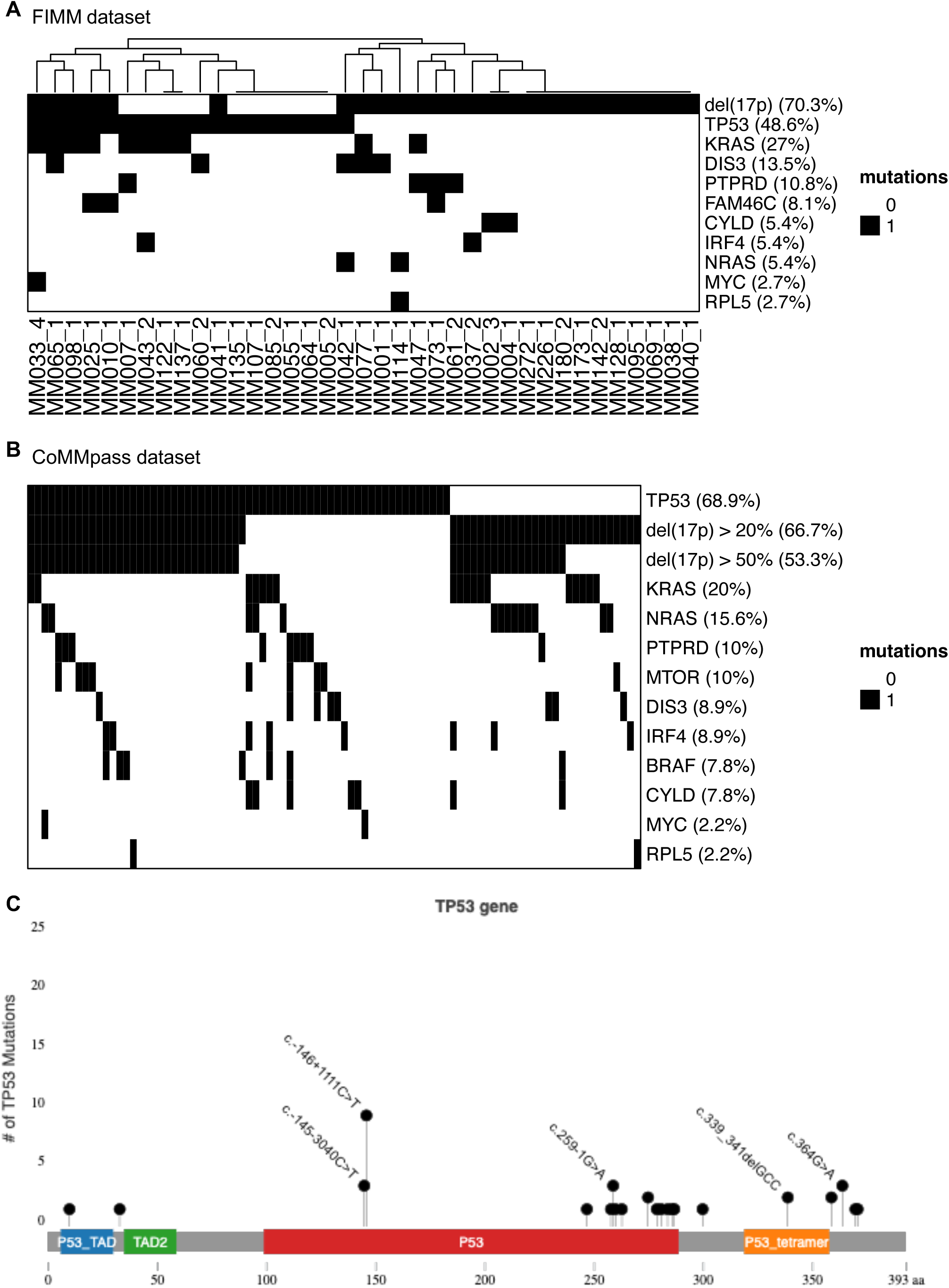
Co-occurrence of other mutations in addition to *TP53* aberrations. (A) Heatmap of common mutations in samples with *TP53* aberrations from the FIMM cohort. *KRAS* mutations in samples with *TP53* mutation are overrepresented compared to del(17p) alone. (B) Heatmap of common mutations in patients containing *TP53* aberrations from the MMRF-CoMMpass study dataset. (C) Frequency of mutations in the *TP53* coding region from 52 patients of the MMRF-CoMMpass study according to available data. The illustration of the functional domains in the p53 protein includes the transcriptional activation domains (TAD, in blue and green), the DNA binding domain (DBD, in red) and the tetramerization domain (TD, in orange).

**Table 2.**
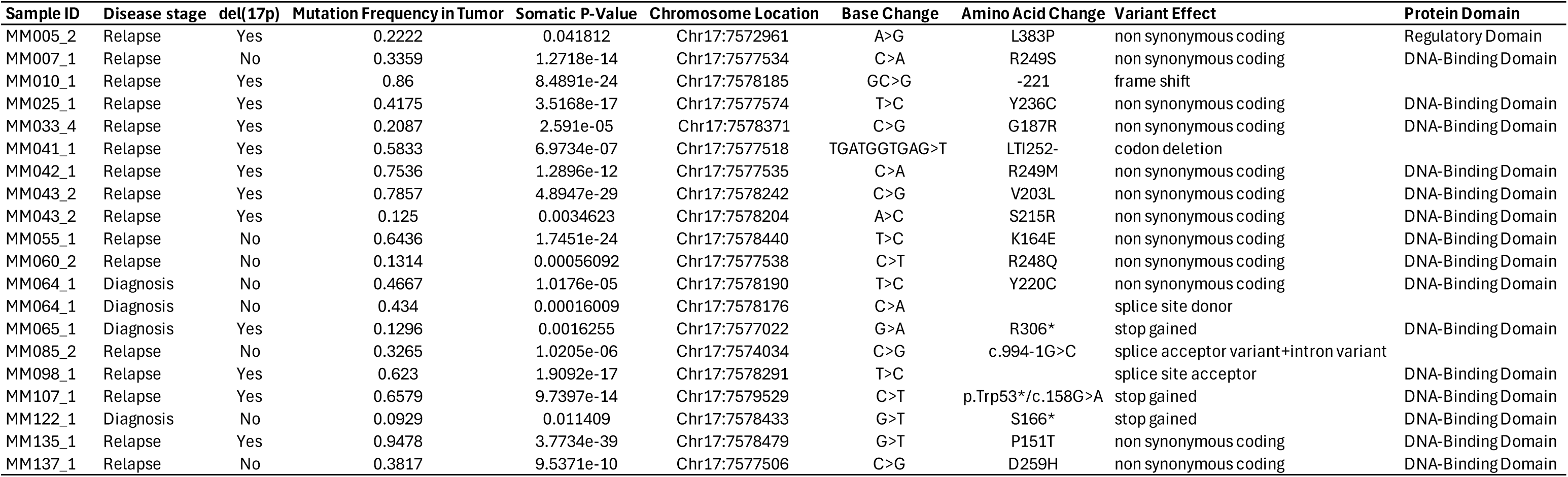
*TP53* mutations detected in patients from the FIMM dataset.

### Differential gene expression analysis reveals a transcriptional profile of enhanced cellular proliferation in TP53 mutated MM

To understand the cellular implications and functional impact of *TP53* mutations in MM, we compared RNA sequencing data of *TP53* mutated to WT *TP53* MM samples and applied differential gene expression (DGE) followed by gene set enrichment analysis (GSEA). To explore the consistency of our analysis with a wider cohort, we followed the same steps using data both from the FIMM cohort as well as the MMRF-CoMMpass study.

In the FIMM cohort, we compared 15 samples with *TP53* mutation against 77 WT *TP53* samples, from which we identified 1043 differentially expressed genes (p-value ≤ 0.05), of which 675 were upregulated and 368 downregulated (**Figure 4A**; **supplemental Tables S5-S7**). From the CoMMpass study samples with RNA sequencing data available, we compared 52 samples exhibiting *TP53* mutation (8.6%) against 552 samples with WT *TP53*. This DGE analysis provided 700 differentially expressed genes (adjusted p-value ≤ 0.01), of which 458 were upregulated and 232 downregulated (**Figure 4B, supplemental Tables S8-S10**). The results from both datasets were consistent and demonstrated that *TP53* mutated MM is associated with cell proliferation gene expression signatures. The G2M checkpoint, E2F targets and mitotic spindle formation were upregulated in both cohorts (**Figure 4C, D**). In addition to the common findings, the CoMMpass dataset showed a significant upregulation of mTORC1 signaling and genes involved in glycolysis. On the other hand, the downregulation of gene sets modulated by NF-κB in response to TNFalpha, hypoxia, response to interferon gamma, and apoptosis were only significant in the FIMM cohort analysis.

**Figure 4.**
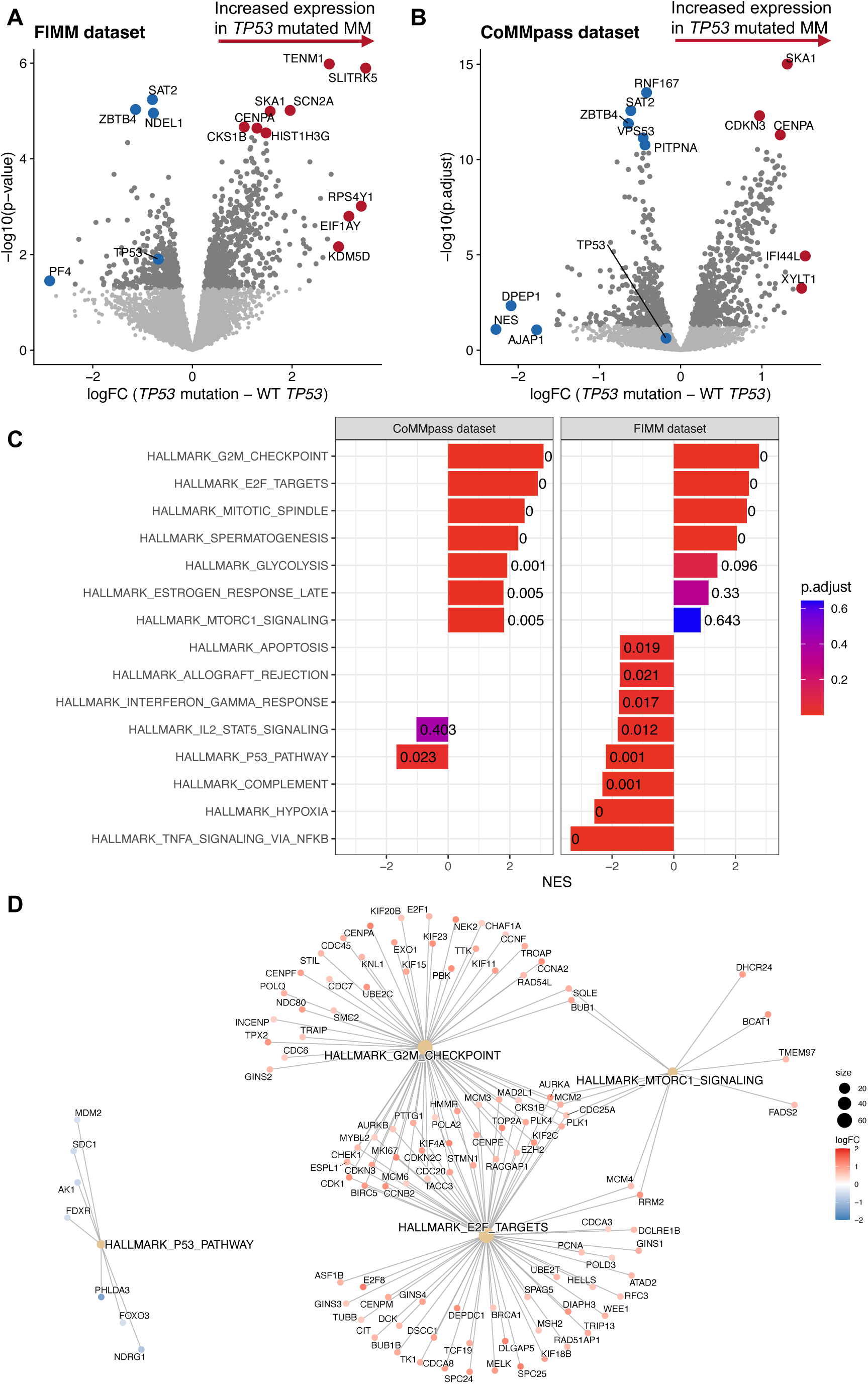
Genes sets associated with cellular proliferation are enriched in TP53 mutated MM. (A) Differential gene expression (DGE) in *TP53* mutated MM samples compared to WT *TP53* samples from the FIMM dataset. The x-axis shows the log2 fold change in expression and the y-axis shows the negative log10 of the p-value. Red dots represent genes with increased expression in *TP53* mutated MM, while decreased expression is represented with blue dots. (B) DGE analysis from the CoMMpass dataset, represented similarly, however the y-axis shows the negative log10 of the adjusted p-value. (C) Gene set enrichment analysis (GSEA) results compared between both datasets, ordered by normalized enrichment score (NES) of the FIMM dataset. The gene sets shown have an adjusted p-value ≤ 0.1 in at least one of the two datasets. (D) GSEA network plot illustrating the relationship between differentially expressed genes from selected hallmark gene sets. The genes shown are differentially expressed in CoMMpass dataset.

Additionally, we explored the transcriptional impact of monoallelic *TP53* on both FIMM and CoMMpass cohorts. DGE analysis of the FIMM cohort identified 724 differentially expressed genes (p-value ≤ 0.05), of which 472 were upregulated and 252 downregulated (**supplemental Figure S4A**). From the CoMMpass study samples, we identified 430 differentially expressed genes (adjusted p-value ≤ 0.05), of which 230 were upregulated and 200 downregulated (**supplemental Figure S4B**). GSEA revealed an increase in the G2M and E2F gene sets for samples with monoallelic *TP53* only in the larger CoMMpass cohort (**supplemental Figure S4C; supplemental Tables S11-S13**).

### Proteomic profiling reveals elevated drug target expression in TP53 mutated MM

To expand on our understanding of the functional molecular landscape associated with *TP53* mutations in MM, we explored proteomic data produced by LC-MS/MS analysis of CD138+ cells from a subset of our samples. We performed proteomic analysis on 34 samples, of which 5 contained *TP53* mutation, 6 had monoallelic *TP53* and 23 were WT *TP53*. Out of the 2753 identified proteins, 430 exhibited significant difference between samples with *TP53* mutation compared to WT *TP53* (Welch Two Sample t-test two sided; p-value ≤ 0.05; **Figure 5A**; **supplemental Table S14**).

**Figure 5.**
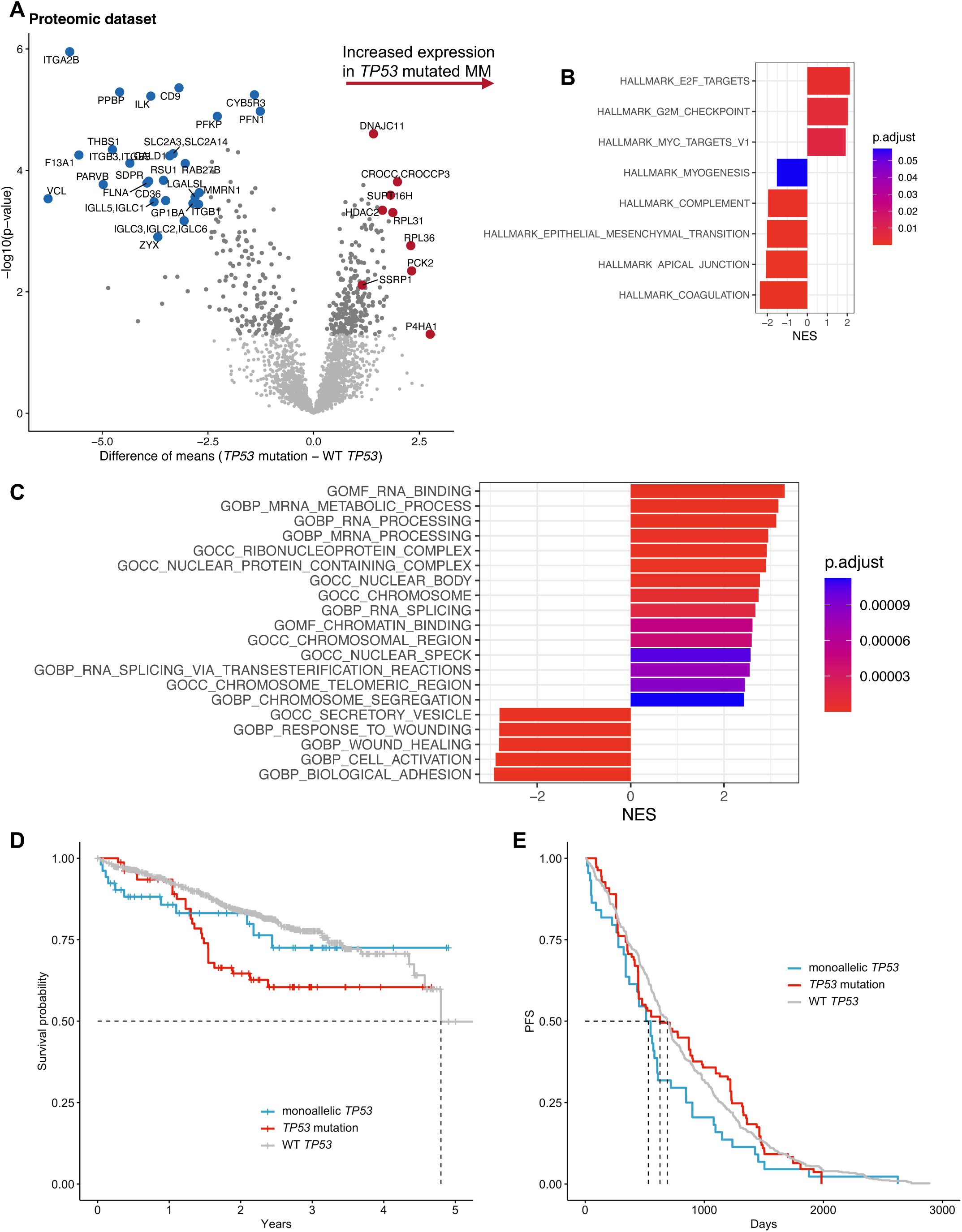
Proteins involved in RNA processing are more prevalent in *TP53* mutated MM cells. (A) Significantly increased expression of proteins from samples with *TP53* mutation compared to WT *TP53* samples is represented in a volcano plot. The x-axis shows the difference in the mean level of protein expression, with the y-axis representing negative log10 of the p-value. Red dots represent genes with increased expression in *TP53* mutated MM, while decreased expression is represented with blue dots. (B) GSEA of significant proteins on the hallmarks database reaffirm the results from the gene expression analysis, which showed gene sets associated with increased proliferation are enriched in *TP53* mutated MM. (C) The ten most positively and five most negatively enriched gene sets on GO C5 database. Gene sets are presented in descending order of normalized enrichment score (NES). (D) Analysis shows significant reduction in survival for patients with *TP53* mutation compared to WT *TP53* (OS: 2.004 [95% CI: 1.314-3.057] p = 0.001). Monoallelic loss of *TP53* (cut-off ≥ 20%) suggests a comparatively better prognosis (OS: HR = 1.264 [95% CI: 0.681-2.343] p = 0.458). (E) Monoallelic loss of *TP53*, with presence of WT *TP53* shows short progression free survival (PFS) of 530 days (PFS: HR = 1.356 [95% CI: 0.997-1.846] p = 0.053). On the other hand, presence of *TP53* mutation shows a delayed median progression 628 days (PFS: HR = 1.024 [95% CI: 0.832-1.260] p = 0.823), WT patients’ median PFS was 689 days.

In alignment with the results from GSEA of RNA sequence data, we observed an enrichment of E2F targets and G2M checkpoint proteins. Additionally, MYC targets were upregulated as shown in **Figure 5B**. Analysis using the Gene Ontology (GO) C5 library revealed significant enrichment of proteins associated with transcription processes, such as RNA binding, RNA/mRNA processing, nuclear speckles, and RNA splicing (**Figure 5C**). Notably, the ribonucleoprotein complex gene set, involved in ribosomal functions, including HURNPA3, HNRNPA2B1, SNRPA1, and RNPS1, along with the FACT complex proteins SSRP1 and SUPT16H, were highly expressed in the *TP53* mutated samples (**Figure 5A**; **supplemental Table S14**).Among the gene sets enriched with the selected proteins, we observed downregulation in the immune response and immune markers CD9, CD36 and CD76 suggesting a mechanism of immune evasion. Other significantly downregulated gene sets included those associated with mechanisms of adhesion and secretion. For the full list of gene sets enriched in this analysis see **supplemental Tables S15-S16**.

Among the significantly expressed proteins identified, some are notable drug targets. We observed increased expression of HDAC2, which is a target of HDAC inhibitors. This supports the observed sensitivity to specific HDAC inhibitors, including the approved non-selective inhibitors panobinostat and quisinostat, and the dual-acting inhibitor CUDC-101, which targets both HDAC and EGFR pathways. Similarly, we observed significant upregulation of HSP90AA1, the target of HSP90 inhibitor tanespimycin (**supplemental Figure S5; supplemental Table S2**). While these single proteins can explain the efficacy of some targeted compounds, the scope is limited by the specificity of the inhibitors and the detectable proteins in our analysis.

In contrast, comparison of monoallelic *TP53* to WT *TP53* proteomic profiles exhibited 96 significantly expressed proteins, with 45 upregulated and 51 downregulated in the monoallelic *TP53* samples. We observed an increase in gene sets related to protein transport and a decrease in mitochondrial matrix and ribonucleoprotein complex (**supplemental Figure S6; supplemental Tables S17-S18**).

### Clinical Implications of TP53 aberrations

To translate our findings to the clinic and evaluate the impact on prognosis of *TP53* mutations, we leveraged the comprehensive dataset provided by the MMRF-CoMMpass study and investigated the clinical prognosis of patients with or without *TP53* mutation.

A significant reduction in overall survival (OS) was notable among patients with *TP53* mutation as compared to those with WT *TP53* (OS: HR = 2.004 [95% CI: 1.314-3.057] p = 0.001; **Figure 5D**). Surprisingly, patients with monoallelic loss of *TP53* showed no significant reduction in OS compared to patients with WT *TP53*, but they clearly had shorter progression free survival (OS: HR = 1.264 [95% CI: 0.681-2.343] p = 0.458; PFS: HR = 1.356 [95% CI: 0.997-1.846] p = 0.053; **Figure 5E**). Patients with monoallelic *TP53* had a median PFS of 530 days (17.4 months) compared to 628 (20.6 months) for patients with *TP53* mutation. This suggests that the presence of *TP53* mutation can lead to a more aggressive disease after the first relapse, possibly because of clonal expansion. Although there is an increase in patients with *TP53* mutation at relapse (**Figure 1**), due to lack of variant allele frequency data in the MMRF-CoMMpass dataset we were not able to explore the clonal evolution of *TP53* mutations.

## DISCUSSION

In our study we provide a comprehensive view and contribute significant insight into the consequences of *TP53* aberrations in MM by applying a multi-omic approach to a large set of patient samples. Functional assessment by *ex vivo* drug sensitivity testing along with genomic, transcriptomic, and proteomic analyses highlight the biological impact of *TP53* mutations and revealed potential treatment vulnerabilities. Notably, our findings discerned distinct differences in *ex vivo* drug responses between MM with *TP53* mutation and MM with monoallelic *TP53* when compared to samples with WT diploid *TP53*. These results were further supported by differences in the genomic, transcriptomic, and proteomic landscapes of these patients. Our research provides novel insights into the vulnerabilities of MM with *TP53* mutations, with or without del(17p), as well as underscoring the need and suggesting potential personalized therapeutic strategies for this high-risk subset of patients for future investigations.

The clinical landscape of MM has long been influenced by the presence of del(17p), a well-documented adverse prognostic factor^2^. With the advent of sequencing technologies, *TP53* mutations have emerged as a novel prognosis marker in MM^6,17^, yet their distinct functional impact compared to del(17p) has not been fully recognized until now. Our study showed that unlike monoallelic *TP53*, *TP53* mutations in CD138+ cells confer increased sensitivity to multiple small molecule inhibitors. This finding presents opportunities to develop effective treatment strategies that includes conventional chemotherapies and other approved drugs. By focusing on patients with *TP53* mutation, we have the potential to mitigate the risk for almost half of the del(17p) cases, including those with double-hit MM and those with *TP53* mutation and intact chromosome 17. However, our results reaffirm the continued challenges of treating del(17p) MM.

Drug sensitivity testing of MM samples from a large cohort of patients demonstrated that MM with *TP53* mutation is vulnerable to multiple approved and investigational drugs with known target profiles. Even at low variant allele frequency (VAF) CD138+ MM cells with a minimum *TP53* mutation VAF of 5% displayed increased sensitivity to a spectrum of compounds, including conventional chemotherapeutics, topoisomerase, RNA synthesis, HDAC, HSP90, IGF1R and PI3K/AKT/mTOR pathway inhibitors. HSP90 and HDAC inhibitors have already been described to have critical roles in the degradation of mutant *TP53* in other types of cancer^18^. The increased expression of proteins HSP90AA1 and HDAC2, together with the increased sensitivity to HSP90 and HDAC inhibitors, suggest a critical role for these factors in *TP53* mutated MM cell survival. *TP53* mutations have been previously reported to upregulate *IGF1R* expression and IGF-1 mediated mitogenesis, possibly through reduced p53 levels and increased demand for new ribosomes, leading to MDM2-p53 interaction^19,20^. Subsequent mTOR pathway activation leads to increased cell proliferation, increased mRNA translation and increased glycolytic activity^19–21^. The observed increase in expression of ribosomal subunits, together with the enrichment of genes involved in glycolysis and mTORC1 signaling may explain the enhanced vulnerability to IGF1R, PI3K/AKT/mTOR inhibitors.

Both gene expression and proteomic analyses showed enrichment of pathways associated with increased cell proliferation in *TP53* mutated MM. Consequently, common chemotherapies including mitotic inhibitors (i.e. paclitaxel and vincristine), intercalating agents (i.e. doxorubicin and gemcitabine) and topoisomerase inhibitors (i.e. topotecan, etoposide and idarubicin) showed increased activity in CD138+ cells with *TP53* mutation. In contrast, CD138+ cells with monoallelic *TP53* did not display an increase in sensitivity to chemotherapies when compared to cells from patients with WT *TP53*.

The enhanced sensitivity of *TP53* mutated samples to doxorubicin, plicamycin and dactinomycin may be explained by their mechanisms of action and association with ribosomal biogenesis and mRNA processing^22,23^. Proteomic analysis revealed that proteins belonging to the ribonucleoprotein complex are upregulated in mutated *TP53* and downregulated in monoallelic *TP53* MM. Doxorubicin has been previously shown to inhibit the synthesis of 47S rRNA precursor^23^, while it also acts as intercalator similarly to daunorubicin and idarubicin. The RNA synthesis inhibitor plicamycin showed the largest difference in activity between *TP53* mutated and WT *TP53* samples. Plicamycin selectively targets genes with GC-rich promoter sequences, activates any remaining wild-type p53 and inhibits the transcriptional regulator SP1 from DNA binding, including to the promoter of *TP53*^23^. The enrichment of pathways associated with mRNA processing and chromatin modulation suggests altered transcriptional regulation and DNA replication in *TP53* mutated MM, which is similar to findings in other cancers^9,18,24,25^.

When exploring potential confounding genomic factors that may contribute to the enhanced drug sensitivity profile in *TP53* mutated MM cells, we noticed that *TP53* mutations were frequently accompanied by *KRAS* mutations, which was not observed in monoallelic *TP53* MM. However, when we compared the drug response profiles of samples with *TP53* mutation and WT *KRAS* to samples with both *TP53* and *KRAS* mutation, we only found 5 drugs were significantly affected by the presence of a *KRAS* mutation, with the *KRAS* mutation conferring a slight negative effect on the activity of these drugs. These results indicate that mutation to *TP53* is the main contributor to the enhanced drug sensitivity and altered transcriptomic and proteomic profiles of *TP53* mutated cells. Further research is needed to understand the mechanisms leading to enhanced sensitivity to drugs such as plicamycin and doxorubicin and their impact on MM cells and disease models with mutated *TP53*. Additionally, other drugs that were not included in our panel could be effective for patients with *TP53* mutation positive MM. For example, our proteomic analysis showed elevated levels of XPO1, the target of approved MM drug selinexor, and its cofactor RANBP3, which are exclusively responsible for transporting p53 out of the nucleus^26,27^. Considering the enhanced activity of several chemotherapies in *TP53* mutated CD138+ MM cells, newer agents such as the antibody-drug conjugate blenrep and peptide-drug conjugate melflufen might also provide increased efficacy for patients with *TP53* mutated MM. In addition, novel therapeutic approaches, such as CAR-T cell therapies, bispecific and monoclonal antibodies have shown promising results for MM in clinical settings^28^. Our drug screening, however, was limited to small molecules, which were tested, depending on cell availability, on isolated CD138+ MM cells without the supporting immune microenvironment. Nevertheless, the results revealed targetable vulnerabilities in *TP53* mutated MM, which could be investigated further for future therapeutic development. While our comprehensive profiling showed that mutation to *TP53* can impact *ex vivo* drug response as well as the transcriptomic and proteomic profiles to cells with the mutation, we have only recently gained understanding of the significance of this aberration on patient outcome. The adverse effect of *TP53* mutation on overall survival suggests that early detection of subclonal *TP53* mutations could potentially predict faster disease progression after relapse and allow for earlier intervention. This would be particularly important for patients without del(17p) but with mutated *TP53*, as these patients may not be correctly stratified and are less likely to receive optimized care.

In conclusion, our study offers translational insights into the implications of *TP53* mutations in MM. Our findings elucidate key molecular aspects from genomic to proteomic landscapes, guiding future research and therapeutic strategies for this subgroup of patients. As the complexities of MM continue to be unraveled with the application of advanced technologies, we will gain better understanding of the impact of genetic mutations like *TP53*, and how these can shape the future of MM risk-stratification and treatment. These findings emphasize the importance of personalized medicine approaches in oncology and provide rationale for the development of multiple therapeutic strategies and treatment of *TP53* mutated MM.

## Data Availability

The ex vivo drug sensitivity, DNA sequencing, RNA sequencing, and proteomic data generated in this study will be deposited in the Zenodo public repository upon acceptance of the manuscript for publication. Additionally, this study was supplemented with publicly available data from the Multiple Myeloma Research Foundation Researcher Gateway (https://research.themmrf.org/).

## ACKNOWLEDGMENTS

The authors thank the patients for their consent and generosity to use their samples and data. We are grateful for the support of the FIMM Technology Center Genomics and High Throughput Biomedicine Units. In addition, we thank the Multiple Myeloma Research Foundation (MMRF) for making their datasets publicly available. The Finnish Hematology Registry and Biobank and the Helsinki Hematology Research Unit are acknowledged for contributions to the sample collection and clinical data.

## AUTHOR CONTRIBUTIONS

C.A.H., D.T., and M.M.M. contributed to the study conceptualization. D.T., J.M., A.P., M.S., and M.M.M. performed experimental work. S.E. performed data normalizations for proteomic and sequencing data. K.D., P.D., D.B. and P.O’G. performed LC-MS/MS experiments and contributed to proteomics data analysis. D.T. performed data analysis and interpretation with N.I.’s contribution. J.L., R.S. and P.A. provided patient samples and contributed clinical data. D.T., J.J.M., I.V., N.I. and C.A.H. contributed to data management. D.T. and C.A.H. wrote the article. C.A.H. supervised the study and provided infrastructure support. All authors critically read and approved the final version of the article.

---

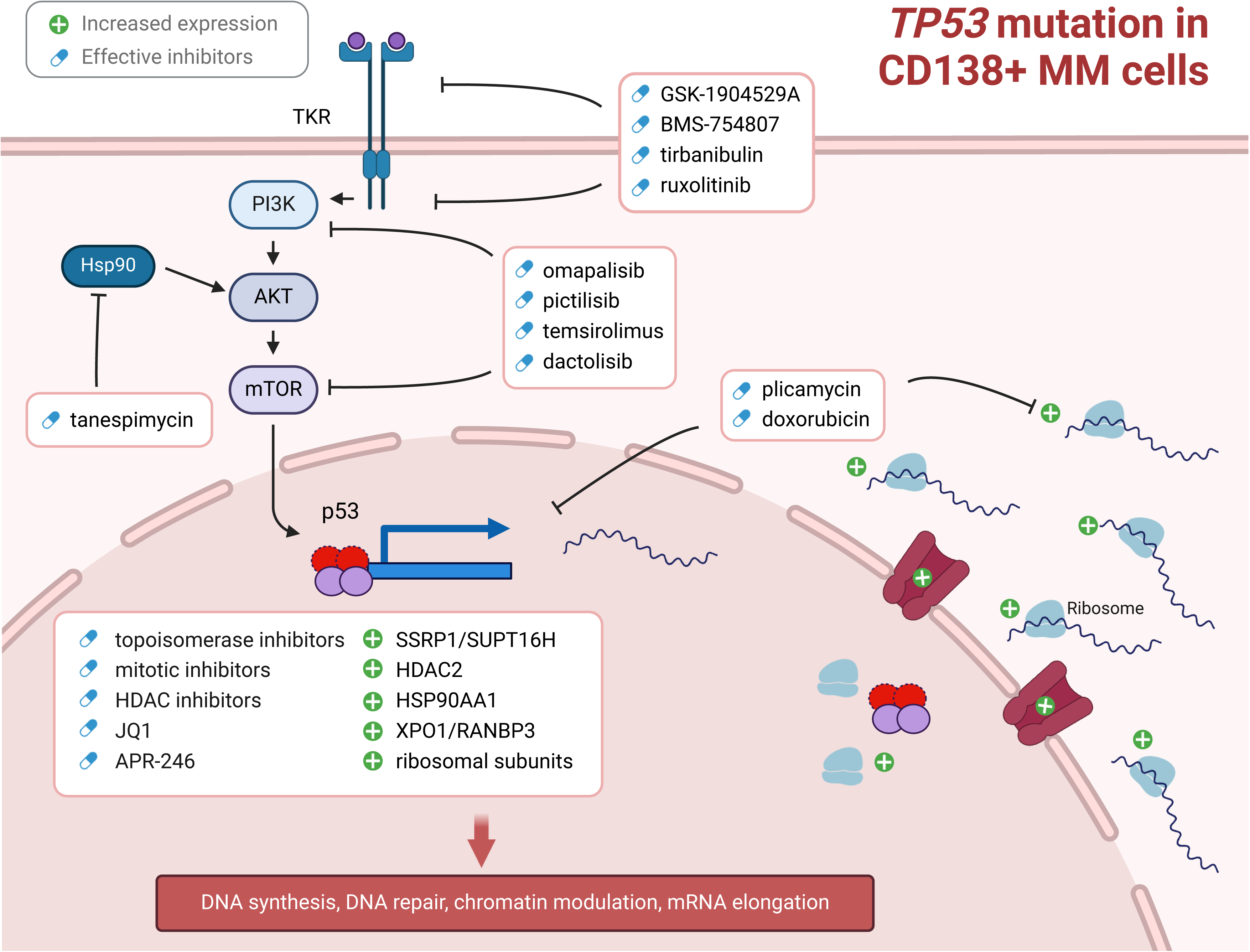

