## Supplemental Materials for "Multi-omic and functional screening reveal targetable vulnerabilities in *TP53* mutated multiple myeloma"

**Table of contents**

**Supplemental Methods**

**Supplemental Tables**

**Supplementary Table S1.** List of compounds tested on Multiple Myeloma samples.

**Supplemental Table S2.** Comparison of ex vivo drug sensitivity for small molecule inhibitors in MM patients with presence of TP53 mutation or monoallelic TP53 compared to WT TP53. (A) p-value and median difference (B) number of samples tested and standard deviation of DSS.

**Supplementary Table S3.** Correlation of *TP53* mutation frequency and drug sensitivity. Results from spearman correlation with compounds tested.

**Supplemental Table S4.** Comparison of ex vivo drug sensitivity for small molecule inhibitors in MM patients with TP53 and KRAS mutations compared to TP53 mutation and WT KRAS. (A) p-values and median difference (B) number of samples and standard deviation of DSS score.

**Supplementary Table S5.** Differentially expressed genes from the comparison between samples with *TP53* mutation compared to WT *TP53 from the FIMM cohort*.

**Supplementary Table S6.** Gene set enrichment analysis of the C5 library from differential gene expression of the comparison between presence of *TP53* mutation to WT *TP53* from the FIMM cohort**.**

**Supplementary Table S7.** Gene set enrichment analysis of the Hallmark library from differential gene expression of the comparison between presence of TP53 mutation to WT *TP53* samples. Data used for this comparison comes from the FIMM dataset.

**Supplementary Table S8.** Differentially expressed genes from the comparison between samples with *TP53* mutation compared to WT *TP53* samples. Data from this table are derived from the comparison of the MMRF-CoMMpass data.

**Supplementary Table S9.** Gene set enrichment analysis of the Hallmark library from differential gene expression of the comparison between *TP53* mutation to WT *TP53* samples. Results from the analysis of the CoMMpass data.

**Supplementary Table S10.** Gene set enrichment analysis of the C5 library from differential gene expression of the comparison between *TP53* mutation to WT *TP53* samples. Results from the CoMMpass dataset.

**Supplementary Table S11**. Differentially expressed genes from the comparison between samples with monoallelic *TP53* compared to WT *TP53*. Data from this table are derived from the FIMM dataset.

**Supplementary Table S12.** Gene set enrichment analysis of the Hallmark library from significantly expressed genes of the comparison between monoallelic *TP53* to WT *TP53* samples of the FIMM cohort.

**Supplementary Table S13.** Gene set enrichment analysis of the GO C5 library from significantly expressed genes of the comparison between monoallelic *TP53* to WT *TP53* samples of the FIMM cohort.

**Supplementary Table S14.** Significantly different proteins between *TP53* mutation compared to WT *TP53*.

**Supplementary Table S15.** Gene set enrichment analysis of the Hallmark library from significantly expressed proteins of the comparison between *TP53* mutation to WT *TP53* samples.

**Supplementary Table S16.** Gene set enrichment analysis of the C5 library from significantly expressed proteins of the comparison between *TP53* mutation to WT *TP53* samples.

**Supplementary Table S17.** Significantly different proteins between monoallelic *TP53* compared to WT *TP53*.

**Supplementary Table S18.** Gene set enrichment analysis of the C5 library from significantly expressed proteins of the comparison between monoallelic *TP53* to WT *TP53* samples of the FIMM cohort.

**Supplemental Figures**

**Supplemental Figure S1.** *Ex vivo* drug screening highlighting the difference in response to treatment between patients with *TP53* mutation and monoallelic *TP53*. Volcano plot illustrating the comparison of drug sensitivity between MM samples with *TP53* mutation and monoallelic *TP53*. Compounds with significant difference are labeled (p-value ≤ 0.05), red dots represent compounds with significant difference below 5 DSS points.

**Supplemental Figure S2.** *TP53* mutation frequency weakly correlates with drug sensitivity of MM cells. (A) Scatter plot summarizing results from the Spearman correlation of Variant Allele Frequency (VAF) for *TP53* mutation to DSS. The x-axis representes the Spearman correlation coefficient (SpearmanRho), indicating the strength and direction of the correlation between *TP53* VAF and DSS. The y-axis represents the negative logarithm (base10) of the p-value (log10(p-value)), indicating the statistical significance of the correlation. Compounds labeled in red show a significant increase in sensitivity in *TP53* mutated MM cells, as presented in **Figure 2B**. The blue lines indicate limits of significance for p-value and spearman Rho at 0.5. (B) Representative correlation between compounds and *TP53* mutation frequency.

**Supplemental Figure S3.** *Ex vivo* drug screening results highlighting the difference in response between MM samples with *KRAS* and *TP53* mutation co-occurrence compared to WT *KRAS* and *TP53* mutation. Volcano plot illustrating the comparison of drug sensitivity between MM samples with *KRAS* and *TP53* mutation co-occurrence compared to WT *KRAS* and *TP53* mutation. Compounds with significant difference are labeled (p-value ≤ 0.05), red dots represent compounds with significant difference below 5 DSS points.

**Supplemental Figure S4.** Monoallelic *TP53* leads to reduced expression of multiple genes in the 17p region. (A) DGE in *TP53* mutated samples compared to WT *TP53* samples from the FIMM dataset. The x-axis shows the log2 fold change in expression and the y-axis shows the negative log10 of the p-value. Red dots represent genes with increased expression in monoallelic *TP53* MM, while decreased expression is represented with blue dots. (B) DGE analysis of the CoMMpass dataset, similarly presented, however the y-axis shows the negative log10 of the adjusted p-value with cut-off for significance for adjusted p-value ≤ 0.05. (C) GSEA results compared between both datasets, ordered by the NES of the FIMM dataset. The gene sets shown have an adjusted p-value ≤ 0.1 in at least one of the two datasets.

**Supplemental Figure S5.** Monoallelic *TP53* MM has minimal impact on the MM proteomic profile. (A) Significantly increased expression of proteins from samples with monoallelic *TP53* compared to WT *TP53* MM samples is represented in a volcano plot. The x-axis shows difference in mean level of protein expression, with the y-axis representing negative log10 of the p-value. Red dots represent proteins with increased expression in monoallelic *TP53* MM, while decreased expression is represented with blue dots. (B) GSEA results, ordered by the NES.

### Supplemental Methods

*Label-Free LC-MS/MS Analysis and Data Processing of CD138+ cells*

500 ng of each digested sample was loaded onto a Q-Exactive (ThermoFisher Scientific, Hemel Hempstead, UK) high-resolution accurate mass spectrometer connected to a Dionex Ultimate 3000 (RSLCnano) chromatography system (ThermoFisher Scientific, Hemel Hempstead, UK). Peptides were separated using a 2% to 40% gradient of acetonitrile on a Biobasic C18 Picofrit column (ThermoFisher Scientific, Hemel Hempstead, UK) (100 mm length, 75 mm ID) over 65 min at a flow rate of 250 nl/min. Data was acquired with the mass spectrometer operating in automatic data dependent switching mode. A full MS scan at 140,000 resolution and a range of 300–1700 m/z was followed by an MS/MS scan, resolution 17,500 and a range of 200–2000 m/z, selecting the ten most intense ions prior to MS/MS. Protein identification and label-free quantification (LFQ) normalisation of MS/MS data was performed using MaxQuant v1.5.2.8 (http://www.maxquant.org). The Andromeda search algorithm incorporated in the MaxQuant software was used to correlate MS/MS data against the Homo sapiens Uniprot reference proteome database and a contaminant sequence set provided by MaxQuant. Perseus v.1.5.6.0 (www.maxquant.org/) was used for data analysis and processing. In our figures, uniprot accession IDs are presented as gene symbol IDs.

### Supplemental Tables

**Supplemental Table S1.** List of compounds tested on Multiple Myeloma samples.

| Drug | Functional class | Mechanism target | Number of samples screened |
|---|---|---|---|
| GSK-J4 | Differentiation/Epigenetic modifier | JMJD3 (histone demethylase) inhibitor | 135 |
| Carfilzomib | Proteasome inhibitor | Proteasome inhibitor (20S subunit) | 133 |
| Navitoclax | Apoptotic modulator | Bcl-2inhibitor, Bcl-XL inhibitor | 131 |
| Quisinostat | Differentiation/Epigenetic modifier | HDAC inhibitor | 131 |
| Omacetaxine | Conventional Chemotherapeutic | Protein synthesis inhib (80 S ribosome) | 131 |
| PF-04691502 | Kinase inhibitor | PI3K/mTOR inhibitor | 130 |
| APR-246 | Apoptotic modulator | p53 activator, thioredoxin reductase 1 inhibitor | 130 |
| JQ1 | Differentiation/Epigenetic modifier; BET inhibitor | BET family inhibitor | 130 |
| Omipalisib | Kinase inhibitor | PI3K/mTOR inhibitor | 129 |
| BMS-754807 | Kinase inhibitor | IGF1R inhibitor | 128 |
| Alvocidib | Kinase inhibitor; CDK inhibitor | CDK inhibitor | 128 |
| Panobinostat | Differentiation/Epigenetic modifier; HDAC inhibitor | HDAC inhibitor | 128 |
| Dactinomycin | Conventional Chemotherapeutic | RNA and DNA synthesis inhibitor | 128 |
| Auranofin | Conventional Chemotherapeutic | Antirheumatic agent | 128 |
| Refametinib | Kinase inhibitor | MEK1/2 inhibitor | 127 |
| Doxorubicin | Conventional Chemotherapeutic; Topoisomerase inhibitor; Anthracycline | Topoisomerase II inhibitor | 127 |
| Plicamycin | Conventional Chemotherapeutic | RNA synthesis inhibitor | 127 |
| Venetoclax | Apoptotic modulator | Bcl-2 inhibitor | 127 |
| Tosedostat | Protease inhibitor | M1 family of aminopeptidase inhibitor (especially LTA4H and PSA) | 126 |
| Ipatasertib | Kinase inhibitor; AKT inhibitor | AKT inhibitor | 126 |
| Clofarabine | Conventional Chemotherapeutic; Antimetabolite | Purine analog | 126 |
| Lestaurtinib | Kinase inhibitor; Tyrosine kinase inhibitor | FLT3, JAK2, TrkA, TrkB, TrkC inhibitor | 125 |
| Daporinad | Metabolic modifier | NAMPT inhibitor | 125 |
| Bortezomib | Proteasome inhibitor | Proteasome inhibitor (26S subunit) | 125 |
| Ruxolitinib | Kinase inhibitor; Tyrosine kinase inhibitor | JAK1&2 inhibitor | 123 |
| Linsitinib | Kinase inhibitor | IGF1R, IR inhibitor | 123 |
| AZD8055 | Kinase inhibitor; mTOR inhibitor | mTOR inhibitor | 123 |
| Idelalisib | Kinase inhibitor | PI3K inhibitor, p110Œ¥-selective | 123 |
| Dexamethasone | Immunomodulator; Steroid | Glucocorticoid | 123 |
| Vistusertib | Kinase inhibitor | mTOR inhibitor, ATP-competitive | 123 |
| Pictilisib | Kinase inhibitor | PI3K inhibitor, pan-class I | 123 |
| Tanespimycin | HSP inhibitor | HSP90 inhibitor | 122 |
| Methylprednisolone | Immunomodulator | Glucocorticoid, immunomodulatory agent | 121 |
| Pentostatin | Conventional Chemotherapeutic | Antimetabolite, Purine analog | 121 |
| GSK269962 | Kinase inhibitor | ROCK1 and ROCK2 inhibitor | 121 |
| Dabrafenib | Kinase inhibitor | B-Raf(V600E) inhibitor | 121 |
| Lenalidomide | Immunomodulator | Immunomodulatory | 120 |
| Dactolisib | Kinase inhibitor; PI3K inhibitor | mTOR/(PI3K) inhibitor | 120 |
| Doramapimod | Kinase inhibitor | p38MAPK inhibitor | 120 |

| | | | |
|---|---|---|---|
| Trametinib | Kinase inhibitor; MEK inhibitor | MEK1/2 inhibitor | 119 |
| Pimasertib | Kinase inhibitor | MEK1/2 inhibitor | 118 |
| Tirbanibulin | Kinase inhibitor; Tyrosine kinase inhibitor | Src inhibitor | 116 |
| Alvespimycin | HSP inhibitor | HSP90 inhibitor | 115 |
| Enzastaurin | Kinase inhibitor | PKCbeta inhibitor | 114 |
| Temsirolimus | Rapalog; mTOR inhibitor | mTORC1 inhibitor | 112 |
| Midostaurin | Kinase inhibitor; Tyrosine kinase inhibitor | Broad TK (FLT3, KIT, RET, JAK, EGFR...) inhibitor | 111 |
| Bryostatin 1 | Kinase inhibitor | PKC activator | 109 |
| SCH772984 | Kinase inhibitor | ERK1 & 2 inhibitor | 109 |
| Ibrutinib | Kinase inhibitor; Tyrosine kinase inhibitor | Btk inhibitor | 108 |
| Cladribine | Conventional Chemotherapeutic | Antimetabolite, Purine analog | 108 |
| (5Z)-7-Oxozeaenol | Kinase inhibitor | TAK1 inhibitor | 106 |
| I-BET151 | Differentiation/Epigenetic modifier | BET family inhibitor | 106 |
| Vinblastine | Conventional Chemotherapeutic | Mitotic inhibitor. Vinca alkaloid microtubule depolymerizer | 106 |
| Idarubicin | Conventional Chemotherapeutic; Anthracycline; Topoisomerase inhibitor | Topoisomerase II inhibitor | 105 |
| Pomalidomide | Immunomodulator | Immunomodulatory agent, anti-angiogenic | 105 |
| Veliparib | Differentiation/Epigenetic modifier | PARP inhibitor | 104 |
| Nutlin-3 | Apoptotic modulator | MDM2 inhibitor | 104 |
| Topotecan | Conventional Chemotherapeutic; Topoisomerase inhibitor | Topoisomerase I inhibitor | 104 |
| Danusertib | Kinase inhibitor | Aurora, Ret, TrkA, FGFR-1 inhibitor | 103 |
| Vorinostat | Differentiation/Epigenetic modifier | HDAC inhibitor | 103 |
| AZD7762 | Kinase inhibitor; CHK inhibitor | Chk1 inhibitor | 103 |
| Mepacrine | Other | Unclear. PLA2 inhibitor. NF-kB inhibitor, p53 activator | 103 |
| Buparlisib | Kinase inhibitor; PI3K inhibitor | PI3K inhibitor, pan-class I | 102 |
| Mocetinostat | Differentiation/Epigenetic modifier; HDAC inhibitor | HDAC1 inhibitor, HDAC2 inhibitor | 102 |
| AT-406 | Apoptotic modulator | XIAP, cIAP1, cIAP2 inhibitor | 102 |
| Stattic | Other | STAT3 SH2 domain inhibitor | 102 |
| Tubacin | Differentiation/Epigenetic modifier | HDAC6 inhibitor | 102 |
| C646 | Differentiation/Epigenetic modifier | p300/CREB-binding protein (CBP) inhibitor | 102 |
| Vemurafenib | Kinase inhibitor | B-Raf(V600E) inhibitor | 102 |
| Vismodegib | Hedgehog inhibitor | Smoothened (Hh) inhibitor | 101 |
| VER 155008 | HSP inhibitor | HSP70 inhibitor | 101 |
| Oprozomib | Proteasome inhibitor | Proteasome inhibitor (20S subunit) | 101 |
| Tepotinib | Kinase inhibitor | c-Met inhibitor | 100 |
| Obatoclax | Apoptotic modulator | Bcl-2 family inhibitor | 100 |
| Entinostat | Differentiation/Epigenetic modifier; HDAC inhibitor | HDAC inhibitor | 100 |
| Regorafenib | Kinase inhibitor; Tyrosine kinase inhibitor | B-Raf, c-Kit, VEGFR2 inhibitor | 100 |
| Apitolisib | Kinase inhibitor | PI3K/mTOR inhibitor | 100 |
| Tamatinib | Kinase inhibitor | Syk inhibitor | 100 |
| Pevonedistat | Metabolic modifier | NAE inhibitor | 100 |
| Iniparib | Differentiation/Epigenetic modifier; PARP inhibitor | PARP inhibitor | 99 |
| Dinaciclib | Kinase inhibitor; CDK inhibitor | CDK inhibitor | 99 |

| | | | |
|---|---|---|---|
| Rocilinostat | Differentiation/Epigenetic modifier; HDAC inhibitor | HDAC-6 selective inhibitor | 99 |
| Everolimus | Rapalog | binds FKBP12, causes inhibition of mTORC1 | 98 |
| Quizartinib | Kinase inhibitor; Tyrosine kinase inhibitor | FLT3 inhibitor | 98 |
| Azacitidine | Antimetabolite | Pyrimidine analog, DNA methyl transferase inhibitor | 98 |
| Momelotinib | Kinase inhibitor; Tyrosine kinase inhibitor | JAK1 & 2 inhibitor | 98 |
| Sapanisertib | Kinase inhibitor | mTOR inhibitor | 98 |
| UNC0638 | Differentiation/Epigenetic modifier | G9a/GLP inhibitor | 98 |
| WEHI-539 | Apoptotic modulator | Bcl-XL inhibitor | 98 |
| TPCA-1 | Kinase inhibitor | IKK-2 inhibitor | 98 |
| Alisertib | Kinase inhibitor | Aurora A inhibitor | 97 |
| Selumetinib | Kinase inhibitor; MEK inhibitor | MEK1/2 inhibitor | 97 |
| Dasatinib | Kinase inhibitor; Tyrosine kinase inhibitor | Abl inhibitor, Src inhibitor, Kit inhibitor, EphR inhibitor | 97 |
| NVP-AEW541 | Kinase inhibitor | IGF1R inhibitor | 97 |
| ONX-0914 | Proteasome inhibitor | LMP7 (immunoproteasome) | 97 |
| Ixazomib | Proteasome inhibitor | Proteasome inhibitor (20S subunit) | 97 |
| Dovitinib | Kinase inhibitor; Tyrosine kinase inhibitor | FGFR inhibitor | 96 |
| PF 431396 | Kinase inhibitor | FAK/PYK2 inhibitor | 96 |
| SGC0946 | Differentiation/Epigenetic modifier | DOT1L inhibitor | 96 |
| Prednisolone | Immunomodulator; Steroid | Glucocorticoid | 96 |
| SGC-CBP30 | Differentiation/Epigenetic modifier | CREBBP/EP300 bromodomain inhibitor | 96 |
| PFI-1 | Differentiation/Epigenetic modifier | BET family inhibitor | 95 |
| PHA 408 | Kinase inhibitor | IKK-2 inhibitor | 95 |
| PS-1145 | Kinase inhibitor | IKK-2 inhibitor | 95 |
| Altretamine | Conventional Chemotherapeutic | Formaldehyde release, alkylating agent | 94 |
| Ralimetinib | Kinase inhibitor | p38MAPK inhibitor | 94 |
| Deferoxamine | Other | Iron chelator | 94 |
| Pinometostat | Differentiation/Epigenetic modifier | DOT1L inhibitor | 94 |
| Luminespib | HSP inhibitor | HSP90 inhibitor | 93 |
| Vincristine | Conventional Chemotherapeutic | Mitotic inhibitor, Vinca alkaloid microtubule depolymerizer | 93 |
| RAF-265 | Kinase inhibitor; RAF inhibitor | C-Raf inhibitor, unclear MoA | 93 |
| Filanesib | Kinesin inhibitor | KSP/Eg5 inhibitor | 93 |
| SGI-1776 | Kinase inhibitor | PIM kinase inhibitor | 93 |
| GSK-1838705A | Kinase inhibitor | IGF1R, INSR, ALK inhib | 93 |
| AGI-5198 | Metabolic modifier | IDH1 R132H/R132C inhibitor | 92 |
| Camptothecin | Conventional Chemotherapeutic | Topoisomerase I inhibitor | 89 |
| Sunitinib | Kinase inhibitor; Tyrosine kinase inhibitor | Broad TK inhibitor | 87 |
| Galunisertib | Kinase inhibitor | TGF-B/Smad inhibitor | 87 |
| Crenolanib | Kinase inhibitor; Tyrosine kinase inhibitor | PDGFRA inhibitor, PDGFRB inhibitor | 87 |
| VGX-1027 | Immunomodulator | Nitric oxide-donating immunomodulator | 87 |
| Foretinib | Kinase inhibitor | MET, VEGFR2 inhibitor | 85 |
| Mitoxantrone | Conventional Chemotherapeutic; Topoisomerase inhibitor | Topoisomerase II inhibitor | 85 |
| Sorafenib | Kinase inhibitor; Tyrosine kinase inhibitor | B-Raf, FGFR-1, VEGFR-2 & -3, PDGFR-beta, KIT, and FLT3 inhib | 83 |

| | | | |
|---|---|---|---|
| Axitinib | Kinase inhibitor; Tyrosine kinase inhibitor | VEGFR, PDGFR, KIT inhibitor | 79 |
| AZD1208 | Kinase inhibitor | PIM1, 2, 3 kinase inhibitor | 76 |
| Idasanutlin | Apoptotic modulator | p53-MDM2 inhibitor | 74 |
| Fedratinib | Kinase inhibitor; Tyrosine kinase inhibitor | JAK2 inhibitor | 72 |
| TH588 | Metabolic modifier | MTH1 inhibitor | 72 |
| CUDC-907 | Differentiation/Epigenetic modifier | HDAC1/2/3/10, PI3Kalpha inhibitor | 72 |
| NMS-873 | Other | p97/VCP inhibitor | 71 |
| URB597 | Metabolic modifier | FAAH inhibitor | 71 |
| KU-60019 | Kinase inhibitor | ATM inhibitor | 70 |
| AZ191 | Kinase inhibitor | DYRK1A inhibitor | 69 |
| GSK-1904529A | Kinase inhibitor | IGF1R, INSR inhib | 69 |
| TAK-580 | Kinase inhibitor | pan-RAF inhibitor | 69 |
| NVP-LGK974 | Other | PORCN inhibitor | 68 |
| Hydroxyfasudil | Kinase inhibitor | ROCK, PKA, PKG, PRK inhibitor | 68 |
| BX-912 | Kinase inhibitor | PDK1 inhibitor | 68 |
| GDC-0919 | Immunomodulator | IDO inhibitor | 57 |
| CEP-32496 | Kinase inhibitor | BRAF inhibitor | 57 |
| Disulfiram | Metabolic modifier | alcohol dehydrogenase inhibitor | 54 |
| PF-670462 | Kinase inhibitor | CK1epsilon and CK1delta inhibitor | 48 |
| MK-0752 | Other | gamma-secretase/notch inhibitor | 46 |
| PF-04708671 | Kinase inhibitor | p70S6K inhibitor | 45 |
| Melphalan | Conventional Chemotherapeutic; Alkylating agent | Nitrogen mustard alkylating agent | 45 |
| Geldanamycin | HSP inhibitor | HSP90 inhibitor | 44 |
| Seliciclib | Kinase inhibitor; CDK inhibitor | CDK2/7/9 inhibitor | 41 |
| Tarenflurbil | Other | Gamma-secretase inhibitor | 40 |
| Sepantronium bromide | Apoptotic modulator | Survivin inhibitor | 33 |
| PF-4800567 | Kinase inhibitor | CK1epsilon inhibitor | 33 |
| Chloroquine | Conventional Chemotherapeutic | Antimalaria agent, chemo/radio sensitizer | 33 |
| 2-methoxyestradiol | Other | Angiogenesis inhibitor | 32 |
| SNS-032 | Kinase inhibitor | CDK inhibitor | 32 |
| Tivantinib | Kinase inhibitor | MET inhibitor | 32 |
| PF-3845 | Other | FAAH inhibitor | 32 |
| Exemestane | Hormone therapy | Aromatase inhibitor | 32 |
| Imiquimod | Immunomodulator | Immunomodulatory agent, TLR7 agonist | 32 |
| PAC-1 | Apoptotic modulator | Procaspase-3 activator | 32 |
| Atorvastatin | Metabolic modifier | HMG-CoA reductase inhibitor | 32 |
| StemRegenin 1 | Differentiation/Epigenetic modifier | AHR antagonist, stem cell regenerating | 32 |
| Belinostat | Differentiation/Epigenetic modifier; HDAC inhibitor | HDAC inhibitor | 31 |
| Nilotinib | Kinase inhibitor; Tyrosine kinase inhibitor | Abl inhibitor | 31 |
| Masitinib | Kinase inhibitor | KIT inhibitor | 31 |
| Gandotinib | Kinase inhibitor | JAK2 inhibitor | 31 |
| Ponatinib | Kinase inhibitor; Tyrosine kinase inhibitor | Broad TK inhibitor | 31 |

| | | | |
|---|---|---|---|
| AZD1480 | Kinase inhibitor | JAK1/2, FGFR inhibitor | 31 |
| Lapatinib | Kinase inhibitor | HER2, EGFR inhibitor | 31 |
| (-)JQ1 | Differentiation/Epigenetic modifier | Inactive stereoisomer of (+)JQ1 | 31 |
| Linifanib | Kinase inhibitor; Tyrosine kinase inhibitor | VEGFR, PDGFR, CSF-1R, FLT3 inhibitor | 31 |
| Levamisole | Immunomodulator | Immunomodulatory agent | 31 |
| Pilocarpine | Other | Non-selective muscarinic receptor agonist | 31 |
| TAK-901 | Kinase inhibitor | Aurora, Src family, JAK3, RTK inhibitor | 31 |
| Bleomycin | Conventional Chemotherapeutic | Glycopeptide antibiotic, causes DNA breaks | 31 |
| Erlotinib | Kinase inhibitor | EGFR inhibitor | 31 |
| Fludarabine | Conventional Chemotherapeutic; Antimetabolite | Purine analog | 31 |
| Mercaptopurine | Conventional Chemotherapeutic; Antimetabolite | Purine analog | 31 |
| CPI-613 | Metabolic modifier | pyruvate dehydrogenase, alpha-ketoglutarate dehydrogenase inhibitor | 31 |
| AVN944 | Metabolic modifier | IMPDH inhibitor | 31 |
| Tubastatin A | Differentiation/Epigenetic modifier | HDAC6 inhibitor | 31 |
| 15D-PGJ2 | Other | Endogenous PPARŒ≥ ligand, prostaglandin, NFkB signaling inhibitor | 31 |
| Bosutinib | Kinase inhibitor; Tyrosine kinase inhibitor | Abl, Src inhibitor | 31 |
| Uramustine | Conventional Chemotherapeutic | Alkylating agent | 31 |
| Valrubicin | Conventional Chemotherapeutic | Topoisomerase II inhibitor | 31 |
| Valproic acid | Differentiation/Epigenetic modifier | HDAC inhibitor | 31 |
| OSI-027 | Kinase inhibitor | mTOR inhibitor | 30 |
| ABT-751 | Conventional Chemotherapeutic | Mitotic inhibitor. Colchicine site binding microtubule depolymerizer. | 30 |
| AT9283 | Kinase inhibitor; Tyrosine kinase inhibitor; Aurora inhibitor | Aurora A & B, Jak2, Flt, Abl inhibitor | 30 |
| CUDC-101 | Kinase inhibitor | HDAC & EGFR, Her2 inhibitor | 30 |
| Palbociclib | Kinase inhibitor; CDK inhibitor | CDK4/6 inhibitor | 30 |
| Irinotecan | Conventional Chemotherapeutic | Topoisomerase I inhibitor. Camptothecin prodrug analog | 30 |
| Saracatinib | Kinase inhibitor | Src, Abl inhibitor | 30 |
| Tofacitinib | Kinase inhibitor | JAK3, JAK2(V617F) inhibitor | 30 |
| UCN-01 | Kinase inhibitor | PKCbeta, PDK1, Chk, Cdk2 inhibitor | 30 |
| Ruboxistaurin | Kinase inhibitor | PKCbeta inhibitor | 30 |
| Aminoglutethimide | Hormone therapy | Anti-steroid, aromatase inhibitor | 30 |
| BI 2536 | Kinase inhibitor | PLK1 inhibitor | 30 |
| Sonolisib | Kinase inhibitor | PI3K inhibitor, pan-class I. Irreversible | 30 |
| Bicalutamide | Hormone therapy | Nonsteroidal antiandrogen | 30 |
| Volasertib | Kinase inhibitor | PLK1 inhibitor | 30 |
| Nintedanib | Kinase inhibitor | VEGFR, PDGFR, FGFR inhibitor | 30 |
| AT 101 | Apoptotic modulator | Bcl-2 family inhibitor | 30 |
| Tacrolimus | Rapalog | Binds FKBP12, causes inhibition of calcineurin | 30 |
| Flutamide | Hormone therapy | Nonsteroidal antiandrogen | 30 |
| Methotrexate | Antimetabolite | Anti-folate agent | 30 |
| TAK-733 | Kinase inhibitor | MEK1/2 inhibitor | 30 |
| Celecoxib | NSAID | Selective COX-2 inhibitor | 30 |
| Chlorambucil | Conventional Chemotherapeutic | Nitrogen mustard alkylating agent | 30 |

| | | | |
|---|---|---|---|
| Dacarbazine | Conventional Chemotherapeutic | Alkylating agent | 30 |
| Daunorubicin | Conventional Chemotherapeutic; Topoisomerase inhibitor; Anthracycline | Topoisomerase II inhibitor | 30 |
| Fulvestrant | Hormone therapy | Estrogen receptor antagonist | 30 |
| Pipobroman | Conventional Chemotherapeutic | Alkylating agent | 30 |
| Streptozocin | Conventional Chemotherapeutic | Alkylating glucosamine-nitrosourea agent | 30 |
| GSK2636771 | Kinase inhibitor | PI3K beta selective inhibitor | 30 |
| Dacomitinib | Kinase inhibitor | pan-HER inhibitor | 30 |
| Simvastatin | Metabolic modifier | HMG-CoA reductase inhibitor | 30 |
| Sirolimus | Rapalog | binds FKBP12, causes inhibition of mTORC1 | 30 |
| VX-11E | Kinase inhibitor | ERK1 & 2 inhibitor | 30 |
| RD162 | Hormone therapy | AR antagonist | 30 |
| Vinorelbine | Conventional Chemotherapeutic | Mitotic inhibitor. Vinca alkaloid microtubule depolymerizer | 30 |
| IOX-2 | Differentiation/Epigenetic modifier | PHD2 inhibitor | 30 |
| 8-chloro-adenosine | Conventional Chemotherapeutic | Nucleoside analog, RNA synthesis inhibitor | 30 |
| 8-amino-adenosine | Conventional Chemotherapeutic | Nucleoside analog, RNA synthesis inhibitor | 30 |
| SB 743921 | Kinesin inhibitor | Mitotic inhibitor. Eg5/KSP inhibitor | 30 |
| Perifosine | Kinase inhibitor | AKT/PI3K inhibitor | 30 |
| Pravastatin | Metabolic modifier | HMG-CoA reductase inhibitor | 30 |
| Estramustine | Conventional Chemotherapeutic | Alkylating agent | 30 |
| UNC0642 | Differentiation/Epigenetic modifier | G9a/GLP inhibitor | 30 |
| Motesanib | Kinase inhibitor | VEGFR, PDGFR, Ret, Kit inhibitor | 29 |
| Serdemetan | Apoptotic modulator | HDM2-p53 antagonist | 29 |
| Tipifarnib | Differentiation/Epigenetic modifier | Farnesyltransferase inhibitor | 29 |
| Abiraterone | Hormone therapy | P450 17alpha-hydroxylase-17,20-lyase inhibitor | 29 |
| Canertinib | Kinase inhibitor | pan-HER inhibitor | 29 |
| Olaparib | Differentiation/Epigenetic modifier; PARP inhibitor | PARP inhibitor | 29 |
| Vatalanib | Kinase inhibitor | VEGFR-1 & -2 inhibitor | 29 |
| Indibulin | Conventional Chemotherapeutic | Mitoric inhibitor. Microtubule depolymerizer | 29 |
| Sotrastaurin | Kinase inhibitor | PKC inhibitor | 29 |
| AZ 3146 | Kinase inhibitor | Mps1 kinase (TTK) inhibitor | 29 |
| Bexarotene | Differentiation/Epigenetic modifier | Antineoplastic agent, retinoid specifically selective for retinoid X receptors | 29 |
| Tandutinib | Kinase inhibitor | FLT3, PDGFR, KIT inhibitor | 29 |
| Ridaforolimus | Rapalog | binds FKBP12, causes inhibition of mTORC1 | 29 |
| Bendamustine | Conventional Chemotherapeutic | Nitrogen mustard alkylating agent | 29 |
| Galiellalactone | Other | STAT3-DNA interaction inhibitor | 29 |
| Lasofoxifene | Hormone therapy | Selective estrogen receptor modulator | 29 |
| Nilutamide | Hormone therapy | Nonsteroidal antiandrogen | 29 |
| Toremifene | Hormone therapy | selective estrogen receptor modulator | 29 |
| Clomifene | Hormone therapy | Selective estrogen receptor modulator | 29 |
| Brivanib | Kinase inhibitor | VEGFR inhibitor | 29 |
| Thio-TEPA | Conventional Chemotherapeutic; Alkylating agent | Alkylating agent | 29 |
| Hydroxyurea | Conventional Chemotherapeutic | Antineoplastic agent | 29 |

| | | | |
|---|---|---|---|
| Mechlorethamine | Conventional Chemotherapeutic | Nitrogen mustard alkylating agent | 29 |
| Allopurinol | Conventional Chemotherapeutic | Xanthine oxidase inhibitor | 29 |
| Ixabepilone | Conventional Chemotherapeutic | Mitotic inhibitor. Epothilone microtubule stabilizer. | 29 |
| Raloxifene | Hormone therapy | Selective estrogen receptor modulator | 29 |
| Tamoxifen | Hormone therapy | Estrogen receptor antagonist | 29 |
| Rebastinib | Kinase inhibitor | Allosteric ABL, FLT3, TIE2, TRKA inhibitor | 29 |
| Lomeguatrib | Differentiation/Epigenetic modifier | O6-methylguanine-DNA methyltransferase inhibitor | 29 |
| GSK650394 | Kinase inhibitor | SGK1 & 2 inhibitor | 29 |
| BMS-911543 | Kinase inhibitor | JAK2 inhibitor | 29 |
| Infigratinib | Kinase inhibitor | FGFR inhibitor | 29 |
| PIK-75 | Kinase inhibitor | p110alpha selective PI3K inhibitor | 29 |
| TGX-221 | Kinase inhibitor | PI3K beta selective inhibitor | 29 |
| Enzalutamide | Hormone therapy | AR antagonist | 29 |
| Tretinoin | Differentiation/Epigenetic modifier | Retinoic acid receptor agonist | 29 |
| Temozolomide | Conventional Chemotherapeutic | Alkylating agent | 29 |
| Thioguanine | Conventional Chemotherapeutic; Antimetabolite | Purine analog | 29 |
| Carboplatin | Conventional Chemotherapeutic | Platinum-based antineoplastic agent | 29 |
| Oxaliplatin | Conventional Chemotherapeutic | Platinum-based antineoplastic agent | 29 |
| Amonafide | Conventional Chemotherapeutic | Topoisomerase II inhibitor / DNA intercalator | 29 |
| Cytarabine | Conventional Chemotherapeutic; Antimetabolite | DNA synthesis | 29 |
| Bimatoprost | Immunomodulator | Prostaglandin analog | 28 |
| BIIB021 | HSP inhibitor | HSP90 inhibitor | 28 |
| Fasudil | Kinase inhibitor | ROCK, PKA, PKG, PRK inhibitor, prodrug | 28 |
| Pazopanib | Kinase inhibitor | VEGFR inhibitor | 28 |
| XAV-939 | Differentiation/Epigenetic modifier | Tankyrase-1 and -2 | 28 |
| S-trityl-L-cysteine | Kinesin inhibitor | Eg5 inhibitor | 28 |
| Cediranib | Kinase inhibitor | KDR/Flt/VEGFR inhibitor | 28 |
| Barasertib | Kinase inhibitor; Aurora inhibitor | Aurora B inhibitor | 28 |
| MK-2206 | Kinase inhibitor; AKT inhibitor | AKT inhibitor | 28 |
| Tacedinaline | Differentiation/Epigenetic modifier | HDAC inhibitor | 28 |
| Niraparib | Differentiation/Epigenetic modifier | PARP inhibitor | 28 |
| Binimetinib | Kinase inhibitor | MEK1/2 inhibitor | 28 |
| Capecitabine | Conventional Chemotherapeutic | 5-FU prodrug | 28 |
| Gefitinib | Kinase inhibitor | EGFR inhibitor | 28 |
| Imatinib | Tyrosine kinase inhibitor; Kinase inhibitor | Bcr-Abl inhibitor, Kit inhibitor, PDGFRB inhibitor | 28 |
| Cyclopamine | Other | Smothened inhibitor | 28 |
| Prednisone | Immunomodulator | Immunomodulatory agent | 28 |
| Rabusertib | Kinase inhibitor | Chk1 inhibitor | 28 |
| Anastrozole | Hormone therapy | Aromatase inhibitor | 28 |
| Etoposide | Conventional Chemotherapeutic; Topoisomerase inhibitor | Topoisomerase II inhibitor | 28 |
| Floxuridine | Conventional Chemotherapeutic | Antimetabolite, Analog of 5-fluorouracil | 28 |
| Fluorouracil | Conventional Chemotherapeutic | Antimetabolite | 28 |

| | | | |
|---|---|---|---|
| Letrozole | Hormone therapy | Aromatase inhibitor | 28 |
| Lomustine | Conventional Chemotherapeutic | Alkylating nitrosourea compound | 28 |
| Roxadustat | Differentiation/Epigenetic modifier | HIF prolyl hydroxylase inhibitor | 28 |
| Neratinib | Kinase inhibitor | HER2, EGFR inhibitor | 28 |
| 4-hydroxytamoxifen | Hormone therapy | Selective estrogen receptor modulator | 28 |
| Thalidomide | Immunomodulator | Immunosuppressant | 28 |
| Ifosfamide | Conventional Chemotherapeutic | Nitrogen mustard alkylating agent | 28 |
| Gemcitabine | Conventional Chemotherapeutic; Antimetabolite | Pyrimidin analog | 28 |
| Teniposide | Conventional Chemotherapeutic | Topoisomerase II inhibitor | 28 |
| ZSTK474 | Kinase inhibitor | PI3K gamma selective inhibitor | 28 |
| Arsenic(III) oxide | Differentiation/Epigenetic modifier | Thioredoxin reductase inhibitor, cytotoxic chemotherapeutic | 28 |
| Cisplatin | Conventional Chemotherapeutic | Platinum-based antineoplastic agent | 28 |
| Crizotinib | Kinase inhibitor | ALK, c-Met inhibitor | 28 |
| GSK343 | Differentiation/Epigenetic modifier | EZH2 inhibitor | 28 |
| Mitomycin | Conventional Chemotherapeutic; Alkylating agent | Antineoplastic anatibiotic, DNA crosslinker | 28 |
| AZD1775 | Kinase inhibitor | Wee1 inhibitor | 28 |
| PF-00477736 | Kinase inhibitor | Chk1 inhibitor | 27 |
| Rucaparib | Differentiation/Epigenetic modifier | PARP inhibitor | 27 |
| Decitabine | Antimetabolite | Pyrimidine analog, DNA methyl transferase inhibitor | 27 |
| Afatinib | Kinase inhibitor | EGFR inhibitor | 27 |
| Docetaxel | Conventional Chemotherapeutic | Mitotic inhibitor, taxane microtubule stabilizer | 27 |
| Patupilone | Conventional Chemotherapeutic | Mitotic inhibitor, epothilone microtubule stabilizer | 27 |
| Busulfan | Conventional Chemotherapeutic; Alkylating agent | Alkylating agent | 27 |
| Megestrol acetate | Hormone therapy | Progestogen | 27 |
| Mitotane | Conventional Chemotherapeutic | Antineoplastic agent | 27 |
| Procarbazine | Conventional Chemotherapeutic | Alkylating agent | 27 |
| Rofecoxib | NSAID | COX-2 inhibitor | 27 |
| Fostamatinib | Kinase inhibitor | Syk inhibitor | 27 |
| Sonidegib | Hedgehog inhibitor | Smoothened (Hh) inhibitor | 27 |
| Carmustine | Conventional Chemotherapeutic; Alkylating agent | Alkylating agent | 27 |
| Paclitaxel | Conventional Chemotherapeutic | Mitotic inhibitor, taxane microtubule stabilizer | 27 |
| Tivozanib | Kinase inhibitor | VEGFR1, 2, 3, c-Kit, PDGFRB inhibitor | 26 |
| Fingolimod | Other | S1PR antagonist | 26 |
| Finasteride | Hormone therapy | type II 5-alpha reductase inhibitor | 26 |
| Goserelin | Hormone therapy | Gonadotropin releasing hormone superagonist | 26 |
| UNC1215 | Differentiation/Epigenetic modifier | L3MBTL3 inhibitor | 26 |
| Pemetrexed | Metabolic modifier | Dihydrofolate reductase inhibitor | 26 |
| 1-methyl-D-tryptophan | Other | Indolamine 2,3-dioxygenase 1 and 2 inhibitor | 26 |
| Nelarabine | Conventional Chemotherapeutic; Antimetabolite | Purine analog, DNA, RNA synth inhibitor | 26 |
| Cabozantinib | Kinase inhibitor; Tyrosine kinase inhibitor | VEGFR2, Met, FLT3, Tie2, Kit and Ret inhibitor | 25 |
| Anagrelide | Other | PDE-3, PLA2 inhibitor | 25 |
| Cyclophosphamide | Conventional Chemotherapeutic | Alkylating agent | 25 |

| | | | |
|---|---|---|---|
| Varespladib | Other | Secretory phospholipase A2 inhibitor | 25 |
| Vandetanib | Kinase inhibitor | VEGFR,EGFR, RET inhibitor | 24 |
| AZD4547 | Kinase inhibitor | FGFR inhibitor | 24 |
| Plerixafor | Other | CXCR4 antagonist | 23 |
| Cilengitide | Other | alphaVbeta3 integrin inhibitor | 16 |
| deltarasin | Other | Ras-PDEdelta inhibitor | 16 |
| IRAK1/4 inhibitor | Kinase inhibitor | IRAK1/4 inhibitor | 16 |
| Pilaralisib | Kinase inhibitor | PI3K inhibitor. Pan-class I | 15 |
| Tasquinimod | Immunomodulator | S100A9 inhibitor, immunomodulatory, anti-angiogenic | 15 |
| Triapine | Metabolic modifier | ribonucleotide reductase inhibitor | 15 |
| Birinapant | Apoptotic modulator | IAP, SMAC mimetic | 14 |
| Metformin | Metabolic modifier | AMPK activator | 13 |
| Voxtalisib | Kinase inhibitor | mTOR/PI3K inhibitor | 7 |

**Supplemental Table S2.** Comparison of *ex vivo* drug sensitivity for small molecule inhibitors in MM patients with presence of *TP53* mutation or monoallelic *TP53* compared to WT *TP53*. (A) p-value and median difference (B) number of samples tested and standard deviation of DSS.

**A.**

| **Drug Name** | **P-value (*TP53* mutation - WT)** | **Median Difference (*TP53* mutation - WT)** | **P-value (monoallelic *TP53* - WT)** | **Median difference (monoallelic *TP53* - WT)** | **Functional Class** | **Mechanism Target** |
|---|---|---|---|---|---|---|
| Doxorubicin | 0.00004 | 5.9 | 0.04360 | 2.2 | Kinase inhibitor | VEGFR-1 & -2 inhibitor |
| Patupilone | 0.00033 | 5.3 | 0.03023 | 2.3 | Rapalog; mTOR inhibitor | mTORC1 inhibitor |
| Quisinostat | 0.00033 | 7.3 | 0.54921 | 0.3 | Differentiation/Epigenetic modifier | PARP inhibitor |
| Dactolisib | 0.00046 | 6.8 | 0.41313 | -1.3 | Kinase inhibitor | PKC activator |
| Tirbanibulin | 0.00095 | 8.7 | 0.11451 | -3.3 | Kinase inhibitor | Syk inhibitor |
| Rabusertib | 0.00115 | 2.5 | 1.00000 | 0.0 | Kinase inhibitor | TGF-B/Smad inhibitor |
| Omipalisib | 0.00194 | 10.7 | 0.84587 | -0.9 | Kinase inhibitor | MEK1/2 inhibitor |
| Vistusertib | 0.00221 | 8.1 | 0.23672 | -2.4 | Kinase inhibitor | TAK1 inhibitor |
| Mitotane | 0.00236 | 0.6 | NA | NA | Conventional Chemotherapeutic; Antimetabolite | Purine analog |
| Paclitaxel | 0.00239 | 9.7 | NA | NA | Kinase inhibitor | PI3K inhibitor. Pan-class I |
| Niraparib | 0.00267 | 2.4 | 0.74459 | 0.0 | Kinase inhibitor | PI3K/mTOR inhibitor |
| Topotecan | 0.00271 | 7.4 | 0.88853 | -0.6 | Conventional Chemotherapeutic | Nitrogen mustard alkylating agent |
| Raloxifene | 0.00276 | 4.5 | 0.18557 | -1.2 | Conventional Chemotherapeutic | Alkylating agent |
| Panobinostat | 0.00305 | 4.1 | 0.69961 | 1.4 | Kinase inhibitor; Tyrosine kinase inhibitor | FLT3, JAK2, TrkA, TrkB, TrkC inhibitor |
| Finasteride | 0.00310 | 0.6 | 0.79683 | 0.0 | Protease inhibitor | M1 family of aminopeptidase inhibitor (especially LTA4H and PSA) |
| Aminoglutethimide | 0.00320 | 0.4 | 0.81015 | 0.0 | Kinesin inhibitor | Eg5 inhibitor |
| GSK-1838705A | 0.00387 | 4.8 | 0.57137 | 1.1 | Kinase inhibitor | ERK1 & 2 inhibitor |
| CUDC-101 | 0.00400 | 10.6 | 0.23006 | -5.4 | Kinase inhibitor | HDAC & EGFR, Her2 inhibitor |
| Temsirolimus | 0.00525 | 6.3 | 0.80675 | 0.6 | Kinase inhibitor; Tyrosine kinase inhibitor | B-Raf, FGFR-1, VEGFR-2 & -3, PDGFR-beta, KIT, and FLT3 inhib |
| Vinorelbine | 0.00583 | 13.5 | 0.17757 | 5.5 | Conventional Chemotherapeutic | Topoisomerase II inhibitor |
| ZSTK474 | 0.00596 | 11.4 | NA | NA | Proteasome inhibitor | Proteasome inhibitor (26S subunit) |
| Etoposide | 0.00780 | 6.9 | 0.44502 | 0.0 | Kinase inhibitor | EGFR inhibitor |
| Erlotinib | 0.00858 | 2.9 | 0.48207 | 0.0 | Conventional Chemotherapeutic; Topoisomerase inhibitor; Anthracycline | Topoisomerase II inhibitor |
| Ruxolitinib | 0.00870 | 5.1 | 0.77624 | 0.2 | Kinase inhibitor; Tyrosine kinase inhibitor | JAK1&2 inhibitor |
| Pictilisib | 0.00878 | 7.8 | 0.78710 | 1.5 | Metabolic modifier | AMPK activator |
| Floxuridine | 0.01048 | 0.7 | 0.88278 | 0.0 | Hormone therapy | Aromatase inhibitor |
| Idarubicin | 0.01049 | 5.0 | 0.42708 | 0.9 | Conventional Chemotherapeutic | Mitotic inhibitor, epothilone microtubule stabilizer |
| MK-2206 | 0.01103 | 5.0 | 0.46640 | -2.4 | Kinase inhibitor; Aurora inhibitor | Aurora B inhibitor |
| PF-04691502 | 0.01134 | 6.4 | 0.51535 | -2.9 | Kinase inhibitor | PI3K/mTOR inhibitor |
| AZ 3146 | 0.01140 | 3.7 | 0.48740 | -0.8 | Kinase inhibitor; Tyrosine kinase inhibitor | Broad TK (FLT3, KIT, RET, JAK, EGFR...) inhibitor |
| APR-246 | 0.01282 | 6.1 | 0.20062 | 0.7 | Conventional Chemotherapeutic | Topoisomerase I inhibitor |
| JQ1 | 0.01300 | 6.8 | 0.76077 | 0.2 | Differentiation/Epigenetic modifier | Inactive stereoisomer of (+)JQ1 |
| Dactinomycin | 0.01310 | 5.3 | 0.20169 | 2.3 | Conventional Chemotherapeutic | Alkylating agent |
| Irinotecan | 0.01466 | 8.9 | 0.08850 | 9.5 | Other | S1PR antagonist |
| BMS-754807 | 0.01528 | 11.8 | 0.71004 | -0.8 | Kinase inhibitor | IGF1R inhibitor |
| Sepantronium bromide | 0.01543 | 7.7 | 0.59059 | -1.5 | Apoptotic modulator | Survivin inhibitor |
| VX-11E | 0.01602 | 2.5 | 0.59279 | -0.6 | Rapalog | binds FKBP12, causes inhibition of mTORC1 |
| Saracatinib | 0.01603 | 1.4 | NA | NA | Differentiation/Epigenetic modifier; PARP inhibitor | PARP inhibitor |
| ABT-751 | 0.01621 | 5.5 | 0.78966 | -0.6 | Conventional Chemotherapeutic | Mitotic inhibitor. Colchicine site binding microtubule depolymerizer. |
| Vinblastine | 0.01630 | 3.6 | 0.42052 | -1.2 | Differentiation/Epigenetic modifier | Retinoic acid receptor agonist |
| Bimatoprost | 0.01649 | 0.6 | 0.80144 | 0.0 | Immunomodulator | Prostaglandin analog |
| Plicamycin | 0.01664 | 16.1 | 0.45082 | 3.9 | Immunomodulator | Glucocorticoid, immunomodulatory agent |
| Daunorubicin | 0.01768 | 8.0 | 0.50484 | -1.8 | Conventional Chemotherapeutic | RNA and DNA synthesis inhibitor |
| Camptothecin | 0.01799 | 7.8 | 0.18016 | -2.3 | Kinase inhibitor; PI3K inhibitor | mTOR/(PI3K) inhibitor |
| 4-hydroxytamoxifen | 0.01802 | 2.1 | NA | NA | Kinase inhibitor | HER2, EGFR inhibitor |
| Valproic acid | 0.01806 | 2.9 | 0.84591 | -1.0 | Conventional Chemotherapeutic | Antimalaria agent, chemo/radio sensitizer |
| GSK-1904529A | 0.01830 | 11.6 | 0.70983 | -0.6 | Kinase inhibitor | DYRK1A inhibitor |
| Ixabepilone | 0.01952 | 1.8 | 0.81015 | 0.0 | Immunomodulator | Immunomodulatory agent, TLR7 agonist |
| Tipifarnib | 0.01981 | 9.4 | NA | NA | HSP inhibitor | HSP90 inhibitor |
| Bicalutamide | 0.02336 | 0.2 | 0.88533 | 0.0 | HSP inhibitor | HSP70 inhibitor |
| Thioguanine | 0.02411 | 3.2 | 0.27579 | -1.0 | Conventional Chemotherapeutic | Alkylating agent |
| Canertinib | 0.02546 | 2.8 | 0.44511 | 1.5 | Kinase inhibitor; Tyrosine kinase inhibitor | VEGFR, PDGFR, KIT inhibitor |
| Lenalidomide | 0.02564 | 3.2 | 0.72491 | -0.9 | Conventional Chemotherapeutic | Topoisomerase I inhibitor. Camptothecin prodrug analog |
| Serdemetan | 0.02584 | 4.1 | 0.59374 | 0.5 | Kinase inhibitor; MEK inhibitor | MEK1/2 inhibitor |
| Tofacitinib | 0.02703 | 8.1 | 1.00000 | 0.0 | Conventional Chemotherapeutic; Topoisomerase inhibitor; Anthracycline | Topoisomerase II inhibitor |
| Thalidomide | 0.02704 | 0.2 | NA | NA | Conventional Chemotherapeutic | Mitotic inhibitor. Vinca alkaloid microtubule depolymerizer |
| Apitolisib | 0.02755 | 5.8 | 0.47062 | -2.2 | Hormone therapy | Nonsteroidal antiandrogen |
| Teniposide | 0.02777 | 11.8 | 0.14498 | 4.6 | Conventional Chemotherapeutic; Antimetabolite | Pyrimidin analog |
| Docetaxel | 0.02809 | 7.6 | NA | NA | Differentiation/Epigenetic modifier | HDAC inhibitor |

| | | | | | | |
|---|---|---|---|---|---|---|
| PFI-1 | 0.02893 | 2.1 | 0.47885 | -1.1 | Conventional Chemotherapeutic | Antirheumatic agent |
| TAK-901 | 0.03196 | 5.1 | 0.45712 | -2.3 | Kinase inhibitor; Tyrosine kinase inhibitor | Btk inhibitor |
| Nintedanib | 0.03365 | 4.4 | 0.89355 | -1.9 | Hormone therapy | selective estrogen receptor modulator |
| Simvastatin | 0.03400 | 5.4 | 0.60171 | 1.0 | Kinase inhibitor; Tyrosine kinase inhibitor | PDGFRA inhibitor, PDGFRB inhibitor |
| Vincristine | 0.03456 | 5.7 | 0.13638 | -2.8 | Apoptotic modulator | MDM2 inhibitor |
| SGC-CBP30 | 0.03479 | 2.9 | 0.97730 | 1.3 | Kinase inhibitor | IKK-2 inhibitor |
| Bexarotene | 0.03528 | 2.7 | 0.51850 | 0.0 | Hormone therapy | Anti-steroid, aromatase inhibitor |
| Toremifene | 0.03530 | 1.7 | 0.18574 | -1.3 | Immunomodulator | Immunomodulatory agent |
| AZD8055 | 0.03690 | 9.4 | 0.03855 | -4.3 | Kinase inhibitor; mTOR inhibitor | mTOR inhibitor |
| Sapanisertib | 0.03768 | 8.2 | 0.77148 | 1.0 | Kinase inhibitor; Tyrosine kinase inhibitor | JAK1 & 2 inhibitor |
| GSK2636771 | 0.03817 | 0.4 | 0.75541 | 0.0 | Hormone therapy | Estrogen receptor antagonist |
| Vorinostat | 0.03855 | 1.5 | 0.62511 | -0.4 | Hedgehog inhibitor | Smoothened (Hh) inhibitor |
| Tivozanib | 0.04002 | 3.5 | 0.31206 | 3.0 | Kinase inhibitor | VEGFR1, 2, 3, c-Kit, PDGFRB inhibitor |
| Cabozantinib | 0.04075 | 2.3 | 0.52219 | 0.0 | Kinase inhibitor | PLK1 inhibitor |
| Gandotinib | 0.04082 | 4.6 | 0.33522 | 2.4 | Apoptotic modulator | p53 activator, thioredoxin reductase 1 inhibitor |
| Tanespimycin | 0.04096 | 4.6 | 0.37421 | -0.8 | Kinase inhibitor | CDK inhibitor |
| Nutlin-3 | 0.04268 | -2.7 | 0.01433 | -2.6 | Differentiation/Epigenetic modifier; HDAC inhibitor | HDAC1 inhibitor, HDAC2 inhibitor |
| Omacetaxine | 0.04579 | 2.7 | 0.39816 | 1.8 | Other | STAT3-DNA interaction inhibitor |
| Mitoxantrone | 0.04614 | 3.6 | 0.59366 | -0.4 | Conventional Chemotherapeutic | Antineoplastic agent |
| NVP-AEW541 | 0.04724 | 3.0 | 0.42168 | -0.5 | Kinase inhibitor; Tyrosine kinase inhibitor | JAK2 inhibitor |
| Clomifene | 0.04890 | 1.4 | 0.25675 | -2.2 | Conventional Chemotherapeutic | Mitotic inhibitor, Vinca alkaloid microtubule depolymerizer |
| Carfilzomib | 0.05025 | 3.4 | 0.48474 | -0.6 | Kinase inhibitor; Tyrosine kinase inhibitor | FLT3 inhibitor |
| Valrubicin | 0.05088 | 6.4 | 1.00000 | 0.1 | Conventional Chemotherapeutic | Alkylating agent |
| Trametinib | 0.05096 | 5.8 | 0.61937 | -1.6 | Kinase inhibitor; CHK inhibitor | Chk1 inhibitor |
| AT9283 | 0.05238 | 4.0 | 0.31799 | -0.6 | Kinase inhibitor; Tyrosine kinase inhibitor; Aurora inhibitor | Aurora A & B, Jak2, Flt, Abl inhibitor |
| Neratinib | 0.05257 | 3.2 | NA | NA | Differentiation/Epigenetic modifier | BET family inhibitor |
| Olaparib | 0.05284 | 1.8 | 0.04687 | 4.2 | Kinase inhibitor; Tyrosine kinase inhibitor | Abl inhibitor |
| Cyclopamine | 0.05517 | 2.4 | 0.54687 | 0.0 | Conventional Chemotherapeutic | Nitrogen mustard alkylating agent |
| Tretinoin | 0.05780 | 2.4 | 0.37363 | 1.9 | Kinase inhibitor; Tyrosine kinase inhibitor | Abl, Src inhibitor |
| Dasatinib | 0.05899 | 3.5 | 0.38274 | 1.9 | Kinase inhibitor | PI3K gamma selective inhibitor |
| Chlorambucil | 0.05932 | 1.9 | 0.56783 | 0.0 | NSAID | Selective COX-2 inhibitor |
| Bortezomib | 0.06100 | 1.6 | 0.18755 | 1.4 | Conventional Chemotherapeutic; Antimetabolite | Purine analog |
| Ridaforolimus | 0.06256 | 11.7 | 0.20247 | -4.3 | Differentiation/Epigenetic modifier | HDAC inhibitor |
| SNS-032 | 0.06340 | 5.5 | 0.38611 | -6.8 | Apoptotic modulator | HDM2-p53 antagonist |
| Veliparib | 0.06535 | 1.4 | 0.56909 | -0.2 | Kinase inhibitor | EGFR inhibitor |
| Infigratinib | 0.06736 | 0.0 | 1.00000 | 0.0 | Kinase inhibitor | ERK1 & 2 inhibitor |
| BI 2536 | 0.06763 | 5.8 | 0.84715 | 0.6 | HSP inhibitor | HSP90 inhibitor |
| Ralimetinib | 0.06782 | 1.9 | 0.58740 | -1.9 | Kinase inhibitor | CK1epsilon and CK1delta inhibitor |
| Midostaurin | 0.06784 | 4.6 | 0.40595 | -1.5 | Kinase inhibitor | PKC inhibitor |
| IRAK1/4 inhibitor | 0.07136 | -0.5 | 0.07136 | -0.5 | Immunomodulator | S100A9 inhibitor, immunomodulatory, anti-angiogenic |
| Linsitinib | 0.07179 | 6.1 | 0.50698 | 0.2 | Kinase inhibitor | IGF1R, IR inhibitor |
| Belinostat | 0.07221 | 9.5 | 0.26507 | -3.7 | Differentiation/Epigenetic modifier; HDAC inhibitor | HDAC inhibitor |
| Sonolisib | 0.07259 | 2.7 | 0.36712 | -2.2 | Proteasome inhibitor | Proteasome inhibitor (20S subunit) |
| Navitoclax | 0.07296 | -5.4 | 0.04097 | -4.9 | Kinase inhibitor | VEGFR, PDGFR, Ret, Kit inhibitor |
| Prednisolone | 0.07668 | 4.4 | 0.68715 | -1.3 | Conventional Chemotherapeutic | Mitotic inhibitor, taxane microtubule stabilizer |
| TPCA-1 | 0.07675 | 3.4 | 0.32877 | -2.1 | Kinase inhibitor | PIM kinase inhibitor |
| Fasudil | 0.07677 | 2.4 | 0.43446 | 0.6 | Differentiation/Epigenetic modifier; HDAC inhibitor | HDAC inhibitor |
| Lapatinib | 0.07697 | 0.2 | 0.65927 | 0.0 | Tyrosine kinase inhibitor; Kinase inhibitor | Bcr-Abl inhibitor, Kit inhibitor, PDGFRB inhibitor |
| AZ191 | 0.07739 | 2.7 | 0.49871 | -0.6 | Other | alphaVbeta3 integrin inhibitor |
| Tivantinib | 0.07991 | 1.1 | 0.76033 | 0.0 | Metabolic modifier | NAMPT inhibitor |
| PHA 408 | 0.08459 | 1.3 | 0.07575 | 0.9 | Kinase inhibitor | PIM1, 2, 3 kinase inhibitor |
| Ipatasertib | 0.09079 | 1.6 | 0.50579 | -2.6 | Kinase inhibitor | JAK2 inhibitor |
| Alvocidib | 0.09145 | 3.5 | 0.79598 | 0.3 | Differentiation/Epigenetic modifier | PARP inhibitor |
| Pomalidomide | 0.09155 | 6.0 | 0.78647 | -1.1 | Conventional Chemotherapeutic | Alkylating agent |
| Axitinib | 0.09232 | 0.6 | 0.51372 | 0.1 | Hormone therapy | P450 17alpha-hydroxylase-17,20-lyase inhibitor |
| AZD1775 | 0.09278 | 6.9 | 0.55282 | 1.7 | Conventional Chemotherapeutic; Antimetabolite | Purine analog, DNA, RNA synth inhibitor |
| Seliciclib | 0.09423 | 1.3 | 0.81830 | 0.2 | Kinase inhibitor | VEGFR inhibitor |
| UCN-01 | 0.09456 | 16.6 | 0.92308 | 1.6 | Kinase inhibitor | JAK2 inhibitor |
| PF-00477736 | 0.09462 | 10.7 | 0.93984 | 1.4 | Kinase inhibitor | Chk1 inhibitor |
| PF-04708671 | 0.09476 | 1.6 | 0.62240 | -0.6 | Kinase inhibitor | VEGFR inhibitor |
| Allopurinol | 0.09632 | 0.0 | 1.00000 | 0.0 | Conventional Chemotherapeutic | Nitrogen mustard alkylating agent |
| GSK-J4 | 0.09682 | 2.4 | 0.49962 | 0.9 | Differentiation/Epigenetic modifier | PHD2 inhibitor |

| | | | | | | |
|---|---|---|---|---|---|---|
| Gemcitabine | 0.09962 | 9.2 | 0.10021 | 5.8 | Conventional Chemotherapeutic; Antimetabolite | Purine analog |
| 8-chloro-adenosine | 0.09980 | 6.5 | 0.78951 | 0.4 | Differentiation/Epigenetic modifier | p300/CREB-binding protein (CBP) inhibitor |
| AZD7762 | 0.10033 | 2.6 | 0.17780 | -1.3 | Kinase inhibitor; AKT inhibitor | AKT inhibitor |
| 2-methoxyestradiol | 0.10755 | 4.2 | 0.94906 | -0.1 | Other | Angiogenesis inhibitor |
| AVN944 | 0.10763 | 4.3 | 0.21363 | 5.4 | Kinase inhibitor | SGK1 & 2 inhibitor |
| Oprozomib | 0.10775 | 4.7 | 0.84083 | 0.9 | Kinase inhibitor | IKK-2 inhibitor |
| PF-670462 | 0.10937 | 0.3 | 0.51196 | 0.0 | Kinase inhibitor | CK1epsilon inhibitor |
| Foretinib | 0.10996 | 3.8 | 0.83008 | 0.9 | Kinase inhibitor | PLK1 inhibitor |
| PS-1145 | 0.11116 | 1.1 | 0.36982 | 1.6 | Immunomodulator | IDO inhibitor |
| PF-3845 | 0.11136 | 1.1 | 0.71012 | -1.1 | Kinase inhibitor; Tyrosine kinase inhibitor | Src inhibitor |
| Clofarabine | 0.11136 | 2.9 | 0.49795 | -2.4 | Conventional Chemotherapeutic | Topoisomerase II inhibitor |
| Mercaptopurine | 0.11193 | 1.4 | 0.47074 | 0.0 | Hormone therapy | Progestogen |
| I-BET151 | 0.11467 | 1.7 | 0.22370 | -2.1 | Kinase inhibitor; AKT inhibitor | AKT inhibitor |
| IOX-2 | 0.12379 | 1.2 | 0.08861 | 3.4 | Kinase inhibitor; Tyrosine kinase inhibitor | Abl inhibitor, Src inhibitor, Kit inhibitor, EphR inhibitor |
| Cilengitide | 0.12508 | 1.1 | 0.67495 | 1.4 | Proteasome inhibitor | LMP7 (immunoproteasome) |
| Alvespimycin | 0.12697 | 1.7 | 0.96678 | -0.2 | Kinase inhibitor; MEK inhibitor | MEK1/2 inhibitor |
| Lestaurtinib | 0.12863 | 2.5 | 0.37580 | -0.3 | Rapalog | binds FKBP12, causes inhibition of mTORC1 |
| Vatalanib | 0.13119 | 1.6 | 0.05933 | 0.7 | Kinase inhibitor; CDK inhibitor | CDK2/7/9 inhibitor |
| TGX-221 | 0.13122 | 2.9 | 0.08535 | 10.5 | Kinase inhibitor | p110alpha selective PI3K inhibitor |
| Pravastatin | 0.13370 | 0.3 | 0.45835 | 0.9 | Other | Indolamine 2,3-dioxygenase 1 and 2 inhibitor |
| Mitomycin | 0.13545 | 3.8 | NA | NA | Kinase inhibitor | mTOR/PI3K inhibitor |
| AZD1480 | 0.13556 | 4.0 | 1.00000 | 0.0 | Kinase inhibitor | MEK1/2 inhibitor |
| AT-406 | 0.14185 | 1.2 | 0.52389 | -1.6 | Kinase inhibitor | Allosteric ABL, FLT3, TIE2, TRKA inhibitor |
| Buparlisib | 0.14241 | 2.8 | 0.49164 | -1.9 | Kinase inhibitor | VEGFR, PDGFR, FGFR inhibitor |
| Lomeguatrib | 0.14328 | 0.1 | 0.74459 | 0.0 | Apoptotic modulator | XIAP, cIAP1, cIAP2 inhibitor |
| Afatinib | 0.14426 | 0.1 | NA | NA | Differentiation/Epigenetic modifier | Farnesyltransferase inhibitor |
| Prednisone | 0.14568 | 0.1 | 0.73849 | 0.0 | Hormone therapy | Nonsteroidal antiandrogen |
| Pentostatin | 0.14756 | 0.2 | 0.92787 | 0.0 | Conventional Chemotherapeutic; Topoisomerase inhibitor | Topoisomerase II inhibitor |
| Doramapimod | 0.14829 | 4.1 | 0.65833 | 0.7 | Kinase inhibitor | IGF1R inhibitor |
| Indibulin | 0.14968 | 5.1 | 0.11496 | 2.1 | Kinase inhibitor | JAK3, JAK2(V617F) inhibitor |
| Masitinib | 0.15766 | 1.9 | 0.33105 | 2.3 | Kinase inhibitor | ROCK, PKA, PKG, PRK inhibitor, prodrug |
| StemRegenin 1 | 0.16033 | 1.7 | 0.33862 | 2.2 | Other | Secretory phospholipase A2 inhibitor |
| Mocetinostat | 0.16591 | 3.0 | 0.73528 | 3.2 | Immunomodulator; Steroid | Glucocorticoid |
| Nilotinib | 0.17022 | 2.9 | 0.08027 | 3.1 | Immunomodulator | Immunomodulatory |
| Palbociclib | 0.17199 | 2.6 | 1.00000 | 0.2 | Apoptotic modulator | Bcl-2 family inhibitor |
| Refametinib | 0.17403 | 5.2 | 0.16822 | -2.6 | Kinase inhibitor | MEK1/2 inhibitor |
| RAF-265 | 0.17539 | -0.8 | 0.00925 | -0.8 | Kinase inhibitor | Chk1 inhibitor |
| Methotrexate | 0.17636 | 1.8 | 0.43702 | 0.0 | Conventional Chemotherapeutic; Alkylating agent | Alkylating agent |
| GSK269962 | 0.17639 | 1.2 | 0.81553 | -0.7 | Differentiation/Epigenetic modifier | O6-methylguanine-DNA methyltransferase inhibitor |
| Crizotinib | 0.18091 | 1.0 | 0.64362 | 0.0 | Conventional Chemotherapeutic; Alkylating agent | Alkylating agent |
| Perifosine | 0.18181 | 0.1 | 0.70480 | 0.0 | Kinase inhibitor | Syk inhibitor |
| Bosutinib | 0.18454 | 4.0 | NA | NA | Hormone therapy | AR antagonist |
| Cladribine | 0.18455 | 0.4 | 0.83675 | -0.5 | Conventional Chemotherapeutic | Nitrogen mustard alkylating agent |
| Luminespib | 0.18866 | 6.4 | 0.16294 | -7.3 | Kinase inhibitor; Tyrosine kinase inhibitor | Broad TK inhibitor |
| Binimetinib | 0.19025 | 9.7 | NA | NA | Kinase inhibitor | MET inhibitor |
| Auranofin | 0.19135 | 3.0 | 0.23844 | 1.8 | Other | Endogenous PPARŒ≥ ligand, prostaglandin, NFkB signaling inhibitor |
| Rofecoxib | 0.19286 | 0.0 | 0.88017 | 0.0 | Kinase inhibitor | B-Raf(V600E) inhibitor |
| Dinaciclib | 0.19292 | 2.0 | 0.69614 | -1.7 | Kinase inhibitor | pan-HER inhibitor |
| PF 431396 | 0.19550 | 2.6 | 0.75242 | -1.0 | Kinase inhibitor | ROCK1 and ROCK2 inhibitor |
| Crenolanib | 0.19892 | 2.3 | 0.85097 | 0.0 | Kinase inhibitor | p38MAPK inhibitor |
| Rocilinostat | 0.20125 | 1.9 | 0.75886 | -0.3 | Apoptotic modulator | Bcl-XL inhibitor |
| Procarbazine | 0.20156 | 0.0 | NA | NA | Conventional Chemotherapeutic | Alkylating agent |
| RD162 | 0.20514 | 0.6 | 0.05577 | 4.8 | Hormone therapy | Selective estrogen receptor modulator |
| PIK-75 | 0.20579 | 4.4 | NA | NA | Kinase inhibitor | mTOR inhibitor |
| Stattic | 0.20762 | 3.4 | 0.85929 | 0.4 | Metabolic modifier | HMG-CoA reductase inhibitor |
| Pimasertib | 0.21364 | 2.6 | 0.73491 | 0.8 | Kinase inhibitor | MEK1/2 inhibitor |
| Tubastatin A | 0.21450 | 3.1 | 0.09288 | 6.6 | Differentiation/Epigenetic modifier | HDAC6 inhibitor |
| CUDC-907 | 0.21555 | 1.5 | 0.58980 | -0.1 | Metabolic modifier | FAAH inhibitor |
| TAK-580 | 0.21823 | 0.9 | 0.06333 | -2.1 | Kinase inhibitor | ROCK, PKA, PKG, PRK inhibitor |
| S-trityl-L-cysteine | 0.21845 | 0.2 | 0.51828 | 0.0 | Kinase inhibitor | Mps1 kinase (TTK) inhibitor |
| Vandetanib | 0.22028 | -0.1 | 0.46687 | 2.8 | Kinase inhibitor | VEGFR,EGFR, RET inhibitor |
| Geldanamycin | 0.22610 | 6.7 | 0.57365 | -3.4 | Other | Smothened inhibitor |

| | | | | | | |
|---|---|---|---|---|---|---|
| Hydroxyfasudil | 0.22833 | 2.9 | 0.04642 | -0.2 | Kinase inhibitor | IGF1R, INSR inhib |
| Capecitabine | 0.23189 | 0.3 | 0.68413 | 0.0 | Kinase inhibitor; Tyrosine kinase inhibitor | VEGFR2, Met, FLT3, Tie2, Kit and Ret inhibitor |
| Galiellalactone | 0.23202 | 2.3 | 0.42044 | -4.5 | HSP inhibitor | HSP90 inhibitor |
| Methylprednisolone | 0.23991 | 1.1 | 0.90360 | -0.6 | Hormone therapy | Selective estrogen receptor modulator |
| OSI-027 | 0.24187 | 5.9 | 0.42355 | -3.2 | Kinase inhibitor | mTOR inhibitor |
| Chloroquine | 0.24260 | 1.9 | 0.59050 | -1.8 | Kinase inhibitor | AKT/PI3K inhibitor |
| Tacrolimus | 0.24442 | 2.0 | 0.65170 | 0.0 | Hormone therapy | type II 5-alpha reductase inhibitor |
| Ibrutinib | 0.24726 | 0.0 | 0.17481 | 0.0 | Kinase inhibitor | PI3K/mTOR inhibitor |
| TH588 | 0.24994 | -0.5 | 0.05297 | -0.6 | Kinase inhibitor | ATM inhibitor |
| Sotrastaurin | 0.25067 | 2.6 | NA | NA | Kinase inhibitor | PKCbeta inhibitor |
| AT 101 | 0.25171 | 2.2 | 0.84724 | 0.4 | Immunomodulator | Immunomodulatory agent |
| CEP-32496 | 0.25407 | 2.5 | 0.61456 | -0.2 | Metabolic modifier | MTH1 inhibitor |
| Pipobroman | 0.25660 | 0.0 | 0.81404 | 0.0 | Conventional Chemotherapeutic | Antimetabolite, Purine analog |
| URB597 | 0.25872 | 0.0 | 0.39953 | 0.0 | Kinase inhibitor | BRAF inhibitor |
| GSK343 | 0.25973 | 2.0 | 0.14839 | 2.7 | Kinase inhibitor | B-Raf(V600E) inhibitor |
| BMS-911543 | 0.26159 | 0.8 | 0.07864 | 2.2 | Apoptotic modulator | Bcl-2 inhibitor |
| Momelotinib | 0.26528 | 1.6 | 0.37978 | 0.0 | Kinase inhibitor | FGFR inhibitor |
| Volasertib | 0.26797 | 1.7 | 1.00000 | -0.1 | Kinase inhibitor | FGFR inhibitor |
| Idasanutlin | 0.27188 | -5.8 | 0.07489 | -6.8 | Differentiation/Epigenetic modifier | G9a/GLP inhibitor |
| TAK-733 | 0.27407 | 6.3 | 1.00000 | -0.2 | Other | gamma-secretase/notch inhibitor |
| Dacomitinib | 0.27921 | 0.6 | 0.38666 | 0.1 | Differentiation/Epigenetic modifier | BET family inhibitor |
| Dabrafenib | 0.27954 | 1.2 | 0.41475 | -0.7 | Kinase inhibitor; CDK inhibitor | CDK inhibitor |
| 8-amino-adenosine | 0.28546 | 6.0 | NA | NA | Conventional Chemotherapeutic | Nucleoside analog, RNA synthesis inhibitor |
| Azacitidine | 0.28862 | 2.2 | 0.10958 | 1.0 | Hormone therapy | Aromatase inhibitor |
| Tosedostat | 0.29185 | 1.8 | 0.17125 | -2.4 | Apoptotic modulator | Bcl-2 family inhibitor |
| ONX-0914 | 0.29560 | 3.3 | 0.98229 | -1.3 | Other | PORCN inhibitor |
| Fingolimod | 0.29719 | -0.5 | 0.36655 | 0.8 | Kinase inhibitor | PKCbeta inhibitor |
| SCH772984 | 0.29918 | 2.8 | 0.25546 | -1.4 | Differentiation/Epigenetic modifier | DOT1L inhibitor |
| AZD1208 | 0.30333 | 1.5 | 0.40784 | -0.1 | Kinase inhibitor | p38MAPK inhibitor |
| GDC-0919 | 0.30413 | 3.5 | 0.00841 | -0.7 | Immunomodulator | Nitric oxide-donating immunomodulator |
| Danusertib | 0.31137 | 0.9 | 0.51240 | -2.0 | Kinase inhibitor | Aurora, Ret, TrkA, FGFR-1 inhibitor |
| PAC-1 | 0.31560 | 1.5 | 0.43671 | -1.8 | Kinase inhibitor | mTOR inhibitor, ATP-competitive |
| Tamoxifen | 0.32594 | 1.1 | 0.56302 | -0.9 | Conventional Chemotherapeutic | Alkylating glucosamine-nitrosourea agent |
| Daporinad | 0.32604 | 2.9 | 0.97935 | -1.5 | Rapalog | binds FKBP12, causes inhibition of mTORC1 |
| Sirolimus | 0.32810 | 7.2 | NA | NA | Other | STAT3 SH2 domain inhibitor |
| BX-912 | 0.32848 | 1.2 | 0.27002 | -1.1 | Differentiation/Epigenetic modifier | HDAC1/2/3/10, PI3Kalpha inhibitor |
| Gefitinib | 0.33426 | 1.7 | 0.16454 | -1.7 | Conventional Chemotherapeutic | 5-FU prodrug |
| Fulvestrant | 0.34510 | 0.4 | 0.54047 | 0.0 | Conventional Chemotherapeutic | Antimetabolite |
| Lasofoxifene | 0.35039 | 0.0 | 0.69149 | 0.0 | Conventional Chemotherapeutic | Protein synthesis inhib (80 S ribosome) |
| Fedratinib | 0.35464 | -0.2 | 0.04485 | -1.7 | Conventional Chemotherapeutic; Antimetabolite | DNA synthesis |
| Rebastinib | 0.35924 | 0.0 | 0.88288 | 0.0 | Metabolic modifier | pyruvate dehydrogenase, alpha-ketoglutarate dehydrogenase inhibitor |
| Tasquinimod | 0.36077 | 0.0 | 0.69696 | 0.0 | Other | Ras-PDEdelta inhibitor |
| Cisplatin | 0.36241 | -0.9 | 0.33169 | -1.2 | Differentiation/Epigenetic modifier | Thioredoxin reductase inhibitor, cytotoxic chemotherapeutic |
| Roxadustat | 0.36752 | 0.0 | 0.64350 | 0.0 | Apoptotic modulator | Procaspase-3 activator |
| Bleomycin | 0.36844 | 1.7 | 0.32153 | -2.7 | Antimetabolite | Pyrimidine analog, DNA methyl transferase inhibitor |
| Metformin | 0.37095 | 0.0 | 0.58274 | 0.0 | Differentiation/Epigenetic modifier | DOT1L inhibitor |
| Amonafide | 0.37512 | 1.2 | 0.88962 | -0.1 | Kinase inhibitor | ALK, c-Met inhibitor |
| Triapine | 0.37576 | -0.8 | 0.15354 | -1.8 | Kinase inhibitor | IRAK1/4 inhibitor |
| Fluorouracil | 0.38140 | 0.0 | 0.88028 | 0.0 | Conventional Chemotherapeutic; Antimetabolite | Purine analog |
| Mepacrine | 0.38329 | 1.3 | 0.12076 | 1.7 | Conventional Chemotherapeutic; Alkylating agent | Nitrogen mustard alkylating agent |
| 1-methyl-D-tryptophan | 0.38393 | 0.0 | 0.16273 | 0.1 | Other | Unclear. PLA2 inhibitor. NF-kB inhibitor, p53 activator |
| NMS-873 | 0.39040 | 2.2 | 0.67505 | -1.2 | Apoptotic modulator | p53-MDM2 inhibitor |
| Atorvastatin | 0.39187 | 4.2 | 0.38546 | 4.2 | Differentiation/Epigenetic modifier | HDAC6 inhibitor |
| Linifanib | 0.39999 | 0.0 | 0.66604 | 0.0 | Differentiation/Epigenetic modifier; BET inhibitor | BET family inhibitor |
| Pevonedistat | 0.40117 | 4.8 | 0.77065 | -1.3 | Kinase inhibitor | PI3K beta selective inhibitor |
| Entinostat | 0.40295 | 1.1 | 0.95929 | 1.5 | Differentiation/Epigenetic modifier | HDAC inhibitor |
| SGC0946 | 0.40980 | 0.0 | 0.92169 | -0.3 | Differentiation/Epigenetic modifier | EZH2 inhibitor |
| C646 | 0.41617 | 0.9 | 0.80697 | -0.7 | Differentiation/Epigenetic modifier | L3MBTL3 inhibitor |
| VER 155008 | 0.42166 | 1.3 | 0.40344 | 0.5 | Kinase inhibitor | PI3K inhibitor, pan-class I. Irreversible |
| KU-60019 | 0.42462 | 1.0 | 0.54953 | -0.9 | Kinase inhibitor | pan-RAF inhibitor |
| Arsenic(III) oxide | 0.42542 | 0.0 | 0.22886 | 0.3 | Conventional Chemotherapeutic | Platinum-based antineoplastic agent |
| SB 743921 | 0.42878 | 0.0 | 0.75007 | 0.0 | Conventional Chemotherapeutic | Nucleoside analog, RNA synthesis inhibitor |

| | | | | | | |
|---|---|---|---|---|---|---|
| Nelarabine | 0.42883 | -3.4 | 1.00000 | 0.2 | Kinase inhibitor | IGF1R, INSR, ALK inhib |
| Ruboxistaurin | 0.43057 | 2.0 | 0.36844 | 1.9 | Differentiation/Epigenetic modifier | Tankyrase-1 and -2 |
| Plerixafor | 0.43283 | 0.0 | 0.77379 | 0.0 | Kinase inhibitor | Wee1 inhibitor |
| Pilocarpine | 0.43880 | 0.0 | 0.75541 | 0.0 | Hormone therapy | Gonadotropin releasing hormone superagonist |
| 15D-PGJ2 | 0.43880 | 0.0 | 0.75541 | 0.0 | Differentiation/Epigenetic modifier | AHR antagonist, stem cell regenerating |
| Abiraterone | 0.45544 | 0.2 | 0.11192 | 0.6 | Kinase inhibitor; CDK inhibitor | CDK inhibitor |
| Anagrelide | 0.45976 | 0.0 | 0.72533 | 0.0 | Other | Non-selective muscarinic receptor agonist |
| Cyclophosphamide | 0.46007 | 0.0 | 0.72467 | 0.0 | Conventional Chemotherapeutic; Alkylating agent | Alkylating agent |
| Fostamatinib | 0.46819 | 0.8 | 0.42445 | -2.1 | Metabolic modifier | Dihydrofolate reductase inhibitor |
| Rucaparib | 0.47511 | 1.2 | 0.51836 | 0.4 | Differentiation/Epigenetic modifier | PARP inhibitor |
| Birinapant | 0.48881 | 2.0 | 0.43910 | 3.0 | Metabolic modifier | ribonucleotide reductase inhibitor |
| Ifosfamide | 0.49161 | 0.0 | 0.80157 | 0.0 | Immunomodulator | Immunosuppressant |
| Quizartinib | 0.49317 | 1.0 | 0.20745 | 0.5 | Conventional Chemotherapeutic; Anthracycline; Topoisomerase inhibitor | Topoisomerase II inhibitor |
| Tarenflurbil | 0.49684 | 0.0 | 0.49684 | 0.0 | Other | Gamma-secretase inhibitor |
| Exemestane | 0.50011 | 1.5 | 0.40240 | -0.1 | Conventional Chemotherapeutic; Topoisomerase inhibitor | Topoisomerase II inhibitor |
| SGI-1776 | 0.50031 | 1.0 | 0.80204 | -0.1 | Proteasome inhibitor | Proteasome inhibitor (20S subunit) |
| Nilutamide | 0.50215 | 0.0 | 0.75017 | 0.0 | Conventional Chemotherapeutic | RNA synthesis inhibitor |
| Temozolomide | 0.50215 | 0.0 | 0.75017 | 0.0 | Conventional Chemotherapeutic; Topoisomerase inhibitor | Topoisomerase I inhibitor |
| NVP-LGK974 | 0.50358 | -0.3 | 0.11490 | -0.3 | Metabolic modifier | alcohol dehydrogenase inhibitor |
| CPI-613 | 0.51918 | 0.0 | 0.81395 | 0.0 | Differentiation/Epigenetic modifier | HIF prolyl hydroxylase inhibitor |
| Levamisole | 0.51935 | 0.0 | 0.81404 | 0.0 | Kinase inhibitor | p70S6K inhibitor |
| Megestrol acetate | 0.52917 | 0.0 | 0.69149 | 0.0 | Conventional Chemotherapeutic | Alkylating nitrosourea compound |
| Pinometostat | 0.53282 | 0.0 | 0.12075 | 0.1 | Kinase inhibitor | IKK-2 inhibitor |
| Decitabine | 0.54459 | 1.0 | NA | NA | | |
| Voxtalisib | 0.54649 | 0.1 | 0.52709 | -0.1 | Differentiation/Epigenetic modifier | G9a/GLP inhibitor |
| Obatoclax | 0.54761 | 1.6 | 0.22856 | -1.3 | Apoptotic modulator | Bcl-2inhibitor, Bcl-XL inhibitor |
| Tubacin | 0.57050 | 1.8 | 0.03807 | -1.9 | Kinase inhibitor | PI3K beta selective inhibitor |
| Tacedinaline | 0.57161 | 0.0 | 0.80589 | 0.0 | Kinase inhibitor; Tyrosine kinase inhibitor | Broad TK inhibitor |
| Enzastaurin | 0.57608 | 0.0 | 0.23730 | 0.0 | Kinase inhibitor | pan-HER inhibitor |
| Motesanib | 0.58012 | 0.0 | 0.81015 | 0.0 | | FF |
| Goserelin | 0.58296 | 0.0 | 0.73862 | 0.0 | Hormone therapy | Nonsteroidal antiandrogen |
| Pazopanib | 0.59897 | 0.0 | 0.02371 | 0.4 | Kinase inhibitor | KIT inhibitor |
| Ponatinib | 0.60178 | 0.6 | 0.51861 | -1.4 | Kinase inhibitor | MET, VEGFR2 inhibitor |
| BIIB021 | 0.60845 | 7.6 | 0.38586 | -3.4 | HSP inhibitor | HSP90 inhibitor |
| UNC0642 | 0.61249 | 0.8 | 0.84124 | 0.2 | Immunomodulator; Steroid | Glucocorticoid |
| PF-4800567 | 0.62004 | 0.0 | 0.76496 | 0.0 | NSAID | COX-2 inhibitor |
| Sunitinib | 0.62470 | 1.0 | 0.56443 | 0.0 | Kinase inhibitor | KDR/Flt/VEGFR inhibitor |
| Dovitinib | 0.63290 | 0.3 | 0.27831 | 0.7 | Antimetabolite | Pyrimidine analog, DNA methyl transferase inhibitor |
| Carboplatin | 0.63859 | -0.4 | 0.36155 | -0.4 | Differentiation/Epigenetic modifier | HDAC inhibitor |
| Cytarabine | 0.64064 | 0.9 | NA | NA | Conventional Chemotherapeutic; Alkylating agent | Antineoplastic anatibiotic, DNA crosslinker |
| Venetoclax | 0.64091 | 0.1 | 0.85344 | -2.6 | Metabolic modifier | IMPDH inhibitor |
| (5Z)-7-Oxozeaenol | 0.64549 | -1.4 | 0.03068 | -3.9 | Metabolic modifier | NAE inhibitor |
| Oxaliplatin | 0.67235 | -1.8 | NA | NA | Other | Iron chelator |
| Sorafenib | 0.67321 | 0.0 | 0.06256 | 0.0 | Kinase inhibitor | FLT3, PDGFR, KIT inhibitor |
| Bryostatin 1 | 0.67414 | 1.9 | 0.30461 | -1.1 | Conventional Chemotherapeutic | Mitoric inhibitor. Microtubule depolymerizer |
| Everolimus | 0.67486 | 0.3 | 0.49531 | -0.3 | Conventional Chemotherapeutic | Mitotic inhibitor, taxane microtubule stabilizer |
| GSK650394 | 0.67888 | 0.0 | 0.20612 | 0.5 | Kinase inhibitor | FAK/PYK2 inhibitor |
| Dexamethasone | 0.69156 | 1.6 | 0.43553 | 3.3 | Kinase inhibitor; PI3K inhibitor | PI3K inhibitor, pan-class I |
| Bendamustine | 0.69452 | 0.7 | 0.44470 | 0.0 | Kinase inhibitor | HER2, EGFR inhibitor |
| Altretamine | 0.71216 | 0.0 | 0.04510 | 0.0 | Conventional Chemotherapeutic | Xanthine oxidase inhibitor |
| Idelalisib | 0.72687 | -0.4 | 0.11543 | -2.7 | Kinase inhibitor | PI3K inhibitor, p110Œ¥-selective |
| Galunisertib | 0.74083 | 0.0 | 0.00133 | 0.0 | Kinase inhibitor; Tyrosine kinase inhibitor | VEGFR, PDGFR, CSF-1R, FLT3 inhibitor |
| Selumetinib | 0.74158 | 1.6 | 0.02378 | -5.9 | Kinase inhibitor; CDK inhibitor | CDK4/6 inhibitor |
| Varespladib | 0.74956 | 0.0 | NA | NA | Metabolic modifier | HMG-CoA reductase inhibitor |
| UNC1215 | 0.74956 | 0.0 | 0.01197 | 1.1 | Differentiation/Epigenetic modifier | JMJD3 (histone demethylase) inhibitor |
| XAV-939 | 0.75445 | 0.0 | 0.00092 | 0.4 | Kinase inhibitor; Tyrosine kinase inhibitor | B-Raf, c-Kit, VEGFR2 inhibitor |
| Anastrozole | 0.75445 | 0.0 | 1.00000 | 0.0 | Conventional Chemotherapeutic | Formaldehyde release, alkylating agent |
| Mechlorethamine | 0.75917 | -0.8 | 0.30309 | -0.8 | Kinase inhibitor | Aurora, Src family, JAK3, RTK inhibitor |
| Alisertib | 0.76815 | 0.5 | 0.37648 | -1.1 | Kinase inhibitor | Aurora A inhibitor |
| Tepotinib | 0.78437 | 0.0 | 0.09628 | 0.3 | Kinase inhibitor | c-Met inhibitor |
| WEHI-539 | 0.79032 | -0.1 | 0.06530 | -2.3 | Metabolic modifier | IDH1 R132H/R132C inhibitor |
| VGX-1027 | 0.79465 | 0.0 | 0.76398 | 0.1 | Proteasome inhibitor | Proteasome inhibitor (20S subunit) |

| | | | | | | |
|---|---|---|---|---|---|---|
| Cediranib | 0.81366 | 0.0 | 1.00000 | 0.0 | Differentiation/Epigenetic modifier | Antineoplastic agent, retinoid specifically selective for retinoid X receptors |
| Barasertib | 0.81366 | 0.0 | 1.00000 | 0.0 | HSP inhibitor | HSP90 inhibitor |
| Tandutinib | 0.81736 | 0.0 | 1.00000 | 0.0 | Differentiation/Epigenetic modifier; HDAC inhibitor | HDAC inhibitor |
| Brivanib | 0.81736 | 0.0 | 1.00000 | 0.0 | Kinase inhibitor; RAF inhibitor | C-Raf inhibitor, unclear MoA |
| Hydroxyurea | 0.81736 | 0.0 | 0.00151 | 0.2 | Conventional Chemotherapeutic | Alkylating agent |
| Dacarbazine | 0.82085 | 0.0 | 1.00000 | 0.0 | Conventional Chemotherapeutic | Antimetabolite, Purine analog |
| Uramustine | 0.82414 | 0.0 | 0.00031 | 2.4 | Conventional Chemotherapeutic | Mitotic inhibitor. Vinca alkaloid microtubule depolymerizer |
| (-)JQ1 | 0.82529 | 0.9 | 0.31854 | -0.7 | Hormone therapy | Selective estrogen receptor modulator |
| UNC0638 | 0.82716 | 0.2 | 0.87680 | 0.4 | Differentiation/Epigenetic modifier | CREBBP/EP300 bromodomain inhibitor |
| AGI-5198 | 0.83115 | 0.0 | 0.40889 | 0.0 | Kinase inhibitor | PDK1 inhibitor |
| Sonidegib | 0.83405 | 0.2 | 0.82232 | 0.1 | Immunomodulator | Immunomodulatory agent, anti-angiogenic |
| Pilaralisib | 0.83873 | 0.0 | 0.08437 | 0.7 | Kinase inhibitor | PI3K inhibitor, pan-class I |
| Thio-TEPA | 0.84501 | 0.0 | 0.65129 | 0.0 | Other | PDE-3, PLA2 inhibitor |
| Fludarabine | 0.85441 | -0.3 | 0.60630 | -1.9 | Conventional Chemotherapeutic | Antimetabolite, Analog of 5-fluorouracil |
| Vemurafenib | 0.85449 | -1.7 | 0.07064 | -3.5 | Conventional Chemotherapeutic | Topoisomerase II inhibitor / DNA intercalator |
| Enzalutamide | 0.87732 | 0.0 | NA | NA | Hormone therapy | AR antagonist |
| AZD4547 | 0.87737 | 0.0 | 1.00000 | 0.0 | Kinase inhibitor | JAK1/2, FGFR inhibitor |
| Deferoxamine | 0.88303 | -0.3 | 0.56150 | -0.3 | Metabolic modifier | HMG-CoA reductase inhibitor |
| Busulfan | 0.88523 | 0.0 | 1.00000 | 0.0 | Conventional Chemotherapeutic | Antineoplastic agent |
| Iniparib | 0.89194 | 1.6 | 0.11175 | -0.7 | Differentiation/Epigenetic modifier; PARP inhibitor | PARP inhibitor |
| Imiquimod | 0.90020 | 0.0 | 0.59389 | 0.0 | Hormone therapy | Estrogen receptor antagonist |
| Disulfiram | 0.91410 | 0.0 | 0.26406 | 0.5 | Other | p97/VCP inhibitor |
| Tamatinib | 0.91771 | 0.6 | 0.82151 | 0.1 | Antimetabolite | Anti-folate agent |
| Vismodegib | 0.93026 | -0.1 | 0.44736 | -0.1 | Kinase inhibitor | Src, Abl inhibitor |
| MK-0752 | 0.93264 | 0.0 | 0.27188 | 0.0 | Other | FAAH inhibitor |
| Ixazomib | 0.94608 | 1.7 | 0.42019 | 1.0 | Kinesin inhibitor | KSP/Eg5 inhibitor |
| Melphalan | 0.94790 | 0.0 | 0.98121 | 0.1 | Conventional Chemotherapeutic | Platinum-based antineoplastic agent |
| Pemetrexed | 0.96327 | 0.0 | 0.45353 | 0.0 | Kinesin inhibitor | Mitotic inhibitor. Eg5/KSP inhibitor |
| Imatinib | 0.96333 | 0.0 | 0.55780 | 0.0 | Kinase inhibitor | EGFR inhibitor |
| Celecoxib | 0.96796 | 0.0 | 0.60822 | 0.0 | Conventional Chemotherapeutic | Glycopeptide antibiotic, causes DNA breaks |
| Estramustine | 0.96798 | 0.0 | 0.60862 | 0.0 | Conventional Chemotherapeutic | Platinum-based antineoplastic agent |
| Filanesib | 0.97504 | 0.1 | 0.44417 | -0.2 | Differentiation/Epigenetic modifier; HDAC inhibitor | HDAC-6 selective inhibitor |
| Regorafenib | 0.99437 | 0.2 | 0.81721 | 0.0 | Kinase inhibitor | PKCbeta, PDK1, Chk, Cdk2 inhibitor |
| Flutamide | 1.00000 | 0.0 | 0.65160 | 0.0 | Rapalog | Binds FKBP12, causes inhibition of calcineurin |
| deltarasin | 1.00000 | -0.3 | 0.28283 | 0.8 | | |
| Letrozole | NA | NA | NA | NA | Conventional Chemotherapeutic | Mitotic inhibitor. Epothilone microtubule stabilizer. |
| Lomustine | NA | NA | NA | NA | Hormone therapy | Aromatase inhibitor |
| Streptozocin | NA | NA | NA | NA | Hormone therapy | Selective estrogen receptor modulator |
| Carmustine | NA | NA | NA | NA | Hedgehog inhibitor | Smoothened (Hh) inhibitor |
| IWP-L6 | NA | NA | NA | NA | Apoptotic modulator | IAP, SMAC mimetic |

**B.**

| **Drug Name** | **Number of samples screened** | **Number of samples screened with *TP53* mutation** | **Number of samples screened with monoallelic *TP53*** | **Standard deviation of samples screened** | **Standard deviation of samples screened with *TP53* mutation** | **Standard deviation of samples screened with monoallelic *TP53*** |
|---|---|---|---|---|---|---|
| Doxorubicin | 127 | 14 | 16 | 4.3 | 4.1 | 3.2 |
| Patupilone | 27 | 4 | 1 | 3.3 | 5.7 | NA |
| Quisinostat | 131 | 14 | 16 | 7.4 | 6.4 | 6.8 |
| Dactolisib | 120 | 12 | 17 | 6.5 | 4.1 | 5.6 |
| Tirbanibulin | 116 | 9 | 13 | 6.3 | 5.2 | 8.0 |
| Rabusertib | 28 | 3 | 1 | 0.9 | 2.2 | NA |
| Omipalisib | 129 | 12 | 16 | 9.5 | 4.7 | 7.6 |
| Vistusertib | 123 | 14 | 15 | 6.9 | 7.6 | 4.9 |
| Mitotane | 27 | 3 | 0 | 0.7 | 1.4 | NA |
| Paclitaxel | 27 | 4 | 0 | 4.2 | 6.5 | NA |
| Niraparib | 28 | 3 | 1 | 1.1 | 1.4 | NA |
| Topotecan | 104 | 8 | 14 | 5.5 | 6.1 | 5.7 |
| Raloxifene | 29 | 4 | 1 | 1.9 | 1.7 | NA |
| Panobinostat | 128 | 14 | 17 | 6.5 | 4.8 | 5.3 |
| Finasteride | 26 | 3 | 1 | 0.7 | 0.1 | NA |
| Aminoglutethimide | 30 | 4 | 1 | 0.4 | 0.9 | NA |
| GSK-1838705A | 93 | 7 | 16 | 5.8 | 2.6 | 4.9 |
| CUDC-101 | 30 | 4 | 1 | 7.3 | 2.0 | NA |
| Temsirolimus | 112 | 13 | 15 | 5.3 | 5.7 | 3.9 |
| Vinorelbine | 30 | 3 | 1 | 5.1 | 3.5 | NA |
| ZSTK474 | 28 | 3 | 0 | 6.3 | 3.3 | NA |
| Etoposide | 28 | 3 | 1 | 2.6 | 5.1 | NA |
| Erlotinib | 31 | 3 | 1 | 1.6 | 0.7 | NA |
| Ruxolitinib | 123 | 13 | 16 | 5.8 | 5.2 | 6.1 |
| Pictilisib | 123 | 13 | 15 | 6.9 | 6.5 | 5.8 |
| Floxuridine | 28 | 3 | 1 | 0.4 | 0.8 | NA |
| Idarubicin | 105 | 9 | 14 | 5.7 | 4.8 | 5.0 |
| MK-2206 | 28 | 4 | 1 | 3.7 | 2.9 | NA |
| PF-04691502 | 130 | 13 | 16 | 8.0 | 6.1 | 6.7 |
| AZ 3146 | 29 | 4 | 1 | 2.1 | 0.4 | NA |
| APR-246 | 130 | 14 | 17 | 5.7 | 6.7 | 6.8 |
| JQ1 | 130 | 14 | 17 | 6.7 | 7.1 | 5.6 |

| | | | | | | |
|---|---|---|---|---|---|---|
| Dactinomycin | 128 | 14 | 16 | 7.8 | 6.3 | 5.3 |
| Irinotecan | 30 | 3 | 1 | 4.0 | 7.3 | NA |
| BMS-754807 | 128 | 13 | 16 | 9.4 | 10.8 | 10.0 |
| Sepantronium bromide | 33 | 4 | 1 | 5.2 | 4.7 | NA |
| VX-11E | 30 | 4 | 1 | 1.7 | 1.5 | NA |
| Saracatinib | 30 | 4 | 0 | 0.7 | 1.0 | NA |
| ABT-751 | 30 | 4 | 1 | 2.8 | 2.9 | NA |
| Vinblastine | 106 | 10 | 11 | 4.4 | 6.0 | 2.9 |
| Bimatoprost | 28 | 4 | 1 | 0.6 | 0.5 | NA |
| Plicamycin | 127 | 14 | 16 | 10.4 | 10.8 | 11.6 |
| Daunorubicin | 30 | 4 | 1 | 5.8 | 5.0 | NA |
| Camptothecin | 89 | 6 | 9 | 5.3 | 7.9 | 3.6 |
| 4-hydroxytamoxifen | 28 | 4 | 0 | 1.9 | 1.2 | NA |
| Valproic acid | 31 | 4 | 1 | 2.2 | 0.6 | NA |
| GSK-1904529A | 69 | 4 | 12 | 8.7 | 3.7 | 6.9 |
| Ixabepilone | 29 | 3 | 1 | 0.8 | 1.1 | NA |
| Tipifarnib | 29 | 3 | 0 | 5.0 | 7.7 | NA |
| Bicalutamide | 30 | 4 | 1 | 0.2 | 0.4 | NA |
| Thioguanine | 29 | 3 | 1 | 2.1 | 3.3 | NA |
| Canertinib | 29 | 4 | 1 | 2.6 | 3.1 | NA |
| Lenalidomide | 120 | 12 | 15 | 6.3 | 6.8 | 6.3 |
| Serdemetan | 29 | 3 | 1 | 3.3 | 4.5 | NA |
| Tofacitinib | 30 | 2 | 1 | 3.1 | 3.3 | NA |
| Thalidomide | 28 | 4 | 0 | 0.6 | 1.2 | NA |
| Apitolisib | 100 | 9 | 12 | 7.2 | 5.6 | 5.2 |
| Teniposide | 28 | 3 | 1 | 5.4 | 7.6 | NA |
| Docetaxel | 27 | 4 | 0 | 5.3 | 10.6 | NA |
| PFI-1 | 95 | 8 | 12 | 3.1 | 1.9 | 2.5 |
| TAK-901 | 31 | 3 | 1 | 3.0 | 1.9 | NA |
| Nintedanib | 30 | 4 | 1 | 4.0 | 4.1 | NA |
| Simvastatin | 30 | 2 | 1 | 2.5 | 1.1 | NA |
| Vincristine | 93 | 8 | 12 | 6.3 | 5.4 | 4.5 |
| SGC-CBP30 | 96 | 10 | 15 | 4.4 | 5.6 | 3.4 |
| Bexarotene | 29 | 4 | 1 | 2.0 | 2.3 | NA |
| Toremifene | 29 | 4 | 1 | 1.5 | 0.9 | NA |
| AZD8055 | 123 | 13 | 15 | 9.3 | 10.3 | 5.3 |

| | | | | | | |
|---|---|---|---|---|---|---|
| Sapanisertib | 98 | 9 | 13 | 7.3 | 5.8 | 6.4 |
| GSK2636771 | 30 | 3 | 1 | 0.8 | 2.3 | NA |
| Vorinostat | 103 | 9 | 14 | 3.8 | 3.7 | 2.6 |
| Tivozanib | 26 | 4 | 1 | 2.5 | 1.8 | NA |
| Cabozantinib | 25 | 3 | 1 | 1.6 | 1.8 | NA |
| Gandotinib | 31 | 4 | 1 | 3.2 | 2.9 | NA |
| Tanespimycin | 122 | 12 | 15 | 6.4 | 4.9 | 6.5 |
| Nutlin-3 | 104 | 7 | 14 | 3.2 | 2.2 | 3.5 |
| Omacetaxine | 131 | 14 | 16 | 8.4 | 4.4 | 6.8 |
| Mitoxantrone | 85 | 5 | 9 | 3.1 | 3.7 | 2.9 |
| NVP-AEW541 | 97 | 10 | 15 | 5.1 | 4.2 | 4.9 |
| Clomifene | 29 | 3 | 1 | 2.0 | 1.4 | NA |
| Carfilzomib | 133 | 14 | 17 | 7.1 | 4.4 | 6.1 |
| Valrubicin | 31 | 4 | 1 | 5.4 | 4.2 | NA |
| Trametinib | 119 | 12 | 13 | 6.9 | 8.6 | 6.3 |
| AT9283 | 30 | 3 | 1 | 2.1 | 2.5 | NA |
| Neratinib | 28 | 3 | 0 | 2.2 | 1.3 | NA |
| Olaparib | 29 | 2 | 1 | 1.3 | 1.1 | NA |
| Cyclopamine | 28 | 4 | 1 | 1.3 | 1.9 | NA |
| Tretinoin | 29 | 3 | 1 | 1.8 | 1.0 | NA |
| Dasatinib | 97 | 8 | 12 | 5.4 | 8.3 | 3.1 |
| Chlorambucil | 30 | 4 | 1 | 1.1 | 2.0 | NA |
| Bortezomib | 125 | 12 | 16 | 7.1 | 3.3 | 3.5 |
| Ridaforolimus | 29 | 3 | 1 | 5.2 | 7.7 | NA |
| SNS-032 | 32 | 4 | 1 | 10.6 | 3.7 | NA |
| Veliparib | 104 | 8 | 14 | 1.9 | 2.2 | 2.2 |
| Infigratinib | 29 | 3 | 1 | 1.5 | 4.2 | NA |
| BI 2536 | 30 | 3 | 1 | 3.9 | 7.1 | NA |
| Ralimetinib | 94 | 6 | 13 | 6.0 | 5.5 | 5.0 |
| Midostaurin | 111 | 9 | 12 | 6.6 | 7.3 | 5.4 |
| IRAK1/4 inhibitor | 16 | 4 | 4 | 0.4 | 0.0 | 0.0 |
| Linsitinib | 123 | 12 | 17 | 4.2 | 5.9 | 3.3 |
| Belinostat | 31 | 3 | 1 | 6.1 | 5.3 | NA |
| Sonolisib | 30 | 3 | 1 | 2.9 | 4.1 | NA |
| Navitoclax | 131 | 13 | 16 | 8.4 | 5.7 | 7.6 |
| Prednisolone | 96 | 7 | 14 | 5.3 | 5.5 | 3.8 |

| | | | | | | |
|---|---|---|---|---|---|---|
| TPCA-1 | 98 | 10 | 15 | 5.1 | 6.0 | 5.4 |
| Fasudil | 28 | 3 | 1 | 1.9 | 1.4 | NA |
| Lapatinib | 31 | 4 | 1 | 1.4 | 1.1 | NA |
| AZ191 | 69 | 4 | 12 | 3.3 | 1.2 | 2.6 |
| Tivantinib | 32 | 4 | 1 | 0.9 | 1.2 | NA |
| PHA 408 | 95 | 10 | 15 | 2.3 | 2.3 | 1.6 |
| Ipatasertib | 126 | 14 | 16 | 5.5 | 6.2 | 5.8 |
| Alvocidib | 128 | 14 | 17 | 6.2 | 3.7 | 4.4 |
| Pomalidomide | 105 | 10 | 14 | 7.1 | 9.1 | 8.0 |
| Axitinib | 79 | 5 | 10 | 1.9 | 1.9 | 2.0 |
| AZD1775 | 28 | 4 | 1 | 4.5 | 7.7 | NA |
| Seliciclib | 41 | 5 | 5 | 1.6 | 1.4 | 1.4 |
| UCN-01 | 30 | 4 | 1 | 9.6 | 12.0 | NA |
| PF-00477736 | 27 | 4 | 1 | 5.8 | 9.3 | NA |
| PF-04708671 | 45 | 6 | 5 | 1.7 | 2.1 | 2.1 |
| Allopurinol | 29 | 3 | 1 | 0.2 | 0.5 | NA |
| GSK-J4 | 135 | 14 | 17 | 4.4 | 4.7 | 4.6 |
| Gemcitabine | 28 | 3 | 1 | 3.8 | 8.4 | NA |
| 8-chloro-adenosine | 30 | 4 | 1 | 5.3 | 6.3 | NA |
| AZD7762 | 103 | 9 | 13 | 3.4 | 4.4 | 2.0 |
| 2-methoxyestradiol | 32 | 4 | 1 | 2.2 | 3.5 | NA |
| AVN944 | 31 | 4 | 1 | 4.1 | 7.5 | NA |
| Oprozomib | 101 | 9 | 15 | 7.5 | 6.1 | 5.8 |
| PF-670462 | 48 | 8 | 5 | 1.7 | 3.2 | 0.1 |
| Foretinib | 85 | 6 | 10 | 3.9 | 3.5 | 2.2 |
| PS-1145 | 95 | 9 | 14 | 2.2 | 2.0 | 2.3 |
| PF-3845 | 32 | 4 | 1 | 2.0 | 0.6 | NA |
| Clofarabine | 126 | 14 | 16 | 7.1 | 4.7 | 6.4 |
| Mercaptopurine | 31 | 4 | 1 | 1.4 | 2.1 | NA |
| I-BET151 | 106 | 9 | 14 | 5.3 | 4.8 | 3.3 |
| IOX-2 | 30 | 3 | 1 | 1.1 | 0.7 | NA |
| Cilengitide | 16 | 3 | 4 | 1.7 | 0.5 | 2.4 |
| Alvespimycin | 115 | 11 | 15 | 5.3 | 4.8 | 5.9 |
| Lestaurtinib | 125 | 13 | 16 | 4.9 | 5.2 | 4.2 |
| Vatalanib | 29 | 4 | 1 | 1.3 | 2.0 | NA |
| TGX-221 | 29 | 4 | 1 | 2.5 | 3.5 | NA |

| | | | | | | |
|---|---|---|---|---|---|---|
| Pravastatin | 30 | 4 | 1 | 1.7 | 1.4 | NA |
| Mitomycin | 28 | 3 | 0 | 3.4 | 7.9 | NA |
| AZD1480 | 31 | 4 | 1 | 2.5 | 3.5 | NA |
| AT-406 | 102 | 7 | 14 | 4.4 | 3.3 | 3.8 |
| Buparlisib | 102 | 9 | 14 | 5.3 | 3.4 | 3.5 |
| Lomeguatrib | 29 | 4 | 1 | 0.5 | 0.9 | NA |
| Afatinib | 27 | 4 | 0 | 0.8 | 0.3 | NA |
| Prednisone | 28 | 4 | 1 | 1.7 | 3.8 | NA |
| Pentostatin | 121 | 13 | 16 | 4.5 | 8.6 | 3.1 |
| Doramapimod | 120 | 11 | 15 | 3.8 | 5.3 | 3.5 |
| Indibulin | 29 | 4 | 1 | 3.5 | 7.8 | NA |
| Masitinib | 31 | 4 | 1 | 3.4 | 1.8 | NA |
| StemRegenin 1 | 32 | 3 | 1 | 1.8 | 0.8 | NA |
| Mocetinostat | 102 | 9 | 12 | 5.3 | 3.8 | 4.5 |
| Nilotinib | 31 | 4 | 1 | 2.7 | 4.1 | NA |
| Palbociclib | 30 | 4 | 1 | 2.9 | 3.2 | NA |
| Refametinib | 127 | 14 | 17 | 8.3 | 9.6 | 7.0 |
| RAF-265 | 93 | 7 | 11 | 3.0 | 3.2 | 1.5 |
| Methotrexate | 30 | 3 | 1 | 3.5 | 10.6 | NA |
| GSK269962 | 121 | 11 | 17 | 3.1 | 2.7 | 3.9 |
| Crizotinib | 28 | 3 | 1 | 1.4 | 1.8 | NA |
| Perifosine | 30 | 3 | 1 | 0.8 | 0.2 | NA |
| Bosutinib | 31 | 4 | 0 | 2.7 | 4.1 | NA |
| Cladribine | 108 | 9 | 12 | 4.9 | 4.1 | 3.9 |
| Luminespib | 93 | 8 | 12 | 9.5 | 8.5 | 10.0 |
| Binimetinib | 28 | 3 | 0 | 4.6 | 8.6 | NA |
| Auranofin | 128 | 14 | 16 | 4.6 | 4.8 | 4.8 |
| Rofecoxib | 27 | 3 | 1 | 1.5 | 4.5 | NA |
| Dinaciclib | 99 | 8 | 14 | 7.5 | 1.8 | 5.9 |
| PF 431396 | 96 | 8 | 11 | 4.9 | 4.6 | 3.7 |
| Crenolanib | 87 | 6 | 9 | 4.8 | 3.0 | 1.6 |
| Rocilinostat | 99 | 10 | 15 | 3.2 | 3.5 | 2.7 |
| Procarbazine | 27 | 4 | 0 | 0.2 | 0.1 | NA |
| RD162 | 30 | 3 | 1 | 1.1 | 0.8 | NA |
| PIK-75 | 29 | 4 | 0 | 8.5 | 0.6 | NA |
| Stattic | 102 | 9 | 14 | 5.3 | 5.7 | 4.4 |

| | | | | | | |
|---|---|---|---|---|---|---|
| Pimasertib | 118 | 14 | 17 | 8.0 | 10.0 | 7.2 |
| Tubastatin A | 31 | 4 | 1 | 2.2 | 3.9 | NA |
| CUDC-907 | 72 | 5 | 13 | 10.2 | 2.4 | 8.0 |
| TAK-580 | 69 | 4 | 13 | 2.6 | 6.0 | 2.6 |
| S-trityl-L-cysteine | 28 | 3 | 1 | 0.8 | 0.2 | NA |
| Vandetanib | 24 | 4 | 1 | 2.8 | 0.5 | NA |
| Geldanamycin | 44 | 7 | 4 | 6.9 | 7.4 | 10.6 |
| Hydroxyfasudil | 68 | 5 | 12 | 1.3 | 1.8 | 0.4 |
| Capecitabine | 28 | 4 | 1 | 1.1 | 1.8 | NA |
| Galiellalactone | 29 | 3 | 1 | 3.1 | 2.4 | NA |
| Methylprednisolone | 121 | 13 | 15 | 6.4 | 7.2 | 6.7 |
| OSI-027 | 30 | 4 | 1 | 4.8 | 6.4 | NA |
| Chloroquine | 33 | 4 | 1 | 3.4 | 2.7 | NA |
| Tacrolimus | 30 | 4 | 1 | 2.4 | 3.0 | NA |
| Ibrutinib | 108 | 10 | 17 | 1.0 | 0.0 | 1.9 |
| TH588 | 72 | 3 | 13 | 1.8 | 0.4 | 0.6 |
| Sotrastaurin | 29 | 4 | 0 | 2.5 | 2.3 | NA |
| AT 101 | 30 | 3 | 1 | 2.8 | 3.0 | NA |
| CEP-32496 | 57 | 2 | 10 | 3.0 | 1.3 | 1.9 |
| Pipobroman | 30 | 3 | 1 | 0.9 | 2.9 | NA |
| URB597 | 71 | 5 | 11 | 1.4 | 0.0 | 0.9 |
| GSK343 | 28 | 4 | 1 | 2.3 | 4.1 | NA |
| BMS-911543 | 29 | 4 | 1 | 1.0 | 1.1 | NA |
| Momelotinib | 98 | 8 | 13 | 3.9 | 2.5 | 3.3 |
| Volasertib | 30 | 4 | 1 | 3.4 | 5.9 | NA |
| Idasanutlin | 74 | 5 | 12 | 7.5 | 6.6 | 8.0 |
| TAK-733 | 30 | 4 | 1 | 6.3 | 11.3 | NA |
| Dacomitinib | 30 | 4 | 1 | 1.7 | 4.1 | NA |
| Dabrafenib | 121 | 11 | 14 | 3.2 | 2.8 | 2.9 |
| 8-amino-adenosine | 30 | 4 | 0 | 5.0 | 7.3 | NA |
| Azacitidine | 98 | 9 | 14 | 3.0 | 2.2 | 2.7 |
| Tosedostat | 126 | 14 | 16 | 7.1 | 3.4 | 7.5 |
| ONX-0914 | 97 | 9 | 15 | 7.2 | 4.6 | 6.2 |
| Fingolimod | 26 | 1 | 1 | 1.2 | NA | NA |
| SCH772984 | 109 | 10 | 14 | 7.4 | 9.2 | 7.1 |
| AZD1208 | 76 | 4 | 13 | 3.8 | 1.3 | 2.6 |

| | | | | | | |
|---|---|---|---|---|---|---|
| GDC-0919 | 57 | 3 | 8 | 4.6 | 7.2 | 0.0 |
| Danusertib | 103 | 8 | 13 | 3.1 | 2.3 | 3.3 |
| PAC-1 | 32 | 3 | 1 | 1.7 | 0.9 | NA |
| Tamoxifen | 29 | 2 | 1 | 1.6 | 0.5 | NA |
| Daporinad | 125 | 14 | 15 | 9.2 | 9.2 | 9.5 |
| Sirolimus | 30 | 4 | 0 | 3.9 | 8.6 | NA |
| BX-912 | 68 | 5 | 12 | 3.1 | 1.3 | 2.5 |
| Gefitinib | 28 | 3 | 1 | 1.8 | 3.0 | NA |
| Fulvestrant | 30 | 3 | 1 | 1.3 | 1.7 | NA |
| Lasofoxifene | 29 | 4 | 1 | 0.7 | 0.0 | NA |
| Fedratinib | 72 | 5 | 13 | 3.9 | 3.2 | 2.6 |
| Rebastinib | 29 | 4 | 1 | 0.4 | 0.5 | NA |
| Tasquinimod | 15 | 4 | 3 | 2.0 | 0.0 | 3.6 |
| Cisplatin | 28 | 2 | 1 | 3.4 | 0.4 | NA |
| Roxadustat | 28 | 3 | 1 | 0.2 | 0.0 | NA |
| Bleomycin | 31 | 3 | 1 | 4.0 | 0.6 | NA |
| Metformin | 13 | 2 | 1 | 2.0 | 0.0 | NA |
| Amonafide | 29 | 4 | 1 | 2.7 | 1.4 | NA |
| Triapine | 15 | 3 | 4 | 1.8 | 1.9 | 1.9 |
| Fluorouracil | 28 | 4 | 1 | 0.6 | 0.8 | NA |
| Mepacrine | 103 | 9 | 14 | 6.7 | 8.6 | 5.4 |
| 1-methyl-D-tryptophan | 26 | 4 | 1 | 1.2 | 0.0 | NA |
| NMS-873 | 71 | 5 | 13 | 6.3 | 3.4 | 4.7 |
| Atorvastatin | 32 | 4 | 1 | 4.8 | 5.0 | NA |
| Linifanib | 31 | 3 | 1 | 0.6 | 0.0 | NA |
| Pevonedistat | 100 | 9 | 14 | 8.0 | 7.9 | 7.7 |
| Entinostat | 100 | 9 | 13 | 3.6 | 4.0 | 2.6 |
| SGC0946 | 96 | 9 | 14 | 2.4 | 1.8 | 2.4 |
| C646 | 102 | 8 | 14 | 3.0 | 3.3 | 3.3 |
| VER 155008 | 101 | 9 | 12 | 2.6 | 2.7 | 1.9 |
| KU-60019 | 70 | 4 | 13 | 2.7 | 1.6 | 2.8 |
| Arsenic(III) oxide | 28 | 3 | 1 | 0.4 | 0.0 | NA |
| SB 743921 | 30 | 4 | 1 | 4.2 | 0.0 | NA |
| Nelarabine | 26 | 4 | 1 | 6.0 | 8.4 | NA |
| Ruboxistaurin | 30 | 3 | 1 | 2.6 | 2.4 | NA |
| Plerixafor | 23 | 4 | 1 | 0.8 | 0.0 | NA |

| | | | | | | |
|---|---|---|---|---|---|---|
| Pilocarpine | 31 | 4 | 1 | 0.6 | 0.0 | NA |
| 15D-PGJ2 | 31 | 4 | 1 | 0.4 | 0.0 | NA |
| Abiraterone | 29 | 2 | 1 | 0.3 | 0.3 | NA |
| Anagrelide | 25 | 3 | 1 | 0.3 | 0.0 | NA |
| Cyclophosphamide | 25 | 3 | 1 | 0.5 | 1.3 | NA |
| Fostamatinib | 27 | 3 | 1 | 2.2 | 3.8 | NA |
| Rucaparib | 27 | 3 | 1 | 1.2 | 0.9 | NA |
| Birinapant | 14 | 3 | 4 | 3.7 | 5.3 | 4.2 |
| Ifosfamide | 28 | 4 | 1 | 0.6 | 0.0 | NA |
| Quizartinib | 98 | 9 | 9 | 4.2 | 2.4 | 4.8 |
| Tarenflurbil | 40 | 5 | 5 | 0.6 | 0.0 | 0.0 |
| Exemestane | 32 | 3 | 1 | 1.9 | 1.2 | NA |
| SGI-1776 | 93 | 9 | 16 | 2.1 | 2.3 | 2.0 |
| Nilutamide | 29 | 3 | 1 | 0.4 | 0.0 | NA |
| Temozolomide | 29 | 3 | 1 | 0.6 | 0.0 | NA |
| NVP-LGK974 | 68 | 3 | 11 | 1.7 | 1.0 | 0.6 |
| CPI-613 | 31 | 4 | 1 | 1.2 | 0.0 | NA |
| Levamisole | 31 | 4 | 1 | 0.5 | 0.0 | NA |
| Megestrol acetate | 27 | 2 | 1 | 1.0 | 0.0 | NA |
| Pinometostat | 94 | 10 | 13 | 1.3 | 0.1 | 1.3 |
| Decitabine | 27 | 3 | 0 | 2.6 | 0.5 | NA |
| Voxtalisib | 7 | 1 | 1 | 0.5 | NA | NA |
| Obatoclax | 100 | 8 | 14 | 2.9 | 3.2 | 2.1 |
| Tubacin | 102 | 8 | 12 | 3.2 | 6.3 | 2.4 |
| Tacedinaline | 28 | 3 | 1 | 0.0 | 0.0 | NA |
| Enzastaurin | 114 | 9 | 15 | 2.9 | 0.6 | 2.9 |
| Motesanib | 29 | 3 | 1 | 0.7 | 0.0 | NA |
| Goserelin | 26 | 2 | 1 | 0.3 | 0.0 | NA |
| Pazopanib | 28 | 4 | 1 | 0.4 | 0.0 | NA |
| Ponatinib | 31 | 4 | 1 | 2.6 | 3.0 | NA |
| BIIB021 | 28 | 4 | 1 | 8.0 | 9.6 | NA |
| UNC0642 | 30 | 4 | 1 | 1.7 | 1.4 | NA |
| PF-4800567 | 33 | 4 | 1 | 0.4 | 0.6 | NA |
| Sunitinib | 87 | 6 | 10 | 1.7 | 0.9 | 1.9 |
| Dovitinib | 96 | 9 | 13 | 1.3 | 0.4 | 1.1 |
| Carboplatin | 29 | 3 | 1 | 1.8 | 2.0 | NA |

| | | | | | | |
|---|---|---|---|---|---|---|
| Cytarabine | 29 | 3 | 0 | 3.6 | 3.3 | NA |
| Venetoclax | 127 | 14 | 17 | 10.0 | 7.4 | 10.2 |
| (5Z)-7-Oxozeaenol | 106 | 9 | 14 | 5.8 | 7.9 | 3.7 |
| Oxaliplatin | 29 | 4 | 0 | 5.9 | 5.2 | NA |
| Sorafenib | 83 | 7 | 10 | 4.4 | 0.2 | 1.2 |
| Bryostatin 1 | 109 | 12 | 15 | 8.3 | 7.3 | 5.8 |
| Everolimus | 98 | 9 | 14 | 4.9 | 6.5 | 2.8 |
| GSK650394 | 29 | 3 | 1 | 1.2 | 1.3 | NA |
| Dexamethasone | 123 | 12 | 15 | 8.3 | 9.5 | 7.9 |
| Bendamustine | 29 | 4 | 1 | 0.9 | 1.0 | NA |
| Altretamine | 94 | 6 | 13 | 0.7 | 0.0 | 0.8 |
| Idelalisib | 123 | 14 | 15 | 5.1 | 4.7 | 3.1 |
| Galunisertib | 87 | 9 | 13 | 0.3 | 0.0 | 0.8 |
| Selumetinib | 97 | 9 | 14 | 8.2 | 9.6 | 3.4 |
| Varespladib | 25 | 2 | 0 | 0.9 | 0.0 | NA |
| UNC1215 | 26 | 2 | 1 | 0.4 | 0.0 | NA |
| XAV-939 | 28 | 4 | 1 | 0.1 | 0.0 | NA |
| Anastrozole | 28 | 4 | 1 | 0.1 | 0.0 | NA |
| Mechlorethamine | 29 | 3 | 1 | 2.0 | 3.1 | NA |
| Alisertib | 97 | 7 | 13 | 3.1 | 2.2 | 1.8 |
| Tepotinib | 100 | 8 | 12 | 3.0 | 1.3 | 2.1 |
| WEHI-539 | 98 | 10 | 15 | 6.2 | 4.0 | 4.1 |
| VGX-1027 | 87 | 7 | 14 | 5.4 | 5.9 | 4.4 |
| Cediranib | 28 | 3 | 1 | 0.1 | 0.0 | NA |
| Barasertib | 28 | 3 | 1 | 0.7 | 0.0 | NA |
| Tandutinib | 29 | 3 | 1 | 0.0 | 0.0 | NA |
| Brivanib | 29 | 3 | 1 | 0.3 | 0.0 | NA |
| Hydroxyurea | 29 | 3 | 1 | 2.2 | 0.0 | NA |
| Dacarbazine | 30 | 3 | 1 | 0.1 | 0.0 | NA |
| Uramustine | 31 | 3 | 1 | 0.4 | 0.0 | NA |
| (-)JQ1 | 31 | 4 | 1 | 1.9 | 2.5 | NA |
| UNC0638 | 98 | 8 | 13 | 1.9 | 1.1 | 2.0 |
| AGI-5198 | 92 | 10 | 16 | 1.0 | 0.7 | 1.8 |
| Sonidegib | 27 | 2 | 1 | 1.7 | 0.2 | NA |
| Pilaralisib | 15 | 1 | 1 | 2.4 | NA | NA |
| Thio-TEPA | 29 | 3 | 1 | 0.3 | 0.4 | NA |

| | | | | | | |
|---|---|---|---|---|---|---|
| Fludarabine | 31 | 4 | 1 | 4.8 | 2.7 | NA |
| Vemurafenib | 102 | 9 | 15 | 4.8 | 4.1 | 3.8 |
| Enzalutamide | 29 | 4 | 0 | 0.9 | 0.5 | NA |
| AZD4547 | 24 | 2 | 1 | 0.0 | 0.0 | NA |
| Deferoxamine | 94 | 6 | 10 | 2.5 | 4.9 | 2.0 |
| Busulfan | 27 | 2 | 1 | 0.1 | 0.0 | NA |
| Iniparib | 99 | 8 | 14 | 3.6 | 2.0 | 1.3 |
| Imiquimod | 32 | 3 | 1 | 0.3 | 0.1 | NA |
| Disulfiram | 54 | 2 | 8 | 4.8 | 0.1 | 2.4 |
| Tamatinib | 100 | 8 | 14 | 2.9 | 2.4 | 2.3 |
| Vismodegib | 101 | 9 | 12 | 1.2 | 1.6 | 0.5 |
| MK-0752 | 46 | 7 | 5 | 0.6 | 1.1 | 0.2 |
| Ixazomib | 97 | 9 | 16 | 5.3 | 4.9 | 3.2 |
| Melphalan | 45 | 6 | 5 | 6.9 | 8.7 | 2.2 |
| Pemetrexed | 26 | 3 | 1 | 2.1 | 5.0 | NA |
| Imatinib | 28 | 3 | 1 | 1.0 | 0.9 | NA |
| Celecoxib | 30 | 4 | 1 | 0.8 | 0.5 | NA |
| Estramustine | 30 | 4 | 1 | 3.7 | 1.7 | NA |
| Filanesib | 93 | 10 | 14 | 2.4 | 0.8 | 1.7 |
| Regorafenib | 100 | 9 | 14 | 2.8 | 2.2 | 2.5 |
| Flutamide | 30 | 4 | 1 | 1.2 | 0.2 | NA |
| deltarasin | 16 | 4 | 4 | 0.9 | 1.2 | 0.8 |
| Letrozole | 28 | 2 | 1 | 0.0 | 0.0 | NA |
| Lomustine | 28 | 3 | 1 | 0.0 | 0.0 | NA |
| Streptozocin | 30 | 4 | 0 | 0.0 | 0.0 | NA |
| Carmustine | 27 | 4 | 1 | 0.0 | 0.0 | NA |
| IWP-L6 | 14 | 2 | 4 | 0.0 | 0.0 | 0.0 |

**Supplemental Table S3.** Correlation of *TP53* mutation frequency and drug sensitivity. Results from spearman correlation with compounds tested.

| Drug Name | SpearmanRho | P-value | N | maximum DSS | average DSS | DSS Standard Deviation |
|---|---|---|---|---|---|---|
| GSK-1904529A | 1.000 | 0.00000 | 4 | 23.1 | 20.3 | 3.7 |
| Masitinib | 1.000 | 0.00000 | 4 | 5.3 | 3.1 | 1.8 |
| OSI-027 | 1.000 | 0.00000 | 4 | 15.6 | 10.9 | 6.4 |
| Volasertib | 1.000 | 0.00000 | 4 | 15.7 | 7.5 | 5.9 |
| Bendamustine | 0.949 | 0.05132 | 4 | 2.0 | 0.9 | 1.0 |
| Dacomitinib | 0.949 | 0.05132 | 4 | 8.6 | 2.5 | 4.1 |
| Tacrolimus | 0.949 | 0.05132 | 4 | 6.1 | 2.5 | 3.0 |
| Deferoxamine | 0.880 | 0.02060 | 6 | 12.2 | 2.4 | 4.9 |
| Alisertib | 0.821 | 0.02345 | 7 | 5.9 | 2.5 | 2.2 |
| 2-methoxyestradiol | 0.800 | 0.20000 | 4 | 7.2 | 4.1 | 3.5 |
| ABT-751 | 0.800 | 0.20000 | 4 | 11.4 | 8.7 | 2.9 |
| AVN944 | 0.800 | 0.20000 | 4 | 17.6 | 7.2 | 7.5 |
| AZD1208 | 0.800 | 0.20000 | 4 | 6.8 | 5.0 | 1.3 |
| AZD1775 | 0.800 | 0.20000 | 4 | 18.5 | 8.9 | 7.7 |
| BIIB021 | 0.800 | 0.20000 | 4 | 21.1 | 14.0 | 9.6 |
| CUDC-101 | 0.800 | 0.20000 | 4 | 27.5 | 24.6 | 2.0 |
| CUDC-907 | 0.800 | 0.10409 | 5 | 45.7 | 42.7 | 2.4 |
| Chloroquine | 0.800 | 0.20000 | 4 | 12.6 | 9.8 | 2.7 |
| Docetaxel | 0.800 | 0.20000 | 4 | 21.4 | 9.2 | 10.6 |
| Gandotinib | 0.800 | 0.20000 | 4 | 11.0 | 7.8 | 2.9 |
| I-BET151 | 0.800 | 0.00963 | 9 | 18.9 | 11.8 | 4.8 |
| PF-00477736 | 0.800 | 0.20000 | 4 | 23.4 | 14.3 | 9.3 |
| Patupilone | 0.800 | 0.20000 | 4 | 13.8 | 6.3 | 5.7 |
| Ponatinib | 0.800 | 0.20000 | 4 | 6.8 | 2.8 | 3.0 |
| Pravastatin | 0.800 | 0.20000 | 4 | 3.0 | 0.9 | 1.4 |
| Sotrastaurin | 0.800 | 0.20000 | 4 | 5.1 | 2.8 | 2.3 |
| TAK-733 | 0.800 | 0.20000 | 4 | 24.8 | 9.5 | 11.3 |
| Fluorouracil | 0.775 | 0.22540 | 4 | 1.5 | 0.4 | 0.8 |
| PF-4800567 | 0.775 | 0.22540 | 4 | 1.2 | 0.3 | 0.6 |

| | | | | | | |
|---|---|---|---|---|---|---|
| Luminespib | 0.762 | 0.02800 | 8 | 26.0 | 20.3 | 8.5 |
| Pevonedistat | 0.762 | 0.01712 | 9 | 20.4 | 11.7 | 7.9 |
| (-)JQ1 | 0.738 | 0.26214 | 4 | 5.2 | 2.1 | 2.5 |
| Bimatoprost | 0.738 | 0.26214 | 4 | 0.9 | 0.5 | 0.5 |
| Nilotinib | 0.738 | 0.26214 | 4 | 8.0 | 3.4 | 4.1 |
| Vatalanib | 0.738 | 0.26214 | 4 | 3.9 | 1.8 | 2.0 |
| Veliparib | 0.714 | 0.04653 | 8 | 7.3 | 2.8 | 2.2 |
| Mocetinostat | 0.683 | 0.04244 | 9 | 19.6 | 14.7 | 3.8 |
| AZD7762 | 0.667 | 0.04987 | 9 | 12.8 | 5.1 | 4.4 |
| Regorafenib | 0.650 | 0.05807 | 9 | 6.2 | 3.4 | 2.2 |
| Vemurafenib | 0.644 | 0.06103 | 9 | 11.0 | 3.5 | 4.1 |
| Obatoclax | 0.635 | 0.09089 | 8 | 9.9 | 3.4 | 3.2 |
| Indibulin | 0.632 | 0.36754 | 4 | 15.6 | 6.4 | 7.8 |
| Lomeguatrib | 0.632 | 0.36754 | 4 | 1.9 | 0.5 | 0.9 |
| Tivantinib | 0.632 | 0.36754 | 4 | 2.2 | 1.1 | 1.2 |
| NVP-AEW541 | 0.624 | 0.05372 | 10 | 16.6 | 11.0 | 4.2 |
| Clofarabine | 0.622 | 0.01755 | 14 | 23.5 | 12.5 | 4.7 |
| SGC-CBP30 | 0.612 | 0.05997 | 10 | 19.9 | 11.1 | 5.6 |
| Cyclopamine | 0.600 | 0.40000 | 4 | 4.4 | 2.3 | 1.9 |
| NMS-873 | 0.600 | 0.28476 | 5 | 19.8 | 15.6 | 3.4 |
| Toremifene | 0.600 | 0.40000 | 4 | 3.8 | 2.8 | 0.9 |
| Cladribine | 0.594 | 0.09158 | 9 | 11.1 | 4.7 | 4.1 |
| JQ1 | 0.582 | 0.02885 | 14 | 28.6 | 17.3 | 7.1 |
| Ralimetinib | 0.543 | 0.26570 | 6 | 17.9 | 9.1 | 5.5 |
| Ixazomib | 0.533 | 0.13923 | 9 | 19.1 | 14.0 | 4.9 |
| Selumetinib | 0.533 | 0.13923 | 9 | 28.3 | 10.4 | 9.6 |
| Topotecan | 0.524 | 0.18272 | 8 | 25.2 | 16.6 | 6.1 |
| Idarubicin | 0.517 | 0.15439 | 9 | 26.1 | 17.7 | 4.8 |
| Mitoxantrone | 0.500 | 0.39100 | 5 | 8.6 | 4.6 | 3.7 |
| Azacitidine | 0.492 | 0.17893 | 9 | 5.7 | 2.4 | 2.2 |
| Stattic | 0.483 | 0.18747 | 9 | 23.3 | 16.6 | 5.7 |

| | | | | | | |
|---|---|---|---|---|---|---|
| Nutlin-3 | 0.464 | 0.29393 | 7 | 5.5 | 2.3 | 2.2 |
| PF 431396 | 0.452 | 0.26040 | 8 | 18.1 | 11.7 | 4.6 |
| PFI-1 | 0.452 | 0.26040 | 8 | 9.4 | 7.0 | 1.9 |
| Dactinomycin | 0.451 | 0.10593 | 14 | 33.2 | 25.7 | 6.3 |
| Refametinib | 0.446 | 0.10981 | 14 | 31.3 | 11.9 | 9.6 |
| Camptothecin | 0.429 | 0.39650 | 6 | 29.9 | 18.2 | 7.9 |
| Crenolanib | 0.429 | 0.39650 | 6 | 15.5 | 10.1 | 3.0 |
| Temsirolimus | 0.423 | 0.14975 | 13 | 20.3 | 9.8 | 5.7 |
| Methylprednisolone | 0.401 | 0.17434 | 13 | 18.9 | 7.0 | 7.2 |
| Ruxolitinib | 0.401 | 0.17436 | 13 | 22.6 | 11.6 | 5.2 |
| 8-amino-adenosine | 0.400 | 0.60000 | 4 | 25.5 | 19.4 | 7.3 |
| 8-chloro-adenosine | 0.400 | 0.60000 | 4 | 23.9 | 16.1 | 6.3 |
| AZD1480 | 0.400 | 0.60000 | 4 | 8.5 | 4.5 | 3.5 |
| Atorvastatin | 0.400 | 0.60000 | 4 | 11.7 | 7.0 | 5.0 |
| Bexarotene | 0.400 | 0.60000 | 4 | 5.8 | 3.0 | 2.3 |
| Bosutinib | 0.400 | 0.60000 | 4 | 12.1 | 8.2 | 4.1 |
| Canertinib | 0.400 | 0.60000 | 4 | 14.5 | 10.0 | 3.1 |
| Chlorambucil | 0.400 | 0.60000 | 4 | 3.7 | 1.9 | 2.0 |
| KU-60019 | 0.400 | 0.60000 | 4 | 5.8 | 3.8 | 1.6 |
| MK-2206 | 0.400 | 0.60000 | 4 | 12.6 | 8.6 | 2.9 |
| Mercaptopurine | 0.400 | 0.60000 | 4 | 4.9 | 1.9 | 2.1 |
| Paclitaxel | 0.400 | 0.60000 | 4 | 17.2 | 9.7 | 6.5 |
| Saracatinib | 0.400 | 0.60000 | 4 | 2.3 | 1.3 | 1.0 |
| Sepantronium bromide | 0.400 | 0.60000 | 4 | 42.5 | 37.8 | 4.7 |
| UCN-01 | 0.400 | 0.60000 | 4 | 37.9 | 28.1 | 12.0 |
| VX-11E | 0.400 | 0.60000 | 4 | 6.4 | 4.4 | 1.5 |
| Valproic acid | 0.400 | 0.60000 | 4 | 5.1 | 4.5 | 0.6 |
| deltarasin | 0.400 | 0.60000 | 4 | 7.5 | 5.7 | 1.2 |
| Pomalidomide | 0.394 | 0.26000 | 10 | 25.8 | 9.5 | 9.1 |
| Rocilinostat | 0.394 | 0.26000 | 10 | 17.1 | 9.0 | 3.5 |
| TPCA-1 | 0.394 | 0.26000 | 10 | 20.6 | 11.5 | 6.0 |

| | | | | | | |
|---|---|---|---|---|---|---|
| VER 155008 | 0.383 | 0.30850 | 9 | 7.8 | 3.6 | 2.7 |
| Tosedostat | 0.368 | 0.19568 | 14 | 13.0 | 6.2 | 3.4 |
| Panobinostat | 0.367 | 0.19674 | 14 | 31.2 | 22.8 | 4.8 |
| Danusertib | 0.357 | 0.38512 | 8 | 6.7 | 3.6 | 2.3 |
| Pentostatin | 0.347 | 0.24574 | 13 | 26.9 | 4.8 | 8.6 |
| C646 | 0.337 | 0.41379 | 8 | 9.3 | 5.2 | 3.3 |
| Buparlisib | 0.333 | 0.38071 | 9 | 14.7 | 10.8 | 3.4 |
| Oprozomib | 0.333 | 0.38071 | 9 | 31.3 | 24.3 | 6.1 |
| SCH772984 | 0.333 | 0.34659 | 10 | 30.8 | 10.4 | 9.2 |
| Sapanisertib | 0.333 | 0.38071 | 9 | 26.1 | 19.3 | 5.8 |
| Daporinad | 0.323 | 0.25987 | 14 | 36.1 | 17.6 | 9.2 |
| Geldanamycin | 0.321 | 0.48207 | 7 | 28.2 | 21.5 | 7.4 |
| Lapatinib | 0.316 | 0.68377 | 4 | 2.3 | 0.7 | 1.1 |
| TAK-580 | 0.316 | 0.68377 | 4 | 14.1 | 5.3 | 6.0 |
| Foretinib | 0.314 | 0.54409 | 6 | 9.8 | 5.9 | 3.5 |
| Apitolisib | 0.310 | 0.41748 | 9 | 24.5 | 18.0 | 5.6 |
| Venetoclax | 0.308 | 0.28398 | 14 | 27.8 | 12.3 | 7.4 |
| Trametinib | 0.301 | 0.34226 | 12 | 28.9 | 10.8 | 8.6 |
| Axitinib | 0.300 | 0.62384 | 5 | 4.5 | 1.5 | 1.9 |
| Idasanutlin | 0.300 | 0.62384 | 5 | 18.4 | 9.7 | 6.6 |
| APR-246 | 0.293 | 0.30941 | 14 | 20.8 | 9.7 | 6.7 |
| AT-406 | 0.286 | 0.53451 | 7 | 12.6 | 5.3 | 3.3 |
| UNC0638 | 0.286 | 0.49273 | 8 | 7.2 | 5.5 | 1.1 |
| ONX-0914 | 0.283 | 0.46003 | 9 | 33.4 | 26.7 | 4.6 |
| Vorinostat | 0.283 | 0.46003 | 9 | 15.7 | 11.0 | 3.7 |
| Omacetaxine | 0.277 | 0.33727 | 14 | 34.5 | 28.7 | 4.4 |
| Doxorubicin | 0.270 | 0.34992 | 14 | 20.7 | 13.5 | 4.1 |
| Pinometostat | 0.270 | 0.45114 | 10 | 0.2 | 0.0 | 0.1 |
| Flutamide | 0.258 | 0.74180 | 4 | 0.3 | 0.1 | 0.2 |
| Rebastinib | 0.258 | 0.74180 | 4 | 1.0 | 0.3 | 0.5 |
| Navitoclax | 0.253 | 0.40477 | 13 | 27.3 | 18.1 | 5.7 |

| | | | | | | |
|---|---|---|---|---|---|---|
| GSK-J4 | 0.248 | 0.39192 | 14 | 30.4 | 22.8 | 4.7 |
| Dasatinib | 0.238 | 0.57016 | 8 | 22.9 | 7.4 | 8.3 |
| Vincristine | 0.238 | 0.57016 | 8 | 25.0 | 16.9 | 5.4 |
| Alvespimycin | 0.236 | 0.48409 | 11 | 16.5 | 9.8 | 4.8 |
| Linsitinib | 0.231 | 0.47053 | 12 | 15.8 | 8.5 | 5.9 |
| Vistusertib | 0.222 | 0.44563 | 14 | 25.4 | 17.9 | 7.6 |
| Vismodegib | 0.219 | 0.57115 | 9 | 4.4 | 0.9 | 1.6 |
| (5Z)-7-Oxozeaenol | 0.217 | 0.57551 | 9 | 22.6 | 9.3 | 7.9 |
| SGI-1776 | 0.217 | 0.57551 | 9 | 10.3 | 7.8 | 2.3 |
| Dinaciclib | 0.214 | 0.61034 | 8 | 30.6 | 28.3 | 1.8 |
| Sirolimus | 0.211 | 0.78918 | 4 | 15.3 | 7.4 | 8.6 |
| Fedratinib | 0.200 | 0.74706 | 5 | 11.4 | 9.0 | 3.2 |
| Nintedanib | 0.200 | 0.80000 | 4 | 13.7 | 9.4 | 4.1 |
| TGX-221 | 0.200 | 0.80000 | 4 | 6.5 | 3.1 | 3.5 |
| Tubastatin A | 0.200 | 0.80000 | 4 | 8.1 | 3.7 | 3.9 |
| Pimasertib | 0.178 | 0.54215 | 14 | 32.1 | 12.1 | 10.0 |
| BMS-754807 | 0.165 | 0.59048 | 13 | 33.2 | 19.3 | 10.8 |
| Bryostatin 1 | 0.154 | 0.63249 | 12 | 24.1 | 9.7 | 7.3 |
| Mepacrine | 0.142 | 0.71503 | 9 | 22.5 | 14.8 | 8.6 |
| Bortezomib | 0.140 | 0.66352 | 12 | 33.3 | 29.1 | 3.3 |
| Quizartinib | 0.139 | 0.72084 | 9 | 6.8 | 1.7 | 2.4 |
| Carfilzomib | 0.138 | 0.63688 | 14 | 38.2 | 31.8 | 4.4 |
| Plicamycin | 0.134 | 0.64772 | 14 | 29.6 | 17.5 | 10.8 |
| Pictilisib | 0.132 | 0.66674 | 13 | 22.5 | 15.6 | 6.5 |
| AZD8055 | 0.132 | 0.66761 | 13 | 38.1 | 22.5 | 10.3 |
| PHA 408 | 0.128 | 0.72525 | 10 | 12.0 | 7.5 | 2.3 |
| BMS-911543 | 0.105 | 0.89459 | 4 | 2.0 | 0.9 | 1.1 |
| Seliciclib | 0.100 | 0.87289 | 5 | 4.0 | 2.5 | 1.4 |
| Doramapimod | 0.092 | 0.78848 | 11 | 14.1 | 6.1 | 5.3 |
| Vinblastine | 0.091 | 0.80277 | 10 | 22.0 | 9.9 | 6.0 |
| Ipatasertib | 0.090 | 0.75908 | 14 | 26.0 | 9.3 | 6.2 |

| | | | | | | |
|---|---|---|---|---|---|---|
| Filanesib | 0.084 | 0.81744 | 10 | 1.9 | 0.7 | 0.8 |
| Tubacin | 0.084 | 0.84355 | 8 | 14.7 | 7.3 | 6.3 |
| VGX-1027 | 0.079 | 0.86662 | 7 | 14.4 | 3.5 | 5.9 |
| Idelalisib | 0.073 | 0.80453 | 14 | 15.2 | 5.2 | 4.7 |
| GSK-1838705A | 0.071 | 0.87905 | 7 | 17.7 | 13.0 | 2.6 |
| Lestaurtinib | 0.071 | 0.81663 | 13 | 20.1 | 10.4 | 5.2 |
| Quisinostat | 0.068 | 0.81698 | 14 | 35.0 | 23.7 | 6.4 |
| Melphalan | 0.068 | 0.89874 | 6 | 17.1 | 5.6 | 8.7 |
| PS-1145 | 0.050 | 0.89835 | 9 | 4.5 | 2.1 | 2.0 |
| PF-04691502 | 0.038 | 0.90072 | 13 | 30.4 | 23.3 | 6.1 |
| Prednisolone | 0.036 | 0.93941 | 7 | 16.5 | 7.9 | 5.5 |
| Everolimus | 0.033 | 0.93216 | 9 | 19.1 | 7.8 | 6.5 |
| Entinostat | 0.017 | 0.96605 | 9 | 15.9 | 9.1 | 4.0 |
| Amonafide | 0.000 | 1.00000 | 4 | 6.6 | 4.7 | 1.4 |
| Sunitinib | 0.000 | 1.00000 | 6 | 2.2 | 1.0 | 0.9 |
| Tivozanib | 0.000 | 1.00000 | 4 | 5.0 | 3.3 | 1.8 |
| Dexamethasone | -0.004 | 0.99124 | 12 | 24.1 | 8.5 | 9.5 |
| Lenalidomide | -0.014 | 0.96559 | 12 | 22.5 | 7.8 | 6.8 |
| Alvocidib | -0.031 | 0.91675 | 14 | 26.0 | 21.8 | 3.7 |
| Tepotinib | -0.031 | 0.94159 | 8 | 3.8 | 0.6 | 1.3 |
| Auranofin | -0.035 | 0.90490 | 14 | 21.1 | 15.6 | 4.8 |
| Iniparib | -0.071 | 0.86653 | 8 | 6.1 | 2.6 | 2.0 |
| Dactolisib | -0.102 | 0.75343 | 12 | 25.2 | 17.9 | 4.1 |
| Afatinib | -0.105 | 0.89459 | 4 | 0.7 | 0.2 | 0.3 |
| Bicalutamide | -0.105 | 0.89459 | 4 | 0.9 | 0.3 | 0.4 |
| Thalidomide | -0.105 | 0.89459 | 4 | 2.6 | 0.8 | 1.2 |
| Tanespimycin | -0.112 | 0.72919 | 12 | 21.7 | 13.9 | 4.9 |
| PF-04708671 | -0.116 | 0.82685 | 6 | 6.3 | 2.8 | 2.1 |
| GSK269962 | -0.118 | 0.72869 | 11 | 9.2 | 4.1 | 2.7 |
| Dovitinib | -0.126 | 0.74658 | 9 | 1.4 | 0.5 | 0.4 |
| Tirbanibulin | -0.133 | 0.73237 | 9 | 23.4 | 18.3 | 5.2 |

| | | | | | | |
|---|---|---|---|---|---|---|
| Tamatinib | -0.140 | 0.74006 | 8 | 6.4 | 1.7 | 2.4 |
| PF-670462 | -0.146 | 0.72942 | 8 | 9.1 | 1.8 | 3.2 |
| Dabrafenib | -0.169 | 0.62028 | 11 | 7.9 | 3.0 | 2.8 |
| Omipalisib | -0.182 | 0.57170 | 12 | 31.3 | 24.6 | 4.7 |
| AZ191 | -0.200 | 0.80000 | 4 | 10.0 | 8.7 | 1.2 |
| Midostaurin | -0.200 | 0.60590 | 9 | 23.9 | 12.0 | 7.3 |
| Raloxifene | -0.200 | 0.80000 | 4 | 7.0 | 5.3 | 1.7 |
| MK-0752 | -0.204 | 0.66064 | 7 | 3.0 | 0.4 | 1.1 |
| Momelotinib | -0.214 | 0.61034 | 8 | 11.7 | 8.5 | 2.5 |
| Enzastaurin | -0.239 | 0.53562 | 9 | 1.3 | 0.4 | 0.6 |
| AGI-5198 | -0.240 | 0.50439 | 10 | 2.3 | 0.4 | 0.7 |
| Celecoxib | -0.258 | 0.74180 | 4 | 1.0 | 0.3 | 0.5 |
| Enzalutamide | -0.258 | 0.74180 | 4 | 1.0 | 0.3 | 0.5 |
| WEHI-539 | -0.298 | 0.40320 | 10 | 13.7 | 5.2 | 4.0 |
| RAF-265 | -0.299 | 0.51507 | 7 | 8.5 | 1.3 | 3.2 |
| Nelarabine | -0.316 | 0.68377 | 4 | 16.8 | 4.3 | 8.4 |
| AZ 3146 | -0.400 | 0.60000 | 4 | 5.6 | 5.2 | 0.4 |
| Fludarabine | -0.400 | 0.60000 | 4 | 7.1 | 3.2 | 2.7 |
| PF-3845 | -0.400 | 0.60000 | 4 | 5.1 | 4.4 | 0.6 |
| PIK-75 | -0.400 | 0.60000 | 4 | 30.4 | 29.8 | 0.6 |
| SNS-032 | -0.400 | 0.60000 | 4 | 40.0 | 34.9 | 3.7 |
| GSK343 | -0.600 | 0.40000 | 4 | 9.8 | 5.3 | 4.1 |
| Sorafenib | -0.612 | 0.14381 | 7 | 0.6 | 0.1 | 0.2 |
| Oxaliplatin | -0.632 | 0.36754 | 4 | 10.9 | 4.1 | 5.2 |
| Prednisone | -0.632 | 0.36754 | 4 | 7.7 | 2.0 | 3.8 |
| SGC0946 | -0.650 | 0.05807 | 9 | 7.1 | 3.1 | 1.8 |
| BX-912 | -0.700 | 0.18812 | 5 | 10.5 | 9.0 | 1.3 |
| Estramustine | -0.775 | 0.22540 | 4 | 3.3 | 0.8 | 1.7 |
| Procarbazine | -0.775 | 0.22540 | 4 | 0.2 | 0.1 | 0.1 |
| Vandetanib | -0.775 | 0.22540 | 4 | 1.0 | 0.3 | 0.5 |
| 4-hydroxytamoxifen | -0.800 | 0.20000 | 4 | 7.5 | 5.8 | 1.2 |

| | | | | | | |
|---|---|---|---|---|---|---|
| Daunorubicin | -0.800 | 0.20000 | 4 | 22.9 | 17.3 | 5.0 |
| Palbociclib | -0.800 | 0.20000 | 4 | 9.0 | 4.4 | 3.2 |
| UNC0642 | -0.800 | 0.20000 | 4 | 6.7 | 5.2 | 1.4 |
| Valrubicin | -0.800 | 0.20000 | 4 | 22.7 | 18.2 | 4.2 |
| Hydroxyfasudil | -0.821 | 0.08859 | 5 | 3.6 | 2.0 | 1.8 |
| Unknown [FIMM003726] | -0.900 | 0.03739 | 5 | 7.3 | 3.2 | 2.8 |
| Capecitabine | -0.949 | 0.05132 | 4 | 3.8 | 1.1 | 1.8 |
| Aminoglutethimide | -1.000 | 0.00000 | 4 | 2.1 | 0.7 | 0.9 |

**Supplemental Table S4.** Comparison of *ex vivo* drug sensitivity for small molecule inhibitors in MM patients with *TP53* and KRAS mutations compared to *TP53* mutation and WT *KRAS*. (A) p-values and median difference (B) number of samples and standard deviation of DSS score.

A.

| Drug Name | *P*-value (*TP53**KRAS* mutation - *TP53* mutation\\WT *KRAS*) | Median Difference (*TP53**KRAS* mutation - *TP53* mutation\\WT *KRAS*) | Functional Class | Mechanism Target |
|---|---|---|---|---|
| AZD7762 | 0.01587 | -6.75 | Kinase inhibitor; CHK inhibitor | Chk1 inhibitor |
| Stattic | 0.01587 | -7.7 | Other | STAT3 SH2 domain inhibitor |
| Azacitidine | 0.01794 | -4.25 | Antimetabolite | Pyrimidine analog, DNA methyl transferase inhibitor |
| Veliparib | 0.03571 | -2.2 | Differentiation/Epigenetic modifier | PARP inhibitor |
| Obatoclax | 0.03577 | -3.6 | Apoptotic modulator | Bcl-2 family inhibitor |
| Deferoxamine | 0.06360 | -1.8 | Other | Iron chelator |
| AGI-5198 | 0.07054 | 0.5 | Metabolic modifier | IDH1 R132H/R132C inhibitor |
| Apitolisib | 0.10986 | -3.7 | Kinase inhibitor | PI3K/mTOR inhibitor |
| Tubacin | 0.11021 | -11.4 | Differentiation/Epigenetic modifier | HDAC6 inhibitor |
| Vorinostat | 0.11111 | -3.6 | Differentiation/Epigenetic modifier | HDAC inhibitor |
| Topotecan | 0.11429 | -10.35 | Conventional Chemotherapeutic; Topoisomerase inhibitor | Topoisomerase I inhibitor |
| Dabrafenib | 0.11985 | 1.45 | Kinase inhibitor | B-Raf(V600E) inhibitor |
| Dactinomycin | 0.12821 | -6.7 | Conventional Chemotherapeutic | RNA and DNA synthesis inhibitor |
| Clofarabine | 0.12821 | -3.3 | Conventional Chemotherapeutic; Antimetabolite | Purine analog |
| Filanesib | 0.13933 | 1.25 | Kinesin inhibitor | KSP/Eg5 inhibitor |
| Pevonedistat | 0.17610 | -4.6 | Metabolic modifier | NAE inhibitor |
| PF-670462 | 0.18315 | 2.4 | Kinase inhibitor | CK1epsilon and CK1delta inhibitor |
| VER 155008 | 0.19048 | -3.4 | HSP inhibitor | HSP70 inhibitor |
| Buparlisib | 0.19048 | -1.4 | Kinase inhibitor; PI3K inhibitor | PI3K inhibitor, pan-class I |
| (5Z)-7-Oxozeaenol | 0.19048 | -9.8 | Kinase inhibitor | TAK1 inhibitor |
| Melphalan | 0.19671 | -16.5 | Conventional Chemotherapeutic; Alkylating agent | Nitrogen mustard alkylating agent |
| Bortezomib | 0.19861 | -1.35 | Proteasome inhibitor | Proteasome inhibitor (26S subunit) |
| Vincristine | 0.20000 | -3.5 | Conventional Chemotherapeutic | Mitotic inhibitor, Vinca alkaloid microtubule depolymerizer |
| Alvocidib | 0.22429 | 0.8 | Kinase inhibitor; CDK inhibitor | CDK inhibitor |
| Doramapimod | 0.23105 | 6.2 | Kinase inhibitor | p38MAPK inhibitor |
| Ipatasertib | 0.24963 | 4.2 | Kinase inhibitor; AKT inhibitor | AKT inhibitor |
| Vinblastine | 0.25714 | -4.6 | Conventional Chemotherapeutic | Mitotic inhibitor. Vinca alkaloid microtubule depolymerizer |
| Tosedostat | 0.27638 | -2.8 | Protease inhibitor | M1 family of aminopeptidase inhibitor (especially LTA4H and PSA) |
| Omacetaxine | 0.27691 | -0.8 | Conventional Chemotherapeutic | Protein synthesis inhib (80 S ribosome) |
| Auranofin | 0.27691 | 3.3 | Conventional Chemotherapeutic | Antirheumatic agent |
| I-BET151 | 0.28571 | -4.1 | Differentiation/Epigenetic modifier | BET family inhibitor |
| SCH772984 | 0.30952 | 5.4 | Kinase inhibitor | ERK1 & 2 inhibitor |
| Daporinad | 0.31760 | -7.3 | Metabolic modifier | NAMPT inhibitor |
| Luminespib | 0.34286 | -2.8 | HSP inhibitor | HSP90 inhibitor |
| Dinaciclib | 0.34286 | -1.7 | Kinase inhibitor; CDK inhibitor | CDK inhibitor |
| GSK269962 | 0.36022 | 1.95 | Kinase inhibitor | ROCK1 and ROCK2 inhibitor |
| BMS-754807 | 0.36597 | -7.1 | Kinase inhibitor | IGF1R inhibitor |
| Midostaurin | 0.38095 | 4.3 | Kinase inhibitor; Tyrosine kinase inhibitor | Broad TK (FLT3, KIT, RET, JAK, EGFR…) inhibitor |
| GSK-J4 | 0.38287 | -2.1 | Differentiation/Epigenetic modifier | JMJD3 (histone demethylase) inhibitor |
| Vemurafenib | 0.38928 | -2.5 | Kinase inhibitor | B-Raf(V600E) inhibitor |
| Pictilisib | 0.39006 | -2.65 | Kinase inhibitor | PI3K inhibitor, pan-class I |
| PHA 408 | 0.39233 | 0.9 | Kinase inhibitor | IKK-2 inhibitor |
| Omipalisib | 0.39394 | -0.95 | Kinase inhibitor | PI3K/mTOR inhibitor |
| AT-406 | 0.40000 | -1.05 | Apoptotic modulator | XIAP, cIAP1, cIAP2 inhibitor |
| Pimasertib | 0.40572 | 6 | Kinase inhibitor | MEK1/2 inhibitor |
| Venetoclax | 0.40572 | 1.8 | Apoptotic modulator | Bcl-2 inhibitor |
| Sapanisertib | 0.41270 | -1.5 | Kinase inhibitor | mTOR inhibitor |
| Tamatinib | 0.42398 | 1.2 | Kinase inhibitor | Syk inhibitor |
| Alvespimycin | 0.42857 | 3.5 | HSP inhibitor | HSP90 inhibitor |
| AZD8055 | 0.44522 | -3.35 | Kinase inhibitor; mTOR inhibitor | mTOR inhibitor |
| Momelotinib | 0.48571 | 1.8 | Kinase inhibitor; Tyrosine kinase inhibitor | JAK1 & 2 inhibitor |
| Quizartinib | 0.50018 | -1.05 | Kinase inhibitor; Tyrosine kinase inhibitor | FLT3 inhibitor |
| Vismodegib | 0.50233 | -1.55 | Hedgehog inhibitor | Smoothened (Hh) inhibitor |
| Lenalidomide | 0.53030 | 1.5 | Immunomodulator | Immunomodulatory |
| Temsirolimus | 0.53380 | -1.6 | Rapalog; mTOR inhibitor | mTORC1 inhibitor |
| Doxorubicin | 0.53497 | -2.5 | Conventional Chemotherapeutic; Topoisomerase inhibitor; Anthracycline | Topoisomerase II inhibitor |
| Mepacrine | 0.53859 | 8.85 | Other | Unclear. PLA2 inhibitor. NF-kB inhibitor, p53 activator |

| | | | | |
|---|---|---|---|---|
| Pomalidomide | 0.54762 | -2.1 | Immunomodulator | Immunomodulatory agent, anti-angiogenic |
| Selumetinib | 0.55556 | -7.35 | Kinase inhibitor; MEK inhibitor | MEK1/2 inhibitor |
| Entinostat | 0.55556 | 3.85 | Differentiation/Epigenetic modifier; HDAC inhibitor | HDAC inhibitor |
| Ixazomib | 0.55556 | -3.55 | Proteasome inhibitor | Proteasome inhibitor (20S subunit) |
| Sorafenib | 0.56370 | 0 | Kinase inhibitor; Tyrosine kinase inhibitor | B-Raf, FGFR-1, VEGFR-2 & -3, PDGFR-beta, KIT, and FLT3 inhib |
| MK-0752 | 0.56370 | 0 | Other | gamma-secretase/notch inhibitor |
| Methylprednisolone | 0.56502 | -6.7 | Immunomodulator | Glucocorticoid, immunomodulatory agent |
| Linsitinib | 0.58874 | -6.45 | Kinase inhibitor | IGF1R, IR inhibitor |
| WEHI-539 | 0.59291 | 2.1 | Apoptotic modulator | Bcl-XL inhibitor |
| Refametinib | 0.62005 | 5.3 | Kinase inhibitor | MEK1/2 inhibitor |
| Panobinostat | 0.62005 | -1.2 | Differentiation/Epigenetic modifier; HDAC inhibitor | HDAC inhibitor |
| Nutlin-3 | 0.62857 | 4.1 | Apoptotic modulator | MDM2 inhibitor |
| Dactolisib | 0.63036 | -3.35 | Kinase inhibitor; PI3K inhibitor | mTOR/(PI3K) inhibitor |
| Tanespimycin | 0.63889 | -0.3 | HSP inhibitor | HSP90 inhibitor |
| Enzastaurin | 0.66055 | 0.5 | Kinase inhibitor | PKCbeta inhibitor |
| Bryostatin 1 | 0.68422 | -3.4 | Kinase inhibitor | PKC activator |
| Iniparib | 0.68571 | 1.3 | Differentiation/Epigenetic modifier; PARP inhibitor | PARP inhibitor |
| PFI-1 | 0.68571 | -0.45 | Differentiation/Epigenetic modifier | BET family inhibitor |
| UNC0638 | 0.68571 | -0.2 | Differentiation/Epigenetic modifier | G9a/GLP inhibitor |
| Idelalisib | 0.70022 | -1 | Kinase inhibitor | PI3K inhibitor, p110Œ¥-selective |
| Dovitinib | 0.71099 | 0.2 | Kinase inhibitor; Tyrosine kinase inhibitor | FGFR inhibitor |
| Tirbanibulin | 0.71429 | -1 | Kinase inhibitor; Tyrosine kinase inhibitor | Src inhibitor |
| PS-1145 | 0.71429 | 0.9 | Kinase inhibitor | IKK-2 inhibitor |
| Everolimus | 0.73016 | -5.6 | Rapalog | binds FKBP12, causes inhibition of mTORC1 |
| SGC0946 | 0.73016 | 0.05 | Differentiation/Epigenetic modifier | DOT1L inhibitor |
| PF-04691502 | 0.73077 | -0.95 | Kinase inhibitor | PI3K/mTOR inhibitor |
| Dexamethasone | 0.74092 | 4.6 | Immunomodulator; Steroid | Glucocorticoid |
| APR-246 | 0.74886 | 2.8 | Apoptotic modulator | p53 activator, thioredoxin reductase 1 inhibitor |
| NVP-AEW541 | 0.76190 | -1.3 | Kinase inhibitor | IGF1R inhibitor |
| C646 | 0.77015 | -0.05 | Differentiation/Epigenetic modifier | p300/CREB-binding protein (CBP) inhibitor |
| PF 431396 | 0.78571 | -0.4 | Kinase inhibitor | FAK/PYK2 inhibitor |
| Quisinostat | 0.80478 | -1.4 | Differentiation/Epigenetic modifier | HDAC inhibitor |
| Navitoclax | 0.83566 | 2 | Apoptotic modulator | Bcl-2inhibitor, Bcl-XL inhibitor |
| Lestaurtinib | 0.83566 | -3.2 | Kinase inhibitor; Tyrosine kinase inhibitor | FLT3, JAK2, TrkA, TrkB, TrkC inhibitor |
| Geldanamycin | 0.85714 | 6.5 | HSP inhibitor | HSP90 inhibitor |
| Prednisolone | 0.85714 | 0.9 | Immunomodulator; Steroid | Glucocorticoid |
| Pentostatin | 0.88056 | -0.1 | Conventional Chemotherapeutic | Antimetabolite, Purine analog |
| Danusertib | 0.88571 | -0.7 | Kinase inhibitor | Aurora, Ret, TrkA, FGFR-1 inhibitor |
| Carfilzomib | 0.90152 | -0.6 | Proteasome inhibitor | Proteasome inhibitor (20S subunit) |
| Regorafenib | 0.90476 | -1.4 | Kinase inhibitor; Tyrosine kinase inhibitor | B-Raf, c-Kit, VEGFR2 inhibitor |
| Idarubicin | 0.90476 | -1.45 | Conventional Chemotherapeutic; Anthracycline; Topoisomerase inhibitor | Topoisomerase II inhibitor |
| Mocetinostat | 0.90476 | -2 | Differentiation/Epigenetic modifier; HDAC inhibitor | HDAC1 inhibitor, HDAC2 inhibitor |
| Oprozomib | 0.90476 | 3.15 | Proteasome inhibitor | Proteasome inhibitor (20S subunit) |
| ONX-0914 | 0.90476 | 2.45 | Proteasome inhibitor | LMP7 (immunoproteasome) |
| SGC-CBP30 | 0.91429 | 2.7 | Differentiation/Epigenetic modifier | CREBBP/EP300 bromodomain inhibitor |
| Rocilinostat | 0.91429 | -0.8 | Differentiation/Epigenetic modifier; HDAC inhibitor | HDAC-6 selective inhibitor |
| Trametinib | 0.93723 | -1.85 | Kinase inhibitor; MEK inhibitor | MEK1/2 inhibitor |
| Ruxolitinib | 0.94522 | 1.45 | Kinase inhibitor; Tyrosine kinase inhibitor | JAK1&2 inhibitor |
| Tepotinib | 1.00000 | 0 | Kinase inhibitor | c-Met inhibitor |
| Plicamycin | 1.00000 | 13.1 | Conventional Chemotherapeutic | RNA synthesis inhibitor |
| JQ1 | 1.00000 | -3.8 | Differentiation/Epigenetic modifier; BET inhibitor | BET family inhibitor |
| RAF-265 | 1.00000 | 0.15 | Kinase inhibitor; RAF inhibitor | C-Raf inhibitor, unclear MoA |
| Cladribine | 1.00000 | -0.9 | Conventional Chemotherapeutic | Antimetabolite, Purine analog |
| Vistusertib | 1.00000 | 0.4 | Kinase inhibitor | mTOR inhibitor, ATP-competitive |
| SGI-1776 | 1.00000 | 0.45 | Kinase inhibitor | PIM kinase inhibitor |
| TPCA-1 | 1.00000 | 0.75 | Kinase inhibitor | IKK-2 inhibitor |
| Pinometostat | 1.00000 | 0 | Differentiation/Epigenetic modifier | DOT1L inhibitor |
| Galunisertib | NaN | 0 | Kinase inhibitor | TGF-B/Smad inhibitor |
| Ibrutinib | NaN | 0 | Kinase inhibitor; Tyrosine kinase inhibitor | Btk inhibitor |

B.

| Drug Name | Number of samples screened | Number of samples screened with *TP53* mutation and WT *KRAS* | Number of samples screened with *TP53 and KRAS* mutations | Standard deviation of DSS from all samples screened | Standard deviation of DSS from samples screened containing *TP53* mutation and WT *KRAS* | Standard deviation of DSS from samples screened containing both *TP53* and *KRAS* mutation |
|---|---|---|---|---|---|---|
| AZD7762 | 103 | 4 | 5 | 3.4 | 3.2 | 1.8 |
| Stattic | 102 | 4 | 5 | 5.3 | 1.8 | 5.2 |
| Azacitidine | 98 | 4 | 5 | 3.0 | 1.0 | 1.3 |
| Veliparib | 104 | 3 | 5 | 1.9 | 2.2 | 1.0 |
| Obatoclax | 100 | 3 | 5 | 2.9 | 3.2 | 1.7 |
| Deferoxamine | 94 | 3 | 3 | 2.5 | 6.6 | 0.0 |
| AGI-5198 | 92 | 4 | 6 | 1.0 | 0.0 | 0.9 |
| Apitolisib | 100 | 4 | 5 | 7.2 | 3.0 | 6.0 |
| Tubacin | 102 | 4 | 4 | 3.2 | 5.8 | 4.1 |
| Vorinostat | 103 | 4 | 5 | 3.8 | 2.5 | 3.8 |
| Topotecan | 104 | 4 | 4 | 5.5 | 5.0 | 4.7 |
| Dabrafenib | 121 | 6 | 5 | 3.2 | 2.3 | 3.1 |
| Dactinomycin | 128 | 7 | 7 | 7.8 | 8.5 | 3.1 |
| Clofarabine | 126 | 7 | 7 | 7.1 | 4.8 | 4.2 |
| Filanesib | 93 | 4 | 6 | 2.4 | 0.3 | 0.9 |
| Pevonedistat | 100 | 4 | 5 | 8.0 | 6.1 | 8.5 |
| PF-670462 | 48 | 4 | 4 | 1.7 | 0.2 | 3.9 |
| VER 155008 | 101 | 4 | 5 | 2.6 | 1.9 | 2.9 |
| Buparlisib | 102 | 4 | 5 | 5.3 | 2.3 | 3.8 |
| (5Z)-7-Oxozeaenol | 106 | 4 | 5 | 5.8 | 9.4 | 4.3 |
| Melphalan | 45 | 3 | 3 | 6.9 | 9.7 | 0.0 |
| Bortezomib | 125 | 6 | 6 | 7.1 | 2.1 | 3.8 |
| Vincristine | 93 | 4 | 4 | 6.3 | 4.4 | 4.9 |
| Alvocidib | 128 | 7 | 7 | 6.2 | 4.3 | 2.9 |
| Doramapimod | 120 | 6 | 5 | 3.8 | 5.5 | 4.6 |
| Ipatasertib | 126 | 7 | 7 | 5.5 | 3.9 | 7.6 |
| Vinblastine | 106 | 6 | 4 | 4.4 | 7.2 | 2.2 |
| Tosedostat | 126 | 7 | 7 | 7.1 | 3.3 | 3.3 |
| Omacetaxine | 131 | 7 | 7 | 8.4 | 5.9 | 2.6 |

| | | | | | | |
|---|---|---|---|---|---|---|
| Auranofin | 128 | 7 | 7 | 4.6 | 5.2 | 4.3 |
| I-BET151 | 106 | 4 | 5 | 5.3 | 3.8 | 4.8 |
| SCH772984 | 109 | 5 | 5 | 7.4 | 4.3 | 11.6 |
| Daporinad | 125 | 7 | 7 | 9.2 | 8.5 | 10.3 |
| Luminespib | 93 | 4 | 4 | 9.5 | 1.9 | 11.5 |
| Dinaciclib | 99 | 4 | 4 | 7.5 | 1.2 | 2.1 |
| GSK269962 | 121 | 6 | 5 | 3.1 | 2.8 | 2.4 |
| BMS-754807 | 128 | 7 | 6 | 9.4 | 11.2 | 10.7 |
| Midostaurin | 111 | 6 | 3 | 6.6 | 7.2 | 7.6 |
| GSK-J4 | 135 | 7 | 7 | 4.4 | 6.0 | 3.3 |
| Vemurafenib | 102 | 5 | 4 | 4.8 | 4.1 | 4.5 |
| Pictilisib | 123 | 7 | 6 | 6.9 | 7.6 | 5.5 |
| PHA 408 | 95 | 4 | 6 | 2.3 | 1.4 | 2.9 |
| Omipalisib | 129 | 6 | 6 | 9.5 | 3.7 | 5.5 |
| AT-406 | 102 | 3 | 4 | 4.4 | 0.8 | 4.6 |
| Pimasertib | 118 | 7 | 7 | 8.0 | 9.7 | 10.6 |
| Venetoclax | 127 | 7 | 7 | 10.0 | 8.7 | 6.5 |
| Sapanisertib | 98 | 4 | 5 | 7.3 | 4.2 | 6.9 |
| Tamatinib | 100 | 3 | 5 | 2.9 | 0.7 | 2.9 |
| Alvespimycin | 115 | 6 | 5 | 5.3 | 4.6 | 5.1 |
| AZD8055 | 123 | 7 | 6 | 9.3 | 10.4 | 10.6 |
| Momelotinib | 98 | 4 | 4 | 3.9 | 3.4 | 1.0 |
| Quizartinib | 98 | 6 | 3 | 4.2 | 2.8 | 1.2 |
| Vismodegib | 101 | 4 | 5 | 1.2 | 2.2 | 0.3 |
| Lenalidomide | 120 | 7 | 5 | 6.3 | 5.4 | 8.5 |
| Temsirolimus | 112 | 6 | 7 | 5.3 | 5.6 | 6.1 |
| Doxorubicin | 127 | 7 | 7 | 4.3 | 4.9 | 3.6 |
| Mepacrine | 103 | 4 | 5 | 6.7 | 11.1 | 2.3 |
| Pomalidomide | 105 | 5 | 5 | 7.1 | 7.4 | 11.2 |
| Selumetinib | 97 | 4 | 5 | 8.2 | 10.9 | 8.9 |
| Entinostat | 100 | 4 | 5 | 3.6 | 4.0 | 4.2 |
| Ixazomib | 97 | 4 | 5 | 5.3 | 2.9 | 6.1 |
| Sorafenib | 83 | 3 | 4 | 4.4 | 0.0 | 0.3 |
| MK-0752 | 46 | 3 | 4 | 0.6 | 0.0 | 1.5 |
| Methylprednisolone | 121 | 6 | 7 | 6.4 | 7.1 | 7.8 |

| | | | | | | |
|---|---|---|---|---|---|---|
| Linsitinib | 123 | 6 | 6 | 4.2 | 6.1 | 5.9 |
| WEHI-539 | 98 | 4 | 6 | 6.2 | 4.3 | 3.8 |
| Refametinib | 127 | 7 | 7 | 8.3 | 8.8 | 10.9 |
| Panobinostat | 128 | 7 | 7 | 6.5 | 5.9 | 3.9 |
| Nutlin-3 | 104 | 4 | 3 | 3.2 | 0.7 | 3.0 |
| Dactolisib | 120 | 6 | 6 | 6.5 | 3.9 | 4.5 |
| Tanespimycin | 122 | 7 | 5 | 6.4 | 5.5 | 4.1 |
| Enzastaurin | 114 | 5 | 4 | 2.9 | 0.6 | 0.6 |
| Bryostatin 1 | 109 | 7 | 5 | 8.3 | 8.3 | 6.2 |
| Iniparib | 99 | 4 | 4 | 3.6 | 2.8 | 1.2 |
| PFI-1 | 95 | 4 | 4 | 3.1 | 1.9 | 2.2 |
| UNC0638 | 98 | 4 | 4 | 1.9 | 0.9 | 1.2 |
| Idelalisib | 123 | 7 | 7 | 5.1 | 3.4 | 6.1 |
| Dovitinib | 96 | 4 | 5 | 1.3 | 0.3 | 0.5 |
| Tirbanibulin | 116 | 6 | 3 | 6.3 | 5.8 | 4.9 |
| PS-1145 | 95 | 3 | 6 | 2.2 | 1.6 | 2.2 |
| Everolimus | 98 | 4 | 5 | 4.9 | 8.5 | 2.8 |
| SGC0946 | 96 | 4 | 5 | 2.4 | 2.6 | 1.1 |
| PF-04691502 | 130 | 7 | 6 | 8.0 | 7.3 | 5.1 |
| Dexamethasone | 123 | 7 | 5 | 8.3 | 8.2 | 11.6 |
| APR-246 | 130 | 7 | 7 | 5.7 | 7.0 | 6.9 |
| NVP-AEW541 | 97 | 4 | 6 | 5.1 | 4.9 | 4.3 |
| C646 | 102 | 4 | 4 | 3.0 | 4.1 | 3.0 |
| PF 431396 | 96 | 3 | 5 | 4.9 | 4.7 | 5.0 |
| Quisinostat | 131 | 7 | 7 | 7.4 | 7.4 | 5.6 |
| Navitoclax | 131 | 6 | 7 | 8.4 | 6.5 | 5.5 |
| Lestaurtinib | 125 | 6 | 7 | 4.9 | 5.8 | 5.1 |
| Geldanamycin | 44 | 4 | 3 | 6.9 | 6.6 | 10.0 |
| Prednisolone | 96 | 3 | 4 | 5.3 | 5.0 | 6.5 |
| Pentostatin | 121 | 6 | 7 | 4.5 | 11.6 | 4.9 |
| Danusertib | 103 | 4 | 4 | 3.1 | 2.2 | 2.8 |
| Carfilzomib | 133 | 7 | 7 | 7.1 | 4.4 | 4.6 |
| Regorafenib | 100 | 4 | 5 | 2.8 | 2.7 | 2.1 |
| Idarubicin | 105 | 4 | 5 | 5.7 | 7.0 | 3.0 |
| Mocetinostat | 102 | 4 | 5 | 5.3 | 4.9 | 3.0 |

| | | | | | | |
|---|---|---|---|---|---|---|
| Oprozomib | 101 | 4 | 5 | 7.5 | 8.1 | 4.6 |
| ONX-0914 | 97 | 4 | 5 | 7.2 | 5.4 | 4.6 |
| SGC-CBP30 | 96 | 4 | 6 | 4.4 | 3.2 | 6.9 |
| Rocilinostat | 99 | 4 | 6 | 3.2 | 2.4 | 4.2 |
| Trametinib | 119 | 6 | 6 | 6.9 | 7.0 | 10.6 |
| Ruxolitinib | 123 | 6 | 7 | 5.8 | 5.4 | 5.4 |
| Tepotinib | 100 | 4 | 4 | 3.0 | 1.9 | 0.5 |
| Plicamycin | 127 | 7 | 7 | 10.4 | 12.8 | 8.8 |
| JQ1 | 130 | 7 | 7 | 6.7 | 5.9 | 8.7 |
| RAF-265 | 93 | 3 | 4 | 3.0 | 4.9 | 0.2 |
| Cladribine | 108 | 6 | 3 | 4.9 | 3.8 | 5.5 |
| Vistusertib | 123 | 7 | 7 | 6.9 | 7.6 | 8.1 |
| SGI-1776 | 93 | 3 | 6 | 2.1 | 3.0 | 2.1 |
| TPCA-1 | 98 | 4 | 6 | 5.1 | 6.0 | 6.6 |
| Pinometostat | 94 | 4 | 6 | 1.3 | 0.1 | 0.1 |
| Galunisertib | 87 | 4 | 5 | 0.3 | 0.0 | 0.0 |
| Ibrutinib | 108 | 5 | 5 | 1.0 | 0.0 | 0.0 |

**Supplemental Table S5.** Differentially expressed genes from the comparison between samples with *TP53* mutation compared to WT *TP53 from the FIMM cohort* .

| ENSEMBL_ID | logFC | AveExpr | t | P.Value | P.adjust | B | External gene name | Description |
|---|---|---|---|---|---|---|---|---|
| ENSG00000009694 | 2.747 | 1.142 | 5.232 | 0.00000 | 0.00812 | 4.937 | TENM1 | teneurin transmembrane protein 1 [Source:HGNC Symbol;Acc:HGNC:8117] |
| ENSG00000165300 | 3.478 | 0.572 | 5.185 | 0.00000 | 0.00812 | 4.627 | SLITRK5 | SLIT and NTRK like family member 5 [Source:HGNC Symbol;Acc:HGNC:20295] |
| ENSG00000141504 | -0.803 | 4.509 | -4.814 | 0.00001 | 0.02019 | 3.722 | SAT2 | spermidine/spermine N1-acetyltransferase family member 2 [Source:HGNC Symbol;Acc:HGNC:23160] |
| ENSG00000174282 | -1.141 | 5.255 | -4.694 | 0.00001 | 0.02019 | 3.325 | ZBTB4 | zinc finger and BTB domain containing 4 [Source:HGNC Symbol;Acc:HGNC:23847] |
| ENSG00000136531 | 1.961 | 2.545 | 4.682 | 0.00001 | 0.02019 | 3.230 | SCN2A | sodium voltage-gated channel alpha subunit 2 [Source:HGNC Symbol;Acc:HGNC:10588] |
| ENSG00000154839 | 1.560 | 2.192 | 4.671 | 0.00001 | 0.02019 | 3.130 | SKA1 | spindle and kinetochore associated complex subunit 1 [Source:HGNC Symbol;Acc:HGNC:28109] |
| ENSG00000166579 | -0.783 | 6.462 | -4.649 | 0.00001 | 0.02019 | 3.181 | NDEL1 | nudE neurodevelopment protein 1 like 1 [Source:HGNC Symbol;Acc:HGNC:17620] |
| ENSG00000173207 | 1.040 | 3.009 | 4.477 | 0.00002 | 0.03236 | 2.529 | CKS1B | CDC28 protein kinase regulatory subunit 1B [Source:HGNC Symbol;Acc:HGNC:19083] |
| ENSG00000115163 | 1.296 | 0.869 | 4.463 | 0.00002 | 0.03236 | 2.004 | CENPA | centromere protein A [Source:HGNC Symbol;Acc:HGNC:1851] |
| ENSG00000273983 | 1.478 | 2.689 | 4.403 | 0.00003 | 0.03670 | 2.273 | HIST1H3G | histone cluster 1 H3 family member g [Source:HGNC Symbol;Acc:HGNC:4772] |
| ENSG00000129810 | 1.192 | 2.066 | 4.344 | 0.00004 | 0.04168 | 1.990 | SGO1 | shugoshin 1 [Source:HGNC Symbol;Acc:HGNC:25088] |
| ENSG00000162073 | 1.245 | 1.404 | 4.302 | 0.00004 | 0.04372 | 1.715 | PAQR4 | progestin and adipoQ receptor family member 4 [Source:HGNC Symbol;Acc:HGNC:26386] |
| ENSG00000177606 | -1.301 | 10.696 | -4.279 | 0.00005 | 0.04372 | 1.881 | JUN | Jun proto-oncogene, AP-1 transcription factor subunit [Source:HGNC Symbol;Acc:HGNC:6204] |
| ENSG00000166851 | 1.477 | 1.029 | 4.267 | 0.00005 | 0.04372 | 1.524 | PLK1 | polo like kinase 1 [Source:HGNC Symbol;Acc:HGNC:9077] |
| ENSG00000100526 | 1.172 | 2.947 | 4.217 | 0.00006 | 0.04921 | 1.664 | CDKN3 | cyclin dependent kinase inhibitor 3 [Source:HGNC Symbol;Acc:HGNC:1791] |
| ENSG00000164611 | 1.157 | 3.670 | 4.194 | 0.00006 | 0.05016 | 1.606 | PTTG1 | pituitary tumor-transforming 1 [Source:HGNC Symbol;Acc:HGNC:9690] |
| ENSG00000143228 | 1.492 | 3.318 | 4.174 | 0.00007 | 0.05082 | 1.539 | NUF2 | NUF2, NDC80 kinetochore complex component [Source:HGNC Symbol;Acc:HGNC:14621] |
| ENSG00000157456 | 1.430 | 2.862 | 4.141 | 0.00008 | 0.05432 | 1.420 | CCNB2 | cyclin B2 [Source:HGNC Symbol;Acc:HGNC:1580] |
| ENSG00000127564 | 1.187 | 1.520 | 4.076 | 0.00010 | 0.05950 | 1.055 | PKMYT1 | protein kinase, membrane associated tyrosine/threonine 1 [Source:HGNC Symbol;Acc:HGNC:29650] |
| ENSG00000112984 | 1.498 | 2.136 | 4.047 | 0.00011 | 0.05950 | 1.080 | KIF20A | kinesin family member 20A [Source:HGNC Symbol;Acc:HGNC:9787] |
| ENSG00000185480 | 1.066 | 3.383 | 4.038 | 0.00011 | 0.05950 | 1.094 | PARPBP | PARP1 binding protein [Source:HGNC Symbol;Acc:HGNC:26074] |
| ENSG00000183395 | 1.086 | 1.363 | 4.037 | 0.00011 | 0.05950 | 0.868 | PMCH | pro-melanin concentrating hormone [Source:HGNC Symbol;Acc:HGNC:9109] |
| ENSG00000080986 | 1.210 | 3.677 | 4.029 | 0.00012 | 0.05950 | 1.065 | NDC80 | NDC80, kinetochore complex component [Source:HGNC Symbol;Acc:HGNC:16909] |
| ENSG00000133328 | 2.359 | 1.448 | 4.027 | 0.00012 | 0.05950 | 0.981 | HRASLS2 | HRAS like suppressor 2 [Source:HGNC Symbol;Acc:HGNC:17824] |
| ENSG00000105254 | 0.595 | 5.108 | 4.025 | 0.00012 | 0.05950 | 1.044 | TBCB | tubulin folding cofactor B [Source:HGNC Symbol;Acc:HGNC:1989] |
| ENSG00000203760 | 1.237 | 1.755 | 4.013 | 0.00012 | 0.05995 | 0.906 | CENPW | centromere protein W [Source:HGNC Symbol;Acc:HGNC:21488] |
| ENSG00000144395 | 1.296 | 1.612 | 3.935 | 0.00016 | 0.07543 | 0.664 | CCDC150 | coiled-coil domain containing 150 [Source:HGNC Symbol;Acc:HGNC:26834] |
| ENSG00000104889 | 0.889 | 3.308 | 3.892 | 0.00019 | 0.07543 | 0.633 | RNASEH2A | ribonuclease H2 subunit A [Source:HGNC Symbol;Acc:HGNC:18518] |
| ENSG00000140525 | 0.908 | 5.271 | 3.892 | 0.00019 | 0.07543 | 0.610 | FANCI | Fanconi anemia complementation group I [Source:HGNC Symbol;Acc:HGNC:25568] |
| ENSG00000071539 | 1.228 | 2.326 | 3.891 | 0.00019 | 0.07543 | 0.606 | TRIP13 | thyroid hormone receptor interactor 13 [Source:HGNC Symbol;Acc:HGNC:12307] |
| ENSG00000117650 | 1.559 | 1.938 | 3.890 | 0.00019 | 0.07543 | 0.583 | NEK2 | NIMA related kinase 2 [Source:HGNC Symbol;Acc:HGNC:7745] |
| ENSG00000090889 | 1.319 | 2.483 | 3.882 | 0.00019 | 0.07543 | 0.591 | KIF4A | kinesin family member 4A [Source:HGNC Symbol;Acc:HGNC:13339] |
| ENSG00000166845 | 1.096 | 1.250 | 3.867 | 0.00020 | 0.07543 | 0.370 | C18orf54 | chromosome 18 open reading frame 54 [Source:HGNC Symbol;Acc:HGNC:13796] |
| ENSG00000089169 | 2.784 | 0.390 | 3.860 | 0.00021 | 0.07543 | 0.328 | RPH3A | rabphilin 3A [Source:HGNC Symbol;Acc:HGNC:17056] |
| ENSG00000091490 | -1.574 | 8.291 | -3.859 | 0.00021 | 0.07543 | 0.511 | SEL1L3 | SEL1L family member 3 [Source:HGNC Symbol;Acc:HGNC:29108] |
| ENSG00000126787 | 1.389 | 3.395 | 3.857 | 0.00021 | 0.07543 | 0.520 | DLGAP5 | DLG associated protein 5 [Source:HGNC Symbol;Acc:HGNC:16864] |
| ENSG00000104381 | 0.965 | 2.592 | 3.827 | 0.00024 | 0.08032 | 0.417 | GDAP1 | ganglioside induced differentiation associated protein 1 [Source:HGNC Symbol;Acc:HGNC:15968] |
| ENSG00000183354 | -0.479 | 6.628 | -3.818 | 0.00024 | 0.08032 | 0.375 | KIAA2026 | KIAA2026 [Source:HGNC Symbol;Acc:HGNC:23378] |
| ENSG00000114405 | 1.479 | 0.914 | 3.816 | 0.00025 | 0.08032 | 0.179 | C3orf14 | chromosome 3 open reading frame 14 [Source:HGNC Symbol;Acc:HGNC:25024] |
| ENSG00000029153 | 1.722 | 2.129 | 3.772 | 0.00029 | 0.08928 | 0.250 | ARNTL2 | aryl hydrocarbon receptor nuclear translocator like 2 [Source:HGNC Symbol;Acc:HGNC:18984] |
| ENSG00000124802 | 0.784 | 4.065 | 3.771 | 0.00029 | 0.08928 | 0.249 | EEF1E1 | eukaryotic translation elongation factor 1 epsilon 1 [Source:HGNC Symbol;Acc:HGNC:3212] |
| ENSG00000168078 | 1.340 | 2.009 | 3.752 | 0.00031 | 0.09305 | 0.173 | PBK | PDZ binding kinase [Source:HGNC Symbol;Acc:HGNC:18282] |
| ENSG00000105011 | 0.961 | 2.014 | 3.740 | 0.00032 | 0.09482 | 0.120 | ASF1B | anti-silencing function 1B histone chaperone [Source:HGNC Symbol;Acc:HGNC:20996] |
| ENSG00000089685 | 1.336 | 2.988 | 3.721 | 0.00034 | 0.09886 | 0.109 | BIRC5 | baculoviral IAP repeat containing 5 [Source:HGNC Symbol;Acc:HGNC:593] |
| ENSG00000136982 | 1.232 | 1.791 | 3.698 | 0.00037 | 0.10173 | -0.005 | DSCC1 | DNA replication and sister chromatid cohesion 1 [Source:HGNC Symbol;Acc:HGNC:24453] |
| ENSG00000139734 | 1.054 | 2.587 | 3.695 | 0.00037 | 0.10173 | 0.027 | DIAPH3 | diaphanous related formin 3 [Source:HGNC Symbol;Acc:HGNC:15480] |
| ENSG00000135451 | 1.677 | 0.922 | 3.694 | 0.00037 | 0.10173 | -0.118 | TROAP | trophinin associated protein [Source:HGNC Symbol;Acc:HGNC:12327] |
| ENSG00000124762 | -1.165 | 7.362 | -3.671 | 0.00040 | 0.10462 | -0.079 | CDKN1A | cyclin dependent kinase inhibitor 1A [Source:HGNC Symbol;Acc:HGNC:1784] |
| ENSG00000140534 | 1.280 | 1.217 | 3.662 | 0.00042 | 0.10462 | -0.178 | TICRR | TOPBP1 interacting checkpoint and replication regulator [Source:HGNC Symbol;Acc:HGNC:28704] |
| ENSG00000165501 | 0.592 | 3.737 | 3.656 | 0.00043 | 0.10462 | -0.091 | LRR1 | leucine rich repeat protein 1 [Source:HGNC Symbol;Acc:HGNC:19742] |
| ENSG00000175063 | 1.468 | 0.888 | 3.646 | 0.00044 | 0.10462 | -0.273 | UBE2C | ubiquitin conjugating enzyme E2 C [Source:HGNC Symbol;Acc:HGNC:15937] |
| ENSG00000163808 | 1.154 | 3.932 | 3.641 | 0.00045 | 0.10462 | -0.148 | KIF15 | kinesin family member 15 [Source:HGNC Symbol;Acc:HGNC:17273] |
| ENSG00000170312 | 1.329 | 3.198 | 3.641 | 0.00045 | 0.10462 | -0.134 | CDK1 | cyclin dependent kinase 1 [Source:HGNC Symbol;Acc:HGNC:1722] |
| ENSG00000183763 | 1.104 | 1.521 | 3.631 | 0.00046 | 0.10462 | -0.244 | TRAIP | TRAF interacting protein [Source:HGNC Symbol;Acc:HGNC:30764] |
| ENSG00000138640 | -1.220 | 6.539 | -3.629 | 0.00047 | 0.10462 | -0.204 | FAM13A | family with sequence similarity 13 member A [Source:HGNC Symbol;Acc:HGNC:19367] |
| ENSG00000104147 | 1.090 | 1.117 | 3.629 | 0.00047 | 0.10462 | -0.321 | OIP5 | Opa interacting protein 5 [Source:HGNC Symbol;Acc:HGNC:20300] |
| ENSG00000184661 | 1.281 | 2.181 | 3.623 | 0.00048 | 0.10462 | -0.192 | CDCA2 | cell division cycle associated 2 [Source:HGNC Symbol;Acc:HGNC:14623] |
| ENSG00000165304 | 1.105 | 2.923 | 3.620 | 0.00048 | 0.10462 | -0.191 | MELK | maternal embryonic leucine zipper kinase [Source:HGNC Symbol;Acc:HGNC:16870] |
| ENSG00000114346 | 0.899 | 4.449 | 3.619 | 0.00048 | 0.10462 | -0.226 | ECT2 | epithelial cell transforming 2 [Source:HGNC Symbol;Acc:HGNC:3155] |
| ENSG00000137812 | 1.091 | 5.198 | 3.612 | 0.00049 | 0.10517 | -0.258 | KNL1 | kinetochore scaffold 1 [Source:HGNC Symbol;Acc:HGNC:24054] |
| ENSG00000146670 | 1.217 | 2.131 | 3.593 | 0.00053 | 0.11033 | -0.283 | CDCA5 | cell division cycle associated 5 [Source:HGNC Symbol;Acc:HGNC:14626] |
| ENSG00000064787 | 1.764 | 2.354 | 3.579 | 0.00055 | 0.11286 | -0.312 | BCAS1 | breast carcinoma amplified sequence 1 [Source:HGNC Symbol;Acc:HGNC:974] |
| ENSG00000138180 | 1.242 | 2.885 | 3.576 | 0.00056 | 0.11286 | -0.320 | CEP55 | centrosomal protein 55 [Source:HGNC Symbol;Acc:HGNC:1161] |
| ENSG00000186193 | 1.306 | 1.816 | 3.570 | 0.00057 | 0.11286 | -0.361 | SAPCD2 | suppressor APC domain containing 2 [Source:HGNC Symbol;Acc:HGNC:28055] |
| ENSG00000112742 | 1.190 | 3.342 | 3.564 | 0.00058 | 0.11286 | -0.363 | TTK | TTK protein kinase [Source:HGNC Symbol;Acc:HGNC:12401] |
| ENSG00000142961 | -0.805 | 4.595 | -3.562 | 0.00058 | 0.11286 | -0.368 | MOB3C | MOB kinase activator 3C [Source:HGNC Symbol;Acc:HGNC:29800] |
| ENSG00000104490 | 1.707 | 1.288 | 3.555 | 0.00060 | 0.11286 | -0.432 | NCALD | neurocalcin delta [Source:HGNC Symbol;Acc:HGNC:7655] |
| ENSG00000213186 | 0.609 | 5.537 | 3.552 | 0.00060 | 0.11286 | -0.439 | TRIM59 | tripartite motif containing 59 [Source:HGNC Symbol;Acc:HGNC:30834] |
| ENSG00000104872 | 0.450 | 5.455 | 3.549 | 0.00061 | 0.11286 | -0.446 | PIH1D1 | PIH1 domain containing 1 [Source:HGNC Symbol;Acc:HGNC:26075] |
| ENSG00000134057 | 1.128 | 3.874 | 3.535 | 0.00064 | 0.11427 | -0.464 | CCNB1 | cyclin B1 [Source:HGNC Symbol;Acc:HGNC:1579] |
| ENSG00000102384 | 1.110 | 1.374 | 3.530 | 0.00065 | 0.11427 | -0.533 | CENPI | centromere protein I [Source:HGNC Symbol;Acc:HGNC:3968] |
| ENSG00000142875 | -1.113 | 6.119 | -3.529 | 0.00065 | 0.11427 | -0.499 | PRKACB | protein kinase cAMP-activated catalytic subunit beta [Source:HGNC Symbol;Acc:HGNC:9381] |
| ENSG00000156970 | 1.146 | 4.192 | 3.529 | 0.00065 | 0.11427 | -0.491 | BUB1B | BUB1 mitotic checkpoint serine/threonine kinase B [Source:HGNC Symbol;Acc:HGNC:1149] |
| ENSG00000169684 | 1.413 | 0.850 | 3.525 | 0.00066 | 0.11427 | -0.634 | CHRNA5 | cholinergic receptor nicotinic alpha 5 subunit [Source:HGNC Symbol;Acc:HGNC:1959] |
| ENSG00000170627 | 2.572 | 0.373 | 3.520 | 0.00067 | 0.11452 | -0.598 | GTSF1 | gametocyte specific factor 1 [Source:HGNC Symbol;Acc:HGNC:26565] |
| ENSG00000068489 | 1.184 | 2.033 | 3.509 | 0.00070 | 0.11560 | -0.521 | PRR11 | proline rich 11 [Source:HGNC Symbol;Acc:HGNC:25619] |
| ENSG00000152253 | 1.332 | 1.065 | 3.509 | 0.00070 | 0.11560 | -0.614 | SPC25 | SPC25, NDC80 kinetochore complex component [Source:HGNC Symbol;Acc:HGNC:24031] |
| ENSG00000079462 | 0.766 | 1.932 | 3.505 | 0.00071 | 0.11560 | -0.556 | PAFAH1B3 | platelet activating factor acetylhydrolase 1b catalytic subunit 3 [Source:HGNC Symbol;Acc:HGNC:8576] |
| ENSG00000184708 | -0.508 | 4.277 | -3.495 | 0.00073 | 0.11698 | -0.560 | EIF4ENIF1 | eukaryotic translation initiation factor 4E nuclear import factor 1 [Source:HGNC Symbol;Acc:HGNC:16687] |
| ENSG00000051341 | 1.139 | 4.175 | 3.494 | 0.00073 | 0.11698 | -0.592 | POLQ | DNA polymerase theta [Source:HGNC Symbol;Acc:HGNC:9186] |
| ENSG00000163781 | 0.579 | 6.590 | 3.489 | 0.00074 | 0.11738 | -0.625 | TOPBP1 | topoisomerase (DNA) II binding protein 1 [Source:HGNC Symbol;Acc:HGNC:17008] |
| ENSG00000196636 | 0.893 | 2.927 | 3.477 | 0.00077 | 0.11951 | -0.603 | SDHAF3 | succinate dehydrogenase complex assembly factor 3 [Source:HGNC Symbol;Acc:HGNC:21752] |
| ENSG00000118193 | 1.297 | 3.611 | 3.476 | 0.00078 | 0.11951 | -0.629 | KIF14 | kinesin family member 14 [Source:HGNC Symbol;Acc:HGNC:19181] |
| ENSG00000129245 | -0.492 | 4.878 | -3.443 | 0.00087 | 0.13166 | -0.729 | FXR2 | FMR1 autosomal homolog 2 [Source:HGNC Symbol;Acc:HGNC:4024] |
| ENSG00000024526 | 1.325 | 2.958 | 3.433 | 0.00089 | 0.13300 | -0.733 | DEPDC1 | DEP domain containing 1 [Source:HGNC Symbol;Acc:HGNC:22949] |
| ENSG00000158402 | 0.761 | 2.098 | 3.432 | 0.00090 | 0.13300 | -0.746 | CDC25C | cell division cycle 25C [Source:HGNC Symbol;Acc:HGNC:1727] |
| ENSG00000122952 | 1.175 | 3.407 | 3.429 | 0.00091 | 0.13300 | -0.756 | ZWINT | ZW10 interacting kinetochore protein [Source:HGNC Symbol;Acc:HGNC:13195] |

| | | | | | | | | |
|---|---|---|---|---|---|---|---|---|
| ENSG00000169679 | 0.950 | 4.638 | 3.419 | 0.00094 | 0.13623 | -0.821 | BUB1 | BUB1 mitotic checkpoint serine/threonine kinase [Source:HGNC Symbol;Acc:HGNC:1148] |
| ENSG00000134690 | 1.128 | 1.699 | 3.415 | 0.00095 | 0.13648 | -0.804 | CDCA8 | cell division cycle associated 8 [Source:HGNC Symbol;Acc:HGNC:14629] |
| ENSG00000129824 | 3.389 | 1.972 | 3.407 | 0.00098 | 0.13839 | -0.801 | RPS4Y1 | ribosomal protein S4, Y-linked 1 [Source:HGNC Symbol;Acc:HGNC:10425] |
| ENSG00000091651 | 0.988 | 2.053 | 3.392 | 0.00102 | 0.13900 | -0.848 | ORC6 | origin recognition complex subunit 6 [Source:HGNC Symbol;Acc:HGNC:17151] |
| ENSG00000072571 | 1.079 | 4.218 | 3.392 | 0.00102 | 0.13900 | -0.889 | HMMR | hyaluronan mediated motility receptor [Source:HGNC Symbol;Acc:HGNC:5012] |
| ENSG00000163923 | 0.861 | 2.123 | 3.391 | 0.00103 | 0.13900 | -0.853 | RPL39L | ribosomal protein L39 like [Source:HGNC Symbol;Acc:HGNC:17094] |
| ENSG00000130338 | -0.593 | 7.135 | -3.389 | 0.00103 | 0.13900 | -0.919 | TULP4 | tubby like protein 4 [Source:HGNC Symbol;Acc:HGNC:15530] |
| ENSG00000137804 | 1.114 | 5.274 | 3.387 | 0.00104 | 0.13900 | -0.924 | NUSAP1 | nucleolar and spindle associated protein 1 [Source:HGNC Symbol;Acc:HGNC:18538] |
| ENSG00000123080 | 1.505 | 3.254 | 3.386 | 0.00104 | 0.13900 | -0.880 | CDKN2C | cyclin dependent kinase inhibitor 2C [Source:HGNC Symbol;Acc:HGNC:1789] |
| ENSG00000186010 | 0.576 | 5.833 | 3.373 | 0.00109 | 0.14263 | -0.965 | NDUFA13 | NADH:ubiquinone oxidoreductase subunit A13 [Source:HGNC Symbol;Acc:HGNC:17194] |
| ENSG00000137807 | 1.125 | 3.242 | 3.371 | 0.00109 | 0.14263 | -0.915 | KIF23 | kinesin family member 23 [Source:HGNC Symbol;Acc:HGNC:6392] |
| ENSG00000106268 | 0.842 | 2.063 | 3.365 | 0.00112 | 0.14386 | -0.921 | NUDT1 | nudix hydrolase 1 [Source:HGNC Symbol;Acc:HGNC:8048] |
| ENSG00000189171 | 0.875 | 3.470 | 3.360 | 0.00113 | 0.14433 | -0.947 | S100A13 | S100 calcium binding protein A13 [Source:HGNC Symbol;Acc:HGNC:10490] |
| ENSG00000164109 | 0.997 | 4.245 | 3.358 | 0.00114 | 0.14433 | -0.984 | MAD2L1 | mitotic arrest deficient 2 like 1 [Source:HGNC Symbol;Acc:HGNC:6763] |
| ENSG00000121152 | 1.059 | 2.977 | 3.353 | 0.00116 | 0.14532 | -0.956 | NCAPH | non-SMC condensin I complex subunit H [Source:HGNC Symbol;Acc:HGNC:1112] |
| ENSG00000174371 | 1.187 | 3.349 | 3.339 | 0.00121 | 0.15034 | -1.009 | EXO1 | exonuclease 1 [Source:HGNC Symbol;Acc:HGNC:3511] |
| ENSG00000238083 | -0.677 | 4.211 | -3.328 | 0.00126 | 0.15229 | -1.028 | LRRC37A2 | leucine rich repeat containing 37 member A2 [Source:HGNC Symbol;Acc:HGNC:32404] |
| ENSG00000153253 | 2.051 | 3.137 | 3.327 | 0.00126 | 0.15229 | -1.050 | SCN3A | sodium voltage-gated channel alpha subunit 3 [Source:HGNC Symbol;Acc:HGNC:10590] |
| ENSG00000258674 | 0.584 | 5.795 | 3.324 | 0.00128 | 0.15229 | -1.107 | AC011448.1 | |
| ENSG00000004468 | -0.847 | 8.364 | -3.324 | 0.00128 | 0.15229 | -1.088 | CD38 | CD38 molecule [Source:HGNC Symbol;Acc:HGNC:1667] |
| ENSG00000166508 | 0.728 | 6.453 | 3.318 | 0.00130 | 0.15369 | -1.120 | MCM7 | minichromosome maintenance complex component 7 [Source:HGNC Symbol;Acc:HGNC:6950] |
| ENSG00000149554 | 0.819 | 4.960 | 3.313 | 0.00132 | 0.15467 | -1.128 | CHEK1 | checkpoint kinase 1 [Source:HGNC Symbol;Acc:HGNC:1925] |
| ENSG00000153993 | 1.612 | 1.884 | 3.309 | 0.00134 | 0.15529 | -1.066 | SEMA3D | semaphorin 3D [Source:HGNC Symbol;Acc:HGNC:10726] |
| ENSG00000160352 | 0.887 | 3.283 | 3.306 | 0.00135 | 0.15545 | -1.093 | ZNF714 | zinc finger protein 714 [Source:HGNC Symbol;Acc:HGNC:27124] |
| ENSG00000065328 | 1.098 | 2.281 | 3.297 | 0.00139 | 0.15739 | -1.097 | MCM10 | minichromosome maintenance 10 replication initiation factor [Source:HGNC Symbol;Acc:HGNC:18043] |
| ENSG00000008277 | 1.291 | 3.522 | 3.291 | 0.00141 | 0.15739 | -1.150 | ADAM22 | ADAM metallopeptidase domain 22 [Source:HGNC Symbol;Acc:HGNC:201] |
| ENSG00000259288 | 0.806 | 1.629 | 3.291 | 0.00142 | 0.15739 | -1.156 | BUB1B-PAK6 | BUB1B-PAK6 readthrough [Source:HGNC Symbol;Acc:HGNC:52276] |
| ENSG00000182022 | -0.548 | 6.895 | -3.289 | 0.00142 | 0.15739 | -1.203 | CHST15 | carbohydrate sulfotransferase 15 [Source:HGNC Symbol;Acc:HGNC:18137] |
| ENSG00000073008 | 1.432 | 0.691 | 3.288 | 0.00143 | 0.15739 | -1.250 | PVR | poliovirus receptor [Source:HGNC Symbol;Acc:HGNC:9705] |
| ENSG00000146410 | 0.781 | 1.693 | 3.284 | 0.00145 | 0.15783 | -1.166 | MTFR2 | mitochondrial fission regulator 2 [Source:HGNC Symbol;Acc:HGNC:21115] |
| ENSG00000121289 | 0.453 | 3.368 | 3.275 | 0.00149 | 0.15853 | -1.170 | CEP89 | centrosomal protein 89 [Source:HGNC Symbol;Acc:HGNC:25907] |
| ENSG00000136156 | -0.850 | 7.767 | -3.272 | 0.00150 | 0.15853 | -1.246 | ITM2B | integral membrane protein 2B [Source:HGNC Symbol;Acc:HGNC:6174] |
| ENSG00000075213 | 1.863 | 2.842 | 3.272 | 0.00151 | 0.15853 | -1.189 | SEMA3A | semaphorin 3A [Source:HGNC Symbol;Acc:HGNC:10723] |
| ENSG00000093009 | 1.118 | 2.761 | 3.266 | 0.00153 | 0.15853 | -1.189 | CDC45 | cell division cycle 45 [Source:HGNC Symbol;Acc:HGNC:1739] |
| ENSG00000152580 | 1.674 | -0.070 | 3.266 | 0.00153 | 0.15853 | -1.446 | IGSF10 | immunoglobulin superfamily member 10 [Source:HGNC Symbol;Acc:HGNC:26384] |
| ENSG00000079616 | 0.719 | 5.232 | 3.265 | 0.00154 | 0.15853 | -1.268 | KIF22 | kinesin family member 22 [Source:HGNC Symbol;Acc:HGNC:6391] |
| ENSG00000141252 | -0.538 | 5.179 | -3.265 | 0.00154 | 0.15853 | -1.246 | VPS53 | VPS53, GARP complex subunit [Source:HGNC Symbol;Acc:HGNC:25608] |
| ENSG00000198692 | 3.141 | 0.627 | 3.255 | 0.00159 | 0.16217 | -1.229 | EIF1AY | eukaryotic translation initiation factor 1A, Y-linked [Source:HGNC Symbol;Acc:HGNC:3252] |
| ENSG00000179958 | 0.774 | 1.966 | 3.250 | 0.00161 | 0.16343 | -1.236 | DCTPP1 | dCTP pyrophosphatase 1 [Source:HGNC Symbol;Acc:HGNC:28777] |
| ENSG00000197746 | -0.713 | 10.317 | -3.244 | 0.00164 | 0.16524 | -1.269 | PSAP | prosaposin [Source:HGNC Symbol;Acc:HGNC:9498] |
| ENSG00000131747 | 1.234 | 5.276 | 3.240 | 0.00166 | 0.16606 | -1.340 | TOP2A | topoisomerase (DNA) II alpha [Source:HGNC Symbol;Acc:HGNC:11989] |
| ENSG00000179873 | 1.838 | 0.949 | 3.235 | 0.00169 | 0.16646 | -1.292 | NLRP11 | NLR family pyrin domain containing 11 [Source:HGNC Symbol;Acc:HGNC:22945] |
| ENSG00000137713 | 0.493 | 4.889 | 3.234 | 0.00169 | 0.16646 | -1.343 | PPP2R1B | protein phosphatase 2 scaffold subunit Abeta [Source:HGNC Symbol;Acc:HGNC:9303] |
| ENSG00000183207 | 0.616 | 5.560 | 3.228 | 0.00172 | 0.16788 | -1.373 | RUVBL2 | RuvB like AAA ATPase 2 [Source:HGNC Symbol;Acc:HGNC:10475] |
| ENSG00000133019 | 2.233 | 0.968 | 3.226 | 0.00174 | 0.16788 | -1.305 | CHRM3 | cholinergic receptor muscarinic 3 [Source:HGNC Symbol;Acc:HGNC:1952] |
| ENSG00000131153 | 0.896 | 1.863 | 3.224 | 0.00175 | 0.16816 | -1.302 | GINS2 | GINS complex subunit 2 [Source:HGNC Symbol;Acc:HGNC:24575] |
| ENSG00000134897 | 1.564 | 0.659 | 3.213 | 0.00181 | 0.17244 | -1.406 | BIVM | basic, immunoglobulin-like variable motif containing [Source:HGNC Symbol;Acc:HGNC:16034] |
| ENSG00000086102 | -0.461 | 6.572 | -3.208 | 0.00184 | 0.17405 | -1.432 | NFX1 | nuclear transcription factor, X-box binding 1 [Source:HGNC Symbol;Acc:HGNC:7803] |
| ENSG00000106588 | 0.646 | 6.535 | 3.204 | 0.00186 | 0.17405 | -1.440 | PSMA2 | proteasome subunit alpha 2 [Source:HGNC Symbol;Acc:HGNC:9531] |
| ENSG00000175048 | -1.310 | 3.937 | -3.197 | 0.00190 | 0.17405 | -1.363 | ZDHHC14 | zinc finger DHHC-type containing 14 [Source:HGNC Symbol;Acc:HGNC:20341] |
| ENSG00000182150 | -0.438 | 5.782 | -3.195 | 0.00192 | 0.17405 | -1.458 | ERCC6L2 | ERCC excision repair 6 like 2 [Source:HGNC Symbol;Acc:HGNC:26922] |
| ENSG00000073111 | 1.087 | 4.579 | 3.195 | 0.00192 | 0.17405 | -1.453 | MCM2 | minichromosome maintenance complex component 2 [Source:HGNC Symbol;Acc:HGNC:6944] |
| ENSG00000131080 | -1.692 | 2.491 | -3.194 | 0.00192 | 0.17405 | -1.538 | EDA2R | ectodysplasin A2 receptor [Source:HGNC Symbol;Acc:HGNC:17756] |
| ENSG00000128789 | 0.481 | 4.598 | 3.194 | 0.00192 | 0.17405 | -1.445 | PSMG2 | proteasome assembly chaperone 2 [Source:HGNC Symbol;Acc:HGNC:24929] |
| ENSG00000166803 | 0.961 | 2.728 | 3.190 | 0.00195 | 0.17415 | -1.389 | PCLAF | PCNA clamp associated factor [Source:HGNC Symbol;Acc:HGNC:28961] |
| ENSG00000112877 | 0.828 | 1.396 | 3.188 | 0.00196 | 0.17415 | -1.433 | CEP72 | centrosomal protein 72 [Source:HGNC Symbol;Acc:HGNC:25547] |
| ENSG00000261915 | -0.409 | 5.109 | -3.187 | 0.00196 | 0.17415 | -1.460 | AC026954.2 | |
| ENSG00000111247 | 0.935 | 3.529 | 3.181 | 0.00200 | 0.17552 | -1.446 | RAD51AP1 | RAD51 associated protein 1 [Source:HGNC Symbol;Acc:HGNC:16956] |
| ENSG00000005189 | 0.803 | 1.929 | 3.180 | 0.00201 | 0.17552 | -1.413 | REXO5 | RNA exonuclease 5 [Source:HGNC Symbol;Acc:HGNC:24661] |
| ENSG00000087586 | 0.994 | 2.282 | 3.173 | 0.00205 | 0.17693 | -1.423 | AURKA | aurora kinase A [Source:HGNC Symbol;Acc:HGNC:11393] |
| ENSG00000133275 | 0.437 | 5.171 | 3.172 | 0.00205 | 0.17693 | -1.519 | CSNK1G2 | casein kinase 1 gamma 2 [Source:HGNC Symbol;Acc:HGNC:2455] |
| ENSG00000087448 | 0.688 | 4.216 | 3.171 | 0.00206 | 0.17693 | -1.497 | KLHL42 | kelch like family member 42 [Source:HGNC Symbol;Acc:HGNC:29252] |
| ENSG00000099795 | 0.569 | 4.931 | 3.161 | 0.00212 | 0.17945 | -1.545 | NDUFB7 | NADH:ubiquinone oxidoreductase subunit B7 [Source:HGNC Symbol;Acc:HGNC:7702] |
| ENSG00000271503 | -1.937 | 4.077 | -3.160 | 0.00214 | 0.17945 | -1.457 | CCL5 | C-C motif chemokine ligand 5 [Source:HGNC Symbol;Acc:HGNC:10632] |
| ENSG00000183647 | 0.562 | 2.051 | 3.158 | 0.00215 | 0.17945 | -1.470 | ZNF530 | zinc finger protein 530 [Source:HGNC Symbol;Acc:HGNC:29297] |
| ENSG00000108654 | -0.473 | 11.029 | -3.157 | 0.00216 | 0.17945 | -1.488 | DDX5 | DEAD-box helicase 5 [Source:HGNC Symbol;Acc:HGNC:2746] |
| ENSG00000040633 | -0.472 | 3.651 | -3.156 | 0.00216 | 0.17945 | -1.478 | PHF23 | PHD finger protein 23 [Source:HGNC Symbol;Acc:HGNC:28428] |
| ENSG00000123219 | 0.877 | 3.598 | 3.153 | 0.00218 | 0.17945 | -1.522 | CENPK | centromere protein K [Source:HGNC Symbol;Acc:HGNC:29479] |
| ENSG00000115310 | -0.564 | 7.317 | -3.144 | 0.00224 | 0.18309 | -1.604 | RTN4 | reticulon 4 [Source:HGNC Symbol;Acc:HGNC:14085] |
| ENSG00000164087 | 0.911 | 1.323 | 3.140 | 0.00227 | 0.18309 | -1.546 | POC1A | POC1 centriolar protein A [Source:HGNC Symbol;Acc:HGNC:24488] |
| ENSG00000075702 | 1.056 | 2.308 | 3.139 | 0.00228 | 0.18309 | -1.512 | WDR62 | WD repeat domain 62 [Source:HGNC Symbol;Acc:HGNC:24502] |
| ENSG00000198826 | 0.944 | 4.369 | 3.136 | 0.00229 | 0.18309 | -1.602 | ARHGAP11A | Rho GTPase activating protein 11A [Source:HGNC Symbol;Acc:HGNC:15783] |
| ENSG00000273604 | 1.243 | 1.119 | 3.135 | 0.00230 | 0.18309 | -1.573 | EPOP | elongin BC and polycomb repressive complex 2 associated protein [Source:HGNC Symbol;Acc:HGNC:34493] |
| ENSG00000146918 | 0.826 | 4.332 | 3.135 | 0.00231 | 0.18309 | -1.603 | NCAPG2 | non-SMC condensin II complex subunit G2 [Source:HGNC Symbol;Acc:HGNC:21904] |
| ENSG00000213145 | 1.498 | 1.858 | 3.131 | 0.00233 | 0.18401 | -1.531 | CRIP1 | cysteine rich protein 1 [Source:HGNC Symbol;Acc:HGNC:2360] |
| ENSG00000265818 | 0.560 | 3.974 | 3.124 | 0.00238 | 0.18671 | -1.609 | EEF1E1-BLOC1S5 | EEF1E1-BLOC1S5 readthrough (NMD candidate) [Source:HGNC Symbol;Acc:HGNC:49187] |
| ENSG00000171320 | 1.136 | 2.244 | 3.122 | 0.00240 | 0.18699 | -1.556 | ESCO2 | establishment of sister chromatid cohesion N-acetyltransferase 2 [Source:HGNC Symbol;Acc:HGNC:27230] |
| ENSG00000127445 | 0.500 | 4.576 | 3.118 | 0.00243 | 0.18787 | -1.650 | PIN1 | peptidylprolyl cis/trans isomerase, NIMA-interacting 1 [Source:HGNC Symbol;Acc:HGNC:8988] |
| ENSG00000079277 | -0.493 | 4.472 | -3.116 | 0.00244 | 0.18787 | -1.620 | MKNK1 | MAP kinase interacting serine/threonine kinase 1 [Source:HGNC Symbol;Acc:HGNC:7110] |
| ENSG00000144366 | 2.459 | 0.595 | 3.110 | 0.00249 | 0.19011 | -1.619 | GULP1 | GULP, engulfment adaptor PTB domain containing 1 [Source:HGNC Symbol;Acc:HGNC:18649] |
| ENSG00000144115 | 2.253 | -1.150 | 3.106 | 0.00252 | 0.19129 | -1.948 | THNSL2 | threonine synthase like 2 [Source:HGNC Symbol;Acc:HGNC:25602] |
| ENSG00000179750 | 1.030 | 2.784 | 3.104 | 0.00253 | 0.19129 | -1.617 | APOBEC3B | apolipoprotein B mRNA editing enzyme catalytic subunit 3B [Source:HGNC Symbol;Acc:HGNC:17352] |
| ENSG00000142731 | 1.012 | 2.983 | 3.103 | 0.00255 | 0.19129 | -1.630 | PLK4 | polo like kinase 4 [Source:HGNC Symbol;Acc:HGNC:11397] |
| ENSG00000276043 | 1.019 | 2.551 | 3.097 | 0.00259 | 0.19301 | -1.626 | UHRF1 | ubiquitin like with PHD and ring finger domains 1 [Source:HGNC Symbol;Acc:HGNC:12556] |
| ENSG00000138160 | 0.919 | 4.644 | 3.096 | 0.00260 | 0.19301 | -1.720 | KIF11 | kinesin family member 11 [Source:HGNC Symbol;Acc:HGNC:6388] |
| ENSG00000197299 | 0.844 | 4.131 | 3.093 | 0.00262 | 0.19327 | -1.706 | BLM | Bloom syndrome RecQ like helicase [Source:HGNC Symbol;Acc:HGNC:1058] |
| ENSG00000123636 | -0.698 | 7.716 | -3.085 | 0.00269 | 0.19621 | -1.758 | BAZ2B | bromodomain adjacent to zinc finger domain 2B [Source:HGNC Symbol;Acc:HGNC:963] |
| ENSG00000140386 | -0.660 | 6.767 | -3.084 | 0.00270 | 0.19621 | -1.769 | SCAPER | S-phase cyclin A associated protein in the ER [Source:HGNC Symbol;Acc:HGNC:13081] |
| ENSG00000138411 | 1.365 | 0.331 | 3.083 | 0.00270 | 0.19621 | -1.775 | HECW2 | HECT, C2 and WW domain containing E3 ubiquitin protein ligase 2 [Source:HGNC Symbol;Acc:HGNC:29853] |
| ENSG00000011426 | 1.036 | 3.467 | 3.080 | 0.00273 | 0.19685 | -1.715 | ANLN | anillin actin binding protein [Source:HGNC Symbol;Acc:HGNC:14082] |

| | | | | | | | | |
|---|---|---|---|---|---|---|---|---|
| ENSG00000188368 | 0.792 | 0.106 | 3.070 | 0.00282 | 0.20018 | -1.920 | PRR19 | proline rich 19 [Source:HGNC Symbol;Acc:HGNC:33728] |
| ENSG00000129173 | 1.168 | 1.276 | 3.066 | 0.00285 | 0.20018 | -1.715 | E2F8 | E2F transcription factor 8 [Source:HGNC Symbol;Acc:HGNC:24727] |
| ENSG00000105364 | 0.677 | 3.270 | 3.066 | 0.00285 | 0.20018 | -1.732 | MRPL4 | mitochondrial ribosomal protein L4 [Source:HGNC Symbol;Acc:HGNC:14276] |
| ENSG00000163535 | 0.785 | 3.178 | 3.065 | 0.00285 | 0.20018 | -1.731 | SGO2 | shugoshin 2 [Source:HGNC Symbol;Acc:HGNC:30812] |
| ENSG00000077152 | 1.035 | 2.365 | 3.064 | 0.00287 | 0.20018 | -1.706 | UBE2T | ubiquitin conjugating enzyme E2 T [Source:HGNC Symbol;Acc:HGNC:25009] |
| ENSG00000117724 | 1.192 | 5.618 | 3.063 | 0.00287 | 0.20018 | -1.821 | CENPF | centromere protein F [Source:HGNC Symbol;Acc:HGNC:1857] |
| ENSG00000140395 | 0.482 | 4.805 | 3.060 | 0.00290 | 0.20146 | -1.813 | WDR61 | WD repeat domain 61 [Source:HGNC Symbol;Acc:HGNC:30300] |
| ENSG00000147536 | 0.828 | 1.915 | 3.054 | 0.00295 | 0.20380 | -1.725 | GINS4 | GINS complex subunit 4 [Source:HGNC Symbol;Acc:HGNC:28226] |
| ENSG00000172590 | 0.520 | 4.184 | 3.050 | 0.00298 | 0.20485 | -1.812 | MRPL52 | mitochondrial ribosomal protein L52 [Source:HGNC Symbol;Acc:HGNC:16655] |
| ENSG00000109805 | 0.712 | 4.877 | 3.047 | 0.00302 | 0.20580 | -1.853 | NCAPG | non-SMC condensin I complex subunit G [Source:HGNC Symbol;Acc:HGNC:24304] |
| ENSG00000185386 | 1.204 | 0.720 | 3.043 | 0.00305 | 0.20580 | -1.824 | MAPK11 | mitogen-activated protein kinase 11 [Source:HGNC Symbol;Acc:HGNC:6873] |
| ENSG00000101624 | 0.444 | 3.560 | 3.042 | 0.00306 | 0.20580 | -1.798 | CEP76 | centrosomal protein 76 [Source:HGNC Symbol;Acc:HGNC:25727] |
| ENSG00000065559 | -0.444 | 5.480 | -3.042 | 0.00306 | 0.20580 | -1.861 | MAP2K4 | mitogen-activated protein kinase kinase 4 [Source:HGNC Symbol;Acc:HGNC:6844] |
| ENSG00000197121 | 1.099 | 2.084 | 3.038 | 0.00310 | 0.20722 | -1.765 | PGAP1 | post-GPI attachment to proteins 1 [Source:HGNC Symbol;Acc:HGNC:25712] |
| ENSG00000136866 | 1.287 | 0.690 | 3.033 | 0.00314 | 0.20892 | -1.847 | ZFP37 | ZFP37 zinc finger protein [Source:HGNC Symbol;Acc:HGNC:12863] |
| ENSG00000123485 | 1.112 | 2.570 | 3.030 | 0.00317 | 0.20892 | -1.799 | HJURP | Holliday junction recognition protein [Source:HGNC Symbol;Acc:HGNC:25444] |
| ENSG00000149548 | 1.066 | 2.123 | 3.030 | 0.00317 | 0.20892 | -1.784 | CCDC15 | coiled-coil domain containing 15 [Source:HGNC Symbol;Acc:HGNC:25798] |
| ENSG00000256646 | 0.621 | 6.461 | 3.028 | 0.00319 | 0.20919 | -1.915 | AC010132.3 | |
| ENSG00000008394 | 1.870 | 0.172 | 3.024 | 0.00323 | 0.20979 | -1.904 | MGST1 | microsomal glutathione S-transferase 1 [Source:HGNC Symbol;Acc:HGNC:7061] |
| ENSG00000143476 | 1.028 | 3.976 | 3.023 | 0.00324 | 0.20979 | -1.887 | DTL | denticleless E3 ubiquitin protein ligase homolog [Source:HGNC Symbol;Acc:HGNC:30288] |
| ENSG00000178852 | -0.894 | 5.146 | -3.015 | 0.00332 | 0.21367 | -1.903 | EFCAB13 | EF-hand calcium binding domain 13 [Source:HGNC Symbol;Acc:HGNC:26864] |
| ENSG00000141968 | 0.561 | 6.736 | 3.014 | 0.00333 | 0.21367 | -1.948 | VAV1 | vav guanine nucleotide exchange factor 1 [Source:HGNC Symbol;Acc:HGNC:12657] |
| ENSG00000088325 | 1.058 | 3.747 | 3.009 | 0.00338 | 0.21582 | -1.916 | TPX2 | TPX2, microtubule nucleation factor [Source:HGNC Symbol;Acc:HGNC:1249] |
| ENSG00000055208 | -0.655 | 6.189 | -3.007 | 0.00340 | 0.21582 | -1.966 | TAB2 | TGF-beta activated kinase 1/MAP3K7 binding protein 2 [Source:HGNC Symbol;Acc:HGNC:17075] |
| ENSG00000066279 | 1.147 | 5.273 | 3.004 | 0.00343 | 0.21632 | -1.978 | ASPM | abnormal spindle microtubule assembly [Source:HGNC Symbol;Acc:HGNC:19048] |
| ENSG00000100462 | 0.552 | 5.248 | 3.003 | 0.00344 | 0.21632 | -1.975 | PRMT5 | protein arginine methyltransferase 5 [Source:HGNC Symbol;Acc:HGNC:10894] |
| ENSG00000152270 | 1.324 | 3.786 | 2.986 | 0.00362 | 0.22655 | -1.982 | PDE3B | phosphodiesterase 3B [Source:HGNC Symbol;Acc:HGNC:8779] |
| ENSG00000026297 | -1.000 | 7.431 | -2.983 | 0.00365 | 0.22752 | -2.032 | RNASET2 | ribonuclease T2 [Source:HGNC Symbol;Acc:HGNC:21686] |
| ENSG00000178125 | 0.512 | 2.968 | 2.976 | 0.00372 | 0.22991 | -1.934 | PPP1R42 | protein phosphatase 1 regulatory subunit 42 [Source:HGNC Symbol;Acc:HGNC:33732] |
| ENSG00000074356 | -0.353 | 5.715 | -2.975 | 0.00374 | 0.22991 | -2.044 | NCBP3 | nuclear cap binding subunit 3 [Source:HGNC Symbol;Acc:HGNC:24612] |
| ENSG00000122966 | 0.975 | 2.758 | 2.975 | 0.00374 | 0.22991 | -1.946 | CIT | citron rho-interacting serine/threonine kinase [Source:HGNC Symbol;Acc:HGNC:1985] |
| ENSG00000198087 | -0.420 | 7.008 | -2.972 | 0.00377 | 0.23055 | -2.062 | CD2AP | CD2 associated protein [Source:HGNC Symbol;Acc:HGNC:14258] |
| ENSG00000257341 | 1.254 | 2.094 | 2.966 | 0.00384 | 0.23349 | -1.942 | AL928654.4 | |
| ENSG00000161800 | 0.871 | 3.794 | 2.963 | 0.00387 | 0.23452 | -2.030 | RACGAP1 | Rac GTPase activating protein 1 [Source:HGNC Symbol;Acc:HGNC:9804] |
| ENSG00000092208 | 0.617 | 2.991 | 2.950 | 0.00403 | 0.24220 | -2.004 | GEMIN2 | gem nuclear organelle associated protein 2 [Source:HGNC Symbol;Acc:HGNC:10884] |
| ENSG00000010818 | -0.738 | 6.095 | -2.948 | 0.00405 | 0.24220 | -2.117 | HIVEP2 | human immunodeficiency virus type I enhancer binding protein 2 [Source:HGNC Symbol;Acc:HGNC:4921] |
| ENSG00000100479 | 0.964 | 1.887 | 2.947 | 0.00406 | 0.24220 | -1.983 | POLE2 | DNA polymerase epsilon 2, accessory subunit [Source:HGNC Symbol;Acc:HGNC:9178] |
| ENSG00000123975 | 0.808 | 4.312 | 2.936 | 0.00419 | 0.24896 | -2.122 | CKS2 | CDC28 protein kinase regulatory subunit 2 [Source:HGNC Symbol;Acc:HGNC:2000] |
| ENSG00000136052 | -0.707 | 5.711 | -2.932 | 0.00424 | 0.25073 | -2.147 | SLC41A2 | solute carrier family 41 member 2 [Source:HGNC Symbol;Acc:HGNC:31045] |
| ENSG00000196550 | 0.756 | 0.663 | 2.929 | 0.00429 | 0.25088 | -2.126 | FAM72A | family with sequence similarity 72 member A [Source:HGNC Symbol;Acc:HGNC:24044] |
| ENSG00000174238 | -0.382 | 5.196 | -2.929 | 0.00429 | 0.25088 | -2.147 | PITPNA | phosphatidylinositol transfer protein alpha [Source:HGNC Symbol;Acc:HGNC:9001] |
| ENSG00000169021 | 0.452 | 5.603 | 2.928 | 0.00430 | 0.25088 | -2.176 | UQCRFS1 | ubiquinol-cytochrome c reductase, Rieske iron-sulfur polypeptide 1 [Source:HGNC Symbol;Acc:HGNC:12587] |
| ENSG00000188610 | 1.056 | -0.229 | 2.926 | 0.00432 | 0.25099 | -2.248 | FAM72B | family with sequence similarity 72 member B [Source:HGNC Symbol;Acc:HGNC:24805] |
| ENSG00000150753 | 0.731 | 7.056 | 2.924 | 0.00434 | 0.25101 | -2.170 | CCT5 | chaperonin containing TCP1 subunit 5 [Source:HGNC Symbol;Acc:HGNC:1618] |
| ENSG00000186185 | 1.365 | -0.136 | 2.912 | 0.00450 | 0.25881 | -2.215 | KIF18B | kinesin family member 18B [Source:HGNC Symbol;Acc:HGNC:27102] |
| ENSG00000005194 | 0.458 | 3.874 | 2.908 | 0.00456 | 0.25887 | -2.160 | CIAPIN1 | cytokine induced apoptosis inhibitor 1 [Source:HGNC Symbol;Acc:HGNC:28050] |
| ENSG00000076706 | 1.514 | 1.498 | 2.908 | 0.00456 | 0.25887 | -2.076 | MCAM | melanoma cell adhesion molecule [Source:HGNC Symbol;Acc:HGNC:6934] |
| ENSG00000178764 | -0.612 | 4.953 | -2.906 | 0.00458 | 0.25887 | -2.184 | ZHX2 | zinc fingers and homeoboxes 2 [Source:HGNC Symbol;Acc:HGNC:18513] |
| ENSG00000256229 | 1.505 | 0.184 | 2.906 | 0.00459 | 0.25887 | -2.157 | ZNF486 | zinc finger protein 486 [Source:HGNC Symbol;Acc:HGNC:20807] |
| ENSG00000070950 | 0.531 | 4.429 | 2.904 | 0.00461 | 0.25887 | -2.202 | RAD18 | RAD18, E3 ubiquitin protein ligase [Source:HGNC Symbol;Acc:HGNC:18278] |
| ENSG00000012048 | 0.765 | 4.440 | 2.903 | 0.00462 | 0.25887 | -2.212 | BRCA1 | BRCA1, DNA repair associated [Source:HGNC Symbol;Acc:HGNC:1100] |
| ENSG00000120129 | -0.902 | 8.960 | -2.901 | 0.00465 | 0.25961 | -2.206 | DUSP1 | dual specificity phosphatase 1 [Source:HGNC Symbol;Acc:HGNC:3064] |
| ENSG00000135476 | 0.909 | 1.941 | 2.896 | 0.00471 | 0.26034 | -2.105 | ESPL1 | extra spindle pole bodies like 1, separase [Source:HGNC Symbol;Acc:HGNC:16856] |
| ENSG00000169607 | 0.977 | 2.727 | 2.896 | 0.00472 | 0.26034 | -2.140 | CKAP2L | cytoskeleton associated protein 2 like [Source:HGNC Symbol;Acc:HGNC:26877] |
| ENSG00000182050 | 2.710 | 0.880 | 2.894 | 0.00475 | 0.26034 | -2.112 | MGAT4C | MGAT4 family member C [Source:HGNC Symbol;Acc:HGNC:30871] |
| ENSG00000160957 | 0.933 | 2.372 | 2.893 | 0.00477 | 0.26034 | -2.127 | RECQL4 | RecQ like helicase 4 [Source:HGNC Symbol;Acc:HGNC:9949] |
| ENSG00000237649 | 0.911 | 2.700 | 2.892 | 0.00477 | 0.26034 | -2.143 | KIFC1 | kinesin family member C1 [Source:HGNC Symbol;Acc:HGNC:6389] |
| ENSG00000214960 | -2.281 | 0.431 | -2.890 | 0.00481 | 0.26118 | -2.647 | ISPD | isoprenoid synthase domain containing [Source:HGNC Symbol;Acc:HGNC:37276] |
| ENSG00000277775 | 1.191 | 1.584 | 2.884 | 0.00488 | 0.26415 | -2.131 | HIST1H3F | histone cluster 1 H3 family member f [Source:HGNC Symbol;Acc:HGNC:4773] |
| ENSG00000105401 | 0.357 | 7.154 | 2.881 | 0.00493 | 0.26517 | -2.284 | CDC37 | cell division cycle 37 [Source:HGNC Symbol;Acc:HGNC:1735] |
| ENSG00000107518 | 2.223 | -0.029 | 2.880 | 0.00494 | 0.26517 | -2.223 | ATRNL1 | attractin like 1 [Source:HGNC Symbol;Acc:HGNC:29063] |
| ENSG00000105197 | 0.466 | 4.344 | 2.877 | 0.00499 | 0.26688 | -2.266 | TIMM50 | translocase of inner mitochondrial membrane 50 [Source:HGNC Symbol;Acc:HGNC:23656] |
| ENSG00000108106 | 0.591 | 4.217 | 2.871 | 0.00507 | 0.26987 | -2.275 | UBE2S | ubiquitin conjugating enzyme E2 S [Source:HGNC Symbol;Acc:HGNC:17895] |
| ENSG00000145386 | 0.775 | 4.208 | 2.865 | 0.00516 | 0.27295 | -2.297 | CCNA2 | cyclin A2 [Source:HGNC Symbol;Acc:HGNC:1578] |
| ENSG00000178971 | -0.623 | 4.805 | -2.864 | 0.00517 | 0.27295 | -2.281 | CTC1 | CST telomere replication complex component 1 [Source:HGNC Symbol;Acc:HGNC:26169] |
| ENSG00000041357 | 0.621 | 6.852 | 2.855 | 0.00532 | 0.27952 | -2.353 | PSMA4 | proteasome subunit alpha 4 [Source:HGNC Symbol;Acc:HGNC:9533] |
| ENSG00000087842 | 0.927 | 0.394 | 2.849 | 0.00541 | 0.28332 | -2.312 | PIR | pirin [Source:HGNC Symbol;Acc:HGNC:30048] |
| ENSG00000170296 | -0.455 | 5.898 | -2.843 | 0.00550 | 0.28567 | -2.383 | GABARAP | GABA type A receptor-associated protein [Source:HGNC Symbol;Acc:HGNC:4067] |
| ENSG00000063241 | 0.659 | 3.207 | 2.843 | 0.00550 | 0.28567 | -2.285 | ISOC2 | isochorismatase domain containing 2 [Source:HGNC Symbol;Acc:HGNC:26278] |
| ENSG00000117399 | 1.063 | 1.498 | 2.841 | 0.00553 | 0.28582 | -2.231 | CDC20 | cell division cycle 20 [Source:HGNC Symbol;Acc:HGNC:1723] |
| ENSG00000182326 | -1.157 | 2.116 | -2.837 | 0.00561 | 0.28816 | -2.297 | C1S | complement C1s [Source:HGNC Symbol;Acc:HGNC:1247] |
| ENSG00000076382 | 0.773 | 4.774 | 2.836 | 0.00562 | 0.28816 | -2.393 | SPAG5 | sperm associated antigen 5 [Source:HGNC Symbol;Acc:HGNC:13452] |
| ENSG00000099341 | 0.460 | 6.264 | 2.833 | 0.00566 | 0.28911 | -2.418 | PSMD8 | proteasome 26S subunit, non-ATPase 8 [Source:HGNC Symbol;Acc:HGNC:9566] |
| ENSG00000155561 | 0.424 | 6.179 | 2.832 | 0.00568 | 0.28911 | -2.421 | NUP205 | nucleoporin 205 [Source:HGNC Symbol;Acc:HGNC:18658] |
| ENSG00000146416 | -0.582 | 6.293 | -2.827 | 0.00577 | 0.28935 | -2.430 | AIG1 | androgen induced 1 [Source:HGNC Symbol;Acc:HGNC:21607] |
| ENSG00000101888 | 0.699 | 4.496 | 2.826 | 0.00579 | 0.28935 | -2.407 | NXT2 | nuclear transport factor 2 like export factor 2 [Source:HGNC Symbol;Acc:HGNC:18151] |
| ENSG00000204315 | 0.603 | 2.274 | 2.825 | 0.00579 | 0.28935 | -2.273 | FKBPL | FK506 binding protein like [Source:HGNC Symbol;Acc:HGNC:13949] |
| ENSG00000117318 | 1.717 | 1.398 | 2.825 | 0.00580 | 0.28935 | -2.269 | ID3 | inhibitor of DNA binding 3, HLH protein [Source:HGNC Symbol;Acc:HGNC:5362] |
| ENSG00000156509 | 1.094 | -0.107 | 2.825 | 0.00580 | 0.28935 | -2.415 | FBXO43 | F-box protein 43 [Source:HGNC Symbol;Acc:HGNC:28521] |
| ENSG00000067646 | 2.232 | 1.835 | 2.818 | 0.00592 | 0.29420 | -2.312 | ZFY | zinc finger protein, Y-linked [Source:HGNC Symbol;Acc:HGNC:12870] |
| ENSG00000176890 | 0.696 | 3.689 | 2.812 | 0.00601 | 0.29736 | -2.395 | TYMS | thymidylate synthetase [Source:HGNC Symbol;Acc:HGNC:12441] |
| ENSG00000004866 | -0.367 | 5.161 | -2.811 | 0.00603 | 0.29736 | -2.441 | ST7 | suppression of tumorigenicity 7 [Source:HGNC Symbol;Acc:HGNC:11351] |
| ENSG00000115380 | 1.954 | 1.225 | 2.807 | 0.00611 | 0.30007 | -2.310 | EFEMP1 | EGF containing fibulin like extracellular matrix protein 1 [Source:HGNC Symbol;Acc:HGNC:3218] |
| ENSG00000167670 | 0.693 | 4.575 | 2.801 | 0.00621 | 0.30373 | -2.472 | CHAF1A | chromatin assembly factor 1 subunit A [Source:HGNC Symbol;Acc:HGNC:1910] |
| ENSG00000051180 | 0.872 | 1.946 | 2.800 | 0.00623 | 0.30373 | -2.329 | RAD51 | RAD51 recombinase [Source:HGNC Symbol;Acc:HGNC:9817] |
| ENSG00000213920 | 0.525 | 3.803 | 2.795 | 0.00632 | 0.30595 | -2.433 | MDP1 | magnesium dependent phosphatase 1 [Source:HGNC Symbol;Acc:HGNC:28781] |
| ENSG00000169258 | 0.872 | 0.461 | 2.794 | 0.00634 | 0.30595 | -2.418 | GPRIN1 | G protein regulated inducer of neurite outgrowth 1 [Source:HGNC Symbol;Acc:HGNC:24835] |
| ENSG00000196655 | 0.450 | 4.266 | 2.789 | 0.00642 | 0.30595 | -2.475 | TRAPPC4 | trafficking protein particle complex 4 [Source:HGNC Symbol;Acc:HGNC:19943] |
| ENSG00000133460 | -0.599 | 4.497 | -2.789 | 0.00643 | 0.30595 | -2.447 | SLC2A11 | solute carrier family 2 member 11 [Source:HGNC Symbol;Acc:HGNC:14239] |
| ENSG00000249141 | -0.983 | 6.464 | -2.788 | 0.00644 | 0.30595 | -2.522 | AL159163.1 | |

| | | | | | | | | |
|---|---|---|---|---|---|---|---|---|
| ENSG00000142230 | 0.461 | 5.778 | 2.788 | 0.00644 | 0.30595 | -2.529 | SAE1 | SUMO1 activating enzyme subunit 1 [Source:HGNC Symbol;Acc:HGNC:30660] |
| ENSG00000135541 | -0.666 | 5.778 | -2.788 | 0.00644 | 0.30595 | -2.513 | AHI1 | Abelson helper integration site 1 [Source:HGNC Symbol;Acc:HGNC:21575] |
| ENSG00000164828 | 0.399 | 6.322 | 2.785 | 0.00649 | 0.30636 | -2.536 | SUN1 | Sad1 and UNC84 domain containing 1 [Source:HGNC Symbol;Acc:HGNC:18587] |
| ENSG00000183336 | 0.668 | 1.124 | 2.785 | 0.00650 | 0.30636 | -2.388 | BOLA2 | bolA family member 2 [Source:HGNC Symbol;Acc:HGNC:29488] |
| ENSG00000142945 | 0.935 | 2.628 | 2.781 | 0.00656 | 0.30691 | -2.407 | KIF2C | kinesin family member 2C [Source:HGNC Symbol;Acc:HGNC:6393] |
| ENSG00000075218 | 1.016 | 0.669 | 2.779 | 0.00660 | 0.30691 | -2.411 | GTSE1 | G2 and S-phase expressed 1 [Source:HGNC Symbol;Acc:HGNC:13698] |
| ENSG00000131067 | -0.916 | 2.662 | -2.778 | 0.00663 | 0.30691 | -2.376 | GGT7 | gamma-glutamyltransferase 7 [Source:HGNC Symbol;Acc:HGNC:4259] |
| ENSG00000115233 | 0.531 | 6.761 | 2.778 | 0.00663 | 0.30691 | -2.547 | PSMD14 | proteasome 26S subunit, non-ATPase 14 [Source:HGNC Symbol;Acc:HGNC:16889] |
| ENSG00000035499 | 1.147 | 2.150 | 2.778 | 0.00663 | 0.30691 | -2.393 | DEPDC1B | DEP domain containing 1B [Source:HGNC Symbol;Acc:HGNC:24902] |
| ENSG00000111581 | 0.449 | 5.687 | 2.776 | 0.00666 | 0.30710 | -2.557 | NUP107 | nucleoporin 107 [Source:HGNC Symbol;Acc:HGNC:29914] |
| ENSG00000142599 | -0.647 | 5.603 | -2.773 | 0.00672 | 0.30886 | -2.544 | RERE | arginine-glutamic acid dipeptide repeats [Source:HGNC Symbol;Acc:HGNC:9965] |
| ENSG00000117632 | 0.984 | 5.505 | 2.770 | 0.00678 | 0.30951 | -2.573 | STMN1 | stathmin 1 [Source:HGNC Symbol;Acc:HGNC:6510] |
| ENSG00000183530 | -0.456 | 5.746 | -2.770 | 0.00679 | 0.30951 | -2.562 | PRR14L | proline rich 14 like [Source:HGNC Symbol;Acc:HGNC:28738] |
| ENSG00000156103 | 2.285 | 0.319 | 2.766 | 0.00686 | 0.31188 | -2.423 | MMP16 | matrix metallopeptidase 16 [Source:HGNC Symbol;Acc:HGNC:7162] |
| ENSG00000012817 | 2.933 | 1.552 | 2.764 | 0.00689 | 0.31188 | -2.434 | KDM5D | lysine demethylase 5D [Source:HGNC Symbol;Acc:HGNC:11115] |
| ENSG00000097046 | 0.676 | 2.849 | 2.760 | 0.00697 | 0.31188 | -2.460 | CDC7 | cell division cycle 7 [Source:HGNC Symbol;Acc:HGNC:1745] |
| ENSG00000150773 | 0.578 | 0.761 | 2.760 | 0.00698 | 0.31188 | -2.481 | PIH1D2 | PIH1 domain containing 2 [Source:HGNC Symbol;Acc:HGNC:25210] |
| ENSG00000262526 | -0.444 | 5.723 | -2.760 | 0.00698 | 0.31188 | -2.585 | AC120057.3 | |
| ENSG00000167674 | 0.487 | 4.500 | 2.759 | 0.00698 | 0.31188 | -2.563 | AC011498.1 | Hepatoma-derived growth factor-related protein 2 [Source:UniProtKB/Swiss-Prot;Acc:Q7Z4V5] |
| ENSG00000145494 | 0.543 | 5.061 | 2.756 | 0.00705 | 0.31270 | -2.594 | NDUFS6 | NADH:ubiquinone oxidoreductase subunit S6 [Source:HGNC Symbol;Acc:HGNC:7713] |
| ENSG00000176619 | 0.558 | 5.443 | 2.756 | 0.00705 | 0.31270 | -2.604 | LMNB2 | lamin B2 [Source:HGNC Symbol;Acc:HGNC:6638] |
| ENSG00000140307 | 0.437 | 4.859 | 2.753 | 0.00711 | 0.31408 | -2.592 | GTF2A2 | general transcription factor IIA subunit 2 [Source:HGNC Symbol;Acc:HGNC:4647] |
| ENSG00000174307 | -1.422 | 0.777 | -2.749 | 0.00720 | 0.31691 | -2.720 | PHLDA3 | pleckstrin homology like domain family A member 3 [Source:HGNC Symbol;Acc:HGNC:8934] |
| ENSG00000072134 | -0.593 | 4.232 | -2.745 | 0.00726 | 0.31883 | -2.533 | EPN2 | epsin 2 [Source:HGNC Symbol;Acc:HGNC:18639] |
| ENSG00000105171 | 0.357 | 5.166 | 2.741 | 0.00735 | 0.32160 | -2.630 | POP4 | POP4 homolog, ribonuclease P/MRP subunit [Source:HGNC Symbol;Acc:HGNC:30081] |
| ENSG00000040275 | 0.617 | 3.236 | 2.740 | 0.00738 | 0.32172 | -2.534 | SPDL1 | spindle apparatus coiled-coil protein 1 [Source:HGNC Symbol;Acc:HGNC:26010] |
| ENSG00000184988 | -0.683 | 2.987 | -2.738 | 0.00741 | 0.32192 | -2.469 | TMEM106A | transmembrane protein 106A [Source:HGNC Symbol;Acc:HGNC:28288] |
| ENSG00000079257 | 0.443 | 3.290 | 2.737 | 0.00744 | 0.32223 | -2.535 | LXN | latexin [Source:HGNC Symbol;Acc:HGNC:13347] |
| ENSG00000174442 | 0.529 | 4.531 | 2.732 | 0.00754 | 0.32387 | -2.632 | ZWILCH | zwilch kinetochore protein [Source:HGNC Symbol;Acc:HGNC:25468] |
| ENSG00000183066 | -0.717 | 2.238 | -2.730 | 0.00758 | 0.32387 | -2.490 | WBP2NL | WBP2 N-terminal like [Source:HGNC Symbol;Acc:HGNC:28389] |
| ENSG00000029725 | -0.518 | 6.666 | -2.730 | 0.00759 | 0.32387 | -2.672 | RABEP1 | rabaptin, RAB GTPase binding effector protein 1 [Source:HGNC Symbol;Acc:HGNC:17677] |
| ENSG00000112511 | -0.639 | 6.165 | -2.730 | 0.00760 | 0.32387 | -2.666 | PHF1 | PHD finger protein 1 [Source:HGNC Symbol;Acc:HGNC:8919] |
| ENSG00000221867 | 2.350 | -0.890 | 2.729 | 0.00761 | 0.32387 | -2.578 | MAGEA3 | MAGE family member A3 [Source:HGNC Symbol;Acc:HGNC:6801] |
| ENSG00000136522 | 0.444 | 5.048 | 2.727 | 0.00766 | 0.32488 | -2.663 | MRPL47 | mitochondrial ribosomal protein L47 [Source:HGNC Symbol;Acc:HGNC:16652] |
| ENSG00000134317 | -1.060 | 3.778 | -2.726 | 0.00768 | 0.32488 | -2.520 | GRHL1 | grainyhead like transcription factor 1 [Source:HGNC Symbol;Acc:HGNC:17923] |
| ENSG00000189180 | -0.435 | 6.350 | -2.723 | 0.00775 | 0.32652 | -2.688 | ZNF33A | zinc finger protein 33A [Source:HGNC Symbol;Acc:HGNC:13096] |
| ENSG00000161692 | 0.660 | 1.234 | 2.717 | 0.00787 | 0.33079 | -2.516 | DBF4B | DBF4 zinc finger B [Source:HGNC Symbol;Acc:HGNC:17883] |
| ENSG00000138674 | -0.363 | 8.121 | -2.711 | 0.00799 | 0.33472 | -2.688 | SEC31A | SEC31 homolog A, COPII coat complex component [Source:HGNC Symbol;Acc:HGNC:17052] |
| ENSG00000136108 | 0.640 | 4.942 | 2.706 | 0.00811 | 0.33698 | -2.714 | CKAP2 | cytoskeleton associated protein 2 [Source:HGNC Symbol;Acc:HGNC:1990] |
| ENSG00000072778 | -0.529 | 7.992 | -2.704 | 0.00815 | 0.33698 | -2.712 | ACADVL | acyl-CoA dehydrogenase, very long chain [Source:HGNC Symbol;Acc:HGNC:92] |
| ENSG00000074755 | -0.470 | 6.485 | -2.704 | 0.00816 | 0.33698 | -2.733 | ZZEF1 | zinc finger ZZ-type and EF-hand domain containing 1 [Source:HGNC Symbol;Acc:HGNC:29027] |
| ENSG00000143995 | 1.741 | 1.674 | 2.704 | 0.00816 | 0.33698 | -2.551 | MEIS1 | Meis homeobox 1 [Source:HGNC Symbol;Acc:HGNC:7000] |
| ENSG00000171241 | 0.920 | 4.065 | 2.703 | 0.00818 | 0.33698 | -2.689 | SHCBP1 | SHC binding and spindle associated 1 [Source:HGNC Symbol;Acc:HGNC:29547] |
| ENSG00000131373 | 0.339 | 5.300 | 2.700 | 0.00825 | 0.33868 | -2.733 | HACL1 | 2-hydroxyacyl-CoA lyase 1 [Source:HGNC Symbol;Acc:HGNC:17856] |
| ENSG00000170871 | -0.488 | 6.058 | -2.695 | 0.00837 | 0.34279 | -2.750 | KIAA0232 | KIAA0232 [Source:HGNC Symbol;Acc:HGNC:28992] |
| ENSG00000165312 | -0.951 | 7.865 | -2.692 | 0.00844 | 0.34441 | -2.752 | OTUD1 | OTU deubiquitinase 1 [Source:HGNC Symbol;Acc:HGNC:27346] |
| ENSG00000093072 | -1.040 | 6.285 | -2.690 | 0.00849 | 0.34538 | -2.757 | ADA2 | adenosine deaminase 2 [Source:HGNC Symbol;Acc:HGNC:1839] |
| ENSG00000142546 | 0.376 | 4.611 | 2.687 | 0.00855 | 0.34686 | -2.738 | NOSIP | nitric oxide synthase interacting protein [Source:HGNC Symbol;Acc:HGNC:17946] |
| ENSG00000144843 | 0.769 | 3.265 | 2.686 | 0.00858 | 0.34695 | -2.667 | ADPRH | ADP-ribosylarginine hydrolase [Source:HGNC Symbol;Acc:HGNC:269] |
| ENSG00000140284 | 1.314 | 0.172 | 2.682 | 0.00867 | 0.34880 | -2.649 | SLC27A2 | solute carrier family 27 member 2 [Source:HGNC Symbol;Acc:HGNC:10996] |
| ENSG00000256100 | 0.383 | 3.702 | 2.681 | 0.00868 | 0.34880 | -2.693 | AP000721.1 | |
| ENSG00000163913 | 0.432 | 4.148 | 2.677 | 0.00879 | 0.35175 | -2.737 | IFT122 | intraflagellar transport 122 [Source:HGNC Symbol;Acc:HGNC:13556] |
| ENSG00000111859 | -0.802 | 6.421 | -2.676 | 0.00882 | 0.35207 | -2.796 | NEDD9 | neural precursor cell expressed, developmentally down-regulated 9 [Source:HGNC Symbol;Acc:HGNC:7733] |
| ENSG00000204519 | 0.520 | 3.932 | 2.670 | 0.00895 | 0.35523 | -2.742 | ZNF551 | zinc finger protein 551 [Source:HGNC Symbol;Acc:HGNC:25108] |
| ENSG00000169231 | 0.608 | 3.166 | 2.669 | 0.00899 | 0.35523 | -2.690 | THBS3 | thrombospondin 3 [Source:HGNC Symbol;Acc:HGNC:11787] |
| ENSG00000125910 | 1.628 | 1.010 | 2.666 | 0.00906 | 0.35523 | -2.617 | S1PR4 | sphingosine-1-phosphate receptor 4 [Source:HGNC Symbol;Acc:HGNC:3170] |
| ENSG00000077348 | 0.603 | 2.823 | 2.665 | 0.00908 | 0.35523 | -2.673 | EXOSC5 | exosome component 5 [Source:HGNC Symbol;Acc:HGNC:24662] |
| ENSG00000164114 | 1.292 | 3.605 | 2.665 | 0.00908 | 0.35523 | -2.761 | MAP9 | microtubule associated protein 9 [Source:HGNC Symbol;Acc:HGNC:26118] |
| ENSG00000196678 | 0.546 | 3.625 | 2.665 | 0.00910 | 0.35523 | -2.731 | ERI2 | ERI1 exoribonuclease family member 2 [Source:HGNC Symbol;Acc:HGNC:30541] |
| ENSG00000176681 | -0.738 | 2.426 | -2.664 | 0.00910 | 0.35523 | -2.622 | LRRC37A | leucine rich repeat containing 37A [Source:HGNC Symbol;Acc:HGNC:29069] |
| ENSG00000049541 | 0.505 | 3.084 | 2.663 | 0.00913 | 0.35523 | -2.696 | RFC2 | replication factor C subunit 2 [Source:HGNC Symbol;Acc:HGNC:9970] |
| ENSG00000101945 | 0.504 | 2.399 | 2.663 | 0.00915 | 0.35523 | -2.648 | SUV39H1 | suppressor of variegation 3-9 homolog 1 [Source:HGNC Symbol;Acc:HGNC:11479] |
| ENSG00000085274 | 0.356 | 5.706 | 2.661 | 0.00918 | 0.35543 | -2.834 | MYNN | myoneurin [Source:HGNC Symbol;Acc:HGNC:14955] |
| ENSG00000072506 | 0.544 | 4.570 | 2.659 | 0.00925 | 0.35694 | -2.809 | HSD17B10 | hydroxysteroid 17-beta dehydrogenase 10 [Source:HGNC Symbol;Acc:HGNC:4800] |
| ENSG00000078596 | -1.424 | 5.538 | -2.657 | 0.00930 | 0.35779 | -2.783 | ITM2A | integral membrane protein 2A [Source:HGNC Symbol;Acc:HGNC:6173] |
| ENSG00000100364 | -0.587 | 5.609 | -2.655 | 0.00933 | 0.35788 | -2.828 | KIAA0930 | KIAA0930 [Source:HGNC Symbol;Acc:HGNC:1314] |
| ENSG00000138771 | 2.258 | 2.291 | 2.653 | 0.00940 | 0.35915 | -2.723 | SHROOM3 | shroom family member 3 [Source:HGNC Symbol;Acc:HGNC:30422] |
| ENSG00000172404 | -0.776 | 0.334 | -2.652 | 0.00942 | 0.35915 | -2.877 | DNAJB7 | DnaJ heat shock protein family (Hsp40) member B7 [Source:HGNC Symbol;Acc:HGNC:24986] |
| ENSG00000105639 | 0.865 | 4.467 | 2.647 | 0.00955 | 0.36215 | -2.840 | JAK3 | Janus kinase 3 [Source:HGNC Symbol;Acc:HGNC:6193] |
| ENSG00000092470 | 0.686 | 4.352 | 2.646 | 0.00957 | 0.36215 | -2.831 | WDR76 | WD repeat domain 76 [Source:HGNC Symbol;Acc:HGNC:25773] |
| ENSG00000162613 | -0.397 | 8.084 | -2.645 | 0.00960 | 0.36215 | -2.846 | FUBP1 | far upstream element binding protein 1 [Source:HGNC Symbol;Acc:HGNC:4004] |
| ENSG00000101811 | 0.457 | 3.277 | 2.644 | 0.00963 | 0.36215 | -2.752 | CSTF2 | cleavage stimulation factor subunit 2 [Source:HGNC Symbol;Acc:HGNC:2484] |
| ENSG00000102900 | 0.526 | 4.303 | 2.644 | 0.00964 | 0.36215 | -2.830 | NUP93 | nucleoporin 93 [Source:HGNC Symbol;Acc:HGNC:28958] |
| ENSG00000178562 | 1.540 | 4.349 | 2.641 | 0.00970 | 0.36316 | -2.860 | CD28 | CD28 molecule [Source:HGNC Symbol;Acc:HGNC:1653] |
| ENSG00000144278 | 1.366 | 1.335 | 2.634 | 0.00989 | 0.36862 | -2.686 | GALNT13 | polypeptide N-acetylgalactosaminyltransferase 13 [Source:HGNC Symbol;Acc:HGNC:23242] |
| ENSG00000013288 | -0.541 | 5.123 | -2.634 | 0.00990 | 0.36862 | -2.858 | MAN2B2 | mannosidase alpha class 2B member 2 [Source:HGNC Symbol;Acc:HGNC:29623] |
| ENSG00000130332 | 0.435 | 3.519 | 2.632 | 0.00994 | 0.36862 | -2.796 | LSM7 | LSM7 homolog, U6 small nuclear RNA and mRNA degradation associated [Source:HGNC Symbol;Acc:HGNC:20470] |
| ENSG00000198258 | 0.486 | 5.410 | 2.629 | 0.01004 | 0.36862 | -2.906 | UBL5 | ubiquitin like 5 [Source:HGNC Symbol;Acc:HGNC:13736] |
| ENSG00000025423 | 0.733 | 1.269 | 2.628 | 0.01005 | 0.36862 | -2.700 | HSD17B6 | hydroxysteroid 17-beta dehydrogenase 6 [Source:HGNC Symbol;Acc:HGNC:23316] |
| ENSG00000172349 | -0.688 | 6.725 | -2.628 | 0.01006 | 0.36862 | -2.914 | IL16 | interleukin 16 [Source:HGNC Symbol;Acc:HGNC:5980] |
| ENSG00000112624 | -0.365 | 5.093 | -2.628 | 0.01006 | 0.36862 | -2.877 | BICRAL | BRD4 interacting chromatin remodelling complex associated protein like [Source:HGNC Symbol;Acc:HGNC:21111] |
| ENSG00000141503 | -0.549 | 4.423 | -2.628 | 0.01007 | 0.36862 | -2.827 | MINK1 | misshapen like kinase 1 [Source:HGNC Symbol;Acc:HGNC:17565] |
| ENSG00000124782 | -0.339 | 6.131 | -2.625 | 0.01015 | 0.36954 | -2.919 | RREB1 | ras responsive element binding protein 1 [Source:HGNC Symbol;Acc:HGNC:10449] |
| ENSG00000163162 | -0.521 | 6.586 | -2.625 | 0.01016 | 0.36954 | -2.922 | RNF149 | ring finger protein 149 [Source:HGNC Symbol;Acc:HGNC:23137] |
| ENSG00000163918 | 0.448 | 4.657 | 2.623 | 0.01019 | 0.36972 | -2.892 | RFC4 | replication factor C subunit 4 [Source:HGNC Symbol;Acc:HGNC:9972] |
| ENSG00000173852 | 0.645 | 6.159 | 2.622 | 0.01022 | 0.36972 | -2.927 | DPY19L1 | dpy-19 like 1 [Source:HGNC Symbol;Acc:HGNC:22205] |
| ENSG00000267059 | 0.455 | 5.272 | 2.621 | 0.01025 | 0.36982 | -2.921 | AC005943.1 | |
| ENSG00000104738 | 0.771 | 5.325 | 2.620 | 0.01029 | 0.37003 | -2.928 | MCM4 | minichromosome maintenance complex component 4 [Source:HGNC Symbol;Acc:HGNC:6947] |
| ENSG00000100567 | 0.432 | 6.530 | 2.617 | 0.01037 | 0.37106 | -2.938 | PSMA3 | proteasome subunit alpha 3 [Source:HGNC Symbol;Acc:HGNC:9532] |
| ENSG00000173011 | -0.464 | 5.045 | -2.616 | 0.01039 | 0.37106 | -2.898 | TADA2B | transcriptional adaptor 2B [Source:HGNC Symbol;Acc:HGNC:30781] |

| | | | | | | | | |
|---|---|---|---|---|---|---|---|---|
| ENSG00000243696 | 0.651 | 2.800 | 2.616 | 0.01040 | 0.37106 | -2.785 | AC006254.1 | |
| ENSG00000105671 | 0.517 | 4.799 | 2.614 | 0.01046 | 0.37115 | -2.924 | DDX49 | DEAD-box helicase 49 [Source:HGNC Symbol;Acc:HGNC:18684] |
| ENSG00000172167 | 0.531 | 3.840 | 2.613 | 0.01048 | 0.37115 | -2.867 | MTBP | MDM2 binding protein [Source:HGNC Symbol;Acc:HGNC:7417] |
| ENSG00000183856 | 1.125 | 1.547 | 2.613 | 0.01049 | 0.37115 | -2.735 | IQGAP3 | IQ motif containing GTPase activating protein 3 [Source:HGNC Symbol;Acc:HGNC:20669] |
| ENSG00000119559 | 0.490 | 2.492 | 2.609 | 0.01059 | 0.37349 | -2.769 | C19orf25 | chromosome 19 open reading frame 25 [Source:HGNC Symbol;Acc:HGNC:26711] |
| ENSG00000267157 | 0.661 | 0.957 | 2.608 | 0.01062 | 0.37349 | -2.756 | AC011499.1 | |
| ENSG00000163312 | -0.444 | 4.818 | -2.604 | 0.01072 | 0.37349 | -2.910 | HELQ | helicase, POLQ-like [Source:HGNC Symbol;Acc:HGNC:18536] |
| ENSG00000153044 | 0.649 | 3.488 | 2.604 | 0.01074 | 0.37349 | -2.868 | CENPH | centromere protein H [Source:HGNC Symbol;Acc:HGNC:17268] |
| ENSG00000119471 | -0.616 | 5.068 | -2.604 | 0.01075 | 0.37349 | -2.920 | HSDL2 | hydroxysteroid dehydrogenase like 2 [Source:HGNC Symbol;Acc:HGNC:18572] |
| ENSG00000013275 | 0.348 | 6.192 | 2.603 | 0.01077 | 0.37349 | -2.973 | PSMC4 | proteasome 26S subunit, ATPase 4 [Source:HGNC Symbol;Acc:HGNC:9551] |
| ENSG00000035720 | -1.722 | 5.100 | -2.601 | 0.01083 | 0.37349 | -2.868 | STAP1 | signal transducing adaptor family member 1 [Source:HGNC Symbol;Acc:HGNC:24133] |
| ENSG00000010803 | -0.459 | 4.405 | -2.601 | 0.01084 | 0.37349 | -2.889 | SCMH1 | sex comb on midleg homolog 1 (Drosophila) [Source:HGNC Symbol;Acc:HGNC:19003] |
| ENSG00000108523 | -0.377 | 4.954 | -2.600 | 0.01087 | 0.37349 | -2.934 | RNF167 | ring finger protein 167 [Source:HGNC Symbol;Acc:HGNC:24544] |
| ENSG00000266953 | 0.440 | 3.715 | 2.598 | 0.01092 | 0.37349 | -2.891 | AC092073.1 | |
| ENSG00000141027 | -0.361 | 7.359 | -2.598 | 0.01093 | 0.37349 | -2.978 | NCOR1 | nuclear receptor corepressor 1 [Source:HGNC Symbol;Acc:HGNC:7672] |
| ENSG00000175643 | 0.883 | 0.844 | 2.596 | 0.01097 | 0.37349 | -2.778 | RMI2 | RecQ mediated genome instability 2 [Source:HGNC Symbol;Acc:HGNC:28349] |
| ENSG00000176994 | -0.366 | 5.956 | -2.596 | 0.01097 | 0.37349 | -2.982 | SMCR8 | Smith-Magenis syndrome chromosome region, candidate 8 [Source:HGNC Symbol;Acc:HGNC:17921] |
| ENSG00000163833 | 0.867 | 0.661 | 2.596 | 0.01098 | 0.37349 | -2.795 | FBXO40 | F-box protein 40 [Source:HGNC Symbol;Acc:HGNC:29816] |
| ENSG00000167645 | 0.626 | 2.750 | 2.595 | 0.01100 | 0.37349 | -2.829 | YIF1B | Yip1 interacting factor homolog B, membrane trafficking protein [Source:HGNC Symbol;Acc:HGNC:30511] |
| ENSG00000261740 | 0.564 | 2.116 | 2.589 | 0.01118 | 0.37762 | -2.791 | AC133555.5 | BOLA2-SMG1P6 protein isoform a [Source:RefSeq peptide;Acc:NP_001307551] |
| ENSG00000196236 | -0.514 | 3.815 | -2.589 | 0.01120 | 0.37762 | -2.869 | XPNPEP3 | X-prolyl aminopeptidase 3 [Source:HGNC Symbol;Acc:HGNC:28052] |
| ENSG00000121022 | 0.329 | 5.707 | 2.587 | 0.01125 | 0.37762 | -3.007 | COPS5 | COP9 signalosome subunit 5 [Source:HGNC Symbol;Acc:HGNC:2240] |
| ENSG00000130816 | 0.592 | 6.084 | 2.586 | 0.01126 | 0.37762 | -3.011 | DNMT1 | DNA methyltransferase 1 [Source:HGNC Symbol;Acc:HGNC:2976] |
| ENSG00000171848 | 1.071 | 4.831 | 2.586 | 0.01126 | 0.37762 | -2.998 | RRM2 | ribonucleotide reductase regulatory subunit M2 [Source:HGNC Symbol;Acc:HGNC:10452] |
| ENSG00000036448 | 1.694 | -0.422 | 2.582 | 0.01140 | 0.38070 | -2.881 | MYOM2 | myomesin 2 [Source:HGNC Symbol;Acc:HGNC:7614] |
| ENSG00000181938 | 0.670 | 1.593 | 2.581 | 0.01142 | 0.38070 | -2.796 | GINS3 | GINS complex subunit 3 [Source:HGNC Symbol;Acc:HGNC:25851] |
| ENSG00000139174 | -0.832 | 3.162 | -2.580 | 0.01146 | 0.38070 | -2.820 | PRICKLE1 | prickle planar cell polarity protein 1 [Source:HGNC Symbol;Acc:HGNC:17019] |
| ENSG00000151725 | 0.757 | 5.137 | 2.579 | 0.01148 | 0.38070 | -3.018 | CENPU | centromere protein U [Source:HGNC Symbol;Acc:HGNC:21348] |
| ENSG00000159388 | -0.703 | 9.646 | -2.578 | 0.01153 | 0.38113 | -2.956 | BTG2 | BTG anti-proliferation factor 2 [Source:HGNC Symbol;Acc:HGNC:1131] |
| ENSG00000183878 | 2.618 | 1.570 | 2.577 | 0.01155 | 0.38113 | -2.852 | UTY | ubiquitously transcribed tetratricopeptide repeat containing, Y-linked [Source:HGNC Symbol;Acc:HGNC:12638] |
| ENSG00000055955 | 0.671 | 2.585 | 2.573 | 0.01167 | 0.38337 | -2.863 | ITIH4 | inter-alpha-trypsin inhibitor heavy chain family member 4 [Source:HGNC Symbol;Acc:HGNC:6169] |
| ENSG00000152240 | 0.416 | 3.710 | 2.573 | 0.01168 | 0.38337 | -2.946 | HAUS1 | HAUS augmin like complex subunit 1 [Source:HGNC Symbol;Acc:HGNC:25174] |
| ENSG00000106246 | 0.339 | 5.073 | 2.570 | 0.01178 | 0.38375 | -3.031 | PTCD1 | pentatricopeptide repeat domain 1 [Source:HGNC Symbol;Acc:HGNC:22198] |
| ENSG00000119979 | -0.452 | 4.170 | -2.570 | 0.01179 | 0.38375 | -2.941 | FAM45A | family with sequence similarity 45 member A [Source:HGNC Symbol;Acc:HGNC:31793] |
| ENSG00000176463 | 1.406 | 2.522 | 2.568 | 0.01183 | 0.38375 | -2.904 | SLCO3A1 | solute carrier organic anion transporter family member 3A1 [Source:HGNC Symbol;Acc:HGNC:10952] |
| ENSG00000100911 | 0.532 | 6.876 | 2.567 | 0.01187 | 0.38375 | -3.045 | PSME2 | proteasome activator subunit 2 [Source:HGNC Symbol;Acc:HGNC:9569] |
| ENSG00000133119 | 0.616 | 3.039 | 2.567 | 0.01188 | 0.38375 | -2.915 | RFC3 | replication factor C subunit 3 [Source:HGNC Symbol;Acc:HGNC:9971] |
| ENSG00000179115 | 0.492 | 4.927 | 2.565 | 0.01194 | 0.38375 | -3.041 | FARSA | phenylalanyl-tRNA synthetase alpha subunit [Source:HGNC Symbol;Acc:HGNC:3592] |
| ENSG00000162063 | 0.833 | 1.667 | 2.564 | 0.01196 | 0.38375 | -2.836 | CCNF | cyclin F [Source:HGNC Symbol;Acc:HGNC:1591] |
| ENSG00000087470 | 0.338 | 6.453 | 2.564 | 0.01196 | 0.38375 | -3.062 | DNM1L | dynamin 1 like [Source:HGNC Symbol;Acc:HGNC:2973] |
| ENSG00000150779 | 0.704 | 4.229 | 2.564 | 0.01198 | 0.38375 | -3.015 | TIMM8B | translocase of inner mitochondrial membrane 8 homolog B [Source:HGNC Symbol;Acc:HGNC:11818] |
| ENSG00000188177 | -0.650 | 5.359 | -2.563 | 0.01199 | 0.38375 | -3.026 | ZC3H6 | zinc finger CCCH-type containing 6 [Source:HGNC Symbol;Acc:HGNC:24762] |
| ENSG00000112584 | -0.412 | 4.660 | -2.560 | 0.01210 | 0.38647 | -3.005 | FAM120B | family with sequence similarity 120B [Source:HGNC Symbol;Acc:HGNC:21109] |
| ENSG00000174842 | 0.478 | 3.109 | 2.556 | 0.01224 | 0.38874 | -2.937 | GLMN | glomulin, FKBP associated protein [Source:HGNC Symbol;Acc:HGNC:14373] |
| ENSG00000105655 | 0.724 | 1.967 | 2.555 | 0.01226 | 0.38874 | -2.864 | ISYNA1 | inositol-3-phosphate synthase 1 [Source:HGNC Symbol;Acc:HGNC:29821] |
| ENSG00000124207 | 0.481 | 6.043 | 2.555 | 0.01227 | 0.38874 | -3.084 | CSE1L | chromosome segregation 1 like [Source:HGNC Symbol;Acc:HGNC:2431] |
| ENSG00000110104 | 0.681 | 2.685 | 2.551 | 0.01237 | 0.39112 | -2.920 | CCDC86 | coiled-coil domain containing 86 [Source:HGNC Symbol;Acc:HGNC:28359] |
| ENSG00000134744 | -0.511 | 7.167 | -2.550 | 0.01243 | 0.39117 | -3.092 | ZCCHC11 | zinc finger CCHC-type containing 11 [Source:HGNC Symbol;Acc:HGNC:28981] |
| ENSG00000145241 | -0.500 | 7.360 | -2.548 | 0.01250 | 0.39117 | -3.094 | CENPC | centromere protein C [Source:HGNC Symbol;Acc:HGNC:1854] |
| ENSG00000106367 | 0.520 | 2.419 | 2.547 | 0.01252 | 0.39117 | -2.902 | AP1S1 | adaptor related protein complex 1 sigma 1 subunit [Source:HGNC Symbol;Acc:HGNC:559] |
| ENSG00000133138 | 1.607 | 0.273 | 2.547 | 0.01253 | 0.39117 | -2.885 | TBC1D8B | TBC1 domain family member 8B [Source:HGNC Symbol;Acc:HGNC:24715] |
| ENSG00000068394 | 0.339 | 4.121 | 2.547 | 0.01254 | 0.39117 | -3.032 | GPKOW | G-patch domain and KOW motifs [Source:HGNC Symbol;Acc:HGNC:30677] |
| ENSG00000141510 | -0.685 | 3.562 | -2.546 | 0.01256 | 0.39117 | -2.931 | TP53 | tumor protein p53 [Source:HGNC Symbol;Acc:HGNC:11998] |
| ENSG00000142687 | -0.351 | 6.145 | -2.543 | 0.01265 | 0.39324 | -3.107 | KIAA0319L | KIAA0319 like [Source:HGNC Symbol;Acc:HGNC:30071] |
| ENSG00000112118 | 0.635 | 5.685 | 2.540 | 0.01274 | 0.39404 | -3.116 | MCM3 | minichromosome maintenance complex component 3 [Source:HGNC Symbol;Acc:HGNC:6945] |
| ENSG00000161956 | -0.389 | 4.904 | -2.540 | 0.01275 | 0.39404 | -3.066 | SENP3 | SUMO1/sentrin/SMT3 specific peptidase 3 [Source:HGNC Symbol;Acc:HGNC:17862] |
| ENSG00000044459 | 1.171 | 4.038 | 2.539 | 0.01279 | 0.39404 | -3.075 | CNTLN | centlein [Source:HGNC Symbol;Acc:HGNC:23432] |
| ENSG00000120738 | -1.129 | 6.269 | -2.539 | 0.01280 | 0.39404 | -3.107 | EGR1 | early growth response 1 [Source:HGNC Symbol;Acc:HGNC:3238] |
| ENSG00000213967 | 1.110 | 1.186 | 2.536 | 0.01288 | 0.39468 | -2.886 | ZNF726 | zinc finger protein 726 [Source:HGNC Symbol;Acc:HGNC:32462] |
| ENSG00000105676 | 0.621 | 3.503 | 2.534 | 0.01295 | 0.39468 | -3.027 | ARMC6 | armadillo repeat containing 6 [Source:HGNC Symbol;Acc:HGNC:25049] |
| ENSG00000062822 | 0.580 | 3.612 | 2.534 | 0.01296 | 0.39468 | -3.036 | POLD1 | DNA polymerase delta 1, catalytic subunit [Source:HGNC Symbol;Acc:HGNC:9175] |
| ENSG00000125731 | 0.586 | 3.045 | 2.534 | 0.01297 | 0.39468 | -2.984 | SH2D3A | SH2 domain containing 3A [Source:HGNC Symbol;Acc:HGNC:16885] |
| ENSG00000145349 | -0.960 | 6.011 | -2.534 | 0.01298 | 0.39468 | -3.114 | CAMK2D | calcium/calmodulin dependent protein kinase II delta [Source:HGNC Symbol;Acc:HGNC:1462] |
| ENSG00000172613 | 0.387 | 4.090 | 2.531 | 0.01305 | 0.39548 | -3.065 | RAD9A | RAD9 checkpoint clamp component A [Source:HGNC Symbol;Acc:HGNC:9827] |
| ENSG00000151503 | 0.501 | 5.385 | 2.530 | 0.01309 | 0.39548 | -3.133 | NCAPD3 | non-SMC condensin II complex subunit D3 [Source:HGNC Symbol;Acc:HGNC:28952] |
| ENSG00000046774 | 1.747 | 0.647 | 2.530 | 0.01310 | 0.39548 | -2.899 | MAGEC2 | MAGE family member C2 [Source:HGNC Symbol;Acc:HGNC:13574] |
| ENSG00000183134 | 0.823 | 0.394 | 2.528 | 0.01318 | 0.39700 | -2.941 | PTGDR2 | prostaglandin D2 receptor 2 [Source:HGNC Symbol;Acc:HGNC:4502] |
| ENSG00000198015 | 0.428 | 6.132 | 2.524 | 0.01329 | 0.39953 | -3.153 | MRPL42 | mitochondrial ribosomal protein L42 [Source:HGNC Symbol;Acc:HGNC:14493] |
| ENSG00000154229 | -1.347 | 4.043 | -2.523 | 0.01336 | 0.40042 | -2.972 | PRKCA | protein kinase C alpha [Source:HGNC Symbol;Acc:HGNC:9393] |
| ENSG00000158406 | 0.582 | 4.561 | 2.514 | 0.01366 | 0.40702 | -3.141 | HIST1H4H | histone cluster 1 H4 family member h [Source:HGNC Symbol;Acc:HGNC:4788] |
| ENSG00000180879 | -0.590 | 8.707 | -2.514 | 0.01367 | 0.40702 | -3.130 | SSR4 | signal sequence receptor subunit 4 [Source:HGNC Symbol;Acc:HGNC:11326] |
| ENSG00000105393 | 0.334 | 5.472 | 2.514 | 0.01369 | 0.40702 | -3.171 | BABAM1 | BRISC and BRCA1 A complex member 1 [Source:HGNC Symbol;Acc:HGNC:25008] |
| ENSG00000126005 | -0.419 | 3.173 | -2.513 | 0.01372 | 0.40702 | -2.984 | MMP24-AS1 | MMP24 antisense RNA 1 [Source:HGNC Symbol;Acc:HGNC:44421] |
| ENSG00000274641 | 0.714 | 3.593 | 2.512 | 0.01374 | 0.40702 | -3.086 | HIST1H2BO | histone cluster 1 H2B family member o [Source:HGNC Symbol;Acc:HGNC:4758] |
| ENSG00000124787 | 0.713 | 2.564 | 2.511 | 0.01377 | 0.40702 | -2.994 | RPP40 | ribonuclease P/MRP subunit p40 [Source:HGNC Symbol;Acc:HGNC:20992] |
| ENSG00000268400 | 0.429 | 6.152 | 2.508 | 0.01388 | 0.40933 | -3.190 | AC008763.2 | |
| ENSG00000064726 | 0.397 | 6.704 | 2.503 | 0.01407 | 0.41414 | -3.196 | BTBD1 | BTB domain containing 1 [Source:HGNC Symbol;Acc:HGNC:1120] |
| ENSG00000147905 | -0.499 | 7.048 | -2.502 | 0.01412 | 0.41448 | -3.202 | ZCCHC7 | zinc finger CCHC-type containing 7 [Source:HGNC Symbol;Acc:HGNC:26209] |
| ENSG00000136518 | 0.432 | 5.584 | 2.501 | 0.01416 | 0.41468 | -3.202 | ACTL6A | actin like 6A [Source:HGNC Symbol;Acc:HGNC:24124] |
| ENSG00000108405 | -1.127 | 5.699 | -2.499 | 0.01422 | 0.41560 | -3.169 | P2RX1 | purinergic receptor P2X 1 [Source:HGNC Symbol;Acc:HGNC:8533] |
| ENSG00000130340 | -2.232 | 4.614 | -2.498 | 0.01427 | 0.41598 | -3.028 | SNX9 | sorting nexin 9 [Source:HGNC Symbol;Acc:HGNC:14973] |
| ENSG00000100354 | -0.453 | 7.163 | -2.495 | 0.01438 | 0.41843 | -3.216 | TNRC6B | trinucleotide repeat containing 6B [Source:HGNC Symbol;Acc:HGNC:29190] |
| ENSG00000138686 | 0.416 | 3.650 | 2.492 | 0.01449 | 0.42071 | -3.120 | BBS7 | Bardet-Biedl syndrome 7 [Source:HGNC Symbol;Acc:HGNC:18758] |
| ENSG00000215712 | -0.527 | 4.210 | -2.490 | 0.01458 | 0.42223 | -3.115 | TMEM242 | transmembrane protein 242 [Source:HGNC Symbol;Acc:HGNC:17206] |
| ENSG00000182670 | -0.422 | 8.478 | -2.486 | 0.01470 | 0.42235 | -3.196 | TTC3 | tetratricopeptide repeat domain 3 [Source:HGNC Symbol;Acc:HGNC:12393] |
| ENSG00000198554 | 0.692 | 3.604 | 2.486 | 0.01471 | 0.42235 | -3.146 | WDHD1 | WD repeat and HMG-box DNA binding protein 1 [Source:HGNC Symbol;Acc:HGNC:23170] |
| ENSG00000178127 | 0.392 | 6.899 | 2.486 | 0.01471 | 0.42235 | -3.230 | NDUFV2 | NADH:ubiquinone oxidoreductase core subunit V2 [Source:HGNC Symbol;Acc:HGNC:7717] |
| ENSG00000143106 | 0.456 | 6.219 | 2.485 | 0.01475 | 0.42235 | -3.241 | PSMA5 | proteasome subunit alpha 5 [Source:HGNC Symbol;Acc:HGNC:9534] |
| ENSG00000254673 | 0.375 | 4.207 | 2.484 | 0.01481 | 0.42235 | -3.179 | AC110275.1 | |
| ENSG00000079387 | 0.289 | 4.997 | 2.483 | 0.01482 | 0.42235 | -3.222 | SENP1 | SUMO1/sentrin specific peptidase 1 [Source:HGNC Symbol;Acc:HGNC:17927] |

| | | | | | | | | |
|---|---|---|---|---|---|---|---|---|
| ENSG00000198901 | 0.655 | 5.234 | 2.480 | 0.01496 | 0.42235 | -3.245 | PRC1 | protein regulator of cytokinesis 1 [Source:HGNC Symbol;Acc:HGNC:9341] |
| ENSG00000144229 | 1.483 | 0.607 | 2.480 | 0.01497 | 0.42235 | -3.002 | THSD7B | thrombospondin type 1 domain containing 7B [Source:HGNC Symbol;Acc:HGNC:29348] |
| ENSG00000274267 | 1.033 | 4.327 | 2.480 | 0.01497 | 0.42235 | -3.221 | HIST1H3B | histone cluster 1 H3 family member b [Source:HGNC Symbol;Acc:HGNC:4776] |
| ENSG00000127804 | -0.311 | 4.931 | -2.478 | 0.01502 | 0.42235 | -3.208 | METTL16 | methyltransferase like 16 [Source:HGNC Symbol;Acc:HGNC:28484] |
| ENSG00000125740 | -0.885 | 8.421 | -2.478 | 0.01502 | 0.42235 | -3.226 | FOSB | FosB proto-oncogene, AP-1 transcription factor subunit [Source:HGNC Symbol;Acc:HGNC:3797] |
| ENSG00000117226 | -0.667 | 5.026 | -2.478 | 0.01504 | 0.42235 | -3.201 | GBP3 | guanylate binding protein 3 [Source:HGNC Symbol;Acc:HGNC:4184] |
| ENSG00000156172 | 0.572 | 0.693 | 2.477 | 0.01506 | 0.42235 | -3.022 | C8orf37 | chromosome 8 open reading frame 37 [Source:HGNC Symbol;Acc:HGNC:27232] |
| ENSG00000123358 | -1.319 | 5.830 | -2.477 | 0.01509 | 0.42235 | -3.226 | NR4A1 | nuclear receptor subfamily 4 group A member 1 [Source:HGNC Symbol;Acc:HGNC:7980] |
| ENSG00000111665 | 0.960 | 1.506 | 2.476 | 0.01511 | 0.42235 | -3.013 | CDCA3 | cell division cycle associated 3 [Source:HGNC Symbol;Acc:HGNC:14624] |
| ENSG00000100650 | -0.487 | 8.873 | -2.476 | 0.01511 | 0.42235 | -3.205 | SRSF5 | serine and arginine rich splicing factor 5 [Source:HGNC Symbol;Acc:HGNC:10787] |
| ENSG00000125772 | -0.551 | 7.553 | -2.475 | 0.01515 | 0.42262 | -3.254 | GPCPD1 | glycerophosphocholine phosphodiesterase 1 [Source:HGNC Symbol;Acc:HGNC:26957] |
| ENSG00000230989 | 0.499 | 5.468 | 2.473 | 0.01523 | 0.42268 | -3.264 | HSBP1 | heat shock factor binding protein 1 [Source:HGNC Symbol;Acc:HGNC:5203] |
| ENSG00000171763 | 0.434 | 3.320 | 2.473 | 0.01524 | 0.42268 | -3.136 | SPATA5L1 | spermatogenesis associated 5 like 1 [Source:HGNC Symbol;Acc:HGNC:28762] |
| ENSG00000174652 | 0.478 | 4.878 | 2.472 | 0.01526 | 0.42268 | -3.247 | ZNF266 | zinc finger protein 266 [Source:HGNC Symbol;Acc:HGNC:13059] |
| ENSG00000148773 | 1.072 | 5.413 | 2.471 | 0.01531 | 0.42300 | -3.270 | MKI67 | marker of proliferation Ki-67 [Source:HGNC Symbol;Acc:HGNC:7107] |
| ENSG00000138735 | -0.722 | 5.097 | -2.470 | 0.01533 | 0.42300 | -3.216 | PDE5A | phosphodiesterase 5A [Source:HGNC Symbol;Acc:HGNC:8784] |
| ENSG00000169432 | 1.353 | 4.287 | 2.467 | 0.01545 | 0.42482 | -3.251 | SCN9A | sodium voltage-gated channel alpha subunit 9 [Source:HGNC Symbol;Acc:HGNC:10597] |
| ENSG00000132646 | 0.655 | 5.586 | 2.467 | 0.01547 | 0.42482 | -3.279 | PCNA | proliferating cell nuclear antigen [Source:HGNC Symbol;Acc:HGNC:8729] |
| ENSG00000125844 | -0.758 | 9.188 | -2.465 | 0.01557 | 0.42523 | -3.226 | RRBP1 | ribosome binding protein 1 [Source:HGNC Symbol;Acc:HGNC:10448] |
| ENSG00000110324 | -0.983 | 6.081 | -2.464 | 0.01558 | 0.42523 | -3.270 | IL10RA | interleukin 10 receptor subunit alpha [Source:HGNC Symbol;Acc:HGNC:5964] |
| ENSG00000172172 | 0.519 | 4.307 | 2.463 | 0.01563 | 0.42523 | -3.237 | MRPL13 | mitochondrial ribosomal protein L13 [Source:HGNC Symbol;Acc:HGNC:14278] |
| ENSG00000114054 | 0.374 | 5.452 | 2.462 | 0.01566 | 0.42523 | -3.285 | PCCB | propionyl-CoA carboxylase beta subunit [Source:HGNC Symbol;Acc:HGNC:8654] |
| ENSG00000132688 | 2.481 | 3.783 | 2.462 | 0.01567 | 0.42523 | -3.255 | NES | nestin [Source:HGNC Symbol;Acc:HGNC:7756] |
| ENSG00000101003 | 0.780 | 2.387 | 2.460 | 0.01574 | 0.42523 | -3.098 | GINS1 | GINS complex subunit 1 [Source:HGNC Symbol;Acc:HGNC:28980] |
| ENSG00000156876 | 0.521 | 3.786 | 2.460 | 0.01574 | 0.42523 | -3.204 | SASS6 | SAS-6 centriolar assembly protein [Source:HGNC Symbol;Acc:HGNC:25403] |
| ENSG00000120889 | -0.801 | 5.063 | -2.460 | 0.01575 | 0.42523 | -3.233 | TNFRSF10B | TNF receptor superfamily member 10b [Source:HGNC Symbol;Acc:HGNC:11905] |
| ENSG00000163655 | 0.394 | 5.571 | 2.459 | 0.01581 | 0.42591 | -3.296 | GMPS | guanine monophosphate synthase [Source:HGNC Symbol;Acc:HGNC:4378] |
| ENSG00000139372 | 0.325 | 5.049 | 2.457 | 0.01588 | 0.42591 | -3.283 | TDG | thymine DNA glycosylase [Source:HGNC Symbol;Acc:HGNC:11700] |
| ENSG00000090581 | -0.487 | 5.309 | -2.456 | 0.01592 | 0.42591 | -3.272 | GNPTG | N-acetylglucosamine-1-phosphate transferase gamma subunit [Source:HGNC Symbol;Acc:HGNC:23026] |
| ENSG00000118816 | -0.362 | 6.872 | -2.455 | 0.01595 | 0.42591 | -3.307 | CCNI | cyclin I [Source:HGNC Symbol;Acc:HGNC:1595] |
| ENSG00000126524 | -0.539 | 7.724 | -2.455 | 0.01597 | 0.42591 | -3.294 | SBDS | SBDS, ribosome assembly guanine nucleotide exchange factor [Source:HGNC Symbol;Acc:HGNC:19440] |
| ENSG00000004455 | 0.352 | 5.744 | 2.455 | 0.01597 | 0.42591 | -3.307 | AK2 | adenylate kinase 2 [Source:HGNC Symbol;Acc:HGNC:362] |
| ENSG00000198732 | -1.578 | 4.455 | -2.442 | 0.01650 | 0.43623 | -3.164 | SMOC1 | SPARC related modular calcium binding 1 [Source:HGNC Symbol;Acc:HGNC:20318] |
| ENSG00000228716 | 0.599 | 4.632 | 2.441 | 0.01655 | 0.43623 | -3.308 | DHFR | dihydrofolate reductase [Source:HGNC Symbol;Acc:HGNC:2861] |
| ENSG00000090621 | -0.526 | 8.782 | -2.441 | 0.01655 | 0.43623 | -3.286 | PABPC4 | poly(A) binding protein cytoplasmic 4 [Source:HGNC Symbol;Acc:HGNC:8557] |
| ENSG00000224383 | -0.572 | 4.435 | -2.441 | 0.01656 | 0.43623 | -3.241 | PRR29 | proline rich 29 [Source:HGNC Symbol;Acc:HGNC:25673] |
| ENSG00000171649 | 1.011 | 1.287 | 2.441 | 0.01657 | 0.43623 | -3.078 | ZIK1 | zinc finger protein interacting with K protein 1 [Source:HGNC Symbol;Acc:HGNC:33104] |
| ENSG00000214265 | -0.697 | 5.842 | -2.440 | 0.01661 | 0.43623 | -3.324 | AC124312.1 | |
| ENSG00000038219 | -0.421 | 7.962 | -2.440 | 0.01661 | 0.43623 | -3.318 | BOD1L1 | biorientation of chromosomes in cell division 1 like 1 [Source:HGNC Symbol;Acc:HGNC:31792] |
| ENSG00000109171 | -0.436 | 6.023 | -2.439 | 0.01663 | 0.43623 | -3.336 | SLAIN2 | SLAIN motif family member 2 [Source:HGNC Symbol;Acc:HGNC:29282] |
| ENSG00000170745 | 2.152 | 1.227 | 2.435 | 0.01679 | 0.43954 | -3.107 | KCNS3 | potassium voltage-gated channel modifier subfamily S member 3 [Source:HGNC Symbol;Acc:HGNC:6302] |
| ENSG00000139323 | -0.365 | 5.225 | -2.433 | 0.01692 | 0.44189 | -3.322 | POC1B | POC1 centriolar protein B [Source:HGNC Symbol;Acc:HGNC:30836] |
| ENSG00000269307 | 0.324 | 5.239 | 2.430 | 0.01705 | 0.44410 | -3.351 | AC010463.1 | |
| ENSG00000164045 | 0.734 | 2.621 | 2.429 | 0.01707 | 0.44410 | -3.177 | CDC25A | cell division cycle 25A [Source:HGNC Symbol;Acc:HGNC:1725] |
| ENSG00000149196 | 0.465 | 4.857 | 2.427 | 0.01717 | 0.44573 | -3.344 | HIKESHI | Hikeshi, heat shock protein nuclear import factor [Source:HGNC Symbol;Acc:HGNC:26938] |
| ENSG00000109881 | 0.705 | 2.753 | 2.422 | 0.01738 | 0.44993 | -3.207 | CCDC34 | coiled-coil domain containing 34 [Source:HGNC Symbol;Acc:HGNC:25079] |
| ENSG00000139350 | 0.387 | 5.004 | 2.421 | 0.01743 | 0.44993 | -3.361 | NEDD1 | neural precursor cell expressed, developmentally down-regulated 1 [Source:HGNC Symbol;Acc:HGNC:7723] |
| ENSG00000119950 | -0.537 | 6.331 | -2.420 | 0.01747 | 0.44993 | -3.382 | MXI1 | MAX interactor 1, dimerization protein [Source:HGNC Symbol;Acc:HGNC:7534] |
| ENSG00000083896 | -0.452 | 8.048 | -2.420 | 0.01747 | 0.44993 | -3.358 | YTHDC1 | YTH domain containing 1 [Source:HGNC Symbol;Acc:HGNC:30626] |
| ENSG00000071575 | -1.670 | 3.707 | -2.418 | 0.01756 | 0.45128 | -3.151 | TRIB2 | tribbles pseudokinase 2 [Source:HGNC Symbol;Acc:HGNC:30809] |
| ENSG00000105778 | 0.303 | 5.838 | 2.417 | 0.01764 | 0.45146 | -3.391 | AVL9 | AVL9 cell migration associated [Source:HGNC Symbol;Acc:HGNC:28994] |
| ENSG00000129595 | -0.613 | 5.785 | -2.417 | 0.01764 | 0.45146 | -3.374 | EPB41L4A | erythrocyte membrane protein band 4.1 like 4A [Source:HGNC Symbol;Acc:HGNC:13278] |
| ENSG00000172244 | 0.627 | 1.519 | 2.414 | 0.01777 | 0.45403 | -3.130 | C5orf34 | chromosome 5 open reading frame 34 [Source:HGNC Symbol;Acc:HGNC:24738] |
| ENSG00000138166 | -0.776 | 9.133 | -2.411 | 0.01788 | 0.45579 | -3.344 | DUSP5 | dual specificity phosphatase 5 [Source:HGNC Symbol;Acc:HGNC:3071] |
| ENSG00000038002 | -0.700 | 6.596 | -2.407 | 0.01806 | 0.45921 | -3.411 | AGA | aspartylglucosaminidase [Source:HGNC Symbol;Acc:HGNC:318] |
| ENSG00000131023 | -0.302 | 5.141 | -2.407 | 0.01808 | 0.45921 | -3.378 | LATS1 | large tumor suppressor kinase 1 [Source:HGNC Symbol;Acc:HGNC:6514] |
| ENSG00000127540 | 0.494 | 4.974 | 2.405 | 0.01817 | 0.45986 | -3.398 | UQCR11 | ubiquinol-cytochrome c reductase, complex III subunit XI [Source:HGNC Symbol;Acc:HGNC:30862] |
| ENSG00000168393 | 0.477 | 2.691 | 2.405 | 0.01818 | 0.45986 | -3.221 | DTYMK | deoxythymidylate kinase [Source:HGNC Symbol;Acc:HGNC:3061] |
| ENSG00000006625 | 0.596 | 4.144 | 2.403 | 0.01826 | 0.46091 | -3.362 | GGCT | gamma-glutamylcyclotransferase [Source:HGNC Symbol;Acc:HGNC:21705] |
| ENSG00000132423 | 0.519 | 1.934 | 2.399 | 0.01844 | 0.46464 | -3.175 | COQ3 | coenzyme Q3, methyltransferase [Source:HGNC Symbol;Acc:HGNC:18175] |
| ENSG00000113810 | 0.485 | 7.583 | 2.396 | 0.01858 | 0.46709 | -3.403 | SMC4 | structural maintenance of chromosomes 4 [Source:HGNC Symbol;Acc:HGNC:14013] |
| ENSG00000101160 | -0.501 | 7.149 | -2.392 | 0.01876 | 0.46929 | -3.442 | CTSZ | cathepsin Z [Source:HGNC Symbol;Acc:HGNC:2547] |
| ENSG00000144036 | -0.781 | 4.623 | -2.392 | 0.01879 | 0.46929 | -3.352 | EXOC6B | exocyst complex component 6B [Source:HGNC Symbol;Acc:HGNC:17085] |
| ENSG00000258472 | 0.596 | 2.918 | 2.392 | 0.01880 | 0.46929 | -3.280 | AC005726.2 | |
| ENSG00000240065 | 0.467 | 5.158 | 2.391 | 0.01881 | 0.46929 | -3.433 | PSMB9 | proteasome subunit beta 9 [Source:HGNC Symbol;Acc:HGNC:9546] |
| ENSG00000111057 | 1.553 | 0.690 | 2.388 | 0.01899 | 0.47290 | -3.173 | KRT18 | keratin 18 [Source:HGNC Symbol;Acc:HGNC:6430] |
| ENSG00000136492 | 0.660 | 3.731 | 2.386 | 0.01909 | 0.47311 | -3.370 | BRIP1 | BRCA1 interacting protein C-terminal helicase 1 [Source:HGNC Symbol;Acc:HGNC:20473] |
| ENSG00000147099 | 0.307 | 4.056 | 2.385 | 0.01913 | 0.47311 | -3.378 | HDAC8 | histone deacetylase 8 [Source:HGNC Symbol;Acc:HGNC:13315] |
| ENSG00000118900 | -0.292 | 5.767 | -2.384 | 0.01918 | 0.47311 | -3.453 | UBN1 | ubinuclein 1 [Source:HGNC Symbol;Acc:HGNC:12506] |
| ENSG00000186951 | -0.515 | 4.694 | -2.382 | 0.01926 | 0.47311 | -3.393 | PPARA | peroxisome proliferator activated receptor alpha [Source:HGNC Symbol;Acc:HGNC:9232] |
| ENSG00000183527 | 0.450 | 4.123 | 2.382 | 0.01928 | 0.47311 | -3.398 | PSMG1 | proteasome assembly chaperone 1 [Source:HGNC Symbol;Acc:HGNC:3043] |
| ENSG00000136319 | 0.388 | 3.908 | 2.382 | 0.01928 | 0.47311 | -3.377 | TTC5 | tetratricopeptide repeat domain 5 [Source:HGNC Symbol;Acc:HGNC:19274] |
| ENSG00000204520 | -0.585 | 3.213 | -2.381 | 0.01932 | 0.47311 | -3.246 | MICA | MHC class I polypeptide-related sequence A [Source:HGNC Symbol;Acc:HGNC:7090] |
| ENSG00000174886 | 0.446 | 4.574 | 2.381 | 0.01934 | 0.47311 | -3.431 | NDUFA11 | NADH:ubiquinone oxidoreductase subunit A11 [Source:HGNC Symbol;Acc:HGNC:20371] |
| ENSG00000177694 | 1.453 | 0.874 | 2.380 | 0.01938 | 0.47311 | -3.189 | NAALADL2 | N-acetylated alpha-linked acidic dipeptidase like 2 [Source:HGNC Symbol;Acc:HGNC:23219] |
| ENSG00000143977 | 0.445 | 4.939 | 2.379 | 0.01940 | 0.47311 | -3.451 | SNRPG | small nuclear ribonucleoprotein polypeptide G [Source:HGNC Symbol;Acc:HGNC:11163] |
| ENSG00000144468 | -0.368 | 6.239 | -2.379 | 0.01941 | 0.47311 | -3.471 | RHBDD1 | rhomboid domain containing 1 [Source:HGNC Symbol;Acc:HGNC:23081] |
| ENSG00000064961 | 0.345 | 4.834 | 2.378 | 0.01948 | 0.47391 | -3.447 | HMG20B | high mobility group 20B [Source:HGNC Symbol;Acc:HGNC:5002] |
| ENSG00000236383 | -0.746 | 1.115 | -2.377 | 0.01952 | 0.47396 | -3.224 | LINC00854 | long intergenic non-protein coding RNA 854 [Source:HGNC Symbol;Acc:HGNC:43658] |
| ENSG00000126457 | 0.457 | 6.441 | 2.375 | 0.01960 | 0.47419 | -3.480 | PRMT1 | protein arginine methyltransferase 1 [Source:HGNC Symbol;Acc:HGNC:5187] |
| ENSG00000165959 | -0.981 | 4.917 | -2.374 | 0.01966 | 0.47419 | -3.401 | CLMN | calmin [Source:HGNC Symbol;Acc:HGNC:19972] |
| ENSG00000074621 | 0.603 | 4.104 | 2.373 | 0.01970 | 0.47419 | -3.421 | SLC24A1 | solute carrier family 24 member 1 [Source:HGNC Symbol;Acc:HGNC:10975] |
| ENSG00000100908 | 0.331 | 3.186 | 2.373 | 0.01971 | 0.47419 | -3.321 | EMC9 | ER membrane protein complex subunit 9 [Source:HGNC Symbol;Acc:HGNC:20273] |
| ENSG00000082512 | -1.022 | 3.488 | -2.373 | 0.01971 | 0.47419 | -3.261 | TRAF5 | TNF receptor associated factor 5 [Source:HGNC Symbol;Acc:HGNC:12035] |
| ENSG00000167077 | -0.720 | 6.225 | -2.370 | 0.01987 | 0.47704 | -3.485 | MEI1 | meiotic double-stranded break formation protein 1 [Source:HGNC Symbol;Acc:HGNC:28613] |
| ENSG00000145908 | 1.115 | 2.842 | 2.367 | 0.02000 | 0.47923 | -3.354 | ZNF300 | zinc finger protein 300 [Source:HGNC Symbol;Acc:HGNC:13091] |
| ENSG00000005436 | 0.347 | 5.054 | 2.366 | 0.02008 | 0.48015 | -3.481 | GCFC2 | GC-rich sequence DNA-binding factor 2 [Source:HGNC Symbol;Acc:HGNC:1317] |
| ENSG00000085871 | 1.072 | 1.455 | 2.363 | 0.02023 | 0.48299 | -3.238 | MGST2 | microsomal glutathione S-transferase 2 [Source:HGNC Symbol;Acc:HGNC:7063] |
| ENSG00000189159 | 0.441 | 4.434 | 2.360 | 0.02039 | 0.48532 | -3.466 | JPT1 | Jupiter microtubule associated homolog 1 [Source:HGNC Symbol;Acc:HGNC:14569] |
| ENSG00000114861 | -0.863 | 5.267 | -2.359 | 0.02042 | 0.48532 | -3.462 | FOXP1 | forkhead box P1 [Source:HGNC Symbol;Acc:HGNC:3823] |

| | | | | | | | | |
|---|---|---|---|---|---|---|---|---|
| ENSG00000124279 | 0.451 | 2.890 | 2.359 | 0.02044 | 0.48532 | -3.332 | FASTKD3 | FAST kinase domains 3 [Source:HGNC Symbol;Acc:HGNC:28758] |
| ENSG00000166451 | 0.417 | 3.687 | 2.356 | 0.02057 | 0.48754 | -3.415 | CENPN | centromere protein N [Source:HGNC Symbol;Acc:HGNC:30873] |
| ENSG00000165704 | 0.485 | 3.063 | 2.355 | 0.02063 | 0.48790 | -3.360 | HPRT1 | hypoxanthine phosphoribosyltransferase 1 [Source:HGNC Symbol;Acc:HGNC:5157] |
| ENSG00000100014 | -0.430 | 5.569 | -2.354 | 0.02072 | 0.48913 | -3.507 | SPECC1L | sperm antigen with calponin homology and coiled-coil domains 1 like [Source:HGNC Symbol;Acc:HGNC:29022] |
| ENSG00000111237 | 0.281 | 5.745 | 2.351 | 0.02086 | 0.49048 | -3.532 | VPS29 | VPS29, retromer complex component [Source:HGNC Symbol;Acc:HGNC:14340] |
| ENSG00000219200 | -0.477 | 4.500 | -2.351 | 0.02086 | 0.49048 | -3.443 | RNASEK | ribonuclease K [Source:HGNC Symbol;Acc:HGNC:33911] |
| ENSG00000249437 | -1.079 | 1.655 | -2.350 | 0.02092 | 0.49048 | -3.250 | NAIP | NLR family apoptosis inhibitory protein [Source:HGNC Symbol;Acc:HGNC:7634] |
| ENSG00000132510 | -0.944 | 4.054 | -2.349 | 0.02094 | 0.49048 | -3.377 | KDM6B | lysine demethylase 6B [Source:HGNC Symbol;Acc:HGNC:29012] |
| ENSG00000101182 | 0.366 | 6.482 | 2.349 | 0.02097 | 0.49048 | -3.537 | PSMA7 | proteasome subunit alpha 7 [Source:HGNC Symbol;Acc:HGNC:9536] |
| ENSG00000100330 | -0.378 | 5.659 | -2.347 | 0.02104 | 0.49066 | -3.525 | MTMR3 | myotubularin related protein 3 [Source:HGNC Symbol;Acc:HGNC:7451] |
| ENSG00000108622 | -0.554 | 4.981 | -2.347 | 0.02105 | 0.49066 | -3.484 | ICAM2 | intercellular adhesion molecule 2 [Source:HGNC Symbol;Acc:HGNC:5345] |
| ENSG00000094804 | 0.684 | 4.708 | 2.345 | 0.02117 | 0.49221 | -3.521 | CDC6 | cell division cycle 6 [Source:HGNC Symbol;Acc:HGNC:1744] |
| ENSG00000177150 | 0.357 | 4.097 | 2.344 | 0.02120 | 0.49221 | -3.470 | FAM210A | family with sequence similarity 210 member A [Source:HGNC Symbol;Acc:HGNC:28346] |
| ENSG00000144554 | 0.590 | 5.048 | 2.344 | 0.02124 | 0.49221 | -3.534 | FANCD2 | Fanconi anemia complementation group D2 [Source:HGNC Symbol;Acc:HGNC:3585] |
| ENSG00000189376 | 0.373 | 3.649 | 2.343 | 0.02127 | 0.49221 | -3.434 | C8orf76 | chromosome 8 open reading frame 76 [Source:HGNC Symbol;Acc:HGNC:25924] |
| ENSG00000170502 | -0.469 | 4.139 | -2.342 | 0.02136 | 0.49330 | -3.429 | NUDT9 | nudix hydrolase 9 [Source:HGNC Symbol;Acc:HGNC:8056] |
| ENSG00000144893 | 1.222 | 2.776 | 2.334 | 0.02174 | 0.50128 | -3.410 | MED12L | mediator complex subunit 12 like [Source:HGNC Symbol;Acc:HGNC:16050] |
| ENSG00000181754 | -1.014 | 0.718 | -2.331 | 0.02195 | 0.50211 | -3.354 | AMIGO1 | adhesion molecule with Ig like domain 1 [Source:HGNC Symbol;Acc:HGNC:20824] |
| ENSG00000145246 | -0.770 | 5.663 | -2.330 | 0.02198 | 0.50211 | -3.549 | ATP10D | ATPase phospholipid transporting 10D (putative) [Source:HGNC Symbol;Acc:HGNC:13549] |
| ENSG00000065268 | 0.582 | 2.511 | 2.330 | 0.02200 | 0.50211 | -3.363 | WDR18 | WD repeat domain 18 [Source:HGNC Symbol;Acc:HGNC:17956] |
| ENSG00000169118 | 0.283 | 5.349 | 2.329 | 0.02203 | 0.50211 | -3.569 | CSNK1G1 | casein kinase 1 gamma 1 [Source:HGNC Symbol;Acc:HGNC:2454] |
| ENSG00000145730 | -0.537 | 6.597 | -2.329 | 0.02203 | 0.50211 | -3.580 | PAM | peptidylglycine alpha-amidating monooxygenase [Source:HGNC Symbol;Acc:HGNC:8596] |
| ENSG00000115365 | 0.406 | 5.268 | 2.329 | 0.02206 | 0.50211 | -3.570 | LANCL1 | LanC like 1 [Source:HGNC Symbol;Acc:HGNC:6508] |
| ENSG00000128581 | 0.826 | 0.694 | 2.328 | 0.02207 | 0.50211 | -3.282 | IFT22 | intraflagellar transport 22 [Source:HGNC Symbol;Acc:HGNC:21895] |
| ENSG00000182054 | 0.578 | 7.067 | 2.328 | 0.02209 | 0.50211 | -3.565 | IDH2 | isocitrate dehydrogenase (NADP(+)) 2, mitochondrial [Source:HGNC Symbol;Acc:HGNC:5383] |
| ENSG00000105669 | 0.477 | 6.224 | 2.326 | 0.02220 | 0.50343 | -3.587 | COPE | coatomer protein complex subunit epsilon [Source:HGNC Symbol;Acc:HGNC:2234] |
| ENSG00000089327 | -0.791 | 6.232 | -2.326 | 0.02223 | 0.50343 | -3.578 | FXYD5 | FXYD domain containing ion transport regulator 5 [Source:HGNC Symbol;Acc:HGNC:4029] |
| ENSG00000171204 | 0.426 | 4.934 | 2.324 | 0.02233 | 0.50399 | -3.567 | TMEM126B | transmembrane protein 126B [Source:HGNC Symbol;Acc:HGNC:30883] |
| ENSG00000069869 | 1.107 | 2.236 | 2.324 | 0.02233 | 0.50399 | -3.378 | NEDD4 | neural precursor cell expressed, developmentally down-regulated 4, E3 ubiquitin protein ligase [Source:HGNC Symbol;Acc:HGNC:7727] |
| ENSG00000092201 | 0.379 | 6.617 | 2.323 | 0.02240 | 0.50452 | -3.591 | SUPT16H | SPT16 homolog, facilitates chromatin remodeling subunit [Source:HGNC Symbol;Acc:HGNC:11465] |
| ENSG00000141258 | -0.543 | 4.355 | -2.320 | 0.02256 | 0.50735 | -3.493 | SGSM2 | small G protein signaling modulator 2 [Source:HGNC Symbol;Acc:HGNC:29026] |
| ENSG00000215041 | -0.429 | 3.847 | -2.317 | 0.02269 | 0.50940 | -3.455 | NEURL4 | neuralized E3 ubiquitin protein ligase 4 [Source:HGNC Symbol;Acc:HGNC:34410] |
| ENSG00000115486 | -0.298 | 5.570 | -2.316 | 0.02277 | 0.50942 | -3.590 | GGCX | gamma-glutamyl carboxylase [Source:HGNC Symbol;Acc:HGNC:4247] |
| ENSG00000112592 | -0.374 | 3.362 | -2.316 | 0.02277 | 0.50942 | -3.411 | TBP | TATA-box binding protein [Source:HGNC Symbol;Acc:HGNC:11588] |
| ENSG00000104529 | -0.506 | 9.533 | -2.314 | 0.02288 | 0.51084 | -3.527 | EEF1D | eukaryotic translation elongation factor 1 delta [Source:HGNC Symbol;Acc:HGNC:3211] |
| ENSG00000119969 | 0.645 | 5.246 | 2.313 | 0.02296 | 0.51172 | -3.606 | HELLS | helicase, lymphoid-specific [Source:HGNC Symbol;Acc:HGNC:4861] |
| ENSG00000259753 | -1.236 | 3.420 | -2.311 | 0.02303 | 0.51172 | -3.376 | AC068234.1 | |
| ENSG00000083223 | -0.361 | 7.711 | -2.311 | 0.02307 | 0.51172 | -3.600 | ZCCHC6 | zinc finger CCHC-type containing 6 [Source:HGNC Symbol;Acc:HGNC:25817] |
| ENSG00000198961 | -0.300 | 8.746 | -2.311 | 0.02308 | 0.51172 | -3.559 | PJA2 | praja ring finger ubiquitin ligase 2 [Source:HGNC Symbol;Acc:HGNC:17481] |
| ENSG00000100804 | 0.485 | 5.116 | 2.309 | 0.02319 | 0.51239 | -3.608 | PSMB5 | proteasome subunit beta 5 [Source:HGNC Symbol;Acc:HGNC:9542] |
| ENSG00000241468 | 0.482 | 5.303 | 2.309 | 0.02319 | 0.51239 | -3.614 | ATP5J2 | ATP synthase, H+ transporting, mitochondrial Fo complex subunit F2 [Source:HGNC Symbol;Acc:HGNC:848] |
| ENSG00000164056 | -1.656 | 1.588 | -2.305 | 0.02338 | 0.51576 | -3.360 | SPRY1 | sprouty RTK signaling antagonist 1 [Source:HGNC Symbol;Acc:HGNC:11269] |
| ENSG00000165480 | 0.666 | 2.164 | 2.303 | 0.02355 | 0.51849 | -3.395 | SKA3 | spindle and kinetochore associated complex subunit 3 [Source:HGNC Symbol;Acc:HGNC:20262] |
| ENSG00000179051 | 0.478 | 5.991 | 2.301 | 0.02366 | 0.51887 | -3.640 | RCC2 | regulator of chromosome condensation 2 [Source:HGNC Symbol;Acc:HGNC:30297] |
| ENSG00000018510 | 0.315 | 6.163 | 2.300 | 0.02368 | 0.51887 | -3.641 | AGPS | alkylglycerone phosphate synthase [Source:HGNC Symbol;Acc:HGNC:327] |
| ENSG00000175455 | 0.616 | 6.001 | 2.300 | 0.02372 | 0.51887 | -3.642 | CCDC14 | coiled-coil domain containing 14 [Source:HGNC Symbol;Acc:HGNC:25766] |
| ENSG00000128567 | 0.762 | 2.097 | 2.300 | 0.02373 | 0.51887 | -3.394 | PODXL | podocalyxin like [Source:HGNC Symbol;Acc:HGNC:9171] |
| ENSG00000197223 | 0.311 | 4.384 | 2.295 | 0.02400 | 0.52400 | -3.592 | C1D | C1D nuclear receptor corepressor [Source:HGNC Symbol;Acc:HGNC:29911] |
| ENSG00000100890 | 0.358 | 5.460 | 2.293 | 0.02409 | 0.52407 | -3.648 | KIAA0391 | KIAA0391 [Source:HGNC Symbol;Acc:HGNC:19958] |
| ENSG00000182973 | 0.270 | 5.126 | 2.293 | 0.02412 | 0.52407 | -3.636 | CNOT10 | CCR4-NOT transcription complex subunit 10 [Source:HGNC Symbol;Acc:HGNC:23817] |
| ENSG00000143155 | 0.430 | 5.643 | 2.293 | 0.02413 | 0.52407 | -3.653 | TIPRL | TOR signaling pathway regulator [Source:HGNC Symbol;Acc:HGNC:30231] |
| ENSG00000072958 | 0.368 | 4.217 | 2.289 | 0.02436 | 0.52742 | -3.597 | AP1M1 | adaptor related protein complex 1 mu 1 subunit [Source:HGNC Symbol;Acc:HGNC:13667] |
| ENSG00000168404 | 0.763 | 4.979 | 2.289 | 0.02436 | 0.52742 | -3.649 | MLKL | mixed lineage kinase domain like pseudokinase [Source:HGNC Symbol;Acc:HGNC:26617] |
| ENSG00000267740 | 0.446 | 4.336 | 2.287 | 0.02451 | 0.52777 | -3.614 | AC024592.3 | |
| ENSG00000128016 | -0.684 | 7.832 | -2.286 | 0.02454 | 0.52777 | -3.654 | ZFP36 | ZFP36 ring finger protein [Source:HGNC Symbol;Acc:HGNC:12862] |
| ENSG00000152455 | 0.452 | 3.464 | 2.285 | 0.02460 | 0.52777 | -3.542 | SUV39H2 | suppressor of variegation 3-9 homolog 2 [Source:HGNC Symbol;Acc:HGNC:17287] |
| ENSG00000078114 | 1.633 | 0.690 | 2.285 | 0.02462 | 0.52777 | -3.358 | NEBL | nebulette [Source:HGNC Symbol;Acc:HGNC:16932] |
| ENSG00000188542 | -0.502 | 4.229 | -2.283 | 0.02473 | 0.52777 | -3.555 | DUSP28 | dual specificity phosphatase 28 [Source:HGNC Symbol;Acc:HGNC:33237] |
| ENSG00000161057 | 0.407 | 7.216 | 2.283 | 0.02473 | 0.52777 | -3.658 | PSMC2 | proteasome 26S subunit, ATPase 2 [Source:HGNC Symbol;Acc:HGNC:9548] |
| ENSG00000075131 | 0.514 | 3.582 | 2.283 | 0.02473 | 0.52777 | -3.562 | TIPIN | TIMELESS interacting protein [Source:HGNC Symbol;Acc:HGNC:30750] |
| ENSG00000111647 | 0.342 | 6.018 | 2.282 | 0.02479 | 0.52777 | -3.679 | UHRF1BP1L | UHRF1 binding protein 1 like [Source:HGNC Symbol;Acc:HGNC:29102] |
| ENSG00000004399 | -1.708 | 1.355 | -2.280 | 0.02489 | 0.52777 | -3.404 | PLXND1 | plexin D1 [Source:HGNC Symbol;Acc:HGNC:9107] |
| ENSG00000185022 | -0.737 | 6.190 | -2.280 | 0.02489 | 0.52777 | -3.673 | MAFF | MAF bZIP transcription factor F [Source:HGNC Symbol;Acc:HGNC:6780] |
| ENSG00000219626 | -0.709 | 3.608 | -2.280 | 0.02493 | 0.52777 | -3.482 | FAM228B | family with sequence similarity 228 member B [Source:HGNC Symbol;Acc:HGNC:24736] |
| ENSG00000089737 | -0.446 | 8.568 | -2.279 | 0.02498 | 0.52777 | -3.636 | DDX24 | DEAD-box helicase 24 [Source:HGNC Symbol;Acc:HGNC:13266] |
| ENSG00000165490 | 0.663 | 3.135 | 2.279 | 0.02500 | 0.52777 | -3.532 | DDIAS | DNA damage induced apoptosis suppressor [Source:HGNC Symbol;Acc:HGNC:26351] |
| ENSG00000180346 | -0.455 | 3.718 | -2.278 | 0.02501 | 0.52777 | -3.514 | TIGD2 | tigger transposable element derived 2 [Source:HGNC Symbol;Acc:HGNC:18333] |
| ENSG00000182621 | 1.243 | 3.783 | 2.278 | 0.02504 | 0.52777 | -3.625 | PLCB1 | phospholipase C beta 1 [Source:HGNC Symbol;Acc:HGNC:15917] |
| ENSG00000099800 | 0.474 | 4.397 | 2.278 | 0.02504 | 0.52777 | -3.637 | TIMM13 | translocase of inner mitochondrial membrane 13 [Source:HGNC Symbol;Acc:HGNC:11816] |
| ENSG00000248592 | 0.344 | 4.697 | 2.274 | 0.02528 | 0.53159 | -3.657 | TMEM110-MUSTN1 | TMEM110-MUSTN1 readthrough [Source:HGNC Symbol;Acc:HGNC:38834] |
| ENSG00000143297 | -0.951 | 7.850 | -2.274 | 0.02531 | 0.53159 | -3.683 | FCRL5 | Fc receptor like 5 [Source:HGNC Symbol;Acc:HGNC:18508] |
| ENSG00000099910 | -0.383 | 3.458 | -2.271 | 0.02545 | 0.53196 | -3.506 | KLHL22 | kelch like family member 22 [Source:HGNC Symbol;Acc:HGNC:25888] |
| ENSG00000166881 | 0.417 | 4.834 | 2.269 | 0.02558 | 0.53196 | -3.676 | NEMP1 | nuclear envelope integral membrane protein 1 [Source:HGNC Symbol;Acc:HGNC:29001] |
| ENSG00000032389 | 0.372 | 3.976 | 2.269 | 0.02559 | 0.53196 | -3.616 | TSSC1 | tumor suppressing subtransferable candidate 1 [Source:HGNC Symbol;Acc:HGNC:12383] |
| ENSG00000147592 | 0.548 | 4.086 | 2.269 | 0.02560 | 0.53196 | -3.633 | LACTB2 | lactamase beta 2 [Source:HGNC Symbol;Acc:HGNC:18512] |
| ENSG00000129460 | 0.348 | 5.762 | 2.268 | 0.02568 | 0.53196 | -3.706 | NGDN | neuroguidin [Source:HGNC Symbol;Acc:HGNC:20271] |
| ENSG00000165792 | 0.331 | 4.631 | 2.268 | 0.02569 | 0.53196 | -3.666 | METTL17 | methyltransferase like 17 [Source:HGNC Symbol;Acc:HGNC:19280] |
| ENSG00000149177 | -1.349 | 4.051 | -2.268 | 0.02569 | 0.53196 | -3.516 | PTPRJ | protein tyrosine phosphatase, receptor type J [Source:HGNC Symbol;Acc:HGNC:9673] |
| ENSG00000064490 | 0.394 | 3.895 | 2.267 | 0.02571 | 0.53196 | -3.616 | RFXANK | regulatory factor X associated ankyrin containing protein [Source:HGNC Symbol;Acc:HGNC:9987] |
| ENSG00000100888 | 0.338 | 6.786 | 2.267 | 0.02575 | 0.53196 | -3.705 | CHD8 | chromodomain helicase DNA binding protein 8 [Source:HGNC Symbol;Acc:HGNC:20153] |
| ENSG00000188687 | 0.465 | 3.702 | 2.266 | 0.02580 | 0.53196 | -3.602 | SLC4A5 | solute carrier family 4 member 5 [Source:HGNC Symbol;Acc:HGNC:18168] |
| ENSG00000112893 | -0.366 | 8.723 | -2.266 | 0.02581 | 0.53196 | -3.655 | MAN2A1 | mannosidase alpha class 2A member 1 [Source:HGNC Symbol;Acc:HGNC:6824] |
| ENSG00000249115 | 0.461 | 3.505 | 2.265 | 0.02588 | 0.53196 | -3.589 | HAUS5 | HAUS augmin like complex subunit 5 [Source:HGNC Symbol;Acc:HGNC:29130] |
| ENSG00000168067 | -0.559 | 3.416 | -2.265 | 0.02588 | 0.53196 | -3.506 | MAP4K2 | mitogen-activated protein kinase kinase kinase kinase 2 [Source:HGNC Symbol;Acc:HGNC:6864] |
| ENSG00000150433 | 0.454 | 4.452 | 2.264 | 0.02595 | 0.53196 | -3.665 | TMEM218 | transmembrane protein 218 [Source:HGNC Symbol;Acc:HGNC:27344] |
| ENSG00000119801 | -0.567 | 9.785 | -2.264 | 0.02595 | 0.53196 | -3.622 | YPEL5 | yippee like 5 [Source:HGNC Symbol;Acc:HGNC:18329] |
| ENSG00000259494 | 0.434 | 3.453 | 2.263 | 0.02600 | 0.53215 | -3.586 | MRPL46 | mitochondrial ribosomal protein L46 [Source:HGNC Symbol;Acc:HGNC:1192] |
| ENSG00000160256 | 0.523 | 2.793 | 2.257 | 0.02637 | 0.53626 | -3.529 | FAM207A | family with sequence similarity 207 member A [Source:HGNC Symbol;Acc:HGNC:15811] |
| ENSG00000138613 | -0.995 | 3.941 | -2.257 | 0.02639 | 0.53626 | -3.545 | APH1B | aph-1 homolog B, gamma-secretase subunit [Source:HGNC Symbol;Acc:HGNC:24080] |
| ENSG00000101447 | 0.830 | 1.073 | 2.256 | 0.02642 | 0.53626 | -3.413 | FAM83D | family with sequence similarity 83 member D [Source:HGNC Symbol;Acc:HGNC:16122] |

| | | | | | | | | |
|---|---|---|---|---|---|---|---|---|
| ENSG00000106462 | 0.521 | 4.977 | 2.256 | 0.02645 | 0.53626 | -3.713 | EZH2 | enhancer of zeste 2 polycomb repressive complex 2 subunit [Source:HGNC Symbol;Acc:HGNC:3527] |
| ENSG00000026751 | -0.567 | 9.064 | -2.255 | 0.02649 | 0.53626 | -3.667 | SLAMF7 | SLAM family member 7 [Source:HGNC Symbol;Acc:HGNC:21394] |
| ENSG00000144908 | 1.016 | 2.224 | 2.255 | 0.02653 | 0.53626 | -3.507 | ALDH1L1 | aldehyde dehydrogenase 1 family member L1 [Source:HGNC Symbol;Acc:HGNC:3978] |
| ENSG00000248919 | 0.418 | 5.042 | 2.255 | 0.02653 | 0.53626 | -3.716 | ATP5J2-PTCD1 | ATP5J2-PTCD1 readthrough [Source:HGNC Symbol;Acc:HGNC:38844] |
| ENSG00000166111 | 1.667 | -0.027 | 2.254 | 0.02658 | 0.53626 | -3.414 | SVOP | SV2 related protein [Source:HGNC Symbol;Acc:HGNC:25417] |
| ENSG00000123143 | 0.456 | 5.249 | 2.254 | 0.02659 | 0.53626 | -3.726 | PKN1 | protein kinase N1 [Source:HGNC Symbol;Acc:HGNC:9405] |
| ENSG00000169255 | 1.398 | 0.124 | 2.253 | 0.02662 | 0.53626 | -3.413 | B3GALNT1 | beta-1,3-N-acetylgalactosaminyltransferase 1 (globoside blood group) [Source:HGNC Symbol;Acc:HGNC:918] |
| ENSG00000214941 | -0.464 | 3.792 | -2.250 | 0.02681 | 0.53694 | -3.581 | ZSWIM7 | zinc finger SWIM-type containing 7 [Source:HGNC Symbol;Acc:HGNC:26993] |
| ENSG00000135604 | -0.650 | 5.695 | -2.249 | 0.02689 | 0.53694 | -3.723 | STX11 | syntaxin 11 [Source:HGNC Symbol;Acc:HGNC:11429] |
| ENSG00000150756 | 0.447 | 3.436 | 2.249 | 0.02690 | 0.53694 | -3.611 | FAM173B | family with sequence similarity 173 member B [Source:HGNC Symbol;Acc:HGNC:27029] |
| ENSG00000070081 | -0.718 | 8.604 | -2.248 | 0.02694 | 0.53694 | -3.704 | NUCB2 | nucleobindin 2 [Source:HGNC Symbol;Acc:HGNC:8044] |
| ENSG00000198642 | -0.508 | 5.514 | -2.247 | 0.02702 | 0.53694 | -3.724 | KLHL9 | kelch like family member 9 [Source:HGNC Symbol;Acc:HGNC:18732] |
| ENSG00000170421 | 1.336 | 0.836 | 2.247 | 0.02702 | 0.53694 | -3.432 | KRT8 | keratin 8 [Source:HGNC Symbol;Acc:HGNC:6446] |
| ENSG00000188486 | 0.504 | 4.022 | 2.246 | 0.02707 | 0.53694 | -3.672 | H2AFX | H2A histone family member X [Source:HGNC Symbol;Acc:HGNC:4739] |
| ENSG00000179029 | -0.870 | 6.919 | -2.246 | 0.02711 | 0.53694 | -3.752 | TMEM107 | transmembrane protein 107 [Source:HGNC Symbol;Acc:HGNC:28128] |
| ENSG00000076248 | 0.559 | 3.516 | 2.246 | 0.02711 | 0.53694 | -3.632 | UNG | uracil DNA glycosylase [Source:HGNC Symbol;Acc:HGNC:12572] |
| ENSG00000146281 | 0.528 | 6.485 | 2.245 | 0.02716 | 0.53694 | -3.752 | PM20D2 | peptidase M20 domain containing 2 [Source:HGNC Symbol;Acc:HGNC:21408] |
| ENSG00000187323 | 1.558 | 2.643 | 2.244 | 0.02720 | 0.53694 | -3.601 | DCC | DCC netrin 1 receptor [Source:HGNC Symbol;Acc:HGNC:2701] |
| ENSG00000079805 | 0.289 | 6.989 | 2.243 | 0.02726 | 0.53694 | -3.748 | DNM2 | dynamin 2 [Source:HGNC Symbol;Acc:HGNC:2974] |
| ENSG00000065000 | 0.365 | 6.956 | 2.243 | 0.02727 | 0.53694 | -3.748 | AP3D1 | adaptor related protein complex 3 delta 1 subunit [Source:HGNC Symbol;Acc:HGNC:568] |
| ENSG00000163956 | -0.414 | 7.360 | -2.243 | 0.02727 | 0.53694 | -3.751 | LRPAP1 | LDL receptor related protein associated protein 1 [Source:HGNC Symbol;Acc:HGNC:6701] |
| ENSG00000100201 | -0.364 | 9.404 | -2.243 | 0.02729 | 0.53694 | -3.673 | DDX17 | DEAD-box helicase 17 [Source:HGNC Symbol;Acc:HGNC:2740] |
| ENSG00000168395 | -0.336 | 4.839 | -2.242 | 0.02733 | 0.53694 | -3.700 | ING5 | inhibitor of growth family member 5 [Source:HGNC Symbol;Acc:HGNC:19421] |
| ENSG00000111206 | 0.609 | 2.679 | 2.242 | 0.02738 | 0.53718 | -3.562 | FOXM1 | forkhead box M1 [Source:HGNC Symbol;Acc:HGNC:3818] |
| ENSG00000172197 | 0.823 | 2.362 | 2.237 | 0.02769 | 0.54234 | -3.536 | MBOAT1 | membrane bound O-acyltransferase domain containing 1 [Source:HGNC Symbol;Acc:HGNC:21579] |
| ENSG00000006451 | 0.403 | 4.403 | 2.236 | 0.02773 | 0.54236 | -3.717 | RALA | RAS like proto-oncogene A [Source:HGNC Symbol;Acc:HGNC:9839] |
| ENSG00000259075 | -0.375 | 5.527 | -2.232 | 0.02803 | 0.54611 | -3.758 | POC1B-GALNT4 | POC1B-GALNT4 readthrough [Source:HGNC Symbol;Acc:HGNC:42957] |
| ENSG00000162616 | -0.669 | 5.589 | -2.232 | 0.02803 | 0.54611 | -3.752 | DNAJB4 | DnaJ heat shock protein family (Hsp40) member B4 [Source:HGNC Symbol;Acc:HGNC:14886] |
| ENSG00000108509 | -0.647 | 2.543 | -2.232 | 0.02805 | 0.54611 | -3.486 | CAMTA2 | calmodulin binding transcription activator 2 [Source:HGNC Symbol;Acc:HGNC:18807] |
| ENSG00000228439 | -0.515 | 1.456 | -2.231 | 0.02812 | 0.54635 | -3.451 | TSTD3 | thiosulfate sulfurtransferase (rhodanese)-like domain containing 3 [Source:HGNC Symbol;Acc:HGNC:40910] |
| ENSG00000172009 | 0.450 | 4.691 | 2.230 | 0.02818 | 0.54635 | -3.752 | THOP1 | thimet oligopeptidase 1 [Source:HGNC Symbol;Acc:HGNC:11793] |
| ENSG00000120802 | 0.541 | 7.533 | 2.229 | 0.02826 | 0.54635 | -3.753 | TMPO | thymopoietin [Source:HGNC Symbol;Acc:HGNC:11875] |
| ENSG00000171488 | -0.786 | 5.752 | -2.228 | 0.02832 | 0.54635 | -3.763 | LRRC8C | leucine rich repeat containing 8 family member C [Source:HGNC Symbol;Acc:HGNC:25075] |
| ENSG00000132286 | -0.325 | 6.032 | -2.227 | 0.02841 | 0.54635 | -3.787 | TIMM10B | translocase of inner mitochondrial membrane 10B [Source:HGNC Symbol;Acc:HGNC:4022] |
| ENSG00000171757 | 0.536 | 2.405 | 2.226 | 0.02846 | 0.54635 | -3.554 | LRRC34 | leucine rich repeat containing 34 [Source:HGNC Symbol;Acc:HGNC:28408] |
| ENSG00000256043 | -0.670 | 6.708 | -2.226 | 0.02848 | 0.54635 | -3.793 | CTSO | cathepsin O [Source:HGNC Symbol;Acc:HGNC:2542] |
| ENSG00000108839 | -0.744 | 1.399 | -2.225 | 0.02849 | 0.54635 | -3.462 | ALOX12 | arachidonate 12-lipoxygenase, 12S type [Source:HGNC Symbol;Acc:HGNC:429] |
| ENSG00000114744 | 0.324 | 5.025 | 2.224 | 0.02858 | 0.54635 | -3.774 | COMMD2 | COMM domain containing 2 [Source:HGNC Symbol;Acc:HGNC:24993] |
| ENSG00000105185 | 0.612 | 5.952 | 2.224 | 0.02859 | 0.54635 | -3.798 | PDCD5 | programmed cell death 5 [Source:HGNC Symbol;Acc:HGNC:8764] |
| ENSG00000112149 | -1.372 | 4.896 | -2.224 | 0.02860 | 0.54635 | -3.681 | CD83 | CD83 molecule [Source:HGNC Symbol;Acc:HGNC:1703] |
| ENSG00000100425 | -0.360 | 5.242 | -2.224 | 0.02860 | 0.54635 | -3.763 | BRD1 | bromodomain containing 1 [Source:HGNC Symbol;Acc:HGNC:1102] |
| ENSG00000128951 | 0.397 | 6.121 | 2.223 | 0.02865 | 0.54635 | -3.800 | DUT | deoxyuridine triphosphatase [Source:HGNC Symbol;Acc:HGNC:3078] |
| ENSG00000198176 | 0.828 | 3.933 | 2.223 | 0.02866 | 0.54635 | -3.729 | TFDP1 | transcription factor Dp-1 [Source:HGNC Symbol;Acc:HGNC:11749] |
| ENSG00000105220 | 0.404 | 6.122 | 2.221 | 0.02882 | 0.54847 | -3.805 | GPI | glucose-6-phosphate isomerase [Source:HGNC Symbol;Acc:HGNC:4458] |
| ENSG00000114656 | 0.593 | 0.695 | 2.219 | 0.02892 | 0.54847 | -3.471 | KIAA1257 | KIAA1257 [Source:HGNC Symbol;Acc:HGNC:29231] |
| ENSG00000170876 | 0.371 | 5.521 | 2.218 | 0.02900 | 0.54847 | -3.803 | TMEM43 | transmembrane protein 43 [Source:HGNC Symbol;Acc:HGNC:28472] |
| ENSG00000151292 | -0.540 | 7.185 | -2.218 | 0.02900 | 0.54847 | -3.806 | CSNK1G3 | casein kinase 1 gamma 3 [Source:HGNC Symbol;Acc:HGNC:2456] |
| ENSG00000105397 | 0.436 | 5.663 | 2.217 | 0.02910 | 0.54847 | -3.810 | TYK2 | tyrosine kinase 2 [Source:HGNC Symbol;Acc:HGNC:12440] |
| ENSG00000105258 | 0.374 | 4.159 | 2.217 | 0.02911 | 0.54847 | -3.737 | POLR2I | RNA polymerase II subunit I [Source:HGNC Symbol;Acc:HGNC:9196] |
| ENSG00000271383 | -0.682 | 3.419 | -2.216 | 0.02917 | 0.54847 | -3.591 | NBPF19 | NBPF member 19 [Source:HGNC Symbol;Acc:HGNC:31999] |
| ENSG00000125633 | -0.354 | 6.094 | -2.215 | 0.02920 | 0.54847 | -3.811 | CCDC93 | coiled-coil domain containing 93 [Source:HGNC Symbol;Acc:HGNC:25611] |
| ENSG00000146143 | 0.378 | 4.039 | 2.215 | 0.02923 | 0.54847 | -3.731 | PRIM2 | primase (DNA) subunit 2 [Source:HGNC Symbol;Acc:HGNC:9370] |
| ENSG00000168522 | 0.365 | 5.784 | 2.214 | 0.02928 | 0.54847 | -3.816 | FNTA | farnesyltransferase, CAAX box, alpha [Source:HGNC Symbol;Acc:HGNC:3782] |
| ENSG00000228696 | -0.850 | 1.682 | -2.214 | 0.02930 | 0.54847 | -3.479 | ARL17B | ADP ribosylation factor like GTPase 17B [Source:HGNC Symbol;Acc:HGNC:32387] |
| ENSG00000131174 | 0.465 | 5.896 | 2.214 | 0.02931 | 0.54847 | -3.818 | COX7B | cytochrome c oxidase subunit 7B [Source:HGNC Symbol;Acc:HGNC:2291] |
| ENSG00000105486 | 0.433 | 5.304 | 2.213 | 0.02933 | 0.54847 | -3.808 | LIG1 | DNA ligase 1 [Source:HGNC Symbol;Acc:HGNC:6598] |
| ENSG00000281039 | 0.395 | 3.597 | 2.212 | 0.02940 | 0.54899 | -3.695 | AC005154.6 | |
| ENSG00000131013 | -0.419 | 6.142 | -2.212 | 0.02945 | 0.54914 | -3.818 | PPIL4 | peptidylprolyl isomerase like 4 [Source:HGNC Symbol;Acc:HGNC:15702] |
| ENSG00000130520 | 0.314 | 5.236 | 2.211 | 0.02952 | 0.54961 | -3.809 | LSM4 | LSM4 homolog, U6 small nuclear RNA and mRNA degradation associated [Source:HGNC Symbol;Acc:HGNC:17259] |
| ENSG00000204130 | -0.316 | 5.415 | -2.209 | 0.02962 | 0.55069 | -3.801 | RUFY2 | RUN and FYVE domain containing 2 [Source:HGNC Symbol;Acc:HGNC:19761] |
| ENSG00000070190 | -0.631 | 6.290 | -2.207 | 0.02979 | 0.55209 | -3.826 | DAPP1 | dual adaptor of phosphotyrosine and 3-phosphoinositides 1 [Source:HGNC Symbol;Acc:HGNC:16500] |
| ENSG00000105058 | 0.362 | 5.965 | 2.207 | 0.02982 | 0.55209 | -3.833 | FAM32A | family with sequence similarity 32 member A [Source:HGNC Symbol;Acc:HGNC:24563] |
| ENSG00000104972 | -0.912 | 4.276 | -2.206 | 0.02985 | 0.55209 | -3.683 | LILRB1 | leukocyte immunoglobulin like receptor B1 [Source:HGNC Symbol;Acc:HGNC:6605] |
| ENSG00000108384 | 0.430 | 3.416 | 2.206 | 0.02988 | 0.55209 | -3.693 | RAD51C | RAD51 paralog C [Source:HGNC Symbol;Acc:HGNC:9820] |
| ENSG00000119203 | 0.341 | 5.527 | 2.205 | 0.02991 | 0.55209 | -3.829 | CPSF3 | cleavage and polyadenylation specific factor 3 [Source:HGNC Symbol;Acc:HGNC:2326] |
| ENSG00000145476 | -0.796 | 4.595 | -2.204 | 0.02998 | 0.55260 | -3.727 | CYP4V2 | cytochrome P450 family 4 subfamily V member 2 [Source:HGNC Symbol;Acc:HGNC:23198] |
| ENSG00000186638 | 0.528 | 2.808 | 2.202 | 0.03017 | 0.55512 | -3.645 | KIF24 | kinesin family member 24 [Source:HGNC Symbol;Acc:HGNC:19916] |
| ENSG00000127586 | 0.603 | 2.892 | 2.201 | 0.03024 | 0.55512 | -3.661 | CHTF18 | chromosome transmission fidelity factor 18 [Source:HGNC Symbol;Acc:HGNC:18435] |
| ENSG00000161791 | -0.623 | 4.284 | -2.199 | 0.03038 | 0.55512 | -3.721 | FMNL3 | formin like 3 [Source:HGNC Symbol;Acc:HGNC:23698] |
| ENSG00000065057 | 0.698 | 1.870 | 2.199 | 0.03038 | 0.55512 | -3.562 | NTHL1 | nth like DNA glycosylase 1 [Source:HGNC Symbol;Acc:HGNC:8028] |
| ENSG00000132793 | 1.273 | 0.619 | 2.199 | 0.03041 | 0.55512 | -3.506 | LPIN3 | lipin 3 [Source:HGNC Symbol;Acc:HGNC:14451] |
| ENSG00000184220 | 0.535 | 5.097 | 2.198 | 0.03044 | 0.55512 | -3.834 | CMSS1 | cms1 ribosomal small subunit homolog (yeast) [Source:HGNC Symbol;Acc:HGNC:28666] |
| ENSG00000113719 | -0.398 | 7.596 | -2.198 | 0.03044 | 0.55512 | -3.835 | ERGIC1 | endoplasmic reticulum-golgi intermediate compartment 1 [Source:HGNC Symbol;Acc:HGNC:29205] |
| ENSG00000133026 | -0.808 | 3.737 | -2.198 | 0.03049 | 0.55512 | -3.655 | MYH10 | myosin heavy chain 10 [Source:HGNC Symbol;Acc:HGNC:7568] |
| ENSG00000022267 | -1.129 | 4.715 | -2.197 | 0.03055 | 0.55512 | -3.736 | FHL1 | four and a half LIM domains 1 [Source:HGNC Symbol;Acc:HGNC:3702] |
| ENSG00000088367 | -1.012 | 2.689 | -2.197 | 0.03056 | 0.55512 | -3.541 | EPB41L1 | erythrocyte membrane protein band 4.1 like 1 [Source:HGNC Symbol;Acc:HGNC:3378] |
| ENSG00000189233 | -0.582 | 6.728 | -2.194 | 0.03077 | 0.55773 | -3.858 | NUGGC | nuclear GTPase, germinal center associated [Source:HGNC Symbol;Acc:HGNC:33550] |
| ENSG00000064199 | 0.720 | 0.811 | 2.193 | 0.03079 | 0.55773 | -3.515 | SPA17 | sperm autoantigenic protein 17 [Source:HGNC Symbol;Acc:HGNC:11210] |
| ENSG00000118596 | -0.545 | 6.611 | -2.191 | 0.03096 | 0.56014 | -3.863 | SLC16A7 | solute carrier family 16 member 7 [Source:HGNC Symbol;Acc:HGNC:10928] |
| ENSG00000111912 | -0.566 | 6.741 | -2.190 | 0.03105 | 0.56033 | -3.866 | NCOA7 | nuclear receptor coactivator 7 [Source:HGNC Symbol;Acc:HGNC:21081] |
| ENSG00000122035 | -0.954 | 0.687 | -2.189 | 0.03109 | 0.56033 | -3.557 | RASL11A | RAS like family 11 member A [Source:HGNC Symbol;Acc:HGNC:23802] |
| ENSG00000114491 | 0.422 | 4.358 | 2.189 | 0.03111 | 0.56033 | -3.809 | UMPS | uridine monophosphate synthetase [Source:HGNC Symbol;Acc:HGNC:12563] |
| ENSG00000213533 | 0.332 | 4.704 | 2.189 | 0.03115 | 0.56039 | -3.829 | TMEM110 | transmembrane protein 110 [Source:HGNC Symbol;Acc:HGNC:30526] |
| ENSG00000185055 | 0.385 | 2.733 | 2.187 | 0.03128 | 0.56114 | -3.650 | EFCAB10 | EF-hand calcium binding domain 10 [Source:HGNC Symbol;Acc:HGNC:34531] |
| ENSG00000116209 | -0.541 | 8.693 | -2.187 | 0.03128 | 0.56114 | -3.819 | TMEM59 | transmembrane protein 59 [Source:HGNC Symbol;Acc:HGNC:1239] |
| ENSG00000160401 | 0.580 | 1.263 | 2.185 | 0.03140 | 0.56165 | -3.538 | CFAP157 | cilia and flagella associated protein 157 [Source:HGNC Symbol;Acc:HGNC:27843] |
| ENSG00000007968 | 0.840 | 3.512 | 2.185 | 0.03145 | 0.56165 | -3.773 | E2F2 | E2F transcription factor 2 [Source:HGNC Symbol;Acc:HGNC:3114] |
| ENSG00000101473 | 0.345 | 3.044 | 2.185 | 0.03146 | 0.56165 | -3.686 | ACOT8 | acyl-CoA thioesterase 8 [Source:HGNC Symbol;Acc:HGNC:15919] |
| ENSG00000165272 | -0.714 | 5.899 | -2.184 | 0.03153 | 0.56165 | -3.861 | AQP3 | aquaporin 3 (Gill blood group) [Source:HGNC Symbol;Acc:HGNC:636] |
| ENSG00000132388 | -0.361 | 7.007 | -2.183 | 0.03155 | 0.56165 | -3.877 | UBE2G1 | ubiquitin conjugating enzyme E2 G1 [Source:HGNC Symbol;Acc:HGNC:12482] |

| | | | | | | | | |
|---|---|---|---|---|---|---|---|---|
| ENSG00000140563 | -0.465 | 7.687 | -2.183 | 0.03157 | 0.56165 | -3.864 | MCTP2 | multiple C2 and transmembrane domain containing 2 [Source:HGNC Symbol;Acc:HGNC:25636] |
| ENSG00000163705 | 0.579 | 1.847 | 2.182 | 0.03164 | 0.56205 | -3.584 | FANCD2OS | FANCD2 opposite strand [Source:HGNC Symbol;Acc:HGNC:28623] |
| ENSG00000184635 | 0.865 | 1.183 | 2.181 | 0.03172 | 0.56205 | -3.557 | ZNF93 | zinc finger protein 93 [Source:HGNC Symbol;Acc:HGNC:13169] |
| ENSG00000158457 | -1.812 | 1.526 | -2.181 | 0.03173 | 0.56205 | -3.551 | TSPAN33 | tetraspanin 33 [Source:HGNC Symbol;Acc:HGNC:28743] |
| ENSG00000151500 | 0.427 | 4.771 | 2.180 | 0.03179 | 0.56238 | -3.853 | THYN1 | thymocyte nuclear protein 1 [Source:HGNC Symbol;Acc:HGNC:29560] |
| ENSG00000164898 | 0.462 | 2.031 | 2.179 | 0.03185 | 0.56257 | -3.592 | FMC1 | formation of mitochondrial complex V assembly factor 1 homolog [Source:HGNC Symbol;Acc:HGNC:26946] |
| ENSG00000167522 | -0.574 | 6.519 | -2.177 | 0.03204 | 0.56290 | -3.891 | ANKRD11 | ankyrin repeat domain 11 [Source:HGNC Symbol;Acc:HGNC:21316] |
| ENSG00000120699 | 0.360 | 4.315 | 2.177 | 0.03205 | 0.56290 | -3.829 | EXOSC8 | exosome component 8 [Source:HGNC Symbol;Acc:HGNC:17035] |
| ENSG00000258790 | 0.451 | 7.560 | 2.176 | 0.03208 | 0.56290 | -3.859 | AL121594.3 | |
| ENSG00000075568 | -0.350 | 6.171 | -2.176 | 0.03210 | 0.56290 | -3.891 | TMEM131 | transmembrane protein 131 [Source:HGNC Symbol;Acc:HGNC:30366] |
| ENSG00000142733 | 0.679 | 1.363 | 2.176 | 0.03211 | 0.56290 | -3.566 | MAP3K6 | mitogen-activated protein kinase kinase kinase 6 [Source:HGNC Symbol;Acc:HGNC:6858] |
| ENSG00000136379 | 0.874 | 0.976 | 2.175 | 0.03215 | 0.56290 | -3.552 | ABHD17C | abhydrolase domain containing 17C [Source:HGNC Symbol;Acc:HGNC:26925] |
| ENSG00000006459 | -0.479 | 6.026 | -2.175 | 0.03219 | 0.56290 | -3.888 | KDM7A | lysine demethylase 7A [Source:HGNC Symbol;Acc:HGNC:22224] |
| ENSG00000153046 | -0.498 | 6.611 | -2.174 | 0.03227 | 0.56290 | -3.898 | CDYL | chromodomain Y like [Source:HGNC Symbol;Acc:HGNC:1811] |
| ENSG00000171574 | 0.494 | 1.254 | 2.174 | 0.03229 | 0.56290 | -3.555 | ZNF584 | zinc finger protein 584 [Source:HGNC Symbol;Acc:HGNC:27318] |
| ENSG00000123473 | 0.568 | 2.995 | 2.173 | 0.03233 | 0.56290 | -3.727 | STIL | SCL/TAL1 interrupting locus [Source:HGNC Symbol;Acc:HGNC:10879] |
| ENSG00000167862 | 0.446 | 3.810 | 2.173 | 0.03236 | 0.56290 | -3.798 | MRPL58 | mitochondrial ribosomal protein L58 [Source:HGNC Symbol;Acc:HGNC:5359] |
| ENSG00000128654 | 0.329 | 4.664 | 2.172 | 0.03239 | 0.56290 | -3.859 | MTX2 | metaxin 2 [Source:HGNC Symbol;Acc:HGNC:7506] |
| ENSG00000126215 | 0.532 | 2.770 | 2.171 | 0.03252 | 0.56343 | -3.697 | XRCC3 | X-ray repair cross complementing 3 [Source:HGNC Symbol;Acc:HGNC:12830] |
| ENSG00000196275 | -0.517 | 1.417 | -2.170 | 0.03256 | 0.56343 | -3.551 | GTF2IRD2 | GTF2I repeat domain containing 2 [Source:HGNC Symbol;Acc:HGNC:30775] |
| ENSG00000171467 | -0.439 | 4.557 | -2.170 | 0.03256 | 0.56343 | -3.820 | ZNF318 | zinc finger protein 318 [Source:HGNC Symbol;Acc:HGNC:13578] |
| ENSG00000110344 | 0.309 | 6.842 | 2.168 | 0.03274 | 0.56469 | -3.903 | UBE4A | ubiquitination factor E4A [Source:HGNC Symbol;Acc:HGNC:12499] |
| ENSG00000197363 | -0.711 | 1.769 | -2.167 | 0.03278 | 0.56469 | -3.557 | ZNF517 | zinc finger protein 517 [Source:HGNC Symbol;Acc:HGNC:27984] |
| ENSG00000106125 | 0.734 | 1.070 | 2.167 | 0.03280 | 0.56469 | -3.565 | MINDY4 | MINDY lysine 48 deubiquitinase 4 [Source:HGNC Symbol;Acc:HGNC:21916] |
| ENSG00000204264 | 0.440 | 6.057 | 2.167 | 0.03281 | 0.56469 | -3.912 | PSMB8 | proteasome subunit beta 8 [Source:HGNC Symbol;Acc:HGNC:9545] |
| ENSG00000143553 | 0.391 | 3.125 | 2.166 | 0.03287 | 0.56509 | -3.735 | SNAPIN | SNAP associated protein [Source:HGNC Symbol;Acc:HGNC:17145] |
| ENSG00000108599 | -0.274 | 5.609 | -2.165 | 0.03297 | 0.56555 | -3.899 | AKAP10 | A-kinase anchoring protein 10 [Source:HGNC Symbol;Acc:HGNC:368] |
| ENSG00000164754 | 0.369 | 7.719 | 2.165 | 0.03300 | 0.56555 | -3.879 | RAD21 | RAD21 cohesin complex component [Source:HGNC Symbol;Acc:HGNC:9811] |
| ENSG00000188171 | 0.815 | 2.944 | 2.164 | 0.03303 | 0.56555 | -3.751 | ZNF626 | zinc finger protein 626 [Source:HGNC Symbol;Acc:HGNC:30461] |
| ENSG00000123384 | -1.319 | 3.631 | -2.162 | 0.03318 | 0.56629 | -3.673 | LRP1 | LDL receptor related protein 1 [Source:HGNC Symbol;Acc:HGNC:6692] |
| ENSG00000118762 | -0.522 | 4.576 | -2.162 | 0.03322 | 0.56629 | -3.827 | PKD2 | polycystin 2, transient receptor potential cation channel [Source:HGNC Symbol;Acc:HGNC:9009] |
| ENSG00000125743 | 0.444 | 5.815 | 2.162 | 0.03322 | 0.56629 | -3.921 | SNRPD2 | small nuclear ribonucleoprotein D2 polypeptide [Source:HGNC Symbol;Acc:HGNC:11159] |
| ENSG00000198937 | 0.445 | 3.054 | 2.161 | 0.03325 | 0.56629 | -3.740 | CCDC167 | coiled-coil domain containing 167 [Source:HGNC Symbol;Acc:HGNC:21239] |
| ENSG00000171202 | 0.387 | 4.316 | 2.160 | 0.03340 | 0.56793 | -3.862 | TMEM126A | transmembrane protein 126A [Source:HGNC Symbol;Acc:HGNC:25382] |
| ENSG00000100139 | -0.463 | 2.978 | -2.159 | 0.03348 | 0.56793 | -3.671 | MICALL1 | MICAL like 1 [Source:HGNC Symbol;Acc:HGNC:29804] |
| ENSG00000012822 | -0.300 | 6.880 | -2.158 | 0.03350 | 0.56793 | -3.928 | CALCOCO1 | calcium binding and coiled-coil domain 1 [Source:HGNC Symbol;Acc:HGNC:29306] |
| ENSG00000265354 | 0.337 | 5.249 | 2.158 | 0.03353 | 0.56793 | -3.915 | TIMM23 | translocase of inner mitochondrial membrane 23 [Source:HGNC Symbol;Acc:HGNC:17312] |
| ENSG00000186532 | -0.351 | 3.308 | -2.157 | 0.03359 | 0.56818 | -3.713 | SMYD4 | SET and MYND domain containing 4 [Source:HGNC Symbol;Acc:HGNC:21067] |
| ENSG00000106591 | 0.347 | 5.997 | 2.155 | 0.03373 | 0.56818 | -3.935 | MRPL32 | mitochondrial ribosomal protein L32 [Source:HGNC Symbol;Acc:HGNC:14035] |
| ENSG00000135297 | -0.293 | 5.765 | -2.155 | 0.03376 | 0.56818 | -3.924 | MTO1 | mitochondrial tRNA translation optimization 1 [Source:HGNC Symbol;Acc:HGNC:19261] |
| ENSG00000267673 | 0.467 | 1.911 | 2.155 | 0.03376 | 0.56818 | -3.629 | FDX2 | ferredoxin 2 [Source:HGNC Symbol;Acc:HGNC:30546] |
| ENSG00000100380 | -0.411 | 7.557 | -2.155 | 0.03377 | 0.56818 | -3.923 | ST13 | ST13, Hsp70 interacting protein [Source:HGNC Symbol;Acc:HGNC:11343] |
| ENSG00000092853 | 0.691 | 3.564 | 2.154 | 0.03382 | 0.56818 | -3.828 | CLSPN | claspin [Source:HGNC Symbol;Acc:HGNC:19715] |
| ENSG00000158050 | -1.036 | 5.647 | -2.154 | 0.03389 | 0.56818 | -3.899 | DUSP2 | dual specificity phosphatase 2 [Source:HGNC Symbol;Acc:HGNC:3068] |
| ENSG00000254959 | 0.705 | 0.815 | 2.153 | 0.03390 | 0.56818 | -3.581 | INMT-MINDY4 | INMT-MINDY4 readthrough (NMD candidate) [Source:HGNC Symbol;Acc:HGNC:41995] |
| ENSG00000108666 | 0.340 | 3.699 | 2.151 | 0.03406 | 0.57008 | -3.822 | C17orf75 | chromosome 17 open reading frame 75 [Source:HGNC Symbol;Acc:HGNC:30173] |
| ENSG00000184162 | 0.392 | 2.796 | 2.150 | 0.03417 | 0.57109 | -3.727 | NR2C2AP | nuclear receptor 2C2 associated protein [Source:HGNC Symbol;Acc:HGNC:30763] |
| ENSG00000154222 | -0.274 | 5.332 | -2.150 | 0.03421 | 0.57109 | -3.919 | CC2D1B | coiled-coil and C2 domain containing 1B [Source:HGNC Symbol;Acc:HGNC:29386] |
| ENSG00000162600 | -0.443 | 6.066 | -2.149 | 0.03429 | 0.57111 | -3.941 | OMA1 | OMA1 zinc metallopeptidase [Source:HGNC Symbol;Acc:HGNC:29661] |
| ENSG00000174130 | -0.851 | 4.997 | -2.148 | 0.03430 | 0.57111 | -3.870 | TLR6 | toll like receptor 6 [Source:HGNC Symbol;Acc:HGNC:16711] |
| ENSG00000091732 | 0.471 | 2.355 | 2.147 | 0.03439 | 0.57113 | -3.692 | ZC3HC1 | zinc finger C3HC-type containing 1 [Source:HGNC Symbol;Acc:HGNC:29913] |
| ENSG00000220205 | -0.567 | 3.674 | -2.147 | 0.03439 | 0.57113 | -3.759 | VAMP2 | vesicle associated membrane protein 2 [Source:HGNC Symbol;Acc:HGNC:12643] |
| ENSG00000139318 | -0.893 | 5.933 | -2.146 | 0.03447 | 0.57169 | -3.930 | DUSP6 | dual specificity phosphatase 6 [Source:HGNC Symbol;Acc:HGNC:3072] |
| ENSG00000151914 | -0.747 | 9.108 | -2.146 | 0.03451 | 0.57169 | -3.888 | DST | dystonin [Source:HGNC Symbol;Acc:HGNC:1090] |
| ENSG00000116670 | 0.356 | 4.048 | 2.144 | 0.03463 | 0.57299 | -3.871 | MAD2L2 | mitotic arrest deficient 2 like 2 [Source:HGNC Symbol;Acc:HGNC:6764] |
| ENSG00000092094 | 0.314 | 4.633 | 2.144 | 0.03469 | 0.57320 | -3.912 | OSGEP | O-sialoglycoprotein endopeptidase [Source:HGNC Symbol;Acc:HGNC:18028] |
| ENSG00000138767 | -0.414 | 7.049 | -2.141 | 0.03488 | 0.57555 | -3.960 | CNOT6L | CCR4-NOT transcription complex subunit 6 like [Source:HGNC Symbol;Acc:HGNC:18042] |
| ENSG00000168958 | 0.306 | 6.161 | 2.138 | 0.03512 | 0.57855 | -3.969 | MFF | mitochondrial fission factor [Source:HGNC Symbol;Acc:HGNC:24858] |
| ENSG00000138641 | -0.274 | 6.200 | -2.138 | 0.03515 | 0.57855 | -3.967 | HERC3 | HECT and RLD domain containing E3 ubiquitin protein ligase 3 [Source:HGNC Symbol;Acc:HGNC:4876] |
| ENSG00000119013 | 0.450 | 4.493 | 2.136 | 0.03535 | 0.58049 | -3.924 | NDUFB3 | NADH:ubiquinone oxidoreductase subunit B3 [Source:HGNC Symbol;Acc:HGNC:7698] |
| ENSG00000113966 | 0.495 | 2.698 | 2.136 | 0.03536 | 0.58049 | -3.745 | ARL6 | ADP ribosylation factor like GTPase 6 [Source:HGNC Symbol;Acc:HGNC:13210] |
| ENSG00000154277 | 2.343 | 1.559 | 2.135 | 0.03543 | 0.58061 | -3.710 | UCHL1 | ubiquitin C-terminal hydrolase L1 [Source:HGNC Symbol;Acc:HGNC:12513] |
| ENSG00000176978 | -0.516 | 6.107 | -2.134 | 0.03549 | 0.58061 | -3.970 | DPP7 | dipeptidyl peptidase 7 [Source:HGNC Symbol;Acc:HGNC:14892] |
| ENSG00000163737 | -2.865 | 0.349 | -2.134 | 0.03550 | 0.58061 | -3.715 | PF4 | platelet factor 4 [Source:HGNC Symbol;Acc:HGNC:8861] |
| ENSG00000273173 | -0.638 | 6.357 | -2.133 | 0.03558 | 0.58064 | -3.975 | SNURF | SNRPN upstream reading frame [Source:HGNC Symbol;Acc:HGNC:11171] |
| ENSG00000198728 | -0.427 | 4.532 | -2.133 | 0.03560 | 0.58064 | -3.891 | LDB1 | LIM domain binding 1 [Source:HGNC Symbol;Acc:HGNC:6532] |
| ENSG00000125434 | -0.360 | 2.446 | -2.132 | 0.03567 | 0.58112 | -3.670 | SLC25A35 | solute carrier family 25 member 35 [Source:HGNC Symbol;Acc:HGNC:31921] |
| ENSG00000126460 | 0.875 | 0.035 | 2.131 | 0.03572 | 0.58120 | -3.618 | PRRG2 | proline rich and Gla domain 2 [Source:HGNC Symbol;Acc:HGNC:9470] |
| ENSG00000041982 | -1.531 | 2.030 | -2.130 | 0.03580 | 0.58177 | -3.615 | TNC | tenascin C [Source:HGNC Symbol;Acc:HGNC:5318] |
| ENSG00000082898 | 0.267 | 7.329 | 2.129 | 0.03592 | 0.58211 | -3.966 | XPO1 | exportin 1 [Source:HGNC Symbol;Acc:HGNC:12825] |
| ENSG00000013364 | 0.449 | 5.870 | 2.129 | 0.03594 | 0.58211 | -3.987 | MVP | major vault protein [Source:HGNC Symbol;Acc:HGNC:7531] |
| ENSG00000256269 | 0.586 | 3.320 | 2.128 | 0.03600 | 0.58211 | -3.846 | HMBS | hydroxymethylbilane synthase [Source:HGNC Symbol;Acc:HGNC:4982] |
| ENSG00000167807 | 0.453 | 1.987 | 2.128 | 0.03604 | 0.58211 | -3.685 | AC011511.1 | |
| ENSG00000172059 | -0.971 | 4.893 | -2.127 | 0.03607 | 0.58211 | -3.892 | KLF11 | Kruppel like factor 11 [Source:HGNC Symbol;Acc:HGNC:11811] |
| ENSG00000132376 | -0.370 | 4.329 | -2.127 | 0.03612 | 0.58211 | -3.884 | INPP5K | inositol polyphosphate-5-phosphatase K [Source:HGNC Symbol;Acc:HGNC:33882] |
| ENSG00000197008 | 0.391 | 3.993 | 2.126 | 0.03614 | 0.58211 | -3.901 | ZNF138 | zinc finger protein 138 [Source:HGNC Symbol;Acc:HGNC:12922] |
| ENSG00000204248 | 0.645 | 0.603 | 2.126 | 0.03620 | 0.58227 | -3.622 | COL11A2 | collagen type XI alpha 2 chain [Source:HGNC Symbol;Acc:HGNC:2187] |
| ENSG00000241106 | -1.091 | 6.002 | -2.123 | 0.03644 | 0.58491 | -3.971 | HLA-DOB | major histocompatibility complex, class II, DO beta [Source:HGNC Symbol;Acc:HGNC:4937] |
| ENSG00000187951 | 0.569 | 3.865 | 2.122 | 0.03649 | 0.58491 | -3.909 | ARHGAP11B | Rho GTPase activating protein 11B [Source:HGNC Symbol;Acc:HGNC:15782] |
| ENSG00000257594 | -0.363 | 5.481 | -2.122 | 0.03650 | 0.58491 | -3.974 | GALNT4 | polypeptide N-acetylgalactosaminyltransferase 4 [Source:HGNC Symbol;Acc:HGNC:4126] |
| ENSG00000119640 | 0.487 | 2.645 | 2.121 | 0.03658 | 0.58536 | -3.766 | ACYP1 | acylphosphatase 1 [Source:HGNC Symbol;Acc:HGNC:179] |
| ENSG00000182224 | -0.489 | 4.418 | -2.121 | 0.03662 | 0.58536 | -3.894 | CYB5D1 | cytochrome b5 domain containing 1 [Source:HGNC Symbol;Acc:HGNC:26516] |
| ENSG00000185404 | -0.404 | 5.338 | -2.120 | 0.03670 | 0.58536 | -3.971 | SP140L | SP140 nuclear body protein like [Source:HGNC Symbol;Acc:HGNC:25105] |
| ENSG00000198356 | 0.439 | 4.664 | 2.119 | 0.03674 | 0.58536 | -3.967 | ASNA1 | arsA arsenite transporter, ATP-binding, homolog 1 (bacterial) [Source:HGNC Symbol;Acc:HGNC:752] |
| ENSG00000075914 | 0.314 | 4.492 | 2.119 | 0.03676 | 0.58536 | -3.950 | EXOSC7 | exosome component 7 [Source:HGNC Symbol;Acc:HGNC:28112] |
| ENSG00000136811 | 0.320 | 5.868 | 2.118 | 0.03685 | 0.58544 | -4.007 | ODF2 | outer dense fiber of sperm tails 2 [Source:HGNC Symbol;Acc:HGNC:8114] |
| ENSG00000163131 | -0.563 | 9.021 | -2.118 | 0.03690 | 0.58544 | -3.941 | CTSS | cathepsin S [Source:HGNC Symbol;Acc:HGNC:2545] |
| ENSG00000259371 | 0.395 | 2.772 | 2.118 | 0.03690 | 0.58544 | -3.785 | AL136295.3 | |
| ENSG00000139133 | 0.345 | 3.593 | 2.117 | 0.03695 | 0.58544 | -3.877 | ALG10 | ALG10, alpha-1,2-glucosyltransferase [Source:HGNC Symbol;Acc:HGNC:23162] |
| ENSG00000205413 | 0.565 | 7.128 | 2.115 | 0.03708 | 0.58544 | -3.992 | SAMD9 | sterile alpha motif domain containing 9 [Source:HGNC Symbol;Acc:HGNC:1348] |

| | | | | | | | | |
|---|---|---|---|---|---|---|---|---|
| ENSG00000117643 | -1.495 | 2.505 | -2.115 | 0.03709 | 0.58544 | -3.647 | MAN1C1 | mannosidase alpha class 1C member 1 [Source:HGNC Symbol;Acc:HGNC:19080] |
| ENSG00000138778 | 0.655 | 4.904 | 2.115 | 0.03711 | 0.58544 | -3.993 | CENPE | centromere protein E [Source:HGNC Symbol;Acc:HGNC:1856] |
| ENSG00000020922 | 0.362 | 5.893 | 2.115 | 0.03713 | 0.58544 | -4.013 | MRE11 | MRE11 homolog, double strand break repair nuclease [Source:HGNC Symbol;Acc:HGNC:7230] |
| ENSG00000163435 | 1.409 | 0.815 | 2.113 | 0.03728 | 0.58587 | -3.669 | ELF3 | E74 like ETS transcription factor 3 [Source:HGNC Symbol;Acc:HGNC:3318] |
| ENSG00000182568 | -1.183 | 4.784 | -2.112 | 0.03741 | 0.58587 | -3.898 | SATB1 | SATB homeobox 1 [Source:HGNC Symbol;Acc:HGNC:10541] |
| ENSG00000187024 | 0.694 | 1.063 | 2.111 | 0.03746 | 0.58587 | -3.657 | PTRH1 | peptidyl-tRNA hydrolase 1 homolog [Source:HGNC Symbol;Acc:HGNC:27039] |
| ENSG00000139579 | 0.374 | 3.651 | 2.111 | 0.03746 | 0.58587 | -3.895 | NABP2 | nucleic acid binding protein 2 [Source:HGNC Symbol;Acc:HGNC:28412] |
| ENSG00000091483 | 0.520 | 5.896 | 2.110 | 0.03759 | 0.58587 | -4.024 | FH | fumarate hydratase [Source:HGNC Symbol;Acc:HGNC:3700] |
| ENSG00000088970 | -0.370 | 5.335 | -2.110 | 0.03760 | 0.58587 | -3.992 | KIZ | kizuna centrosomal protein [Source:HGNC Symbol;Acc:HGNC:15865] |
| ENSG00000166289 | 0.974 | 1.378 | 2.108 | 0.03771 | 0.58587 | -3.687 | PLEKHF1 | pleckstrin homology and FYVE domain containing 1 [Source:HGNC Symbol;Acc:HGNC:20764] |
| ENSG00000147231 | 1.092 | 2.276 | 2.107 | 0.03782 | 0.58587 | -3.797 | CXorf57 | chromosome X open reading frame 57 [Source:HGNC Symbol;Acc:HGNC:25486] |
| ENSG00000107341 | -0.292 | 5.626 | -2.107 | 0.03785 | 0.58587 | -4.013 | UBE2R2 | ubiquitin conjugating enzyme E2 R2 [Source:HGNC Symbol;Acc:HGNC:19907] |
| ENSG00000162972 | 0.249 | 3.879 | 2.107 | 0.03785 | 0.58587 | -3.918 | MAIP1 | matrix AAA peptidase interacting protein 1 [Source:HGNC Symbol;Acc:HGNC:26198] |
| ENSG00000115266 | 0.444 | 1.331 | 2.107 | 0.03786 | 0.58587 | -3.669 | APC2 | APC2, WNT signaling pathway regulator [Source:HGNC Symbol;Acc:HGNC:24036] |
| ENSG00000152818 | -0.542 | 8.921 | -2.106 | 0.03790 | 0.58587 | -3.967 | UTRN | utrophin [Source:HGNC Symbol;Acc:HGNC:12635] |
| ENSG00000161011 | -0.437 | 8.131 | -2.106 | 0.03793 | 0.58587 | -3.998 | SQSTM1 | sequestosome 1 [Source:HGNC Symbol;Acc:HGNC:11280] |
| ENSG00000148841 | -0.791 | 5.404 | -2.106 | 0.03793 | 0.58587 | -3.986 | ITPRIP | inositol 1,4,5-trisphosphate receptor interacting protein [Source:HGNC Symbol;Acc:HGNC:29370] |
| ENSG00000198885 | 1.085 | 0.537 | 2.105 | 0.03798 | 0.58587 | -3.658 | ITPRIPL1 | inositol 1,4,5-trisphosphate receptor interacting protein-like 1 [Source:HGNC Symbol;Acc:HGNC:29371] |
| ENSG00000178078 | 0.779 | 0.778 | 2.105 | 0.03801 | 0.58587 | -3.658 | STAP2 | signal transducing adaptor family member 2 [Source:HGNC Symbol;Acc:HGNC:30430] |
| ENSG00000151748 | -0.502 | 4.478 | -2.105 | 0.03802 | 0.58587 | -3.930 | SAV1 | salvador family WW domain containing protein 1 [Source:HGNC Symbol;Acc:HGNC:17795] |
| ENSG00000259316 | 0.258 | 4.288 | 2.105 | 0.03802 | 0.58587 | -3.961 | AC087632.1 | |
| ENSG00000267534 | 0.773 | 2.455 | 2.105 | 0.03803 | 0.58587 | -3.806 | S1PR2 | sphingosine-1-phosphate receptor 2 [Source:HGNC Symbol;Acc:HGNC:3169] |
| ENSG00000139351 | -1.045 | 2.067 | -2.104 | 0.03809 | 0.58597 | -3.665 | SYCP3 | synaptonemal complex protein 3 [Source:HGNC Symbol;Acc:HGNC:18130] |
| ENSG00000136514 | 0.797 | 1.065 | 2.104 | 0.03813 | 0.58597 | -3.674 | RTP4 | receptor transporter protein 4 [Source:HGNC Symbol;Acc:HGNC:23992] |
| ENSG00000139354 | 0.925 | 1.398 | 2.103 | 0.03822 | 0.58674 | -3.708 | GAS2L3 | growth arrest specific 2 like 3 [Source:HGNC Symbol;Acc:HGNC:27475] |
| ENSG00000100320 | -0.540 | 5.189 | -2.102 | 0.03827 | 0.58685 | -3.991 | RBFOX2 | RNA binding protein, fox-1 homolog 2 [Source:HGNC Symbol;Acc:HGNC:9906] |
| ENSG00000183722 | -1.778 | -0.091 | -2.101 | 0.03837 | 0.58714 | -3.758 | LHFP | lipoma HMGIC fusion partner [Source:HGNC Symbol;Acc:HGNC:6586] |
| ENSG00000101412 | 0.732 | 2.078 | 2.101 | 0.03839 | 0.58714 | -3.762 | E2F1 | E2F transcription factor 1 [Source:HGNC Symbol;Acc:HGNC:3113] |
| ENSG00000121211 | 0.723 | 1.755 | 2.100 | 0.03849 | 0.58801 | -3.731 | MND1 | meiotic nuclear divisions 1 [Source:HGNC Symbol;Acc:HGNC:24839] |
| ENSG00000188641 | -0.421 | 6.526 | -2.099 | 0.03854 | 0.58803 | -4.044 | DPYD | dihydropyrimidine dehydrogenase [Source:HGNC Symbol;Acc:HGNC:3012] |
| ENSG00000110880 | -0.513 | 6.442 | -2.097 | 0.03870 | 0.58924 | -4.046 | CORO1C | coronin 1C [Source:HGNC Symbol;Acc:HGNC:2254] |
| ENSG00000130724 | 0.343 | 5.831 | 2.097 | 0.03871 | 0.58924 | -4.047 | CHMP2A | charged multivesicular body protein 2A [Source:HGNC Symbol;Acc:HGNC:30216] |
| ENSG00000162430 | -0.831 | 2.285 | -2.095 | 0.03893 | 0.59007 | -3.691 | SELENON | selenoprotein N [Source:HGNC Symbol;Acc:HGNC:15999] |
| ENSG00000146263 | 0.526 | 4.265 | 2.095 | 0.03894 | 0.59007 | -3.993 | MMS22L | MMS22 like, DNA repair protein [Source:HGNC Symbol;Acc:HGNC:21475] |
| ENSG00000069493 | -0.793 | 5.046 | -2.093 | 0.03910 | 0.59007 | -3.982 | CLEC2D | C-type lectin domain family 2 member D [Source:HGNC Symbol;Acc:HGNC:14351] |
| ENSG00000159131 | 0.377 | 5.939 | 2.093 | 0.03910 | 0.59007 | -4.056 | GART | phosphoribosylglycinamide formyltransferase, phosphoribosylglycinamide synthetase, phosphoribosylaminoimidazole synthetase [Source:HGNC Symbol;Acc:HGNC:4163] |
| ENSG00000198832 | -0.902 | 3.748 | -2.093 | 0.03914 | 0.59007 | -3.828 | SELENOM | selenoprotein M [Source:HGNC Symbol;Acc:HGNC:30397] |
| ENSG00000127423 | 0.435 | 2.230 | 2.092 | 0.03920 | 0.59007 | -3.774 | AUNIP | aurora kinase A and ninein interacting protein [Source:HGNC Symbol;Acc:HGNC:28363] |
| ENSG00000196365 | 0.431 | 6.141 | 2.092 | 0.03920 | 0.59007 | -4.059 | LONP1 | lon peptidase 1, mitochondrial [Source:HGNC Symbol;Acc:HGNC:9479] |
| ENSG00000100207 | -0.306 | 5.956 | -2.092 | 0.03921 | 0.59007 | -4.052 | TCF20 | transcription factor 20 [Source:HGNC Symbol;Acc:HGNC:11631] |
| ENSG00000149054 | -0.683 | 6.228 | -2.092 | 0.03922 | 0.59007 | -4.051 | ZNF215 | zinc finger protein 215 [Source:HGNC Symbol;Acc:HGNC:13007] |
| ENSG00000173406 | -0.444 | 3.922 | -2.092 | 0.03922 | 0.59007 | -3.896 | DAB1 | DAB1, reelin adaptor protein [Source:HGNC Symbol;Acc:HGNC:2661] |
| ENSG00000085840 | 0.663 | 2.637 | 2.090 | 0.03934 | 0.59098 | -3.846 | ORC1 | origin recognition complex subunit 1 [Source:HGNC Symbol;Acc:HGNC:8487] |
| ENSG00000125347 | -0.620 | 8.896 | -2.090 | 0.03939 | 0.59098 | -4.000 | IRF1 | interferon regulatory factor 1 [Source:HGNC Symbol;Acc:HGNC:6116] |
| ENSG00000139289 | -1.322 | 5.012 | -2.089 | 0.03942 | 0.59098 | -3.955 | PHLDA1 | pleckstrin homology like domain family A member 1 [Source:HGNC Symbol;Acc:HGNC:8933] |
| ENSG00000187815 | 0.676 | 2.388 | 2.089 | 0.03948 | 0.59107 | -3.808 | ZFP69 | ZFP69 zinc finger protein [Source:HGNC Symbol;Acc:HGNC:24708] |
| ENSG00000277791 | 0.376 | 5.342 | 2.088 | 0.03955 | 0.59130 | -4.055 | PSMB3 | proteasome subunit beta 3 [Source:HGNC Symbol;Acc:HGNC:9540] |
| ENSG00000187240 | 0.887 | 2.782 | 2.088 | 0.03958 | 0.59130 | -3.871 | DYNC2H1 | dynein cytoplasmic 2 heavy chain 1 [Source:HGNC Symbol;Acc:HGNC:2962] |
| ENSG00000205683 | 1.228 | 1.042 | 2.086 | 0.03970 | 0.59231 | -3.715 | DPF3 | double PHD fingers 3 [Source:HGNC Symbol;Acc:HGNC:17427] |
| ENSG00000084693 | -0.347 | 5.416 | -2.084 | 0.03995 | 0.59545 | -4.047 | AGBL5 | ATP/GTP binding protein like 5 [Source:HGNC Symbol;Acc:HGNC:26147] |
| ENSG00000250312 | -1.246 | 2.140 | -2.082 | 0.04007 | 0.59611 | -3.699 | ZNF718 | zinc finger protein 718 [Source:HGNC Symbol;Acc:HGNC:26889] |
| ENSG00000115677 | -0.376 | 10.417 | -2.082 | 0.04013 | 0.59611 | -3.946 | HDLBP | high density lipoprotein binding protein [Source:HGNC Symbol;Acc:HGNC:4857] |
| ENSG00000138092 | 0.420 | 3.560 | 2.081 | 0.04018 | 0.59611 | -3.951 | CENPO | centromere protein O [Source:HGNC Symbol;Acc:HGNC:28152] |
| ENSG00000047056 | -0.320 | 4.208 | -2.081 | 0.04019 | 0.59611 | -3.963 | WDR37 | WD repeat domain 37 [Source:HGNC Symbol;Acc:HGNC:31406] |
| ENSG00000165914 | -1.360 | 2.044 | -2.081 | 0.04024 | 0.59611 | -3.696 | TTC7B | tetratricopeptide repeat domain 7B [Source:HGNC Symbol;Acc:HGNC:19858] |
| ENSG00000146674 | -1.440 | 0.665 | -2.080 | 0.04028 | 0.59611 | -3.735 | IGFBP3 | insulin like growth factor binding protein 3 [Source:HGNC Symbol;Acc:HGNC:5472] |
| ENSG00000141401 | 0.655 | 2.040 | 2.077 | 0.04062 | 0.59885 | -3.799 | IMPA2 | inositol monophosphatase 2 [Source:HGNC Symbol;Acc:HGNC:6051] |
| ENSG00000162804 | -0.388 | 3.511 | -2.076 | 0.04065 | 0.59885 | -3.889 | SNED1 | sushi, nidogen and EGF like domains 1 [Source:HGNC Symbol;Acc:HGNC:24696] |
| ENSG00000129625 | -0.451 | 7.311 | -2.076 | 0.04070 | 0.59885 | -4.082 | REEP5 | receptor accessory protein 5 [Source:HGNC Symbol;Acc:HGNC:30077] |
| ENSG00000144034 | 0.423 | 4.450 | 2.076 | 0.04070 | 0.59885 | -4.035 | TPRKB | TP53RK binding protein [Source:HGNC Symbol;Acc:HGNC:24259] |
| ENSG00000118046 | 0.374 | 4.947 | 2.075 | 0.04074 | 0.59885 | -4.064 | STK11 | serine/threonine kinase 11 [Source:HGNC Symbol;Acc:HGNC:11389] |
| ENSG00000163507 | 0.592 | 5.123 | 2.075 | 0.04078 | 0.59885 | -4.076 | KIAA1524 | KIAA1524 [Source:HGNC Symbol;Acc:HGNC:29302] |
| ENSG00000099995 | -0.361 | 5.088 | -2.075 | 0.04081 | 0.59885 | -4.047 | SF3A1 | splicing factor 3a subunit 1 [Source:HGNC Symbol;Acc:HGNC:10765] |
| ENSG00000170234 | -0.283 | 5.224 | -2.074 | 0.04084 | 0.59885 | -4.057 | PWWP2A | PWWP domain containing 2A [Source:HGNC Symbol;Acc:HGNC:29406] |
| ENSG00000198056 | 0.479 | 3.565 | 2.073 | 0.04098 | 0.59961 | -3.968 | PRIM1 | primase (DNA) subunit 1 [Source:HGNC Symbol;Acc:HGNC:9369] |
| ENSG00000173041 | 0.809 | 3.026 | 2.073 | 0.04099 | 0.59961 | -3.931 | ZNF680 | zinc finger protein 680 [Source:HGNC Symbol;Acc:HGNC:26897] |
| ENSG00000116032 | 0.661 | 0.165 | 2.070 | 0.04125 | 0.60280 | -3.710 | GRIN3B | glutamate ionotropic receptor NMDA type subunit 3B [Source:HGNC Symbol;Acc:HGNC:16768] |
| ENSG00000127311 | 0.442 | 6.073 | 2.069 | 0.04135 | 0.60311 | -4.102 | HELB | DNA helicase B [Source:HGNC Symbol;Acc:HGNC:17196] |
| ENSG00000168763 | -0.368 | 3.920 | -2.069 | 0.04137 | 0.60311 | -3.952 | CNNM3 | cyclin and CBS domain divalent metal cation transport mediator 3 [Source:HGNC Symbol;Acc:HGNC:104] |
| ENSG00000277462 | 0.661 | 0.925 | 2.067 | 0.04157 | 0.60466 | -3.723 | ZNF670 | zinc finger protein 670 [Source:HGNC Symbol;Acc:HGNC:28167] |
| ENSG00000072422 | -2.353 | 1.349 | -2.066 | 0.04162 | 0.60466 | -3.743 | RHOBTB1 | Rho related BTB domain containing 1 [Source:HGNC Symbol;Acc:HGNC:18738] |
| ENSG00000173930 | -1.304 | 1.938 | -2.065 | 0.04174 | 0.60466 | -3.718 | SLCO4C1 | solute carrier organic anion transporter family member 4C1 [Source:HGNC Symbol;Acc:HGNC:23612] |
| ENSG00000176208 | 0.512 | 4.427 | 2.065 | 0.04175 | 0.60466 | -4.059 | ATAD5 | ATPase family, AAA domain containing 5 [Source:HGNC Symbol;Acc:HGNC:25752] |
| ENSG00000123159 | 0.621 | 2.062 | 2.064 | 0.04182 | 0.60466 | -3.818 | GIPC1 | GIPC PDZ domain containing family member 1 [Source:HGNC Symbol;Acc:HGNC:1226] |
| ENSG00000196866 | 0.713 | 4.550 | 2.064 | 0.04185 | 0.60466 | -4.076 | HIST1H2AD | histone cluster 1 H2A family member d [Source:HGNC Symbol;Acc:HGNC:4729] |
| ENSG00000196976 | 0.713 | 1.578 | 2.064 | 0.04186 | 0.60466 | -3.773 | LAGE3 | L antigen family member 3 [Source:HGNC Symbol;Acc:HGNC:26058] |
| ENSG00000148459 | 0.390 | 2.878 | 2.063 | 0.04188 | 0.60466 | -3.898 | PDSS1 | decaprenyl diphosphate synthase subunit 1 [Source:HGNC Symbol;Acc:HGNC:17759] |
| ENSG00000129084 | 0.341 | 6.734 | 2.061 | 0.04208 | 0.60466 | -4.111 | PSMA1 | proteasome subunit alpha 1 [Source:HGNC Symbol;Acc:HGNC:9530] |
| ENSG00000105889 | 1.114 | 0.887 | 2.061 | 0.04212 | 0.60466 | -3.752 | STEAP1B | STEAP family member 1B [Source:HGNC Symbol;Acc:HGNC:41907] |
| ENSG00000105607 | 0.363 | 4.690 | 2.058 | 0.04236 | 0.60466 | -4.081 | GCDH | glutaryl-CoA dehydrogenase [Source:HGNC Symbol;Acc:HGNC:4189] |
| ENSG00000104980 | 0.345 | 5.263 | 2.058 | 0.04240 | 0.60466 | -4.109 | TIMM44 | translocase of inner mitochondrial membrane 44 [Source:HGNC Symbol;Acc:HGNC:17316] |
| ENSG00000177105 | -0.700 | 2.129 | -2.058 | 0.04240 | 0.60466 | -3.760 | RHOG | ras homolog family member G [Source:HGNC Symbol;Acc:HGNC:672] |
| ENSG00000100519 | 0.306 | 7.015 | 2.057 | 0.04248 | 0.60466 | -4.112 | PSMC6 | proteasome 26S subunit, ATPase 6 [Source:HGNC Symbol;Acc:HGNC:9553] |
| ENSG00000121621 | 0.558 | 3.908 | 2.057 | 0.04253 | 0.60466 | -4.034 | KIF18A | kinesin family member 18A [Source:HGNC Symbol;Acc:HGNC:29441] |
| ENSG00000119906 | -0.348 | 5.634 | -2.056 | 0.04257 | 0.60466 | -4.107 | SLF2 | SMC5-SMC6 complex localization factor 2 [Source:HGNC Symbol;Acc:HGNC:17814] |
| ENSG00000139329 | -1.783 | 0.076 | -2.055 | 0.04267 | 0.60466 | -3.808 | LUM | lumican [Source:HGNC Symbol;Acc:HGNC:6724] |
| ENSG00000138111 | 0.667 | -0.121 | 2.055 | 0.04270 | 0.60466 | -3.737 | MFSD13A | major facilitator superfamily domain containing 13A [Source:HGNC Symbol;Acc:HGNC:26196] |
| ENSG00000128578 | 0.777 | 0.314 | 2.055 | 0.04272 | 0.60466 | -3.732 | STRIP2 | striatin interacting protein 2 [Source:HGNC Symbol;Acc:HGNC:22209] |
| ENSG00000113569 | 0.298 | 6.025 | 2.055 | 0.04275 | 0.60466 | -4.130 | NUP155 | nucleoporin 155 [Source:HGNC Symbol;Acc:HGNC:8063] |
| ENSG00000221963 | -0.530 | 7.993 | -2.054 | 0.04276 | 0.60466 | -4.103 | APOL6 | apolipoprotein L6 [Source:HGNC Symbol;Acc:HGNC:14870] |

| | | | | | | | | |
|---|---|---|---|---|---|---|---|---|
| ENSG00000258555 | -0.363 | 5.725 | -2.054 | 0.04285 | 0.60466 | -4.116 | SPECC1L-ADORA2A | SPECC1L-ADORA2A readthrough (NMD candidate) [Source:HGNC Symbol;Acc:HGNC:49185] |
| ENSG00000166169 | -0.391 | 3.426 | -2.053 | 0.04290 | 0.60466 | -3.922 | POLL | DNA polymerase lambda [Source:HGNC Symbol;Acc:HGNC:9184] |
| ENSG00000166340 | -0.495 | 4.436 | -2.053 | 0.04292 | 0.60466 | -4.028 | TPP1 | tripeptidyl peptidase 1 [Source:HGNC Symbol;Acc:HGNC:2073] |
| ENSG00000128739 | -0.608 | 6.672 | -2.052 | 0.04296 | 0.60466 | -4.133 | SNRPN | small nuclear ribonucleoprotein polypeptide N [Source:HGNC Symbol;Acc:HGNC:11164] |
| ENSG00000185883 | -0.608 | 2.716 | -2.051 | 0.04312 | 0.60466 | -3.830 | ATP6V0C | ATPase H+ transporting V0 subunit c [Source:HGNC Symbol;Acc:HGNC:855] |
| ENSG00000115286 | 0.425 | 3.780 | 2.050 | 0.04316 | 0.60466 | -4.027 | NDUFS7 | NADH:ubiquinone oxidoreductase core subunit S7 [Source:HGNC Symbol;Acc:HGNC:7714] |
| ENSG00000158604 | -0.311 | 7.184 | -2.050 | 0.04319 | 0.60466 | -4.132 | TMED4 | transmembrane p24 trafficking protein 4 [Source:HGNC Symbol;Acc:HGNC:22301] |
| ENSG00000136824 | 0.461 | 5.798 | 2.049 | 0.04326 | 0.60466 | -4.138 | SMC2 | structural maintenance of chromosomes 2 [Source:HGNC Symbol;Acc:HGNC:14011] |
| ENSG00000197050 | 0.620 | 2.410 | 2.049 | 0.04331 | 0.60466 | -3.883 | ZNF420 | zinc finger protein 420 [Source:HGNC Symbol;Acc:HGNC:20649] |
| ENSG00000165060 | -0.490 | 3.813 | -2.049 | 0.04331 | 0.60466 | -3.964 | FXN | frataxin [Source:HGNC Symbol;Acc:HGNC:3951] |
| ENSG00000154310 | -1.563 | 1.812 | -2.048 | 0.04336 | 0.60466 | -3.741 | TNIK | TRAF2 and NCK interacting kinase [Source:HGNC Symbol;Acc:HGNC:30765] |
| ENSG00000273734 | 0.387 | 5.399 | 2.048 | 0.04337 | 0.60466 | -4.132 | AC005258.1 | |
| ENSG00000174243 | 0.271 | 6.697 | 2.047 | 0.04347 | 0.60466 | -4.139 | DDX23 | DEAD-box helicase 23 [Source:HGNC Symbol;Acc:HGNC:17347] |
| ENSG00000126254 | 0.331 | 3.891 | 2.047 | 0.04349 | 0.60466 | -4.040 | RBM42 | RNA binding motif protein 42 [Source:HGNC Symbol;Acc:HGNC:28117] |
| ENSG00000160408 | -0.591 | 4.498 | -2.046 | 0.04355 | 0.60466 | -4.039 | ST6GALNAC6 | ST6 N-acetylgalactosaminide alpha-2,6-sialyltransferase 6 [Source:HGNC Symbol;Acc:HGNC:23364] |
| ENSG00000116678 | -0.696 | 4.496 | -2.046 | 0.04357 | 0.60466 | -4.030 | LEPR | leptin receptor [Source:HGNC Symbol;Acc:HGNC:6554] |
| ENSG00000103051 | 0.376 | 4.772 | 2.045 | 0.04370 | 0.60466 | -4.111 | COG4 | component of oligomeric golgi complex 4 [Source:HGNC Symbol;Acc:HGNC:18620] |
| ENSG00000145016 | -0.398 | 6.347 | -2.045 | 0.04372 | 0.60466 | -4.146 | RUBCN | RUN and cysteine rich domain containing beclin 1 interacting protein [Source:HGNC Symbol;Acc:HGNC:28991] |
| ENSG00000172650 | -0.419 | 2.688 | -2.044 | 0.04376 | 0.60466 | -3.842 | AGAP5 | ArfGAP with GTPase domain, ankyrin repeat and PH domain 5 [Source:HGNC Symbol;Acc:HGNC:23467] |
| ENSG00000145545 | 0.384 | 2.532 | 2.044 | 0.04384 | 0.60466 | -3.890 | SRD5A1 | steroid 5 alpha-reductase 1 [Source:HGNC Symbol;Acc:HGNC:11284] |
| ENSG00000186310 | 1.091 | 0.044 | 2.043 | 0.04391 | 0.60466 | -3.749 | NAP1L3 | nucleosome assembly protein 1 like 3 [Source:HGNC Symbol;Acc:HGNC:7639] |
| ENSG00000128513 | 0.303 | 4.930 | 2.042 | 0.04400 | 0.60466 | -4.123 | POT1 | protection of telomeres 1 [Source:HGNC Symbol;Acc:HGNC:17284] |
| ENSG00000179965 | 0.457 | 1.583 | 2.042 | 0.04402 | 0.60466 | -3.792 | ZNF771 | zinc finger protein 771 [Source:HGNC Symbol;Acc:HGNC:29653] |
| ENSG00000198162 | -0.347 | 9.061 | -2.041 | 0.04412 | 0.60466 | -4.078 | MAN1A2 | mannosidase alpha class 1A member 2 [Source:HGNC Symbol;Acc:HGNC:6822] |
| ENSG00000079246 | 0.290 | 8.038 | 2.040 | 0.04420 | 0.60466 | -4.106 | XRCC5 | X-ray repair cross complementing 5 [Source:HGNC Symbol;Acc:HGNC:12833] |
| ENSG00000086827 | 0.305 | 5.012 | 2.040 | 0.04421 | 0.60466 | -4.130 | ZW10 | zw10 kinetochore protein [Source:HGNC Symbol;Acc:HGNC:13194] |
| ENSG00000138346 | 0.451 | 3.764 | 2.040 | 0.04422 | 0.60466 | -4.047 | DNA2 | DNA replication helicase/nuclease 2 [Source:HGNC Symbol;Acc:HGNC:2939] |
| ENSG00000162889 | -0.507 | 7.771 | -2.040 | 0.04422 | 0.60466 | -4.138 | MAPKAPK2 | mitogen-activated protein kinase-activated protein kinase 2 [Source:HGNC Symbol;Acc:HGNC:6887] |
| ENSG00000188895 | -0.291 | 6.074 | -2.040 | 0.04422 | 0.60466 | -4.153 | MSL1 | male specific lethal 1 homolog [Source:HGNC Symbol;Acc:HGNC:27905] |
| ENSG00000127989 | 0.318 | 3.760 | 2.039 | 0.04428 | 0.60466 | -4.036 | MTERF1 | mitochondrial transcription termination factor 1 [Source:HGNC Symbol;Acc:HGNC:21463] |
| ENSG00000134265 | 0.278 | 6.125 | 2.039 | 0.04431 | 0.60466 | -4.159 | NAPG | NSF attachment protein gamma [Source:HGNC Symbol;Acc:HGNC:7642] |
| ENSG00000160179 | 0.975 | 2.034 | 2.039 | 0.04436 | 0.60466 | -3.883 | ABCG1 | ATP binding cassette subfamily G member 1 [Source:HGNC Symbol;Acc:HGNC:73] |
| ENSG00000107902 | -0.498 | 2.387 | -2.038 | 0.04443 | 0.60466 | -3.820 | LHPP | phospholysine phosphohistidine inorganic pyrophosphate phosphatase [Source:HGNC Symbol;Acc:HGNC:30042] |
| ENSG00000048828 | -0.254 | 6.757 | -2.038 | 0.04444 | 0.60466 | -4.161 | FAM120A | family with sequence similarity 120A [Source:HGNC Symbol;Acc:HGNC:13247] |
| ENSG00000187699 | -0.560 | 6.286 | -2.037 | 0.04449 | 0.60466 | -4.157 | C2orf88 | chromosome 2 open reading frame 88 [Source:HGNC Symbol;Acc:HGNC:28191] |
| ENSG00000158258 | 1.370 | 0.715 | 2.035 | 0.04467 | 0.60466 | -3.792 | CLSTN2 | calsyntenin 2 [Source:HGNC Symbol;Acc:HGNC:17448] |
| ENSG00000138587 | 0.861 | 3.121 | 2.035 | 0.04470 | 0.60466 | -4.010 | MNS1 | meiosis specific nuclear structural 1 [Source:HGNC Symbol;Acc:HGNC:29636] |
| ENSG00000181991 | 0.353 | 4.227 | 2.035 | 0.04475 | 0.60466 | -4.092 | MRPS11 | mitochondrial ribosomal protein S11 [Source:HGNC Symbol;Acc:HGNC:14050] |
| ENSG00000175826 | -0.316 | 4.189 | -2.034 | 0.04478 | 0.60466 | -4.047 | CTDNEP1 | CTD nuclear envelope phosphatase 1 [Source:HGNC Symbol;Acc:HGNC:19085] |
| ENSG00000176222 | 0.718 | 1.172 | 2.034 | 0.04479 | 0.60466 | -3.790 | ZNF404 | zinc finger protein 404 [Source:HGNC Symbol;Acc:HGNC:19417] |
| ENSG00000204592 | -0.456 | 9.977 | -2.034 | 0.04479 | 0.60466 | -4.054 | HLA-E | major histocompatibility complex, class I, E [Source:HGNC Symbol;Acc:HGNC:4962] |
| ENSG00000100902 | 0.431 | 7.435 | 2.034 | 0.04487 | 0.60466 | -4.139 | PSMA6 | proteasome subunit alpha 6 [Source:HGNC Symbol;Acc:HGNC:9535] |
| ENSG00000094880 | 0.314 | 4.729 | 2.033 | 0.04491 | 0.60466 | -4.130 | CDC23 | cell division cycle 23 [Source:HGNC Symbol;Acc:HGNC:1724] |
| ENSG00000119705 | 0.402 | 5.193 | 2.033 | 0.04496 | 0.60466 | -4.154 | SLIRP | SRA stem-loop interacting RNA binding protein [Source:HGNC Symbol;Acc:HGNC:20495] |
| ENSG00000243710 | 0.476 | 1.539 | 2.032 | 0.04501 | 0.60466 | -3.807 | CFAP57 | cilia and flagella associated protein 57 [Source:HGNC Symbol;Acc:HGNC:26485] |
| ENSG00000070366 | -0.364 | 5.189 | -2.032 | 0.04501 | 0.60466 | -4.130 | SMG6 | SMG6, nonsense mediated mRNA decay factor [Source:HGNC Symbol;Acc:HGNC:17809] |
| ENSG00000164105 | 0.494 | 3.735 | 2.032 | 0.04505 | 0.60466 | -4.058 | SAP30 | Sin3A associated protein 30 [Source:HGNC Symbol;Acc:HGNC:10532] |
| ENSG00000116830 | 0.392 | 4.876 | 2.032 | 0.04507 | 0.60466 | -4.143 | TTF2 | transcription termination factor 2 [Source:HGNC Symbol;Acc:HGNC:12398] |
| ENSG00000126249 | 0.518 | 3.102 | 2.031 | 0.04511 | 0.60466 | -3.993 | PDCD2L | programmed cell death 2 like [Source:HGNC Symbol;Acc:HGNC:28194] |
| ENSG00000153815 | -0.639 | 3.216 | -2.030 | 0.04527 | 0.60466 | -3.923 | CMIP | c-Maf inducing protein [Source:HGNC Symbol;Acc:HGNC:24319] |
| ENSG00000260007 | 0.681 | 3.799 | 2.029 | 0.04530 | 0.60466 | -4.076 | AC107871.1 | |
| ENSG00000100219 | -0.459 | 10.090 | -2.029 | 0.04531 | 0.60466 | -4.059 | XBP1 | X-box binding protein 1 [Source:HGNC Symbol;Acc:HGNC:12801] |
| ENSG00000125249 | 0.619 | 4.760 | 2.029 | 0.04537 | 0.60466 | -4.149 | RAP2A | RAP2A, member of RAS oncogene family [Source:HGNC Symbol;Acc:HGNC:9861] |
| ENSG00000164442 | -0.582 | 7.412 | -2.028 | 0.04543 | 0.60466 | -4.171 | CITED2 | Cbp/p300 interacting transactivator with Glu/Asp rich carboxy-terminal domain 2 [Source:HGNC Symbol;Acc:HGNC:1987] |
| ENSG00000106992 | -0.691 | 2.516 | -2.028 | 0.04547 | 0.60466 | -3.836 | AK1 | adenylate kinase 1 [Source:HGNC Symbol;Acc:HGNC:361] |
| ENSG00000135968 | -0.356 | 7.334 | -2.027 | 0.04551 | 0.60466 | -4.172 | GCC2 | GRIP and coiled-coil domain containing 2 [Source:HGNC Symbol;Acc:HGNC:23218] |
| ENSG00000278311 | -0.336 | 7.082 | -2.027 | 0.04551 | 0.60466 | -4.177 | GGNBP2 | gametogenetin binding protein 2 [Source:HGNC Symbol;Acc:HGNC:19357] |
| ENSG00000123091 | -0.449 | 7.119 | -2.027 | 0.04551 | 0.60466 | -4.177 | RNF11 | ring finger protein 11 [Source:HGNC Symbol;Acc:HGNC:10056] |
| ENSG00000197852 | -0.715 | 2.118 | -2.027 | 0.04554 | 0.60466 | -3.812 | FAM212B | family with sequence similarity 212 member B [Source:HGNC Symbol;Acc:HGNC:28045] |
| ENSG00000129007 | 0.538 | 5.425 | 2.027 | 0.04555 | 0.60466 | -4.174 | CALML4 | calmodulin like 4 [Source:HGNC Symbol;Acc:HGNC:18445] |
| ENSG00000183479 | 0.401 | 3.313 | 2.027 | 0.04556 | 0.60466 | -4.017 | TREX2 | three prime repair exonuclease 2 [Source:HGNC Symbol;Acc:HGNC:12270] |
| ENSG00000095059 | 0.295 | 5.099 | 2.027 | 0.04556 | 0.60466 | -4.159 | DHPS | deoxyhypusine synthase [Source:HGNC Symbol;Acc:HGNC:2869] |
| ENSG00000096717 | -0.333 | 5.820 | -2.027 | 0.04558 | 0.60466 | -4.170 | SIRT1 | sirtuin 1 [Source:HGNC Symbol;Acc:HGNC:14929] |
| ENSG00000144130 | 0.643 | 0.034 | 2.027 | 0.04560 | 0.60466 | -3.775 | NT5DC4 | 5'-nucleotidase domain containing 4 [Source:HGNC Symbol;Acc:HGNC:27678] |
| ENSG00000132792 | 0.271 | 4.609 | 2.026 | 0.04563 | 0.60466 | -4.133 | CTNNBL1 | catenin beta like 1 [Source:HGNC Symbol;Acc:HGNC:15879] |
| ENSG00000185347 | 0.625 | 1.884 | 2.026 | 0.04563 | 0.60466 | -3.860 | C14orf80 | chromosome 14 open reading frame 80 [Source:HGNC Symbol;Acc:HGNC:20127] |
| ENSG00000163558 | 0.312 | 5.938 | 2.026 | 0.04568 | 0.60466 | -4.183 | PRKCI | protein kinase C iota [Source:HGNC Symbol;Acc:HGNC:9404] |
| ENSG00000159055 | 0.540 | 2.258 | 2.024 | 0.04583 | 0.60466 | -3.897 | MIS18A | MIS18 kinetochore protein A [Source:HGNC Symbol;Acc:HGNC:1286] |
| ENSG00000014641 | 0.300 | 5.972 | 2.022 | 0.04603 | 0.60466 | -4.189 | MDH1 | malate dehydrogenase 1 [Source:HGNC Symbol;Acc:HGNC:6970] |
| ENSG00000204406 | -0.391 | 4.504 | -2.022 | 0.04604 | 0.60466 | -4.095 | MBD5 | methyl-CpG binding domain protein 5 [Source:HGNC Symbol;Acc:HGNC:20444] |
| ENSG00000140740 | 0.308 | 6.530 | 2.022 | 0.04611 | 0.60466 | -4.189 | UQCRC2 | ubiquinol-cytochrome c reductase core protein II [Source:HGNC Symbol;Acc:HGNC:12586] |
| ENSG00000104324 | -1.123 | 4.401 | -2.022 | 0.04612 | 0.60466 | -4.018 | CPQ | carboxypeptidase Q [Source:HGNC Symbol;Acc:HGNC:16910] |
| ENSG00000111726 | 0.408 | 3.751 | 2.021 | 0.04614 | 0.60466 | -4.077 | CMAS | cytidine monophosphate N-acetylneuraminic acid synthetase [Source:HGNC Symbol;Acc:HGNC:18290] |
| ENSG00000160298 | 0.796 | 0.846 | 2.020 | 0.04633 | 0.60466 | -3.802 | C21orf58 | chromosome 21 open reading frame 58 [Source:HGNC Symbol;Acc:HGNC:1300] |
| ENSG00000184792 | -1.168 | 1.187 | -2.019 | 0.04637 | 0.60466 | -3.785 | OSBP2 | oxysterol binding protein 2 [Source:HGNC Symbol;Acc:HGNC:8504] |
| ENSG00000075975 | 0.238 | 5.440 | 2.019 | 0.04641 | 0.60466 | -4.186 | MKRN2 | makorin ring finger protein 2 [Source:HGNC Symbol;Acc:HGNC:7113] |
| ENSG00000166337 | -0.316 | 5.992 | -2.018 | 0.04645 | 0.60466 | -4.191 | TAF10 | TATA-box binding protein associated factor 10 [Source:HGNC Symbol;Acc:HGNC:11543] |
| ENSG00000117280 | 0.906 | 3.219 | 2.018 | 0.04651 | 0.60466 | -4.058 | RAB29 | RAB29, member RAS oncogene family [Source:HGNC Symbol;Acc:HGNC:9789] |
| ENSG00000151612 | -0.460 | 4.903 | -2.018 | 0.04652 | 0.60466 | -4.135 | ZNF827 | zinc finger protein 827 [Source:HGNC Symbol;Acc:HGNC:27193] |
| ENSG00000181924 | 0.414 | 4.270 | 2.017 | 0.04657 | 0.60466 | -4.129 | COA4 | cytochrome c oxidase assembly factor 4 homolog [Source:HGNC Symbol;Acc:HGNC:24604] |
| ENSG00000125388 | -0.609 | 2.774 | -2.017 | 0.04658 | 0.60466 | -3.877 | GRK4 | G protein-coupled receptor kinase 4 [Source:HGNC Symbol;Acc:HGNC:4543] |
| ENSG00000204599 | -0.318 | 4.970 | -2.017 | 0.04664 | 0.60466 | -4.148 | TRIM39 | tripartite motif containing 39 [Source:HGNC Symbol;Acc:HGNC:10065] |
| ENSG00000160404 | -0.376 | 3.138 | -2.016 | 0.04672 | 0.60466 | -3.948 | TOR2A | torsin family 2 member A [Source:HGNC Symbol;Acc:HGNC:11996] |
| ENSG00000116205 | -0.419 | 3.418 | -2.016 | 0.04674 | 0.60466 | -3.983 | TCEANC2 | transcription elongation factor A N-terminal and central domain containing 2 [Source:HGNC Symbol;Acc:HGNC:26494] |
| ENSG00000107951 | 0.286 | 5.123 | 2.016 | 0.04675 | 0.60466 | -4.181 | MTPAP | mitochondrial poly(A) polymerase [Source:HGNC Symbol;Acc:HGNC:25532] |
| ENSG00000148719 | -0.224 | 5.767 | -2.015 | 0.04678 | 0.60466 | -4.192 | DNAJB12 | DnaJ heat shock protein family (Hsp40) member B12 [Source:HGNC Symbol;Acc:HGNC:14891] |
| ENSG00000150991 | -0.385 | 11.772 | -2.015 | 0.04684 | 0.60466 | -4.015 | UBC | ubiquitin C [Source:HGNC Symbol;Acc:HGNC:12468] |
| ENSG00000067191 | 0.839 | 0.589 | 2.014 | 0.04693 | 0.60466 | -3.802 | CACNB1 | calcium voltage-gated channel auxiliary subunit beta 1 [Source:HGNC Symbol;Acc:HGNC:1401] |
| ENSG00000141385 | 0.287 | 6.365 | 2.013 | 0.04701 | 0.60466 | -4.207 | AFG3L2 | AFG3 like matrix AAA peptidase subunit 2 [Source:HGNC Symbol;Acc:HGNC:315] |
| ENSG00000137692 | 0.443 | 4.694 | 2.013 | 0.04705 | 0.60466 | -4.169 | DCUN1D5 | defective in cullin neddylation 1 domain containing 5 [Source:HGNC Symbol;Acc:HGNC:28409] |

| | | | | | | | | |
|---|---|---|---|---|---|---|---|---|
| ENSG00000138757 | -0.329 | 7.758 | -2.012 | 0.04712 | 0.60466 | -4.186 | G3BP2 | G3BP stress granule assembly factor 2 [Source:HGNC Symbol;Acc:HGNC:30291] |
| ENSG00000186472 | 1.402 | 0.510 | 2.012 | 0.04713 | 0.60466 | -3.808 | PCLO | piccolo presynaptic cytomatrix protein [Source:HGNC Symbol;Acc:HGNC:13406] |
| ENSG00000170100 | -0.483 | 3.112 | -2.010 | 0.04731 | 0.60466 | -3.961 | ZNF778 | zinc finger protein 778 [Source:HGNC Symbol;Acc:HGNC:26479] |
| ENSG00000145882 | 0.684 | 1.771 | 2.010 | 0.04736 | 0.60466 | -3.887 | PCYOX1L | prenylcysteine oxidase 1 like [Source:HGNC Symbol;Acc:HGNC:28477] |
| ENSG00000176401 | 0.407 | 1.896 | 2.010 | 0.04737 | 0.60466 | -3.875 | EID2B | EP300 interacting inhibitor of differentiation 2B [Source:HGNC Symbol;Acc:HGNC:26796] |
| ENSG00000139180 | 0.293 | 5.470 | 2.009 | 0.04744 | 0.60466 | -4.205 | NDUFA9 | NADH:ubiquinone oxidoreductase subunit A9 [Source:HGNC Symbol;Acc:HGNC:7693] |
| ENSG00000079950 | -0.341 | 5.918 | -2.009 | 0.04751 | 0.60466 | -4.206 | STX7 | syntaxin 7 [Source:HGNC Symbol;Acc:HGNC:11442] |
| ENSG00000175279 | 0.404 | 2.513 | 2.008 | 0.04754 | 0.60466 | -3.950 | CENPS | centromere protein S [Source:HGNC Symbol;Acc:HGNC:23163] |
| ENSG00000149313 | 0.350 | 6.086 | 2.008 | 0.04759 | 0.60466 | -4.217 | AASDHPPT | aminoadipate-semialdehyde dehydrogenase-phosphopantetheinyl transferase [Source:HGNC Symbol;Acc:HGNC:14235] |
| ENSG00000242247 | -0.383 | 7.476 | -2.008 | 0.04760 | 0.60466 | -4.204 | ARFGAP3 | ADP ribosylation factor GTPase activating protein 3 [Source:HGNC Symbol;Acc:HGNC:661] |
| ENSG00000074370 | -0.591 | 6.708 | -2.007 | 0.04764 | 0.60466 | -4.217 | ATP2A3 | ATPase sarcoplasmic/endoplasmic reticulum Ca2+ transporting 3 [Source:HGNC Symbol;Acc:HGNC:813] |
| ENSG00000100348 | -0.396 | 3.887 | -2.007 | 0.04769 | 0.60466 | -4.061 | TXN2 | thioredoxin 2 [Source:HGNC Symbol;Acc:HGNC:17772] |
| ENSG00000115484 | 0.287 | 6.792 | 2.006 | 0.04779 | 0.60466 | -4.214 | CCT4 | chaperonin containing TCP1 subunit 4 [Source:HGNC Symbol;Acc:HGNC:1617] |
| ENSG00000166483 | 0.757 | 4.724 | 2.006 | 0.04780 | 0.60466 | -4.193 | WEE1 | WEE1 G2 checkpoint kinase [Source:HGNC Symbol;Acc:HGNC:12761] |
| ENSG00000162407 | -1.467 | 2.411 | -2.005 | 0.04786 | 0.60466 | -3.822 | PLPP3 | phospholipid phosphatase 3 [Source:HGNC Symbol;Acc:HGNC:9229] |
| ENSG00000162704 | 0.445 | 6.457 | 2.005 | 0.04792 | 0.60466 | -4.220 | ARPC5 | actin related protein 2/3 complex subunit 5 [Source:HGNC Symbol;Acc:HGNC:708] |
| ENSG00000101298 | -0.761 | 0.584 | -2.004 | 0.04795 | 0.60466 | -3.815 | SNPH | syntaphilin [Source:HGNC Symbol;Acc:HGNC:15931] |
| ENSG00000255292 | 0.320 | 3.344 | 2.004 | 0.04799 | 0.60466 | -4.052 | SDHD | succinate dehydrogenase complex subunit D [Source:HGNC Symbol;Acc:HGNC:10683] |
| ENSG00000250317 | -0.457 | 4.731 | -2.004 | 0.04801 | 0.60466 | -4.139 | SMIM20 | small integral membrane protein 20 [Source:HGNC Symbol;Acc:HGNC:37260] |
| ENSG00000175305 | 0.470 | 5.258 | 2.004 | 0.04803 | 0.60466 | -4.211 | CCNE2 | cyclin E2 [Source:HGNC Symbol;Acc:HGNC:1590] |
| ENSG00000142528 | 0.338 | 3.075 | 2.004 | 0.04803 | 0.60466 | -4.024 | ZNF473 | zinc finger protein 473 [Source:HGNC Symbol;Acc:HGNC:23239] |
| ENSG00000105750 | 1.149 | 0.880 | 2.004 | 0.04805 | 0.60466 | -3.842 | ZNF85 | zinc finger protein 85 [Source:HGNC Symbol;Acc:HGNC:13160] |
| ENSG00000101871 | 0.887 | 0.692 | 2.003 | 0.04815 | 0.60466 | -3.820 | MID1 | midline 1 [Source:HGNC Symbol;Acc:HGNC:7095] |
| ENSG00000136161 | -0.725 | 4.744 | -2.002 | 0.04817 | 0.60466 | -4.129 | RCBTB2 | RCC1 and BTB domain containing protein 2 [Source:HGNC Symbol;Acc:HGNC:1914] |
| ENSG00000122335 | -0.508 | 2.499 | -2.002 | 0.04818 | 0.60466 | -3.897 | SERAC1 | serine active site containing 1 [Source:HGNC Symbol;Acc:HGNC:21061] |
| ENSG00000133265 | 0.425 | 2.777 | 2.002 | 0.04820 | 0.60466 | -4.000 | HSPBP1 | HSPA (Hsp70) binding protein 1 [Source:HGNC Symbol;Acc:HGNC:24989] |
| ENSG00000139192 | -0.525 | 5.370 | -2.002 | 0.04822 | 0.60466 | -4.190 | TAPBPL | TAP binding protein like [Source:HGNC Symbol;Acc:HGNC:30683] |
| ENSG00000020577 | -1.250 | 2.169 | -2.002 | 0.04826 | 0.60466 | -3.820 | SAMD4A | sterile alpha motif domain containing 4A [Source:HGNC Symbol;Acc:HGNC:23023] |
| ENSG00000115364 | 0.306 | 5.038 | 2.001 | 0.04829 | 0.60466 | -4.204 | MRPL19 | mitochondrial ribosomal protein L19 [Source:HGNC Symbol;Acc:HGNC:14052] |
| ENSG00000213203 | -1.387 | -0.167 | -2.001 | 0.04830 | 0.60466 | -3.870 | GIMAP1 | GTPase, IMAP family member 1 [Source:HGNC Symbol;Acc:HGNC:23237] |
| ENSG00000168496 | 0.470 | 4.613 | 2.001 | 0.04830 | 0.60466 | -4.187 | FEN1 | flap structure-specific endonuclease 1 [Source:HGNC Symbol;Acc:HGNC:3650] |
| ENSG00000101057 | 0.676 | 3.791 | 2.001 | 0.04834 | 0.60466 | -4.138 | MYBL2 | MYB proto-oncogene like 2 [Source:HGNC Symbol;Acc:HGNC:7548] |
| ENSG00000135503 | -0.737 | 3.241 | -2.001 | 0.04835 | 0.60466 | -3.966 | ACVR1B | activin A receptor type 1B [Source:HGNC Symbol;Acc:HGNC:172] |
| ENSG00000174013 | 0.382 | 4.668 | 2.001 | 0.04835 | 0.60466 | -4.187 | FBXO45 | F-box protein 45 [Source:HGNC Symbol;Acc:HGNC:29148] |
| ENSG00000125875 | -0.294 | 4.799 | -2.000 | 0.04844 | 0.60466 | -4.168 | TBC1D20 | TBC1 domain family member 20 [Source:HGNC Symbol;Acc:HGNC:16133] |
| ENSG00000185420 | 0.524 | 4.117 | 2.000 | 0.04847 | 0.60466 | -4.157 | SMYD3 | SET and MYND domain containing 3 [Source:HGNC Symbol;Acc:HGNC:15513] |
| ENSG00000249915 | 0.328 | 5.423 | 2.000 | 0.04849 | 0.60466 | -4.222 | PDCD6 | programmed cell death 6 [Source:HGNC Symbol;Acc:HGNC:8765] |
| ENSG00000023839 | -0.656 | 2.004 | -1.999 | 0.04853 | 0.60466 | -3.841 | ABCC2 | ATP binding cassette subfamily C member 2 [Source:HGNC Symbol;Acc:HGNC:53] |
| ENSG00000168397 | 0.252 | 4.435 | 1.998 | 0.04866 | 0.60493 | -4.171 | ATG4B | autophagy related 4B cysteine peptidase [Source:HGNC Symbol;Acc:HGNC:20790] |
| ENSG00000132471 | -0.423 | 4.535 | -1.998 | 0.04868 | 0.60493 | -4.144 | WBP2 | WW domain binding protein 2 [Source:HGNC Symbol;Acc:HGNC:12738] |
| ENSG00000120708 | -1.561 | 3.180 | -1.998 | 0.04869 | 0.60493 | -3.903 | TGFBI | transforming growth factor beta induced [Source:HGNC Symbol;Acc:HGNC:11771] |
| ENSG00000213397 | 0.394 | 3.377 | 1.997 | 0.04877 | 0.60532 | -4.077 | HAUS7 | HAUS augmin like complex subunit 7 [Source:HGNC Symbol;Acc:HGNC:32979] |
| ENSG00000107758 | -0.235 | 5.213 | -1.997 | 0.04882 | 0.60534 | -4.203 | PPP3CB | protein phosphatase 3 catalytic subunit beta [Source:HGNC Symbol;Acc:HGNC:9315] |
| ENSG00000123689 | 1.221 | 5.003 | 1.993 | 0.04918 | 0.60865 | -4.234 | G0S2 | G0/G1 switch 2 [Source:HGNC Symbol;Acc:HGNC:30229] |
| ENSG00000100307 | -0.651 | 5.006 | -1.993 | 0.04921 | 0.60865 | -4.173 | CBX7 | chromobox 7 [Source:HGNC Symbol;Acc:HGNC:1557] |
| ENSG00000105173 | 0.596 | 3.262 | 1.993 | 0.04923 | 0.60865 | -4.085 | CCNE1 | cyclin E1 [Source:HGNC Symbol;Acc:HGNC:1589] |
| ENSG00000159259 | 0.531 | 1.780 | 1.992 | 0.04929 | 0.60878 | -3.906 | CHAF1B | chromatin assembly factor 1 subunit B [Source:HGNC Symbol;Acc:HGNC:1911] |
| ENSG00000135951 | -0.464 | 3.898 | -1.992 | 0.04938 | 0.60928 | -4.077 | TSGA10 | testis specific 10 [Source:HGNC Symbol;Acc:HGNC:14927] |
| ENSG00000169629 | -0.844 | 2.359 | -1.991 | 0.04946 | 0.60928 | -3.870 | RGPD8 | RANBP2-like and GRIP domain containing 8 [Source:HGNC Symbol;Acc:HGNC:9849] |
| ENSG00000172500 | 0.365 | 4.213 | 1.991 | 0.04948 | 0.60928 | -4.173 | FIBP | FGF1 intracellular binding protein [Source:HGNC Symbol;Acc:HGNC:3705] |
| ENSG00000261739 | 1.682 | 3.132 | 1.990 | 0.04954 | 0.60928 | -4.141 | GOLGA8S | golgin A8 family member S [Source:HGNC Symbol;Acc:HGNC:44409] |
| ENSG00000103018 | 0.342 | 4.590 | 1.990 | 0.04959 | 0.60928 | -4.202 | CYB5B | cytochrome b5 type B [Source:HGNC Symbol;Acc:HGNC:24374] |
| ENSG00000182534 | -0.930 | 2.326 | -1.989 | 0.04961 | 0.60928 | -3.874 | MXRA7 | matrix remodeling associated 7 [Source:HGNC Symbol;Acc:HGNC:7541] |
| ENSG00000101577 | -0.654 | 3.986 | -1.989 | 0.04970 | 0.60978 | -4.090 | LPIN2 | lipin 2 [Source:HGNC Symbol;Acc:HGNC:14450] |
| ENSG00000111666 | -1.122 | 3.974 | -1.987 | 0.04984 | 0.61047 | -4.042 | CHPT1 | choline phosphotransferase 1 [Source:HGNC Symbol;Acc:HGNC:17852] |
| ENSG00000112245 | -0.400 | 7.716 | -1.987 | 0.04985 | 0.61047 | -4.235 | PTP4A1 | protein tyrosine phosphatase type IVA, member 1 [Source:HGNC Symbol;Acc:HGNC:9634] |

**Supplemental Table S6.** Gene set enrichment analysis of the C5 library from differential gene expression of the comparison between presence of *TP53* mutation to WT *TP53* from the FIMM cohort.

| ID | setSize | enrichmentScore | NES | pvalue | p.adjust | qvalue | rank |
|---|---|---|---|---|---|---|---|
| GOBP_MITOTIC_NUCLEAR_DIVISION | 77 | 0.575 | 2.602 | 0.00000 | 0.00000 | 0.00000 | 316 |
| GOBP_MITOTIC_CELL_CYCLE | 168 | 0.504 | 2.535 | 0.00000 | 0.00000 | 0.00000 | 326 |
| GOBP_ORGANELLE_FISSION | 102 | 0.532 | 2.512 | 0.00000 | 0.00000 | 0.00000 | 316 |
| GOBP_CELL_DIVISION | 110 | 0.526 | 2.511 | 0.00000 | 0.00000 | 0.00000 | 337 |
| GOBP_CHROMOSOME_SEGREGATION | 90 | 0.531 | 2.464 | 0.00000 | 0.00000 | 0.00000 | 319 |
| GOBP_MITOTIC_SISTER_CHROMATID_SEGREGATION | 58 | 0.569 | 2.457 | 0.00000 | 0.00000 | 0.00000 | 316 |
| GOBP_MITOTIC_CELL_CYCLE_PROCESS | 159 | 0.491 | 2.456 | 0.00000 | 0.00000 | 0.00000 | 326 |
| GOBP_SISTER_CHROMATID_SEGREGATION | 66 | 0.552 | 2.435 | 0.00000 | 0.00000 | 0.00001 | 279 |
| GOBP_CELL_CYCLE_PROCESS | 198 | 0.475 | 2.426 | 0.00000 | 0.00000 | 0.00000 | 326 |
| GOBP_SPINDLE_ORGANIZATION | 39 | 0.607 | 2.416 | 0.00000 | 0.00000 | 0.00002 | 319 |
| GOBP_MITOTIC_SPINDLE_ORGANIZATION | 27 | 0.659 | 2.400 | 0.00000 | 0.00000 | 0.00006 | 216 |
| GOBP_NUCLEAR_CHROMOSOME_SEGREGATION | 77 | 0.529 | 2.393 | 0.00000 | 0.00000 | 0.00000 | 316 |
| GOBP_CELL_CYCLE | 250 | 0.458 | 2.379 | 0.00000 | 0.00000 | 0.00000 | 326 |
| GOBP_MICROTUBULE_CYTOSKELETON_ORGANIZATION_INVOLVED_IN_MITOSIS | 36 | 0.607 | 2.374 | 0.00000 | 0.00000 | 0.00008 | 216 |
| GOBP_REGULATION_OF_NUCLEAR_DIVISION | 38 | 0.598 | 2.368 | 0.00000 | 0.00000 | 0.00016 | 199 |
| GOBP_REGULATION_OF_CELL_CYCLE_PROCESS | 116 | 0.487 | 2.342 | 0.00000 | 0.00000 | 0.00000 | 316 |
| GOBP_MICROTUBULE_BASED_PROCESS | 96 | 0.498 | 2.330 | 0.00000 | 0.00000 | 0.00000 | 325 |
| GOBP_MICROTUBULE_CYTOSKELETON_ORGANIZATION | 84 | 0.507 | 2.329 | 0.00000 | 0.00000 | 0.00001 | 262 |
| GOCC_SPINDLE | 79 | 0.506 | 2.299 | 0.00000 | 0.00000 | 0.00001 | 325 |
| GOBP_NON_MEMBRANE_BOUNDED_ORGANELLE_ASSEMBLY | 52 | 0.533 | 2.251 | 0.00000 | 0.00000 | 0.00021 | 319 |
| GOBP_POSITIVE_REGULATION_OF_CELL_CYCLE_PROCESS | 44 | 0.544 | 2.224 | 0.00001 | 0.00001 | 0.00050 | 419 |
| GOBP_REGULATION_OF_MITOTIC_NUCLEAR_DIVISION | 35 | 0.572 | 2.223 | 0.00002 | 0.00002 | 0.00075 | 199 |
| GOBP_SPINDLE_ASSEMBLY | 24 | 0.629 | 2.223 | 0.00003 | 0.00003 | 0.00112 | 216 |
| GOCC_MITOTIC_SPINDLE | 36 | 0.565 | 2.209 | 0.00003 | 0.00003 | 0.00095 | 316 |
| GOBP_REGULATION_OF_CELL_DIVISION | 20 | 0.649 | 2.180 | 0.00007 | 0.00007 | 0.00196 | 222 |
| GOCC_CONDENSED_CHROMOSOME_CENTROMERIC_REGION | 45 | 0.531 | 2.178 | 0.00002 | 0.00002 | 0.00082 | 319 |
| GOBP_REGULATION_OF_CHROMOSOME_SEGREGATION | 33 | 0.568 | 2.177 | 0.00006 | 0.00006 | 0.00177 | 209 |
| GOMF_TUBULIN_BINDING | 46 | 0.525 | 2.164 | 0.00002 | 0.00002 | 0.00075 | 273 |
| GOBP_CHROMOSOME_SEPARATION | 30 | 0.577 | 2.159 | 0.00006 | 0.00006 | 0.00177 | 199 |
| GOBP_REGULATION_OF_CELL_CYCLE | 148 | 0.434 | 2.154 | 0.00000 | 0.00000 | 0.00001 | 316 |
| GOCC_POLYMERIC_CYTOSKELETAL_FIBER | 63 | 0.491 | 2.148 | 0.00001 | 0.00001 | 0.00045 | 221 |
| GOBP_CYTOSKELETON_ORGANIZATION | 123 | 0.440 | 2.136 | 0.00000 | 0.00000 | 0.00004 | 262 |
| GOBP_POSITIVE_REGULATION_OF_CELL_CYCLE | 50 | 0.508 | 2.129 | 0.00001 | 0.00001 | 0.00051 | 311 |
| GOCC_SUPRAMOLECULAR_COMPLEX | 118 | 0.441 | 2.127 | 0.00000 | 0.00000 | 0.00004 | 221 |
| GOCC_CHROMOSOMAL_REGION | 87 | 0.458 | 2.112 | 0.00000 | 0.00000 | 0.00021 | 424 |
| GOMF_MICROTUBULE_BINDING | 39 | 0.528 | 2.104 | 0.00008 | 0.00008 | 0.00205 | 227 |
| GOBP_FEMALE_GAMETE_GENERATION | 16 | 0.663 | 2.097 | 0.00032 | 0.00032 | 0.00653 | 176 |
| GOCC_CONDENSED_CHROMOSOME | 59 | 0.482 | 2.085 | 0.00006 | 0.00006 | 0.00171 | 424 |
| GOBP_MITOTIC_SPINDLE_ASSEMBLY | 14 | 0.685 | 2.081 | 0.00030 | 0.00030 | 0.00614 | 216 |

| | | | | | | | |
|---|---|---|---|---|---|---|---|
| GOCC_CHROMOSOME_CENTROMERIC_REGION | 63 | 0.474 | 2.074 | 0.00004 | 0.00004 | 0.00117 | 424 |
| GOBP_CELL_CYCLE_PHASE_TRANSITION | 100 | 0.439 | 2.068 | 0.00002 | 0.00002 | 0.00065 | 423 |
| GOCC_MICROTUBULE | 55 | 0.483 | 2.065 | 0.00008 | 0.00008 | 0.00200 | 325 |
| GOBP_MEIOTIC_CELL_CYCLE_PROCESS | 41 | 0.513 | 2.063 | 0.00012 | 0.00012 | 0.00305 | 271 |
| GOBP_MITOTIC_CELL_CYCLE_PHASE_TRANSITION | 90 | 0.444 | 2.060 | 0.00002 | 0.00002 | 0.00071 | 409 |
| GOBP_REGULATION_OF_CHROMOSOME_SEPARATION | 27 | 0.565 | 2.057 | 0.00030 | 0.00030 | 0.00614 | 199 |
| GOBP_MEIOTIC_CELL_CYCLE | 50 | 0.491 | 2.057 | 0.00005 | 0.00005 | 0.00146 | 271 |
| GOCC_MICROTUBULE_CYTOSKELETON | 152 | 0.411 | 2.045 | 0.00000 | 0.00000 | 0.00005 | 340 |
| GOBP_REGULATION_OF_MITOTIC_CELL_CYCLE | 85 | 0.443 | 2.038 | 0.00002 | 0.00002 | 0.00075 | 425 |
| GOCC_SUPRAMOLECULAR_POLYMER | 73 | 0.451 | 2.021 | 0.00007 | 0.00007 | 0.00183 | 221 |
| GOBP_DNA_UNWINDING_INVOLVED_IN_DNA_REPLICATION | 12 | 0.695 | 2.015 | 0.00057 | 0.00057 | 0.00996 | 326 |
| GOCC_AXON | 25 | 0.563 | 2.015 | 0.00125 | 0.00125 | 0.01799 | 88 |
| GOBP_DNA_CONFORMATION_CHANGE | 62 | 0.461 | 2.012 | 0.00019 | 0.00019 | 0.00437 | 424 |
| GOBP_CYTOKINESIS | 27 | 0.551 | 2.006 | 0.00072 | 0.00072 | 0.01159 | 216 |
| GOBP_NEGATIVE_REGULATION_OF_CHROMOSOME_ORGANIZATION | 26 | 0.554 | 2.003 | 0.00105 | 0.00105 | 0.01574 | 199 |
| GOCC_MICROTUBULE_ASSOCIATED_COMPLEX | 21 | 0.586 | 1.995 | 0.00051 | 0.00051 | 0.00936 | 208 |
| GOBP_DNA_METABOLIC_PROCESS | 128 | 0.409 | 1.993 | 0.00001 | 0.00001 | 0.00045 | 423 |
| GOBP_METAPHASE_ANAPHASE_TRANSITION_OF_CELL_CYCLE | 26 | 0.551 | 1.992 | 0.00122 | 0.00122 | 0.01760 | 199 |
| GOBP_CHROMOSOME_ORGANIZATION | 169 | 0.396 | 1.991 | 0.00000 | 0.00000 | 0.00016 | 359 |
| GOBP_REGULATION_OF_CELL_CYCLE_PHASE_TRANSITION | 78 | 0.439 | 1.991 | 0.00010 | 0.00010 | 0.00255 | 423 |
| GOBP_ORGANELLE_ASSEMBLY | 75 | 0.440 | 1.983 | 0.00018 | 0.00018 | 0.00404 | 257 |
| GOCC_CHROMOSOME | 205 | 0.387 | 1.981 | 0.00000 | 0.00000 | 0.00003 | 424 |
| GOBP_REGULATION_OF_CYTOKINESIS | 16 | 0.626 | 1.979 | 0.00183 | 0.00183 | 0.02384 | 332 |
| GOBP_ATTACHMENT_OF_SPINDLE_MICROTUBULES_TO_KINETOCHORE | 14 | 0.649 | 1.973 | 0.00141 | 0.00141 | 0.01950 | 253 |
| GOBP_CYTOSKELETON_DEPENDENT_CYTOKINESIS | 18 | 0.602 | 1.971 | 0.00054 | 0.00054 | 0.00962 | 216 |
| GOBP_MITOTIC_CYTOKINESIS | 18 | 0.602 | 1.971 | 0.00054 | 0.00054 | 0.00962 | 216 |
| GOCC_SYNAPTIC_MEMBRANE | 16 | 0.621 | 1.965 | 0.00212 | 0.00212 | 0.02571 | 88 |
| GOBP_CELL_CYCLE_G2_M_PHASE_TRANSITION | 43 | 0.483 | 1.963 | 0.00096 | 0.00096 | 0.01483 | 311 |
| GOBP_REGULATION_OF_MITOTIC_CELL_CYCLE_PHASE_TRANSITION | 68 | 0.442 | 1.962 | 0.00012 | 0.00012 | 0.00297 | 419 |
| GOMF_CYTOSKELETAL_PROTEIN_BINDING | 67 | 0.443 | 1.962 | 0.00013 | 0.00013 | 0.00315 | 273 |
| GOBP_DNA_REPLICATION | 69 | 0.441 | 1.961 | 0.00026 | 0.00026 | 0.00557 | 421 |
| GOCC_MIDBODY | 37 | 0.490 | 1.929 | 0.00154 | 0.00154 | 0.02113 | 216 |
| GOBP_DNA_DEPENDENT_DNA_REPLICATION | 49 | 0.461 | 1.923 | 0.00051 | 0.00051 | 0.00936 | 454 |
| GOMF_MICROTUBULE_MOTOR_ACTIVITY | 14 | 0.632 | 1.922 | 0.00278 | 0.00278 | 0.03051 | 206 |
| GOCC_NEURON_PROJECTION | 54 | 0.450 | 1.915 | 0.00098 | 0.00098 | 0.01510 | 178 |
| GOBP_METAPHASE_PLATE_CONGRESSION | 20 | 0.569 | 1.912 | 0.00194 | 0.00194 | 0.02438 | 337 |
| GOBP_REGULATION_OF_MITOTIC_SISTER_CHROMATID_SEGREGATION | 20 | 0.569 | 1.911 | 0.00196 | 0.00196 | 0.02450 | 188 |
| GOBP_POSITIVE_REGULATION_OF_CELL_CYCLE_PHASE_TRANSITION | 25 | 0.532 | 1.903 | 0.00438 | 0.00438 | 0.04138 | 419 |
| GOBP_REGULATION_OF_CHROMOSOME_ORGANIZATION | 50 | 0.454 | 1.901 | 0.00063 | 0.00063 | 0.01057 | 209 |
| HP_ABNORMAL_VAGINA_MORPHOLOGY | 11 | 0.664 | 1.880 | 0.00296 | 0.00296 | 0.03156 | 219 |
| GOCC_POSTSYNAPTIC_MEMBRANE | 13 | 0.632 | 1.877 | 0.00305 | 0.00305 | 0.03193 | 88 |

| | | | | | | | |
|---|---|---|---|---|---|---|---|
| GOBP_SISTER_CHROMATID_COHESION | 17 | 0.583 | 1.871 | 0.00280 | 0.00280 | 0.03051 | 273 |
| GOBP_NUCLEOSOME_ASSEMBLY | 15 | 0.603 | 1.869 | 0.00342 | 0.00342 | 0.03538 | 423 |
| GOBP_DNA_PACKAGING | 35 | 0.480 | 1.864 | 0.00193 | 0.00193 | 0.02438 | 439 |
| GOBP_POSITIVE_REGULATION_OF_MITOTIC_CELL_CYCLE_PHASE_TRANSITION | 22 | 0.536 | 1.852 | 0.00350 | 0.00350 | 0.03565 | 311 |
| GOCC_NUCLEAR_CHROMOSOME | 42 | 0.457 | 1.848 | 0.00129 | 0.00129 | 0.01838 | 423 |
| HP_SLOPING_FOREHEAD | 29 | 0.495 | 1.837 | 0.00261 | 0.00261 | 0.02955 | 215 |
| GOBP_MICROTUBULE_ORGANIZING_CENTER_ORGANIZATION | 31 | 0.486 | 1.837 | 0.00245 | 0.00245 | 0.02851 | 257 |
| GOBP_CHROMOSOME_LOCALIZATION | 23 | 0.525 | 1.835 | 0.00416 | 0.00416 | 0.03989 | 337 |
| GOBP_REGULATION_OF_ORGANELLE_ORGANIZATION | 119 | 0.378 | 1.825 | 0.00032 | 0.00032 | 0.00653 | 268 |
| GOBP_REGULATION_OF_MEIOTIC_CELL_CYCLE | 10 | 0.666 | 1.824 | 0.00508 | 0.00508 | 0.04660 | 264 |
| GOBP_NEGATIVE_REGULATION_OF_METAPHASE_ANAPHASE_TRANSITION_OF_CELL_ | 19 | 0.549 | 1.824 | 0.00372 | 0.00372 | 0.03629 | 188 |
| GOCC_SPINDLE_POLE | 37 | 0.457 | 1.799 | 0.00378 | 0.00378 | 0.03656 | 337 |
| GOCC_KINESIN_COMPLEX | 11 | 0.634 | 1.795 | 0.00636 | 0.00636 | 0.05506 | 197 |
| GOBP_DNA_REPAIR | 87 | 0.389 | 1.794 | 0.00095 | 0.00095 | 0.01483 | 423 |
| GOBP_REPRODUCTION | 108 | 0.377 | 1.794 | 0.00103 | 0.00103 | 0.01564 | 272 |
| GOBP_DNA_GEOMETRIC_CHANGE | 29 | 0.483 | 1.792 | 0.00417 | 0.00417 | 0.03989 | 347 |
| GOBP_REGULATION_OF_MICROTUBULE_BASED_PROCESS | 27 | 0.490 | 1.785 | 0.00527 | 0.00527 | 0.04746 | 408 |
| GOBP_NEGATIVE_REGULATION_OF_ORGANELLE_ORGANIZATION | 44 | 0.435 | 1.779 | 0.00369 | 0.00369 | 0.03629 | 257 |
| GOCC_SPINDLE_MIDZONE | 12 | 0.613 | 1.775 | 0.00735 | 0.00735 | 0.06043 | 316 |
| GOBP_NEGATIVE_REGULATION_OF_NUCLEAR_DIVISION | 22 | 0.514 | 1.775 | 0.00686 | 0.00686 | 0.05811 | 188 |
| GOCC_CENTROSOME | 83 | 0.387 | 1.774 | 0.00165 | 0.00165 | 0.02217 | 268 |
| GOCC_CENTRIOLAR_SATELLITE | 13 | 0.597 | 1.774 | 0.00799 | 0.00799 | 0.06282 | 261 |
| GOMF_CYTOSKELETAL_MOTOR_ACTIVITY | 16 | 0.561 | 1.774 | 0.00803 | 0.00803 | 0.06288 | 206 |
| GOCC_INTRINSIC_COMPONENT_OF_PLASMA_MEMBRANE | 46 | 0.430 | 1.773 | 0.00367 | 0.00367 | 0.03629 | 88 |
| GOBP_REGULATION_OF_CYCLIN_DEPENDENT_PROTEIN_KINASE_ACTIVITY | 21 | 0.520 | 1.771 | 0.00749 | 0.00749 | 0.06109 | 292 |
| GOCC_PROTEIN_DNA_COMPLEX | 27 | 0.484 | 1.764 | 0.00685 | 0.00685 | 0.05811 | 365 |
| GOBP_NEGATIVE_REGULATION_OF_CELL_CYCLE_PROCESS | 58 | 0.408 | 1.759 | 0.00297 | 0.00297 | 0.03156 | 423 |
| GOCC_MICROTUBULE_ORGANIZING_CENTER | 100 | 0.373 | 1.757 | 0.00279 | 0.00279 | 0.03051 | 425 |
| GOBP_ESTABLISHMENT_OF_ORGANELLE_LOCALIZATION | 47 | 0.423 | 1.753 | 0.00428 | 0.00428 | 0.04067 | 227 |
| GOBP_MITOTIC_METAPHASE_PLATE_CONGRESSION | 17 | 0.544 | 1.749 | 0.00951 | 0.00951 | 0.06994 | 337 |
| GOBP_REGULATION_OF_REPRODUCTIVE_PROCESS | 18 | 0.534 | 1.749 | 0.00957 | 0.00957 | 0.06994 | 264 |
| GOBP_REGULATION_OF_UBIQUITIN_PROTEIN_TRANSFERASE_ACTIVITY | 11 | 0.616 | 1.745 | 0.01065 | 0.01065 | 0.07401 | 175 |
| HP_NARROW_PALATE | 12 | 0.601 | 1.741 | 0.01029 | 0.01029 | 0.07339 | 332 |
| GOBP_PROTEIN_DEPOLYMERIZATION | 11 | 0.611 | 1.730 | 0.01243 | 0.01243 | 0.08263 | 208 |
| GOBP_DNA_REPLICATION_INITIATION | 16 | 0.547 | 1.729 | 0.01114 | 0.01114 | 0.07635 | 367 |
| GOBP_MICROTUBULE_BASED_MOVEMENT | 28 | 0.468 | 1.723 | 0.00888 | 0.00888 | 0.06688 | 273 |
| HP_SIMPLIFIED_GYRAL_PATTERN | 11 | 0.607 | 1.718 | 0.01346 | 0.01346 | 0.08801 | 382 |
| GOBP_ORGANELLE_LOCALIZATION | 55 | 0.402 | 1.717 | 0.00539 | 0.00539 | 0.04787 | 227 |
| HP_AGENESIS_OF_CORPUS_CALLOSUM | 31 | 0.454 | 1.715 | 0.00966 | 0.00966 | 0.07027 | 215 |
| GOBP_MEIOSIS_I_CELL_CYCLE_PROCESS | 28 | 0.464 | 1.709 | 0.01045 | 0.01045 | 0.07393 | 271 |
| GOCC_SPINDLE_MICROTUBULE | 20 | 0.508 | 1.708 | 0.01209 | 0.01209 | 0.08110 | 316 |

| | | | | | | | |
|---|---|---|---|---|---|---|---|
| GOBP_MEIOTIC_CHROMOSOME_SEGREGATION | 24 | 0.482 | 1.706 | 0.01199 | 0.01199 | 0.08076 | 199 |
| GOBP_DNA_RECOMBINATION | 49 | 0.407 | 1.697 | 0.00754 | 0.00754 | 0.06117 | 529 |
| GOBP_CHROMOSOME_CONDENSATION | 11 | 0.598 | 1.695 | 0.01686 | 0.01686 | 0.10416 | 279 |
| GOMF_CATION_CHANNEL_ACTIVITY | 12 | 0.583 | 1.688 | 0.01837 | 0.01837 | 0.10918 | 73 |
| GOBP_NEGATIVE_REGULATION_OF_CELLULAR_COMPONENT_ORGANIZATION | 61 | 0.387 | 1.686 | 0.00662 | 0.00662 | 0.05668 | 235 |
| GOBP_REGULATION_OF_DNA_DEPENDENT_DNA_REPLICATION | 17 | 0.525 | 1.686 | 0.01594 | 0.01594 | 0.09998 | 421 |
| GOBP_EMBRYONIC_ORGAN_DEVELOPMENT | 14 | 0.554 | 1.684 | 0.01961 | 0.01961 | 0.11364 | 173 |
| GOBP_GAMETE_GENERATION | 50 | 0.402 | 1.682 | 0.00788 | 0.00788 | 0.06263 | 272 |
| HP_HIGH_NARROW_PALATE | 11 | 0.594 | 1.681 | 0.01849 | 0.01849 | 0.10935 | 332 |
| GOBP_BEHAVIOR | 25 | 0.469 | 1.680 | 0.01521 | 0.01521 | 0.09657 | 96 |
| GOBP_EMBRYO_DEVELOPMENT | 65 | 0.381 | 1.678 | 0.00797 | 0.00797 | 0.06282 | 261 |
| GOBP_POSITIVE_REGULATION_OF_MITOTIC_CELL_CYCLE | 23 | 0.479 | 1.677 | 0.01529 | 0.01529 | 0.09667 | 311 |
| GOBP_CENTROMERE_COMPLEX_ASSEMBLY | 16 | 0.530 | 1.675 | 0.01790 | 0.01790 | 0.10850 | 321 |
| GOBP_CELL_CYCLE_CHECKPOINT_SIGNALING | 45 | 0.408 | 1.672 | 0.00937 | 0.00937 | 0.06977 | 423 |
| GOBP_REGULATION_OF_MICROTUBULE_CYTOSKELETON_ORGANIZATION | 14 | 0.549 | 1.668 | 0.02175 | 0.02175 | 0.12134 | 325 |
| GOCC_TRANSPORT_VESICLE_MEMBRANE | 12 | 0.574 | 1.664 | 0.02233 | 0.02233 | 0.12320 | 38 |
| GOBP_PROTEIN_DNA_COMPLEX_ASSEMBLY | 37 | 0.423 | 1.663 | 0.01435 | 0.01435 | 0.09196 | 321 |
| GOBP_REGULATION_OF_DNA_REPLICATION | 29 | 0.448 | 1.662 | 0.01421 | 0.01421 | 0.09171 | 347 |
| HP_DISINHIBITION | 14 | 0.546 | 1.660 | 0.02333 | 0.02333 | 0.12728 | 161 |
| HP_ABNORMAL_URETER_PHYSIOLOGY | 20 | 0.494 | 1.659 | 0.01814 | 0.01814 | 0.10850 | 232 |
| GOBP_REGULATION_OF_DNA_RECOMBINATION | 16 | 0.521 | 1.649 | 0.02199 | 0.02199 | 0.12213 | 235 |
| GOCC_POSTSYNAPSE | 27 | 0.452 | 1.648 | 0.01712 | 0.01712 | 0.10529 | 138 |
| GOBP_NEURAL_PRECURSOR_CELL_PROLIFERATION | 11 | 0.579 | 1.639 | 0.02515 | 0.02515 | 0.13428 | 216 |
| GOBP_PROTEIN_LOCALIZATION_TO_CYTOSKELETON | 10 | 0.597 | 1.636 | 0.02511 | 0.02511 | 0.13428 | 375 |
| GOBP_KINETOCHORE_ORGANIZATION | 14 | 0.538 | 1.636 | 0.02834 | 0.02834 | 0.14350 | 108 |
| GOCC_DNA_PACKAGING_COMPLEX | 15 | 0.526 | 1.629 | 0.02811 | 0.02811 | 0.14293 | 463 |
| GOBP_CELLULAR_COMPONENT_MORPHOGENESIS | 38 | 0.411 | 1.627 | 0.01802 | 0.01802 | 0.10850 | 178 |
| GOBP_MITOTIC_CELL_CYCLE_CHECKPOINT_SIGNALING | 38 | 0.410 | 1.623 | 0.01839 | 0.01839 | 0.10918 | 418 |
| GOBP_CELLULAR_PROCESS_INVOLVED_IN_REPRODUCTION_IN_MULTICELLULAR_C | 28 | 0.438 | 1.614 | 0.02155 | 0.02155 | 0.12099 | 234 |
| GOCC_CELL_DIVISION_SITE | 11 | 0.570 | 1.613 | 0.03136 | 0.03136 | 0.15205 | 216 |
| HP_PHENOTYPIC_VARIABILITY | 22 | 0.467 | 1.613 | 0.02543 | 0.02543 | 0.13496 | 215 |
| GOBP_CHROMATIN_ASSEMBLY_OR_DISASSEMBLY | 26 | 0.446 | 1.611 | 0.02522 | 0.02522 | 0.13428 | 423 |
| GOBP_NUCLEOSOME_ORGANIZATION | 19 | 0.485 | 1.611 | 0.02813 | 0.02813 | 0.14293 | 423 |
| GOBP_DNA_MODIFICATION | 11 | 0.566 | 1.602 | 0.03402 | 0.03402 | 0.16221 | 409 |
| GOBP_SPINDLE_LOCALIZATION | 12 | 0.552 | 1.601 | 0.03438 | 0.03438 | 0.16282 | 199 |
| GOMF_METAL_ION_TRANSMEMBRANE_TRANSPORTER_ACTIVITY | 11 | 0.565 | 1.599 | 0.03447 | 0.03447 | 0.16282 | 73 |
| HP_ABNORMAL_CORTICAL_BONE_MORPHOLOGY | 10 | 0.582 | 1.595 | 0.03677 | 0.03677 | 0.16782 | 181 |
| HP_RENAL_HYPOPLASIA_APLASIA | 31 | 0.422 | 1.594 | 0.02460 | 0.02460 | 0.13282 | 257 |
| HP_ABNORMAL_MALE_REPRODUCTIVE_SYSTEM_PHYSIOLOGY | 23 | 0.456 | 1.594 | 0.02695 | 0.02695 | 0.13966 | 259 |
| HP_CRANIOSYNOSTOSIS | 13 | 0.536 | 1.594 | 0.03660 | 0.03660 | 0.16782 | 316 |
| HP_GASTROINTESTINAL_ATRESIA | 10 | 0.580 | 1.589 | 0.03827 | 0.03827 | 0.17038 | 206 |

| | | | | | | | |
|---|---|---|---|---|---|---|---|
| HP_CEREBRAL_HYPOPLASIA | 11 | 0.561 | 1.589 | 0.03669 | 0.03669 | 0.16782 | 181 |
| HP_ABNORMALITY_OF_CHROMOSOME_STABILITY | 11 | 0.561 | 1.589 | 0.03698 | 0.03698 | 0.16782 | 359 |
| GOBP_CELLULAR_RESPONSE_TO_DNA_DAMAGE_STIMULUS | 115 | 0.330 | 1.586 | 0.00836 | 0.00836 | 0.06480 | 423 |
| GOBP_PROTEIN_LOCALIZATION_TO_CHROMOSOME_CENTROMERIC_REGION | 10 | 0.577 | 1.580 | 0.04006 | 0.04006 | 0.17436 | 146 |
| GOMF_PASSIVE_TRANSMEMBRANE_TRANSPORTER_ACTIVITY | 14 | 0.519 | 1.577 | 0.04165 | 0.04165 | 0.17876 | 73 |
| HP_ABNORMALITY_OF_THE_URETER | 35 | 0.404 | 1.570 | 0.02688 | 0.02688 | 0.13966 | 257 |
| HP_HEMATOLOGICAL_NEOPLASM | 28 | 0.426 | 1.569 | 0.03056 | 0.03056 | 0.15034 | 312 |
| GOBP_NEGATIVE_REGULATION_OF_CELL_CYCLE | 66 | 0.354 | 1.561 | 0.02177 | 0.02177 | 0.12134 | 423 |
| GOBP_NEGATIVE_REGULATION_OF_CELL_CYCLE_PHASE_TRANSITION | 50 | 0.372 | 1.560 | 0.02425 | 0.02425 | 0.13139 | 423 |
| GOBP_NEGATIVE_REGULATION_OF_MITOTIC_CELL_CYCLE_PHASE_TRANSITION | 40 | 0.389 | 1.559 | 0.02889 | 0.02889 | 0.14486 | 306 |
| GOBP_MICROTUBULE_DEPOLYMERIZATION | 10 | 0.568 | 1.558 | 0.04544 | 0.04544 | 0.19290 | 402 |
| GOBP_RECOMBINATIONAL_REPAIR | 36 | 0.398 | 1.556 | 0.03030 | 0.03030 | 0.14960 | 521 |
| HP_ABNORMALITY_OF_EPIPHYSIS_MORPHOLOGY | 24 | 0.440 | 1.555 | 0.03703 | 0.03703 | 0.16782 | 323 |
| GOMF_GLYCOSYLTRANSFERASE_ACTIVITY | 10 | 0.567 | 1.555 | 0.04619 | 0.04619 | 0.19448 | 70 |
| GOBP_POSITIVE_REGULATION_OF_CELL_CYCLE_G1_S_PHASE_TRANSITION | 10 | 0.565 | 1.549 | 0.04798 | 0.04798 | 0.19826 | 409 |
| GOBP_REGULATION_OF_CELL_CYCLE_G2_M_PHASE_TRANSITION | 31 | 0.409 | 1.544 | 0.03632 | 0.03632 | 0.16782 | 311 |
| HP_RIGIDITY | 14 | 0.507 | 1.540 | 0.05367 | 0.05367 | 0.21134 | 259 |
| GOBP_CELL_CYCLE_DNA_REPLICATION | 18 | 0.470 | 1.538 | 0.04576 | 0.04576 | 0.19321 | 514 |
| GOBP_POSITIVE_REGULATION_OF_DNA_BIOSYNTHETIC_PROCESS | 15 | 0.496 | 1.537 | 0.05308 | 0.05308 | 0.21036 | 347 |
| HP_ABNORMAL_DUODENUM_MORPHOLOGY | 17 | 0.477 | 1.532 | 0.04991 | 0.04991 | 0.20351 | 257 |
| GOMF_CATION_TRANSMEMBRANE_TRANSPORTER_ACTIVITY | 17 | 0.477 | 1.531 | 0.05061 | 0.05061 | 0.20474 | 73 |
| GOMF_CATALYTIC_ACTIVITY_ACTING_ON_DNA | 39 | 0.384 | 1.531 | 0.03704 | 0.03704 | 0.16782 | 564 |
| GOBP_ACTOMYOSIN_STRUCTURE_ORGANIZATION | 11 | 0.540 | 1.530 | 0.05725 | 0.05725 | 0.21897 | 216 |
| GOBP_REGULATION_OF_DEVELOPMENTAL_GROWTH | 18 | 0.467 | 1.529 | 0.04881 | 0.04881 | 0.20034 | 107 |
| GOBP_DOUBLE_STRAND_BREAK_REPAIR | 48 | 0.366 | 1.521 | 0.03251 | 0.03251 | 0.15690 | 521 |
| GOBP_REGULATION_OF_ANATOMICAL_STRUCTURE_SIZE | 16 | 0.480 | 1.518 | 0.05751 | 0.05751 | 0.21897 | 46 |
| GOBP_SODIUM_ION_TRANSMEMBRANE_TRANSPORT | 12 | 0.524 | 1.517 | 0.05965 | 0.05965 | 0.22146 | 138 |
| GOBP_REGULATION_OF_DOUBLE_STRAND_BREAK_REPAIR_VIA_HOMOLOGOUS_RE | 12 | 0.524 | 1.517 | 0.05980 | 0.05980 | 0.22146 | 235 |
| GOMF_ATP_HYDROLYSIS_ACTIVITY | 43 | 0.373 | 1.515 | 0.03672 | 0.03672 | 0.16782 | 518 |
| HP_DECREASED_FERTILITY_IN_MALES | 10 | 0.553 | 1.515 | 0.05740 | 0.05740 | 0.21897 | 359 |
| GOBP_PROTEIN_LOCALIZATION_TO_CHROMOSOME | 26 | 0.418 | 1.512 | 0.04885 | 0.04885 | 0.20034 | 146 |
| GOCC_INTERCELLULAR_BRIDGE | 13 | 0.508 | 1.510 | 0.06128 | 0.06128 | 0.22239 | 314 |
| GOBP_SODIUM_ION_TRANSPORT | 13 | 0.508 | 1.509 | 0.06143 | 0.06143 | 0.22239 | 138 |
| GOBP_PROTEIN_DNA_COMPLEX_SUBUNIT_ORGANIZATION | 41 | 0.375 | 1.508 | 0.04029 | 0.04029 | 0.17437 | 423 |
| GOCC_SOMATODENDRITIC_COMPARTMENT | 29 | 0.406 | 1.505 | 0.04887 | 0.04887 | 0.20034 | 213 |
| HP_RENAL_AGENESIS | 14 | 0.494 | 1.503 | 0.06340 | 0.06340 | 0.22481 | 181 |
| GOMF_DNA_HELICASE_ACTIVITY | 21 | 0.439 | 1.496 | 0.05925 | 0.05925 | 0.22146 | 347 |
| GOBP_CELL_MORPHOGENESIS | 44 | 0.366 | 1.496 | 0.04388 | 0.04388 | 0.18784 | 84 |
| HP_ANAL_ATRESIA | 18 | 0.457 | 1.495 | 0.05991 | 0.05991 | 0.22146 | 257 |
| HP_ABNORMAL_LUNG_DEVELOPMENT | 13 | 0.503 | 1.493 | 0.06724 | 0.06724 | 0.23307 | 206 |
| GOBP_KINETOCHORE_ASSEMBLY | 10 | 0.545 | 1.493 | 0.06592 | 0.06592 | 0.23139 | 319 |

| | | | | | | | |
|---|---|---|---|---|---|---|---|
| HP_ABNORMALITY_OF_THE_PALPEBRAL_FISSURES | 65 | 0.338 | 1.490 | 0.03529 | 0.03529 | 0.16571 | 215 |
| GOBP_ESTABLISHMENT_OF_MITOTIC_SPINDLE_LOCALIZATION | 11 | 0.526 | 1.489 | 0.07263 | 0.07263 | 0.24099 | 199 |
| HP_ABNORMAL_URETER_MORPHOLOGY | 16 | 0.471 | 1.489 | 0.06950 | 0.06950 | 0.23614 | 257 |
| HP_DECREASED_FERTILITY | 20 | 0.443 | 1.488 | 0.06265 | 0.06265 | 0.22381 | 257 |
| HP_SLANTING_OF_THE_PALPEBRAL_FISSURE | 58 | 0.345 | 1.487 | 0.03937 | 0.03937 | 0.17261 | 215 |
| HP_TRIANGULAR_FACE | 14 | 0.489 | 1.486 | 0.06856 | 0.06856 | 0.23432 | 200 |
| GOCC_CLEAVAGE_FURROW | 10 | 0.542 | 1.485 | 0.06981 | 0.06981 | 0.23668 | 216 |
| GOCC_REPLICATION_FORK | 23 | 0.424 | 1.483 | 0.06020 | 0.06020 | 0.22146 | 444 |
| GOBP_REGULATION_OF_DNA_DIRECTED_DNA_POLYMERASE_ACTIVITY | 10 | 0.540 | 1.480 | 0.07280 | 0.07280 | 0.24101 | 347 |
| HP_THIN_UPPER_LIP_VERMILION | 22 | 0.428 | 1.479 | 0.06244 | 0.06244 | 0.22381 | 208 |
| HP_ABNORMALITY_OF_THE_CLITORIS | 10 | 0.539 | 1.477 | 0.07369 | 0.07369 | 0.24243 | 306 |
| HP_RENAL_HYPOPLASIA | 11 | 0.522 | 1.477 | 0.07766 | 0.07766 | 0.24958 | 232 |
| HP_PROPTOSIS | 24 | 0.417 | 1.476 | 0.06100 | 0.06100 | 0.22239 | 365 |
| HP_BULBOUS_NOSE | 11 | 0.521 | 1.475 | 0.07796 | 0.07796 | 0.24958 | 181 |
| GOBP_CELL_MORPHOGENESIS_INVOLVED_IN_DIFFERENTIATION | 35 | 0.380 | 1.475 | 0.05401 | 0.05401 | 0.21134 | 84 |
| HP_DIARRHEA | 21 | 0.432 | 1.473 | 0.06660 | 0.06660 | 0.23139 | 259 |
| HP_EXTERNAL_EAR_MALFORMATION | 10 | 0.537 | 1.473 | 0.07549 | 0.07549 | 0.24624 | 375 |
| GOBP_MITOTIC_G2_M_TRANSITION_CHECKPOINT | 16 | 0.465 | 1.472 | 0.07584 | 0.07584 | 0.24687 | 306 |
| GOBP_PROTEIN_CONTAINING_COMPLEX_DISASSEMBLY | 18 | 0.449 | 1.468 | 0.07225 | 0.07225 | 0.24099 | 208 |
| GOBP_NEGATIVE_REGULATION_OF_MITOTIC_CELL_CYCLE | 45 | 0.358 | 1.466 | 0.05460 | 0.05460 | 0.21269 | 418 |
| GOBP_SYNAPTIC_SIGNALING | 27 | 0.403 | 1.466 | 0.06204 | 0.06204 | 0.22355 | 158 |
| GOBP_AXON_DEVELOPMENT | 20 | 0.436 | 1.465 | 0.07104 | 0.07104 | 0.23773 | 46 |
| GOBP_REGULATION_OF_ION_TRANSMEMBRANE_TRANSPORT | 22 | 0.424 | 1.463 | 0.06904 | 0.06904 | 0.23509 | 73 |
| HP_ABNORMALITY_OF_THE_ANKLES | 11 | 0.516 | 1.461 | 0.08417 | 0.08417 | 0.26084 | 126 |
| HP_MICROPENIS | 19 | 0.440 | 1.461 | 0.07502 | 0.07502 | 0.24577 | 206 |
| HP_ABNORMALITY_OF_THE_PERIPHERAL_NERVOUS_SYSTEM | 29 | 0.394 | 1.461 | 0.06269 | 0.06269 | 0.22381 | 268 |
| GOBP_DEVELOPMENTAL_PROCESS_INVOLVED_IN_REPRODUCTION | 55 | 0.342 | 1.461 | 0.05045 | 0.05045 | 0.20474 | 272 |
| GOMF_SINGLE_STRANDED_DNA_HELICASE_ACTIVITY | 14 | 0.480 | 1.460 | 0.07986 | 0.07986 | 0.25410 | 502 |
| GOBP_MULTICELLULAR_ORGANISM_REPRODUCTION | 55 | 0.341 | 1.459 | 0.05189 | 0.05189 | 0.20793 | 272 |
| GOBP_ANIMAL_ORGAN_MORPHOGENESIS | 37 | 0.370 | 1.457 | 0.06141 | 0.06141 | 0.22239 | 83 |
| GOBP_DEVELOPMENTAL_CELL_GROWTH | 10 | 0.531 | 1.456 | 0.08132 | 0.08132 | 0.25677 | 46 |
| GOBP_DNA_INTEGRITY_CHECKPOINT_SIGNALING | 27 | 0.399 | 1.455 | 0.06625 | 0.06625 | 0.23139 | 367 |
| HP_ABNORMALITY_OF_CRANIAL_SUTURES | 17 | 0.453 | 1.455 | 0.08025 | 0.08025 | 0.25479 | 316 |
| GOMF_PROTEIN_SERINE_KINASE_ACTIVITY | 32 | 0.382 | 1.454 | 0.06474 | 0.06474 | 0.22849 | 188 |
| HP_ABNORMAL_FEMALE_REPRODUCTIVE_SYSTEM_PHYSIOLOGY | 15 | 0.469 | 1.454 | 0.08376 | 0.08376 | 0.26084 | 50 |
| HP_ABNORMALITY_OF_THE_FONTANELLES_OR_CRANIAL_SUTURES | 19 | 0.438 | 1.453 | 0.07820 | 0.07820 | 0.24982 | 323 |
| GOMF_ATP_DEPENDENT_ACTIVITY_ACTING_ON_DNA | 26 | 0.402 | 1.452 | 0.06866 | 0.06866 | 0.23432 | 521 |
| HP_ABNORMALITY_OF_HINDBRAIN_MORPHOLOGY | 60 | 0.334 | 1.451 | 0.05056 | 0.05056 | 0.20474 | 206 |
| HP_ABNORMALITY_OF_THE_PHALANGES_OF_THE_TOES | 12 | 0.500 | 1.448 | 0.08654 | 0.08654 | 0.26185 | 359 |
| GOBP_POSITIVE_REGULATION_OF_DNA_METABOLIC_PROCESS | 32 | 0.380 | 1.447 | 0.06781 | 0.06781 | 0.23432 | 434 |
| GOBP_REGULATION_OF_TRANSMEMBRANE_TRANSPORT | 24 | 0.409 | 1.447 | 0.07365 | 0.07365 | 0.24243 | 73 |

| | | | | | | | |
|---|---|---|---|---|---|---|---|
| GOCC_CYTOPLASMIC_REGION | 16 | 0.457 | 1.447 | 0.08669 | 0.08669 | 0.26185 | 221 |
| HP_ABNORMAL_FOREARM_BONE_MORPHOLOGY | 13 | 0.487 | 1.446 | 0.09018 | 0.09018 | 0.26769 | 376 |
| GOBP_BLASTOCYST_DEVELOPMENT | 15 | 0.467 | 1.446 | 0.08804 | 0.08804 | 0.26387 | 247 |
| GOBP_MICROTUBULE_POLYMERIZATION_OR_DEPOLYMERIZATION | 15 | 0.467 | 1.446 | 0.08804 | 0.08804 | 0.26387 | 425 |
| GOBP_POSITIVE_REGULATION_OF_INTRACELLULAR_TRANSPORT | 16 | 0.457 | 1.445 | 0.08853 | 0.08853 | 0.26429 | 253 |
| GOMF_ION_TRANSMEMBRANE_TRANSPORTER_ACTIVITY | 23 | 0.413 | 1.445 | 0.07656 | 0.07656 | 0.24766 | 73 |
| GOBP_REGULATION_OF_CELLULAR_COMPONENT_SIZE | 13 | 0.486 | 1.445 | 0.09105 | 0.09105 | 0.26925 | 181 |
| GOBP_REGULATION_OF_DNA_METABOLIC_PROCESS | 51 | 0.344 | 1.444 | 0.05793 | 0.05793 | 0.21947 | 434 |
| GOBP_CHROMOSOME_ORGANIZATION_INVOLVED_IN_MEIOTIC_CELL_CYCLE | 18 | 0.441 | 1.443 | 0.08321 | 0.08321 | 0.26002 | 591 |
| HP_ABNORMALITY_OF_THE_SMALL_INTESTINE | 19 | 0.434 | 1.442 | 0.08275 | 0.08275 | 0.26002 | 257 |
| GOMF_CYCLIN_DEPENDENT_PROTEIN_SERINE_THREONINE_KINASE_REGULATOR_ | 11 | 0.508 | 1.439 | 0.09364 | 0.09364 | 0.27349 | 250 |
| HP_ABNORMALITY_OF_THE_EXTERNAL_NOSE | 55 | 0.336 | 1.437 | 0.06028 | 0.06028 | 0.22146 | 208 |
| GOBP_MULTI_ORGANISM_REPRODUCTIVE_PROCESS | 64 | 0.327 | 1.436 | 0.05399 | 0.05399 | 0.21134 | 272 |
| GOBP_MUSCLE_CELL_DIFFERENTIATION | 17 | 0.446 | 1.432 | 0.08975 | 0.08975 | 0.26744 | 107 |
| GOBP_POSITIVE_REGULATION_OF_CELL_CYCLE_G2_M_PHASE_TRANSITION | 10 | 0.522 | 1.431 | 0.09043 | 0.09043 | 0.26792 | 311 |
| GOCC_CENTRIOLE | 21 | 0.420 | 1.431 | 0.08472 | 0.08472 | 0.26097 | 226 |
| HP_COLOR_VISION_DEFECT | 12 | 0.494 | 1.430 | 0.09580 | 0.09580 | 0.27570 | 46 |
| GOCC_TRANSPORTER_COMPLEX | 24 | 0.404 | 1.430 | 0.08258 | 0.08258 | 0.26002 | 73 |
| GOBP_CELL_PART_MORPHOGENESIS | 34 | 0.370 | 1.430 | 0.07413 | 0.07413 | 0.24334 | 71 |
| HP_ABNORMALITY_OF_THE_FEMALE_GENITALIA | 55 | 0.334 | 1.429 | 0.06303 | 0.06303 | 0.22400 | 222 |
| HP_UPPER_LIMB_UNDERGROWTH | 12 | 0.493 | 1.428 | 0.09712 | 0.09712 | 0.27612 | 359 |
| HP_ABNORMAL_CORPUS_CALLOSUM_MORPHOLOGY | 62 | 0.327 | 1.428 | 0.05962 | 0.05962 | 0.22146 | 215 |
| GOBP_NEGATIVE_REGULATION_OF_DEVELOPMENTAL_GROWTH | 10 | 0.521 | 1.427 | 0.09163 | 0.09163 | 0.27043 | 107 |
| GOBP_SYNAPSE_ORGANIZATION | 16 | 0.451 | 1.426 | 0.09600 | 0.09600 | 0.27570 | 225 |
| GOMF_ATP_DEPENDENT_ACTIVITY | 68 | 0.321 | 1.424 | 0.05823 | 0.05823 | 0.22005 | 347 |
| GOBP_SENSORY_SYSTEM_DEVELOPMENT | 17 | 0.443 | 1.424 | 0.09311 | 0.09311 | 0.27349 | 30 |
| GOBP_REGULATION_OF_MEMBRANE_POTENTIAL | 10 | 0.519 | 1.423 | 0.09372 | 0.09372 | 0.27349 | 138 |
| GOBP_NEUROGENESIS | 75 | 0.316 | 1.423 | 0.05752 | 0.05752 | 0.21897 | 216 |
| HP_ABNORMAL_BONE_STRUCTURE | 53 | 0.336 | 1.422 | 0.06807 | 0.06807 | 0.23432 | 376 |
| HP_WIDE_NOSE | 10 | 0.518 | 1.421 | 0.09522 | 0.09522 | 0.27570 | 206 |
| HP_ABNORMAL_SPERMATOGENESIS | 15 | 0.458 | 1.417 | 0.09946 | 0.09946 | 0.27867 | 257 |
| HP_MYELODYSPLASIA | 18 | 0.433 | 1.417 | 0.09638 | 0.09638 | 0.27570 | 376 |
| GOBP_NEURON_DIFFERENTIATION | 60 | 0.326 | 1.416 | 0.06654 | 0.06654 | 0.23139 | 214 |
| GOBP_MITOTIC_G2_DNA_DAMAGE_CHECKPOINT_SIGNALING | 13 | 0.476 | 1.415 | 0.10529 | 0.10529 | 0.28571 | 289 |
| GOCC_DENDRITIC_TREE | 19 | 0.426 | 1.415 | 0.09612 | 0.09612 | 0.27570 | 138 |
| HP_LEUKEMIA | 15 | 0.457 | 1.414 | 0.10060 | 0.10060 | 0.27885 | 222 |
| GOBP_VIRAL_LIFE_CYCLE | 14 | 0.465 | 1.414 | 0.10419 | 0.10419 | 0.28423 | 201 |
| GOMF_PROTEIN_SERINE_THREONINE_TYROSINE_KINASE_ACTIVITY | 35 | 0.364 | 1.413 | 0.08064 | 0.08064 | 0.25552 | 300 |
| GOCC_CULLIN_RING_UBIQUITIN_LIGASE_COMPLEX | 15 | 0.456 | 1.412 | 0.10174 | 0.10174 | 0.28001 | 290 |
| GOBP_HOMOLOGOUS_CHROMOSOME_SEGREGATION | 14 | 0.463 | 1.409 | 0.10734 | 0.10734 | 0.28826 | 359 |
| HP_ABNORMALITY_OF_THE_AUTONOMIC_NERVOUS_SYSTEM | 23 | 0.403 | 1.409 | 0.09372 | 0.09372 | 0.27349 | 268 |

| | | | | | | | |
|---|---|---|---|---|---|---|---|
| HP_UPSLANTED_PALPEBRAL_FISSURE | 27 | 0.386 | 1.407 | 0.08654 | 0.08654 | 0.26185 | 424 |
| HP_HYDROURETER | 10 | 0.512 | 1.404 | 0.10419 | 0.10419 | 0.28423 | 359 |
| HP_ABNORMAL_UTERUS_MORPHOLOGY | 21 | 0.412 | 1.402 | 0.09847 | 0.09847 | 0.27742 | 365 |
| HP_ABNORMALITY_OF_THE_NASAL_ALAE | 39 | 0.352 | 1.401 | 0.08618 | 0.08618 | 0.26185 | 208 |
| HP_DECREASED_TESTICULAR_SIZE | 11 | 0.494 | 1.398 | 0.11124 | 0.11124 | 0.29314 | 46 |
| HP_ABNORMAL_CARDIAC_ATRIUM_MORPHOLOGY | 41 | 0.348 | 1.398 | 0.08779 | 0.08779 | 0.26387 | 232 |
| HP_ABNORMALITY_OF_THE_AMNIOTIC_FLUID | 28 | 0.380 | 1.398 | 0.09339 | 0.09339 | 0.27349 | 215 |
| HP_POLYHYDRAMNIOS | 10 | 0.510 | 1.397 | 0.10822 | 0.10822 | 0.28862 | 188 |
| HP_PARAPLEGIA | 14 | 0.459 | 1.394 | 0.11464 | 0.11464 | 0.30006 | 376 |
| GOCC_CYCLIN_DEPENDENT_PROTEIN_KINASE_HOLOENZYME_COMPLEX | 12 | 0.480 | 1.390 | 0.11593 | 0.11593 | 0.30088 | 351 |
| HP_PES_PLANUS | 25 | 0.388 | 1.390 | 0.10024 | 0.10024 | 0.27884 | 215 |
| GOBP_SEX_DIFFERENTIATION | 12 | 0.480 | 1.389 | 0.11622 | 0.11622 | 0.30114 | 136 |
| GOBP_GOLGI_ORGANIZATION | 13 | 0.467 | 1.388 | 0.12126 | 0.12126 | 0.30526 | 206 |
| GOCC_HETEROCHROMATIN | 11 | 0.490 | 1.388 | 0.11538 | 0.11538 | 0.30083 | 424 |
| GOBP_MITOTIC_DNA_INTEGRITY_CHECKPOINT_SIGNALING | 20 | 0.412 | 1.383 | 0.10800 | 0.10800 | 0.28851 | 367 |
| HP_TREMOR | 35 | 0.355 | 1.381 | 0.09636 | 0.09636 | 0.27570 | 73 |
| GOBP_DNA_BIOSYNTHETIC_PROCESS | 35 | 0.355 | 1.379 | 0.09763 | 0.09763 | 0.27655 | 365 |
| HP_ENCEPHALOPATHY | 18 | 0.420 | 1.375 | 0.12051 | 0.12051 | 0.30526 | 96 |
| GOCC_CHROMOSOME_TELOMERIC_REGION | 26 | 0.380 | 1.375 | 0.10760 | 0.10760 | 0.28846 | 480 |
| GOMF_INORGANIC_MOLECULAR_ENTITY_TRANSMEMBRANE_TRANSPORTER_ACTIV | 21 | 0.402 | 1.370 | 0.11700 | 0.11700 | 0.30227 | 73 |
| HP_ABNORMAL_UVULA_MORPHOLOGY | 19 | 0.412 | 1.369 | 0.12150 | 0.12150 | 0.30526 | 359 |
| HP_NON_MOTOR_SEIZURE | 11 | 0.482 | 1.366 | 0.12781 | 0.12781 | 0.31286 | 141 |
| HP_COGNITIVE_IMPAIRMENT | 31 | 0.361 | 1.366 | 0.10948 | 0.10948 | 0.29020 | 73 |
| HP_ABNORMAL_ORAL_PHYSIOLOGY | 13 | 0.459 | 1.365 | 0.13317 | 0.13317 | 0.32090 | 129 |
| HP_APLASIA_HYPOPLASIA_OF_THE_CEREBELLUM | 32 | 0.358 | 1.363 | 0.11076 | 0.11076 | 0.29235 | 232 |
| GOCC_SYNAPSE | 51 | 0.324 | 1.361 | 0.10023 | 0.10023 | 0.27884 | 150 |
| GOMF_SINGLE_STRANDED_DNA_BINDING | 20 | 0.405 | 1.361 | 0.12201 | 0.12201 | 0.30526 | 269 |
| HP_VARIABLE_EXPRESSIVITY | 16 | 0.429 | 1.356 | 0.13561 | 0.13561 | 0.32326 | 196 |
| GOMF_ACTIN_FILAMENT_BINDING | 11 | 0.478 | 1.354 | 0.13536 | 0.13536 | 0.32326 | 194 |
| HP_ACTION_TREMOR | 12 | 0.467 | 1.353 | 0.13606 | 0.13606 | 0.32383 | 73 |
| HP_PARAPLEGIA_PARAPARESIS | 18 | 0.413 | 1.352 | 0.13382 | 0.13382 | 0.32148 | 376 |
| GOBP_CENTRAL_NERVOUS_SYSTEM_DEVELOPMENT | 51 | 0.322 | 1.351 | 0.10556 | 0.10556 | 0.28596 | 161 |
| GOCC_PERINUCLEAR_REGION_OF_CYTOPLASM | 45 | 0.330 | 1.351 | 0.10956 | 0.10956 | 0.29020 | 226 |
| GOMF_ACTIN_BINDING | 20 | 0.402 | 1.351 | 0.12874 | 0.12874 | 0.31415 | 194 |
| GOMF_STRUCTURAL_MOLECULE_ACTIVITY | 32 | 0.354 | 1.349 | 0.12088 | 0.12088 | 0.30526 | 66 |
| GOBP_LIPID_CATABOLIC_PROCESS | 20 | 0.401 | 1.348 | 0.13026 | 0.13026 | 0.31591 | 154 |
| HP_DIALEPTIC_SEIZURE | 12 | 0.465 | 1.346 | 0.14164 | 0.14164 | 0.33102 | 141 |
| GOBP_SIGNAL_TRANSDUCTION_IN_RESPONSE_TO_DNA_DAMAGE | 29 | 0.363 | 1.345 | 0.12186 | 0.12186 | 0.30526 | 423 |
| HP_ABNORMALITY_OF_THE_CHOANAE | 16 | 0.425 | 1.344 | 0.14251 | 0.14251 | 0.33256 | 375 |
| HP_SHORT_ATTENTION_SPAN | 34 | 0.348 | 1.343 | 0.12028 | 0.12028 | 0.30526 | 141 |
| HP_BICORNUATE_UTERUS | 10 | 0.490 | 1.343 | 0.14051 | 0.14051 | 0.33071 | 359 |

| | | | | | | | |
|---|---|---|---|---|---|---|---|
| GOBP_POSITIVE_REGULATION_OF_ORGANELLE_ORGANIZATION | 41 | 0.334 | 1.342 | 0.11776 | 0.11776 | 0.30360 | 279 |
| HP_ABNORMALITY_OF_THE_ANUS | 22 | 0.389 | 1.342 | 0.13524 | 0.13524 | 0.32326 | 257 |
| HP_CEREBELLAR_HYPOPLASIA | 28 | 0.364 | 1.341 | 0.12657 | 0.12657 | 0.31179 | 232 |
| GOCC_MITOCHONDRIAL_PROTEIN_CONTAINING_COMPLEX | 32 | 0.352 | 1.341 | 0.12498 | 0.12498 | 0.30986 | 663 |
| GOBP_REPRODUCTIVE_SYSTEM_DEVELOPMENT | 20 | 0.398 | 1.338 | 0.13685 | 0.13685 | 0.32522 | 226 |
| GOBP_NERVOUS_SYSTEM_PROCESS | 34 | 0.346 | 1.337 | 0.12537 | 0.12537 | 0.31030 | 96 |
| HP_BILATERAL_TONIC_CLONIC_SEIZURE | 20 | 0.398 | 1.337 | 0.13740 | 0.13740 | 0.32553 | 141 |
| GOBP_CELL_MORPHOGENESIS_INVOLVED_IN_NEURON_DIFFERENTIATION | 27 | 0.367 | 1.337 | 0.12816 | 0.12816 | 0.31322 | 71 |
| GOBP_MALE_GAMETE_GENERATION | 37 | 0.340 | 1.336 | 0.12333 | 0.12333 | 0.30773 | 272 |
| GOCC_NUCLEOLUS | 92 | 0.288 | 1.336 | 0.09557 | 0.09557 | 0.27570 | 295 |
| GOBP_CAMERA_TYPE_EYE_DEVELOPMENT | 13 | 0.449 | 1.336 | 0.15089 | 0.15089 | 0.34585 | 30 |
| HP_ABNORMAL_CORTICAL_GYRATION | 35 | 0.344 | 1.335 | 0.12578 | 0.12578 | 0.31052 | 196 |
| GOBP_PROTEASOME_MEDIATED_UBIQUITIN_DEPENDENT_PROTEIN_CATABOLIC_PI | 37 | 0.339 | 1.335 | 0.12434 | 0.12434 | 0.30906 | 176 |
| GOCC_NUCLEAR_PERIPHERY | 17 | 0.414 | 1.331 | 0.14847 | 0.14847 | 0.34233 | 222 |
| GOBP_VIRAL_PROCESS | 20 | 0.395 | 1.329 | 0.14345 | 0.14345 | 0.33364 | 201 |
| HP_APLASIA_HYPOPLASIA_OF_THE_ULNA | 11 | 0.469 | 1.328 | 0.15311 | 0.15311 | 0.34979 | 359 |
| HP_ABNORMAL_RENAL_MORPHOLOGY | 76 | 0.294 | 1.328 | 0.10692 | 0.10692 | 0.28763 | 215 |
| HP_ABNORMAL_EYELID_MORPHOLOGY | 89 | 0.287 | 1.328 | 0.09892 | 0.09892 | 0.27816 | 232 |
| HP_AZOOSPERMIA | 14 | 0.437 | 1.327 | 0.15930 | 0.15930 | 0.35617 | 257 |
| HP_ABNORMALITY_OF_SALIVATION | 11 | 0.468 | 1.325 | 0.15488 | 0.15488 | 0.35188 | 120 |
| HP_ORAL_CLEFT | 44 | 0.324 | 1.323 | 0.13067 | 0.13067 | 0.31591 | 215 |
| HP_OSTEOLYSIS | 10 | 0.482 | 1.322 | 0.15366 | 0.15366 | 0.35014 | 188 |
| HP_ABNORMAL_INTERNAL_GENITALIA | 50 | 0.315 | 1.320 | 0.12756 | 0.12756 | 0.31286 | 222 |
| GOBP_REGULATION_OF_DOUBLE_STRAND_BREAK_REPAIR | 16 | 0.417 | 1.319 | 0.15915 | 0.15915 | 0.35617 | 342 |
| GOBP_REGULATION_OF_NERVOUS_SYSTEM_DEVELOPMENT | 20 | 0.392 | 1.318 | 0.15403 | 0.15403 | 0.35045 | 161 |
| GOMF_PROTEIN_HETERODIMERIZATION_ACTIVITY | 24 | 0.373 | 1.318 | 0.14931 | 0.14931 | 0.34376 | 276 |
| GOMF_METALLOPEPTIDASE_ACTIVITY | 12 | 0.455 | 1.318 | 0.16089 | 0.16089 | 0.35839 | 88 |
| GOCC_UBIQUITIN_LIGASE_COMPLEX | 24 | 0.372 | 1.315 | 0.15024 | 0.15024 | 0.34488 | 290 |
| GOBP_REGULATION_OF_CYTOSKELETON_ORGANIZATION | 34 | 0.340 | 1.312 | 0.14367 | 0.14367 | 0.33364 | 257 |
| GOBP_REGULATION_OF_DNA_REPAIR | 21 | 0.385 | 1.311 | 0.15786 | 0.15786 | 0.35582 | 517 |
| HP_AMENORRHEA | 10 | 0.478 | 1.310 | 0.16188 | 0.16188 | 0.35903 | 50 |
| GOMF_PROTEIN_SERINE_THREONINE_KINASE_ACTIVITY | 36 | 0.335 | 1.309 | 0.14672 | 0.14672 | 0.33981 | 188 |
| GOMF_DAMAGED_DNA_BINDING | 15 | 0.420 | 1.302 | 0.17180 | 0.17180 | 0.37284 | 453 |
| HP_DOWNSLANTED_PALPEBRAL_FISSURES | 37 | 0.331 | 1.302 | 0.14812 | 0.14812 | 0.34205 | 196 |
| HP_NEOPLASM_OF_THE_STOMACH | 11 | 0.459 | 1.300 | 0.17337 | 0.17337 | 0.37519 | 222 |
| HP_ABNORMALITY_OF_THE_LOWER_URINARY_TRACT | 56 | 0.303 | 1.299 | 0.14114 | 0.14114 | 0.33086 | 460 |
| GOBP_HOMOLOGOUS_RECOMBINATION | 11 | 0.458 | 1.298 | 0.17559 | 0.17559 | 0.37769 | 359 |
| GOBP_SUPRAMOLECULAR_FIBER_ORGANIZATION | 34 | 0.336 | 1.296 | 0.15575 | 0.15575 | 0.35283 | 208 |
| GOCC_NEURON_TO_NEURON_SYNAPSE | 12 | 0.447 | 1.295 | 0.17646 | 0.17646 | 0.37870 | 88 |
| GOBP_ELECTRON_TRANSPORT_CHAIN | 14 | 0.426 | 1.295 | 0.18234 | 0.18234 | 0.38916 | 567 |
| GOCC_CHROMATIN | 92 | 0.279 | 1.294 | 0.12061 | 0.12061 | 0.30526 | 424 |

| | | | | | | | |
|---|---|---|---|---|---|---|---|
| GOBP_RESPONSE_TO_RADIATION | 36 | 0.330 | 1.293 | 0.15884 | 0.15884 | 0.35617 | 367 |
| GOMF_S_ADENOSYLMETHIONINE_DEPENDENT_METHYLTRANSFERASE_ACTIVITY | 10 | 0.472 | 1.293 | 0.17474 | 0.17474 | 0.37663 | 489 |
| HP_CONSTIPATION | 28 | 0.350 | 1.289 | 0.16510 | 0.16510 | 0.36235 | 103 |
| HP_ABNORMAL_SHAPE_OF_THE_PALPEBRAL_FISSURE | 10 | 0.470 | 1.289 | 0.17848 | 0.17848 | 0.38144 | 359 |
| GOMF_ADENYL_NUCLEOTIDE_BINDING | 132 | 0.264 | 1.289 | 0.10616 | 0.10616 | 0.28667 | 326 |
| GOBP_NEGATIVE_REGULATION_OF_CELL_GROWTH | 14 | 0.423 | 1.287 | 0.18663 | 0.18663 | 0.39291 | 50 |
| GOMF_CALCIUM_ION_BINDING | 28 | 0.350 | 1.287 | 0.16693 | 0.16693 | 0.36481 | 96 |
| GOBP_TRANSMEMBRANE_TRANSPORT | 52 | 0.305 | 1.287 | 0.15295 | 0.15295 | 0.34979 | 73 |
| HP_HYPERREFLEXIA | 54 | 0.302 | 1.285 | 0.15328 | 0.15328 | 0.34979 | 459 |
| GOBP_PROTEIN_MODIFICATION_BY_SMALL_PROTEIN_REMOVAL | 12 | 0.442 | 1.281 | 0.18778 | 0.18778 | 0.39351 | 95 |
| HP_ABNORMALITY_OF_THE_BLADDER | 32 | 0.336 | 1.280 | 0.17126 | 0.17126 | 0.37218 | 232 |
| HP_INTERICTAL_EEG_ABNORMALITY | 16 | 0.405 | 1.280 | 0.19199 | 0.19199 | 0.39985 | 141 |
| GOBP_EMBRYO_DEVELOPMENT_ENDING_IN_BIRTH_OR_EGG_HATCHING | 44 | 0.313 | 1.277 | 0.16411 | 0.16411 | 0.36119 | 261 |
| GOMF_TRANSPORTER_ACTIVITY | 42 | 0.316 | 1.277 | 0.16644 | 0.16644 | 0.36425 | 82 |
| GOBP_DEVELOPMENTAL_GROWTH_INVOLVED_IN_MORPHOGENESIS | 10 | 0.465 | 1.275 | 0.18924 | 0.18924 | 0.39476 | 46 |
| GOCC_TRANSPORT_VESICLE | 21 | 0.374 | 1.274 | 0.18333 | 0.18333 | 0.38924 | 38 |
| HP_FUNCTIONAL_ABNORMALITY_OF_MALE_INTERNAL_GENITALIA | 18 | 0.389 | 1.274 | 0.18818 | 0.18818 | 0.39351 | 257 |
| GOBP_CELL_CYCLE_G1_S_PHASE_TRANSITION | 32 | 0.334 | 1.274 | 0.17575 | 0.17575 | 0.37769 | 409 |
| GOMF_GTPASE_ACTIVATOR_ACTIVITY | 18 | 0.389 | 1.273 | 0.18929 | 0.18929 | 0.39476 | 255 |
| GOBP_NEGATIVE_REGULATION_OF_MOLECULAR_FUNCTION | 68 | 0.287 | 1.272 | 0.15687 | 0.15687 | 0.35483 | 181 |
| GOMF_ELECTRON_TRANSFER_ACTIVITY | 14 | 0.418 | 1.270 | 0.19737 | 0.19737 | 0.40667 | 567 |
| HP_ABNORMALITY_OF_THE_MENSTRUAL_CYCLE | 12 | 0.438 | 1.268 | 0.19880 | 0.19880 | 0.40761 | 50 |
| GOMF_UBIQUITIN_LIKE_PROTEIN_LIGASE_ACTIVITY | 17 | 0.395 | 1.267 | 0.19544 | 0.19544 | 0.40486 | 217 |
| GOBP_ION_TRANSMEMBRANE_TRANSPORT | 39 | 0.317 | 1.264 | 0.17698 | 0.17698 | 0.37928 | 73 |
| GOCC_PLASMA_MEMBRANE_PROTEIN_COMPLEX | 18 | 0.386 | 1.263 | 0.19484 | 0.19484 | 0.40416 | 73 |
| GOBP_DEVELOPMENT_OF_PRIMARY_SEXUAL_CHARACTERISTICS | 11 | 0.446 | 1.262 | 0.20178 | 0.20178 | 0.41138 | 122 |
| GOBP_NEGATIVE_REGULATION_OF_CELL_CYCLE_G2_M_PHASE_TRANSITION | 18 | 0.385 | 1.261 | 0.19692 | 0.19692 | 0.40629 | 306 |
| GOCC_EXOCYTIC_VESICLE | 13 | 0.424 | 1.260 | 0.20651 | 0.20651 | 0.41718 | 38 |
| HP_ABNORMALITY_OF_NEURONAL_MIGRATION | 37 | 0.319 | 1.256 | 0.18689 | 0.18689 | 0.39291 | 196 |
| GOCC_PLASMA_MEMBRANE_REGION | 55 | 0.294 | 1.256 | 0.17424 | 0.17424 | 0.37653 | 216 |
| GOMF_PROTEIN_C_TERMINUS_BINDING | 15 | 0.405 | 1.253 | 0.21204 | 0.21204 | 0.42614 | 215 |
| GOBP_CELLULAR_COMPONENT_DISASSEMBLY | 32 | 0.329 | 1.253 | 0.19485 | 0.19485 | 0.40416 | 208 |
| GOBP_IN_UTERO_EMBRYONIC_DEVELOPMENT | 32 | 0.329 | 1.252 | 0.19600 | 0.19600 | 0.40493 | 247 |
| HP_DYSKINESIA | 18 | 0.382 | 1.252 | 0.20469 | 0.20469 | 0.41567 | 141 |
| GOBP_REGULATION_OF_CELL_CYCLE_G1_S_PHASE_TRANSITION | 21 | 0.367 | 1.250 | 0.20621 | 0.20621 | 0.41713 | 409 |
| GOBP_REGULATION_OF_CENTROSOME_CYCLE | 12 | 0.431 | 1.249 | 0.21364 | 0.21364 | 0.42766 | 408 |
| GOBP_RESPONSE_TO_TOXIC_SUBSTANCE | 10 | 0.454 | 1.245 | 0.21390 | 0.21390 | 0.42766 | 349 |
| HP_ABNORMALITY_OF_THE_UTERUS | 28 | 0.338 | 1.245 | 0.20167 | 0.20167 | 0.41138 | 365 |
| GOBP_OSSIFICATION | 11 | 0.439 | 1.242 | 0.21568 | 0.21568 | 0.42910 | 107 |
| GOBP_PROTEIN_HOMOOLIGOMERIZATION | 11 | 0.439 | 1.242 | 0.21568 | 0.21568 | 0.42910 | 258 |
| HP_GENERALIZED_ONSET_SEIZURE | 22 | 0.359 | 1.242 | 0.21182 | 0.21182 | 0.42614 | 141 |

| | | | | | | | |
|---|---|---|---|---|---|---|---|
| HP_POOR_HEAD_CONTROL | 15 | 0.400 | 1.241 | 0.22160 | 0.22160 | 0.43486 | 66 |
| HP_GENERALIZED_MYOCLONIC_SEIZURE | 12 | 0.428 | 1.240 | 0.22172 | 0.22172 | 0.43486 | 124 |
| GOBP_TELENCEPHALON_DEVELOPMENT | 16 | 0.392 | 1.239 | 0.22526 | 0.22526 | 0.43959 | 161 |
| GOBP_DNA_STRAND_ELONGATION | 13 | 0.417 | 1.238 | 0.22611 | 0.22611 | 0.43959 | 517 |
| GOBP_CIRCADIAN_RHYTHM | 16 | 0.390 | 1.235 | 0.22893 | 0.22893 | 0.44229 | 99 |
| HP_ABNORMAL_SOFT_PALATE_MORPHOLOGY | 20 | 0.367 | 1.235 | 0.22012 | 0.22012 | 0.43337 | 323 |
| GOCC_ENDOPEPTIDASE_COMPLEX | 17 | 0.384 | 1.233 | 0.22592 | 0.22592 | 0.43959 | 649 |
| GOCC_PROTEASOME_COMPLEX | 17 | 0.384 | 1.233 | 0.22592 | 0.22592 | 0.43959 | 649 |
| HP_ABNORMALITY_OF_GLOBE_SIZE | 31 | 0.326 | 1.233 | 0.21522 | 0.21522 | 0.42910 | 257 |
| GOBP_REGULATION_OF_PROTEIN_SERINE_THREONINE_KINASE_ACTIVITY | 35 | 0.317 | 1.233 | 0.20794 | 0.20794 | 0.41952 | 264 |
| HP_URINARY_TRACT_NEOPLASM | 12 | 0.424 | 1.228 | 0.23185 | 0.23185 | 0.44628 | 284 |
| HP_ABNORMALITY_OF_THE_URETHRA | 33 | 0.320 | 1.226 | 0.21789 | 0.21789 | 0.43118 | 312 |
| GOBP_CELLULAR_MACROMOLECULE_BIOSYNTHETIC_PROCESS | 145 | 0.247 | 1.225 | 0.16049 | 0.16049 | 0.35832 | 367 |
| HP_STATUS_EPILEPTICUS | 19 | 0.369 | 1.224 | 0.22811 | 0.22811 | 0.44127 | 103 |
| HP_ABNORMAL_PALATE_MORPHOLOGY | 67 | 0.276 | 1.223 | 0.20072 | 0.20072 | 0.41044 | 215 |
| HP_ABNORMAL_ESOPHAGUS_MORPHOLOGY | 22 | 0.354 | 1.223 | 0.22729 | 0.22729 | 0.44078 | 421 |
| HP_GONOSOMAL_INHERITANCE | 13 | 0.411 | 1.221 | 0.24121 | 0.24121 | 0.45916 | 81 |
| HP_ABNORMALITY_OF_EXTRAPYRAMIDAL_MOTOR_FUNCTION | 16 | 0.385 | 1.217 | 0.24471 | 0.24471 | 0.46121 | 73 |
| GOBP_SECOND_MESSENGER_MEDIATED_SIGNALING | 10 | 0.444 | 1.217 | 0.24036 | 0.24036 | 0.45909 | 80 |
| HP_ABNORMALITY_OF_REPRODUCTIVE_SYSTEM_PHYSIOLOGY | 52 | 0.288 | 1.216 | 0.21354 | 0.21354 | 0.42766 | 222 |
| HP_HYPSARRHYTHMIA | 13 | 0.408 | 1.212 | 0.24775 | 0.24775 | 0.46349 | 141 |
| GOCC_GLUTAMATERGIC_SYNAPSE | 14 | 0.399 | 1.212 | 0.24360 | 0.24360 | 0.46033 | 96 |
| GOBP_NEGATIVE_REGULATION_OF_CELL_PROJECTION_ORGANIZATION | 11 | 0.428 | 1.211 | 0.24497 | 0.24497 | 0.46121 | 46 |
| GOBP_REGULATION_OF_ION_TRANSPORT | 28 | 0.329 | 1.210 | 0.23707 | 0.23707 | 0.45350 | 73 |
| GOMF_HISTONE_BINDING | 25 | 0.338 | 1.209 | 0.24319 | 0.24319 | 0.46033 | 238 |
| HP_ABNORMAL_NEURAL_TUBE_MORPHOLOGY | 15 | 0.390 | 1.209 | 0.24829 | 0.24829 | 0.46349 | 421 |
| HP_THICK_VERMILION_BORDER | 19 | 0.364 | 1.208 | 0.24107 | 0.24107 | 0.45916 | 306 |
| GOBP_NEUROTRANSMITTER_SECRETION | 11 | 0.426 | 1.206 | 0.24867 | 0.24867 | 0.46349 | 66 |
| GOBP_NEUROTRANSMITTER_TRANSPORT | 11 | 0.426 | 1.206 | 0.24867 | 0.24867 | 0.46349 | 66 |
| GOBP_REGULATION_OF_NEUROTRANSMITTER_LEVELS | 11 | 0.426 | 1.206 | 0.24867 | 0.24867 | 0.46349 | 66 |
| HP_OLIGOHYDRAMNIOS | 19 | 0.362 | 1.202 | 0.24700 | 0.24700 | 0.46349 | 257 |
| HP_APLASIA_HYPOPLASIA_INVOLVING_BONES_OF_THE_THORAX | 12 | 0.415 | 1.202 | 0.25507 | 0.25507 | 0.47275 | 323 |
| GOMF_SIGNALING_RECEPTOR_REGULATOR_ACTIVITY | 14 | 0.395 | 1.201 | 0.25590 | 0.25590 | 0.47374 | 41 |
| GOBP_PEPTIDYL_THREONINE_MODIFICATION | 14 | 0.394 | 1.199 | 0.25748 | 0.25748 | 0.47551 | 252 |
| GOMF_GTPASE_BINDING | 20 | 0.356 | 1.197 | 0.25295 | 0.25295 | 0.47052 | 202 |
| GOCC_CONDENSED_NUCLEAR_CHROMOSOME | 12 | 0.413 | 1.197 | 0.25904 | 0.25904 | 0.47689 | 423 |
| HP_NEOPLASM_OF_THE_LARGE_INTESTINE | 13 | 0.402 | 1.195 | 0.26358 | 0.26358 | 0.48142 | 284 |
| GOBP_MUSCLE_STRUCTURE_DEVELOPMENT | 36 | 0.304 | 1.189 | 0.25341 | 0.25341 | 0.47080 | 108 |
| GOBP_ACTIN_FILAMENT_ORGANIZATION | 18 | 0.363 | 1.189 | 0.26293 | 0.26293 | 0.48142 | 194 |
| HP_INTRAUTERINE_GROWTH_RETARDATION | 57 | 0.276 | 1.188 | 0.24059 | 0.24059 | 0.45909 | 328 |
| HP_MYOCLONUS | 22 | 0.344 | 1.188 | 0.25945 | 0.25945 | 0.47689 | 103 |

| | | | | | | | |
|---|---|---|---|---|---|---|---|
| HP_INCREASED_SUSCEPTIBILITY_TO_FRACTURES | 14 | 0.390 | 1.187 | 0.27007 | 0.27007 | 0.48568 | 73 |
| GOCC_PRESYNAPSE | 24 | 0.335 | 1.186 | 0.25919 | 0.25919 | 0.47689 | 66 |
| GOMF_HELICASE_ACTIVITY | 29 | 0.320 | 1.186 | 0.25844 | 0.25844 | 0.47672 | 347 |
| GOBP_G_PROTEIN_COUPLED_RECEPTOR_SIGNALING_PATHWAY | 22 | 0.343 | 1.184 | 0.26363 | 0.26363 | 0.48142 | 96 |
| HP_SOFT_TISSUE_SARCOMA | 11 | 0.418 | 1.183 | 0.26805 | 0.26805 | 0.48488 | 188 |
| HP_GLAUCOMA | 27 | 0.325 | 1.182 | 0.26409 | 0.26409 | 0.48142 | 232 |
| HP_DOWNTURNED_CORNERS_OF_MOUTH | 10 | 0.431 | 1.181 | 0.27130 | 0.27130 | 0.48598 | 120 |
| GOBP_PROTEIN_MODIFICATION_BY_SMALL_PROTEIN_CONJUGATION_OR_REMOVA | 77 | 0.261 | 1.181 | 0.23304 | 0.23304 | 0.44745 | 226 |
| HP_ABNORMALITY_OF_FEMALE_EXTERNAL_GENITALIA | 14 | 0.389 | 1.181 | 0.27322 | 0.27322 | 0.48829 | 306 |
| GOBP_FOREBRAIN_DEVELOPMENT | 21 | 0.346 | 1.179 | 0.26818 | 0.26818 | 0.48488 | 206 |
| GOBP_DENDRITE_DEVELOPMENT | 15 | 0.380 | 1.178 | 0.27554 | 0.27554 | 0.49083 | 158 |
| GOBP_PROTEIN_PROCESSING | 13 | 0.396 | 1.177 | 0.27621 | 0.27621 | 0.49085 | 15 |
| GOMF_CHROMATIN_BINDING | 62 | 0.270 | 1.176 | 0.24367 | 0.24367 | 0.46033 | 186 |
| HP_APLASIA_HYPOPLASIA_OF_THE_BREASTS | 10 | 0.429 | 1.176 | 0.27818 | 0.27818 | 0.49350 | 306 |
| HP_THIN_VERMILION_BORDER | 28 | 0.319 | 1.176 | 0.27299 | 0.27299 | 0.48829 | 208 |
| GOCC_PROTEASOME_CORE_COMPLEX | 11 | 0.415 | 1.174 | 0.27559 | 0.27559 | 0.49083 | 615 |
| GOBP_NEGATIVE_REGULATION_OF_GROWTH | 19 | 0.354 | 1.174 | 0.27486 | 0.27486 | 0.49065 | 107 |
| GOBP_REGULATION_OF_RESPONSE_TO_DNA_DAMAGE_STIMULUS | 29 | 0.316 | 1.173 | 0.27069 | 0.27069 | 0.48568 | 342 |
| HP_ABNORMALITY_OF_FLUID_REGULATION | 39 | 0.295 | 1.173 | 0.26578 | 0.26578 | 0.48335 | 206 |
| GOBP_PEPTIDYL_SERINE_MODIFICATION | 24 | 0.331 | 1.171 | 0.27624 | 0.27624 | 0.49085 | 111 |
| GOBP_ADAPTIVE_IMMUNE_RESPONSE_BASED_ON_SOMATIC_RECOMBINATION_OF | 17 | 0.364 | 1.170 | 0.28799 | 0.28799 | 0.50195 | 112 |
| HP_ABNORMALITY_OF_THE_PERIORBITAL_REGION | 18 | 0.357 | 1.169 | 0.28401 | 0.28401 | 0.49951 | 196 |
| HP_DISPLACEMENT_OF_THE_URETHRAL_MEATUS | 28 | 0.317 | 1.168 | 0.28174 | 0.28174 | 0.49664 | 312 |
| GOBP_ION_TRANSPORT | 56 | 0.272 | 1.167 | 0.26152 | 0.26152 | 0.48012 | 73 |
| GOMF_LIGASE_ACTIVITY | 10 | 0.425 | 1.165 | 0.28789 | 0.28789 | 0.50195 | 82 |
| GOBP_NEGATIVE_REGULATION_OF_DNA_METABOLIC_PROCESS | 16 | 0.368 | 1.164 | 0.29165 | 0.29165 | 0.50236 | 235 |
| GOBP_ORGANIC_HYDROXY_COMPOUND_BIOSYNTHETIC_PROCESS | 10 | 0.425 | 1.164 | 0.28924 | 0.28924 | 0.50236 | 387 |
| GOBP_CENTROSOME_DUPLICATION | 15 | 0.375 | 1.161 | 0.29095 | 0.29095 | 0.50236 | 408 |
| GOBP_REGULATION_OF_INTRACELLULAR_PROTEIN_TRANSPORT | 13 | 0.391 | 1.161 | 0.28899 | 0.28899 | 0.50236 | 202 |
| GOBP_CEREBRAL_CORTEX_DEVELOPMENT | 12 | 0.400 | 1.160 | 0.29165 | 0.29165 | 0.50236 | 161 |
| HP_FACIAL_EDEMA | 11 | 0.410 | 1.160 | 0.28772 | 0.28772 | 0.50195 | 192 |
| GOMF_TRANSFERASE_ACTIVITY_TRANSFERRING_ONE_CARBON_GROUPS | 16 | 0.366 | 1.159 | 0.29645 | 0.29645 | 0.50285 | 489 |
| GOBP_REGULATION_OF_TRANS_SYNAPTIC_SIGNALING | 15 | 0.374 | 1.158 | 0.29409 | 0.29409 | 0.50236 | 158 |
| GOBP_REGULATION_OF_GROWTH | 41 | 0.288 | 1.158 | 0.28115 | 0.28115 | 0.49617 | 107 |
| HP_LISSENCEPHALY | 16 | 0.366 | 1.157 | 0.29828 | 0.29828 | 0.50285 | 181 |
| HP_ABNORMALITY_OF_SKELETAL_MATURATION | 28 | 0.314 | 1.157 | 0.29284 | 0.29284 | 0.50236 | 192 |
| GOMF_STRUCTURAL_CONSTITUENT_OF_RIBOSOME | 11 | 0.408 | 1.157 | 0.29157 | 0.29157 | 0.50236 | 2 |
| GOBP_LYMPHOCYTE_MEDIATED_IMMUNITY | 18 | 0.353 | 1.155 | 0.29954 | 0.29954 | 0.50285 | 112 |
| GOBP_RESPONSE_TO_UV | 17 | 0.359 | 1.153 | 0.30561 | 0.30561 | 0.50808 | 365 |
| HP_NEOPLASM_OF_THE_BREAST | 14 | 0.379 | 1.152 | 0.30042 | 0.30042 | 0.50323 | 395 |
| GOCC_CELL_CELL_JUNCTION | 22 | 0.333 | 1.152 | 0.29471 | 0.29471 | 0.50236 | 202 |

| | | | | | | | |
|---|---|---|---|---|---|---|---|
| GOBP_ESTABLISHMENT_OF_CELL_POLARITY | 13 | 0.388 | 1.152 | 0.29814 | 0.29814 | 0.50285 | 337 |
| GOCC_MICROBODY | 10 | 0.420 | 1.150 | 0.30374 | 0.30374 | 0.50715 | 183 |
| GOBP_ORGANIC_ACID_CATABOLIC_PROCESS | 15 | 0.371 | 1.149 | 0.30479 | 0.30479 | 0.50801 | 171 |
| GOBP_CHROMATIN_REMODELING | 31 | 0.304 | 1.149 | 0.29985 | 0.29985 | 0.50285 | 185 |
| HP_FEMALE_REPRODUCTIVE_SYSTEM_NEOPLASM | 11 | 0.405 | 1.148 | 0.30178 | 0.30178 | 0.50496 | 188 |
| HP_SELF_INJURIOUS_BEHAVIOR | 13 | 0.386 | 1.147 | 0.30366 | 0.30366 | 0.50715 | 120 |
| HP_ABNORMAL_IMMUNE_SYSTEM_MORPHOLOGY | 46 | 0.278 | 1.147 | 0.29198 | 0.29198 | 0.50236 | 257 |
| GOMF_PROTEIN_KINASE_ACTIVITY | 43 | 0.282 | 1.146 | 0.29361 | 0.29361 | 0.50236 | 262 |
| GOBP_REGULATION_OF_DNA_BIOSYNTHETIC_PROCESS | 23 | 0.327 | 1.143 | 0.30491 | 0.30491 | 0.50801 | 347 |
| HP_PALLOR | 10 | 0.416 | 1.140 | 0.31375 | 0.31375 | 0.51884 | 200 |
| GOBP_PROTEIN_CONTAINING_COMPLEX_ORGANIZATION | 137 | 0.231 | 1.137 | 0.26909 | 0.26909 | 0.48568 | 222 |
| GOBP_NEGATIVE_REGULATION_OF_RESPONSE_TO_DNA_DAMAGE_STIMULUS | 10 | 0.414 | 1.136 | 0.31749 | 0.31749 | 0.52223 | 510 |
| GOMF_MOLECULAR_FUNCTION_REGULATOR | 97 | 0.242 | 1.136 | 0.28325 | 0.28325 | 0.49874 | 255 |
| HP_RECURRENT_URINARY_TRACT_INFECTIONS | 17 | 0.353 | 1.135 | 0.32001 | 0.32001 | 0.52359 | 376 |
| HP_MULTIPLE_CAFE_AU_LAIT_SPOTS | 13 | 0.382 | 1.135 | 0.31470 | 0.31470 | 0.51935 | 359 |
| HP_ABNORMALITY_OF_DIGESTIVE_SYSTEM_MORPHOLOGY | 71 | 0.254 | 1.134 | 0.29090 | 0.29090 | 0.50236 | 222 |
| GOBP_NEGATIVE_REGULATION_OF_PROTEIN_SERINE_THREONINE_KINASE_ACTIVIT | 10 | 0.414 | 1.134 | 0.31898 | 0.31898 | 0.52335 | 58 |
| HP_ABNORMAL_CARDIAC_SEPTUM_MORPHOLOGY | 55 | 0.265 | 1.134 | 0.29599 | 0.29599 | 0.50285 | 232 |
| GOBP_POSITIVE_REGULATION_OF_PROTEIN_LOCALIZATION | 26 | 0.314 | 1.133 | 0.31872 | 0.31872 | 0.52335 | 213 |
| HP_ABNORMALITY_OF_THE_NASAL_DORSUM | 16 | 0.358 | 1.133 | 0.32140 | 0.32140 | 0.52415 | 205 |
| HP_ABNORMALITY_OF_THE_RIBS | 16 | 0.358 | 1.132 | 0.32295 | 0.32295 | 0.52503 | 323 |
| GOBP_POSITIVE_REGULATION_OF_PROTEIN_MODIFICATION_BY_SMALL_PROTEIN_( | 16 | 0.358 | 1.131 | 0.32408 | 0.32408 | 0.52503 | 257 |
| HP_ABNORMALITY_OF_THE_CEREBRAL_SUBCORTEX | 79 | 0.248 | 1.130 | 0.29329 | 0.29329 | 0.50236 | 215 |
| HP_ABNORMALITY_OF_THE_KIDNEY | 92 | 0.243 | 1.128 | 0.29426 | 0.29426 | 0.50236 | 232 |
| GOMF_ENZYME_ACTIVATOR_ACTIVITY | 42 | 0.279 | 1.128 | 0.31473 | 0.31473 | 0.51935 | 255 |
| GOCC_PEPTIDASE_COMPLEX | 19 | 0.339 | 1.125 | 0.32975 | 0.32975 | 0.52903 | 696 |
| GOBP_HEAD_DEVELOPMENT | 40 | 0.281 | 1.125 | 0.31740 | 0.31740 | 0.52223 | 161 |
| HP_ANORECTAL_ANOMALY | 30 | 0.300 | 1.125 | 0.32608 | 0.32608 | 0.52630 | 365 |
| HP_POSTNATAL_GROWTH_RETARDATION | 19 | 0.339 | 1.125 | 0.33016 | 0.33016 | 0.52903 | 254 |
| HP_RESPIRATORY_FAILURE | 12 | 0.388 | 1.123 | 0.32427 | 0.32427 | 0.52503 | 306 |
| GOBP_PROTEIN_POLYMERIZATION | 15 | 0.362 | 1.123 | 0.32934 | 0.32934 | 0.52903 | 178 |
| GOBP_NEURON_DEVELOPMENT | 47 | 0.271 | 1.122 | 0.32042 | 0.32042 | 0.52371 | 71 |
| GOBP_HISTONE_METHYLATION | 12 | 0.387 | 1.122 | 0.32574 | 0.32574 | 0.52630 | 482 |
| GOCC_RECEPTOR_COMPLEX | 11 | 0.396 | 1.122 | 0.32988 | 0.32988 | 0.52903 | 63 |
| HP_ABNORMALITY_OF_UPPER_LIP_VERMILLION | 27 | 0.307 | 1.120 | 0.33325 | 0.33325 | 0.53174 | 257 |
| HP_NEOPLASM_OF_THE_COLON | 10 | 0.408 | 1.119 | 0.33543 | 0.33543 | 0.53174 | 284 |
| GOMF_UBIQUITIN_LIKE_PROTEIN_TRANSFERASE_ACTIVITY | 29 | 0.301 | 1.118 | 0.33299 | 0.33299 | 0.53174 | 358 |
| HP_DERMATOLOGICAL_MANIFESTATIONS_OF_SYSTEMIC_DISORDERS | 16 | 0.353 | 1.117 | 0.33634 | 0.33634 | 0.53174 | 76 |
| HP_APLASIA_HYPOPLASIA_OF_THE_RADIUS | 11 | 0.394 | 1.117 | 0.33624 | 0.33624 | 0.53174 | 359 |
| GOCC_APICAL_PART_OF_CELL | 10 | 0.407 | 1.115 | 0.34006 | 0.34006 | 0.53277 | 259 |
| HP_SARCOMA | 15 | 0.360 | 1.114 | 0.33861 | 0.33861 | 0.53174 | 192 |

| | | | | | | | |
|---|---|---|---|---|---|---|---|
| GOCC_GOLGI_APPARATUS | 91 | 0.240 | 1.113 | 0.31696 | 0.31696 | 0.52223 | 138 |
| GOBP_REGULATION_OF_INTRACELLULAR_TRANSPORT | 23 | 0.318 | 1.113 | 0.33655 | 0.33655 | 0.53174 | 202 |
| HP_LIMB_UNDERGROWTH | 22 | 0.322 | 1.111 | 0.33872 | 0.33872 | 0.53174 | 370 |
| HP_VENTRICULOMEGALY | 51 | 0.265 | 1.111 | 0.33172 | 0.33172 | 0.53043 | 215 |
| GOCC_OXIDOREDUCTASE_COMPLEX | 13 | 0.374 | 1.111 | 0.33779 | 0.33779 | 0.53174 | 658 |
| HP_WEIGHT_LOSS | 23 | 0.317 | 1.110 | 0.33950 | 0.33950 | 0.53244 | 359 |
| GOCC_PROTEIN_KINASE_COMPLEX | 17 | 0.345 | 1.108 | 0.35090 | 0.35090 | 0.53886 | 351 |
| HP_ABNORMAL_CEREBRAL_VENTRICLE_MORPHOLOGY | 66 | 0.251 | 1.108 | 0.32678 | 0.32678 | 0.52688 | 215 |
| HP_MULTICYSTIC_KIDNEY_DYSPLASIA | 13 | 0.373 | 1.108 | 0.34069 | 0.34069 | 0.53322 | 188 |
| GOBP_NEURON_APOPTOTIC_PROCESS | 13 | 0.372 | 1.107 | 0.34229 | 0.34229 | 0.53402 | 84 |
| HP_NEPHROTIC_SYNDROME | 13 | 0.372 | 1.106 | 0.34258 | 0.34258 | 0.53402 | 535 |
| HP_BREAST_CARCINOMA | 12 | 0.382 | 1.106 | 0.34426 | 0.34426 | 0.53479 | 395 |
| HP_ABNORMALITY_OF_THE_FOREHEAD | 81 | 0.242 | 1.105 | 0.32960 | 0.32960 | 0.52903 | 215 |
| HP_ABNORMALITY_OF_THE_SPINAL_CORD | 18 | 0.338 | 1.105 | 0.35446 | 0.35446 | 0.54164 | 421 |
| GOBP_RESPONSE_TO_LIGHT_STIMULUS | 20 | 0.328 | 1.102 | 0.35243 | 0.35243 | 0.54014 | 365 |
| HP_RENAL_INSUFFICIENCY | 32 | 0.289 | 1.102 | 0.34970 | 0.34970 | 0.53886 | 529 |
| GOBP_REGULATION_OF_SUPRAMOLECULAR_FIBER_ORGANIZATION | 12 | 0.380 | 1.102 | 0.34837 | 0.34837 | 0.53886 | 208 |
| GOBP_TELOMERE_ORGANIZATION | 27 | 0.302 | 1.101 | 0.35564 | 0.35564 | 0.54237 | 529 |
| HP_HYPERACTIVITY | 32 | 0.289 | 1.100 | 0.35444 | 0.35444 | 0.54164 | 196 |
| HP_ABNORMAL_INTESTINE_MORPHOLOGY | 54 | 0.258 | 1.099 | 0.34198 | 0.34198 | 0.53402 | 222 |
| GOBP_CELL_PROJECTION_ORGANIZATION | 74 | 0.244 | 1.097 | 0.33596 | 0.33596 | 0.53174 | 262 |
| GOMF_OXIDOREDUCTASE_ACTIVITY | 42 | 0.271 | 1.097 | 0.35053 | 0.35053 | 0.53886 | 27 |
| HP_GENITAL_NEOPLASM | 19 | 0.330 | 1.096 | 0.36147 | 0.36147 | 0.54652 | 370 |
| HP_SPARSE_HAIR | 19 | 0.330 | 1.096 | 0.36188 | 0.36188 | 0.54652 | 257 |
| GOBP_CELLULAR_RESPONSE_TO_UV | 13 | 0.369 | 1.096 | 0.35231 | 0.35231 | 0.54014 | 365 |
| GOBP_CELLULAR_LIPID_CATABOLIC_PROCESS | 14 | 0.360 | 1.093 | 0.35738 | 0.35738 | 0.54396 | 154 |
| HP_BIFID_UVULA | 11 | 0.386 | 1.093 | 0.36524 | 0.36524 | 0.54665 | 323 |
| HP_ABNORMAL_5TH_FINGER_MORPHOLOGY | 46 | 0.265 | 1.093 | 0.35799 | 0.35799 | 0.54436 | 316 |
| GOBP_PROTEIN_MODIFICATION_BY_SMALL_PROTEIN_CONJUGATION | 65 | 0.248 | 1.091 | 0.34951 | 0.34951 | 0.53886 | 226 |
| GOBP_PATTERN_SPECIFICATION_PROCESS | 22 | 0.316 | 1.091 | 0.36307 | 0.36307 | 0.54665 | 221 |
| GOCC_RIBOSOMAL_SUBUNIT | 13 | 0.367 | 1.090 | 0.35899 | 0.35899 | 0.54534 | 2 |
| HP_ABNORMAL_NASAL_BRIDGE_MORPHOLOGY | 62 | 0.250 | 1.089 | 0.35309 | 0.35309 | 0.54061 | 208 |
| HP_ABNORMAL_PENIS_MORPHOLOGY | 51 | 0.259 | 1.088 | 0.36020 | 0.36020 | 0.54652 | 215 |
| GOBP_CELLULAR_RESPONSE_TO_STRESS | 176 | 0.215 | 1.088 | 0.33365 | 0.33365 | 0.53174 | 426 |
| GOBP_NEGATIVE_REGULATION_OF_CELL_DIFFERENTIATION | 36 | 0.278 | 1.087 | 0.36742 | 0.36742 | 0.54718 | 46 |
| HP_DELAYED_PUBERTY | 11 | 0.383 | 1.086 | 0.37175 | 0.37175 | 0.55021 | 46 |
| HP_APLASIA_HYPOPLASIA_AFFECTING_THE_ANTERIOR_SEGMENT_OF_THE_EYE | 13 | 0.365 | 1.085 | 0.36422 | 0.36422 | 0.54665 | 421 |
| GOMF_CELL_ADHESION_MOLECULE_BINDING | 29 | 0.292 | 1.084 | 0.37195 | 0.37195 | 0.55021 | 165 |
| GOBP_POSITIVE_REGULATION_OF_ESTABLISHMENT_OF_PROTEIN_LOCALIZATION | 17 | 0.337 | 1.083 | 0.37816 | 0.37816 | 0.55383 | 202 |
| HP_CONGENITAL_ONSET | 23 | 0.310 | 1.083 | 0.37316 | 0.37316 | 0.55066 | 427 |
| GOBP_MODIFICATION_DEPENDENT_MACROMOLECULE_CATABOLIC_PROCESS | 54 | 0.255 | 1.083 | 0.36406 | 0.36406 | 0.54665 | 176 |

| | | | | | | | |
|---|---|---|---|---|---|---|---|
| GOBP_NEGATIVE_REGULATION_OF_CATALYTIC_ACTIVITY | 47 | 0.261 | 1.083 | 0.37225 | 0.37225 | 0.55021 | 181 |
| GOBP_PROTEIN_POLYUBIQUITINATION | 18 | 0.331 | 1.082 | 0.37526 | 0.37526 | 0.55217 | 290 |
| HP_NEOPLASM | 65 | 0.246 | 1.082 | 0.36159 | 0.36159 | 0.54652 | 376 |
| GOCC_RESPIRASOME | 12 | 0.373 | 1.082 | 0.36953 | 0.36953 | 0.54844 | 658 |
| HP_APLASIA_HYPOPLASIA_INVOLVING_FOREARM_BONES | 13 | 0.364 | 1.082 | 0.36756 | 0.36756 | 0.54718 | 359 |
| HP_ABNORMALITY_OF_MOUTH_SHAPE | 17 | 0.336 | 1.080 | 0.38278 | 0.38278 | 0.55757 | 124 |
| HP_ABNORMAL_CEREBRAL_CORTEX_MORPHOLOGY | 52 | 0.255 | 1.079 | 0.36999 | 0.36999 | 0.54860 | 196 |
| HP_DECREASED_HEAD_CIRCUMFERENCE | 107 | 0.227 | 1.078 | 0.36069 | 0.36069 | 0.54652 | 257 |
| HP_SYNOSTOSIS_OF_JOINTS | 11 | 0.381 | 1.077 | 0.38092 | 0.38092 | 0.55629 | 215 |
| HP_APLASIA_HYPOPLASIA_AFFECTING_THE_EYE | 33 | 0.281 | 1.076 | 0.38288 | 0.38288 | 0.55757 | 215 |
| GOBP_GLYCOSYLATION | 13 | 0.362 | 1.075 | 0.37351 | 0.37351 | 0.55066 | 70 |
| HP_HYPERPIGMENTATION_OF_THE_SKIN | 34 | 0.278 | 1.074 | 0.38436 | 0.38436 | 0.55921 | 257 |
| GOCC_NUCLEAR_BODY | 70 | 0.241 | 1.074 | 0.36435 | 0.36435 | 0.54665 | 277 |
| GOBP_REGULATION_OF_ORGANELLE_ASSEMBLY | 20 | 0.320 | 1.074 | 0.38582 | 0.38582 | 0.56028 | 257 |
| HP_HYPOPIGMENTED_SKIN_PATCHES | 15 | 0.346 | 1.072 | 0.38028 | 0.38028 | 0.55629 | 359 |
| HP_AGANGLIONIC_MEGACOLON | 13 | 0.361 | 1.071 | 0.37671 | 0.37671 | 0.55274 | 359 |
| HP_NAUSEA_AND_VOMITING | 30 | 0.286 | 1.071 | 0.38647 | 0.38647 | 0.56064 | 50 |
| GOBP_SENSORY_ORGAN_DEVELOPMENT | 20 | 0.318 | 1.070 | 0.39159 | 0.39159 | 0.56548 | 30 |
| HP_ABNORMALITY_OF_THE_NAIL | 27 | 0.293 | 1.069 | 0.39344 | 0.39344 | 0.56657 | 222 |
| HP_OBSESSIVE_COMPULSIVE_BEHAVIOR | 10 | 0.390 | 1.069 | 0.38759 | 0.38759 | 0.56128 | 73 |
| HP_ABNORMAL_EXTERNAL_GENITALIA | 80 | 0.235 | 1.068 | 0.38155 | 0.38155 | 0.55668 | 222 |
| HP_GENERALIZED_ONSET_MOTOR_SEIZURE | 14 | 0.351 | 1.066 | 0.38557 | 0.38557 | 0.56028 | 124 |
| GOBP_REGULATION_OF_PROTEIN_MODIFICATION_BY_SMALL_PROTEIN_CONJUGAT | 25 | 0.298 | 1.066 | 0.39513 | 0.39513 | 0.56795 | 200 |
| HP_DEVELOPMENTAL_REGRESSION | 20 | 0.317 | 1.065 | 0.39777 | 0.39777 | 0.56952 | 205 |
| HP_SKELETAL_DYSPLASIA | 11 | 0.376 | 1.064 | 0.39438 | 0.39438 | 0.56739 | 323 |
| HP_ABNORMAL_PATELLA_MORPHOLOGY | 12 | 0.367 | 1.064 | 0.39068 | 0.39068 | 0.56522 | 306 |
| GOBP_SKELETAL_SYSTEM_DEVELOPMENT | 17 | 0.331 | 1.064 | 0.39997 | 0.39997 | 0.57014 | 30 |
| GOBP_EMBRYONIC_MORPHOGENESIS | 24 | 0.300 | 1.062 | 0.39664 | 0.39664 | 0.56952 | 24 |
| GOBP_DNA_DEPENDENT_DNA_REPLICATION_MAINTENANCE_OF_FIDELITY | 11 | 0.374 | 1.060 | 0.39882 | 0.39882 | 0.56958 | 591 |
| HP_SPINA_BIFIDA | 13 | 0.356 | 1.059 | 0.39268 | 0.39268 | 0.56653 | 421 |
| GOBP_PROTEASOMAL_PROTEIN_CATABOLIC_PROCESS | 49 | 0.254 | 1.059 | 0.40124 | 0.40124 | 0.57042 | 176 |
| GOMF_RIBONUCLEOTIDE_BINDING | 147 | 0.213 | 1.058 | 0.38679 | 0.38679 | 0.56064 | 273 |
| GOBP_PROTEIN_COMPLEX_OLIGOMERIZATION | 16 | 0.334 | 1.057 | 0.39921 | 0.39921 | 0.56958 | 258 |
| GOBP_REGULATION_OF_TRANSPORT | 72 | 0.236 | 1.057 | 0.39335 | 0.39335 | 0.56657 | 78 |
| HP_INVOLUNTARY_MOVEMENTS | 64 | 0.241 | 1.057 | 0.40148 | 0.40148 | 0.57042 | 141 |
| HP_EXTERNAL_GENITAL_HYPOPLASIA | 41 | 0.262 | 1.055 | 0.40164 | 0.40164 | 0.57042 | 206 |
| HP_SOMATIC_MUTATION | 15 | 0.341 | 1.055 | 0.39912 | 0.39912 | 0.56958 | 257 |
| GOBP_REGULATION_OF_CELL_DEVELOPMENT | 29 | 0.284 | 1.054 | 0.40779 | 0.40779 | 0.57632 | 176 |
| GOBP_BASE_EXCISION_REPAIR | 10 | 0.385 | 1.054 | 0.40149 | 0.40149 | 0.57042 | 514 |
| HP_MOTOR_SEIZURE | 24 | 0.298 | 1.053 | 0.40703 | 0.40703 | 0.57632 | 141 |
| GOMF_KINASE_ACTIVITY | 47 | 0.254 | 1.053 | 0.40966 | 0.40966 | 0.57731 | 262 |

| | | | | | | | |
|---|---|---|---|---|---|---|---|
| GOBP_INORGANIC_ION_TRANSMEMBRANE_TRANSPORT | 32 | 0.276 | 1.053 | 0.41174 | 0.41174 | 0.57788 | 73 |
| HP_ABNORMALITY_OF_BONE_MINERAL_DENSITY | 30 | 0.280 | 1.050 | 0.41239 | 0.41239 | 0.57827 | 215 |
| GOBP_SENSORY_PERCEPTION | 19 | 0.316 | 1.050 | 0.41318 | 0.41318 | 0.57834 | 75 |
| GOBP_BONE_DEVELOPMENT | 12 | 0.362 | 1.049 | 0.40802 | 0.40802 | 0.57632 | 107 |
| HP_PERIORBITAL_EDEMA | 10 | 0.382 | 1.047 | 0.41001 | 0.41001 | 0.57731 | 165 |
| GOBP_ADAPTIVE_IMMUNE_RESPONSE | 21 | 0.307 | 1.046 | 0.41841 | 0.41841 | 0.58313 | 112 |
| HP_ABNORMAL_TRACHEAL_MORPHOLOGY | 15 | 0.337 | 1.045 | 0.41025 | 0.41025 | 0.57731 | 446 |
| GOMF_CATALYTIC_ACTIVITY_ACTING_ON_A_NUCLEIC_ACID | 68 | 0.235 | 1.044 | 0.40783 | 0.40783 | 0.57632 | 597 |
| GOBP_AEROBIC_RESPIRATION | 17 | 0.325 | 1.044 | 0.42234 | 0.42234 | 0.58434 | 555 |
| GOBP_CELLULAR_RESPIRATION | 17 | 0.325 | 1.044 | 0.42234 | 0.42234 | 0.58434 | 555 |
| HP_MYOCLONIC_SEIZURE | 15 | 0.337 | 1.044 | 0.41139 | 0.41139 | 0.57788 | 124 |
| HP_LETHARGY | 14 | 0.343 | 1.044 | 0.41005 | 0.41005 | 0.57731 | 76 |
| GOBP_CELLULAR_RESPONSE_TO_RADIATION | 25 | 0.292 | 1.044 | 0.42158 | 0.42158 | 0.58434 | 365 |
| GOBP_REGULATION_OF_NEURON_PROJECTION_DEVELOPMENT | 20 | 0.310 | 1.041 | 0.42567 | 0.42567 | 0.58571 | 233 |
| GOBP_CELLULAR_RESPONSE_TO_ORGANIC_CYCLIC_COMPOUND | 35 | 0.268 | 1.041 | 0.42424 | 0.42424 | 0.58512 | 257 |
| GOCC_ORGANELLAR_RIBOSOME | 10 | 0.379 | 1.040 | 0.41943 | 0.41943 | 0.58339 | 651 |
| HP_ABNORMAL_AGGRESSIVE_IMPULSIVE_OR_VIOLENT_BEHAVIOR | 24 | 0.294 | 1.040 | 0.42621 | 0.42621 | 0.58571 | 196 |
| HP_RENAL_CYST | 25 | 0.290 | 1.039 | 0.42727 | 0.42727 | 0.58640 | 206 |
| GOBP_REGULATION_OF_TRANSFERASE_ACTIVITY | 89 | 0.224 | 1.038 | 0.42107 | 0.42107 | 0.58434 | 317 |
| GOBP_POSITIVE_REGULATION_OF_CELLULAR_COMPONENT_ORGANIZATION | 59 | 0.240 | 1.038 | 0.42222 | 0.42222 | 0.58434 | 199 |
| HP_ABNORMALITY_OF_THE_CALVARIA | 63 | 0.237 | 1.038 | 0.42293 | 0.42293 | 0.58459 | 232 |
| GOMF_METAL_CLUSTER_BINDING | 14 | 0.341 | 1.036 | 0.41663 | 0.41663 | 0.58158 | 571 |
| GOBP_CELL_JUNCTION_ORGANIZATION | 25 | 0.289 | 1.036 | 0.43071 | 0.43071 | 0.58803 | 225 |
| HP_ONSET | 73 | 0.231 | 1.036 | 0.42445 | 0.42445 | 0.58512 | 232 |
| GOBP_DEVELOPMENTAL_MATURATION | 13 | 0.348 | 1.035 | 0.41868 | 0.41868 | 0.58313 | 176 |
| GOBP_PROTEIN_LOCALIZATION_TO_MITOCHONDRION | 15 | 0.334 | 1.034 | 0.42237 | 0.42237 | 0.58434 | 700 |
| HP_VISUAL_LOSS | 12 | 0.357 | 1.033 | 0.42345 | 0.42345 | 0.58478 | 24 |
| GOBP_PEPTIDYL_LYSINE_METHYLATION | 11 | 0.365 | 1.032 | 0.42944 | 0.42944 | 0.58781 | 489 |
| HP_ABNORMALITY_OF_PRENATAL_DEVELOPMENT_OR_BIRTH | 59 | 0.238 | 1.031 | 0.43111 | 0.43111 | 0.58803 | 215 |
| GOCC_RIBOSOME | 14 | 0.339 | 1.030 | 0.42493 | 0.42493 | 0.58526 | 2 |
| HP_IMPAIRMENT_IN_PERSONALITY_FUNCTIONING | 50 | 0.246 | 1.030 | 0.43567 | 0.43567 | 0.59268 | 161 |
| HP_HYPOPLASIA_OF_THE_CORPUS_CALLOSUM | 31 | 0.272 | 1.028 | 0.44062 | 0.44062 | 0.59632 | 161 |
| HP_ABNORMAL_EMOTION_AFFECT_BEHAVIOR | 46 | 0.249 | 1.026 | 0.44752 | 0.44752 | 0.60088 | 226 |
| HP_POINTED_CHIN | 11 | 0.362 | 1.025 | 0.43905 | 0.43905 | 0.59521 | 198 |
| GOBP_POSITIVE_REGULATION_OF_PROTEIN_SERINE_THREONINE_KINASE_ACTIVITY | 13 | 0.345 | 1.025 | 0.43015 | 0.43015 | 0.58803 | 311 |
| HP_PES_CAVUS | 22 | 0.296 | 1.024 | 0.44422 | 0.44422 | 0.60009 | 46 |
| HP_FINGER_CLINODACTYLY | 42 | 0.253 | 1.024 | 0.44189 | 0.44189 | 0.59747 | 316 |
| HP_ABNORMAL_CSF_METABOLITE_LEVEL | 12 | 0.353 | 1.022 | 0.43726 | 0.43726 | 0.59432 | 555 |
| GOBP_REGULATION_OF_NEUROGENESIS | 18 | 0.312 | 1.022 | 0.44599 | 0.44599 | 0.60088 | 161 |
| GOBP_REGULATION_OF_CELL_PROJECTION_ORGANIZATION | 35 | 0.263 | 1.022 | 0.44630 | 0.44630 | 0.60088 | 233 |
| HP_CEREBELLAR_MALFORMATION | 12 | 0.353 | 1.022 | 0.43829 | 0.43829 | 0.59520 | 206 |

| | | | | | | | |
|---|---|---|---|---|---|---|---|
| GOBP_REGULATION_OF_LYMPHOCYTE_MEDIATED_IMMUNITY | 10 | 0.372 | 1.019 | 0.44738 | 0.44738 | 0.60088 | 61 |
| HP_ABNORMALITY_OF_THE_TESTIS_SIZE | 20 | 0.303 | 1.018 | 0.45205 | 0.45205 | 0.60549 | 257 |
| GOBP_POSITIVE_REGULATION_OF_CHROMOSOME_ORGANIZATION | 15 | 0.328 | 1.017 | 0.43907 | 0.43907 | 0.59521 | 662 |
| GOBP_REGULATION_OF_MULTICELLULAR_ORGANISMAL_DEVELOPMENT | 63 | 0.232 | 1.016 | 0.45475 | 0.45475 | 0.60693 | 122 |
| HP_FOCAL_ONSET_SEIZURE | 19 | 0.306 | 1.016 | 0.45414 | 0.45414 | 0.60693 | 141 |
| GOBP_REGULATION_OF_AUTOPHAGY | 20 | 0.302 | 1.014 | 0.45672 | 0.45672 | 0.60784 | 14 |
| HP_SHORT_LONG_BONE | 20 | 0.301 | 1.013 | 0.45837 | 0.45837 | 0.60784 | 359 |
| HP_EPILEPTIC_SPASM | 15 | 0.327 | 1.013 | 0.44592 | 0.44592 | 0.60088 | 141 |
| GOBP_CATION_TRANSMEMBRANE_TRANSPORT | 34 | 0.262 | 1.012 | 0.45887 | 0.45887 | 0.60784 | 73 |
| GOBP_GERM_CELL_DEVELOPMENT | 18 | 0.309 | 1.011 | 0.45833 | 0.45833 | 0.60784 | 221 |
| GOBP_DEVELOPMENTAL_GROWTH | 41 | 0.251 | 1.008 | 0.46245 | 0.46245 | 0.61035 | 247 |
| GOBP_PALLIUM_DEVELOPMENT | 14 | 0.331 | 1.007 | 0.45213 | 0.45213 | 0.60549 | 161 |
| GOBP_RHYTHMIC_PROCESS | 21 | 0.295 | 1.006 | 0.46677 | 0.46677 | 0.61344 | 99 |
| GOBP_GLYCEROLIPID_METABOLIC_PROCESS | 15 | 0.325 | 1.006 | 0.45477 | 0.45477 | 0.60693 | 141 |
| HP_ABNORMALITY_OF_LOWER_LIMB_JOINT | 46 | 0.244 | 1.006 | 0.47471 | 0.47471 | 0.61707 | 312 |
| HP_ABNORMALITY_OF_THE_BREAST | 39 | 0.252 | 1.005 | 0.47082 | 0.47082 | 0.61626 | 312 |
| HP_ABNORMALITY_OF_CENTRAL_NERVOUS_SYSTEM_ELECTROPHYSIOLOGY | 33 | 0.262 | 1.004 | 0.47054 | 0.47054 | 0.61626 | 163 |
| GOCC_CATALYTIC_COMPLEX | 127 | 0.206 | 1.004 | 0.47364 | 0.47364 | 0.61672 | 663 |
| GOCC_FIBRILLAR_CENTER | 10 | 0.366 | 1.003 | 0.46547 | 0.46547 | 0.61225 | 4 |
| GOBP_TRNA_METABOLIC_PROCESS | 12 | 0.346 | 1.003 | 0.46209 | 0.46209 | 0.61035 | 686 |
| HP_RETINAL_DEGENERATION | 14 | 0.330 | 1.003 | 0.45799 | 0.45799 | 0.60784 | 24 |
| HP_NEOPLASM_OF_THE_GENITOURINARY_TRACT | 22 | 0.290 | 1.001 | 0.47598 | 0.47598 | 0.61764 | 370 |
| GOMF_CIS_REGULATORY_REGION_SEQUENCE_SPECIFIC_DNA_BINDING | 59 | 0.231 | 1.000 | 0.47190 | 0.47190 | 0.61652 | 237 |
| GOBP_CELLULAR_RESPONSE_TO_LIGHT_STIMULUS | 14 | 0.328 | 0.998 | 0.46286 | 0.46286 | 0.61037 | 365 |
| GOBP_CYTOSKELETON_DEPENDENT_INTRACELLULAR_TRANSPORT | 11 | 0.352 | 0.998 | 0.47130 | 0.47130 | 0.61626 | 81 |
| GOBP_TRANSPORT_ALONG_MICROTUBULE | 11 | 0.352 | 0.998 | 0.47130 | 0.47130 | 0.61626 | 81 |
| HP_ABNORMALITY_OF_THE_NASAL_TIP | 20 | 0.297 | 0.997 | 0.47664 | 0.47664 | 0.61764 | 196 |
| HP_ABNORMAL_REFLEX | 72 | 0.222 | 0.995 | 0.48233 | 0.48233 | 0.62280 | 215 |
| HP_ABNORMAL_NUMBER_OF_TEETH | 23 | 0.284 | 0.995 | 0.48445 | 0.48445 | 0.62501 | 222 |
| GOBP_ACTIN_FILAMENT_BASED_PROCESS | 40 | 0.248 | 0.994 | 0.48736 | 0.48736 | 0.62720 | 216 |
| GOBP_PROTEIN_TARGETING_TO_MITOCHONDRION | 12 | 0.343 | 0.993 | 0.47443 | 0.47443 | 0.61707 | 462 |
| HP_FAILURE_TO_THRIVE_IN_INFANCY | 10 | 0.362 | 0.993 | 0.47833 | 0.47833 | 0.61918 | 219 |
| GOBP_NEGATIVE_REGULATION_OF_PROTEOLYSIS | 16 | 0.314 | 0.992 | 0.47237 | 0.47237 | 0.61657 | 119 |
| GOBP_CELL_JUNCTION_ASSEMBLY | 14 | 0.326 | 0.991 | 0.47073 | 0.47073 | 0.61626 | 69 |
| GOBP_REGULATION_OF_CELLULAR_COMPONENT_BIOGENESIS | 53 | 0.234 | 0.990 | 0.48950 | 0.48950 | 0.62891 | 259 |
| HP_ABNORMAL_CIRCULATING_HORMONE_CONCENTRATION | 24 | 0.279 | 0.987 | 0.49534 | 0.49534 | 0.63260 | 50 |
| GOBP_METHYLATION | 23 | 0.282 | 0.987 | 0.49410 | 0.49410 | 0.63168 | 489 |
| GOBP_POSITIVE_REGULATION_OF_TRANSFERASE_ACTIVITY | 52 | 0.233 | 0.986 | 0.49215 | 0.49215 | 0.63127 | 358 |
| HP_ABNORMALITY_OF_SKIN_PIGMENTATION | 42 | 0.244 | 0.985 | 0.49347 | 0.49347 | 0.63168 | 257 |
| HP_ABNORMAL_VASCULAR_PHYSIOLOGY | 14 | 0.324 | 0.985 | 0.47674 | 0.47674 | 0.61764 | 76 |
| HP_MENTAL_DETERIORATION | 16 | 0.311 | 0.985 | 0.47970 | 0.47970 | 0.62044 | 22 |

| | | | | | | | |
|---|---|---|---|---|---|---|---|
| GOCC_MITOCHONDRIAL_MATRIX | 41 | 0.245 | 0.984 | 0.49677 | 0.49677 | 0.63300 | 658 |
| GOMF_CADHERIN_BINDING | 19 | 0.296 | 0.983 | 0.49579 | 0.49579 | 0.63260 | 165 |
| GOBP_POSITIVE_REGULATION_OF_CELL_POPULATION_PROLIFERATION | 52 | 0.233 | 0.982 | 0.49783 | 0.49783 | 0.63331 | 161 |
| GOBP_PROTEIN_MATURATION | 18 | 0.299 | 0.980 | 0.49605 | 0.49605 | 0.63260 | 15 |
| GOBP_CHROMATIN_ORGANIZATION | 59 | 0.226 | 0.980 | 0.50534 | 0.50534 | 0.63700 | 185 |
| GOBP_REGULATION_OF_INTRACELLULAR_SIGNAL_TRANSDUCTION | 90 | 0.211 | 0.979 | 0.51476 | 0.51476 | 0.64427 | 245 |
| HP_IMPAIRED_SOCIAL_INTERACTIONS | 11 | 0.345 | 0.978 | 0.49749 | 0.49749 | 0.63331 | 555 |
| HP_IRREGULAR_HYPERPIGMENTATION | 27 | 0.268 | 0.977 | 0.50566 | 0.50566 | 0.63700 | 222 |
| GOBP_HOMEOSTASIS_OF_NUMBER_OF_CELLS | 17 | 0.304 | 0.977 | 0.50035 | 0.50035 | 0.63547 | 232 |
| HP_HIGH_FOREHEAD | 19 | 0.294 | 0.975 | 0.50545 | 0.50545 | 0.63700 | 188 |
| HP_HYDROCEPHALUS | 25 | 0.272 | 0.973 | 0.51283 | 0.51283 | 0.64299 | 376 |
| HP_HYPOPIGMENTATION_OF_THE_SKIN | 19 | 0.293 | 0.973 | 0.50738 | 0.50738 | 0.63864 | 257 |
| GOMF_EXONUCLEASE_ACTIVITY | 13 | 0.327 | 0.972 | 0.49390 | 0.49390 | 0.63168 | 642 |
| HP_ABNORMALITY_OF_TEMPERATURE_REGULATION | 29 | 0.262 | 0.972 | 0.51297 | 0.51297 | 0.64299 | 205 |
| HP_NEOPLASM_OF_THE_GASTROINTESTINAL_TRACT | 27 | 0.266 | 0.968 | 0.51844 | 0.51844 | 0.64652 | 365 |
| GOBP_REGULATION_OF_GENE_EXPRESSION_EPIGENETIC | 15 | 0.312 | 0.968 | 0.50128 | 0.50128 | 0.63562 | 446 |
| GOMF_ENDOPEPTIDASE_ACTIVITY | 25 | 0.270 | 0.968 | 0.51865 | 0.51865 | 0.64652 | 88 |
| HP_CEREBRAL_VISUAL_IMPAIRMENT | 15 | 0.312 | 0.967 | 0.50214 | 0.50214 | 0.63588 | 141 |
| GOMF_ENZYME_REGULATOR_ACTIVITY | 74 | 0.215 | 0.966 | 0.52841 | 0.52841 | 0.65415 | 255 |
| GOBP_GLAND_DEVELOPMENT | 30 | 0.258 | 0.965 | 0.52320 | 0.52320 | 0.64959 | 181 |
| HP_SHORT_PALPEBRAL_FISSURE | 12 | 0.332 | 0.963 | 0.50926 | 0.50926 | 0.64049 | 359 |
| HP_HETEROGENEOUS | 13 | 0.324 | 0.962 | 0.50988 | 0.50988 | 0.64074 | 312 |
| GOBP_REGULATION_OF_PROTEIN_CONTAINING_COMPLEX_ASSEMBLY | 14 | 0.316 | 0.961 | 0.50408 | 0.50408 | 0.63656 | 178 |
| GOBP_PHOSPHORYLATION | 125 | 0.198 | 0.960 | 0.54867 | 0.54867 | 0.66836 | 269 |
| HP_GLOMERULAR_SCLEROSIS | 11 | 0.338 | 0.957 | 0.52160 | 0.52160 | 0.64941 | 529 |
| HP_ABNORMALITY_OF_THE_OUTER_EAR | 85 | 0.208 | 0.957 | 0.54451 | 0.54451 | 0.66749 | 222 |
| GOBP_NEGATIVE_REGULATION_OF_DEVELOPMENTAL_PROCESS | 47 | 0.231 | 0.956 | 0.53839 | 0.53839 | 0.66208 | 122 |
| GOBP_OXIDATIVE_PHOSPHORYLATION | 13 | 0.322 | 0.955 | 0.51844 | 0.51844 | 0.64652 | 555 |
| GOBP_PYRIMIDINE_CONTAINING_COMPOUND_METABOLIC_PROCESS | 10 | 0.349 | 0.955 | 0.52317 | 0.52317 | 0.64959 | 548 |
| GOBP_CELLULAR_RESPONSE_TO_ABIOTIC_STIMULUS | 31 | 0.252 | 0.953 | 0.53838 | 0.53838 | 0.66208 | 264 |
| HP_ABNORMAL_RESPIRATORY_SYSTEM_MORPHOLOGY | 82 | 0.209 | 0.953 | 0.55268 | 0.55268 | 0.67061 | 226 |
| GOBP_NEGATIVE_REGULATION_OF_PEPTIDASE_ACTIVITY | 10 | 0.348 | 0.953 | 0.52511 | 0.52511 | 0.65144 | 119 |
| GOBP_IMMUNOGLOBULIN_PRODUCTION | 13 | 0.320 | 0.950 | 0.52643 | 0.52643 | 0.65255 | 125 |
| HP_FEVER | 24 | 0.269 | 0.950 | 0.54089 | 0.54089 | 0.66463 | 205 |
| HP_CAFE_AU_LAIT_SPOT | 21 | 0.279 | 0.949 | 0.53773 | 0.53773 | 0.66208 | 257 |
| HP_ABNORMAL_INVOLUNTARY_EYE_MOVEMENTS | 80 | 0.208 | 0.948 | 0.55633 | 0.55633 | 0.67226 | 215 |
| GOBP_REGULATION_OF_CELLULAR_LOCALIZATION | 49 | 0.227 | 0.946 | 0.55449 | 0.55449 | 0.67175 | 268 |
| GOBP_DENDRITE_MORPHOGENESIS | 10 | 0.345 | 0.946 | 0.53423 | 0.53423 | 0.65958 | 138 |
| GOBP_PROGRAMMED_CELL_DEATH | 120 | 0.196 | 0.946 | 0.57407 | 0.57407 | 0.68318 | 258 |
| HP_ABNORMALITY_OF_LONG_BONE_MORPHOLOGY | 56 | 0.221 | 0.945 | 0.55101 | 0.55101 | 0.66963 | 323 |
| GOBP_HISTONE_MODIFICATION | 36 | 0.242 | 0.945 | 0.54836 | 0.54836 | 0.66836 | 257 |

| | | | | | | | |
|---|---|---|---|---|---|---|---|
| HP_ABNORMALITY_OF_FEMUR_MORPHOLOGY | 27 | 0.259 | 0.945 | 0.54544 | 0.54544 | 0.66782 | 323 |
| GOBP_REGULATION_OF_CELLULAR_MACROMOLECULE_BIOSYNTHETIC_PROCESS | 70 | 0.212 | 0.943 | 0.56721 | 0.56721 | 0.67866 | 349 |
| GOBP_PROTEIN_METHYLATION | 14 | 0.310 | 0.941 | 0.53127 | 0.53127 | 0.65698 | 489 |
| HP_CLEFT_SOFT_PALATE | 12 | 0.325 | 0.941 | 0.53379 | 0.53379 | 0.65957 | 323 |
| GOBP_FATTY_ACID_METABOLIC_PROCESS | 19 | 0.283 | 0.940 | 0.54792 | 0.54792 | 0.66836 | 92 |
| GOBP_POSITIVE_REGULATION_OF_DNA_REPAIR | 12 | 0.324 | 0.939 | 0.53688 | 0.53688 | 0.66180 | 709 |
| HP_HYPOPLASIA_OF_PENIS | 35 | 0.242 | 0.939 | 0.55966 | 0.55966 | 0.67435 | 206 |
| GOMF_SEQUENCE_SPECIFIC_DNA_BINDING | 92 | 0.202 | 0.938 | 0.57836 | 0.57836 | 0.68605 | 308 |
| GOBP_PROTEIN_INSERTION_INTO_MEMBRANE | 10 | 0.342 | 0.936 | 0.54589 | 0.54589 | 0.66782 | 506 |
| GOMF_KINASE_REGULATOR_ACTIVITY | 23 | 0.267 | 0.935 | 0.55524 | 0.55524 | 0.67214 | 250 |
| GOBP_MESENCHYMAL_CELL_DIFFERENTIATION | 12 | 0.322 | 0.932 | 0.54628 | 0.54628 | 0.66782 | 41 |
| HP_TOE_SYNDACTYLY | 20 | 0.277 | 0.930 | 0.56499 | 0.56499 | 0.67758 | 257 |
| HP_APLASIA_HYPOPLASIA_OF_THE_CEREBELLAR_VERMIS | 18 | 0.284 | 0.930 | 0.55665 | 0.55665 | 0.67226 | 232 |
| HP_GRAY_MATTER_HETEROTOPIA | 12 | 0.321 | 0.929 | 0.55040 | 0.55040 | 0.66941 | 181 |
| GOBP_NEURON_DEATH | 22 | 0.269 | 0.929 | 0.56170 | 0.56170 | 0.67573 | 84 |
| GOMF_NUCLEAR_RECEPTOR_BINDING | 13 | 0.312 | 0.928 | 0.55591 | 0.55591 | 0.67226 | 4 |
| GOBP_MICROTUBULE_BASED_TRANSPORT | 12 | 0.319 | 0.924 | 0.55789 | 0.55789 | 0.67324 | 81 |
| GOBP_REGULATION_OF_BINDING | 22 | 0.267 | 0.923 | 0.56829 | 0.56829 | 0.67866 | 178 |
| GOBP_TELOMERE_MAINTENANCE | 23 | 0.264 | 0.922 | 0.56852 | 0.56852 | 0.67866 | 662 |
| GOMF_HYDROLASE_ACTIVITY_ACTING_ON_ACID_ANHYDRIDES | 63 | 0.210 | 0.921 | 0.58873 | 0.58873 | 0.69004 | 554 |
| HP_ATROPHY_DEGENERATION_AFFECTING_THE_CEREBRUM | 54 | 0.216 | 0.919 | 0.58840 | 0.58840 | 0.69004 | 103 |
| GOCC_TRANSFERASE_COMPLEX_TRANSFERRING_PHOSPHORUS_CONTAINING_GI | 28 | 0.249 | 0.918 | 0.58386 | 0.58386 | 0.68766 | 365 |
| GOBP_CELLULAR_MODIFIED_AMINO_ACID_METABOLIC_PROCESS | 12 | 0.316 | 0.917 | 0.56671 | 0.56671 | 0.67859 | 401 |
| GOBP_LEUKOCYTE_MEDIATED_IMMUNITY | 21 | 0.269 | 0.916 | 0.58227 | 0.58227 | 0.68740 | 112 |
| HP_ABNORMAL_LARGE_INTESTINE_MORPHOLOGY | 24 | 0.259 | 0.915 | 0.58205 | 0.58205 | 0.68740 | 284 |
| HP_THICKENED_SKIN | 18 | 0.279 | 0.915 | 0.57704 | 0.57704 | 0.68513 | 48 |
| GOBP_REGULATION_OF_INTRINSIC_APOPTOTIC_SIGNALING_PATHWAY | 11 | 0.322 | 0.911 | 0.57544 | 0.57544 | 0.68417 | 419 |
| GOBP_PEPTIDE_BIOSYNTHETIC_PROCESS | 41 | 0.226 | 0.910 | 0.60109 | 0.60109 | 0.69576 | 48 |
| HP_ABNORMALITY_OF_GLOBE_LOCATION | 76 | 0.202 | 0.910 | 0.61632 | 0.61632 | 0.70166 | 215 |
| HP_FUNCTIONAL_ABNORMALITY_OF_THE_INNER_EAR | 49 | 0.218 | 0.910 | 0.60496 | 0.60496 | 0.69640 | 46 |
| GOBP_NEGATIVE_REGULATION_OF_NEURON_DEATH | 11 | 0.321 | 0.909 | 0.57944 | 0.57944 | 0.68605 | 84 |
| HP_ABNORMALITY_OF_THE_NECK | 35 | 0.234 | 0.908 | 0.60467 | 0.60467 | 0.69640 | 141 |
| GOMF_KINASE_BINDING | 72 | 0.203 | 0.908 | 0.61916 | 0.61916 | 0.70439 | 267 |
| HP_ABNORMAL_NERVOUS_SYSTEM_ELECTROPHYSIOLOGY | 39 | 0.228 | 0.907 | 0.60215 | 0.60215 | 0.69576 | 183 |
| GOBP_HORMONE_MEDIATED_SIGNALING_PATHWAY | 14 | 0.298 | 0.907 | 0.58294 | 0.58294 | 0.68740 | 4 |
| GOBP_STEROID_HORMONE_MEDIATED_SIGNALING_PATHWAY | 14 | 0.298 | 0.907 | 0.58294 | 0.58294 | 0.68740 | 4 |
| HP_ABNORMAL_FEAR_ANXIETY_RELATED_BEHAVIOR | 27 | 0.249 | 0.905 | 0.59997 | 0.59997 | 0.69576 | 103 |
| HP_TUBE_FEEDING | 19 | 0.273 | 0.905 | 0.59068 | 0.59068 | 0.69179 | 222 |
| GOBP_REGULATION_OF_PROTEIN_LOCALIZATION | 43 | 0.223 | 0.904 | 0.60675 | 0.60675 | 0.69640 | 229 |
| GOMF_NUCLEOSIDE_TRIPHOSPHATASE_REGULATOR_ACTIVITY | 26 | 0.250 | 0.904 | 0.60021 | 0.60021 | 0.69576 | 255 |
| GOBP_REGULATION_OF_ADAPTIVE_IMMUNE_RESPONSE | 12 | 0.312 | 0.903 | 0.58448 | 0.58448 | 0.68766 | 61 |

| | | | | | | | |
|---|---|---|---|---|---|---|---|
| HP_HYPERTROPHIC_CARDIOMYOPATHY | 29 | 0.243 | 0.903 | 0.60263 | 0.60263 | 0.69580 | 599 |
| GOMF_TRANSFERASE_ACTIVITY_TRANSFERRING_PHOSPHORUS_CONTAINING_GR( | 61 | 0.207 | 0.902 | 0.62190 | 0.62190 | 0.70491 | 188 |
| HP_ABNORMAL_CARDIAC_VENTRICLE_MORPHOLOGY | 42 | 0.223 | 0.902 | 0.61206 | 0.61206 | 0.69929 | 200 |
| GOBP_MESENCHYME_DEVELOPMENT | 13 | 0.303 | 0.901 | 0.58844 | 0.58844 | 0.69004 | 41 |
| HP_ABNORMAL_RENAL_PHYSIOLOGY | 49 | 0.216 | 0.901 | 0.61615 | 0.61615 | 0.70166 | 535 |
| GOBP_EPITHELIAL_CELL_DIFFERENTIATION | 30 | 0.240 | 0.900 | 0.60562 | 0.60562 | 0.69640 | 118 |
| GOCC_TRANSCRIPTION_REGULATOR_COMPLEX | 20 | 0.268 | 0.900 | 0.60030 | 0.60030 | 0.69576 | 267 |
| HP_ABDOMINAL_PAIN | 20 | 0.268 | 0.899 | 0.60071 | 0.60071 | 0.69576 | 165 |
| HP_ABNORMALITY_OF_MALAR_BONES | 11 | 0.317 | 0.898 | 0.59482 | 0.59482 | 0.69350 | 323 |
| GOMF_CALMODULIN_BINDING | 10 | 0.327 | 0.897 | 0.59716 | 0.59716 | 0.69570 | 130 |
| HP_ABNORMALITY_OF_RETINAL_PIGMENTATION | 22 | 0.259 | 0.896 | 0.60315 | 0.60315 | 0.69588 | 232 |
| GOCC_METHYLTRANSFERASE_COMPLEX | 12 | 0.309 | 0.895 | 0.59345 | 0.59345 | 0.69271 | 8 |
| HP_ATTENTION_DEFICIT_HYPERACTIVITY_DISORDER | 21 | 0.263 | 0.895 | 0.60801 | 0.60801 | 0.69733 | 122 |
| GOBP_GLYCEROPHOSPHOLIPID_METABOLIC_PROCESS | 11 | 0.315 | 0.893 | 0.60163 | 0.60163 | 0.69576 | 141 |
| GOBP_NEGATIVE_REGULATION_OF_BINDING | 14 | 0.293 | 0.892 | 0.59969 | 0.59969 | 0.69576 | 45 |
| HP_ABNORMALITY_OF_THE_KNEE | 23 | 0.255 | 0.891 | 0.60861 | 0.60861 | 0.69750 | 306 |
| GOBP_REGULATION_OF_PROTEASOMAL_UBIQUITIN_DEPENDENT_PROTEIN_CATAB( | 14 | 0.293 | 0.890 | 0.60198 | 0.60198 | 0.69576 | 176 |
| GOMF_EXONUCLEASE_ACTIVITY_ACTIVE_WITH_EITHER_RIBO_OR_DEOXYRIBONUC | 11 | 0.314 | 0.889 | 0.60666 | 0.60666 | 0.69640 | 642 |
| HP_CLINODACTYLY | 55 | 0.207 | 0.885 | 0.63966 | 0.63966 | 0.71177 | 316 |
| HP_APLASIA_HYPOPLASIA_OF_THE_NAILS | 15 | 0.285 | 0.884 | 0.60616 | 0.60616 | 0.69640 | 198 |
| GOCC_CELL_BODY | 16 | 0.279 | 0.884 | 0.60544 | 0.60544 | 0.69640 | 348 |
| HP_ABNORMALITY_OF_THE_LYMPH_NODES | 14 | 0.290 | 0.883 | 0.61228 | 0.61228 | 0.69929 | 257 |
| HP_ABNORMAL_LYMPHOCYTE_MORPHOLOGY | 14 | 0.290 | 0.882 | 0.61242 | 0.61242 | 0.69929 | 382 |
| HP_SHORT_THUMB | 10 | 0.322 | 0.882 | 0.61525 | 0.61525 | 0.70166 | 382 |
| HP_SPECIFIC_LEARNING_DISABILITY | 21 | 0.259 | 0.881 | 0.62531 | 0.62531 | 0.70722 | 306 |
| GOBP_NEGATIVE_REGULATION_OF_NUCLEOBASE_CONTAINING_COMPOUND_MET | 94 | 0.188 | 0.879 | 0.67026 | 0.67026 | 0.73011 | 170 |
| HP_DEPRESSED_NASAL_BRIDGE | 33 | 0.229 | 0.879 | 0.63553 | 0.63553 | 0.71151 | 206 |
| GOBP_ORGANIC_ACID_BIOSYNTHETIC_PROCESS | 14 | 0.289 | 0.879 | 0.61571 | 0.61571 | 0.70166 | 154 |
| GOBP_CELLULAR_PROTEIN_CATABOLIC_PROCESS | 70 | 0.197 | 0.879 | 0.65970 | 0.65970 | 0.72495 | 176 |
| GOBP_SMALL_MOLECULE_CATABOLIC_PROCESS | 23 | 0.251 | 0.878 | 0.62644 | 0.62644 | 0.70799 | 266 |
| GOBP_PROCESS_UTILIZING_AUTOPHAGIC_MECHANISM | 29 | 0.237 | 0.878 | 0.63795 | 0.63795 | 0.71177 | 14 |
| HP_NEOPLASM_BY_HISTOLOGY | 28 | 0.238 | 0.877 | 0.63754 | 0.63754 | 0.71177 | 257 |
| HP_CEREBELLAR_ATROPHY | 27 | 0.240 | 0.875 | 0.63751 | 0.63751 | 0.71177 | 205 |
| HP_ABNORMAL_UVEA_MORPHOLOGY | 27 | 0.240 | 0.874 | 0.63923 | 0.63923 | 0.71177 | 257 |
| GOBP_PROTEIN_LOCALIZATION_TO_ORGANELLE | 86 | 0.190 | 0.873 | 0.67247 | 0.67247 | 0.73011 | 462 |
| HP_GASTROINTESTINAL_CARCINOMA | 10 | 0.318 | 0.871 | 0.63154 | 0.63154 | 0.70897 | 188 |
| HP_ABNORMALITY_OF_THE_ABDOMINAL_WALL | 28 | 0.236 | 0.869 | 0.64668 | 0.64668 | 0.71624 | 421 |
| GOBP_REGULATION_OF_UBIQUITIN_DEPENDENT_PROTEIN_CATABOLIC_PROCESS | 15 | 0.280 | 0.867 | 0.62828 | 0.62828 | 0.70800 | 176 |
| GOMF_ACYLTRANSFERASE_ACTIVITY | 10 | 0.316 | 0.867 | 0.63632 | 0.63632 | 0.71177 | 256 |
| GOBP_MITOCHONDRIAL_RESPIRATORY_CHAIN_COMPLEX_ASSEMBLY | 14 | 0.285 | 0.866 | 0.63189 | 0.63189 | 0.70897 | 563 |
| HP_DEVIATION_OF_THE_HAND_OR_OF_FINGERS_OF_THE_HAND | 47 | 0.208 | 0.864 | 0.66430 | 0.66430 | 0.72924 | 316 |

| | | | | | | | |
|---|---|---|---|---|---|---|---|
| HP_ABNORMAL_CEREBELLAR_VERMIS_MORPHOLOGY | 20 | 0.257 | 0.863 | 0.64647 | 0.64647 | 0.71624 | 232 |
| HP_INCOORDINATION | 18 | 0.263 | 0.862 | 0.63902 | 0.63902 | 0.71177 | 73 |
| GOBP_REGULATION_OF_PROTEIN_CATABOLIC_PROCESS | 31 | 0.228 | 0.862 | 0.65495 | 0.65495 | 0.72263 | 176 |
| HP_UNUSUAL_INFECTION | 52 | 0.204 | 0.859 | 0.67166 | 0.67166 | 0.73011 | 257 |
| HP_ABNORMAL_AORTIC_VALVE_MORPHOLOGY | 15 | 0.277 | 0.858 | 0.64155 | 0.64155 | 0.71285 | 359 |
| HP_ABNORMAL_ODONTOID_TISSUE_MORPHOLOGY | 12 | 0.296 | 0.857 | 0.64179 | 0.64179 | 0.71285 | 192 |
| HP_INFANTILE_ONSET | 38 | 0.216 | 0.856 | 0.67021 | 0.67021 | 0.73011 | 268 |
| HP_PREMATURELY_AGED_APPEARANCE | 10 | 0.312 | 0.856 | 0.65007 | 0.65007 | 0.71948 | 421 |
| HP_ABNORMALITY_OF_THE_THYROID_GLAND | 23 | 0.245 | 0.855 | 0.65715 | 0.65715 | 0.72266 | 205 |
| HP_GAIT_DISTURBANCE | 58 | 0.198 | 0.855 | 0.68146 | 0.68146 | 0.73377 | 73 |
| HP_ABNORMALITY_OF_THE_ABDOMINAL_ORGANS | 90 | 0.184 | 0.853 | 0.70651 | 0.70651 | 0.74392 | 591 |
| HP_MUSCULAR_HYPOTONIA_OF_THE_TRUNK | 14 | 0.280 | 0.852 | 0.65278 | 0.65278 | 0.72093 | 205 |
| HP_ABNORMALITY_OF_THYROID_PHYSIOLOGY | 16 | 0.269 | 0.852 | 0.64590 | 0.64590 | 0.71624 | 205 |
| GOCC_TRANSFERASE_COMPLEX | 68 | 0.192 | 0.851 | 0.69931 | 0.69931 | 0.74319 | 517 |
| HP_APLASIA_HYPOPLASIA_AFFECTING_BONES_OF_THE_AXIAL_SKELETON | 75 | 0.189 | 0.850 | 0.70351 | 0.70351 | 0.74321 | 323 |
| HP_ABNORMAL_LIP_MORPHOLOGY | 71 | 0.190 | 0.849 | 0.70446 | 0.70446 | 0.74321 | 232 |
| HP_TALIPES_EQUINOVARUS | 23 | 0.242 | 0.847 | 0.66720 | 0.66720 | 0.73011 | 206 |
| HP_TRACHEOESOPHAGEAL_FISTULA | 12 | 0.292 | 0.846 | 0.65619 | 0.65619 | 0.72263 | 421 |
| HP_ABNORMAL_TRACHEOBRONCHIAL_MORPHOLOGY | 19 | 0.255 | 0.845 | 0.66791 | 0.66791 | 0.73011 | 446 |
| HP_WEAKNESS_DUE_TO_UPPER_MOTOR_NEURON_DYSFUNCTION | 33 | 0.220 | 0.844 | 0.67744 | 0.67744 | 0.73149 | 205 |
| GOBP_VESICLE_MEDIATED_TRANSPORT_IN_SYNAPSE | 11 | 0.298 | 0.843 | 0.66464 | 0.66464 | 0.72924 | 66 |
| GOCC_INTRACELLULAR_PROTEIN_CONTAINING_COMPLEX | 74 | 0.187 | 0.842 | 0.71242 | 0.71242 | 0.74651 | 603 |
| HP_PROMINENT_NASAL_BRIDGE | 19 | 0.254 | 0.842 | 0.67108 | 0.67108 | 0.73011 | 257 |
| GOMF_PROTEIN_DOMAIN_SPECIFIC_BINDING | 38 | 0.212 | 0.840 | 0.68873 | 0.68873 | 0.73648 | 287 |
| HP_ABNORMALITY_OF_UPPER_LIP | 53 | 0.199 | 0.840 | 0.70034 | 0.70034 | 0.74321 | 208 |
| HP_IMPAIRMENT_OF_ACTIVITIES_OF_DAILY_LIVING | 11 | 0.297 | 0.840 | 0.66879 | 0.66879 | 0.73011 | 73 |
| HP_MACULE | 26 | 0.232 | 0.839 | 0.67798 | 0.67798 | 0.73155 | 257 |
| HP_PHOTOPHOBIA | 14 | 0.276 | 0.839 | 0.66824 | 0.66824 | 0.73011 | 45 |
| GOMF_DNA_BINDING_TRANSCRIPTION_FACTOR_ACTIVITY | 60 | 0.193 | 0.838 | 0.70673 | 0.70673 | 0.74392 | 308 |
| HP_AUTISTIC_BEHAVIOR | 47 | 0.202 | 0.837 | 0.70000 | 0.70000 | 0.74321 | 103 |
| GOCC_ACTIN_CYTOSKELETON | 14 | 0.275 | 0.837 | 0.67067 | 0.67067 | 0.73011 | 194 |
| GOBP_AMIDE_BIOSYNTHETIC_PROCESS | 45 | 0.204 | 0.837 | 0.69842 | 0.69842 | 0.74275 | 48 |
| HP_ABNORMALITY_OF_TOE | 51 | 0.199 | 0.835 | 0.70379 | 0.70379 | 0.74321 | 215 |
| HP_ABNORMAL_FACIAL_EXPRESSION | 11 | 0.295 | 0.835 | 0.67382 | 0.67382 | 0.73011 | 73 |
| HP_PANCYTOPENIA | 10 | 0.304 | 0.835 | 0.67519 | 0.67519 | 0.73106 | 376 |
| GOBP_CONNECTIVE_TISSUE_DEVELOPMENT | 10 | 0.304 | 0.834 | 0.67564 | 0.67564 | 0.73106 | 52 |
| HP_DYSMETRIA | 12 | 0.288 | 0.834 | 0.67646 | 0.67646 | 0.73144 | 14 |
| GOBP_INTRINSIC_APOPTOTIC_SIGNALING_PATHWAY | 24 | 0.235 | 0.832 | 0.68434 | 0.68434 | 0.73483 | 321 |
| HP_HEADACHE | 17 | 0.259 | 0.832 | 0.67370 | 0.67370 | 0.73011 | 205 |
| HP_ABNORMAL_EYELASH_MORPHOLOGY | 13 | 0.279 | 0.829 | 0.68008 | 0.68008 | 0.73279 | 306 |
| HP_INCREASED_BLOOD_PRESSURE | 21 | 0.243 | 0.828 | 0.69082 | 0.69082 | 0.73720 | 102 |

| | | | | | | | |
|---|---|---|---|---|---|---|---|
| GOCC_LARGE_RIBOSOMAL_SUBUNIT | 11 | 0.292 | 0.826 | 0.68506 | 0.68506 | 0.73510 | 523 |
| HP_DOLICHOCEPHALY | 19 | 0.249 | 0.825 | 0.68956 | 0.68956 | 0.73687 | 257 |
| GOBP_FATTY_ACID_CATABOLIC_PROCESS | 11 | 0.291 | 0.825 | 0.68698 | 0.68698 | 0.73605 | 92 |
| GOBP_MONOCARBOXYLIC_ACID_CATABOLIC_PROCESS | 11 | 0.291 | 0.825 | 0.68698 | 0.68698 | 0.73605 | 92 |
| HP_CRYPTORCHIDISM | 58 | 0.191 | 0.822 | 0.72582 | 0.72582 | 0.75323 | 222 |
| GOBP_CELLULAR_RESPONSE_TO_STEROID_HORMONE_STIMULUS | 17 | 0.256 | 0.822 | 0.68433 | 0.68433 | 0.73483 | 4 |
| HP_IMMUNODEFICIENCY | 14 | 0.270 | 0.821 | 0.69214 | 0.69214 | 0.73810 | 382 |
| GOBP_CELLULAR_AMIDE_METABOLIC_PROCESS | 61 | 0.188 | 0.818 | 0.73776 | 0.73776 | 0.75655 | 12 |
| HP_ABNORMALITY_OF_THE_CERVICAL_SPINE | 23 | 0.234 | 0.817 | 0.70394 | 0.70394 | 0.74321 | 141 |
| HP_JAUNDICE | 11 | 0.289 | 0.817 | 0.69749 | 0.69749 | 0.74242 | 257 |
| GOBP_MONOCARBOXYLIC_ACID_METABOLIC_PROCESS | 23 | 0.233 | 0.816 | 0.70555 | 0.70555 | 0.74370 | 92 |
| GOCC_MEMBRANE_PROTEIN_COMPLEX | 60 | 0.188 | 0.816 | 0.73869 | 0.73869 | 0.75674 | 73 |
| HP_GAIT_ATAXIA | 16 | 0.258 | 0.816 | 0.69072 | 0.69072 | 0.73720 | 73 |
| HP_ABNORMAL_CELLULAR_PHENOTYPE | 51 | 0.194 | 0.816 | 0.73288 | 0.73288 | 0.75504 | 555 |
| HP_MUSCLE_WEAKNESS | 46 | 0.198 | 0.815 | 0.72701 | 0.72701 | 0.75323 | 66 |
| GOCC_NUCLEAR_SPECK | 30 | 0.217 | 0.814 | 0.71890 | 0.71890 | 0.75042 | 273 |
| GOBP_CELL_SUBSTRATE_ADHESION | 14 | 0.268 | 0.814 | 0.70202 | 0.70202 | 0.74321 | 84 |
| GOMF_LYASE_ACTIVITY | 15 | 0.263 | 0.814 | 0.69763 | 0.69763 | 0.74242 | 154 |
| HP_FAILURE_TO_THRIVE | 63 | 0.186 | 0.812 | 0.74994 | 0.74994 | 0.75981 | 257 |
| GOBP_CELLULAR_RESPONSE_TO_INSULIN_STIMULUS | 11 | 0.287 | 0.812 | 0.70266 | 0.70266 | 0.74321 | 92 |
| HP_LEUKOPENIA | 13 | 0.273 | 0.810 | 0.70258 | 0.70258 | 0.74321 | 376 |
| HP_ABNORMALITY_OF_COORDINATION | 62 | 0.186 | 0.810 | 0.75053 | 0.75053 | 0.75981 | 73 |
| HP_HYPOPITUITARISM | 12 | 0.279 | 0.810 | 0.70746 | 0.70746 | 0.74419 | 200 |
| GOMF_NUCLEASE_ACTIVITY | 25 | 0.226 | 0.807 | 0.71476 | 0.71476 | 0.74831 | 616 |
| GOBP_RESPONSE_TO_INORGANIC_SUBSTANCE | 27 | 0.221 | 0.805 | 0.72511 | 0.72511 | 0.75323 | 250 |
| GOBP_MITOCHONDRIAL_TRANSPORT | 17 | 0.250 | 0.804 | 0.70823 | 0.70823 | 0.74423 | 700 |
| HP_REDUCED_CONSCIOUSNESS_CONFUSION | 21 | 0.236 | 0.802 | 0.72296 | 0.72296 | 0.75323 | 76 |
| HP_WIDE_MOUTH | 12 | 0.277 | 0.802 | 0.71878 | 0.71878 | 0.75042 | 141 |
| HP_ABNORMAL_LOCATION_OF_EARS | 51 | 0.190 | 0.800 | 0.75373 | 0.75373 | 0.76083 | 205 |
| HP_EPICANTHUS | 36 | 0.204 | 0.800 | 0.73965 | 0.73965 | 0.75674 | 215 |
| GOBP_REGULATION_OF_CELLULAR_RESPONSE_TO_STRESS | 53 | 0.189 | 0.799 | 0.75344 | 0.75344 | 0.76083 | 349 |
| HP_POSITIONAL_FOOT_DEFORMITY | 30 | 0.213 | 0.798 | 0.73717 | 0.73717 | 0.75644 | 206 |
| HP_ABNORMALITY_OF_THE_FOREARM | 21 | 0.234 | 0.796 | 0.73086 | 0.73086 | 0.75497 | 376 |
| HP_ABNORMAL_UMBILICUS_MORPHOLOGY | 19 | 0.240 | 0.796 | 0.72542 | 0.72542 | 0.75323 | 421 |
| GOBP_ENDOPLASMIC_RETICULUM_TO_GOLGI_VESICLE_MEDIATED_TRANSPORT | 11 | 0.281 | 0.796 | 0.71967 | 0.71967 | 0.75042 | 753 |
| HP_ABNORMALITY_OF_THE_LYMPHATIC_SYSTEM | 40 | 0.198 | 0.795 | 0.74922 | 0.74922 | 0.75981 | 257 |
| HP_OCULAR_ANTERIOR_SEGMENT_DYSGENESIS | 24 | 0.225 | 0.795 | 0.73242 | 0.73242 | 0.75504 | 257 |
| HP_ABNORMAL_SHAPE_OF_THE_FRONTAL_REGION | 31 | 0.210 | 0.793 | 0.74356 | 0.74356 | 0.75848 | 312 |
| HP_DELAYED_MYELINATION | 10 | 0.289 | 0.792 | 0.72601 | 0.72601 | 0.75323 | 141 |
| GOCC_RNA_POLYMERASE_II_TRANSCRIPTION_REGULATOR_COMPLEX | 14 | 0.260 | 0.792 | 0.72606 | 0.72606 | 0.75323 | 267 |
| HP_APLASIA_HYPOPLASIA_INVOLVING_THE_SKELETON | 87 | 0.172 | 0.791 | 0.79095 | 0.79095 | 0.77649 | 323 |

| | | | | | | | |
|---|---|---|---|---|---|---|---|
| GOBP_REGULATED_EXOCYTOSIS | 10 | 0.288 | 0.789 | 0.72945 | 0.72945 | 0.75435 | 66 |
| HP_ABNORMAL_SCALP_MORPHOLOGY | 16 | 0.249 | 0.789 | 0.72723 | 0.72723 | 0.75323 | 257 |
| HP_SMALL_FOR_GESTATIONAL_AGE | 18 | 0.241 | 0.789 | 0.73208 | 0.73208 | 0.75504 | 306 |
| HP_ABNORMAL_RIB_CAGE_MORPHOLOGY | 20 | 0.234 | 0.787 | 0.73990 | 0.73990 | 0.75674 | 323 |
| GOBP_REGULATION_OF_SMALL_GTPASE_MEDIATED_SIGNAL_TRANSDUCTION | 13 | 0.264 | 0.786 | 0.73410 | 0.73410 | 0.75529 | 216 |
| GOBP_PROTEOLYSIS | 116 | 0.163 | 0.786 | 0.81304 | 0.81304 | 0.77808 | 95 |
| GOCC_NUCLEAR_PROTEIN_CONTAINING_COMPLEX | 116 | 0.163 | 0.786 | 0.81337 | 0.81337 | 0.77808 | 659 |
| HP_ABNORMALITY_OF_THE_CEREBROSPINAL_FLUID | 40 | 0.196 | 0.784 | 0.76429 | 0.76429 | 0.76502 | 388 |
| HP_AGGRESSIVE_BEHAVIOR | 15 | 0.253 | 0.784 | 0.73545 | 0.73545 | 0.75567 | 196 |
| GOBP_POSITIVE_REGULATION_OF_CATALYTIC_ACTIVITY | 81 | 0.172 | 0.783 | 0.79680 | 0.79680 | 0.77649 | 358 |
| GOMF_PEPTIDASE_ACTIVITY | 38 | 0.198 | 0.783 | 0.76392 | 0.76392 | 0.76502 | 88 |
| GOBP_MITOCHONDRION_ORGANIZATION | 49 | 0.186 | 0.778 | 0.77645 | 0.77645 | 0.77031 | 674 |
| GOBP_NEGATIVE_REGULATION_OF_TRANSCRIPTION_BY_RNA_POLYMERASE_II | 50 | 0.186 | 0.778 | 0.77628 | 0.77628 | 0.77031 | 138 |
| HP_ABNORMAL_MYELINATION | 30 | 0.208 | 0.777 | 0.76374 | 0.76374 | 0.76502 | 205 |
| HP_ABNORMALITY_OF_THE_LENS | 49 | 0.186 | 0.777 | 0.77767 | 0.77767 | 0.77031 | 215 |
| HP_ABNORMALITY_OF_THE_CALF | 22 | 0.225 | 0.777 | 0.75158 | 0.75158 | 0.76015 | 323 |
| HP_ABNORMAL_HAND_MORPHOLOGY | 23 | 0.222 | 0.775 | 0.75556 | 0.75556 | 0.76170 | 200 |
| GOBP_CELLULAR_RESPONSE_TO_CHEMICAL_STRESS | 22 | 0.224 | 0.775 | 0.75347 | 0.75347 | 0.76083 | 139 |
| HP_CLUBBING | 13 | 0.260 | 0.773 | 0.75065 | 0.75065 | 0.75981 | 359 |
| HP_DELAYED_GROSS_MOTOR_DEVELOPMENT | 23 | 0.221 | 0.773 | 0.75771 | 0.75771 | 0.76237 | 141 |
| HP_ABNORMALITY_OF_BLOOD_CIRCULATION | 17 | 0.240 | 0.771 | 0.74752 | 0.74752 | 0.75981 | 129 |
| HP_UPPER_MOTOR_NEURON_DYSFUNCTION | 80 | 0.169 | 0.769 | 0.81398 | 0.81398 | 0.77808 | 205 |
| HP_BABINSKI_SIGN | 12 | 0.265 | 0.769 | 0.75727 | 0.75727 | 0.76237 | 141 |
| HP_ATROPHY_DEGENERATION_AFFECTING_THE_CENTRAL_NERVOUS_SYSTEM | 64 | 0.175 | 0.769 | 0.80332 | 0.80332 | 0.77776 | 73 |
| GOMF_CYSTEINE_TYPE_PEPTIDASE_ACTIVITY | 12 | 0.264 | 0.766 | 0.76080 | 0.76080 | 0.76427 | 14 |
| HP_REGIONAL_ABNORMALITY_OF_SKIN | 33 | 0.199 | 0.764 | 0.77853 | 0.77853 | 0.77031 | 222 |
| HP_HYPERTONIA | 82 | 0.167 | 0.762 | 0.82249 | 0.82249 | 0.77808 | 232 |
| HP_ABNORMALITY_OF_EARLOBE | 10 | 0.278 | 0.761 | 0.76173 | 0.76173 | 0.76443 | 316 |
| GOBP_PROTEIN_CATABOLIC_PROCESS | 78 | 0.168 | 0.761 | 0.81793 | 0.81793 | 0.77808 | 176 |
| GOMF_TRANSCRIPTION_REGULATOR_ACTIVITY | 97 | 0.162 | 0.761 | 0.83184 | 0.83184 | 0.77840 | 137 |
| GOBP_CELLULAR_RESPONSE_TO_PEPTIDE_HORMONE_STIMULUS | 18 | 0.232 | 0.760 | 0.76938 | 0.76938 | 0.76844 | 312 |
| HP_ABNORMAL_STOMACH_MORPHOLOGY | 23 | 0.217 | 0.759 | 0.77353 | 0.77353 | 0.76979 | 222 |
| HP_CHOLESTASIS | 13 | 0.255 | 0.758 | 0.76881 | 0.76881 | 0.76844 | 411 |
| HP_APNEA | 18 | 0.231 | 0.757 | 0.77229 | 0.77229 | 0.76975 | 188 |
| HP_MIDFACE_RETRUSION | 13 | 0.255 | 0.757 | 0.76968 | 0.76968 | 0.76844 | 124 |
| HP_ABNORMAL_VERTEBRAL_MORPHOLOGY | 24 | 0.214 | 0.757 | 0.77677 | 0.77677 | 0.77031 | 375 |
| HP_ABNORMAL_HAIR_MORPHOLOGY | 67 | 0.171 | 0.756 | 0.81722 | 0.81722 | 0.77808 | 222 |
| HP_ABNORMAL_FACIAL_SHAPE | 79 | 0.166 | 0.755 | 0.82719 | 0.82719 | 0.77808 | 222 |
| HP_SPASTICITY | 69 | 0.170 | 0.755 | 0.82599 | 0.82599 | 0.77808 | 205 |
| GOBP_INTRACELLULAR_RECEPTOR_SIGNALING_PATHWAY | 18 | 0.230 | 0.754 | 0.77520 | 0.77520 | 0.77031 | 4 |
| HP_ABNORMAL_JOINT_MORPHOLOGY | 67 | 0.170 | 0.752 | 0.82141 | 0.82141 | 0.77808 | 215 |

| | | | | | | | |
|---|---|---|---|---|---|---|---|
| GOCC_CILIARY_BASAL_BODY | 11 | 0.265 | 0.751 | 0.77115 | 0.77115 | 0.76940 | 198 |
| HP_ABNORMAL_LEUKOCYTE_COUNT | 31 | 0.198 | 0.750 | 0.79353 | 0.79353 | 0.77649 | 382 |
| GOBP_POSITIVE_REGULATION_OF_APOPTOTIC_SIGNALING_PATHWAY | 10 | 0.273 | 0.749 | 0.77250 | 0.77250 | 0.76975 | 245 |
| HP_ABNORMALITY_OF_URINE_HOMEOSTASIS | 42 | 0.185 | 0.749 | 0.81057 | 0.81057 | 0.77808 | 102 |
| HP_ABNORMALITY_OF_THE_PINNA | 53 | 0.177 | 0.748 | 0.82030 | 0.82030 | 0.77808 | 198 |
| GOBP_ANION_TRANSPORT | 12 | 0.258 | 0.746 | 0.78519 | 0.78519 | 0.77443 | 65 |
| HP_PRENATAL_MOVEMENT_ABNORMALITY | 13 | 0.251 | 0.745 | 0.78173 | 0.78173 | 0.77200 | 161 |
| HP_ANEMIA_OF_INADEQUATE_PRODUCTION | 20 | 0.221 | 0.743 | 0.78840 | 0.78840 | 0.77633 | 622 |
| GOBP_PRODUCTION_OF_MOLECULAR_MEDIATOR_OF_IMMUNE_RESPONSE | 16 | 0.235 | 0.743 | 0.78066 | 0.78066 | 0.77193 | 125 |
| HP_ABNORMAL_RECTUM_MORPHOLOGY | 11 | 0.261 | 0.740 | 0.78254 | 0.78254 | 0.77231 | 580 |
| GOBP_PEPTIDE_METABOLIC_PROCESS | 51 | 0.176 | 0.739 | 0.82548 | 0.82548 | 0.77808 | 3 |
| GOBP_GENERATION_OF_PRECURSOR_METABOLITES_AND_ENERGY | 31 | 0.196 | 0.739 | 0.80294 | 0.80294 | 0.77776 | 567 |
| GOBP_REGULATION_OF_LEUKOCYTE_MEDIATED_IMMUNITY | 12 | 0.255 | 0.739 | 0.79268 | 0.79268 | 0.77649 | 61 |
| HP_VOMITING | 19 | 0.222 | 0.737 | 0.79630 | 0.79630 | 0.77649 | 50 |
| HP_SMALL_NAIL | 11 | 0.260 | 0.737 | 0.78654 | 0.78654 | 0.77527 | 421 |
| GOCC_MITOCHONDRIAL_ENVELOPE | 58 | 0.171 | 0.736 | 0.83799 | 0.83799 | 0.78079 | 663 |
| GOBP_NEGATIVE_REGULATION_OF_CATABOLIC_PROCESS | 26 | 0.204 | 0.736 | 0.80341 | 0.80341 | 0.77776 | 80 |
| HP_PUBERTY_AND_GONADAL_DISORDERS | 45 | 0.179 | 0.735 | 0.82795 | 0.82795 | 0.77808 | 46 |
| HP_ABNORMAL_VENTRICULAR_SEPTUM_MORPHOLOGY | 35 | 0.189 | 0.735 | 0.81374 | 0.81374 | 0.77808 | 200 |
| GOBP_POSITIVE_REGULATION_OF_CELL_PROJECTION_ORGANIZATION | 13 | 0.247 | 0.734 | 0.79422 | 0.79422 | 0.77649 | 6 |
| GOCC_GOLGI_MEMBRANE | 39 | 0.184 | 0.734 | 0.82015 | 0.82015 | 0.77808 | 113 |
| HP_HERNIA | 36 | 0.188 | 0.734 | 0.81730 | 0.81730 | 0.77808 | 529 |
| HP_ABNORMALITY_OF_THE_SPLEEN | 29 | 0.198 | 0.733 | 0.81402 | 0.81402 | 0.77808 | 257 |
| HP_ABNORMALITY_OF_THE_HYPOTHALAMUS_PITUITARY_AXIS | 28 | 0.199 | 0.733 | 0.80904 | 0.80904 | 0.77808 | 257 |
| HP_JOINT_DISLOCATION | 33 | 0.191 | 0.733 | 0.81623 | 0.81623 | 0.77808 | 312 |
| HP_SLEEP_DISTURBANCE | 32 | 0.192 | 0.732 | 0.81502 | 0.81502 | 0.77808 | 141 |
| GOBP_PROTEIN_ACETYLATION | 15 | 0.236 | 0.732 | 0.79680 | 0.79680 | 0.77649 | 800 |
| GOMF_MAGNESIUM_ION_BINDING | 22 | 0.212 | 0.732 | 0.80406 | 0.80406 | 0.77776 | 633 |
| HP_ABNORMALITY_OF_THE_ULNA | 19 | 0.220 | 0.732 | 0.80086 | 0.80086 | 0.77776 | 257 |
| HP_WIDE_NASAL_BRIDGE | 34 | 0.189 | 0.732 | 0.81602 | 0.81602 | 0.77808 | 208 |
| HP_LANGUAGE_IMPAIRMENT | 56 | 0.171 | 0.731 | 0.84121 | 0.84121 | 0.78140 | 177 |
| HP_VISUAL_IMPAIRMENT | 80 | 0.161 | 0.731 | 0.85628 | 0.85628 | 0.78545 | 200 |
| HP_LOWER_LIMB_SPASTICITY | 15 | 0.235 | 0.729 | 0.80023 | 0.80023 | 0.77776 | 268 |
| HP_ABNORMAL_CORNEA_MORPHOLOGY | 51 | 0.173 | 0.728 | 0.83869 | 0.83869 | 0.78096 | 232 |
| GOBP_RESPONSE_TO_METAL_ION | 16 | 0.230 | 0.727 | 0.79884 | 0.79884 | 0.77750 | 223 |
| GOBP_ORGANONITROGEN_COMPOUND_CATABOLIC_PROCESS | 99 | 0.154 | 0.726 | 0.87166 | 0.87166 | 0.79122 | 227 |
| GOBP_RESPONSE_TO_XENOBIOTIC_STIMULUS | 22 | 0.210 | 0.725 | 0.81066 | 0.81066 | 0.77808 | 335 |
| GOBP_MACROAUTOPHAGY | 16 | 0.229 | 0.724 | 0.80166 | 0.80166 | 0.77776 | 14 |
| HP_ABNORMAL_MORPHOLOGY_OF_THE_GREAT_VESSELS | 42 | 0.179 | 0.723 | 0.83704 | 0.83704 | 0.78037 | 257 |
| HP_ABNORMALITY_OF_THE_LARYNX | 11 | 0.255 | 0.721 | 0.80089 | 0.80089 | 0.77776 | 332 |
| HP_PATENT_DUCTUS_ARTERIOSUS | 24 | 0.202 | 0.715 | 0.82379 | 0.82379 | 0.77808 | 257 |

| | | | | | | | |
|---|---|---|---|---|---|---|---|
| GOBP_REGULATION_OF_PROTEASOMAL_PROTEIN_CATABOLIC_PROCESS | 19 | 0.215 | 0.715 | 0.81837 | 0.81837 | 0.77808 | 176 |
| GOMF_ZINC_ION_BINDING | 55 | 0.167 | 0.714 | 0.85668 | 0.85668 | 0.78545 | 222 |
| GOBP_AGING | 15 | 0.230 | 0.713 | 0.81664 | 0.81664 | 0.77808 | 285 |
| HP_FEEDING_DIFFICULTIES_IN_INFANCY | 35 | 0.183 | 0.713 | 0.83619 | 0.83619 | 0.78004 | 141 |
| HP_ABNORMAL_URINE_PROTEIN_LEVEL | 17 | 0.222 | 0.713 | 0.81183 | 0.81183 | 0.77808 | 529 |
| HP_MYOPIA | 28 | 0.193 | 0.712 | 0.83098 | 0.83098 | 0.77840 | 200 |
| HP_ABNORMALITY_OF_BONE_MARROW_CELL_MORPHOLOGY | 19 | 0.215 | 0.712 | 0.82127 | 0.82127 | 0.77808 | 376 |
| HP_ABNORMAL_SOCIAL_BEHAVIOR | 16 | 0.225 | 0.710 | 0.81858 | 0.81858 | 0.77808 | 14 |
| GOCC_MITOCHONDRION | 124 | 0.146 | 0.710 | 0.90177 | 0.90177 | 0.80302 | 667 |
| HP_HIGH_PALATE | 49 | 0.170 | 0.710 | 0.85466 | 0.85466 | 0.78452 | 312 |
| HP_ABNORMAL_HEART_VALVE_PHYSIOLOGY | 24 | 0.200 | 0.709 | 0.83085 | 0.83085 | 0.77840 | 232 |
| HP_PNEUMONIA | 11 | 0.250 | 0.708 | 0.81479 | 0.81479 | 0.77808 | 376 |
| HP_PEDIATRIC_ONSET | 45 | 0.172 | 0.707 | 0.85741 | 0.85741 | 0.78565 | 88 |
| GOBP_REGULATION_OF_TELOMERE_MAINTENANCE | 10 | 0.257 | 0.705 | 0.82302 | 0.82302 | 0.77808 | 45 |
| HP_ABNORMALITY_OF_THE_ZYGOMATIC_BONE | 14 | 0.232 | 0.705 | 0.82482 | 0.82482 | 0.77808 | 421 |
| HP_ABNORMAL_RENAL_CORTEX_MORPHOLOGY | 14 | 0.232 | 0.705 | 0.82510 | 0.82510 | 0.77808 | 529 |
| HP_PERIPHERAL_NEUROPATHY | 33 | 0.183 | 0.703 | 0.84677 | 0.84677 | 0.78281 | 73 |
| HP_ABNORMAL_NEPHRON_MORPHOLOGY | 17 | 0.219 | 0.703 | 0.82343 | 0.82343 | 0.77808 | 529 |
| GOBP_PEPTIDYL_LYSINE_ACETYLATION | 13 | 0.236 | 0.701 | 0.82994 | 0.82994 | 0.77840 | 800 |
| HP_OPTIC_ATROPHY | 50 | 0.167 | 0.701 | 0.86177 | 0.86177 | 0.78642 | 196 |
| GOBP_NUCLEIC_ACID_PHOSPHODIESTER_BOND_HYDROLYSIS | 27 | 0.192 | 0.701 | 0.84694 | 0.84694 | 0.78281 | 616 |
| GOBP_POSITIVE_REGULATION_OF_PROTEOLYSIS_INVOLVED_IN_CELLULAR_PROTEII | 12 | 0.242 | 0.700 | 0.83206 | 0.83206 | 0.77840 | 176 |
| HP_ABNORMAL_TESTIS_MORPHOLOGY | 70 | 0.157 | 0.699 | 0.88161 | 0.88161 | 0.79652 | 222 |
| HP_COLOBOMA | 13 | 0.235 | 0.699 | 0.83241 | 0.83241 | 0.77840 | 126 |
| GOBP_REGULATION_OF_CELL_PROJECTION_ASSEMBLY | 15 | 0.225 | 0.698 | 0.83105 | 0.83105 | 0.77840 | 6 |
| HP_ABNORMALITY_OF_MOUTH_SIZE | 32 | 0.183 | 0.697 | 0.85156 | 0.85156 | 0.78400 | 192 |
| GOBP_POSITIVE_REGULATION_OF_RESPONSE_TO_DNA_DAMAGE_STIMULUS | 16 | 0.220 | 0.696 | 0.83479 | 0.83479 | 0.77968 | 817 |
| GOBP_REGULATION_OF_ESTABLISHMENT_OF_PROTEIN_LOCALIZATION | 24 | 0.197 | 0.696 | 0.84137 | 0.84137 | 0.78140 | 202 |
| GOBP_ENDOMEMBRANE_SYSTEM_ORGANIZATION | 39 | 0.174 | 0.695 | 0.86231 | 0.86231 | 0.78642 | 206 |
| HP_EARLY_ONSET_OF_SEXUAL_MATURATION | 11 | 0.245 | 0.693 | 0.82929 | 0.82929 | 0.77840 | 23 |
| HP_ABNORMALITY_OF_THE_CURVATURE_OF_THE_VERTEBRAL_COLUMN | 69 | 0.156 | 0.693 | 0.88473 | 0.88473 | 0.79664 | 382 |
| HP_ABNORMALITY_OF_THE_LIVER | 72 | 0.155 | 0.693 | 0.89074 | 0.89074 | 0.79918 | 590 |
| HP_ABNORMAL_BRAINSTEM_MORPHOLOGY | 19 | 0.208 | 0.690 | 0.84292 | 0.84292 | 0.78162 | 97 |
| HP_APLASIA_HYPOPLASIA_OF_THE_THUMB | 12 | 0.238 | 0.689 | 0.84117 | 0.84117 | 0.78140 | 382 |
| HP_RECURRENT_RESPIRATORY_INFECTIONS | 29 | 0.185 | 0.687 | 0.85977 | 0.85977 | 0.78595 | 254 |
| HP_ABNORMAL_FEMORAL_NECK_HEAD_MORPHOLOGY | 14 | 0.226 | 0.686 | 0.84385 | 0.84385 | 0.78201 | 323 |
| GOBP_CELLULAR_RESPONSE_TO_HORMONE_STIMULUS | 36 | 0.175 | 0.686 | 0.86301 | 0.86301 | 0.78658 | 138 |
| GOBP_NUCLEUS_ORGANIZATION | 20 | 0.204 | 0.684 | 0.85010 | 0.85010 | 0.78312 | 97 |
| HP_ABNORMAL_PYRAMIDAL_SIGN | 19 | 0.206 | 0.683 | 0.84926 | 0.84926 | 0.78281 | 268 |
| HP_HYPERTELORISM | 54 | 0.161 | 0.683 | 0.88765 | 0.88765 | 0.79733 | 215 |
| HP_NEOPLASM_OF_THE_PANCREAS | 12 | 0.235 | 0.682 | 0.84910 | 0.84910 | 0.78281 | 312 |

| | | | | | | | |
|---|---|---|---|---|---|---|---|
| HP_ABNORMAL_SKIN_MORPHOLOGY_OF_THE_PALM | 19 | 0.205 | 0.680 | 0.85271 | 0.85271 | 0.78426 | 179 |
| HP_ABNORMAL_THROMBOCYTE_MORPHOLOGY | 29 | 0.183 | 0.679 | 0.86628 | 0.86628 | 0.78819 | 376 |
| GOCC_OUTER_MEMBRANE | 10 | 0.247 | 0.678 | 0.84918 | 0.84918 | 0.78281 | 25 |
| HP_RESPIRATORY_TRACT_INFECTION | 41 | 0.168 | 0.675 | 0.87864 | 0.87864 | 0.79616 | 222 |
| HP_ABNORMAL_CAROTID_ARTERY_MORPHOLOGY | 11 | 0.238 | 0.674 | 0.84911 | 0.84911 | 0.78281 | 421 |
| HP_ABNORMALITY_OF_THE_OPTIC_NERVE | 67 | 0.152 | 0.673 | 0.90051 | 0.90051 | 0.80264 | 200 |
| HP_RETINAL_DYSTROPHY | 18 | 0.206 | 0.673 | 0.85841 | 0.85841 | 0.78595 | 45 |
| HP_ABSENT_SPEECH | 19 | 0.203 | 0.673 | 0.86043 | 0.86043 | 0.78610 | 141 |
| HP_ABNORMAL_ABDOMEN_MORPHOLOGY | 42 | 0.166 | 0.672 | 0.88154 | 0.88154 | 0.79652 | 590 |
| GOMF_ENDONUCLEASE_ACTIVITY | 16 | 0.212 | 0.672 | 0.85932 | 0.85932 | 0.78595 | 616 |
| HP_APLASIA_HYPOPLASIA_OF_THE_PHALANGES_OF_THE_HAND | 14 | 0.220 | 0.669 | 0.85888 | 0.85888 | 0.78595 | 200 |
| HP_DEPRESSION | 20 | 0.199 | 0.668 | 0.86700 | 0.86700 | 0.78837 | 50 |
| HP_SECONDARY_MICROCEPHALY | 12 | 0.230 | 0.666 | 0.86350 | 0.86350 | 0.78658 | 205 |
| HP_LONG_FINGERS | 13 | 0.223 | 0.664 | 0.86523 | 0.86523 | 0.78770 | 103 |
| HP_HYPODONTIA | 11 | 0.234 | 0.663 | 0.86198 | 0.86198 | 0.78642 | 22 |
| GOMF_HYDROLASE_ACTIVITY_ACTING_ON_ESTER_BONDS | 60 | 0.152 | 0.661 | 0.91085 | 0.91085 | 0.80764 | 286 |
| HP_PROGRESSIVE | 23 | 0.189 | 0.660 | 0.87946 | 0.87946 | 0.79643 | 66 |
| HP_ABNORMAL_VASCULAR_MORPHOLOGY | 49 | 0.158 | 0.659 | 0.90258 | 0.90258 | 0.80309 | 232 |
| GOMF_DNA_BINDING_TRANSCRIPTION_FACTOR_BINDING | 32 | 0.173 | 0.658 | 0.88655 | 0.88655 | 0.79727 | 32 |
| GOBP_RESPONSE_TO_STEROID_HORMONE | 23 | 0.188 | 0.658 | 0.88107 | 0.88107 | 0.79652 | 4 |
| HP_JOINT_HYPERFLEXIBILITY | 17 | 0.204 | 0.656 | 0.87586 | 0.87586 | 0.79410 | 421 |
| GOCC_ORGANELLE_ENVELOPE_LUMEN | 13 | 0.221 | 0.655 | 0.87337 | 0.87337 | 0.79231 | 624 |
| HP_ABNORMALITY_OF_THE_SYNOVIA | 26 | 0.180 | 0.652 | 0.88474 | 0.88474 | 0.79664 | 273 |
| HP_HIGHLY_ARCHED_EYEBROW | 16 | 0.205 | 0.647 | 0.88483 | 0.88483 | 0.79664 | 141 |
| HP_HIP_DISLOCATION | 23 | 0.185 | 0.645 | 0.89233 | 0.89233 | 0.80014 | 312 |
| GOBP_CELL_AGING | 12 | 0.222 | 0.643 | 0.88304 | 0.88304 | 0.79664 | 245 |
| HP_ABNORMAL_PHALANGEAL_JOINT_MORPHOLOGY_OF_THE_HAND | 15 | 0.207 | 0.642 | 0.88713 | 0.88713 | 0.79732 | 529 |
| GOCC_RIBONUCLEOPROTEIN_COMPLEX | 52 | 0.151 | 0.639 | 0.92045 | 0.92045 | 0.81173 | 739 |
| HP_HYPOGONADISM | 33 | 0.166 | 0.637 | 0.90288 | 0.90288 | 0.80309 | 46 |
| HP_GASTROESOPHAGEAL_REFLUX | 30 | 0.170 | 0.637 | 0.89943 | 0.89943 | 0.80264 | 222 |
| HP_SKIN_RASH | 10 | 0.230 | 0.631 | 0.88924 | 0.88924 | 0.79829 | 222 |
| HP_NARROW_MOUTH | 20 | 0.188 | 0.630 | 0.89489 | 0.89489 | 0.80150 | 365 |
| HP_STAGE_5_CHRONIC_KIDNEY_DISEASE | 14 | 0.207 | 0.630 | 0.89480 | 0.89480 | 0.80150 | 529 |
| HP_ABNORMALITY_IRIS_MORPHOLOGY | 22 | 0.182 | 0.629 | 0.90419 | 0.90419 | 0.80379 | 421 |
| GOBP_CELLULAR_RESPONSE_TO_IONIZING_RADIATION | 14 | 0.204 | 0.621 | 0.90053 | 0.90053 | 0.80264 | 257 |
| HP_DEEPLY_SET_EYE | 23 | 0.177 | 0.619 | 0.91445 | 0.91445 | 0.80875 | 198 |
| HP_SYNDACTYLY | 33 | 0.160 | 0.614 | 0.92153 | 0.92153 | 0.81195 | 222 |
| GOBP_ALCOHOL_METABOLIC_PROCESS | 12 | 0.210 | 0.610 | 0.91067 | 0.91067 | 0.80764 | 415 |
| HP_CHRONIC_KIDNEY_DISEASE | 16 | 0.193 | 0.609 | 0.91415 | 0.91415 | 0.80875 | 529 |
| HP_ABNORMAL_MYOCARDIUM_MORPHOLOGY | 40 | 0.152 | 0.609 | 0.92753 | 0.92753 | 0.81427 | 583 |
| HP_SINGLE_TRANSVERSE_PALMAR_CREASE | 12 | 0.210 | 0.607 | 0.91243 | 0.91243 | 0.80788 | 179 |

| | | | | | | | |
|---|---|---|---|---|---|---|---|
| GOBP_RESPONSE_TO_IONIZING_RADIATION | 20 | 0.181 | 0.607 | 0.91151 | 0.91151 | 0.80764 | 257 |
| HP_ABNORMAL_FINGER_PHALANX_MORPHOLOGY | 55 | 0.142 | 0.606 | 0.94524 | 0.94524 | 0.81702 | 323 |
| HP_ABNORMALITY_OF_PELVIC_GIRDLE_BONE_MORPHOLOGY | 50 | 0.143 | 0.599 | 0.94628 | 0.94628 | 0.81702 | 323 |
| HP_ABNORMAL_HAIR_QUANTITY | 42 | 0.148 | 0.599 | 0.93785 | 0.93785 | 0.81547 | 222 |
| GOBP_ORGANELLE_FUSION | 10 | 0.217 | 0.596 | 0.92033 | 0.92033 | 0.81173 | 183 |
| GOBP_CELLULAR_RESPONSE_TO_OXYGEN_LEVELS | 11 | 0.209 | 0.591 | 0.92249 | 0.92249 | 0.81195 | 23 |
| GOCC_SITE_OF_DNA_DAMAGE | 17 | 0.183 | 0.587 | 0.93010 | 0.93010 | 0.81509 | 599 |
| HP_ABNORMAL_SIZE_OF_THE_PALPEBRAL_FISSURES | 29 | 0.158 | 0.587 | 0.93679 | 0.93679 | 0.81547 | 312 |
| HP_DYSTONIA | 38 | 0.148 | 0.586 | 0.94345 | 0.94345 | 0.81644 | 23 |
| HP_TETRALOGY_OF_FALLOT | 19 | 0.177 | 0.586 | 0.93077 | 0.93077 | 0.81509 | 375 |
| HP_LIMITATION_OF_JOINT_MOBILITY | 27 | 0.161 | 0.585 | 0.93757 | 0.93757 | 0.81547 | 192 |
| HP_ABNORMAL_LOCALIZATION_OF_KIDNEY | 20 | 0.174 | 0.585 | 0.92745 | 0.92745 | 0.81427 | 372 |
| HP_ABNORMALITY_OF_THE_BILIARY_SYSTEM | 25 | 0.162 | 0.581 | 0.93811 | 0.93811 | 0.81547 | 553 |
| HP_CONGENITAL_MALFORMATION_OF_THE_GREAT_ARTERIES | 28 | 0.158 | 0.580 | 0.94070 | 0.94070 | 0.81556 | 376 |
| HP_NARROW_FACE | 14 | 0.191 | 0.580 | 0.93273 | 0.93273 | 0.81509 | 222 |
| HP_SMALL_FACE | 14 | 0.191 | 0.580 | 0.93273 | 0.93273 | 0.81509 | 222 |
| HP_HYPERTRICHOSIS | 16 | 0.183 | 0.578 | 0.93657 | 0.93657 | 0.81547 | 222 |
| HP_BLINDNESS | 22 | 0.167 | 0.578 | 0.93837 | 0.93837 | 0.81547 | 555 |
| HP_ABNORMALITY_OF_THE_PALM | 28 | 0.156 | 0.574 | 0.94370 | 0.94370 | 0.81644 | 206 |
| HP_ABNORMALITY_OF_THE_URINARY_SYSTEM_PHYSIOLOGY | 74 | 0.127 | 0.573 | 0.96794 | 0.96794 | 0.82429 | 535 |
| HP_ABNORMALITY_OF_DENTAL_STRUCTURE | 17 | 0.178 | 0.572 | 0.94030 | 0.94030 | 0.81556 | 192 |
| HP_HEMIPLEGIA_HEMIPARESIS | 13 | 0.192 | 0.571 | 0.93320 | 0.93320 | 0.81509 | 277 |
| GOBP_CELLULAR_MACROMOLECULE_LOCALIZATION | 131 | 0.116 | 0.569 | 0.98186 | 0.98186 | 0.83044 | 231 |
| HP_DISTAL_AMYOTROPHY | 10 | 0.207 | 0.568 | 0.94111 | 0.94111 | 0.81556 | 829 |
| GOBP_PROTEIN_TARGETING | 27 | 0.154 | 0.562 | 0.95166 | 0.95166 | 0.81966 | 650 |
| GOBP_INTRACELLULAR_PROTEIN_TRANSPORT | 65 | 0.128 | 0.562 | 0.97198 | 0.97198 | 0.82524 | 6 |
| HP_LIPODYSTROPHY | 10 | 0.204 | 0.558 | 0.94649 | 0.94649 | 0.81702 | 630 |
| HP_ABNORMALITY_OF_THE_PHILTRUM | 38 | 0.139 | 0.552 | 0.95934 | 0.95934 | 0.82188 | 208 |
| GOBP_REGULATION_OF_PRODUCTION_OF_MOLECULAR_MEDIATOR_OF_IMMUNE_ | 10 | 0.199 | 0.546 | 0.95247 | 0.95247 | 0.81990 | 48 |
| HP_ABNORMAL_URINE_METABOLITE_LEVEL | 36 | 0.139 | 0.545 | 0.96288 | 0.96288 | 0.82307 | 99 |
| GOBP_CARBOHYDRATE_BIOSYNTHETIC_PROCESS | 12 | 0.188 | 0.543 | 0.94916 | 0.94916 | 0.81842 | 315 |
| HP_ABNORMAL_RETINAL_VASCULAR_MORPHOLOGY | 16 | 0.171 | 0.541 | 0.95546 | 0.95546 | 0.82143 | 108 |
| HP_BONE_MARROW_HYPOCELLULARITY | 10 | 0.197 | 0.541 | 0.95605 | 0.95605 | 0.82143 | 376 |
| GOBP_ENERGY_DERIVATION_BY_OXIDATION_OF_ORGANIC_COMPOUNDS | 21 | 0.158 | 0.537 | 0.95873 | 0.95873 | 0.82188 | 555 |
| HP_ABNORMAL_CONNECTION_OF_THE_CARDIAC_SEGMENTS | 20 | 0.159 | 0.535 | 0.95493 | 0.95493 | 0.82143 | 375 |
| HP_ABNORMAL_FEMORAL_NECK_MORPHOLOGY | 11 | 0.188 | 0.534 | 0.95947 | 0.95947 | 0.82188 | 306 |
| GOBP_ORGANIC_ACID_METABOLIC_PROCESS | 37 | 0.135 | 0.533 | 0.96816 | 0.96816 | 0.82429 | 204 |
| GOMF_CATALYTIC_ACTIVITY_ACTING_ON_RNA | 30 | 0.142 | 0.532 | 0.96358 | 0.96358 | 0.82307 | 832 |
| GOBP_ESTABLISHMENT_OF_PROTEIN_LOCALIZATION | 116 | 0.110 | 0.530 | 0.98979 | 0.98979 | 0.83396 | 108 |
| GOBP_ERYTHROCYTE_HOMEOSTASIS | 11 | 0.187 | 0.528 | 0.96109 | 0.96109 | 0.82230 | 232 |
| GOBP_MYELOID_CELL_HOMEOSTASIS | 11 | 0.187 | 0.528 | 0.96109 | 0.96109 | 0.82230 | 232 |

| | | | | | | | |
|---|---|---|---|---|---|---|---|
| GOMF_TRANSCRIPTION_COREGULATOR_ACTIVITY | 36 | 0.135 | 0.527 | 0.97033 | 0.97033 | 0.82429 | 103 |
| HP_ABNORMALITY_OF_THE_MIDDLE_EAR | 37 | 0.133 | 0.525 | 0.97030 | 0.97030 | 0.82429 | 222 |
| HP_ABNORMALITY_OF_THE_BASAL_GANGLIA | 17 | 0.163 | 0.524 | 0.96309 | 0.96309 | 0.82307 | 658 |
| HP_ABNORMALITY_OF_REFRACTION | 41 | 0.130 | 0.522 | 0.97588 | 0.97588 | 0.82764 | 215 |
| GOBP_INTRACELLULAR_TRANSPORT | 102 | 0.110 | 0.519 | 0.99076 | 0.99076 | 0.83433 | 14 |
| HP_RESPIRATORY_INSUFFICIENCY | 37 | 0.132 | 0.518 | 0.97319 | 0.97319 | 0.82582 | 388 |
| GOBP_HEPATICOBILIARY_SYSTEM_DEVELOPMENT | 15 | 0.166 | 0.515 | 0.96975 | 0.96975 | 0.82429 | 409 |
| HP_IRRITABILITY | 15 | 0.166 | 0.515 | 0.96975 | 0.96975 | 0.82429 | 205 |
| GOBP_RNA_PHOSPHODIESTER_BOND_HYDROLYSIS | 18 | 0.157 | 0.513 | 0.96810 | 0.96810 | 0.82429 | 642 |
| HP_ABNORMAL_ELECTRORETINOGRAM | 11 | 0.181 | 0.512 | 0.96879 | 0.96879 | 0.82429 | 45 |
| GOMF_RNA_BINDING | 110 | 0.104 | 0.495 | 0.99414 | 0.99414 | 0.83472 | 47 |
| GOBP_REGULATION_OF_PROTEIN_STABILITY | 19 | 0.145 | 0.482 | 0.97918 | 0.97918 | 0.82953 | 332 |
| GOBP_SMALL_MOLECULE_BIOSYNTHETIC_PROCESS | 30 | 0.125 | 0.470 | 0.98419 | 0.98419 | 0.83105 | 270 |
| HP_MACROTIA | 26 | 0.130 | 0.470 | 0.98376 | 0.98376 | 0.83105 | 181 |
| GOBP_ORGANOPHOSPHATE_CATABOLIC_PROCESS | 13 | 0.157 | 0.467 | 0.98083 | 0.98083 | 0.83002 | 437 |
| HP_ABNORMAL_CONJUGATE_EYE_MOVEMENT | 69 | 0.105 | 0.466 | 0.99464 | 0.99464 | 0.83472 | 277 |
| GOBP_PROTEIN_ACYLATION | 17 | 0.141 | 0.451 | 0.98728 | 0.98728 | 0.83275 | 257 |
| GOBP_ESTABLISHMENT_OF_PROTEIN_LOCALIZATION_TO_MEMBRANE | 20 | 0.132 | 0.443 | 0.98928 | 0.98928 | 0.83396 | 451 |
| GOBP_SULFUR_COMPOUND_METABOLIC_PROCESS | 10 | 0.157 | 0.429 | 0.99297 | 0.99297 | 0.83444 | 183 |
| GOMF_RIBONUCLEASE_ACTIVITY | 16 | 0.134 | 0.425 | 0.99239 | 0.99239 | 0.83444 | 642 |
| GOBP_RNA_PHOSPHODIESTER_BOND_HYDROLYSIS_ENDONUCLEOLYTIC | 11 | 0.150 | 0.424 | 0.99305 | 0.99305 | 0.83444 | 278 |
| GOMF_ENDORIBONUCLEASE_ACTIVITY | 11 | 0.150 | 0.424 | 0.99305 | 0.99305 | 0.83444 | 278 |
| GOBP_PROTEIN_LOCALIZATION_TO_NUCLEUS | 24 | 0.116 | 0.412 | 0.99560 | 0.99560 | 0.83472 | 78 |
| GOCC_SITE_OF_DOUBLE_STRAND_BREAK | 13 | 0.136 | 0.404 | 0.99608 | 0.99608 | 0.83472 | 599 |
| HP_FUNCTIONAL_ABNORMALITY_OF_THE_MIDDLE_EAR | 25 | 0.103 | 0.367 | 0.99907 | 0.99907 | 0.83587 | 200 |
| HP_ABNORMALITY_OF_THE_SCROTUM | 12 | -0.121 | -0.420 | 0.99812 | 0.99812 | 0.83553 | 40 |
| GOBP_ESTABLISHMENT_OF_PROTEIN_LOCALIZATION_TO_ORGANELLE | 41 | -0.084 | -0.442 | 0.99745 | 0.99745 | 0.83542 | 86 |
| HP_ABNORMALITY_OF_THE_CURVATURE_OF_THE_CORNEA | 30 | -0.107 | -0.512 | 0.98600 | 0.98600 | 0.83213 | 174 |
| GOCC_CILIUM | 33 | -0.106 | -0.523 | 0.98254 | 0.98254 | 0.83057 | 937 |
| GOMF_METHYLATED_HISTONE_BINDING | 10 | -0.164 | -0.525 | 0.98037 | 0.98037 | 0.83002 | 875 |
| HP_PREMATURE_BIRTH | 10 | -0.173 | -0.555 | 0.96679 | 0.96679 | 0.82429 | 40 |
| GOBP_MACROMOLECULE_CATABOLIC_PROCESS | 112 | -0.085 | -0.557 | 0.99579 | 0.99579 | 0.83472 | 60 |
| GOBP_CARBOHYDRATE_DERIVATIVE_CATABOLIC_PROCESS | 12 | -0.162 | -0.564 | 0.95713 | 0.95713 | 0.82164 | 877 |
| HP_PARALYSIS | 10 | -0.176 | -0.565 | 0.95954 | 0.95954 | 0.82188 | 862 |
| GOBP_ORGANOPHOSPHATE_METABOLIC_PROCESS | 39 | -0.111 | -0.572 | 0.96673 | 0.96673 | 0.82429 | 215 |
| HP_ABNORMAL_BLOOD_ION_CONCENTRATION | 16 | -0.150 | -0.581 | 0.94842 | 0.94842 | 0.81823 | 207 |
| GOMF_MOLECULAR_ADAPTOR_ACTIVITY | 23 | -0.133 | -0.585 | 0.95637 | 0.95637 | 0.82143 | 61 |
| HP_HYPERMETROPIA | 12 | -0.168 | -0.586 | 0.94587 | 0.94587 | 0.81702 | 207 |
| GOBP_SMALL_MOLECULE_METABOLIC_PROCESS | 79 | -0.096 | -0.588 | 0.97902 | 0.97902 | 0.82953 | 217 |
| HP_PROMINENT_NOSE | 10 | -0.184 | -0.591 | 0.93992 | 0.93992 | 0.81556 | 162 |
| HP_ABNORMAL_CIRCULATING_CARBOXYLIC_ACID_CONCENTRATION | 12 | -0.171 | -0.596 | 0.94055 | 0.94055 | 0.81556 | 675 |

| | | | | | | | |
|---|---|---|---|---|---|---|---|
| GOMF_MODIFICATION_DEPENDENT_PROTEIN_BINDING | 14 | -0.164 | -0.602 | 0.93765 | 0.93765 | 0.81547 | 875 |
| GOMF_ACTIVE_TRANSMEMBRANE_TRANSPORTER_ACTIVITY | 18 | -0.150 | -0.604 | 0.93837 | 0.93837 | 0.81547 | 188 |
| HP_ABNORMAL_LEUKOCYTE_PHYSIOLOGY | 16 | -0.158 | -0.611 | 0.93157 | 0.93157 | 0.81509 | 147 |
| GOBP_ORGANOPHOSPHATE_BIOSYNTHETIC_PROCESS | 24 | -0.137 | -0.613 | 0.93464 | 0.93464 | 0.81547 | 904 |
| GOBP_ATP_METABOLIC_PROCESS | 21 | -0.144 | -0.613 | 0.93008 | 0.93008 | 0.81509 | 672 |
| HP_HYPOTELORISM | 11 | -0.185 | -0.617 | 0.92196 | 0.92196 | 0.81195 | 853 |
| HP_HIRSUTISM | 11 | -0.187 | -0.623 | 0.91826 | 0.91826 | 0.81094 | 673 |
| HP_OTITIS_MEDIA | 16 | -0.161 | -0.625 | 0.92400 | 0.92400 | 0.81209 | 207 |
| HP_ABNORMAL_PALMAR_DERMATOGLYPHICS | 16 | -0.161 | -0.625 | 0.92331 | 0.92331 | 0.81195 | 80 |
| HP_APLASIA_HYPOPLASIA_INVOLVING_BONES_OF_THE_LOWER_LIMBS | 29 | -0.132 | -0.628 | 0.92314 | 0.92314 | 0.81195 | 162 |
| HP_ELEVATED_HEPATIC_TRANSAMINASE | 16 | -0.163 | -0.631 | 0.91850 | 0.91850 | 0.81094 | 200 |
| HP_LONG_PHILTRUM | 24 | -0.143 | -0.638 | 0.91540 | 0.91540 | 0.80912 | 40 |
| HP_ABNORMAL_SYSTEMIC_ARTERIAL_MORPHOLOGY | 32 | -0.131 | -0.639 | 0.92867 | 0.92867 | 0.81481 | 148 |
| HP_ABNORMAL_ERYTHROCYTE_MORPHOLOGY | 47 | -0.119 | -0.643 | 0.94347 | 0.94347 | 0.81644 | 925 |
| HP_SMALL_HAND | 12 | -0.185 | -0.645 | 0.89737 | 0.89737 | 0.80188 | 853 |
| HP_ABNORMALITY_OF_UPPER_LIMB_JOINT | 29 | -0.138 | -0.654 | 0.90726 | 0.90726 | 0.80606 | 99 |
| HP_ABNORMAL_AORTIC_MORPHOLOGY | 30 | -0.137 | -0.655 | 0.91164 | 0.91164 | 0.80764 | 55 |
| GOBP_NUCLEAR_TRANSPORT | 20 | -0.157 | -0.660 | 0.89978 | 0.89978 | 0.80264 | 131 |
| HP_REDUCED_VISUAL_ACUITY | 29 | -0.139 | -0.662 | 0.90082 | 0.90082 | 0.80264 | 155 |
| HP_ABNORMAL_DERMATOGLYPHICS | 20 | -0.158 | -0.662 | 0.89684 | 0.89684 | 0.80188 | 292 |
| HP_HEPATOMEGALY | 31 | -0.138 | -0.663 | 0.90840 | 0.90840 | 0.80661 | 200 |
| GOBP_CILIUM_ORGANIZATION | 17 | -0.168 | -0.664 | 0.89540 | 0.89540 | 0.80150 | 872 |
| GOBP_REGULATION_OF_CANONICAL_WNT_SIGNALING_PATHWAY | 14 | -0.182 | -0.667 | 0.88027 | 0.88027 | 0.79652 | 548 |
| HP_JOINT_LAXITY | 13 | -0.189 | -0.676 | 0.87035 | 0.87035 | 0.79049 | 242 |
| HP_ABNORMAL_STERNUM_MORPHOLOGY | 24 | -0.152 | -0.676 | 0.88412 | 0.88412 | 0.79664 | 125 |
| HP_ABNORMAL_HEART_VALVE_MORPHOLOGY | 24 | -0.152 | -0.677 | 0.88412 | 0.88412 | 0.79664 | 207 |
| GOCC_ENVELOPE | 104 | -0.105 | -0.679 | 0.95080 | 0.95080 | 0.81938 | 674 |
| HP_ABNORMAL_NIPPLE_MORPHOLOGY | 12 | -0.198 | -0.690 | 0.85357 | 0.85357 | 0.78426 | 148 |
| HP_FEEDING_DIFFICULTIES | 73 | -0.115 | -0.690 | 0.93294 | 0.93294 | 0.81509 | 174 |
| HP_SEVERE_GLOBAL_DEVELOPMENTAL_DELAY | 13 | -0.195 | -0.697 | 0.84531 | 0.84531 | 0.78281 | 60 |
| GOBP_STEROID_METABOLIC_PROCESS | 12 | -0.201 | -0.700 | 0.84168 | 0.84168 | 0.78140 | 128 |
| GOCC_PML_BODY | 11 | -0.211 | -0.705 | 0.83405 | 0.83405 | 0.77946 | 454 |
| HP_CEREBRAL_CORTICAL_ATROPHY | 19 | -0.171 | -0.706 | 0.85387 | 0.85387 | 0.78426 | 461 |
| GOBP_CELLULAR_CARBOHYDRATE_METABOLIC_PROCESS | 13 | -0.197 | -0.706 | 0.83537 | 0.83537 | 0.77975 | 841 |
| GOBP_NUCLEOBASE_CONTAINING_SMALL_MOLECULE_METABOLIC_PROCESS | 28 | -0.152 | -0.711 | 0.84825 | 0.84825 | 0.78281 | 890 |
| HP_ASTIGMATISM | 23 | -0.162 | -0.711 | 0.84080 | 0.84080 | 0.78140 | 879 |
| HP_ABNORMAL_CNS_MYELINATION | 20 | -0.170 | -0.712 | 0.84288 | 0.84288 | 0.78162 | 155 |
| HP_EMOTIONAL_LABILITY | 10 | -0.222 | -0.713 | 0.82277 | 0.82277 | 0.77808 | 519 |
| GOCC_MOTILE_CILIUM | 13 | -0.199 | -0.713 | 0.82702 | 0.82702 | 0.77808 | 839 |
| HP_ABNORMAL_UPPER_LIMB_BONE_MORPHOLOGY | 37 | -0.140 | -0.714 | 0.86868 | 0.86868 | 0.78944 | 40 |
| GOBP_CELLULAR_RESPONSE_TO_REACTIVE_OXYGEN_SPECIES | 11 | -0.215 | -0.716 | 0.81863 | 0.81863 | 0.77808 | 70 |

| | | | | | | | |
|---|---|---|---|---|---|---|---|
| GOBP_RESPONSE_TO_REACTIVE_OXYGEN_SPECIES | 11 | -0.215 | -0.716 | 0.81863 | 0.81863 | 0.77808 | 70 |
| HP_SHORT_FOOT | 10 | -0.224 | -0.717 | 0.81582 | 0.81582 | 0.77808 | 162 |
| HP_ABNORMAL_NEUTROPHIL_COUNT | 16 | -0.186 | -0.720 | 0.82806 | 0.82806 | 0.77808 | 853 |
| HP_ABNORMAL_LIVER_MORPHOLOGY | 50 | -0.132 | -0.724 | 0.88604 | 0.88604 | 0.79727 | 204 |
| GOBP_REGULATION_OF_LIPID_METABOLIC_PROCESS | 14 | -0.198 | -0.727 | 0.81426 | 0.81426 | 0.77808 | 348 |
| GOBP_DNA_TEMPLATED_TRANSCRIPTION_INITIATION | 10 | -0.227 | -0.727 | 0.80405 | 0.80405 | 0.77776 | 131 |
| GOCC_P_BODY | 12 | -0.209 | -0.729 | 0.80413 | 0.80413 | 0.77776 | 269 |
| GOBP_REGULATION_OF_APOPTOTIC_SIGNALING_PATHWAY | 22 | -0.169 | -0.730 | 0.82769 | 0.82769 | 0.77808 | 2 |
| GOBP_SPERMATID_DIFFERENTIATION | 12 | -0.210 | -0.733 | 0.79819 | 0.79819 | 0.77735 | 54 |
| GOMF_PROTEIN_MACROMOLECULE_ADAPTOR_ACTIVITY | 20 | -0.175 | -0.733 | 0.81865 | 0.81865 | 0.77808 | 224 |
| HP_ABNORMAL_CEREBRAL_VASCULAR_MORPHOLOGY | 13 | -0.205 | -0.735 | 0.79525 | 0.79525 | 0.77649 | 207 |
| HP_ABNORMALITY_OF_HUMORAL_IMMUNITY | 15 | -0.194 | -0.735 | 0.81764 | 0.81764 | 0.77808 | 147 |
| HP_APLASIA_HYPOPLASIA_INVOLVING_THE_METACARPAL_BONES | 10 | -0.229 | -0.735 | 0.79469 | 0.79469 | 0.77649 | 11 |
| HP_RESPIRATORY_DISTRESS | 11 | -0.220 | -0.735 | 0.79395 | 0.79395 | 0.77649 | 200 |
| HP_ABNORMAL_GRANULOCYTE_COUNT | 17 | -0.186 | -0.737 | 0.82239 | 0.82239 | 0.77808 | 853 |
| GOBP_NUCLEOSIDE_PHOSPHATE_BIOSYNTHETIC_PROCESS | 15 | -0.195 | -0.738 | 0.81496 | 0.81496 | 0.77808 | 608 |
| GOBP_PEPTIDYL_AMINO_ACID_MODIFICATION | 78 | -0.122 | -0.744 | 0.89698 | 0.89698 | 0.80188 | 45 |
| HP_DELAYED_ABILITY_TO_WALK | 14 | -0.204 | -0.747 | 0.79204 | 0.79204 | 0.77649 | 162 |
| HP_LEUKODYSTROPHY | 12 | -0.215 | -0.750 | 0.77597 | 0.77597 | 0.77031 | 674 |
| HP_FUNCTIONAL_MOTOR_DEFICIT | 27 | -0.162 | -0.751 | 0.81162 | 0.81162 | 0.77808 | 269 |
| GOBP_PROTEIN_IMPORT | 18 | -0.187 | -0.754 | 0.79470 | 0.79470 | 0.77649 | 131 |
| HP_BOWING_OF_THE_LEGS | 18 | -0.187 | -0.755 | 0.79183 | 0.79183 | 0.77649 | 242 |
| HP_ABNORMAL_ATRIOVENTRICULAR_VALVE_PHYSIOLOGY | 17 | -0.192 | -0.759 | 0.79291 | 0.79291 | 0.77649 | 242 |
| HP_ABNORMAL_DIAPHYSIS_MORPHOLOGY | 21 | -0.178 | -0.760 | 0.79098 | 0.79098 | 0.77649 | 242 |
| HP_GENERALIZED_HYPOTONIA | 62 | -0.132 | -0.760 | 0.85336 | 0.85336 | 0.78426 | 168 |
| HP_ABNORMAL_ESOPHAGUS_PHYSIOLOGY | 50 | -0.139 | -0.762 | 0.84729 | 0.84729 | 0.78281 | 174 |
| HP_SOMATIC_SENSORY_DYSFUNCTION | 14 | -0.209 | -0.766 | 0.76418 | 0.76418 | 0.76502 | 308 |
| HP_ABNORMALITY_OF_THE_VASCULATURE_OF_THE_EYE | 22 | -0.179 | -0.769 | 0.77791 | 0.77791 | 0.77031 | 155 |
| HP_INTELLECTUAL_DISABILITY_MILD | 31 | -0.160 | -0.771 | 0.80742 | 0.80742 | 0.77808 | 109 |
| HP_PERIPHERAL_AXONAL_DEGENERATION | 10 | -0.241 | -0.775 | 0.73490 | 0.73490 | 0.75562 | 385 |
| GOBP_MAINTENANCE_OF_LOCATION | 11 | -0.233 | -0.776 | 0.74244 | 0.74244 | 0.75834 | 354 |
| HP_PECTUS_EXCAVATUM | 16 | -0.201 | -0.777 | 0.75447 | 0.75447 | 0.76109 | 87 |
| HP_SMOOTH_PHILTRUM | 14 | -0.212 | -0.778 | 0.74892 | 0.74892 | 0.75981 | 562 |
| GOBP_ACTIVATION_OF_GTPASE_ACTIVITY | 10 | -0.244 | -0.782 | 0.72615 | 0.72615 | 0.75323 | 212 |
| GOMF_PRIMARY_ACTIVE_TRANSMEMBRANE_TRANSPORTER_ACTIVITY | 13 | -0.219 | -0.784 | 0.72978 | 0.72978 | 0.75435 | 674 |
| HP_ABNORMAL_ELASTICITY_OF_SKIN | 11 | -0.235 | -0.784 | 0.73381 | 0.73381 | 0.75529 | 131 |
| GOBP_DEFENSE_RESPONSE_TO_SYMBIONT | 11 | -0.235 | -0.786 | 0.73226 | 0.73226 | 0.75504 | 170 |
| HP_APLASIA_HYPOPLASIA_INVOLVING_BONES_OF_THE_FEET | 20 | -0.187 | -0.786 | 0.76065 | 0.76065 | 0.76427 | 40 |
| HP_ABNORMALITY_OF_THE_METAPHYSIS | 16 | -0.203 | -0.787 | 0.74106 | 0.74106 | 0.75742 | 195 |
| HP_ABNORMALITY_OF_THE_UPPER_RESPIRATORY_TRACT | 26 | -0.173 | -0.788 | 0.77348 | 0.77348 | 0.76979 | 155 |
| HP_TYPE_II_DIABETES_MELLITUS | 12 | -0.227 | -0.790 | 0.71965 | 0.71965 | 0.75042 | 207 |

| | | | | | | | |
|---|---|---|---|---|---|---|---|
| GOBP_REGULATION_OF_CELLULAR_PROTEIN_CATABOLIC_PROCESS | 21 | -0.186 | -0.793 | 0.75075 | 0.75075 | 0.75981 | 86 |
| GOBP_NUCLEAR_ENVELOPE_ORGANIZATION | 10 | -0.247 | -0.794 | 0.71256 | 0.71256 | 0.74651 | 98 |
| GOBP_REGULATION_OF_SMALL_MOLECULE_METABOLIC_PROCESS | 14 | -0.217 | -0.794 | 0.72902 | 0.72902 | 0.75435 | 209 |
| HP_DYSARTHRIA | 35 | -0.158 | -0.795 | 0.77825 | 0.77825 | 0.77031 | 321 |
| HP_ABNORMALITY_OF_NEUTROPHILS | 21 | -0.187 | -0.796 | 0.74624 | 0.74624 | 0.75981 | 853 |
| HP_ABNORMAL_MYELOID_LEUKOCYTE_MORPHOLOGY | 21 | -0.187 | -0.796 | 0.74624 | 0.74624 | 0.75981 | 853 |
| HP_DYSPNEA | 17 | -0.201 | -0.796 | 0.74693 | 0.74693 | 0.75981 | 200 |
| GOBP_RESPONSE_TO_VIRUS | 17 | -0.202 | -0.798 | 0.74342 | 0.74342 | 0.75848 | 170 |
| GOBP_VESICLE_ORGANIZATION | 20 | -0.191 | -0.803 | 0.73899 | 0.73899 | 0.75674 | 486 |
| HP_CONTRACTURES_OF_THE_JOINTS_OF_THE_UPPER_LIMBS | 23 | -0.183 | -0.806 | 0.73703 | 0.73703 | 0.75644 | 99 |
| GOBP_CELL_PROJECTION_ASSEMBLY | 33 | -0.164 | -0.808 | 0.75057 | 0.75057 | 0.75981 | 140 |
| HP_ABNORMAL_RETINAL_MORPHOLOGY | 56 | -0.143 | -0.809 | 0.78862 | 0.78862 | 0.77633 | 155 |
| HP_NEOPLASM_OF_THE_NERVOUS_SYSTEM | 14 | -0.221 | -0.809 | 0.70846 | 0.70846 | 0.74423 | 148 |
| GOBP_CELLULAR_RESPONSE_TO_EXTRACELLULAR_STIMULUS | 16 | -0.209 | -0.811 | 0.70461 | 0.70461 | 0.74321 | 303 |
| HP_ABNORMALITY_OF_MUSCLE_SIZE | 38 | -0.159 | -0.814 | 0.76108 | 0.76108 | 0.76427 | 46 |
| HP_FUNCTIONAL_ABNORMALITY_OF_THE_GASTROINTESTINAL_TRACT | 63 | -0.141 | -0.816 | 0.78166 | 0.78166 | 0.77200 | 174 |
| HP_ABNORMAL_CIRCULATING_METABOLITE_CONCENTRATION | 76 | -0.135 | -0.816 | 0.79592 | 0.79592 | 0.77649 | 685 |
| GOBP_RESPONSE_TO_PEPTIDE_HORMONE | 22 | -0.190 | -0.819 | 0.71567 | 0.71567 | 0.74876 | 249 |
| HP_ABNORMAL_CIRCULATING_LIPID_CONCENTRATION | 18 | -0.203 | -0.820 | 0.71014 | 0.71014 | 0.74549 | 207 |
| HP_LOW_SET_POSTERIORLY_ROTATED_EARS | 19 | -0.199 | -0.822 | 0.70084 | 0.70084 | 0.74321 | 148 |
| GOBP_REGULATION_OF_HISTONE_MODIFICATION | 14 | -0.224 | -0.824 | 0.68789 | 0.68789 | 0.73610 | 813 |
| HP_HYPERLIPIDEMIA | 10 | -0.257 | -0.826 | 0.67331 | 0.67331 | 0.73011 | 147 |
| HP_HYPERTRIGLYCERIDEMIA | 10 | -0.257 | -0.826 | 0.67331 | 0.67331 | 0.73011 | 147 |
| GOBP_POSITIVE_REGULATION_OF_HISTONE_MODIFICATION | 11 | -0.248 | -0.828 | 0.67088 | 0.67088 | 0.73011 | 788 |
| GOMF_UNFOLDED_PROTEIN_BINDING | 10 | -0.258 | -0.829 | 0.66878 | 0.66878 | 0.73011 | 777 |
| HP_ABNORMAL_VISUAL_ELECTROPHYSIOLOGY | 15 | -0.219 | -0.830 | 0.68737 | 0.68737 | 0.73605 | 229 |
| HP_PTOSIS | 56 | -0.147 | -0.831 | 0.75689 | 0.75689 | 0.76237 | 148 |
| GOBP_GOLGI_VESICLE_TRANSPORT | 25 | -0.185 | -0.832 | 0.71189 | 0.71189 | 0.74651 | 413 |
| HP_GENERALIZED_ABNORMALITY_OF_SKIN | 55 | -0.149 | -0.834 | 0.74955 | 0.74955 | 0.75981 | 207 |
| GOMF_TRANSCRIPTION_FACTOR_BINDING | 37 | -0.164 | -0.835 | 0.72714 | 0.72714 | 0.75323 | 209 |
| HP_JOINT_CONTRACTURE_OF_THE_HAND | 18 | -0.207 | -0.836 | 0.68219 | 0.68219 | 0.73381 | 87 |
| GOBP_LIPID_MODIFICATION | 14 | -0.229 | -0.840 | 0.66501 | 0.66501 | 0.72924 | 209 |
| HP_TALL_STATURE | 14 | -0.231 | -0.848 | 0.65605 | 0.65605 | 0.72263 | 42 |
| HP_PROTRUDING_EAR | 10 | -0.266 | -0.853 | 0.63043 | 0.63043 | 0.70837 | 4 |
| HP_ABNORMALITY_OF_THE_SKULL_BASE | 10 | -0.267 | -0.855 | 0.62802 | 0.62802 | 0.70800 | 79 |
| HP_OPTIC_DISC_PALLOR | 15 | -0.226 | -0.856 | 0.65097 | 0.65097 | 0.71996 | 673 |
| HP_CENTRAL_NERVOUS_SYSTEM_CYST | 10 | -0.267 | -0.857 | 0.62742 | 0.62742 | 0.70800 | 79 |
| HP_VASCULAR_SKIN_ABNORMALITY | 30 | -0.180 | -0.857 | 0.67935 | 0.67935 | 0.73253 | 196 |
| HP_PIGMENTARY_RETINOPATHY | 13 | -0.239 | -0.857 | 0.63864 | 0.63864 | 0.71177 | 674 |
| HP_ABNORMAL_INFLAMMATORY_RESPONSE | 56 | -0.152 | -0.858 | 0.71617 | 0.71617 | 0.74877 | 211 |
| HP_FINGER_SYNDACTYLY | 20 | -0.204 | -0.859 | 0.65675 | 0.65675 | 0.72266 | 15 |

| | | | | | | | |
|---|---|---|---|---|---|---|---|
| HP_ABNORMAL_MITRAL_VALVE_PHYSIOLOGY | 12 | -0.247 | -0.862 | 0.62985 | 0.62985 | 0.70837 | 242 |
| GOBP_PROTEIN_FOLDING | 11 | -0.259 | -0.863 | 0.62461 | 0.62461 | 0.70696 | 777 |
| HP_NYCTALOPIA | 13 | -0.241 | -0.865 | 0.62965 | 0.62965 | 0.70837 | 141 |
| GOBP_CELLULAR_KETONE_METABOLIC_PROCESS | 12 | -0.248 | -0.865 | 0.62735 | 0.62735 | 0.70800 | 788 |
| GOBP_POSITIVE_REGULATION_OF_NUCLEOBASE_CONTAINING_COMPOUND_META | 125 | -0.130 | -0.866 | 0.76969 | 0.76969 | 0.76844 | 156 |
| HP_JOINT_STIFFNESS | 14 | -0.236 | -0.867 | 0.63018 | 0.63018 | 0.70837 | 675 |
| HP_TRIANGULAR_SHAPED_PHALANGES_OF_THE_HAND | 21 | -0.204 | -0.868 | 0.65564 | 0.65564 | 0.72263 | 55 |
| GOCC_COATED_VESICLE | 15 | -0.229 | -0.868 | 0.63427 | 0.63427 | 0.71062 | 413 |
| HP_VISUAL_FIELD_DEFECT | 12 | -0.249 | -0.868 | 0.62140 | 0.62140 | 0.70487 | 93 |
| GOMF_TRANSITION_METAL_ION_BINDING | 64 | -0.150 | -0.872 | 0.69620 | 0.69620 | 0.74192 | 269 |
| HP_BRACHYCEPHALY | 16 | -0.225 | -0.872 | 0.62070 | 0.62070 | 0.70487 | 162 |
| GOBP_VESICLE_LOCALIZATION | 14 | -0.238 | -0.873 | 0.62454 | 0.62454 | 0.70696 | 510 |
| GOCC_RECYCLING_ENDOSOME | 14 | -0.239 | -0.875 | 0.61990 | 0.61990 | 0.70471 | 260 |
| HP_DEMENTIA | 12 | -0.252 | -0.877 | 0.60920 | 0.60920 | 0.69766 | 119 |
| HP_SHORT_PHILTRUM | 10 | -0.274 | -0.878 | 0.60054 | 0.60054 | 0.69576 | 143 |
| HP_ABNORMALITY_OF_THE_OVARY | 20 | -0.209 | -0.879 | 0.63326 | 0.63326 | 0.71000 | 830 |
| HP_LOWER_EXTREMITY_JOINT_DISLOCATION | 27 | -0.190 | -0.880 | 0.65270 | 0.65270 | 0.72093 | 99 |
| GOBP_NUCLEOSIDE_MONOPHOSPHATE_METABOLIC_PROCESS | 10 | -0.275 | -0.882 | 0.59360 | 0.59360 | 0.69271 | 760 |
| HP_ABNORMALITY_OF_CORNEAL_SIZE | 12 | -0.254 | -0.884 | 0.60075 | 0.60075 | 0.69576 | 79 |
| GOBP_REGULATION_OF_CATABOLIC_PROCESS | 76 | -0.146 | -0.886 | 0.67725 | 0.67725 | 0.73149 | 217 |
| HP_ABNORMAL_ANTERIOR_EYE_SEGMENT_MORPHOLOGY | 70 | -0.149 | -0.887 | 0.68244 | 0.68244 | 0.73381 | 174 |
| HP_ABNORMAL_LOWER_LIMB_BONE_MORPHOLOGY | 57 | -0.157 | -0.888 | 0.67278 | 0.67278 | 0.73011 | 162 |
| HP_HYPERLORDOSIS | 12 | -0.255 | -0.889 | 0.59168 | 0.59168 | 0.69195 | 233 |
| HP_ABNORMAL_ADIPOSE_TISSUE_MORPHOLOGY | 17 | -0.225 | -0.890 | 0.60969 | 0.60969 | 0.69771 | 55 |
| GOBP_IMPORT_INTO_NUCLEUS | 14 | -0.244 | -0.896 | 0.59370 | 0.59370 | 0.69271 | 131 |
| GOBP_INTRINSIC_APOPTOTIC_SIGNALING_PATHWAY_IN_RESPONSE_TO_DNA_DAM | 10 | -0.279 | -0.896 | 0.57579 | 0.57579 | 0.68417 | 42 |
| GOBP_RESPONSE_TO_ORGANIC_CYCLIC_COMPOUND | 49 | -0.165 | -0.897 | 0.63989 | 0.63989 | 0.71177 | 132 |
| HP_INTELLECTUAL_DISABILITY_PROFOUND | 14 | -0.245 | -0.898 | 0.59171 | 0.59171 | 0.69195 | 109 |
| HP_LIMITED_ELBOW_MOVEMENT | 11 | -0.270 | -0.902 | 0.57958 | 0.57958 | 0.68605 | 99 |
| HP_SANDAL_GAP | 11 | -0.270 | -0.902 | 0.57896 | 0.57896 | 0.68605 | 40 |
| GOBP_NEGATIVE_REGULATION_OF_APOPTOTIC_SIGNALING_PATHWAY | 13 | -0.252 | -0.902 | 0.58537 | 0.58537 | 0.68817 | 2 |
| HP_COARSE_FACIAL_FEATURES | 10 | -0.283 | -0.907 | 0.56341 | 0.56341 | 0.67621 | 296 |
| GOBP_MITOCHONDRIAL_MEMBRANE_ORGANIZATION | 11 | -0.272 | -0.908 | 0.57033 | 0.57033 | 0.68030 | 763 |
| HP_DIFFICULTY_WALKING | 14 | -0.248 | -0.909 | 0.57347 | 0.57347 | 0.68299 | 46 |
| GOBP_RESPONSE_TO_ABIOTIC_STIMULUS | 71 | -0.153 | -0.910 | 0.64561 | 0.64561 | 0.71624 | 218 |
| GOBP_RESPONSE_TO_OXYGEN_LEVELS | 17 | -0.230 | -0.912 | 0.57318 | 0.57318 | 0.68299 | 209 |
| HP_SHORT_NOSE | 21 | -0.215 | -0.915 | 0.58045 | 0.58045 | 0.68656 | 358 |
| GOBP_ORGANIC_HYDROXY_COMPOUND_METABOLIC_PROCESS | 24 | -0.206 | -0.917 | 0.58581 | 0.58581 | 0.68817 | 131 |
| GOCC_EARLY_ENDOSOME | 15 | -0.243 | -0.919 | 0.56012 | 0.56012 | 0.67435 | 339 |
| HP_BRACHYDACTYLY | 19 | -0.222 | -0.919 | 0.56307 | 0.56307 | 0.67621 | 173 |
| HP_ABNORMALITY_OF_THE_FRONTAL_HAIRLINE | 12 | -0.265 | -0.921 | 0.54693 | 0.54693 | 0.66782 | 174 |

| | | | | | | | |
|---|---|---|---|---|---|---|---|
| HP_ABNORMALITY_OF_THE_HAIRLINE | 12 | -0.265 | -0.921 | 0.54693 | 0.54693 | 0.66782 | 174 |
| HP_ABNORMAL_PHARYNX_MORPHOLOGY | 13 | -0.258 | -0.922 | 0.55231 | 0.55231 | 0.67061 | 131 |
| HP_ABNORMALITY_OF_FACIAL_SKELETON | 89 | -0.148 | -0.925 | 0.62129 | 0.62129 | 0.70487 | 99 |
| HP_JOINT_HYPERMOBILITY | 26 | -0.204 | -0.928 | 0.56796 | 0.56796 | 0.67866 | 70 |
| HP_ABNORMALITY_OF_THE_ELBOW | 18 | -0.230 | -0.929 | 0.54389 | 0.54389 | 0.66726 | 99 |
| HP_SKELETAL_MUSCLE_ATROPHY | 31 | -0.195 | -0.938 | 0.55407 | 0.55407 | 0.67175 | 46 |
| HP_MICROCORNEA | 10 | -0.293 | -0.939 | 0.52204 | 0.52204 | 0.64941 | 79 |
| GOBP_POSITIVE_REGULATION_OF_NERVOUS_SYSTEM_DEVELOPMENT | 12 | -0.270 | -0.941 | 0.52222 | 0.52222 | 0.64941 | 98 |
| HP_ABNORMAL_CIRCULATING_PROTEIN_CONCENTRATION | 35 | -0.187 | -0.941 | 0.56548 | 0.56548 | 0.67765 | 65 |
| HP_ABNORMALITY_OF_THE_VOICE | 36 | -0.186 | -0.941 | 0.56244 | 0.56244 | 0.67609 | 87 |
| HP_MYALGIA | 13 | -0.263 | -0.941 | 0.52856 | 0.52856 | 0.65415 | 55 |
| GOBP_NEURAL_TUBE_DEVELOPMENT | 10 | -0.295 | -0.945 | 0.51600 | 0.51600 | 0.64530 | 204 |
| HP_REDUCED_TENDON_REFLEXES | 28 | -0.202 | -0.945 | 0.54987 | 0.54987 | 0.66929 | 310 |
| GOBP_DEPHOSPHORYLATION | 31 | -0.196 | -0.946 | 0.54200 | 0.54200 | 0.66547 | 153 |
| HP_APLASIA_HYPOPLASIA_OF_THE_EXTREMITIES | 52 | -0.172 | -0.947 | 0.55995 | 0.55995 | 0.67435 | 55 |
| HP_LOCALIZED_SKIN_LESION | 68 | -0.161 | -0.948 | 0.58445 | 0.58445 | 0.68766 | 174 |
| HP_ABNORMAL_ENZYME_COENZYME_ACTIVITY | 30 | -0.199 | -0.949 | 0.53500 | 0.53500 | 0.66000 | 685 |
| GOCC_RIBONUCLEOPROTEIN_GRANULE | 18 | -0.236 | -0.951 | 0.51129 | 0.51129 | 0.64200 | 269 |
| GOBP_GLIOGENESIS | 12 | -0.274 | -0.955 | 0.50313 | 0.50313 | 0.63588 | 31 |
| HP_ABNORMAL_OVARIAN_MORPHOLOGY | 12 | -0.274 | -0.956 | 0.50094 | 0.50094 | 0.63562 | 761 |
| GOBP_PHOSPHOLIPID_METABOLIC_PROCESS | 13 | -0.268 | -0.961 | 0.50289 | 0.50289 | 0.63588 | 49 |
| HP_CHOREA | 21 | -0.226 | -0.962 | 0.51692 | 0.51692 | 0.64593 | 119 |
| HP_ACUTE_PHASE_RESPONSE | 10 | -0.300 | -0.963 | 0.49004 | 0.49004 | 0.62908 | 147 |
| HP_HYPOREFLEXIA | 18 | -0.240 | -0.967 | 0.48692 | 0.48692 | 0.62716 | 162 |
| GOMF_NUCLEOTIDYLTRANSFERASE_ACTIVITY | 11 | -0.291 | -0.972 | 0.48489 | 0.48489 | 0.62506 | 4 |
| HP_ABNORMAL_LATERAL_VENTRICLE_MORPHOLOGY | 14 | -0.265 | -0.973 | 0.48922 | 0.48922 | 0.62891 | 174 |
| HP_DYSPHAGIA | 31 | -0.202 | -0.975 | 0.49866 | 0.49866 | 0.63385 | 334 |
| HP_ABNORMAL_PATTERN_OF_RESPIRATION | 37 | -0.191 | -0.977 | 0.50292 | 0.50292 | 0.63588 | 200 |
| GOBP_HINDBRAIN_DEVELOPMENT | 10 | -0.305 | -0.977 | 0.47313 | 0.47313 | 0.61657 | 274 |
| GOBP_REGULATION_OF_CYSTEINE_TYPE_ENDOPEPTIDASE_ACTIVITY | 14 | -0.266 | -0.977 | 0.48060 | 0.48060 | 0.62108 | 52 |
| GOBP_RNA_LOCALIZATION | 15 | -0.260 | -0.983 | 0.46827 | 0.46827 | 0.61489 | 777 |
| HP_ABNORMALITY_OF_FINGER | 90 | -0.157 | -0.985 | 0.51332 | 0.51332 | 0.64299 | 80 |
| GOBP_LYMPHOCYTE_ACTIVATION_INVOLVED_IN_IMMUNE_RESPONSE | 12 | -0.283 | -0.987 | 0.45713 | 0.45713 | 0.60784 | 176 |
| HP_APHASIA | 11 | -0.296 | -0.987 | 0.46514 | 0.46514 | 0.61225 | 55 |
| HP_BROAD_FINGER | 10 | -0.308 | -0.987 | 0.45984 | 0.45984 | 0.60795 | 40 |
| HP_BROAD_LONG_BONES | 10 | -0.308 | -0.987 | 0.45984 | 0.45984 | 0.60795 | 40 |
| GOMF_TRANSCRIPTION_COACTIVATOR_ACTIVITY | 18 | -0.245 | -0.988 | 0.45898 | 0.45898 | 0.60784 | 356 |
| HP_DEVIATION_OF_TOES | 11 | -0.296 | -0.988 | 0.46391 | 0.46391 | 0.61124 | 46 |
| GOCC_FICOLIN_1_RICH_GRANULE_LUMEN | 10 | -0.309 | -0.991 | 0.45592 | 0.45592 | 0.60784 | 725 |
| HP_ABNORMALITY_OF_SKIN_PHYSIOLOGY | 24 | -0.222 | -0.991 | 0.47273 | 0.47273 | 0.61657 | 200 |
| HP_ABNORMALITY_OF_THE_PHARYNX | 15 | -0.262 | -0.991 | 0.45324 | 0.45324 | 0.60646 | 155 |

| | | | | | | | |
|---|---|---|---|---|---|---|---|
| GOBP_RESPONSE_TO_OXIDATIVE_STRESS | 26 | -0.218 | -0.992 | 0.47652 | 0.47652 | 0.61764 | 87 |
| HP_OSTEOPOROSIS | 11 | -0.298 | -0.993 | 0.45898 | 0.45898 | 0.60784 | 207 |
| GOBP_REGULATION_OF_IMMUNE_EFFECTOR_PROCESS | 17 | -0.251 | -0.994 | 0.44717 | 0.44717 | 0.60088 | 73 |
| HP_ABNORMAL_JAW_MORPHOLOGY | 82 | -0.163 | -1.001 | 0.49291 | 0.49291 | 0.63168 | 174 |
| GOBP_PROTEIN_AUTOPHOSPHORYLATION | 15 | -0.266 | -1.006 | 0.43454 | 0.43454 | 0.59165 | 227 |
| HP_APLASIA_HYPOPLASIA_OF_THE_EAR | 12 | -0.290 | -1.011 | 0.42804 | 0.42804 | 0.58693 | 148 |
| GOBP_LIPID_METABOLIC_PROCESS | 59 | -0.177 | -1.013 | 0.44707 | 0.44707 | 0.60088 | 226 |
| GOBP_RESPONSE_TO_HORMONE | 46 | -0.189 | -1.014 | 0.45182 | 0.45182 | 0.60549 | 356 |
| HP_ABNORMALITY_OF_FACIAL_SOFT_TISSUE | 24 | -0.228 | -1.016 | 0.44066 | 0.44066 | 0.59632 | 87 |
| HP_POOR_SPEECH | 19 | -0.246 | -1.017 | 0.42930 | 0.42930 | 0.58781 | 162 |
| HP_PACE_OF_PROGRESSION | 36 | -0.201 | -1.019 | 0.43084 | 0.43084 | 0.58803 | 87 |
| HP_ABNORMAL_HAIR_PATTERN | 19 | -0.247 | -1.019 | 0.42639 | 0.42639 | 0.58571 | 174 |
| HP_ABNORMALITY_OF_THE_MIDFACE | 33 | -0.207 | -1.023 | 0.43179 | 0.43179 | 0.58843 | 174 |
| HP_ABNORMALITY_OF_THE_PITUITARY_GLAND | 20 | -0.243 | -1.023 | 0.41887 | 0.41887 | 0.58313 | 204 |
| GOBP_ESTABLISHMENT_OR_MAINTENANCE_OF_CELL_POLARITY | 18 | -0.254 | -1.026 | 0.41491 | 0.41491 | 0.58022 | 124 |
| GOCC_FICOLIN_1_RICH_GRANULE | 11 | -0.309 | -1.031 | 0.41302 | 0.41302 | 0.57834 | 725 |
| HP_APLASIA_HYPOPLASIA_INVOLVING_BONES_OF_THE_UPPER_LIMBS | 37 | -0.203 | -1.036 | 0.41537 | 0.41537 | 0.58035 | 55 |
| GOBP_CARBOHYDRATE_METABOLIC_PROCESS | 24 | -0.233 | -1.037 | 0.41059 | 0.41059 | 0.57731 | 343 |
| HP_ABNORMALITY_OF_LOWER_LIP | 18 | -0.258 | -1.040 | 0.39807 | 0.39807 | 0.56952 | 46 |
| GOBP_CARBOHYDRATE_DERIVATIVE_METABOLIC_PROCESS | 52 | -0.189 | -1.040 | 0.40629 | 0.40629 | 0.57632 | 192 |
| HP_ABNORMAL_THORAX_MORPHOLOGY | 47 | -0.194 | -1.044 | 0.40779 | 0.40779 | 0.57632 | 99 |
| GOBP_MEMBRANE_ORGANIZATION | 55 | -0.186 | -1.044 | 0.39710 | 0.39710 | 0.56952 | 29 |
| HP_INABILITY_TO_WALK | 18 | -0.259 | -1.044 | 0.39126 | 0.39126 | 0.56548 | 229 |
| GOBP_ORGANIC_CYCLIC_COMPOUND_CATABOLIC_PROCESS | 52 | -0.190 | -1.048 | 0.39756 | 0.39756 | 0.56952 | 285 |
| HP_BLEPHAROPHIMOSIS | 17 | -0.265 | -1.050 | 0.38084 | 0.38084 | 0.55629 | 256 |
| HP_ABNORMALITY_OF_THE_METACARPAL_BONES | 14 | -0.288 | -1.055 | 0.37612 | 0.37612 | 0.55240 | 15 |
| HP_HIP_DYSPLASIA | 18 | -0.262 | -1.055 | 0.37764 | 0.37764 | 0.55359 | 174 |
| GOBP_POSITIVE_REGULATION_OF_PEPTIDASE_ACTIVITY | 10 | -0.329 | -1.056 | 0.37560 | 0.37560 | 0.55217 | 704 |
| GOBP_POSITIVE_REGULATION_OF_CELLULAR_BIOSYNTHETIC_PROCESS | 117 | -0.160 | -1.058 | 0.36585 | 0.36585 | 0.54665 | 156 |
| HP_ABNORMALITY_OF_THE_DENTITION | 56 | -0.189 | -1.064 | 0.36766 | 0.36766 | 0.54718 | 174 |
| GOBP_VESICLE_MEDIATED_TRANSPORT_TO_THE_PLASMA_MEMBRANE | 11 | -0.320 | -1.066 | 0.36582 | 0.36582 | 0.54665 | 385 |
| GOBP_CELLULAR_RESPONSE_TO_NITROGEN_COMPOUND | 37 | -0.209 | -1.067 | 0.37549 | 0.37549 | 0.55217 | 144 |
| GOBP_REGENERATION | 14 | -0.291 | -1.067 | 0.36252 | 0.36252 | 0.54665 | 98 |
| HP_INCREASED_HEAD_CIRCUMFERENCE | 24 | -0.239 | -1.067 | 0.37249 | 0.37249 | 0.55021 | 148 |
| HP_ABNORMAL_GLUCOSE_HOMEOSTASIS | 44 | -0.202 | -1.067 | 0.36797 | 0.36797 | 0.54718 | 229 |
| GOBP_NEGATIVE_REGULATION_OF_INTRACELLULAR_SIGNAL_TRANSDUCTION | 31 | -0.222 | -1.068 | 0.36640 | 0.36640 | 0.54694 | 100 |
| GOBP_REGULATION_OF_PROTEIN_BINDING | 14 | -0.291 | -1.068 | 0.36119 | 0.36119 | 0.54652 | 42 |
| GOBP_PEPTIDYL_LYSINE_MODIFICATION | 27 | -0.230 | -1.068 | 0.36473 | 0.36473 | 0.54665 | 810 |
| GOBP_PROTEIN_DEPHOSPHORYLATION | 22 | -0.248 | -1.068 | 0.36951 | 0.36951 | 0.54844 | 100 |
| GOMF_ENZYME_INHIBITOR_ACTIVITY | 17 | -0.270 | -1.070 | 0.35556 | 0.35556 | 0.54237 | 117 |
| GOBP_CELLULAR_RESPONSE_TO_EXTERNAL_STIMULUS | 20 | -0.255 | -1.070 | 0.36344 | 0.36344 | 0.54665 | 303 |

| | | | | | | | |
|---|---|---|---|---|---|---|---|
| HP_SLOWLY_PROGRESSIVE | 13 | -0.299 | -1.071 | 0.35655 | 0.35655 | 0.54323 | 60 |
| GOBP_CELLULAR_RESPONSE_TO_PEPTIDE | 23 | -0.244 | -1.072 | 0.36557 | 0.36557 | 0.54665 | 176 |
| GOBP_REGULATION_OF_DNA_BINDING_TRANSCRIPTION_FACTOR_ACTIVITY | 19 | -0.260 | -1.075 | 0.35005 | 0.35005 | 0.53886 | 81 |
| GOBP_APOPTOTIC_SIGNALING_PATHWAY | 46 | -0.201 | -1.076 | 0.35003 | 0.35003 | 0.53886 | 111 |
| GOBP_RESPONSE_TO_INSULIN | 13 | -0.301 | -1.078 | 0.35077 | 0.35077 | 0.53886 | 249 |
| HP_MOTOR_DELAY | 46 | -0.201 | -1.078 | 0.34676 | 0.34676 | 0.53783 | 87 |
| HP_INFLAMMATORY_ABNORMALITY_OF_THE_EYE | 11 | -0.324 | -1.081 | 0.34917 | 0.34917 | 0.53886 | 155 |
| HP_TOOTH_MALPOSITION | 18 | -0.269 | -1.083 | 0.34360 | 0.34360 | 0.53454 | 207 |
| GOBP_POSITIVE_REGULATION_OF_CATABOLIC_PROCESS | 38 | -0.212 | -1.084 | 0.34445 | 0.34445 | 0.53479 | 36 |
| HP_VASCULAR_DILATATION | 13 | -0.303 | -1.085 | 0.34307 | 0.34307 | 0.53425 | 256 |
| HP_MORTALITY_AGING | 17 | -0.274 | -1.086 | 0.33801 | 0.33801 | 0.53174 | 40 |
| HP_PROMINENT_FOREHEAD | 18 | -0.269 | -1.086 | 0.34110 | 0.34110 | 0.53332 | 80 |
| HP_APLASIA_HYPOPLASIA_OF_FINGERS | 24 | -0.244 | -1.088 | 0.34723 | 0.34723 | 0.53803 | 55 |
| HP_MYOPATHY | 20 | -0.259 | -1.089 | 0.33847 | 0.33847 | 0.53174 | 310 |
| GOBP_RESPONSE_TO_EXTRACELLULAR_STIMULUS | 20 | -0.259 | -1.090 | 0.33847 | 0.33847 | 0.53174 | 303 |
| GOBP_POSITIVE_REGULATION_OF_CELLULAR_PROTEIN_CATABOLIC_PROCESS | 13 | -0.304 | -1.090 | 0.33729 | 0.33729 | 0.53174 | 86 |
| HP_OSTEOPENIA | 11 | -0.327 | -1.091 | 0.33683 | 0.33683 | 0.53174 | 207 |
| GOMF_DNA_BINDING_TRANSCRIPTION_ACTIVATOR_ACTIVITY | 21 | -0.256 | -1.092 | 0.33534 | 0.33534 | 0.53174 | 156 |
| HP_POLYMICROGYRIA | 18 | -0.271 | -1.092 | 0.33680 | 0.33680 | 0.53174 | 162 |
| GOBP_POSITIVE_REGULATION_OF_PROTEIN_MODIFICATION_PROCESS | 71 | -0.184 | -1.093 | 0.31082 | 0.31082 | 0.51564 | 150 |
| HP_POLYDACTYLY | 17 | -0.276 | -1.094 | 0.32748 | 0.32748 | 0.52747 | 148 |
| HP_ABNORMALITY_OF_THE_PANCREAS | 21 | -0.257 | -1.096 | 0.33083 | 0.33083 | 0.52955 | 211 |
| HP_CONSTITUTIONAL_SYMPTOM | 58 | -0.193 | -1.099 | 0.32414 | 0.32414 | 0.52503 | 144 |
| GOBP_POSITIVE_REGULATION_OF_NEUROGENESIS | 11 | -0.330 | -1.099 | 0.32542 | 0.32542 | 0.52630 | 98 |
| GOBP_NEGATIVE_REGULATION_OF_CELLULAR_CATABOLIC_PROCESS | 19 | -0.266 | -1.101 | 0.32170 | 0.32170 | 0.52415 | 351 |
| GOBP_RAS_PROTEIN_SIGNAL_TRANSDUCTION | 19 | -0.267 | -1.103 | 0.31952 | 0.31952 | 0.52335 | 131 |
| GOMF_PROTEIN_DIMERIZATION_ACTIVITY | 70 | -0.186 | -1.104 | 0.29534 | 0.29534 | 0.50285 | 81 |
| GOBP_PEPTIDYL_TYROSINE_MODIFICATION | 16 | -0.286 | -1.107 | 0.32221 | 0.32221 | 0.52444 | 28 |
| GOBP_RESPONSE_TO_NITROGEN_COMPOUND | 52 | -0.201 | -1.108 | 0.31141 | 0.31141 | 0.51606 | 144 |
| GOBP_RESPONSE_TO_ENDOPLASMIC_RETICULUM_STRESS | 12 | -0.318 | -1.109 | 0.32416 | 0.32416 | 0.52503 | 414 |
| GOMF_ACTIVE_ION_TRANSMEMBRANE_TRANSPORTER_ACTIVITY | 10 | -0.346 | -1.109 | 0.31944 | 0.31944 | 0.52335 | 188 |
| HP_SHORT_DIGIT | 38 | -0.217 | -1.111 | 0.31060 | 0.31060 | 0.51564 | 59 |
| GOBP_NEGATIVE_REGULATION_OF_TRANSFERASE_ACTIVITY | 26 | -0.244 | -1.111 | 0.32084 | 0.32084 | 0.52385 | 132 |
| GOBP_NITROGEN_COMPOUND_TRANSPORT | 110 | -0.169 | -1.113 | 0.26996 | 0.26996 | 0.48568 | 147 |
| GOBP_CELLULAR_LIPID_METABOLIC_PROCESS | 47 | -0.207 | -1.113 | 0.28760 | 0.28760 | 0.50195 | 215 |
| HP_INTELLECTUAL_DISABILITY_SEVERE | 32 | -0.229 | -1.115 | 0.30532 | 0.30532 | 0.50808 | 109 |
| GOCC_NUCLEAR_ENVELOPE | 47 | -0.207 | -1.117 | 0.28485 | 0.28485 | 0.49995 | 115 |
| HP_ADDUCTED_THUMB | 10 | -0.349 | -1.120 | 0.31190 | 0.31190 | 0.51632 | 59 |
| HP_POSTERIORLY_ROTATED_EARS | 32 | -0.230 | -1.121 | 0.29986 | 0.29986 | 0.50285 | 148 |
| GOBP_REGULATION_OF_CELLULAR_RESPONSE_TO_GROWTH_FACTOR_STIMULUS | 13 | -0.314 | -1.124 | 0.29910 | 0.29910 | 0.50285 | 209 |
| HP_FATIGUE | 13 | -0.314 | -1.125 | 0.29910 | 0.29910 | 0.50285 | 173 |

| | | | | | | | |
|---|---|---|---|---|---|---|---|
| HP_ABNORMALITY_OF_CARDIOVASCULAR_SYSTEM_ELECTROPHYSIOLOGY | 32 | -0.231 | -1.125 | 0.29441 | 0.29441 | 0.50236 | 207 |
| HP_ABNORMAL_SYSTEMIC_BLOOD_PRESSURE | 24 | -0.252 | -1.125 | 0.29751 | 0.29751 | 0.50285 | 211 |
| GOMF_G_PROTEIN_COUPLED_RECEPTOR_BINDING | 13 | -0.314 | -1.125 | 0.29846 | 0.29846 | 0.50285 | 55 |
| GOBP_CANONICAL_WNT_SIGNALING_PATHWAY | 15 | -0.298 | -1.127 | 0.29292 | 0.29292 | 0.50236 | 86 |
| HP_LEUKOENCEPHALOPATHY | 18 | -0.280 | -1.130 | 0.29631 | 0.29631 | 0.50285 | 168 |
| GOBP_SIGNAL_TRANSDUCTION_BY_P53_CLASS_MEDIATOR | 16 | -0.292 | -1.131 | 0.29711 | 0.29711 | 0.50285 | 42 |
| HP_OPHTHALMOPARESIS | 20 | -0.269 | -1.131 | 0.29148 | 0.29148 | 0.50236 | 141 |
| GOBP_POSITIVE_REGULATION_OF_TRANSCRIPTION_BY_RNA_POLYMERASE_II | 66 | -0.193 | -1.132 | 0.26680 | 0.26680 | 0.48465 | 179 |
| HP_ABNORMALITY_OF_COAGULATION | 12 | -0.325 | -1.132 | 0.29819 | 0.29819 | 0.50285 | 215 |
| GOBP_POSITIVE_REGULATION_OF_IMMUNE_EFFECTOR_PROCESS | 13 | -0.317 | -1.135 | 0.29236 | 0.29236 | 0.50236 | 73 |
| HP_AORTIC_ANEURYSM | 12 | -0.326 | -1.137 | 0.29380 | 0.29380 | 0.50236 | 256 |
| GOBP_CELLULAR_NITROGEN_COMPOUND_CATABOLIC_PROCESS | 50 | -0.208 | -1.138 | 0.26724 | 0.26724 | 0.48487 | 285 |
| GOBP_LIPID_LOCALIZATION | 16 | -0.295 | -1.141 | 0.28680 | 0.28680 | 0.50195 | 215 |
| GOBP_CELL_ACTIVATION_INVOLVED_IN_IMMUNE_RESPONSE | 14 | -0.312 | -1.144 | 0.28491 | 0.28491 | 0.49995 | 176 |
| HP_GLUCOSE_INTOLERANCE | 35 | -0.227 | -1.145 | 0.26761 | 0.26761 | 0.48488 | 229 |
| HP_ABNORMAL_BLEEDING | 20 | -0.273 | -1.145 | 0.27937 | 0.27937 | 0.49359 | 155 |
| GOMF_PROTEIN_CONTAINING_COMPLEX_BINDING | 76 | -0.190 | -1.146 | 0.24263 | 0.24263 | 0.46033 | 132 |
| GOBP_STRESS_ACTIVATED_PROTEIN_KINASE_SIGNALING_CASCADE | 22 | -0.266 | -1.147 | 0.27927 | 0.27927 | 0.49359 | 74 |
| HP_ABNORMAL_INSULIN_LEVEL | 11 | -0.344 | -1.148 | 0.27853 | 0.27853 | 0.49350 | 162 |
| HP_ABNORMAL_EYEBROW_MORPHOLOGY | 33 | -0.233 | -1.148 | 0.27056 | 0.27056 | 0.48568 | 174 |
| GOBP_NEGATIVE_REGULATION_OF_BIOSYNTHETIC_PROCESS | 106 | -0.176 | -1.148 | 0.21602 | 0.21602 | 0.42910 | 274 |
| HP_ABNORMAL_ACTIVITY_OF_MITOCHONDRIAL_RESPIRATORY_CHAIN | 10 | -0.358 | -1.149 | 0.28895 | 0.28895 | 0.50236 | 674 |
| HP_GENERALIZED_HIRSUTISM | 10 | -0.359 | -1.152 | 0.28593 | 0.28593 | 0.50118 | 673 |
| GOBP_RRNA_METABOLIC_PROCESS | 21 | -0.271 | -1.155 | 0.26955 | 0.26955 | 0.48568 | 767 |
| GOBP_ORGAN_GROWTH | 11 | -0.346 | -1.155 | 0.27082 | 0.27082 | 0.48568 | 229 |
| GOBP_REGULATION_OF_T_CELL_PROLIFERATION | 10 | -0.361 | -1.158 | 0.27868 | 0.27868 | 0.49350 | 156 |
| GOCC_NUCLEAR_MEMBRANE | 31 | -0.241 | -1.162 | 0.24844 | 0.24844 | 0.46349 | 96 |
| HP_ABNORMAL_METACARPAL_MORPHOLOGY | 12 | -0.335 | -1.167 | 0.26408 | 0.26408 | 0.48142 | 15 |
| GOMF_HYDROLASE_ACTIVITY_ACTING_ON_GLYCOSYL_BONDS | 11 | -0.351 | -1.171 | 0.25386 | 0.25386 | 0.47107 | 84 |
| GOBP_REACTIVE_OXYGEN_SPECIES_METABOLIC_PROCESS | 10 | -0.365 | -1.172 | 0.26570 | 0.26570 | 0.48335 | 131 |
| HP_FLEXION_CONTRACTURE_OF_DIGIT | 21 | -0.275 | -1.173 | 0.24887 | 0.24887 | 0.46349 | 87 |
| HP_ABNORMALITY_OF_THE_GINGIVA | 10 | -0.366 | -1.174 | 0.26208 | 0.26208 | 0.48057 | 207 |
| GOBP_CELLULAR_RESPONSE_TO_ENDOGENOUS_STIMULUS | 67 | -0.201 | -1.177 | 0.20310 | 0.20310 | 0.41300 | 209 |
| GOBP_CARBOHYDRATE_DERIVATIVE_BIOSYNTHETIC_PROCESS | 37 | -0.231 | -1.178 | 0.22811 | 0.22811 | 0.44127 | 209 |
| GOMF_RIBONUCLEOPROTEIN_COMPLEX_BINDING | 10 | -0.368 | -1.180 | 0.25664 | 0.25664 | 0.47454 | 664 |
| GOBP_PROTEIN_LOCALIZATION_TO_EXTRACELLULAR_REGION | 11 | -0.354 | -1.180 | 0.24769 | 0.24769 | 0.46349 | 88 |
| GOBP_POSITIVE_REGULATION_OF_PROTEIN_CATABOLIC_PROCESS | 19 | -0.286 | -1.182 | 0.24391 | 0.24391 | 0.46033 | 31 |
| GOMF_LIPID_BINDING | 36 | -0.234 | -1.184 | 0.22430 | 0.22430 | 0.43883 | 49 |
| GOBP_REGULATION_OF_CELLULAR_AMIDE_METABOLIC_PROCESS | 26 | -0.261 | -1.187 | 0.23023 | 0.23023 | 0.44425 | 256 |
| HP_SPLENOMEGALY | 18 | -0.295 | -1.189 | 0.24221 | 0.24221 | 0.46033 | 155 |
| HP_ABNORMALITY_OF_THE_PULMONARY_VASCULATURE | 10 | -0.372 | -1.192 | 0.24275 | 0.24275 | 0.46033 | 256 |

| | | | | | | | |
|---|---|---|---|---|---|---|---|
| GOBP_RIBOSOME_BIOGENESIS | 23 | -0.272 | -1.194 | 0.22484 | 0.22484 | 0.43933 | 767 |
| GOBP_RNA_3_END_PROCESSING | 13 | -0.334 | -1.196 | 0.23492 | 0.23492 | 0.45017 | 700 |
| HP_INCREASED_CSF_LACTATE | 11 | -0.359 | -1.196 | 0.23504 | 0.23504 | 0.45017 | 674 |
| GOCC_NUCLEAR_OUTER_MEMBRANE_ENDOPLASMIC_RETICULUM_MEMBRANE_NI | 55 | -0.214 | -1.196 | 0.19252 | 0.19252 | 0.40040 | 204 |
| HP_LIMB_JOINT_CONTRACTURE | 25 | -0.266 | -1.198 | 0.22008 | 0.22008 | 0.43337 | 99 |
| GOBP_REGIONALIZATION | 14 | -0.327 | -1.201 | 0.23085 | 0.23085 | 0.44489 | 173 |
| GOMF_IDENTICAL_PROTEIN_BINDING | 120 | -0.180 | -1.202 | 0.14081 | 0.14081 | 0.33071 | 98 |
| HP_PAIN | 47 | -0.223 | -1.203 | 0.18661 | 0.18661 | 0.39291 | 144 |
| GOMF_TRANSCRIPTION_COREPRESSOR_ACTIVITY | 14 | -0.328 | -1.205 | 0.22687 | 0.22687 | 0.44050 | 706 |
| GOBP_REGULATION_OF_PEPTIDASE_ACTIVITY | 23 | -0.274 | -1.207 | 0.21030 | 0.21030 | 0.42374 | 61 |
| HP_ARTHRALGIA | 10 | -0.376 | -1.207 | 0.23219 | 0.23219 | 0.44636 | 233 |
| GOBP_POSITIVE_REGULATION_OF_PROTEIN_METABOLIC_PROCESS | 100 | -0.188 | -1.209 | 0.12989 | 0.12989 | 0.31591 | 92 |
| GOBP_POSITIVE_REGULATION_OF_RNA_METABOLIC_PROCESS | 105 | -0.186 | -1.209 | 0.13347 | 0.13347 | 0.32113 | 275 |
| HP_GENU_VALGUM | 11 | -0.363 | -1.210 | 0.22270 | 0.22270 | 0.43625 | 242 |
| GOBP_POSITIVE_REGULATION_OF_CELL_DEVELOPMENT | 17 | -0.306 | -1.212 | 0.21657 | 0.21657 | 0.42911 | 113 |
| GOCC_ORGANELLE_SUBCOMPARTMENT | 73 | -0.202 | -1.213 | 0.15816 | 0.15816 | 0.35582 | 204 |
| GOBP_CELLULAR_RESPONSE_TO_LIPID | 34 | -0.244 | -1.214 | 0.19888 | 0.19888 | 0.40761 | 80 |
| HP_ABNORMAL_FINGERTIP_MORPHOLOGY | 11 | -0.364 | -1.214 | 0.22085 | 0.22085 | 0.43427 | 80 |
| GOMF_PHOSPHATIDYLINOSITOL_BINDING | 10 | -0.381 | -1.222 | 0.21981 | 0.21981 | 0.43337 | 30 |
| GOBP_POSITIVE_REGULATION_OF_CELLULAR_COMPONENT_BIOGENESIS | 25 | -0.271 | -1.222 | 0.20205 | 0.20205 | 0.41139 | 131 |
| GOBP_NEGATIVE_REGULATION_OF_CELLULAR_AMIDE_METABOLIC_PROCESS | 19 | -0.296 | -1.223 | 0.21628 | 0.21628 | 0.42910 | 403 |
| HP_BRAIN_IMAGING_ABNORMALITY | 10 | -0.381 | -1.223 | 0.21830 | 0.21830 | 0.43144 | 292 |
| GOBP_MUSCLE_CONTRACTION | 10 | -0.382 | -1.226 | 0.21588 | 0.21588 | 0.42910 | 113 |
| GOBP_NEGATIVE_REGULATION_OF_CELL_DEATH | 54 | -0.221 | -1.227 | 0.16098 | 0.16098 | 0.35839 | 179 |
| GOBP_ACTIVATION_OF_PROTEIN_KINASE_ACTIVITY | 10 | -0.385 | -1.234 | 0.21377 | 0.21377 | 0.42766 | 92 |
| GOBP_RIBONUCLEOPROTEIN_COMPLEX_SUBUNIT_ORGANIZATION | 15 | -0.326 | -1.234 | 0.20608 | 0.20608 | 0.41713 | 709 |
| GOBP_B_CELL_ACTIVATION | 14 | -0.337 | -1.237 | 0.20498 | 0.20498 | 0.41571 | 131 |
| HP_ABNORMAL_TENDON_MORPHOLOGY | 53 | -0.223 | -1.240 | 0.15559 | 0.15559 | 0.35283 | 105 |
| GOBP_METAL_ION_TRANSPORT | 32 | -0.255 | -1.240 | 0.18401 | 0.18401 | 0.38930 | 204 |
| GOBP_EXOCYTOSIS | 15 | -0.328 | -1.241 | 0.19840 | 0.19840 | 0.40761 | 176 |
| GOBP_NEGATIVE_REGULATION_OF_RESPONSE_TO_EXTERNAL_STIMULUS | 15 | -0.328 | -1.243 | 0.19572 | 0.19572 | 0.40490 | 209 |
| GOBP_REGULATION_OF_CELLULAR_RESPONSE_TO_TRANSFORMING_GROWTH_FA( | 10 | -0.389 | -1.249 | 0.20079 | 0.20079 | 0.41044 | 209 |
| HP_KYPHOSIS | 28 | -0.267 | -1.250 | 0.17477 | 0.17477 | 0.37663 | 174 |
| HP_ABNORMALITY_OF_DENTAL_ERUPTION | 10 | -0.390 | -1.251 | 0.19837 | 0.19837 | 0.40761 | 40 |
| HP_DUPLICATION_OF_HAND_BONES | 17 | -0.316 | -1.253 | 0.18743 | 0.18743 | 0.39351 | 141 |
| GOCC_VACUOLAR_MEMBRANE | 16 | -0.324 | -1.254 | 0.18363 | 0.18363 | 0.38924 | 254 |
| GOMF_DNA_BINDING_TRANSCRIPTION_REPRESSOR_ACTIVITY | 16 | -0.324 | -1.255 | 0.18363 | 0.18363 | 0.38924 | 247 |
| GOBP_LOCALIZATION_WITHIN_MEMBRANE | 44 | -0.237 | -1.256 | 0.14087 | 0.14087 | 0.33071 | 33 |
| GOBP_REGULATION_OF_WNT_SIGNALING_PATHWAY | 18 | -0.311 | -1.257 | 0.18416 | 0.18416 | 0.38930 | 86 |
| GOBP_NEGATIVE_REGULATION_OF_HYDROLASE_ACTIVITY | 15 | -0.332 | -1.259 | 0.18303 | 0.18303 | 0.38924 | 52 |
| HP_AUTISM | 13 | -0.353 | -1.265 | 0.18806 | 0.18806 | 0.39351 | 207 |

| | | | | | | | |
|---|---|---|---|---|---|---|---|
| GOBP_PROTEIN_STABILIZATION | 11 | -0.379 | -1.266 | 0.18630 | 0.18630 | 0.39291 | 383 |
| GOBP_EXTRINSIC_APOPTOTIC_SIGNALING_PATHWAY | 11 | -0.381 | -1.273 | 0.18291 | 0.18291 | 0.38924 | 111 |
| GOBP_GROWTH | 61 | -0.222 | -1.277 | 0.11269 | 0.11269 | 0.29646 | 131 |
| GOBP_SECRETION | 33 | -0.259 | -1.277 | 0.14975 | 0.14975 | 0.34426 | 103 |
| HP_ABNORMAL_FORM_OF_THE_VERTEBRAL_BODIES | 16 | -0.331 | -1.280 | 0.16575 | 0.16575 | 0.36326 | 207 |
| HP_ABNORMAL_CRANIAL_NERVE_PHYSIOLOGY | 20 | -0.305 | -1.281 | 0.16263 | 0.16263 | 0.35964 | 148 |
| GOBP_CALCIUM_ION_TRANSMEMBRANE_TRANSPORT | 11 | -0.384 | -1.281 | 0.17736 | 0.17736 | 0.37958 | 204 |
| GOMF_PHOSPHOLIPID_BINDING | 22 | -0.297 | -1.282 | 0.14780 | 0.14780 | 0.34181 | 30 |
| GOBP_POSITIVE_REGULATION_OF_GTPASE_ACTIVITY | 18 | -0.318 | -1.284 | 0.16446 | 0.16446 | 0.36145 | 124 |
| HP_ABNORMALITY_OF_THE_TONGUE | 20 | -0.306 | -1.285 | 0.15822 | 0.15822 | 0.35582 | 162 |
| HP_ABNORMALITY_OF_CIRCULATING_ENZYME_LEVEL | 16 | -0.332 | -1.286 | 0.16197 | 0.16197 | 0.35903 | 310 |
| GOBP_REGULATION_OF_SECRETION | 20 | -0.306 | -1.286 | 0.15749 | 0.15749 | 0.35572 | 88 |
| GOBP_CELL_KILLING | 11 | -0.387 | -1.291 | 0.17057 | 0.17057 | 0.37121 | 178 |
| GOBP_LEUKOCYTE_MEDIATED_CYTOTOXICITY | 11 | -0.387 | -1.291 | 0.17057 | 0.17057 | 0.37121 | 178 |
| GOCC_SPLICEOSOMAL_COMPLEX | 13 | -0.361 | -1.291 | 0.17234 | 0.17234 | 0.37347 | 552 |
| GOBP_NCRNA_PROCESSING | 29 | -0.272 | -1.292 | 0.14384 | 0.14384 | 0.33364 | 768 |
| GOMF_PHOSPHORIC_ESTER_HYDROLASE_ACTIVITY | 28 | -0.276 | -1.292 | 0.14152 | 0.14152 | 0.33102 | 113 |
| GOMF_UBIQUITIN_LIKE_PROTEIN_LIGASE_BINDING | 31 | -0.268 | -1.293 | 0.13986 | 0.13986 | 0.33071 | 57 |
| GOBP_REGULATION_OF_INFLAMMATORY_RESPONSE | 15 | -0.342 | -1.294 | 0.15865 | 0.15865 | 0.35617 | 209 |
| GOBP_REGULATION_OF_RESPONSE_TO_STRESS | 79 | -0.212 | -1.297 | 0.08469 | 0.08469 | 0.26097 | 186 |
| GOCC_EXTERNAL_SIDE_OF_PLASMA_MEMBRANE | 12 | -0.373 | -1.298 | 0.16802 | 0.16802 | 0.36669 | 178 |
| GOBP_ESTABLISHMENT_OF_RNA_LOCALIZATION | 11 | -0.391 | -1.303 | 0.16317 | 0.16317 | 0.35964 | 641 |
| GOBP_MRNA_TRANSPORT | 11 | -0.391 | -1.303 | 0.16317 | 0.16317 | 0.35964 | 641 |
| HP_DEVIATION_OF_THE_HALLUX | 10 | -0.407 | -1.304 | 0.16274 | 0.16274 | 0.35964 | 624 |
| GOCC_INTRINSIC_COMPONENT_OF_ORGANELLE_MEMBRANE | 21 | -0.306 | -1.306 | 0.13722 | 0.13722 | 0.32553 | 421 |
| HP_OPEN_MOUTH | 10 | -0.407 | -1.307 | 0.16153 | 0.16153 | 0.35903 | 162 |
| GOBP_CELLULAR_HOMEOSTASIS | 27 | -0.282 | -1.307 | 0.12780 | 0.12780 | 0.31286 | 249 |
| GOBP_POSITIVE_REGULATION_OF_MOLECULAR_FUNCTION | 97 | -0.205 | -1.308 | 0.05701 | 0.05701 | 0.21897 | 124 |
| GOBP_NCRNA_METABOLIC_PROCESS | 31 | -0.273 | -1.313 | 0.12377 | 0.12377 | 0.30833 | 768 |
| GOBP_RESPONSE_TO_PEPTIDE | 27 | -0.284 | -1.317 | 0.12158 | 0.12158 | 0.30526 | 249 |
| GOBP_POSITIVE_REGULATION_OF_HYDROLASE_ACTIVITY | 32 | -0.271 | -1.318 | 0.12585 | 0.12585 | 0.31052 | 124 |
| GOBP_UROGENITAL_SYSTEM_DEVELOPMENT | 16 | -0.341 | -1.322 | 0.14065 | 0.14065 | 0.33071 | 45 |
| GOBP_MORPHOGENESIS_OF_AN_EPITHELIUM | 22 | -0.307 | -1.324 | 0.12447 | 0.12447 | 0.30906 | 86 |
| GOBP_GLYCOPROTEIN_BIOSYNTHETIC_PROCESS | 20 | -0.316 | -1.326 | 0.13069 | 0.13069 | 0.31591 | 192 |
| GOBP_RESPONSE_TO_ENDOGENOUS_STIMULUS | 75 | -0.220 | -1.331 | 0.07054 | 0.07054 | 0.23773 | 209 |
| GOBP_LIPID_BIOSYNTHETIC_PROCESS | 34 | -0.268 | -1.331 | 0.10950 | 0.10950 | 0.29020 | 49 |
| GOBP_IMMUNE_SYSTEM_DEVELOPMENT | 56 | -0.236 | -1.331 | 0.08323 | 0.08323 | 0.26002 | 202 |
| GOBP_CELL_CELL_SIGNALING | 58 | -0.234 | -1.333 | 0.07712 | 0.07712 | 0.24842 | 86 |
| GOMF_PHOSPHATASE_ACTIVITY | 22 | -0.310 | -1.337 | 0.11591 | 0.11591 | 0.30088 | 100 |
| GOCC_CELL_CORTEX | 22 | -0.310 | -1.337 | 0.11552 | 0.11552 | 0.30083 | 221 |
| GOBP_FAT_CELL_DIFFERENTIATION | 13 | -0.373 | -1.337 | 0.14345 | 0.14345 | 0.33364 | 343 |

| | | | | | | | |
|---|---|---|---|---|---|---|---|
| GOBP_T_CELL_PROLIFERATION | 12 | -0.386 | -1.343 | 0.14049 | 0.14049 | 0.33071 | 156 |
| GOBP_POSITIVE_REGULATION_OF_SIGNALING | 83 | -0.218 | -1.343 | 0.05849 | 0.05849 | 0.22052 | 193 |
| GOBP_REGULATION_OF_PROTEIN_MODIFICATION_PROCESS | 116 | -0.203 | -1.344 | 0.03135 | 0.03135 | 0.15205 | 92 |
| GOBP_PURINE_CONTAINING_COMPOUND_METABOLIC_PROCESS | 18 | -0.334 | -1.348 | 0.12110 | 0.12110 | 0.30526 | 608 |
| HP_ABNORMALITY_OF_THE_MUSCULATURE_OF_THE_LIMBS | 27 | -0.291 | -1.348 | 0.10041 | 0.10041 | 0.27884 | 119 |
| GOBP_REGULATION_OF_STRESS_ACTIVATED_PROTEIN_KINASE_SIGNALING_CASCA | 17 | -0.341 | -1.350 | 0.12110 | 0.12110 | 0.30526 | 74 |
| HP_MALABSORPTION | 11 | -0.405 | -1.350 | 0.13448 | 0.13448 | 0.32257 | 207 |
| GOBP_REGULATION_OF_SYSTEM_PROCESS | 14 | -0.368 | -1.351 | 0.12968 | 0.12968 | 0.31591 | 113 |
| GOBP_REGULATION_OF_VESICLE_MEDIATED_TRANSPORT | 24 | -0.303 | -1.351 | 0.10626 | 0.10626 | 0.28667 | 73 |
| HP_HAND_POLYDACTYLY | 13 | -0.378 | -1.354 | 0.13543 | 0.13543 | 0.32326 | 141 |
| GOBP_REGULATION_OF_TRANSPORTER_ACTIVITY | 17 | -0.342 | -1.355 | 0.11829 | 0.11829 | 0.30446 | 204 |
| HP_LIMB_MUSCLE_WEAKNESS | 12 | -0.389 | -1.356 | 0.13267 | 0.13267 | 0.32019 | 87 |
| GOBP_NEGATIVE_REGULATION_OF_CELLULAR_MACROMOLECULE_BIOSYNTHETIC_ | 31 | -0.282 | -1.358 | 0.09830 | 0.09830 | 0.27742 | 228 |
| GOBP_NUCLEOBASE_CONTAINING_COMPOUND_TRANSPORT | 12 | -0.391 | -1.361 | 0.13048 | 0.13048 | 0.31591 | 641 |
| GOBP_POSITIVE_REGULATION_OF_DEVELOPMENTAL_PROCESS | 55 | -0.243 | -1.362 | 0.06639 | 0.06639 | 0.23139 | 140 |
| GOBP_PROTEIN_LOCALIZATION_TO_CELL_PERIPHERY | 19 | -0.330 | -1.364 | 0.11705 | 0.11705 | 0.30227 | 31 |
| GOBP_CELL_CELL_SIGNALING_BY_WNT | 25 | -0.303 | -1.364 | 0.09754 | 0.09754 | 0.27655 | 86 |
| GOMF_RNA_POLYMERASE_II_SPECIFIC_DNA_BINDING_TRANSCRIPTION_FACTOR_I | 24 | -0.306 | -1.365 | 0.10024 | 0.10024 | 0.27884 | 354 |
| HP_ABNORMALITY_OF_THE_CHEEK | 12 | -0.393 | -1.367 | 0.12703 | 0.12703 | 0.31244 | 174 |
| HP_ABNORMALITY_OF_THE_CHIN | 26 | -0.300 | -1.368 | 0.09308 | 0.09308 | 0.27349 | 174 |
| GOBP_T_CELL_DIFFERENTIATION | 14 | -0.373 | -1.370 | 0.12139 | 0.12139 | 0.30526 | 156 |
| GOBP_NEGATIVE_REGULATION_OF_KINASE_ACTIVITY | 19 | -0.332 | -1.370 | 0.11305 | 0.11305 | 0.29689 | 74 |
| HP_LARGE_FACE | 16 | -0.354 | -1.371 | 0.11520 | 0.11520 | 0.30083 | 174 |
| GOBP_EPITHELIAL_CELL_PROLIFERATION | 14 | -0.374 | -1.372 | 0.12139 | 0.12139 | 0.30526 | 249 |
| HP_AREFLEXIA | 17 | -0.347 | -1.372 | 0.10916 | 0.10916 | 0.29020 | 310 |
| GOBP_REGULATION_OF_TRANSMEMBRANE_RECEPTOR_PROTEIN_SERINE_THREON | 14 | -0.374 | -1.373 | 0.12106 | 0.12106 | 0.30526 | 209 |
| GOBP_RNA_SPLICING_VIA_TRANSESTERIFICATION_REACTIONS | 20 | -0.327 | -1.376 | 0.10095 | 0.10095 | 0.27933 | 709 |
| GOBP_MUSCLE_TISSUE_DEVELOPMENT | 21 | -0.323 | -1.376 | 0.09925 | 0.09925 | 0.27859 | 238 |
| HP_LACTIC_ACIDOSIS | 14 | -0.375 | -1.376 | 0.11907 | 0.11907 | 0.30526 | 658 |
| HP_ABNORMAL_ATRIOVENTRICULAR_VALVE_MORPHOLOGY | 11 | -0.413 | -1.378 | 0.12215 | 0.12215 | 0.30526 | 242 |
| GOBP_CELL_CELL_ADHESION | 34 | -0.277 | -1.379 | 0.08610 | 0.08610 | 0.26185 | 186 |
| HP_ABNORMALITY_OF_THE_SEPTUM_PELLUCIDUM | 11 | -0.414 | -1.380 | 0.12122 | 0.12122 | 0.30526 | 17 |
| GOBP_REGULATION_OF_PROTEOLYSIS | 47 | -0.256 | -1.380 | 0.07245 | 0.07245 | 0.24099 | 61 |
| GOBP_RESPONSE_TO_TRANSFORMING_GROWTH_FACTOR_BETA | 13 | -0.385 | -1.380 | 0.11970 | 0.11970 | 0.30526 | 209 |
| GOBP_CATION_TRANSPORT | 42 | -0.262 | -1.380 | 0.07358 | 0.07358 | 0.24243 | 204 |
| GOBP_REGULATION_OF_DEFENSE_RESPONSE | 22 | -0.321 | -1.383 | 0.09646 | 0.09646 | 0.27570 | 176 |
| GOBP_HEART_DEVELOPMENT | 31 | -0.288 | -1.385 | 0.08311 | 0.08311 | 0.26002 | 220 |
| GOBP_ORGANONITROGEN_COMPOUND_BIOSYNTHETIC_PROCESS | 89 | -0.222 | -1.388 | 0.03752 | 0.03752 | 0.16901 | 229 |
| HP_MANDIBULAR_PROGNATHIA | 12 | -0.399 | -1.389 | 0.11421 | 0.11421 | 0.29942 | 162 |
| GOBP_CELL_SURFACE_RECEPTOR_SIGNALING_PATHWAY_INVOLVED_IN_CELL_CEL | 28 | -0.297 | -1.391 | 0.08525 | 0.08525 | 0.26106 | 86 |
| GOBP_REGULATION_OF_PROTEIN_KINASE_ACTIVITY | 58 | -0.245 | -1.392 | 0.05643 | 0.05643 | 0.21803 | 92 |

| | | | | | | | |
|---|---|---|---|---|---|---|---|
| HP_INCREASED_SERUM_LACTATE | 16 | -0.360 | -1.395 | 0.10282 | 0.10282 | 0.28248 | 674 |
| GOBP_REGULATION_OF_CELL_POPULATION_PROLIFERATION | 91 | -0.223 | -1.396 | 0.02669 | 0.02669 | 0.13966 | 84 |
| GOBP_CIRCULATORY_SYSTEM_PROCESS | 19 | -0.339 | -1.399 | 0.09706 | 0.09706 | 0.27612 | 221 |
| GOBP_CELL_GROWTH | 30 | -0.293 | -1.400 | 0.08136 | 0.08136 | 0.25677 | 131 |
| GOBP_REGULATION_OF_MAP_KINASE_ACTIVITY | 11 | -0.420 | -1.400 | 0.11073 | 0.11073 | 0.29235 | 113 |
| GOBP_POSITIVE_REGULATION_OF_PHOSPHORUS_METABOLIC_PROCESS | 51 | -0.255 | -1.403 | 0.05882 | 0.05882 | 0.22123 | 143 |
| HP_INGUINAL_HERNIA | 12 | -0.404 | -1.406 | 0.10638 | 0.10638 | 0.28667 | 340 |
| GOBP_PURINE_CONTAINING_COMPOUND_BIOSYNTHETIC_PROCESS | 11 | -0.422 | -1.409 | 0.10518 | 0.10518 | 0.28571 | 608 |
| GOBP_RIBOSE_PHOSPHATE_BIOSYNTHETIC_PROCESS | 11 | -0.422 | -1.409 | 0.10518 | 0.10518 | 0.28571 | 608 |
| GOBP_RIBONUCLEOPROTEIN_COMPLEX_BIOGENESIS | 34 | -0.283 | -1.410 | 0.06832 | 0.06832 | 0.23432 | 709 |
| HP_ABNORMAL_THUMB_MORPHOLOGY | 37 | -0.276 | -1.410 | 0.07004 | 0.07004 | 0.23695 | 79 |
| HP_INCREASED_BODY_WEIGHT | 34 | -0.284 | -1.410 | 0.06832 | 0.06832 | 0.23432 | 207 |
| GOBP_RNA_SPLICING | 22 | -0.328 | -1.414 | 0.08635 | 0.08635 | 0.26185 | 709 |
| GOBP_NEGATIVE_REGULATION_OF_CELL_CELL_ADHESION | 10 | -0.441 | -1.414 | 0.10779 | 0.10779 | 0.28846 | 209 |
| HP_EMG_ABNORMALITY | 12 | -0.406 | -1.414 | 0.10357 | 0.10357 | 0.28352 | 292 |
| GOBP_MONOSACCHARIDE_METABOLIC_PROCESS | 12 | -0.406 | -1.415 | 0.10357 | 0.10357 | 0.28352 | 343 |
| HP_ABNORMALITY_OF_THE_HALLUX | 12 | -0.407 | -1.419 | 0.10169 | 0.10169 | 0.28001 | 624 |
| GOBP_GLYCOPROTEIN_METABOLIC_PROCESS | 21 | -0.334 | -1.422 | 0.07707 | 0.07707 | 0.24842 | 192 |
| HP_CORNEAL_OPACITY | 18 | -0.352 | -1.422 | 0.08599 | 0.08599 | 0.26185 | 105 |
| HP_STEREOTYPY | 16 | -0.368 | -1.423 | 0.08838 | 0.08838 | 0.26429 | 176 |
| GOBP_JNK_CASCADE | 14 | -0.388 | -1.424 | 0.09652 | 0.09652 | 0.27570 | 57 |
| GOCC_VESICLE_LUMEN | 16 | -0.368 | -1.425 | 0.08700 | 0.08700 | 0.26227 | 60 |
| GOBP_POSITIVE_REGULATION_OF_STRESS_ACTIVATED_PROTEIN_KINASE_SIGNALIN | 11 | -0.427 | -1.425 | 0.10025 | 0.10025 | 0.27884 | 57 |
| GOBP_RESPONSE_TO_LIPID | 48 | -0.264 | -1.427 | 0.05257 | 0.05257 | 0.20941 | 84 |
| GOBP_CELLULAR_RESPONSE_TO_STARVATION | 10 | -0.445 | -1.429 | 0.10115 | 0.10115 | 0.27937 | 484 |
| HP_PROXIMAL_MUSCLE_WEAKNESS | 11 | -0.429 | -1.430 | 0.09809 | 0.09809 | 0.27734 | 87 |
| GOBP_HOMEOSTATIC_PROCESS | 67 | -0.244 | -1.431 | 0.03879 | 0.03879 | 0.17122 | 316 |
| GOBP_POSITIVE_REGULATION_OF_PROTEOLYSIS | 24 | -0.321 | -1.433 | 0.07257 | 0.07257 | 0.24099 | 86 |
| GOBP_EPITHELIUM_DEVELOPMENT | 53 | -0.258 | -1.434 | 0.04779 | 0.04779 | 0.19803 | 226 |
| GOBP_NEGATIVE_REGULATION_OF_LOCOMOTION | 13 | -0.400 | -1.434 | 0.09596 | 0.09596 | 0.27570 | 74 |
| GOBP_POSITIVE_REGULATION_OF_RESPONSE_TO_EXTERNAL_STIMULUS | 12 | -0.412 | -1.435 | 0.09543 | 0.09543 | 0.27570 | 130 |
| GOBP_SMALL_GTPASE_MEDIATED_SIGNAL_TRANSDUCTION | 29 | -0.303 | -1.436 | 0.06655 | 0.06655 | 0.23139 | 131 |
| GOMF_PROTEIN_SERINE_THREONINE_PHOSPHATASE_ACTIVITY | 10 | -0.448 | -1.438 | 0.09662 | 0.09662 | 0.27570 | 100 |
| GOBP_NEGATIVE_REGULATION_OF_GENE_EXPRESSION | 65 | -0.246 | -1.438 | 0.03870 | 0.03870 | 0.17122 | 256 |
| HP_ABNORMAL_BLOOD_GLUCOSE_CONCENTRATION | 17 | -0.365 | -1.443 | 0.07898 | 0.07898 | 0.25179 | 670 |
| GOCC_CELL_PROJECTION_MEMBRANE | 11 | -0.433 | -1.446 | 0.09007 | 0.09007 | 0.26769 | 212 |
| GOBP_POSITIVE_REGULATION_OF_INTRACELLULAR_SIGNAL_TRANSDUCTION | 44 | -0.274 | -1.449 | 0.05463 | 0.05463 | 0.21269 | 186 |
| GOMF_PHOSPHOPROTEIN_PHOSPHATASE_ACTIVITY | 15 | -0.383 | -1.450 | 0.08517 | 0.08517 | 0.26106 | 100 |
| HP_ABNORMALITY_OF_THE_MITOCHONDRION | 20 | -0.346 | -1.454 | 0.07085 | 0.07085 | 0.23773 | 690 |
| GOBP_IMMUNE_EFFECTOR_PROCESS | 30 | -0.305 | -1.456 | 0.06037 | 0.06037 | 0.22146 | 80 |
| GOCC_INNER_MITOCHONDRIAL_MEMBRANE_PROTEIN_COMPLEX | 18 | -0.361 | -1.456 | 0.07058 | 0.07058 | 0.23773 | 674 |

| | | | | | | | |
|---|---|---|---|---|---|---|---|
| GOMF_PROTEIN_HOMODIMERIZATION_ACTIVITY | 46 | -0.273 | -1.463 | 0.04573 | 0.04573 | 0.19321 | 73 |
| GOBP_TISSUE_MORPHOGENESIS | 24 | -0.328 | -1.463 | 0.06415 | 0.06415 | 0.22695 | 86 |
| GOBP_POSITIVE_REGULATION_OF_TRANSPORT | 41 | -0.280 | -1.465 | 0.04541 | 0.04541 | 0.19290 | 88 |
| HP_ABNORMAL_ORAL_MUCOSA_MORPHOLOGY | 15 | -0.387 | -1.465 | 0.07782 | 0.07782 | 0.24958 | 207 |
| GOBP_REGULATION_OF_T_CELL_ACTIVATION | 16 | -0.379 | -1.467 | 0.07634 | 0.07634 | 0.24748 | 156 |
| GOBP_PEPTIDYL_TYROSINE_DEPHOSPHORYLATION | 13 | -0.410 | -1.467 | 0.08408 | 0.08408 | 0.26084 | 100 |
| GOMF_PROTEIN_TYROSINE_PHOSPHATASE_ACTIVITY | 13 | -0.410 | -1.467 | 0.08408 | 0.08408 | 0.26084 | 100 |
| HP_CONTRACTURES_OF_THE_JOINTS_OF_THE_LOWER_LIMBS | 10 | -0.457 | -1.467 | 0.08514 | 0.08514 | 0.26106 | 87 |
| HP_MICRODONTIA | 10 | -0.458 | -1.469 | 0.08454 | 0.08454 | 0.26097 | 43 |
| GOBP_NEGATIVE_REGULATION_OF_CELL_POPULATION_PROLIFERATION | 35 | -0.294 | -1.478 | 0.04681 | 0.04681 | 0.19530 | 131 |
| GOCC_ANCHORING_JUNCTION | 38 | -0.289 | -1.479 | 0.04480 | 0.04480 | 0.19122 | 142 |
| GOCC_SECRETORY_VESICLE | 47 | -0.275 | -1.482 | 0.03952 | 0.03952 | 0.17261 | 221 |
| GOCC_CELL_SURFACE | 27 | -0.321 | -1.488 | 0.05104 | 0.05104 | 0.20594 | 198 |
| GOBP_POSITIVE_REGULATION_OF_PROTEIN_PHOSPHORYLATION | 48 | -0.276 | -1.492 | 0.04027 | 0.04027 | 0.17437 | 143 |
| GOBP_REGULATION_OF_JNK_CASCADE | 12 | -0.429 | -1.493 | 0.07635 | 0.07635 | 0.24748 | 57 |
| GOBP_REGULATION_OF_LEUKOCYTE_PROLIFERATION | 14 | -0.407 | -1.493 | 0.07098 | 0.07098 | 0.23773 | 156 |
| GOBP_EXPORT_FROM_CELL | 28 | -0.319 | -1.493 | 0.04689 | 0.04689 | 0.19530 | 103 |
| GOBP_CARBOHYDRATE_HOMEOSTASIS | 10 | -0.466 | -1.494 | 0.07548 | 0.07548 | 0.24624 | 563 |
| GOBP_REGULATION_OF_HEMOPOIESIS | 21 | -0.351 | -1.494 | 0.05564 | 0.05564 | 0.21607 | 156 |
| GOBP_SIGNAL_RELEASE | 17 | -0.378 | -1.498 | 0.05897 | 0.05897 | 0.22123 | 176 |
| GOBP_TRANSFORMING_GROWTH_FACTOR_BETA_RECEPTOR_SIGNALING_PATHWA | 12 | -0.433 | -1.508 | 0.06852 | 0.06852 | 0.23432 | 209 |
| GOBP_LEUKOCYTE_PROLIFERATION | 18 | -0.374 | -1.508 | 0.05733 | 0.05733 | 0.21897 | 156 |
| GOBP_MUSCLE_SYSTEM_PROCESS | 14 | -0.411 | -1.509 | 0.06501 | 0.06501 | 0.22893 | 209 |
| GOMF_PEPTIDE_BINDING | 13 | -0.422 | -1.512 | 0.06290 | 0.06290 | 0.22400 | 128 |
| GOBP_EPITHELIAL_TUBE_MORPHOGENESIS | 17 | -0.382 | -1.513 | 0.05370 | 0.05370 | 0.21134 | 221 |
| HP_INTELLECTUAL_DISABILITY_MODERATE | 15 | -0.401 | -1.518 | 0.06079 | 0.06079 | 0.22215 | 174 |
| GOBP_REGULATION_OF_PHOSPHORUS_METABOLIC_PROCESS | 92 | -0.242 | -1.528 | 0.00720 | 0.00720 | 0.06037 | 92 |
| GOBP_TISSUE_DEVELOPMENT | 93 | -0.243 | -1.529 | 0.01065 | 0.01065 | 0.07401 | 226 |
| GOBP_MRNA_PROCESSING | 27 | -0.330 | -1.529 | 0.03942 | 0.03942 | 0.17261 | 709 |
| GOBP_POSITIVE_REGULATION_OF_LOCOMOTION | 21 | -0.360 | -1.533 | 0.04662 | 0.04662 | 0.19524 | 130 |
| GOBP_CELLULAR_RESPONSE_TO_OXYGEN_CONTAINING_COMPOUND | 61 | -0.268 | -1.536 | 0.02137 | 0.02137 | 0.12043 | 81 |
| GOCC_ORGANELLE_INNER_MEMBRANE | 49 | -0.283 | -1.537 | 0.02753 | 0.02753 | 0.14124 | 763 |
| GOBP_REGULATION_OF_EPITHELIAL_CELL_MIGRATION | 11 | -0.461 | -1.539 | 0.06262 | 0.06262 | 0.22381 | 80 |
| GOBP_RESPONSE_TO_STARVATION | 11 | -0.462 | -1.541 | 0.06169 | 0.06169 | 0.22281 | 343 |
| HP_ABNORMAL_TONGUE_MORPHOLOGY | 17 | -0.390 | -1.544 | 0.04914 | 0.04914 | 0.20090 | 242 |
| GOBP_TRANSMEMBRANE_RECEPTOR_PROTEIN_TYROSINE_KINASE_SIGNALING_PA | 24 | -0.346 | -1.544 | 0.04050 | 0.04050 | 0.17479 | 180 |
| GOBP_NEGATIVE_REGULATION_OF_MAPK_CASCADE | 13 | -0.432 | -1.547 | 0.05167 | 0.05167 | 0.20793 | 100 |
| HP_SKELETAL_MUSCLE_HYPERTROPHY | 11 | -0.464 | -1.548 | 0.06046 | 0.06046 | 0.22146 | 207 |
| GOBP_NEGATIVE_REGULATION_OF_PROTEIN_METABOLIC_PROCESS | 76 | -0.256 | -1.549 | 0.02279 | 0.02279 | 0.12524 | 82 |
| GOBP_INTRINSIC_APOPTOTIC_SIGNALING_PATHWAY_BY_P53_CLASS_MEDIATOR | 10 | -0.483 | -1.550 | 0.06008 | 0.06008 | 0.22146 | 42 |
| GOBP_RNA_CATABOLIC_PROCESS | 33 | -0.314 | -1.552 | 0.03032 | 0.03032 | 0.14960 | 403 |

| | | | | | | | |
|---|---|---|---|---|---|---|---|
| HP_ECLABION | 11 | -0.466 | -1.555 | 0.05768 | 0.05768 | 0.21905 | 46 |
| GOBP_LEUKOCYTE_CELL_CELL_ADHESION | 16 | -0.404 | -1.563 | 0.04780 | 0.04780 | 0.19803 | 253 |
| GOBP_VASCULAR_PROCESS_IN_CIRCULATORY_SYSTEM | 10 | -0.489 | -1.568 | 0.05616 | 0.05616 | 0.21754 | 188 |
| GOBP_POSITIVE_REGULATION_OF_LEUKOCYTE_CELL_CELL_ADHESION | 12 | -0.450 | -1.568 | 0.05382 | 0.05382 | 0.21134 | 73 |
| GOCC_ENDOSOME | 49 | -0.290 | -1.576 | 0.02135 | 0.02135 | 0.12043 | 299 |
| GOMF_SIGNALING_RECEPTOR_BINDING | 52 | -0.287 | -1.580 | 0.01804 | 0.01804 | 0.10850 | 131 |
| GOBP_MYELOID_CELL_DIFFERENTIATION | 21 | -0.371 | -1.581 | 0.03797 | 0.03797 | 0.17004 | 202 |
| GOBP_POSITIVE_REGULATION_OF_KINASE_ACTIVITY | 31 | -0.328 | -1.582 | 0.02592 | 0.02592 | 0.13612 | 113 |
| GOBP_NEGATIVE_REGULATION_OF_CYTOKINE_PRODUCTION | 10 | -0.494 | -1.584 | 0.05284 | 0.05284 | 0.20993 | 132 |
| GOBP_REGULATION_OF_CELL_DIFFERENTIATION | 76 | -0.262 | -1.585 | 0.01776 | 0.01776 | 0.10834 | 159 |
| GOBP_EXTERNAL_ENCAPSULATING_STRUCTURE_ORGANIZATION | 10 | -0.494 | -1.585 | 0.05254 | 0.05254 | 0.20941 | 31 |
| GOBP_REGULATION_OF_GTPASE_ACTIVITY | 22 | -0.368 | -1.587 | 0.03656 | 0.03656 | 0.16782 | 124 |
| GOBP_METAL_ION_HOMEOSTASIS | 17 | -0.401 | -1.588 | 0.04107 | 0.04107 | 0.17676 | 229 |
| GOCC_GOLGI_APPARATUS_SUBCOMPARTMENT | 22 | -0.369 | -1.589 | 0.03656 | 0.03656 | 0.16782 | 33 |
| GOBP_POSITIVE_REGULATION_OF_DNA_BINDING_TRANSCRIPTION_FACTOR_ACTIV | 10 | -0.496 | -1.590 | 0.05193 | 0.05193 | 0.20793 | 81 |
| GOBP_VESICLE_MEDIATED_TRANSPORT | 80 | -0.260 | -1.591 | 0.00484 | 0.00484 | 0.04536 | 217 |
| GOBP_CELLULAR_ION_HOMEOSTASIS | 19 | -0.385 | -1.592 | 0.03671 | 0.03671 | 0.16782 | 237 |
| GOBP_MUSCLE_ORGAN_DEVELOPMENT | 19 | -0.386 | -1.593 | 0.03635 | 0.03635 | 0.16782 | 249 |
| GOBP_REGULATION_OF_CELL_MORPHOGENESIS | 12 | -0.458 | -1.595 | 0.04662 | 0.04662 | 0.19524 | 154 |
| GOBP_POSITIVE_REGULATION_OF_PROTEIN_KINASE_ACTIVITY | 30 | -0.334 | -1.595 | 0.02012 | 0.02012 | 0.11591 | 113 |
| GOMF_MRNA_BINDING | 22 | -0.371 | -1.598 | 0.03423 | 0.03423 | 0.16268 | 344 |
| GOMF_AMIDE_BINDING | 21 | -0.377 | -1.604 | 0.03346 | 0.03346 | 0.16001 | 128 |
| HP_RETROGNATHIA | 19 | -0.389 | -1.607 | 0.03272 | 0.03272 | 0.15743 | 87 |
| GOCC_SECRETORY_GRANULE | 38 | -0.314 | -1.609 | 0.01941 | 0.01941 | 0.11308 | 221 |
| GOBP_I_KAPPAB_KINASE_NF_KAPPAB_SIGNALING | 14 | -0.439 | -1.609 | 0.03847 | 0.03847 | 0.17082 | 143 |
| GOBP_REGULATION_OF_ACTIN_FILAMENT_BASED_PROCESS | 16 | -0.416 | -1.611 | 0.03955 | 0.03955 | 0.17261 | 66 |
| GOBP_REGULATION_OF_MYELOID_CELL_DIFFERENTIATION | 14 | -0.440 | -1.613 | 0.03781 | 0.03781 | 0.16982 | 132 |
| HP_ABNORMALITY_OF_ACID_BASE_HOMEOSTASIS | 24 | -0.362 | -1.615 | 0.02686 | 0.02686 | 0.13966 | 675 |
| GOMF_GTPASE_ACTIVITY | 13 | -0.451 | -1.617 | 0.03723 | 0.03723 | 0.16817 | 172 |
| GOBP_RNA_PROCESSING | 59 | -0.284 | -1.621 | 0.01687 | 0.01687 | 0.10416 | 709 |
| GOBP_NUCLEAR_TRANSCRIBED_MRNA_CATABOLIC_PROCESS_DEADENYLATION_D | 11 | -0.486 | -1.623 | 0.03825 | 0.03825 | 0.17038 | 325 |
| GOBP_NEGATIVE_REGULATION_OF_CELL_ADHESION | 18 | -0.404 | -1.629 | 0.03117 | 0.03117 | 0.15205 | 212 |
| GOBP_POSITIVE_REGULATION_OF_MRNA_METABOLIC_PROCESS | 13 | -0.455 | -1.629 | 0.03466 | 0.03466 | 0.16324 | 484 |
| GOBP_REGULATION_OF_HYDROLASE_ACTIVITY | 54 | -0.293 | -1.631 | 0.01436 | 0.01436 | 0.09196 | 128 |
| GOBP_BIOLOGICAL_ADHESION | 59 | -0.287 | -1.642 | 0.01380 | 0.01380 | 0.08988 | 113 |
| GOCC_SIDE_OF_MEMBRANE | 17 | -0.415 | -1.642 | 0.02913 | 0.02913 | 0.14562 | 204 |
| HP_APLASIA_HYPOPLASIA_OF_THE_SKIN | 12 | -0.475 | -1.654 | 0.03285 | 0.03285 | 0.15760 | 155 |
| GOBP_REGULATION_OF_PROTEIN_PHOSPHORYLATION | 80 | -0.271 | -1.658 | 0.00212 | 0.00212 | 0.02571 | 92 |
| GOBP_ENDOSOMAL_TRANSPORT | 14 | -0.453 | -1.660 | 0.02985 | 0.02985 | 0.14859 | 463 |
| GOBP_NEGATIVE_REGULATION_OF_PROTEIN_MODIFICATION_PROCESS | 41 | -0.318 | -1.661 | 0.01122 | 0.01122 | 0.07662 | 77 |
| GOBP_REGULATION_OF_CELL_DEATH | 93 | -0.265 | -1.671 | 0.00301 | 0.00301 | 0.03172 | 140 |

| | | | | | | | |
|---|---|---|---|---|---|---|---|
| GOBP_RNA_DESTABILIZATION | 12 | -0.480 | -1.672 | 0.03129 | 0.03129 | 0.15205 | 452 |
| GOBP_AMEBOIDAL_TYPE_CELL_MIGRATION | 19 | -0.405 | -1.673 | 0.02399 | 0.02399 | 0.13043 | 111 |
| GOBP_PROTEIN_KINASE_B_SIGNALING | 11 | -0.502 | -1.673 | 0.02992 | 0.02992 | 0.14859 | 28 |
| GOBP_NEGATIVE_REGULATION_OF_MULTICELLULAR_ORGANISMAL_PROCESS | 36 | -0.331 | -1.674 | 0.01393 | 0.01393 | 0.09030 | 229 |
| GOBP_IMMUNE_RESPONSE_REGULATING_CELL_SURFACE_RECEPTOR_SIGNALING_ | 10 | -0.523 | -1.677 | 0.03140 | 0.03140 | 0.15205 | 84 |
| GOBP_BIOLOGICAL_PROCESS_INVOLVED_IN_SYMBIOTIC_INTERACTION | 11 | -0.503 | -1.678 | 0.02869 | 0.02869 | 0.14450 | 35 |
| GOBP_DIVALENT_INORGANIC_CATION_HOMEOSTASIS | 15 | -0.443 | -1.679 | 0.02872 | 0.02872 | 0.14450 | 204 |
| GOBP_REGULATION_OF_ANATOMICAL_STRUCTURE_MORPHOGENESIS | 46 | -0.313 | -1.679 | 0.00877 | 0.00877 | 0.06661 | 180 |
| GOBP_POSTTRANSCRIPTIONAL_REGULATION_OF_GENE_EXPRESSION | 33 | -0.340 | -1.681 | 0.01148 | 0.01148 | 0.07804 | 258 |
| GOBP_RIBOSE_PHOSPHATE_METABOLIC_PROCESS | 17 | -0.425 | -1.683 | 0.02317 | 0.02317 | 0.12684 | 608 |
| GOBP_CELL_MIGRATION | 65 | -0.288 | -1.683 | 0.00494 | 0.00494 | 0.04560 | 195 |
| GOBP_CIRCULATORY_SYSTEM_DEVELOPMENT | 54 | -0.303 | -1.687 | 0.00854 | 0.00854 | 0.06519 | 209 |
| GOBP_REGULATION_OF_VASCULATURE_DEVELOPMENT | 11 | -0.506 | -1.688 | 0.02745 | 0.02745 | 0.14124 | 29 |
| GOMF_SULFUR_COMPOUND_BINDING | 20 | -0.402 | -1.688 | 0.02056 | 0.02056 | 0.11754 | 55 |
| GOBP_PROTEIN_LOCALIZATION_TO_PLASMA_MEMBRANE | 15 | -0.447 | -1.692 | 0.02705 | 0.02705 | 0.13973 | 31 |
| GOBP_RESPONSE_TO_GROWTH_FACTOR | 33 | -0.344 | -1.697 | 0.01056 | 0.01056 | 0.07401 | 238 |
| GOBP_REGULATION_OF_RESPONSE_TO_EXTERNAL_STIMULUS | 32 | -0.349 | -1.698 | 0.01227 | 0.01227 | 0.08193 | 176 |
| GOBP_MUSCLE_CELL_PROLIFERATION | 12 | -0.488 | -1.700 | 0.02785 | 0.02785 | 0.14240 | 80 |
| GOBP_ION_HOMEOSTASIS | 21 | -0.400 | -1.705 | 0.02068 | 0.02068 | 0.11754 | 237 |
| GOBP_REGULATION_OF_RESPONSE_TO_BIOTIC_STIMULUS | 11 | -0.511 | -1.706 | 0.02591 | 0.02591 | 0.13612 | 170 |
| GOBP_POSITIVE_REGULATION_OF_CELL_CELL_ADHESION | 15 | -0.452 | -1.713 | 0.02505 | 0.02505 | 0.13428 | 73 |
| GOCC_EXTERNAL_ENCAPSULATING_STRUCTURE | 17 | -0.433 | -1.716 | 0.02071 | 0.02071 | 0.11754 | 119 |
| HP_ABNORMAL_PERIPHERAL_NERVOUS_SYSTEM_MORPHOLOGY | 26 | -0.377 | -1.717 | 0.01112 | 0.01112 | 0.07635 | 119 |
| HP_DILATED_CARDIOMYOPATHY | 10 | -0.536 | -1.718 | 0.02566 | 0.02566 | 0.13572 | 200 |
| GOBP_ENZYME_LINKED_RECEPTOR_PROTEIN_SIGNALING_PATHWAY | 39 | -0.333 | -1.718 | 0.00979 | 0.00979 | 0.07090 | 180 |
| GOBP_REGULATION_OF_HORMONE_LEVELS | 13 | -0.480 | -1.721 | 0.01926 | 0.01926 | 0.11259 | 84 |
| GOBP_CALCIUM_ION_TRANSPORT | 17 | -0.435 | -1.722 | 0.01966 | 0.01966 | 0.11364 | 204 |
| GOBP_REGULATION_OF_LYMPHOCYTE_ACTIVATION | 22 | -0.400 | -1.723 | 0.01478 | 0.01478 | 0.09425 | 170 |
| GOBP_LOCOMOTION | 74 | -0.287 | -1.728 | 0.00167 | 0.00167 | 0.02217 | 195 |
| GOBP_TRANSMEMBRANE_RECEPTOR_PROTEIN_SERINE_THREONINE_KINASE_SIGN | 18 | -0.429 | -1.729 | 0.01863 | 0.01863 | 0.10935 | 238 |
| GOBP_NUCLEAR_TRANSCRIBED_MRNA_CATABOLIC_PROCESS | 16 | -0.447 | -1.731 | 0.01754 | 0.01754 | 0.10741 | 403 |
| GOBP_CHEMICAL_HOMEOSTASIS | 40 | -0.335 | -1.736 | 0.01568 | 0.01568 | 0.09874 | 249 |
| GOCC_CELL_SUBSTRATE_JUNCTION | 17 | -0.438 | -1.736 | 0.01860 | 0.01860 | 0.10935 | 161 |
| GOBP_ACTIVATION_OF_IMMUNE_RESPONSE | 10 | -0.542 | -1.740 | 0.02234 | 0.02234 | 0.12320 | 84 |
| GOCC_ENDOSOME_MEMBRANE | 22 | -0.405 | -1.744 | 0.01167 | 0.01167 | 0.07895 | 253 |
| GOCC_TRANS_GOLGI_NETWORK | 16 | -0.451 | -1.744 | 0.01685 | 0.01685 | 0.10416 | 21 |
| GOCC_SECRETORY_GRANULE_MEMBRANE | 12 | -0.503 | -1.753 | 0.02065 | 0.02065 | 0.11754 | 198 |
| GOBP_MAPK_CASCADE | 45 | -0.332 | -1.775 | 0.00357 | 0.00357 | 0.03565 | 133 |
| HP_APRAXIA | 15 | -0.469 | -1.775 | 0.01603 | 0.01603 | 0.10017 | 562 |
| GOBP_REGULATION_OF_IMMUNE_RESPONSE | 34 | -0.359 | -1.784 | 0.00889 | 0.00889 | 0.06688 | 265 |
| GOBP_POSITIVE_REGULATION_OF_I_KAPPAB_KINASE_NF_KAPPAB_SIGNALING | 10 | -0.556 | -1.784 | 0.01812 | 0.01812 | 0.10850 | 143 |

| | | | | | | | |
|---|---|---|---|---|---|---|---|
| GOBP_INFLAMMATORY_RESPONSE | 28 | -0.381 | -1.785 | 0.00767 | 0.00767 | 0.06189 | 132 |
| GOMF_MOLECULAR_TRANSDUCER_ACTIVITY | 29 | -0.377 | -1.789 | 0.00988 | 0.00988 | 0.07110 | 128 |
| GOBP_ANATOMICAL_STRUCTURE_FORMATION_INVOLVED_IN_MORPHOGENESIS | 49 | -0.331 | -1.799 | 0.00183 | 0.00183 | 0.02384 | 180 |
| HP_SCARRING | 13 | -0.503 | -1.800 | 0.01059 | 0.01059 | 0.07401 | 141 |
| GOBP_PHAGOCYTOSIS | 11 | -0.541 | -1.806 | 0.01265 | 0.01265 | 0.08374 | 128 |
| GOBP_REGULATION_OF_BODY_FLUID_LEVELS | 20 | -0.431 | -1.811 | 0.00991 | 0.00991 | 0.07110 | 202 |
| GOBP_REGULATION_OF_CATION_CHANNEL_ACTIVITY | 10 | -0.566 | -1.816 | 0.01329 | 0.01329 | 0.08722 | 204 |
| GOBP_REGULATION_OF_CALCIUM_ION_TRANSPORT | 12 | -0.523 | -1.823 | 0.01283 | 0.01283 | 0.08459 | 130 |
| GOBP_RESPONSE_TO_TUMOR_NECROSIS_FACTOR | 12 | -0.526 | -1.832 | 0.01095 | 0.01095 | 0.07575 | 139 |
| GOCC_ENDOPLASMIC_RETICULUM | 88 | -0.293 | -1.833 | 0.00058 | 0.00058 | 0.01001 | 144 |
| GOBP_REGULATION_OF_LEUKOCYTE_DIFFERENTIATION | 11 | -0.552 | -1.842 | 0.00956 | 0.00956 | 0.06994 | 80 |
| GOBP_REGULATION_OF_MRNA_CATABOLIC_PROCESS | 20 | -0.439 | -1.845 | 0.00771 | 0.00771 | 0.06189 | 285 |
| GOBP_RECEPTOR_MEDIATED_ENDOCYTOSIS | 12 | -0.531 | -1.849 | 0.01033 | 0.01033 | 0.07339 | 284 |
| GOCC_VESICLE_MEMBRANE | 52 | -0.336 | -1.850 | 0.00261 | 0.00261 | 0.02955 | 212 |
| GOBP_ERK1_AND_ERK2_CASCADE | 11 | -0.556 | -1.854 | 0.00956 | 0.00956 | 0.06994 | 100 |
| GOBP_RESPONSE_TO_OXYGEN_CONTAINING_COMPOUND | 78 | -0.305 | -1.858 | 0.00090 | 0.00090 | 0.01430 | 144 |
| GOBP_RESPONSE_TO_MECHANICAL_STIMULUS | 10 | -0.579 | -1.859 | 0.00936 | 0.00936 | 0.06977 | 265 |
| GOBP_IMMUNE_RESPONSE_REGULATING_SIGNALING_PATHWAY | 16 | -0.480 | -1.859 | 0.00653 | 0.00653 | 0.05624 | 217 |
| GOBP_POSITIVE_REGULATION_OF_ION_TRANSPORT | 10 | -0.582 | -1.866 | 0.00845 | 0.00845 | 0.06517 | 58 |
| GOBP_TISSUE_MIGRATION | 16 | -0.482 | -1.867 | 0.00619 | 0.00619 | 0.05388 | 80 |
| GOBP_POSITIVE_REGULATION_OF_CELL_DIFFERENTIATION | 36 | -0.370 | -1.872 | 0.00372 | 0.00372 | 0.03629 | 140 |
| GOBP_NEGATIVE_REGULATION_OF_IMMUNE_SYSTEM_PROCESS | 17 | -0.473 | -1.875 | 0.00737 | 0.00737 | 0.06043 | 203 |
| GOBP_TUBE_DEVELOPMENT | 45 | -0.350 | -1.876 | 0.00136 | 0.00136 | 0.01921 | 86 |
| GOBP_REGULATION_OF_CATION_TRANSMEMBRANE_TRANSPORT | 17 | -0.475 | -1.882 | 0.00737 | 0.00737 | 0.06043 | 204 |
| HP_ABNORMALITY_OF_FACIAL_MUSCULATURE | 10 | -0.590 | -1.891 | 0.00725 | 0.00725 | 0.06037 | 87 |
| HP_ABNORMALITY_OF_DENTAL_MORPHOLOGY | 22 | -0.440 | -1.895 | 0.00828 | 0.00828 | 0.06444 | 50 |
| GOBP_NEGATIVE_REGULATION_OF_RESPONSE_TO_STIMULUS | 83 | -0.307 | -1.896 | 0.00060 | 0.00060 | 0.01024 | 132 |
| GOCC_LATE_ENDOSOME | 10 | -0.591 | -1.897 | 0.00725 | 0.00725 | 0.06037 | 433 |
| GOBP_MRNA_METABOLIC_PROCESS | 51 | -0.345 | -1.899 | 0.00217 | 0.00217 | 0.02610 | 578 |
| GOBP_IMMUNE_RESPONSE | 63 | -0.330 | -1.911 | 0.00028 | 0.00028 | 0.00593 | 183 |
| GOBP_POSITIVE_REGULATION_OF_IMMUNE_RESPONSE | 19 | -0.463 | -1.914 | 0.00854 | 0.00854 | 0.06519 | 84 |
| GOBP_POSITIVE_REGULATION_OF_MAPK_CASCADE | 21 | -0.451 | -1.921 | 0.00488 | 0.00488 | 0.04536 | 133 |
| GOBP_DEFENSE_RESPONSE | 55 | -0.344 | -1.925 | 0.00070 | 0.00070 | 0.01149 | 183 |
| GOBP_REGULATION_OF_CELLULAR_COMPONENT_MOVEMENT | 42 | -0.368 | -1.938 | 0.00139 | 0.00139 | 0.01934 | 124 |
| GOCC_BASOLATERAL_PLASMA_MEMBRANE | 10 | -0.604 | -1.939 | 0.00604 | 0.00604 | 0.05316 | 204 |
| GOBP_EPIDERMIS_DEVELOPMENT | 12 | -0.557 | -1.940 | 0.00532 | 0.00532 | 0.04764 | 220 |
| GOCC_ENDOPLASMIC_RETICULUM_LUMEN | 11 | -0.582 | -1.941 | 0.00524 | 0.00524 | 0.04746 | 75 |
| GOBP_VASCULATURE_DEVELOPMENT | 30 | -0.407 | -1.942 | 0.00311 | 0.00311 | 0.03229 | 45 |
| GOCC_COLLAGEN_CONTAINING_EXTRACELLULAR_MATRIX | 15 | -0.513 | -1.944 | 0.00618 | 0.00618 | 0.05388 | 119 |
| HP_ABNORMALITY_OF_THE_MUSCULATURE_OF_THE_UPPER_LIMBS | 16 | -0.503 | -1.945 | 0.00783 | 0.00783 | 0.06250 | 119 |
| HP_ABNORMAL_CRANIAL_NERVE_MORPHOLOGY | 10 | -0.608 | -1.952 | 0.00513 | 0.00513 | 0.04678 | 87 |

| | | | | | | | |
|---|---|---|---|---|---|---|---|
| GOBP_T_CELL_ACTIVATION | 25 | -0.434 | -1.957 | 0.00355 | 0.00355 | 0.03565 | 178 |
| GOBP_MULTICELLULAR_ORGANISMAL_HOMEOSTASIS | 22 | -0.455 | -1.959 | 0.00541 | 0.00541 | 0.04787 | 316 |
| GOBP_MRNA_CATABOLIC_PROCESS | 27 | -0.423 | -1.961 | 0.00257 | 0.00257 | 0.02945 | 403 |
| GOBP_REGULATION_OF_METAL_ION_TRANSPORT | 20 | -0.467 | -1.964 | 0.00254 | 0.00254 | 0.02936 | 204 |
| GOMF_GUANYL_NUCLEOTIDE_BINDING | 15 | -0.521 | -1.972 | 0.00489 | 0.00489 | 0.04536 | 172 |
| GOBP_BLOOD_VESSEL_MORPHOGENESIS | 23 | -0.450 | -1.977 | 0.00347 | 0.00347 | 0.03565 | 35 |
| GOBP_MONONUCLEAR_CELL_DIFFERENTIATION | 20 | -0.475 | -1.995 | 0.00193 | 0.00193 | 0.02438 | 156 |
| GOBP_TUBE_MORPHOGENESIS | 35 | -0.398 | -2.003 | 0.00163 | 0.00163 | 0.02211 | 173 |
| GOBP_INNATE_IMMUNE_RESPONSE | 25 | -0.446 | -2.009 | 0.00265 | 0.00265 | 0.02972 | 183 |
| GOBP_POSITIVE_REGULATION_OF_GENE_EXPRESSION | 52 | -0.365 | -2.011 | 0.00065 | 0.00065 | 0.01077 | 253 |
| GOBP_REGULATION_OF_MRNA_METABOLIC_PROCESS | 25 | -0.451 | -2.030 | 0.00235 | 0.00235 | 0.02754 | 325 |
| GOBP_POSITIVE_REGULATION_OF_CYTOKINE_PRODUCTION | 22 | -0.472 | -2.031 | 0.00354 | 0.00354 | 0.03565 | 156 |
| GOBP_CYTOKINE_PRODUCTION | 29 | -0.429 | -2.035 | 0.00167 | 0.00167 | 0.02217 | 156 |
| GOBP_POSITIVE_REGULATION_OF_MULTICELLULAR_ORGANISMAL_PROCESS | 61 | -0.356 | -2.045 | 0.00014 | 0.00014 | 0.00338 | 133 |
| GOBP_BIOLOGICAL_PROCESS_INVOLVED_IN_INTERSPECIES_INTERACTION_BETWE | 59 | -0.358 | -2.046 | 0.00026 | 0.00026 | 0.00566 | 183 |
| GOBP_REGULATION_OF_ENDOCYTOSIS | 10 | -0.638 | -2.046 | 0.00272 | 0.00272 | 0.03037 | 385 |
| GOBP_RESPONSE_TO_WOUNDING | 26 | -0.452 | -2.057 | 0.00191 | 0.00191 | 0.02438 | 132 |
| GOBP_SKELETAL_MUSCLE_ORGAN_DEVELOPMENT | 11 | -0.618 | -2.063 | 0.00287 | 0.00287 | 0.03086 | 238 |
| GOCC_BASAL_PART_OF_CELL | 11 | -0.619 | -2.064 | 0.00287 | 0.00287 | 0.03086 | 204 |
| GOBP_LEUKOCYTE_MIGRATION | 10 | -0.648 | -2.078 | 0.00220 | 0.00220 | 0.02621 | 130 |
| GOBP_POSITIVE_REGULATION_OF_CELL_ADHESION | 22 | -0.485 | -2.088 | 0.00230 | 0.00230 | 0.02718 | 91 |
| GOBP_REGULATION_OF_CELL_CELL_ADHESION | 24 | -0.473 | -2.109 | 0.00211 | 0.00211 | 0.02571 | 253 |
| GOBP_LYMPHOCYTE_ACTIVATION | 38 | -0.415 | -2.121 | 0.00035 | 0.00035 | 0.00677 | 178 |
| GOBP_REGULATION_OF_IMMUNE_SYSTEM_PROCESS | 61 | -0.370 | -2.124 | 0.00006 | 0.00006 | 0.00173 | 203 |
| GOCC_VACUOLE | 32 | -0.438 | -2.134 | 0.00064 | 0.00064 | 0.01073 | 300 |
| GOCC_CELL_LEADING_EDGE | 18 | -0.530 | -2.139 | 0.00118 | 0.00118 | 0.01731 | 105 |
| GOBP_GENE_SILENCING_BY_RNA | 11 | -0.648 | -2.163 | 0.00112 | 0.00112 | 0.01654 | 375 |
| GOBP_WOUND_HEALING | 16 | -0.561 | -2.174 | 0.00112 | 0.00112 | 0.01654 | 107 |
| GOBP_NEGATIVE_REGULATION_OF_PHOSPHORUS_METABOLIC_PROCESS | 31 | -0.452 | -2.177 | 0.00057 | 0.00057 | 0.00997 | 77 |
| GOBP_ENDOCYTOSIS | 28 | -0.466 | -2.182 | 0.00044 | 0.00044 | 0.00833 | 291 |
| GOBP_DEFENSE_RESPONSE_TO_OTHER_ORGANISM | 34 | -0.440 | -2.188 | 0.00042 | 0.00042 | 0.00813 | 183 |
| GOBP_ORGANIC_HYDROXY_COMPOUND_TRANSPORT | 11 | -0.659 | -2.197 | 0.00085 | 0.00085 | 0.01359 | 215 |
| GOBP_NEGATIVE_REGULATION_OF_SIGNALING | 66 | -0.375 | -2.198 | 0.00003 | 0.00003 | 0.00117 | 100 |
| GOCC_ENDOCYTIC_VESICLE | 14 | -0.599 | -2.199 | 0.00202 | 0.00202 | 0.02504 | 339 |
| GOBP_TAXIS | 18 | -0.552 | -2.226 | 0.00052 | 0.00052 | 0.00940 | 130 |
| GOBP_POSITIVE_REGULATION_OF_CELL_ACTIVATION | 18 | -0.556 | -2.241 | 0.00046 | 0.00046 | 0.00867 | 84 |
| GOBP_NEGATIVE_REGULATION_OF_PHOSPHORYLATION | 28 | -0.481 | -2.255 | 0.00023 | 0.00023 | 0.00511 | 77 |
| GOBP_RESPONSE_TO_CYTOKINE | 42 | -0.430 | -2.266 | 0.00006 | 0.00006 | 0.00175 | 156 |
| GOBP_REGULATION_OF_CELL_ACTIVATION | 27 | -0.491 | -2.278 | 0.00017 | 0.00017 | 0.00396 | 170 |
| GOBP_HEART_MORPHOGENESIS | 11 | -0.689 | -2.299 | 0.00033 | 0.00033 | 0.00653 | 220 |
| GOBP_REGULATION_OF_CELL_ADHESION | 40 | -0.444 | -2.304 | 0.00008 | 0.00008 | 0.00210 | 212 |

| | | | | | | | |
|---|---|---|---|---|---|---|---|
| GOBP_POSITIVE_REGULATION_OF_CELL_DEATH | 40 | -0.453 | -2.347 | 0.00005 | 0.00005 | 0.00155 | 92 |
| GOBP_LEUKOCYTE_DIFFERENTIATION | 24 | -0.535 | -2.386 | 0.00013 | 0.00013 | 0.00315 | 173 |
| GOBP_CELL_CHEMOTAXIS | 10 | -0.758 | -2.430 | 0.00006 | 0.00006 | 0.00171 | 130 |
| GOBP_CYTOKINE_MEDIATED_SIGNALING_PATHWAY | 20 | -0.600 | -2.522 | 0.00001 | 0.00001 | 0.00062 | 156 |
| GOBP_CELLULAR_RESPONSE_TO_BIOTIC_STIMULUS | 10 | -0.805 | -2.583 | 0.00000 | 0.00000 | 0.00021 | 172 |
| GOBP_POSITIVE_REGULATION_OF_IMMUNE_SYSTEM_PROCESS | 32 | -0.545 | -2.655 | 0.00000 | 0.00000 | 0.00018 | 84 |
| GOBP_RESPONSE_TO_BACTERIUM | 20 | -0.643 | -2.700 | 0.00000 | 0.00000 | 0.00009 | 172 |
| GOBP_CELL_ACTIVATION | 53 | -0.500 | -2.776 | 0.00000 | 0.00000 | 0.00001 | 178 |
| GOBP_RESPONSE_TO_MOLECULE_OF_BACTERIAL_ORIGIN | 14 | -0.784 | -2.876 | 0.00000 | 0.00000 | 0.00002 | 172 |

**Supplemental Table S7.** Gene set enrichment analysis of the Hallmark library from differential gene expression of the comparison between presence of TP53 mutation to WT *TP53* samples. Data used for this comparison comes from the FIMM dataset.

| ID | setSize | enrichmentScore | NES | pvalue | p.adjust | qvalue | rank |
|---|---|---|---|---|---|---|---|
| HALLMARK_G2M_CHECKPOINT | 76 | 0.611 | 2.762 | 0.00000 | 0.00000 | 0.00000 | 316 |
| HALLMARK_E2F_TARGETS | 88 | 0.525 | 2.435 | 0.00000 | 0.00000 | 0.00000 | 423 |
| HALLMARK_MITOTIC_SPINDLE | 37 | 0.602 | 2.368 | 0.00000 | 0.00000 | 0.00000 | 229 |
| HALLMARK_SPERMATOGENESIS | 16 | 0.645 | 2.044 | 0.00027 | 0.00027 | 0.00052 | 220 |
| HALLMARK_GLYCOLYSIS | 18 | 0.432 | 1.416 | 0.09621 | 0.09621 | 0.08347 | 178 |
| HALLMARK_MYC_TARGETS_V1 | 36 | 0.316 | 1.238 | 0.20234 | 0.20234 | 0.14483 | 678 |
| HALLMARK_ESTROGEN_RESPONSE_LATE | 18 | 0.344 | 1.127 | 0.32960 | 0.32960 | 0.21202 | 395 |
| HALLMARK_MTORC1_SIGNALING | 32 | 0.228 | 0.867 | 0.64327 | 0.64327 | 0.39202 | 567 |
| HALLMARK_DNA_REPAIR | 24 | 0.222 | 0.787 | 0.73614 | 0.73614 | 0.42619 | 577 |
| HALLMARK_ADIPOGENESIS | 11 | 0.249 | 0.708 | 0.82005 | 0.82005 | 0.44342 | 786 |
| HALLMARK_PROTEIN_SECRETION | 10 | 0.247 | 0.681 | 0.84249 | 0.84249 | 0.44342 | 47 |
| HALLMARK_FATTY_ACID_METABOLISM | 10 | 0.150 | 0.413 | 0.99474 | 0.99474 | 0.50079 | 888 |
| HALLMARK_TNFA_SIGNALING_VIA_NFKB | 21 | -0.792 | -3.347 | 0.00000 | 0.00000 | 0.00000 | 156 |
| HALLMARK_HYPOXIA | 15 | -0.682 | -2.578 | 0.00004 | 0.00004 | 0.00009 | 198 |
| HALLMARK_COMPLEMENT | 15 | -0.612 | -2.315 | 0.00054 | 0.00054 | 0.00090 | 181 |
| HALLMARK_P53_PATHWAY | 11 | -0.661 | -2.202 | 0.00063 | 0.00063 | 0.00091 | 131 |
| HALLMARK_IL2_STAT5_SIGNALING | 14 | -0.496 | -1.819 | 0.01248 | 0.01248 | 0.01606 | 110 |
| HALLMARK_INTERFERON_GAMMA_RESPONSE | 16 | -0.460 | -1.778 | 0.01689 | 0.01689 | 0.01956 | 199 |
| HALLMARK_ALLOGRAFT_REJECTION | 10 | -0.543 | -1.750 | 0.02095 | 0.02095 | 0.02021 | 181 |
| HALLMARK_APOPTOSIS | 15 | -0.461 | -1.744 | 0.01946 | 0.01946 | 0.02021 | 156 |
| HALLMARK_HEME_METABOLISM | 12 | -0.411 | -1.418 | 0.10092 | 0.10092 | 0.08347 | 256 |
| HALLMARK_OXIDATIVE_PHOSPHORYLATION | 25 | -0.300 | -1.348 | 0.11384 | 0.11384 | 0.08788 | 675 |
| HALLMARK_UV_RESPONSE_UP | 12 | -0.356 | -1.230 | 0.21263 | 0.21263 | 0.14483 | 156 |

**Supplemental Table S8.** Differentially expressed genes from the comparison between samples with *TP53* mutation compared to WT *TP53* samples. Data from this table are derived from the comparison of the MMRF-CoMMpass data.

| ENSEMBL_ID | logFC | AveExpr | t | P.Value | P.adjust | B | external_gene_name | Description |
|---|---|---|---|---|---|---|---|---|
| ENSG00000000460 | 0.513 | 2.085 | 4.383 | 0.00001 | 0.00048 | 2.777 | C1orf112 | chromosome 1 open reading frame 112 [Source:HGNC Symbol;Acc:HGNC:25565] |
| ENSG00000004455 | 0.260 | 6.321 | 3.570 | 0.00039 | 0.00713 | -0.688 | AK2 | adenylate kinase 2 [Source:HGNC Symbol;Acc:HGNC:362] |
| ENSG00000004660 | -0.862 | 3.723 | -5.633 | 0.00000 | 0.00000 | 8.601 | CAMKK1 | calcium/calmodulin dependent protein kinase kinase 1 [Source:HGNC Symbol;Acc:HGNC:1469] |
| ENSG00000004961 | 0.276 | 4.281 | 4.769 | 0.00000 | 0.00011 | 4.201 | HCCS | holocytochrome c synthase [Source:HGNC Symbol;Acc:HGNC:4837] |
| ENSG00000004975 | -0.381 | 5.043 | -6.292 | 0.00000 | 0.00000 | 12.120 | DVL2 | dishevelled segment polarity protein 2 [Source:HGNC Symbol;Acc:HGNC:3086] |
| ENSG00000005189 | 0.659 | 1.291 | 5.330 | 0.00000 | 0.00001 | 7.132 | REXO5 | RNA exonuclease 5 [Source:HGNC Symbol;Acc:HGNC:24661] |
| ENSG00000006282 | -0.293 | 6.554 | -3.790 | 0.00017 | 0.00371 | 0.094 | SPATA20 | spermatogenesis associated 20 [Source:HGNC Symbol;Acc:HGNC:26125] |
| ENSG00000006576 | 0.330 | 4.875 | 3.726 | 0.00021 | 0.00460 | -0.112 | PHTF2 | putative homeodomain transcription factor 2 [Source:HGNC Symbol;Acc:HGNC:13411] |
| ENSG00000006625 | 0.337 | 2.366 | 3.642 | 0.00029 | 0.00587 | -0.082 | GGCT | gamma-glutamylcyclotransferase [Source:HGNC Symbol;Acc:HGNC:21705] |
| ENSG00000006744 | -0.347 | 6.365 | -6.164 | 0.00000 | 0.00000 | 11.330 | ELAC2 | elaC ribonuclease Z 2 [Source:HGNC Symbol;Acc:HGNC:14198] |
| ENSG00000007168 | -0.405 | 6.789 | -7.440 | 0.00000 | 0.00000 | 19.326 | PAFAH1B1 | platelet activating factor acetylhydrolase 1b regulatory subunit 1 [Source:HGNC Symbol;Acc:HGNC:8574] |
| ENSG00000007392 | -0.339 | 6.437 | -3.830 | 0.00014 | 0.00327 | 0.240 | LUC7L | LUC7 like [Source:HGNC Symbol;Acc:HGNC:6723] |
| ENSG00000011295 | -0.239 | 5.760 | -4.488 | 0.00001 | 0.00033 | 2.888 | TTC19 | tetratricopeptide repeat domain 19 [Source:HGNC Symbol;Acc:HGNC:26006] |
| ENSG00000011426 | 0.944 | 2.036 | 6.100 | 0.00000 | 0.00000 | 11.162 | ANLN | anillin actin binding protein [Source:HGNC Symbol;Acc:HGNC:14082] |
| ENSG00000012048 | 0.557 | 3.291 | 4.133 | 0.00004 | 0.00118 | 1.575 | BRCA1 | BRCA1, DNA repair associated [Source:HGNC Symbol;Acc:HGNC:1100] |
| ENSG00000012963 | 0.375 | 4.940 | 5.165 | 0.00000 | 0.00002 | 6.022 | UBR7 | ubiquitin protein ligase E3 component n-recognin 7 (putative) [Source:HGNC Symbol;Acc:HGNC:20344] |
| ENSG00000013810 | 0.522 | 4.072 | 4.423 | 0.00001 | 0.00043 | 2.675 | TACC3 | transforming acidic coiled-coil containing protein 3 [Source:HGNC Symbol;Acc:HGNC:11524] |
| ENSG00000014138 | 0.465 | 1.853 | 5.648 | 0.00000 | 0.00000 | 8.739 | POLA2 | DNA polymerase alpha 2, accessory subunit [Source:HGNC Symbol;Acc:HGNC:30073] |
| ENSG00000015413 | -2.087 | 4.740 | -3.721 | 0.00022 | 0.00466 | 0.126 | DPEP1 | dipeptidase 1 (renal) [Source:HGNC Symbol;Acc:HGNC:3002] |
| ENSG00000020922 | 0.276 | 5.318 | 3.562 | 0.00040 | 0.00731 | -0.714 | MRE11 | MRE11 homolog, double strand break repair nuclease [Source:HGNC Symbol;Acc:HGNC:7230] |
| ENSG00000022277 | 0.196 | 5.527 | 4.002 | 0.00007 | 0.00183 | 0.898 | RTFDC1 | replication termination factor 2 domain containing 1 [Source:HGNC Symbol;Acc:HGNC:15890] |
| ENSG00000022976 | -0.281 | 3.982 | -3.682 | 0.00025 | 0.00524 | -0.127 | ZNF839 | zinc finger protein 839 [Source:HGNC Symbol;Acc:HGNC:20345] |
| ENSG00000023909 | 0.416 | 2.896 | 4.138 | 0.00004 | 0.00116 | 1.678 | GCLM | glutamate-cysteine ligase modifier subunit [Source:HGNC Symbol;Acc:HGNC:4312] |
| ENSG00000024526 | 1.124 | 1.436 | 6.575 | 0.00000 | 0.00000 | 13.931 | DEPDC1 | DEP domain containing 1 [Source:HGNC Symbol;Acc:HGNC:22949] |
| ENSG00000027847 | -0.211 | 5.861 | -3.521 | 0.00046 | 0.00816 | -0.857 | B4GALT7 | beta-1,4-galactosyltransferase 7 [Source:HGNC Symbol;Acc:HGNC:930] |
| ENSG00000028116 | 0.236 | 4.627 | 4.072 | 0.00005 | 0.00145 | 1.215 | VRK2 | vaccinia related kinase 2 [Source:HGNC Symbol;Acc:HGNC:12719] |
| ENSG00000029153 | 1.166 | 0.225 | 4.501 | 0.00001 | 0.00032 | 3.376 | ARNTL2 | aryl hydrocarbon receptor nuclear translocator like 2 [Source:HGNC Symbol;Acc:HGNC:18984] |
| ENSG00000029725 | -0.335 | 5.510 | -5.495 | 0.00000 | 0.00000 | 7.686 | RABEP1 | rabaptin, RAB GTPase binding effector protein 1 [Source:HGNC Symbol;Acc:HGNC:17677] |
| ENSG00000031081 | 0.511 | 4.619 | 3.558 | 0.00040 | 0.00739 | -0.698 | ARHGAP31 | Rho GTPase activating protein 31 [Source:HGNC Symbol;Acc:HGNC:29216] |
| ENSG00000031691 | 0.529 | 1.904 | 4.086 | 0.00005 | 0.00138 | 1.609 | CENPQ | centromere protein Q [Source:HGNC Symbol;Acc:HGNC:21347] |
| ENSG00000035499 | 0.776 | 1.993 | 4.987 | 0.00000 | 0.00004 | 5.430 | DEPDC1B | DEP domain containing 1B [Source:HGNC Symbol;Acc:HGNC:24902] |
| ENSG00000037042 | -0.333 | 5.194 | -3.703 | 0.00023 | 0.00492 | -0.181 | TUBG2 | tubulin gamma 2 [Source:HGNC Symbol;Acc:HGNC:12419] |
| ENSG00000038210 | 0.289 | 5.933 | 3.804 | 0.00016 | 0.00355 | 0.146 | PI4K2B | phosphatidylinositol 4-kinase type 2 beta [Source:HGNC Symbol;Acc:HGNC:18215] |
| ENSG00000039650 | -0.233 | 6.410 | -3.940 | 0.00009 | 0.00225 | 0.654 | PNKP | polynucleotide kinase 3'-phosphatase [Source:HGNC Symbol;Acc:HGNC:9154] |
| ENSG00000040275 | 0.372 | 3.224 | 4.026 | 0.00006 | 0.00170 | 1.186 | SPDL1 | spindle apparatus coiled-coil protein 1 [Source:HGNC Symbol;Acc:HGNC:26010] |
| ENSG00000040531 | -0.416 | 4.506 | -3.839 | 0.00014 | 0.00317 | 0.390 | CTNS | cystinosin, lysosomal cystine transporter [Source:HGNC Symbol;Acc:HGNC:2518] |
| ENSG00000040633 | -0.409 | 4.357 | -4.143 | 0.00004 | 0.00114 | 1.575 | PHF23 | PHD finger protein 23 [Source:HGNC Symbol;Acc:HGNC:28428] |
| ENSG00000048028 | 0.325 | 5.183 | 3.776 | 0.00018 | 0.00390 | 0.053 | USP28 | ubiquitin specific peptidase 28 [Source:HGNC Symbol;Acc:HGNC:12625] |
| ENSG00000049541 | 0.395 | 3.586 | 5.475 | 0.00000 | 0.00000 | 7.686 | RFC2 | replication factor C subunit 2 [Source:HGNC Symbol;Acc:HGNC:9970] |
| ENSG00000051180 | 0.767 | 1.484 | 6.909 | 0.00000 | 0.00000 | 15.956 | RAD51 | RAD51 recombinase [Source:HGNC Symbol;Acc:HGNC:9817] |
| ENSG00000051341 | 0.787 | 2.857 | 6.048 | 0.00000 | 0.00000 | 10.818 | POLQ | DNA polymerase theta [Source:HGNC Symbol;Acc:HGNC:9186] |
| ENSG00000051523 | -0.547 | 9.630 | -4.613 | 0.00000 | 0.00021 | 3.572 | CYBA | cytochrome b-245 alpha chain [Source:HGNC Symbol;Acc:HGNC:2577] |
| ENSG00000054965 | 0.241 | 5.003 | 3.770 | 0.00018 | 0.00398 | 0.042 | FAM168A | family with sequence similarity 168 member A [Source:HGNC Symbol;Acc:HGNC:28999] |
| ENSG00000055070 | 0.223 | 5.344 | 3.683 | 0.00025 | 0.00524 | -0.285 | SZRD1 | SUZ RNA binding domain containing 1 [Source:HGNC Symbol;Acc:HGNC:30232] |
| ENSG00000056050 | 0.361 | 3.887 | 3.664 | 0.00027 | 0.00548 | -0.246 | HPF1 | histone PARylation factor 1 [Source:HGNC Symbol;Acc:HGNC:26051] |
| ENSG00000057608 | 0.264 | 7.557 | 3.669 | 0.00027 | 0.00543 | -0.282 | GDI2 | GDP dissociation inhibitor 2 [Source:HGNC Symbol;Acc:HGNC:4227] |
| ENSG00000058804 | 0.284 | 4.607 | 3.450 | 0.00060 | 0.01000 | -1.052 | NDC1 | NDC1 transmembrane nucleoporin [Source:HGNC Symbol;Acc:HGNC:25525] |
| ENSG00000060688 | 0.228 | 4.510 | 4.180 | 0.00003 | 0.00100 | 1.653 | SNRNP40 | small nuclear ribonucleoprotein U5 subunit 40 [Source:HGNC Symbol;Acc:HGNC:30857] |
| ENSG00000060982 | 1.001 | 2.608 | 4.055 | 0.00006 | 0.00154 | 1.326 | BCAT1 | branched chain amino acid transaminase 1 [Source:HGNC Symbol;Acc:HGNC:976] |
| ENSG00000061794 | 0.285 | 4.763 | 3.451 | 0.00060 | 0.00998 | -1.060 | MRPS35 | mitochondrial ribosomal protein S35 [Source:HGNC Symbol;Acc:HGNC:16635] |
| ENSG00000063180 | -0.555 | 2.659 | -3.656 | 0.00028 | 0.00563 | 0.031 | CA11 | carbonic anhydrase 11 [Source:HGNC Symbol;Acc:HGNC:1370] |
| ENSG00000063241 | 0.389 | 4.776 | 4.306 | 0.00002 | 0.00064 | 2.141 | ISOC2 | isochorismatase domain containing 2 [Source:HGNC Symbol;Acc:HGNC:26278] |
| ENSG00000064666 | -0.511 | 7.029 | -4.933 | 0.00000 | 0.00005 | 4.891 | CNN2 | calponin 2 [Source:HGNC Symbol;Acc:HGNC:2156] |

| | | | | | | | | |
|---|---|---|---|---|---|---|---|---|
| ENSG00000064687 | -0.317 | 7.156 | -3.505 | 0.00049 | 0.00853 | -0.900 | ABCA7 | ATP binding cassette subfamily A member 7 [Source:HGNC Symbol;Acc:HGNC:37] |
| ENSG00000065328 | 1.033 | 1.097 | 6.552 | 0.00000 | 0.00000 | 13.771 | MCM10 | minichromosome maintenance 10 replication initiation factor [Source:HGNC Symbol;Acc:HGNC:18043] |
| ENSG00000065559 | -0.298 | 5.003 | -5.547 | 0.00000 | 0.00000 | 7.984 | MAP2K4 | mitogen-activated protein kinase kinase 4 [Source:HGNC Symbol;Acc:HGNC:6844] |
| ENSG00000066279 | 0.945 | 3.626 | 5.622 | 0.00000 | 0.00000 | 8.411 | ASPM | abnormal spindle microtubule assembly [Source:HGNC Symbol;Acc:HGNC:19048] |
| ENSG00000068394 | 0.264 | 4.229 | 4.405 | 0.00001 | 0.00045 | 2.607 | GPKOW | G-patch domain and KOW motifs [Source:HGNC Symbol;Acc:HGNC:30677] |
| ENSG00000068489 | 0.969 | 1.712 | 6.711 | 0.00000 | 0.00000 | 14.758 | PRR11 | proline rich 11 [Source:HGNC Symbol;Acc:HGNC:25619] |
| ENSG00000070366 | -0.371 | 4.734 | -4.823 | 0.00000 | 0.00009 | 4.454 | SMG6 | SMG6, nonsense mediated mRNA decay factor [Source:HGNC Symbol;Acc:HGNC:17809] |
| ENSG00000070444 | -0.326 | 4.822 | -4.242 | 0.00003 | 0.00082 | 1.918 | MNT | MAX network transcriptional repressor [Source:HGNC Symbol;Acc:HGNC:7188] |
| ENSG00000070950 | 0.454 | 3.672 | 6.136 | 0.00000 | 0.00000 | 11.264 | RAD18 | RAD18, E3 ubiquitin protein ligase [Source:HGNC Symbol;Acc:HGNC:18278] |
| ENSG00000071539 | 0.816 | 1.707 | 5.705 | 0.00000 | 0.00000 | 9.034 | TRIP13 | thyroid hormone receptor interactor 13 [Source:HGNC Symbol;Acc:HGNC:12307] |
| ENSG00000072501 | 0.280 | 6.451 | 3.637 | 0.00030 | 0.00593 | -0.447 | SMC1A | structural maintenance of chromosomes 1A [Source:HGNC Symbol;Acc:HGNC:11111] |
| ENSG00000072506 | 0.313 | 5.506 | 4.565 | 0.00001 | 0.00025 | 3.217 | HSD17B10 | hydroxysteroid 17-beta dehydrogenase 10 [Source:HGNC Symbol;Acc:HGNC:4800] |
| ENSG00000072571 | 0.951 | 2.584 | 6.971 | 0.00000 | 0.00000 | 16.356 | HMMR | hyaluronan mediated motility receptor [Source:HGNC Symbol;Acc:HGNC:5012] |
| ENSG00000072778 | -0.687 | 9.306 | -6.712 | 0.00000 | 0.00000 | 14.690 | ACADVL | acyl-CoA dehydrogenase, very long chain [Source:HGNC Symbol;Acc:HGNC:92] |
| ENSG00000072849 | -0.436 | 6.606 | -6.213 | 0.00000 | 0.00000 | 11.614 | DERL2 | derlin 2 [Source:HGNC Symbol;Acc:HGNC:17943] |
| ENSG00000072864 | 0.350 | 3.293 | 4.826 | 0.00000 | 0.00009 | 4.564 | NDE1 | nudE neurodevelopment protein 1 [Source:HGNC Symbol;Acc:HGNC:17619] |
| ENSG00000073111 | 0.783 | 3.549 | 5.890 | 0.00000 | 0.00000 | 9.876 | MCM2 | minichromosome maintenance complex component 2 [Source:HGNC Symbol;Acc:HGNC:6944] |
| ENSG00000074356 | -0.457 | 5.937 | -7.724 | 0.00000 | 0.00000 | 21.278 | NCBP3 | nuclear cap binding subunit 3 [Source:HGNC Symbol;Acc:HGNC:24612] |
| ENSG00000074370 | -0.782 | 9.460 | -7.507 | 0.00000 | 0.00000 | 19.844 | ATP2A3 | ATPase sarcoplasmic/endoplasmic reticulum Ca2+ transporting 3 [Source:HGNC Symbol;Acc:HGNC:813] |
| ENSG00000074755 | -0.428 | 7.192 | -5.636 | 0.00000 | 0.00000 | 8.420 | ZZEF1 | zinc finger ZZ-type and EF-hand domain containing 1 [Source:HGNC Symbol;Acc:HGNC:29027] |
| ENSG00000074800 | 0.321 | 8.111 | 3.516 | 0.00047 | 0.00827 | -0.772 | ENO1 | enolase 1 [Source:HGNC Symbol;Acc:HGNC:3350] |
| ENSG00000075131 | 0.424 | 2.553 | 5.073 | 0.00000 | 0.00003 | 5.803 | TIPIN | TIMELESS interacting protein [Source:HGNC Symbol;Acc:HGNC:30750] |
| ENSG00000075213 | 1.152 | 0.926 | 3.546 | 0.00042 | 0.00763 | -0.265 | SEMA3A | semaphorin 3A [Source:HGNC Symbol;Acc:HGNC:10723] |
| ENSG00000075218 | 0.989 | 1.731 | 7.100 | 0.00000 | 0.00000 | 17.185 | GTSE1 | G2 and S-phase expressed 1 [Source:HGNC Symbol;Acc:HGNC:13698] |
| ENSG00000075240 | -0.452 | 5.772 | -4.553 | 0.00001 | 0.00026 | 3.174 | GRAMD4 | GRAM domain containing 4 [Source:HGNC Symbol;Acc:HGNC:29113] |
| ENSG00000075336 | 0.286 | 4.448 | 3.524 | 0.00046 | 0.00811 | -0.789 | TIMM21 | translocase of inner mitochondrial membrane 21 [Source:HGNC Symbol;Acc:HGNC:25010] |
| ENSG00000075399 | -0.370 | 3.950 | -3.917 | 0.00010 | 0.00242 | 0.753 | VPS9D1 | VPS9 domain containing 1 [Source:HGNC Symbol;Acc:HGNC:13526] |
| ENSG00000075702 | 0.599 | 2.925 | 4.669 | 0.00000 | 0.00016 | 3.875 | WDR62 | WD repeat domain 62 [Source:HGNC Symbol;Acc:HGNC:24502] |
| ENSG00000076003 | 0.473 | 5.110 | 4.711 | 0.00000 | 0.00014 | 3.874 | MCM6 | minichromosome maintenance complex component 6 [Source:HGNC Symbol;Acc:HGNC:6949] |
| ENSG00000076382 | 0.606 | 3.948 | 5.266 | 0.00000 | 0.00001 | 6.577 | SPAG5 | sperm associated antigen 5 [Source:HGNC Symbol;Acc:HGNC:13452] |
| ENSG00000077152 | 0.723 | 2.161 | 4.670 | 0.00000 | 0.00016 | 3.969 | UBE2T | ubiquitin conjugating enzyme E2 T [Source:HGNC Symbol;Acc:HGNC:25009] |
| ENSG00000077514 | 0.501 | 3.406 | 5.764 | 0.00000 | 0.00000 | 9.219 | POLD3 | DNA polymerase delta 3, accessory subunit [Source:HGNC Symbol;Acc:HGNC:20932] |
| ENSG00000079387 | 0.203 | 4.367 | 3.768 | 0.00018 | 0.00398 | 0.090 | SENP1 | SUMO1/sentrin specific peptidase 1 [Source:HGNC Symbol;Acc:HGNC:17927] |
| ENSG00000080986 | 0.973 | 2.655 | 6.970 | 0.00000 | 0.00000 | 16.347 | NDC80 | NDC80, kinetochore complex component [Source:HGNC Symbol;Acc:HGNC:16909] |
| ENSG00000085840 | 0.708 | 1.628 | 6.075 | 0.00000 | 0.00000 | 11.044 | ORC1 | origin recognition complex subunit 1 [Source:HGNC Symbol;Acc:HGNC:8487] |
| ENSG00000085999 | 0.702 | 1.200 | 5.969 | 0.00000 | 0.00000 | 10.447 | RAD54L | RAD54 like (S. cerevisiae) [Source:HGNC Symbol;Acc:HGNC:9826] |
| ENSG00000086827 | 0.370 | 4.222 | 5.333 | 0.00000 | 0.00001 | 6.905 | ZW10 | zw10 kinetochore protein [Source:HGNC Symbol;Acc:HGNC:13194] |
| ENSG00000087053 | 0.342 | 4.645 | 4.999 | 0.00000 | 0.00004 | 5.237 | MTMR2 | myotubularin related protein 2 [Source:HGNC Symbol;Acc:HGNC:7450] |
| ENSG00000087088 | -0.278 | 5.400 | -4.375 | 0.00001 | 0.00050 | 2.423 | BAX | BCL2 associated X, apoptosis regulator [Source:HGNC Symbol;Acc:HGNC:959] |
| ENSG00000087301 | 0.417 | 3.919 | 4.944 | 0.00000 | 0.00005 | 5.032 | TXNDC16 | thioredoxin domain containing 16 [Source:HGNC Symbol;Acc:HGNC:19965] |
| ENSG00000087303 | 1.038 | 2.493 | 4.183 | 0.00003 | 0.00100 | 1.848 | NID2 | nidogen 2 [Source:HGNC Symbol;Acc:HGNC:13389] |
| ENSG00000087586 | 1.000 | 1.387 | 6.423 | 0.00000 | 0.00000 | 13.029 | AURKA | aurora kinase A [Source:HGNC Symbol;Acc:HGNC:11393] |
| ENSG00000087842 | 0.660 | 0.596 | 4.034 | 0.00006 | 0.00166 | 1.528 | PIR | pirin [Source:HGNC Symbol;Acc:HGNC:30048] |
| ENSG00000088325 | 1.049 | 2.993 | 6.459 | 0.00000 | 0.00000 | 13.171 | TPX2 | TPX2, microtubule nucleation factor [Source:HGNC Symbol;Acc:HGNC:1249] |
| ENSG00000089685 | 1.019 | 2.373 | 6.524 | 0.00000 | 0.00000 | 13.609 | BIRC5 | baculoviral IAP repeat containing 5 [Source:HGNC Symbol;Acc:HGNC:593] |
| ENSG00000090889 | 1.183 | 1.544 | 7.401 | 0.00000 | 0.00000 | 19.148 | KIF4A | kinesin family member 4A [Source:HGNC Symbol;Acc:HGNC:13339] |
| ENSG00000091164 | 0.257 | 6.292 | 4.759 | 0.00000 | 0.00011 | 4.086 | TXNL1 | thioredoxin like 1 [Source:HGNC Symbol;Acc:HGNC:12436] |
| ENSG00000091542 | -0.325 | 7.074 | -4.573 | 0.00001 | 0.00024 | 3.260 | ALKBH5 | alkB homolog 5, RNA demethylase [Source:HGNC Symbol;Acc:HGNC:25996] |
| ENSG00000091622 | -0.540 | 1.276 | -4.000 | 0.00007 | 0.00184 | 1.418 | PITPNM3 | PITPNM family member 3 [Source:HGNC Symbol;Acc:HGNC:21043] |
| ENSG00000091640 | -0.470 | 5.307 | -7.605 | 0.00000 | 0.00000 | 20.475 | SPAG7 | sperm associated antigen 7 [Source:HGNC Symbol;Acc:HGNC:11216] |
| ENSG00000092421 | -0.491 | 1.502 | -4.441 | 0.00001 | 0.00040 | 3.131 | SEMA6A | semaphorin 6A [Source:HGNC Symbol;Acc:HGNC:10738] |
| ENSG00000092470 | 0.708 | 2.974 | 6.533 | 0.00000 | 0.00000 | 13.635 | WDR76 | WD repeat domain 76 [Source:HGNC Symbol;Acc:HGNC:25773] |
| ENSG00000092853 | 0.815 | 1.820 | 6.083 | 0.00000 | 0.00000 | 11.083 | CLSPN | claspin [Source:HGNC Symbol;Acc:HGNC:19715] |
| ENSG00000093009 | 0.778 | 2.068 | 5.239 | 0.00000 | 0.00001 | 6.633 | CDC45 | cell division cycle 45 [Source:HGNC Symbol;Acc:HGNC:1739] |
| ENSG00000093072 | -0.595 | 8.957 | -3.572 | 0.00038 | 0.00709 | -0.573 | ADA2 | adenosine deaminase 2 [Source:HGNC Symbol;Acc:HGNC:1839] |
| ENSG00000093217 | 0.534 | 1.802 | 3.986 | 0.00008 | 0.00193 | 1.234 | XYLB | xylulokinase [Source:HGNC Symbol;Acc:HGNC:12839] |

| | | | | | | | | |
|---|---|---|---|---|---|---|---|---|
| ENSG00000094804 | 0.512 | 3.910 | 3.518 | 0.00047 | 0.00822 | -0.770 | CDC6 | cell division cycle 6 [Source:HGNC Symbol;Acc:HGNC:1744] |
| ENSG00000095002 | 0.558 | 3.671 | 4.180 | 0.00003 | 0.00100 | 1.711 | MSH2 | mutS homolog 2 [Source:HGNC Symbol;Acc:HGNC:7325] |
| ENSG00000095139 | 0.356 | 8.034 | 5.078 | 0.00000 | 0.00003 | 5.667 | ARCN1 | archain 1 [Source:HGNC Symbol;Acc:HGNC:649] |
| ENSG00000097046 | 0.570 | 2.682 | 4.744 | 0.00000 | 0.00012 | 4.255 | CDC7 | cell division cycle 7 [Source:HGNC Symbol;Acc:HGNC:1745] |
| ENSG00000099901 | 0.252 | 5.507 | 3.620 | 0.00032 | 0.00621 | -0.515 | RANBP1 | RAN binding protein 1 [Source:HGNC Symbol;Acc:HGNC:9847] |
| ENSG00000100162 | 0.860 | 1.664 | 6.883 | 0.00000 | 0.00000 | 15.809 | CENPM | centromere protein M [Source:HGNC Symbol;Acc:HGNC:18352] |
| ENSG00000100281 | 0.359 | 4.036 | 5.506 | 0.00000 | 0.00000 | 7.806 | HMGXB4 | HMG-box containing 4 [Source:HGNC Symbol;Acc:HGNC:5003] |
| ENSG00000100296 | 0.244 | 4.073 | 3.799 | 0.00016 | 0.00361 | 0.230 | THOC5 | THO complex 5 [Source:HGNC Symbol;Acc:HGNC:19074] |
| ENSG00000100321 | -1.490 | 2.682 | -3.480 | 0.00054 | 0.00920 | -0.471 | SYNGR1 | synaptogyrin 1 [Source:HGNC Symbol;Acc:HGNC:11498] |
| ENSG00000100350 | 0.552 | 3.587 | 3.630 | 0.00031 | 0.00605 | -0.342 | FOXRED2 | FAD dependent oxidoreductase domain containing 2 [Source:HGNC Symbol;Acc:HGNC:26264] |
| ENSG00000100479 | 0.806 | 0.945 | 5.859 | 0.00000 | 0.00000 | 9.848 | POLE2 | DNA polymerase epsilon 2, accessory subunit [Source:HGNC Symbol;Acc:HGNC:9178] |
| ENSG00000100526 | 0.971 | 2.050 | 8.481 | 0.00000 | 0.00000 | 26.766 | CDKN3 | cyclin dependent kinase inhibitor 3 [Source:HGNC Symbol;Acc:HGNC:1791] |
| ENSG00000100628 | 1.157 | 0.415 | 4.336 | 0.00002 | 0.00058 | 2.699 | ASB2 | ankyrin repeat and SOCS box containing 2 [Source:HGNC Symbol;Acc:HGNC:16012] |
| ENSG00000101003 | 0.802 | 1.434 | 4.981 | 0.00000 | 0.00005 | 5.450 | GINS1 | GINS complex subunit 1 [Source:HGNC Symbol;Acc:HGNC:28980] |
| ENSG00000101057 | 0.473 | 4.605 | 3.914 | 0.00010 | 0.00245 | 0.596 | MYBL2 | MYB proto-oncogene like 2 [Source:HGNC Symbol;Acc:HGNC:7548] |
| ENSG00000101126 | 0.185 | 6.459 | 3.619 | 0.00032 | 0.00622 | -0.516 | ADNP | activity dependent neuroprotector homeobox [Source:HGNC Symbol;Acc:HGNC:15766] |
| ENSG00000101412 | 0.699 | 3.449 | 4.905 | 0.00000 | 0.00006 | 4.876 | E2F1 | E2F transcription factor 1 [Source:HGNC Symbol;Acc:HGNC:3113] |
| ENSG00000101447 | 0.736 | 0.390 | 4.271 | 0.00002 | 0.00073 | 2.449 | FAM83D | family with sequence similarity 83 member D [Source:HGNC Symbol;Acc:HGNC:16122] |
| ENSG00000101473 | 0.195 | 3.632 | 3.548 | 0.00042 | 0.00763 | -0.598 | ACOT8 | acyl-CoA thioesterase 8 [Source:HGNC Symbol;Acc:HGNC:15919] |
| ENSG00000101639 | 0.272 | 5.494 | 3.603 | 0.00034 | 0.00648 | -0.574 | CEP192 | centrosomal protein 192 [Source:HGNC Symbol;Acc:HGNC:25515] |
| ENSG00000101773 | 0.510 | 4.686 | 6.130 | 0.00000 | 0.00000 | 11.161 | RBBP8 | RB binding protein 8, endonuclease [Source:HGNC Symbol;Acc:HGNC:9891] |
| ENSG00000101811 | 0.402 | 3.440 | 5.194 | 0.00000 | 0.00002 | 6.288 | CSTF2 | cleavage stimulation factor subunit 2 [Source:HGNC Symbol;Acc:HGNC:2484] |
| ENSG00000101843 | 0.334 | 5.166 | 4.401 | 0.00001 | 0.00046 | 2.521 | PSMD10 | proteasome 26S subunit, non-ATPase 10 [Source:HGNC Symbol;Acc:HGNC:9555] |
| ENSG00000101868 | 0.462 | 3.805 | 5.746 | 0.00000 | 0.00000 | 9.088 | POLA1 | DNA polymerase alpha 1, catalytic subunit [Source:HGNC Symbol;Acc:HGNC:9173] |
| ENSG00000101888 | 0.604 | 3.940 | 6.054 | 0.00000 | 0.00000 | 10.769 | NXT2 | nuclear transport factor 2 like export factor 2 [Source:HGNC Symbol;Acc:HGNC:18151] |
| ENSG00000101911 | 0.441 | 4.570 | 3.666 | 0.00027 | 0.00546 | -0.311 | PRPS2 | phosphoribosyl pyrophosphate synthetase 2 [Source:HGNC Symbol;Acc:HGNC:9465] |
| ENSG00000101935 | 0.372 | 4.093 | 4.467 | 0.00001 | 0.00036 | 2.876 | AMMECR1 | Alport syndrome, mental retardation, midface hypoplasia and elliptocytosis chromosomal region gene 1 [Source:HGNC Symbol;Acc:HGNC:467] |
| ENSG00000101945 | 0.375 | 2.877 | 5.157 | 0.00000 | 0.00002 | 6.180 | SUV39H1 | suppressor of variegation 3-9 homolog 1 [Source:HGNC Symbol;Acc:HGNC:11479] |
| ENSG00000102172 | 0.288 | 4.110 | 3.922 | 0.00010 | 0.00239 | 0.684 | SMS | spermine synthase [Source:HGNC Symbol;Acc:HGNC:11123] |
| ENSG00000102384 | 1.099 | 0.131 | 6.664 | 0.00000 | 0.00000 | 14.297 | CENPI | centromere protein I [Source:HGNC Symbol;Acc:HGNC:3968] |
| ENSG00000102781 | 0.465 | 2.148 | 3.467 | 0.00056 | 0.00955 | -0.656 | KATNAL1 | katanin catalytic subunit A1 like 1 [Source:HGNC Symbol;Acc:HGNC:28361] |
| ENSG00000103044 | -0.515 | 1.485 | -3.983 | 0.00008 | 0.00195 | 1.341 | HAS3 | hyaluronan synthase 3 [Source:HGNC Symbol;Acc:HGNC:4820] |
| ENSG00000103064 | -0.312 | 4.865 | -3.610 | 0.00033 | 0.00639 | -0.485 | SLC7A6 | solute carrier family 7 member 6 [Source:HGNC Symbol;Acc:HGNC:11064] |
| ENSG00000103202 | -0.628 | 5.325 | -3.947 | 0.00009 | 0.00220 | 0.733 | NME4 | NME/NM23 nucleoside diphosphate kinase 4 [Source:HGNC Symbol;Acc:HGNC:7852] |
| ENSG00000103227 | -0.656 | 5.736 | -4.436 | 0.00001 | 0.00041 | 2.682 | LMF1 | lipase maturation factor 1 [Source:HGNC Symbol;Acc:HGNC:14154] |
| ENSG00000103489 | 1.489 | 3.523 | 4.343 | 0.00002 | 0.00056 | 2.334 | XYLT1 | xylosyltransferase 1 [Source:HGNC Symbol;Acc:HGNC:15516] |
| ENSG00000104324 | -0.986 | 5.184 | -4.405 | 0.00001 | 0.00045 | 2.614 | CPQ | carboxypeptidase Q [Source:HGNC Symbol;Acc:HGNC:16910] |
| ENSG00000104356 | 0.392 | 2.304 | 3.852 | 0.00013 | 0.00303 | 0.677 | POP1 | POP1 homolog, ribonuclease P/MRP subunit [Source:HGNC Symbol;Acc:HGNC:30129] |
| ENSG00000104419 | -0.862 | 7.119 | -4.218 | 0.00003 | 0.00089 | 1.752 | NDRG1 | N-myc downstream regulated 1 [Source:HGNC Symbol;Acc:HGNC:7679] |
| ENSG00000104442 | 0.392 | 5.158 | 4.020 | 0.00007 | 0.00173 | 0.974 | ARMC1 | armadillo repeat containing 1 [Source:HGNC Symbol;Acc:HGNC:17684] |
| ENSG00000104549 | 0.780 | 4.254 | 4.831 | 0.00000 | 0.00008 | 4.456 | SQLE | squalene epoxidase [Source:HGNC Symbol;Acc:HGNC:11279] |
| ENSG00000104626 | 0.375 | 3.033 | 4.124 | 0.00004 | 0.00121 | 1.601 | ERI1 | exoribonuclease 1 [Source:HGNC Symbol;Acc:HGNC:23994] |
| ENSG00000104738 | 0.647 | 4.612 | 5.299 | 0.00000 | 0.00001 | 6.691 | MCM4 | minichromosome maintenance complex component 4 [Source:HGNC Symbol;Acc:HGNC:6947] |
| ENSG00000104859 | -0.243 | 6.676 | -3.462 | 0.00057 | 0.00968 | -1.059 | CLASRP | CLK4 associating serine/arginine rich protein [Source:HGNC Symbol;Acc:HGNC:17731] |
| ENSG00000104863 | -0.459 | 3.094 | -4.793 | 0.00000 | 0.00010 | 4.528 | LIN7B | lin-7 homolog B, crumbs cell polarity complex component [Source:HGNC Symbol;Acc:HGNC:17788] |
| ENSG00000104946 | -0.236 | 6.401 | -3.815 | 0.00015 | 0.00344 | 0.184 | TBC1D17 | TBC1 domain family member 17 [Source:HGNC Symbol;Acc:HGNC:25699] |
| ENSG00000104960 | -0.248 | 6.114 | -3.522 | 0.00046 | 0.00814 | -0.858 | PTOV1 | prostate tumor overexpressed 1 [Source:HGNC Symbol;Acc:HGNC:9632] |
| ENSG00000105011 | 0.763 | 3.131 | 6.013 | 0.00000 | 0.00000 | 10.597 | ASF1B | anti-silencing function 1B histone chaperone [Source:HGNC Symbol;Acc:HGNC:20996] |
| ENSG00000105287 | -0.551 | 8.423 | -5.085 | 0.00000 | 0.00003 | 5.676 | PRKD2 | protein kinase D2 [Source:HGNC Symbol;Acc:HGNC:17293] |
| ENSG00000105976 | 0.705 | 3.740 | 3.606 | 0.00034 | 0.00645 | -0.460 | MET | MET proto-oncogene, receptor tyrosine kinase [Source:HGNC Symbol;Acc:HGNC:7029] |
| ENSG00000106462 | 0.701 | 4.430 | 6.164 | 0.00000 | 0.00000 | 11.360 | EZH2 | enhancer of zeste 2 polycomb repressive complex 2 subunit [Source:HGNC Symbol;Acc:HGNC:3527] |
| ENSG00000106554 | 0.298 | 4.986 | 5.352 | 0.00000 | 0.00001 | 6.952 | CHCHD3 | coiled-coil-helix-coiled-coil-helix domain containing 3 [Source:HGNC Symbol;Acc:HGNC:21906] |
| ENSG00000106992 | -0.583 | 0.937 | -3.631 | 0.00031 | 0.00604 | 0.131 | AK1 | adenylate kinase 1 [Source:HGNC Symbol;Acc:HGNC:361] |
| ENSG00000107140 | -0.315 | 4.762 | -3.902 | 0.00011 | 0.00255 | 0.592 | TESK1 | testis-specific kinase 1 [Source:HGNC Symbol;Acc:HGNC:11731] |
| ENSG00000108055 | 0.288 | 6.099 | 4.180 | 0.00003 | 0.00100 | 1.597 | SMC3 | structural maintenance of chromosomes 3 [Source:HGNC Symbol;Acc:HGNC:2468] |
| ENSG00000108064 | 0.264 | 5.621 | 4.314 | 0.00002 | 0.00062 | 2.147 | TFAM | transcription factor A, mitochondrial [Source:HGNC Symbol;Acc:HGNC:11741] |

| | | | | | | | | |
|---|---|---|---|---|---|---|---|---|
| ENSG00000108094 | 0.182 | 5.039 | 3.942 | 0.00009 | 0.00224 | 0.685 | CUL2 | cullin 2 [Source:HGNC Symbol;Acc:HGNC:2552] |
| ENSG00000108106 | 0.327 | 4.705 | 3.535 | 0.00044 | 0.00784 | -0.773 | UBE2S | ubiquitin conjugating enzyme E2 S [Source:HGNC Symbol;Acc:HGNC:17895] |
| ENSG00000108384 | 0.337 | 3.214 | 3.675 | 0.00026 | 0.00535 | -0.109 | RAD51C | RAD51 paralog C [Source:HGNC Symbol;Acc:HGNC:9820] |
| ENSG00000108474 | -0.418 | 4.223 | -4.336 | 0.00002 | 0.00058 | 2.382 | PIGL | phosphatidylinositol glycan anchor biosynthesis class L [Source:HGNC Symbol;Acc:HGNC:8966] |
| ENSG00000108509 | -0.476 | 5.321 | -7.302 | 0.00000 | 0.00000 | 18.425 | CAMTA2 | calmodulin binding transcription activator 2 [Source:HGNC Symbol;Acc:HGNC:18807] |
| ENSG00000108515 | -0.476 | 4.220 | -4.402 | 0.00001 | 0.00046 | 2.665 | ENO3 | enolase 3 [Source:HGNC Symbol;Acc:HGNC:3354] |
| ENSG00000108518 | -0.310 | 7.379 | -4.282 | 0.00002 | 0.00070 | 2.037 | PFN1 | profilin 1 [Source:HGNC Symbol;Acc:HGNC:8881] |
| ENSG00000108523 | -0.419 | 6.997 | -8.926 | 0.00000 | 0.00000 | 30.178 | RNF167 | ring finger protein 167 [Source:HGNC Symbol;Acc:HGNC:24544] |
| ENSG00000108551 | -1.506 | 3.646 | -4.109 | 0.00005 | 0.00128 | 1.689 | RASD1 | ras related dexamethasone induced 1 [Source:HGNC Symbol;Acc:HGNC:15828] |
| ENSG00000108559 | -0.328 | 5.335 | -5.641 | 0.00000 | 0.00000 | 8.463 | NUP88 | nucleoporin 88 [Source:HGNC Symbol;Acc:HGNC:8067] |
| ENSG00000108561 | -0.340 | 6.197 | -3.722 | 0.00022 | 0.00466 | -0.155 | C1QBP | complement C1q binding protein [Source:HGNC Symbol;Acc:HGNC:1243] |
| ENSG00000108590 | -0.380 | 2.439 | -4.969 | 0.00000 | 0.00005 | 5.393 | MED31 | mediator complex subunit 31 [Source:HGNC Symbol;Acc:HGNC:24260] |
| ENSG00000108591 | -0.234 | 4.905 | -4.987 | 0.00000 | 0.00004 | 5.192 | DRG2 | developmentally regulated GTP binding protein 2 [Source:HGNC Symbol;Acc:HGNC:3030] |
| ENSG00000108599 | -0.266 | 5.330 | -5.476 | 0.00000 | 0.00000 | 7.597 | AKAP10 | A-kinase anchoring protein 10 [Source:HGNC Symbol;Acc:HGNC:368] |
| ENSG00000108654 | -0.301 | 11.409 | -4.037 | 0.00006 | 0.00164 | 1.295 | DDX5 | DEAD-box helicase 5 [Source:HGNC Symbol;Acc:HGNC:2746] |
| ENSG00000108963 | -0.501 | 4.977 | -7.375 | 0.00000 | 0.00000 | 18.925 | DPH1 | diphthamide biosynthesis 1 [Source:HGNC Symbol;Acc:HGNC:3003] |
| ENSG00000109063 | -0.444 | 2.598 | -4.200 | 0.00003 | 0.00095 | 2.067 | MYH3 | myosin heavy chain 3 [Source:HGNC Symbol;Acc:HGNC:7573] |
| ENSG00000109084 | 0.713 | 3.437 | 6.143 | 0.00000 | 0.00000 | 11.311 | TMEM97 | transmembrane protein 97 [Source:HGNC Symbol;Acc:HGNC:28106] |
| ENSG00000109103 | -0.361 | 5.392 | -3.875 | 0.00012 | 0.00279 | 0.438 | UNC119 | unc-119 lipid binding chaperone [Source:HGNC Symbol;Acc:HGNC:12565] |
| ENSG00000109805 | 0.945 | 2.678 | 7.824 | 0.00000 | 0.00000 | 22.055 | NCAPG | non-SMC condensin I complex subunit G [Source:HGNC Symbol;Acc:HGNC:24304] |
| ENSG00000109881 | 0.804 | 2.081 | 6.593 | 0.00000 | 0.00000 | 14.038 | CCDC34 | coiled-coil domain containing 34 [Source:HGNC Symbol;Acc:HGNC:25079] |
| ENSG00000109911 | 0.286 | 3.948 | 3.873 | 0.00012 | 0.00281 | 0.513 | ELP4 | elongator acetyltransferase complex subunit 4 [Source:HGNC Symbol;Acc:HGNC:1171] |
| ENSG00000110063 | 0.323 | 5.370 | 3.629 | 0.00031 | 0.00605 | -0.481 | DCPS | decapping enzyme, scavenger [Source:HGNC Symbol;Acc:HGNC:29812] |
| ENSG00000110107 | 0.279 | 6.798 | 3.769 | 0.00018 | 0.00398 | 0.038 | PRPF19 | pre-mRNA processing factor 19 [Source:HGNC Symbol;Acc:HGNC:17896] |
| ENSG00000110344 | 0.324 | 6.262 | 4.146 | 0.00004 | 0.00113 | 1.466 | UBE4A | ubiquitination factor E4A [Source:HGNC Symbol;Acc:HGNC:12499] |
| ENSG00000110435 | 0.203 | 4.296 | 3.520 | 0.00046 | 0.00819 | -0.783 | PDHX | pyruvate dehydrogenase complex component X [Source:HGNC Symbol;Acc:HGNC:21350] |
| ENSG00000110442 | 0.257 | 4.432 | 4.200 | 0.00003 | 0.00095 | 1.739 | COMMD9 | COMM domain containing 9 [Source:HGNC Symbol;Acc:HGNC:25014] |
| ENSG00000110925 | -0.239 | 4.739 | -3.573 | 0.00038 | 0.00708 | -0.604 | CSRNP2 | cysteine and serine rich nuclear protein 2 [Source:HGNC Symbol;Acc:HGNC:16006] |
| ENSG00000111206 | 0.843 | 2.587 | 6.316 | 0.00000 | 0.00000 | 12.371 | FOXM1 | forkhead box M1 [Source:HGNC Symbol;Acc:HGNC:3818] |
| ENSG00000111247 | 0.653 | 2.309 | 5.474 | 0.00000 | 0.00000 | 7.794 | RAD51AP1 | RAD51 associated protein 1 [Source:HGNC Symbol;Acc:HGNC:16956] |
| ENSG00000111554 | -0.233 | 4.759 | -3.872 | 0.00012 | 0.00281 | 0.470 | MDM1 | Mdm1 nuclear protein [Source:HGNC Symbol;Acc:HGNC:29917] |
| ENSG00000111665 | 0.465 | 2.475 | 4.171 | 0.00003 | 0.00103 | 1.860 | CDCA3 | cell division cycle associated 3 [Source:HGNC Symbol;Acc:HGNC:14624] |
| ENSG00000111676 | -0.423 | 6.564 | -5.222 | 0.00000 | 0.00002 | 6.281 | ATN1 | atrophin 1 [Source:HGNC Symbol;Acc:HGNC:3033] |
| ENSG00000112118 | 0.537 | 5.316 | 4.425 | 0.00001 | 0.00042 | 2.613 | MCM3 | minichromosome maintenance complex component 3 [Source:HGNC Symbol;Acc:HGNC:6945] |
| ENSG00000112137 | 0.976 | 4.541 | 3.702 | 0.00023 | 0.00493 | -0.206 | PHACTR1 | phosphatase and actin regulator 1 [Source:HGNC Symbol;Acc:HGNC:20990] |
| ENSG00000112312 | 0.440 | 3.387 | 4.690 | 0.00000 | 0.00015 | 3.927 | GMNN | geminin, DNA replication inhibitor [Source:HGNC Symbol;Acc:HGNC:17493] |
| ENSG00000112511 | -0.415 | 8.004 | -4.132 | 0.00004 | 0.00118 | 1.457 | PHF1 | PHD finger protein 1 [Source:HGNC Symbol;Acc:HGNC:8919] |
| ENSG00000112742 | 0.881 | 1.748 | 5.857 | 0.00000 | 0.00000 | 9.838 | TTK | TTK protein kinase [Source:HGNC Symbol;Acc:HGNC:12401] |
| ENSG00000112787 | -0.288 | 5.673 | -3.589 | 0.00036 | 0.00675 | -0.614 | FBRSL1 | fibrosin like 1 [Source:HGNC Symbol;Acc:HGNC:29308] |
| ENSG00000112984 | 1.057 | 1.239 | 6.111 | 0.00000 | 0.00000 | 11.241 | KIF20A | kinesin family member 20A [Source:HGNC Symbol;Acc:HGNC:9787] |
| ENSG00000113108 | -0.288 | 5.632 | -3.469 | 0.00056 | 0.00951 | -1.026 | APBB3 | amyloid beta precursor protein binding family B member 3 [Source:HGNC Symbol;Acc:HGNC:20708] |
| ENSG00000113368 | 0.399 | 4.346 | 3.539 | 0.00043 | 0.00776 | -0.735 | LMNB1 | lamin B1 [Source:HGNC Symbol;Acc:HGNC:6637] |
| ENSG00000113742 | -0.460 | 8.731 | -3.634 | 0.00030 | 0.00598 | -0.364 | CPEB4 | cytoplasmic polyadenylation element binding protein 4 [Source:HGNC Symbol;Acc:HGNC:21747] |
| ENSG00000113810 | 0.413 | 6.401 | 4.195 | 0.00003 | 0.00096 | 1.669 | SMC4 | structural maintenance of chromosomes 4 [Source:HGNC Symbol;Acc:HGNC:14013] |
| ENSG00000114013 | 0.849 | 3.681 | 3.645 | 0.00029 | 0.00583 | -0.325 | CD86 | CD86 molecule [Source:HGNC Symbol;Acc:HGNC:1705] |
| ENSG00000114209 | 0.253 | 4.754 | 3.959 | 0.00008 | 0.00211 | 0.767 | PDCD10 | programmed cell death 10 [Source:HGNC Symbol;Acc:HGNC:8761] |
| ENSG00000114346 | 0.488 | 3.409 | 4.400 | 0.00001 | 0.00046 | 2.657 | ECT2 | epithelial cell transforming 2 [Source:HGNC Symbol;Acc:HGNC:3155] |
| ENSG00000114491 | 0.374 | 4.230 | 3.817 | 0.00015 | 0.00342 | 0.268 | UMPS | uridine monophosphate synthetase [Source:HGNC Symbol;Acc:HGNC:12563] |
| ENSG00000114520 | 0.218 | 4.791 | 3.572 | 0.00038 | 0.00709 | -0.646 | SNX4 | sorting nexin 4 [Source:HGNC Symbol;Acc:HGNC:11175] |
| ENSG00000114744 | 0.201 | 4.476 | 3.554 | 0.00041 | 0.00749 | -0.681 | COMMD2 | COMM domain containing 2 [Source:HGNC Symbol;Acc:HGNC:24993] |
| ENSG00000115163 | 1.226 | 0.540 | 8.120 | 0.00000 | 0.00000 | 23.865 | CENPA | centromere protein A [Source:HGNC Symbol;Acc:HGNC:1851] |
| ENSG00000115267 | 0.463 | 5.828 | 3.989 | 0.00007 | 0.00192 | 0.843 | IFIH1 | interferon induced with helicase C domain 1 [Source:HGNC Symbol;Acc:HGNC:18873] |
| ENSG00000115392 | 0.246 | 3.807 | 3.879 | 0.00012 | 0.00276 | 0.560 | FANCL | Fanconi anemia complementation group L [Source:HGNC Symbol;Acc:HGNC:20748] |
| ENSG00000115484 | 0.234 | 6.433 | 3.706 | 0.00023 | 0.00488 | -0.206 | CCT4 | chaperonin containing TCP1 subunit 4 [Source:HGNC Symbol;Acc:HGNC:1617] |
| ENSG00000115525 | -0.471 | 6.177 | -3.985 | 0.00008 | 0.00193 | 0.829 | ST3GAL5 | ST3 beta-galactoside alpha-2,3-sialyltransferase 5 [Source:HGNC Symbol;Acc:HGNC:10872] |
| ENSG00000115884 | -0.552 | 10.196 | -3.956 | 0.00009 | 0.00213 | 0.904 | SDC1 | syndecan 1 [Source:HGNC Symbol;Acc:HGNC:10658] |

| | | | | | | | | |
|---|---|---|---|---|---|---|---|---|
| ENSG00000116133 | 0.979 | 2.881 | 4.502 | 0.00001 | 0.00032 | 3.117 | DHCR24 | 24-dehydrocholesterol reductase [Source:HGNC Symbol;Acc:HGNC:2859] |
| ENSG00000116793 | 0.302 | 4.807 | 3.507 | 0.00049 | 0.00847 | -0.873 | PHTF1 | putative homeodomain transcription factor 1 [Source:HGNC Symbol;Acc:HGNC:8939] |
| ENSG00000116830 | 0.276 | 4.828 | 3.547 | 0.00042 | 0.00763 | -0.738 | TTF2 | transcription termination factor 2 [Source:HGNC Symbol;Acc:HGNC:12398] |
| ENSG00000117010 | 0.335 | 1.218 | 4.427 | 0.00001 | 0.00042 | 3.059 | ZNF684 | zinc finger protein 684 [Source:HGNC Symbol;Acc:HGNC:28418] |
| ENSG00000117174 | 0.332 | 4.246 | 4.472 | 0.00001 | 0.00036 | 2.887 | ZNHIT6 | zinc finger HIT-type containing 6 [Source:HGNC Symbol;Acc:HGNC:26089] |
| ENSG00000117266 | -0.769 | 3.601 | -3.542 | 0.00043 | 0.00772 | -0.486 | CDK18 | cyclin dependent kinase 18 [Source:HGNC Symbol;Acc:HGNC:8751] |
| ENSG00000117399 | 0.906 | 2.122 | 6.135 | 0.00000 | 0.00000 | 11.360 | CDC20 | cell division cycle 20 [Source:HGNC Symbol;Acc:HGNC:1723] |
| ENSG00000117632 | 0.843 | 5.286 | 5.283 | 0.00000 | 0.00001 | 6.588 | STMN1 | stathmin 1 [Source:HGNC Symbol;Acc:HGNC:6510] |
| ENSG00000117650 | 1.137 | 1.012 | 5.913 | 0.00000 | 0.00000 | 10.151 | NEK2 | NIMA related kinase 2 [Source:HGNC Symbol;Acc:HGNC:7745] |
| ENSG00000117724 | 0.958 | 3.154 | 5.344 | 0.00000 | 0.00001 | 7.017 | CENPF | centromere protein F [Source:HGNC Symbol;Acc:HGNC:1857] |
| ENSG00000117751 | 0.231 | 5.028 | 4.156 | 0.00004 | 0.00109 | 1.523 | PPP1R8 | protein phosphatase 1 regulatory subunit 8 [Source:HGNC Symbol;Acc:HGNC:9296] |
| ENSG00000118193 | 0.877 | 1.471 | 5.127 | 0.00000 | 0.00002 | 6.133 | KIF14 | kinesin family member 14 [Source:HGNC Symbol;Acc:HGNC:19181] |
| ENSG00000118513 | 1.093 | 0.769 | 4.496 | 0.00001 | 0.00032 | 3.341 | MYB | MYB proto-oncogene, transcription factor [Source:HGNC Symbol;Acc:HGNC:7545] |
| ENSG00000118655 | 0.658 | 1.363 | 6.351 | 0.00000 | 0.00000 | 12.594 | DCLRE1B | DNA cross-link repair 1B [Source:HGNC Symbol;Acc:HGNC:17641] |
| ENSG00000118689 | -0.455 | 7.810 | -4.228 | 0.00003 | 0.00087 | 1.829 | FOXO3 | forkhead box O3 [Source:HGNC Symbol;Acc:HGNC:3821] |
| ENSG00000118690 | -0.988 | 4.792 | -4.879 | 0.00000 | 0.00007 | 4.752 | ARMC2 | armadillo repeat containing 2 [Source:HGNC Symbol;Acc:HGNC:23045] |
| ENSG00000119203 | 0.235 | 5.040 | 3.716 | 0.00022 | 0.00474 | -0.154 | CPSF3 | cleavage and polyadenylation specific factor 3 [Source:HGNC Symbol;Acc:HGNC:2326] |
| ENSG00000119242 | -0.317 | 3.509 | -3.493 | 0.00051 | 0.00888 | -0.692 | CCDC92 | coiled-coil domain containing 92 [Source:HGNC Symbol;Acc:HGNC:29563] |
| ENSG00000119403 | 0.589 | 3.842 | 3.662 | 0.00027 | 0.00553 | -0.273 | PHF19 | PHD finger protein 19 [Source:HGNC Symbol;Acc:HGNC:24566] |
| ENSG00000119616 | 0.245 | 4.743 | 3.531 | 0.00044 | 0.00793 | -0.784 | FCF1 | FCF1 rRNA-processing protein [Source:HGNC Symbol;Acc:HGNC:20220] |
| ENSG00000119969 | 0.564 | 2.958 | 4.650 | 0.00000 | 0.00018 | 3.793 | HELLS | helicase, lymphoid-specific [Source:HGNC Symbol;Acc:HGNC:4861] |
| ENSG00000120334 | 0.392 | 2.290 | 4.287 | 0.00002 | 0.00069 | 2.364 | CENPL | centromere protein L [Source:HGNC Symbol;Acc:HGNC:17879] |
| ENSG00000120509 | 0.260 | 4.233 | 4.330 | 0.00002 | 0.00059 | 2.293 | PDZD11 | PDZ domain containing 11 [Source:HGNC Symbol;Acc:HGNC:28034] |
| ENSG00000120539 | 0.352 | 3.895 | 4.414 | 0.00001 | 0.00044 | 2.672 | MASTL | microtubule associated serine/threonine kinase like [Source:HGNC Symbol;Acc:HGNC:19042] |
| ENSG00000120802 | 0.373 | 6.273 | 3.450 | 0.00060 | 0.01000 | -1.097 | TMPO | thymopoietin [Source:HGNC Symbol;Acc:HGNC:11875] |
| ENSG00000120889 | -0.578 | 5.497 | -3.656 | 0.00028 | 0.00563 | -0.355 | TNFRSF10B | TNF receptor superfamily member 10b [Source:HGNC Symbol;Acc:HGNC:11905] |
| ENSG00000120899 | -0.433 | 9.332 | -3.691 | 0.00024 | 0.00509 | -0.120 | PTK2B | protein tyrosine kinase 2 beta [Source:HGNC Symbol;Acc:HGNC:9612] |
| ENSG00000121152 | 0.935 | 1.884 | 6.555 | 0.00000 | 0.00000 | 13.816 | NCAPH | non-SMC condensin I complex subunit H [Source:HGNC Symbol;Acc:HGNC:1112] |
| ENSG00000121211 | 0.969 | 0.453 | 6.493 | 0.00000 | 0.00000 | 13.342 | MND1 | meiotic nuclear divisions 1 [Source:HGNC Symbol;Acc:HGNC:24839] |
| ENSG00000121281 | -0.367 | 5.348 | -3.515 | 0.00047 | 0.00829 | -0.848 | ADCY7 | adenylate cyclase 7 [Source:HGNC Symbol;Acc:HGNC:238] |
| ENSG00000121621 | 0.658 | 2.020 | 4.899 | 0.00000 | 0.00006 | 5.024 | KIF18A | kinesin family member 18A [Source:HGNC Symbol;Acc:HGNC:29441] |
| ENSG00000121774 | 0.214 | 6.659 | 3.796 | 0.00016 | 0.00364 | 0.130 | KHDRBS1 | KH RNA binding domain containing, signal transduction associated 1 [Source:HGNC Symbol;Acc:HGNC:18116] |
| ENSG00000121957 | 0.258 | 3.223 | 3.461 | 0.00058 | 0.00971 | -0.832 | GPSM2 | G protein signaling modulator 2 [Source:HGNC Symbol;Acc:HGNC:29501] |
| ENSG00000122483 | 0.399 | 3.190 | 4.387 | 0.00001 | 0.00048 | 2.648 | CCDC18 | coiled-coil domain containing 18 [Source:HGNC Symbol;Acc:HGNC:30370] |
| ENSG00000122952 | 0.917 | 3.040 | 6.114 | 0.00000 | 0.00000 | 11.163 | ZWINT | ZW10 interacting kinetochore protein [Source:HGNC Symbol;Acc:HGNC:13195] |
| ENSG00000122966 | 0.710 | 1.918 | 4.569 | 0.00001 | 0.00024 | 3.566 | CIT | citron rho-interacting serine/threonine kinase [Source:HGNC Symbol;Acc:HGNC:1985] |
| ENSG00000123080 | 0.976 | 2.705 | 5.198 | 0.00000 | 0.00002 | 6.350 | CDKN2C | cyclin dependent kinase inhibitor 2C [Source:HGNC Symbol;Acc:HGNC:1789] |
| ENSG00000123219 | 0.751 | 2.367 | 5.724 | 0.00000 | 0.00000 | 9.088 | CENPK | centromere protein K [Source:HGNC Symbol;Acc:HGNC:29479] |
| ENSG00000123358 | -1.244 | 7.309 | -4.767 | 0.00000 | 0.00011 | 4.118 | NR4A1 | nuclear receptor subfamily 4 group A member 1 [Source:HGNC Symbol;Acc:HGNC:7980] |
| ENSG00000123416 | 0.373 | 5.768 | 4.018 | 0.00007 | 0.00174 | 0.956 | TUBA1B | tubulin alpha 1b [Source:HGNC Symbol;Acc:HGNC:18809] |
| ENSG00000123473 | 0.693 | 2.399 | 6.569 | 0.00000 | 0.00000 | 13.889 | STIL | SCL/TAL1 interrupting locus [Source:HGNC Symbol;Acc:HGNC:10879] |
| ENSG00000123485 | 0.895 | 2.245 | 5.349 | 0.00000 | 0.00001 | 7.150 | HJURP | Holliday junction recognition protein [Source:HGNC Symbol;Acc:HGNC:25444] |
| ENSG00000123609 | 0.409 | 3.831 | 4.439 | 0.00001 | 0.00040 | 2.779 | NMI | N-myc and STAT interactor [Source:HGNC Symbol;Acc:HGNC:7854] |
| ENSG00000123636 | -0.440 | 6.314 | -4.644 | 0.00000 | 0.00018 | 3.565 | BAZ2B | bromodomain adjacent to zinc finger domain 2B [Source:HGNC Symbol;Acc:HGNC:963] |
| ENSG00000123737 | 0.226 | 5.010 | 4.002 | 0.00007 | 0.00183 | 0.916 | EXOSC9 | exosome component 9 [Source:HGNC Symbol;Acc:HGNC:9137] |
| ENSG00000124207 | 0.316 | 5.773 | 4.410 | 0.00001 | 0.00045 | 2.548 | CSE1L | chromosome segregation 1 like [Source:HGNC Symbol;Acc:HGNC:2431] |
| ENSG00000124256 | -0.406 | 7.829 | -3.707 | 0.00023 | 0.00486 | -0.161 | ZBP1 | Z-DNA binding protein 1 [Source:HGNC Symbol;Acc:HGNC:16176] |
| ENSG00000125319 | 0.599 | 0.874 | 4.556 | 0.00001 | 0.00026 | 3.601 | C17orf53 | chromosome 17 open reading frame 53 [Source:HGNC Symbol;Acc:HGNC:28460] |
| ENSG00000125434 | -0.514 | 2.608 | -4.670 | 0.00000 | 0.00016 | 4.035 | SLC25A35 | solute carrier family 25 member 35 [Source:HGNC Symbol;Acc:HGNC:31921] |
| ENSG00000125482 | 0.230 | 4.082 | 3.488 | 0.00052 | 0.00899 | -0.868 | TTF1 | transcription termination factor 1 [Source:HGNC Symbol;Acc:HGNC:12397] |
| ENSG00000125870 | 0.246 | 5.375 | 3.539 | 0.00043 | 0.00776 | -0.792 | SNRPB2 | small nuclear ribonucleoprotein polypeptide B2 [Source:HGNC Symbol;Acc:HGNC:11155] |
| ENSG00000125885 | 0.417 | 2.771 | 4.223 | 0.00003 | 0.00088 | 2.031 | MCM8 | minichromosome maintenance 8 homologous recombination repair factor [Source:HGNC Symbol;Acc:HGNC:16147] |
| ENSG00000126067 | 0.238 | 6.402 | 3.972 | 0.00008 | 0.00201 | 0.784 | PSMB2 | proteasome subunit beta 2 [Source:HGNC Symbol;Acc:HGNC:9539] |
| ENSG00000126698 | 0.247 | 6.028 | 4.394 | 0.00001 | 0.00047 | 2.480 | DNAJC8 | DnaJ heat shock protein family (Hsp40) member C8 [Source:HGNC Symbol;Acc:HGNC:15470] |
| ENSG00000126767 | 0.300 | 4.760 | 4.622 | 0.00000 | 0.00020 | 3.501 | ELK1 | ELK1, ETS transcription factor [Source:HGNC Symbol;Acc:HGNC:3321] |
| ENSG00000126787 | 1.220 | 1.470 | 6.928 | 0.00000 | 0.00000 | 16.096 | DLGAP5 | DLG associated protein 5 [Source:HGNC Symbol;Acc:HGNC:16864] |

| | | | | | | | | |
|---|---|---|---|---|---|---|---|---|
| ENSG00000126953 | 0.496 | 2.511 | 5.157 | 0.00000 | 0.00002 | 6.210 | TIMM8A | translocase of inner mitochondrial membrane 8A [Source:HGNC Symbol;Acc:HGNC:11817] |
| ENSG00000127191 | 0.332 | 4.039 | 3.645 | 0.00029 | 0.00583 | -0.330 | TRAF2 | TNF receptor associated factor 2 [Source:HGNC Symbol;Acc:HGNC:12032] |
| ENSG00000127314 | -0.311 | 7.004 | -3.470 | 0.00056 | 0.00949 | -1.026 | RAP1B | RAP1B, member of RAS oncogene family [Source:HGNC Symbol;Acc:HGNC:9857] |
| ENSG00000127399 | 0.560 | 1.821 | 5.488 | 0.00000 | 0.00000 | 7.907 | LRRC61 | leucine rich repeat containing 61 [Source:HGNC Symbol;Acc:HGNC:21704] |
| ENSG00000127564 | 0.768 | 2.010 | 5.190 | 0.00000 | 0.00002 | 6.397 | PKMYT1 | protein kinase, membrane associated tyrosine/threonine 1 [Source:HGNC Symbol;Acc:HGNC:29650] |
| ENSG00000127804 | -0.272 | 5.113 | -4.183 | 0.00003 | 0.00100 | 1.650 | METTL16 | methyltransferase like 16 [Source:HGNC Symbol;Acc:HGNC:28484] |
| ENSG00000128708 | 0.252 | 5.124 | 3.593 | 0.00035 | 0.00669 | -0.595 | HAT1 | histone acetyltransferase 1 [Source:HGNC Symbol;Acc:HGNC:4821] |
| ENSG00000129083 | 0.219 | 7.635 | 3.814 | 0.00015 | 0.00344 | 0.248 | COPB1 | coatomer protein complex subunit beta 1 [Source:HGNC Symbol;Acc:HGNC:2231] |
| ENSG00000129173 | 1.273 | 1.041 | 7.202 | 0.00000 | 0.00000 | 17.807 | E2F8 | E2F transcription factor 8 [Source:HGNC Symbol;Acc:HGNC:24727] |
| ENSG00000129197 | -0.420 | 4.928 | -7.426 | 0.00000 | 0.00000 | 19.268 | RPAIN | RPA interacting protein [Source:HGNC Symbol;Acc:HGNC:28641] |
| ENSG00000129235 | -0.333 | 4.505 | -4.167 | 0.00004 | 0.00105 | 1.649 | TXNDC17 | thioredoxin domain containing 17 [Source:HGNC Symbol;Acc:HGNC:28218] |
| ENSG00000129245 | -0.462 | 5.055 | -6.610 | 0.00000 | 0.00000 | 14.024 | FXR2 | FMR1 autosomal homolog 2 [Source:HGNC Symbol;Acc:HGNC:4024] |
| ENSG00000129250 | -0.329 | 6.340 | -3.867 | 0.00012 | 0.00286 | 0.378 | KIF1C | kinesin family member 1C [Source:HGNC Symbol;Acc:HGNC:6317] |
| ENSG00000129255 | -0.356 | 5.241 | -4.211 | 0.00003 | 0.00091 | 1.760 | MPDU1 | mannose-P-dolichol utilization defect 1 [Source:HGNC Symbol;Acc:HGNC:7207] |
| ENSG00000129657 | -0.316 | 8.761 | -4.295 | 0.00002 | 0.00067 | 2.172 | SEC14L1 | SEC14 like lipid binding 1 [Source:HGNC Symbol;Acc:HGNC:10698] |
| ENSG00000129667 | -0.335 | 6.841 | -4.184 | 0.00003 | 0.00100 | 1.615 | RHBDF2 | rhomboid 5 homolog 2 [Source:HGNC Symbol;Acc:HGNC:20788] |
| ENSG00000129810 | 0.865 | 0.574 | 6.711 | 0.00000 | 0.00000 | 14.630 | SGO1 | shugoshin 1 [Source:HGNC Symbol;Acc:HGNC:25088] |
| ENSG00000129993 | -0.483 | 5.737 | -3.807 | 0.00016 | 0.00353 | 0.170 | CBFA2T3 | CBFA2/RUNX1 translocation partner 3 [Source:HGNC Symbol;Acc:HGNC:1537] |
| ENSG00000130270 | 0.542 | 1.580 | 4.113 | 0.00004 | 0.00126 | 1.757 | ATP8B3 | ATPase phospholipid transporting 8B3 [Source:HGNC Symbol;Acc:HGNC:13535] |
| ENSG00000130695 | 0.333 | 4.420 | 3.590 | 0.00036 | 0.00674 | -0.559 | CEP85 | centrosomal protein 85 [Source:HGNC Symbol;Acc:HGNC:25309] |
| ENSG00000130787 | -0.444 | 5.873 | -4.025 | 0.00006 | 0.00170 | 0.991 | HIP1R | huntingtin interacting protein 1 related [Source:HGNC Symbol;Acc:HGNC:18415] |
| ENSG00000130816 | 0.336 | 6.317 | 4.212 | 0.00003 | 0.00091 | 1.733 | DNMT1 | DNA methyltransferase 1 [Source:HGNC Symbol;Acc:HGNC:2976] |
| ENSG00000131080 | -1.340 | 3.268 | -4.931 | 0.00000 | 0.00006 | 5.222 | EDA2R | ectodysplasin A2 receptor [Source:HGNC Symbol;Acc:HGNC:17756] |
| ENSG00000131153 | 0.698 | 1.902 | 5.073 | 0.00000 | 0.00003 | 5.853 | GINS2 | GINS complex subunit 2 [Source:HGNC Symbol;Acc:HGNC:24575] |
| ENSG00000131269 | 0.222 | 4.291 | 4.385 | 0.00001 | 0.00048 | 2.520 | ABCB7 | ATP binding cassette subfamily B member 7 [Source:HGNC Symbol;Acc:HGNC:48] |
| ENSG00000131351 | 0.308 | 2.455 | 3.584 | 0.00037 | 0.00687 | -0.293 | HAUS8 | HAUS augmin like complex subunit 8 [Source:HGNC Symbol;Acc:HGNC:30532] |
| ENSG00000131747 | 1.128 | 3.512 | 5.890 | 0.00000 | 0.00000 | 9.854 | TOP2A | topoisomerase (DNA) II alpha [Source:HGNC Symbol;Acc:HGNC:11989] |
| ENSG00000131944 | 0.412 | 0.761 | 4.195 | 0.00003 | 0.00096 | 2.148 | FAAP24 | Fanconi anemia core complex associated protein 24 [Source:HGNC Symbol;Acc:HGNC:28467] |
| ENSG00000132275 | 0.273 | 4.060 | 3.814 | 0.00015 | 0.00344 | 0.282 | RRP8 | ribosomal RNA processing 8, methyltransferase, homolog (yeast) [Source:HGNC Symbol;Acc:HGNC:29030] |
| ENSG00000132361 | -0.341 | 6.569 | -4.869 | 0.00000 | 0.00007 | 4.587 | CLUH | clustered mitochondria homolog [Source:HGNC Symbol;Acc:HGNC:29094] |
| ENSG00000132376 | -0.418 | 5.617 | -6.236 | 0.00000 | 0.00000 | 11.763 | INPP5K | inositol polyphosphate-5-phosphatase K [Source:HGNC Symbol;Acc:HGNC:33882] |
| ENSG00000132382 | -0.441 | 6.852 | -5.759 | 0.00000 | 0.00000 | 9.073 | MYBBP1A | MYB binding protein 1a [Source:HGNC Symbol;Acc:HGNC:7546] |
| ENSG00000132388 | -0.461 | 7.229 | -5.756 | 0.00000 | 0.00000 | 9.063 | UBE2G1 | ubiquitin conjugating enzyme E2 G1 [Source:HGNC Symbol;Acc:HGNC:12482] |
| ENSG00000132436 | 0.692 | 3.067 | 6.509 | 0.00000 | 0.00000 | 13.487 | FIGNL1 | fidgetin like 1 [Source:HGNC Symbol;Acc:HGNC:13286] |
| ENSG00000132471 | -0.298 | 6.988 | -5.008 | 0.00000 | 0.00004 | 5.252 | WBP2 | WW domain binding protein 2 [Source:HGNC Symbol;Acc:HGNC:12738] |
| ENSG00000132507 | -0.317 | 7.761 | -4.535 | 0.00001 | 0.00028 | 3.125 | EIF5A | eukaryotic translation initiation factor 5A [Source:HGNC Symbol;Acc:HGNC:3300] |
| ENSG00000132510 | -0.544 | 5.552 | -4.192 | 0.00003 | 0.00097 | 1.673 | KDM6B | lysine demethylase 6B [Source:HGNC Symbol;Acc:HGNC:29012] |
| ENSG00000132522 | -0.643 | 1.194 | -7.325 | 0.00000 | 0.00000 | 18.235 | GPS2 | G protein pathway suppressor 2 [Source:HGNC Symbol;Acc:HGNC:4550] |
| ENSG00000132535 | -0.817 | 3.220 | -3.681 | 0.00025 | 0.00524 | 0.076 | DLG4 | discs large MAGUK scaffold protein 4 [Source:HGNC Symbol;Acc:HGNC:2903] |
| ENSG00000132646 | 0.625 | 5.095 | 4.733 | 0.00000 | 0.00013 | 3.969 | PCNA | proliferating cell nuclear antigen [Source:HGNC Symbol;Acc:HGNC:8729] |
| ENSG00000133026 | -0.709 | 3.215 | -4.950 | 0.00000 | 0.00005 | 5.257 | MYH10 | myosin heavy chain 10 [Source:HGNC Symbol;Acc:HGNC:7568] |
| ENSG00000133028 | -0.253 | 4.979 | -3.902 | 0.00011 | 0.00255 | 0.562 | SCO1 | SCO1, cytochrome c oxidase assembly protein [Source:HGNC Symbol;Acc:HGNC:10603] |
| ENSG00000133030 | -0.223 | 7.354 | -3.640 | 0.00030 | 0.00590 | -0.421 | MPRIP | myosin phosphatase Rho interacting protein [Source:HGNC Symbol;Acc:HGNC:30321] |
| ENSG00000133119 | 0.615 | 2.656 | 5.610 | 0.00000 | 0.00000 | 8.472 | RFC3 | replication factor C subunit 3 [Source:HGNC Symbol;Acc:HGNC:9971] |
| ENSG00000133318 | 0.325 | 7.472 | 3.699 | 0.00024 | 0.00497 | -0.175 | RTN3 | reticulon 3 [Source:HGNC Symbol;Acc:HGNC:10469] |
| ENSG00000133321 | 0.603 | 3.479 | 3.575 | 0.00038 | 0.00707 | -0.526 | RARRES3 | retinoic acid receptor responder 3 [Source:HGNC Symbol;Acc:HGNC:9869] |
| ENSG00000133393 | 0.310 | 5.007 | 4.621 | 0.00000 | 0.00020 | 3.481 | FOPNL | FGFR1OP N-terminal like [Source:HGNC Symbol;Acc:HGNC:26435] |
| ENSG00000134001 | 0.276 | 6.218 | 3.600 | 0.00034 | 0.00652 | -0.584 | EIF2S1 | eukaryotic translation initiation factor 2 subunit alpha [Source:HGNC Symbol;Acc:HGNC:3265] |
| ENSG00000134057 | 0.860 | 2.797 | 6.347 | 0.00000 | 0.00000 | 12.534 | CCNB1 | cyclin B1 [Source:HGNC Symbol;Acc:HGNC:1579] |
| ENSG00000134146 | 0.340 | 2.575 | 3.944 | 0.00009 | 0.00222 | 0.983 | DPH6 | diphthamine biosynthesis 6 [Source:HGNC Symbol;Acc:HGNC:30543] |
| ENSG00000134222 | 0.575 | 0.038 | 3.487 | 0.00052 | 0.00902 | -0.344 | PSRC1 | proline and serine rich coiled-coil 1 [Source:HGNC Symbol;Acc:HGNC:24472] |
| ENSG00000134242 | 0.837 | 2.183 | 3.736 | 0.00020 | 0.00446 | 0.219 | PTPN22 | protein tyrosine phosphatase, non-receptor type 22 [Source:HGNC Symbol;Acc:HGNC:9652] |
| ENSG00000134291 | 0.381 | 5.249 | 3.973 | 0.00008 | 0.00201 | 0.787 | TMEM106C | transmembrane protein 106C [Source:HGNC Symbol;Acc:HGNC:28775] |
| ENSG00000134597 | 0.291 | 4.227 | 3.632 | 0.00030 | 0.00601 | -0.391 | RBMX2 | RNA binding motif protein, X-linked 2 [Source:HGNC Symbol;Acc:HGNC:24282] |
| ENSG00000134690 | 1.000 | 1.960 | 7.257 | 0.00000 | 0.00000 | 18.212 | CDCA8 | cell division cycle associated 8 [Source:HGNC Symbol;Acc:HGNC:14629] |
| ENSG00000134779 | 0.353 | 4.638 | 4.091 | 0.00005 | 0.00136 | 1.283 | TPGS2 | tubulin polyglutamylase complex subunit 2 [Source:HGNC Symbol;Acc:HGNC:24561] |

| | | | | | | | | |
|---|---|---|---|---|---|---|---|---|
| ENSG00000134809 | 0.350 | 4.364 | 4.222 | 0.00003 | 0.00088 | 1.829 | TIMM10 | translocase of inner mitochondrial membrane 10 [Source:HGNC Symbol;Acc:HGNC:11814] |
| ENSG00000134824 | 0.658 | 3.090 | 3.526 | 0.00045 | 0.00805 | -0.637 | FADS2 | fatty acid desaturase 2 [Source:HGNC Symbol;Acc:HGNC:3575] |
| ENSG00000135045 | 0.428 | 2.501 | 3.800 | 0.00016 | 0.00360 | 0.448 | C9orf40 | chromosome 9 open reading frame 40 [Source:HGNC Symbol;Acc:HGNC:23433] |
| ENSG00000135404 | -0.525 | 8.543 | -4.238 | 0.00003 | 0.00083 | 1.917 | CD63 | CD63 molecule [Source:HGNC Symbol;Acc:HGNC:1692] |
| ENSG00000135451 | 0.836 | 2.003 | 5.249 | 0.00000 | 0.00001 | 6.683 | TROAP | trophinin associated protein [Source:HGNC Symbol;Acc:HGNC:12327] |
| ENSG00000135476 | 0.734 | 2.596 | 5.202 | 0.00000 | 0.00002 | 6.397 | ESPL1 | extra spindle pole bodies like 1, separase [Source:HGNC Symbol;Acc:HGNC:16856] |
| ENSG00000135679 | -0.409 | 6.933 | -3.931 | 0.00009 | 0.00233 | 0.625 | MDM2 | MDM2 proto-oncogene [Source:HGNC Symbol;Acc:HGNC:6973] |
| ENSG00000136108 | 0.474 | 4.376 | 4.266 | 0.00002 | 0.00074 | 1.999 | CKAP2 | cytoskeleton associated protein 2 [Source:HGNC Symbol;Acc:HGNC:1990] |
| ENSG00000136193 | -0.499 | 7.064 | -3.773 | 0.00018 | 0.00395 | 0.036 | SCRN1 | secernin 1 [Source:HGNC Symbol;Acc:HGNC:22192] |
| ENSG00000136492 | 0.585 | 2.004 | 4.240 | 0.00003 | 0.00083 | 2.193 | BRIP1 | BRCA1 interacting protein C-terminal helicase 1 [Source:HGNC Symbol;Acc:HGNC:20473] |
| ENSG00000136514 | 0.749 | 1.640 | 3.778 | 0.00017 | 0.00389 | 0.454 | RTP4 | receptor transporter protein 4 [Source:HGNC Symbol;Acc:HGNC:23992] |
| ENSG00000136824 | 0.530 | 4.626 | 4.692 | 0.00000 | 0.00015 | 3.808 | SMC2 | structural maintenance of chromosomes 2 [Source:HGNC Symbol;Acc:HGNC:14011] |
| ENSG00000136840 | -0.347 | 7.417 | -3.457 | 0.00058 | 0.00982 | -1.052 | ST6GALNAC4 | ST6 N-acetylgalactosaminide alpha-2,6-sialyltransferase 4 [Source:HGNC Symbol;Acc:HGNC:17846] |
| ENSG00000136982 | 0.960 | 0.569 | 6.084 | 0.00000 | 0.00000 | 11.045 | DSCC1 | DNA replication and sister chromatid cohesion 1 [Source:HGNC Symbol;Acc:HGNC:24453] |
| ENSG00000137310 | 0.814 | 4.129 | 5.492 | 0.00000 | 0.00000 | 7.697 | TCF19 | transcription factor 19 [Source:HGNC Symbol;Acc:HGNC:11629] |
| ENSG00000137547 | 0.348 | 4.733 | 3.672 | 0.00026 | 0.00538 | -0.298 | MRPL15 | mitochondrial ribosomal protein L15 [Source:HGNC Symbol;Acc:HGNC:14054] |
| ENSG00000137563 | 0.583 | 5.122 | 3.495 | 0.00051 | 0.00881 | -0.941 | GGH | gamma-glutamyl hydrolase [Source:HGNC Symbol;Acc:HGNC:4248] |
| ENSG00000137574 | 0.295 | 4.382 | 4.645 | 0.00000 | 0.00018 | 3.633 | TGS1 | trimethylguanosine synthase 1 [Source:HGNC Symbol;Acc:HGNC:17843] |
| ENSG00000137656 | 0.320 | 4.051 | 4.482 | 0.00001 | 0.00034 | 2.946 | BUD13 | BUD13 homolog [Source:HGNC Symbol;Acc:HGNC:28199] |
| ENSG00000137713 | 0.261 | 4.689 | 3.625 | 0.00031 | 0.00612 | -0.456 | PPP2R1B | protein phosphatase 2 scaffold subunit Abeta [Source:HGNC Symbol;Acc:HGNC:9303] |
| ENSG00000137807 | 1.102 | 2.223 | 6.885 | 0.00000 | 0.00000 | 15.826 | KIF23 | kinesin family member 23 [Source:HGNC Symbol;Acc:HGNC:6392] |
| ENSG00000137812 | 0.712 | 3.263 | 5.222 | 0.00000 | 0.00002 | 6.417 | KNL1 | kinetochore scaffold 1 [Source:HGNC Symbol;Acc:HGNC:24054] |
| ENSG00000137959 | 1.533 | 2.157 | 5.287 | 0.00000 | 0.00001 | 6.802 | IFI44L | interferon induced protein 44 like [Source:HGNC Symbol;Acc:HGNC:17817] |
| ENSG00000138031 | 0.646 | 3.181 | 4.272 | 0.00002 | 0.00073 | 2.147 | ADCY3 | adenylate cyclase 3 [Source:HGNC Symbol;Acc:HGNC:234] |
| ENSG00000138092 | 0.557 | 1.922 | 6.357 | 0.00000 | 0.00000 | 12.650 | CENPO | centromere protein O [Source:HGNC Symbol;Acc:HGNC:28152] |
| ENSG00000138160 | 0.985 | 3.251 | 7.224 | 0.00000 | 0.00000 | 17.952 | KIF11 | kinesin family member 11 [Source:HGNC Symbol;Acc:HGNC:6388] |
| ENSG00000138180 | 1.193 | 1.928 | 7.417 | 0.00000 | 0.00000 | 19.273 | CEP55 | centrosomal protein 55 [Source:HGNC Symbol;Acc:HGNC:1161] |
| ENSG00000138182 | 0.584 | 3.214 | 5.620 | 0.00000 | 0.00000 | 8.470 | KIF20B | kinesin family member 20B [Source:HGNC Symbol;Acc:HGNC:7212] |
| ENSG00000138346 | 0.371 | 2.620 | 3.837 | 0.00014 | 0.00320 | 0.568 | DNA2 | DNA replication helicase/nuclease 2 [Source:HGNC Symbol;Acc:HGNC:2939] |
| ENSG00000138594 | 0.242 | 4.651 | 3.455 | 0.00059 | 0.00987 | -1.037 | TMOD3 | tropomodulin 3 [Source:HGNC Symbol;Acc:HGNC:11873] |
| ENSG00000138663 | 0.245 | 4.875 | 3.643 | 0.00029 | 0.00586 | -0.407 | COPS4 | COP9 signalosome subunit 4 [Source:HGNC Symbol;Acc:HGNC:16702] |
| ENSG00000138756 | 0.616 | 3.276 | 3.507 | 0.00049 | 0.00847 | -0.723 | BMP2K | BMP2 inducible kinase [Source:HGNC Symbol;Acc:HGNC:18041] |
| ENSG00000138778 | 0.852 | 2.237 | 6.024 | 0.00000 | 0.00000 | 10.734 | CENPE | centromere protein E [Source:HGNC Symbol;Acc:HGNC:1856] |
| ENSG00000138796 | 0.395 | 5.166 | 4.103 | 0.00005 | 0.00130 | 1.298 | HADH | hydroxyacyl-CoA dehydrogenase [Source:HGNC Symbol;Acc:HGNC:4799] |
| ENSG00000139190 | -0.532 | 5.537 | -3.668 | 0.00027 | 0.00543 | -0.317 | VAMP1 | vesicle associated membrane protein 1 [Source:HGNC Symbol;Acc:HGNC:12642] |
| ENSG00000139631 | -0.342 | 4.824 | -3.901 | 0.00011 | 0.00255 | 0.582 | CSAD | cysteine sulfinic acid decarboxylase [Source:HGNC Symbol;Acc:HGNC:18966] |
| ENSG00000139722 | -0.481 | 7.084 | -3.764 | 0.00018 | 0.00404 | 0.005 | VPS37B | VPS37B, ESCRT-I subunit [Source:HGNC Symbol;Acc:HGNC:25754] |
| ENSG00000139734 | 1.053 | 0.714 | 6.634 | 0.00000 | 0.00000 | 14.220 | DIAPH3 | diaphanous related formin 3 [Source:HGNC Symbol;Acc:HGNC:15480] |
| ENSG00000140398 | -0.870 | 5.111 | -4.896 | 0.00000 | 0.00006 | 4.795 | NEIL1 | nei like DNA glycosylase 1 [Source:HGNC Symbol;Acc:HGNC:18448] |
| ENSG00000140400 | -0.454 | 7.505 | -4.949 | 0.00000 | 0.00005 | 4.986 | MAN2C1 | mannosidase alpha class 2C member 1 [Source:HGNC Symbol;Acc:HGNC:6827] |
| ENSG00000140525 | 0.587 | 4.608 | 4.985 | 0.00000 | 0.00004 | 5.160 | FANCI | Fanconi anemia complementation group I [Source:HGNC Symbol;Acc:HGNC:25568] |
| ENSG00000140534 | 1.075 | 0.819 | 6.393 | 0.00000 | 0.00000 | 12.822 | TICRR | TOPBP1 interacting checkpoint and replication regulator [Source:HGNC Symbol;Acc:HGNC:28704] |
| ENSG00000141127 | -0.275 | 5.460 | -3.772 | 0.00018 | 0.00395 | 0.050 | PRPSAP2 | phosphoribosyl pyrophosphate synthetase associated protein 2 [Source:HGNC Symbol;Acc:HGNC:9467] |
| ENSG00000141252 | -0.461 | 5.402 | -8.052 | 0.00000 | 0.00000 | 23.618 | VPS53 | VPS53, GARP complex subunit [Source:HGNC Symbol;Acc:HGNC:25608] |
| ENSG00000141258 | -0.633 | 5.661 | -6.704 | 0.00000 | 0.00000 | 14.580 | SGSM2 | small G protein signaling modulator 2 [Source:HGNC Symbol;Acc:HGNC:29026] |
| ENSG00000141447 | 1.195 | 2.124 | 3.709 | 0.00023 | 0.00485 | 0.096 | OSBPL1A | oxysterol binding protein like 1A [Source:HGNC Symbol;Acc:HGNC:16398] |
| ENSG00000141456 | -0.352 | 5.834 | -5.045 | 0.00000 | 0.00003 | 5.428 | PELP1 | proline, glutamate and leucine rich protein 1 [Source:HGNC Symbol;Acc:HGNC:30134] |
| ENSG00000141499 | -0.366 | 3.657 | -5.818 | 0.00000 | 0.00000 | 9.557 | WRAP53 | WD repeat containing antisense to TP53 [Source:HGNC Symbol;Acc:HGNC:25522] |
| ENSG00000141503 | -0.490 | 6.142 | -7.032 | 0.00000 | 0.00000 | 16.636 | MINK1 | misshapen like kinase 1 [Source:HGNC Symbol;Acc:HGNC:17565] |
| ENSG00000141504 | -0.613 | 5.932 | -8.597 | 0.00000 | 0.00000 | 27.644 | SAT2 | spermidine/spermine N1-acetyltransferase family member 2 [Source:HGNC Symbol;Acc:HGNC:23160] |
| ENSG00000141642 | 0.324 | 3.280 | 4.431 | 0.00001 | 0.00042 | 2.825 | ELAC1 | elaC ribonuclease Z 1 [Source:HGNC Symbol;Acc:HGNC:14197] |
| ENSG00000142230 | 0.348 | 5.676 | 4.426 | 0.00001 | 0.00042 | 2.614 | SAE1 | SUMO1 activating enzyme subunit 1 [Source:HGNC Symbol;Acc:HGNC:30660] |
| ENSG00000142507 | -0.311 | 6.299 | -4.970 | 0.00000 | 0.00005 | 5.060 | PSMB6 | proteasome subunit beta 6 [Source:HGNC Symbol;Acc:HGNC:9543] |
| ENSG00000142731 | 0.857 | 2.260 | 6.781 | 0.00000 | 0.00000 | 15.183 | PLK4 | polo like kinase 4 [Source:HGNC Symbol;Acc:HGNC:11397] |
| ENSG00000142856 | 0.347 | 4.087 | 4.079 | 0.00005 | 0.00142 | 1.283 | ITGB3BP | integrin subunit beta 3 binding protein [Source:HGNC Symbol;Acc:HGNC:6157] |
| ENSG00000142945 | 1.049 | 1.567 | 7.567 | 0.00000 | 0.00000 | 20.249 | KIF2C | kinesin family member 2C [Source:HGNC Symbol;Acc:HGNC:6393] |

| | | | | | | | | |
|---|---|---|---|---|---|---|---|---|
| ENSG00000143228 | 0.870 | 2.132 | 5.655 | 0.00000 | 0.00000 | 8.735 | NUF2 | NUF2, NDC80 kinetochore complex component [Source:HGNC Symbol;Acc:HGNC:14621] |
| ENSG00000143344 | 1.093 | 2.183 | 4.110 | 0.00004 | 0.00127 | 1.620 | RGL1 | ral guanine nucleotide dissociation stimulator like 1 [Source:HGNC Symbol;Acc:HGNC:30281] |
| ENSG00000143476 | 0.860 | 2.728 | 5.455 | 0.00000 | 0.00001 | 7.636 | DTL | denticleless E3 ubiquitin protein ligase homolog [Source:HGNC Symbol;Acc:HGNC:30288] |
| ENSG00000143977 | 0.309 | 5.041 | 4.268 | 0.00002 | 0.00074 | 1.974 | SNRPG | small nuclear ribonucleoprotein polypeptide G [Source:HGNC Symbol;Acc:HGNC:11163] |
| ENSG00000144034 | 0.252 | 4.538 | 3.958 | 0.00008 | 0.00212 | 0.781 | TPRKB | TP53RK binding protein [Source:HGNC Symbol;Acc:HGNC:24259] |
| ENSG00000144395 | 0.814 | 0.281 | 5.193 | 0.00000 | 0.00002 | 6.448 | CCDC150 | coiled-coil domain containing 150 [Source:HGNC Symbol;Acc:HGNC:26834] |
| ENSG00000145147 | 1.289 | 1.292 | 4.844 | 0.00000 | 0.00008 | 4.816 | SLIT2 | slit guidance ligand 2 [Source:HGNC Symbol;Acc:HGNC:11086] |
| ENSG00000145375 | 0.321 | 2.960 | 4.187 | 0.00003 | 0.00099 | 1.869 | SPATA5 | spermatogenesis associated 5 [Source:HGNC Symbol;Acc:HGNC:18119] |
| ENSG00000145386 | 1.065 | 1.834 | 7.579 | 0.00000 | 0.00000 | 20.349 | CCNA2 | cyclin A2 [Source:HGNC Symbol;Acc:HGNC:1578] |
| ENSG00000146063 | -0.227 | 5.804 | -3.701 | 0.00023 | 0.00493 | -0.224 | TRIM41 | tripartite motif containing 41 [Source:HGNC Symbol;Acc:HGNC:19013] |
| ENSG00000146143 | 0.377 | 3.008 | 5.065 | 0.00000 | 0.00003 | 5.717 | PRIM2 | primase (DNA) subunit 2 [Source:HGNC Symbol;Acc:HGNC:9370] |
| ENSG00000146416 | -0.387 | 6.532 | -3.529 | 0.00045 | 0.00797 | -0.835 | AIG1 | androgen induced 1 [Source:HGNC Symbol;Acc:HGNC:21607] |
| ENSG00000146670 | 1.059 | 2.952 | 7.764 | 0.00000 | 0.00000 | 21.621 | CDCA5 | cell division cycle associated 5 [Source:HGNC Symbol;Acc:HGNC:14626] |
| ENSG00000146842 | 0.309 | 4.920 | 3.672 | 0.00026 | 0.00538 | -0.309 | TMEM209 | transmembrane protein 209 [Source:HGNC Symbol;Acc:HGNC:21898] |
| ENSG00000146859 | 0.322 | 4.973 | 3.544 | 0.00042 | 0.00766 | -0.760 | TMEM140 | transmembrane protein 140 [Source:HGNC Symbol;Acc:HGNC:21870] |
| ENSG00000146918 | 0.681 | 3.966 | 5.376 | 0.00000 | 0.00001 | 7.121 | NCAPG2 | non-SMC condensin II complex subunit G2 [Source:HGNC Symbol;Acc:HGNC:21904] |
| ENSG00000147155 | 0.314 | 3.439 | 4.686 | 0.00000 | 0.00015 | 3.916 | EBP | emopamil binding protein (sterol isomerase) [Source:HGNC Symbol;Acc:HGNC:3133] |
| ENSG00000147164 | 0.217 | 4.686 | 4.329 | 0.00002 | 0.00059 | 2.253 | SNX12 | sorting nexin 12 [Source:HGNC Symbol;Acc:HGNC:14976] |
| ENSG00000147536 | 0.918 | 0.791 | 7.098 | 0.00000 | 0.00000 | 17.053 | GINS4 | GINS complex subunit 4 [Source:HGNC Symbol;Acc:HGNC:28226] |
| ENSG00000147570 | 0.815 | 0.555 | 3.911 | 0.00010 | 0.00247 | 1.073 | DNAJC5B | DnaJ heat shock protein family (Hsp40) member C5 beta [Source:HGNC Symbol;Acc:HGNC:24138] |
| ENSG00000147883 | 1.158 | 1.447 | 7.094 | 0.00000 | 0.00000 | 17.131 | CDKN2B | cyclin dependent kinase inhibitor 2B [Source:HGNC Symbol;Acc:HGNC:1788] |
| ENSG00000148153 | 0.396 | 4.366 | 3.601 | 0.00034 | 0.00652 | -0.524 | INIP | INTS3 and NABP interacting protein [Source:HGNC Symbol;Acc:HGNC:24994] |
| ENSG00000148688 | 0.203 | 4.505 | 4.037 | 0.00006 | 0.00164 | 1.092 | RPP30 | ribonuclease P/MRP subunit p30 [Source:HGNC Symbol;Acc:HGNC:17688] |
| ENSG00000148773 | 1.153 | 4.326 | 6.024 | 0.00000 | 0.00000 | 10.554 | MKI67 | marker of proliferation Ki-67 [Source:HGNC Symbol;Acc:HGNC:7107] |
| ENSG00000148908 | 0.780 | 1.994 | 3.450 | 0.00060 | 0.01000 | -0.724 | RGS10 | regulator of G protein signaling 10 [Source:HGNC Symbol;Acc:HGNC:9992] |
| ENSG00000148925 | 0.303 | 4.009 | 4.443 | 0.00001 | 0.00040 | 2.787 | BTBD10 | BTB domain containing 10 [Source:HGNC Symbol;Acc:HGNC:21445] |
| ENSG00000148950 | 0.225 | 2.713 | 3.558 | 0.00040 | 0.00739 | -0.419 | IMMP1L | inner mitochondrial membrane peptidase subunit 1 [Source:HGNC Symbol;Acc:HGNC:26317] |
| ENSG00000149136 | 0.251 | 6.984 | 4.979 | 0.00000 | 0.00005 | 5.132 | SSRP1 | structure specific recognition protein 1 [Source:HGNC Symbol;Acc:HGNC:11327] |
| ENSG00000149503 | 0.479 | 3.904 | 5.840 | 0.00000 | 0.00000 | 9.591 | INCENP | inner centromere protein [Source:HGNC Symbol;Acc:HGNC:6058] |
| ENSG00000149548 | 1.162 | 0.330 | 7.132 | 0.00000 | 0.00000 | 17.212 | CCDC15 | coiled-coil domain containing 15 [Source:HGNC Symbol;Acc:HGNC:25798] |
| ENSG00000149554 | 0.651 | 3.628 | 6.338 | 0.00000 | 0.00000 | 12.430 | CHEK1 | checkpoint kinase 1 [Source:HGNC Symbol;Acc:HGNC:1925] |
| ENSG00000150477 | 0.324 | 2.575 | 3.881 | 0.00012 | 0.00276 | 0.750 | KIAA1328 | KIAA1328 [Source:HGNC Symbol;Acc:HGNC:29248] |
| ENSG00000150768 | 0.317 | 5.506 | 3.495 | 0.00051 | 0.00883 | -0.949 | DLAT | dihydrolipoamide S-acetyltransferase [Source:HGNC Symbol;Acc:HGNC:2896] |
| ENSG00000150776 | 0.288 | 5.261 | 3.782 | 0.00017 | 0.00383 | 0.074 | C11orf57 | chromosome 11 open reading frame 57 [Source:HGNC Symbol;Acc:HGNC:25569] |
| ENSG00000151287 | 0.403 | 0.903 | 3.530 | 0.00045 | 0.00796 | -0.256 | TEX30 | testis expressed 30 [Source:HGNC Symbol;Acc:HGNC:25188] |
| ENSG00000151465 | 0.193 | 4.864 | 3.646 | 0.00029 | 0.00581 | -0.391 | CDC123 | cell division cycle 123 [Source:HGNC Symbol;Acc:HGNC:16827] |
| ENSG00000151466 | 0.271 | 3.777 | 3.887 | 0.00011 | 0.00270 | 0.590 | SCLT1 | sodium channel and clathrin linker 1 [Source:HGNC Symbol;Acc:HGNC:26406] |
| ENSG00000151503 | 0.464 | 5.354 | 5.632 | 0.00000 | 0.00000 | 8.390 | NCAPD3 | non-SMC condensin II complex subunit D3 [Source:HGNC Symbol;Acc:HGNC:28952] |
| ENSG00000151611 | 0.270 | 1.915 | 3.576 | 0.00038 | 0.00706 | -0.223 | MMAA | methylmalonic aciduria (cobalamin deficiency) cblA type [Source:HGNC Symbol;Acc:HGNC:18871] |
| ENSG00000151725 | 0.702 | 3.773 | 5.909 | 0.00000 | 0.00000 | 9.967 | CENPU | centromere protein U [Source:HGNC Symbol;Acc:HGNC:21348] |
| ENSG00000152219 | 0.274 | 5.457 | 3.472 | 0.00055 | 0.00943 | -1.024 | ARL14EP | ADP ribosylation factor like GTPase 14 effector protein [Source:HGNC Symbol;Acc:HGNC:26798] |
| ENSG00000152240 | 0.366 | 3.501 | 4.704 | 0.00000 | 0.00014 | 3.985 | HAUS1 | HAUS augmin like complex subunit 1 [Source:HGNC Symbol;Acc:HGNC:25174] |
| ENSG00000152253 | 1.264 | 0.356 | 7.369 | 0.00000 | 0.00000 | 18.753 | SPC25 | SPC25, NDC80 kinetochore complex component [Source:HGNC Symbol;Acc:HGNC:24031] |
| ENSG00000153044 | 0.459 | 2.788 | 4.371 | 0.00001 | 0.00051 | 2.629 | CENPH | centromere protein H [Source:HGNC Symbol;Acc:HGNC:17268] |
| ENSG00000153944 | -0.328 | 7.685 | -3.824 | 0.00015 | 0.00334 | 0.258 | MSI2 | musashi RNA binding protein 2 [Source:HGNC Symbol;Acc:HGNC:18585] |
| ENSG00000154719 | 0.258 | 4.103 | 3.639 | 0.00030 | 0.00591 | -0.351 | MRPL39 | mitochondrial ribosomal protein L39 [Source:HGNC Symbol;Acc:HGNC:14027] |
| ENSG00000154839 | 1.312 | 1.486 | 9.434 | 0.00000 | 0.00000 | 34.073 | SKA1 | spindle and kinetochore associated complex subunit 1 [Source:HGNC Symbol;Acc:HGNC:28109] |
| ENSG00000154957 | -0.324 | 3.821 | -5.048 | 0.00000 | 0.00003 | 5.606 | ZNF18 | zinc finger protein 18 [Source:HGNC Symbol;Acc:HGNC:12969] |
| ENSG00000155085 | -0.375 | 3.704 | -3.679 | 0.00025 | 0.00528 | -0.079 | AK9 | adenylate kinase 9 [Source:HGNC Symbol;Acc:HGNC:33814] |
| ENSG00000155438 | 0.271 | 4.366 | 3.603 | 0.00034 | 0.00648 | -0.506 | NIFK | nucleolar protein interacting with the FHA domain of MKI67 [Source:HGNC Symbol;Acc:HGNC:17838] |
| ENSG00000156136 | 0.696 | 2.995 | 5.441 | 0.00000 | 0.00001 | 7.551 | DCK | deoxycytidine kinase [Source:HGNC Symbol;Acc:HGNC:2704] |
| ENSG00000156531 | 0.418 | 5.290 | 3.683 | 0.00025 | 0.00524 | -0.290 | PHF6 | PHD finger protein 6 [Source:HGNC Symbol;Acc:HGNC:18145] |
| ENSG00000156603 | 0.260 | 3.238 | 3.741 | 0.00020 | 0.00438 | 0.134 | MED19 | mediator complex subunit 19 [Source:HGNC Symbol;Acc:HGNC:29600] |
| ENSG00000156787 | 0.465 | 2.315 | 4.388 | 0.00001 | 0.00048 | 2.769 | TBC1D31 | TBC1 domain family member 31 [Source:HGNC Symbol;Acc:HGNC:30888] |
| ENSG00000156802 | 0.714 | 4.179 | 5.905 | 0.00000 | 0.00000 | 9.914 | ATAD2 | ATPase family, AAA domain containing 2 [Source:HGNC Symbol;Acc:HGNC:30123] |
| ENSG00000156876 | 0.415 | 2.545 | 4.920 | 0.00000 | 0.00006 | 5.096 | SASS6 | SAS-6 centriolar assembly protein [Source:HGNC Symbol;Acc:HGNC:25403] |

| | | | | | | | | |
|---|---|---|---|---|---|---|---|---|
| ENSG00000156970 | 0.850 | 2.964 | 5.670 | 0.00000 | 0.00000 | 8.735 | BUB1B | BUB1 mitotic checkpoint serine/threonine kinase B [Source:HGNC Symbol;Acc:HGNC:1149] |
| ENSG00000157191 | 0.355 | 5.136 | 4.062 | 0.00006 | 0.00150 | 1.139 | NECAP2 | NECAP endocytosis associated 2 [Source:HGNC Symbol;Acc:HGNC:25528] |
| ENSG00000157212 | 0.286 | 4.108 | 4.071 | 0.00005 | 0.00145 | 1.255 | PAXIP1 | PAX interacting protein 1 [Source:HGNC Symbol;Acc:HGNC:8624] |
| ENSG00000157353 | -0.392 | 4.827 | -4.648 | 0.00000 | 0.00018 | 3.652 | FUK | fucokinase [Source:HGNC Symbol;Acc:HGNC:29500] |
| ENSG00000157456 | 1.074 | 2.289 | 7.045 | 0.00000 | 0.00000 | 16.834 | CCNB2 | cyclin B2 [Source:HGNC Symbol;Acc:HGNC:1580] |
| ENSG00000158161 | 0.222 | 4.431 | 3.486 | 0.00053 | 0.00903 | -0.910 | EYA3 | EYA transcriptional coactivator and phosphatase 3 [Source:HGNC Symbol;Acc:HGNC:3521] |
| ENSG00000158402 | 0.893 | 0.513 | 6.152 | 0.00000 | 0.00000 | 11.413 | CDC25C | cell division cycle 25C [Source:HGNC Symbol;Acc:HGNC:1727] |
| ENSG00000158526 | 0.225 | 4.523 | 3.851 | 0.00013 | 0.00303 | 0.379 | TSR2 | TSR2, ribosome maturation factor [Source:HGNC Symbol;Acc:HGNC:25455] |
| ENSG00000158717 | -0.509 | 4.996 | -4.855 | 0.00000 | 0.00008 | 4.587 | RNF166 | ring finger protein 166 [Source:HGNC Symbol;Acc:HGNC:28856] |
| ENSG00000158769 | -0.505 | 4.938 | -4.593 | 0.00001 | 0.00022 | 3.410 | F11R | F11 receptor [Source:HGNC Symbol;Acc:HGNC:14685] |
| ENSG00000158773 | -0.354 | 7.526 | -3.927 | 0.00010 | 0.00235 | 0.634 | USF1 | upstream transcription factor 1 [Source:HGNC Symbol;Acc:HGNC:12593] |
| ENSG00000158792 | -0.409 | 2.920 | -4.392 | 0.00001 | 0.00047 | 2.804 | SPATA2L | spermatogenesis associated 2 like [Source:HGNC Symbol;Acc:HGNC:28393] |
| ENSG00000158805 | -0.351 | 5.430 | -4.053 | 0.00006 | 0.00155 | 1.117 | ZNF276 | zinc finger protein 276 [Source:HGNC Symbol;Acc:HGNC:23330] |
| ENSG00000159055 | 0.444 | 2.618 | 5.421 | 0.00000 | 0.00001 | 7.514 | MIS18A | MIS18 kinetochore protein A [Source:HGNC Symbol;Acc:HGNC:1286] |
| ENSG00000159147 | 0.278 | 4.231 | 3.856 | 0.00013 | 0.00299 | 0.421 | DONSON | downstream neighbor of SON [Source:HGNC Symbol;Acc:HGNC:2993] |
| ENSG00000159259 | 0.732 | 1.849 | 5.870 | 0.00000 | 0.00000 | 9.913 | CHAF1B | chromatin assembly factor 1 subunit B [Source:HGNC Symbol;Acc:HGNC:1911] |
| ENSG00000160298 | 0.596 | 2.421 | 4.909 | 0.00000 | 0.00006 | 5.034 | C21orf58 | chromosome 21 open reading frame 58 [Source:HGNC Symbol;Acc:HGNC:1300] |
| ENSG00000160408 | -0.578 | 1.814 | -3.747 | 0.00020 | 0.00430 | 0.462 | ST6GALNAC6 | ST6 N-acetylgalactosaminide alpha-2,6-sialyltransferase 6 [Source:HGNC Symbol;Acc:HGNC:23364] |
| ENSG00000160678 | -0.605 | 0.789 | -4.226 | 0.00003 | 0.00087 | 2.276 | S100A1 | S100 calcium binding protein A1 [Source:HGNC Symbol;Acc:HGNC:10486] |
| ENSG00000160957 | 0.465 | 3.600 | 3.876 | 0.00012 | 0.00279 | 0.550 | RECQL4 | RecQ like helicase 4 [Source:HGNC Symbol;Acc:HGNC:9949] |
| ENSG00000161010 | -0.401 | 6.192 | -3.932 | 0.00009 | 0.00232 | 0.625 | MRNIP | MRN complex interacting protein [Source:HGNC Symbol;Acc:HGNC:30817] |
| ENSG00000161057 | 0.311 | 6.662 | 4.736 | 0.00000 | 0.00013 | 3.993 | PSMC2 | proteasome 26S subunit, ATPase 2 [Source:HGNC Symbol;Acc:HGNC:9548] |
| ENSG00000161513 | -0.504 | 3.205 | -3.977 | 0.00008 | 0.00199 | 1.119 | FDXR | ferredoxin reductase [Source:HGNC Symbol;Acc:HGNC:3642] |
| ENSG00000161618 | -0.250 | 5.506 | -3.498 | 0.00050 | 0.00876 | -0.924 | ALDH16A1 | aldehyde dehydrogenase 16 family member A1 [Source:HGNC Symbol;Acc:HGNC:28114] |
| ENSG00000161692 | 0.364 | 2.294 | 3.565 | 0.00039 | 0.00723 | -0.331 | DBF4B | DBF4 zinc finger B [Source:HGNC Symbol;Acc:HGNC:17883] |
| ENSG00000161800 | 0.724 | 3.468 | 5.790 | 0.00000 | 0.00000 | 9.341 | RACGAP1 | Rac GTPase activating protein 1 [Source:HGNC Symbol;Acc:HGNC:9804] |
| ENSG00000161888 | 1.008 | 1.586 | 6.710 | 0.00000 | 0.00000 | 14.751 | SPC24 | SPC24, NDC80 kinetochore complex component [Source:HGNC Symbol;Acc:HGNC:26913] |
| ENSG00000161920 | -0.405 | 2.510 | -3.892 | 0.00011 | 0.00264 | 0.894 | MED11 | mediator complex subunit 11 [Source:HGNC Symbol;Acc:HGNC:32687] |
| ENSG00000161956 | -0.393 | 3.694 | -7.734 | 0.00000 | 0.00000 | 21.429 | SENP3 | SUMO1/sentrin/SMT3 specific peptidase 3 [Source:HGNC Symbol;Acc:HGNC:17862] |
| ENSG00000161960 | -0.677 | 4.318 | -4.974 | 0.00000 | 0.00005 | 5.228 | EIF4A1 | eukaryotic translation initiation factor 4A1 [Source:HGNC Symbol;Acc:HGNC:3282] |
| ENSG00000162062 | 0.564 | 1.104 | 3.722 | 0.00022 | 0.00466 | 0.351 | C16orf59 | chromosome 16 open reading frame 59 [Source:HGNC Symbol;Acc:HGNC:25849] |
| ENSG00000162063 | 0.521 | 1.549 | 3.980 | 0.00008 | 0.00197 | 1.243 | CCNF | cyclin F [Source:HGNC Symbol;Acc:HGNC:1591] |
| ENSG00000162073 | 0.649 | 0.632 | 4.319 | 0.00002 | 0.00062 | 2.634 | PAQR4 | progestin and adipoQ receptor family member 4 [Source:HGNC Symbol;Acc:HGNC:26386] |
| ENSG00000162174 | 0.465 | 1.758 | 4.031 | 0.00006 | 0.00168 | 1.421 | ASRGL1 | asparaginase like 1 [Source:HGNC Symbol;Acc:HGNC:16448] |
| ENSG00000162377 | 0.334 | 3.983 | 3.649 | 0.00029 | 0.00575 | -0.307 | COA7 | cytochrome c oxidase assembly factor 7 (putative) [Source:HGNC Symbol;Acc:HGNC:25716] |
| ENSG00000162607 | 0.355 | 5.430 | 4.422 | 0.00001 | 0.00043 | 2.601 | USP1 | ubiquitin specific peptidase 1 [Source:HGNC Symbol;Acc:HGNC:12607] |
| ENSG00000162927 | 0.341 | 2.508 | 4.147 | 0.00004 | 0.00112 | 1.781 | PUS10 | pseudouridylate synthase 10 [Source:HGNC Symbol;Acc:HGNC:26505] |
| ENSG00000162929 | 0.444 | 1.522 | 3.583 | 0.00037 | 0.00690 | -0.156 | KIAA1841 | KIAA1841 [Source:HGNC Symbol;Acc:HGNC:29387] |
| ENSG00000163354 | -0.611 | 0.825 | -4.018 | 0.00007 | 0.00174 | 1.491 | DCST2 | DC-STAMP domain containing 2 [Source:HGNC Symbol;Acc:HGNC:26562] |
| ENSG00000163507 | 0.509 | 4.001 | 3.988 | 0.00007 | 0.00192 | 0.928 | KIAA1524 | KIAA1524 [Source:HGNC Symbol;Acc:HGNC:29302] |
| ENSG00000163535 | 0.861 | 2.165 | 6.410 | 0.00000 | 0.00000 | 12.947 | SGO2 | shugoshin 2 [Source:HGNC Symbol;Acc:HGNC:30812] |
| ENSG00000163655 | 0.283 | 5.418 | 4.360 | 0.00002 | 0.00053 | 2.341 | GMPS | guanine monophosphate synthase [Source:HGNC Symbol;Acc:HGNC:4378] |
| ENSG00000163755 | 0.284 | 4.831 | 3.693 | 0.00024 | 0.00507 | -0.227 | HPS3 | HPS3, biogenesis of lysosomal organelles complex 2 subunit 1 [Source:HGNC Symbol;Acc:HGNC:15597] |
| ENSG00000163781 | 0.300 | 6.067 | 4.538 | 0.00001 | 0.00027 | 3.099 | TOPBP1 | topoisomerase (DNA) II binding protein 1 [Source:HGNC Symbol;Acc:HGNC:17008] |
| ENSG00000163808 | 0.877 | 2.344 | 6.845 | 0.00000 | 0.00000 | 15.579 | KIF15 | kinesin family member 15 [Source:HGNC Symbol;Acc:HGNC:17273] |
| ENSG00000163918 | 0.394 | 4.288 | 4.990 | 0.00000 | 0.00004 | 5.219 | RFC4 | replication factor C subunit 4 [Source:HGNC Symbol;Acc:HGNC:9972] |
| ENSG00000163950 | 0.283 | 4.980 | 3.928 | 0.00010 | 0.00234 | 0.633 | SLBP | stem-loop binding protein [Source:HGNC Symbol;Acc:HGNC:10904] |
| ENSG00000164010 | -0.277 | 3.115 | -3.547 | 0.00042 | 0.00763 | -0.448 | ERMAP | erythroblast membrane associated protein (Scianna blood group) [Source:HGNC Symbol;Acc:HGNC:15743] |
| ENSG00000164045 | 0.603 | 2.516 | 5.401 | 0.00000 | 0.00001 | 7.408 | CDC25A | cell division cycle 25A [Source:HGNC Symbol;Acc:HGNC:1725] |
| ENSG00000164087 | 0.594 | 2.277 | 5.488 | 0.00000 | 0.00000 | 7.875 | POC1A | POC1 centriolar protein A [Source:HGNC Symbol;Acc:HGNC:24488] |
| ENSG00000164105 | 0.334 | 3.486 | 4.015 | 0.00007 | 0.00176 | 1.108 | SAP30 | Sin3A associated protein 30 [Source:HGNC Symbol;Acc:HGNC:10532] |
| ENSG00000164109 | 0.752 | 3.496 | 6.544 | 0.00000 | 0.00000 | 13.662 | MAD2L1 | mitotic arrest deficient 2 like 1 [Source:HGNC Symbol;Acc:HGNC:6763] |
| ENSG00000164535 | -0.248 | 3.443 | -3.481 | 0.00054 | 0.00918 | -0.741 | DAGLB | diacylglycerol lipase beta [Source:HGNC Symbol;Acc:HGNC:28923] |
| ENSG00000164611 | 0.813 | 3.463 | 5.727 | 0.00000 | 0.00000 | 8.988 | PTTG1 | pituitary tumor-transforming 1 [Source:HGNC Symbol;Acc:HGNC:9690] |
| ENSG00000164754 | 0.276 | 7.069 | 3.573 | 0.00038 | 0.00709 | -0.648 | RAD21 | RAD21 cohesin complex component [Source:HGNC Symbol;Acc:HGNC:9811] |
| ENSG00000165115 | -0.351 | 2.971 | -3.929 | 0.00010 | 0.00233 | 0.955 | KIF27 | kinesin family member 27 [Source:HGNC Symbol;Acc:HGNC:18632] |

| | | | | | | | | |
|---|---|---|---|---|---|---|---|---|
| ENSG00000165244 | 0.584 | 3.593 | 5.167 | 0.00000 | 0.00002 | 6.123 | ZNF367 | zinc finger protein 367 [Source:HGNC Symbol;Acc:HGNC:18320] |
| ENSG00000165304 | 0.987 | 2.039 | 6.770 | 0.00000 | 0.00000 | 15.120 | MELK | maternal embryonic leucine zipper kinase [Source:HGNC Symbol;Acc:HGNC:16870] |
| ENSG00000165480 | 1.159 | 0.799 | 7.361 | 0.00000 | 0.00000 | 18.785 | SKA3 | spindle and kinetochore associated complex subunit 3 [Source:HGNC Symbol;Acc:HGNC:20262] |
| ENSG00000165490 | 0.528 | 2.858 | 3.701 | 0.00023 | 0.00493 | 0.022 | DDIAS | DNA damage induced apoptosis suppressor [Source:HGNC Symbol;Acc:HGNC:26351] |
| ENSG00000165501 | 0.402 | 2.723 | 4.798 | 0.00000 | 0.00010 | 4.505 | LRR1 | leucine rich repeat protein 1 [Source:HGNC Symbol;Acc:HGNC:19742] |
| ENSG00000165916 | 0.263 | 6.574 | 3.876 | 0.00012 | 0.00279 | 0.424 | PSMC3 | proteasome 26S subunit, ATPase 3 [Source:HGNC Symbol;Acc:HGNC:9549] |
| ENSG00000166037 | 0.258 | 6.071 | 3.484 | 0.00053 | 0.00909 | -0.988 | CEP57 | centrosomal protein 57 [Source:HGNC Symbol;Acc:HGNC:30794] |
| ENSG00000166181 | 0.274 | 6.653 | 4.636 | 0.00000 | 0.00019 | 3.542 | API5 | apoptosis inhibitor 5 [Source:HGNC Symbol;Acc:HGNC:594] |
| ENSG00000166451 | 0.370 | 2.312 | 4.107 | 0.00005 | 0.00128 | 1.644 | CENPN | centromere protein N [Source:HGNC Symbol;Acc:HGNC:30873] |
| ENSG00000166483 | 0.755 | 3.212 | 3.545 | 0.00042 | 0.00765 | -0.615 | WEE1 | WEE1 G2 checkpoint kinase [Source:HGNC Symbol;Acc:HGNC:12761] |
| ENSG00000166579 | -0.390 | 5.714 | -5.113 | 0.00000 | 0.00003 | 5.759 | NDEL1 | nudE neurodevelopment protein 1 like 1 [Source:HGNC Symbol;Acc:HGNC:17620] |
| ENSG00000166788 | 0.283 | 4.198 | 3.803 | 0.00016 | 0.00357 | 0.225 | SAAL1 | serum amyloid A like 1 [Source:HGNC Symbol;Acc:HGNC:25158] |
| ENSG00000166803 | 0.684 | 0.987 | 4.400 | 0.00001 | 0.00046 | 2.943 | PCLAF | PCNA clamp associated factor [Source:HGNC Symbol;Acc:HGNC:28961] |
| ENSG00000166845 | 0.973 | 0.826 | 6.447 | 0.00000 | 0.00000 | 13.120 | C18orf54 | chromosome 18 open reading frame 54 [Source:HGNC Symbol;Acc:HGNC:13796] |
| ENSG00000166851 | 0.802 | 1.931 | 6.399 | 0.00000 | 0.00000 | 12.890 | PLK1 | polo like kinase 1 [Source:HGNC Symbol;Acc:HGNC:9077] |
| ENSG00000166881 | 0.312 | 5.340 | 3.709 | 0.00023 | 0.00485 | -0.196 | NEMP1 | nuclear envelope integral membrane protein 1 [Source:HGNC Symbol;Acc:HGNC:29001] |
| ENSG00000166896 | 0.346 | 2.716 | 4.003 | 0.00007 | 0.00183 | 1.187 | ATP23 | ATP23 metallopeptidase and ATP synthase assembly factor homolog [Source:HGNC Symbol;Acc:HGNC:29452] |
| ENSG00000167081 | 0.491 | 2.361 | 4.073 | 0.00005 | 0.00145 | 1.492 | PBX3 | PBX homeobox 3 [Source:HGNC Symbol;Acc:HGNC:8634] |
| ENSG00000167088 | 0.301 | 5.291 | 4.094 | 0.00005 | 0.00135 | 1.260 | SNRPD1 | small nuclear ribonucleoprotein D1 polypeptide [Source:HGNC Symbol;Acc:HGNC:11158] |
| ENSG00000167193 | -0.287 | 5.642 | -4.054 | 0.00006 | 0.00154 | 1.110 | CRK | CRK proto-oncogene, adaptor protein [Source:HGNC Symbol;Acc:HGNC:2362] |
| ENSG00000167325 | 0.587 | 4.815 | 7.009 | 0.00000 | 0.00000 | 16.494 | RRM1 | ribonucleotide reductase catalytic subunit M1 [Source:HGNC Symbol;Acc:HGNC:10451] |
| ENSG00000167333 | 0.366 | 3.177 | 3.724 | 0.00021 | 0.00464 | 0.066 | TRIM68 | tripartite motif containing 68 [Source:HGNC Symbol;Acc:HGNC:21161] |
| ENSG00000167377 | -0.346 | 0.557 | -3.541 | 0.00043 | 0.00773 | -0.156 | ZNF23 | zinc finger protein 23 [Source:HGNC Symbol;Acc:HGNC:13023] |
| ENSG00000167513 | 0.755 | 2.137 | 5.287 | 0.00000 | 0.00001 | 6.867 | CDT1 | chromatin licensing and DNA replication factor 1 [Source:HGNC Symbol;Acc:HGNC:24576] |
| ENSG00000167525 | -0.413 | 2.639 | -3.534 | 0.00044 | 0.00788 | -0.399 | PROCA1 | protein interacting with cyclin A1 [Source:HGNC Symbol;Acc:HGNC:28600] |
| ENSG00000167670 | 0.467 | 4.109 | 4.882 | 0.00000 | 0.00007 | 4.721 | CHAF1A | chromatin assembly factor 1 subunit A [Source:HGNC Symbol;Acc:HGNC:1910] |
| ENSG00000167674 | 0.267 | 4.999 | 4.160 | 0.00004 | 0.00108 | 1.538 | AC011498.1 | Hepatoma-derived growth factor-related protein 2 [Source:UniProtKB/Swiss-Prot;Acc:Q7Z4V5] |
| ENSG00000167699 | -0.298 | 4.925 | -4.710 | 0.00000 | 0.00014 | 3.917 | GLOD4 | glyoxalase domain containing 4 [Source:HGNC Symbol;Acc:HGNC:14111] |
| ENSG00000167702 | -0.416 | 5.496 | -4.021 | 0.00007 | 0.00173 | 0.991 | KIFC2 | kinesin family member C2 [Source:HGNC Symbol;Acc:HGNC:29530] |
| ENSG00000167705 | -0.714 | 2.015 | -4.444 | 0.00001 | 0.00040 | 3.132 | RILP | Rab interacting lysosomal protein [Source:HGNC Symbol;Acc:HGNC:30266] |
| ENSG00000167716 | -0.343 | 5.302 | -4.344 | 0.00002 | 0.00056 | 2.304 | WDR81 | WD repeat domain 81 [Source:HGNC Symbol;Acc:HGNC:26600] |
| ENSG00000167740 | -0.422 | 4.603 | -4.786 | 0.00000 | 0.00010 | 4.298 | CYB5D2 | cytochrome b5 domain containing 2 [Source:HGNC Symbol;Acc:HGNC:28471] |
| ENSG00000167775 | 0.379 | 5.578 | 3.519 | 0.00047 | 0.00819 | -0.867 | CD320 | CD320 molecule [Source:HGNC Symbol;Acc:HGNC:16692] |
| ENSG00000167842 | -0.375 | 5.892 | -4.612 | 0.00000 | 0.00021 | 3.429 | MIS12 | MIS12, kinetochore complex component [Source:HGNC Symbol;Acc:HGNC:24967] |
| ENSG00000167861 | -0.474 | 7.883 | -3.634 | 0.00030 | 0.00598 | -0.422 | HID1 | HID1 domain containing [Source:HGNC Symbol;Acc:HGNC:15736] |
| ENSG00000167900 | 0.826 | 2.880 | 5.965 | 0.00000 | 0.00000 | 10.350 | TK1 | thymidine kinase 1 [Source:HGNC Symbol;Acc:HGNC:11830] |
| ENSG00000168002 | 0.223 | 5.109 | 3.768 | 0.00018 | 0.00398 | 0.031 | POLR2G | RNA polymerase II subunit G [Source:HGNC Symbol;Acc:HGNC:9194] |
| ENSG00000168061 | 0.444 | 2.773 | 3.851 | 0.00013 | 0.00303 | 0.591 | SAC3D1 | SAC3 domain containing 1 [Source:HGNC Symbol;Acc:HGNC:30179] |
| ENSG00000168078 | 1.120 | 1.301 | 7.013 | 0.00000 | 0.00000 | 16.617 | PBK | PDZ binding kinase [Source:HGNC Symbol;Acc:HGNC:18282] |
| ENSG00000168395 | -0.228 | 5.951 | -3.760 | 0.00019 | 0.00408 | -0.014 | ING5 | inhibitor of growth family member 5 [Source:HGNC Symbol;Acc:HGNC:19421] |
| ENSG00000168496 | 0.620 | 4.739 | 5.799 | 0.00000 | 0.00000 | 9.303 | FEN1 | flap structure-specific endonuclease 1 [Source:HGNC Symbol;Acc:HGNC:3650] |
| ENSG00000169188 | 0.253 | 3.921 | 3.664 | 0.00027 | 0.00548 | -0.238 | APEX2 | apurinic/apyrimidinic endodeoxyribonuclease 2 [Source:HGNC Symbol;Acc:HGNC:17889] |
| ENSG00000169288 | 0.287 | 3.846 | 3.473 | 0.00055 | 0.00943 | -0.896 | MRPL1 | mitochondrial ribosomal protein L1 [Source:HGNC Symbol;Acc:HGNC:14275] |
| ENSG00000169607 | 1.133 | 1.600 | 7.530 | 0.00000 | 0.00000 | 20.011 | CKAP2L | cytoskeleton associated protein 2 like [Source:HGNC Symbol;Acc:HGNC:26877] |
| ENSG00000169621 | 0.305 | 2.357 | 3.491 | 0.00052 | 0.00890 | -0.583 | APLF | aprataxin and PNKP like factor [Source:HGNC Symbol;Acc:HGNC:28724] |
| ENSG00000169679 | 0.891 | 4.056 | 6.533 | 0.00000 | 0.00000 | 13.550 | BUB1 | BUB1 mitotic checkpoint serine/threonine kinase [Source:HGNC Symbol;Acc:HGNC:1148] |
| ENSG00000169718 | -0.238 | 7.545 | -3.639 | 0.00030 | 0.00590 | -0.412 | DUS1L | dihydrouridine synthase 1 like [Source:HGNC Symbol;Acc:HGNC:30086] |
| ENSG00000169814 | -0.383 | 5.588 | -4.343 | 0.00002 | 0.00056 | 2.283 | BTD | biotinidase [Source:HGNC Symbol;Acc:HGNC:1122] |
| ENSG00000169891 | 0.550 | 3.622 | 3.791 | 0.00017 | 0.00371 | 0.232 | REPS2 | RALBP1 associated Eps domain containing 2 [Source:HGNC Symbol;Acc:HGNC:9963] |
| ENSG00000170037 | -0.379 | 4.796 | -4.729 | 0.00000 | 0.00013 | 4.017 | CNTROB | centrobin, centriole duplication and spindle assembly protein [Source:HGNC Symbol;Acc:HGNC:29616] |
| ENSG00000170043 | -0.339 | 4.798 | -4.317 | 0.00002 | 0.00062 | 2.231 | TRAPPC1 | trafficking protein particle complex 1 [Source:HGNC Symbol;Acc:HGNC:19894] |
| ENSG00000170222 | -0.346 | 2.438 | -4.114 | 0.00004 | 0.00126 | 1.737 | ADPRM | ADP-ribose/CDP-alcohol diphosphatase, manganese dependent [Source:HGNC Symbol;Acc:HGNC:30925] |
| ENSG00000170291 | -0.365 | 5.031 | -6.194 | 0.00000 | 0.00000 | 11.548 | ELP5 | elongator acetyltransferase complex subunit 5 [Source:HGNC Symbol;Acc:HGNC:30617] |
| ENSG00000170296 | -0.671 | 0.323 | -4.474 | 0.00001 | 0.00035 | 3.213 | GABARAP | GABA type A receptor-associated protein [Source:HGNC Symbol;Acc:HGNC:4067] |
| ENSG00000170310 | -0.260 | 4.055 | -3.608 | 0.00033 | 0.00641 | -0.396 | STX8 | syntaxin 8 [Source:HGNC Symbol;Acc:HGNC:11443] |
| ENSG00000170312 | 1.082 | 2.365 | 6.571 | 0.00000 | 0.00000 | 13.883 | CDK1 | cyclin dependent kinase 1 [Source:HGNC Symbol;Acc:HGNC:1722] |

| | | | | | | | | |
|---|---|---|---|---|---|---|---|---|
| ENSG00000170540 | 0.412 | 5.369 | 4.581 | 0.00001 | 0.00023 | 3.288 | ARL6IP1 | ADP ribosylation factor like GTPase 6 interacting protein 1 [Source:HGNC Symbol;Acc:HGNC:697] |
| ENSG00000170779 | 0.489 | 2.461 | 5.312 | 0.00000 | 0.00001 | 6.979 | CDCA4 | cell division cycle associated 4 [Source:HGNC Symbol;Acc:HGNC:14625] |
| ENSG00000171044 | -0.554 | 3.089 | -3.654 | 0.00028 | 0.00566 | -0.054 | XKR6 | XK related 6 [Source:HGNC Symbol;Acc:HGNC:27806] |
| ENSG00000171241 | 0.702 | 3.071 | 4.542 | 0.00001 | 0.00027 | 3.287 | SHCBP1 | SHC binding and spindle associated 1 [Source:HGNC Symbol;Acc:HGNC:29547] |
| ENSG00000171262 | 0.380 | 4.525 | 3.551 | 0.00041 | 0.00757 | -0.710 | FAM98B | family with sequence similarity 98 member B [Source:HGNC Symbol;Acc:HGNC:26773] |
| ENSG00000171310 | -0.390 | 6.502 | -3.590 | 0.00036 | 0.00674 | -0.623 | CHST11 | carbohydrate sulfotransferase 11 [Source:HGNC Symbol;Acc:HGNC:17422] |
| ENSG00000171320 | 1.218 | 0.561 | 7.657 | 0.00000 | 0.00000 | 20.700 | ESCO2 | establishment of sister chromatid cohesion N-acetyltransferase 2 [Source:HGNC Symbol;Acc:HGNC:27230] |
| ENSG00000171497 | 0.315 | 3.950 | 5.003 | 0.00000 | 0.00004 | 5.317 | PPID | peptidylprolyl isomerase D [Source:HGNC Symbol;Acc:HGNC:9257] |
| ENSG00000171848 | 1.051 | 4.533 | 5.018 | 0.00000 | 0.00004 | 5.306 | RRM2 | ribonucleotide reductase regulatory subunit M2 [Source:HGNC Symbol;Acc:HGNC:10452] |
| ENSG00000171861 | -0.311 | 2.899 | -4.779 | 0.00000 | 0.00011 | 4.467 | MRM3 | mitochondrial rRNA methyltransferase 3 [Source:HGNC Symbol;Acc:HGNC:18485] |
| ENSG00000171953 | -0.252 | 3.364 | -4.460 | 0.00001 | 0.00037 | 3.004 | ATPAF2 | ATP synthase mitochondrial F1 complex assembly factor 2 [Source:HGNC Symbol;Acc:HGNC:18802] |
| ENSG00000171962 | -0.359 | 1.760 | -4.029 | 0.00006 | 0.00168 | 1.484 | DRC3 | dynein regulatory complex subunit 3 [Source:HGNC Symbol;Acc:HGNC:25384] |
| ENSG00000172167 | 0.403 | 2.384 | 4.600 | 0.00001 | 0.00022 | 3.671 | MTBP | MDM2 binding protein [Source:HGNC Symbol;Acc:HGNC:7417] |
| ENSG00000172172 | 0.333 | 3.796 | 4.189 | 0.00003 | 0.00098 | 1.754 | MRPL13 | mitochondrial ribosomal protein L13 [Source:HGNC Symbol;Acc:HGNC:14278] |
| ENSG00000172244 | 0.495 | 0.610 | 4.501 | 0.00001 | 0.00032 | 3.381 | C5orf34 | chromosome 5 open reading frame 34 [Source:HGNC Symbol;Acc:HGNC:24738] |
| ENSG00000172638 | 0.449 | 2.031 | 3.510 | 0.00048 | 0.00843 | -0.487 | EFEMP2 | EGF containing fibulin like extracellular matrix protein 2 [Source:HGNC Symbol;Acc:HGNC:3219] |
| ENSG00000172893 | 0.366 | 5.237 | 3.807 | 0.00016 | 0.00353 | 0.164 | DHCR7 | 7-dehydrocholesterol reductase [Source:HGNC Symbol;Acc:HGNC:2860] |
| ENSG00000173163 | 0.214 | 4.255 | 3.606 | 0.00034 | 0.00645 | -0.479 | COMMD1 | copper metabolism domain containing 1 [Source:HGNC Symbol;Acc:HGNC:23024] |
| ENSG00000173207 | 0.483 | 3.600 | 4.108 | 0.00005 | 0.00128 | 1.444 | CKS1B | CDC28 protein kinase regulatory subunit 1B [Source:HGNC Symbol;Acc:HGNC:19083] |
| ENSG00000173456 | 0.320 | 4.345 | 4.941 | 0.00000 | 0.00005 | 4.989 | RNF26 | ring finger protein 26 [Source:HGNC Symbol;Acc:HGNC:14646] |
| ENSG00000173692 | 0.206 | 6.202 | 3.545 | 0.00042 | 0.00765 | -0.778 | PSMD1 | proteasome 26S subunit, non-ATPase 1 [Source:HGNC Symbol;Acc:HGNC:9554] |
| ENSG00000174231 | -0.346 | 8.436 | -6.417 | 0.00000 | 0.00000 | 12.875 | PRPF8 | pre-mRNA processing factor 8 [Source:HGNC Symbol;Acc:HGNC:17340] |
| ENSG00000174238 | -0.438 | 6.164 | -7.916 | 0.00000 | 0.00000 | 22.634 | PITPNA | phosphatidylinositol transfer protein alpha [Source:HGNC Symbol;Acc:HGNC:9001] |
| ENSG00000174282 | -0.643 | 6.768 | -8.327 | 0.00000 | 0.00000 | 25.612 | ZBTB4 | zinc finger and BTB domain containing 4 [Source:HGNC Symbol;Acc:HGNC:23847] |
| ENSG00000174292 | -0.574 | 2.256 | -3.991 | 0.00007 | 0.00190 | 1.311 | TNK1 | tyrosine kinase non receptor 1 [Source:HGNC Symbol;Acc:HGNC:11940] |
| ENSG00000174307 | -1.390 | 2.133 | -4.985 | 0.00000 | 0.00004 | 5.483 | PHLDA3 | pleckstrin homology like domain family A member 3 [Source:HGNC Symbol;Acc:HGNC:8934] |
| ENSG00000174371 | 0.933 | 1.731 | 6.109 | 0.00000 | 0.00000 | 11.230 | EXO1 | exonuclease 1 [Source:HGNC Symbol;Acc:HGNC:3511] |
| ENSG00000174442 | 0.342 | 3.649 | 4.017 | 0.00007 | 0.00174 | 1.095 | ZWILCH | zwilch kinetochore protein [Source:HGNC Symbol;Acc:HGNC:25468] |
| ENSG00000174547 | 0.293 | 4.920 | 4.079 | 0.00005 | 0.00142 | 1.221 | MRPL11 | mitochondrial ribosomal protein L11 [Source:HGNC Symbol;Acc:HGNC:14042] |
| ENSG00000174842 | 0.307 | 2.684 | 3.605 | 0.00034 | 0.00645 | -0.255 | GLMN | glomulin, FKBP associated protein [Source:HGNC Symbol;Acc:HGNC:14373] |
| ENSG00000175063 | 1.051 | 1.460 | 6.869 | 0.00000 | 0.00000 | 15.722 | UBE2C | ubiquitin conjugating enzyme E2 C [Source:HGNC Symbol;Acc:HGNC:15937] |
| ENSG00000175216 | 0.418 | 5.923 | 5.042 | 0.00000 | 0.00003 | 5.405 | CKAP5 | cytoskeleton associated protein 5 [Source:HGNC Symbol;Acc:HGNC:28959] |
| ENSG00000175305 | 0.413 | 2.840 | 3.616 | 0.00032 | 0.00627 | -0.251 | CCNE2 | cyclin E2 [Source:HGNC Symbol;Acc:HGNC:1590] |
| ENSG00000175643 | 0.619 | 0.827 | 3.618 | 0.00032 | 0.00622 | 0.022 | RMI2 | RecQ mediated genome instability 2 [Source:HGNC Symbol;Acc:HGNC:28349] |
| ENSG00000175662 | -0.283 | 5.453 | -4.181 | 0.00003 | 0.00100 | 1.621 | TOM1L2 | target of myb1 like 2 membrane trafficking protein [Source:HGNC Symbol;Acc:HGNC:11984] |
| ENSG00000175826 | -0.379 | 5.438 | -7.170 | 0.00000 | 0.00000 | 17.548 | CTDNEP1 | CTD nuclear envelope phosphatase 1 [Source:HGNC Symbol;Acc:HGNC:19085] |
| ENSG00000176155 | -0.313 | 5.684 | -3.947 | 0.00009 | 0.00220 | 0.693 | CCDC57 | coiled-coil domain containing 57 [Source:HGNC Symbol;Acc:HGNC:27564] |
| ENSG00000176208 | 0.379 | 2.798 | 3.618 | 0.00032 | 0.00622 | -0.238 | ATAD5 | ATPase family, AAA domain containing 5 [Source:HGNC Symbol;Acc:HGNC:25752] |
| ENSG00000176619 | 0.386 | 5.839 | 4.580 | 0.00001 | 0.00023 | 3.282 | LMNB2 | lamin B2 [Source:HGNC Symbol;Acc:HGNC:6638] |
| ENSG00000176890 | 0.563 | 2.269 | 4.768 | 0.00000 | 0.00011 | 4.413 | TYMS | thymidylate synthetase [Source:HGNC Symbol;Acc:HGNC:12441] |
| ENSG00000177150 | 0.275 | 3.773 | 3.918 | 0.00010 | 0.00242 | 0.709 | FAM210A | family with sequence similarity 210 member A [Source:HGNC Symbol;Acc:HGNC:28346] |
| ENSG00000177370 | -0.264 | 3.872 | -4.886 | 0.00000 | 0.00007 | 4.827 | TIMM22 | translocase of inner mitochondrial membrane 22 [Source:HGNC Symbol;Acc:HGNC:17317] |
| ENSG00000177427 | -0.366 | 3.363 | -4.584 | 0.00001 | 0.00023 | 3.553 | MIEF2 | mitochondrial elongation factor 2 [Source:HGNC Symbol;Acc:HGNC:17920] |
| ENSG00000177731 | -0.371 | 7.230 | -4.503 | 0.00001 | 0.00032 | 2.960 | FLII | FLII, actin remodeling protein [Source:HGNC Symbol;Acc:HGNC:3750] |
| ENSG00000178105 | 0.332 | 4.963 | 3.730 | 0.00021 | 0.00455 | -0.103 | DDX10 | DEAD-box helicase 10 [Source:HGNC Symbol;Acc:HGNC:2735] |
| ENSG00000178149 | -0.306 | 4.352 | -4.127 | 0.00004 | 0.00120 | 1.503 | DALRD3 | DALR anticodon binding domain containing 3 [Source:HGNC Symbol;Acc:HGNC:25536] |
| ENSG00000178921 | -0.366 | 5.193 | -3.941 | 0.00009 | 0.00224 | 0.702 | PFAS | phosphoribosylformylglycinamidine synthase [Source:HGNC Symbol;Acc:HGNC:8863] |
| ENSG00000178971 | -0.561 | 6.481 | -6.656 | 0.00000 | 0.00000 | 14.263 | CTC1 | CST telomere replication complex component 1 [Source:HGNC Symbol;Acc:HGNC:26169] |
| ENSG00000178999 | 0.555 | 1.684 | 3.507 | 0.00049 | 0.00847 | -0.458 | AURKB | aurora kinase B [Source:HGNC Symbol;Acc:HGNC:11390] |
| ENSG00000179029 | -0.839 | 4.375 | -4.701 | 0.00000 | 0.00014 | 3.983 | TMEM107 | transmembrane protein 107 [Source:HGNC Symbol;Acc:HGNC:28128] |
| ENSG00000179750 | 0.952 | 1.847 | 4.732 | 0.00000 | 0.00013 | 4.276 | APOBEC3B | apolipoprotein B mRNA editing enzyme catalytic subunit 3B [Source:HGNC Symbol;Acc:HGNC:17352] |
| ENSG00000180198 | 0.398 | 4.433 | 4.214 | 0.00003 | 0.00090 | 1.789 | RCC1 | regulator of chromosome condensation 1 [Source:HGNC Symbol;Acc:HGNC:1913] |
| ENSG00000180425 | 0.379 | 3.933 | 3.453 | 0.00059 | 0.00995 | -0.985 | C11orf71 | chromosome 11 open reading frame 71 [Source:HGNC Symbol;Acc:HGNC:25937] |
| ENSG00000181222 | -0.358 | 7.237 | -4.560 | 0.00001 | 0.00025 | 3.208 | POLR2A | RNA polymerase II subunit A [Source:HGNC Symbol;Acc:HGNC:9187] |
| ENSG00000181284 | -0.579 | 1.229 | -4.419 | 0.00001 | 0.00043 | 3.043 | TMEM102 | transmembrane protein 102 [Source:HGNC Symbol;Acc:HGNC:26722] |
| ENSG00000181544 | 0.592 | 1.023 | 5.349 | 0.00000 | 0.00001 | 7.226 | FANCB | Fanconi anemia complementation group B [Source:HGNC Symbol;Acc:HGNC:3583] |

| | | | | | | | | |
|---|---|---|---|---|---|---|---|---|
| ENSG00000181938 | 0.550 | 0.757 | 3.761 | 0.00019 | 0.00407 | 0.529 | GINS3 | GINS complex subunit 3 [Source:HGNC Symbol;Acc:HGNC:25851] |
| ENSG00000183087 | -0.761 | 7.895 | -4.317 | 0.00002 | 0.00062 | 2.190 | GAS6 | growth arrest specific 6 [Source:HGNC Symbol;Acc:HGNC:4168] |
| ENSG00000183401 | -0.271 | 5.104 | -3.459 | 0.00058 | 0.00978 | -1.027 | CCDC159 | coiled-coil domain containing 159 [Source:HGNC Symbol;Acc:HGNC:26996] |
| ENSG00000183431 | 0.233 | 5.869 | 4.148 | 0.00004 | 0.00112 | 1.467 | SF3A3 | splicing factor 3a subunit 3 [Source:HGNC Symbol;Acc:HGNC:10767] |
| ENSG00000183520 | 0.233 | 4.642 | 4.134 | 0.00004 | 0.00118 | 1.458 | UTP11 | UTP11, small subunit processome component homolog (S. cerevisiae) [Source:HGNC Symbol;Acc:HGNC:24329] |
| ENSG00000183624 | -0.322 | 7.280 | -3.714 | 0.00022 | 0.00477 | -0.163 | HMCES | 5-hydroxymethylcytosine binding, ES cell specific [Source:HGNC Symbol;Acc:HGNC:24446] |
| ENSG00000183682 | -0.717 | 5.071 | -4.704 | 0.00000 | 0.00014 | 3.904 | BMP8A | bone morphogenetic protein 8a [Source:HGNC Symbol;Acc:HGNC:21650] |
| ENSG00000183723 | -0.673 | 3.656 | -4.732 | 0.00000 | 0.00013 | 4.197 | CMTM4 | CKLF like MARVEL transmembrane domain containing 4 [Source:HGNC Symbol;Acc:HGNC:19175] |
| ENSG00000183763 | 0.573 | 1.698 | 5.379 | 0.00000 | 0.00001 | 7.364 | TRAIP | TRAF interacting protein [Source:HGNC Symbol;Acc:HGNC:30764] |
| ENSG00000183856 | 0.716 | 1.678 | 3.831 | 0.00014 | 0.00326 | 0.654 | IQGAP3 | IQ motif containing GTPase activating protein 3 [Source:HGNC Symbol;Acc:HGNC:20669] |
| ENSG00000184220 | 0.327 | 5.002 | 3.817 | 0.00015 | 0.00342 | 0.212 | CMSS1 | cms1 ribosomal small subunit homolog (yeast) [Source:HGNC Symbol;Acc:HGNC:28666] |
| ENSG00000184271 | -0.438 | 4.897 | -4.392 | 0.00001 | 0.00047 | 2.542 | POU6F1 | POU class 6 homeobox 1 [Source:HGNC Symbol;Acc:HGNC:9224] |
| ENSG00000184445 | -0.445 | 7.516 | -3.569 | 0.00039 | 0.00713 | -0.669 | KNTC1 | kinetochore associated 1 [Source:HGNC Symbol;Acc:HGNC:17255] |
| ENSG00000184465 | -0.467 | 4.992 | -4.072 | 0.00005 | 0.00145 | 1.232 | WDR27 | WD repeat domain 27 [Source:HGNC Symbol;Acc:HGNC:21248] |
| ENSG00000184661 | 1.017 | 0.896 | 6.208 | 0.00000 | 0.00000 | 11.771 | CDCA2 | cell division cycle associated 2 [Source:HGNC Symbol;Acc:HGNC:14623] |
| ENSG00000185245 | -0.470 | 0.449 | -3.569 | 0.00039 | 0.00713 | -0.058 | GP1BA | glycoprotein Ib platelet alpha subunit [Source:HGNC Symbol;Acc:HGNC:4439] |
| ENSG00000185480 | 0.791 | 1.559 | 7.132 | 0.00000 | 0.00000 | 17.370 | PARPBP | PARP1 binding protein [Source:HGNC Symbol;Acc:HGNC:26074] |
| ENSG00000185504 | -0.258 | 5.881 | -3.674 | 0.00026 | 0.00536 | -0.323 | FAAP100 | Fanconi anemia core complex associated protein 100 [Source:HGNC Symbol;Acc:HGNC:26171] |
| ENSG00000185515 | 0.357 | 4.409 | 4.079 | 0.00005 | 0.00142 | 1.251 | BRCC3 | BRCA1/BRCA2-containing complex subunit 3 [Source:HGNC Symbol;Acc:HGNC:24185] |
| ENSG00000185627 | 0.241 | 6.926 | 4.268 | 0.00002 | 0.00074 | 1.978 | PSMD13 | proteasome 26S subunit, non-ATPase 13 [Source:HGNC Symbol;Acc:HGNC:9558] |
| ENSG00000185721 | 0.292 | 5.057 | 4.333 | 0.00002 | 0.00058 | 2.242 | DRG1 | developmentally regulated GTP binding protein 1 [Source:HGNC Symbol;Acc:HGNC:3029] |
| ENSG00000185722 | -0.438 | 5.937 | -6.413 | 0.00000 | 0.00000 | 12.793 | ANKFY1 | ankyrin repeat and FYVE domain containing 1 [Source:HGNC Symbol;Acc:HGNC:20763] |
| ENSG00000185869 | 1.043 | -0.157 | 4.173 | 0.00003 | 0.00103 | 2.072 | ZNF829 | zinc finger protein 829 [Source:HGNC Symbol;Acc:HGNC:34032] |
| ENSG00000185989 | -0.636 | 5.996 | -3.627 | 0.00031 | 0.00608 | -0.482 | RASA3 | RAS p21 protein activator 3 [Source:HGNC Symbol;Acc:HGNC:20331] |
| ENSG00000186185 | 0.938 | 1.605 | 5.324 | 0.00000 | 0.00001 | 7.079 | KIF18B | kinesin family member 18B [Source:HGNC Symbol;Acc:HGNC:27102] |
| ENSG00000186193 | 0.622 | 2.278 | 3.880 | 0.00012 | 0.00276 | 0.749 | SAPCD2 | suppressor APC domain containing 2 [Source:HGNC Symbol;Acc:HGNC:28055] |
| ENSG00000186638 | 0.620 | 1.580 | 4.597 | 0.00001 | 0.00022 | 3.732 | KIF24 | kinesin family member 24 [Source:HGNC Symbol;Acc:HGNC:19916] |
| ENSG00000186908 | -0.260 | 5.699 | -3.538 | 0.00043 | 0.00778 | -0.795 | ZDHHC17 | zinc finger DHHC-type containing 17 [Source:HGNC Symbol;Acc:HGNC:18412] |
| ENSG00000187531 | -0.216 | 5.123 | -3.625 | 0.00031 | 0.00612 | -0.461 | SIRT7 | sirtuin 7 [Source:HGNC Symbol;Acc:HGNC:14935] |
| ENSG00000187735 | 0.343 | 6.825 | 4.443 | 0.00001 | 0.00040 | 2.713 | TCEA1 | transcription elongation factor A1 [Source:HGNC Symbol;Acc:HGNC:11612] |
| ENSG00000187741 | 0.488 | 3.556 | 4.030 | 0.00006 | 0.00168 | 1.145 | FANCA | Fanconi anemia complementation group A [Source:HGNC Symbol;Acc:HGNC:3582] |
| ENSG00000187951 | 0.594 | 2.238 | 4.784 | 0.00000 | 0.00010 | 4.483 | ARHGAP11B | Rho GTPase activating protein 11B [Source:HGNC Symbol;Acc:HGNC:15782] |
| ENSG00000188486 | 0.372 | 4.761 | 3.624 | 0.00031 | 0.00613 | -0.472 | H2AFX | H2A histone family member X [Source:HGNC Symbol;Acc:HGNC:4739] |
| ENSG00000188732 | -0.497 | 4.132 | -4.289 | 0.00002 | 0.00069 | 2.206 | FAM221A | family with sequence similarity 221 member A [Source:HGNC Symbol;Acc:HGNC:27977] |
| ENSG00000188779 | -0.364 | 0.693 | -3.464 | 0.00057 | 0.00964 | -0.410 | SKOR1 | SKI family transcriptional corepressor 1 [Source:HGNC Symbol;Acc:HGNC:21326] |
| ENSG00000189057 | 1.205 | 1.903 | 7.227 | 0.00000 | 0.00000 | 18.014 | FAM111B | family with sequence similarity 111 member B [Source:HGNC Symbol;Acc:HGNC:24200] |
| ENSG00000189266 | 0.312 | 5.756 | 4.002 | 0.00007 | 0.00183 | 0.892 | PNRC2 | proline rich nuclear receptor coactivator 2 [Source:HGNC Symbol;Acc:HGNC:23158] |
| ENSG00000196187 | -0.409 | 6.471 | -3.682 | 0.00025 | 0.00524 | -0.300 | TMEM63A | transmembrane protein 63A [Source:HGNC Symbol;Acc:HGNC:29118] |
| ENSG00000196230 | 0.520 | 7.399 | 4.503 | 0.00001 | 0.00032 | 3.008 | TUBB | tubulin beta class I [Source:HGNC Symbol;Acc:HGNC:20778] |
| ENSG00000196247 | 0.484 | 3.820 | 4.147 | 0.00004 | 0.00112 | 1.572 | ZNF107 | zinc finger protein 107 [Source:HGNC Symbol;Acc:HGNC:12887] |
| ENSG00000196584 | 0.655 | 2.255 | 4.597 | 0.00001 | 0.00022 | 3.646 | XRCC2 | X-ray repair cross complementing 2 [Source:HGNC Symbol;Acc:HGNC:12829] |
| ENSG00000196586 | -0.824 | 6.318 | -5.401 | 0.00000 | 0.00001 | 7.190 | MYO6 | myosin VI [Source:HGNC Symbol;Acc:HGNC:7605] |
| ENSG00000196655 | 0.279 | 4.409 | 4.219 | 0.00003 | 0.00089 | 1.817 | TRAPPC4 | trafficking protein particle complex 4 [Source:HGNC Symbol;Acc:HGNC:19943] |
| ENSG00000196664 | 1.384 | 0.586 | 4.312 | 0.00002 | 0.00062 | 2.581 | TLR7 | toll like receptor 7 [Source:HGNC Symbol;Acc:HGNC:15631] |
| ENSG00000196689 | -0.400 | 2.704 | -3.973 | 0.00008 | 0.00201 | 1.168 | TRPV1 | transient receptor potential cation channel subfamily V member 1 [Source:HGNC Symbol;Acc:HGNC:12716] |
| ENSG00000197299 | 0.498 | 3.455 | 3.547 | 0.00042 | 0.00763 | -0.614 | BLM | Bloom syndrome RecQ like helicase [Source:HGNC Symbol;Acc:HGNC:1058] |
| ENSG00000197417 | -0.465 | 1.519 | -6.123 | 0.00000 | 0.00000 | 11.234 | SHPK | sedoheptulokinase [Source:HGNC Symbol;Acc:HGNC:1492] |
| ENSG00000197771 | 0.259 | 5.548 | 3.974 | 0.00008 | 0.00201 | 0.788 | MCMBP | minichromosome maintenance complex binding protein [Source:HGNC Symbol;Acc:HGNC:25782] |
| ENSG00000197774 | -0.338 | 5.471 | -4.135 | 0.00004 | 0.00117 | 1.438 | EME2 | essential meiotic structure-specific endonuclease subunit 2 [Source:HGNC Symbol;Acc:HGNC:27289] |
| ENSG00000197879 | -0.582 | 6.732 | -5.879 | 0.00000 | 0.00000 | 9.728 | MYO1C | myosin IC [Source:HGNC Symbol;Acc:HGNC:7597] |
| ENSG00000197912 | -0.388 | 6.650 | -4.355 | 0.00002 | 0.00054 | 2.314 | SPG7 | SPG7, paraplegin matrix AAA peptidase subunit [Source:HGNC Symbol;Acc:HGNC:11237] |
| ENSG00000197943 | -0.402 | 8.022 | -3.575 | 0.00038 | 0.00707 | -0.617 | PLCG2 | phospholipase C gamma 2 [Source:HGNC Symbol;Acc:HGNC:9066] |
| ENSG00000198055 | -0.382 | 7.713 | -3.677 | 0.00026 | 0.00532 | -0.276 | GRK6 | G protein-coupled receptor kinase 6 [Source:HGNC Symbol;Acc:HGNC:4545] |
| ENSG00000198331 | 0.412 | 2.601 | 4.012 | 0.00007 | 0.00177 | 1.226 | HYLS1 | HYLS1, centriolar and ciliogenesis associated [Source:HGNC Symbol;Acc:HGNC:26558] |
| ENSG00000198453 | 0.858 | 0.796 | 3.471 | 0.00056 | 0.00945 | -0.462 | ZNF568 | zinc finger protein 568 [Source:HGNC Symbol;Acc:HGNC:25392] |
| ENSG00000198554 | 0.736 | 1.685 | 5.258 | 0.00000 | 0.00001 | 6.762 | WDHD1 | WD repeat and HMG-box DNA binding protein 1 [Source:HGNC Symbol;Acc:HGNC:23170] |

| | | | | | | | | |
|---|---|---|---|---|---|---|---|---|
| ENSG00000198563 | -0.325 | 7.582 | -4.047 | 0.00006 | 0.00158 | 1.104 | DDX39B | DExD-box helicase 39B [Source:HGNC Symbol;Acc:HGNC:13917] |
| ENSG00000198730 | 0.348 | 6.147 | 4.207 | 0.00003 | 0.00092 | 1.708 | CTR9 | CTR9 homolog, Paf1/RNA polymerase II complex component [Source:HGNC Symbol;Acc:HGNC:16850] |
| ENSG00000198814 | 0.305 | 2.897 | 3.669 | 0.00026 | 0.00543 | -0.067 | GK | glycerol kinase [Source:HGNC Symbol;Acc:HGNC:4289] |
| ENSG00000198826 | 0.785 | 3.181 | 6.229 | 0.00000 | 0.00000 | 11.819 | ARHGAP11A | Rho GTPase activating protein 11A [Source:HGNC Symbol;Acc:HGNC:15783] |
| ENSG00000198920 | -0.469 | 4.011 | -7.039 | 0.00000 | 0.00000 | 16.774 | KIAA0753 | KIAA0753 [Source:HGNC Symbol;Acc:HGNC:29110] |
| ENSG00000203760 | 0.627 | 1.551 | 5.233 | 0.00000 | 0.00001 | 6.647 | CENPW | centromere protein W [Source:HGNC Symbol;Acc:HGNC:21488] |
| ENSG00000204370 | 0.280 | 5.773 | 3.574 | 0.00038 | 0.00707 | -0.678 | SDHD | succinate dehydrogenase complex subunit D [Source:HGNC Symbol;Acc:HGNC:10683] |
| ENSG00000204599 | -0.265 | 4.339 | -3.602 | 0.00034 | 0.00649 | -0.454 | TRIM39 | tripartite motif containing 39 [Source:HGNC Symbol;Acc:HGNC:10065] |
| ENSG00000205208 | 0.367 | 3.856 | 3.723 | 0.00022 | 0.00465 | -0.028 | C4orf46 | chromosome 4 open reading frame 46 [Source:HGNC Symbol;Acc:HGNC:27320] |
| ENSG00000205544 | -0.376 | 3.028 | -4.219 | 0.00003 | 0.00089 | 2.073 | TMEM256 | transmembrane protein 256 [Source:HGNC Symbol;Acc:HGNC:28618] |
| ENSG00000213186 | 0.454 | 2.805 | 5.478 | 0.00000 | 0.00000 | 7.783 | TRIM59 | tripartite motif containing 59 [Source:HGNC Symbol;Acc:HGNC:30834] |
| ENSG00000213551 | 0.177 | 3.800 | 3.491 | 0.00052 | 0.00890 | -0.814 | DNAJC9 | DnaJ heat shock protein family (Hsp40) member C9 [Source:HGNC Symbol;Acc:HGNC:19123] |
| ENSG00000213859 | -0.403 | 3.189 | -4.314 | 0.00002 | 0.00062 | 2.440 | KCTD11 | potassium channel tetramerization domain containing 11 [Source:HGNC Symbol;Acc:HGNC:21302] |
| ENSG00000214941 | -0.397 | 3.944 | -5.943 | 0.00000 | 0.00000 | 10.217 | ZSWIM7 | zinc finger SWIM-type containing 7 [Source:HGNC Symbol;Acc:HGNC:26993] |
| ENSG00000215041 | -0.541 | 4.953 | -7.690 | 0.00000 | 0.00000 | 21.079 | NEURL4 | neuralized E3 ubiquitin protein ligase 4 [Source:HGNC Symbol;Acc:HGNC:34410] |
| ENSG00000215114 | 0.299 | 4.876 | 3.609 | 0.00033 | 0.00640 | -0.526 | UBXN2B | UBX domain protein 2B [Source:HGNC Symbol;Acc:HGNC:27035] |
| ENSG00000219200 | -0.449 | 1.568 | -3.730 | 0.00021 | 0.00456 | 0.424 | RNASEK | ribonuclease K [Source:HGNC Symbol;Acc:HGNC:33911] |
| ENSG00000220205 | -0.454 | 6.818 | -6.277 | 0.00000 | 0.00000 | 11.992 | VAMP2 | vesicle associated membrane protein 2 [Source:HGNC Symbol;Acc:HGNC:12643] |
| ENSG00000221823 | 0.213 | 4.788 | 3.629 | 0.00031 | 0.00605 | -0.445 | PPP3R1 | protein phosphatase 3 regulatory subunit B, alpha [Source:HGNC Symbol;Acc:HGNC:9317] |
| ENSG00000228486 | -0.565 | 3.881 | -4.717 | 0.00000 | 0.00014 | 4.089 | LINC01125 | long intergenic non-protein coding RNA 1125 [Source:HGNC Symbol;Acc:HGNC:49272] |
| ENSG00000236104 | -0.355 | 5.224 | -4.677 | 0.00000 | 0.00016 | 3.751 | ZBTB22 | zinc finger and BTB domain containing 22 [Source:HGNC Symbol;Acc:HGNC:13085] |
| ENSG00000237441 | -0.457 | 5.889 | -4.503 | 0.00001 | 0.00032 | 2.954 | RGL2 | ral guanine nucleotide dissociation stimulator like 2 [Source:HGNC Symbol;Acc:HGNC:9769] |
| ENSG00000237649 | 0.872 | 2.810 | 5.935 | 0.00000 | 0.00000 | 10.185 | KIFC1 | kinesin family member C1 [Source:HGNC Symbol;Acc:HGNC:6389] |
| ENSG00000239264 | -0.466 | 7.383 | -4.210 | 0.00003 | 0.00091 | 1.738 | TXNDC5 | thioredoxin domain containing 5 [Source:HGNC Symbol;Acc:HGNC:21073] |
| ENSG00000240038 | -0.484 | 4.735 | -3.902 | 0.00011 | 0.00255 | 0.607 | AMY2B | amylase, alpha 2B (pancreatic) [Source:HGNC Symbol;Acc:HGNC:478] |
| ENSG00000242612 | -0.481 | 2.822 | -4.119 | 0.00004 | 0.00124 | 1.710 | DECR2 | 2,4-dienoyl-CoA reductase 2 [Source:HGNC Symbol;Acc:HGNC:2754] |
| ENSG00000250091 | -0.841 | 0.191 | -4.390 | 0.00001 | 0.00047 | 2.853 | DNAH10OS | dynein axonemal heavy chain 10 opposite strand [Source:HGNC Symbol;Acc:HGNC:37121] |
| ENSG00000254004 | 0.557 | 3.381 | 3.706 | 0.00023 | 0.00488 | -0.046 | ZNF260 | zinc finger protein 260 [Source:HGNC Symbol;Acc:HGNC:13499] |
| ENSG00000256806 | -0.510 | 2.352 | -4.912 | 0.00000 | 0.00006 | 5.141 | C17orf100 | chromosome 17 open reading frame 100 [Source:HGNC Symbol;Acc:HGNC:34494] |
| ENSG00000269743 | 0.302 | 1.653 | 3.464 | 0.00057 | 0.00964 | -0.559 | SLC25A53 | solute carrier family 25 member 53 [Source:HGNC Symbol;Acc:HGNC:31894] |
| ENSG00000276043 | 0.844 | 1.837 | 5.466 | 0.00000 | 0.00000 | 7.783 | UHRF1 | ubiquitin like with PHD and ring finger domains 1 [Source:HGNC Symbol;Acc:HGNC:12556] |

**Supplemental Table S9.** Gene set enrichment analysis of the Hallmark library from differential gene expression of the comparison between *TP53* mutation to WT *TP53* samples. Results from the analysis of the CoMMpass data.

| ID | setSize | enrichmentScore | NES | pvalue | p.adjust | qvalue | rank |
|---|---|---|---|---|---|---|---|
| HALLMARK_G2M_CHECKPOINT | 74 | 0.654 | 2.958 | 0.00000 | 0.00000 | 0.00000 | 230 |
| HALLMARK_E2F_TARGETS | 94 | 0.597 | 2.786 | 0.00000 | 0.00000 | 0.00000 | 231 |
| HALLMARK_MITOTIC_SPINDLE | 38 | 0.603 | 2.408 | 0.00000 | 0.00000 | 0.00000 | 129 |
| HALLMARK_SPERMATOGENESIS | 12 | 0.766 | 2.272 | 0.00001 | 0.00001 | 0.00001 | 78 |
| HALLMARK_GLYCOLYSIS | 13 | 0.635 | 1.935 | 0.00096 | 0.00096 | 0.00061 | 138 |
| HALLMARK_ESTROGEN_RESPONSE_LATE | 11 | 0.618 | 1.791 | 0.00643 | 0.00643 | 0.00321 | 213 |
| HALLMARK_MTORC1_SIGNALING | 21 | 0.520 | 1.810 | 0.00711 | 0.00711 | 0.00321 | 172 |
| HALLMARK_P53_PATHWAY | 15 | -0.420 | -1.695 | 0.02450 | 0.02450 | 0.00967 | 115 |
| HALLMARK_OXIDATIVE_PHOSPHORYLATION | 11 | -0.448 | -1.585 | 0.04891 | 0.04891 | 0.01716 | 392 |
| HALLMARK_MYC_TARGETS_V1 | 24 | 0.327 | 1.179 | 0.26921 | 0.26921 | 0.08501 | 272 |
| HALLMARK_DNA_REPAIR | 21 | 0.331 | 1.151 | 0.30347 | 0.30347 | 0.08712 | 287 |
| HALLMARK_IL2_STAT5_SIGNALING | 10 | -0.304 | -1.036 | 0.40064 | 0.40064 | 0.10543 | 72 |

**Supplemental Table S10.** Gene set enrichment analysis of the C5 library from differential gene expression of the comparison between *TP53* mutation to WT *TP53* samples. Results from the CoMMpass dataset.

| ID | setSize | enrichmentScore | NES | pvalue | p.adjust | qvalue | rank |
|---|---|---|---|---|---|---|---|
| GOBP_CELL_CYCLE | 234 | 0.592 | 3.004 | 0.00000 | 0.00000 | 0.00000 | 257 |
| GOBP_CELL_CYCLE_PROCESS | 201 | 0.580 | 2.916 | 0.00000 | 0.00000 | 0.00000 | 228 |
| GOBP_MITOTIC_CELL_CYCLE | 175 | 0.589 | 2.944 | 0.00000 | 0.00000 | 0.00000 | 228 |
| GOBP_MITOTIC_CELL_CYCLE_PROCESS | 162 | 0.568 | 2.816 | 0.00000 | 0.00000 | 0.00000 | 228 |
| GOBP_CELL_DIVISION | 108 | 0.590 | 2.820 | 0.00000 | 0.00000 | 0.00000 | 257 |
| GOBP_CHROMOSOME_ORGANIZATION | 132 | 0.557 | 2.715 | 0.00000 | 0.00000 | 0.00000 | 211 |
| GOBP_CHROMOSOME_SEGREGATION | 88 | 0.591 | 2.754 | 0.00000 | 0.00000 | 0.00000 | 243 |
| GOBP_ORGANELLE_FISSION | 99 | 0.577 | 2.733 | 0.00000 | 0.00000 | 0.00000 | 157 |
| GOBP_MITOTIC_NUCLEAR_DIVISION | 79 | 0.602 | 2.761 | 0.00000 | 0.00000 | 0.00000 | 129 |
| GOBP_NUCLEAR_CHROMOSOME_SEGREGATION | 73 | 0.614 | 2.785 | 0.00000 | 0.00000 | 0.00000 | 211 |
| GOCC_CHROMOSOME | 182 | 0.495 | 2.476 | 0.00000 | 0.00000 | 0.00000 | 302 |
| GOBP_SISTER_CHROMATID_SEGREGATION | 61 | 0.637 | 2.802 | 0.00000 | 0.00000 | 0.00000 | 211 |
| GOBP_REGULATION_OF_CELL_CYCLE | 137 | 0.522 | 2.555 | 0.00000 | 0.00000 | 0.00000 | 197 |
| GOBP_REGULATION_OF_CELL_CYCLE_PROCESS | 112 | 0.551 | 2.647 | 0.00000 | 0.00000 | 0.00000 | 216 |
| GOBP_MITOTIC_SISTER_CHROMATID_SEGREGATION | 54 | 0.645 | 2.775 | 0.00000 | 0.00000 | 0.00000 | 152 |
| GOCC_CHROMOSOMAL_REGION | 90 | 0.529 | 2.471 | 0.00000 | 0.00000 | 0.00000 | 295 |
| GOBP_MITOTIC_CELL_CYCLE_PHASE_TRANSITION | 84 | 0.534 | 2.470 | 0.00000 | 0.00000 | 0.00000 | 216 |
| GOCC_SPINDLE | 73 | 0.540 | 2.452 | 0.00000 | 0.00000 | 0.00000 | 232 |
| GOBP_MICROTUBULE_BASED_PROCESS | 92 | 0.502 | 2.354 | 0.00000 | 0.00000 | 0.00000 | 232 |
| GOBP_CELL_CYCLE_PHASE_TRANSITION | 99 | 0.503 | 2.380 | 0.00000 | 0.00000 | 0.00000 | 216 |
| GOBP_MICROTUBULE_CYTOSKELETON_ORGANIZATION | 86 | 0.507 | 2.353 | 0.00000 | 0.00000 | 0.00000 | 232 |
| GOBP_CATION_TRANSPORT | 25 | -0.640 | -3.136 | 0.00000 | 0.00000 | 0.00000 | 252 |
| GOBP_REGULATION_OF_MITOTIC_CELL_CYCLE_PHASE_TRANSITION | 62 | 0.550 | 2.427 | 0.00000 | 0.00000 | 0.00000 | 197 |
| GOBP_CATION_TRANSMEMBRANE_TRANSPORT | 20 | -0.710 | -3.219 | 0.00000 | 0.00000 | 0.00000 | 143 |
| GOCC_CHROMOSOME_CENTROMERIC_REGION | 67 | 0.542 | 2.426 | 0.00000 | 0.00000 | 0.00000 | 169 |
| GOBP_REGULATION_OF_MITOTIC_CELL_CYCLE | 76 | 0.526 | 2.401 | 0.00000 | 0.00000 | 0.00000 | 197 |
| GOCC_MICROTUBULE_CYTOSKELETON | 132 | 0.463 | 2.254 | 0.00000 | 0.00000 | 0.00000 | 211 |
| GOBP_TRANSMEMBRANE_TRANSPORT | 26 | -0.617 | -3.059 | 0.00000 | 0.00000 | 0.00000 | 214 |
| GOBP_ION_TRANSMEMBRANE_TRANSPORT | 21 | -0.672 | -3.122 | 0.00000 | 0.00000 | 0.00000 | 143 |
| GOCC_CONDENSED_CHROMOSOME_CENTROMERIC_REGION | 53 | 0.559 | 2.395 | 0.00000 | 0.00000 | 0.00000 | 169 |
| GOBP_REGULATION_OF_CELL_CYCLE_PHASE_TRANSITION | 77 | 0.510 | 2.334 | 0.00000 | 0.00000 | 0.00000 | 197 |
| GOCC_CONDENSED_CHROMOSOME | 67 | 0.525 | 2.346 | 0.00000 | 0.00000 | 0.00000 | 205 |
| GOBP_REPRODUCTION | 89 | 0.483 | 2.252 | 0.00000 | 0.00000 | 0.00000 | 236 |

| | | | | | | | |
|---|---|---|---|---|---|---|---|
| GOBP_DNA_METABOLIC_PROCESS | 124 | 0.449 | 2.178 | 0.00000 | 0.00000 | 0.00000 | 272 |
| GOBP_REGULATION_OF_ORGANELLE_ORGANIZATION | 99 | 0.474 | 2.245 | 0.00000 | 0.00000 | 0.00001 | 211 |
| GOCC_SUPRAMOLECULAR_COMPLEX | 109 | 0.454 | 2.169 | 0.00000 | 0.00000 | 0.00001 | 139 |
| GOBP_REGULATION_OF_NUCLEAR_DIVISION | 32 | 0.627 | 2.422 | 0.00000 | 0.00000 | 0.00001 | 157 |
| GOBP_SECRETION | 22 | -0.625 | -2.940 | 0.00000 | 0.00000 | 0.00002 | 124 |
| GOBP_CHROMOSOME_SEPARATION | 27 | 0.651 | 2.413 | 0.00000 | 0.00000 | 0.00002 | 127 |
| GOBP_DNA_REPLICATION | 66 | 0.510 | 2.276 | 0.00000 | 0.00000 | 0.00002 | 283 |
| GOBP_REGULATION_OF_MITOTIC_NUCLEAR_DIVISION | 30 | 0.627 | 2.384 | 0.00000 | 0.00000 | 0.00002 | 127 |
| GOCC_MITOTIC_SPINDLE | 31 | 0.621 | 2.376 | 0.00000 | 0.00000 | 0.00002 | 222 |
| GOBP_POSITIVE_REGULATION_OF_CELL_CYCLE | 53 | 0.531 | 2.278 | 0.00000 | 0.00000 | 0.00002 | 224 |
| GOBP_CYTOSKELETON_ORGANIZATION | 112 | 0.446 | 2.140 | 0.00000 | 0.00000 | 0.00002 | 173 |
| GOBP_MEIOTIC_CELL_CYCLE | 50 | 0.545 | 2.315 | 0.00000 | 0.00000 | 0.00002 | 225 |
| GOBP_INORGANIC_ION_TRANSMEMBRANE_TRANSPORT | 17 | -0.683 | -2.919 | 0.00000 | 0.00000 | 0.00002 | 143 |
| GOBP_REGULATION_OF_CHROMOSOME_SEGREGATION | 29 | 0.634 | 2.395 | 0.00000 | 0.00000 | 0.00002 | 127 |
| GOBP_ION_TRANSPORT | 28 | -0.556 | -2.852 | 0.00000 | 0.00000 | 0.00003 | 143 |
| GOBP_DNA_CONFORMATION_CHANGE | 48 | 0.545 | 2.296 | 0.00000 | 0.00000 | 0.00003 | 293 |
| GOBP_SPINDLE_ORGANIZATION | 44 | 0.565 | 2.342 | 0.00000 | 0.00000 | 0.00003 | 251 |
| GOBP_RNA_PROCESSING | 54 | -0.407 | -2.534 | 0.00000 | 0.00000 | 0.00003 | 438 |
| GOBP_POSITIVE_REGULATION_OF_CELL_CYCLE_PROCESS | 47 | 0.553 | 2.323 | 0.00000 | 0.00000 | 0.00004 | 197 |
| GOBP_REGULATION_OF_CHROMOSOME_ORGANIZATION | 40 | 0.570 | 2.318 | 0.00000 | 0.00000 | 0.00004 | 211 |
| GOCC_MIDBODY | 27 | 0.636 | 2.360 | 0.00000 | 0.00000 | 0.00004 | 225 |
| GOBP_MEIOTIC_CELL_CYCLE_PROCESS | 41 | 0.564 | 2.302 | 0.00000 | 0.00000 | 0.00005 | 182 |
| GOBP_CALCIUM_ION_TRANSPORT | 10 | -0.780 | -2.681 | 0.00000 | 0.00000 | 0.00005 | 124 |
| GOBP_MITOTIC_SPINDLE_ORGANIZATION | 35 | 0.571 | 2.252 | 0.00000 | 0.00000 | 0.00006 | 207 |
| GOBP_VESICLE_MEDIATED_TRANSPORT | 48 | -0.423 | -2.562 | 0.00000 | 0.00000 | 0.00008 | 192 |
| GOBP_REGULATION_OF_TRANSPORT | 45 | -0.423 | -2.511 | 0.00000 | 0.00000 | 0.00008 | 150 |
| GOCC_VESICLE_MEMBRANE | 34 | -0.471 | -2.587 | 0.00001 | 0.00001 | 0.00009 | 117 |
| GOBP_REGULATION_OF_CHROMOSOME_SEPARATION | 24 | 0.642 | 2.318 | 0.00001 | 0.00001 | 0.00009 | 127 |
| GOMF_MICROTUBULE_BINDING | 39 | 0.554 | 2.238 | 0.00001 | 0.00001 | 0.00009 | 186 |
| GOBP_NEGATIVE_REGULATION_OF_CELL_CYCLE_PROCESS | 59 | 0.495 | 2.161 | 0.00001 | 0.00001 | 0.00010 | 216 |
| GOCC_MICROTUBULE_ORGANIZING_CENTER | 94 | 0.437 | 2.054 | 0.00001 | 0.00001 | 0.00010 | 258 |
| GOCC_CENTROSOME | 81 | 0.457 | 2.105 | 0.00001 | 0.00001 | 0.00016 | 258 |
| GOBP_MICROTUBULE_CYTOSKELETON_ORGANIZATION_INVOLVED_IN_MITOSIS | 45 | 0.529 | 2.207 | 0.00001 | 0.00001 | 0.00019 | 207 |
| GOBP_DNA_REPAIR | 89 | 0.437 | 2.040 | 0.00001 | 0.00001 | 0.00019 | 240 |
| GOBP_METAL_ION_TRANSPORT | 16 | -0.662 | -2.759 | 0.00002 | 0.00002 | 0.00024 | 124 |
| GOBP_DNA_DEPENDENT_DNA_REPLICATION | 49 | 0.510 | 2.156 | 0.00002 | 0.00002 | 0.00025 | 300 |
| GOBP_MEIOSIS_I_CELL_CYCLE_PROCESS | 25 | 0.612 | 2.231 | 0.00002 | 0.00002 | 0.00025 | 182 |
| GOMF_TUBULIN_BINDING | 44 | 0.526 | 2.182 | 0.00002 | 0.00002 | 0.00026 | 196 |

| | | | | | | | |
|---|---|---|---|---|---|---|---|
| GOBP_REGULATION_OF_CELL_DIVISION | 16 | 0.699 | 2.263 | 0.00002 | 0.00002 | 0.00027 | 222 |
| GOBP_CYTOKINESIS | 25 | 0.609 | 2.218 | 0.00002 | 0.00002 | 0.00029 | 226 |
| GOBP_FEMALE_MEIOTIC_NUCLEAR_DIVISION | 10 | 0.774 | 2.176 | 0.00002 | 0.00002 | 0.00030 | 107 |
| GOBP_CELLULAR_ION_HOMEOSTASIS | 12 | -0.702 | -2.604 | 0.00003 | 0.00003 | 0.00035 | 124 |
| GOBP_NEGATIVE_REGULATION_OF_CHROMOSOME_ORGANIZATION | 23 | 0.631 | 2.255 | 0.00003 | 0.00003 | 0.00036 | 127 |
| GOBP_METAPHASE_ANAPHASE_TRANSITION_OF_CELL_CYCLE | 23 | 0.631 | 2.255 | 0.00003 | 0.00003 | 0.00036 | 127 |
| GOBP_METAPHASE_PLATE_CONGRESSION | 22 | 0.629 | 2.219 | 0.00003 | 0.00003 | 0.00037 | 225 |
| GOBP_CHROMOSOME_LOCALIZATION | 25 | 0.598 | 2.180 | 0.00003 | 0.00003 | 0.00043 | 205 |
| GOCC_NUCLEAR_CHROMOSOME | 40 | 0.530 | 2.156 | 0.00004 | 0.00004 | 0.00050 | 291 |
| GOBP_CELL_CYCLE_G2_M_PHASE_TRANSITION | 43 | 0.514 | 2.120 | 0.00005 | 0.00005 | 0.00063 | 216 |
| GOBP_REGULATION_OF_MITOTIC_SISTER_CHROMATID_SEGREGATION | 18 | 0.652 | 2.175 | 0.00005 | 0.00005 | 0.00063 | 122 |
| GOBP_EXPORT_FROM_CELL | 17 | -0.600 | -2.564 | 0.00005 | 0.00005 | 0.00065 | 124 |
| GOBP_NON_MEMBRANE_BOUNDED_ORGANELLE_ASSEMBLY | 56 | 0.466 | 2.020 | 0.00007 | 0.00007 | 0.00082 | 173 |
| GOBP_NEGATIVE_REGULATION_OF_CELL_CYCLE_PHASE_TRANSITION | 51 | 0.481 | 2.049 | 0.00007 | 0.00007 | 0.00088 | 216 |
| GOBP_ATTACHMENT_OF_SPINDLE_MICROTUBULES_TO_KINETOCHORE | 16 | 0.669 | 2.166 | 0.00009 | 0.00009 | 0.00107 | 222 |
| GOBP_NEGATIVE_REGULATION_OF_MITOTIC_CELL_CYCLE_PHASE_TRANSITION | 38 | 0.531 | 2.134 | 0.00009 | 0.00009 | 0.00107 | 216 |
| GOBP_RNA_SPLICING | 24 | -0.503 | -2.430 | 0.00012 | 0.00012 | 0.00135 | 361 |
| GOBP_MITOTIC_METAPHASE_PLATE_CONGRESSION | 17 | 0.648 | 2.131 | 0.00012 | 0.00012 | 0.00142 | 197 |
| HP_ABNORMAL_DUODENUM_MORPHOLOGY | 19 | 0.608 | 2.061 | 0.00013 | 0.00013 | 0.00142 | 221 |
| GOCC_VACUOLE | 27 | -0.460 | -2.320 | 0.00014 | 0.00014 | 0.00158 | 117 |
| GOMF_MICROTUBULE_MOTOR_ACTIVITY | 17 | 0.643 | 2.116 | 0.00016 | 0.00016 | 0.00172 | 184 |
| GOBP_METAL_ION_HOMEOSTASIS | 12 | -0.656 | -2.432 | 0.00017 | 0.00017 | 0.00180 | 124 |
| GOBP_REGULATION_OF_MICROTUBULE_BASED_PROCESS | 24 | 0.577 | 2.085 | 0.00017 | 0.00017 | 0.00180 | 255 |
| GOCC_MICROTUBULE | 53 | 0.460 | 1.972 | 0.00017 | 0.00017 | 0.00182 | 107 |
| GOBP_REGULATION_OF_CYTOKINESIS | 13 | 0.696 | 2.128 | 0.00018 | 0.00018 | 0.00185 | 222 |
| GOBP_NEGATIVE_REGULATION_OF_MITOTIC_CELL_CYCLE | 43 | 0.490 | 2.021 | 0.00018 | 0.00018 | 0.00185 | 216 |
| HP_WEIGHT_LOSS | 22 | 0.592 | 2.088 | 0.00020 | 0.00020 | 0.00201 | 221 |
| GOBP_NEGATIVE_REGULATION_OF_METAPHASE_ANAPHASE_TRANSITION_OF_CELL_CYCLE | 17 | 0.638 | 2.098 | 0.00020 | 0.00020 | 0.00202 | 122 |
| GOBP_REGULATION_OF_SYSTEM_PROCESS | 16 | -0.598 | -2.493 | 0.00022 | 0.00022 | 0.00218 | 124 |
| GOBP_REGULATION_OF_TRANSFERASE_ACTIVITY | 68 | 0.442 | 1.984 | 0.00022 | 0.00022 | 0.00218 | 225 |
| GOBP_REGULATION_OF_VESICLE_MEDIATED_TRANSPORT | 17 | -0.561 | -2.399 | 0.00022 | 0.00022 | 0.00220 | 192 |
| GOBP_REGULATION_OF_CYCLIN_DEPENDENT_PROTEIN_KINASE_ACTIVITY | 17 | 0.634 | 2.086 | 0.00023 | 0.00023 | 0.00229 | 224 |
| GOBP_NEGATIVE_REGULATION_OF_CELL_CYCLE | 65 | 0.446 | 1.985 | 0.00026 | 0.00026 | 0.00249 | 216 |
| GOBP_CARBOHYDRATE_METABOLIC_PROCESS | 22 | -0.499 | -2.349 | 0.00026 | 0.00026 | 0.00254 | 159 |
| GOBP_SISTER_CHROMATID_COHESION | 17 | 0.625 | 2.056 | 0.00031 | 0.00031 | 0.00295 | 127 |
| GOBP_REGULATION_OF_REPRODUCTIVE_PROCESS | 10 | 0.722 | 2.032 | 0.00031 | 0.00031 | 0.00295 | 172 |
| GOBP_INTRACELLULAR_TRANSPORT | 65 | -0.289 | -1.894 | 0.00032 | 0.00032 | 0.00295 | 180 |
| GOBP_ION_HOMEOSTASIS | 14 | -0.615 | -2.419 | 0.00033 | 0.00033 | 0.00310 | 124 |

| | | | | | | | |
|---|---|---|---|---|---|---|---|
| HP_SLOPING_FOREHEAD | 30 | 0.533 | 2.028 | 0.00034 | 0.00034 | 0.00310 | 196 |
| GOBP_REGULATION_OF_TRANSMEMBRANE_TRANSPORT | 11 | -0.655 | -2.349 | 0.00034 | 0.00034 | 0.00310 | 122 |
| GOCC_POSTSYNAPSE | 11 | -0.653 | -2.344 | 0.00035 | 0.00035 | 0.00319 | 124 |
| GOBP_IN_UTERO_EMBRYONIC_DEVELOPMENT | 21 | 0.591 | 2.061 | 0.00037 | 0.00037 | 0.00326 | 157 |
| GOBP_CELLULAR_HOMEOSTASIS | 17 | -0.543 | -2.320 | 0.00037 | 0.00037 | 0.00326 | 124 |
| GOBP_MITOTIC_CELL_CYCLE_CHECKPOINT_SIGNALING | 37 | 0.495 | 1.978 | 0.00038 | 0.00038 | 0.00333 | 216 |
| GOBP_DNA_GEOMETRIC_CHANGE | 23 | 0.585 | 2.089 | 0.00038 | 0.00038 | 0.00333 | 272 |
| GOCC_INTRINSIC_COMPONENT_OF_PLASMA_MEMBRANE | 23 | -0.469 | -2.230 | 0.00039 | 0.00039 | 0.00333 | 102 |
| GOMF_CYTOSKELETAL_PROTEIN_BINDING | 60 | 0.436 | 1.911 | 0.00039 | 0.00039 | 0.00334 | 131 |
| GOBP_DNA_UNWINDING_INVOLVED_IN_DNA_REPLICATION | 12 | 0.679 | 2.027 | 0.00040 | 0.00040 | 0.00337 | 233 |
| GOCC_PRESYNAPSE | 15 | -0.574 | -2.319 | 0.00042 | 0.00042 | 0.00356 | 90 |
| HP_ABNORMALITY_OF_GLOBE_SIZE | 30 | 0.527 | 2.005 | 0.00047 | 0.00047 | 0.00393 | 233 |
| GOCC_MEMBRANE_PROTEIN_COMPLEX | 30 | -0.413 | -2.174 | 0.00048 | 0.00048 | 0.00393 | 398 |
| GOBP_CELLULAR_LIPID_METABOLIC_PROCESS | 27 | -0.431 | -2.169 | 0.00048 | 0.00048 | 0.00393 | 159 |
| GOBP_REGULATION_OF_ION_TRANSPORT | 12 | -0.620 | -2.301 | 0.00049 | 0.00049 | 0.00399 | 122 |
| GOBP_MITOTIC_SPINDLE_ASSEMBLY | 17 | 0.615 | 2.022 | 0.00051 | 0.00051 | 0.00409 | 129 |
| GOBP_MRNA_PROCESSING | 29 | -0.418 | -2.171 | 0.00052 | 0.00052 | 0.00409 | 361 |
| GOBP_MEIOTIC_CHROMOSOME_SEGREGATION | 22 | 0.571 | 2.013 | 0.00052 | 0.00052 | 0.00409 | 127 |
| GOBP_DNA_RECOMBINATION | 48 | 0.461 | 1.943 | 0.00052 | 0.00052 | 0.00409 | 233 |
| GOBP_RNA_SPLICING_VIA_TRANSESTERIFICATION_REACTIONS | 19 | -0.499 | -2.225 | 0.00053 | 0.00053 | 0.00411 | 361 |
| GOCC_MICROTUBULE_ASSOCIATED_COMPLEX | 25 | 0.536 | 1.951 | 0.00054 | 0.00054 | 0.00416 | 81 |
| GOBP_CELLULAR_AMIDE_METABOLIC_PROCESS | 37 | -0.376 | -2.123 | 0.00057 | 0.00057 | 0.00438 | 382 |
| GOBP_CELL_CYCLE_CHECKPOINT_SIGNALING | 47 | 0.464 | 1.950 | 0.00059 | 0.00059 | 0.00446 | 216 |
| GOBP_MITOTIC_CYTOKINESIS | 19 | 0.571 | 1.937 | 0.00059 | 0.00059 | 0.00446 | 226 |
| GOBP_EMBRYO_DEVELOPMENT | 42 | 0.472 | 1.940 | 0.00065 | 0.00065 | 0.00486 | 219 |
| GOBP_NITROGEN_COMPOUND_TRANSPORT | 72 | -0.277 | -1.858 | 0.00066 | 0.00066 | 0.00494 | 256 |
| GOBP_VESICLE_ORGANIZATION | 13 | -0.594 | -2.268 | 0.00067 | 0.00067 | 0.00495 | 120 |
| GOBP_RECOMBINATIONAL_REPAIR | 34 | 0.502 | 1.967 | 0.00068 | 0.00068 | 0.00498 | 233 |
| GOCC_SPINDLE_POLE | 41 | 0.471 | 1.923 | 0.00070 | 0.00070 | 0.00513 | 232 |
| GOBP_EMBRYO_DEVELOPMENT_ENDING_IN_BIRTH_OR_EGG_HATCHING | 29 | 0.527 | 1.989 | 0.00072 | 0.00072 | 0.00523 | 213 |
| GOBP_REGULATION_OF_ION_TRANSMEMBRANE_TRANSPORT | 10 | -0.637 | -2.189 | 0.00073 | 0.00073 | 0.00528 | 122 |
| GOBP_CELL_CELL_SIGNALING | 32 | -0.412 | -2.219 | 0.00076 | 0.00076 | 0.00542 | 150 |
| GOCC_SYNAPSE | 32 | -0.411 | -2.217 | 0.00079 | 0.00079 | 0.00558 | 129 |
| GOBP_FEMALE_GAMETE_GENERATION | 15 | 0.624 | 1.992 | 0.00080 | 0.00080 | 0.00562 | 107 |
| GOBP_REGULATION_OF_SECRETION | 14 | -0.582 | -2.288 | 0.00081 | 0.00081 | 0.00566 | 201 |
| GOMF_SINGLE_STRANDED_DNA_BINDING | 22 | 0.560 | 1.975 | 0.00082 | 0.00082 | 0.00566 | 257 |
| GOBP_CYTOSKELETON_DEPENDENT_CYTOKINESIS | 20 | 0.577 | 1.983 | 0.00082 | 0.00082 | 0.00566 | 226 |
| HP_CLINODACTYLY | 48 | 0.453 | 1.909 | 0.00082 | 0.00082 | 0.00566 | 221 |

| | | | | | | | |
|---|---|---|---|---|---|---|---|
| GOBP_SPINDLE_ASSEMBLY | 29 | 0.524 | 1.981 | 0.00083 | 0.00083 | 0.00566 | 129 |
| GOMF_ADENYL_NUCLEOTIDE_BINDING | 123 | 0.357 | 1.728 | 0.00086 | 0.00086 | 0.00582 | 204 |
| GOBP_DOUBLE_STRAND_BREAK_REPAIR | 52 | 0.436 | 1.862 | 0.00095 | 0.00095 | 0.00640 | 238 |
| GOCC_CELL_LEADING_EDGE | 14 | -0.575 | -2.260 | 0.00096 | 0.00096 | 0.00642 | 187 |
| HP_SOFT_TISSUE_SARCOMA | 12 | 0.655 | 1.957 | 0.00097 | 0.00097 | 0.00646 | 99 |
| GOBP_RIBONUCLEOPROTEIN_COMPLEX_BIOGENESIS | 26 | -0.420 | -2.082 | 0.00099 | 0.00099 | 0.00650 | 418 |
| HP_ABNORMALITY_OF_THE_PALPEBRAL_FISSURES | 63 | 0.409 | 1.813 | 0.00102 | 0.00102 | 0.00664 | 297 |
| GOCC_SPINDLE_MIDZONE | 13 | 0.651 | 1.992 | 0.00102 | 0.00102 | 0.00664 | 225 |
| GOMF_PHOSPHOLIPID_BINDING | 17 | -0.509 | -2.175 | 0.00103 | 0.00103 | 0.00664 | 142 |
| GOBP_ESTABLISHMENT_OF_ORGANELLE_LOCALIZATION | 42 | 0.465 | 1.910 | 0.00103 | 0.00103 | 0.00664 | 205 |
| HP_ABNORMALITY_OF_THE_SMALL_INTESTINE | 22 | 0.554 | 1.956 | 0.00104 | 0.00104 | 0.00664 | 242 |
| GOBP_LIPID_METABOLIC_PROCESS | 43 | -0.325 | -1.911 | 0.00112 | 0.00112 | 0.00706 | 159 |
| GOMF_TRANSPORTER_ACTIVITY | 19 | -0.480 | -2.138 | 0.00112 | 0.00112 | 0.00706 | 214 |
| GOBP_NEGATIVE_REGULATION_OF_TRANSFERASE_ACTIVITY | 11 | 0.663 | 1.923 | 0.00121 | 0.00121 | 0.00760 | 149 |
| GOBP_POSITIVE_REGULATION_OF_MITOTIC_CELL_CYCLE_PHASE_TRANSITION | 22 | 0.550 | 1.941 | 0.00122 | 0.00122 | 0.00760 | 211 |
| GOBP_CELL_CYCLE_DNA_REPLICATION | 22 | 0.549 | 1.939 | 0.00124 | 0.00124 | 0.00770 | 300 |
| GOBP_CHEMICAL_HOMEOSTASIS | 23 | -0.433 | -2.058 | 0.00132 | 0.00132 | 0.00809 | 124 |
| GOBP_PROTEIN_DNA_COMPLEX_SUBUNIT_ORGANIZATION | 28 | 0.512 | 1.916 | 0.00132 | 0.00132 | 0.00809 | 247 |
| GOBP_PROTEIN_DNA_COMPLEX_ASSEMBLY | 27 | 0.525 | 1.946 | 0.00135 | 0.00135 | 0.00820 | 247 |
| HP_SOMATIC_MUTATION | 11 | 0.657 | 1.908 | 0.00143 | 0.00143 | 0.00863 | 196 |
| GOBP_CELL_CYCLE_G1_S_PHASE_TRANSITION | 30 | 0.506 | 1.924 | 0.00147 | 0.00147 | 0.00881 | 268 |
| GOBP_PEPTIDE_METABOLIC_PROCESS | 29 | -0.394 | -2.050 | 0.00151 | 0.00151 | 0.00901 | 382 |
| GOCC_VACUOLAR_MEMBRANE | 16 | -0.536 | -2.236 | 0.00157 | 0.00157 | 0.00934 | 117 |
| GOCC_SPINDLE_MICROTUBULE | 20 | 0.562 | 1.932 | 0.00159 | 0.00159 | 0.00936 | 107 |
| HP_JOINT_HYPERFLEXIBILITY | 15 | 0.605 | 1.930 | 0.00163 | 0.00163 | 0.00950 | 270 |
| GOBP_ORGANONITROGEN_COMPOUND_BIOSYNTHETIC_PROCESS | 59 | -0.278 | -1.781 | 0.00163 | 0.00163 | 0.00950 | 257 |
| GOBP_RESPONSE_TO_TUMOR_NECROSIS_FACTOR | 14 | -0.552 | -2.170 | 0.00165 | 0.00165 | 0.00957 | 185 |
| GOBP_SYNAPTIC_SIGNALING | 19 | -0.460 | -2.052 | 0.00172 | 0.00172 | 0.00989 | 124 |
| GOBP_GLAND_DEVELOPMENT | 15 | 0.602 | 1.921 | 0.00182 | 0.00182 | 0.01032 | 191 |
| GOMF_PHOSPHATIDYLINOSITOL_BINDING | 13 | -0.560 | -2.141 | 0.00183 | 0.00183 | 0.01032 | 249 |
| GOBP_NCRNA_METABOLIC_PROCESS | 25 | -0.403 | -1.974 | 0.00183 | 0.00183 | 0.01032 | 429 |
| GOBP_NCRNA_PROCESSING | 25 | -0.403 | -1.974 | 0.00183 | 0.00183 | 0.01032 | 429 |
| GOBP_PHOSPHORYLATION | 98 | 0.366 | 1.730 | 0.00186 | 0.00186 | 0.01039 | 225 |
| HP_ABNORMAL_BONE_STRUCTURE | 42 | 0.459 | 1.885 | 0.00195 | 0.00195 | 0.01086 | 237 |
| HP_INTRAUTERINE_GROWTH_RETARDATION | 58 | 0.417 | 1.815 | 0.00200 | 0.00200 | 0.01097 | 247 |
| GOCC_POLYMERIC_CYTOSKELETAL_FIBER | 58 | 0.416 | 1.812 | 0.00200 | 0.00200 | 0.01097 | 107 |
| HP_SLANTING_OF_THE_PALPEBRAL_FISSURE | 57 | 0.419 | 1.820 | 0.00200 | 0.00200 | 0.01097 | 297 |
| GOBP_NEGATIVE_REGULATION_OF_NUCLEAR_DIVISION | 20 | 0.556 | 1.910 | 0.00205 | 0.00205 | 0.01106 | 122 |

| | | | | | | | |
|---|---|---|---|---|---|---|---|
| GOBP_POSITIVE_REGULATION_OF_CELL_CYCLE_PHASE_TRANSITION | 28 | 0.503 | 1.884 | 0.00206 | 0.00206 | 0.01106 | 188 |
| GOBP_REGULATION_OF_DNA_REPLICATION | 28 | 0.498 | 1.862 | 0.00206 | 0.00206 | 0.01106 | 300 |
| GOMF_RIBONUCLEOTIDE_BINDING | 131 | 0.339 | 1.653 | 0.00206 | 0.00206 | 0.01106 | 211 |
| GOBP_ORGANELLE_LOCALIZATION | 47 | 0.439 | 1.843 | 0.00208 | 0.00208 | 0.01109 | 205 |
| GOBP_DNA_PACKAGING | 27 | 0.513 | 1.903 | 0.00211 | 0.00211 | 0.01120 | 323 |
| GOBP_REGULATION_OF_DNA_DEPENDENT_DNA_REPLICATION | 18 | 0.569 | 1.900 | 0.00219 | 0.00219 | 0.01154 | 272 |
| GOMF_LIPID_BINDING | 22 | -0.434 | -2.041 | 0.00227 | 0.00227 | 0.01191 | 150 |
| GOBP_NEGATIVE_REGULATION_OF_ORGANELLE_ORGANIZATION | 45 | 0.440 | 1.835 | 0.00230 | 0.00230 | 0.01199 | 164 |
| GOBP_DNA_REPLICATION_INITIATION | 17 | 0.581 | 1.913 | 0.00233 | 0.00233 | 0.01212 | 256 |
| GOCC_KINESIN_COMPLEX | 16 | 0.583 | 1.889 | 0.00235 | 0.00235 | 0.01213 | 81 |
| GOBP_NEGATIVE_REGULATION_OF_TRANSPORT | 14 | -0.536 | -2.107 | 0.00249 | 0.00249 | 0.01279 | 192 |
| GOBP_CELLULAR_RESPONSE_TO_DNA_DAMAGE_STIMULUS | 116 | 0.347 | 1.669 | 0.00250 | 0.00250 | 0.01279 | 240 |
| GOCC_SUPRAMOLECULAR_POLYMER | 65 | 0.397 | 1.768 | 0.00251 | 0.00251 | 0.01279 | 107 |
| GOBP_MICROTUBULE_BASED_MOVEMENT | 23 | 0.539 | 1.926 | 0.00258 | 0.00258 | 0.01305 | 184 |
| GOBP_REGULATION_OF_PROTEIN_SERINE_THREONINE_KINASE_ACTIVITY | 27 | 0.502 | 1.861 | 0.00267 | 0.00267 | 0.01345 | 225 |
| GOBP_AMIDE_BIOSYNTHETIC_PROCESS | 29 | -0.378 | -1.963 | 0.00268 | 0.00268 | 0.01345 | 382 |
| HP_PROPTOSIS | 24 | 0.520 | 1.878 | 0.00270 | 0.00270 | 0.01346 | 242 |
| GOBP_REPRODUCTIVE_SYSTEM_DEVELOPMENT | 15 | 0.583 | 1.859 | 0.00288 | 0.00288 | 0.01430 | 221 |
| HP_ABNORMALITY_OF_THE_NASAL_ALAE | 32 | 0.475 | 1.835 | 0.00298 | 0.00298 | 0.01471 | 91 |
| HP_EXTERNAL_EAR_MALFORMATION | 13 | 0.621 | 1.899 | 0.00315 | 0.00315 | 0.01541 | 221 |
| GOBP_MRNA_METABOLIC_PROCESS | 36 | -0.334 | -1.871 | 0.00315 | 0.00315 | 0.01541 | 389 |
| HP_ABNORMALITY_OF_CHROMOSOME_STABILITY | 13 | 0.614 | 1.877 | 0.00321 | 0.00321 | 0.01565 | 221 |
| HP_MULTIPLE_CAFE_AU_LAIT_SPOTS | 16 | 0.572 | 1.854 | 0.00326 | 0.00326 | 0.01576 | 221 |
| HP_NEOPLASM | 51 | 0.415 | 1.768 | 0.00327 | 0.00327 | 0.01576 | 237 |
| HP_SINGLE_TRANSVERSE_PALMAR_CREASE | 13 | 0.613 | 1.876 | 0.00335 | 0.00335 | 0.01602 | 23 |
| GOCC_SECRETORY_VESICLE | 29 | -0.368 | -1.913 | 0.00336 | 0.00336 | 0.01602 | 129 |
| GOMF_DNA_HELICASE_ACTIVITY | 17 | 0.566 | 1.861 | 0.00337 | 0.00337 | 0.01602 | 293 |
| HP_ABNORMAL_UVULA_MORPHOLOGY | 26 | 0.502 | 1.847 | 0.00340 | 0.00340 | 0.01608 | 258 |
| HP_FINGER_CLINODACTYLY | 42 | 0.440 | 1.810 | 0.00344 | 0.00344 | 0.01620 | 221 |
| HP_FINGER_SYNDACTYLY | 17 | 0.564 | 1.856 | 0.00350 | 0.00350 | 0.01637 | 221 |
| GOMF_PROTEIN_SERINE_THREONINE_TYROSINE_KINASE_ACTIVITY | 28 | 0.487 | 1.824 | 0.00351 | 0.00351 | 0.01637 | 197 |
| GOBP_REGULATION_OF_CELL_CYCLE_G2_M_PHASE_TRANSITION | 32 | 0.469 | 1.809 | 0.00357 | 0.00357 | 0.01660 | 216 |
| GOCC_SPLICEOSOMAL_COMPLEX | 16 | -0.504 | -2.103 | 0.00363 | 0.00363 | 0.01681 | 356 |
| GOBP_POSITIVE_REGULATION_OF_MITOTIC_CELL_CYCLE | 24 | 0.511 | 1.844 | 0.00368 | 0.00368 | 0.01693 | 211 |
| HP_SYNDACTYLY | 30 | 0.474 | 1.802 | 0.00372 | 0.00372 | 0.01704 | 221 |
| GOBP_MEMBRANE_ORGANIZATION | 40 | -0.316 | -1.817 | 0.00397 | 0.00397 | 0.01813 | 147 |
| GOMF_PROTEIN_SERINE_KINASE_ACTIVITY | 24 | 0.509 | 1.838 | 0.00404 | 0.00404 | 0.01837 | 197 |
| HP_ABNORMAL_PALMAR_DERMATOGLYPHICS | 14 | 0.588 | 1.839 | 0.00410 | 0.00410 | 0.01855 | 23 |

| | | | | | | | |
|---|---|---|---|---|---|---|---|
| GOMF_ATP_DEPENDENT_ACTIVITY_ACTING_ON_DNA | 24 | 0.508 | 1.833 | 0.00429 | 0.00429 | 0.01931 | 293 |
| GOBP_MULTICELLULAR_ORGANISM_REPRODUCTION | 44 | 0.429 | 1.779 | 0.00434 | 0.00434 | 0.01944 | 193 |
| HP_ABNORMALITY_OF_THE_FEMALE_GENITALIA | 47 | 0.415 | 1.744 | 0.00477 | 0.00477 | 0.02123 | 277 |
| GOBP_CELLULAR_PROCESS_INVOLVED_IN_REPRODUCTION_IN_MULTICELLULAR_ORGANISM | 24 | 0.502 | 1.814 | 0.00478 | 0.00478 | 0.02123 | 193 |
| GOBP_POSITIVE_REGULATION_OF_TRANSPORT | 28 | -0.366 | -1.874 | 0.00484 | 0.00484 | 0.02143 | 124 |
| GOBP_REGULATION_OF_PROTEIN_PHOSPHORYLATION | 55 | 0.402 | 1.735 | 0.00491 | 0.00491 | 0.02162 | 225 |
| GOBP_CHROMOSOME_CONDENSATION | 10 | 0.642 | 1.806 | 0.00502 | 0.00502 | 0.02184 | 257 |
| GOBP_DEVELOPMENTAL_PROCESS_INVOLVED_IN_REPRODUCTION | 37 | 0.444 | 1.774 | 0.00502 | 0.00502 | 0.02184 | 152 |
| GOBP_DNA_BIOSYNTHETIC_PROCESS | 31 | 0.465 | 1.779 | 0.00502 | 0.00502 | 0.02184 | 215 |
| GOBP_NEGATIVE_REGULATION_OF_CELLULAR_COMPONENT_ORGANIZATION | 53 | 0.406 | 1.741 | 0.00505 | 0.00505 | 0.02184 | 164 |
| GOBP_REGULATION_OF_MICROTUBULE_CYTOSKELETON_ORGANIZATION | 16 | 0.565 | 1.829 | 0.00521 | 0.00521 | 0.02247 | 251 |
| HP_ABNORMAL_5TH_FINGER_MORPHOLOGY | 45 | 0.418 | 1.742 | 0.00524 | 0.00524 | 0.02247 | 221 |
| GOBP_LOCALIZATION_WITHIN_MEMBRANE | 23 | -0.394 | -1.874 | 0.00533 | 0.00533 | 0.02269 | 249 |
| GOMF_ATP_DEPENDENT_ACTIVITY | 67 | 0.380 | 1.697 | 0.00533 | 0.00533 | 0.02269 | 237 |
| GOBP_HOMEOSTATIC_PROCESS | 36 | -0.324 | -1.815 | 0.00539 | 0.00539 | 0.02285 | 124 |
| GOBP_MITOTIC_DNA_REPLICATION | 10 | 0.635 | 1.788 | 0.00572 | 0.00572 | 0.02404 | 238 |
| GOMF_CATALYTIC_ACTIVITY_ACTING_ON_DNA | 37 | 0.441 | 1.761 | 0.00572 | 0.00572 | 0.02404 | 280 |
| GOMF_CYTOSKELETAL_MOTOR_ACTIVITY | 21 | 0.510 | 1.777 | 0.00588 | 0.00588 | 0.02462 | 90 |
| HP_ABNORMAL_UTERUS_MORPHOLOGY | 19 | 0.526 | 1.783 | 0.00648 | 0.00648 | 0.02698 | 221 |
| GOBP_ENDOCYTOSIS | 18 | -0.457 | -2.000 | 0.00650 | 0.00650 | 0.02698 | 183 |
| GOMF_PROTEIN_KINASE_ACTIVITY | 30 | 0.460 | 1.749 | 0.00671 | 0.00671 | 0.02771 | 197 |
| GOCC_ACTIN_CYTOSKELETON | 17 | -0.453 | -1.934 | 0.00673 | 0.00673 | 0.02771 | 94 |
| GOBP_REGULATION_OF_PROTEIN_MODIFICATION_PROCESS | 86 | 0.348 | 1.617 | 0.00726 | 0.00726 | 0.02967 | 225 |
| HP_CAFE_AU_LAIT_SPOT | 23 | 0.498 | 1.779 | 0.00727 | 0.00727 | 0.02967 | 221 |
| HP_ABNORMALITY_OF_EPIPHYSIS_MORPHOLOGY | 18 | 0.533 | 1.777 | 0.00733 | 0.00733 | 0.02979 | 247 |
| GOBP_CYTOSOLIC_TRANSPORT | 10 | -0.569 | -1.955 | 0.00742 | 0.00742 | 0.03005 | 248 |
| GOBP_ESTABLISHMENT_OF_PROTEIN_LOCALIZATION | 67 | -0.242 | -1.611 | 0.00748 | 0.00748 | 0.03018 | 214 |
| GOCC_ENDOCYTIC_VESICLE | 12 | -0.535 | -1.985 | 0.00766 | 0.00766 | 0.03073 | 171 |
| GOBP_REGULATION_OF_PROTEIN_KINASE_ACTIVITY | 42 | 0.416 | 1.710 | 0.00768 | 0.00768 | 0.03073 | 225 |
| GOBP_REGULATION_OF_CELL_CYCLE_G1_S_PHASE_TRANSITION | 18 | 0.529 | 1.764 | 0.00784 | 0.00784 | 0.03126 | 211 |
| HP_APLASIA_HYPOPLASIA_AFFECTING_THE_EYE | 34 | 0.447 | 1.749 | 0.00792 | 0.00792 | 0.03143 | 233 |
| GOCC_PROTEIN_DNA_COMPLEX | 18 | 0.528 | 1.761 | 0.00797 | 0.00797 | 0.03152 | 238 |
| HP_GLAUCOMA | 26 | 0.479 | 1.764 | 0.00801 | 0.00801 | 0.03154 | 99 |
| GOBP_REGULATION_OF_NERVOUS_SYSTEM_DEVELOPMENT | 15 | 0.555 | 1.771 | 0.00811 | 0.00811 | 0.03183 | 173 |
| GOMF_SINGLE_STRANDED_DNA_HELICASE_ACTIVITY | 12 | 0.592 | 1.766 | 0.00825 | 0.00825 | 0.03226 | 293 |
| HP_SEVERE_SHORT_STATURE | 12 | 0.591 | 1.765 | 0.00853 | 0.00853 | 0.03319 | 137 |
| GOBP_NEGATIVE_REGULATION_OF_MOLECULAR_FUNCTION | 43 | 0.415 | 1.711 | 0.00859 | 0.00859 | 0.03331 | 149 |
| GOBP_REGULATION_OF_CYSTEINE_TYPE_ENDOPEPTIDASE_ACTIVITY | 11 | -0.542 | -1.946 | 0.00881 | 0.00881 | 0.03403 | 19 |

| | | | | | | | |
|---|---|---|---|---|---|---|---|
| GOBP_MICROTUBULE_POLYMERIZATION_OR_DEPOLYMERIZATION | 19 | 0.518 | 1.755 | 0.00903 | 0.00903 | 0.03462 | 186 |
| HP_SARCOMA | 15 | 0.551 | 1.757 | 0.00903 | 0.00903 | 0.03462 | 99 |
| GOCC_INTERCELLULAR_BRIDGE | 10 | 0.623 | 1.751 | 0.00906 | 0.00906 | 0.03462 | 211 |
| HP_CONGENITAL_MALFORMATION_OF_THE_GREAT_ARTERIES | 23 | 0.491 | 1.752 | 0.00912 | 0.00912 | 0.03469 | 297 |
| HP_DEVIATION_OF_THE_HAND_OR_OF_FINGERS_OF_THE_HAND | 47 | 0.402 | 1.690 | 0.00931 | 0.00931 | 0.03528 | 221 |
| HP_REDUCED_TENDON_REFLEXES | 16 | -0.446 | -1.858 | 0.00944 | 0.00944 | 0.03565 | 194 |
| GOBP_MULTI_ORGANISM_REPRODUCTIVE_PROCESS | 47 | 0.402 | 1.687 | 0.00953 | 0.00953 | 0.03587 | 201 |
| GOCC_SECRETORY_GRANULE | 24 | -0.381 | -1.840 | 0.00977 | 0.00977 | 0.03663 | 90 |
| HP_RENAL_HYPOPLASIA_APLASIA | 33 | 0.442 | 1.719 | 0.00984 | 0.00984 | 0.03676 | 196 |
| HP_ABNORMAL_SHAPE_OF_THE_PALPEBRAL_FISSURE | 11 | 0.601 | 1.746 | 0.00989 | 0.00989 | 0.03682 | 221 |
| GOBP_GAMETE_GENERATION | 40 | 0.419 | 1.702 | 0.01005 | 0.01005 | 0.03728 | 193 |
| HP_DUODENAL_STENOSIS | 11 | 0.600 | 1.742 | 0.01017 | 0.01017 | 0.03752 | 221 |
| HP_SHORT_LONG_BONE | 20 | 0.505 | 1.736 | 0.01020 | 0.01020 | 0.03752 | 233 |
| GOBP_NEGATIVE_REGULATION_OF_PHOSPHORYLATION | 12 | 0.583 | 1.742 | 0.01028 | 0.01028 | 0.03759 | 149 |
| HP_UPSLANTED_PALPEBRAL_FISSURE | 31 | 0.447 | 1.711 | 0.01029 | 0.01029 | 0.03759 | 266 |
| HP_PEDIATRIC_ONSET | 28 | -0.352 | -1.804 | 0.01040 | 0.01040 | 0.03788 | 122 |
| HP_DECREASED_HEAD_CIRCUMFERENCE | 98 | 0.331 | 1.562 | 0.01045 | 0.01045 | 0.03790 | 255 |
| GOBP_ORGANELLE_ASSEMBLY | 76 | 0.353 | 1.611 | 0.01063 | 0.01063 | 0.03843 | 177 |
| GOBP_REGULATION_OF_PHOSPHORUS_METABOLIC_PROCESS | 66 | 0.366 | 1.635 | 0.01069 | 0.01069 | 0.03849 | 149 |
| HP_ABNORMALITY_OF_SKELETAL_MATURATION | 26 | 0.468 | 1.723 | 0.01092 | 0.01092 | 0.03919 | 273 |
| HP_ABNORMAL_SOFT_PALATE_MORPHOLOGY | 28 | 0.456 | 1.708 | 0.01100 | 0.01100 | 0.03934 | 258 |
| GOCC_CHROMATIN | 65 | 0.365 | 1.628 | 0.01136 | 0.01136 | 0.04047 | 302 |
| GOBP_REGULATION_OF_CYTOSKELETON_ORGANIZATION | 35 | 0.433 | 1.708 | 0.01152 | 0.01152 | 0.04089 | 226 |
| GOBP_MRNA_TRANSPORT | 10 | -0.555 | -1.907 | 0.01166 | 0.01166 | 0.04126 | 318 |
| HP_HETEROGENEOUS | 10 | 0.615 | 1.730 | 0.01171 | 0.01171 | 0.04130 | 196 |
| GOBP_REGULATION_OF_NEUROGENESIS | 13 | 0.564 | 1.726 | 0.01205 | 0.01205 | 0.04234 | 173 |
| HP_ABNORMAL_INTERNAL_GENITALIA | 40 | 0.413 | 1.678 | 0.01213 | 0.01213 | 0.04238 | 277 |
| GOBP_RIBOSOME_BIOGENESIS | 18 | -0.415 | -1.815 | 0.01215 | 0.01215 | 0.04238 | 418 |
| HP_HEMATOLOGICAL_NEOPLASM | 29 | 0.449 | 1.695 | 0.01249 | 0.01249 | 0.04343 | 237 |
| GOMF_PROTEIN_SERINE_THREONINE_KINASE_ACTIVITY | 27 | 0.459 | 1.703 | 0.01261 | 0.01261 | 0.04370 | 197 |
| GOBP_KINETOCHORE_ORGANIZATION | 15 | 0.539 | 1.719 | 0.01295 | 0.01295 | 0.04474 | 161 |
| GOMF_ATP_HYDROLYSIS_ACTIVITY | 46 | 0.395 | 1.653 | 0.01306 | 0.01306 | 0.04495 | 280 |
| HP_RECURRENT_URINARY_TRACT_INFECTIONS | 16 | 0.529 | 1.712 | 0.01317 | 0.01317 | 0.04516 | 270 |
| GOCC_AZUROPHIL_GRANULE | 11 | -0.513 | -1.840 | 0.01321 | 0.01321 | 0.04516 | 79 |
| HP_ABNORMALITY_OF_THE_BREAST | 37 | 0.420 | 1.677 | 0.01343 | 0.01343 | 0.04575 | 216 |
| GOCC_RIBONUCLEOPROTEIN_COMPLEX | 41 | -0.291 | -1.685 | 0.01350 | 0.01350 | 0.04576 | 390 |
| HP_ABNORMAL_FOREARM_BONE_MORPHOLOGY | 13 | 0.560 | 1.714 | 0.01352 | 0.01352 | 0.04576 | 233 |
| HP_MYELODYSPLASIA | 20 | 0.495 | 1.703 | 0.01372 | 0.01372 | 0.04625 | 233 |

| | | | | | | | |
|---|---|---|---|---|---|---|---|
| HP_ABNORMALITY_OF_THE_KIDNEY | 75 | 0.349 | 1.594 | 0.01376 | 0.01376 | 0.04625 | 297 |
| HP_ABNORMALITY_OF_THE_TESTIS_SIZE | 15 | 0.535 | 1.708 | 0.01413 | 0.01413 | 0.04735 | 253 |
| GOBP_CENTROMERE_COMPLEX_ASSEMBLY | 16 | 0.525 | 1.700 | 0.01434 | 0.01434 | 0.04782 | 243 |
| HP_PATENT_DUCTUS_ARTERIOSUS | 19 | 0.505 | 1.711 | 0.01437 | 0.01437 | 0.04782 | 297 |
| GOBP_REGULATION_OF_CELL_POPULATION_PROLIFERATION | 64 | 0.360 | 1.600 | 0.01444 | 0.01444 | 0.04790 | 188 |
| HP_INFANTILE_ONSET | 19 | -0.394 | -1.757 | 0.01451 | 0.01451 | 0.04796 | 122 |
| GOCC_TRANSPORT_VESICLE | 10 | -0.540 | -1.856 | 0.01484 | 0.01484 | 0.04891 | 129 |
| HP_AGENESIS_OF_CORPUS_CALLOSUM | 25 | 0.462 | 1.681 | 0.01500 | 0.01500 | 0.04928 | 258 |
| GOBP_CELLULAR_MACROMOLECULE_BIOSYNTHETIC_PROCESS | 111 | 0.319 | 1.529 | 0.01525 | 0.01525 | 0.04980 | 256 |
| HP_ANAL_ATRESIA | 13 | 0.554 | 1.693 | 0.01526 | 0.01526 | 0.04980 | 233 |
| GOBP_REGULATION_OF_PEPTIDASE_ACTIVITY | 12 | -0.494 | -1.833 | 0.01531 | 0.01531 | 0.04980 | 19 |
| GOBP_CELLULAR_RESPONSE_TO_STRESS | 151 | 0.300 | 1.481 | 0.01536 | 0.01536 | 0.04980 | 240 |
| GOCC_CHROMOSOME_TELOMERIC_REGION | 24 | 0.467 | 1.685 | 0.01593 | 0.01593 | 0.05149 | 233 |
| GOBP_REGULATION_OF_HYDROLASE_ACTIVITY | 36 | -0.294 | -1.644 | 0.01634 | 0.01634 | 0.05247 | 134 |
| GOBP_RESPONSE_TO_CYTOKINE | 36 | -0.292 | -1.636 | 0.01634 | 0.01634 | 0.05247 | 124 |
| GOMF_CHROMATIN_BINDING | 57 | 0.371 | 1.614 | 0.01646 | 0.01646 | 0.05270 | 268 |
| GOMF_CALMODULIN_BINDING | 12 | -0.488 | -1.810 | 0.01685 | 0.01685 | 0.05376 | 124 |
| HP_APLASIA_HYPOPLASIA_OF_THE_ULNA | 11 | 0.578 | 1.677 | 0.01704 | 0.01704 | 0.05421 | 233 |
| GOBP_NUCLEAR_EXPORT | 10 | -0.534 | -1.835 | 0.01731 | 0.01731 | 0.05491 | 270 |
| GOBP_SMALL_GTPASE_MEDIATED_SIGNAL_TRANSDUCTION | 24 | 0.464 | 1.675 | 0.01752 | 0.01752 | 0.05540 | 138 |
| HP_ABNORMAL_CONNECTION_OF_THE_CARDIAC_SEGMENTS | 15 | 0.527 | 1.682 | 0.01793 | 0.01793 | 0.05649 | 221 |
| GOBP_NEGATIVE_REGULATION_OF_HYDROLASE_ACTIVITY | 10 | -0.529 | -1.816 | 0.01802 | 0.01802 | 0.05655 | 19 |
| GOBP_REGULATION_OF_DOUBLE_STRAND_BREAK_REPAIR | 15 | 0.527 | 1.681 | 0.01806 | 0.01806 | 0.05655 | 213 |
| GOBP_NEGATIVE_REGULATION_OF_PHOSPHORUS_METABOLIC_PROCESS | 14 | 0.533 | 1.668 | 0.01826 | 0.01826 | 0.05700 | 149 |
| HP_ABNORMAL_CORTICAL_GYRATION | 29 | 0.435 | 1.643 | 0.01837 | 0.01837 | 0.05719 | 277 |
| HP_HYPERPIGMENTATION_OF_THE_SKIN | 35 | 0.416 | 1.641 | 0.01845 | 0.01845 | 0.05724 | 238 |
| HP_ABNORMALITY_OF_FLUID_REGULATION | 31 | 0.430 | 1.645 | 0.01866 | 0.01866 | 0.05772 | 99 |
| GOBP_NEGATIVE_REGULATION_OF_PROTEIN_MODIFICATION_PROCESS | 23 | 0.464 | 1.657 | 0.01885 | 0.01885 | 0.05812 | 149 |
| HP_UPPER_LIMB_UNDERGROWTH | 14 | 0.531 | 1.662 | 0.01918 | 0.01918 | 0.05897 | 258 |
| GOCC_REPLICATION_FORK | 24 | 0.461 | 1.664 | 0.01924 | 0.01924 | 0.05897 | 323 |
| GOBP_MAINTENANCE_OF_LOCATION_IN_CELL | 10 | -0.521 | -1.791 | 0.01943 | 0.01943 | 0.05899 | 124 |
| HP_BICORNUATE_UTERUS | 12 | 0.559 | 1.668 | 0.01962 | 0.01962 | 0.05899 | 221 |
| HP_ABNORMAL_EYELID_MORPHOLOGY | 84 | 0.332 | 1.538 | 0.01966 | 0.01966 | 0.05899 | 255 |
| GOBP_RHYTHMIC_PROCESS | 13 | 0.542 | 1.658 | 0.01968 | 0.01968 | 0.05899 | 191 |
| GOBP_FOREBRAIN_DEVELOPMENT | 16 | 0.514 | 1.666 | 0.01968 | 0.01968 | 0.05899 | 207 |
| GOMF_SIGNALING_RECEPTOR_BINDING | 29 | -0.320 | -1.664 | 0.01970 | 0.01970 | 0.05899 | 65 |
| GOBP_PROTEIN_DEPOLYMERIZATION | 11 | 0.573 | 1.662 | 0.01979 | 0.01979 | 0.05899 | 164 |
| GOCC_MITOTIC_SPINDLE_POLE | 10 | 0.588 | 1.655 | 0.01980 | 0.01980 | 0.05899 | 197 |

| | | | | | | | |
|---|---|---|---|---|---|---|---|
| GOBP_DEVELOPMENTAL_GROWTH | 23 | 0.463 | 1.653 | 0.01983 | 0.01983 | 0.05899 | 157 |
| HP_ABNORMAL_MORPHOLOGY_OF_THE_GREAT_VESSELS | 37 | 0.408 | 1.630 | 0.01985 | 0.01985 | 0.05899 | 297 |
| GOBP_REGULATION_OF_DOUBLE_STRAND_BREAK_REPAIR_VIA_HOMOLOGOUS_RECOMBINATION | 12 | 0.558 | 1.665 | 0.02003 | 0.02003 | 0.05899 | 213 |
| HP_ABNORMAL_DERMATOGLYPHICS | 18 | 0.495 | 1.652 | 0.02005 | 0.02005 | 0.05899 | 23 |
| HP_AZOOSPERMIA | 14 | 0.529 | 1.656 | 0.02011 | 0.02011 | 0.05899 | 253 |
| HP_ABNORMALITY_OF_THE_CHOANAE | 18 | 0.494 | 1.649 | 0.02018 | 0.02018 | 0.05899 | 233 |
| HP_ABNORMAL_ILEUM_MORPHOLOGY | 10 | 0.586 | 1.649 | 0.02036 | 0.02036 | 0.05899 | 221 |
| HP_ABNORMAL_PREPUTIUM_MORPHOLOGY | 10 | 0.586 | 1.649 | 0.02036 | 0.02036 | 0.05899 | 221 |
| HP_ABSENT_TESTIS | 10 | 0.586 | 1.649 | 0.02036 | 0.02036 | 0.05899 | 221 |
| HP_APLASIA_HYPOPLASIA_OF_THE_UVULA | 10 | 0.586 | 1.649 | 0.02036 | 0.02036 | 0.05899 | 221 |
| HP_PYRIDOXINE_RESPONSIVE_SIDEROBLASTIC_ANEMIA | 10 | 0.586 | 1.649 | 0.02036 | 0.02036 | 0.05899 | 221 |
| HP_HYPOREFLEXIA | 13 | -0.469 | -1.791 | 0.02053 | 0.02053 | 0.05932 | 379 |
| HP_POSTNATAL_GROWTH_RETARDATION | 19 | 0.490 | 1.661 | 0.02085 | 0.02085 | 0.06008 | 108 |
| HP_APLASIA_HYPOPLASIA_AFFECTING_BONES_OF_THE_AXIAL_SKELETON | 67 | 0.350 | 1.563 | 0.02101 | 0.02101 | 0.06035 | 247 |
| HP_MACULE | 29 | 0.431 | 1.627 | 0.02114 | 0.02114 | 0.06054 | 221 |
| GOBP_DNA_INTEGRITY_CHECKPOINT_SIGNALING | 32 | 0.420 | 1.622 | 0.02119 | 0.02119 | 0.06054 | 216 |
| GOBP_RESPONSE_TO_METAL_ION | 11 | -0.493 | -1.770 | 0.02128 | 0.02128 | 0.06055 | 144 |
| GOBP_CHROMOSOME_ORGANIZATION_INVOLVED_IN_MEIOTIC_CELL_CYCLE | 17 | 0.499 | 1.642 | 0.02138 | 0.02138 | 0.06055 | 257 |
| HP_SIMPLIFIED_GYRAL_PATTERN | 15 | 0.519 | 1.657 | 0.02146 | 0.02146 | 0.06055 | 277 |
| GOBP_ORGANOPHOSPHATE_METABOLIC_PROCESS | 38 | -0.288 | -1.636 | 0.02152 | 0.02152 | 0.06055 | 146 |
| GOBP_POSITIVE_REGULATION_OF_TRANSFERASE_ACTIVITY | 48 | 0.378 | 1.595 | 0.02152 | 0.02152 | 0.06055 | 225 |
| HP_ABNORMAL_LEUKOCYTE_COUNT | 23 | 0.461 | 1.646 | 0.02168 | 0.02168 | 0.06055 | 233 |
| HP_ABNORMALITY_OF_THE_EXTERNAL_NOSE | 49 | 0.376 | 1.589 | 0.02170 | 0.02170 | 0.06055 | 99 |
| GOBP_HOMOLOGOUS_RECOMBINATION | 11 | 0.567 | 1.645 | 0.02171 | 0.02171 | 0.06055 | 182 |
| HP_ABNORMALITY_OF_THE_HYPOTHALAMUS_PITUITARY_AXIS | 29 | 0.430 | 1.624 | 0.02174 | 0.02174 | 0.06055 | 258 |
| GOBP_LOCOMOTION | 42 | -0.279 | -1.617 | 0.02191 | 0.02191 | 0.06086 | 66 |
| HP_PROPORTIONATE_SHORT_STATURE | 14 | 0.526 | 1.647 | 0.02223 | 0.02223 | 0.06150 | 137 |
| GOBP_PEPTIDE_BIOSYNTHETIC_PROCESS | 24 | -0.353 | -1.705 | 0.02226 | 0.02226 | 0.06150 | 382 |
| GOBP_BLASTOCYST_DEVELOPMENT | 11 | 0.565 | 1.641 | 0.02240 | 0.02240 | 0.06155 | 157 |
| GOBP_REGULATION_OF_CELLULAR_AMIDE_METABOLIC_PROCESS | 19 | -0.381 | -1.699 | 0.02246 | 0.02246 | 0.06155 | 382 |
| HP_ARTERIOVENOUS_MALFORMATION | 12 | 0.552 | 1.648 | 0.02246 | 0.02246 | 0.06155 | 221 |
| HP_ABNORMALITY_OF_LONG_BONE_MORPHOLOGY | 48 | 0.377 | 1.588 | 0.02277 | 0.02277 | 0.06223 | 304 |
| GOCC_SM_LIKE_PROTEIN_FAMILY_COMPLEX | 10 | -0.514 | -1.767 | 0.02297 | 0.02297 | 0.06260 | 346 |
| GOCC_MITOCHONDRIAL_ENVELOPE | 36 | -0.285 | -1.594 | 0.02314 | 0.02314 | 0.06291 | 392 |
| HP_ABNORMALITY_OF_THE_URETER | 35 | 0.408 | 1.611 | 0.02327 | 0.02327 | 0.06307 | 255 |
| GOBP_SEX_DIFFERENTIATION | 12 | 0.550 | 1.641 | 0.02355 | 0.02355 | 0.06366 | 221 |
| GOBP_IMMUNE_SYSTEM_DEVELOPMENT | 33 | 0.413 | 1.608 | 0.02383 | 0.02383 | 0.06427 | 150 |
| GOBP_REGULATION_OF_CELL_DEVELOPMENT | 20 | 0.476 | 1.637 | 0.02391 | 0.02391 | 0.06431 | 173 |

| | | | | | | | |
|---|---|---|---|---|---|---|---|
| GOBP_PROTEIN_LOCALIZATION_TO_CHROMOSOME | 24 | 0.450 | 1.625 | 0.02426 | 0.02426 | 0.06491 | 205 |
| GOMF_KINASE_BINDING | 50 | 0.372 | 1.579 | 0.02427 | 0.02427 | 0.06491 | 143 |
| HP_IRREGULAR_HYPERPIGMENTATION | 31 | 0.422 | 1.615 | 0.02464 | 0.02464 | 0.06574 | 238 |
| HP_WIDE_NOSE | 11 | 0.560 | 1.626 | 0.02487 | 0.02487 | 0.06618 | 99 |
| GOBP_REGULATION_OF_MULTICELLULAR_ORGANISMAL_DEVELOPMENT | 30 | 0.422 | 1.604 | 0.02566 | 0.02566 | 0.06809 | 173 |
| HP_ABNORMALITY_OF_THE_PERIORBITAL_REGION | 10 | 0.573 | 1.612 | 0.02649 | 0.02649 | 0.07012 | 96 |
| GOBP_BIOLOGICAL_ADHESION | 25 | -0.333 | -1.631 | 0.02662 | 0.02662 | 0.07028 | 102 |
| HP_APLASIA_HYPOPLASIA_INVOLVING_BONES_OF_THE_THORAX | 10 | 0.573 | 1.611 | 0.02677 | 0.02677 | 0.07030 | 142 |
| GOBP_POSITIVE_REGULATION_OF_CELL_CYCLE_G2_M_PHASE_TRANSITION | 10 | 0.572 | 1.611 | 0.02677 | 0.02677 | 0.07030 | 188 |
| HP_ABNORMAL_CARDIAC_ATRIUM_MORPHOLOGY | 40 | 0.388 | 1.578 | 0.02716 | 0.02716 | 0.07113 | 297 |
| HP_ABNORMAL_IMMUNE_SYSTEM_MORPHOLOGY | 38 | 0.392 | 1.574 | 0.02916 | 0.02916 | 0.07584 | 237 |
| HP_ABNORMALITY_OF_THE_MITOCHONDRION | 16 | -0.395 | -1.645 | 0.02917 | 0.02917 | 0.07584 | 431 |
| GOBP_RRNA_METABOLIC_PROCESS | 15 | -0.413 | -1.670 | 0.02924 | 0.02924 | 0.07584 | 418 |
| GOCC_NEURON_PROJECTION | 29 | -0.307 | -1.593 | 0.02925 | 0.02925 | 0.07584 | 130 |
| GOMF_MRNA_BINDING | 17 | -0.385 | -1.644 | 0.02935 | 0.02935 | 0.07588 | 270 |
| HP_ABNORMAL_VASCULAR_MORPHOLOGY | 43 | 0.378 | 1.561 | 0.02954 | 0.02954 | 0.07605 | 309 |
| HP_ABNORMALITY_OF_THE_UTERUS | 23 | 0.450 | 1.606 | 0.02956 | 0.02956 | 0.07605 | 221 |
| GOCC_U2_TYPE_SPLICEOSOMAL_COMPLEX | 10 | -0.500 | -1.718 | 0.03004 | 0.03004 | 0.07705 | 356 |
| HP_NEOPLASM_BY_HISTOLOGY | 23 | 0.449 | 1.604 | 0.03018 | 0.03018 | 0.07705 | 233 |
| HP_SIDEROBLASTIC_ANEMIA | 11 | 0.551 | 1.600 | 0.03023 | 0.03023 | 0.07705 | 221 |
| HP_NEOPLASM_OF_THE_STOMACH | 10 | 0.566 | 1.592 | 0.03026 | 0.03026 | 0.07705 | 99 |
| GOBP_GLYCEROLIPID_METABOLIC_PROCESS | 12 | -0.455 | -1.689 | 0.03063 | 0.03063 | 0.07780 | 137 |
| GOBP_GROWTH | 37 | 0.392 | 1.564 | 0.03083 | 0.03083 | 0.07811 | 191 |
| GOBP_SMALL_MOLECULE_METABOLIC_PROCESS | 62 | -0.223 | -1.444 | 0.03108 | 0.03108 | 0.07854 | 146 |
| HP_ABNORMAL_MALE_REPRODUCTIVE_SYSTEM_PHYSIOLOGY | 22 | 0.451 | 1.592 | 0.03163 | 0.03163 | 0.07932 | 270 |
| HP_ABNORMAL_URETER_PHYSIOLOGY | 21 | 0.459 | 1.599 | 0.03168 | 0.03168 | 0.07932 | 270 |
| GOBP_ESTABLISHMENT_OF_RNA_LOCALIZATION | 12 | -0.455 | -1.686 | 0.03178 | 0.03178 | 0.07932 | 318 |
| GOBP_NUCLEOBASE_CONTAINING_COMPOUND_TRANSPORT | 12 | -0.455 | -1.686 | 0.03178 | 0.03178 | 0.07932 | 318 |
| GOBP_RNA_LOCALIZATION | 15 | -0.409 | -1.652 | 0.03178 | 0.03178 | 0.07932 | 318 |
| HP_ABNORMALITY_OF_THE_OUTER_EAR | 84 | 0.320 | 1.480 | 0.03216 | 0.03216 | 0.08008 | 250 |
| GOBP_HOMOLOGOUS_CHROMOSOME_SEGREGATION | 12 | 0.535 | 1.596 | 0.03248 | 0.03248 | 0.08053 | 182 |
| GOBP_EPITHELIUM_DEVELOPMENT | 32 | 0.405 | 1.562 | 0.03250 | 0.03250 | 0.08053 | 191 |
| GOMF_KINASE_REGULATOR_ACTIVITY | 16 | 0.494 | 1.600 | 0.03272 | 0.03272 | 0.08087 | 225 |
| GOBP_REGULATION_OF_DNA_METABOLIC_PROCESS | 51 | 0.361 | 1.536 | 0.03288 | 0.03288 | 0.08094 | 306 |
| HP_ABNORMAL_CARDIAC_SEPTUM_MORPHOLOGY | 47 | 0.365 | 1.534 | 0.03291 | 0.03291 | 0.08094 | 304 |
| HP_CRANIOSYNOSTOSIS | 17 | 0.484 | 1.591 | 0.03316 | 0.03316 | 0.08137 | 247 |
| HP_FOCAL_ONSET_SEIZURE | 10 | -0.490 | -1.683 | 0.03357 | 0.03357 | 0.08216 | 363 |
| HP_TOE_SYNDACTYLY | 23 | 0.443 | 1.583 | 0.03424 | 0.03424 | 0.08362 | 221 |

| | | | | | | | |
|---|---|---|---|---|---|---|---|
| HP_TETRALOGY_OF_FALLOT | 14 | 0.508 | 1.589 | 0.03440 | 0.03440 | 0.08378 | 221 |
| GOBP_DNA_STRAND_ELONGATION | 11 | 0.542 | 1.574 | 0.03463 | 0.03463 | 0.08415 | 293 |
| HP_CONTRACTURES_OF_THE_JOINTS_OF_THE_UPPER_LIMBS | 15 | 0.496 | 1.582 | 0.03507 | 0.03507 | 0.08501 | 142 |
| HP_HYDROURETER | 11 | 0.541 | 1.571 | 0.03573 | 0.03573 | 0.08640 | 221 |
| HP_ABNORMAL_UVEA_MORPHOLOGY | 27 | 0.421 | 1.560 | 0.03589 | 0.03589 | 0.08641 | 221 |
| GOBP_NEGATIVE_REGULATION_OF_CATALYTIC_ACTIVITY | 28 | 0.416 | 1.558 | 0.03590 | 0.03590 | 0.08641 | 149 |
| HP_ABNORMALITY_OF_SKIN_PIGMENTATION | 43 | 0.372 | 1.533 | 0.03630 | 0.03630 | 0.08715 | 238 |
| GOBP_CELL_SURFACE_RECEPTOR_SIGNALING_PATHWAY_INVOLVED_IN_CELL_CELL_SIGNALING | 14 | -0.417 | -1.642 | 0.03643 | 0.03643 | 0.08726 | 137 |
| GOCC_CELL_DIVISION_SITE | 10 | 0.556 | 1.564 | 0.03667 | 0.03667 | 0.08755 | 135 |
| HP_DOWNSLANTED_PALPEBRAL_FISSURES | 31 | 0.404 | 1.548 | 0.03672 | 0.03672 | 0.08755 | 99 |
| HP_GASTROINTESTINAL_HEMORRHAGE | 11 | 0.540 | 1.567 | 0.03683 | 0.03683 | 0.08759 | 96 |
| GOBP_CELL_MIGRATION | 40 | -0.262 | -1.504 | 0.03781 | 0.03781 | 0.08950 | 66 |
| GOBP_REGULATION_OF_ENDOCYTOSIS | 10 | -0.484 | -1.663 | 0.03781 | 0.03781 | 0.08950 | 183 |
| GOBP_POSITIVE_REGULATION_OF_ORGANELLE_ORGANIZATION | 34 | 0.391 | 1.532 | 0.03911 | 0.03911 | 0.09236 | 204 |
| HP_ABNORMAL_HEART_VALVE_MORPHOLOGY | 22 | 0.442 | 1.561 | 0.03932 | 0.03932 | 0.09264 | 238 |
| GOMF_ENZYME_REGULATOR_ACTIVITY | 54 | 0.351 | 1.511 | 0.03977 | 0.03977 | 0.09348 | 167 |
| GOBP_REGULATION_OF_BODY_FLUID_LEVELS | 11 | -0.459 | -1.645 | 0.04000 | 0.04000 | 0.09374 | 155 |
| HP_TRIANGULAR_SHAPED_PHALANGES_OF_THE_HAND | 22 | 0.441 | 1.557 | 0.04006 | 0.04006 | 0.09374 | 221 |
| GOBP_MITOTIC_DNA_INTEGRITY_CHECKPOINT_SIGNALING | 21 | 0.449 | 1.564 | 0.04044 | 0.04044 | 0.09440 | 216 |
| HP_ABNORMAL_RENAL_MORPHOLOGY | 64 | 0.333 | 1.481 | 0.04058 | 0.04058 | 0.09451 | 182 |
| HP_NEOPLASM_OF_THE_GASTROINTESTINAL_TRACT | 22 | 0.440 | 1.554 | 0.04118 | 0.04118 | 0.09569 | 216 |
| GOBP_NEGATIVE_REGULATION_OF_CELL_CYCLE_G2_M_PHASE_TRANSITION | 19 | 0.460 | 1.558 | 0.04145 | 0.04145 | 0.09609 | 216 |
| GOCC_ENDOSOME_MEMBRANE | 16 | -0.375 | -1.565 | 0.04247 | 0.04247 | 0.09813 | 249 |
| GOBP_KINETOCHORE_ASSEMBLY | 10 | 0.549 | 1.544 | 0.04253 | 0.04253 | 0.09813 | 161 |
| GOBP_MICROTUBULE_ORGANIZING_CENTER_ORGANIZATION | 31 | 0.399 | 1.527 | 0.04270 | 0.04270 | 0.09832 | 255 |
| GOCC_HETEROCHROMATIN | 11 | 0.532 | 1.545 | 0.04287 | 0.04287 | 0.09849 | 236 |
| HP_ABNORMAL_SKIN_MORPHOLOGY_OF_THE_PALM | 20 | 0.451 | 1.549 | 0.04330 | 0.04330 | 0.09911 | 23 |
| HP_LOCALIZED_SKIN_LESION | 54 | 0.349 | 1.500 | 0.04334 | 0.04334 | 0.09911 | 221 |
| HP_JOINT_HYPERMOBILITY | 26 | 0.418 | 1.540 | 0.04356 | 0.04356 | 0.09938 | 292 |
| HP_ABNORMAL_CIRCULATING_HORMONE_CONCENTRATION | 17 | 0.473 | 1.555 | 0.04379 | 0.04379 | 0.09968 | 56 |
| GOBP_REGULATION_OF_BINDING | 16 | 0.481 | 1.556 | 0.04393 | 0.04393 | 0.09978 | 143 |
| HP_CORNEAL_OPACITY | 16 | 0.480 | 1.554 | 0.04419 | 0.04419 | 0.09998 | 99 |
| GOBP_RESPONSE_TO_PEPTIDE | 17 | -0.363 | -1.549 | 0.04424 | 0.04424 | 0.09998 | 155 |
| GOBP_INTRACELLULAR_PROTEIN_TRANSPORT | 32 | -0.279 | -1.505 | 0.04432 | 0.04432 | 0.09998 | 108 |
| GOBP_APOPTOTIC_SIGNALING_PATHWAY | 35 | -0.271 | -1.500 | 0.04491 | 0.04491 | 0.10108 | 96 |
| HP_ABNORMAL_UMBILICUS_MORPHOLOGY | 18 | 0.463 | 1.545 | 0.04551 | 0.04551 | 0.10219 | 270 |
| GOBP_ENZYME_LINKED_RECEPTOR_PROTEIN_SIGNALING_PATHWAY | 24 | -0.316 | -1.525 | 0.04560 | 0.04560 | 0.10219 | 214 |
| GOBP_POSITIVE_REGULATION_OF_CELL_POPULATION_PROLIFERATION | 36 | 0.378 | 1.502 | 0.04606 | 0.04606 | 0.10298 | 188 |

| | | | | | | | |
|---|---|---|---|---|---|---|---|
| GOBP_NERVOUS_SYSTEM_PROCESS | 20 | -0.338 | -1.531 | 0.04667 | 0.04667 | 0.10378 | 124 |
| HP_GENITAL_NEOPLASM | 14 | 0.492 | 1.540 | 0.04670 | 0.04670 | 0.10378 | 196 |
| HP_ABNORMALITY_OF_FEMUR_MORPHOLOGY | 22 | 0.436 | 1.540 | 0.04676 | 0.04676 | 0.10378 | 321 |
| GOMF_KINASE_ACTIVITY | 43 | 0.362 | 1.495 | 0.04683 | 0.04683 | 0.10378 | 203 |
| HP_ABNORMAL_ORAL_MUCOSA_MORPHOLOGY | 16 | 0.477 | 1.546 | 0.04693 | 0.04693 | 0.10378 | 56 |
| HP_ABNORMALITY_OF_THE_ULNA | 19 | 0.453 | 1.533 | 0.04755 | 0.04755 | 0.10487 | 233 |
| GOBP_CELLULAR_COMPONENT_DISASSEMBLY | 22 | 0.436 | 1.537 | 0.04763 | 0.04763 | 0.10487 | 164 |
| GOBP_GOLGI_VESICLE_TRANSPORT | 12 | -0.427 | -1.585 | 0.04786 | 0.04786 | 0.10514 | 407 |
| HP_LEUKOPENIA | 15 | 0.483 | 1.542 | 0.04815 | 0.04815 | 0.10545 | 221 |
| HP_ABNORMAL_FACIAL_SHAPE | 62 | 0.331 | 1.461 | 0.04824 | 0.04824 | 0.10545 | 258 |
| HP_ABNORMAL_JAW_MORPHOLOGY | 77 | 0.314 | 1.435 | 0.04838 | 0.04838 | 0.10545 | 247 |
| HP_LISSENCEPHALY | 15 | 0.483 | 1.540 | 0.04842 | 0.04842 | 0.10545 | 266 |
| GOBP_PROTEIN_LOCALIZATION_TO_CELL_PERIPHERY | 12 | -0.427 | -1.583 | 0.04862 | 0.04862 | 0.10554 | 108 |
| HP_DECREASED_FERTILITY | 15 | 0.482 | 1.538 | 0.04868 | 0.04868 | 0.10554 | 221 |
| HP_NEOPLASM_OF_THE_BREAST | 12 | 0.514 | 1.534 | 0.04885 | 0.04885 | 0.10554 | 216 |
| HP_SHORT_PALPEBRAL_FISSURE | 16 | 0.476 | 1.541 | 0.04889 | 0.04889 | 0.10554 | 221 |
| HP_ABNORMALITY_OF_CRANIAL_SUTURES | 18 | 0.460 | 1.533 | 0.04898 | 0.04898 | 0.10554 | 247 |
| HP_CLUBBING | 14 | 0.490 | 1.534 | 0.04935 | 0.04935 | 0.10610 | 238 |
| GOBP_MITOTIC_G2_M_TRANSITION_CHECKPOINT | 17 | 0.467 | 1.536 | 0.05040 | 0.05040 | 0.10813 | 216 |
| GOBP_INTRINSIC_APOPTOTIC_SIGNALING_PATHWAY_BY_P53_CLASS_MEDIATOR | 11 | -0.442 | -1.585 | 0.05064 | 0.05064 | 0.10843 | 205 |
| HP_HYDROCEPHALUS | 27 | 0.406 | 1.506 | 0.05080 | 0.05080 | 0.10849 | 233 |
| HP_ABNORMALITY_OF_THE_CALVARIA | 65 | 0.326 | 1.451 | 0.05100 | 0.05100 | 0.10849 | 266 |
| GOBP_REGULATION_OF_ESTABLISHMENT_OF_PROTEIN_LOCALIZATION | 22 | -0.322 | -1.514 | 0.05103 | 0.05103 | 0.10849 | 147 |
| HP_BIFID_UVULA | 16 | 0.473 | 1.531 | 0.05110 | 0.05110 | 0.10849 | 258 |
| GOCC_MITOCHONDRIAL_PROTEIN_CONTAINING_COMPLEX | 17 | -0.356 | -1.520 | 0.05125 | 0.05125 | 0.10857 | 392 |
| GOBP_SIGNAL_TRANSDUCTION_BY_P53_CLASS_MEDIATOR | 21 | -0.326 | -1.515 | 0.05161 | 0.05161 | 0.10911 | 15 |
| HP_ABNORMAL_CAROTID_ARTERY_MORPHOLOGY | 12 | 0.510 | 1.524 | 0.05223 | 0.05223 | 0.11019 | 321 |
| HP_ABNORMALITY_OF_CARDIOVASCULAR_SYSTEM_ELECTROPHYSIOLOGY | 16 | 0.472 | 1.528 | 0.05241 | 0.05241 | 0.11020 | 96 |
| HP_CEREBRAL_HYPOPLASIA | 11 | 0.521 | 1.512 | 0.05249 | 0.05249 | 0.11020 | 255 |
| GOBP_TRANSMEMBRANE_RECEPTOR_PROTEIN_TYROSINE_KINASE_SIGNALING_PATHWAY | 17 | -0.354 | -1.512 | 0.05256 | 0.05256 | 0.11020 | 129 |
| HP_ABNORMAL_SYSTEMIC_ARTERIAL_MORPHOLOGY | 30 | 0.393 | 1.496 | 0.05287 | 0.05287 | 0.11062 | 321 |
| GOBP_REGULATION_OF_DNA_REPAIR | 22 | 0.431 | 1.521 | 0.05309 | 0.05309 | 0.11084 | 213 |
| GOBP_PURINE_CONTAINING_COMPOUND_METABOLIC_PROCESS | 19 | -0.339 | -1.510 | 0.05335 | 0.05335 | 0.11115 | 146 |
| GOMF_HELICASE_ACTIVITY | 24 | 0.420 | 1.516 | 0.05355 | 0.05355 | 0.11135 | 272 |
| GOBP_REGULATION_OF_PROTEOLYSIS | 28 | -0.288 | -1.477 | 0.05434 | 0.05434 | 0.11274 | 115 |
| GOMF_MOLECULAR_TRANSDUCER_ACTIVITY | 13 | -0.408 | -1.559 | 0.05488 | 0.05488 | 0.11363 | 124 |
| GOBP_SPINDLE_LOCALIZATION | 14 | 0.485 | 1.518 | 0.05517 | 0.05517 | 0.11382 | 163 |
| HP_ABNORMAL_LIP_MORPHOLOGY | 50 | 0.344 | 1.459 | 0.05519 | 0.05519 | 0.11382 | 303 |

| | | | | | | | |
|---|---|---|---|---|---|---|---|
| GOCC_ENDOPLASMIC_RETICULUM | 52 | -0.229 | -1.421 | 0.05535 | 0.05535 | 0.11382 | 109 |
| GOBP_CELLULAR_RESPONSE_TO_EXTERNAL_STIMULUS | 20 | -0.330 | -1.497 | 0.05542 | 0.05542 | 0.11382 | 96 |
| HP_ABNORMAL_CEREBRAL_VENTRICLE_MORPHOLOGY | 62 | 0.327 | 1.444 | 0.05593 | 0.05593 | 0.11463 | 237 |
| GOBP_RESPONSE_TO_UV | 18 | 0.453 | 1.513 | 0.05605 | 0.05605 | 0.11465 | 249 |
| GOMF_TRANSFERASE_ACTIVITY_TRANSFERRING_PHOSPHORUS_CONTAINING_GROUPS | 50 | 0.343 | 1.456 | 0.05621 | 0.05621 | 0.11474 | 203 |
| HP_ABNORMAL_URETER_MORPHOLOGY | 15 | 0.475 | 1.517 | 0.05653 | 0.05653 | 0.11516 | 221 |
| GOCC_ORGANELLE_INNER_MEMBRANE | 28 | -0.286 | -1.464 | 0.05723 | 0.05723 | 0.11635 | 436 |
| GOBP_TELENCEPHALON_DEVELOPMENT | 14 | 0.482 | 1.509 | 0.05794 | 0.05794 | 0.11757 | 207 |
| HP_SPINA_BIFIDA | 14 | 0.482 | 1.508 | 0.05861 | 0.05861 | 0.11867 | 270 |
| HP_APLASIA_HYPOPLASIA_INVOLVING_FOREARM_BONES | 15 | 0.473 | 1.510 | 0.05889 | 0.05889 | 0.11876 | 233 |
| HP_APLASIA_HYPOPLASIA_OF_THE_RADIUS | 15 | 0.473 | 1.510 | 0.05889 | 0.05889 | 0.11876 | 233 |
| GOCC_DENDRITIC_TREE | 16 | -0.359 | -1.498 | 0.05963 | 0.05963 | 0.11979 | 175 |
| GOBP_CYTOKINE_MEDIATED_SIGNALING_PATHWAY | 16 | -0.359 | -1.495 | 0.05963 | 0.05963 | 0.11979 | 120 |
| HP_ABNORMAL_NEURAL_TUBE_MORPHOLOGY | 16 | 0.465 | 1.505 | 0.05984 | 0.05984 | 0.11996 | 270 |
| GOBP_PROTEIN_CONTAINING_COMPLEX_DISASSEMBLY | 14 | 0.480 | 1.502 | 0.06112 | 0.06112 | 0.12229 | 164 |
| GOBP_NEUROGENESIS | 44 | 0.350 | 1.453 | 0.06185 | 0.06185 | 0.12351 | 184 |
| HP_ORAL_CLEFT | 55 | 0.334 | 1.444 | 0.06215 | 0.06215 | 0.12386 | 258 |
| GOBP_REGULATION_OF_CELLULAR_COMPONENT_MOVEMENT | 24 | -0.301 | -1.456 | 0.06243 | 0.06243 | 0.12407 | 102 |
| GOBP_POSITIVE_REGULATION_OF_SIGNALING | 55 | -0.222 | -1.387 | 0.06250 | 0.06250 | 0.12407 | 137 |
| GOBP_RESPONSE_TO_WOUNDING | 14 | -0.392 | -1.541 | 0.06263 | 0.06263 | 0.12408 | 205 |
| GOBP_RAS_PROTEIN_SIGNAL_TRANSDUCTION | 18 | 0.447 | 1.493 | 0.06363 | 0.06363 | 0.12582 | 138 |
| GOBP_NUCLEOSOME_ASSEMBLY | 10 | 0.526 | 1.481 | 0.06470 | 0.06470 | 0.12767 | 314 |
| HP_APLASIA_HYPOPLASIA_INVOLVING_THE_SKELETON | 80 | 0.303 | 1.394 | 0.06485 | 0.06485 | 0.12772 | 297 |
| GOBP_NEURAL_PRECURSOR_CELL_PROLIFERATION | 10 | 0.526 | 1.480 | 0.06511 | 0.06511 | 0.12800 | 173 |
| HP_ABDOMINAL_PAIN | 13 | 0.486 | 1.485 | 0.06601 | 0.06601 | 0.12950 | 228 |
| HP_ABNORMAL_CORNEA_MORPHOLOGY | 42 | 0.350 | 1.438 | 0.06706 | 0.06706 | 0.13130 | 238 |
| HP_URINARY_TRACT_NEOPLASM | 11 | 0.509 | 1.479 | 0.06734 | 0.06734 | 0.13159 | 216 |
| HP_ABNORMALITY_OF_FACIAL_SKELETON | 81 | 0.301 | 1.387 | 0.06768 | 0.06768 | 0.13202 | 247 |
| GOBP_DEFENSE_RESPONSE_TO_SYMBIONT | 13 | 0.484 | 1.480 | 0.06788 | 0.06788 | 0.13215 | 124 |
| GOBP_CELLULAR_RESPONSE_TO_PEPTIDE | 15 | -0.368 | -1.490 | 0.06822 | 0.06822 | 0.13255 | 155 |
| HP_RENAL_CYST | 13 | 0.484 | 1.479 | 0.06842 | 0.06842 | 0.13264 | 99 |
| GOBP_NEGATIVE_REGULATION_OF_PROTEOLYSIS | 10 | -0.437 | -1.502 | 0.06855 | 0.06855 | 0.13264 | 19 |
| GOBP_PROTEIN_CONTAINING_COMPLEX_ORGANIZATION | 106 | 0.287 | 1.371 | 0.06877 | 0.06877 | 0.13264 | 128 |
| HP_RENAL_AGENESIS | 17 | 0.450 | 1.480 | 0.06879 | 0.06879 | 0.13264 | 255 |
| GOBP_MULTICELLULAR_ORGANISMAL_HOMEOSTASIS | 13 | -0.395 | -1.508 | 0.06948 | 0.06948 | 0.13372 | 124 |
| GOBP_RESPONSE_TO_PEPTIDE_HORMONE | 13 | -0.394 | -1.507 | 0.07067 | 0.07067 | 0.13571 | 155 |
| HP_ABNORMAL_LOWER_LIMB_BONE_MORPHOLOGY | 52 | 0.332 | 1.419 | 0.07079 | 0.07079 | 0.13571 | 304 |
| GOBP_POSTTRANSCRIPTIONAL_REGULATION_OF_GENE_EXPRESSION | 22 | -0.307 | -1.444 | 0.07216 | 0.07216 | 0.13809 | 383 |

| | | | | | | | |
|---|---|---|---|---|---|---|---|
| HP_ABNORMALITY_OF_THE_LOWER_URINARY_TRACT | 56 | 0.328 | 1.420 | 0.07266 | 0.07266 | 0.13878 | 270 |
| HP_CRYPTORCHIDISM | 49 | 0.334 | 1.414 | 0.07312 | 0.07312 | 0.13939 | 286 |
| GOBP_REGULATION_OF_DNA_RECOMBINATION | 19 | 0.431 | 1.461 | 0.07349 | 0.07349 | 0.13983 | 216 |
| GOBP_MORPHOGENESIS_OF_AN_EPITHELIUM | 13 | 0.480 | 1.468 | 0.07377 | 0.07377 | 0.14010 | 184 |
| GOBP_INFLAMMATORY_RESPONSE | 19 | -0.325 | -1.448 | 0.07394 | 0.07394 | 0.14015 | 144 |
| GOBP_PROTEASOME_MEDIATED_UBIQUITIN_DEPENDENT_PROTEIN_CATABOLIC_PROCESS | 26 | 0.395 | 1.455 | 0.07413 | 0.07413 | 0.14026 | 107 |
| HP_LIMB_UNDERGROWTH | 23 | 0.408 | 1.458 | 0.07428 | 0.07428 | 0.14027 | 393 |
| GOBP_MITOTIC_G2_DNA_DAMAGE_CHECKPOINT_SIGNALING | 14 | 0.468 | 1.465 | 0.07541 | 0.07541 | 0.14213 | 216 |
| GOBP_CHROMATIN_ASSEMBLY_OR_DISASSEMBLY | 18 | 0.437 | 1.460 | 0.07572 | 0.07572 | 0.14245 | 323 |
| HP_ABNORMAL_INTESTINE_MORPHOLOGY | 51 | 0.331 | 1.410 | 0.07624 | 0.07624 | 0.14317 | 242 |
| GOCC_TRANSFERASE_COMPLEX_TRANSFERRING_PHOSPHORUS_CONTAINING_GROUPS | 21 | 0.421 | 1.466 | 0.07675 | 0.07675 | 0.14386 | 316 |
| GOBP_RESPONSE_TO_GROWTH_FACTOR | 24 | -0.294 | -1.420 | 0.07763 | 0.07763 | 0.14524 | 128 |
| GOCC_CULLIN_RING_UBIQUITIN_LIGASE_COMPLEX | 10 | 0.517 | 1.455 | 0.07794 | 0.07794 | 0.14555 | 91 |
| HP_ABNORMAL_AORTIC_MORPHOLOGY | 26 | 0.393 | 1.445 | 0.07814 | 0.07814 | 0.14565 | 297 |
| HP_ABNORMAL_PATELLA_MORPHOLOGY | 13 | 0.477 | 1.459 | 0.07832 | 0.07832 | 0.14573 | 286 |
| GOBP_RESPONSE_TO_EXTRACELLULAR_STIMULUS | 18 | -0.328 | -1.436 | 0.07872 | 0.07872 | 0.14601 | 124 |
| HP_ABNORMAL_AORTIC_VALVE_MORPHOLOGY | 17 | 0.443 | 1.458 | 0.07877 | 0.07877 | 0.14601 | 292 |
| HP_WIDE_NASAL_BRIDGE | 26 | 0.392 | 1.441 | 0.08032 | 0.08032 | 0.14862 | 89 |
| GOBP_TRNA_METABOLIC_PROCESS | 12 | -0.395 | -1.466 | 0.08078 | 0.08078 | 0.14920 | 429 |
| GOBP_PROTEIN_LOCALIZATION_TO_CHROMOSOME_CENTROMERIC_REGION | 14 | 0.464 | 1.454 | 0.08110 | 0.08110 | 0.14950 | 317 |
| GOCC_SIDE_OF_MEMBRANE | 12 | -0.394 | -1.461 | 0.08193 | 0.08193 | 0.15047 | 124 |
| HP_EXTERNAL_GENITAL_HYPOPLASIA | 36 | 0.356 | 1.415 | 0.08215 | 0.08215 | 0.15047 | 304 |
| HP_APLASIA_HYPOPLASIA_INVOLVING_BONES_OF_THE_LOWER_LIMBS | 28 | 0.381 | 1.426 | 0.08221 | 0.08221 | 0.15047 | 297 |
| GOMF_ZINC_ION_BINDING | 33 | -0.252 | -1.374 | 0.08222 | 0.08222 | 0.15047 | 45 |
| HP_DIARRHEA | 14 | 0.463 | 1.450 | 0.08295 | 0.08295 | 0.15153 | 238 |
| HP_ABNORMAL_LUNG_DEVELOPMENT | 14 | 0.463 | 1.448 | 0.08348 | 0.08348 | 0.15222 | 178 |
| GOBP_REGULATION_OF_INTRACELLULAR_SIGNAL_TRANSDUCTION | 70 | 0.304 | 1.373 | 0.08389 | 0.08389 | 0.15271 | 157 |
| HP_HEADACHE | 13 | 0.472 | 1.445 | 0.08435 | 0.08435 | 0.15326 | 96 |
| HP_ABNORMALITY_OF_BONE_MINERAL_DENSITY | 25 | 0.392 | 1.428 | 0.08464 | 0.08464 | 0.15347 | 237 |
| GOBP_CARBOHYDRATE_DERIVATIVE_METABOLIC_PROCESS | 41 | -0.238 | -1.377 | 0.08477 | 0.08477 | 0.15347 | 159 |
| HP_THICK_VERMILION_BORDER | 16 | 0.447 | 1.449 | 0.08539 | 0.08539 | 0.15427 | 292 |
| HP_ABNORMAL_SPERMATOGENESIS | 16 | 0.447 | 1.448 | 0.08552 | 0.08552 | 0.15427 | 221 |
| GOBP_MAINTENANCE_OF_LOCATION | 12 | -0.390 | -1.448 | 0.08576 | 0.08576 | 0.15436 | 124 |
| GOBP_CELLULAR_RESPONSE_TO_PEPTIDE_HORMONE_STIMULUS | 11 | -0.406 | -1.458 | 0.08587 | 0.08587 | 0.15436 | 155 |
| HP_MORTALITY_AGING | 11 | 0.496 | 1.439 | 0.08602 | 0.08602 | 0.15436 | 63 |
| HP_ABNORMALITY_OF_DIGESTIVE_SYSTEM_MORPHOLOGY | 57 | 0.319 | 1.388 | 0.08632 | 0.08632 | 0.15461 | 242 |
| GOBP_POSITIVE_REGULATION_OF_DNA_BIOSYNTHETIC_PROCESS | 16 | 0.445 | 1.443 | 0.08747 | 0.08747 | 0.15640 | 200 |
| HP_ABNORMALITY_OF_FINGER | 75 | 0.297 | 1.355 | 0.08890 | 0.08890 | 0.15866 | 250 |

| | | | | | | | |
|---|---|---|---|---|---|---|---|
| GOBP_POSITIVE_REGULATION_OF_CHROMOSOME_ORGANIZATION | 13 | 0.469 | 1.435 | 0.08957 | 0.08957 | 0.15936 | 204 |
| HP_NEOPLASM_OF_THE_GENITOURINARY_TRACT | 18 | 0.429 | 1.432 | 0.08960 | 0.08960 | 0.15936 | 216 |
| GOBP_RIBOSE_PHOSPHATE_METABOLIC_PROCESS | 18 | -0.323 | -1.413 | 0.09042 | 0.09042 | 0.16015 | 146 |
| GOBP_GENERATION_OF_PRECURSOR_METABOLITES_AND_ENERGY | 16 | -0.338 | -1.409 | 0.09052 | 0.09052 | 0.16015 | 139 |
| HP_THIN_VERMILION_BORDER | 21 | 0.412 | 1.435 | 0.09052 | 0.09052 | 0.16015 | 173 |
| GOBP_RESPONSE_TO_VIRUS | 18 | 0.429 | 1.430 | 0.09076 | 0.09076 | 0.16028 | 124 |
| HP_LIMB_JOINT_CONTRACTURE | 18 | 0.428 | 1.429 | 0.09114 | 0.09114 | 0.16068 | 142 |
| GOMF_MAGNESIUM_ION_BINDING | 11 | 0.492 | 1.430 | 0.09166 | 0.09166 | 0.16131 | 246 |
| GOBP_DEVELOPMENT_OF_PRIMARY_SEXUAL_CHARACTERISTICS | 10 | 0.505 | 1.421 | 0.09230 | 0.09230 | 0.16208 | 221 |
| GOCC_PLASMA_MEMBRANE_REGION | 33 | -0.247 | -1.346 | 0.09242 | 0.09242 | 0.16208 | 124 |
| HP_CLUBBING_OF_TOES | 12 | 0.478 | 1.427 | 0.09283 | 0.09283 | 0.16252 | 221 |
| HP_ABNORMALITY_OF_THE_BLADDER | 24 | 0.392 | 1.415 | 0.09412 | 0.09412 | 0.16449 | 270 |
| HP_HYPOPLASIA_OF_THE_FRONTAL_LOBES | 10 | 0.504 | 1.416 | 0.09439 | 0.09439 | 0.16469 | 255 |
| HP_ABNORMALITY_OF_THE_FOREARM | 22 | 0.402 | 1.417 | 0.09613 | 0.09613 | 0.16728 | 237 |
| HP_ABNORMALITY_OF_EARLOBE | 12 | 0.476 | 1.420 | 0.09621 | 0.09621 | 0.16728 | 250 |
| HP_ABNORMALITY_OF_THE_ANUS | 17 | 0.431 | 1.419 | 0.09742 | 0.09742 | 0.16909 | 233 |
| GOCC_EARLY_ENDOSOME | 12 | -0.379 | -1.405 | 0.09763 | 0.09763 | 0.16916 | 249 |
| HP_PHENOTYPIC_VARIABILITY | 13 | 0.463 | 1.416 | 0.09867 | 0.09867 | 0.17068 | 138 |
| HP_RESPIRATORY_FAILURE | 11 | 0.488 | 1.416 | 0.09935 | 0.09935 | 0.17156 | 247 |
| HP_AMBIGUOUS_GENITALIA | 11 | 0.487 | 1.414 | 0.09977 | 0.09977 | 0.17177 | 99 |
| GOMF_MOLECULAR_FUNCTION_REGULATOR | 65 | 0.304 | 1.353 | 0.09981 | 0.09981 | 0.17177 | 167 |
| HP_ABNORMAL_UPPER_LIMB_BONE_MORPHOLOGY | 32 | 0.360 | 1.389 | 0.10000 | 0.10000 | 0.17179 | 297 |
| GOBP_TRNA_PROCESSING | 11 | -0.395 | -1.416 | 0.10055 | 0.10055 | 0.17229 | 429 |
| GOBP_REGULATION_OF_ANATOMICAL_STRUCTURE_MORPHOGENESIS | 20 | -0.303 | -1.375 | 0.10063 | 0.10063 | 0.17229 | 202 |
| GOBP_PROTEIN_TARGETING | 14 | -0.362 | -1.422 | 0.10111 | 0.10111 | 0.17281 | 108 |
| GOBP_ANATOMICAL_STRUCTURE_MATURATION | 10 | 0.498 | 1.402 | 0.10220 | 0.10220 | 0.17432 | 99 |
| HP_ABNORMALITY_OF_THE_ABDOMINAL_ORGANS | 57 | 0.312 | 1.354 | 0.10234 | 0.10234 | 0.17432 | 196 |
| GOMF_DAMAGED_DNA_BINDING | 13 | 0.461 | 1.408 | 0.10336 | 0.10336 | 0.17577 | 240 |
| HP_JOINT_CONTRACTURE_OF_THE_HAND | 14 | 0.450 | 1.407 | 0.10385 | 0.10385 | 0.17607 | 142 |
| GOBP_NUCLEOSOME_ORGANIZATION | 11 | 0.484 | 1.406 | 0.10389 | 0.10389 | 0.17607 | 314 |
| GOCC_CENTRIOLAR_SATELLITE | 14 | 0.450 | 1.407 | 0.10451 | 0.10451 | 0.17683 | 207 |
| HP_TRIPHALANGEAL_THUMB | 12 | 0.470 | 1.402 | 0.10514 | 0.10514 | 0.17760 | 221 |
| HP_ABNORMAL_HEART_VALVE_PHYSIOLOGY | 19 | 0.412 | 1.398 | 0.10553 | 0.10553 | 0.17784 | 99 |
| GOBP_PROTEASOMAL_PROTEIN_CATABOLIC_PROCESS | 29 | 0.365 | 1.378 | 0.10580 | 0.10580 | 0.17784 | 107 |
| HP_ABNORMALITY_OF_THE_CLITORIS | 11 | 0.483 | 1.401 | 0.10581 | 0.10581 | 0.17784 | 247 |
| HP_NEONATAL_HYPOTONIA | 11 | -0.392 | -1.406 | 0.10606 | 0.10606 | 0.17787 | 431 |
| GOBP_INTRINSIC_APOPTOTIC_SIGNALING_PATHWAY | 22 | -0.290 | -1.364 | 0.10619 | 0.10619 | 0.17787 | 41 |
| HP_ABNORMAL_NUMBER_OF_TEETH | 21 | 0.402 | 1.402 | 0.10655 | 0.10655 | 0.17818 | 297 |

| | | | | | | | |
|---|---|---|---|---|---|---|---|
| GOMF_ENZYME_ACTIVATOR_ACTIVITY | 26 | 0.376 | 1.386 | 0.10677 | 0.10677 | 0.17826 | 225 |
| HP_SYNOSTOSIS_INVOLVING_BONES_OF_THE_UPPER_LIMBS | 10 | 0.495 | 1.392 | 0.10750 | 0.10750 | 0.17918 | 10 |
| GOCC_CENTRIOLE | 22 | 0.395 | 1.394 | 0.10878 | 0.10878 | 0.18102 | 208 |
| HP_ABNORMALITY_OF_PELVIC_GIRDLE_BONE_MORPHOLOGY | 44 | 0.326 | 1.352 | 0.10944 | 0.10944 | 0.18181 | 297 |
| GOBP_REGULATION_OF_LIPID_METABOLIC_PROCESS | 10 | -0.405 | -1.391 | 0.11025 | 0.11025 | 0.18285 | 219 |
| GOCC_ENDOSOME | 24 | -0.281 | -1.356 | 0.11075 | 0.11075 | 0.18339 | 249 |
| HP_ABNORMALITY_OF_UPPER_LIP | 44 | 0.326 | 1.351 | 0.11126 | 0.11126 | 0.18393 | 303 |
| HP_ABNORMALITY_OF_THE_CEREBROSPINAL_FLUID | 33 | 0.351 | 1.365 | 0.11242 | 0.11242 | 0.18532 | 277 |
| HP_HYPERREFLEXIA | 37 | 0.340 | 1.360 | 0.11256 | 0.11256 | 0.18532 | 270 |
| HP_ABNORMALITY_OF_THE_CURVATURE_OF_THE_CORNEA | 19 | 0.409 | 1.385 | 0.11265 | 0.11265 | 0.18532 | 321 |
| HP_ABNORMALITY_OF_NEURONAL_MIGRATION | 32 | 0.354 | 1.367 | 0.11321 | 0.11321 | 0.18573 | 277 |
| HP_ABNORMALITY_OF_THE_GINGIVA | 13 | 0.455 | 1.391 | 0.11327 | 0.11327 | 0.18573 | 56 |
| HP_THIN_UPPER_LIP_VERMILION | 18 | 0.415 | 1.384 | 0.11351 | 0.11351 | 0.18582 | 173 |
| HP_ABNORMALITY_OF_UPPER_LIMB_JOINT | 25 | 0.377 | 1.375 | 0.11440 | 0.11440 | 0.18698 | 155 |
| GOCC_NUCLEOLUS | 68 | 0.296 | 1.327 | 0.11502 | 0.11502 | 0.18746 | 251 |
| HP_FUNCTIONAL_ABNORMALITY_OF_MALE_INTERNAL_GENITALIA | 19 | 0.407 | 1.380 | 0.11507 | 0.11507 | 0.18746 | 270 |
| HP_ABNORMAL_PHALANGEAL_JOINT_MORPHOLOGY_OF_THE_HAND | 14 | 0.444 | 1.388 | 0.11536 | 0.11536 | 0.18763 | 142 |
| HP_ABNORMAL_SHAPE_OF_THE_FRONTAL_REGION | 31 | 0.355 | 1.357 | 0.11699 | 0.11699 | 0.18979 | 247 |
| HP_DYSMETRIA | 11 | -0.385 | -1.380 | 0.11706 | 0.11706 | 0.18979 | 436 |
| HP_ABNORMAL_ABDOMEN_MORPHOLOGY | 32 | 0.352 | 1.359 | 0.11726 | 0.11726 | 0.18981 | 99 |
| GOBP_POSITIVE_REGULATION_OF_CELL_DIFFERENTIATION | 26 | 0.371 | 1.366 | 0.11757 | 0.11757 | 0.19000 | 173 |
| HP_ABNORMALITY_OF_THE_PINNA | 49 | 0.316 | 1.339 | 0.11799 | 0.11799 | 0.19038 | 270 |
| HP_ASTIGMATISM | 16 | 0.427 | 1.383 | 0.11824 | 0.11824 | 0.19047 | 221 |
| HP_ABNORMALITY_OF_THE_LYMPHATIC_SYSTEM | 26 | 0.371 | 1.364 | 0.11878 | 0.11878 | 0.19104 | 159 |
| HP_AGANGLIONIC_MEGACOLON | 13 | 0.450 | 1.377 | 0.11983 | 0.11983 | 0.19193 | 321 |
| HP_DOLICHOCEPHALY | 18 | 0.412 | 1.374 | 0.11994 | 0.11994 | 0.19193 | 221 |
| GOBP_RESPONSE_TO_INORGANIC_SUBSTANCE | 23 | -0.283 | -1.345 | 0.11996 | 0.11996 | 0.19193 | 50 |
| GOBP_NEGATIVE_REGULATION_OF_CELL_DIFFERENTIATION | 13 | 0.450 | 1.375 | 0.12010 | 0.12010 | 0.19193 | 143 |
| GOBP_PROTEIN_LOCALIZATION_TO_CONDENSED_CHROMOSOME | 13 | 0.449 | 1.374 | 0.12117 | 0.12117 | 0.19325 | 317 |
| GOCC_CONDENSED_NUCLEAR_CHROMOSOME | 15 | 0.432 | 1.379 | 0.12130 | 0.12130 | 0.19325 | 404 |
| GOBP_CELLULAR_RESPONSE_TO_RADIATION | 22 | 0.388 | 1.369 | 0.12292 | 0.12292 | 0.19530 | 238 |
| HP_HYPOPIGMENTED_SKIN_PATCHES | 17 | 0.416 | 1.370 | 0.12307 | 0.12307 | 0.19530 | 221 |
| GOBP_PALLIUM_DEVELOPMENT | 13 | 0.448 | 1.371 | 0.12318 | 0.12318 | 0.19530 | 207 |
| GOBP_ESTABLISHMENT_OF_MITOTIC_SPINDLE_LOCALIZATION | 13 | 0.448 | 1.370 | 0.12358 | 0.12358 | 0.19531 | 163 |
| HP_MICROPENIS | 16 | 0.423 | 1.371 | 0.12371 | 0.12371 | 0.19531 | 304 |
| HP_POSTERIORLY_ROTATED_EARS | 28 | 0.362 | 1.353 | 0.12379 | 0.12379 | 0.19531 | 142 |
| HP_NEOPLASM_OF_THE_NERVOUS_SYSTEM | 10 | 0.487 | 1.370 | 0.12395 | 0.12395 | 0.19531 | 209 |
| HP_ABNORMALITY_OF_THE_ELBOW | 16 | 0.423 | 1.370 | 0.12476 | 0.12476 | 0.19627 | 273 |

| | | | | | | | |
|---|---|---|---|---|---|---|---|
| GOBP_TAXIS | 11 | -0.380 | -1.364 | 0.12550 | 0.12550 | 0.19694 | 102 |
| GOBP_CELLULAR_RESPONSE_TO_INORGANIC_SUBSTANCE | 12 | -0.364 | -1.351 | 0.12557 | 0.12557 | 0.19694 | 16 |
| GOBP_CHROMATIN_ORGANIZATION | 34 | 0.342 | 1.339 | 0.12595 | 0.12595 | 0.19721 | 196 |
| HP_APLASIA_HYPOPLASIA_OF_THE_EAR | 14 | 0.438 | 1.370 | 0.12727 | 0.12727 | 0.19872 | 247 |
| HP_BREAST_CARCINOMA | 10 | 0.485 | 1.363 | 0.12730 | 0.12730 | 0.19872 | 196 |
| GOBP_POSITIVE_REGULATION_OF_CATALYTIC_ACTIVITY | 68 | 0.291 | 1.306 | 0.12805 | 0.12805 | 0.19959 | 225 |
| HP_ABNORMALITY_OF_REPRODUCTIVE_SYSTEM_PHYSIOLOGY | 35 | 0.339 | 1.339 | 0.12832 | 0.12832 | 0.19969 | 270 |
| GOCC_GOLGI_APPARATUS | 53 | -0.209 | -1.296 | 0.13088 | 0.13088 | 0.20323 | 90 |
| HP_CLEFT_SOFT_PALATE | 18 | 0.405 | 1.352 | 0.13099 | 0.13099 | 0.20323 | 258 |
| HP_SPLENOMEGALY | 11 | -0.377 | -1.351 | 0.13138 | 0.13138 | 0.20333 | 145 |
| GOMF_NUCLEOSIDE_TRIPHOSPHATASE_REGULATOR_ACTIVITY | 21 | 0.390 | 1.358 | 0.13146 | 0.13146 | 0.20333 | 131 |
| GOBP_RESPONSE_TO_OXYGEN_LEVELS | 14 | -0.341 | -1.340 | 0.13181 | 0.13181 | 0.20355 | 179 |
| GOBP_LIPID_CATABOLIC_PROCESS | 10 | -0.392 | -1.348 | 0.13216 | 0.13216 | 0.20377 | 131 |
| HP_HIRSUTISM | 10 | 0.482 | 1.355 | 0.13330 | 0.13330 | 0.20522 | 23 |
| GOMF_HYDROLASE_ACTIVITY_ACTING_ON_ACID_ANHYDRIDES | 58 | 0.300 | 1.305 | 0.13384 | 0.13384 | 0.20575 | 293 |
| GOMF_ENZYME_INHIBITOR_ACTIVITY | 14 | 0.434 | 1.357 | 0.13507 | 0.13507 | 0.20709 | 149 |
| HP_ABNORMAL_PENIS_MORPHOLOGY | 48 | 0.312 | 1.314 | 0.13515 | 0.13515 | 0.20709 | 304 |
| GOBP_REGULATION_OF_CELL_DIFFERENTIATION | 37 | 0.331 | 1.324 | 0.13533 | 0.13533 | 0.20709 | 173 |
| GOBP_REGULATION_OF_GROWTH | 22 | 0.382 | 1.349 | 0.13570 | 0.13570 | 0.20733 | 79 |
| HP_INABILITY_TO_WALK | 14 | -0.339 | -1.333 | 0.13631 | 0.13631 | 0.20795 | 361 |
| GOBP_REGULATION_OF_SMALL_GTPASE_MEDIATED_SIGNAL_TRANSDUCTION | 15 | 0.423 | 1.350 | 0.13727 | 0.13727 | 0.20910 | 138 |
| GOBP_NEGATIVE_REGULATION_OF_RESPONSE_TO_EXTERNAL_STIMULUS | 13 | 0.440 | 1.344 | 0.13790 | 0.13790 | 0.20959 | 26 |
| HP_ABNORMAL_SIZE_OF_THE_PALPEBRAL_FISSURES | 30 | 0.349 | 1.328 | 0.13811 | 0.13811 | 0.20959 | 286 |
| HP_ABNORMAL_THROMBOCYTE_MORPHOLOGY | 23 | 0.376 | 1.343 | 0.13821 | 0.13821 | 0.20959 | 237 |
| GOBP_CELL_PART_MORPHOGENESIS | 16 | 0.416 | 1.348 | 0.13870 | 0.13870 | 0.21002 | 121 |
| GOBP_POSITIVE_REGULATION_OF_DNA_METABOLIC_PROCESS | 36 | 0.332 | 1.319 | 0.13993 | 0.13993 | 0.21155 | 213 |
| GOBP_CEREBRAL_CORTEX_DEVELOPMENT | 12 | 0.452 | 1.349 | 0.14019 | 0.14019 | 0.21161 | 79 |
| HP_ABNORMALITY_OF_THE_PALM | 25 | 0.367 | 1.337 | 0.14038 | 0.14038 | 0.21161 | 23 |
| GOBP_CELLULAR_NITROGEN_COMPOUND_CATABOLIC_PROCESS | 22 | -0.277 | -1.302 | 0.14124 | 0.14124 | 0.21258 | 36 |
| HP_ABNORMALITY_OF_THE_AMNIOTIC_FLUID | 35 | 0.334 | 1.319 | 0.14160 | 0.14160 | 0.21280 | 196 |
| HP_EPICANTHUS | 40 | 0.322 | 1.309 | 0.14457 | 0.14457 | 0.21695 | 233 |
| GOBP_POSITIVE_REGULATION_OF_CELL_CYCLE_G1_S_PHASE_TRANSITION | 11 | 0.460 | 1.335 | 0.14525 | 0.14525 | 0.21765 | 263 |
| HP_ABNORMALITY_OF_THE_RIBS | 12 | 0.448 | 1.337 | 0.14614 | 0.14614 | 0.21837 | 247 |
| HP_SPECIFIC_LEARNING_DISABILITY | 15 | 0.419 | 1.337 | 0.14617 | 0.14617 | 0.21837 | 247 |
| GOBP_REGULATION_OF_DNA_BIOSYNTHETIC_PROCESS | 20 | 0.387 | 1.329 | 0.14663 | 0.14663 | 0.21874 | 200 |
| GOBP_REGULATION_OF_PROTEIN_CATABOLIC_PROCESS | 20 | 0.386 | 1.328 | 0.14701 | 0.14701 | 0.21898 | 126 |
| GOBP_NEGATIVE_REGULATION_OF_DEVELOPMENTAL_PROCESS | 16 | 0.412 | 1.333 | 0.14809 | 0.14809 | 0.22026 | 143 |
| GOBP_MALE_GAMETE_GENERATION | 27 | 0.355 | 1.315 | 0.14961 | 0.14961 | 0.22220 | 182 |

| | | | | | | | |
|---|---|---|---|---|---|---|---|
| HP_OCULAR_ANTERIOR_SEGMENT_DYSGENESIS | 22 | 0.376 | 1.327 | 0.15009 | 0.15009 | 0.22257 | 237 |
| HP_ABNORMAL_EXTERNAL_GENITALIA | 75 | 0.278 | 1.270 | 0.15038 | 0.15038 | 0.22267 | 258 |
| HP_ABNORMALITY_OF_GLOBE_LOCATION | 65 | 0.288 | 1.283 | 0.15060 | 0.15060 | 0.22267 | 196 |
| HP_ABNORMAL_LOCALIZATION_OF_KIDNEY | 20 | 0.384 | 1.321 | 0.15217 | 0.15217 | 0.22467 | 297 |
| HP_ABNORMALITY_OF_BLOOD_CIRCULATION | 16 | 0.410 | 1.327 | 0.15304 | 0.15304 | 0.22551 | 96 |
| HP_NARROW_PALATE | 10 | 0.471 | 1.326 | 0.15351 | 0.15351 | 0.22551 | 247 |
| GOBP_CELLULAR_RESPONSE_TO_LIGHT_STIMULUS | 11 | 0.456 | 1.322 | 0.15363 | 0.15363 | 0.22551 | 238 |
| GOBP_CELLULAR_RESPONSE_TO_UV | 11 | 0.456 | 1.322 | 0.15363 | 0.15363 | 0.22551 | 238 |
| GOBP_EMBRYONIC_ORGAN_DEVELOPMENT | 10 | 0.471 | 1.324 | 0.15491 | 0.15491 | 0.22705 | 146 |
| HP_ABNORMALITY_OF_THE_FONTANELLES_OR_CRANIAL_SUTURES | 21 | 0.377 | 1.315 | 0.15650 | 0.15650 | 0.22906 | 247 |
| GOBP_CENTROSOME_SEPARATION | 10 | 0.468 | 1.317 | 0.15979 | 0.15979 | 0.23353 | 53 |
| HP_ABNORMALITY_OF_THE_SPINAL_CORD | 20 | 0.380 | 1.307 | 0.16111 | 0.16111 | 0.23460 | 293 |
| HP_UNUSUAL_INFECTION | 44 | 0.309 | 1.282 | 0.16113 | 0.16113 | 0.23460 | 297 |
| GOBP_LIPID_LOCALIZATION | 10 | -0.378 | -1.298 | 0.16148 | 0.16148 | 0.23460 | 113 |
| HP_HYPOPLASIA_OF_PENIS | 28 | 0.347 | 1.298 | 0.16163 | 0.16163 | 0.23460 | 304 |
| GOBP_POSITIVE_REGULATION_OF_CELLULAR_COMPONENT_ORGANIZATION | 52 | 0.298 | 1.274 | 0.16169 | 0.16169 | 0.23460 | 204 |
| HP_VENTRICULOMEGALY | 55 | 0.294 | 1.269 | 0.16235 | 0.16235 | 0.23467 | 266 |
| GOBP_RESPONSE_TO_RADIATION | 42 | 0.313 | 1.286 | 0.16243 | 0.16243 | 0.23467 | 249 |
| GOBP_CELL_MORPHOGENESIS_INVOLVED_IN_NEURON_DIFFERENTIATION | 11 | 0.452 | 1.311 | 0.16243 | 0.16243 | 0.23467 | 95 |
| GOBP_DEVELOPMENTAL_MATURATION | 14 | 0.420 | 1.315 | 0.16312 | 0.16312 | 0.23532 | 99 |
| GOBP_STRESS_ACTIVATED_PROTEIN_KINASE_SIGNALING_CASCADE | 13 | 0.427 | 1.307 | 0.16388 | 0.16388 | 0.23608 | 96 |
| HP_ABNORMAL_SIZE_OF_THE_CLITORIS | 10 | 0.465 | 1.308 | 0.16662 | 0.16662 | 0.23969 | 247 |
| GOMF_PEPTIDASE_ACTIVITY | 22 | -0.270 | -1.271 | 0.16701 | 0.16701 | 0.23991 | 190 |
| GOCC_NUCLEAR_MEMBRANE | 18 | -0.291 | -1.272 | 0.16779 | 0.16779 | 0.24069 | 139 |
| HP_CHOREA | 12 | -0.344 | -1.276 | 0.16845 | 0.16845 | 0.24130 | 363 |
| HP_ABNORMAL_FINGER_PHALANX_MORPHOLOGY | 51 | 0.297 | 1.266 | 0.16869 | 0.16869 | 0.24130 | 258 |
| GOBP_REGULATION_OF_INTRACELLULAR_PROTEIN_TRANSPORT | 13 | -0.332 | -1.268 | 0.16936 | 0.16936 | 0.24191 | 147 |
| HP_ABNORMAL_TESTIS_MORPHOLOGY | 57 | 0.291 | 1.264 | 0.16974 | 0.16974 | 0.24211 | 303 |
| GOBP_CHROMATIN_REMODELING | 15 | 0.408 | 1.301 | 0.17090 | 0.17090 | 0.24341 | 243 |
| GOCC_VESICLE_LUMEN | 13 | -0.331 | -1.265 | 0.17134 | 0.17134 | 0.24370 | 79 |
| HP_DECREASED_FERTILITY_IN_MALES | 13 | 0.424 | 1.297 | 0.17191 | 0.17191 | 0.24390 | 221 |
| GOBP_CILIUM_ORGANIZATION | 15 | -0.316 | -1.277 | 0.17203 | 0.17203 | 0.24390 | 21 |
| GOBP_CELL_MORPHOGENESIS_INVOLVED_IN_DIFFERENTIATION | 16 | 0.402 | 1.301 | 0.17221 | 0.17221 | 0.24390 | 138 |
| GOBP_RESPONSE_TO_ALCOHOL | 11 | 0.447 | 1.296 | 0.17274 | 0.17274 | 0.24431 | 216 |
| HP_ABNORMAL_CARDIAC_VENTRICLE_MORPHOLOGY | 37 | 0.318 | 1.270 | 0.17433 | 0.17433 | 0.24622 | 309 |
| HP_ABNORMALITY_OF_THE_KNEE | 22 | 0.366 | 1.292 | 0.17477 | 0.17477 | 0.24649 | 286 |
| HP_ABNORMALITY_OF_THE_MUSCULATURE_OF_THE_LIMBS | 16 | -0.303 | -1.262 | 0.17503 | 0.17503 | 0.24652 | 494 |
| HP_FACIAL_ASYMMETRY | 14 | 0.414 | 1.296 | 0.17661 | 0.17661 | 0.24824 | 221 |

| | | | | | | | |
|---|---|---|---|---|---|---|---|
| GOBP_POSITIVE_REGULATION_OF_CELL_DEVELOPMENT | 14 | 0.414 | 1.295 | 0.17701 | 0.17701 | 0.24824 | 173 |
| GOBP_POSITIVE_REGULATION_OF_DEVELOPMENTAL_PROCESS | 42 | 0.309 | 1.269 | 0.17721 | 0.17721 | 0.24824 | 196 |
| HP_OLIGOHYDRAMNIOS | 21 | 0.369 | 1.286 | 0.17741 | 0.17741 | 0.24824 | 242 |
| GOBP_POSITIVE_REGULATION_OF_RESPONSE_TO_EXTERNAL_STIMULUS | 16 | -0.301 | -1.256 | 0.17761 | 0.17761 | 0.24824 | 167 |
| HP_ABNORMAL_VERTEBRAL_MORPHOLOGY | 17 | 0.392 | 1.288 | 0.17774 | 0.17774 | 0.24824 | 297 |
| HP_ABNORMAL_CRANIAL_NERVE_PHYSIOLOGY | 18 | 0.385 | 1.286 | 0.17817 | 0.17817 | 0.24824 | 221 |
| HP_REGIONAL_ABNORMALITY_OF_SKIN | 27 | 0.345 | 1.280 | 0.17823 | 0.17823 | 0.24824 | 23 |
| HP_ABNORMALITY_OF_THE_PITUITARY_GLAND | 19 | 0.381 | 1.290 | 0.17877 | 0.17877 | 0.24865 | 253 |
| HP_ABNORMAL_BLEEDING | 17 | 0.391 | 1.287 | 0.17917 | 0.17917 | 0.24887 | 96 |
| HP_ABNORMALITY_OF_LOWER_LIMB_JOINT | 46 | 0.301 | 1.258 | 0.18045 | 0.18045 | 0.25027 | 297 |
| GOBP_CELL_GROWTH | 19 | 0.380 | 1.286 | 0.18067 | 0.18067 | 0.25027 | 79 |
| HP_INCREASED_BODY_WEIGHT | 17 | 0.390 | 1.283 | 0.18137 | 0.18137 | 0.25089 | 22 |
| GOCC_TRANSFERASE_COMPLEX | 48 | 0.298 | 1.257 | 0.18183 | 0.18183 | 0.25118 | 256 |
| HP_ABNORMAL_FEMALE_REPRODUCTIVE_SYSTEM_PHYSIOLOGY | 10 | 0.456 | 1.282 | 0.18307 | 0.18307 | 0.25252 | 56 |
| HP_ABNORMALITY_IRIS_MORPHOLOGY | 21 | 0.367 | 1.277 | 0.18330 | 0.18330 | 0.25252 | 270 |
| HP_ABNORMALITY_OF_THE_PHALANGES_OF_THE_TOES | 14 | 0.411 | 1.286 | 0.18428 | 0.18428 | 0.25353 | 258 |
| GOBP_RESPONSE_TO_NITROGEN_COMPOUND | 42 | -0.211 | -1.223 | 0.18623 | 0.18623 | 0.25585 | 129 |
| GOCC_NUCLEAR_PERIPHERY | 16 | 0.394 | 1.277 | 0.18694 | 0.18694 | 0.25620 | 285 |
| GOBP_NEURON_DEVELOPMENT | 23 | 0.357 | 1.276 | 0.18699 | 0.18699 | 0.25620 | 142 |
| HP_LEUKEMIA | 15 | 0.401 | 1.279 | 0.18830 | 0.18830 | 0.25765 | 237 |
| GOBP_LIPID_BIOSYNTHETIC_PROCESS | 26 | -0.245 | -1.217 | 0.18864 | 0.18864 | 0.25776 | 137 |
| GOBP_REGULATION_OF_PROTEASOMAL_UBIQUITIN_DEPENDENT_PROTEIN_CATABOLIC_PROCESS | 11 | 0.438 | 1.273 | 0.18923 | 0.18923 | 0.25821 | 107 |
| GOBP_EPITHELIAL_CELL_DIFFERENTIATION | 17 | 0.387 | 1.274 | 0.19031 | 0.19031 | 0.25934 | 209 |
| GOBP_PROTEIN_DEPHOSPHORYLATION | 13 | 0.416 | 1.272 | 0.19092 | 0.19092 | 0.25951 | 96 |
| GOBP_ACTIN_FILAMENT_BASED_PROCESS | 29 | 0.334 | 1.262 | 0.19095 | 0.19095 | 0.25951 | 138 |
| HP_HYPOGONADISM | 23 | 0.356 | 1.272 | 0.19130 | 0.19130 | 0.25964 | 270 |
| GOBP_POSITIVE_REGULATION_OF_PROTEOLYSIS | 16 | -0.297 | -1.240 | 0.19176 | 0.19176 | 0.25992 | 115 |
| GOMF_SEQUENCE_SPECIFIC_DNA_BINDING | 51 | 0.291 | 1.240 | 0.19279 | 0.19279 | 0.26096 | 238 |
| HP_APLASIA_HYPOPLASIA_AFFECTING_THE_ANTERIOR_SEGMENT_OF_THE_EYE | 15 | 0.398 | 1.271 | 0.19419 | 0.19419 | 0.26250 | 221 |
| HP_ABNORMAL_RIB_CAGE_MORPHOLOGY | 14 | 0.406 | 1.272 | 0.19526 | 0.19526 | 0.26360 | 297 |
| GOCC_CELL_BODY | 16 | -0.296 | -1.234 | 0.19691 | 0.19691 | 0.26547 | 149 |
| GOCC_NUCLEAR_BODY | 54 | 0.286 | 1.233 | 0.19772 | 0.19772 | 0.26620 | 225 |
| HP_DISPLACEMENT_OF_THE_URETHRAL_MEATUS | 34 | 0.319 | 1.248 | 0.19849 | 0.19849 | 0.26644 | 221 |
| HP_ABNORMALITY_OF_THE_AUTONOMIC_NERVOUS_SYSTEM | 16 | 0.390 | 1.262 | 0.19867 | 0.19867 | 0.26644 | 321 |
| HP_NEOPLASM_OF_THE_LARGE_INTESTINE | 13 | 0.412 | 1.261 | 0.19869 | 0.19869 | 0.26644 | 216 |
| GOBP_PROTEIN_AUTOPHOSPHORYLATION | 13 | 0.412 | 1.259 | 0.20043 | 0.20043 | 0.26841 | 78 |
| GOBP_REGULATION_OF_NEURON_PROJECTION_DEVELOPMENT | 10 | 0.449 | 1.263 | 0.20106 | 0.20106 | 0.26890 | 142 |
| HP_INCREASED_BLOOD_PRESSURE | 13 | 0.411 | 1.257 | 0.20137 | 0.20137 | 0.26896 | 99 |

| | | | | | | | |
|---|---|---|---|---|---|---|---|
| HP_ABNORMAL_LOCATION_OF_EARS | 54 | 0.285 | 1.229 | 0.20197 | 0.20197 | 0.26940 | 142 |
| GOBP_PROTEIN_MONOUBIQUITINATION | 10 | 0.447 | 1.257 | 0.20399 | 0.20399 | 0.27174 | 240 |
| HP_ABNORMALITY_OF_THE_CALF | 11 | 0.431 | 1.252 | 0.20599 | 0.20599 | 0.27405 | 270 |
| HP_JOINT_DISLOCATION | 33 | 0.318 | 1.238 | 0.20693 | 0.20693 | 0.27459 | 247 |
| GOBP_POSITIVE_REGULATION_OF_HYDROLASE_ACTIVITY | 21 | -0.262 | -1.215 | 0.20695 | 0.20695 | 0.27459 | 134 |
| GOBP_REGULATION_OF_RESPONSE_TO_DNA_DAMAGE_STIMULUS | 32 | 0.320 | 1.235 | 0.20762 | 0.20762 | 0.27512 | 213 |
| GOBP_MODIFICATION_DEPENDENT_MACROMOLECULE_CATABOLIC_PROCESS | 37 | 0.310 | 1.237 | 0.20796 | 0.20796 | 0.27522 | 107 |
| GOBP_CELLULAR_RESPONSE_TO_NITROGEN_COMPOUND | 31 | -0.225 | -1.199 | 0.20828 | 0.20828 | 0.27528 | 129 |
| GOBP_CELL_PROJECTION_ASSEMBLY | 23 | -0.256 | -1.216 | 0.20966 | 0.20966 | 0.27674 | 193 |
| HP_ABNORMALITY_OF_THE_ABDOMINAL_WALL | 28 | 0.331 | 1.240 | 0.21132 | 0.21132 | 0.27806 | 270 |
| HP_UPPER_EXTREMITY_JOINT_DISLOCATION | 11 | 0.429 | 1.244 | 0.21135 | 0.21135 | 0.27806 | 247 |
| HP_ABNORMALITY_OF_ACID_BASE_HOMEOSTASIS | 16 | -0.293 | -1.220 | 0.21150 | 0.21150 | 0.27806 | 355 |
| HP_ABNORMAL_PALATE_MORPHOLOGY | 64 | 0.273 | 1.212 | 0.21177 | 0.21177 | 0.27806 | 247 |
| GOBP_NEGATIVE_REGULATION_OF_CELL_POPULATION_PROLIFERATION | 23 | 0.349 | 1.245 | 0.21212 | 0.21212 | 0.27816 | 263 |
| HP_SPARSE_SCALP_HAIR | 10 | 0.443 | 1.245 | 0.21319 | 0.21319 | 0.27895 | 233 |
| GOBP_AGING | 16 | 0.384 | 1.243 | 0.21327 | 0.21327 | 0.27895 | 197 |
| HP_ABNORMALITY_OF_THE_DIAPHRAGM | 10 | 0.442 | 1.244 | 0.21403 | 0.21403 | 0.27947 | 395 |
| GOCC_UBIQUITIN_LIGASE_COMPLEX | 20 | 0.361 | 1.242 | 0.21498 | 0.21498 | 0.27947 | 240 |
| HP_APRAXIA | 13 | -0.314 | -1.201 | 0.21516 | 0.21516 | 0.27947 | 485 |
| HP_ABNORMALITY_OF_FEMALE_EXTERNAL_GENITALIA | 16 | 0.383 | 1.242 | 0.21523 | 0.21523 | 0.27947 | 211 |
| GOMF_TRANSITION_METAL_ION_BINDING | 41 | -0.204 | -1.180 | 0.21530 | 0.21530 | 0.27947 | 45 |
| GOBP_CELLULAR_COMPONENT_MORPHOGENESIS | 21 | 0.355 | 1.238 | 0.21560 | 0.21560 | 0.27947 | 134 |
| HP_NEOPLASM_OF_THE_PANCREAS | 11 | 0.427 | 1.238 | 0.21561 | 0.21561 | 0.27947 | 196 |
| GOBP_POSITIVE_REGULATION_OF_MAPK_CASCADE | 15 | -0.302 | -1.219 | 0.21653 | 0.21653 | 0.28029 | 134 |
| HP_ABNORMAL_TENDON_MORPHOLOGY | 43 | 0.296 | 1.223 | 0.21802 | 0.21802 | 0.28187 | 162 |
| GOBP_RESPONSE_TO_LIGHT_STIMULUS | 22 | 0.350 | 1.234 | 0.21918 | 0.21918 | 0.28300 | 249 |
| HP_ABNORMALITY_OF_THE_METACARPAL_BONES | 15 | 0.387 | 1.236 | 0.22350 | 0.22350 | 0.28821 | 237 |
| HP_ABNORMALITY_OF_THE_FOREHEAD | 73 | 0.264 | 1.200 | 0.22459 | 0.22459 | 0.28925 | 247 |
| GOBP_HISTONE_MODIFICATION | 28 | 0.327 | 1.225 | 0.22498 | 0.22498 | 0.28938 | 197 |
| HP_REDUCED_VISUAL_ACUITY | 19 | 0.363 | 1.228 | 0.22568 | 0.22568 | 0.28957 | 96 |
| GOBP_POSITIVE_REGULATION_OF_MULTICELLULAR_ORGANISMAL_PROCESS | 42 | 0.295 | 1.213 | 0.22570 | 0.22570 | 0.28957 | 196 |
| GOBP_ENDOMEMBRANE_SYSTEM_ORGANIZATION | 25 | -0.243 | -1.192 | 0.22684 | 0.22684 | 0.29067 | 137 |
| GOMF_GTPASE_ACTIVATOR_ACTIVITY | 15 | 0.386 | 1.231 | 0.22795 | 0.22795 | 0.29171 | 131 |
| GOBP_MITOCHONDRION_ORGANIZATION | 28 | -0.230 | -1.177 | 0.22832 | 0.22832 | 0.29182 | 448 |
| GOBP_PURINE_CONTAINING_COMPOUND_BIOSYNTHETIC_PROCESS | 16 | -0.287 | -1.195 | 0.22909 | 0.22909 | 0.29242 | 146 |
| GOBP_ESTABLISHMENT_OF_SPINDLE_ORIENTATION | 12 | 0.412 | 1.230 | 0.22950 | 0.22950 | 0.29255 | 163 |
| GOBP_CELLULAR_RESPONSE_TO_EXTRACELLULAR_STIMULUS | 13 | -0.310 | -1.186 | 0.22977 | 0.22977 | 0.29255 | 96 |
| HP_ABNORMALITY_OF_THE_NAIL | 23 | 0.343 | 1.223 | 0.23035 | 0.23035 | 0.29293 | 287 |

| | | | | | | | |
|---|---|---|---|---|---|---|---|
| GOBP_CELLULAR_RESPONSE_TO_HORMONE_STIMULUS | 19 | -0.265 | -1.182 | 0.23210 | 0.23210 | 0.29478 | 202 |
| HP_ABNORMAL_CEREBRAL_CORTEX_MORPHOLOGY | 44 | 0.291 | 1.207 | 0.23360 | 0.23360 | 0.29614 | 393 |
| GOBP_CELLULAR_AMINO_ACID_METABOLIC_PROCESS | 11 | -0.332 | -1.192 | 0.23376 | 0.23376 | 0.29614 | 2 |
| GOBP_CENTRAL_NERVOUS_SYSTEM_DEVELOPMENT | 31 | 0.315 | 1.206 | 0.23708 | 0.23708 | 0.29997 | 146 |
| HP_APLASIA_HYPOPLASIA_OF_THE_EXTREMITIES | 54 | 0.278 | 1.195 | 0.23771 | 0.23771 | 0.30039 | 304 |
| HP_NEOPLASM_OF_THE_COLON | 10 | 0.433 | 1.218 | 0.23801 | 0.23801 | 0.30039 | 216 |
| HP_ABNORMALITY_OF_THE_OVARY | 13 | 0.397 | 1.215 | 0.23939 | 0.23939 | 0.30140 | 277 |
| GOBP_CELLULAR_RESPONSE_TO_CHEMICAL_STRESS | 11 | 0.418 | 1.212 | 0.23966 | 0.23966 | 0.30140 | 162 |
| HP_KYPHOSIS | 21 | -0.254 | -1.178 | 0.23970 | 0.23970 | 0.30140 | 469 |
| HP_ABNORMAL_SYSTEMIC_BLOOD_PRESSURE | 14 | 0.389 | 1.218 | 0.24011 | 0.24011 | 0.30154 | 99 |
| GOCC_CYTOPLASMIC_REGION | 12 | -0.316 | -1.172 | 0.24196 | 0.24196 | 0.30291 | 175 |
| GOBP_CELLULAR_RESPONSE_TO_OXYGEN_CONTAINING_COMPOUND | 47 | -0.193 | -1.162 | 0.24202 | 0.24202 | 0.30291 | 129 |
| GOBP_RESPONSE_TO_ORGANIC_CYCLIC_COMPOUND | 29 | 0.320 | 1.208 | 0.24210 | 0.24210 | 0.30291 | 216 |
| GOBP_TISSUE_MORPHOGENESIS | 16 | 0.373 | 1.209 | 0.24286 | 0.24286 | 0.30348 | 184 |
| HP_RENAL_HYPOPLASIA | 11 | 0.416 | 1.208 | 0.24337 | 0.24337 | 0.30365 | 180 |
| HP_CEREBELLAR_HYPOPLASIA | 32 | 0.311 | 1.200 | 0.24369 | 0.24369 | 0.30365 | 258 |
| GOMF_HISTONE_BINDING | 23 | 0.338 | 1.207 | 0.24390 | 0.24390 | 0.30365 | 301 |
| GOBP_CARBOHYDRATE_DERIVATIVE_BIOSYNTHETIC_PROCESS | 34 | -0.212 | -1.165 | 0.24447 | 0.24447 | 0.30365 | 159 |
| GOBP_CELLULAR_PROTEIN_CATABOLIC_PROCESS | 39 | 0.295 | 1.192 | 0.24450 | 0.24450 | 0.30365 | 107 |
| HP_GONOSOMAL_INHERITANCE | 15 | 0.380 | 1.211 | 0.24549 | 0.24549 | 0.30432 | 395 |
| GOCC_CELL_CORTEX | 14 | -0.295 | -1.162 | 0.24601 | 0.24601 | 0.30432 | 120 |
| GOBP_PRODUCTION_OF_MOLECULAR_MEDIATOR_OF_IMMUNE_RESPONSE | 11 | 0.415 | 1.204 | 0.24612 | 0.24612 | 0.30432 | 110 |
| HP_SPARSE_HAIR | 28 | 0.321 | 1.202 | 0.24625 | 0.24625 | 0.30432 | 250 |
| HP_LOWER_LIMB_SPASTICITY | 10 | -0.342 | -1.175 | 0.24664 | 0.24664 | 0.30443 | 465 |
| GOBP_RESPONSE_TO_OXYGEN_CONTAINING_COMPOUND | 54 | -0.187 | -1.167 | 0.24762 | 0.24762 | 0.30468 | 129 |
| GOBP_POSITIVE_REGULATION_OF_MOLECULAR_FUNCTION | 78 | 0.256 | 1.175 | 0.24764 | 0.24764 | 0.30468 | 225 |
| HP_SEVERE_GLOBAL_DEVELOPMENTAL_DELAY | 10 | 0.430 | 1.208 | 0.24777 | 0.24777 | 0.30468 | 91 |
| HP_MYOCLONUS | 14 | -0.295 | -1.160 | 0.24806 | 0.24806 | 0.30468 | 325 |
| GOBP_TELOMERE_ORGANIZATION | 20 | 0.350 | 1.204 | 0.24858 | 0.24858 | 0.30494 | 256 |
| HP_ABNORMAL_RESPIRATORY_SYSTEM_MORPHOLOGY | 73 | 0.260 | 1.178 | 0.24927 | 0.24927 | 0.30494 | 247 |
| HP_NARROW_FACE | 11 | 0.414 | 1.201 | 0.24955 | 0.24955 | 0.30494 | 286 |
| HP_PNEUMONIA | 12 | -0.313 | -1.159 | 0.24962 | 0.24962 | 0.30494 | 486 |
| HP_ABNORMAL_JOINT_MORPHOLOGY | 52 | 0.277 | 1.183 | 0.24978 | 0.24978 | 0.30494 | 286 |
| HP_ABNORMALITY_OF_SKIN_PHYSIOLOGY | 16 | 0.371 | 1.203 | 0.25042 | 0.25042 | 0.30536 | 108 |
| HP_SKELETAL_MUSCLE_ATROPHY | 25 | -0.239 | -1.169 | 0.25125 | 0.25125 | 0.30599 | 233 |
| HP_ABNORMALITY_OF_THE_NASAL_TIP | 14 | 0.385 | 1.204 | 0.25215 | 0.25215 | 0.30672 | 146 |
| GOBP_NEGATIVE_REGULATION_OF_CELLULAR_AMIDE_METABOLIC_PROCESS | 11 | -0.324 | -1.163 | 0.25431 | 0.25431 | 0.30898 | 374 |
| GOBP_REGULATION_OF_RESPONSE_TO_BIOTIC_STIMULUS | 11 | -0.324 | -1.161 | 0.25541 | 0.25541 | 0.30994 | 230 |

| | | | | | | | |
|---|---|---|---|---|---|---|---|
| GOBP_CELL_JUNCTION_ORGANIZATION | 12 | -0.311 | -1.152 | 0.25651 | 0.25651 | 0.31065 | 193 |
| GOCC_CELL_SURFACE | 13 | -0.303 | -1.158 | 0.25661 | 0.25661 | 0.31065 | 206 |
| GOBP_REGULATION_OF_PROTEASOMAL_PROTEIN_CATABOLIC_PROCESS | 13 | 0.390 | 1.194 | 0.25894 | 0.25894 | 0.31137 | 107 |
| HP_MENTAL_DETERIORATION | 13 | -0.301 | -1.151 | 0.25898 | 0.25898 | 0.31137 | 494 |
| GOBP_ESTABLISHMENT_OF_CELL_POLARITY | 17 | 0.363 | 1.193 | 0.25910 | 0.25910 | 0.31137 | 184 |
| HP_ABNORMALITY_OF_THE_LIVER | 43 | 0.286 | 1.180 | 0.25925 | 0.25925 | 0.31137 | 237 |
| HP_ABNORMALITY_OF_REFRACTION | 32 | 0.307 | 1.185 | 0.25940 | 0.25940 | 0.31137 | 393 |
| GOBP_ORGANIC_HYDROXY_COMPOUND_METABOLIC_PROCESS | 16 | -0.279 | -1.162 | 0.25955 | 0.25955 | 0.31137 | 122 |
| GOMF_GTPASE_BINDING | 18 | -0.266 | -1.165 | 0.25956 | 0.25956 | 0.31137 | 143 |
| GOBP_REGULATION_OF_UBIQUITIN_DEPENDENT_PROTEIN_CATABOLIC_PROCESS | 12 | 0.402 | 1.199 | 0.25968 | 0.25968 | 0.31137 | 107 |
| HP_INCREASED_SUSCEPTIBILITY_TO_FRACTURES | 12 | 0.401 | 1.198 | 0.26089 | 0.26089 | 0.31246 | 56 |
| HP_ABNORMALITY_OF_THE_URINARY_SYSTEM_PHYSIOLOGY | 50 | 0.277 | 1.176 | 0.26230 | 0.26230 | 0.31377 | 292 |
| HP_ABNORMAL_LABIA_MORPHOLOGY | 10 | 0.425 | 1.195 | 0.26352 | 0.26352 | 0.31465 | 247 |
| GOBP_REGULATION_OF_PROTEIN_CONTAINING_COMPLEX_ASSEMBLY | 18 | 0.355 | 1.186 | 0.26366 | 0.26366 | 0.31465 | 138 |
| HP_ABNORMAL_MYOCARDIUM_MORPHOLOGY | 28 | 0.316 | 1.184 | 0.26402 | 0.26402 | 0.31471 | 303 |
| GOBP_BIOLOGICAL_PROCESS_INVOLVED_IN_INTERSPECIES_INTERACTION_BETWEEN_ORGANISMS | 48 | -0.189 | -1.144 | 0.26468 | 0.26468 | 0.31476 | 122 |
| HP_ABNORMALITY_OF_THE_HALLUX | 12 | -0.308 | -1.143 | 0.26493 | 0.26493 | 0.31476 | 489 |
| HP_ABNORMALITY_OF_DENTAL_ERUPTION | 10 | -0.336 | -1.155 | 0.26502 | 0.26502 | 0.31476 | 469 |
| GOBP_G_PROTEIN_COUPLED_RECEPTOR_SIGNALING_PATHWAY | 12 | -0.308 | -1.141 | 0.26531 | 0.26531 | 0.31476 | 144 |
| HP_ABNORMAL_STOMACH_MORPHOLOGY | 18 | 0.355 | 1.184 | 0.26610 | 0.26610 | 0.31533 | 99 |
| GOBP_POSITIVE_REGULATION_OF_GROWTH | 10 | -0.335 | -1.152 | 0.26820 | 0.26820 | 0.31717 | 129 |
| HP_GRAY_MATTER_HETEROTOPIA | 14 | 0.379 | 1.186 | 0.26829 | 0.26829 | 0.31717 | 309 |
| GOBP_CENTROSOME_DUPLICATION | 15 | 0.372 | 1.186 | 0.26956 | 0.26956 | 0.31802 | 255 |
| HP_ABNORMALITY_OF_THE_URETHRA | 38 | 0.292 | 1.175 | 0.26963 | 0.26963 | 0.31802 | 270 |
| GOCC_AXON | 18 | 0.353 | 1.178 | 0.27176 | 0.27176 | 0.32015 | 49 |
| HP_ABNORMALITY_OF_THE_LENS | 49 | 0.275 | 1.164 | 0.27294 | 0.27294 | 0.32117 | 233 |
| GOBP_REGULATION_OF_DNA_BINDING_TRANSCRIPTION_FACTOR_ACTIVITY | 15 | -0.282 | -1.141 | 0.27331 | 0.27331 | 0.32122 | 48 |
| GOBP_SULFUR_COMPOUND_METABOLIC_PROCESS | 13 | -0.297 | -1.136 | 0.27477 | 0.27477 | 0.32257 | 2 |
| GOBP_CELL_CELL_ADHESION | 10 | -0.332 | -1.142 | 0.27597 | 0.27597 | 0.32360 | 88 |
| HP_MYOPIA | 17 | 0.356 | 1.170 | 0.27724 | 0.27724 | 0.32471 | 393 |
| GOBP_PROTEIN_CATABOLIC_PROCESS | 44 | 0.280 | 1.162 | 0.27776 | 0.27776 | 0.32494 | 126 |
| HP_INGUINAL_HERNIA | 12 | -0.304 | -1.127 | 0.27871 | 0.27871 | 0.32568 | 152 |
| HP_ABNORMAL_NASAL_BRIDGE_MORPHOLOGY | 51 | 0.272 | 1.158 | 0.28041 | 0.28041 | 0.32728 | 91 |
| GOBP_ESTABLISHMENT_OR_MAINTENANCE_OF_CELL_POLARITY | 21 | 0.335 | 1.169 | 0.28108 | 0.28108 | 0.32748 | 184 |
| HP_RETROGNATHIA | 23 | 0.327 | 1.168 | 0.28197 | 0.28197 | 0.32748 | 258 |
| HP_APLASIA_HYPOPLASIA_OF_THE_CEREBELLUM | 38 | 0.289 | 1.164 | 0.28206 | 0.28206 | 0.32748 | 149 |
| HP_ABNORMAL_FEAR_ANXIETY_RELATED_BEHAVIOR | 15 | -0.280 | -1.133 | 0.28220 | 0.28220 | 0.32748 | 509 |
| HP_ABNORMAL_PYRAMIDAL_SIGN | 15 | -0.280 | -1.133 | 0.28220 | 0.28220 | 0.32748 | 509 |

| | | | | | | | |
|---|---|---|---|---|---|---|---|
| GOBP_BEHAVIOR | 11 | -0.315 | -1.130 | 0.28330 | 0.28330 | 0.32833 | 124 |
| GOMF_PROTEIN_MACROMOLECULE_ADAPTOR_ACTIVITY | 13 | -0.295 | -1.127 | 0.28385 | 0.28385 | 0.32833 | 247 |
| GOBP_CELL_ACTIVATION | 29 | -0.219 | -1.136 | 0.28418 | 0.28418 | 0.32833 | 124 |
| HP_X_LINKED_RECESSIVE_INHERITANCE | 13 | 0.383 | 1.171 | 0.28424 | 0.28424 | 0.32833 | 388 |
| GOBP_POSITIVE_REGULATION_OF_CELLULAR_BIOSYNTHETIC_PROCESS | 82 | 0.247 | 1.141 | 0.28464 | 0.28464 | 0.32842 | 227 |
| GOBP_PROTEIN_POLYMERIZATION | 21 | 0.334 | 1.165 | 0.28534 | 0.28534 | 0.32885 | 41 |
| HP_SHORT_ATTENTION_SPAN | 23 | 0.326 | 1.165 | 0.28640 | 0.28640 | 0.32890 | 79 |
| HP_SYNOSTOSIS_OF_JOINTS | 12 | 0.392 | 1.172 | 0.28660 | 0.28660 | 0.32890 | 10 |
| HP_BABINSKI_SIGN | 11 | -0.313 | -1.125 | 0.28697 | 0.28697 | 0.32890 | 485 |
| HP_CEREBRAL_VISUAL_IMPAIRMENT | 11 | -0.313 | -1.125 | 0.28697 | 0.28697 | 0.32890 | 485 |
| GOCC_CELL_CELL_JUNCTION | 11 | -0.313 | -1.124 | 0.28734 | 0.28734 | 0.32890 | 85 |
| HP_POLYHYDRAMNIOS | 17 | 0.353 | 1.160 | 0.28734 | 0.28734 | 0.32890 | 99 |
| HP_VARIABLE_EXPRESSIVITY | 10 | 0.415 | 1.167 | 0.28876 | 0.28876 | 0.32925 | 135 |
| GOBP_PEPTIDYL_SERINE_MODIFICATION | 15 | 0.365 | 1.164 | 0.28880 | 0.28880 | 0.32925 | 107 |
| GOBP_PROTEIN_COMPLEX_OLIGOMERIZATION | 11 | 0.399 | 1.160 | 0.28886 | 0.28886 | 0.32925 | 222 |
| HP_DELAYED_PUBERTY | 11 | 0.399 | 1.159 | 0.28927 | 0.28927 | 0.32925 | 21 |
| HP_AUTISTIC_BEHAVIOR | 23 | -0.238 | -1.132 | 0.28928 | 0.28928 | 0.32925 | 464 |
| GOBP_REGULATION_OF_MRNA_METABOLIC_PROCESS | 13 | -0.292 | -1.115 | 0.29135 | 0.29135 | 0.33124 | 389 |
| HP_OPHTHALMOPARESIS | 11 | -0.312 | -1.119 | 0.29284 | 0.29284 | 0.33243 | 167 |
| HP_HYPERTELORISM | 49 | 0.271 | 1.145 | 0.29306 | 0.29306 | 0.33243 | 242 |
| HP_DEPRESSION | 15 | 0.362 | 1.156 | 0.29730 | 0.29730 | 0.33676 | 341 |
| HP_AGGRESSIVE_BEHAVIOR | 10 | 0.412 | 1.158 | 0.29755 | 0.29755 | 0.33676 | 351 |
| GOBP_NUCLEOSIDE_DIPHOSPHATE_METABOLIC_PROCESS | 10 | -0.325 | -1.117 | 0.29929 | 0.29929 | 0.33836 | 72 |
| HP_PARAPLEGIA_PARAPARESIS | 10 | 0.410 | 1.154 | 0.30103 | 0.30103 | 0.33994 | 23 |
| GOBP_CELL_CELL_SIGNALING_BY_WNT | 10 | -0.324 | -1.113 | 0.30212 | 0.30212 | 0.34079 | 137 |
| GOBP_NEURON_DIFFERENTIATION | 32 | 0.297 | 1.148 | 0.30310 | 0.30310 | 0.34116 | 184 |
| GOBP_DEFENSE_RESPONSE | 40 | -0.193 | -1.110 | 0.30319 | 0.30319 | 0.34116 | 219 |
| GOBP_PROTEIN_LOCALIZATION_TO_ORGANELLE | 53 | 0.264 | 1.134 | 0.30347 | 0.30347 | 0.34116 | 222 |
| HP_HIP_DISLOCATION | 23 | 0.321 | 1.147 | 0.30451 | 0.30451 | 0.34195 | 242 |
| HP_FLEXION_CONTRACTURE_OF_DIGIT | 18 | 0.344 | 1.147 | 0.30531 | 0.30531 | 0.34213 | 142 |
| GOBP_ACTIVATION_OF_PROTEIN_KINASE_ACTIVITY | 11 | 0.394 | 1.144 | 0.30535 | 0.30535 | 0.34213 | 100 |
| GOMF_RNA_BINDING | 88 | -0.155 | -1.096 | 0.30632 | 0.30632 | 0.34284 | 429 |
| HP_ABNORMAL_HAIR_MORPHOLOGY | 63 | 0.255 | 1.130 | 0.30727 | 0.30727 | 0.34352 | 303 |
| GOBP_NEGATIVE_REGULATION_OF_CATABOLIC_PROCESS | 13 | 0.375 | 1.146 | 0.30901 | 0.30901 | 0.34509 | 143 |
| GOMF_CIS_REGULATORY_REGION_SEQUENCE_SPECIFIC_DNA_BINDING | 26 | 0.311 | 1.143 | 0.30951 | 0.30951 | 0.34526 | 227 |
| HP_SMALL_FACE | 12 | 0.383 | 1.145 | 0.31015 | 0.31015 | 0.34554 | 286 |
| HP_APLASIA_HYPOPLASIA_OF_FINGERS | 29 | 0.301 | 1.137 | 0.31068 | 0.31068 | 0.34554 | 250 |
| GOBP_POSITIVE_REGULATION_OF_NUCLEOBASE_CONTAINING_COMPOUND_METABOLIC_PROCESS | 89 | 0.240 | 1.121 | 0.31078 | 0.31078 | 0.34554 | 227 |

| | | | | | | | |
|---|---|---|---|---|---|---|---|
| HP_ABNORMALITY_OF_THE_SYNOVIA | 26 | 0.310 | 1.142 | 0.31145 | 0.31145 | 0.34590 | 242 |
| HP_FUNCTIONAL_MOTOR_DEFICIT | 20 | -0.244 | -1.107 | 0.31259 | 0.31259 | 0.34641 | 465 |
| GOBP_REGULATION_OF_CELLULAR_MACROMOLECULE_BIOSYNTHETIC_PROCESS | 56 | 0.261 | 1.130 | 0.31260 | 0.31260 | 0.34641 | 216 |
| HP_ANEMIA_OF_INADEQUATE_PRODUCTION | 15 | 0.357 | 1.139 | 0.31392 | 0.31392 | 0.34749 | 221 |
| HP_HYPERTROPHIC_CARDIOMYOPATHY | 21 | 0.325 | 1.133 | 0.31439 | 0.31439 | 0.34762 | 271 |
| HP_ABNORMAL_AORTIC_VALVE_PHYSIOLOGY | 11 | 0.392 | 1.137 | 0.31510 | 0.31510 | 0.34803 | 270 |
| GOBP_NEGATIVE_REGULATION_OF_CYTOSKELETON_ORGANIZATION | 14 | 0.364 | 1.139 | 0.31790 | 0.31790 | 0.35074 | 226 |
| GOBP_CELLULAR_RESPONSE_TO_ABIOTIC_STIMULUS | 28 | 0.303 | 1.132 | 0.32048 | 0.32048 | 0.35320 | 238 |
| GOBP_NEGATIVE_REGULATION_OF_SUPRAMOLECULAR_FIBER_ORGANIZATION | 11 | 0.390 | 1.132 | 0.32101 | 0.32101 | 0.35340 | 138 |
| GOBP_IMMUNE_RESPONSE_REGULATING_SIGNALING_PATHWAY | 15 | -0.269 | -1.088 | 0.32246 | 0.32246 | 0.35434 | 206 |
| HP_SMALL_NAIL | 10 | 0.403 | 1.134 | 0.32264 | 0.32264 | 0.35434 | 273 |
| GOBP_TISSUE_DEVELOPMENT | 52 | 0.262 | 1.121 | 0.32292 | 0.32292 | 0.35434 | 191 |
| HP_ABNORMAL_GLUCOSE_HOMEOSTASIS | 25 | -0.225 | -1.104 | 0.32446 | 0.32446 | 0.35564 | 44 |
| HP_ABNORMAL_HAIR_PATTERN | 13 | -0.284 | -1.085 | 0.32531 | 0.32531 | 0.35618 | 506 |
| GOBP_ACTIN_FILAMENT_ORGANIZATION | 18 | 0.338 | 1.127 | 0.32575 | 0.32575 | 0.35628 | 138 |
| HP_ABNORMAL_ANTERIOR_EYE_SEGMENT_MORPHOLOGY | 62 | 0.253 | 1.115 | 0.32645 | 0.32645 | 0.35645 | 304 |
| HP_LONG_FINGERS | 11 | -0.302 | -1.083 | 0.32661 | 0.32661 | 0.35645 | 493 |
| GOBP_SKELETAL_SYSTEM_DEVELOPMENT | 10 | -0.316 | -1.086 | 0.32721 | 0.32721 | 0.35672 | 128 |
| GOBP_ORGANIC_ACID_METABOLIC_PROCESS | 27 | -0.216 | -1.086 | 0.32782 | 0.32782 | 0.35700 | 146 |
| HP_HYPOPIGMENTATION_OF_THE_SKIN | 22 | 0.319 | 1.126 | 0.32845 | 0.32845 | 0.35730 | 233 |
| GOBP_CELL_MORPHOGENESIS | 25 | 0.309 | 1.126 | 0.32894 | 0.32894 | 0.35745 | 138 |
| GOMF_CALCIUM_ION_BINDING | 11 | 0.387 | 1.122 | 0.32967 | 0.32967 | 0.35785 | 52 |
| GOBP_HEAD_DEVELOPMENT | 29 | 0.296 | 1.120 | 0.33061 | 0.33061 | 0.35849 | 146 |
| HP_POLYMICROGYRIA | 11 | 0.386 | 1.121 | 0.33173 | 0.33173 | 0.35901 | 341 |
| GOBP_RESPONSE_TO_BACTERIUM | 10 | -0.315 | -1.082 | 0.33180 | 0.33180 | 0.35901 | 163 |
| GOBP_MAPK_CASCADE | 28 | 0.300 | 1.121 | 0.33233 | 0.33233 | 0.35919 | 142 |
| GOBP_REGULATION_OF_CELLULAR_PROTEIN_CATABOLIC_PROCESS | 14 | 0.359 | 1.124 | 0.33364 | 0.33364 | 0.36023 | 107 |
| HP_ABNORMAL_FEMORAL_NECK_HEAD_MORPHOLOGY | 11 | 0.385 | 1.117 | 0.33654 | 0.33654 | 0.36201 | 304 |
| HP_TRACHEOESOPHAGEAL_FISTULA | 14 | 0.358 | 1.121 | 0.33668 | 0.33668 | 0.36201 | 221 |
| HP_ABNORMALITY_OF_URINE_HOMEOSTASIS | 25 | 0.307 | 1.119 | 0.33687 | 0.33687 | 0.36201 | 99 |
| GOBP_SUPRAMOLECULAR_FIBER_ORGANIZATION | 39 | 0.276 | 1.113 | 0.33700 | 0.33700 | 0.36201 | 164 |
| HP_ATROPHY_DEGENERATION_AFFECTING_THE_CEREBRUM | 37 | -0.192 | -1.086 | 0.33727 | 0.33727 | 0.36201 | 485 |
| GOBP_TELOMERE_MAINTENANCE | 19 | 0.328 | 1.113 | 0.33744 | 0.33744 | 0.36201 | 256 |
| HP_BILATERAL_TONIC_CLONIC_SEIZURE | 12 | -0.290 | -1.076 | 0.33882 | 0.33882 | 0.36238 | 132 |
| GOBP_MITOCHONDRIAL_TRANSPORT | 10 | -0.313 | -1.075 | 0.33887 | 0.33887 | 0.36238 | 485 |
| HP_ABNORMALITY_OF_THE_BASAL_GANGLIA | 10 | -0.313 | -1.075 | 0.33887 | 0.33887 | 0.36238 | 485 |
| GOBP_NEGATIVE_REGULATION_OF_PROTEIN_METABOLIC_PROCESS | 45 | 0.266 | 1.110 | 0.33986 | 0.33986 | 0.36305 | 122 |
| HP_TUBE_FEEDING | 10 | -0.312 | -1.070 | 0.34382 | 0.34382 | 0.36689 | 486 |

| | | | | | | | |
|---|---|---|---|---|---|---|---|
| GOBP_DEPHOSPHORYLATION | 16 | 0.343 | 1.111 | 0.34428 | 0.34428 | 0.36700 | 96 |
| GOBP_POSITIVE_REGULATION_OF_PROTEIN_MODIFICATION_BY_SMALL_PROTEIN_CONJUGATION_OR_REMOVAL | 16 | 0.342 | 1.108 | 0.34780 | 0.34780 | 0.37036 | 334 |
| GOBP_POSITIVE_REGULATION_OF_ESTABLISHMENT_OF_PROTEIN_LOCALIZATION | 14 | -0.270 | -1.062 | 0.35039 | 0.35039 | 0.37272 | 147 |
| GOBP_REGULATION_OF_GTPASE_ACTIVITY | 19 | 0.325 | 1.101 | 0.35207 | 0.35207 | 0.37411 | 142 |
| HP_ABNORMAL_ADIPOSE_TISSUE_MORPHOLOGY | 12 | 0.368 | 1.100 | 0.35386 | 0.35386 | 0.37540 | 56 |
| HP_ABNORMALITY_OF_UPPER_LIP_VERMILLION | 23 | 0.308 | 1.100 | 0.35403 | 0.35403 | 0.37540 | 196 |
| GOBP_POSITIVE_REGULATION_OF_IMMUNE_EFFECTOR_PROCESS | 10 | 0.392 | 1.103 | 0.35443 | 0.35443 | 0.37544 | 96 |
| HP_GLUCOSE_INTOLERANCE | 17 | -0.249 | -1.063 | 0.35567 | 0.35567 | 0.37636 | 44 |
| GOBP_POSITIVE_REGULATION_OF_LOCOMOTION | 17 | -0.249 | -1.062 | 0.35699 | 0.35699 | 0.37725 | 202 |
| GOBP_CIRCULATORY_SYSTEM_DEVELOPMENT | 22 | 0.311 | 1.098 | 0.35785 | 0.35785 | 0.37725 | 31 |
| GOBP_REGULATION_OF_CATABOLIC_PROCESS | 37 | 0.274 | 1.096 | 0.35789 | 0.35789 | 0.37725 | 143 |
| GOBP_VIRAL_LIFE_CYCLE | 13 | 0.360 | 1.100 | 0.35801 | 0.35801 | 0.37725 | 92 |
| HP_ABNORMALITY_OF_TOE | 49 | 0.258 | 1.091 | 0.36053 | 0.36053 | 0.37914 | 221 |
| HP_PROMINENT_FOREHEAD | 13 | 0.359 | 1.099 | 0.36056 | 0.36056 | 0.37914 | 54 |
| GOBP_RESPONSE_TO_LIPID | 25 | 0.301 | 1.095 | 0.36224 | 0.36224 | 0.38019 | 196 |
| GOBP_POSITIVE_REGULATION_OF_CYTOSKELETON_ORGANIZATION | 12 | 0.366 | 1.093 | 0.36252 | 0.36252 | 0.38019 | 255 |
| GOBP_ANATOMICAL_STRUCTURE_FORMATION_INVOLVED_IN_MORPHOGENESIS | 23 | -0.226 | -1.075 | 0.36306 | 0.36306 | 0.38019 | 142 |
| GOBP_RESPONSE_TO_HORMONE | 26 | -0.216 | -1.074 | 0.36307 | 0.36307 | 0.38019 | 156 |
| HP_ABNORMAL_ERYTHROCYTE_MORPHOLOGY | 27 | 0.295 | 1.095 | 0.36372 | 0.36372 | 0.38048 | 233 |
| HP_ABNORMALITY_OF_THE_PULMONARY_VASCULATURE | 10 | 0.389 | 1.093 | 0.36475 | 0.36475 | 0.38092 | 286 |
| HP_INCREASED_NUCHAL_TRANSLUCENCY | 10 | 0.389 | 1.093 | 0.36489 | 0.36489 | 0.38092 | 99 |
| GOBP_PROCESS_UTILIZING_AUTOPHAGIC_MECHANISM | 13 | -0.275 | -1.049 | 0.36636 | 0.36636 | 0.38206 | 116 |
| GOBP_CYTOSKELETON_DEPENDENT_INTRACELLULAR_TRANSPORT | 10 | -0.305 | -1.047 | 0.36714 | 0.36714 | 0.38247 | 130 |
| HP_ABNORMALITY_OF_PRENATAL_DEVELOPMENT_OR_BIRTH | 58 | 0.249 | 1.086 | 0.36788 | 0.36788 | 0.38284 | 196 |
| GOBP_POSITIVE_REGULATION_OF_CELL_DEATH | 26 | -0.216 | -1.070 | 0.36875 | 0.36875 | 0.38336 | 124 |
| HP_BLEPHAROPHIMOSIS | 12 | 0.364 | 1.088 | 0.36942 | 0.36942 | 0.38366 | 286 |
| GOBP_ORGANIC_HYDROXY_COMPOUND_BIOSYNTHETIC_PROCESS | 10 | 0.387 | 1.089 | 0.37033 | 0.37033 | 0.38421 | 114 |
| HP_PUBERTY_AND_GONADAL_DISORDERS | 34 | 0.277 | 1.086 | 0.37134 | 0.37134 | 0.38486 | 303 |
| GOBP_RIBOSE_PHOSPHATE_BIOSYNTHETIC_PROCESS | 15 | -0.260 | -1.052 | 0.37246 | 0.37246 | 0.38551 | 146 |
| HP_GAIT_DISTURBANCE | 43 | -0.181 | -1.060 | 0.37273 | 0.37273 | 0.38551 | 540 |
| HP_PES_PLANUS | 21 | 0.310 | 1.079 | 0.37323 | 0.37323 | 0.38558 | 221 |
| HP_ABNORMALITY_OF_RETINAL_PIGMENTATION | 11 | 0.373 | 1.082 | 0.37378 | 0.37378 | 0.38558 | 149 |
| GOBP_ESTABLISHMENT_OF_PROTEIN_LOCALIZATION_TO_MEMBRANE | 11 | -0.290 | -1.040 | 0.37394 | 0.37394 | 0.38558 | 212 |
| HP_RENAL_INSUFFICIENCY | 22 | 0.306 | 1.080 | 0.37484 | 0.37484 | 0.38611 | 221 |
| HP_APLASIA_HYPOPLASIA_INVOLVING_BONES_OF_THE_UPPER_LIMBS | 40 | 0.266 | 1.081 | 0.37617 | 0.37617 | 0.38708 | 258 |
| GOBP_NUCLEOBASE_CONTAINING_SMALL_MOLECULE_METABOLIC_PROCESS | 27 | -0.209 | -1.052 | 0.37742 | 0.37742 | 0.38797 | 146 |
| HP_ABNORMALITY_OF_THE_NASAL_DORSUM | 21 | 0.308 | 1.073 | 0.37999 | 0.37999 | 0.39022 | 247 |
| GOBP_REGENERATION | 11 | 0.371 | 1.076 | 0.38065 | 0.38065 | 0.39050 | 191 |

| | | | | | | | |
|---|---|---|---|---|---|---|---|
| HP_ABNORMAL_VENTRICULAR_SEPTUM_MORPHOLOGY | 27 | 0.291 | 1.078 | 0.38118 | 0.38118 | 0.39050 | 304 |
| GOCC_PERINUCLEAR_REGION_OF_CYTOPLASM | 35 | 0.272 | 1.073 | 0.38143 | 0.38143 | 0.39050 | 208 |
| GOMF_EXONUCLEASE_ACTIVITY | 11 | 0.370 | 1.074 | 0.38230 | 0.38230 | 0.39074 | 445 |
| GOBP_REGULATION_OF_STRESS_ACTIVATED_PROTEIN_KINASE_SIGNALING_CASCADE | 11 | 0.370 | 1.074 | 0.38244 | 0.38244 | 0.39074 | 96 |
| HP_LIMITATION_OF_JOINT_MOBILITY | 13 | 0.353 | 1.079 | 0.38332 | 0.38332 | 0.39124 | 142 |
| HP_LOW_SET_POSTERIORLY_ROTATED_EARS | 20 | 0.312 | 1.072 | 0.38414 | 0.38414 | 0.39168 | 297 |
| HP_ABNORMALITY_OF_THE_PHILTRUM | 27 | 0.289 | 1.073 | 0.38652 | 0.38652 | 0.39352 | 303 |
| GOBP_POSITIVE_REGULATION_OF_RNA_METABOLIC_PROCESS | 61 | 0.243 | 1.069 | 0.38700 | 0.38700 | 0.39352 | 227 |
| GOMF_CATALYTIC_ACTIVITY_ACTING_ON_A_NUCLEIC_ACID | 63 | 0.242 | 1.070 | 0.38711 | 0.38711 | 0.39352 | 341 |
| GOCC_CILIUM | 17 | -0.242 | -1.034 | 0.38809 | 0.38809 | 0.39411 | 37 |
| GOBP_SIGNAL_TRANSDUCTION_IN_RESPONSE_TO_DNA_DAMAGE | 35 | 0.270 | 1.065 | 0.39295 | 0.39295 | 0.39842 | 196 |
| HP_ABNORMAL_CEREBRAL_VASCULAR_MORPHOLOGY | 11 | 0.367 | 1.065 | 0.39316 | 0.39316 | 0.39842 | 36 |
| GOBP_POSITIVE_REGULATION_OF_PROTEIN_MODIFICATION_PROCESS | 56 | 0.246 | 1.065 | 0.39351 | 0.39351 | 0.39842 | 225 |
| HP_ABNORMALITY_OF_THE_CERVICAL_SPINE | 26 | 0.289 | 1.065 | 0.39651 | 0.39651 | 0.40105 | 180 |
| GOMF_MOLECULAR_ADAPTOR_ACTIVITY | 14 | -0.261 | -1.028 | 0.39828 | 0.39828 | 0.40244 | 247 |
| GOBP_RESPONSE_TO_IONIZING_RADIATION | 22 | 0.300 | 1.060 | 0.40139 | 0.40139 | 0.40485 | 222 |
| HP_DEPRESSED_NASAL_RIDGE | 11 | -0.284 | -1.018 | 0.40147 | 0.40147 | 0.40485 | 14 |
| HP_HOLOPROSENCEPHALY | 10 | 0.374 | 1.053 | 0.41049 | 0.41049 | 0.41353 | 146 |
| HP_PREMATURELY_AGED_APPEARANCE | 13 | 0.344 | 1.052 | 0.41210 | 0.41210 | 0.41431 | 303 |
| HP_ABNORMAL_ESOPHAGUS_MORPHOLOGY | 19 | 0.310 | 1.050 | 0.41233 | 0.41233 | 0.41431 | 221 |
| HP_TREMOR | 25 | -0.210 | -1.029 | 0.41265 | 0.41265 | 0.41431 | 485 |
| HP_PERIPHERAL_NEUROPATHY | 21 | -0.222 | -1.029 | 0.41290 | 0.41290 | 0.41431 | 485 |
| HP_ABNORMAL_MYELINATION | 22 | -0.216 | -1.015 | 0.41546 | 0.41546 | 0.41646 | 228 |
| HP_ABNORMAL_TRACHEAL_MORPHOLOGY | 19 | 0.309 | 1.046 | 0.41716 | 0.41716 | 0.41750 | 359 |
| HP_BLINDNESS | 12 | 0.350 | 1.044 | 0.41732 | 0.41732 | 0.41750 | 79 |
| HP_ANORECTAL_ANOMALY | 24 | 0.290 | 1.048 | 0.41826 | 0.41826 | 0.41801 | 233 |
| HP_PRENATAL_MOVEMENT_ABNORMALITY | 15 | -0.251 | -1.015 | 0.41907 | 0.41907 | 0.41801 | 529 |
| GOBP_GERM_CELL_DEVELOPMENT | 12 | 0.349 | 1.043 | 0.41908 | 0.41908 | 0.41801 | 193 |
| GOBP_NEGATIVE_REGULATION_OF_RESPONSE_TO_DNA_DAMAGE_STIMULUS | 10 | 0.371 | 1.044 | 0.41997 | 0.41997 | 0.41848 | 206 |
| GOBP_REGULATION_OF_INTRACELLULAR_TRANSPORT | 17 | -0.236 | -1.007 | 0.42050 | 0.42050 | 0.41860 | 147 |
| HP_HYPERTRICHOSIS | 16 | 0.321 | 1.040 | 0.42224 | 0.42224 | 0.41992 | 410 |
| HP_IMPAIRMENT_IN_PERSONALITY_FUNCTIONING | 29 | 0.276 | 1.042 | 0.42356 | 0.42356 | 0.42082 | 393 |
| HP_ABNORMALITY_OF_COORDINATION | 42 | -0.176 | -1.022 | 0.42410 | 0.42410 | 0.42094 | 194 |
| HP_SMALL_HAND | 11 | 0.358 | 1.039 | 0.42586 | 0.42586 | 0.42227 | 177 |
| GOBP_POSITIVE_REGULATION_OF_PROTEIN_CATABOLIC_PROCESS | 12 | 0.347 | 1.036 | 0.42747 | 0.42747 | 0.42345 | 107 |
| GOBP_NEGATIVE_REGULATION_OF_NUCLEOBASE_CONTAINING_COMPOUND_METABOLIC_PROCESS | 57 | 0.239 | 1.037 | 0.42803 | 0.42803 | 0.42359 | 216 |
| GOBP_CELL_SUBSTRATE_ADHESION | 10 | -0.290 | -0.997 | 0.42898 | 0.42898 | 0.42411 | 202 |
| GOCC_SITE_OF_DNA_DAMAGE | 17 | 0.314 | 1.034 | 0.42972 | 0.42972 | 0.42433 | 301 |

| | | | | | | | |
|---|---|---|---|---|---|---|---|
| GOBP_REGULATION_OF_INFLAMMATORY_RESPONSE | 12 | 0.346 | 1.034 | 0.43004 | 0.43004 | 0.42433 | 26 |
| HP_FACIAL_HYPERTRICHOSIS | 10 | 0.368 | 1.035 | 0.43070 | 0.43070 | 0.42456 | 146 |
| HP_ABNORMAL_FEMORAL_NECK_MORPHOLOGY | 10 | 0.368 | 1.034 | 0.43154 | 0.43154 | 0.42497 | 304 |
| GOBP_RESPONSE_TO_XENOBIOTIC_STIMULUS | 16 | -0.241 | -1.005 | 0.43200 | 0.43200 | 0.42502 | 102 |
| GOBP_REGULATION_OF_PROTEIN_MODIFICATION_BY_SMALL_PROTEIN_CONJUGATION_OR_REMOVAL | 22 | 0.293 | 1.034 | 0.43339 | 0.43339 | 0.42537 | 334 |
| GOBP_POSITIVE_REGULATION_OF_PROTEIN_SERINE_THREONINE_KINASE_ACTIVITY | 12 | 0.345 | 1.030 | 0.43424 | 0.43424 | 0.42537 | 331 |
| HP_ABNORMALITY_OF_THE_OPTIC_NERVE | 38 | 0.256 | 1.031 | 0.43471 | 0.43471 | 0.42537 | 96 |
| GOBP_POSITIVE_REGULATION_OF_KINASE_ACTIVITY | 27 | 0.278 | 1.032 | 0.43477 | 0.43477 | 0.42537 | 225 |
| HP_ABNORMAL_RENAL_PHYSIOLOGY | 33 | 0.265 | 1.033 | 0.43484 | 0.43484 | 0.42537 | 292 |
| GOBP_RESPONSE_TO_ENDOGENOUS_STIMULUS | 48 | -0.169 | -1.021 | 0.43489 | 0.43489 | 0.42537 | 156 |
| GOBP_POSITIVE_REGULATION_OF_PHOSPHORUS_METABOLIC_PROCESS | 39 | 0.256 | 1.033 | 0.43610 | 0.43610 | 0.42580 | 142 |
| HP_ABNORMALITY_OF_THYROID_PHYSIOLOGY | 13 | 0.337 | 1.032 | 0.43634 | 0.43634 | 0.42580 | 286 |
| HP_HERNIA | 34 | 0.264 | 1.033 | 0.43679 | 0.43679 | 0.42580 | 270 |
| GOBP_POSITIVE_REGULATION_OF_IMMUNE_SYSTEM_PROCESS | 24 | -0.209 | -1.012 | 0.43702 | 0.43702 | 0.42580 | 122 |
| GOBP_REGULATION_OF_ACTIN_FILAMENT_ORGANIZATION | 12 | 0.344 | 1.027 | 0.43870 | 0.43870 | 0.42687 | 138 |
| GOBP_POSITIVE_REGULATION_OF_TRANSCRIPTION_BY_RNA_POLYMERASE_II | 38 | 0.256 | 1.028 | 0.43971 | 0.43971 | 0.42687 | 143 |
| HP_PROGRESSIVE | 12 | -0.268 | -0.993 | 0.43989 | 0.43989 | 0.42687 | 108 |
| GOMF_DNA_BINDING_TRANSCRIPTION_FACTOR_ACTIVITY | 28 | 0.276 | 1.031 | 0.44028 | 0.44028 | 0.42687 | 227 |
| HP_ABNORMAL_CIRCULATING_METABOLITE_CONCENTRATION | 41 | 0.252 | 1.029 | 0.44054 | 0.44054 | 0.42687 | 160 |
| HP_ABNORMAL_INVOLUNTARY_EYE_MOVEMENTS | 55 | 0.239 | 1.030 | 0.44075 | 0.44075 | 0.42687 | 270 |
| GOBP_INNATE_IMMUNE_RESPONSE | 23 | -0.211 | -1.003 | 0.44108 | 0.44108 | 0.42687 | 213 |
| HP_ABNORMAL_RETINAL_MORPHOLOGY | 33 | 0.264 | 1.027 | 0.44184 | 0.44184 | 0.42708 | 162 |
| GOBP_REGULATION_OF_GENE_EXPRESSION_EPIGENETIC | 10 | 0.364 | 1.025 | 0.44241 | 0.44241 | 0.42708 | 401 |
| HP_INTERICTAL_EEG_ABNORMALITY | 11 | -0.276 | -0.992 | 0.44257 | 0.44257 | 0.42708 | 363 |
| HP_ATROPHY_DEGENERATION_AFFECTING_THE_CENTRAL_NERVOUS_SYSTEM | 46 | -0.170 | -1.019 | 0.44314 | 0.44314 | 0.42722 | 494 |
| HP_ABNORMAL_HAIR_QUANTITY | 46 | 0.246 | 1.028 | 0.44572 | 0.44572 | 0.42864 | 216 |
| GOBP_REGULATION_OF_ORGANELLE_ASSEMBLY | 18 | 0.307 | 1.024 | 0.44633 | 0.44633 | 0.42864 | 196 |
| HP_POSITIONAL_FOOT_DEFORMITY | 26 | 0.278 | 1.024 | 0.44637 | 0.44637 | 0.42864 | 393 |
| GOBP_REGULATION_OF_SUPRAMOLECULAR_FIBER_ORGANIZATION | 18 | 0.307 | 1.024 | 0.44646 | 0.44646 | 0.42864 | 251 |
| HP_ABNORMAL_CORPUS_CALLOSUM_MORPHOLOGY | 53 | 0.239 | 1.025 | 0.44746 | 0.44746 | 0.42864 | 266 |
| GOCC_GOLGI_MEMBRANE | 19 | 0.301 | 1.019 | 0.44768 | 0.44768 | 0.42864 | 54 |
| HP_SOMATIC_SENSORY_DYSFUNCTION | 12 | -0.266 | -0.986 | 0.44793 | 0.44793 | 0.42864 | 142 |
| GOMF_PHOSPHATASE_ACTIVITY | 11 | 0.351 | 1.020 | 0.44812 | 0.44812 | 0.42864 | 96 |
| GOBP_INTRACELLULAR_RECEPTOR_SIGNALING_PATHWAY | 11 | -0.275 | -0.986 | 0.44844 | 0.44844 | 0.42864 | 28 |
| GOBP_ORGANOPHOSPHATE_BIOSYNTHETIC_PROCESS | 30 | -0.190 | -1.002 | 0.45515 | 0.45515 | 0.43464 | 146 |
| GOBP_CELLULAR_RESPONSE_TO_IONIZING_RADIATION | 11 | 0.349 | 1.014 | 0.45609 | 0.45609 | 0.43513 | 303 |
| HP_ABNORMALITY_OF_THE_THYROID_GLAND | 20 | 0.295 | 1.012 | 0.45689 | 0.45689 | 0.43530 | 99 |
| GOBP_LYMPHOCYTE_ACTIVATION_INVOLVED_IN_IMMUNE_RESPONSE | 11 | 0.349 | 1.013 | 0.45747 | 0.45747 | 0.43530 | 92 |

| | | | | | | | |
|---|---|---|---|---|---|---|---|
| HP_LARGE_FACE | 13 | -0.256 | -0.979 | 0.45756 | 0.45756 | 0.43530 | 525 |
| HP_RESPIRATORY_INSUFFICIENCY | 24 | 0.281 | 1.014 | 0.45944 | 0.45944 | 0.43667 | 247 |
| HP_ABNORMALITY_OF_TEMPERATURE_REGULATION | 13 | 0.331 | 1.011 | 0.46231 | 0.46231 | 0.43888 | 99 |
| GOBP_POSITIVE_REGULATION_OF_CYTOKINE_PRODUCTION | 10 | 0.358 | 1.008 | 0.46277 | 0.46277 | 0.43888 | 96 |
| HP_HEPATOMEGALY | 19 | 0.297 | 1.007 | 0.46306 | 0.46306 | 0.43888 | 36 |
| HP_INTELLECTUAL_DISABILITY_MILD | 17 | 0.306 | 1.006 | 0.46405 | 0.46405 | 0.43940 | 418 |
| GOBP_ACTOMYOSIN_STRUCTURE_ORGANIZATION | 18 | 0.302 | 1.006 | 0.46574 | 0.46574 | 0.44058 | 138 |
| HP_ABNORMAL_METACARPAL_MORPHOLOGY | 10 | 0.357 | 1.004 | 0.46640 | 0.46640 | 0.44079 | 237 |
| GOBP_TISSUE_MIGRATION | 12 | 0.335 | 1.000 | 0.46820 | 0.46820 | 0.44208 | 68 |
| GOBP_NEURON_MIGRATION | 12 | 0.334 | 0.998 | 0.47077 | 0.47077 | 0.44409 | 66 |
| HP_NARROW_MOUTH | 20 | 0.291 | 0.999 | 0.47212 | 0.47212 | 0.44463 | 292 |
| HP_LONG_PHILTRUM | 19 | 0.296 | 1.001 | 0.47222 | 0.47222 | 0.44463 | 303 |
| HP_APLASIA_HYPOPLASIA_OF_THE_PHALANGES_OF_THE_HAND | 15 | 0.311 | 0.993 | 0.47540 | 0.47540 | 0.44714 | 250 |
| GOBP_NEGATIVE_REGULATION_OF_MULTICELLULAR_ORGANISMAL_PROCESS | 19 | 0.294 | 0.998 | 0.47578 | 0.47578 | 0.44714 | 21 |
| GOCC_ENVELOPE | 68 | -0.151 | -1.002 | 0.47925 | 0.47925 | 0.44980 | 63 |
| HP_TALIPES_EQUINOVARUS | 18 | 0.298 | 0.995 | 0.47950 | 0.47950 | 0.44980 | 318 |
| GOBP_NEGATIVE_REGULATION_OF_RESPONSE_TO_STIMULUS | 54 | 0.232 | 1.000 | 0.48034 | 0.48034 | 0.45001 | 206 |
| HP_ABNORMAL_THUMB_MORPHOLOGY | 32 | 0.258 | 0.997 | 0.48083 | 0.48083 | 0.45001 | 297 |
| HP_ABNORMALITY_OF_THE_PERIPHERAL_NERVOUS_SYSTEM | 23 | 0.279 | 0.997 | 0.48165 | 0.48165 | 0.45001 | 321 |
| GOBP_REGULATION_OF_CELL_PROJECTION_ORGANIZATION | 20 | 0.288 | 0.991 | 0.48169 | 0.48169 | 0.45001 | 74 |
| HP_ABNORMAL_FORM_OF_THE_VERTEBRAL_BODIES | 13 | 0.325 | 0.994 | 0.48226 | 0.48226 | 0.45001 | 297 |
| HP_CONSTITUTIONAL_SYMPTOM | 34 | 0.255 | 0.999 | 0.48240 | 0.48240 | 0.45001 | 138 |
| GOBP_CELLULAR_RESPONSE_TO_LIPID | 18 | 0.297 | 0.991 | 0.48348 | 0.48348 | 0.45002 | 196 |
| GOBP_EMBRYONIC_MORPHOGENESIS | 14 | -0.245 | -0.963 | 0.48383 | 0.48383 | 0.45002 | 134 |
| HP_ABNORMALITY_OF_CENTRAL_NERVOUS_SYSTEM_ELECTROPHYSIOLOGY | 25 | -0.199 | -0.977 | 0.48419 | 0.48419 | 0.45002 | 142 |
| HP_ABNORMAL_NERVOUS_SYSTEM_ELECTROPHYSIOLOGY | 25 | -0.199 | -0.977 | 0.48419 | 0.48419 | 0.45002 | 142 |
| HP_ABNORMALITY_OF_THE_SPLEEN | 19 | 0.292 | 0.990 | 0.48493 | 0.48493 | 0.45029 | 159 |
| HP_ABNORMAL_DIAPHYSIS_MORPHOLOGY | 12 | 0.330 | 0.985 | 0.48606 | 0.48606 | 0.45092 | 137 |
| GOBP_REGULATION_OF_RESPONSE_TO_EXTERNAL_STIMULUS | 31 | 0.259 | 0.992 | 0.48923 | 0.48923 | 0.45345 | 26 |
| GOMF_TRANSCRIPTION_COREPRESSOR_ACTIVITY | 11 | -0.266 | -0.956 | 0.48991 | 0.48991 | 0.45366 | 104 |
| GOBP_REGULATION_OF_CELLULAR_RESPONSE_TO_STRESS | 48 | 0.236 | 0.994 | 0.49122 | 0.49122 | 0.45427 | 213 |
| GOBP_PROTEIN_METHYLATION | 12 | 0.329 | 0.981 | 0.49147 | 0.49147 | 0.45427 | 323 |
| GOBP_POSITIVE_REGULATION_OF_PROTEIN_METABOLIC_PROCESS | 75 | 0.219 | 0.997 | 0.49199 | 0.49199 | 0.45434 | 107 |
| GOBP_TUBE_MORPHOGENESIS | 19 | 0.290 | 0.984 | 0.49282 | 0.49282 | 0.45468 | 19 |
| GOCC_TRANSCRIPTION_REGULATOR_COMPLEX | 20 | 0.285 | 0.980 | 0.49515 | 0.49515 | 0.45642 | 25 |
| GOBP_PEPTIDYL_AMINO_ACID_MODIFICATION | 55 | 0.229 | 0.988 | 0.49643 | 0.49643 | 0.45667 | 197 |
| GOBP_REGULATION_OF_ACTIN_FILAMENT_BASED_PROCESS | 16 | 0.302 | 0.978 | 0.49655 | 0.49655 | 0.45667 | 138 |
| HP_ABNORMALITY_OF_THE_UPPER_RESPIRATORY_TRACT | 16 | -0.231 | -0.962 | 0.49678 | 0.49678 | 0.45667 | 150 |

| | | | | | | | |
|---|---|---|---|---|---|---|---|
| GOBP_PROTEIN_MODIFICATION_BY_SMALL_PROTEIN_CONJUGATION_OR_REMOVAL | 61 | 0.225 | 0.991 | 0.49735 | 0.49735 | 0.45678 | 345 |
| GOBP_POSITIVE_REGULATION_OF_PROTEIN_KINASE_ACTIVITY | 25 | 0.269 | 0.981 | 0.49945 | 0.49945 | 0.45829 | 225 |
| GOMF_PROTEIN_C_TERMINUS_BINDING | 21 | 0.280 | 0.974 | 0.50282 | 0.50282 | 0.46096 | 122 |
| GOBP_REGULATION_OF_IMMUNE_SYSTEM_PROCESS | 39 | -0.171 | -0.978 | 0.50440 | 0.50440 | 0.46166 | 206 |
| GOMF_RIBONUCLEOPROTEIN_COMPLEX_BINDING | 16 | -0.229 | -0.957 | 0.50450 | 0.50450 | 0.46166 | 465 |
| HP_MALABSORPTION | 10 | -0.274 | -0.941 | 0.50601 | 0.50601 | 0.46262 | 512 |
| GOBP_ERK1_AND_ERK2_CASCADE | 11 | 0.335 | 0.972 | 0.50763 | 0.50763 | 0.46368 | 131 |
| GOBP_REGULATION_OF_RAS_PROTEIN_SIGNAL_TRANSDUCTION | 10 | 0.344 | 0.967 | 0.50990 | 0.50990 | 0.46533 | 138 |
| HP_APLASIA_HYPOPLASIA_OF_THE_SKIN | 13 | -0.247 | -0.944 | 0.51125 | 0.51125 | 0.46610 | 44 |
| GOBP_ATP_METABOLIC_PROCESS | 14 | -0.240 | -0.946 | 0.51167 | 0.51167 | 0.46610 | 143 |
| GOBP_I_KAPPAB_KINASE_NF_KAPPAB_SIGNALING | 10 | -0.272 | -0.934 | 0.51237 | 0.51237 | 0.46632 | 41 |
| HP_LOWER_EXTREMITY_JOINT_DISLOCATION | 26 | 0.264 | 0.970 | 0.51359 | 0.51359 | 0.46701 | 242 |
| HP_ABNORMAL_EYELASH_MORPHOLOGY | 20 | 0.281 | 0.964 | 0.51454 | 0.51454 | 0.46745 | 233 |
| HP_BULBOUS_NOSE | 10 | 0.341 | 0.960 | 0.51854 | 0.51854 | 0.47032 | 135 |
| GOBP_ANIMAL_ORGAN_MORPHOGENESIS | 19 | 0.284 | 0.962 | 0.51863 | 0.51863 | 0.47032 | 21 |
| GOBP_PEPTIDYL_TYROSINE_MODIFICATION | 17 | 0.292 | 0.960 | 0.52053 | 0.52053 | 0.47162 | 149 |
| GOBP_BLOOD_VESSEL_MORPHOGENESIS | 13 | 0.314 | 0.960 | 0.52256 | 0.52256 | 0.47303 | 6 |
| GOBP_MONONUCLEAR_CELL_DIFFERENTIATION | 14 | 0.305 | 0.956 | 0.52441 | 0.52441 | 0.47406 | 221 |
| HP_ABNORMALITY_OF_DENTAL_STRUCTURE | 12 | 0.320 | 0.955 | 0.52463 | 0.52463 | 0.47406 | 297 |
| GOBP_REGULATION_OF_IMMUNE_EFFECTOR_PROCESS | 13 | 0.312 | 0.955 | 0.52671 | 0.52671 | 0.47529 | 96 |
| GOBP_REGULATION_OF_CELLULAR_COMPONENT_BIOGENESIS | 47 | 0.230 | 0.966 | 0.52712 | 0.52712 | 0.47529 | 196 |
| HP_MICROCORNEA | 12 | 0.319 | 0.952 | 0.52774 | 0.52774 | 0.47529 | 233 |
| GOMF_IDENTICAL_PROTEIN_BINDING | 107 | 0.204 | 0.977 | 0.52816 | 0.52816 | 0.47529 | 246 |
| GOBP_POSITIVE_REGULATION_OF_DNA_REPAIR | 14 | 0.304 | 0.952 | 0.52851 | 0.52851 | 0.47529 | 213 |
| GOBP_VIRAL_PROCESS | 17 | 0.290 | 0.952 | 0.52882 | 0.52882 | 0.47529 | 92 |
| HP_ABNORMALITY_OF_MOUTH_SIZE | 26 | 0.260 | 0.958 | 0.52997 | 0.52997 | 0.47590 | 297 |
| HP_VISUAL_IMPAIRMENT | 54 | 0.224 | 0.965 | 0.53105 | 0.53105 | 0.47645 | 196 |
| HP_PAIN | 26 | 0.260 | 0.957 | 0.53191 | 0.53191 | 0.47679 | 293 |
| GOBP_NEGATIVE_REGULATION_OF_INTRACELLULAR_SIGNAL_TRANSDUCTION | 29 | 0.253 | 0.956 | 0.53333 | 0.53333 | 0.47763 | 147 |
| HP_HYPOPITUITARISM | 10 | 0.336 | 0.946 | 0.53430 | 0.53430 | 0.47808 | 21 |
| GOBP_POSITIVE_REGULATION_OF_PROTEIN_LOCALIZATION | 22 | -0.199 | -0.935 | 0.53557 | 0.53557 | 0.47879 | 38 |
| GOCC_ANCHORING_JUNCTION | 21 | -0.203 | -0.942 | 0.53648 | 0.53648 | 0.47918 | 102 |
| GOBP_REGULATION_OF_CELLULAR_COMPONENT_SIZE | 14 | 0.302 | 0.944 | 0.53711 | 0.53711 | 0.47932 | 135 |
| GOMF_LYASE_ACTIVITY | 11 | -0.257 | -0.922 | 0.53761 | 0.53761 | 0.47935 | 165 |
| GOBP_PROTEIN_MODIFICATION_BY_SMALL_PROTEIN_CONJUGATION | 54 | 0.223 | 0.961 | 0.53843 | 0.53843 | 0.47943 | 345 |
| HP_ABNORMAL_CEREBELLAR_VERMIS_MORPHOLOGY | 22 | -0.198 | -0.934 | 0.53866 | 0.53866 | 0.47943 | 120 |
| GOBP_NUCLEAR_TRANSPORT | 14 | -0.235 | -0.924 | 0.53950 | 0.53950 | 0.47976 | 180 |
| HP_ABNORMAL_RENAL_PELVIS_MORPHOLOGY | 13 | 0.309 | 0.944 | 0.54104 | 0.54104 | 0.48057 | 350 |

| | | | | | | | |
|---|---|---|---|---|---|---|---|
| GOBP_CELLULAR_MACROMOLECULE_LOCALIZATION | 79 | -0.141 | -0.970 | 0.54136 | 0.54136 | 0.48057 | 147 |
| HP_ABNORMALITY_OF_HINDBRAIN_MORPHOLOGY | 58 | 0.219 | 0.956 | 0.54282 | 0.54282 | 0.48143 | 180 |
| GOBP_PROTEOLYSIS | 69 | 0.213 | 0.960 | 0.54406 | 0.54406 | 0.48183 | 107 |
| GOBP_REGULATION_OF_CELL_DEATH | 67 | -0.146 | -0.968 | 0.54423 | 0.54423 | 0.48183 | 128 |
| GOBP_CELLULAR_RESPONSE_TO_ORGANIC_CYCLIC_COMPOUND | 20 | 0.274 | 0.940 | 0.54537 | 0.54537 | 0.48202 | 254 |
| GOBP_REGULATION_OF_CELL_PROJECTION_ASSEMBLY | 10 | 0.333 | 0.937 | 0.54559 | 0.54559 | 0.48202 | 68 |
| HP_CEREBELLAR_MALFORMATION | 16 | 0.290 | 0.939 | 0.54608 | 0.54608 | 0.48202 | 297 |
| HP_APNEA | 15 | -0.230 | -0.929 | 0.54661 | 0.54661 | 0.48202 | 132 |
| GOCC_NUCLEAR_PROTEIN_CONTAINING_COMPLEX | 107 | 0.202 | 0.966 | 0.54773 | 0.54773 | 0.48202 | 448 |
| GOMF_PROTEIN_HETERODIMERIZATION_ACTIVITY | 16 | 0.290 | 0.938 | 0.54778 | 0.54778 | 0.48202 | 49 |
| HP_OTITIS_MEDIA | 12 | -0.247 | -0.917 | 0.54824 | 0.54824 | 0.48202 | 150 |
| GOBP_POSITIVE_REGULATION_OF_CELLULAR_PROTEIN_CATABOLIC_PROCESS | 10 | 0.332 | 0.934 | 0.54922 | 0.54922 | 0.48202 | 107 |
| GOBP_POSITIVE_REGULATION_OF_PROTEOLYSIS_INVOLVED_IN_CELLULAR_PROTEIN_CATABOLIC_PROCESS | 10 | 0.332 | 0.934 | 0.54922 | 0.54922 | 0.48202 | 107 |
| GOMF_PHOSPHATASE_BINDING | 10 | 0.332 | 0.934 | 0.54922 | 0.54922 | 0.48202 | 138 |
| GOBP_BIOLOGICAL_PROCESS_INVOLVED_IN_SYMBIOTIC_INTERACTION | 14 | -0.233 | -0.917 | 0.55342 | 0.55342 | 0.48498 | 108 |
| HP_ABNORMAL_TRACHEOBRONCHIAL_MORPHOLOGY | 24 | 0.259 | 0.936 | 0.55355 | 0.55355 | 0.48498 | 270 |
| GOBP_CELL_PROJECTION_ORGANIZATION | 41 | 0.231 | 0.943 | 0.55416 | 0.55416 | 0.48509 | 184 |
| GOBP_IMMUNE_EFFECTOR_PROCESS | 18 | 0.280 | 0.933 | 0.55560 | 0.55560 | 0.48593 | 96 |
| GOBP_POSITIVE_REGULATION_OF_PROTEIN_PHOSPHORYLATION | 34 | 0.239 | 0.937 | 0.55659 | 0.55659 | 0.48638 | 225 |
| HP_MACROTIA | 13 | 0.304 | 0.929 | 0.55791 | 0.55791 | 0.48710 | 423 |
| GOBP_CELL_ACTIVATION_INVOLVED_IN_IMMUNE_RESPONSE | 13 | 0.303 | 0.928 | 0.56018 | 0.56018 | 0.48867 | 92 |
| HP_ABNORMAL_CELLULAR_PHENOTYPE | 40 | 0.230 | 0.937 | 0.56189 | 0.56189 | 0.48973 | 221 |
| GOBP_CELLULAR_RESPONSE_TO_ENDOGENOUS_STIMULUS | 40 | -0.164 | -0.942 | 0.56338 | 0.56338 | 0.49045 | 156 |
| GOBP_POSITIVE_REGULATION_OF_INTRACELLULAR_SIGNAL_TRANSDUCTION | 32 | 0.241 | 0.929 | 0.56369 | 0.56369 | 0.49045 | 142 |
| HP_ABNORMAL_ENZYME_COENZYME_ACTIVITY | 21 | -0.198 | -0.921 | 0.56427 | 0.56427 | 0.49049 | 132 |
| GOBP_MRNA_CATABOLIC_PROCESS | 12 | -0.244 | -0.906 | 0.56470 | 0.56470 | 0.49049 | 418 |
| GOBP_REGULATION_OF_APOPTOTIC_SIGNALING_PATHWAY | 17 | -0.214 | -0.913 | 0.56592 | 0.56592 | 0.49113 | 555 |
| GOBP_NEGATIVE_REGULATION_OF_TRANSCRIPTION_BY_RNA_POLYMERASE_II | 33 | 0.238 | 0.928 | 0.56789 | 0.56789 | 0.49215 | 197 |
| GOBP_BIOLOGICAL_PROCESS_INVOLVED_IN_INTERACTION_WITH_HOST | 11 | -0.250 | -0.898 | 0.56807 | 0.56807 | 0.49215 | 92 |
| HP_ABNORMALITY_OF_THE_MIDFACE | 19 | 0.271 | 0.919 | 0.57025 | 0.57025 | 0.49357 | 91 |
| GOBP_JNK_CASCADE | 10 | 0.326 | 0.918 | 0.57069 | 0.57069 | 0.49357 | 96 |
| GOBP_VASCULATURE_DEVELOPMENT | 15 | 0.287 | 0.915 | 0.57276 | 0.57276 | 0.49493 | 6 |
| HP_ABNORMALITY_OF_THE_METAPHYSIS | 11 | 0.317 | 0.921 | 0.57373 | 0.57373 | 0.49534 | 304 |
| HP_ACUTE_LEUKEMIA | 11 | 0.317 | 0.921 | 0.57482 | 0.57482 | 0.49586 | 216 |
| HP_ABNORMALITY_OF_THE_NECK | 32 | 0.238 | 0.920 | 0.57548 | 0.57548 | 0.49596 | 13 |
| HP_CONSTIPATION | 16 | 0.282 | 0.915 | 0.57620 | 0.57620 | 0.49596 | 297 |
| GOBP_TELOMERE_MAINTENANCE_VIA_TELOMERE_LENGTHENING | 10 | 0.325 | 0.913 | 0.57641 | 0.57641 | 0.49596 | 197 |
| HP_DYSPHAGIA | 16 | -0.216 | -0.902 | 0.58215 | 0.58215 | 0.50047 | 553 |

| | | | | | | | |
|---|---|---|---|---|---|---|---|
| HP_ABNORMALITY_OF_THE_PANCREAS | 16 | 0.281 | 0.909 | 0.58441 | 0.58441 | 0.50187 | 196 |
| GOBP_ORGANIC_ACID_BIOSYNTHETIC_PROCESS | 10 | 0.323 | 0.908 | 0.58477 | 0.58477 | 0.50187 | 196 |
| GOBP_RESPONSE_TO_OXIDATIVE_STRESS | 18 | 0.273 | 0.911 | 0.58568 | 0.58568 | 0.50214 | 162 |
| HP_SMALL_FOR_GESTATIONAL_AGE | 17 | 0.276 | 0.907 | 0.58609 | 0.58609 | 0.50214 | 247 |
| GOBP_TUBE_DEVELOPMENT | 25 | 0.250 | 0.911 | 0.58775 | 0.58775 | 0.50314 | 196 |
| GOBP_REGULATION_OF_PROTEIN_POLYMERIZATION | 11 | 0.312 | 0.906 | 0.59186 | 0.59186 | 0.50580 | 186 |
| GOBP_HISTONE_METHYLATION | 11 | 0.312 | 0.906 | 0.59186 | 0.59186 | 0.50580 | 323 |
| GOBP_REGULATION_OF_CELL_ADHESION | 16 | 0.278 | 0.901 | 0.59314 | 0.59314 | 0.50647 | 96 |
| HP_SLEEP_DISTURBANCE | 23 | -0.188 | -0.894 | 0.59448 | 0.59448 | 0.50706 | 432 |
| HP_BRACHYDACTYLY | 20 | 0.262 | 0.899 | 0.59484 | 0.59484 | 0.50706 | 213 |
| GOCC_SOMATODENDRITIC_COMPARTMENT | 21 | 0.257 | 0.897 | 0.59922 | 0.59922 | 0.51036 | 73 |
| GOBP_RNA_MODIFICATION | 12 | -0.237 | -0.879 | 0.60222 | 0.60222 | 0.51248 | 383 |
| GOBP_MICROTUBULE_POLYMERIZATION | 11 | 0.308 | 0.895 | 0.60533 | 0.60533 | 0.51470 | 186 |
| GOMF_PROTEIN_HOMODIMERIZATION_ACTIVITY | 34 | 0.230 | 0.900 | 0.60905 | 0.60905 | 0.51719 | 246 |
| GOBP_RNA_CATABOLIC_PROCESS | 17 | -0.208 | -0.888 | 0.60929 | 0.60929 | 0.51719 | 559 |
| HP_ABNORMALITY_OF_MUSCLE_SIZE | 28 | -0.175 | -0.899 | 0.61156 | 0.61156 | 0.51868 | 512 |
| GOBP_REGULATION_OF_ANATOMICAL_STRUCTURE_SIZE | 15 | 0.276 | 0.882 | 0.61267 | 0.61267 | 0.51886 | 135 |
| HP_ABNORMALITY_OF_FACIAL_SOFT_TISSUE | 15 | 0.276 | 0.882 | 0.61306 | 0.61306 | 0.51886 | 318 |
| GOBP_AMEBOIDAL_TYPE_CELL_MIGRATION | 15 | 0.276 | 0.882 | 0.61332 | 0.61332 | 0.51886 | 68 |
| HP_HIGHLY_ARCHED_EYEBROW | 12 | 0.295 | 0.880 | 0.61624 | 0.61624 | 0.52089 | 393 |
| GOBP_METHYLATION | 20 | 0.256 | 0.881 | 0.62102 | 0.62102 | 0.52450 | 401 |
| GOMF_PROTEIN_DOMAIN_SPECIFIC_BINDING | 33 | 0.227 | 0.883 | 0.62528 | 0.62528 | 0.52704 | 229 |
| GOMF_CADHERIN_BINDING | 17 | 0.267 | 0.878 | 0.62560 | 0.62560 | 0.52704 | 68 |
| GOMF_CELL_ADHESION_MOLECULE_BINDING | 17 | 0.267 | 0.878 | 0.62560 | 0.62560 | 0.52704 | 68 |
| HP_ABNORMALITY_OF_CORNEAL_SIZE | 13 | 0.285 | 0.873 | 0.62632 | 0.62632 | 0.52721 | 270 |
| HP_LETHARGY | 11 | 0.301 | 0.874 | 0.62883 | 0.62883 | 0.52853 | 56 |
| HP_ABNORMALITY_OF_THE_CURVATURE_OF_THE_VERTEBRAL_COLUMN | 62 | 0.204 | 0.899 | 0.62894 | 0.62894 | 0.52853 | 297 |
| HP_PROMINENT_NASAL_BRIDGE | 14 | 0.277 | 0.868 | 0.63064 | 0.63064 | 0.52940 | 388 |
| HP_ABNORMALITY_OF_DENTAL_MORPHOLOGY | 16 | -0.208 | -0.868 | 0.63192 | 0.63192 | 0.52940 | 145 |
| GOCC_NUCLEAR_ENVELOPE | 34 | -0.161 | -0.887 | 0.63264 | 0.63264 | 0.52940 | 38 |
| GOBP_MUSCLE_CELL_DIFFERENTIATION | 13 | 0.284 | 0.869 | 0.63275 | 0.63275 | 0.52940 | 31 |
| GOMF_PHOSPHORIC_ESTER_HYDROLASE_ACTIVITY | 14 | 0.277 | 0.865 | 0.63395 | 0.63395 | 0.52940 | 96 |
| HP_ABNORMAL_URINE_PROTEIN_LEVEL | 10 | 0.309 | 0.869 | 0.63399 | 0.63399 | 0.52940 | 292 |
| GOBP_SMALL_MOLECULE_BIOSYNTHETIC_PROCESS | 18 | 0.260 | 0.867 | 0.63414 | 0.63414 | 0.52940 | 54 |
| HP_PTOSIS | 39 | 0.219 | 0.883 | 0.63417 | 0.63417 | 0.52940 | 303 |
| HP_CONGENITAL_ONSET | 26 | 0.237 | 0.873 | 0.63528 | 0.63528 | 0.52989 | 209 |
| GOBP_ORGANONITROGEN_COMPOUND_CATABOLIC_PROCESS | 52 | 0.208 | 0.889 | 0.63787 | 0.63787 | 0.53160 | 126 |
| GOBP_REGULATION_OF_RESPONSE_TO_CYTOKINE_STIMULUS | 10 | -0.248 | -0.854 | 0.63887 | 0.63887 | 0.53200 | 120 |

| | | | | | | | |
|---|---|---|---|---|---|---|---|
| GOBP_REGULATION_OF_HISTONE_MODIFICATION | 14 | 0.275 | 0.860 | 0.63963 | 0.63963 | 0.53220 | 323 |
| HP_MUSCLE_WEAKNESS | 32 | -0.165 | -0.888 | 0.64170 | 0.64170 | 0.53265 | 147 |
| GOBP_IMMUNE_RESPONSE | 44 | 0.213 | 0.883 | 0.64179 | 0.64179 | 0.53265 | 96 |
| HP_APLASIA_HYPOPLASIA_INVOLVING_BONES_OF_THE_FEET | 15 | 0.268 | 0.856 | 0.64303 | 0.64303 | 0.53265 | 393 |
| GOMF_ACTIN_BINDING | 15 | 0.268 | 0.855 | 0.64316 | 0.64316 | 0.53265 | 152 |
| HP_GAIT_ATAXIA | 11 | -0.238 | -0.853 | 0.64330 | 0.64330 | 0.53265 | 436 |
| HP_APLASIA_HYPOPLASIA_OF_THE_THUMB | 17 | 0.263 | 0.864 | 0.64335 | 0.64335 | 0.53265 | 393 |
| HP_CEREBELLAR_ATROPHY | 21 | -0.186 | -0.863 | 0.64665 | 0.64665 | 0.53494 | 175 |
| GOBP_NUCLEUS_ORGANIZATION | 15 | 0.267 | 0.851 | 0.64931 | 0.64931 | 0.53670 | 157 |
| GOBP_POSITIVE_REGULATION_OF_CELL_ACTIVATION | 12 | 0.285 | 0.852 | 0.65034 | 0.65034 | 0.53711 | 96 |
| GOCC_CELL_SUBSTRATE_JUNCTION | 11 | -0.237 | -0.849 | 0.65248 | 0.65248 | 0.53844 | 302 |
| HP_LANGUAGE_IMPAIRMENT | 37 | 0.217 | 0.867 | 0.65402 | 0.65402 | 0.53927 | 397 |
| GOBP_RESPONSE_TO_ABIOTIC_STIMULUS | 61 | 0.199 | 0.875 | 0.65681 | 0.65681 | 0.54113 | 222 |
| HP_PACE_OF_PROGRESSION | 17 | -0.201 | -0.858 | 0.65834 | 0.65834 | 0.54195 | 108 |
| HP_ABNORMALITY_OF_THE_CEREBRAL_SUBCORTEX | 60 | 0.199 | 0.873 | 0.65893 | 0.65893 | 0.54199 | 266 |
| HP_TETRAPARESIS | 10 | 0.302 | 0.848 | 0.66174 | 0.66174 | 0.54386 | 390 |
| HP_HIGH_PALATE | 42 | 0.209 | 0.858 | 0.66735 | 0.66735 | 0.54789 | 247 |
| GOBP_POSITIVE_REGULATION_OF_CELL_PROJECTION_ORGANIZATION | 10 | 0.300 | 0.843 | 0.66774 | 0.66774 | 0.54789 | 142 |
| HP_INTELLECTUAL_DISABILITY_SEVERE | 27 | 0.229 | 0.850 | 0.66840 | 0.66840 | 0.54799 | 393 |
| GOBP_POSITIVE_REGULATION_OF_CELL_ADHESION | 12 | 0.280 | 0.837 | 0.66969 | 0.66969 | 0.54860 | 96 |
| HP_ABNORMAL_SCALP_MORPHOLOGY | 19 | 0.248 | 0.841 | 0.67273 | 0.67273 | 0.54999 | 237 |
| HP_ABNORMAL_TONGUE_MORPHOLOGY | 11 | 0.289 | 0.838 | 0.67322 | 0.67322 | 0.54999 | 297 |
| HP_ABNORMAL_CIRCULATING_PROTEIN_CONCENTRATION | 20 | 0.245 | 0.842 | 0.67325 | 0.67325 | 0.54999 | 138 |
| GOMF_TRANSCRIPTION_COREGULATOR_ACTIVITY | 23 | -0.178 | -0.846 | 0.67357 | 0.67357 | 0.54999 | 135 |
| HP_JOINT_LAXITY | 11 | 0.288 | 0.837 | 0.67445 | 0.67445 | 0.55027 | 137 |
| HP_ABNORMAL_AGGRESSIVE_IMPULSIVE_OR_VIOLENT_BEHAVIOR | 17 | 0.254 | 0.836 | 0.67574 | 0.67574 | 0.55056 | 351 |
| HP_FUNCTIONAL_ABNORMALITY_OF_THE_GASTROINTESTINAL_TRACT | 46 | 0.205 | 0.858 | 0.67590 | 0.67590 | 0.55056 | 270 |
| GOBP_ALCOHOL_METABOLIC_PROCESS | 12 | 0.278 | 0.830 | 0.67767 | 0.67767 | 0.55156 | 114 |
| HP_FLAT_FACE | 10 | 0.297 | 0.835 | 0.67903 | 0.67903 | 0.55181 | 2 |
| HP_ABNORMALITY_OF_THE_CHIN | 13 | 0.272 | 0.832 | 0.67907 | 0.67907 | 0.55181 | 270 |
| HP_INCREASED_HEAD_CIRCUMFERENCE | 18 | 0.249 | 0.832 | 0.68274 | 0.68274 | 0.55434 | 190 |
| HP_APLASIA_HYPOPLASIA_OF_THE_NAILS | 13 | 0.270 | 0.827 | 0.68376 | 0.68376 | 0.55441 | 273 |
| HP_ABNORMAL_REFLEX | 47 | 0.203 | 0.853 | 0.68395 | 0.68395 | 0.55441 | 270 |
| GOMF_PROTEIN_DIMERIZATION_ACTIVITY | 49 | 0.202 | 0.853 | 0.68479 | 0.68479 | 0.55441 | 222 |
| GOBP_REGULATION_OF_PROTEIN_STABILITY | 15 | 0.258 | 0.823 | 0.68503 | 0.68503 | 0.55441 | 107 |
| HP_ABNORMAL_TOENAIL_MORPHOLOGY | 10 | 0.294 | 0.828 | 0.68809 | 0.68809 | 0.55645 | 287 |
| HP_APLASIA_HYPOPLASIA_OF_THE_CEREBELLAR_VERMIS | 20 | 0.240 | 0.826 | 0.69037 | 0.69037 | 0.55784 | 149 |
| HP_VASCULAR_SKIN_ABNORMALITY | 25 | -0.171 | -0.836 | 0.69107 | 0.69107 | 0.55796 | 490 |

| | | | | | | | |
|---|---|---|---|---|---|---|---|
| HP_HIGH_FOREHEAD | 13 | 0.269 | 0.821 | 0.69206 | 0.69206 | 0.55831 | 99 |
| GOBP_POSITIVE_REGULATION_OF_GTPASE_ACTIVITY | 13 | 0.268 | 0.819 | 0.69447 | 0.69447 | 0.55981 | 142 |
| HP_HYPERACTIVITY | 25 | 0.226 | 0.822 | 0.69765 | 0.69765 | 0.56192 | 393 |
| GOBP_CENTRIOLE_ASSEMBLY | 10 | 0.290 | 0.814 | 0.70120 | 0.70120 | 0.56433 | 255 |
| HP_ABNORMALITY_OF_NEUTROPHILS | 13 | -0.214 | -0.816 | 0.70193 | 0.70193 | 0.56447 | 49 |
| GOBP_RESPONSE_TO_HYDROGEN_PEROXIDE | 11 | 0.279 | 0.810 | 0.70386 | 0.70386 | 0.56512 | 162 |
| GOBP_RESPONSE_TO_REACTIVE_OXYGEN_SPECIES | 11 | 0.279 | 0.810 | 0.70386 | 0.70386 | 0.56512 | 162 |
| GOBP_SMALL_MOLECULE_CATABOLIC_PROCESS | 15 | -0.202 | -0.815 | 0.70551 | 0.70551 | 0.56599 | 165 |
| GOBP_REGULATION_OF_PROTEIN_LOCALIZATION | 38 | -0.149 | -0.844 | 0.71377 | 0.71377 | 0.57217 | 38 |
| GOBP_NEGATIVE_REGULATION_OF_CELLULAR_MACROMOLECULE_BIOSYNTHETIC_PROCESS | 21 | -0.175 | -0.814 | 0.71861 | 0.71861 | 0.57530 | 582 |
| GOBP_LEUKOCYTE_DIFFERENTIATION | 16 | 0.245 | 0.795 | 0.71972 | 0.71972 | 0.57530 | 221 |
| GOMF_UBIQUITIN_LIKE_PROTEIN_LIGASE_BINDING | 22 | 0.226 | 0.798 | 0.72017 | 0.72017 | 0.57530 | 130 |
| GOBP_REGULATION_OF_LYMPHOCYTE_ACTIVATION | 13 | 0.261 | 0.797 | 0.72018 | 0.72018 | 0.57530 | 96 |
| HP_ONSET | 64 | -0.132 | -0.863 | 0.72093 | 0.72093 | 0.57530 | 130 |
| HP_ADULT_ONSET | 11 | -0.224 | -0.802 | 0.72110 | 0.72110 | 0.57530 | 547 |
| HP_DYSTONIA | 22 | -0.173 | -0.815 | 0.72320 | 0.72320 | 0.57633 | 363 |
| GOBP_NEGATIVE_REGULATION_OF_APOPTOTIC_SIGNALING_PATHWAY | 12 | 0.266 | 0.794 | 0.72355 | 0.72355 | 0.57633 | 344 |
| HP_POLYDACTYLY | 14 | 0.250 | 0.784 | 0.73369 | 0.73369 | 0.58396 | 216 |
| GOCC_MITOCHONDRION | 79 | -0.125 | -0.860 | 0.73730 | 0.73730 | 0.58636 | 392 |
| HP_ABNORMAL_EYEBROW_MORPHOLOGY | 30 | 0.209 | 0.793 | 0.74020 | 0.74020 | 0.58821 | 146 |
| HP_MYOCLONIC_SEIZURE | 10 | -0.228 | -0.784 | 0.74488 | 0.74488 | 0.59021 | 393 |
| GOMF_ENDOPEPTIDASE_ACTIVITY | 10 | -0.228 | -0.784 | 0.74488 | 0.74488 | 0.59021 | 396 |
| GOMF_TRANSCRIPTION_REGULATOR_ACTIVITY | 50 | 0.189 | 0.803 | 0.74515 | 0.74515 | 0.59021 | 227 |
| GOBP_MYELOID_CELL_DIFFERENTIATION | 10 | 0.276 | 0.777 | 0.74526 | 0.74526 | 0.59021 | 150 |
| HP_IMMUNODEFICIENCY | 12 | -0.213 | -0.789 | 0.74732 | 0.74732 | 0.59021 | 49 |
| HP_RESPIRATORY_TRACT_INFECTION | 33 | -0.150 | -0.819 | 0.74761 | 0.74761 | 0.59021 | 152 |
| HP_ABNORMAL_PATTERN_OF_RESPIRATION | 28 | 0.210 | 0.786 | 0.74807 | 0.74807 | 0.59021 | 137 |
| HP_ABNORMALITY_OF_THE_DENTITION | 43 | 0.193 | 0.796 | 0.74820 | 0.74820 | 0.59021 | 297 |
| GOCC_INTRACELLULAR_PROTEIN_CONTAINING_COMPLEX | 51 | 0.189 | 0.804 | 0.74820 | 0.74820 | 0.59021 | 339 |
| GOBP_MONOCARBOXYLIC_ACID_METABOLIC_PROCESS | 17 | -0.187 | -0.798 | 0.74858 | 0.74858 | 0.59021 | 132 |
| HP_ATTENTION_DEFICIT_HYPERACTIVITY_DISORDER | 18 | 0.233 | 0.777 | 0.75048 | 0.75048 | 0.59055 | 297 |
| GOCC_GOLGI_APPARATUS_SUBCOMPARTMENT | 11 | 0.265 | 0.769 | 0.75058 | 0.75058 | 0.59055 | 2 |
| GOBP_REGULATION_OF_CELL_ACTIVATION | 17 | -0.186 | -0.795 | 0.75077 | 0.75077 | 0.59055 | 88 |
| HP_TALL_STATURE | 11 | 0.264 | 0.766 | 0.75457 | 0.75457 | 0.59308 | 56 |
| HP_MIDFACE_RETRUSION | 11 | 0.263 | 0.764 | 0.75636 | 0.75636 | 0.59402 | 395 |
| HP_CEREBELLAR_CYST | 13 | 0.249 | 0.762 | 0.75793 | 0.75793 | 0.59480 | 146 |
| HP_GENERALIZED_ONSET_MOTOR_SEIZURE | 10 | -0.224 | -0.770 | 0.75972 | 0.75972 | 0.59573 | 393 |
| GOMF_CATALYTIC_ACTIVITY_ACTING_ON_RNA | 28 | -0.154 | -0.789 | 0.76127 | 0.76127 | 0.59649 | 264 |

| | | | | | | | |
|---|---|---|---|---|---|---|---|
| HP_ABNORMAL_NIPPLE_MORPHOLOGY | 16 | 0.235 | 0.760 | 0.76248 | 0.76248 | 0.59697 | 393 |
| HP_DYSPNEA | 16 | 0.234 | 0.757 | 0.76457 | 0.76457 | 0.59770 | 247 |
| HP_THICKENED_SKIN | 15 | 0.238 | 0.759 | 0.76459 | 0.76459 | 0.59770 | 238 |
| HP_SHORT_THUMB | 14 | 0.241 | 0.755 | 0.76783 | 0.76783 | 0.59947 | 393 |
| HP_WEAKNESS_DUE_TO_UPPER_MOTOR_NEURON_DYSFUNCTION | 24 | 0.209 | 0.754 | 0.76838 | 0.76838 | 0.59947 | 266 |
| GOBP_OSSIFICATION | 10 | 0.268 | 0.755 | 0.76924 | 0.76924 | 0.59947 | 167 |
| GOBP_RESPONSE_TO_STEROID_HORMONE | 10 | 0.268 | 0.754 | 0.76924 | 0.76924 | 0.59947 | 196 |
| GOBP_REGULATION_OF_DEFENSE_RESPONSE | 20 | 0.221 | 0.760 | 0.77168 | 0.77168 | 0.60091 | 26 |
| GOBP_RESPONSE_TO_KETONE | 10 | 0.267 | 0.751 | 0.77273 | 0.77273 | 0.60126 | 191 |
| GOBP_NEGATIVE_REGULATION_OF_CELL_DEATH | 45 | 0.185 | 0.772 | 0.77635 | 0.77635 | 0.60361 | 206 |
| HP_OPTIC_ATROPHY | 27 | 0.205 | 0.759 | 0.77764 | 0.77764 | 0.60415 | 96 |
| HP_TETRAPLEGIA_TETRAPARESIS | 11 | 0.256 | 0.742 | 0.77889 | 0.77889 | 0.60466 | 390 |
| HP_ABNORMALITY_OF_BONE_MARROW_CELL_MORPHOLOGY | 12 | 0.248 | 0.741 | 0.78214 | 0.78214 | 0.60652 | 233 |
| GOMF_GUANYL_NUCLEOTIDE_BINDING | 10 | 0.264 | 0.742 | 0.78249 | 0.78249 | 0.60652 | 1 |
| GOBP_DEFENSE_RESPONSE_TO_OTHER_ORGANISM | 27 | 0.203 | 0.753 | 0.78395 | 0.78395 | 0.60693 | 124 |
| HP_CENTRAL_NERVOUS_SYSTEM_CYST | 14 | 0.236 | 0.739 | 0.78423 | 0.78423 | 0.60693 | 146 |
| GOCC_RIBONUCLEOPROTEIN_GRANULE | 11 | 0.254 | 0.738 | 0.78563 | 0.78563 | 0.60755 | 448 |
| HP_ABNORMAL_LARGE_INTESTINE_MORPHOLOGY | 22 | 0.210 | 0.743 | 0.78628 | 0.78628 | 0.60759 | 238 |
| GOBP_LYMPHOCYTE_ACTIVATION | 21 | 0.213 | 0.744 | 0.78841 | 0.78841 | 0.60876 | 221 |
| GOBP_POSITIVE_REGULATION_OF_CATABOLIC_PROCESS | 21 | 0.213 | 0.741 | 0.79204 | 0.79204 | 0.61110 | 107 |
| HP_MYOPATHY | 12 | -0.204 | -0.755 | 0.79518 | 0.79518 | 0.61305 | 432 |
| GOBP_MUSCLE_STRUCTURE_DEVELOPMENT | 19 | 0.217 | 0.736 | 0.79682 | 0.79682 | 0.61385 | 63 |
| HP_DILATED_FOURTH_VENTRICLE | 14 | 0.232 | 0.725 | 0.79984 | 0.79984 | 0.61486 | 146 |
| HP_GENERALIZED_ABNORMALITY_OF_SKIN | 43 | -0.134 | -0.787 | 0.79984 | 0.79984 | 0.61486 | 613 |
| HP_SHORT_DIGIT | 36 | 0.187 | 0.744 | 0.80007 | 0.80007 | 0.61486 | 297 |
| GOBP_MACROMOLECULE_CATABOLIC_PROCESS | 62 | 0.172 | 0.761 | 0.80057 | 0.80057 | 0.61486 | 126 |
| HP_JOINT_STIFFNESS | 10 | 0.257 | 0.723 | 0.80312 | 0.80312 | 0.61635 | 393 |
| GOBP_PROGRAMMED_CELL_DEATH | 93 | 0.166 | 0.779 | 0.80452 | 0.80452 | 0.61695 | 157 |
| HP_ABNORMAL_URINE_METABOLITE_LEVEL | 18 | 0.217 | 0.725 | 0.80614 | 0.80614 | 0.61773 | 292 |
| HP_INVOLUNTARY_MOVEMENTS | 46 | -0.129 | -0.773 | 0.81104 | 0.81104 | 0.62101 | 117 |
| HP_DUPLICATION_OF_HAND_BONES | 14 | 0.227 | 0.709 | 0.81505 | 0.81505 | 0.62361 | 258 |
| HP_ELEVATED_HEPATIC_TRANSAMINASE | 10 | -0.212 | -0.728 | 0.81590 | 0.81590 | 0.62378 | 36 |
| GOBP_NEURON_APOPTOTIC_PROCESS | 13 | 0.232 | 0.709 | 0.81791 | 0.81791 | 0.62485 | 254 |
| GOBP_POSITIVE_REGULATION_OF_RESPONSE_TO_DNA_DAMAGE_STIMULUS | 17 | 0.216 | 0.709 | 0.82277 | 0.82277 | 0.62809 | 213 |
| GOBP_PEPTIDYL_LYSINE_MODIFICATION | 19 | 0.210 | 0.710 | 0.82352 | 0.82352 | 0.62818 | 323 |
| HP_ABNORMAL_LIVER_MORPHOLOGY | 28 | 0.192 | 0.718 | 0.82447 | 0.82447 | 0.62843 | 36 |
| GOBP_NEGATIVE_REGULATION_OF_GENE_EXPRESSION | 36 | 0.182 | 0.721 | 0.82574 | 0.82574 | 0.62892 | 191 |
| HP_ABNORMALITY_OF_THE_TONGUE | 12 | 0.234 | 0.700 | 0.82652 | 0.82652 | 0.62904 | 297 |

| | | | | | | | |
|---|---|---|---|---|---|---|---|
| HP_GENERALIZED_ONSET_SEIZURE | 13 | 0.228 | 0.698 | 0.82929 | 0.82929 | 0.63036 | 91 |
| GOCC_ORGANELLE_SUBCOMPARTMENT | 43 | 0.175 | 0.722 | 0.82950 | 0.82950 | 0.63036 | 154 |
| HP_FAILURE_TO_THRIVE | 51 | 0.171 | 0.728 | 0.83029 | 0.83029 | 0.63049 | 304 |
| GOBP_PATTERN_RECOGNITION_RECEPTOR_SIGNALING_PATHWAY | 10 | 0.247 | 0.695 | 0.83324 | 0.83324 | 0.63184 | 96 |
| GOBP_POSITIVE_REGULATION_OF_CELLULAR_COMPONENT_BIOGENESIS | 24 | 0.193 | 0.699 | 0.83333 | 0.83333 | 0.63184 | 186 |
| HP_RECURRENT_RESPIRATORY_INFECTIONS | 25 | 0.191 | 0.695 | 0.83510 | 0.83510 | 0.63271 | 287 |
| HP_ABNORMAL_INFLAMMATORY_RESPONSE | 41 | 0.175 | 0.716 | 0.83851 | 0.83851 | 0.63481 | 108 |
| GOCC_SITE_OF_DOUBLE_STRAND_BREAK | 13 | 0.225 | 0.689 | 0.83920 | 0.83920 | 0.63486 | 291 |
| HP_NAUSEA_AND_VOMITING | 24 | 0.191 | 0.691 | 0.83983 | 0.83983 | 0.63486 | 271 |
| HP_REDUCED_CONSCIOUSNESS_CONFUSION | 16 | 0.211 | 0.685 | 0.84057 | 0.84057 | 0.63494 | 56 |
| GOMF_UBIQUITIN_LIKE_PROTEIN_TRANSFERASE_ACTIVITY | 26 | 0.188 | 0.693 | 0.84251 | 0.84251 | 0.63593 | 345 |
| HP_ABNORMAL_CONJUGATE_EYE_MOVEMENT | 54 | 0.165 | 0.711 | 0.84718 | 0.84718 | 0.63898 | 297 |
| GOBP_REGULATION_OF_CELLULAR_LOCALIZATION | 39 | -0.129 | -0.740 | 0.84824 | 0.84824 | 0.63930 | 147 |
| GOBP_CYTOKINE_PRODUCTION | 20 | 0.199 | 0.684 | 0.84921 | 0.84921 | 0.63956 | 261 |
| HP_ABNORMALITY_OF_MOUTH_SHAPE | 12 | 0.223 | 0.667 | 0.86157 | 0.86157 | 0.64838 | 149 |
| HP_VOMITING | 16 | 0.203 | 0.656 | 0.86716 | 0.86716 | 0.65192 | 56 |
| HP_RESPIRATORY_DISTRESS | 11 | 0.227 | 0.658 | 0.86918 | 0.86918 | 0.65192 | 247 |
| HP_PECTUS_EXCAVATUM | 16 | 0.202 | 0.653 | 0.86964 | 0.86964 | 0.65192 | 2 |
| HP_HYPERTONIA | 55 | 0.160 | 0.689 | 0.86978 | 0.86978 | 0.65192 | 397 |
| HP_BRACHYCEPHALY | 13 | 0.214 | 0.655 | 0.87000 | 0.87000 | 0.65192 | 91 |
| HP_HIP_DYSPLASIA | 16 | 0.202 | 0.653 | 0.87016 | 0.87016 | 0.65192 | 393 |
| GOBP_NEGATIVE_REGULATION_OF_BIOSYNTHETIC_PROCESS | 67 | 0.156 | 0.697 | 0.87190 | 0.87190 | 0.65274 | 216 |
| HP_ABNORMAL_MYELOID_LEUKOCYTE_MORPHOLOGY | 14 | 0.208 | 0.652 | 0.87419 | 0.87419 | 0.65397 | 233 |
| HP_FUNCTIONAL_ABNORMALITY_OF_THE_MIDDLE_EAR | 20 | 0.190 | 0.653 | 0.87565 | 0.87565 | 0.65457 | 96 |
| HP_COGNITIVE_IMPAIRMENT | 30 | -0.131 | -0.691 | 0.87718 | 0.87718 | 0.65524 | 613 |
| GOBP_POSITIVE_REGULATION_OF_DNA_RECOMBINATION | 10 | 0.231 | 0.649 | 0.87828 | 0.87828 | 0.65525 | 213 |
| HP_ABNORMAL_EMOTION_AFFECT_BEHAVIOR | 28 | 0.176 | 0.659 | 0.87875 | 0.87875 | 0.65525 | 351 |
| HP_TOOTH_MALPOSITION | 16 | 0.199 | 0.644 | 0.87916 | 0.87916 | 0.65525 | 80 |
| GOBP_REGULATION_OF_IMMUNE_RESPONSE | 29 | 0.174 | 0.658 | 0.88135 | 0.88135 | 0.65640 | 96 |
| GOBP_NEURON_DEATH | 18 | 0.193 | 0.643 | 0.88340 | 0.88340 | 0.65745 | 254 |
| HP_ABNORMAL_CIRCULATING_LIPID_CONCENTRATION | 14 | 0.203 | 0.637 | 0.88729 | 0.88729 | 0.65971 | 54 |
| GOCC_CATALYTIC_COMPLEX | 87 | 0.150 | 0.696 | 0.88775 | 0.88775 | 0.65971 | 359 |
| GOMF_UBIQUITIN_LIKE_PROTEIN_LIGASE_ACTIVITY | 20 | 0.186 | 0.638 | 0.88899 | 0.88899 | 0.66014 | 93 |
| HP_ABNORMAL_CNS_MYELINATION | 13 | -0.176 | -0.672 | 0.89222 | 0.89222 | 0.66206 | 212 |
| HP_GENERALIZED_HYPOTONIA | 41 | -0.118 | -0.686 | 0.89422 | 0.89422 | 0.66238 | 623 |
| GOBP_NEGATIVE_REGULATION_OF_PROTEIN_LOCALIZATION | 11 | -0.185 | -0.665 | 0.89541 | 0.89541 | 0.66238 | 108 |
| GOBP_ORGANIC_CYCLIC_COMPOUND_CATABOLIC_PROCESS | 21 | -0.143 | -0.666 | 0.89578 | 0.89578 | 0.66238 | 260 |
| GOBP_POSITIVE_REGULATION_OF_IMMUNE_RESPONSE | 19 | 0.186 | 0.630 | 0.89587 | 0.89587 | 0.66238 | 96 |

| | | | | | | | |
|---|---|---|---|---|---|---|---|
| HP_UPPER_MOTOR_NEURON_DYSFUNCTION | 51 | 0.154 | 0.656 | 0.89595 | 0.89595 | 0.66238 | 270 |
| GOBP_NUCLEIC_ACID_PHOSPHODIESTER_BOND_HYDROLYSIS | 27 | -0.136 | -0.683 | 0.89681 | 0.89681 | 0.66253 | 551 |
| HP_ABNORMAL_HAND_MORPHOLOGY | 27 | 0.168 | 0.624 | 0.90373 | 0.90373 | 0.66716 | 250 |
| HP_DELAYED_GROSS_MOTOR_DEVELOPMENT | 16 | 0.190 | 0.615 | 0.90601 | 0.90601 | 0.66835 | 79 |
| HP_FUNCTIONAL_ABNORMALITY_OF_THE_INNER_EAR | 31 | 0.163 | 0.625 | 0.90969 | 0.90969 | 0.67058 | 138 |
| HP_CEREBRAL_CORTICAL_ATROPHY | 16 | 0.188 | 0.607 | 0.91292 | 0.91292 | 0.67222 | 407 |
| GOBP_PROTEIN_POLYUBIQUITINATION | 18 | 0.181 | 0.603 | 0.91413 | 0.91413 | 0.67222 | 130 |
| GOBP_NEGATIVE_REGULATION_OF_SIGNALING | 49 | 0.149 | 0.629 | 0.91490 | 0.91490 | 0.67222 | 206 |
| HP_ENLARGED_POSTERIOR_FOSSA | 15 | 0.190 | 0.607 | 0.91534 | 0.91534 | 0.67222 | 146 |
| GOCC_NUCLEAR_SPECK | 20 | 0.175 | 0.602 | 0.91668 | 0.91668 | 0.67222 | 539 |
| GOBP_B_CELL_ACTIVATION | 13 | 0.195 | 0.597 | 0.91686 | 0.91686 | 0.67222 | 193 |
| HP_ABNORMAL_BRAINSTEM_MORPHOLOGY | 12 | 0.200 | 0.597 | 0.91732 | 0.91732 | 0.67222 | 13 |
| HP_ABNORMALITY_OF_CIRCULATING_ENZYME_LEVEL | 12 | 0.200 | 0.596 | 0.91786 | 0.91786 | 0.67222 | 11 |
| HP_ABNORMAL_PERIPHERAL_NERVOUS_SYSTEM_MORPHOLOGY | 18 | 0.179 | 0.598 | 0.91850 | 0.91850 | 0.67222 | 56 |
| GOBP_REGULATION_OF_RESPONSE_TO_STRESS | 67 | 0.144 | 0.642 | 0.91859 | 0.91859 | 0.67222 | 221 |
| GOMF_OXIDOREDUCTASE_ACTIVITY | 24 | 0.167 | 0.602 | 0.91998 | 0.91998 | 0.67275 | 198 |
| HP_ABSENT_SPEECH | 13 | 0.193 | 0.590 | 0.92141 | 0.92141 | 0.67331 | 388 |
| GOCC_NUCLEAR_OUTER_MEMBRANE_ENDOPLASMIC_RETICULUM_MEMBRANE_NETWORK | 34 | 0.156 | 0.609 | 0.92309 | 0.92309 | 0.67383 | 154 |
| HP_MANDIBULAR_PROGNATHIA | 10 | 0.210 | 0.592 | 0.92345 | 0.92345 | 0.67383 | 233 |
| GOMF_HYDROLASE_ACTIVITY_ACTING_ON_ESTER_BONDS | 40 | 0.151 | 0.613 | 0.92511 | 0.92511 | 0.67455 | 172 |
| GOBP_POSITIVE_REGULATION_OF_INTRACELLULAR_TRANSPORT | 12 | -0.171 | -0.633 | 0.92611 | 0.92611 | 0.67459 | 90 |
| GOMF_CYSTEINE_TYPE_PEPTIDASE_ACTIVITY | 10 | -0.183 | -0.627 | 0.92650 | 0.92650 | 0.67459 | 575 |
| HP_MOTOR_SEIZURE | 14 | 0.186 | 0.583 | 0.92777 | 0.92777 | 0.67502 | 91 |
| HP_ABNORMAL_THORAX_MORPHOLOGY | 42 | 0.146 | 0.599 | 0.93122 | 0.93122 | 0.67644 | 297 |
| GOBP_NUCLEOSIDE_PHOSPHATE_BIOSYNTHETIC_PROCESS | 21 | 0.168 | 0.585 | 0.93151 | 0.93151 | 0.67644 | 191 |
| HP_SHORT_NOSE | 17 | 0.176 | 0.579 | 0.93173 | 0.93173 | 0.67644 | 270 |
| GOBP_POSITIVE_REGULATION_OF_PROTEIN_CONTAINING_COMPLEX_ASSEMBLY | 11 | -0.173 | -0.620 | 0.93394 | 0.93394 | 0.67710 | 582 |
| GOMF_DNA_BINDING_TRANSCRIPTION_FACTOR_BINDING | 14 | 0.183 | 0.574 | 0.93399 | 0.93399 | 0.67710 | 247 |
| HP_ABNORMAL_NEPHRON_MORPHOLOGY | 11 | 0.201 | 0.582 | 0.93486 | 0.93486 | 0.67719 | 96 |
| GOMF_NUCLEASE_ACTIVITY | 23 | 0.160 | 0.572 | 0.93545 | 0.93545 | 0.67719 | 474 |
| HP_DEPRESSED_NASAL_BRIDGE | 31 | 0.153 | 0.585 | 0.93684 | 0.93684 | 0.67746 | 91 |
| HP_ABNORMALITY_OF_THE_SKULL_BASE | 16 | 0.177 | 0.573 | 0.93717 | 0.93717 | 0.67746 | 146 |
| HP_FEEDING_DIFFICULTIES_IN_INFANCY | 30 | -0.118 | -0.620 | 0.93799 | 0.93799 | 0.67757 | 622 |
| HP_ABNORMAL_STERNUM_MORPHOLOGY | 22 | 0.161 | 0.568 | 0.94021 | 0.94021 | 0.67869 | 138 |
| GOBP_POSITIVE_REGULATION_OF_GENE_EXPRESSION | 36 | 0.145 | 0.577 | 0.94269 | 0.94269 | 0.67999 | 196 |
| GOBP_NUCLEOSIDE_MONOPHOSPHATE_METABOLIC_PROCESS | 10 | 0.200 | 0.562 | 0.94367 | 0.94367 | 0.68021 | 287 |
| GOMF_TRANSCRIPTION_FACTOR_BINDING | 27 | -0.121 | -0.609 | 0.94755 | 0.94755 | 0.68252 | 567 |
| GOBP_ADAPTIVE_IMMUNE_RESPONSE | 12 | 0.184 | 0.550 | 0.94871 | 0.94871 | 0.68287 | 92 |

| | | | | | | | |
|---|---|---|---|---|---|---|---|
| HP_MOTOR_DELAY | 36 | -0.110 | -0.616 | 0.95099 | 0.95099 | 0.68402 | 120 |
| HP_SPASTICITY | 42 | 0.137 | 0.564 | 0.95392 | 0.95392 | 0.68564 | 55 |
| HP_ABNORMALITY_OF_THE_MIDDLE_EAR | 30 | 0.142 | 0.540 | 0.95756 | 0.95756 | 0.68777 | 96 |
| HP_ABNORMALITY_OF_THE_VOICE | 24 | 0.147 | 0.532 | 0.95833 | 0.95833 | 0.68783 | 393 |
| GOBP_PROTEIN_LOCALIZATION_TO_NUCLEUS | 13 | 0.174 | 0.531 | 0.95997 | 0.95997 | 0.68835 | 222 |
| GOBP_POSITIVE_REGULATION_OF_HISTONE_MODIFICATION | 11 | 0.184 | 0.534 | 0.96042 | 0.96042 | 0.68835 | 323 |
| GOBP_DOUBLE_STRAND_BREAK_REPAIR_VIA_NONHOMOLOGOUS_END_JOINING | 12 | 0.176 | 0.525 | 0.96238 | 0.96238 | 0.68927 | 579 |
| HP_DEVELOPMENTAL_REGRESSION | 15 | -0.137 | -0.555 | 0.96398 | 0.96398 | 0.68992 | 74 |
| GOMF_STRUCTURAL_MOLECULE_ACTIVITY | 16 | 0.159 | 0.514 | 0.96884 | 0.96884 | 0.69291 | 384 |
| GOMF_PROTEIN_CONTAINING_COMPLEX_BINDING | 60 | 0.121 | 0.530 | 0.97325 | 0.97325 | 0.69553 | 254 |
| HP_PES_CAVUS | 12 | 0.167 | 0.499 | 0.97388 | 0.97388 | 0.69553 | 162 |
| HP_DEEPLY_SET_EYE | 15 | 0.154 | 0.492 | 0.97632 | 0.97632 | 0.69677 | 423 |
| GOMF_MODIFICATION_DEPENDENT_PROTEIN_BINDING | 14 | 0.155 | 0.486 | 0.97725 | 0.97725 | 0.69694 | 240 |
| HP_ABNORMAL_ESOPHAGUS_PHYSIOLOGY | 33 | 0.122 | 0.476 | 0.98008 | 0.98008 | 0.69847 | 270 |
| GOBP_LYMPHOCYTE_MEDIATED_IMMUNITY | 11 | 0.163 | 0.472 | 0.98557 | 0.98557 | 0.70178 | 379 |
| GOMF_ENDONUCLEASE_ACTIVITY | 18 | 0.136 | 0.453 | 0.98612 | 0.98612 | 0.70178 | 561 |
| GOBP_NEGATIVE_REGULATION_OF_NEURON_DEATH | 11 | 0.160 | 0.465 | 0.98791 | 0.98791 | 0.70256 | 193 |
| HP_GASTROESOPHAGEAL_REFLUX | 23 | 0.119 | 0.424 | 0.99224 | 0.99224 | 0.70478 | 146 |
| GOBP_RNA_PHOSPHODIESTER_BOND_HYDROLYSIS | 14 | 0.134 | 0.418 | 0.99286 | 0.99286 | 0.70478 | 341 |
| HP_DYSARTHRIA | 25 | -0.102 | -0.502 | 0.99390 | 0.99390 | 0.70478 | 560 |
| HP_FEEDING_DIFFICULTIES | 53 | 0.095 | 0.409 | 0.99551 | 0.99551 | 0.70478 | 271 |
| GOMF_RIBONUCLEASE_ACTIVITY | 13 | 0.131 | 0.401 | 0.99625 | 0.99625 | 0.70478 | 610 |
| GOBP_ESTABLISHMENT_OF_PROTEIN_LOCALIZATION_TO_ORGANELLE | 16 | 0.121 | 0.391 | 0.99635 | 0.99635 | 0.70478 | 31 |
| HP_HYPOPLASIA_OF_THE_CORPUS_CALLOSUM | 27 | 0.101 | 0.375 | 0.99709 | 0.99709 | 0.70478 | 350 |
| GOCC_MITOCHONDRIAL_MATRIX | 24 | -0.092 | -0.446 | 0.99729 | 0.99729 | 0.70478 | 88 |
| GOBP_LEUKOCYTE_MEDIATED_IMMUNITY | 13 | 0.128 | 0.392 | 0.99732 | 0.99732 | 0.70478 | 612 |

**Supplemental Table S11**. Differentially expressed genes from the comparison between samples with monoallelic *TP53* compared to WT *TP53* . Data from this table are derived from the FIMM dataset.

| ENSEMBL_ID | logFC | AveExpr | t | P.Value | p.adjust | B | External gene name | Description |
|---|---|---|---|---|---|---|---|---|
| ENSG00000145391 | -1.676 | 6.874 | -4.688 | 0.00000 | 0.00000 | 2.617 | SETD7 | SET domain containing lysine methyltransferase 7 [Source:HGNC Symbol;Acc:HGNC:30412] |
| ENSG00000135925 | 2.711 | 2.612 | 4.57 | 0.00000 | 0.00000 | 2.128 | WNT10A | Wnt family member 10A [Source:HGNC Symbol;Acc:HGNC:13829] |
| ENSG00000142552 | 2.541 | 0.861 | 4.438 | 0.00000 | 0.00000 | 0.966 | RCN3 | reticulocalbin 3 [Source:HGNC Symbol;Acc:HGNC:21145] |
| ENSG00000078401 | 3.267 | 1.475 | 4.433 | 0.00000 | 0.00000 | 1.503 | EDN1 | endothelin 1 [Source:HGNC Symbol;Acc:HGNC:3176] |
| ENSG00000158286 | 2.087 | 1.976 | 4.423 | 0.00000 | 0.00000 | 1.455 | RNF207 | ring finger protein 207 [Source:HGNC Symbol;Acc:HGNC:32947] |
| ENSG00000134184 | 2.715 | -0.27 | 4.202 | 0.00000 | 0.00000 | -0.257 | GSTM1 | glutathione S-transferase mu 1 [Source:HGNC Symbol;Acc:HGNC:4632] |
| ENSG00000087494 | 2.662 | 1.112 | 4.05 | 0.00000 | 0.00000 | 0.182 | PTHLH | parathyroid hormone like hormone [Source:HGNC Symbol;Acc:HGNC:9607] |
| ENSG00000114867 | -0.802 | 8.299 | -4.021 | 0.00000 | 0.00000 | 0.638 | EIF4G1 | eukaryotic translation initiation factor 4 gamma 1 [Source:HGNC Symbol;Acc:HGNC:3296] |
| ENSG00000169126 | 3.188 | 0.897 | 3.983 | 0.00000 | 0.00000 | 0.066 | ARMC4 | armadillo repeat containing 4 [Source:HGNC Symbol;Acc:HGNC:25583] |
| ENSG00000141252 | -1.055 | 5.171 | -3.945 | 0.00000 | 0.00000 | 0.341 | VPS53 | VPS53, GARP complex subunit [Source:HGNC Symbol;Acc:HGNC:25608] |
| ENSG00000253250 | 1.942 | 2.45 | 3.807 | 0.00000 | 0.00000 | -0.07 | C8orf88 | chromosome 8 open reading frame 88 [Source:HGNC Symbol;Acc:HGNC:44672] |
| ENSG00000168447 | 2.179 | 2.066 | 3.774 | 0.00000 | 0.00000 | -0.205 | SCNN1B | sodium channel epithelial 1 beta subunit [Source:HGNC Symbol;Acc:HGNC:10600] |
| ENSG00000154611 | 2.414 | 0.128 | 3.715 | 0.00000 | 0.00000 | -1.082 | PSMA8 | proteasome subunit alpha 8 [Source:HGNC Symbol;Acc:HGNC:22985] |
| ENSG00000154655 | 1.999 | 3.391 | 3.708 | 0.00000 | 0.00000 | -0.204 | L3MBTL4 | l(3)mbt-like 4 (Drosophila) [Source:HGNC Symbol;Acc:HGNC:26677] |
| ENSG00000091592 | -1.379 | 6.252 | -3.697 | 0.00000 | 0.00000 | -0.218 | NLRP1 | NLR family pyrin domain containing 1 [Source:HGNC Symbol;Acc:HGNC:14374] |
| ENSG00000173166 | 1.696 | 4.103 | 3.606 | 0.00100 | 0.00100 | -0.438 | RAPH1 | Ras association (RalGDS/AF-6) and pleckstrin homology domains 1 [Source:HGNC Symbol;Acc:HGNC:14436] |
| ENSG00000132382 | -1.232 | 5.578 | -3.58 | 0.00100 | 0.00100 | -0.566 | MYBBP1A | MYB binding protein 1a [Source:HGNC Symbol;Acc:HGNC:7546] |
| ENSG00000174231 | -0.939 | 7.297 | -3.532 | 0.00100 | 0.00100 | -0.616 | PRPF8 | pre-mRNA processing factor 8 [Source:HGNC Symbol;Acc:HGNC:17340] |
| ENSG00000123094 | 1.928 | 5.047 | 3.519 | 0.00100 | 0.00100 | -0.657 | RASSF8 | Ras association domain family member 8 [Source:HGNC Symbol;Acc:HGNC:13232] |
| ENSG00000123096 | 2.249 | 4.71 | 3.509 | 0.00100 | 0.00100 | -0.683 | SSPN | sarcospan [Source:HGNC Symbol;Acc:HGNC:11322] |
| ENSG00000174238 | -0.775 | 5.184 | -3.488 | 0.00100 | 0.00100 | -0.792 | PITPNA | phosphatidylinositol transfer protein alpha [Source:HGNC Symbol;Acc:HGNC:9001] |
| ENSG00000221909 | 0.846 | 2.061 | 3.427 | 0.00100 | 0.00100 | -1.293 | FAM200A | family with sequence similarity 200 member A [Source:HGNC Symbol;Acc:HGNC:25401] |
| ENSG00000165118 | 0.898 | 2.587 | 3.423 | 0.00100 | 0.00100 | -1.127 | C9orf64 | chromosome 9 open reading frame 64 [Source:HGNC Symbol;Acc:HGNC:28144] |
| ENSG00000150961 | -0.871 | 7.553 | -3.423 | 0.00100 | 0.00100 | -0.897 | SEC24D | SEC24 homolog D, COPII coat complex component [Source:HGNC Symbol;Acc:HGNC:10706] |
| ENSG00000175826 | -0.902 | 4.16 | -3.399 | 0.00100 | 0.00100 | -1.169 | CTDNEP1 | CTD nuclear envelope phosphatase 1 [Source:HGNC Symbol;Acc:HGNC:19085] |
| ENSG00000118777 | 3.077 | 3.748 | 3.393 | 0.00100 | 0.00100 | -0.982 | ABCG2 | ATP binding cassette subfamily G member 2 (Junior blood group) [Source:HGNC Symbol;Acc:HGNC:74] |
| ENSG00000182504 | 1.284 | 6.561 | 3.387 | 0.00100 | 0.00100 | -1.015 | CEP97 | centrosomal protein 97 [Source:HGNC Symbol;Acc:HGNC:26244] |
| ENSG00000221994 | 0.766 | 2.518 | 3.377 | 0.00100 | 0.00100 | -1.283 | ZNF630 | zinc finger protein 630 [Source:HGNC Symbol;Acc:HGNC:28855] |
| ENSG00000154217 | 1.605 | 3.026 | 3.362 | 0.00100 | 0.00100 | -1.117 | PITPNC1 | phosphatidylinositol transfer protein, cytoplasmic 1 [Source:HGNC Symbol;Acc:HGNC:21045] |
| ENSG00000172301 | 0.874 | 3.639 | 3.36 | 0.00100 | 0.00100 | -1.113 | COPRS | coordinator of PRMT5 and differentiation stimulator [Source:HGNC Symbol;Acc:HGNC:28848] |
| ENSG00000197362 | 0.89 | 2.373 | 3.351 | 0.00100 | 0.00100 | -1.383 | ZNF786 | zinc finger protein 786 [Source:HGNC Symbol;Acc:HGNC:21806] |
| ENSG00000174437 | -1.174 | 7.192 | -3.344 | 0.00100 | 0.00100 | -1.087 | ATP2A2 | ATPase sarcoplasmic/endoplasmic reticulum Ca2+ transporting 2 [Source:HGNC Symbol;Acc:HGNC:812] |
| ENSG00000139160 | 0.936 | 2.905 | 3.344 | 0.00100 | 0.00100 | -1.241 | ETFBKMT | electron transfer flavoprotein beta subunit lysine methyltransferase [Source:HGNC Symbol;Acc:HGNC:28739] |
| ENSG00000163235 | 2.269 | 1.254 | 3.33 | 0.00100 | 0.00100 | -1.405 | TGFA | transforming growth factor alpha [Source:HGNC Symbol;Acc:HGNC:11765] |
| ENSG00000108523 | -0.784 | 4.938 | -3.329 | 0.00100 | 0.00100 | -1.202 | RNF167 | ring finger protein 167 [Source:HGNC Symbol;Acc:HGNC:24544] |
| ENSG00000166579 | -0.782 | 6.521 | -3.309 | 0.00100 | 0.00100 | -1.175 | NDEL1 | nudE neurodevelopment protein 1 like 1 [Source:HGNC Symbol;Acc:HGNC:17620] |
| ENSG00000074755 | -0.929 | 6.479 | -3.306 | 0.00100 | 0.00100 | -1.183 | ZZEF1 | zinc finger ZZ-type and EF-hand domain containing 1 [Source:HGNC Symbol;Acc:HGNC:29027] |
| ENSG00000168246 | 0.94 | 3.09 | 3.305 | 0.00100 | 0.00100 | -1.3 | UBTD2 | ubiquitin domain containing 2 [Source:HGNC Symbol;Acc:HGNC:24463] |
| ENSG00000155868 | 0.632 | 4.215 | 3.3 | 0.00100 | 0.00100 | -1.229 | MED7 | mediator complex subunit 7 [Source:HGNC Symbol;Acc:HGNC:2378] |
| ENSG00000044574 | -1.113 | 10.683 | -3.3 | 0.00100 | 0.00100 | -1.338 | HSPA5 | heat shock protein family A (Hsp70) member 5 [Source:HGNC Symbol;Acc:HGNC:5238] |
| ENSG00000132507 | -0.994 | 6.479 | -3.3 | 0.00100 | 0.00100 | -1.199 | EIF5A | eukaryotic translation initiation factor 5A [Source:HGNC Symbol;Acc:HGNC:3300] |
| ENSG00000272602 | 2.271 | 2.24 | 3.3 | 0.00100 | 0.00100 | -1.303 | ZNF595 | zinc finger protein 595 [Source:HGNC Symbol;Acc:HGNC:27196] |
| ENSG00000155858 | 0.861 | 2.307 | 3.275 | 0.00200 | 0.00200 | -1.541 | LSM11 | LSM11, U7 small nuclear RNA associated [Source:HGNC Symbol;Acc:HGNC:30860] |
| ENSG00000132522 | -0.634 | 6.09 | -3.274 | 0.00200 | 0.00200 | -1.263 | GPS2 | G protein pathway suppressor 2 [Source:HGNC Symbol;Acc:HGNC:4550] |

| | | | | | | | | |
|---|---|---|---|---|---|---|---|---|
| ENSG00000078237 | 1.059 | 3.246 | 3.268 | 0.00200 | 0.00200 | -1.349 | TIGAR | TP53 induced glycolysis regulatory phosphatase [Source:HGNC Symbol;Acc:HGNC:1185] |
| ENSG00000064652 | 1.089 | 2.608 | 3.264 | 0.00200 | 0.00200 | -1.454 | SNX24 | sorting nexin 24 [Source:HGNC Symbol;Acc:HGNC:21533] |
| ENSG00000182944 | -0.77 | 8.173 | -3.261 | 0.00200 | 0.00200 | -1.314 | EWSR1 | EWS RNA binding protein 1 [Source:HGNC Symbol;Acc:HGNC:3508] |
| ENSG00000121931 | 0.89 | 5.327 | 3.253 | 0.00200 | 0.00200 | -1.31 | LRIF1 | ligand dependent nuclear receptor interacting factor 1 [Source:HGNC Symbol;Acc:HGNC:30299] |
| ENSG00000087448 | 1.009 | 4.159 | 3.25 | 0.00200 | 0.00200 | -1.338 | KLHL42 | kelch like family member 42 [Source:HGNC Symbol;Acc:HGNC:29252] |
| ENSG00000119408 | 1.585 | 3.919 | 3.221 | 0.00200 | 0.00200 | -1.404 | NEK6 | NIMA related kinase 6 [Source:HGNC Symbol;Acc:HGNC:7749] |
| ENSG00000123119 | 1.72 | 3.075 | 3.203 | 0.00200 | 0.00200 | -1.481 | NECAB1 | N-terminal EF-hand calcium binding protein 1 [Source:HGNC Symbol;Acc:HGNC:20983] |
| ENSG00000205707 | 0.939 | 5.135 | 3.191 | 0.00200 | 0.00200 | -1.461 | ETFRF1 | electron transfer flavoprotein regulatory factor 1 [Source:HGNC Symbol;Acc:HGNC:27052] |
| ENSG00000088247 | -0.831 | 5.83 | -3.188 | 0.00200 | 0.00200 | -1.481 | KHSRP | KH-type splicing regulatory protein [Source:HGNC Symbol;Acc:HGNC:6316] |
| ENSG00000118454 | 0.617 | 5.733 | 3.175 | 0.00200 | 0.00200 | -1.498 | ANKRD13C | ankyrin repeat domain 13C [Source:HGNC Symbol;Acc:HGNC:25374] |
| ENSG00000130714 | 0.812 | 4.129 | 3.155 | 0.00200 | 0.00200 | -1.57 | POMT1 | protein O-mannosyltransferase 1 [Source:HGNC Symbol;Acc:HGNC:9202] |
| ENSG00000172375 | -0.796 | 4.143 | -3.15 | 0.00200 | 0.00200 | -1.711 | C2CD2L | C2CD2 like [Source:HGNC Symbol;Acc:HGNC:29000] |
| ENSG00000225190 | -0.972 | 4.438 | -3.135 | 0.00200 | 0.00200 | -1.725 | PLEKHM1 | pleckstrin homology and RUN domain containing M1 [Source:HGNC Symbol;Acc:HGNC:29017] |
| ENSG00000124151 | 0.891 | 8.635 | 3.106 | 0.00300 | 0.00300 | -1.752 | NCOA3 | nuclear receptor coactivator 3 [Source:HGNC Symbol;Acc:HGNC:7670] |
| ENSG00000101400 | 1.152 | 1.774 | 3.091 | 0.00300 | 0.00300 | -1.994 | SNTA1 | syntrophin alpha 1 [Source:HGNC Symbol;Acc:HGNC:11167] |
| ENSG00000242616 | 1.113 | 4.047 | 3.087 | 0.00300 | 0.00300 | -1.72 | GNG10 | G protein subunit gamma 10 [Source:HGNC Symbol;Acc:HGNC:4402] |
| ENSG00000108963 | -0.599 | 5.211 | -3.084 | 0.00300 | 0.00300 | -1.738 | DPH1 | diphthamide biosynthesis 1 [Source:HGNC Symbol;Acc:HGNC:3003] |
| ENSG00000179873 | 2.204 | 0.734 | 3.076 | 0.00300 | 0.00300 | -2.16 | NLRP11 | NLR family pyrin domain containing 11 [Source:HGNC Symbol;Acc:HGNC:22945] |
| ENSG00000173258 | 2.046 | 0.047 | 3.054 | 0.00300 | 0.00300 | -2.387 | ZNF483 | zinc finger protein 483 [Source:HGNC Symbol;Acc:HGNC:23384] |
| ENSG00000241697 | 2.26 | 3.76 | 3.041 | 0.00300 | 0.00300 | -1.817 | TMEFF1 | transmembrane protein with EGF like and two follistatin like domains 1 [Source:HGNC Symbol;Acc:HGNC:11866] |
| ENSG00000244115 | 0.808 | 4.525 | 3.028 | 0.00300 | 0.00300 | -1.849 | DNAJC25-GNG10 | DNAJC25-GNG10 readthrough [Source:HGNC Symbol;Acc:HGNC:37501] |
| ENSG00000070366 | -0.901 | 5.166 | -3.021 | 0.00300 | 0.00300 | -1.896 | SMG6 | SMG6, nonsense mediated mRNA decay factor [Source:HGNC Symbol;Acc:HGNC:17809] |
| ENSG00000116791 | 1.036 | 4.286 | 3.013 | 0.00300 | 0.00300 | -1.884 | CRYZ | crystallin zeta [Source:HGNC Symbol;Acc:HGNC:2419] |
| ENSG00000251349 | 2.02 | 4.228 | 3.01 | 0.00300 | 0.00300 | -1.884 | MSANTD3-TMEFF1 | MSANTD3-TMEFF1 readthrough [Source:HGNC Symbol;Acc:HGNC:38838] |
| ENSG00000103522 | 2.128 | 1.111 | 3.009 | 0.00300 | 0.00300 | -2.145 | IL21R | interleukin 21 receptor [Source:HGNC Symbol;Acc:HGNC:6006] |
| ENSG00000164440 | 1.6 | 4.267 | 3.004 | 0.00400 | 0.00400 | -1.898 | TXLNB | taxilin beta [Source:HGNC Symbol;Acc:HGNC:21617] |
| ENSG00000182463 | 2.631 | 2.525 | 3.004 | 0.00400 | 0.00400 | -1.935 | TSHZ2 | teashirt zinc finger homeobox 2 [Source:HGNC Symbol;Acc:HGNC:13010] |
| ENSG00000167074 | 0.941 | 2.805 | 3.004 | 0.00400 | 0.00400 | -2.01 | TEF | TEF, PAR bZIP transcription factor [Source:HGNC Symbol;Acc:HGNC:11722] |
| ENSG00000170681 | 1.769 | 0.541 | 2.999 | 0.00400 | 0.00400 | -2.44 | CAVIN4 | caveolae associated protein 4 [Source:HGNC Symbol;Acc:HGNC:33742] |
| ENSG00000005436 | 0.626 | 5.031 | 2.997 | 0.00400 | 0.00400 | -1.914 | GCFC2 | GC-rich sequence DNA-binding factor 2 [Source:HGNC Symbol;Acc:HGNC:1317] |
| ENSG00000176714 | 0.931 | 2.036 | 2.986 | 0.00400 | 0.00400 | -2.173 | CCDC121 | coiled-coil domain containing 121 [Source:HGNC Symbol;Acc:HGNC:25833] |
| ENSG00000134202 | 2.39 | 0.761 | 2.983 | 0.00400 | 0.00400 | -2.313 | GSTM3 | glutathione S-transferase mu 3 [Source:HGNC Symbol;Acc:HGNC:4635] |
| ENSG00000058799 | 0.775 | 4.315 | 2.979 | 0.00400 | 0.00400 | -1.965 | YIPF1 | Yip1 domain family member 1 [Source:HGNC Symbol;Acc:HGNC:25231] |
| ENSG00000118307 | 1.229 | 0.168 | 2.971 | 0.00400 | 0.00400 | -2.754 | CASC1 | cancer susceptibility 1 [Source:HGNC Symbol;Acc:HGNC:29599] |
| ENSG00000198839 | 0.844 | 5.483 | 2.956 | 0.00400 | 0.00400 | -2.004 | ZNF277 | zinc finger protein 277 [Source:HGNC Symbol;Acc:HGNC:13070] |
| ENSG00000161036 | 0.88 | 3.527 | 2.955 | 0.00400 | 0.00400 | -2.047 | LRWD1 | leucine rich repeats and WD repeat domain containing 1 [Source:HGNC Symbol;Acc:HGNC:21769] |
| ENSG00000181315 | 0.853 | 3.615 | 2.95 | 0.00400 | 0.00400 | -2.052 | ZNF322 | zinc finger protein 322 [Source:HGNC Symbol;Acc:HGNC:23640] |
| ENSG00000169213 | 2.02 | 3.296 | 2.934 | 0.00400 | 0.00400 | -2.07 | RAB3B | RAB3B, member RAS oncogene family [Source:HGNC Symbol;Acc:HGNC:9778] |
| ENSG00000262302 | -1.097 | 1.753 | -2.93 | 0.00400 | 0.00400 | -2.952 | AC003688.1 | |
| ENSG00000022840 | -0.695 | 6.6 | -2.928 | 0.00400 | 0.00400 | -2.066 | RNF10 | ring finger protein 10 [Source:HGNC Symbol;Acc:HGNC:10055] |
| ENSG00000109762 | 0.828 | 6.329 | 2.921 | 0.00400 | 0.00400 | -2.087 | SNX25 | sorting nexin 25 [Source:HGNC Symbol;Acc:HGNC:21883] |
| ENSG00000070444 | -1.348 | 3.858 | -2.908 | 0.00500 | 0.00500 | -2.367 | MNT | MAX network transcriptional repressor [Source:HGNC Symbol;Acc:HGNC:7188] |
| ENSG00000170291 | -0.741 | 4.101 | -2.905 | 0.00500 | 0.00500 | -2.231 | ELP5 | elongator acetyltransferase complex subunit 5 [Source:HGNC Symbol;Acc:HGNC:30617] |
| ENSG00000108953 | -0.946 | 7.46 | -2.9 | 0.00500 | 0.00500 | -2.129 | YWHAE | tyrosine 3-monooxygenase/tryptophan 5-monooxygenase activation protein epsilon [Source:HGNC Symbol;Acc:HGNC:12851] |
| ENSG00000128731 | -0.901 | 6.713 | -2.897 | 0.00500 | 0.00500 | -2.134 | HERC2 | HECT and RLD domain containing E3 ubiquitin protein ligase 2 [Source:HGNC Symbol;Acc:HGNC:4868] |
| ENSG00000219438 | 2.749 | 1.532 | 2.891 | 0.00500 | 0.00500 | -2.241 | FAM19A5 | family with sequence similarity 19 member A5, C-C motif chemokine like [Source:HGNC Symbol;Acc:HGNC:21592] |

| | | | | | | | | |
|---|---|---|---|---|---|---|---|---|
| ENSG00000107779 | 1.511 | 2.201 | 2.887 | 0.00500 | 0.00500 | -2.279 | BMPR1A | bone morphogenetic protein receptor type 1A [Source:HGNC Symbol;Acc:HGNC:1076] |
| ENSG00000167721 | -0.639 | 6.345 | -2.886 | 0.00500 | 0.00500 | -2.16 | TSR1 | TSR1, ribosome maturation factor [Source:HGNC Symbol;Acc:HGNC:25542] |
| ENSG00000267561 | 0.942 | 4.344 | 2.885 | 0.00500 | 0.00500 | -2.168 | AC093155.3 | |
| ENSG00000128266 | 1.912 | 1.503 | 2.88 | 0.00500 | 0.00500 | -2.355 | GNAZ | G protein subunit alpha z [Source:HGNC Symbol;Acc:HGNC:4395] |
| ENSG00000134684 | -0.802 | 6.675 | -2.872 | 0.00500 | 0.00500 | -2.19 | YARS | tyrosyl-tRNA synthetase [Source:HGNC Symbol;Acc:HGNC:12840] |
| ENSG00000177599 | 1.182 | 1.037 | 2.872 | 0.00500 | 0.00500 | -2.602 | ZNF491 | zinc finger protein 491 [Source:HGNC Symbol;Acc:HGNC:23706] |
| ENSG00000161960 | -0.712 | 9.119 | -2.864 | 0.00500 | 0.00500 | -2.241 | EIF4A1 | eukaryotic translation initiation factor 4A1 [Source:HGNC Symbol;Acc:HGNC:3282] |
| ENSG00000118432 | 2.556 | 0.431 | 2.859 | 0.00500 | 0.00500 | -2.525 | CNR1 | cannabinoid receptor 1 [Source:HGNC Symbol;Acc:HGNC:2159] |
| ENSG00000277632 | -3.128 | 7.161 | -2.858 | 0.00500 | 0.00500 | -2.242 | CCL3 | C-C motif chemokine ligand 3 [Source:HGNC Symbol;Acc:HGNC:10627] |
| ENSG00000180543 | 1.894 | 2.323 | 2.849 | 0.00600 | 0.00600 | -2.31 | TSPYL5 | TSPY like 5 [Source:HGNC Symbol;Acc:HGNC:29367] |
| ENSG00000157191 | 0.737 | 4.819 | 2.84 | 0.00600 | 0.00600 | -2.263 | NECAP2 | NECAP endocytosis associated 2 [Source:HGNC Symbol;Acc:HGNC:25528] |
| ENSG00000182095 | -1.057 | 4.176 | -2.84 | 0.00600 | 0.00600 | -2.385 | TNRC18 | trinucleotide repeat containing 18 [Source:HGNC Symbol;Acc:HGNC:11962] |
| ENSG00000272414 | 1.585 | 3.958 | 2.835 | 0.00600 | 0.00600 | -2.278 | FAM47E-STBD1 | FAM47E-STBD1 readthrough [Source:HGNC Symbol;Acc:HGNC:44667] |
| ENSG00000073350 | 1.058 | 3.174 | 2.834 | 0.00600 | 0.00600 | -2.317 | LLGL2 | LLGL2, scribble cell polarity complex component [Source:HGNC Symbol;Acc:HGNC:6629] |
| ENSG00000124193 | -0.688 | 8.322 | -2.832 | 0.00600 | 0.00600 | -2.29 | SRSF6 | serine and arginine rich splicing factor 6 [Source:HGNC Symbol;Acc:HGNC:10788] |
| ENSG00000146833 | 0.663 | 5.116 | 2.828 | 0.00600 | 0.00600 | -2.287 | TRIM4 | tripartite motif containing 4 [Source:HGNC Symbol;Acc:HGNC:16275] |
| ENSG00000125844 | -1.324 | 9.203 | -2.828 | 0.00600 | 0.00600 | -2.307 | RRBP1 | ribosome binding protein 1 [Source:HGNC Symbol;Acc:HGNC:10448] |
| ENSG00000167522 | -1.09 | 6.516 | -2.824 | 0.00600 | 0.00600 | -2.297 | ANKRD11 | ankyrin repeat domain 11 [Source:HGNC Symbol;Acc:HGNC:21316] |
| ENSG00000162755 | 1.115 | 0.704 | 2.816 | 0.00600 | 0.00600 | -2.847 | KLHDC9 | kelch domain containing 9 [Source:HGNC Symbol;Acc:HGNC:28489] |
| ENSG00000171574 | 0.848 | 1.217 | 2.814 | 0.00600 | 0.00600 | -2.734 | ZNF584 | zinc finger protein 584 [Source:HGNC Symbol;Acc:HGNC:27318] |
| ENSG00000118513 | 1.604 | 2.739 | 2.812 | 0.00600 | 0.00600 | -2.365 | MYB | MYB proto-oncogene, transcription factor [Source:HGNC Symbol;Acc:HGNC:7545] |
| ENSG00000136068 | 1.656 | 4.584 | 2.809 | 0.00600 | 0.00600 | -2.328 | FLNB | filamin B [Source:HGNC Symbol;Acc:HGNC:3755] |
| ENSG00000134461 | 0.757 | 3.011 | 2.786 | 0.00700 | 0.00700 | -2.444 | ANKRD16 | ankyrin repeat domain 16 [Source:HGNC Symbol;Acc:HGNC:23471] |
| ENSG00000127191 | 0.785 | 3.048 | 2.784 | 0.00700 | 0.00700 | -2.443 | TRAF2 | TNF receptor associated factor 2 [Source:HGNC Symbol;Acc:HGNC:12032] |
| ENSG00000169564 | -0.709 | 5.859 | -2.777 | 0.00700 | 0.00700 | -2.4 | PCBP1 | poly(rC) binding protein 1 [Source:HGNC Symbol;Acc:HGNC:8647] |
| ENSG00000108559 | -0.557 | 6.173 | -2.772 | 0.00700 | 0.00700 | -2.409 | NUP88 | nucleoporin 88 [Source:HGNC Symbol;Acc:HGNC:8067] |
| ENSG00000178904 | -0.738 | 5.523 | -2.763 | 0.00700 | 0.00700 | -2.434 | DPY19L3 | dpy-19 like 3 (C. elegans) [Source:HGNC Symbol;Acc:HGNC:27120] |
| ENSG00000141503 | -0.935 | 4.418 | -2.758 | 0.00700 | 0.00700 | -2.503 | MINK1 | misshapen like kinase 1 [Source:HGNC Symbol;Acc:HGNC:17565] |
| ENSG00000148840 | -0.955 | 5.044 | -2.756 | 0.00700 | 0.00700 | -2.468 | PPRC1 | peroxisome proliferator-activated receptor gamma, coactivator-related 1 [Source:HGNC Symbol;Acc:HGNC:30025] |
| ENSG00000085491 | 0.827 | 4.951 | 2.75 | 0.00700 | 0.00700 | -2.455 | SLC25A24 | solute carrier family 25 member 24 [Source:HGNC Symbol;Acc:HGNC:20662] |
| ENSG00000241973 | -0.775 | 5.505 | -2.745 | 0.00700 | 0.00700 | -2.473 | PI4KA | phosphatidylinositol 4-kinase alpha [Source:HGNC Symbol;Acc:HGNC:8983] |
| ENSG00000144026 | 0.981 | 4.124 | 2.743 | 0.00700 | 0.00700 | -2.476 | ZNF514 | zinc finger protein 514 [Source:HGNC Symbol;Acc:HGNC:25894] |
| ENSG00000133818 | 2.502 | 3.139 | 2.743 | 0.00700 | 0.00700 | -2.477 | RRAS2 | related RAS viral (r-ras) oncogene homolog 2 [Source:HGNC Symbol;Acc:HGNC:17271] |
| ENSG00000119471 | 0.875 | 5.221 | 2.742 | 0.00700 | 0.00700 | -2.472 | HSDL2 | hydroxysteroid dehydrogenase like 2 [Source:HGNC Symbol;Acc:HGNC:18572] |
| ENSG00000108179 | -0.979 | 5.134 | -2.738 | 0.00800 | 0.00800 | -2.502 | PPIF | peptidylprolyl isomerase F [Source:HGNC Symbol;Acc:HGNC:9259] |
| ENSG00000002919 | 0.572 | 3.176 | 2.737 | 0.00800 | 0.00800 | -2.539 | SNX11 | sorting nexin 11 [Source:HGNC Symbol;Acc:HGNC:14975] |
| ENSG00000076984 | -0.709 | 4.506 | -2.731 | 0.00800 | 0.00800 | -2.533 | MAP2K7 | mitogen-activated protein kinase kinase 7 [Source:HGNC Symbol;Acc:HGNC:6847] |
| ENSG00000076928 | -0.809 | 5.335 | -2.73 | 0.00800 | 0.00800 | -2.509 | ARHGEF1 | Rho guanine nucleotide exchange factor 1 [Source:HGNC Symbol;Acc:HGNC:681] |
| ENSG00000170448 | -0.671 | 5.217 | -2.726 | 0.00800 | 0.00800 | -2.515 | NFXL1 | nuclear transcription factor, X-box binding like 1 [Source:HGNC Symbol;Acc:HGNC:18726] |
| ENSG00000175854 | 0.666 | 2.942 | 2.725 | 0.00800 | 0.00800 | -2.569 | SWI5 | SWI5 homologous recombination repair protein [Source:HGNC Symbol;Acc:HGNC:31412] |
| ENSG00000105499 | 1.395 | 1.067 | 2.718 | 0.00800 | 0.00800 | -2.806 | PLA2G4C | phospholipase A2 group IVC [Source:HGNC Symbol;Acc:HGNC:9037] |
| ENSG00000160856 | 2.026 | 2.654 | 2.718 | 0.00800 | 0.00800 | -2.551 | FCRL3 | Fc receptor like 3 [Source:HGNC Symbol;Acc:HGNC:18506] |
| ENSG00000018869 | 1.627 | 0.44 | 2.716 | 0.00800 | 0.00800 | -2.929 | ZNF582 | zinc finger protein 582 [Source:HGNC Symbol;Acc:HGNC:26421] |
| ENSG00000186130 | 0.771 | 4.713 | 2.703 | 0.00800 | 0.00800 | -2.556 | ZBTB6 | zinc finger and BTB domain containing 6 [Source:HGNC Symbol;Acc:HGNC:16764] |
| ENSG00000132361 | -1.097 | 4.617 | -2.702 | 0.00800 | 0.00800 | -2.608 | CLUH | clustered mitochondria homolog [Source:HGNC Symbol;Acc:HGNC:29094] |
| ENSG00000106701 | 1.154 | 2.87 | 2.699 | 0.00800 | 0.00800 | -2.602 | FSD1L | fibronectin type III and SPRY domain containing 1 like [Source:HGNC Symbol;Acc:HGNC:13753] |

| | | | | | | | | |
|---|---|---|---|---|---|---|---|---|
| ENSG00000180938 | 0.915 | 1.435 | 2.698 | 0.00800 | 0.00800 | -2.824 | ZNF572 | zinc finger protein 572 [Source:HGNC Symbol;Acc:HGNC:26758] |
| ENSG00000106636 | -0.6 | 6.181 | -2.696 | 0.00800 | 0.00800 | -2.568 | YKT6 | YKT6 v-SNARE homolog [Source:HGNC Symbol;Acc:HGNC:16959] |
| ENSG00000059588 | -0.976 | 5.315 | -2.695 | 0.00900 | 0.00900 | -2.583 | TARBP1 | TAR (HIV-1) RNA binding protein 1 [Source:HGNC Symbol;Acc:HGNC:11568] |
| ENSG00000186625 | 0.529 | 3.585 | 2.689 | 0.00900 | 0.00900 | -2.608 | KATNA1 | katanin catalytic subunit A1 [Source:HGNC Symbol;Acc:HGNC:6216] |
| ENSG00000127995 | 0.751 | 5.312 | 2.686 | 0.00900 | 0.00900 | -2.589 | CASD1 | CAS1 domain containing 1 [Source:HGNC Symbol;Acc:HGNC:16014] |
| ENSG00000141447 | 2.322 | 3.051 | 2.685 | 0.00900 | 0.00900 | -2.6 | OSBPL1A | oxysterol binding protein like 1A [Source:HGNC Symbol;Acc:HGNC:16398] |
| ENSG00000181222 | -1.066 | 5.98 | -2.684 | 0.00900 | 0.00900 | -2.597 | POLR2A | RNA polymerase II subunit A [Source:HGNC Symbol;Acc:HGNC:9187] |
| ENSG00000149657 | 0.46 | 4.457 | 2.683 | 0.00900 | 0.00900 | -2.602 | LSM14B | LSM family member 14B [Source:HGNC Symbol;Acc:HGNC:15887] |
| ENSG00000100219 | 0.927 | 10.218 | 2.676 | 0.00900 | 0.00900 | -2.692 | XBP1 | X-box binding protein 1 [Source:HGNC Symbol;Acc:HGNC:12801] |
| ENSG00000163923 | 1.008 | 2.037 | 2.675 | 0.00900 | 0.00900 | -2.747 | RPL39L | ribosomal protein L39 like [Source:HGNC Symbol;Acc:HGNC:17094] |
| ENSG00000108468 | 0.729 | 4.595 | 2.675 | 0.00900 | 0.00900 | -2.614 | CBX1 | chromobox 1 [Source:HGNC Symbol;Acc:HGNC:1551] |
| ENSG00000153936 | 0.95 | 6.378 | 2.674 | 0.00900 | 0.00900 | -2.617 | HS2ST1 | heparan sulfate 2-O-sulfotransferase 1 [Source:HGNC Symbol;Acc:HGNC:5193] |
| ENSG00000166716 | -0.991 | 5.187 | -2.672 | 0.00900 | 0.00900 | -2.634 | ZNF592 | zinc finger protein 592 [Source:HGNC Symbol;Acc:HGNC:28986] |
| ENSG00000196652 | 0.444 | 4.584 | 2.667 | 0.00900 | 0.00900 | -2.632 | ZKSCAN5 | zinc finger with KRAB and SCAN domains 5 [Source:HGNC Symbol;Acc:HGNC:12867] |
| ENSG00000125945 | 0.63 | 3.417 | 2.664 | 0.00900 | 0.00900 | -2.662 | ZNF436 | zinc finger protein 436 [Source:HGNC Symbol;Acc:HGNC:20814] |
| ENSG00000138443 | 0.788 | 5.669 | 2.66 | 0.00900 | 0.00900 | -2.643 | ABI2 | abl interactor 2 [Source:HGNC Symbol;Acc:HGNC:24011] |
| ENSG00000115806 | -0.662 | 6.931 | -2.66 | 0.00900 | 0.00900 | -2.643 | GORASP2 | golgi reassembly stacking protein 2 [Source:HGNC Symbol;Acc:HGNC:17500] |
| ENSG00000105227 | 1.275 | 1.998 | 2.659 | 0.00900 | 0.00900 | -2.757 | PRX | periaxin [Source:HGNC Symbol;Acc:HGNC:13797] |
| ENSG00000197841 | 0.876 | 4.195 | 2.653 | 0.01000 | 0.01000 | -2.661 | ZNF181 | zinc finger protein 181 [Source:HGNC Symbol;Acc:HGNC:12971] |
| ENSG00000129757 | 1.564 | 1.347 | 2.648 | 0.01000 | 0.01000 | -2.814 | CDKN1C | cyclin dependent kinase inhibitor 1C [Source:HGNC Symbol;Acc:HGNC:1786] |
| ENSG00000120008 | 0.896 | 6.626 | 2.64 | 0.01000 | 0.01000 | -2.687 | WDR11 | WD repeat domain 11 [Source:HGNC Symbol;Acc:HGNC:13831] |
| ENSG00000168389 | -1.757 | 3.303 | -2.636 | 0.01000 | 0.01000 | -2.985 | MFSD2A | major facilitator superfamily domain containing 2A [Source:HGNC Symbol;Acc:HGNC:25897] |
| ENSG00000177058 | 0.631 | 4.992 | 2.635 | 0.01000 | 0.01000 | -2.695 | SLC38A9 | solute carrier family 38 member 9 [Source:HGNC Symbol;Acc:HGNC:26907] |
| ENSG00000275111 | 0.746 | 1.681 | 2.634 | 0.01000 | 0.01000 | -2.897 | ZNF2 | zinc finger protein 2 [Source:HGNC Symbol;Acc:HGNC:12991] |
| ENSG00000100151 | -0.809 | 3.332 | -2.634 | 0.01000 | 0.01000 | -2.844 | PICK1 | protein interacting with PRKCA 1 [Source:HGNC Symbol;Acc:HGNC:9394] |
| ENSG00000168502 | 1.653 | 1.48 | 2.632 | 0.01000 | 0.01000 | -2.84 | MTCL1 | microtubule crosslinking factor 1 [Source:HGNC Symbol;Acc:HGNC:29121] |
| ENSG00000069424 | -0.962 | 4.35 | -2.631 | 0.01000 | 0.01000 | -2.748 | KCNAB2 | potassium voltage-gated channel subfamily A regulatory beta subunit 2 [Source:HGNC Symbol;Acc:HGNC:6229] |
| ENSG00000050748 | 0.479 | 4.732 | 2.63 | 0.01000 | 0.01000 | -2.706 | MAPK9 | mitogen-activated protein kinase 9 [Source:HGNC Symbol;Acc:HGNC:6886] |
| ENSG00000149658 | -0.586 | 5.361 | -2.628 | 0.01000 | 0.01000 | -2.712 | YTHDF1 | YTH N6-methyladenosine RNA binding protein 1 [Source:HGNC Symbol;Acc:HGNC:15867] |
| ENSG00000214530 | 0.92 | 3.566 | 2.624 | 0.01000 | 0.01000 | -2.73 | STARD10 | StAR related lipid transfer domain containing 10 [Source:HGNC Symbol;Acc:HGNC:10666] |
| ENSG00000138131 | 1.38 | 1.489 | 2.617 | 0.01100 | 0.01100 | -2.961 | LOXL4 | lysyl oxidase like 4 [Source:HGNC Symbol;Acc:HGNC:17171] |
| ENSG00000134253 | 0.85 | 1.374 | 2.616 | 0.01100 | 0.01100 | -3.025 | TRIM45 | tripartite motif containing 45 [Source:HGNC Symbol;Acc:HGNC:19018] |
| ENSG00000161551 | 0.96 | 3.311 | 2.611 | 0.01100 | 0.01100 | -2.76 | ZNF577 | zinc finger protein 577 [Source:HGNC Symbol;Acc:HGNC:28673] |
| ENSG00000165966 | 1.996 | 3.987 | 2.601 | 0.01100 | 0.01100 | -2.764 | PDZRN4 | PDZ domain containing ring finger 4 [Source:HGNC Symbol;Acc:HGNC:30552] |
| ENSG00000151746 | 0.756 | 5.382 | 2.595 | 0.01100 | 0.01100 | -2.775 | BICD1 | BICD cargo adaptor 1 [Source:HGNC Symbol;Acc:HGNC:1049] |
| ENSG00000169131 | 0.693 | 3.363 | 2.593 | 0.01100 | 0.01100 | -2.801 | ZNF354A | zinc finger protein 354A [Source:HGNC Symbol;Acc:HGNC:11628] |
| ENSG00000130584 | 1.408 | 1.679 | 2.589 | 0.01100 | 0.01100 | -2.907 | ZBTB46 | zinc finger and BTB domain containing 46 [Source:HGNC Symbol;Acc:HGNC:16094] |
| ENSG00000071967 | 1.876 | 2.094 | 2.588 | 0.01100 | 0.01100 | -2.836 | CYBRD1 | cytochrome b reductase 1 [Source:HGNC Symbol;Acc:HGNC:20797] |
| ENSG00000186446 | 1.181 | 0.79 | 2.587 | 0.01100 | 0.01100 | -3.103 | ZNF501 | zinc finger protein 501 [Source:HGNC Symbol;Acc:HGNC:23717] |
| ENSG00000152944 | 0.585 | 5.284 | 2.586 | 0.01100 | 0.01100 | -2.793 | MED21 | mediator complex subunit 21 [Source:HGNC Symbol;Acc:HGNC:11473] |
| ENSG00000277957 | -0.677 | 9.021 | -2.578 | 0.01200 | 0.01200 | -2.82 | SENP3-EIF4A1 | SENP3-EIF4A1 readthrough (NMD candidate) [Source:HGNC Symbol;Acc:HGNC:49182] |
| ENSG00000173588 | -0.634 | 5.65 | -2.578 | 0.01200 | 0.01200 | -2.812 | CEP83 | centrosomal protein 83 [Source:HGNC Symbol;Acc:HGNC:17966] |
| ENSG00000148396 | -0.829 | 6.198 | -2.575 | 0.01200 | 0.01200 | -2.817 | SEC16A | SEC16 homolog A, endoplasmic reticulum export factor [Source:HGNC Symbol;Acc:HGNC:29006] |
| ENSG00000138685 | 1.542 | 4.292 | 2.574 | 0.01200 | 0.01200 | -2.818 | FGF2 | fibroblast growth factor 2 [Source:HGNC Symbol;Acc:HGNC:3676] |
| ENSG00000121749 | -0.739 | 6.7 | -2.573 | 0.01200 | 0.01200 | -2.82 | TBC1D15 | TBC1 domain family member 15 [Source:HGNC Symbol;Acc:HGNC:25694] |
| ENSG00000143653 | 1.679 | 2.949 | 2.573 | 0.01200 | 0.01200 | -2.831 | SCCPDH | saccharopine dehydrogenase (putative) [Source:HGNC Symbol;Acc:HGNC:24275] |

| | | | | | | | | |
|---|---|---|---|---|---|---|---|---|
| ENSG00000172269 | -0.694 | 4.356 | -2.57 | 0.01200 | 0.01200 | -2.852 | DPAGT1 | dolichyl-phosphate N-acetylglucosaminephosphotransferase 1 [Source:HGNC Symbol;Acc:HGNC:2995] |
| ENSG00000204574 | -0.77 | 6.8 | -2.569 | 0.01200 | 0.01200 | -2.828 | ABCF1 | ATP binding cassette subfamily F member 1 [Source:HGNC Symbol;Acc:HGNC:70] |
| ENSG00000174718 | 0.81 | 7.579 | 2.568 | 0.01200 | 0.01200 | -2.837 | KIAA1551 | KIAA1551 [Source:HGNC Symbol;Acc:HGNC:25559] |
| ENSG00000197885 | 0.766 | 4.354 | 2.568 | 0.01200 | 0.01200 | -2.832 | NKIRAS1 | NFKB inhibitor interacting Ras like 1 [Source:HGNC Symbol;Acc:HGNC:17899] |
| ENSG00000259522 | -0.691 | 5.204 | -2.566 | 0.01200 | 0.01200 | -2.839 | AL136295.4 | |
| ENSG00000111203 | -0.7 | 4.816 | -2.559 | 0.01200 | 0.01200 | -2.859 | ITFG2 | integrin alpha FG-GAP repeat containing 2 [Source:HGNC Symbol;Acc:HGNC:30879] |
| ENSG00000274349 | 0.94 | 2.32 | 2.558 | 0.01200 | 0.01200 | -2.921 | ZNF658 | zinc finger protein 658 [Source:HGNC Symbol;Acc:HGNC:25226] |
| ENSG00000182712 | 0.807 | 3.602 | 2.558 | 0.01200 | 0.01200 | -2.859 | CMC4 | C-X9-C motif containing 4 [Source:HGNC Symbol;Acc:HGNC:35428] |
| ENSG00000137497 | -0.545 | 7.938 | -2.555 | 0.01200 | 0.01200 | -2.857 | NUMA1 | nuclear mitotic apparatus protein 1 [Source:HGNC Symbol;Acc:HGNC:8059] |
| ENSG00000123064 | -0.701 | 5.48 | -2.551 | 0.01300 | 0.01300 | -2.865 | DDX54 | DEAD-box helicase 54 [Source:HGNC Symbol;Acc:HGNC:20084] |
| ENSG00000185869 | 1.254 | 1.825 | 2.551 | 0.01300 | 0.01300 | -2.957 | ZNF829 | zinc finger protein 829 [Source:HGNC Symbol;Acc:HGNC:34032] |
| ENSG00000162976 | 0.962 | 4.716 | 2.549 | 0.01300 | 0.01300 | -2.869 | PQLC3 | PQ loop repeat containing 3 [Source:HGNC Symbol;Acc:HGNC:28503] |
| ENSG00000167720 | -0.56 | 4.345 | -2.548 | 0.01300 | 0.01300 | -2.89 | SRR | serine racemase [Source:HGNC Symbol;Acc:HGNC:14398] |
| ENSG00000111424 | -1.079 | 4.468 | -2.546 | 0.01300 | 0.01300 | -2.903 | VDR | vitamin D (1,25- dihydroxyvitamin D3) receptor [Source:HGNC Symbol;Acc:HGNC:12679] |
| ENSG00000148154 | 0.911 | 6.439 | 2.545 | 0.01300 | 0.01300 | -2.877 | UGCG | UDP-glucose ceramide glucosyltransferase [Source:HGNC Symbol;Acc:HGNC:12524] |
| ENSG00000176853 | -0.614 | 6.582 | -2.542 | 0.01300 | 0.01300 | -2.881 | FAM91A1 | family with sequence similarity 91 member A1 [Source:HGNC Symbol;Acc:HGNC:26306] |
| ENSG00000160299 | -0.7 | 6.433 | -2.54 | 0.01300 | 0.01300 | -2.886 | PCNT | pericentrin [Source:HGNC Symbol;Acc:HGNC:16068] |
| ENSG00000111863 | 2.661 | 3.518 | 2.537 | 0.01300 | 0.01300 | -2.892 | ADTRP | androgen dependent TFPI regulating protein [Source:HGNC Symbol;Acc:HGNC:21214] |
| ENSG00000156171 | 0.681 | 5.276 | 2.537 | 0.01300 | 0.01300 | -2.892 | DRAM2 | DNA damage regulated autophagy modulator 2 [Source:HGNC Symbol;Acc:HGNC:28769] |
| ENSG00000142002 | -0.627 | 4.76 | -2.536 | 0.01300 | 0.01300 | -2.902 | DPP9 | dipeptidyl peptidase 9 [Source:HGNC Symbol;Acc:HGNC:18648] |
| ENSG00000164011 | 0.603 | 1.782 | 2.533 | 0.01300 | 0.01300 | -3.082 | ZNF691 | zinc finger protein 691 [Source:HGNC Symbol;Acc:HGNC:28028] |
| ENSG00000214827 | 0.782 | 3.764 | 2.532 | 0.01300 | 0.01300 | -2.907 | MTCP1 | mature T-cell proliferation 1 [Source:HGNC Symbol;Acc:HGNC:7423] |
| ENSG00000141522 | -0.965 | 5.613 | -2.531 | 0.01300 | 0.01300 | -2.906 | ARHGDIA | Rho GDP dissociation inhibitor alpha [Source:HGNC Symbol;Acc:HGNC:678] |
| ENSG00000186272 | 0.69 | 3.457 | 2.53 | 0.01300 | 0.01300 | -2.917 | ZNF17 | zinc finger protein 17 [Source:HGNC Symbol;Acc:HGNC:12958] |
| ENSG00000178921 | -0.935 | 4.166 | -2.529 | 0.01300 | 0.01300 | -2.954 | PFAS | phosphoribosylformylglycinamidine synthase [Source:HGNC Symbol;Acc:HGNC:8863] |
| ENSG00000164978 | 0.649 | 3.931 | 2.525 | 0.01300 | 0.01300 | -2.919 | NUDT2 | nudix hydrolase 2 [Source:HGNC Symbol;Acc:HGNC:8049] |
| ENSG00000148481 | 0.753 | 5.707 | 2.522 | 0.01400 | 0.01400 | -2.922 | MINDY3 | MINDY lysine 48 deubiquitinase 3 [Source:HGNC Symbol;Acc:HGNC:23578] |
| ENSG00000173890 | 1.105 | 6.535 | 2.52 | 0.01400 | 0.01400 | -2.927 | GPR160 | G protein-coupled receptor 160 [Source:HGNC Symbol;Acc:HGNC:23693] |
| ENSG00000141510 | -1.03 | 3.569 | -2.518 | 0.01400 | 0.01400 | -3.044 | TP53 | tumor protein p53 [Source:HGNC Symbol;Acc:HGNC:11998] |
| ENSG00000072501 | -0.748 | 6.109 | -2.517 | 0.01400 | 0.01400 | -2.932 | SMC1A | structural maintenance of chromosomes 1A [Source:HGNC Symbol;Acc:HGNC:11111] |
| ENSG00000198440 | 1.13 | 2.962 | 2.51 | 0.01400 | 0.01400 | -2.96 | ZNF583 | zinc finger protein 583 [Source:HGNC Symbol;Acc:HGNC:26427] |
| ENSG00000117620 | 0.56 | 5.593 | 2.504 | 0.01400 | 0.01400 | -2.957 | SLC35A3 | solute carrier family 35 member A3 [Source:HGNC Symbol;Acc:HGNC:11023] |
| ENSG00000111186 | 1.599 | 3.365 | 2.504 | 0.01400 | 0.01400 | -2.959 | WNT5B | Wnt family member 5B [Source:HGNC Symbol;Acc:HGNC:16265] |
| ENSG00000196497 | -0.775 | 4.906 | -2.502 | 0.01400 | 0.01400 | -2.967 | IPO4 | importin 4 [Source:HGNC Symbol;Acc:HGNC:19426] |
| ENSG00000167562 | 1.416 | 2.659 | 2.502 | 0.01400 | 0.01400 | -2.977 | ZNF701 | zinc finger protein 701 [Source:HGNC Symbol;Acc:HGNC:25597] |
| ENSG00000156973 | 0.507 | 3.199 | 2.499 | 0.01400 | 0.01400 | -2.988 | PDE6D | phosphodiesterase 6D [Source:HGNC Symbol;Acc:HGNC:8788] |
| ENSG00000126773 | 0.549 | 6.279 | 2.497 | 0.01400 | 0.01400 | -2.97 | PCNX4 | pecanex homolog 4 (Drosophila) [Source:HGNC Symbol;Acc:HGNC:20349] |
| ENSG00000109066 | -0.786 | 3.265 | -2.495 | 0.01500 | 0.01500 | -3.085 | TMEM104 | transmembrane protein 104 [Source:HGNC Symbol;Acc:HGNC:25984] |
| ENSG00000139182 | -1.09 | 3.262 | -2.495 | 0.01500 | 0.01500 | -3.143 | CLSTN3 | calsyntenin 3 [Source:HGNC Symbol;Acc:HGNC:18371] |
| ENSG00000131467 | -0.695 | 4.974 | -2.492 | 0.01500 | 0.01500 | -2.985 | PSME3 | proteasome activator subunit 3 [Source:HGNC Symbol;Acc:HGNC:9570] |
| ENSG00000141456 | -1.141 | 3.368 | -2.491 | 0.01500 | 0.01500 | -3.161 | PELP1 | proline, glutamate and leucine rich protein 1 [Source:HGNC Symbol;Acc:HGNC:30134] |
| ENSG00000132254 | -0.574 | 5.778 | -2.487 | 0.01500 | 0.01500 | -2.99 | ARFIP2 | ADP ribosylation factor interacting protein 2 [Source:HGNC Symbol;Acc:HGNC:17160] |
| ENSG00000262664 | -0.489 | 4.64 | -2.487 | 0.01500 | 0.01500 | -2.995 | OVCA2 | OVCA2 serine hydrolase domain containing [Source:HGNC Symbol;Acc:HGNC:24203] |
| ENSG00000106733 | 0.79 | 5.728 | 2.487 | 0.01500 | 0.01500 | -2.992 | NMRK1 | nicotinamide riboside kinase 1 [Source:HGNC Symbol;Acc:HGNC:26057] |
| ENSG00000161204 | -0.549 | 5.544 | -2.484 | 0.01500 | 0.01500 | -2.996 | ABCF3 | ATP binding cassette subfamily F member 3 [Source:HGNC Symbol;Acc:HGNC:72] |
| ENSG00000109775 | 0.703 | 5.909 | 2.481 | 0.01500 | 0.01500 | -3.002 | UFSP2 | UFM1 specific peptidase 2 [Source:HGNC Symbol;Acc:HGNC:25640] |

| | | | | | | | | |
|---|---|---|---|---|---|---|---|---|
| ENSG00000122378 | 1.061 | 4.018 | 2.478 | 0.01500 | 0.01500 | -3.007 | FAM213A | family with sequence similarity 213 member A [Source:HGNC Symbol;Acc:HGNC:28651] |
| ENSG00000141985 | -0.863 | 4.097 | -2.473 | 0.01500 | 0.01500 | -3.051 | SH3GL1 | SH3 domain containing GRB2 like 1, endophilin A2 [Source:HGNC Symbol;Acc:HGNC:10830] |
| ENSG00000175224 | -0.519 | 6.02 | -2.472 | 0.01500 | 0.01500 | -3.02 | ATG13 | autophagy related 13 [Source:HGNC Symbol;Acc:HGNC:29091] |
| ENSG00000149532 | -0.634 | 5.177 | -2.468 | 0.01600 | 0.01600 | -3.027 | CPSF7 | cleavage and polyadenylation specific factor 7 [Source:HGNC Symbol;Acc:HGNC:30098] |
| ENSG00000118804 | 1.647 | 4.012 | 2.466 | 0.01600 | 0.01600 | -3.03 | STBD1 | starch binding domain 1 [Source:HGNC Symbol;Acc:HGNC:24854] |
| ENSG00000147138 | 1.312 | 1.388 | 2.464 | 0.01600 | 0.01600 | -3.151 | GPR174 | G protein-coupled receptor 174 [Source:HGNC Symbol;Acc:HGNC:30245] |
| ENSG00000139263 | 2.01 | 2.819 | 2.46 | 0.01600 | 0.01600 | -3.047 | LRIG3 | leucine rich repeats and immunoglobulin like domains 3 [Source:HGNC Symbol;Acc:HGNC:30991] |
| ENSG00000119004 | 0.689 | 6.436 | 2.457 | 0.01600 | 0.01600 | -3.048 | CYP20A1 | cytochrome P450 family 20 subfamily A member 1 [Source:HGNC Symbol;Acc:HGNC:20576] |
| ENSG00000126107 | -0.835 | 4.155 | -2.455 | 0.01600 | 0.01600 | -3.077 | HECTD3 | HECT domain E3 ubiquitin protein ligase 3 [Source:HGNC Symbol;Acc:HGNC:26117] |
| ENSG00000137818 | -1.052 | 9.201 | -2.455 | 0.01600 | 0.01600 | -3.054 | RPLP1 | ribosomal protein lateral stalk subunit P1 [Source:HGNC Symbol;Acc:HGNC:10372] |
| ENSG00000134532 | 1.721 | 1.757 | 2.454 | 0.01600 | 0.01600 | -3.094 | SOX5 | SRY-box 5 [Source:HGNC Symbol;Acc:HGNC:11201] |
| ENSG00000158545 | -0.811 | 4.698 | -2.454 | 0.01600 | 0.01600 | -3.062 | ZC3H18 | zinc finger CCCH-type containing 18 [Source:HGNC Symbol;Acc:HGNC:25091] |
| ENSG00000177272 | 1.22 | 5.705 | 2.453 | 0.01600 | 0.01600 | -3.056 | KCNA3 | potassium voltage-gated channel subfamily A member 3 [Source:HGNC Symbol;Acc:HGNC:6221] |
| ENSG00000137274 | 0.799 | 2.319 | 2.453 | 0.01600 | 0.01600 | -3.12 | BPHL | biphenyl hydrolase like [Source:HGNC Symbol;Acc:HGNC:1094] |
| ENSG00000184743 | 0.697 | 7.055 | 2.448 | 0.01600 | 0.01600 | -3.064 | ATL3 | atlastin GTPase 3 [Source:HGNC Symbol;Acc:HGNC:24526] |
| ENSG00000177565 | 0.69 | 8.311 | 2.448 | 0.01600 | 0.01600 | -3.07 | TBL1XR1 | transducin beta like 1 X-linked receptor 1 [Source:HGNC Symbol;Acc:HGNC:29529] |
| ENSG00000109111 | -0.639 | 6.981 | -2.447 | 0.01600 | 0.01600 | -3.068 | SUPT6H | SPT6 homolog, histone chaperone [Source:HGNC Symbol;Acc:HGNC:11470] |
| ENSG00000134250 | 0.989 | 6.059 | 2.447 | 0.01700 | 0.01700 | -3.068 | NOTCH2 | notch 2 [Source:HGNC Symbol;Acc:HGNC:7882] |
| ENSG00000197619 | 0.84 | 2.823 | 2.445 | 0.01700 | 0.01700 | -3.09 | ZNF615 | zinc finger protein 615 [Source:HGNC Symbol;Acc:HGNC:24740] |
| ENSG00000172845 | 0.46 | 8.298 | 2.445 | 0.01700 | 0.01700 | -3.075 | SP3 | Sp3 transcription factor [Source:HGNC Symbol;Acc:HGNC:11208] |
| ENSG00000184162 | 0.58 | 2.759 | 2.444 | 0.01700 | 0.01700 | -3.106 | NR2C2AP | nuclear receptor 2C2 associated protein [Source:HGNC Symbol;Acc:HGNC:30763] |
| ENSG00000138760 | 0.894 | 8.128 | 2.444 | 0.01700 | 0.01700 | -3.077 | SCARB2 | scavenger receptor class B member 2 [Source:HGNC Symbol;Acc:HGNC:1665] |
| ENSG00000122224 | -1.28 | 5.749 | -2.444 | 0.01700 | 0.01700 | -3.073 | LY9 | lymphocyte antigen 9 [Source:HGNC Symbol;Acc:HGNC:6730] |
| ENSG00000184232 | -1.86 | 3.528 | -2.443 | 0.01700 | 0.01700 | -3.233 | OAF | out at first homolog [Source:HGNC Symbol;Acc:HGNC:28752] |
| ENSG00000138363 | -0.88 | 3.945 | -2.442 | 0.01700 | 0.01700 | -3.113 | ATIC | 5-aminoimidazole-4-carboxamide ribonucleotide formyltransferase/IMP cyclohydrolase [Source:HGNC Symbol;Acc:HGNC:794] |
| ENSG00000151012 | -2.812 | 4.604 | -2.43 | 0.01700 | 0.01700 | -3.25 | SLC7A11 | solute carrier family 7 member 11 [Source:HGNC Symbol;Acc:HGNC:11059] |
| ENSG00000005100 | -0.596 | 4.1 | -2.424 | 0.01800 | 0.01800 | -3.127 | DHX33 | DEAH-box helicase 33 [Source:HGNC Symbol;Acc:HGNC:16718] |
| ENSG00000131115 | 0.661 | 4.442 | 2.424 | 0.01800 | 0.01800 | -3.111 | ZNF227 | zinc finger protein 227 [Source:HGNC Symbol;Acc:HGNC:13020] |
| ENSG00000095209 | 0.81 | 5.179 | 2.421 | 0.01800 | 0.01800 | -3.118 | TMEM38B | transmembrane protein 38B [Source:HGNC Symbol;Acc:HGNC:25535] |
| ENSG00000151148 | -0.529 | 4.873 | -2.421 | 0.01800 | 0.01800 | -3.117 | UBE3B | ubiquitin protein ligase E3B [Source:HGNC Symbol;Acc:HGNC:13478] |
| ENSG00000148153 | 0.694 | 4.683 | 2.421 | 0.01800 | 0.01800 | -3.117 | INIP | INTS3 and NABP interacting protein [Source:HGNC Symbol;Acc:HGNC:24994] |
| ENSG00000131069 | 0.997 | 3.315 | 2.415 | 0.01800 | 0.01800 | -3.13 | ACSS2 | acyl-CoA synthetase short-chain family member 2 [Source:HGNC Symbol;Acc:HGNC:15814] |
| ENSG00000183309 | 0.574 | 4.377 | 2.415 | 0.01800 | 0.01800 | -3.127 | ZNF623 | zinc finger protein 623 [Source:HGNC Symbol;Acc:HGNC:29084] |
| ENSG00000169740 | 0.809 | 3.522 | 2.415 | 0.01800 | 0.01800 | -3.129 | ZNF32 | zinc finger protein 32 [Source:HGNC Symbol;Acc:HGNC:13095] |
| ENSG00000101190 | 1.149 | 2.284 | 2.414 | 0.01800 | 0.01800 | -3.161 | TCFL5 | transcription factor like 5 [Source:HGNC Symbol;Acc:HGNC:11646] |
| ENSG00000072818 | -0.804 | 4.836 | -2.408 | 0.01800 | 0.01800 | -3.143 | ACAP1 | ArfGAP with coiled-coil, ankyrin repeat and PH domains 1 [Source:HGNC Symbol;Acc:HGNC:16467] |
| ENSG00000171806 | 0.685 | 3.265 | 2.4 | 0.01900 | 0.01900 | -3.161 | METTL18 | methyltransferase like 18 [Source:HGNC Symbol;Acc:HGNC:28793] |
| ENSG00000076108 | -0.945 | 6.587 | -2.397 | 0.01900 | 0.01900 | -3.163 | BAZ2A | bromodomain adjacent to zinc finger domain 2A [Source:HGNC Symbol;Acc:HGNC:962] |
| ENSG00000148362 | 0.886 | 2.427 | 2.395 | 0.01900 | 0.01900 | -3.197 | C9orf142 | chromosome 9 open reading frame 142 [Source:HGNC Symbol;Acc:HGNC:27849] |
| ENSG00000070814 | -0.642 | 5.581 | -2.395 | 0.01900 | 0.01900 | -3.166 | TCOF1 | treacle ribosome biogenesis factor 1 [Source:HGNC Symbol;Acc:HGNC:11654] |
| ENSG00000068654 | -0.784 | 5.09 | -2.386 | 0.01900 | 0.01900 | -3.182 | POLR1A | RNA polymerase I subunit A [Source:HGNC Symbol;Acc:HGNC:17264] |
| ENSG00000125551 | 1.528 | -0.104 | 2.384 | 0.01900 | 0.01900 | -3.534 | PLGLB2 | plasminogen-like B2 [Source:HGNC Symbol;Acc:HGNC:9073] |
| ENSG00000198429 | 1.354 | 1.571 | 2.384 | 0.01900 | 0.01900 | -3.272 | ZNF69 | zinc finger protein 69 [Source:HGNC Symbol;Acc:HGNC:13138] |
| ENSG00000079432 | -0.957 | 5.222 | -2.382 | 0.01900 | 0.01900 | -3.188 | CIC | capicua transcriptional repressor [Source:HGNC Symbol;Acc:HGNC:14214] |
| ENSG00000179335 | -0.697 | 6.373 | -2.381 | 0.02000 | 0.02000 | -3.194 | CLK3 | CDC like kinase 3 [Source:HGNC Symbol;Acc:HGNC:2071] |
| ENSG00000061936 | -0.536 | 6.101 | -2.381 | 0.02000 | 0.02000 | -3.194 | SFSWAP | splicing factor SWAP homolog [Source:HGNC Symbol;Acc:HGNC:10790] |

| | | | | | | | | |
|---|---|---|---|---|---|---|---|---|
| ENSG00000048462 | 1.097 | 7.832 | 2.381 | 0.02000 | 0.02000 | -3.192 | TNFRSF17 | TNF receptor superfamily member 17 [Source:HGNC Symbol;Acc:HGNC:11913] |
| ENSG00000153237 | 1.468 | 0.34 | 2.381 | 0.02000 | 0.02000 | -3.521 | CCDC148 | coiled-coil domain containing 148 [Source:HGNC Symbol;Acc:HGNC:25191] |
| ENSG00000115568 | -0.84 | 3.887 | -2.379 | 0.02000 | 0.02000 | -3.227 | ZNF142 | zinc finger protein 142 [Source:HGNC Symbol;Acc:HGNC:12927] |
| ENSG00000112159 | -0.744 | 7.481 | -2.375 | 0.02000 | 0.02000 | -3.204 | MDN1 | midasin AAA ATPase 1 [Source:HGNC Symbol;Acc:HGNC:18302] |
| ENSG00000164086 | -1.002 | 2.104 | -2.374 | 0.02000 | 0.02000 | -3.504 | DUSP7 | dual specificity phosphatase 7 [Source:HGNC Symbol;Acc:HGNC:3073] |
| ENSG00000169989 | 1.286 | 0.027 | 2.374 | 0.02000 | 0.02000 | -3.565 | TIGD4 | tigger transposable element derived 4 [Source:HGNC Symbol;Acc:HGNC:18335] |
| ENSG00000117335 | 0.832 | 8.693 | 2.374 | 0.02000 | 0.02000 | -3.209 | CD46 | CD46 molecule [Source:HGNC Symbol;Acc:HGNC:6953] |
| ENSG00000125970 | 0.68 | 6.814 | 2.372 | 0.02000 | 0.02000 | -3.208 | RALY | RALY heterogeneous nuclear ribonucleoprotein [Source:HGNC Symbol;Acc:HGNC:15921] |
| ENSG00000166881 | 0.622 | 4.794 | 2.372 | 0.02000 | 0.02000 | -3.209 | NEMP1 | nuclear envelope integral membrane protein 1 [Source:HGNC Symbol;Acc:HGNC:29001] |
| ENSG00000153391 | -0.825 | 1.891 | -2.372 | 0.02000 | 0.02000 | -3.522 | INO80C | INO80 complex subunit C [Source:HGNC Symbol;Acc:HGNC:26994] |
| ENSG00000072849 | -0.605 | 6.561 | -2.371 | 0.02000 | 0.02000 | -3.213 | DERL2 | derlin 2 [Source:HGNC Symbol;Acc:HGNC:17943] |
| ENSG00000198252 | 0.537 | 5.665 | 2.371 | 0.02000 | 0.02000 | -3.213 | STYX | serine/threonine/tyrosine interacting protein [Source:HGNC Symbol;Acc:HGNC:11447] |
| ENSG00000172869 | 0.492 | 7.28 | 2.369 | 0.02000 | 0.02000 | -3.214 | DMXL1 | Dmx like 1 [Source:HGNC Symbol;Acc:HGNC:2937] |
| ENSG00000122359 | -0.963 | 5.819 | -2.366 | 0.02000 | 0.02000 | -3.218 | ANXA11 | annexin A11 [Source:HGNC Symbol;Acc:HGNC:535] |
| ENSG00000145743 | 0.495 | 5.158 | 2.366 | 0.02000 | 0.02000 | -3.222 | FBXL17 | F-box and leucine rich repeat protein 17 [Source:HGNC Symbol;Acc:HGNC:13615] |
| ENSG00000196620 | 2.244 | -0.228 | 2.364 | 0.02000 | 0.02000 | -3.506 | UGT2B15 | UDP glucuronosyltransferase family 2 member B15 [Source:HGNC Symbol;Acc:HGNC:12546] |
| ENSG00000105866 | 0.574 | 5.276 | 2.364 | 0.02000 | 0.02000 | -3.226 | SP4 | Sp4 transcription factor [Source:HGNC Symbol;Acc:HGNC:11209] |
| ENSG00000166704 | 0.906 | 2.945 | 2.363 | 0.02000 | 0.02000 | -3.231 | ZNF606 | zinc finger protein 606 [Source:HGNC Symbol;Acc:HGNC:25879] |
| ENSG00000162999 | 1.076 | 1.502 | 2.363 | 0.02000 | 0.02000 | -3.316 | DUSP19 | dual specificity phosphatase 19 [Source:HGNC Symbol;Acc:HGNC:18894] |
| ENSG00000124164 | 0.574 | 4.514 | 2.361 | 0.02100 | 0.02100 | -3.228 | VAPB | VAMP associated protein B and C [Source:HGNC Symbol;Acc:HGNC:12649] |
| ENSG00000108666 | 0.541 | 3.669 | 2.358 | 0.02100 | 0.02100 | -3.233 | C17orf75 | chromosome 17 open reading frame 75 [Source:HGNC Symbol;Acc:HGNC:30173] |
| ENSG00000224470 | -0.701 | 6.304 | -2.358 | 0.02100 | 0.02100 | -3.237 | ATXN1L | ataxin 1 like [Source:HGNC Symbol;Acc:HGNC:33279] |
| ENSG00000180233 | 0.594 | 5.952 | 2.357 | 0.02100 | 0.02100 | -3.239 | ZNRF2 | zinc and ring finger 2 [Source:HGNC Symbol;Acc:HGNC:22316] |
| ENSG00000006744 | -0.541 | 5.614 | -2.357 | 0.02100 | 0.02100 | -3.237 | ELAC2 | elaC ribonuclease Z 2 [Source:HGNC Symbol;Acc:HGNC:14198] |
| ENSG00000204217 | 0.74 | 6.097 | 2.356 | 0.02100 | 0.02100 | -3.24 | BMPR2 | bone morphogenetic protein receptor type 2 [Source:HGNC Symbol;Acc:HGNC:1078] |
| ENSG00000106723 | 0.63 | 4.62 | 2.355 | 0.02100 | 0.02100 | -3.24 | SPIN1 | spindlin 1 [Source:HGNC Symbol;Acc:HGNC:11243] |
| ENSG00000079337 | 1.268 | 3.207 | 2.355 | 0.02100 | 0.02100 | -3.238 | RAPGEF3 | Rap guanine nucleotide exchange factor 3 [Source:HGNC Symbol;Acc:HGNC:16629] |
| ENSG00000057935 | 0.619 | 3.993 | 2.354 | 0.02100 | 0.02100 | -3.239 | MTA3 | metastasis associated 1 family member 3 [Source:HGNC Symbol;Acc:HGNC:23784] |
| ENSG00000125386 | -0.616 | 4.78 | -2.354 | 0.02100 | 0.02100 | -3.24 | FAM193A | family with sequence similarity 193 member A [Source:HGNC Symbol;Acc:HGNC:16822] |
| ENSG00000182253 | 2.003 | 2.829 | 2.354 | 0.02100 | 0.02100 | -3.241 | SYNM | synemin [Source:HGNC Symbol;Acc:HGNC:24466] |
| ENSG00000128694 | 0.579 | 2.832 | 2.352 | 0.02100 | 0.02100 | -3.261 | OSGEPL1 | O-sialoglycoprotein endopeptidase like 1 [Source:HGNC Symbol;Acc:HGNC:23075] |
| ENSG00000159314 | -0.649 | 5.115 | -2.35 | 0.02100 | 0.02100 | -3.248 | ARHGAP27 | Rho GTPase activating protein 27 [Source:HGNC Symbol;Acc:HGNC:31813] |
| ENSG00000139926 | 1.499 | 4.65 | 2.349 | 0.02100 | 0.02100 | -3.254 | FRMD6 | FERM domain containing 6 [Source:HGNC Symbol;Acc:HGNC:19839] |
| ENSG00000165417 | 0.648 | 6.674 | 2.347 | 0.02100 | 0.02100 | -3.256 | GTF2A1 | general transcription factor IIA subunit 1 [Source:HGNC Symbol;Acc:HGNC:4646] |
| ENSG00000172006 | 0.582 | 2.263 | 2.342 | 0.02200 | 0.02200 | -3.316 | ZNF554 | zinc finger protein 554 [Source:HGNC Symbol;Acc:HGNC:26629] |
| ENSG00000167716 | -0.857 | 3.836 | -2.341 | 0.02200 | 0.02200 | -3.291 | WDR81 | WD repeat domain 81 [Source:HGNC Symbol;Acc:HGNC:26600] |
| ENSG00000167978 | -1.015 | 7.909 | -2.34 | 0.02200 | 0.02200 | -3.269 | SRRM2 | serine/arginine repetitive matrix 2 [Source:HGNC Symbol;Acc:HGNC:16639] |
| ENSG00000164241 | -1.166 | 3.016 | -2.34 | 0.02200 | 0.02200 | -3.388 | C5orf63 | chromosome 5 open reading frame 63 [Source:HGNC Symbol;Acc:HGNC:40051] |
| ENSG00000197136 | -0.798 | 4.823 | -2.339 | 0.02200 | 0.02200 | -3.267 | PCNX3 | pecanex homolog 3 (Drosophila) [Source:HGNC Symbol;Acc:HGNC:18760] |
| ENSG00000002587 | 1.872 | -0.473 | 2.336 | 0.02200 | 0.02200 | -3.598 | HS3ST1 | heparan sulfate-glucosamine 3-sulfotransferase 1 [Source:HGNC Symbol;Acc:HGNC:5194] |
| ENSG00000137494 | 0.711 | 4.977 | 2.336 | 0.02200 | 0.02200 | -3.278 | ANKRD42 | ankyrin repeat domain 42 [Source:HGNC Symbol;Acc:HGNC:26752] |
| ENSG00000164953 | 0.939 | 2.377 | 2.335 | 0.02200 | 0.02200 | -3.301 | TMEM67 | transmembrane protein 67 [Source:HGNC Symbol;Acc:HGNC:28396] |
| ENSG00000120519 | -0.704 | 5.17 | -2.334 | 0.02200 | 0.02200 | -3.276 | SLC10A7 | solute carrier family 10 member 7 [Source:HGNC Symbol;Acc:HGNC:23088] |
| ENSG00000099917 | -0.911 | 3.996 | -2.332 | 0.02200 | 0.02200 | -3.299 | MED15 | mediator complex subunit 15 [Source:HGNC Symbol;Acc:HGNC:14248] |
| ENSG00000151689 | 0.755 | 3.045 | 2.332 | 0.02200 | 0.02200 | -3.284 | INPP1 | inositol polyphosphate-1-phosphatase [Source:HGNC Symbol;Acc:HGNC:6071] |
| ENSG00000130402 | -0.816 | 6.387 | -2.331 | 0.02200 | 0.02200 | -3.288 | ACTN4 | actinin alpha 4 [Source:HGNC Symbol;Acc:HGNC:166] |

| | | | | | | | | |
|---|---|---|---|---|---|---|---|---|
| ENSG00000254685 | 0.675 | 2.759 | 2.329 | 0.02200 | 0.02200 | -3.299 | FPGT | fucose-1-phosphate guanylyltransferase [Source:HGNC Symbol;Acc:HGNC:3825] |
| ENSG00000031823 | -0.53 | 4.957 | -2.328 | 0.02200 | 0.02200 | -3.288 | RANBP3 | RAN binding protein 3 [Source:HGNC Symbol;Acc:HGNC:9850] |
| ENSG00000197451 | -0.638 | 6.294 | -2.327 | 0.02200 | 0.02200 | -3.294 | HNRNPAB | heterogeneous nuclear ribonucleoprotein A/B [Source:HGNC Symbol;Acc:HGNC:5034] |
| ENSG00000126461 | -0.778 | 3.963 | -2.327 | 0.02200 | 0.02200 | -3.306 | SCAF1 | SR-related CTD associated factor 1 [Source:HGNC Symbol;Acc:HGNC:30403] |
| ENSG00000222011 | 0.607 | 2.966 | 2.326 | 0.02200 | 0.02200 | -3.299 | FAM185A | family with sequence similarity 185 member A [Source:HGNC Symbol;Acc:HGNC:22412] |
| ENSG00000163946 | 0.51 | 7.144 | 2.324 | 0.02300 | 0.02300 | -3.296 | FAM208A | family with sequence similarity 208 member A [Source:HGNC Symbol;Acc:HGNC:30314] |
| ENSG00000142892 | 0.744 | 6.881 | 2.324 | 0.02300 | 0.02300 | -3.296 | PIGK | phosphatidylinositol glycan anchor biosynthesis class K [Source:HGNC Symbol;Acc:HGNC:8965] |
| ENSG00000198373 | -0.703 | 4.499 | -2.324 | 0.02300 | 0.02300 | -3.295 | WWP2 | WW domain containing E3 ubiquitin protein ligase 2 [Source:HGNC Symbol;Acc:HGNC:16804] |
| ENSG00000167965 | -0.833 | 3.896 | -2.324 | 0.02300 | 0.02300 | -3.317 | MLST8 | MTOR associated protein, LST8 homolog [Source:HGNC Symbol;Acc:HGNC:24825] |
| ENSG00000133878 | 1.687 | 2.786 | 2.322 | 0.02300 | 0.02300 | -3.298 | DUSP26 | dual specificity phosphatase 26 [Source:HGNC Symbol;Acc:HGNC:28161] |
| ENSG00000118965 | 0.632 | 3.906 | 2.321 | 0.02300 | 0.02300 | -3.299 | WDR35 | WD repeat domain 35 [Source:HGNC Symbol;Acc:HGNC:29250] |
| ENSG00000214160 | -1.003 | 4.034 | -2.317 | 0.02300 | 0.02300 | -3.322 | ALG3 | ALG3, alpha-1,3- mannosyltransferase [Source:HGNC Symbol;Acc:HGNC:23056] |
| ENSG00000180304 | -0.601 | 4.051 | -2.317 | 0.02300 | 0.02300 | -3.312 | OAZ2 | ornithine decarboxylase antizyme 2 [Source:HGNC Symbol;Acc:HGNC:8096] |
| ENSG00000121486 | 0.628 | 4.651 | 2.317 | 0.02300 | 0.02300 | -3.311 | TRMT1L | tRNA methyltransferase 1 like [Source:HGNC Symbol;Acc:HGNC:16782] |
| ENSG00000062725 | 0.596 | 5.243 | 2.315 | 0.02300 | 0.02300 | -3.317 | APPBP2 | amyloid beta precursor protein binding protein 2 [Source:HGNC Symbol;Acc:HGNC:622] |
| ENSG00000136010 | -1.39 | 7.187 | -2.315 | 0.02300 | 0.02300 | -3.317 | ALDH1L2 | aldehyde dehydrogenase 1 family member L2 [Source:HGNC Symbol;Acc:HGNC:26777] |
| ENSG00000181192 | 0.628 | 5.68 | 2.315 | 0.02300 | 0.02300 | -3.318 | DHTKD1 | dehydrogenase E1 and transketolase domain containing 1 [Source:HGNC Symbol;Acc:HGNC:23537] |
| ENSG00000103966 | -1.18 | 5.794 | -2.311 | 0.02300 | 0.02300 | -3.318 | EHD4 | EH domain containing 4 [Source:HGNC Symbol;Acc:HGNC:3245] |
| ENSG00000128791 | 0.851 | 5.066 | 2.311 | 0.02300 | 0.02300 | -3.324 | TWSG1 | twisted gastrulation BMP signaling modulator 1 [Source:HGNC Symbol;Acc:HGNC:12429] |
| ENSG00000152465 | 0.617 | 3.984 | 2.311 | 0.02300 | 0.02300 | -3.318 | NMT2 | N-myristoyltransferase 2 [Source:HGNC Symbol;Acc:HGNC:7858] |
| ENSG00000128536 | 1.744 | 1.207 | 2.31 | 0.02300 | 0.02300 | -3.393 | CDHR3 | cadherin related family member 3 [Source:HGNC Symbol;Acc:HGNC:26308] |
| ENSG00000072518 | -0.644 | 4.038 | -2.31 | 0.02300 | 0.02300 | -3.326 | MARK2 | microtubule affinity regulating kinase 2 [Source:HGNC Symbol;Acc:HGNC:3332] |
| ENSG00000104413 | -5.282 | -0.234 | -2.31 | 0.02300 | 0.02300 | -4.046 | ESRP1 | epithelial splicing regulatory protein 1 [Source:HGNC Symbol;Acc:HGNC:25966] |
| ENSG00000113328 | 0.726 | 7.472 | 2.31 | 0.02300 | 0.02300 | -3.32 | CCNG1 | cyclin G1 [Source:HGNC Symbol;Acc:HGNC:1592] |
| ENSG00000127804 | -0.465 | 4.934 | -2.309 | 0.02300 | 0.02300 | -3.32 | METTL16 | methyltransferase like 16 [Source:HGNC Symbol;Acc:HGNC:28484] |
| ENSG00000259224 | -1.522 | 0.127 | -2.309 | 0.02300 | 0.02300 | -4.017 | SLC35G6 | solute carrier family 35 member G6 [Source:HGNC Symbol;Acc:HGNC:31351] |
| ENSG00000170142 | 0.553 | 6.279 | 2.308 | 0.02300 | 0.02300 | -3.329 | UBE2E1 | ubiquitin conjugating enzyme E2 E1 [Source:HGNC Symbol;Acc:HGNC:12477] |
| ENSG00000116478 | 0.368 | 6.055 | 2.308 | 0.02300 | 0.02300 | -3.33 | HDAC1 | histone deacetylase 1 [Source:HGNC Symbol;Acc:HGNC:4852] |
| ENSG00000139372 | -0.498 | 4.943 | -2.306 | 0.02400 | 0.02400 | -3.327 | TDG | thymine DNA glycosylase [Source:HGNC Symbol;Acc:HGNC:11700] |
| ENSG00000172175 | 0.797 | 7.56 | 2.304 | 0.02400 | 0.02400 | -3.33 | MALT1 | MALT1 paracaspase [Source:HGNC Symbol;Acc:HGNC:6819] |
| ENSG00000166887 | -0.471 | 6.423 | -2.302 | 0.02400 | 0.02400 | -3.34 | VPS39 | VPS39, HOPS complex subunit [Source:HGNC Symbol;Acc:HGNC:20593] |
| ENSG00000143153 | 1.319 | 4.542 | 2.302 | 0.02400 | 0.02400 | -3.34 | ATP1B1 | ATPase Na+/K+ transporting subunit beta 1 [Source:HGNC Symbol;Acc:HGNC:804] |
| ENSG00000164291 | 0.687 | 3.306 | 2.302 | 0.02400 | 0.02400 | -3.334 | ARSK | arylsulfatase family member K [Source:HGNC Symbol;Acc:HGNC:25239] |
| ENSG00000117533 | 0.555 | 4.991 | 2.3 | 0.02400 | 0.02400 | -3.343 | VAMP4 | vesicle associated membrane protein 4 [Source:HGNC Symbol;Acc:HGNC:12645] |
| ENSG00000243646 | 0.614 | 4.433 | 2.295 | 0.02400 | 0.02400 | -3.349 | IL10RB | interleukin 10 receptor subunit beta [Source:HGNC Symbol;Acc:HGNC:5965] |
| ENSG00000152223 | -0.687 | 5.592 | -2.294 | 0.02400 | 0.02400 | -3.351 | EPG5 | ectopic P-granules autophagy protein 5 homolog [Source:HGNC Symbol;Acc:HGNC:29331] |
| ENSG00000000003 | 1.873 | 0.472 | 2.292 | 0.02400 | 0.02400 | -3.477 | TSPAN6 | tetraspanin 6 [Source:HGNC Symbol;Acc:HGNC:11858] |
| ENSG00000154945 | 0.494 | 5.864 | 2.292 | 0.02400 | 0.02400 | -3.359 | ANKRD40 | ankyrin repeat domain 40 [Source:HGNC Symbol;Acc:HGNC:28233] |
| ENSG00000105865 | 0.469 | 2.957 | 2.292 | 0.02400 | 0.02400 | -3.36 | DUS4L | dihydrouridine synthase 4 like [Source:HGNC Symbol;Acc:HGNC:21517] |
| ENSG00000197888 | 2.922 | 2.394 | 2.29 | 0.02500 | 0.02500 | -3.353 | UGT2B17 | UDP glucuronosyltransferase family 2 member B17 [Source:HGNC Symbol;Acc:HGNC:12547] |
| ENSG00000154511 | 0.733 | 5.279 | 2.29 | 0.02500 | 0.02500 | -3.363 | FAM69A | family with sequence similarity 69 member A [Source:HGNC Symbol;Acc:HGNC:32213] |
| ENSG00000137145 | 1.26 | 5.543 | 2.289 | 0.02500 | 0.02500 | -3.364 | DENND4C | DENN domain containing 4C [Source:HGNC Symbol;Acc:HGNC:26079] |
| ENSG00000040633 | -0.518 | 3.676 | -2.288 | 0.02500 | 0.02500 | -3.37 | PHF23 | PHD finger protein 23 [Source:HGNC Symbol;Acc:HGNC:28428] |
| ENSG00000108509 | -1.263 | 2.514 | -2.288 | 0.02500 | 0.02500 | -3.597 | CAMTA2 | calmodulin binding transcription activator 2 [Source:HGNC Symbol;Acc:HGNC:18807] |
| ENSG00000120280 | 1 | 4.674 | 2.286 | 0.02500 | 0.02500 | -3.369 | CXorf21 | chromosome X open reading frame 21 [Source:HGNC Symbol;Acc:HGNC:25667] |
| ENSG00000100393 | -0.741 | 6.653 | -2.283 | 0.02500 | 0.02500 | -3.377 | EP300 | E1A binding protein p300 [Source:HGNC Symbol;Acc:HGNC:3373] |

| | | | | | | | | |
|---|---|---|---|---|---|---|---|---|
| ENSG00000183018 | -1.094 | 1.342 | -2.282 | 0.02500 | 0.02500 | -3.811 | SPNS2 | sphingolipid transporter 2 [Source:HGNC Symbol;Acc:HGNC:26992] |
| ENSG00000181817 | -0.86 | 2.195 | -2.282 | 0.02500 | 0.02500 | -3.607 | LSM10 | LSM10, U7 small nuclear RNA associated [Source:HGNC Symbol;Acc:HGNC:17562] |
| ENSG00000243725 | 0.449 | 3.833 | 2.282 | 0.02500 | 0.02500 | -3.368 | TTC4 | tetratricopeptide repeat domain 4 [Source:HGNC Symbol;Acc:HGNC:12394] |
| ENSG00000113356 | -0.764 | 3.334 | -2.282 | 0.02500 | 0.02500 | -3.407 | POLR3G | RNA polymerase III subunit G [Source:HGNC Symbol;Acc:HGNC:30075] |
| ENSG00000130803 | -0.575 | 5.506 | -2.281 | 0.02500 | 0.02500 | -3.374 | ZNF317 | zinc finger protein 317 [Source:HGNC Symbol;Acc:HGNC:13507] |
| ENSG00000090615 | -0.491 | 7.423 | -2.281 | 0.02500 | 0.02500 | -3.377 | GOLGA3 | golgin A3 [Source:HGNC Symbol;Acc:HGNC:4426] |
| ENSG00000161847 | -1.004 | 3.457 | -2.279 | 0.02500 | 0.02500 | -3.425 | RAVER1 | ribonucleoprotein, PTB binding 1 [Source:HGNC Symbol;Acc:HGNC:30296] |
| ENSG00000113441 | 0.418 | 7.481 | 2.277 | 0.02500 | 0.02500 | -3.379 | LNPEP | leucyl and cystinyl aminopeptidase [Source:HGNC Symbol;Acc:HGNC:6656] |
| ENSG00000089154 | -0.662 | 6.718 | -2.273 | 0.02600 | 0.02600 | -3.394 | GCN1 | GCN1, eIF2 alpha kinase activator homolog [Source:HGNC Symbol;Acc:HGNC:4199] |
| ENSG00000198420 | 0.722 | 3.442 | 2.273 | 0.02600 | 0.02600 | -3.383 | TCAF1 | TRPM8 channel associated factor 1 [Source:HGNC Symbol;Acc:HGNC:22201] |
| ENSG00000267303 | -1.06 | 2.152 | -2.273 | 0.02600 | 0.02600 | -3.63 | AC011511.4 | |
| ENSG00000130119 | -0.51 | 4.842 | -2.273 | 0.02600 | 0.02600 | -3.385 | GNL3L | G protein nucleolar 3 like [Source:HGNC Symbol;Acc:HGNC:25553] |
| ENSG00000076944 | -0.812 | 4.366 | -2.271 | 0.02600 | 0.02600 | -3.389 | STXBP2 | syntaxin binding protein 2 [Source:HGNC Symbol;Acc:HGNC:11445] |
| ENSG00000102349 | 0.881 | 2.833 | 2.269 | 0.02600 | 0.02600 | -3.394 | KLF8 | Kruppel like factor 8 [Source:HGNC Symbol;Acc:HGNC:6351] |
| ENSG00000065665 | 0.486 | 3.796 | 2.269 | 0.02600 | 0.02600 | -3.39 | SEC61A2 | Sec61 translocon alpha 2 subunit [Source:HGNC Symbol;Acc:HGNC:17702] |
| ENSG00000137500 | 0.546 | 6.296 | 2.269 | 0.02600 | 0.02600 | -3.4 | CCDC90B | coiled-coil domain containing 90B [Source:HGNC Symbol;Acc:HGNC:28108] |
| ENSG00000011638 | 0.768 | 1.698 | 2.268 | 0.02600 | 0.02600 | -3.464 | TMEM159 | transmembrane protein 159 [Source:HGNC Symbol;Acc:HGNC:30136] |
| ENSG00000139722 | -0.703 | 7.075 | -2.266 | 0.02600 | 0.02600 | -3.406 | VPS37B | VPS37B, ESCRT-I subunit [Source:HGNC Symbol;Acc:HGNC:25754] |
| ENSG00000176390 | 0.491 | 5.78 | 2.262 | 0.02600 | 0.02600 | -3.414 | CRLF3 | cytokine receptor like factor 3 [Source:HGNC Symbol;Acc:HGNC:17177] |
| ENSG00000077254 | 0.482 | 6.44 | 2.262 | 0.02600 | 0.02600 | -3.413 | USP33 | ubiquitin specific peptidase 33 [Source:HGNC Symbol;Acc:HGNC:20059] |
| ENSG00000184261 | -2.643 | 0.491 | -2.262 | 0.02600 | 0.02600 | -4.07 | KCNK12 | potassium two pore domain channel subfamily K member 12 [Source:HGNC Symbol;Acc:HGNC:6274] |
| ENSG00000264187 | 0.8 | 2.939 | 2.26 | 0.02600 | 0.02600 | -3.407 | AC055811.2 | |
| ENSG00000204469 | -0.835 | 5.873 | -2.26 | 0.02600 | 0.02600 | -3.414 | PRRC2A | proline rich coiled-coil 2A [Source:HGNC Symbol;Acc:HGNC:13918] |
| ENSG00000164134 | -0.512 | 6.84 | -2.259 | 0.02700 | 0.02700 | -3.42 | NAA15 | N(alpha)-acetyltransferase 15, NatA auxiliary subunit [Source:HGNC Symbol;Acc:HGNC:30782] |
| ENSG00000151092 | 0.55 | 7.401 | 2.257 | 0.02700 | 0.02700 | -3.415 | NGLY1 | N-glycanase 1 [Source:HGNC Symbol;Acc:HGNC:17646] |
| ENSG00000182534 | 1.037 | 2.524 | 2.255 | 0.02700 | 0.02700 | -3.419 | MXRA7 | matrix remodeling associated 7 [Source:HGNC Symbol;Acc:HGNC:7541] |
| ENSG00000205129 | 1.139 | 0.478 | 2.253 | 0.02700 | 0.02700 | -3.661 | C4orf47 | chromosome 4 open reading frame 47 [Source:HGNC Symbol;Acc:HGNC:34346] |
| ENSG00000164975 | 0.574 | 5.532 | 2.252 | 0.02700 | 0.02700 | -3.431 | SNAPC3 | small nuclear RNA activating complex polypeptide 3 [Source:HGNC Symbol;Acc:HGNC:11136] |
| ENSG00000154803 | 0.833 | 3.701 | 2.251 | 0.02700 | 0.02700 | -3.422 | FLCN | folliculin [Source:HGNC Symbol;Acc:HGNC:27310] |
| ENSG00000170234 | 0.429 | 5.29 | 2.249 | 0.02700 | 0.02700 | -3.437 | PWWP2A | PWWP domain containing 2A [Source:HGNC Symbol;Acc:HGNC:29406] |
| ENSG00000108840 | -0.582 | 5.653 | -2.248 | 0.02700 | 0.02700 | -3.435 | HDAC5 | histone deacetylase 5 [Source:HGNC Symbol;Acc:HGNC:14068] |
| ENSG00000121410 | 1.519 | 0.989 | 2.247 | 0.02700 | 0.02700 | -3.52 | A1BG | alpha-1-B glycoprotein [Source:HGNC Symbol;Acc:HGNC:5] |
| ENSG00000172534 | -0.839 | 5.431 | -2.246 | 0.02700 | 0.02700 | -3.434 | HCFC1 | host cell factor C1 [Source:HGNC Symbol;Acc:HGNC:4839] |
| ENSG00000204219 | 2.021 | 1.516 | 2.246 | 0.02700 | 0.02700 | -3.466 | TCEA3 | transcription elongation factor A3 [Source:HGNC Symbol;Acc:HGNC:11615] |
| ENSG00000119661 | 0.577 | 3.785 | 2.242 | 0.02800 | 0.02800 | -3.437 | DNAL1 | dynein axonemal light chain 1 [Source:HGNC Symbol;Acc:HGNC:23247] |
| ENSG00000135372 | -0.672 | 5.31 | -2.242 | 0.02800 | 0.02800 | -3.442 | NAT10 | N-acetyltransferase 10 [Source:HGNC Symbol;Acc:HGNC:29830] |
| ENSG00000113532 | 0.765 | 7.378 | 2.238 | 0.02800 | 0.02800 | -3.447 | ST8SIA4 | ST8 alpha-N-acetyl-neuraminide alpha-2,8-sialyltransferase 4 [Source:HGNC Symbol;Acc:HGNC:10871] |
| ENSG00000163558 | 0.54 | 5.911 | 2.238 | 0.02800 | 0.02800 | -3.458 | PRKCI | protein kinase C iota [Source:HGNC Symbol;Acc:HGNC:9404] |
| ENSG00000105193 | -1.262 | 6.597 | -2.237 | 0.02800 | 0.02800 | -3.456 | RPS16 | ribosomal protein S16 [Source:HGNC Symbol;Acc:HGNC:10396] |
| ENSG00000175727 | -0.875 | 5.359 | -2.236 | 0.02800 | 0.02800 | -3.451 | MLXIP | MLX interacting protein [Source:HGNC Symbol;Acc:HGNC:17055] |
| ENSG00000196547 | -0.855 | 5.555 | -2.236 | 0.02800 | 0.02800 | -3.453 | MAN2A2 | mannosidase alpha class 2A member 2 [Source:HGNC Symbol;Acc:HGNC:6825] |
| ENSG00000152689 | 1.638 | 7.119 | 2.235 | 0.02800 | 0.02800 | -3.45 | RASGRP3 | RAS guanyl releasing protein 3 [Source:HGNC Symbol;Acc:HGNC:14545] |
| ENSG00000281593 | 0.593 | 4.677 | 2.234 | 0.02800 | 0.02800 | -3.46 | AC006978.2 | |
| ENSG00000136870 | 0.713 | 5.16 | 2.232 | 0.02800 | 0.02800 | -3.466 | ZNF189 | zinc finger protein 189 [Source:HGNC Symbol;Acc:HGNC:12980] |
| ENSG00000133313 | -0.871 | 6.905 | -2.232 | 0.02800 | 0.02800 | -3.467 | CNDP2 | carnosine dipeptidase 2 [Source:HGNC Symbol;Acc:HGNC:24437] |
| ENSG00000171121 | 0.718 | 4.508 | 2.232 | 0.02800 | 0.02800 | -3.463 | KCNMB3 | potassium calcium-activated channel subfamily M regulatory beta subunit 3 [Source:HGNC Symbol;Acc:HGNC:6287] |

| | | | | | | | | |
|---|---|---|---|---|---|---|---|---|
| ENSG00000136816 | 0.559 | 4.812 | 2.231 | 0.02800 | 0.02800 | -3.466 | TOR1B | torsin family 1 member B [Source:HGNC Symbol;Acc:HGNC:11995] |
| ENSG00000065413 | 0.763 | 6.118 | 2.229 | 0.02900 | 0.02900 | -3.472 | ANKRD44 | ankyrin repeat domain 44 [Source:HGNC Symbol;Acc:HGNC:25259] |
| ENSG00000010818 | -0.854 | 6.135 | -2.227 | 0.02900 | 0.02900 | -3.474 | HIVEP2 | human immunodeficiency virus type I enhancer binding protein 2 [Source:HGNC Symbol;Acc:HGNC:4921] |
| ENSG00000243749 | 0.685 | 3.341 | 2.227 | 0.02900 | 0.02900 | -3.462 | TMEM35B | transmembrane protein 35B [Source:HGNC Symbol;Acc:HGNC:40021] |
| ENSG00000179262 | -0.651 | 5.247 | -2.224 | 0.02900 | 0.02900 | -3.472 | RAD23A | RAD23 homolog A, nucleotide excision repair protein [Source:HGNC Symbol;Acc:HGNC:9812] |
| ENSG00000271723 | 0.438 | 3.755 | 2.222 | 0.02900 | 0.02900 | -3.47 | MROH7-TTC4 | MROH7-TTC4 readthrough (NMD candidate) [Source:HGNC Symbol;Acc:HGNC:49180] |
| ENSG00000060237 | -0.641 | 6.204 | -2.221 | 0.02900 | 0.02900 | -3.486 | WNK1 | WNK lysine deficient protein kinase 1 [Source:HGNC Symbol;Acc:HGNC:14540] |
| ENSG00000078061 | -0.602 | 4.432 | -2.221 | 0.02900 | 0.02900 | -3.472 | ARAF | A-Raf proto-oncogene, serine/threonine kinase [Source:HGNC Symbol;Acc:HGNC:646] |
| ENSG00000123159 | 0.871 | 1.995 | 2.22 | 0.02900 | 0.02900 | -3.509 | GIPC1 | GIPC PDZ domain containing family member 1 [Source:HGNC Symbol;Acc:HGNC:1226] |
| ENSG00000152763 | 1.034 | 1.974 | 2.219 | 0.02900 | 0.02900 | -3.499 | WDR78 | WD repeat domain 78 [Source:HGNC Symbol;Acc:HGNC:26252] |
| ENSG00000141644 | -0.485 | 5.95 | -2.218 | 0.02900 | 0.02900 | -3.491 | MBD1 | methyl-CpG binding domain protein 1 [Source:HGNC Symbol;Acc:HGNC:6916] |
| ENSG00000066583 | 0.47 | 4.492 | 2.214 | 0.03000 | 0.03000 | -3.492 | ISOC1 | isochorismatase domain containing 1 [Source:HGNC Symbol;Acc:HGNC:24254] |
| ENSG00000140464 | -0.91 | 3.292 | -2.214 | 0.03000 | 0.03000 | -3.525 | PML | promyelocytic leukemia [Source:HGNC Symbol;Acc:HGNC:9113] |
| ENSG00000179361 | -1.032 | 5.61 | -2.213 | 0.03000 | 0.03000 | -3.491 | ARID3B | AT-rich interaction domain 3B [Source:HGNC Symbol;Acc:HGNC:14350] |
| ENSG00000091640 | -0.643 | 5.136 | -2.213 | 0.03000 | 0.03000 | -3.491 | SPAG7 | sperm associated antigen 7 [Source:HGNC Symbol;Acc:HGNC:11216] |
| ENSG00000121057 | -0.777 | 4.869 | -2.212 | 0.03000 | 0.03000 | -3.488 | AKAP1 | A-kinase anchoring protein 1 [Source:HGNC Symbol;Acc:HGNC:367] |
| ENSG00000258724 | 0.648 | 2.957 | 2.212 | 0.03000 | 0.03000 | -3.487 | AC105001.2 | |
| ENSG00000119514 | 1.073 | 3.014 | 2.209 | 0.03000 | 0.03000 | -3.491 | GALNT12 | polypeptide N-acetylgalactosaminyltransferase 12 [Source:HGNC Symbol;Acc:HGNC:19877] |
| ENSG00000126746 | -0.593 | 4.319 | -2.208 | 0.03000 | 0.03000 | -3.492 | ZNF384 | zinc finger protein 384 [Source:HGNC Symbol;Acc:HGNC:11955] |
| ENSG00000153767 | 0.501 | 4.141 | 2.208 | 0.03000 | 0.03000 | -3.499 | GTF2E1 | general transcription factor IIE subunit 1 [Source:HGNC Symbol;Acc:HGNC:4650] |
| ENSG00000266202 | 0.726 | -0.046 | 2.204 | 0.03000 | 0.03000 | -3.83 | AC005697.1 | |
| ENSG00000183020 | -0.587 | 4.694 | -2.203 | 0.03000 | 0.03000 | -3.502 | AP2A2 | adaptor related protein complex 2 alpha 2 subunit [Source:HGNC Symbol;Acc:HGNC:562] |
| ENSG00000163399 | -0.805 | 6.107 | -2.203 | 0.03000 | 0.03000 | -3.517 | ATP1A1 | ATPase Na+/K+ transporting subunit alpha 1 [Source:HGNC Symbol;Acc:HGNC:799] |
| ENSG00000073614 | -0.4 | 7.413 | -2.202 | 0.03000 | 0.03000 | -3.517 | KDM5A | lysine demethylase 5A [Source:HGNC Symbol;Acc:HGNC:9886] |
| ENSG00000105708 | 0.671 | 3.637 | 2.202 | 0.03000 | 0.03000 | -3.506 | ZNF14 | zinc finger protein 14 [Source:HGNC Symbol;Acc:HGNC:12924] |
| ENSG00000128016 | -1.035 | 7.855 | -2.202 | 0.03000 | 0.03000 | -3.519 | ZFP36 | ZFP36 ring finger protein [Source:HGNC Symbol;Acc:HGNC:12862] |
| ENSG00000117868 | 0.533 | 6.817 | 2.201 | 0.03100 | 0.03100 | -3.518 | ESYT2 | extended synaptotagmin 2 [Source:HGNC Symbol;Acc:HGNC:22211] |
| ENSG00000011295 | -0.381 | 6.124 | -2.201 | 0.03100 | 0.03100 | -3.523 | TTC19 | tetratricopeptide repeat domain 19 [Source:HGNC Symbol;Acc:HGNC:26006] |
| ENSG00000250741 | 0.421 | 2.876 | 2.199 | 0.03100 | 0.03100 | -3.513 | NT5C1B-RDH14 | NT5C1B-RDH14 readthrough [Source:HGNC Symbol;Acc:HGNC:38831] |
| ENSG00000134909 | -1.844 | 3.304 | -2.198 | 0.03100 | 0.03100 | -3.636 | ARHGAP32 | Rho GTPase activating protein 32 [Source:HGNC Symbol;Acc:HGNC:17399] |
| ENSG00000173928 | -0.678 | 0.892 | -2.196 | 0.03100 | 0.03100 | -3.879 | SWSAP1 | SWIM-type zinc finger 7 associated protein 1 [Source:HGNC Symbol;Acc:HGNC:26638] |
| ENSG00000177192 | -0.731 | 3.078 | -2.196 | 0.03100 | 0.03100 | -3.553 | PUS1 | pseudouridylate synthase 1 [Source:HGNC Symbol;Acc:HGNC:15508] |
| ENSG00000182872 | -0.361 | 5.506 | -2.196 | 0.03100 | 0.03100 | -3.527 | RBM10 | RNA binding motif protein 10 [Source:HGNC Symbol;Acc:HGNC:9896] |
| ENSG00000114354 | -0.503 | 6.492 | -2.195 | 0.03100 | 0.03100 | -3.533 | TFG | TRK-fused gene [Source:HGNC Symbol;Acc:HGNC:11758] |
| ENSG00000155506 | -0.656 | 7.023 | -2.195 | 0.03100 | 0.03100 | -3.532 | LARP1 | La ribonucleoprotein domain family member 1 [Source:HGNC Symbol;Acc:HGNC:29531] |
| ENSG00000254093 | 0.607 | 4.052 | 2.191 | 0.03100 | 0.03100 | -3.528 | PINX1 | PIN2/TERF1 interacting, telomerase inhibitor 1 [Source:HGNC Symbol;Acc:HGNC:30046] |
| ENSG00000049860 | 0.875 | 7.251 | 2.19 | 0.03100 | 0.03100 | -3.529 | HEXB | hexosaminidase subunit beta [Source:HGNC Symbol;Acc:HGNC:4879] |
| ENSG00000075790 | 0.849 | 4.93 | 2.189 | 0.03100 | 0.03100 | -3.543 | BCAP29 | B-cell receptor associated protein 29 [Source:HGNC Symbol;Acc:HGNC:24131] |
| ENSG00000106615 | 0.614 | 6.429 | 2.187 | 0.03200 | 0.03200 | -3.545 | RHEB | Ras homolog enriched in brain [Source:HGNC Symbol;Acc:HGNC:10011] |
| ENSG00000100226 | -0.689 | 6.645 | -2.187 | 0.03200 | 0.03200 | -3.548 | GTPBP1 | GTP binding protein 1 [Source:HGNC Symbol;Acc:HGNC:4669] |
| ENSG00000160888 | -1.523 | 6.861 | -2.186 | 0.03200 | 0.03200 | -3.546 | IER2 | immediate early response 2 [Source:HGNC Symbol;Acc:HGNC:28871] |
| ENSG00000137221 | -0.531 | 4.382 | -2.185 | 0.03200 | 0.03200 | -3.532 | TJAP1 | tight junction associated protein 1 [Source:HGNC Symbol;Acc:HGNC:17949] |
| ENSG00000132846 | -1.545 | 0.932 | -2.184 | 0.03200 | 0.03200 | -4.023 | ZBED3 | zinc finger BED-type containing 3 [Source:HGNC Symbol;Acc:HGNC:20711] |
| ENSG00000099381 | -1.195 | 3.606 | -2.183 | 0.03200 | 0.03200 | -3.567 | SETD1A | SET domain containing 1A [Source:HGNC Symbol;Acc:HGNC:29010] |
| ENSG00000163517 | 0.999 | -0.011 | 2.182 | 0.03200 | 0.03200 | -3.779 | HDAC11 | histone deacetylase 11 [Source:HGNC Symbol;Acc:HGNC:19086] |
| ENSG00000247595 | -0.632 | 4.401 | -2.182 | 0.03200 | 0.03200 | -3.537 | SPTY2D1-AS1 | SPTY2D1 antisense RNA 1 [Source:HGNC Symbol;Acc:HGNC:44122] |

| | | | | | | | | | |
|---|---|---|---|---|---|---|---|---|---|
| ENSG00000198189 | 0.819 | 5.505 | 2.179 | 0.03200 | 0.03200 | -3.561 | HSD17B11 | | hydroxysteroid 17-beta dehydrogenase 11 [Source:HGNC Symbol;Acc:HGNC:22960] |
| ENSG00000052723 | 0.455 | 5.807 | 2.178 | 0.03200 | 0.03200 | -3.564 | SIKE1 | | suppressor of IKBKE 1 [Source:HGNC Symbol;Acc:HGNC:26119] |
| ENSG00000198932 | 1.302 | 3.149 | 2.176 | 0.03200 | 0.03200 | -3.548 | GPRASP1 | | G protein-coupled receptor associated sorting protein 1 [Source:HGNC Symbol;Acc:HGNC:24834] |
| ENSG00000107862 | -0.651 | 6.477 | -2.176 | 0.03200 | 0.03200 | -3.566 | GBF1 | | golgi brefeldin A resistant guanine nucleotide exchange factor 1 [Source:HGNC Symbol;Acc:HGNC:4181] |
| ENSG00000015285 | -1.043 | 3.131 | -2.176 | 0.03200 | 0.03200 | -3.606 | WAS | | Wiskott-Aldrich syndrome [Source:HGNC Symbol;Acc:HGNC:12731] |
| ENSG00000101052 | 0.536 | 3.182 | 2.175 | 0.03200 | 0.03200 | -3.547 | IFT52 | | intraflagellar transport 52 [Source:HGNC Symbol;Acc:HGNC:15901] |
| ENSG00000155090 | -0.952 | 7.22 | -2.175 | 0.03200 | 0.03200 | -3.569 | KLF10 | | Kruppel like factor 10 [Source:HGNC Symbol;Acc:HGNC:11810] |
| ENSG00000099954 | 2.044 | 1.058 | 2.173 | 0.03300 | 0.03300 | -3.585 | CECR2 | | CECR2, histone acetyl-lysine reader [Source:HGNC Symbol;Acc:HGNC:1840] |
| ENSG00000179841 | 1.16 | 3.4 | 2.173 | 0.03300 | 0.03300 | -3.555 | AKAP5 | | A-kinase anchoring protein 5 [Source:HGNC Symbol;Acc:HGNC:375] |
| ENSG00000215421 | -0.449 | 5.908 | -2.171 | 0.03300 | 0.03300 | -3.573 | ZNF407 | | zinc finger protein 407 [Source:HGNC Symbol;Acc:HGNC:19904] |
| ENSG00000141076 | -0.694 | 3.92 | -2.171 | 0.03300 | 0.03300 | -3.555 | UTP4 | | UTP4, small subunit processome component [Source:HGNC Symbol;Acc:HGNC:1983] |
| ENSG00000136813 | 0.387 | 8.448 | 2.171 | 0.03300 | 0.03300 | -3.557 | KIAA0368 | | KIAA0368 [Source:HGNC Symbol;Acc:HGNC:29020] |
| ENSG00000123349 | 0.663 | 7.854 | 2.17 | 0.03300 | 0.03300 | -3.56 | PFDN5 | | prefoldin subunit 5 [Source:HGNC Symbol;Acc:HGNC:8869] |
| ENSG00000166889 | -0.647 | 5 | -2.17 | 0.03300 | 0.03300 | -3.562 | PATL1 | | PAT1 homolog 1, processing body mRNA decay factor [Source:HGNC Symbol;Acc:HGNC:26721] |
| ENSG00000167670 | -0.941 | 4.366 | -2.17 | 0.03300 | 0.03300 | -3.556 | CHAF1A | | chromatin assembly factor 1 subunit A [Source:HGNC Symbol;Acc:HGNC:1910] |
| ENSG00000177707 | 1.557 | 3.473 | 2.169 | 0.03300 | 0.03300 | -3.566 | NECTIN3 | | nectin cell adhesion molecule 3 [Source:HGNC Symbol;Acc:HGNC:17664] |
| ENSG00000117791 | 1.291 | 2.612 | 2.166 | 0.03300 | 0.03300 | -3.561 | | Mar.02 | mitochondrial amidoxime reducing component 2 [Source:HGNC Symbol;Acc:HGNC:26064] |
| ENSG00000197168 | -1.349 | 0.005 | -2.166 | 0.03300 | 0.03300 | -4.102 | NEK5 | | NIMA related kinase 5 [Source:HGNC Symbol;Acc:HGNC:7748] |
| ENSG00000213588 | 0.537 | 1.802 | 2.166 | 0.03300 | 0.03300 | -3.62 | ZBTB9 | | zinc finger and BTB domain containing 9 [Source:HGNC Symbol;Acc:HGNC:28323] |
| ENSG00000104427 | 0.708 | 3.646 | 2.165 | 0.03300 | 0.03300 | -3.567 | ZC2HC1A | | zinc finger C2HC-type containing 1A [Source:HGNC Symbol;Acc:HGNC:24277] |
| ENSG00000023445 | 2.197 | 8.176 | 2.165 | 0.03300 | 0.03300 | -3.563 | BIRC3 | | baculoviral IAP repeat containing 3 [Source:HGNC Symbol;Acc:HGNC:591] |
| ENSG00000167004 | -0.7 | 9.026 | -2.165 | 0.03300 | 0.03300 | -3.569 | PDIA3 | | protein disulfide isomerase family A member 3 [Source:HGNC Symbol;Acc:HGNC:4606] |
| ENSG00000135392 | 0.38 | 5.615 | 2.163 | 0.03300 | 0.03300 | -3.588 | DNAJC14 | | DnaJ heat shock protein family (Hsp40) member C14 [Source:HGNC Symbol;Acc:HGNC:24581] |
| ENSG00000135249 | -0.567 | 5.071 | -2.163 | 0.03300 | 0.03300 | -3.576 | RINT1 | | RAD50 interactor 1 [Source:HGNC Symbol;Acc:HGNC:21876] |
| ENSG00000183281 | 1.031 | 0.265 | 2.163 | 0.03300 | 0.03300 | -3.792 | PLGLB1 | | plasminogen-like B1 [Source:HGNC Symbol;Acc:HGNC:9072] |
| ENSG00000205089 | 1.297 | 1.738 | 2.16 | 0.03400 | 0.03400 | -3.585 | CCNI2 | | cyclin I family member 2 [Source:HGNC Symbol;Acc:HGNC:33869] |
| ENSG00000146802 | 0.507 | 4.96 | 2.16 | 0.03400 | 0.03400 | -3.591 | TMEM168 | | transmembrane protein 168 [Source:HGNC Symbol;Acc:HGNC:25826] |
| ENSG00000178234 | 0.868 | 4.612 | 2.159 | 0.03400 | 0.03400 | -3.592 | GALNT11 | | polypeptide N-acetylgalactosaminyltransferase 11 [Source:HGNC Symbol;Acc:HGNC:19875] |
| ENSG00000124225 | 1.161 | 4.898 | 2.158 | 0.03400 | 0.03400 | -3.596 | PMEPA1 | | prostate transmembrane protein, androgen induced 1 [Source:HGNC Symbol;Acc:HGNC:14107] |
| ENSG00000240857 | 0.486 | 4.868 | 2.158 | 0.03400 | 0.03400 | -3.594 | RDH14 | | retinol dehydrogenase 14 (all-trans/9-cis/11-cis) [Source:HGNC Symbol;Acc:HGNC:19979] |
| ENSG00000055070 | -0.661 | 4.441 | -2.157 | 0.03400 | 0.03400 | -3.577 | SZRD1 | | SUZ RNA binding domain containing 1 [Source:HGNC Symbol;Acc:HGNC:30232] |
| ENSG00000233757 | 0.913 | 0.042 | 2.156 | 0.03400 | 0.03400 | -3.819 | AC092835.1 | | |
| ENSG00000105855 | 1.459 | 3.502 | 2.156 | 0.03400 | 0.03400 | -3.588 | ITGB8 | | integrin subunit beta 8 [Source:HGNC Symbol;Acc:HGNC:6163] |
| ENSG00000130723 | -0.643 | 6.75 | -2.155 | 0.03400 | 0.03400 | -3.603 | PRRC2B | | proline rich coiled-coil 2B [Source:HGNC Symbol;Acc:HGNC:28121] |
| ENSG00000133740 | 0.536 | 5.539 | 2.155 | 0.03400 | 0.03400 | -3.602 | E2F5 | | E2F transcription factor 5 [Source:HGNC Symbol;Acc:HGNC:3119] |
| ENSG00000140525 | -0.827 | 5.038 | -2.153 | 0.03400 | 0.03400 | -3.589 | FANCI | | Fanconi anemia complementation group I [Source:HGNC Symbol;Acc:HGNC:25568] |
| ENSG00000160877 | -0.812 | 4.01 | -2.151 | 0.03400 | 0.03400 | -3.588 | NACC1 | | nucleus accumbens associated 1 [Source:HGNC Symbol;Acc:HGNC:20967] |
| ENSG00000099991 | -0.754 | 5.041 | -2.151 | 0.03400 | 0.03400 | -3.594 | CABIN1 | | calcineurin binding protein 1 [Source:HGNC Symbol;Acc:HGNC:24187] |
| ENSG00000132305 | -0.456 | 5.532 | -2.148 | 0.03500 | 0.03500 | -3.609 | IMMT | | inner membrane mitochondrial protein [Source:HGNC Symbol;Acc:HGNC:6047] |
| ENSG00000175592 | -1.897 | 0.328 | -2.148 | 0.03500 | 0.03500 | -4.117 | FOSL1 | | FOS like 1, AP-1 transcription factor subunit [Source:HGNC Symbol;Acc:HGNC:13718] |
| ENSG00000077943 | 2.19 | 6.733 | 2.147 | 0.03500 | 0.03500 | -3.597 | ITGA8 | | integrin subunit alpha 8 [Source:HGNC Symbol;Acc:HGNC:6144] |
| ENSG00000116350 | -0.529 | 6.326 | -2.144 | 0.03500 | 0.03500 | -3.622 | SRSF4 | | serine and arginine rich splicing factor 4 [Source:HGNC Symbol;Acc:HGNC:10786] |
| ENSG00000130684 | 0.495 | 4.403 | 2.144 | 0.03500 | 0.03500 | -3.612 | ZNF337 | | zinc finger protein 337 [Source:HGNC Symbol;Acc:HGNC:15809] |
| ENSG00000249624 | 0.588 | 4.856 | 2.142 | 0.03500 | 0.03500 | -3.621 | AP000295.1 | | |
| ENSG00000282034 | -0.782 | 5.485 | -2.142 | 0.03500 | 0.03500 | -3.615 | AC106886.5 | | |
| ENSG00000138031 | -1.199 | 3.376 | -2.14 | 0.03500 | 0.03500 | -3.647 | ADCY3 | | adenylate cyclase 3 [Source:HGNC Symbol;Acc:HGNC:234] |

| | | | | | | | | |
|---|---|---|---|---|---|---|---|---|
| ENSG00000101846 | 1.299 | 4.448 | 2.14 | 0.03500 | 0.03500 | -3.626 | STS | steroid sulfatase [Source:HGNC Symbol;Acc:HGNC:11425] |
| ENSG00000125520 | 0.755 | 2.617 | 2.139 | 0.03500 | 0.03500 | -3.608 | SLC2A4RG | SLC2A4 regulator [Source:HGNC Symbol;Acc:HGNC:15930] |
| ENSG00000203995 | 1.599 | 1.041 | 2.139 | 0.03500 | 0.03500 | -3.662 | ZYG11A | zyg-11 family member A, cell cycle regulator [Source:HGNC Symbol;Acc:HGNC:32058] |
| ENSG00000197056 | 0.701 | 2.629 | 2.139 | 0.03500 | 0.03500 | -3.608 | ZMYM1 | zinc finger MYM-type containing 1 [Source:HGNC Symbol;Acc:HGNC:26253] |
| ENSG00000129255 | -0.673 | 4.027 | -2.138 | 0.03500 | 0.03500 | -3.607 | MPDU1 | mannose-P-dolichol utilization defect 1 [Source:HGNC Symbol;Acc:HGNC:7207] |
| ENSG00000131389 | -0.968 | 4.114 | -2.137 | 0.03600 | 0.03600 | -3.609 | SLC6A6 | solute carrier family 6 member 6 [Source:HGNC Symbol;Acc:HGNC:11052] |
| ENSG00000177302 | -0.579 | 4.004 | -2.137 | 0.03600 | 0.03600 | -3.608 | TOP3A | topoisomerase (DNA) III alpha [Source:HGNC Symbol;Acc:HGNC:11992] |
| ENSG00000101574 | 0.542 | 3.774 | 2.137 | 0.03600 | 0.03600 | -3.615 | METTL4 | methyltransferase like 4 [Source:HGNC Symbol;Acc:HGNC:24726] |
| ENSG00000050344 | 0.909 | 6.54 | 2.134 | 0.03600 | 0.03600 | -3.632 | NFE2L3 | nuclear factor, erythroid 2 like 3 [Source:HGNC Symbol;Acc:HGNC:7783] |
| ENSG00000168958 | 0.434 | 6.127 | 2.134 | 0.03600 | 0.03600 | -3.639 | MFF | mitochondrial fission factor [Source:HGNC Symbol;Acc:HGNC:24858] |
| ENSG00000017797 | 0.483 | 6.416 | 2.134 | 0.03600 | 0.03600 | -3.637 | RALBP1 | ralA binding protein 1 [Source:HGNC Symbol;Acc:HGNC:9841] |
| ENSG00000179119 | -0.587 | 7.514 | -2.134 | 0.03600 | 0.03600 | -3.636 | SPTY2D1 | SPT2 chromatin protein domain containing 1 [Source:HGNC Symbol;Acc:HGNC:26818] |
| ENSG00000137944 | 0.502 | 4.201 | 2.134 | 0.03600 | 0.03600 | -3.626 | KYAT3 | kynurenine aminotransferase 3 [Source:HGNC Symbol;Acc:HGNC:33238] |
| ENSG00000113272 | 0.711 | 4.395 | 2.133 | 0.03600 | 0.03600 | -3.632 | THG1L | tRNA-histidine guanylyltransferase 1 like [Source:HGNC Symbol;Acc:HGNC:26053] |
| ENSG00000132704 | 1.982 | 4.788 | 2.133 | 0.03600 | 0.03600 | -3.639 | FCRL2 | Fc receptor like 2 [Source:HGNC Symbol;Acc:HGNC:14875] |
| ENSG00000198815 | -0.46 | 6.083 | -2.133 | 0.03600 | 0.03600 | -3.64 | FOXJ3 | forkhead box J3 [Source:HGNC Symbol;Acc:HGNC:29178] |
| ENSG00000198538 | 0.964 | 3.014 | 2.133 | 0.03600 | 0.03600 | -3.617 | ZNF28 | zinc finger protein 28 [Source:HGNC Symbol;Acc:HGNC:13073] |
| ENSG00000198093 | 0.666 | 2.979 | 2.132 | 0.03600 | 0.03600 | -3.615 | ZNF649 | zinc finger protein 649 [Source:HGNC Symbol;Acc:HGNC:25741] |
| ENSG00000178971 | -0.714 | 4.828 | -2.132 | 0.03600 | 0.03600 | -3.621 | CTC1 | CST telomere replication complex component 1 [Source:HGNC Symbol;Acc:HGNC:26169] |
| ENSG00000110046 | -1.103 | 4.778 | -2.131 | 0.03600 | 0.03600 | -3.618 | ATG2A | autophagy related 2A [Source:HGNC Symbol;Acc:HGNC:29028] |
| ENSG00000100084 | -0.64 | 4.361 | -2.129 | 0.03600 | 0.03600 | -3.622 | HIRA | histone cell cycle regulator [Source:HGNC Symbol;Acc:HGNC:4916] |
| ENSG00000113971 | 0.574 | 5.096 | 2.128 | 0.03600 | 0.03600 | -3.647 | NPHP3 | nephrocystin 3 [Source:HGNC Symbol;Acc:HGNC:7907] |
| ENSG00000113580 | 0.816 | 8.365 | 2.127 | 0.03600 | 0.03600 | -3.627 | NR3C1 | nuclear receptor subfamily 3 group C member 1 [Source:HGNC Symbol;Acc:HGNC:7978] |
| ENSG00000186792 | -0.854 | 1.168 | -2.127 | 0.03600 | 0.03600 | -3.913 | HYAL3 | hyaluronoglucosaminidase 3 [Source:HGNC Symbol;Acc:HGNC:5322] |
| ENSG00000196267 | 0.742 | 2.771 | 2.125 | 0.03700 | 0.03700 | -3.626 | ZNF836 | zinc finger protein 836 [Source:HGNC Symbol;Acc:HGNC:34333] |
| ENSG00000052795 | -1.108 | 4.778 | -2.125 | 0.03700 | 0.03700 | -3.628 | FNIP2 | folliculin interacting protein 2 [Source:HGNC Symbol;Acc:HGNC:29280] |
| ENSG00000136444 | 0.376 | 4.617 | 2.122 | 0.03700 | 0.03700 | -3.651 | RSAD1 | radical S-adenosyl methionine domain containing 1 [Source:HGNC Symbol;Acc:HGNC:25634] |
| ENSG00000109118 | -0.741 | 4.877 | -2.118 | 0.03700 | 0.03700 | -3.645 | PHF12 | PHD finger protein 12 [Source:HGNC Symbol;Acc:HGNC:20816] |
| ENSG00000080603 | -0.762 | 5.602 | -2.118 | 0.03700 | 0.03700 | -3.657 | SRCAP | Snf2 related CREBBP activator protein [Source:HGNC Symbol;Acc:HGNC:16974] |
| ENSG00000104852 | -0.599 | 7.385 | -2.118 | 0.03700 | 0.03700 | -3.664 | SNRNP70 | small nuclear ribonucleoprotein U1 subunit 70 [Source:HGNC Symbol;Acc:HGNC:11150] |
| ENSG00000197822 | 1.235 | 1.794 | 2.118 | 0.03700 | 0.03700 | -3.657 | OCLN | occludin [Source:HGNC Symbol;Acc:HGNC:8104] |
| ENSG00000115942 | 0.427 | 5.128 | 2.117 | 0.03700 | 0.03700 | -3.665 | ORC2 | origin recognition complex subunit 2 [Source:HGNC Symbol;Acc:HGNC:8488] |
| ENSG00000137309 | -1.178 | 4.871 | -2.117 | 0.03700 | 0.03700 | -3.642 | HMGA1 | high mobility group AT-hook 1 [Source:HGNC Symbol;Acc:HGNC:5010] |
| ENSG00000087087 | -0.513 | 6.08 | -2.116 | 0.03700 | 0.03700 | -3.669 | SRRT | serrate, RNA effector molecule [Source:HGNC Symbol;Acc:HGNC:24101] |
| ENSG00000164603 | 0.468 | 4.063 | 2.115 | 0.03700 | 0.03700 | -3.655 | BMT2 | base methyltransferase of 25S rRNA 2 homolog [Source:HGNC Symbol;Acc:HGNC:26475] |
| ENSG00000095574 | -0.604 | 4.395 | -2.115 | 0.03700 | 0.03700 | -3.646 | IKZF5 | IKAROS family zinc finger 5 [Source:HGNC Symbol;Acc:HGNC:14283] |
| ENSG00000130997 | -0.58 | 4.099 | -2.114 | 0.03800 | 0.03800 | -3.645 | POLN | DNA polymerase nu [Source:HGNC Symbol;Acc:HGNC:18870] |
| ENSG00000090661 | 1.356 | 2.55 | 2.113 | 0.03800 | 0.03800 | -3.646 | CERS4 | ceramide synthase 4 [Source:HGNC Symbol;Acc:HGNC:23747] |
| ENSG00000115884 | 1.037 | 9.272 | 2.113 | 0.03800 | 0.03800 | -3.646 | SDC1 | syndecan 1 [Source:HGNC Symbol;Acc:HGNC:10658] |
| ENSG00000180098 | 0.416 | 4.308 | 2.113 | 0.03800 | 0.03800 | -3.662 | TRNAU1AP | tRNA selenocysteine 1 associated protein 1 [Source:HGNC Symbol;Acc:HGNC:30813] |
| ENSG00000123643 | -0.893 | 2.31 | -2.109 | 0.03800 | 0.03800 | -3.765 | SLC36A1 | solute carrier family 36 member 1 [Source:HGNC Symbol;Acc:HGNC:18761] |
| ENSG00000067704 | 0.718 | 6.276 | 2.109 | 0.03800 | 0.03800 | -3.679 | IARS2 | isoleucyl-tRNA synthetase 2, mitochondrial [Source:HGNC Symbol;Acc:HGNC:29685] |
| ENSG00000198792 | -1.08 | 4.821 | -2.108 | 0.03800 | 0.03800 | -3.656 | TMEM184B | transmembrane protein 184B [Source:HGNC Symbol;Acc:HGNC:1310] |
| ENSG00000122218 | -0.525 | 7.917 | -2.107 | 0.03800 | 0.03800 | -3.676 | COPA | coatomer protein complex subunit alpha [Source:HGNC Symbol;Acc:HGNC:2230] |
| ENSG00000168491 | 0.928 | 4.38 | 2.107 | 0.03800 | 0.03800 | -3.679 | CCDC110 | coiled-coil domain containing 110 [Source:HGNC Symbol;Acc:HGNC:28504] |
| ENSG00000204256 | -0.555 | 8.326 | -2.103 | 0.03900 | 0.03900 | -3.679 | BRD2 | bromodomain containing 2 [Source:HGNC Symbol;Acc:HGNC:1103] |

| | | | | | | | | |
|---|---|---|---|---|---|---|---|---|
| ENSG00000113790 | 0.994 | 2.456 | 2.102 | 0.03900 | 0.03900 | -3.663 | EHHADH | enoyl-CoA hydratase and 3-hydroxyacyl CoA dehydrogenase [Source:HGNC Symbol;Acc:HGNC:3247] |
| ENSG00000141499 | -0.794 | 2.81 | -2.101 | 0.03900 | 0.03900 | -3.718 | WRAP53 | WD repeat containing antisense to TP53 [Source:HGNC Symbol;Acc:HGNC:25522] |
| ENSG00000108039 | -0.463 | 5.861 | -2.101 | 0.03900 | 0.03900 | -3.693 | XPNPEP1 | X-prolyl aminopeptidase 1 [Source:HGNC Symbol;Acc:HGNC:12822] |
| ENSG00000130749 | -0.638 | 4.325 | -2.099 | 0.03900 | 0.03900 | -3.668 | ZC3H4 | zinc finger CCCH-type containing 4 [Source:HGNC Symbol;Acc:HGNC:17808] |
| ENSG00000050327 | -2.875 | 1.255 | -2.096 | 0.03900 | 0.03900 | -4.139 | ARHGEF5 | Rho guanine nucleotide exchange factor 5 [Source:HGNC Symbol;Acc:HGNC:13209] |
| ENSG00000112473 | -0.542 | 7.103 | -2.095 | 0.03900 | 0.03900 | -3.704 | SLC39A7 | solute carrier family 39 member 7 [Source:HGNC Symbol;Acc:HGNC:4927] |
| ENSG00000074319 | -0.404 | 5.277 | -2.093 | 0.03900 | 0.03900 | -3.698 | TSG101 | tumor susceptibility 101 [Source:HGNC Symbol;Acc:HGNC:15971] |
| ENSG00000170632 | 0.495 | 4.765 | 2.093 | 0.03900 | 0.03900 | -3.703 | ARMC10 | armadillo repeat containing 10 [Source:HGNC Symbol;Acc:HGNC:21706] |
| ENSG00000170917 | 0.628 | 3.52 | 2.092 | 0.03900 | 0.03900 | -3.684 | NUDT6 | nudix hydrolase 6 [Source:HGNC Symbol;Acc:HGNC:8053] |
| ENSG00000100207 | -0.462 | 5.955 | -2.092 | 0.04000 | 0.04000 | -3.709 | TCF20 | transcription factor 20 [Source:HGNC Symbol;Acc:HGNC:11631] |
| ENSG00000148450 | 0.815 | 1.713 | 2.091 | 0.04000 | 0.04000 | -3.706 | MSRB2 | methionine sulfoxide reductase B2 [Source:HGNC Symbol;Acc:HGNC:17061] |
| ENSG00000001629 | 0.38 | 6.733 | 2.091 | 0.04000 | 0.04000 | -3.707 | ANKIB1 | ankyrin repeat and IBR domain containing 1 [Source:HGNC Symbol;Acc:HGNC:22215] |
| ENSG00000075151 | -0.581 | 6.495 | -2.091 | 0.04000 | 0.04000 | -3.713 | EIF4G3 | eukaryotic translation initiation factor 4 gamma 3 [Source:HGNC Symbol;Acc:HGNC:3298] |
| ENSG00000197808 | 0.886 | 1.157 | 2.09 | 0.04000 | 0.04000 | -3.745 | ZNF461 | zinc finger protein 461 [Source:HGNC Symbol;Acc:HGNC:21629] |
| ENSG00000076067 | -0.943 | 2.332 | -2.09 | 0.04000 | 0.04000 | -3.802 | RBMS2 | RNA binding motif single stranded interacting protein 2 [Source:HGNC Symbol;Acc:HGNC:9909] |
| ENSG00000117713 | -0.836 | 5.645 | -2.09 | 0.04000 | 0.04000 | -3.704 | ARID1A | AT-rich interaction domain 1A [Source:HGNC Symbol;Acc:HGNC:11110] |
| ENSG00000168778 | 0.742 | 1.904 | 2.089 | 0.04000 | 0.04000 | -3.712 | TCTN2 | tectonic family member 2 [Source:HGNC Symbol;Acc:HGNC:25774] |
| ENSG00000159788 | 1.01 | 1.756 | 2.089 | 0.04000 | 0.04000 | -3.699 | RGS12 | regulator of G protein signaling 12 [Source:HGNC Symbol;Acc:HGNC:9994] |
| ENSG00000105323 | -0.481 | 6.989 | -2.089 | 0.04000 | 0.04000 | -3.715 | HNRNPUL1 | heterogeneous nuclear ribonucleoprotein U like 1 [Source:HGNC Symbol;Acc:HGNC:17011] |
| ENSG00000118507 | 0.834 | 3.087 | 2.088 | 0.04000 | 0.04000 | -3.687 | AKAP7 | A-kinase anchoring protein 7 [Source:HGNC Symbol;Acc:HGNC:377] |
| ENSG00000187147 | -0.454 | 5.41 | -2.088 | 0.04000 | 0.04000 | -3.709 | RNF220 | ring finger protein 220 [Source:HGNC Symbol;Acc:HGNC:25552] |
| ENSG00000109184 | 0.566 | 5.303 | 2.087 | 0.04000 | 0.04000 | -3.718 | DCUN1D4 | defective in cullin neddylation 1 domain containing 4 [Source:HGNC Symbol;Acc:HGNC:28998] |
| ENSG00000174007 | 0.779 | 1.284 | 2.086 | 0.04000 | 0.04000 | -3.743 | CEP19 | centrosomal protein 19 [Source:HGNC Symbol;Acc:HGNC:28209] |
| ENSG00000197062 | 0.479 | 3.948 | 2.085 | 0.04000 | 0.04000 | -3.701 | ZSCAN26 | zinc finger and SCAN domain containing 26 [Source:HGNC Symbol;Acc:HGNC:12978] |
| ENSG00000214189 | 0.924 | 2.098 | 2.085 | 0.04000 | 0.04000 | -3.694 | ZNF788 | zinc finger family member 788 [Source:HGNC Symbol;Acc:HGNC:33112] |
| ENSG00000162688 | 0.52 | 4.785 | 2.084 | 0.04000 | 0.04000 | -3.719 | AGL | amylo-alpha-1, 6-glucosidase, 4-alpha-glucanotransferase [Source:HGNC Symbol;Acc:HGNC:321] |
| ENSG00000187257 | 0.435 | 5.981 | 2.083 | 0.04000 | 0.04000 | -3.725 | RSBN1L | round spermatid basic protein 1 like [Source:HGNC Symbol;Acc:HGNC:24765] |
| ENSG00000124459 | 0.57 | 3.915 | 2.082 | 0.04000 | 0.04000 | -3.707 | ZNF45 | zinc finger protein 45 [Source:HGNC Symbol;Acc:HGNC:13111] |
| ENSG00000196110 | 1.054 | 2.101 | 2.082 | 0.04000 | 0.04000 | -3.695 | ZNF699 | zinc finger protein 699 [Source:HGNC Symbol;Acc:HGNC:24750] |
| ENSG00000161956 | -0.471 | 4.916 | -2.08 | 0.04100 | 0.04100 | -3.712 | SENP3 | SUMO1/sentrin/SMT3 specific peptidase 3 [Source:HGNC Symbol;Acc:HGNC:17862] |
| ENSG00000178951 | -0.659 | 4.675 | -2.08 | 0.04100 | 0.04100 | -3.704 | ZBTB7A | zinc finger and BTB domain containing 7A [Source:HGNC Symbol;Acc:HGNC:18078] |
| ENSG00000161509 | -1.533 | 0.663 | -2.08 | 0.04100 | 0.04100 | -4.114 | GRIN2C | glutamate ionotropic receptor NMDA type subunit 2C [Source:HGNC Symbol;Acc:HGNC:4587] |
| ENSG00000104859 | -0.538 | 5.129 | -2.079 | 0.04100 | 0.04100 | -3.717 | CLASRP | CLK4 associating serine/arginine rich protein [Source:HGNC Symbol;Acc:HGNC:17731] |
| ENSG00000164236 | 1.929 | 2.505 | 2.079 | 0.04100 | 0.04100 | -3.705 | ANKRD33B | ankyrin repeat domain 33B [Source:HGNC Symbol;Acc:HGNC:35240] |
| ENSG00000074356 | -0.399 | 5.73 | -2.078 | 0.04100 | 0.04100 | -3.73 | NCBP3 | nuclear cap binding subunit 3 [Source:HGNC Symbol;Acc:HGNC:24612] |
| ENSG00000161542 | 0.397 | 4.077 | 2.077 | 0.04100 | 0.04100 | -3.715 | PRPSAP1 | phosphoribosyl pyrophosphate synthetase associated protein 1 [Source:HGNC Symbol;Acc:HGNC:9466] |
| ENSG00000154645 | 0.821 | 1.372 | 2.077 | 0.04100 | 0.04100 | -3.748 | CHODL | chondrolectin [Source:HGNC Symbol;Acc:HGNC:17807] |
| ENSG00000178307 | -0.646 | 4.416 | -2.076 | 0.04100 | 0.04100 | -3.707 | TMEM11 | transmembrane protein 11 [Source:HGNC Symbol;Acc:HGNC:16823] |
| ENSG00000081870 | 0.598 | 4.412 | 2.076 | 0.04100 | 0.04100 | -3.727 | HSPB11 | heat shock protein family B (small) member 11 [Source:HGNC Symbol;Acc:HGNC:25019] |
| ENSG00000170037 | -0.633 | 4.116 | -2.076 | 0.04100 | 0.04100 | -3.704 | CNTROB | centrobin, centriole duplication and spindle assembly protein [Source:HGNC Symbol;Acc:HGNC:29616] |
| ENSG00000105971 | 1.251 | 3.706 | 2.076 | 0.04100 | 0.04100 | -3.725 | CAV2 | caveolin 2 [Source:HGNC Symbol;Acc:HGNC:1528] |
| ENSG00000129535 | -0.694 | 1.06 | -2.075 | 0.04100 | 0.04100 | -3.967 | NRL | neural retina leucine zipper [Source:HGNC Symbol;Acc:HGNC:8002] |
| ENSG00000141867 | -0.72 | 5.102 | -2.075 | 0.04100 | 0.04100 | -3.719 | BRD4 | bromodomain containing 4 [Source:HGNC Symbol;Acc:HGNC:13575] |
| ENSG00000213077 | -3.443 | -0.54 | -2.074 | 0.04100 | 0.04100 | -4.181 | FAM106A | family with sequence similarity 106 member A [Source:HGNC Symbol;Acc:HGNC:25682] |
| ENSG00000140853 | -1.065 | 5.047 | -2.074 | 0.04100 | 0.04100 | -3.715 | NLRC5 | NLR family CARD domain containing 5 [Source:HGNC Symbol;Acc:HGNC:29933] |
| ENSG00000130382 | -0.708 | 3.609 | -2.073 | 0.04100 | 0.04100 | -3.709 | MLLT1 | MLLT1, super elongation complex subunit [Source:HGNC Symbol;Acc:HGNC:7134] |

| | | | | | | | | |
|---|---|---|---|---|---|---|---|---|
| ENSG00000152219 | 0.52 | 5.834 | 2.073 | 0.04100 | 0.04100 | -3.742 | ARL14EP | ADP ribosylation factor like GTPase 14 effector protein [Source:HGNC Symbol;Acc:HGNC:26798] |
| ENSG00000168906 | -0.723 | 6.801 | -2.072 | 0.04100 | 0.04100 | -3.744 | MAT2A | methionine adenosyltransferase 2A [Source:HGNC Symbol;Acc:HGNC:6904] |
| ENSG00000140534 | -2.052 | 0.807 | -2.072 | 0.04100 | 0.04100 | -4.144 | TICRR | TOPBP1 interacting checkpoint and replication regulator [Source:HGNC Symbol;Acc:HGNC:28704] |
| ENSG00000169964 | 0.505 | 2.445 | 2.07 | 0.04200 | 0.04200 | -3.717 | TMEM42 | transmembrane protein 42 [Source:HGNC Symbol;Acc:HGNC:28444] |
| ENSG00000143569 | -0.573 | 6.753 | -2.068 | 0.04200 | 0.04200 | -3.75 | UBAP2L | ubiquitin associated protein 2 like [Source:HGNC Symbol;Acc:HGNC:29877] |
| ENSG00000204323 | 1.064 | 0.098 | 2.068 | 0.04200 | 0.04200 | -3.877 | SMIM5 | small integral membrane protein 5 [Source:HGNC Symbol;Acc:HGNC:40030] |
| ENSG00000119777 | -0.494 | 5.907 | -2.067 | 0.04200 | 0.04200 | -3.749 | TMEM214 | transmembrane protein 214 [Source:HGNC Symbol;Acc:HGNC:25983] |
| ENSG00000170043 | -1.418 | 1.738 | -2.067 | 0.04200 | 0.04200 | -3.96 | TRAPPC1 | trafficking protein particle complex 1 [Source:HGNC Symbol;Acc:HGNC:19894] |
| ENSG00000168066 | -0.633 | 6.919 | -2.065 | 0.04200 | 0.04200 | -3.755 | SF1 | splicing factor 1 [Source:HGNC Symbol;Acc:HGNC:12950] |
| ENSG00000114841 | -0.93 | 5.079 | -2.065 | 0.04200 | 0.04200 | -3.733 | DNAH1 | dynein axonemal heavy chain 1 [Source:HGNC Symbol;Acc:HGNC:2940] |
| ENSG00000169375 | -0.554 | 6.674 | -2.065 | 0.04200 | 0.04200 | -3.757 | SIN3A | SIN3 transcription regulator family member A [Source:HGNC Symbol;Acc:HGNC:19353] |
| ENSG00000023734 | -0.454 | 6.122 | -2.065 | 0.04200 | 0.04200 | -3.755 | STRAP | serine/threonine kinase receptor associated protein [Source:HGNC Symbol;Acc:HGNC:30796] |
| ENSG00000043143 | 0.657 | 5.408 | 2.064 | 0.04200 | 0.04200 | -3.757 | JADE2 | jade family PHD finger 2 [Source:HGNC Symbol;Acc:HGNC:22984] |
| ENSG00000164904 | 1.394 | 0.969 | 2.062 | 0.04200 | 0.04200 | -3.765 | ALDH7A1 | aldehyde dehydrogenase 7 family member A1 [Source:HGNC Symbol;Acc:HGNC:877] |
| ENSG00000111300 | -0.58 | 5.826 | -2.062 | 0.04200 | 0.04200 | -3.757 | NAA25 | N(alpha)-acetyltransferase 25, NatB auxiliary subunit [Source:HGNC Symbol;Acc:HGNC:25783] |
| ENSG00000005339 | -0.649 | 5.99 | -2.061 | 0.04200 | 0.04200 | -3.759 | CREBBP | CREB binding protein [Source:HGNC Symbol;Acc:HGNC:2348] |
| ENSG00000196704 | 0.53 | 6.497 | 2.06 | 0.04200 | 0.04200 | -3.76 | AMZ2 | archaelysin family metallopeptidase 2 [Source:HGNC Symbol;Acc:HGNC:28041] |
| ENSG00000108518 | -0.515 | 5.939 | -2.06 | 0.04300 | 0.04300 | -3.761 | PFN1 | profilin 1 [Source:HGNC Symbol;Acc:HGNC:8881] |
| ENSG00000171649 | 1.161 | 1.183 | 2.06 | 0.04300 | 0.04300 | -3.763 | ZIK1 | zinc finger protein interacting with K protein 1 [Source:HGNC Symbol;Acc:HGNC:33104] |
| ENSG00000143761 | -0.611 | 8.328 | -2.058 | 0.04300 | 0.04300 | -3.752 | ARF1 | ADP ribosylation factor 1 [Source:HGNC Symbol;Acc:HGNC:652] |
| ENSG00000163155 | 1.069 | 0.938 | 2.058 | 0.04300 | 0.04300 | -3.802 | LYSMD1 | LysM domain containing 1 [Source:HGNC Symbol;Acc:HGNC:32070] |
| ENSG00000186472 | -3.665 | 0.003 | -2.057 | 0.04300 | 0.04300 | -4.187 | PCLO | piccolo presynaptic cytomatrix protein [Source:HGNC Symbol;Acc:HGNC:13406] |
| ENSG00000111361 | -0.449 | 5.567 | -2.057 | 0.04300 | 0.04300 | -3.763 | EIF2B1 | eukaryotic translation initiation factor 2B subunit alpha [Source:HGNC Symbol;Acc:HGNC:3257] |
| ENSG00000140497 | -0.621 | 4.719 | -2.056 | 0.04300 | 0.04300 | -3.745 | SCAMP2 | secretory carrier membrane protein 2 [Source:HGNC Symbol;Acc:HGNC:10564] |
| ENSG00000102225 | -0.561 | 5.146 | -2.056 | 0.04300 | 0.04300 | -3.755 | CDK16 | cyclin dependent kinase 16 [Source:HGNC Symbol;Acc:HGNC:8749] |
| ENSG00000112715 | -0.797 | 6.048 | -2.055 | 0.04300 | 0.04300 | -3.769 | VEGFA | vascular endothelial growth factor A [Source:HGNC Symbol;Acc:HGNC:12680] |
| ENSG00000088832 | -0.545 | 4.318 | -2.054 | 0.04300 | 0.04300 | -3.742 | FKBP1A | FK506 binding protein 1A [Source:HGNC Symbol;Acc:HGNC:3711] |
| ENSG00000151743 | 0.57 | 2.8 | 2.053 | 0.04300 | 0.04300 | -3.737 | AMN1 | antagonist of mitotic exit network 1 homolog [Source:HGNC Symbol;Acc:HGNC:27281] |
| ENSG00000185163 | -0.579 | 4.279 | -2.052 | 0.04300 | 0.04300 | -3.744 | DDX51 | DEAD-box helicase 51 [Source:HGNC Symbol;Acc:HGNC:20082] |
| ENSG00000198563 | -0.549 | 6.674 | -2.051 | 0.04300 | 0.04300 | -3.779 | DDX39B | DExD-box helicase 39B [Source:HGNC Symbol;Acc:HGNC:13917] |
| ENSG00000051009 | -0.653 | 3.817 | -2.05 | 0.04400 | 0.04400 | -3.741 | FAM160A2 | family with sequence similarity 160 member A2 [Source:HGNC Symbol;Acc:HGNC:25378] |
| ENSG00000099840 | 0.966 | 2.328 | 2.049 | 0.04400 | 0.04400 | -3.744 | IZUMO4 | IZUMO family member 4 [Source:HGNC Symbol;Acc:HGNC:26950] |
| ENSG00000243207 | -0.687 | 3.95 | -2.048 | 0.04400 | 0.04400 | -3.745 | PPAN-P2RY11 | PPAN-P2RY11 readthrough [Source:HGNC Symbol;Acc:HGNC:33526] |
| ENSG00000160799 | -0.473 | 4.938 | -2.047 | 0.04400 | 0.04400 | -3.766 | CCDC12 | coiled-coil domain containing 12 [Source:HGNC Symbol;Acc:HGNC:28332] |
| ENSG00000142065 | 1.105 | 1.91 | 2.046 | 0.04400 | 0.04400 | -3.751 | ZFP14 | ZFP14 zinc finger protein [Source:HGNC Symbol;Acc:HGNC:29312] |
| ENSG00000013503 | -0.597 | 4.485 | -2.046 | 0.04400 | 0.04400 | -3.758 | POLR3B | RNA polymerase III subunit B [Source:HGNC Symbol;Acc:HGNC:30348] |
| ENSG00000242028 | -0.784 | 5.486 | -2.045 | 0.04400 | 0.04400 | -3.776 | HYPK | huntingtin interacting protein K [Source:HGNC Symbol;Acc:HGNC:18418] |
| ENSG00000079313 | -0.902 | 3.725 | -2.045 | 0.04400 | 0.04400 | -3.752 | REXO1 | RNA exonuclease 1 homolog [Source:HGNC Symbol;Acc:HGNC:24616] |
| ENSG00000114107 | 0.616 | 5.798 | 2.044 | 0.04400 | 0.04400 | -3.79 | CEP70 | centrosomal protein 70 [Source:HGNC Symbol;Acc:HGNC:29972] |
| ENSG00000140993 | 0.771 | 4.154 | 2.044 | 0.04400 | 0.04400 | -3.777 | TIGD7 | tigger transposable element derived 7 [Source:HGNC Symbol;Acc:HGNC:18331] |
| ENSG00000150556 | 1.817 | 2.599 | 2.044 | 0.04400 | 0.04400 | -3.76 | LYPD6B | LY6/PLAUR domain containing 6B [Source:HGNC Symbol;Acc:HGNC:27018] |
| ENSG00000132849 | 1.493 | 4.539 | 2.043 | 0.04400 | 0.04400 | -3.79 | PATJ | PATJ, crumbs cell polarity complex component [Source:HGNC Symbol;Acc:HGNC:28881] |
| ENSG00000166762 | -1.651 | 1.355 | -2.042 | 0.04400 | 0.04400 | -4.037 | CATSPER2 | cation channel sperm associated 2 [Source:HGNC Symbol;Acc:HGNC:18810] |
| ENSG00000165591 | 1.045 | 3.987 | 2.041 | 0.04400 | 0.04400 | -3.781 | FAAH2 | fatty acid amide hydrolase 2 [Source:HGNC Symbol;Acc:HGNC:26440] |
| ENSG00000090266 | 0.464 | 5.284 | 2.041 | 0.04400 | 0.04400 | -3.794 | NDUFB2 | NADH:ubiquinone oxidoreductase subunit B2 [Source:HGNC Symbol;Acc:HGNC:7697] |
| ENSG00000186815 | -0.734 | 3.915 | -2.04 | 0.04400 | 0.04400 | -3.756 | TPCN1 | two pore segment channel 1 [Source:HGNC Symbol;Acc:HGNC:18182] |

| | | | | | | | | |
|---|---|---|---|---|---|---|---|---|
| ENSG00000086758 | -0.503 | 7.791 | -2.04 | 0.04400 | 0.04400 | -3.788 | HUWE1 | HECT, UBA and WWE domain containing 1, E3 ubiquitin protein ligase [Source:HGNC Symbol;Acc:HGNC:30892] |
| ENSG00000168488 | -0.878 | 5.298 | -2.039 | 0.04500 | 0.04500 | -3.779 | ATXN2L | ataxin 2 like [Source:HGNC Symbol;Acc:HGNC:31326] |
| ENSG00000164985 | 0.516 | 6.365 | 2.039 | 0.04500 | 0.04500 | -3.797 | PSIP1 | PC4 and SFRS1 interacting protein 1 [Source:HGNC Symbol;Acc:HGNC:9527] |
| ENSG00000176896 | 0.516 | 3.995 | 2.038 | 0.04500 | 0.04500 | -3.779 | TCEANC | transcription elongation factor A N-terminal and central domain containing [Source:HGNC Symbol;Acc:HGNC:28277] |
| ENSG00000128563 | 0.428 | 5.962 | 2.038 | 0.04500 | 0.04500 | -3.801 | PRKRIP1 | PRKR interacting protein 1 (IL11 inducible) [Source:HGNC Symbol;Acc:HGNC:21894] |
| ENSG00000130816 | -0.682 | 5.918 | -2.036 | 0.04500 | 0.04500 | -3.798 | DNMT1 | DNA methyltransferase 1 [Source:HGNC Symbol;Acc:HGNC:2976] |
| ENSG00000159917 | 0.608 | 3.558 | 2.036 | 0.04500 | 0.04500 | -3.774 | ZNF235 | zinc finger protein 235 [Source:HGNC Symbol;Acc:HGNC:12866] |
| ENSG00000072682 | 1.065 | 3.713 | 2.036 | 0.04500 | 0.04500 | -3.784 | P4HA2 | prolyl 4-hydroxylase subunit alpha 2 [Source:HGNC Symbol;Acc:HGNC:8547] |
| ENSG00000220205 | -0.857 | 3.681 | -2.035 | 0.04500 | 0.04500 | -3.766 | VAMP2 | vesicle associated membrane protein 2 [Source:HGNC Symbol;Acc:HGNC:12643] |
| ENSG00000169230 | 0.57 | 6.511 | 2.035 | 0.04500 | 0.04500 | -3.802 | PRELID1 | PRELI domain containing 1 [Source:HGNC Symbol;Acc:HGNC:30255] |
| ENSG00000081665 | 1.087 | 4.497 | 2.034 | 0.04500 | 0.04500 | -3.803 | ZNF506 | zinc finger protein 506 [Source:HGNC Symbol;Acc:HGNC:23780] |
| ENSG00000119655 | 0.97 | 5.193 | 2.031 | 0.04500 | 0.04500 | -3.812 | NPC2 | NPC intracellular cholesterol transporter 2 [Source:HGNC Symbol;Acc:HGNC:14537] |
| ENSG00000140323 | -1.477 | 0.108 | -2.029 | 0.04600 | 0.04600 | -4.18 | DISP2 | dispatched RND transporter family member 2 [Source:HGNC Symbol;Acc:HGNC:19712] |
| ENSG00000100991 | -0.428 | 6.231 | -2.028 | 0.04600 | 0.04600 | -3.816 | TRPC4AP | transient receptor potential cation channel subfamily C member 4 associated protein [Source:HGNC Symbol;Acc:HGNC:16181] |
| ENSG00000184840 | -0.562 | 8.071 | -2.028 | 0.04600 | 0.04600 | -3.804 | TMED9 | transmembrane p24 trafficking protein 9 [Source:HGNC Symbol;Acc:HGNC:24878] |
| ENSG00000101745 | 0.652 | 8.144 | 2.028 | 0.04600 | 0.04600 | -3.789 | ANKRD12 | ankyrin repeat domain 12 [Source:HGNC Symbol;Acc:HGNC:29135] |
| ENSG00000160741 | -0.98 | 3.802 | -2.028 | 0.04600 | 0.04600 | -3.776 | CRTC2 | CREB regulated transcription coactivator 2 [Source:HGNC Symbol;Acc:HGNC:27301] |
| ENSG00000137331 | -1.678 | 4.994 | -2.027 | 0.04600 | 0.04600 | -3.777 | IER3 | immediate early response 3 [Source:HGNC Symbol;Acc:HGNC:5392] |
| ENSG00000101294 | -0.521 | 7.124 | -2.027 | 0.04600 | 0.04600 | -3.818 | HM13 | histocompatibility minor 13 [Source:HGNC Symbol;Acc:HGNC:16435] |
| ENSG00000162735 | 0.501 | 5.15 | 2.026 | 0.04600 | 0.04600 | -3.818 | PEX19 | peroxisomal biogenesis factor 19 [Source:HGNC Symbol;Acc:HGNC:9713] |
| ENSG00000184178 | -0.536 | 5.101 | -2.026 | 0.04600 | 0.04600 | -3.804 | SCFD2 | sec1 family domain containing 2 [Source:HGNC Symbol;Acc:HGNC:30676] |
| ENSG00000099995 | -0.55 | 5.083 | -2.025 | 0.04600 | 0.04600 | -3.805 | SF3A1 | splicing factor 3a subunit 1 [Source:HGNC Symbol;Acc:HGNC:10765] |
| ENSG00000129562 | 0.727 | 6.589 | 2.025 | 0.04600 | 0.04600 | -3.816 | DAD1 | defender against cell death 1 [Source:HGNC Symbol;Acc:HGNC:2664] |
| ENSG00000177374 | -0.789 | 2.18 | -2.023 | 0.04600 | 0.04600 | -3.876 | HIC1 | HIC ZBTB transcriptional repressor 1 [Source:HGNC Symbol;Acc:HGNC:4909] |
| ENSG00000131876 | -0.745 | 5.51 | -2.022 | 0.04600 | 0.04600 | -3.814 | SNRPA1 | small nuclear ribonucleoprotein polypeptide A' [Source:HGNC Symbol;Acc:HGNC:11152] |
| ENSG00000173145 | 0.536 | 5.874 | 2.022 | 0.04600 | 0.04600 | -3.827 | NOC3L | NOC3 like DNA replication regulator [Source:HGNC Symbol;Acc:HGNC:24034] |
| ENSG00000184154 | -0.418 | 4.313 | -2.022 | 0.04600 | 0.04600 | -3.794 | LRTOMT | leucine rich transmembrane and O-methyltransferase domain containing [Source:HGNC Symbol;Acc:HGNC:25033] |
| ENSG00000177076 | 0.791 | 1.89 | 2.021 | 0.04600 | 0.04600 | -3.793 | ACER2 | alkaline ceramidase 2 [Source:HGNC Symbol;Acc:HGNC:23675] |
| ENSG00000198246 | -0.769 | 1.082 | -2.02 | 0.04700 | 0.04700 | -3.992 | SLC29A3 | solute carrier family 29 member 3 [Source:HGNC Symbol;Acc:HGNC:23096] |
| ENSG00000046651 | 0.549 | 6.465 | 2.02 | 0.04700 | 0.04700 | -3.827 | OFD1 | OFD1, centriole and centriolar satellite protein [Source:HGNC Symbol;Acc:HGNC:2567] |
| ENSG00000198342 | 1.233 | 1.574 | 2.02 | 0.04700 | 0.04700 | -3.8 | ZNF442 | zinc finger protein 442 [Source:HGNC Symbol;Acc:HGNC:20877] |
| ENSG00000096070 | -0.629 | 4.737 | -2.02 | 0.04700 | 0.04700 | -3.801 | BRPF3 | bromodomain and PHD finger containing 3 [Source:HGNC Symbol;Acc:HGNC:14256] |
| ENSG00000180423 | -0.461 | 3.548 | -2.019 | 0.04700 | 0.04700 | -3.786 | HARBI1 | harbinger transposase derived 1 [Source:HGNC Symbol;Acc:HGNC:26522] |
| ENSG00000073756 | 2.29 | 5.94 | 2.019 | 0.04700 | 0.04700 | -3.813 | PTGS2 | prostaglandin-endoperoxide synthase 2 [Source:HGNC Symbol;Acc:HGNC:9605] |
| ENSG00000135338 | -1.512 | 3.73 | -2.017 | 0.04700 | 0.04700 | -3.8 | LCA5 | LCA5, lebercilin [Source:HGNC Symbol;Acc:HGNC:31923] |
| ENSG00000073060 | -1.609 | 2.48 | -2.016 | 0.04700 | 0.04700 | -3.933 | SCARB1 | scavenger receptor class B member 1 [Source:HGNC Symbol;Acc:HGNC:1664] |
| ENSG00000111790 | 0.626 | 7.665 | 2.016 | 0.04700 | 0.04700 | -3.815 | FGFR1OP2 | FGFR1 oncogene partner 2 [Source:HGNC Symbol;Acc:HGNC:23098] |
| ENSG00000153140 | 0.778 | 4.453 | 2.016 | 0.04700 | 0.04700 | -3.829 | CETN3 | centrin 3 [Source:HGNC Symbol;Acc:HGNC:1868] |
| ENSG00000106524 | 0.59 | 4.138 | 2.015 | 0.04700 | 0.04700 | -3.821 | ANKMY2 | ankyrin repeat and MYND domain containing 2 [Source:HGNC Symbol;Acc:HGNC:25370] |
| ENSG00000151353 | 0.559 | 4.964 | 2.014 | 0.04700 | 0.04700 | -3.835 | TMEM18 | transmembrane protein 18 [Source:HGNC Symbol;Acc:HGNC:25257] |
| ENSG00000165730 | 1.546 | 0.756 | 2.014 | 0.04700 | 0.04700 | -3.859 | STOX1 | storkhead box 1 [Source:HGNC Symbol;Acc:HGNC:23508] |
| ENSG00000227124 | 0.727 | 3.344 | 2.013 | 0.04700 | 0.04700 | -3.807 | ZNF717 | zinc finger protein 717 [Source:HGNC Symbol;Acc:HGNC:29448] |
| ENSG00000068354 | -0.725 | 2.868 | -2.013 | 0.04700 | 0.04700 | -3.827 | TBC1D25 | TBC1 domain family member 25 [Source:HGNC Symbol;Acc:HGNC:8092] |
| ENSG00000111696 | -0.596 | 4.177 | -2.012 | 0.04700 | 0.04700 | -3.804 | NT5DC3 | 5'-nucleotidase domain containing 3 [Source:HGNC Symbol;Acc:HGNC:30826] |
| ENSG00000189292 | -1.215 | 0.09 | -2.009 | 0.04800 | 0.04800 | -4.183 | ALKAL2 | ALK and LTK ligand 2 [Source:HGNC Symbol;Acc:HGNC:27683] |
| ENSG00000065802 | 0.436 | 3.208 | 2.009 | 0.04800 | 0.04800 | -3.806 | ASB1 | ankyrin repeat and SOCS box containing 1 [Source:HGNC Symbol;Acc:HGNC:16011] |

| | | | | | | | | |
|---|---|---|---|---|---|---|---|---|
| ENSG00000112893 | 0.496 | 8.805 | 2.009 | 0.04800 | 0.04800 | -3.813 | MAN2A1 | mannosidase alpha class 2A member 1 [Source:HGNC Symbol;Acc:HGNC:6824] |
| ENSG00000162623 | 0.437 | 5.601 | 2.009 | 0.04800 | 0.04800 | -3.848 | TYW3 | tRNA-yW synthesizing protein 3 homolog [Source:HGNC Symbol;Acc:HGNC:24757] |
| ENSG00000104835 | -0.868 | 2.224 | -2.008 | 0.04800 | 0.04800 | -3.887 | SARS2 | seryl-tRNA synthetase 2, mitochondrial [Source:HGNC Symbol;Acc:HGNC:17697] |
| ENSG00000275126 | 1.438 | 0.458 | 2.008 | 0.04800 | 0.04800 | -3.871 | HIST1H4L | histone cluster 1 H4 family member l [Source:HGNC Symbol;Acc:HGNC:4791] |
| ENSG00000177733 | -0.475 | 5.58 | -2.007 | 0.04800 | 0.04800 | -3.843 | HNRNPA0 | heterogeneous nuclear ribonucleoprotein A0 [Source:HGNC Symbol;Acc:HGNC:5030] |
| ENSG00000173715 | 0.746 | 5.13 | 2.006 | 0.04800 | 0.04800 | -3.851 | C11orf80 | chromosome 11 open reading frame 80 [Source:HGNC Symbol;Acc:HGNC:26197] |
| ENSG00000136026 | -0.694 | 7.067 | -2.005 | 0.04800 | 0.04800 | -3.854 | CKAP4 | cytoskeleton associated protein 4 [Source:HGNC Symbol;Acc:HGNC:16991] |
| ENSG00000171970 | 1.457 | -0.063 | 2.004 | 0.04800 | 0.04800 | -3.918 | ZNF57 | zinc finger protein 57 [Source:HGNC Symbol;Acc:HGNC:13125] |
| ENSG00000196659 | 0.542 | 2.715 | 2.003 | 0.04800 | 0.04800 | -3.81 | TTC30B | tetratricopeptide repeat domain 30B [Source:HGNC Symbol;Acc:HGNC:26425] |
| ENSG00000133739 | 0.678 | 5.104 | 2.003 | 0.04800 | 0.04800 | -3.856 | LRRCC1 | leucine rich repeat and coiled-coil centrosomal protein 1 [Source:HGNC Symbol;Acc:HGNC:29373] |
| ENSG00000127663 | -0.488 | 4.986 | -2.002 | 0.04800 | 0.04800 | -3.839 | KDM4B | lysine demethylase 4B [Source:HGNC Symbol;Acc:HGNC:29136] |
| ENSG00000149972 | 2.07 | 4.922 | 2.002 | 0.04900 | 0.04900 | -3.855 | CNTN5 | contactin 5 [Source:HGNC Symbol;Acc:HGNC:2175] |
| ENSG00000179598 | 0.987 | 3.006 | 2.002 | 0.04900 | 0.04900 | -3.819 | PLD6 | phospholipase D family member 6 [Source:HGNC Symbol;Acc:HGNC:30447] |
| ENSG00000151164 | -0.794 | 0.943 | -2.001 | 0.04900 | 0.04900 | -4.062 | RAD9B | RAD9 checkpoint clamp component B [Source:HGNC Symbol;Acc:HGNC:21700] |
| ENSG00000122012 | 1.766 | 2.127 | 2.001 | 0.04900 | 0.04900 | -3.814 | SV2C | synaptic vesicle glycoprotein 2C [Source:HGNC Symbol;Acc:HGNC:30670] |
| ENSG00000149179 | -0.793 | 3.001 | -2 | 0.04900 | 0.04900 | -3.835 | C11orf49 | chromosome 11 open reading frame 49 [Source:HGNC Symbol;Acc:HGNC:28720] |
| ENSG00000186496 | 0.934 | 0.72 | 2 | 0.04900 | 0.04900 | -3.901 | ZNF396 | zinc finger protein 396 [Source:HGNC Symbol;Acc:HGNC:18824] |
| ENSG00000272647 | 0.484 | 2.204 | 1.999 | 0.04900 | 0.04900 | -3.822 | AC005020.2 | |
| ENSG00000184787 | -0.512 | 6.741 | -1.996 | 0.04900 | 0.04900 | -3.869 | UBE2G2 | ubiquitin conjugating enzyme E2 G2 [Source:HGNC Symbol;Acc:HGNC:12483] |
| ENSG00000122642 | 1.156 | 2.413 | 1.993 | 0.05000 | 0.05000 | -3.827 | FKBP9 | FK506 binding protein 9 [Source:HGNC Symbol;Acc:HGNC:3725] |
| ENSG00000164687 | -1.923 | 4.106 | -1.993 | 0.05000 | 0.05000 | -3.83 | FABP5 | fatty acid binding protein 5 [Source:HGNC Symbol;Acc:HGNC:3560] |
| ENSG00000118971 | 3.42 | 5.769 | 1.992 | 0.05000 | 0.05000 | -3.844 | CCND2 | cyclin D2 [Source:HGNC Symbol;Acc:HGNC:1583] |
| ENSG00000185055 | -0.672 | 2.595 | -1.991 | 0.05000 | 0.05000 | -3.869 | EFCAB10 | EF-hand calcium binding domain 10 [Source:HGNC Symbol;Acc:HGNC:34531] |
| ENSG00000187260 | 0.895 | 1.157 | 1.991 | 0.05000 | 0.05000 | -3.877 | WDR86 | WD repeat domain 86 [Source:HGNC Symbol;Acc:HGNC:28020] |
| ENSG00000054219 | 0.604 | 6.893 | 1.99 | 0.05000 | 0.05000 | -3.869 | LY75 | lymphocyte antigen 75 [Source:HGNC Symbol;Acc:HGNC:6729] |

**Supplemental Table S12.** Gene set enrichment analysis of the Hallmark library from significantly expressed genes of the comparison between monoallelic *TP53* to WT *TP53* samples of the FIMM cohort.

| ID | setSize | enrichmentScore | NES | pvalue | p.adjust | qvalue | rank |
|---|---|---|---|---|---|---|---|
| HALLMARK_MTORC1_SIGNALING | 10 | -0.496 | -1.606 | 0.054 | 0.054 | 0.411 | 73 |
| HALLMARK_IL2_STAT5_SIGNALING | 10 | 0.496 | 1.427 | 0.103 | 0.103 | 0.411 | 194 |
| HALLMARK_TNFA_SIGNALING_VIA_NFKB | 10 | 0.430 | 1.239 | 0.225 | 0.225 | 0.444 | 25 |
| HALLMARK_HYPOXIA | 12 | -0.342 | -1.191 | 0.251 | 0.251 | 0.444 | 73 |
| HALLMARK_G2M_CHECKPOINT | 10 | -0.361 | -1.168 | 0.278 | 0.278 | 0.444 | 153 |
| HALLMARK_GLYCOLYSIS | 10 | 0.375 | 1.080 | 0.371 | 0.371 | 0.493 | 156 |
| HALLMARK_MITOTIC_SPINDLE | 13 | -0.277 | -0.996 | 0.441 | 0.441 | 0.493 | 266 |
| HALLMARK_ESTROGEN_RESPONSE_EARLY | 11 | -0.285 | -0.955 | 0.493 | 0.493 | 0.493 | 79 |

**Supplemental Table S13.** Gene set enrichment analysis of the GO C5 library from significantly expressed genes of the comparison between monoallelic *TP53* to WT *TP53* samples of the FIMM cohort.

| ID | setSize | enrichmentScore | NES | pvalue | p.adjust | qvalue | rank |
|---|---|---|---|---|---|---|---|
| GOMF_RNA_BINDING | 112 | -0.443 | -3.045 | 0.00000 | 0.00000 | 0.00000 | 314 |
| GOBP_MRNA_METABOLIC_PROCESS | 42 | -0.471 | -2.538 | 0.00000 | 0.00000 | 0.00600 | 314 |
| GOCC_PLASMA_MEMBRANE_REGION | 41 | 0.517 | 2.228 | 0.00000 | 0.00000 | 0.01400 | 115 |
| GOCC_MEMBRANE_MICRODOMAIN | 15 | 0.697 | 2.283 | 0.00000 | 0.00000 | 0.01400 | 112 |
| GOCC_RIBONUCLEOPROTEIN_COMPLEX | 34 | -0.468 | -2.346 | 0.00000 | 0.00000 | 0.02200 | 295 |
| GOMF_MRNA_BINDING | 26 | -0.528 | -2.425 | 0.00000 | 0.00000 | 0.02400 | 247 |
| GOCC_NUCLEAR_BODY | 49 | -0.394 | -2.209 | 0.00000 | 0.00000 | 0.02400 | 314 |
| GOBP_TUBE_MORPHOGENESIS | 23 | 0.593 | 2.193 | 0.00000 | 0.00000 | 0.02400 | 100 |
| GOBP_PROTEIN_CONTAINING_COMPLEX_ORGANIZATION | 75 | -0.322 | -2.007 | 0.00000 | 0.00000 | 0.03800 | 286 |
| GOBP_CONNECTIVE_TISSUE_DEVELOPMENT | 11 | 0.705 | 2.102 | 0.00000 | 0.00000 | 0.03800 | 81 |
| GOBP_MUSCLE_CONTRACTION | 11 | 0.703 | 2.095 | 0.00000 | 0.00000 | 0.03800 | 115 |
| GOBP_BLOOD_VESSEL_MORPHOGENESIS | 16 | 0.646 | 2.158 | 0.00000 | 0.00000 | 0.03800 | 100 |
| GOCC_NUCLEAR_PROTEIN_CONTAINING_COMPLEX | 71 | -0.316 | -1.937 | 0.00100 | 0.00100 | 0.04400 | 264 |
| GOBP_CELLULAR_RESPONSE_TO_DNA_DAMAGE_STIMULUS | 34 | -0.432 | -2.169 | 0.00100 | 0.00100 | 0.04800 | 204 |
| GOBP_LIPID_METABOLIC_PROCESS | 48 | 0.443 | 1.986 | 0.00100 | 0.00100 | 0.05700 | 172 |
| GOBP_RNA_PROCESSING | 57 | -0.339 | -1.978 | 0.00100 | 0.00100 | 0.05700 | 314 |
| GOMF_SIGNALING_RECEPTOR_REGULATOR_ACTIVITY | 12 | 0.688 | 2.103 | 0.00100 | 0.00100 | 0.05700 | 74 |
| GOBP_EMBRYONIC_MORPHOGENESIS | 23 | 0.558 | 2.063 | 0.00100 | 0.00100 | 0.05700 | 77 |
| GOBP_MRNA_PROCESSING | 30 | -0.451 | -2.184 | 0.00100 | 0.00100 | 0.05700 | 325 |
| HP_DECREASED_HEAD_CIRCUMFERENCE | 54 | -0.346 | -1.993 | 0.00100 | 0.00100 | 0.06600 | 194 |
| GOBP_ANIMAL_ORGAN_MORPHOGENESIS | 34 | 0.487 | 2.01 | 0.00100 | 0.00100 | 0.06700 | 77 |
| GOBP_RESPONSE_TO_XENOBIOTIC_STIMULUS | 18 | 0.589 | 2.037 | 0.00100 | 0.00100 | 0.06900 | 27 |
| GOMF_CHROMATIN_BINDING | 38 | -0.397 | -2.059 | 0.00100 | 0.00100 | 0.06900 | 216 |
| GOBP_PEPTIDE_BIOSYNTHETIC_PROCESS | 30 | -0.433 | -2.099 | 0.00200 | 0.00200 | 0.07400 | 247 |
| GOBP_MONOCARBOXYLIC_ACID_METABOLIC_PROCESS | 23 | 0.545 | 2.015 | 0.00200 | 0.00200 | 0.07400 | 167 |
| GOBP_CIRCULATORY_SYSTEM_DEVELOPMENT | 28 | 0.507 | 1.986 | 0.00200 | 0.00200 | 0.07400 | 100 |
| HP_ABNORMALITY_OF_THE_CEREBRAL_SUBCORTEX | 47 | -0.358 | -1.979 | 0.00200 | 0.00200 | 0.07400 | 191 |
| GOBP_TISSUE_MORPHOGENESIS | 19 | 0.571 | 2.007 | 0.00200 | 0.00200 | 0.07800 | 77 |
| GOBP_POSITIVE_REGULATION_OF_CELL_POPULATION_PROLIFERATION | 28 | 0.504 | 1.971 | 0.00200 | 0.00200 | 0.07800 | 77 |
| HP_ABNORMAL_EMOTION_AFFECT_BEHAVIOR | 32 | -0.434 | -2.147 | 0.00200 | 0.00200 | 0.07900 | 207 |
| GOCC_RIBONUCLEOPROTEIN_GRANULE | 10 | -0.654 | -2.107 | 0.00200 | 0.00200 | 0.07900 | 254 |
| GOBP_REGULATION_OF_MRNA_METABOLIC_PROCESS | 20 | -0.516 | -2.159 | 0.00200 | 0.00200 | 0.08200 | 314 |
| GOBP_VASCULATURE_DEVELOPMENT | 18 | 0.572 | 1.979 | 0.00200 | 0.00200 | 0.08200 | 100 |
| GOBP_MUSCLE_SYSTEM_PROCESS | 12 | 0.651 | 1.99 | 0.00300 | 0.00300 | 0.09100 | 115 |
| GOCC_SIDE_OF_MEMBRANE | 19 | 0.544 | 1.913 | 0.00300 | 0.00300 | 0.09300 | 137 |
| GOBP_EPITHELIAL_CELL_DEVELOPMENT | 11 | 0.64 | 1.907 | 0.00300 | 0.00300 | 0.09300 | 148 |
| GOCC_PLASMA_MEMBRANE_PROTEIN_COMPLEX | 22 | 0.522 | 1.907 | 0.00300 | 0.00300 | 0.09300 | 120 |
| HP_HYPOPLASIA_OF_THE_CORPUS_CALLOSUM | 28 | -0.429 | -2.03 | 0.00300 | 0.00300 | 0.09300 | 183 |
| GOCC_APICAL_PLASMA_MEMBRANE | 12 | 0.634 | 1.938 | 0.00300 | 0.00300 | 0.09500 | 104 |
| GOBP_ORGANIC_ACID_METABOLIC_PROCESS | 33 | 0.463 | 1.898 | 0.00300 | 0.00300 | 0.09500 | 23 |
| GOBP_EMBRYONIC_ORGAN_DEVELOPMENT | 14 | 0.598 | 1.917 | 0.00300 | 0.00300 | 0.09500 | 77 |

| | | | | | | | |
|---|---|---|---|---|---|---|---|
| GOBP_ANATOMICAL_STRUCTURE_FORMATION_INVOLVED_IN_MORPHOGENESIS | 30 | 0.477 | 1.902 | 0.00400 | 0.00400 | 0.09500 | 100 |
| GOBP_RNA_SPLICING | 29 | -0.413 | -1.969 | 0.00400 | 0.00400 | 0.09600 | 325 |
| GOMF_SIGNALING_RECEPTOR_BINDING | 31 | 0.47 | 1.891 | 0.00400 | 0.00400 | 0.09800 | 81 |
| GOBP_CELLULAR_RESPONSE_TO_EXTRACELLULAR_STIMULUS | 16 | -0.52 | -2.013 | 0.00500 | 0.00500 | 0.12100 | 128 |
| GOBP_MORPHOGENESIS_OF_AN_EPITHELIUM | 14 | 0.584 | 1.875 | 0.00500 | 0.00500 | 0.12100 | 74 |
| GOBP_ORGANELLE_FISSION | 18 | 0.539 | 1.864 | 0.00500 | 0.00500 | 0.12900 | 103 |
| GOBP_CELLULAR_RESPONSE_TO_ENDOGENOUS_STIMULUS | 54 | 0.388 | 1.787 | 0.00600 | 0.00600 | 0.13000 | 212 |
| GOCC_CELL_SURFACE | 24 | 0.491 | 1.84 | 0.00600 | 0.00600 | 0.13000 | 81 |
| GOBP_AXON_DEVELOPMENT | 14 | 0.577 | 1.85 | 0.00600 | 0.00600 | 0.13000 | 56 |
| HP_INTELLECTUAL_DISABILITY_SEVERE | 16 | -0.514 | -1.99 | 0.00600 | 0.00600 | 0.13000 | 183 |
| HP_ATROPHY_DEGENERATION_AFFECTING_THE_CEREBRUM | 29 | -0.398 | -1.896 | 0.00600 | 0.00600 | 0.13600 | 194 |
| GOBP_MITOTIC_NUCLEAR_DIVISION | 12 | 0.605 | 1.849 | 0.00700 | 0.00700 | 0.13600 | 103 |
| HP_INCREASED_BODY_WEIGHT | 22 | -0.428 | -1.856 | 0.00700 | 0.00700 | 0.13600 | 215 |
| GOBP_TUBE_DEVELOPMENT | 36 | 0.439 | 1.836 | 0.00700 | 0.00700 | 0.13600 | 148 |
| HP_ABNORMAL_CORPUS_CALLOSUM_MORPHOLOGY | 43 | -0.339 | -1.838 | 0.00700 | 0.00700 | 0.13800 | 190 |
| GOBP_SMALL_MOLECULE_METABOLIC_PROCESS | 55 | 0.384 | 1.772 | 0.00700 | 0.00700 | 0.13800 | 27 |
| GOMF_ATP_HYDROLYSIS_ACTIVITY | 13 | -0.527 | -1.876 | 0.00700 | 0.00700 | 0.14000 | 264 |
| HP_ATROPHY_DEGENERATION_AFFECTING_THE_CENTRAL_NERVOUS_SYSTEM | 37 | -0.352 | -1.819 | 0.00700 | 0.00700 | 0.14000 | 194 |
| GOBP_REGULATION_OF_MRNA_PROCESSING | 11 | -0.57 | -1.897 | 0.00800 | 0.00800 | 0.14000 | 314 |
| GOBP_RESPONSE_TO_TRANSFORMING_GROWTH_FACTOR_BETA | 22 | 0.494 | 1.808 | 0.00800 | 0.00800 | 0.14300 | 208 |
| GOMF_DNA_BINDING_TRANSCRIPTION_FACTOR_ACTIVITY | 84 | 0.334 | 1.677 | 0.00800 | 0.00800 | 0.14900 | 280 |
| GOBP_AGING | 17 | 0.532 | 1.805 | 0.00900 | 0.00900 | 0.15200 | 77 |
| GOBP_RESPONSE_TO_NITROGEN_COMPOUND | 43 | 0.405 | 1.764 | 0.00900 | 0.00900 | 0.15900 | 103 |
| HP_INFANTILE_ONSET | 29 | -0.38 | -1.815 | 0.00900 | 0.00900 | 0.16100 | 121 |
| GOBP_NEGATIVE_REGULATION_OF_TRANSPORT | 13 | 0.571 | 1.794 | 0.01000 | 0.01000 | 0.16600 | 18 |
| GOBP_CELLULAR_RESPONSE_TO_STRESS | 86 | -0.245 | -1.578 | 0.01000 | 0.01000 | 0.17100 | 207 |
| GOBP_DNA_REPAIR | 21 | -0.422 | -1.796 | 0.01100 | 0.01100 | 0.17200 | 287 |
| GOBP_REGULATION_OF_CELLULAR_COMPONENT_MOVEMENT | 26 | 0.459 | 1.762 | 0.01100 | 0.01100 | 0.17500 | 77 |
| HP_ABNORMAL_ERYTHROCYTE_MORPHOLOGY | 27 | -0.385 | -1.791 | 0.01100 | 0.01100 | 0.17500 | 183 |
| GOCC_INTRINSIC_COMPONENT_OF_PLASMA_MEMBRANE | 40 | 0.41 | 1.761 | 0.01100 | 0.01100 | 0.17500 | 107 |
| GOBP_TRANSMEMBRANE_TRANSPORT | 49 | -0.314 | -1.763 | 0.01100 | 0.01100 | 0.17500 | 105 |
| GOBP_REGULATION_OF_HORMONE_LEVELS | 14 | 0.553 | 1.775 | 0.01200 | 0.01200 | 0.17500 | 27 |
| HP_ABNORMAL_ANTERIOR_EYE_SEGMENT_MORPHOLOGY | 48 | -0.302 | -1.674 | 0.01200 | 0.01200 | 0.17500 | 215 |
| GOBP_ION_TRANSMEMBRANE_TRANSPORT | 33 | -0.356 | -1.769 | 0.01200 | 0.01200 | 0.17800 | 125 |
| GOBP_FATTY_ACID_METABOLIC_PROCESS | 13 | 0.563 | 1.77 | 0.01200 | 0.01200 | 0.17800 | 167 |
| GOBP_POSITIVE_REGULATION_OF_CATABOLIC_PROCESS | 17 | -0.463 | -1.837 | 0.01300 | 0.01300 | 0.17800 | 341 |
| GOBP_CELLULAR_RESPONSE_TO_BIOTIC_STIMULUS | 11 | -0.552 | -1.839 | 0.01300 | 0.01300 | 0.17800 | 84 |
| GOBP_TISSUE_DEVELOPMENT | 66 | 0.345 | 1.648 | 0.01300 | 0.01300 | 0.17800 | 103 |
| GOCC_CHROMOSOME | 82 | -0.248 | -1.581 | 0.01300 | 0.01300 | 0.17800 | 216 |
| GOBP_SENSORY_ORGAN_MORPHOGENESIS | 15 | 0.54 | 1.768 | 0.01300 | 0.01300 | 0.17800 | 74 |
| GOCC_APICAL_PART_OF_CELL | 15 | 0.54 | 1.767 | 0.01300 | 0.01300 | 0.17800 | 104 |
| GOMF_RIBONUCLEOPROTEIN_COMPLEX_BINDING | 12 | -0.526 | -1.82 | 0.01300 | 0.01300 | 0.17800 | 305 |
| HP_ABNORMAL_AGGRESSIVE_IMPULSIVE_OR_VIOLENT_BEHAVIOR | 17 | -0.46 | -1.825 | 0.01300 | 0.01300 | 0.17800 | 207 |

| | | | | | | | |
|---|---|---|---|---|---|---|---|
| GOBP_POSITIVE_REGULATION_OF_NUCLEOBASE_CONTAINING_COMPOUND_ME | 91 | -0.231 | -1.516 | 0.01400 | 0.01400 | 0.17800 | 246 |
| GOBP_REGULATION_OF_TRANSPORT | 66 | -0.265 | -1.596 | 0.01400 | 0.01400 | 0.18100 | 131 |
| GOBP_POSITIVE_REGULATION_OF_RNA_METABOLIC_PROCESS | 89 | -0.233 | -1.521 | 0.01400 | 0.01400 | 0.18100 | 246 |
| GOBP_RNA_CATABOLIC_PROCESS | 14 | -0.487 | -1.795 | 0.01400 | 0.01400 | 0.18100 | 314 |
| GOCC_ENDOPLASMIC_RETICULUM_LUMEN | 11 | 0.587 | 1.75 | 0.01500 | 0.01500 | 0.18100 | 129 |
| GOBP_G_PROTEIN_COUPLED_RECEPTOR_SIGNALING_PATHWAY | 20 | 0.486 | 1.732 | 0.01500 | 0.01500 | 0.18100 | 141 |
| HP_ABNORMAL_MYELOID_LEUKOCYTE_MORPHOLOGY | 11 | -0.543 | -1.808 | 0.01500 | 0.01500 | 0.18100 | 166 |
| GOBP_MUSCLE_TISSUE_DEVELOPMENT | 12 | 0.572 | 1.75 | 0.01500 | 0.01500 | 0.18100 | 103 |
| HP_BILATERAL_TONIC_CLONIC_SEIZURE | 10 | -0.559 | -1.8 | 0.01500 | 0.01500 | 0.18100 | 153 |
| HP_GLUCOSE_INTOLERANCE | 13 | -0.501 | -1.78 | 0.01600 | 0.01600 | 0.18100 | 185 |
| GOBP_POSITIVE_REGULATION_OF_CELLULAR_BIOSYNTHETIC_PROCESS | 93 | -0.227 | -1.495 | 0.01600 | 0.01600 | 0.18100 | 246 |
| HP_CARIOUS_TEETH | 12 | -0.52 | -1.8 | 0.01600 | 0.01600 | 0.18100 | 215 |
| GOBP_GLAND_DEVELOPMENT | 16 | 0.521 | 1.739 | 0.01600 | 0.01600 | 0.18100 | 77 |
| GOMF_PROTEIN_HOMODIMERIZATION_ACTIVITY | 25 | 0.453 | 1.718 | 0.01600 | 0.01600 | 0.18100 | 103 |
| GOBP_CIRCULATORY_SYSTEM_PROCESS | 20 | 0.483 | 1.721 | 0.01700 | 0.01700 | 0.18500 | 115 |
| HP_ABNORMALITY_OF_THE_OPTIC_NERVE | 40 | -0.321 | -1.695 | 0.01700 | 0.01700 | 0.18500 | 194 |
| GOBP_RESPONSE_TO_STARVATION | 15 | -0.469 | -1.769 | 0.01700 | 0.01700 | 0.18700 | 202 |
| GOMF_TRANSCRIPTION_COREGULATOR_ACTIVITY | 33 | -0.34 | -1.69 | 0.01800 | 0.01800 | 0.19400 | 223 |
| GOCC_ENDOSOME_MEMBRANE | 23 | -0.394 | -1.73 | 0.01800 | 0.01800 | 0.19400 | 177 |
| HP_AUTISTIC_BEHAVIOR | 29 | -0.357 | -1.704 | 0.01800 | 0.01800 | 0.19400 | 215 |
| GOBP_RNA_SPLICING_VIA_TRANSESTERIFICATION_REACTIONS | 19 | -0.425 | -1.741 | 0.01900 | 0.01900 | 0.19600 | 314 |
| HP_FUNCTIONAL_ABNORMALITY_OF_THE_INNER_EAR | 33 | -0.335 | -1.665 | 0.01900 | 0.01900 | 0.19600 | 191 |
| GOBP_POSITIVE_REGULATION_OF_EPITHELIAL_CELL_PROLIFERATION | 10 | 0.591 | 1.704 | 0.01900 | 0.01900 | 0.19700 | 167 |
| HP_SINGLE_TRANSVERSE_PALMAR_CREASE | 14 | -0.476 | -1.754 | 0.02000 | 0.02000 | 0.20300 | 381 |
| GOBP_REPRODUCTION | 49 | 0.365 | 1.644 | 0.02000 | 0.02000 | 0.20400 | 149 |
| GOBP_REGULATION_OF_ANATOMICAL_STRUCTURE_SIZE | 12 | 0.559 | 1.71 | 0.02000 | 0.02000 | 0.20600 | 35 |
| GOBP_POSITIVE_REGULATION_OF_PROTEIN_MODIFICATION_PROCESS | 46 | 0.372 | 1.653 | 0.02100 | 0.02100 | 0.20600 | 77 |
| GOBP_MULTI_MULTICELLULAR_ORGANISM_PROCESS | 14 | 0.528 | 1.695 | 0.02100 | 0.02100 | 0.21100 | 94 |
| GOBP_SMALL_MOLECULE_BIOSYNTHETIC_PROCESS | 11 | 0.574 | 1.709 | 0.02100 | 0.02100 | 0.21100 | 74 |
| HP_PEDIATRIC_ONSET | 34 | -0.33 | -1.653 | 0.02200 | 0.02200 | 0.21200 | 121 |
| GOBP_I_KAPPAB_KINASE_NF_KAPPAB_SIGNALING | 11 | 0.571 | 1.703 | 0.02200 | 0.02200 | 0.21200 | 70 |
| GOBP_CELL_DIVISION | 25 | 0.437 | 1.657 | 0.02200 | 0.02200 | 0.21200 | 225 |
| GOBP_AMIDE_BIOSYNTHETIC_PROCESS | 34 | -0.328 | -1.647 | 0.02200 | 0.02200 | 0.21200 | 247 |
| GOCC_NUCLEAR_SPECK | 25 | -0.373 | -1.701 | 0.02300 | 0.02300 | 0.21200 | 314 |
| HP_SELF_INJURIOUS_BEHAVIOR | 10 | -0.532 | -1.711 | 0.02300 | 0.02300 | 0.21400 | 207 |
| GOBP_RESPONSE_TO_ENDOGENOUS_STIMULUS | 63 | 0.335 | 1.588 | 0.02400 | 0.02400 | 0.21700 | 212 |
| GOBP_REGULATION_OF_CELLULAR_AMIDE_METABOLIC_PROCESS | 21 | -0.396 | -1.685 | 0.02400 | 0.02400 | 0.21700 | 247 |
| GOBP_STEROID_METABOLIC_PROCESS | 10 | 0.579 | 1.67 | 0.02400 | 0.02400 | 0.21700 | 27 |
| GOMF_TRANSPORTER_ACTIVITY | 43 | -0.295 | -1.599 | 0.02400 | 0.02400 | 0.21700 | 76 |
| HP_UPPER_MOTOR_NEURON_DYSFUNCTION | 39 | -0.31 | -1.628 | 0.02400 | 0.02400 | 0.21700 | 120 |
| GOBP_CELLULAR_RESPONSE_TO_EXTERNAL_STIMULUS | 17 | -0.431 | -1.708 | 0.02400 | 0.02400 | 0.21700 | 128 |
| GOBP_NEURON_DEATH | 11 | -0.517 | -1.721 | 0.02500 | 0.02500 | 0.21800 | 62 |
| GOBP_CELL_MORPHOGENESIS_INVOLVED_IN_DIFFERENTIATION | 21 | 0.459 | 1.659 | 0.02500 | 0.02500 | 0.22000 | 78 |

| | | | | | | | |
|---|---|---|---|---|---|---|---|
| GOBP_CIRCADIAN_RHYTHM | 11 | -0.516 | -1.717 | 0.02500 | 0.02500 | 0.22200 | 352 |
| GOBP_MEMBRANE_ORGANIZATION | 38 | -0.309 | -1.602 | 0.02700 | 0.02700 | 0.22900 | 145 |
| GOBP_REGULATION_OF_ANATOMICAL_STRUCTURE_MORPHOGENESIS | 27 | 0.422 | 1.636 | 0.02700 | 0.02700 | 0.22900 | 100 |
| GOBP_REGULATION_OF_CELL_POPULATION_PROLIFERATION | 49 | 0.356 | 1.603 | 0.02800 | 0.02800 | 0.23400 | 77 |
| GOBP_REGULATION_OF_PROTEIN_CONTAINING_COMPLEX_ASSEMBLY | 17 | -0.425 | -1.686 | 0.02800 | 0.02800 | 0.23400 | 156 |
| GOBP_NEGATIVE_REGULATION_OF_MULTICELLULAR_ORGANISMAL_PROCESS | 25 | 0.428 | 1.622 | 0.02800 | 0.02800 | 0.23400 | 85 |
| HP_ABNORMALITY_OF_UPPER_LIP | 44 | -0.288 | -1.576 | 0.02900 | 0.02900 | 0.23700 | 165 |
| GOBP_DEVELOPMENTAL_PROCESS_INVOLVED_IN_REPRODUCTION | 34 | 0.395 | 1.627 | 0.02900 | 0.02900 | 0.23700 | 149 |
| GOMF_ADENYL_NUCLEOTIDE_BINDING | 54 | -0.267 | -1.535 | 0.02900 | 0.02900 | 0.23700 | 287 |
| GOMF_NUCLEAR_RECEPTOR_BINDING | 10 | -0.523 | -1.684 | 0.02900 | 0.02900 | 0.23700 | 201 |
| HP_ABNORMAL_BRONCHUS_MORPHOLOGY | 11 | 0.554 | 1.651 | 0.03000 | 0.03000 | 0.24000 | 59 |
| GOBP_REGULATION_OF_RESPONSE_TO_EXTERNAL_STIMULUS | 28 | 0.414 | 1.62 | 0.03000 | 0.03000 | 0.24000 | 74 |
| GOCC_ANCHORING_JUNCTION | 34 | 0.391 | 1.614 | 0.03100 | 0.03100 | 0.24000 | 104 |
| GOCC_CELL_CELL_JUNCTION | 15 | 0.498 | 1.629 | 0.03100 | 0.03100 | 0.24000 | 104 |
| HP_PACE_OF_PROGRESSION | 16 | -0.434 | -1.68 | 0.03100 | 0.03100 | 0.24000 | 239 |
| GOCC_NEURON_TO_NEURON_SYNAPSE | 16 | -0.434 | -1.679 | 0.03100 | 0.03100 | 0.24000 | 125 |
| GOBP_CATION_TRANSMEMBRANE_TRANSPORT | 30 | -0.336 | -1.627 | 0.03100 | 0.03100 | 0.24000 | 125 |
| GOBP_POSITIVE_REGULATION_OF_CELL_CYCLE_PROCESS | 13 | 0.523 | 1.645 | 0.03200 | 0.03200 | 0.24000 | 73 |
| HP_VISUAL_IMPAIRMENT | 44 | -0.286 | -1.566 | 0.03200 | 0.03200 | 0.24000 | 127 |
| HP_GLAUCOMA | 12 | -0.486 | -1.685 | 0.03200 | 0.03200 | 0.24000 | 215 |
| GOBP_SKELETAL_SYSTEM_DEVELOPMENT | 24 | 0.43 | 1.611 | 0.03200 | 0.03200 | 0.24000 | 81 |
| HP_ABNORMALITY_OF_UPPER_LIP_VERMILLION | 21 | -0.381 | -1.621 | 0.03300 | 0.03300 | 0.24000 | 156 |
| HP_ABNORMAL_TRACHEOBRONCHIAL_MORPHOLOGY | 17 | 0.48 | 1.63 | 0.03300 | 0.03300 | 0.24000 | 59 |
| GOMF_PROTEIN_N_TERMINUS_BINDING | 10 | -0.517 | -1.666 | 0.03300 | 0.03300 | 0.24000 | 137 |
| HP_UPSLANTED_PALPEBRAL_FISSURE | 16 | -0.432 | -1.672 | 0.03300 | 0.03300 | 0.24000 | 291 |
| GOBP_NEGATIVE_REGULATION_OF_RESPONSE_TO_STIMULUS | 44 | 0.361 | 1.582 | 0.03300 | 0.03300 | 0.24300 | 204 |
| GOBP_POSITIVE_REGULATION_OF_PROTEIN_CONTAINING_COMPLEX_ASSEMBLY | 12 | -0.482 | -1.669 | 0.03400 | 0.03400 | 0.24900 | 130 |
| HP_HYPERREFLEXIA | 25 | -0.355 | -1.615 | 0.03500 | 0.03500 | 0.24900 | 207 |
| GOBP_HOMEOSTATIC_PROCESS | 61 | -0.249 | -1.478 | 0.03500 | 0.03500 | 0.24900 | 80 |
| HP_ABNORMALITY_OF_THE_VASCULATURE_OF_THE_EYE | 11 | -0.493 | -1.641 | 0.03500 | 0.03500 | 0.24900 | 215 |
| GOBP_WOUND_HEALING | 16 | 0.482 | 1.609 | 0.03500 | 0.03500 | 0.24900 | 167 |
| GOBP_POSITIVE_REGULATION_OF_PROTEIN_METABOLIC_PROCESS | 59 | 0.331 | 1.551 | 0.03600 | 0.03600 | 0.24900 | 77 |
| GOBP_CELLULAR_LIPID_METABOLIC_PROCESS | 34 | 0.384 | 1.582 | 0.03700 | 0.03700 | 0.25300 | 242 |
| HP_BRACHYCEPHALY | 15 | -0.438 | -1.652 | 0.03700 | 0.03700 | 0.25300 | 300 |
| GOBP_TISSUE_MIGRATION | 10 | 0.557 | 1.605 | 0.03800 | 0.03800 | 0.25600 | 74 |
| GOBP_HEAD_DEVELOPMENT | 27 | -0.343 | -1.597 | 0.03800 | 0.03800 | 0.25600 | 96 |
| GOMF_OXIDOREDUCTASE_ACTIVITY | 25 | 0.418 | 1.584 | 0.03800 | 0.03800 | 0.25800 | 209 |
| GOBP_HEART_DEVELOPMENT | 16 | 0.479 | 1.6 | 0.03800 | 0.03800 | 0.25800 | 77 |
| GOBP_REGULATION_OF_RNA_SPLICING | 14 | -0.443 | -1.629 | 0.03800 | 0.03800 | 0.25800 | 314 |
| GOCC_TRANSPORT_VESICLE | 20 | 0.45 | 1.603 | 0.03900 | 0.03900 | 0.25800 | 52 |
| HP_SPASTICITY | 30 | -0.326 | -1.581 | 0.03900 | 0.03900 | 0.25800 | 166 |
| GOBP_PATTERN_SPECIFICATION_PROCESS | 17 | 0.471 | 1.6 | 0.03900 | 0.03900 | 0.25800 | 77 |
| GOBP_RIBONUCLEOPROTEIN_COMPLEX_BIOGENESIS | 25 | -0.35 | -1.594 | 0.04000 | 0.04000 | 0.26100 | 305 |

| | | | | | | | |
|---|---|---|---|---|---|---|---|
| GOBP_MRNA_CATABOLIC_PROCESS | 12 | -0.472 | -1.634 | 0.04100 | 0.04100 | 0.26500 | 314 |
| HP_ONSET | 59 | -0.25 | -1.471 | 0.04200 | 0.04200 | 0.27300 | 110 |
| HP_ABNORMALITY_OF_FACIAL_SOFT_TISSUE | 15 | -0.427 | -1.612 | 0.04300 | 0.04300 | 0.27700 | 183 |
| HP_HYPERTONIA | 40 | -0.289 | -1.526 | 0.04400 | 0.04400 | 0.27700 | 194 |
| HP_GASTROESOPHAGEAL_REFLUX | 21 | -0.368 | -1.565 | 0.04400 | 0.04400 | 0.27700 | 251 |
| GOMF_G_PROTEIN_COUPLED_RECEPTOR_BINDING | 12 | 0.516 | 1.579 | 0.04400 | 0.04400 | 0.27700 | 68 |
| GOBP_POSTTRANSCRIPTIONAL_REGULATION_OF_GENE_EXPRESSION | 24 | -0.353 | -1.579 | 0.04400 | 0.04400 | 0.27700 | 335 |
| GOBP_EPIDERMIS_DEVELOPMENT | 17 | -0.403 | -1.596 | 0.04500 | 0.04500 | 0.28200 | 60 |
| GOBP_POSITIVE_REGULATION_OF_PHOSPHORUS_METABOLIC_PROCESS | 33 | 0.376 | 1.539 | 0.04600 | 0.04600 | 0.28200 | 132 |
| GOBP_INORGANIC_ION_TRANSMEMBRANE_TRANSPORT | 28 | -0.33 | -1.565 | 0.04600 | 0.04600 | 0.28200 | 125 |
| GOBP_REGULATION_OF_INTRACELLULAR_SIGNAL_TRANSDUCTION | 61 | 0.318 | 1.499 | 0.04600 | 0.04600 | 0.28200 | 112 |
| GOBP_VASCULAR_PROCESS_IN_CIRCULATORY_SYSTEM | 11 | 0.526 | 1.566 | 0.04700 | 0.04700 | 0.28200 | 115 |
| GOBP_REGULATION_OF_PROTEIN_SERINE_THREONINE_KINASE_ACTIVITY | 18 | 0.454 | 1.572 | 0.04700 | 0.04700 | 0.28200 | 95 |
| GOBP_BEHAVIOR | 18 | -0.392 | -1.581 | 0.04700 | 0.04700 | 0.28200 | 77 |
| GOBP_EPIDERMAL_CELL_DIFFERENTIATION | 10 | -0.496 | -1.595 | 0.04700 | 0.04700 | 0.28200 | 60 |
| GOBP_MULTI_ORGANISM_REPRODUCTIVE_PROCESS | 37 | 0.364 | 1.532 | 0.04800 | 0.04800 | 0.28600 | 94 |
| HP_ABNORMALITY_OF_THE_LENS | 31 | -0.312 | -1.531 | 0.04900 | 0.04900 | 0.29100 | 215 |
| HP_RECURRENT_RESPIRATORY_INFECTIONS | 24 | 0.41 | 1.536 | 0.05000 | 0.05000 | 0.29300 | 28 |
| GOBP_POSITIVE_REGULATION_OF_CELLULAR_COMPONENT_BIOGENESIS | 24 | -0.346 | -1.546 | 0.05000 | 0.05000 | 0.29300 | 141 |
| HP_OPTIC_ATROPHY | 32 | -0.309 | -1.526 | 0.05000 | 0.05000 | 0.29300 | 194 |
| GOMF_TRANSCRIPTION_FACTOR_BINDING | 31 | -0.311 | -1.527 | 0.05000 | 0.05000 | 0.29300 | 261 |
| GOBP_REGULATION_OF_BINDING | 16 | -0.406 | -1.572 | 0.05100 | 0.05100 | 0.29400 | 431 |
| HP_ABNORMAL_TONGUE_MORPHOLOGY | 20 | 0.436 | 1.554 | 0.05100 | 0.05100 | 0.29400 | 210 |
| GOBP_SENSORY_ORGAN_DEVELOPMENT | 23 | 0.416 | 1.54 | 0.05100 | 0.05100 | 0.29400 | 74 |
| GOBP_AMEBOIDAL_TYPE_CELL_MIGRATION | 11 | 0.519 | 1.545 | 0.05200 | 0.05200 | 0.29500 | 74 |
| HP_ABNORMALITY_OF_CENTRAL_NERVOUS_SYSTEM_ELECTROPHYSIOLOGY | 23 | -0.351 | -1.542 | 0.05200 | 0.05200 | 0.29600 | 207 |
| GOBP_REGULATION_OF_BLOOD_CIRCULATION | 10 | 0.537 | 1.548 | 0.05300 | 0.05300 | 0.29800 | 115 |
| HP_DELAYED_ERUPTION_OF_TEETH | 11 | 0.518 | 1.542 | 0.05300 | 0.05300 | 0.29800 | 20 |
| GOCC_EARLY_ENDOSOME | 12 | -0.458 | -1.586 | 0.05300 | 0.05300 | 0.29800 | 161 |
| GOBP_REGULATION_OF_MICROTUBULE_BASED_PROCESS | 11 | 0.517 | 1.539 | 0.05400 | 0.05400 | 0.29900 | 162 |
| GOBP_LIPID_BIOSYNTHETIC_PROCESS | 26 | 0.398 | 1.53 | 0.05500 | 0.05500 | 0.29900 | 242 |
| HP_ABNORMALITY_OF_THE_BLADDER | 14 | -0.426 | -1.568 | 0.05500 | 0.05500 | 0.29900 | 238 |
| GOBP_RHYTHMIC_PROCESS | 14 | -0.426 | -1.567 | 0.05500 | 0.05500 | 0.29900 | 352 |
| GOMF_MOLECULAR_TRANSDUCER_ACTIVITY | 20 | 0.432 | 1.541 | 0.05500 | 0.05500 | 0.29900 | 208 |
| GOBP_EPITHELIUM_DEVELOPMENT | 41 | 0.349 | 1.503 | 0.05500 | 0.05500 | 0.30000 | 78 |
| GOBP_CELLULAR_RESPONSE_TO_STARVATION | 13 | -0.434 | -1.543 | 0.05700 | 0.05700 | 0.30600 | 199 |
| GOBP_REGULATION_OF_CELLULAR_RESPONSE_TO_STRESS | 25 | -0.334 | -1.521 | 0.05700 | 0.05700 | 0.30600 | 168 |
| GOBP_RRNA_METABOLIC_PROCESS | 12 | -0.454 | -1.572 | 0.05700 | 0.05700 | 0.30600 | 250 |
| GOMF_INORGANIC_MOLECULAR_ENTITY_TRANSMEMBRANE_TRANSPORTER_ACT | 18 | -0.382 | -1.542 | 0.05800 | 0.05800 | 0.30600 | 95 |
| HP_ABNORMAL_EYELID_MORPHOLOGY | 57 | -0.246 | -1.433 | 0.05800 | 0.05800 | 0.30600 | 166 |
| GOBP_RESPONSE_TO_GROWTH_FACTOR | 31 | 0.376 | 1.51 | 0.05800 | 0.05800 | 0.30600 | 208 |
| HP_INTELLECTUAL_DISABILITY_MODERATE | 10 | -0.484 | -1.557 | 0.05800 | 0.05800 | 0.30600 | 183 |
| HP_ABNORMALITY_OF_THE_TONGUE | 23 | 0.412 | 1.521 | 0.05900 | 0.05900 | 0.30600 | 210 |

| | | | | | | | |
|---|---|---|---|---|---|---|---|
| GOBP_CELL_MORPHOGENESIS | 25 | 0.396 | 1.502 | 0.06000 | 0.06000 | 0.31100 | 78 |
| GOMF_TRANSFERASE_ACTIVITY_TRANSFERRING_ONE_CARBON_GROUPS | 13 | -0.43 | -1.528 | 0.06000 | 0.06000 | 0.31100 | 96 |
| GOCC_SYNAPTIC_MEMBRANE | 14 | 0.477 | 1.53 | 0.06100 | 0.06100 | 0.31100 | 115 |
| GOMF_CATION_TRANSMEMBRANE_TRANSPORTER_ACTIVITY | 19 | -0.368 | -1.51 | 0.06100 | 0.06100 | 0.31100 | 95 |
| GOBP_REGULATION_OF_MRNA_CATABOLIC_PROCESS | 11 | -0.459 | -1.528 | 0.06100 | 0.06100 | 0.31100 | 314 |
| HP_ABNORMAL_PENIS_MORPHOLOGY | 32 | -0.3 | -1.484 | 0.06200 | 0.06200 | 0.31100 | 215 |
| GOMF_METAL_ION_TRANSMEMBRANE_TRANSPORTER_ACTIVITY | 17 | -0.388 | -1.539 | 0.06200 | 0.06200 | 0.31100 | 76 |
| GOMF_UBIQUITIN_LIKE_PROTEIN_LIGASE_BINDING | 12 | -0.447 | -1.547 | 0.06200 | 0.06200 | 0.31100 | 132 |
| GOBP_BIOLOGICAL_ADHESION | 27 | 0.389 | 1.508 | 0.06200 | 0.06200 | 0.31100 | 91 |
| HP_THIN_UPPER_LIP_VERMILION | 19 | -0.367 | -1.505 | 0.06300 | 0.06300 | 0.31100 | 156 |
| GOBP_PROTEIN_STABILIZATION | 13 | -0.425 | -1.513 | 0.06300 | 0.06300 | 0.31100 | 192 |
| GOMF_ION_TRANSMEMBRANE_TRANSPORTER_ACTIVITY | 23 | -0.344 | -1.513 | 0.06300 | 0.06300 | 0.31200 | 95 |
| GOMF_PROTEIN_DIMERIZATION_ACTIVITY | 41 | 0.342 | 1.475 | 0.06300 | 0.06300 | 0.31200 | 182 |
| GOBP_RESPONSE_TO_CYTOKINE | 35 | -0.289 | -1.462 | 0.06500 | 0.06500 | 0.31800 | 119 |
| GOBP_REGULATION_OF_PROTEIN_PHOSPHORYLATION | 43 | 0.334 | 1.456 | 0.06600 | 0.06600 | 0.32300 | 111 |
| GOMF_HEXOSYLTRANSFERASE_ACTIVITY | 10 | 0.522 | 1.506 | 0.06700 | 0.06700 | 0.32600 | 210 |
| GOBP_EXPORT_FROM_CELL | 26 | 0.386 | 1.483 | 0.07000 | 0.07000 | 0.33500 | 35 |
| GOBP_POSITIVE_REGULATION_OF_PROTEIN_SERINE_THREONINE_KINASE_ACTIVI | 13 | 0.48 | 1.508 | 0.07000 | 0.07000 | 0.33500 | 77 |
| GOMF_TRANSFERASE_ACTIVITY_TRANSFERRING_PHOSPHORUS_CONTAINING_G | 28 | -0.312 | -1.476 | 0.07100 | 0.07100 | 0.33500 | 260 |
| GOBP_GAMETE_GENERATION | 24 | 0.394 | 1.476 | 0.07200 | 0.07200 | 0.33500 | 25 |
| GOBP_CELLULAR_RESPONSE_TO_NITROGEN_COMPOUND | 22 | 0.405 | 1.48 | 0.07200 | 0.07200 | 0.33500 | 103 |
| GOBP_CELL_CELL_ADHESION | 15 | 0.455 | 1.491 | 0.07200 | 0.07200 | 0.33500 | 72 |
| HP_ABNORMAL_SIZE_OF_THE_PALPEBRAL_FISSURES | 18 | -0.373 | -1.504 | 0.07200 | 0.07200 | 0.33500 | 314 |
| GOBP_EPITHELIAL_TUBE_MORPHOGENESIS | 12 | 0.487 | 1.488 | 0.07200 | 0.07200 | 0.33500 | 74 |
| HP_AUTISM | 12 | -0.434 | -1.505 | 0.07200 | 0.07200 | 0.33500 | 215 |
| HP_INTELLECTUAL_DISABILITY_PROFOUND | 10 | -0.471 | -1.516 | 0.07200 | 0.07200 | 0.33500 | 120 |
| GOBP_MUSCLE_STRUCTURE_DEVELOPMENT | 22 | 0.404 | 1.478 | 0.07200 | 0.07200 | 0.33500 | 206 |
| GOBP_CENTRAL_NERVOUS_SYSTEM_DEVELOPMENT | 27 | -0.315 | -1.463 | 0.07300 | 0.07300 | 0.33500 | 116 |
| HP_WEAKNESS_DUE_TO_UPPER_MOTOR_NEURON_DYSFUNCTION | 14 | -0.406 | -1.496 | 0.07300 | 0.07300 | 0.33500 | 166 |
| GOMF_CIS_REGULATORY_REGION_SEQUENCE_SPECIFIC_DNA_BINDING | 74 | 0.287 | 1.409 | 0.07300 | 0.07300 | 0.33600 | 265 |
| GOBP_REGULATION_OF_CELLULAR_COMPONENT_BIOGENESIS | 38 | -0.273 | -1.418 | 0.07400 | 0.07400 | 0.33800 | 159 |
| GOBP_APOPTOTIC_SIGNALING_PATHWAY | 20 | -0.352 | -1.474 | 0.07500 | 0.07500 | 0.33800 | 180 |
| HP_ABNORMALITY_OF_PELVIC_GIRDLE_BONE_MORPHOLOGY | 34 | -0.287 | -1.437 | 0.07500 | 0.07500 | 0.33800 | 176 |
| GOBP_CELLULAR_MACROMOLECULE_BIOSYNTHETIC_PROCESS | 70 | -0.223 | -1.364 | 0.07600 | 0.07600 | 0.33800 | 249 |
| GOBP_POSITIVE_REGULATION_OF_PROTEIN_PHOSPHORYLATION | 30 | 0.366 | 1.461 | 0.07600 | 0.07600 | 0.33800 | 77 |
| GOMF_STRUCTURAL_MOLECULE_ACTIVITY | 11 | -0.447 | -1.487 | 0.07600 | 0.07600 | 0.33800 | 58 |
| GOBP_HOMEOSTASIS_OF_NUMBER_OF_CELLS | 11 | -0.446 | -1.486 | 0.07600 | 0.07600 | 0.33800 | 60 |
| GOBP_REGULATION_OF_CELL_CYCLE | 53 | 0.313 | 1.431 | 0.07700 | 0.07700 | 0.33800 | 103 |
| GOMF_MOLECULAR_FUNCTION_REGULATOR | 71 | 0.29 | 1.405 | 0.07700 | 0.07700 | 0.33800 | 101 |
| GOBP_SYNAPSE_ORGANIZATION | 10 | -0.468 | -1.505 | 0.07700 | 0.07700 | 0.33800 | 128 |
| GOBP_REGULATION_OF_PEPTIDASE_ACTIVITY | 11 | 0.496 | 1.477 | 0.07800 | 0.07800 | 0.33800 | 25 |
| HP_ABNORMALITY_OF_THE_LOWER_URINARY_TRACT | 29 | -0.305 | -1.453 | 0.07800 | 0.07800 | 0.33800 | 215 |
| GOCC_MEMBRANE_PROTEIN_COMPLEX | 42 | 0.33 | 1.432 | 0.07800 | 0.07800 | 0.33800 | 120 |

| | | | | | | | |
|---|---|---|---|---|---|---|---|
| GOBP_DEVELOPMENTAL_GROWTH | 18 | 0.426 | 1.474 | 0.07800 | 0.07800 | 0.33800 | 77 |
| GOBP_SYNAPTIC_SIGNALING | 22 | -0.338 | -1.465 | 0.07900 | 0.07900 | 0.33800 | 123 |
| GOBP_POSITIVE_REGULATION_OF_CELL_DEATH | 22 | -0.338 | -1.463 | 0.07900 | 0.07900 | 0.33800 | 90 |
| GOBP_CELLULAR_COMPONENT_DISASSEMBLY | 21 | -0.343 | -1.458 | 0.08000 | 0.08000 | 0.34000 | 285 |
| GOBP_REGULATION_OF_PROTEIN_MODIFICATION_PROCESS | 71 | 0.288 | 1.4 | 0.08000 | 0.08000 | 0.34000 | 115 |
| GOBP_CANONICAL_WNT_SIGNALING_PATHWAY | 13 | 0.47 | 1.477 | 0.08100 | 0.08100 | 0.34200 | 74 |
| GOBP_POSITIVE_REGULATION_OF_TRANSMEMBRANE_RECEPTOR_PROTEIN_SERI | 11 | 0.494 | 1.471 | 0.08100 | 0.08100 | 0.34200 | 245 |
| GOBP_VESICLE_MEDIATED_TRANSPORT | 67 | -0.223 | -1.354 | 0.08100 | 0.08100 | 0.34200 | 130 |
| HP_APLASIA_HYPOPLASIA_OF_THE_CEREBELLUM | 26 | -0.316 | -1.449 | 0.08200 | 0.08200 | 0.34300 | 190 |
| GOMF_CATALYTIC_ACTIVITY_ACTING_ON_RNA | 23 | -0.333 | -1.463 | 0.08200 | 0.08200 | 0.34400 | 268 |
| GOBP_REGULATION_OF_PROTEIN_KINASE_ACTIVITY | 26 | 0.376 | 1.446 | 0.08300 | 0.08300 | 0.34500 | 95 |
| GOMF_TRANSCRIPTION_COACTIVATOR_ACTIVITY | 20 | -0.347 | -1.453 | 0.08400 | 0.08400 | 0.34700 | 221 |
| GOBP_REGULATION_OF_VESICLE_MEDIATED_TRANSPORT | 19 | -0.354 | -1.452 | 0.08500 | 0.08500 | 0.35300 | 130 |
| GOBP_POSITIVE_REGULATION_OF_DEVELOPMENTAL_PROCESS | 40 | 0.328 | 1.412 | 0.08700 | 0.08700 | 0.35800 | 218 |
| GOBP_RAS_PROTEIN_SIGNAL_TRANSDUCTION | 18 | 0.42 | 1.454 | 0.08700 | 0.08700 | 0.35800 | 101 |
| GOBP_NERVOUS_SYSTEM_PROCESS | 29 | -0.3 | -1.429 | 0.08700 | 0.08700 | 0.35800 | 36 |
| HP_POSTNATAL_GROWTH_RETARDATION | 10 | -0.457 | -1.471 | 0.08800 | 0.08800 | 0.35800 | 207 |
| HP_ABNORMAL_FACIAL_SHAPE | 46 | 0.316 | 1.402 | 0.08900 | 0.08900 | 0.35900 | 77 |
| GOBP_RNA_LOCALIZATION | 12 | -0.42 | -1.454 | 0.08900 | 0.08900 | 0.36000 | 264 |
| GOBP_PEPTIDE_METABOLIC_PROCESS | 35 | -0.277 | -1.405 | 0.09000 | 0.09000 | 0.36000 | 247 |
| HP_ABNORMALITY_OF_DENTAL_ERUPTION | 15 | 0.44 | 1.442 | 0.09000 | 0.09000 | 0.36000 | 59 |
| GOBP_INTRACELLULAR_TRANSPORT | 82 | -0.207 | -1.319 | 0.09000 | 0.09000 | 0.36000 | 286 |
| HP_MACROTIA | 13 | -0.404 | -1.439 | 0.09000 | 0.09000 | 0.36000 | 165 |
| GOMF_GTPASE_ACTIVITY | 11 | 0.487 | 1.45 | 0.09100 | 0.09100 | 0.36000 | 141 |
| GOBP_REGULATION_OF_CELL_CYCLE_PROCESS | 34 | 0.344 | 1.417 | 0.09100 | 0.09100 | 0.36100 | 103 |
| HP_TUBE_FEEDING | 12 | -0.419 | -1.451 | 0.09200 | 0.09200 | 0.36200 | 207 |
| GOBP_RESPONSE_TO_RADIATION | 18 | -0.359 | -1.446 | 0.09200 | 0.09200 | 0.36200 | 168 |
| GOBP_VIRAL_PROCESS | 25 | -0.314 | -1.429 | 0.09200 | 0.09200 | 0.36200 | 204 |
| HP_IRRITABILITY | 10 | -0.453 | -1.459 | 0.09400 | 0.09400 | 0.36600 | 191 |
| GOBP_REGULATION_OF_CATABOLIC_PROCESS | 46 | -0.25 | -1.375 | 0.09400 | 0.09400 | 0.36600 | 247 |
| GOBP_SECRETION | 28 | 0.361 | 1.415 | 0.09500 | 0.09500 | 0.36600 | 35 |
| GOBP_POSITIVE_REGULATION_OF_PROTEIN_KINASE_ACTIVITY | 19 | 0.409 | 1.439 | 0.09500 | 0.09500 | 0.36700 | 77 |
| GOMF_ENZYME_ACTIVATOR_ACTIVITY | 27 | -0.302 | -1.403 | 0.09600 | 0.09600 | 0.36900 | 214 |
| GOCC_CELL_LEADING_EDGE | 12 | 0.47 | 1.437 | 0.09600 | 0.09600 | 0.37000 | 112 |
| GOBP_ACTIN_FILAMENT_ORGANIZATION | 12 | -0.415 | -1.436 | 0.09700 | 0.09700 | 0.37000 | 123 |
| GOBP_CELLULAR_NITROGEN_COMPOUND_CATABOLIC_PROCESS | 17 | -0.36 | -1.427 | 0.09800 | 0.09800 | 0.37200 | 314 |
| HP_MACULE | 15 | -0.375 | -1.414 | 0.09800 | 0.09800 | 0.37200 | 300 |
| HP_AGENESIS_OF_CORPUS_CALLOSUM | 17 | -0.359 | -1.424 | 0.09900 | 0.09900 | 0.37400 | 207 |
| HP_ABNORMALITY_OF_THE_BILIARY_SYSTEM | 19 | 0.406 | 1.429 | 0.09900 | 0.09900 | 0.37500 | 178 |
| HP_ABNORMAL_IMMUNE_SYSTEM_MORPHOLOGY | 23 | -0.322 | -1.417 | 0.10100 | 0.10100 | 0.37900 | 207 |
| HP_CEREBELLAR_ATROPHY | 13 | -0.396 | -1.407 | 0.10300 | 0.10300 | 0.38500 | 104 |
| HP_ABNORMALITY_OF_THE_CALVARIA | 50 | -0.238 | -1.347 | 0.10300 | 0.10300 | 0.38500 | 215 |
| HP_HYPERPIGMENTATION_OF_THE_SKIN | 15 | -0.371 | -1.4 | 0.10400 | 0.10400 | 0.38500 | 300 |

| | | | | | | | |
|---|---|---|---|---|---|---|---|
| GOBP_RESPONSE_TO_TOXIC_SUBSTANCE | 11 | 0.477 | 1.42 | 0.10400 | 0.10400 | 0.38500 | 18 |
| GOMF_CATALYTIC_ACTIVITY_ACTING_ON_A_NUCLEIC_ACID | 29 | -0.293 | -1.399 | 0.10400 | 0.10400 | 0.38500 | 298 |
| HP_DISPLACEMENT_OF_THE_URETHRAL_MEATUS | 20 | -0.338 | -1.414 | 0.10500 | 0.10500 | 0.38700 | 215 |
| GOBP_ENZYME_LINKED_RECEPTOR_PROTEIN_SIGNALING_PATHWAY | 27 | 0.361 | 1.399 | 0.10500 | 0.10500 | 0.38700 | 204 |
| GOCC_POSTSYNAPSE | 21 | -0.329 | -1.398 | 0.10700 | 0.10700 | 0.38900 | 125 |
| HP_PNEUMONIA | 13 | 0.454 | 1.425 | 0.10700 | 0.10700 | 0.38900 | 28 |
| HP_ABNORMALITY_OF_THE_METAPHYSIS | 15 | -0.369 | -1.392 | 0.10700 | 0.10700 | 0.38900 | 176 |
| GOMF_RNA_POLYMERASE_II_SPECIFIC_DNA_BINDING_TRANSCRIPTION_FACTOI | 22 | -0.322 | -1.393 | 0.10800 | 0.10800 | 0.39200 | 215 |
| HP_THIN_VERMILION_BORDER | 25 | -0.306 | -1.393 | 0.10900 | 0.10900 | 0.39300 | 165 |
| GOBP_NEGATIVE_REGULATION_OF_MRNA_METABOLIC_PROCESS | 10 | -0.44 | -1.418 | 0.11000 | 0.11000 | 0.39300 | 314 |
| GOBP_CELL_CELL_SIGNALING | 45 | 0.31 | 1.364 | 0.11000 | 0.11000 | 0.39300 | 74 |
| GOBP_POSITIVE_REGULATION_OF_CELL_CYCLE | 16 | 0.419 | 1.4 | 0.11100 | 0.11100 | 0.39300 | 73 |
| GOBP_LIPID_CATABOLIC_PROCESS | 15 | 0.427 | 1.398 | 0.11100 | 0.11100 | 0.39300 | 219 |
| GOBP_REGULATION_OF_RESPONSE_TO_STRESS | 45 | 0.309 | 1.363 | 0.11100 | 0.11100 | 0.39300 | 77 |
| GOBP_RESPONSE_TO_WOUNDING | 19 | 0.4 | 1.405 | 0.11100 | 0.11100 | 0.39300 | 167 |
| GOBP_EMBRYO_DEVELOPMENT | 46 | 0.307 | 1.364 | 0.11200 | 0.11200 | 0.39300 | 77 |
| GOMF_GLYCOSYLTRANSFERASE_ACTIVITY | 12 | 0.46 | 1.408 | 0.11200 | 0.11200 | 0.39300 | 232 |
| GOBP_POSITIVE_REGULATION_OF_INTRACELLULAR_SIGNAL_TRANSDUCTION | 35 | 0.33 | 1.372 | 0.11200 | 0.11200 | 0.39300 | 103 |
| GOCC_SYNAPSE | 41 | -0.251 | -1.342 | 0.11300 | 0.11300 | 0.39300 | 125 |
| HP_ABNORMAL_REFLEX | 34 | -0.271 | -1.361 | 0.11300 | 0.11300 | 0.39300 | 214 |
| GOBP_GROWTH | 27 | 0.356 | 1.383 | 0.11300 | 0.11300 | 0.39300 | 77 |
| GOBP_RIBONUCLEOPROTEIN_COMPLEX_SUBUNIT_ORGANIZATION | 11 | -0.419 | -1.394 | 0.11300 | 0.11300 | 0.39300 | 420 |
| GOBP_REGULATION_OF_PROTEIN_CATABOLIC_PROCESS | 13 | -0.39 | -1.386 | 0.11400 | 0.11400 | 0.39300 | 373 |
| GOBP_MUSCLE_CELL_DIFFERENTIATION | 16 | 0.417 | 1.393 | 0.11400 | 0.11400 | 0.39300 | 167 |
| GOBP_DNA_REPLICATION | 14 | -0.377 | -1.389 | 0.11500 | 0.11500 | 0.39300 | 261 |
| GOBP_MACROAUTOPHAGY | 18 | -0.343 | -1.385 | 0.11500 | 0.11500 | 0.39400 | 310 |
| GOMF_LIPID_BINDING | 39 | -0.255 | -1.339 | 0.11600 | 0.11600 | 0.39500 | 161 |
| GOBP_REGULATION_OF_MULTICELLULAR_ORGANISMAL_DEVELOPMENT | 43 | 0.31 | 1.354 | 0.11600 | 0.11600 | 0.39600 | 100 |
| GOBP_POSITIVE_REGULATION_OF_SIGNALING | 69 | 0.276 | 1.33 | 0.11800 | 0.11800 | 0.40000 | 77 |
| GOMF_PHOSPHOLIPID_BINDING | 25 | -0.304 | -1.383 | 0.11800 | 0.11800 | 0.40000 | 161 |
| GOBP_MITOTIC_CELL_CYCLE | 42 | 0.311 | 1.349 | 0.12000 | 0.12000 | 0.40400 | 103 |
| GOBP_POSITIVE_REGULATION_OF_GTPASE_ACTIVITY | 11 | -0.413 | -1.376 | 0.12100 | 0.12100 | 0.40600 | 6 |
| GOBP_MALE_GAMETE_GENERATION | 17 | 0.407 | 1.382 | 0.12100 | 0.12100 | 0.40600 | 149 |
| GOBP_RESPONSE_TO_METAL_ION | 12 | 0.454 | 1.387 | 0.12200 | 0.12200 | 0.40700 | 160 |
| GOBP_RESPONSE_TO_BACTERIUM | 14 | 0.432 | 1.387 | 0.12200 | 0.12200 | 0.40700 | 27 |
| GOBP_NEGATIVE_REGULATION_OF_SIGNALING | 43 | 0.307 | 1.339 | 0.12400 | 0.12400 | 0.41100 | 204 |
| GOBP_CELL_MORPHOGENESIS_INVOLVED_IN_NEURON_DIFFERENTIATION | 16 | 0.41 | 1.37 | 0.12400 | 0.12400 | 0.41100 | 56 |
| HP_INCREASED_HEAD_CIRCUMFERENCE | 24 | 0.365 | 1.369 | 0.12500 | 0.12500 | 0.41100 | 77 |
| GOBP_CELL_FATE_COMMITMENT | 11 | 0.464 | 1.383 | 0.12500 | 0.12500 | 0.41100 | 77 |
| HP_ABNORMALITY_OF_THE_PALPEBRAL_FISSURES | 37 | -0.256 | -1.321 | 0.12500 | 0.12500 | 0.41100 | 156 |
| GOCC_OUTER_MEMBRANE | 11 | 0.464 | 1.381 | 0.12600 | 0.12600 | 0.41400 | 149 |
| GOBP_POSITIVE_REGULATION_OF_KINASE_ACTIVITY | 21 | 0.378 | 1.365 | 0.12800 | 0.12800 | 0.41600 | 77 |
| GOBP_MULTICELLULAR_ORGANISM_REPRODUCTION | 29 | 0.341 | 1.352 | 0.12800 | 0.12800 | 0.41600 | 25 |

| | | | | | | | |
|---|---|---|---|---|---|---|---|
| HP_HIP_DYSPLASIA | 16 | -0.35 | -1.357 | 0.12900 | 0.12900 | 0.41800 | 207 |
| GOBP_EXOCYTOSIS | 15 | -0.358 | -1.35 | 0.13000 | 0.13000 | 0.42200 | 121 |
| GOBP_CALCIUM_ION_TRANSMEMBRANE_TRANSPORT | 12 | -0.388 | -1.345 | 0.13200 | 0.13200 | 0.42300 | 40 |
| GOMF_TRANSCRIPTION_COREPRESSOR_ACTIVITY | 13 | -0.379 | -1.346 | 0.13200 | 0.13200 | 0.42300 | 449 |
| GOBP_NEGATIVE_REGULATION_OF_INTRACELLULAR_SIGNAL_TRANSDUCTION | 16 | 0.406 | 1.355 | 0.13200 | 0.13200 | 0.42300 | 202 |
| GOBP_CELL_CELL_SIGNALING_BY_WNT | 16 | 0.405 | 1.351 | 0.13300 | 0.13300 | 0.42300 | 74 |
| GOBP_EPITHELIAL_CELL_PROLIFERATION | 15 | 0.414 | 1.354 | 0.13300 | 0.13300 | 0.42300 | 167 |
| GOBP_REGULATION_OF_PHOSPHORUS_METABOLIC_PROCESS | 57 | 0.284 | 1.322 | 0.13300 | 0.13300 | 0.42300 | 140 |
| GOBP_INTRINSIC_APOPTOTIC_SIGNALING_PATHWAY | 14 | -0.367 | -1.351 | 0.13400 | 0.13400 | 0.42300 | 156 |
| GOBP_CELLULAR_COMPONENT_MORPHOGENESIS | 17 | 0.399 | 1.356 | 0.13500 | 0.13500 | 0.42300 | 56 |
| GOBP_CELL_PART_MORPHOGENESIS | 17 | 0.399 | 1.356 | 0.13500 | 0.13500 | 0.42300 | 56 |
| GOBP_TRANSFORMING_GROWTH_FACTOR_BETA_RECEPTOR_SIGNALING_PATHW | 18 | 0.393 | 1.36 | 0.13500 | 0.13500 | 0.42300 | 204 |
| GOBP_PROTEIN_LOCALIZATION_TO_PLASMA_MEMBRANE | 11 | 0.46 | 1.371 | 0.13500 | 0.13500 | 0.42300 | 112 |
| GOBP_IMMUNE_RESPONSE | 51 | 0.291 | 1.324 | 0.13500 | 0.13500 | 0.42300 | 81 |
| HP_AGGRESSIVE_BEHAVIOR | 12 | -0.387 | -1.341 | 0.13600 | 0.13600 | 0.42300 | 313 |
| GOBP_METAL_ION_HOMEOSTASIS | 24 | 0.36 | 1.35 | 0.13600 | 0.13600 | 0.42400 | 103 |
| HP_RESPIRATORY_TRACT_INFECTION | 31 | 0.332 | 1.334 | 0.13700 | 0.13700 | 0.42400 | 28 |
| GOBP_CATION_TRANSPORT | 39 | 0.311 | 1.326 | 0.13700 | 0.13700 | 0.42400 | 115 |
| GOCC_SPLICEOSOMAL_COMPLEX | 12 | -0.387 | -1.339 | 0.13800 | 0.13800 | 0.42600 | 443 |
| HP_FAILURE_TO_THRIVE | 40 | -0.247 | -1.304 | 0.13900 | 0.13900 | 0.42600 | 215 |
| GOBP_REGULATION_OF_MITOTIC_CELL_CYCLE | 27 | 0.345 | 1.339 | 0.13900 | 0.13900 | 0.42600 | 73 |
| GOBP_SODIUM_ION_TRANSPORT | 10 | 0.471 | 1.359 | 0.13900 | 0.13900 | 0.42600 | 115 |
| GOBP_REGULATION_OF_GTPASE_ACTIVITY | 13 | -0.375 | -1.332 | 0.14100 | 0.14100 | 0.42600 | 6 |
| GOBP_MITOTIC_CELL_CYCLE_PROCESS | 38 | 0.312 | 1.324 | 0.14100 | 0.14100 | 0.42600 | 103 |
| GOBP_PROGRAMMED_CELL_DEATH | 66 | -0.208 | -1.255 | 0.14100 | 0.14100 | 0.42600 | 74 |
| HP_ABNORMALITY_OF_THE_PHILTRUM | 28 | -0.28 | -1.326 | 0.14100 | 0.14100 | 0.42600 | 165 |
| GOBP_CELL_MIGRATION | 37 | 0.312 | 1.317 | 0.14200 | 0.14200 | 0.42800 | 81 |
| HP_GENERALIZED_ONSET_SEIZURE | 13 | -0.374 | -1.329 | 0.14300 | 0.14300 | 0.43000 | 183 |
| HP_ABNORMAL_ORAL_PHYSIOLOGY | 14 | 0.422 | 1.355 | 0.14400 | 0.14400 | 0.43000 | 27 |
| HP_ABNORMAL_PALMAR_DERMATOGLYPHICS | 17 | -0.336 | -1.331 | 0.14400 | 0.14400 | 0.43000 | 381 |
| HP_SMALL_FACE | 11 | 0.455 | 1.356 | 0.14400 | 0.14400 | 0.43000 | 20 |
| HP_POSTAXIAL_POLYDACTYLY | 10 | 0.468 | 1.349 | 0.14500 | 0.14500 | 0.43000 | 355 |
| HP_ABNORMAL_RESPIRATORY_SYSTEM_MORPHOLOGY | 68 | 0.27 | 1.295 | 0.14500 | 0.14500 | 0.43000 | 28 |
| GOBP_RESPONSE_TO_STEROID_HORMONE | 16 | 0.4 | 1.336 | 0.14600 | 0.14600 | 0.43200 | 18 |
| GOBP_RESPONSE_TO_OXYGEN_LEVELS | 12 | 0.441 | 1.349 | 0.14700 | 0.14700 | 0.43200 | 132 |
| GOBP_REGULATION_OF_ION_TRANSPORT | 26 | 0.346 | 1.328 | 0.14700 | 0.14700 | 0.43200 | 35 |
| GOBP_GLYCEROLIPID_METABOLIC_PROCESS | 10 | -0.417 | -1.344 | 0.14800 | 0.14800 | 0.43500 | 18 |
| GOBP_POSITIVE_REGULATION_OF_CELL_DIFFERENTIATION | 29 | 0.333 | 1.318 | 0.14900 | 0.14900 | 0.43500 | 85 |
| GOBP_POSITIVE_REGULATION_OF_TRANSPORT | 34 | 0.318 | 1.312 | 0.15000 | 0.15000 | 0.43700 | 112 |
| GOBP_POSITIVE_REGULATION_OF_CATALYTIC_ACTIVITY | 43 | 0.299 | 1.303 | 0.15000 | 0.15000 | 0.43700 | 132 |
| HP_LIMB_JOINT_CONTRACTURE | 12 | -0.382 | -1.322 | 0.15100 | 0.15100 | 0.43700 | 183 |
| HP_ABNORMAL_PHARYNX_MORPHOLOGY | 14 | 0.418 | 1.343 | 0.15200 | 0.15200 | 0.43900 | 28 |
| GOBP_HEART_MORPHOGENESIS | 10 | 0.463 | 1.336 | 0.15300 | 0.15300 | 0.43900 | 77 |

| | | | | | | | |
|---|---|---|---|---|---|---|---|
| GOBP_POSITIVE_REGULATION_OF_LOCOMOTION | 15 | 0.405 | 1.326 | 0.15300 | 0.15300 | 0.43900 | 74 |
| GOBP_PHOSPHOLIPID_METABOLIC_PROCESS | 10 | -0.414 | -1.333 | 0.15300 | 0.15300 | 0.43900 | 18 |
| HP_DELAYED_GROSS_MOTOR_DEVELOPMENT | 11 | -0.396 | -1.318 | 0.15400 | 0.15400 | 0.44100 | 300 |
| GOBP_REGIONALIZATION | 12 | 0.437 | 1.338 | 0.15400 | 0.15400 | 0.44100 | 149 |
| GOBP_ANATOMICAL_STRUCTURE_HOMEOSTASIS | 12 | 0.436 | 1.335 | 0.15500 | 0.15500 | 0.44100 | 18 |
| HP_ABNORMAL_NERVOUS_SYSTEM_ELECTROPHYSIOLOGY | 27 | -0.279 | -1.298 | 0.15500 | 0.15500 | 0.44100 | 214 |
| GOBP_NUCLEOBASE_CONTAINING_COMPOUND_TRANSPORT | 13 | -0.368 | -1.309 | 0.15800 | 0.15800 | 0.44700 | 264 |
| GOBP_RESPONSE_TO_LIGHT_STIMULUS | 10 | -0.411 | -1.322 | 0.15900 | 0.15900 | 0.44900 | 207 |
| GOBP_ION_TRANSPORT | 46 | 0.291 | 1.294 | 0.16000 | 0.16000 | 0.44900 | 47 |
| HP_ABNORMAL_TENDON_MORPHOLOGY | 24 | -0.291 | -1.302 | 0.16000 | 0.16000 | 0.44900 | 286 |
| GOBP_DOUBLE_STRAND_BREAK_REPAIR | 11 | -0.392 | -1.304 | 0.16000 | 0.16000 | 0.44900 | 251 |
| HP_IRREGULAR_HYPERPIGMENTATION | 14 | -0.354 | -1.305 | 0.16000 | 0.16000 | 0.44900 | 300 |
| HP_ABNORMALITY_OF_THE_PULMONARY_VASCULATURE | 12 | 0.433 | 1.326 | 0.16100 | 0.16100 | 0.44900 | 14 |
| GOBP_POSITIVE_REGULATION_OF_TRANSFERASE_ACTIVITY | 23 | 0.355 | 1.314 | 0.16100 | 0.16100 | 0.45000 | 77 |
| HP_ABNORMAL_JOINT_MORPHOLOGY | 46 | -0.229 | -1.261 | 0.16500 | 0.16500 | 0.45500 | 215 |
| GOBP_NEGATIVE_REGULATION_OF_PHOSPHORYLATION | 14 | 0.412 | 1.322 | 0.16500 | 0.16500 | 0.45500 | 140 |
| GOBP_PEPTIDYL_TYROSINE_MODIFICATION | 10 | -0.408 | -1.313 | 0.16500 | 0.16500 | 0.45500 | 179 |
| GOBP_PROTEIN_ACETYLATION | 17 | -0.328 | -1.299 | 0.16600 | 0.16600 | 0.45500 | 283 |
| HP_DYSTONIA | 16 | -0.335 | -1.296 | 0.16700 | 0.16700 | 0.45500 | 239 |
| HP_FLEXION_CONTRACTURE_OF_DIGIT | 10 | -0.407 | -1.312 | 0.16700 | 0.16700 | 0.45500 | 285 |
| HP_ABNORMALITY_OF_FEMALE_EXTERNAL_GENITALIA | 10 | -0.407 | -1.311 | 0.16700 | 0.16700 | 0.45500 | 176 |
| GOMF_RIBONUCLEOTIDE_BINDING | 67 | -0.202 | -1.227 | 0.16700 | 0.16700 | 0.45500 | 264 |
| GOBP_NEGATIVE_REGULATION_OF_CELL_DEATH | 33 | 0.315 | 1.291 | 0.16700 | 0.16700 | 0.45500 | 25 |
| HP_ABNORMALITY_OF_THE_ABDOMINAL_ORGANS | 45 | 0.29 | 1.276 | 0.16800 | 0.16800 | 0.45500 | 77 |
| GOBP_FOREBRAIN_DEVELOPMENT | 10 | -0.406 | -1.307 | 0.16900 | 0.16900 | 0.45500 | 141 |
| GOBP_REGULATION_OF_CELLULAR_RESPONSE_TO_GROWTH_FACTOR_STIMULUS | 19 | 0.371 | 1.306 | 0.16900 | 0.16900 | 0.45500 | 204 |
| GOBP_INNATE_IMMUNE_RESPONSE | 22 | -0.298 | -1.289 | 0.16900 | 0.16900 | 0.45500 | 156 |
| HP_HYPERTRICHOSIS | 11 | -0.387 | -1.29 | 0.17000 | 0.17000 | 0.45500 | 183 |
| GOBP_RESPONSE_TO_ABIOTIC_STIMULUS | 29 | -0.269 | -1.283 | 0.17000 | 0.17000 | 0.45500 | 168 |
| HP_NEOPLASM | 29 | 0.325 | 1.286 | 0.17000 | 0.17000 | 0.45500 | 94 |
| GOBP_REGULATION_OF_TRANSMEMBRANE_RECEPTOR_PROTEIN_SERINE_THREO | 19 | 0.371 | 1.304 | 0.17100 | 0.17100 | 0.45500 | 204 |
| GOBP_RESPONSE_TO_MOLECULE_OF_BACTERIAL_ORIGIN | 11 | 0.44 | 1.311 | 0.17100 | 0.17100 | 0.45500 | 18 |
| GOCC_NUCLEAR_MEMBRANE | 11 | 0.44 | 1.31 | 0.17100 | 0.17100 | 0.45500 | 18 |
| GOBP_CELLULAR_ION_HOMEOSTASIS | 26 | 0.337 | 1.293 | 0.17200 | 0.17200 | 0.45600 | 103 |
| GOBP_TRANSMEMBRANE_RECEPTOR_PROTEIN_SERINE_THREONINE_KINASE_SIG | 22 | 0.355 | 1.296 | 0.17300 | 0.17300 | 0.45700 | 204 |
| GOBP_GOLGI_VESICLE_TRANSPORT | 19 | -0.312 | -1.28 | 0.17300 | 0.17300 | 0.45700 | 286 |
| GOBP_REGULATION_OF_SECRETION | 18 | 0.376 | 1.301 | 0.17400 | 0.17400 | 0.45700 | 35 |
| GOBP_SENSORY_PERCEPTION | 16 | 0.387 | 1.293 | 0.17400 | 0.17400 | 0.45700 | 30 |
| HP_DEEPLY_SET_EYE | 15 | -0.34 | -1.283 | 0.17500 | 0.17500 | 0.45800 | 156 |
| GOBP_MULTICELLULAR_ORGANISMAL_HOMEOSTASIS | 19 | 0.369 | 1.296 | 0.17600 | 0.17600 | 0.46100 | 27 |
| GOBP_TAXIS | 16 | -0.331 | -1.282 | 0.17700 | 0.17700 | 0.46100 | 11 |
| HP_ABNORMAL_PULMONARY_INTERSTITIAL_MORPHOLOGY | 11 | 0.436 | 1.3 | 0.17700 | 0.17700 | 0.46100 | 16 |
| GOBP_RESPONSE_TO_EXTRACELLULAR_STIMULUS | 24 | -0.285 | -1.274 | 0.17900 | 0.17900 | 0.46100 | 79 |

| | | | | | | | |
|---|---|---|---|---|---|---|---|
| HP_ABNORMAL_NUMBER_OF_TEETH | 21 | 0.355 | 1.284 | 0.18000 | 0.18000 | 0.46100 | 59 |
| GOBP_CELLULAR_RESPONSE_TO_ABIOTIC_STIMULUS | 11 | 0.435 | 1.295 | 0.18100 | 0.18100 | 0.46100 | 46 |
| GOMF_PROTEIN_KINASE_ACTIVITY | 16 | -0.329 | -1.273 | 0.18100 | 0.18100 | 0.46100 | 260 |
| GOMF_PROTEIN_SERINE_THREONINE_KINASE_ACTIVITY | 16 | -0.329 | -1.273 | 0.18100 | 0.18100 | 0.46100 | 260 |
| GOBP_RESPONSE_TO_OXYGEN_CONTAINING_COMPOUND | 48 | -0.222 | -1.233 | 0.18200 | 0.18200 | 0.46100 | 70 |
| HP_STEREOTYPY | 16 | -0.328 | -1.27 | 0.18300 | 0.18300 | 0.46100 | 207 |
| GOBP_CALCIUM_ION_TRANSPORT | 16 | -0.328 | -1.27 | 0.18300 | 0.18300 | 0.46100 | 90 |
| GOBP_REGULATION_OF_BODY_FLUID_LEVELS | 12 | 0.424 | 1.296 | 0.18300 | 0.18300 | 0.46100 | 27 |
| GOBP_ALCOHOL_METABOLIC_PROCESS | 10 | 0.448 | 1.293 | 0.18300 | 0.18300 | 0.46100 | 236 |
| HP_LOW_SET_POSTERIORLY_ROTATED_EARS | 13 | 0.413 | 1.298 | 0.18300 | 0.18300 | 0.46100 | 14 |
| GOMF_ATP_DEPENDENT_ACTIVITY | 23 | -0.29 | -1.276 | 0.18300 | 0.18300 | 0.46100 | 192 |
| GOBP_REGULATION_OF_DNA_BINDING_TRANSCRIPTION_FACTOR_ACTIVITY | 18 | -0.315 | -1.27 | 0.18300 | 0.18300 | 0.46100 | 78 |
| HP_INFLAMMATORY_ABNORMALITY_OF_THE_EYE | 10 | -0.4 | -1.286 | 0.18400 | 0.18400 | 0.46100 | 166 |
| GOMF_DNA_BINDING_TRANSCRIPTION_FACTOR_BINDING | 28 | -0.267 | -1.264 | 0.18400 | 0.18400 | 0.46100 | 261 |
| GOCC_COATED_VESICLE | 12 | 0.423 | 1.293 | 0.18400 | 0.18400 | 0.46100 | 20 |
| GOCC_RNA_POLYMERASE_II_TRANSCRIPTION_REGULATOR_COMPLEX | 12 | -0.365 | -1.265 | 0.18400 | 0.18400 | 0.46100 | 89 |
| HP_ABNORMALITY_OF_THE_CURVATURE_OF_THE_CORNEA | 13 | -0.356 | -1.266 | 0.18400 | 0.18400 | 0.46100 | 300 |
| HP_ABNORMALITY_OF_THE_GINGIVA | 10 | -0.399 | -1.285 | 0.18600 | 0.18600 | 0.46200 | 166 |
| HP_ABNORMAL_ORAL_MUCOSA_MORPHOLOGY | 10 | -0.399 | -1.285 | 0.18600 | 0.18600 | 0.46200 | 166 |
| GOCC_GLUTAMATERGIC_SYNAPSE | 10 | -0.399 | -1.284 | 0.18700 | 0.18700 | 0.46200 | 50 |
| GOBP_CELLULAR_RESPONSE_TO_ORGANIC_CYCLIC_COMPOUND | 27 | 0.328 | 1.272 | 0.18700 | 0.18700 | 0.46200 | 111 |
| HP_ORAL_CLEFT | 32 | 0.312 | 1.267 | 0.18700 | 0.18700 | 0.46200 | 71 |
| GOMF_KINASE_ACTIVITY | 18 | -0.312 | -1.258 | 0.18900 | 0.18900 | 0.46600 | 260 |
| HP_ABNORMALITY_OF_FEMUR_MORPHOLOGY | 18 | -0.312 | -1.257 | 0.18900 | 0.18900 | 0.46600 | 176 |
| GOBP_CELLULAR_RESPONSE_TO_STEROID_HORMONE_STIMULUS | 12 | 0.42 | 1.283 | 0.19000 | 0.19000 | 0.46800 | 10 |
| GOBP_IMMUNE_RESPONSE_REGULATING_SIGNALING_PATHWAY | 13 | 0.409 | 1.285 | 0.19200 | 0.19200 | 0.47000 | 48 |
| HP_ABNORMALITY_OF_THE_INCISOR | 10 | 0.444 | 1.28 | 0.19300 | 0.19300 | 0.47100 | 20 |
| GOBP_POSITIVE_REGULATION_OF_CELLULAR_COMPONENT_ORGANIZATION | 47 | -0.221 | -1.222 | 0.19400 | 0.19400 | 0.47300 | 141 |
| HP_VASCULAR_SKIN_ABNORMALITY | 19 | -0.306 | -1.253 | 0.19500 | 0.19500 | 0.47300 | 166 |
| GOBP_NCRNA_METABOLIC_PROCESS | 31 | -0.253 | -1.242 | 0.19500 | 0.19500 | 0.47300 | 481 |
| GOCC_TRANSCRIPTION_REGULATOR_COMPLEX | 24 | -0.28 | -1.252 | 0.19600 | 0.19600 | 0.47300 | 449 |
| GOBP_REGULATION_OF_SMALL_MOLECULE_METABOLIC_PROCESS | 11 | -0.377 | -1.255 | 0.19600 | 0.19600 | 0.47300 | 14 |
| GOBP_REGULATION_OF_CELLULAR_MACROMOLECULE_BIOSYNTHETIC_PROCESS | 37 | -0.239 | -1.233 | 0.19700 | 0.19700 | 0.47300 | 247 |
| GOBP_MAINTENANCE_OF_LOCATION | 12 | -0.361 | -1.25 | 0.19700 | 0.19700 | 0.47300 | 43 |
| GOBP_NEURON_DIFFERENTIATION | 37 | 0.296 | 1.248 | 0.19800 | 0.19800 | 0.47300 | 71 |
| HP_WIDELY_SPACED_TEETH | 10 | 0.442 | 1.274 | 0.19800 | 0.19800 | 0.47300 | 20 |
| GOBP_COGNITION | 12 | -0.36 | -1.247 | 0.19800 | 0.19800 | 0.47300 | 36 |
| GOBP_NEURON_DEVELOPMENT | 30 | 0.314 | 1.252 | 0.19900 | 0.19900 | 0.47300 | 71 |
| GOBP_ERK1_AND_ERK2_CASCADE | 11 | -0.375 | -1.249 | 0.19900 | 0.19900 | 0.47300 | 66 |
| GOCC_PERINUCLEAR_REGION_OF_CYTOPLASM | 26 | 0.327 | 1.255 | 0.19900 | 0.19900 | 0.47300 | 103 |
| GOCC_RECEPTOR_COMPLEX | 12 | 0.415 | 1.269 | 0.20000 | 0.20000 | 0.47400 | 148 |
| HP_ABNORMALITY_OF_MOUTH_SHAPE | 10 | -0.391 | -1.259 | 0.20200 | 0.20200 | 0.47700 | 165 |
| HP_DUPLICATION_OF_HAND_BONES | 11 | 0.424 | 1.263 | 0.20300 | 0.20300 | 0.48000 | 243 |

| | | | | | | | |
|---|---|---|---|---|---|---|---|
| GOBP_REGULATION_OF_MEMBRANE_POTENTIAL | 15 | 0.386 | 1.265 | 0.20400 | 0.20400 | 0.48100 | 115 |
| HP_ABNORMALITY_OF_BONE_MARROW_CELL_MORPHOLOGY | 10 | -0.389 | -1.252 | 0.20600 | 0.20600 | 0.48500 | 166 |
| GOCC_MITOCHONDRIAL_ENVELOPE | 20 | 0.352 | 1.253 | 0.20700 | 0.20700 | 0.48500 | 217 |
| GOBP_OSSIFICATION | 16 | 0.375 | 1.251 | 0.21000 | 0.21000 | 0.49200 | 77 |
| GOBP_REGULATION_OF_CELL_ADHESION | 15 | 0.384 | 1.256 | 0.21000 | 0.21000 | 0.49200 | 219 |
| GOMF_LIPID_TRANSPORTER_ACTIVITY | 11 | -0.37 | -1.233 | 0.21200 | 0.21200 | 0.49400 | 48 |
| GOBP_RIBOSOME_BIOGENESIS | 15 | -0.327 | -1.234 | 0.21300 | 0.21300 | 0.49500 | 298 |
| HP_ABNORMAL_LUNG_DEVELOPMENT | 15 | 0.382 | 1.251 | 0.21300 | 0.21300 | 0.49500 | 59 |
| HP_PROMINENT_FOREHEAD | 13 | 0.399 | 1.255 | 0.21400 | 0.21400 | 0.49600 | 71 |
| GOBP_LOCOMOTION | 50 | 0.27 | 1.223 | 0.21500 | 0.21500 | 0.49800 | 81 |
| HP_ABNORMALITY_OF_THE_PHARYNX | 17 | 0.366 | 1.242 | 0.21600 | 0.21600 | 0.49900 | 28 |
| GOBP_PROTEIN_COMPLEX_OLIGOMERIZATION | 10 | -0.385 | -1.241 | 0.21700 | 0.21700 | 0.49900 | 69 |
| HP_ABNORMALITY_OF_SKIN_PIGMENTATION | 22 | -0.284 | -1.231 | 0.21700 | 0.21700 | 0.49900 | 300 |
| GOBP_RESPONSE_TO_PEPTIDE_HORMONE | 13 | 0.398 | 1.249 | 0.21800 | 0.21800 | 0.49900 | 103 |
| GOBP_CELL_SURFACE_RECEPTOR_SIGNALING_PATHWAY_INVOLVED_IN_CELL_C | 17 | 0.365 | 1.239 | 0.21900 | 0.21900 | 0.50100 | 74 |
| HP_TOOTH_MALPOSITION | 23 | 0.335 | 1.239 | 0.21900 | 0.21900 | 0.50100 | 54 |
| GOMF_PEPTIDE_BINDING | 10 | 0.431 | 1.242 | 0.22100 | 0.22100 | 0.50400 | 16 |
| GOMF_CYTOSKELETAL_PROTEIN_BINDING | 27 | 0.316 | 1.227 | 0.22200 | 0.22200 | 0.50500 | 235 |
| GOMF_CELL_ADHESION_MOLECULE_BINDING | 30 | 0.306 | 1.222 | 0.22300 | 0.22300 | 0.50600 | 81 |
| GOBP_NEUROGENESIS | 51 | 0.267 | 1.214 | 0.22400 | 0.22400 | 0.50800 | 77 |
| HP_EARLY_ONSET_OF_SEXUAL_MATURATION | 11 | -0.365 | -1.216 | 0.22500 | 0.22500 | 0.50900 | 300 |
| GOBP_REGULATION_OF_TRANSFERASE_ACTIVITY | 39 | 0.285 | 1.217 | 0.22600 | 0.22600 | 0.51000 | 140 |
| GOBP_REGULATION_OF_PROTEOLYSIS | 19 | 0.35 | 1.23 | 0.22800 | 0.22800 | 0.51300 | 25 |
| GOMF_PEPTIDASE_ACTIVITY | 16 | -0.311 | -1.205 | 0.22900 | 0.22900 | 0.51300 | 497 |
| GOBP_POSITIVE_REGULATION_OF_CYTOKINE_PRODUCTION | 12 | -0.35 | -1.213 | 0.23000 | 0.23000 | 0.51300 | 31 |
| GOBP_NEGATIVE_REGULATION_OF_CATALYTIC_ACTIVITY | 28 | 0.311 | 1.217 | 0.23000 | 0.23000 | 0.51300 | 163 |
| HP_FUNCTIONAL_MOTOR_DEFICIT | 15 | -0.32 | -1.208 | 0.23000 | 0.23000 | 0.51300 | 183 |
| HP_ABNORMALITY_OF_THE_UPPER_RESPIRATORY_TRACT | 30 | 0.304 | 1.213 | 0.23000 | 0.23000 | 0.51300 | 59 |
| GOCC_CHROMATIN | 58 | -0.2 | -1.174 | 0.23100 | 0.23100 | 0.51400 | 216 |
| HP_LONG_PHILTRUM | 21 | -0.285 | -1.214 | 0.23200 | 0.23200 | 0.51400 | 165 |
| GOBP_REGULATION_OF_IMMUNE_SYSTEM_PROCESS | 41 | 0.281 | 1.213 | 0.23200 | 0.23200 | 0.51500 | 85 |
| HP_SHORT_FOOT | 10 | -0.377 | -1.214 | 0.23500 | 0.23500 | 0.51800 | 449 |
| HP_CLINODACTYLY | 32 | -0.241 | -1.192 | 0.23500 | 0.23500 | 0.51800 | 183 |
| HP_GENERALIZED_ABNORMALITY_OF_SKIN | 35 | -0.237 | -1.199 | 0.23500 | 0.23500 | 0.51900 | 183 |
| GOBP_CELL_ACTIVATION | 29 | 0.306 | 1.212 | 0.23600 | 0.23600 | 0.51900 | 81 |
| GOBP_ENDOSOMAL_TRANSPORT | 10 | -0.376 | -1.212 | 0.23600 | 0.23600 | 0.51900 | 296 |
| GOBP_INFLAMMATORY_RESPONSE | 19 | 0.346 | 1.218 | 0.23900 | 0.23900 | 0.52300 | 27 |
| GOBP_CELL_CYCLE_PROCESS | 54 | 0.26 | 1.196 | 0.23900 | 0.23900 | 0.52300 | 103 |
| HP_ABNORMALITY_OF_TEMPERATURE_REGULATION | 19 | -0.291 | -1.194 | 0.24000 | 0.24000 | 0.52300 | 153 |
| GOMF_ACTIVE_TRANSMEMBRANE_TRANSPORTER_ACTIVITY | 12 | -0.347 | -1.2 | 0.24000 | 0.24000 | 0.52300 | 125 |
| HP_ABNORMAL_HEART_VALVE_PHYSIOLOGY | 16 | 0.364 | 1.216 | 0.24100 | 0.24100 | 0.52300 | 54 |
| GOBP_POSITIVE_REGULATION_OF_TRANSCRIPTION_BY_RNA_POLYMERASE_II | 61 | -0.196 | -1.16 | 0.24100 | 0.24100 | 0.52300 | 79 |
| GOBP_PROTEIN_ACYLATION | 18 | -0.294 | -1.187 | 0.24100 | 0.24100 | 0.52300 | 283 |

| | | | | | | | |
|---|---|---|---|---|---|---|---|
| GOBP_METAL_ION_TRANSPORT | 32 | 0.294 | 1.196 | 0.24300 | 0.24300 | 0.52500 | 115 |
| HP_ABNORMALITY_OF_THE_PALM | 25 | -0.262 | -1.195 | 0.24400 | 0.24400 | 0.52600 | 291 |
| GOMF_MODIFICATION_DEPENDENT_PROTEIN_BINDING | 13 | -0.336 | -1.194 | 0.24500 | 0.24500 | 0.52700 | 204 |
| GOBP_POSITIVE_REGULATION_OF_MULTICELLULAR_ORGANISMAL_PROCESS | 42 | 0.275 | 1.195 | 0.24600 | 0.24600 | 0.52800 | 100 |
| HP_ABNORMAL_URINE_PROTEIN_LEVEL | 10 | 0.419 | 1.209 | 0.24700 | 0.24700 | 0.52800 | 203 |
| HP_ABNORMALITY_OF_CARDIOVASCULAR_SYSTEM_ELECTROPHYSIOLOGY | 13 | 0.386 | 1.214 | 0.24700 | 0.24700 | 0.52800 | 134 |
| HP_ABNORMALITY_OF_THE_URETHRA | 22 | -0.276 | -1.195 | 0.24700 | 0.24700 | 0.52800 | 215 |
| GOCC_VACUOLE | 36 | -0.23 | -1.18 | 0.24800 | 0.24800 | 0.52900 | 190 |
| GOBP_REGULATION_OF_CELL_DEATH | 55 | 0.257 | 1.187 | 0.24900 | 0.24900 | 0.53000 | 25 |
| GOBP_REGULATION_OF_TRANSMEMBRANE_TRANSPORT | 23 | -0.271 | -1.192 | 0.25000 | 0.25000 | 0.53000 | 119 |
| GOBP_NEGATIVE_REGULATION_OF_CELL_CYCLE_PHASE_TRANSITION | 11 | -0.356 | -1.186 | 0.25100 | 0.25100 | 0.53000 | 464 |
| GOBP_REGULATION_OF_CYTOSOLIC_CALCIUM_ION_CONCENTRATION | 12 | 0.396 | 1.21 | 0.25100 | 0.25100 | 0.53000 | 103 |
| GOBP_REGULATION_OF_CELLULAR_LOCALIZATION | 40 | -0.221 | -1.17 | 0.25100 | 0.25100 | 0.53000 | 285 |
| GOBP_PROTEIN_DNA_COMPLEX_SUBUNIT_ORGANIZATION | 13 | -0.333 | -1.184 | 0.25400 | 0.25400 | 0.53000 | 151 |
| GOBP_RESPONSE_TO_ORGANIC_CYCLIC_COMPOUND | 33 | 0.29 | 1.187 | 0.25500 | 0.25500 | 0.53000 | 45 |
| GOMF_CATION_CHANNEL_ACTIVITY | 11 | -0.355 | -1.181 | 0.25500 | 0.25500 | 0.53000 | 20 |
| GOMF_PASSIVE_TRANSMEMBRANE_TRANSPORTER_ACTIVITY | 11 | -0.355 | -1.181 | 0.25500 | 0.25500 | 0.53000 | 20 |
| GOBP_CELLULAR_HOMEOSTASIS | 30 | 0.297 | 1.186 | 0.25500 | 0.25500 | 0.53000 | 103 |
| GOCC_ENVELOPE | 39 | 0.278 | 1.186 | 0.25500 | 0.25500 | 0.53000 | 45 |
| HP_UNUSUAL_INFECTION | 37 | 0.28 | 1.182 | 0.25600 | 0.25600 | 0.53000 | 28 |
| GOMF_CALCIUM_ION_BINDING | 20 | -0.281 | -1.178 | 0.25600 | 0.25600 | 0.53000 | 88 |
| HP_NARROW_MOUTH | 19 | 0.34 | 1.196 | 0.25600 | 0.25600 | 0.53000 | 2 |
| GOBP_CELL_JUNCTION_ORGANIZATION | 16 | -0.303 | -1.174 | 0.25700 | 0.25700 | 0.53000 | 7 |
| HP_ABNORMAL_GROWTH_HORMONE_LEVEL | 10 | 0.415 | 1.197 | 0.25700 | 0.25700 | 0.53000 | 71 |
| HP_NEOPLASM_BY_HISTOLOGY | 12 | -0.341 | -1.18 | 0.25700 | 0.25700 | 0.53000 | 165 |
| HP_ABNORMALITY_OF_THE_NASAL_ALAE | 37 | -0.227 | -1.172 | 0.25700 | 0.25700 | 0.53000 | 185 |
| GOBP_POSITIVE_REGULATION_OF_INTRACELLULAR_PROTEIN_TRANSPORT | 12 | 0.393 | 1.2 | 0.25900 | 0.25900 | 0.53300 | 112 |
| GOBP_REGULATION_OF_PROTEIN_STABILITY | 20 | -0.28 | -1.171 | 0.26000 | 0.26000 | 0.53400 | 207 |
| HP_ABNORMALITY_OF_THE_LIVER | 38 | 0.277 | 1.177 | 0.26200 | 0.26200 | 0.53400 | 188 |
| HP_ABNORMALITY_OF_LOWER_LIMB_JOINT | 31 | -0.237 | -1.162 | 0.26200 | 0.26200 | 0.53400 | 231 |
| HP_DELAYED_PUBERTY | 10 | -0.365 | -1.177 | 0.26200 | 0.26200 | 0.53400 | 165 |
| HP_HIRSUTISM | 13 | -0.33 | -1.175 | 0.26200 | 0.26200 | 0.53400 | 207 |
| GOBP_DNA_RECOMBINATION | 10 | -0.364 | -1.171 | 0.26800 | 0.26800 | 0.54400 | 251 |
| GOBP_VESICLE_LOCALIZATION | 14 | -0.319 | -1.173 | 0.26800 | 0.26800 | 0.54400 | 256 |
| HP_ABNORMAL_CONNECTION_OF_THE_CARDIAC_SEGMENTS | 14 | 0.37 | 1.188 | 0.26900 | 0.26900 | 0.54500 | 14 |
| HP_LOW_ANTERIOR_HAIRLINE | 10 | -0.362 | -1.167 | 0.27000 | 0.27000 | 0.54500 | 213 |
| HP_ABNORMAL_ESOPHAGUS_PHYSIOLOGY | 33 | -0.231 | -1.148 | 0.27000 | 0.27000 | 0.54500 | 251 |
| HP_HEMATOLOGICAL_NEOPLASM | 13 | -0.327 | -1.162 | 0.27000 | 0.27000 | 0.54500 | 166 |
| GOBP_GASTRULATION | 10 | 0.41 | 1.182 | 0.27100 | 0.27100 | 0.54600 | 77 |
| GOMF_PROTEIN_SERINE_KINASE_ACTIVITY | 11 | -0.349 | -1.161 | 0.27200 | 0.27200 | 0.54600 | 257 |
| GOMF_PROTEIN_SERINE_THREONINE_TYROSINE_KINASE_ACTIVITY | 11 | -0.349 | -1.161 | 0.27200 | 0.27200 | 0.54600 | 257 |
| GOBP_POSITIVE_REGULATION_OF_DNA_BINDING_TRANSCRIPTION_FACTOR_ACT | 14 | -0.317 | -1.166 | 0.27400 | 0.27400 | 0.54700 | 78 |
| GOMF_NUCLEOSIDE_TRIPHOSPHATASE_REGULATOR_ACTIVITY | 23 | -0.263 | -1.157 | 0.27600 | 0.27600 | 0.55100 | 221 |

| | | | | | | | |
|---|---|---|---|---|---|---|---|
| HP_DEVIATION_OF_THE_HAND_OR_OF_FINGERS_OF_THE_HAND | 25 | -0.255 | -1.162 | 0.27700 | 0.27700 | 0.55200 | 183 |
| GOBP_NCRNA_PROCESSING | 21 | -0.273 | -1.161 | 0.27800 | 0.27800 | 0.55200 | 481 |
| GOBP_IMPORT_INTO_CELL | 10 | 0.408 | 1.175 | 0.27800 | 0.27800 | 0.55200 | 91 |
| HP_OTITIS_MEDIA | 11 | 0.395 | 1.177 | 0.27900 | 0.27900 | 0.55300 | 28 |
| GOBP_RESPONSE_TO_HORMONE | 31 | 0.289 | 1.163 | 0.27900 | 0.27900 | 0.55300 | 18 |
| HP_FOCAL_ONSET_SEIZURE | 10 | -0.358 | -1.152 | 0.28100 | 0.28100 | 0.55500 | 231 |
| GOBP_PEPTIDYL_SERINE_MODIFICATION | 13 | 0.374 | 1.175 | 0.28200 | 0.28200 | 0.55500 | 100 |
| HP_ABNORMALITY_OF_HINDBRAIN_MORPHOLOGY | 41 | -0.213 | -1.142 | 0.28200 | 0.28200 | 0.55500 | 191 |
| GOMF_AMIDE_BINDING | 12 | 0.384 | 1.173 | 0.28200 | 0.28200 | 0.55500 | 16 |
| GOCC_CELL_BODY | 12 | 0.383 | 1.172 | 0.28300 | 0.28300 | 0.55500 | 77 |
| HP_ABNORMALITY_OF_THE_PITUITARY_GLAND | 14 | 0.366 | 1.173 | 0.28400 | 0.28400 | 0.55500 | 77 |
| HP_ABNORMAL_LIP_MORPHOLOGY | 48 | -0.204 | -1.134 | 0.28400 | 0.28400 | 0.55600 | 165 |
| HP_ABNORMAL_CEREBRAL_VENTRICLE_MORPHOLOGY | 52 | -0.197 | -1.134 | 0.28500 | 0.28500 | 0.55600 | 165 |
| HP_ABNORMAL_ODONTOID_TISSUE_MORPHOLOGY | 15 | 0.358 | 1.172 | 0.28600 | 0.28600 | 0.55700 | 59 |
| GOMF_CADHERIN_BINDING | 24 | -0.256 | -1.146 | 0.28700 | 0.28700 | 0.55800 | 289 |
| HP_ABDOMINAL_PAIN | 16 | 0.348 | 1.162 | 0.28800 | 0.28800 | 0.55900 | 218 |
| HP_DEVIATION_OF_TOES | 10 | -0.354 | -1.14 | 0.28800 | 0.28800 | 0.55900 | 300 |
| GOBP_REGULATION_OF_ORGANELLE_ASSEMBLY | 10 | -0.354 | -1.139 | 0.28900 | 0.28900 | 0.55900 | 45 |
| HP_PATENT_DUCTUS_ARTERIOSUS | 19 | -0.279 | -1.144 | 0.29000 | 0.29000 | 0.55900 | 231 |
| HP_ABNORMAL_SHAPE_OF_THE_FRONTAL_REGION | 21 | 0.32 | 1.157 | 0.29100 | 0.29100 | 0.55900 | 77 |
| GOBP_REGULATION_OF_EXOCYTOSIS | 10 | -0.353 | -1.137 | 0.29100 | 0.29100 | 0.55900 | 121 |
| HP_VENTRICULOMEGALY | 41 | -0.212 | -1.135 | 0.29200 | 0.29200 | 0.55900 | 127 |
| HP_HAND_POLYDACTYLY | 10 | 0.402 | 1.161 | 0.29200 | 0.29200 | 0.55900 | 355 |
| HP_ABNORMAL_CONJUGATE_EYE_MOVEMENT | 40 | -0.213 | -1.127 | 0.29200 | 0.29200 | 0.55900 | 222 |
| GOBP_ORGANONITROGEN_COMPOUND_BIOSYNTHETIC_PROCESS | 69 | -0.184 | -1.12 | 0.29500 | 0.29500 | 0.56300 | 247 |
| GOBP_REGULATION_OF_DEFENSE_RESPONSE | 17 | 0.341 | 1.159 | 0.29500 | 0.29500 | 0.56300 | 48 |
| GOBP_PEPTIDYL_LYSINE_ACETYLATION | 14 | -0.311 | -1.144 | 0.29600 | 0.29600 | 0.56400 | 261 |
| GOBP_PROTEIN_LOCALIZATION_TO_CELL_PERIPHERY | 13 | 0.369 | 1.16 | 0.29700 | 0.29700 | 0.56400 | 112 |
| GOBP_REGULATION_OF_IMMUNE_RESPONSE | 27 | 0.297 | 1.151 | 0.29700 | 0.29700 | 0.56400 | 65 |
| GOBP_ORGANELLE_LOCALIZATION | 34 | -0.224 | -1.126 | 0.29900 | 0.29900 | 0.56400 | 121 |
| GOBP_REGULATION_OF_CELL_DIFFERENTIATION | 50 | 0.251 | 1.137 | 0.30000 | 0.30000 | 0.56400 | 85 |
| GOBP_POSITIVE_REGULATION_OF_INTRACELLULAR_TRANSPORT | 13 | 0.368 | 1.156 | 0.30000 | 0.30000 | 0.56400 | 112 |
| GOCC_CELL_PROJECTION_MEMBRANE | 10 | 0.399 | 1.152 | 0.30000 | 0.30000 | 0.56400 | 47 |
| HP_DOLICHOCEPHALY | 11 | -0.341 | -1.134 | 0.30000 | 0.30000 | 0.56400 | 144 |
| HP_ABNORMALITY_OF_THE_CURVATURE_OF_THE_VERTEBRAL_COLUMN | 50 | -0.199 | -1.122 | 0.30000 | 0.30000 | 0.56400 | 215 |
| GOBP_CELL_CYCLE | 81 | 0.226 | 1.125 | 0.30100 | 0.30100 | 0.56500 | 103 |
| HP_HYPERHIDROSIS | 10 | 0.399 | 1.15 | 0.30200 | 0.30200 | 0.56500 | 25 |
| GOBP_POSITIVE_REGULATION_OF_RESPONSE_TO_EXTERNAL_STIMULUS | 17 | 0.339 | 1.151 | 0.30200 | 0.30200 | 0.56500 | 18 |
| HP_RENAL_INSUFFICIENCY | 23 | 0.311 | 1.149 | 0.30300 | 0.30300 | 0.56600 | 203 |
| GOBP_ORGANIC_ANION_TRANSPORT | 11 | 0.385 | 1.146 | 0.30400 | 0.30400 | 0.56700 | 4 |
| GOMF_SEQUENCE_SPECIFIC_DNA_BINDING | 107 | 0.213 | 1.113 | 0.30700 | 0.30700 | 0.57000 | 327 |
| HP_INTRAUTERINE_GROWTH_RETARDATION | 23 | -0.255 | -1.122 | 0.30700 | 0.30700 | 0.57000 | 261 |
| GOBP_NEGATIVE_REGULATION_OF_DEVELOPMENTAL_PROCESS | 26 | 0.296 | 1.138 | 0.30700 | 0.30700 | 0.57000 | 100 |

| | | | | | | | |
|---|---|---|---|---|---|---|---|
| GOBP_ION_HOMEOSTASIS | 28 | 0.29 | 1.136 | 0.30900 | 0.30900 | 0.57000 | 103 |
| GOCC_ACTIN_CYTOSKELETON | 10 | -0.348 | -1.121 | 0.30900 | 0.30900 | 0.57000 | 119 |
| GOBP_PHOSPHORYLATION | 66 | 0.235 | 1.125 | 0.30900 | 0.30900 | 0.57000 | 140 |
| GOBP_RESPONSE_TO_INORGANIC_SUBSTANCE | 16 | 0.342 | 1.142 | 0.31100 | 0.31100 | 0.57200 | 222 |
| GOBP_ENDOPLASMIC_RETICULUM_TO_GOLGI_VESICLE_MEDIATED_TRANSPORT | 13 | -0.315 | -1.12 | 0.31200 | 0.31200 | 0.57200 | 286 |
| HP_HYDROCEPHALUS | 21 | 0.314 | 1.136 | 0.31200 | 0.31200 | 0.57200 | 3 |
| HP_EPILEPTIC_SPASM | 11 | -0.337 | -1.121 | 0.31200 | 0.31200 | 0.57200 | 183 |
| GOBP_LIPID_LOCALIZATION | 22 | 0.312 | 1.141 | 0.31300 | 0.31300 | 0.57200 | 188 |
| GOMF_KINASE_BINDING | 34 | 0.273 | 1.126 | 0.31300 | 0.31300 | 0.57200 | 245 |
| GOBP_PROCESS_UTILIZING_AUTOPHAGIC_MECHANISM | 31 | -0.227 | -1.115 | 0.31400 | 0.31400 | 0.57300 | 296 |
| GOCC_CELL_CORTEX | 14 | -0.304 | -1.117 | 0.31600 | 0.31600 | 0.57400 | 13 |
| GOBP_ORGANIC_CYCLIC_COMPOUND_CATABOLIC_PROCESS | 21 | -0.262 | -1.114 | 0.31700 | 0.31700 | 0.57400 | 298 |
| HP_ABNORMAL_AORTIC_MORPHOLOGY | 18 | -0.276 | -1.113 | 0.31700 | 0.31700 | 0.57400 | 300 |
| GOBP_REGULATION_OF_HEMOPOIESIS | 13 | 0.362 | 1.139 | 0.31700 | 0.31700 | 0.57400 | 218 |
| HP_COUGH | 14 | 0.353 | 1.133 | 0.31800 | 0.31800 | 0.57600 | 27 |
| GOBP_DEFENSE_RESPONSE | 42 | 0.257 | 1.116 | 0.32000 | 0.32000 | 0.57800 | 59 |
| HP_ABSENT_SPEECH | 19 | -0.272 | -1.113 | 0.32100 | 0.32100 | 0.57800 | 207 |
| HP_RENAL_HYPOPLASIA_APLASIA | 14 | 0.353 | 1.131 | 0.32100 | 0.32100 | 0.57800 | 176 |
| HP_HIGH_FOREHEAD | 17 | 0.333 | 1.131 | 0.32200 | 0.32200 | 0.57800 | 14 |
| GOBP_RESPONSE_TO_OXIDATIVE_STRESS | 18 | -0.275 | -1.108 | 0.32200 | 0.32200 | 0.57900 | 90 |
| GOBP_RESPONSE_TO_TUMOR_NECROSIS_FACTOR | 15 | 0.346 | 1.133 | 0.32400 | 0.32400 | 0.58100 | 25 |
| GOBP_ANION_TRANSPORT | 12 | 0.368 | 1.127 | 0.32600 | 0.32600 | 0.58300 | 4 |
| GOBP_MICROTUBULE_CYTOSKELETON_ORGANIZATION | 27 | 0.289 | 1.123 | 0.32600 | 0.32600 | 0.58300 | 162 |
| GOMF_PHOSPHATIDYLINOSITOL_BINDING | 17 | -0.278 | -1.103 | 0.32700 | 0.32700 | 0.58300 | 161 |
| GOBP_CELL_AGING | 10 | -0.342 | -1.102 | 0.32900 | 0.32900 | 0.58600 | 168 |
| HP_ABNORMALITY_OF_THE_HYPOTHALAMUS_PITUITARY_AXIS | 15 | 0.344 | 1.126 | 0.33000 | 0.33000 | 0.58600 | 77 |
| GOBP_REGULATION_OF_DNA_METABOLIC_PROCESS | 20 | -0.264 | -1.105 | 0.33000 | 0.33000 | 0.58600 | 216 |
| HP_ABNORMALITY_OF_SKIN_ADNEXA_PHYSIOLOGY | 12 | 0.366 | 1.121 | 0.33200 | 0.33200 | 0.58600 | 218 |
| GOBP_OSTEOBLAST_DIFFERENTIATION | 12 | 0.366 | 1.12 | 0.33200 | 0.33200 | 0.58600 | 77 |
| GOBP_DNA_METABOLIC_PROCESS | 44 | -0.202 | -1.102 | 0.33300 | 0.33300 | 0.58600 | 216 |
| HP_LONG_FINGERS | 14 | -0.299 | -1.101 | 0.33300 | 0.33300 | 0.58600 | 300 |
| HP_ABNORMAL_UVEA_MORPHOLOGY | 22 | -0.254 | -1.1 | 0.33300 | 0.33300 | 0.58600 | 213 |
| GOBP_CYTOKINE_MEDIATED_SIGNALING_PATHWAY | 15 | 0.342 | 1.121 | 0.33400 | 0.33400 | 0.58700 | 28 |
| GOBP_REGULATION_OF_LEUKOCYTE_DIFFERENTIATION | 11 | 0.376 | 1.119 | 0.33500 | 0.33500 | 0.58700 | 218 |
| GOBP_CARBOHYDRATE_DERIVATIVE_METABOLIC_PROCESS | 41 | 0.257 | 1.109 | 0.33500 | 0.33500 | 0.58700 | 250 |
| HP_ABNORMAL_INTERNAL_GENITALIA | 25 | 0.294 | 1.114 | 0.33700 | 0.33700 | 0.58700 | 77 |
| HP_ABNORMAL_LIVER_MORPHOLOGY | 32 | 0.272 | 1.105 | 0.33700 | 0.33700 | 0.58700 | 188 |
| GOBP_LOCALIZATION_WITHIN_MEMBRANE | 24 | -0.243 | -1.089 | 0.33700 | 0.33700 | 0.58700 | 123 |
| HP_ABNORMALITY_OF_THE_DENTITION | 44 | 0.252 | 1.106 | 0.33700 | 0.33700 | 0.58700 | 59 |
| GOBP_CELL_PROJECTION_ORGANIZATION | 56 | 0.238 | 1.102 | 0.33800 | 0.33800 | 0.58700 | 30 |
| HP_CORNEAL_OPACITY | 12 | 0.364 | 1.113 | 0.33800 | 0.33800 | 0.58700 | 243 |
| HP_ABNORMAL_LEUKOCYTE_COUNT | 12 | -0.316 | -1.093 | 0.33900 | 0.33900 | 0.58700 | 121 |
| GOBP_REGULATION_OF_ION_TRANSMEMBRANE_TRANSPORT | 19 | -0.267 | -1.095 | 0.34000 | 0.34000 | 0.58700 | 119 |

| | | | | | | | |
|---|---|---|---|---|---|---|---|
| HP_THICK_EYEBROW | 10 | -0.339 | -1.09 | 0.34000 | 0.34000 | 0.58700 | 207 |
| GOBP_NITROGEN_COMPOUND_TRANSPORT | 98 | -0.163 | -1.085 | 0.34200 | 0.34200 | 0.58900 | 285 |
| GOCC_NUCLEAR_ENVELOPE | 20 | 0.311 | 1.107 | 0.34200 | 0.34200 | 0.58900 | 86 |
| HP_ABNORMAL_EXTERNAL_GENITALIA | 53 | -0.189 | -1.086 | 0.34300 | 0.34300 | 0.58900 | 215 |
| GOBP_CYTOKINE_PRODUCTION | 17 | -0.274 | -1.087 | 0.34300 | 0.34300 | 0.58900 | 90 |
| GOBP_REGULATION_OF_RESPONSE_TO_BIOTIC_STIMULUS | 10 | 0.385 | 1.109 | 0.34500 | 0.34500 | 0.59100 | 144 |
| GOBP_CARBOHYDRATE_DERIVATIVE_BIOSYNTHETIC_PROCESS | 27 | 0.284 | 1.103 | 0.34700 | 0.34700 | 0.59200 | 250 |
| GOBP_POSITIVE_REGULATION_OF_IMMUNE_RESPONSE | 17 | 0.326 | 1.106 | 0.34700 | 0.34700 | 0.59200 | 222 |
| GOBP_CYTOSKELETON_ORGANIZATION | 49 | 0.244 | 1.1 | 0.34800 | 0.34800 | 0.59300 | 104 |
| GOBP_REGULATION_OF_CELLULAR_RESPONSE_TO_TRANSFORMING_GROWTH_F | 14 | 0.344 | 1.105 | 0.34800 | 0.34800 | 0.59300 | 204 |
| HP_ABNORMAL_SKIN_MORPHOLOGY_OF_THE_PALM | 22 | -0.251 | -1.086 | 0.34900 | 0.34900 | 0.59300 | 291 |
| GOBP_NEGATIVE_REGULATION_OF_CELL_POPULATION_PROLIFERATION | 25 | 0.291 | 1.103 | 0.34900 | 0.34900 | 0.59300 | 103 |
| GOBP_RESPONSE_TO_LIPID | 30 | 0.274 | 1.095 | 0.35000 | 0.35000 | 0.59300 | 18 |
| GOCC_CYTOPLASMIC_SIDE_OF_MEMBRANE | 11 | 0.371 | 1.105 | 0.35000 | 0.35000 | 0.59300 | 120 |
| GOBP_REGULATION_OF_SMALL_GTPASE_MEDIATED_SIGNAL_TRANSDUCTION | 12 | -0.313 | -1.083 | 0.35100 | 0.35100 | 0.59300 | 13 |
| GOBP_REGULATION_OF_HYDROLASE_ACTIVITY | 31 | 0.272 | 1.095 | 0.35100 | 0.35100 | 0.59300 | 103 |
| HP_POSTERIORLY_ROTATED_EARS | 23 | 0.298 | 1.101 | 0.35300 | 0.35300 | 0.59500 | 266 |
| GOCC_VESICLE_MEMBRANE | 55 | -0.186 | -1.077 | 0.35500 | 0.35500 | 0.59500 | 126 |
| HP_HEPATOMEGALY | 25 | 0.29 | 1.097 | 0.35500 | 0.35500 | 0.59500 | 188 |
| HP_INCREASED_BLOOD_PRESSURE | 17 | 0.324 | 1.098 | 0.35600 | 0.35600 | 0.59500 | 245 |
| HP_ABNORMAL_SYSTEMIC_BLOOD_PRESSURE | 19 | 0.311 | 1.094 | 0.35600 | 0.35600 | 0.59500 | 245 |
| GOBP_CELLULAR_LIPID_CATABOLIC_PROCESS | 10 | 0.381 | 1.098 | 0.35600 | 0.35600 | 0.59500 | 219 |
| GOBP_REGULATION_OF_TRANSPORTER_ACTIVITY | 12 | 0.357 | 1.093 | 0.35700 | 0.35700 | 0.59500 | 91 |
| GOBP_POSITIVE_REGULATION_OF_MOLECULAR_FUNCTION | 64 | 0.229 | 1.088 | 0.35700 | 0.35700 | 0.59500 | 132 |
| GOBP_REGULATION_OF_MAP_KINASE_ACTIVITY | 10 | 0.38 | 1.097 | 0.35700 | 0.35700 | 0.59500 | 126 |
| HP_ABNORMAL_VENTRICULAR_SEPTUM_MORPHOLOGY | 18 | 0.317 | 1.096 | 0.35800 | 0.35800 | 0.59500 | 59 |
| GOBP_REGULATION_OF_GROWTH | 18 | 0.316 | 1.094 | 0.36000 | 0.36000 | 0.59800 | 77 |
| GOBP_DIVALENT_INORGANIC_CATION_HOMEOSTASIS | 20 | 0.305 | 1.088 | 0.36100 | 0.36100 | 0.59900 | 103 |
| HP_SPLENOMEGALY | 17 | -0.269 | -1.068 | 0.36200 | 0.36200 | 0.60000 | 166 |
| HP_ABNORMALITY_OF_THE_PANCREAS | 16 | 0.327 | 1.092 | 0.36400 | 0.36400 | 0.60200 | 77 |
| HP_ABNORMALITY_OF_THE_AMNIOTIC_FLUID | 23 | 0.295 | 1.089 | 0.36500 | 0.36500 | 0.60200 | 162 |
| HP_ABNORMAL_LARYNX_MORPHOLOGY | 10 | 0.377 | 1.088 | 0.36600 | 0.36600 | 0.60200 | 59 |
| GOBP_POSITIVE_REGULATION_OF_GENE_EXPRESSION | 41 | 0.25 | 1.08 | 0.36600 | 0.36600 | 0.60200 | 81 |
| HP_ABNORMAL_BLOOD_ION_CONCENTRATION | 12 | 0.355 | 1.085 | 0.36600 | 0.36600 | 0.60200 | 147 |
| HP_ABNORMALITY_OF_THE_COAGULATION_CASCADE | 10 | -0.332 | -1.068 | 0.36700 | 0.36700 | 0.60200 | 266 |
| GOBP_NUCLEUS_ORGANIZATION | 10 | -0.332 | -1.067 | 0.36800 | 0.36800 | 0.60200 | 141 |
| GOBP_REGULATION_OF_TRANS_SYNAPTIC_SIGNALING | 16 | 0.326 | 1.089 | 0.36800 | 0.36800 | 0.60200 | 18 |
| HP_TELECANTHUS | 11 | 0.365 | 1.087 | 0.36800 | 0.36800 | 0.60200 | 59 |
| HP_LIMITATION_OF_JOINT_MOBILITY | 10 | -0.331 | -1.067 | 0.36900 | 0.36900 | 0.60200 | 190 |
| HP_HYPODONTIA | 10 | 0.375 | 1.081 | 0.37300 | 0.37300 | 0.60700 | 59 |
| HP_SLANTING_OF_THE_PALPEBRAL_FISSURE | 34 | -0.213 | -1.066 | 0.37400 | 0.37400 | 0.60900 | 215 |
| GOCC_SECRETORY_VESICLE | 35 | 0.258 | 1.075 | 0.37500 | 0.37500 | 0.60900 | 103 |
| HP_ABNORMAL_INFLAMMATORY_RESPONSE | 45 | 0.243 | 1.068 | 0.38000 | 0.38000 | 0.61500 | 28 |

| | | | | | | | |
|---|---|---|---|---|---|---|---|
| HP_DYSPHAGIA | 20 | -0.254 | -1.063 | 0.38000 | 0.38000 | 0.61500 | 153 |
| GOMF_IDENTICAL_PROTEIN_BINDING | 81 | 0.213 | 1.064 | 0.38100 | 0.38100 | 0.61500 | 77 |
| GOBP_PROTEIN_LOCALIZATION_TO_NUCLEUS | 15 | -0.28 | -1.058 | 0.38100 | 0.38100 | 0.61500 | 146 |
| HP_CEREBELLAR_HYPOPLASIA | 22 | -0.244 | -1.056 | 0.38300 | 0.38300 | 0.61700 | 14 |
| HP_HIGH_PALATE | 39 | -0.201 | -1.058 | 0.38400 | 0.38400 | 0.61800 | 213 |
| GOBP_POSITIVE_REGULATION_OF_MAPK_CASCADE | 15 | 0.327 | 1.072 | 0.38500 | 0.38500 | 0.61800 | 148 |
| GOBP_TELOMERE_ORGANIZATION | 10 | -0.326 | -1.049 | 0.38600 | 0.38600 | 0.61800 | 196 |
| GOBP_POSITIVE_REGULATION_OF_PROTEIN_MODIFICATION_BY_SMALL_PROTEIN | 10 | 0.371 | 1.069 | 0.38600 | 0.38600 | 0.61800 | 46 |
| HP_BRACHYDACTYLY | 18 | 0.309 | 1.069 | 0.38600 | 0.38600 | 0.61800 | 59 |
| HP_ABNORMAL_HAIR_QUANTITY | 33 | -0.211 | -1.052 | 0.38800 | 0.38800 | 0.61900 | 213 |
| HP_ABNORMAL_UPPER_LIMB_BONE_MORPHOLOGY | 24 | 0.286 | 1.073 | 0.38800 | 0.38800 | 0.61900 | 162 |
| HP_ABNORMALITY_OF_THE_PINNA | 32 | -0.214 | -1.06 | 0.38900 | 0.38900 | 0.61900 | 215 |
| GOMF_ENZYME_REGULATOR_ACTIVITY | 52 | -0.183 | -1.053 | 0.38900 | 0.38900 | 0.61900 | 126 |
| GOMF_TRANSCRIPTION_REGULATOR_ACTIVITY | 117 | 0.2 | 1.061 | 0.39000 | 0.39000 | 0.61900 | 280 |
| HP_ABNORMALITY_OF_SKIN_PHYSIOLOGY | 11 | 0.357 | 1.065 | 0.39100 | 0.39100 | 0.61900 | 27 |
| GOBP_CELLULAR_RESPONSE_TO_HORMONE_STIMULUS | 22 | 0.291 | 1.064 | 0.39100 | 0.39100 | 0.61900 | 179 |
| GOBP_NEGATIVE_REGULATION_OF_GENE_EXPRESSION | 43 | 0.243 | 1.061 | 0.39100 | 0.39100 | 0.61900 | 81 |
| GOBP_REGULATION_OF_LIPID_METABOLIC_PROCESS | 15 | 0.324 | 1.062 | 0.39300 | 0.39300 | 0.62000 | 163 |
| GOBP_REGULATION_OF_CATION_TRANSMEMBRANE_TRANSPORT | 14 | 0.33 | 1.058 | 0.39300 | 0.39300 | 0.62000 | 115 |
| HP_COGNITIVE_IMPAIRMENT | 18 | -0.258 | -1.04 | 0.39400 | 0.39400 | 0.62000 | 207 |
| HP_ABNORMALITY_OF_THE_URETER | 13 | -0.292 | -1.04 | 0.39600 | 0.39600 | 0.62200 | 215 |
| GOBP_PROTEIN_METHYLATION | 11 | -0.313 | -1.043 | 0.39700 | 0.39700 | 0.62200 | 216 |
| GOBP_MAPK_CASCADE | 25 | 0.279 | 1.059 | 0.39700 | 0.39700 | 0.62200 | 74 |
| HP_JOINT_DISLOCATION | 19 | -0.254 | -1.042 | 0.39700 | 0.39700 | 0.62200 | 231 |
| GOMF_KINASE_REGULATOR_ACTIVITY | 17 | 0.312 | 1.06 | 0.39900 | 0.39900 | 0.62200 | 126 |
| GOCC_INTRINSIC_COMPONENT_OF_ORGANELLE_MEMBRANE | 18 | 0.306 | 1.058 | 0.39900 | 0.39900 | 0.62200 | 52 |
| GOCC_CYTOPLASMIC_REGION | 13 | -0.292 | -1.037 | 0.39900 | 0.39900 | 0.62200 | 23 |
| GOBP_RESPONSE_TO_PEPTIDE | 15 | 0.323 | 1.056 | 0.40000 | 0.40000 | 0.62200 | 103 |
| GOBP_REGULATION_OF_ACTIN_FILAMENT_BASED_PROCESS | 14 | -0.281 | -1.036 | 0.40000 | 0.40000 | 0.62200 | 123 |
| GOBP_HISTONE_MODIFICATION | 31 | -0.214 | -1.052 | 0.40100 | 0.40100 | 0.62200 | 226 |
| GOBP_REGULATION_OF_SYSTEM_PROCESS | 14 | 0.327 | 1.05 | 0.40100 | 0.40100 | 0.62200 | 29 |
| GOBP_METHYLATION | 20 | -0.249 | -1.043 | 0.40200 | 0.40200 | 0.62200 | 216 |
| HP_ABNORMAL_METACARPAL_MORPHOLOGY | 10 | 0.366 | 1.054 | 0.40200 | 0.40200 | 0.62200 | 59 |
| HP_ABNORMALITY_OF_URINE_HOMEOSTASIS | 21 | 0.291 | 1.052 | 0.40400 | 0.40400 | 0.62400 | 203 |
| GOBP_BIOLOGICAL_PROCESS_INVOLVED_IN_INTERSPECIES_INTERACTION_BETW | 53 | 0.23 | 1.051 | 0.40500 | 0.40500 | 0.62400 | 59 |
| GOBP_SIGNAL_RELEASE | 10 | 0.365 | 1.052 | 0.40500 | 0.40500 | 0.62400 | 12 |
| GOBP_ORGANOPHOSPHATE_BIOSYNTHETIC_PROCESS | 17 | -0.259 | -1.025 | 0.40600 | 0.40600 | 0.62500 | 96 |
| HP_HERNIA | 25 | -0.227 | -1.033 | 0.40700 | 0.40700 | 0.62500 | 238 |
| GOBP_SMALL_GTPASE_MEDIATED_SIGNAL_TRANSDUCTION | 23 | 0.284 | 1.049 | 0.40800 | 0.40800 | 0.62500 | 101 |
| GOBP_CELLULAR_RESPONSE_TO_OXYGEN_CONTAINING_COMPOUND | 35 | -0.206 | -1.041 | 0.40800 | 0.40800 | 0.62500 | 84 |
| GOBP_REGULATION_OF_CELL_PROJECTION_ORGANIZATION | 20 | -0.247 | -1.036 | 0.40900 | 0.40900 | 0.62600 | 159 |
| HP_ABNORMALITY_OF_FLUID_REGULATION | 18 | 0.303 | 1.047 | 0.40900 | 0.40900 | 0.62600 | 77 |
| GOBP_DEFENSE_RESPONSE_TO_OTHER_ORGANISM | 28 | 0.268 | 1.048 | 0.41100 | 0.41100 | 0.62700 | 59 |

| | | | | | | | |
|---|---|---|---|---|---|---|---|
| GOBP_REGULATION_OF_HISTONE_MODIFICATION | 13 | -0.288 | -1.025 | 0.41100 | 0.41100 | 0.62700 | 261 |
| GOBP_NEGATIVE_REGULATION_OF_PHOSPHORUS_METABOLIC_PROCESS | 18 | 0.301 | 1.043 | 0.41300 | 0.41300 | 0.62700 | 204 |
| HP_ABNORMAL_HEART_VALVE_MORPHOLOGY | 12 | -0.296 | -1.025 | 0.41300 | 0.41300 | 0.62700 | 231 |
| HP_ABNORMALITY_OF_THE_MIDDLE_EAR | 31 | 0.259 | 1.041 | 0.41300 | 0.41300 | 0.62700 | 162 |
| GOBP_ORGANIC_ACID_TRANSPORT | 11 | 0.35 | 1.044 | 0.41400 | 0.41400 | 0.62700 | 4 |
| GOBP_REGULATION_OF_CELL_ACTIVATION | 11 | 0.35 | 1.043 | 0.41500 | 0.41500 | 0.62700 | 218 |
| GOBP_NEGATIVE_REGULATION_OF_CELL_CYCLE_PROCESS | 12 | -0.296 | -1.024 | 0.41600 | 0.41600 | 0.62700 | 133 |
| GOBP_REGULATION_OF_APOPTOTIC_SIGNALING_PATHWAY | 15 | -0.272 | -1.027 | 0.41600 | 0.41600 | 0.62700 | 90 |
| GOCC_GOLGI_APPARATUS_SUBCOMPARTMENT | 17 | -0.257 | -1.018 | 0.41700 | 0.41700 | 0.62700 | 383 |
| GOBP_CELLULAR_RESPONSE_TO_LIPID | 21 | -0.242 | -1.031 | 0.41800 | 0.41800 | 0.62700 | 125 |
| GOBP_BIOLOGICAL_PROCESS_INVOLVED_IN_SYMBIOTIC_INTERACTION | 14 | -0.278 | -1.022 | 0.41800 | 0.41800 | 0.62700 | 18 |
| GOCC_CILIUM | 30 | 0.26 | 1.037 | 0.41800 | 0.41800 | 0.62700 | 355 |
| HP_HYPOPITUITARISM | 12 | 0.34 | 1.039 | 0.41900 | 0.41900 | 0.62700 | 71 |
| HP_ABNORMAL_MYOCARDIUM_MORPHOLOGY | 14 | -0.277 | -1.02 | 0.41900 | 0.41900 | 0.62700 | 190 |
| GOCC_CELL_SUBSTRATE_JUNCTION | 19 | 0.295 | 1.038 | 0.41900 | 0.41900 | 0.62700 | 103 |
| GOBP_POSITIVE_REGULATION_OF_DEFENSE_RESPONSE | 11 | -0.307 | -1.023 | 0.42000 | 0.42000 | 0.62700 | 54 |
| GOBP_REGULATION_OF_INTRACELLULAR_PROTEIN_TRANSPORT | 15 | 0.315 | 1.032 | 0.42200 | 0.42200 | 0.62800 | 112 |
| HP_ABNORMALITY_OF_THE_HALLUX | 11 | 0.348 | 1.036 | 0.42200 | 0.42200 | 0.62800 | 54 |
| HP_ABNORMAL_THROMBOCYTE_MORPHOLOGY | 20 | -0.245 | -1.024 | 0.42300 | 0.42300 | 0.62800 | 132 |
| GOBP_CHROMATIN_ASSEMBLY_OR_DISASSEMBLY | 19 | -0.248 | -1.018 | 0.42300 | 0.42300 | 0.62800 | 261 |
| GOCC_PRESYNAPSE | 18 | 0.298 | 1.032 | 0.42400 | 0.42400 | 0.62900 | 52 |
| HP_ABNORMALITY_OF_THE_EXTERNAL_NOSE | 47 | -0.186 | -1.027 | 0.42500 | 0.42500 | 0.62900 | 190 |
| HP_GAIT_DISTURBANCE | 39 | -0.196 | -1.031 | 0.42500 | 0.42500 | 0.62900 | 156 |
| HP_ABNORMALITY_OF_UPPER_LIMB_JOINT | 18 | -0.251 | -1.014 | 0.42700 | 0.42700 | 0.63000 | 176 |
| HP_POLYMICROGYRIA | 10 | 0.356 | 1.028 | 0.42700 | 0.42700 | 0.63100 | 260 |
| HP_CONGENITAL_ONSET | 22 | -0.235 | -1.017 | 0.42900 | 0.42900 | 0.63300 | 194 |
| GOCC_ENDOPLASMIC_RETICULUM_PROTEIN_CONTAINING_COMPLEX | 10 | -0.314 | -1.012 | 0.43000 | 0.43000 | 0.63300 | 493 |
| HP_INVOLUNTARY_MOVEMENTS | 38 | -0.197 | -1.025 | 0.43400 | 0.43400 | 0.63700 | 191 |
| HP_CEREBRAL_CORTICAL_ATROPHY | 17 | -0.253 | -1.004 | 0.43400 | 0.43400 | 0.63700 | 300 |
| GOBP_ORGANOPHOSPHATE_METABOLIC_PROCESS | 26 | -0.22 | -1.009 | 0.43500 | 0.43500 | 0.63800 | 96 |
| HP_IMMUNODEFICIENCY | 12 | 0.335 | 1.023 | 0.43900 | 0.43900 | 0.64200 | 28 |
| GOBP_GLYCOPROTEIN_BIOSYNTHETIC_PROCESS | 15 | 0.309 | 1.012 | 0.44600 | 0.44600 | 0.65000 | 250 |
| GOBP_POSITIVE_REGULATION_OF_IMMUNE_SYSTEM_PROCESS | 24 | 0.273 | 1.022 | 0.44600 | 0.44600 | 0.65000 | 222 |
| GOMF_GUANYL_NUCLEOTIDE_BINDING | 16 | 0.305 | 1.018 | 0.44600 | 0.44600 | 0.65000 | 45 |
| HP_SKELETAL_DYSPLASIA | 12 | 0.332 | 1.015 | 0.44700 | 0.44700 | 0.65100 | 286 |
| GOBP_CELL_GROWTH | 15 | -0.264 | -0.997 | 0.45000 | 0.45000 | 0.65400 | 141 |
| HP_ABNORMAL_5TH_FINGER_MORPHOLOGY | 27 | 0.262 | 1.015 | 0.45100 | 0.45100 | 0.65500 | 71 |
| GOCC_ENDOSOME | 37 | -0.196 | -1.012 | 0.45200 | 0.45200 | 0.65500 | 177 |
| HP_INCREASED_SUSCEPTIBILITY_TO_FRACTURES | 15 | -0.264 | -0.996 | 0.45200 | 0.45200 | 0.65500 | 166 |
| GOMF_DNA_BINDING_TRANSCRIPTION_REPRESSOR_ACTIVITY | 20 | 0.283 | 1.01 | 0.45300 | 0.45300 | 0.65500 | 228 |
| GOBP_DNA_CONFORMATION_CHANGE | 19 | -0.242 | -0.993 | 0.45300 | 0.45300 | 0.65500 | 261 |
| GOBP_CELL_ACTIVATION_INVOLVED_IN_IMMUNE_RESPONSE | 14 | -0.268 | -0.987 | 0.45500 | 0.45500 | 0.65700 | 121 |
| HP_LANGUAGE_IMPAIRMENT | 43 | -0.185 | -1.004 | 0.46000 | 0.46000 | 0.66200 | 156 |

| | | | | | | | |
|---|---|---|---|---|---|---|---|
| HP_IMPAIRMENT_IN_PERSONALITY_FUNCTIONING | 28 | -0.209 | -0.991 | 0.46300 | 0.46300 | 0.66600 | 313 |
| GOBP_EPITHELIAL_CELL_DIFFERENTIATION | 25 | 0.264 | 1.001 | 0.46500 | 0.46500 | 0.66700 | 179 |
| GOMF_TUBULIN_BINDING | 10 | 0.346 | 0.996 | 0.46600 | 0.46600 | 0.66700 | 349 |
| GOBP_ORGANIC_HYDROXY_COMPOUND_METABOLIC_PROCESS | 14 | 0.309 | 0.992 | 0.46600 | 0.46600 | 0.66700 | 236 |
| GOBP_ESTABLISHMENT_OF_PROTEIN_LOCALIZATION | 87 | -0.155 | -1.004 | 0.46600 | 0.46600 | 0.66700 | 285 |
| HP_NAUSEA_AND_VOMITING | 13 | 0.317 | 0.996 | 0.46700 | 0.46700 | 0.66800 | 77 |
| GOBP_POTASSIUM_ION_TRANSPORT | 12 | -0.283 | -0.982 | 0.46800 | 0.46800 | 0.66800 | 125 |
| HP_ABNORMALITY_OF_FACIAL_SKELETON | 63 | 0.211 | 0.998 | 0.46900 | 0.46900 | 0.66800 | 71 |
| HP_ABNORMAL_THORAX_MORPHOLOGY | 36 | -0.195 | -1 | 0.46900 | 0.46900 | 0.66800 | 183 |
| HP_MOTOR_SEIZURE | 12 | -0.283 | -0.979 | 0.47100 | 0.47100 | 0.66800 | 183 |
| HP_FEVER | 17 | -0.247 | -0.978 | 0.47100 | 0.47100 | 0.66800 | 149 |
| HP_WEIGHT_LOSS | 15 | 0.303 | 0.991 | 0.47100 | 0.47100 | 0.66800 | 218 |
| GOCC_UBIQUITIN_LIGASE_COMPLEX | 15 | 0.303 | 0.991 | 0.47100 | 0.47100 | 0.66800 | 407 |
| GOBP_REGULATION_OF_MITOTIC_CELL_CYCLE_PHASE_TRANSITION | 21 | 0.275 | 0.994 | 0.47200 | 0.47200 | 0.66800 | 73 |
| HP_ABNORMALITY_OF_THE_ELBOW | 10 | -0.303 | -0.975 | 0.47400 | 0.47400 | 0.66900 | 207 |
| GOBP_IN_UTERO_EMBRYONIC_DEVELOPMENT | 20 | 0.278 | 0.99 | 0.47500 | 0.47500 | 0.67000 | 148 |
| GOBP_CELLULAR_AMIDE_METABOLIC_PROCESS | 45 | -0.181 | -0.995 | 0.47700 | 0.47700 | 0.67300 | 247 |
| GOBP_NEGATIVE_REGULATION_OF_MITOTIC_CELL_CYCLE | 11 | -0.292 | -0.973 | 0.47800 | 0.47800 | 0.67300 | 133 |
| HP_ABNORMALITY_OF_THE_CEREBROSPINAL_FLUID | 26 | 0.258 | 0.99 | 0.48000 | 0.48000 | 0.67500 | 3 |
| HP_FINGER_CLINODACTYLY | 22 | -0.225 | -0.973 | 0.48100 | 0.48100 | 0.67500 | 207 |
| HP_ABNORMAL_BONE_STRUCTURE | 35 | -0.195 | -0.987 | 0.48100 | 0.48100 | 0.67500 | 214 |
| GOCC_EXOCYTIC_VESICLE | 10 | 0.34 | 0.979 | 0.48400 | 0.48400 | 0.67800 | 52 |
| HP_ABNORMALITY_OF_THE_METACARPAL_BONES | 11 | 0.33 | 0.984 | 0.48400 | 0.48400 | 0.67800 | 59 |
| HP_OLIGOHYDRAMNIOS | 10 | 0.338 | 0.975 | 0.48900 | 0.48900 | 0.68200 | 243 |
| GOCC_SPINDLE | 17 | 0.289 | 0.981 | 0.49000 | 0.49000 | 0.68200 | 305 |
| HP_SHORT_DIGIT | 28 | 0.25 | 0.98 | 0.49000 | 0.49000 | 0.68200 | 162 |
| HP_TRIANGULAR_FACE | 13 | 0.31 | 0.974 | 0.49000 | 0.49000 | 0.68200 | 205 |
| GOBP_PEPTIDYL_LYSINE_MODIFICATION | 25 | -0.214 | -0.977 | 0.49100 | 0.49100 | 0.68200 | 261 |
| GOBP_ORGANOPHOSPHATE_ESTER_TRANSPORT | 10 | -0.299 | -0.964 | 0.49100 | 0.49100 | 0.68200 | 18 |
| GOBP_MITOTIC_CELL_CYCLE_PHASE_TRANSITION | 24 | 0.261 | 0.979 | 0.49200 | 0.49200 | 0.68200 | 95 |
| GOBP_SMALL_MOLECULE_CATABOLIC_PROCESS | 11 | 0.328 | 0.977 | 0.49300 | 0.49300 | 0.68200 | 12 |
| GOCC_NEURON_PROJECTION | 36 | 0.234 | 0.981 | 0.49300 | 0.49300 | 0.68200 | 112 |
| GOBP_CELLULAR_RESPONSE_TO_CHEMICAL_STRESS | 13 | 0.309 | 0.971 | 0.49400 | 0.49400 | 0.68200 | 73 |
| HP_INTERICTAL_EEG_ABNORMALITY | 11 | -0.29 | -0.965 | 0.49400 | 0.49400 | 0.68200 | 183 |
| GOBP_RESPONSE_TO_VIRUS | 21 | -0.227 | -0.966 | 0.49600 | 0.49600 | 0.68400 | 150 |
| GOBP_NUCLEOSOME_ORGANIZATION | 10 | -0.298 | -0.959 | 0.49800 | 0.49800 | 0.68400 | 260 |
| HP_ABNORMALITY_OF_THE_LYMPHATIC_SYSTEM | 38 | 0.23 | 0.975 | 0.49800 | 0.49800 | 0.68400 | 27 |
| HP_ABNORMALITY_OF_NEURONAL_MIGRATION | 23 | -0.219 | -0.962 | 0.49800 | 0.49800 | 0.68400 | 144 |
| GOBP_SKIN_DEVELOPMENT | 11 | 0.326 | 0.973 | 0.49800 | 0.49800 | 0.68400 | 179 |
| GOBP_MICROTUBULE_BASED_MOVEMENT | 13 | -0.268 | -0.955 | 0.49900 | 0.49900 | 0.68400 | 36 |
| GOCC_ENDOPLASMIC_RETICULUM | 88 | 0.194 | 0.979 | 0.50000 | 0.50000 | 0.68400 | 94 |
| HP_ABNORMALITY_OF_THE_LYMPH_NODES | 11 | -0.288 | -0.959 | 0.50100 | 0.50100 | 0.68400 | 149 |
| HP_APLASIA_HYPOPLASIA_OF_THE_NAILS | 14 | 0.3 | 0.964 | 0.50200 | 0.50200 | 0.68400 | 71 |

| | | | | | | | |
|---|---|---|---|---|---|---|---|
| GOBP_EMBRYO_DEVELOPMENT_ENDING_IN_BIRTH_OR_EGG_HATCHING | 29 | 0.246 | 0.975 | 0.50200 | 0.50200 | 0.68400 | 77 |
| HP_ABNORMALITY_OF_FACIAL_MUSCULATURE | 10 | -0.297 | -0.955 | 0.50200 | 0.50200 | 0.68400 | 183 |
| HP_ABNORMALITY_OF_DENTAL_STRUCTURE | 20 | 0.271 | 0.967 | 0.50300 | 0.50300 | 0.68500 | 59 |
| HP_INTESTINAL_MALROTATION | 13 | 0.306 | 0.963 | 0.50400 | 0.50400 | 0.68500 | 59 |
| HP_NARROW_PALATE | 11 | -0.287 | -0.957 | 0.50500 | 0.50500 | 0.68500 | 3 |
| HP_ABNORMAL_LOCATION_OF_EARS | 43 | 0.223 | 0.974 | 0.50700 | 0.50700 | 0.68700 | 77 |
| HP_SLEEP_DISTURBANCE | 26 | -0.21 | -0.964 | 0.50700 | 0.50700 | 0.68700 | 300 |
| HP_ABNORMAL_CARDIAC_VENTRICLE_MORPHOLOGY | 26 | 0.252 | 0.967 | 0.50800 | 0.50800 | 0.68700 | 77 |
| HP_TREMOR | 19 | -0.232 | -0.951 | 0.50900 | 0.50900 | 0.68700 | 553 |
| HP_FUNCTIONAL_ABNORMALITY_OF_THE_GASTROINTESTINAL_TRACT | 43 | -0.179 | -0.971 | 0.50900 | 0.50900 | 0.68700 | 239 |
| HP_ABNORMALITY_OF_COORDINATION | 36 | -0.189 | -0.969 | 0.51200 | 0.51200 | 0.69000 | 586 |
| HP_ABNORMAL_TESTIS_MORPHOLOGY | 46 | -0.177 | -0.975 | 0.51300 | 0.51300 | 0.69000 | 215 |
| GOBP_REGULATION_OF_NEURON_PROJECTION_DEVELOPMENT | 13 | -0.266 | -0.944 | 0.51500 | 0.51500 | 0.69100 | 156 |
| GOCC_TRANS_GOLGI_NETWORK | 15 | -0.251 | -0.946 | 0.51500 | 0.51500 | 0.69100 | 539 |
| GOBP_REGULATION_OF_METAL_ION_TRANSPORT | 16 | 0.286 | 0.955 | 0.51600 | 0.51600 | 0.69200 | 115 |
| GOBP_CELL_CYCLE_G1_S_PHASE_TRANSITION | 10 | 0.33 | 0.953 | 0.51900 | 0.51900 | 0.69200 | 1 |
| GOBP_CELLULAR_RESPONSE_TO_RADIATION | 10 | 0.33 | 0.953 | 0.51900 | 0.51900 | 0.69200 | 46 |
| HP_MIDFACE_RETRUSION | 16 | 0.285 | 0.952 | 0.51900 | 0.51900 | 0.69200 | 71 |
| HP_LOCALIZED_SKIN_LESION | 41 | -0.18 | -0.962 | 0.52000 | 0.52000 | 0.69200 | 215 |
| GOBP_TRNA_METABOLIC_PROCESS | 15 | -0.249 | -0.941 | 0.52000 | 0.52000 | 0.69200 | 540 |
| GOBP_POSITIVE_REGULATION_OF_NERVOUS_SYSTEM_DEVELOPMENT | 10 | -0.292 | -0.939 | 0.52200 | 0.52200 | 0.69400 | 509 |
| GOMF_GTPASE_ACTIVATOR_ACTIVITY | 13 | -0.264 | -0.939 | 0.52200 | 0.52200 | 0.69400 | 163 |
| GOCC_SECRETORY_GRANULE | 25 | 0.252 | 0.954 | 0.52400 | 0.52400 | 0.69600 | 103 |
| HP_PUBERTY_AND_GONADAL_DISORDERS | 30 | -0.196 | -0.948 | 0.52800 | 0.52800 | 0.69900 | 185 |
| HP_ABNORMAL_VENOUS_MORPHOLOGY | 10 | 0.327 | 0.944 | 0.52900 | 0.52900 | 0.69900 | 3 |
| GOBP_NEGATIVE_REGULATION_OF_CATABOLIC_PROCESS | 20 | -0.225 | -0.941 | 0.53000 | 0.53000 | 0.69900 | 90 |
| HP_CLEFT_LIP | 10 | 0.327 | 0.943 | 0.53000 | 0.53000 | 0.69900 | 59 |
| HP_CLEFT_UPPER_LIP | 10 | 0.327 | 0.943 | 0.53000 | 0.53000 | 0.69900 | 59 |
| HP_PHENOTYPIC_VARIABILITY | 12 | -0.269 | -0.932 | 0.53100 | 0.53100 | 0.69900 | 207 |
| HP_ABNORMAL_GLUCOSE_HOMEOSTASIS | 24 | -0.21 | -0.938 | 0.53100 | 0.53100 | 0.69900 | 185 |
| GOBP_GLYCOPROTEIN_METABOLIC_PROCESS | 16 | 0.282 | 0.941 | 0.53300 | 0.53300 | 0.69900 | 250 |
| HP_ABNORMALITY_OF_PRENATAL_DEVELOPMENT_OR_BIRTH | 43 | 0.218 | 0.953 | 0.53300 | 0.53300 | 0.69900 | 162 |
| GOBP_ESTABLISHMENT_OF_ORGANELLE_LOCALIZATION | 23 | -0.212 | -0.934 | 0.53300 | 0.53300 | 0.69900 | 286 |
| HP_ABNORMAL_VASCULAR_PHYSIOLOGY | 10 | 0.326 | 0.94 | 0.53400 | 0.53400 | 0.69900 | 245 |
| GOBP_MONONUCLEAR_CELL_DIFFERENTIATION | 18 | 0.272 | 0.941 | 0.53600 | 0.53600 | 0.70100 | 218 |
| HP_PES_PLANUS | 15 | -0.245 | -0.925 | 0.53800 | 0.53800 | 0.70100 | 238 |
| HP_ABNORMALITY_OF_HEPATOBILIARY_SYSTEM_PHYSIOLOGY | 10 | 0.325 | 0.938 | 0.53800 | 0.53800 | 0.70100 | 162 |
| GOCC_SUPRAMOLECULAR_POLYMER | 22 | 0.258 | 0.943 | 0.53900 | 0.53900 | 0.70100 | 70 |
| GOBP_NON_MEMBRANE_BOUNDED_ORGANELLE_ASSEMBLY | 17 | 0.276 | 0.938 | 0.54000 | 0.54000 | 0.70100 | 98 |
| HP_ABNORMALITY_OF_DENTAL_MORPHOLOGY | 19 | 0.267 | 0.94 | 0.54000 | 0.54000 | 0.70100 | 20 |
| HP_SMALL_FOR_GESTATIONAL_AGE | 11 | 0.314 | 0.936 | 0.54000 | 0.54000 | 0.70100 | 205 |
| GOBP_NEGATIVE_REGULATION_OF_CELL_DIFFERENTIATION | 21 | 0.259 | 0.937 | 0.54200 | 0.54200 | 0.70100 | 85 |
| GOBP_CHEMICAL_HOMEOSTASIS | 41 | 0.219 | 0.946 | 0.54200 | 0.54200 | 0.70100 | 103 |

| | | | | | | | |
|---|---|---|---|---|---|---|---|
| GOCC_SUPRAMOLECULAR_COMPLEX | 38 | -0.182 | -0.945 | 0.54200 | 0.54200 | 0.70100 | 266 |
| HP_ABNORMALITY_OF_MOUTH_SIZE | 28 | 0.241 | 0.942 | 0.54300 | 0.54300 | 0.70100 | 2 |
| HP_FUNCTIONAL_ABNORMALITY_OF_THE_MIDDLE_EAR | 29 | 0.237 | 0.939 | 0.54300 | 0.54300 | 0.70100 | 162 |
| GOBP_NEGATIVE_REGULATION_OF_PROTEIN_METABOLIC_PROCESS | 42 | 0.217 | 0.944 | 0.54300 | 0.54300 | 0.70100 | 126 |
| GOBP_ACTIN_FILAMENT_BASED_PROCESS | 24 | 0.25 | 0.938 | 0.54400 | 0.54400 | 0.70200 | 148 |
| GOBP_REGULATION_OF_ESTABLISHMENT_OF_PROTEIN_LOCALIZATION | 26 | 0.244 | 0.938 | 0.54600 | 0.54600 | 0.70300 | 133 |
| HP_ABNORMAL_EYELASH_MORPHOLOGY | 16 | -0.237 | -0.916 | 0.54800 | 0.54800 | 0.70300 | 238 |
| HP_ABNORMAL_DISTAL_PHALANX_MORPHOLOGY_OF_FINGER | 12 | 0.304 | 0.93 | 0.54800 | 0.54800 | 0.70300 | 205 |
| GOCC_TRANSFERASE_COMPLEX_TRANSFERRING_PHOSPHORUS_CONTAINING_( | 12 | 0.304 | 0.929 | 0.55000 | 0.55000 | 0.70400 | 1 |
| HP_POLYDACTYLY | 16 | 0.278 | 0.927 | 0.55100 | 0.55100 | 0.70400 | 355 |
| GOBP_RNA_MODIFICATION | 14 | -0.249 | -0.916 | 0.55100 | 0.55100 | 0.70400 | 540 |
| HP_ABNORMALITY_OF_BLOOD_CIRCULATION | 11 | -0.276 | -0.919 | 0.55100 | 0.55100 | 0.70400 | 276 |
| GOBP_REGULATION_OF_CELL_CYCLE_PHASE_TRANSITION | 23 | 0.253 | 0.934 | 0.55100 | 0.55100 | 0.70400 | 73 |
| HP_EPICANTHUS | 27 | -0.2 | -0.929 | 0.55700 | 0.55700 | 0.71100 | 156 |
| GOBP_IMMUNE_EFFECTOR_PROCESS | 18 | 0.266 | 0.922 | 0.56000 | 0.56000 | 0.71300 | 65 |
| HP_ABNORMALITY_OF_THE_FOREARM | 12 | 0.3 | 0.916 | 0.56400 | 0.56400 | 0.71700 | 162 |
| GOBP_ESTABLISHMENT_OF_PROTEIN_LOCALIZATION_TO_MEMBRANE | 15 | 0.28 | 0.916 | 0.56500 | 0.56500 | 0.71700 | 112 |
| GOBP_CELL_CYCLE_PHASE_TRANSITION | 26 | 0.24 | 0.922 | 0.56500 | 0.56500 | 0.71700 | 95 |
| HP_CAFE_AU_LAIT_SPOT | 11 | -0.273 | -0.91 | 0.56600 | 0.56600 | 0.71800 | 300 |
| GOCC_CHROMOSOMAL_REGION | 16 | -0.233 | -0.902 | 0.56700 | 0.56700 | 0.71800 | 196 |
| GOBP_GLIOGENESIS | 11 | -0.273 | -0.908 | 0.56800 | 0.56800 | 0.71800 | 5 |
| HP_FULL_CHEEKS | 15 | -0.24 | -0.905 | 0.56800 | 0.56800 | 0.71800 | 37 |
| HP_HETEROTROPIA | 11 | 0.306 | 0.913 | 0.56900 | 0.56900 | 0.71800 | 54 |
| HP_ABNORMALITY_OF_BONE_MINERAL_DENSITY | 28 | -0.194 | -0.919 | 0.57000 | 0.57000 | 0.71800 | 581 |
| HP_NEOPLASM_OF_THE_SKIN | 10 | -0.281 | -0.905 | 0.57000 | 0.57000 | 0.71800 | 165 |
| GOBP_REGULATION_OF_PROTEIN_LOCALIZATION | 40 | -0.176 | -0.928 | 0.57100 | 0.57100 | 0.71800 | 141 |
| HP_HYPERACTIVITY | 28 | -0.194 | -0.918 | 0.57100 | 0.57100 | 0.71800 | 300 |
| GOBP_REGULATION_OF_PROTEIN_MODIFICATION_BY_SMALL_PROTEIN_CONJUG | 14 | 0.283 | 0.907 | 0.57300 | 0.57300 | 0.71900 | 46 |
| HP_ABNORMAL_ABDOMEN_MORPHOLOGY | 28 | 0.235 | 0.918 | 0.57400 | 0.57400 | 0.71900 | 188 |
| HP_ABNORMAL_CIRCULATING_PROTEIN_CONCENTRATION | 13 | -0.253 | -0.901 | 0.57400 | 0.57400 | 0.71900 | 183 |
| GOBP_INTRACELLULAR_PROTEIN_TRANSPORT | 48 | -0.168 | -0.932 | 0.57400 | 0.57400 | 0.71900 | 285 |
| GOBP_REGULATION_OF_INTRACELLULAR_TRANSPORT | 18 | 0.261 | 0.902 | 0.58300 | 0.58300 | 0.72800 | 112 |
| HP_RETINAL_DEGENERATION | 12 | -0.259 | -0.897 | 0.58300 | 0.58300 | 0.72800 | 127 |
| HP_ABNORMALITY_OF_DIGESTIVE_SYSTEM_MORPHOLOGY | 45 | 0.208 | 0.916 | 0.58300 | 0.58300 | 0.72800 | 94 |
| GOBP_REGULATION_OF_CHROMOSOME_ORGANIZATION | 15 | -0.237 | -0.894 | 0.58600 | 0.58600 | 0.72900 | 284 |
| HP_DYSPNEA | 21 | 0.25 | 0.905 | 0.58600 | 0.58600 | 0.72900 | 27 |
| HP_ABNORMALITY_OF_THE_CHEEK | 20 | -0.214 | -0.897 | 0.58700 | 0.58700 | 0.72900 | 49 |
| GOBP_RESPONSE_TO_ENDOPLASMIC_RETICULUM_STRESS | 19 | -0.217 | -0.891 | 0.58800 | 0.58800 | 0.73100 | 198 |
| HP_DYSARTHRIA | 19 | 0.255 | 0.897 | 0.59100 | 0.59100 | 0.73300 | 190 |
| GOBP_INTRACELLULAR_RECEPTOR_SIGNALING_PATHWAY | 15 | -0.236 | -0.891 | 0.59100 | 0.59100 | 0.73300 | 221 |
| HP_ABNORMAL_FEAR_ANXIETY_RELATED_BEHAVIOR | 22 | -0.207 | -0.895 | 0.59200 | 0.59200 | 0.73300 | 313 |
| HP_ABNORMAL_CORNEA_MORPHOLOGY | 33 | -0.183 | -0.909 | 0.59300 | 0.59300 | 0.73300 | 215 |
| GOBP_POSITIVE_REGULATION_OF_HYDROLASE_ACTIVITY | 21 | 0.248 | 0.898 | 0.59400 | 0.59400 | 0.73400 | 103 |

| | | | | | | | |
|---|---|---|---|---|---|---|---|
| HP_PTOSIS | 26 | 0.235 | 0.901 | 0.59500 | 0.59500 | 0.73400 | 190 |
| GOBP_REGULATION_OF_CYTOSKELETON_ORGANIZATION | 20 | 0.251 | 0.896 | 0.59600 | 0.59600 | 0.73400 | 162 |
| HP_ABNORMAL_RIB_CAGE_MORPHOLOGY | 10 | 0.31 | 0.894 | 0.59800 | 0.59800 | 0.73600 | 59 |
| GOCC_ENDOCYTIC_VESICLE | 15 | 0.27 | 0.885 | 0.60000 | 0.60000 | 0.73800 | 104 |
| HP_RETINAL_ATROPHY | 10 | -0.274 | -0.883 | 0.60100 | 0.60100 | 0.73800 | 300 |
| HP_ABNORMALITY_OF_FINGER | 54 | 0.196 | 0.899 | 0.60100 | 0.60100 | 0.73800 | 77 |
| HP_APLASIA_HYPOPLASIA_OF_FINGERS | 18 | 0.256 | 0.887 | 0.60300 | 0.60300 | 0.73800 | 148 |
| HP_JOINT_HYPERMOBILITY | 22 | 0.243 | 0.887 | 0.60300 | 0.60300 | 0.73800 | 59 |
| GOBP_MACROMOLECULE_DEACYLATION | 11 | -0.265 | -0.883 | 0.60400 | 0.60400 | 0.73800 | 315 |
| HP_PERIPHERAL_NEUROPATHY | 16 | -0.226 | -0.877 | 0.60500 | 0.60500 | 0.73800 | 251 |
| GOBP_MYELOID_CELL_DIFFERENTIATION | 14 | 0.274 | 0.879 | 0.60500 | 0.60500 | 0.73800 | 148 |
| GOMF_ENZYME_INHIBITOR_ACTIVITY | 13 | 0.281 | 0.883 | 0.60500 | 0.60500 | 0.73800 | 126 |
| GOBP_REPRODUCTIVE_SYSTEM_DEVELOPMENT | 14 | 0.274 | 0.878 | 0.60600 | 0.60600 | 0.73800 | 148 |
| GOBP_PROTEASOME_MEDIATED_UBIQUITIN_DEPENDENT_PROTEIN_CATABOLIC_ | 22 | -0.204 | -0.884 | 0.60800 | 0.60800 | 0.73800 | 573 |
| GOCC_NUCLEOLUS | 51 | -0.159 | -0.907 | 0.60800 | 0.60800 | 0.73800 | 250 |
| HP_RESPIRATORY_FAILURE | 12 | 0.288 | 0.879 | 0.60900 | 0.60900 | 0.73800 | 3 |
| HP_WIDE_NASAL_BRIDGE | 25 | -0.194 | -0.882 | 0.61000 | 0.61000 | 0.73800 | 215 |
| GOCC_VESICLE_LUMEN | 10 | -0.272 | -0.875 | 0.61000 | 0.61000 | 0.73800 | 25 |
| GOBP_CHROMATIN_REMODELING | 22 | 0.241 | 0.882 | 0.61000 | 0.61000 | 0.73800 | 83 |
| HP_ABNORMALITY_OF_THE_CERVICAL_SPINE | 20 | -0.21 | -0.879 | 0.61100 | 0.61100 | 0.73800 | 215 |
| HP_FATIGUE | 16 | 0.262 | 0.875 | 0.61300 | 0.61300 | 0.73900 | 218 |
| HP_ABNORMAL_PERIPHERAL_NERVOUS_SYSTEM_MORPHOLOGY | 17 | -0.219 | -0.869 | 0.61300 | 0.61300 | 0.73900 | 214 |
| HP_DEPRESSED_NASAL_BRIDGE | 30 | 0.222 | 0.887 | 0.61400 | 0.61400 | 0.73900 | 77 |
| GOBP_IMMUNE_SYSTEM_DEVELOPMENT | 35 | -0.177 | -0.897 | 0.61500 | 0.61500 | 0.74000 | 90 |
| HP_ABNORMAL_NEURAL_TUBE_MORPHOLOGY | 10 | -0.271 | -0.873 | 0.61600 | 0.61600 | 0.74100 | 261 |
| HP_ABNORMAL_CELLULAR_PHENOTYPE | 14 | 0.27 | 0.868 | 0.61900 | 0.61900 | 0.74300 | 28 |
| HP_ABNORMAL_HAIR_MORPHOLOGY | 49 | -0.161 | -0.9 | 0.62100 | 0.62100 | 0.74500 | 190 |
| HP_SHORT_ATTENTION_SPAN | 25 | -0.192 | -0.874 | 0.62200 | 0.62200 | 0.74500 | 300 |
| HP_INABILITY_TO_WALK | 10 | -0.269 | -0.866 | 0.62400 | 0.62400 | 0.74700 | 525 |
| HP_KYPHOSIS | 21 | -0.204 | -0.867 | 0.62700 | 0.62700 | 0.74900 | 171 |
| GOBP_PROTEIN_LOCALIZATION_TO_ORGANELLE | 36 | -0.174 | -0.891 | 0.63200 | 0.63200 | 0.75500 | 537 |
| GOBP_TRNA_PROCESSING | 12 | -0.248 | -0.86 | 0.63300 | 0.63300 | 0.75500 | 540 |
| GOBP_DEPHOSPHORYLATION | 13 | 0.273 | 0.857 | 0.63400 | 0.63400 | 0.75600 | 132 |
| HP_ABNORMALITY_OF_THE_CALF | 16 | -0.219 | -0.849 | 0.64000 | 0.64000 | 0.76100 | 176 |
| HP_APLASIA_HYPOPLASIA_OF_THE_PHALANGES_OF_THE_HAND | 12 | 0.279 | 0.854 | 0.64000 | 0.64000 | 0.76100 | 59 |
| GOBP_FAT_CELL_DIFFERENTIATION | 10 | 0.297 | 0.857 | 0.64100 | 0.64100 | 0.76100 | 68 |
| HP_HYPERMETROPIA | 16 | -0.218 | -0.845 | 0.64700 | 0.64700 | 0.76500 | 207 |
| GOBP_NEGATIVE_REGULATION_OF_CELLULAR_MACROMOLECULE_BIOSYNTHETI( | 16 | -0.218 | -0.845 | 0.64700 | 0.64700 | 0.76500 | 208 |
| GOBP_DEFENSE_RESPONSE_TO_SYMBIONT | 13 | 0.27 | 0.848 | 0.64700 | 0.64700 | 0.76500 | 48 |
| GOMF_MOLECULAR_ADAPTOR_ACTIVITY | 21 | 0.237 | 0.855 | 0.64700 | 0.64700 | 0.76500 | 235 |
| HP_PROMINENT_NASAL_BRIDGE | 14 | -0.23 | -0.848 | 0.64900 | 0.64900 | 0.76500 | 215 |
| HP_ABNORMALITY_OF_REPRODUCTIVE_SYSTEM_PHYSIOLOGY | 31 | 0.214 | 0.862 | 0.64900 | 0.64900 | 0.76500 | 26 |
| GOBP_CAMERA_TYPE_EYE_DEVELOPMENT | 16 | 0.254 | 0.848 | 0.64900 | 0.64900 | 0.76500 | 74 |

| | | | | | | | |
|---|---|---|---|---|---|---|---|
| HP_ABNORMAL_STERNUM_MORPHOLOGY | 19 | 0.242 | 0.852 | 0.65000 | 0.65000 | 0.76500 | 59 |
| GOBP_NEGATIVE_REGULATION_OF_TRANSFERASE_ACTIVITY | 12 | 0.276 | 0.844 | 0.65100 | 0.65100 | 0.76500 | 140 |
| GOBP_T_CELL_ACTIVATION | 11 | -0.255 | -0.848 | 0.65100 | 0.65100 | 0.76500 | 62 |
| HP_ABNORMALITY_OF_THE_VOICE | 22 | -0.197 | -0.851 | 0.65300 | 0.65300 | 0.76500 | 176 |
| GOMF_PROTEIN_CONTAINING_COMPLEX_BINDING | 56 | 0.186 | 0.86 | 0.65300 | 0.65300 | 0.76500 | 91 |
| HP_ABNORMAL_URINE_METABOLITE_LEVEL | 14 | 0.262 | 0.841 | 0.65400 | 0.65400 | 0.76500 | 203 |
| GOCC_EXTRINSIC_COMPONENT_OF_MEMBRANE | 16 | 0.253 | 0.844 | 0.65400 | 0.65400 | 0.76500 | 120 |
| GOBP_VESICLE_ORGANIZATION | 18 | -0.209 | -0.845 | 0.65600 | 0.65600 | 0.76600 | 394 |
| HP_PROPTOSIS | 11 | 0.281 | 0.838 | 0.65700 | 0.65700 | 0.76600 | 71 |
| HP_CONSTITUTIONAL_SYMPTOM | 35 | 0.206 | 0.856 | 0.65700 | 0.65700 | 0.76600 | 218 |
| GOBP_CELLULAR_MACROMOLECULE_LOCALIZATION | 82 | -0.14 | -0.896 | 0.65800 | 0.65800 | 0.76600 | 285 |
| GOCC_MIDBODY | 10 | 0.291 | 0.838 | 0.66200 | 0.66200 | 0.77000 | 60 |
| GOBP_REGULATION_OF_ORGANELLE_ORGANIZATION | 59 | 0.184 | 0.861 | 0.66300 | 0.66300 | 0.77000 | 149 |
| HP_ABNORMALITY_OF_THE_MENSTRUAL_CYCLE | 14 | -0.227 | -0.837 | 0.66300 | 0.66300 | 0.77000 | 165 |
| HP_ABNORMALITY_OF_THE_KNEE | 17 | -0.21 | -0.835 | 0.66400 | 0.66400 | 0.77000 | 231 |
| GOBP_CHROMOSOME_ORGANIZATION | 60 | -0.15 | -0.882 | 0.66600 | 0.66600 | 0.77100 | 266 |
| GOBP_PROTEIN_FOLDING | 11 | -0.251 | -0.835 | 0.66700 | 0.66700 | 0.77100 | 436 |
| GOBP_LEUKOCYTE_MEDIATED_IMMUNITY | 10 | 0.289 | 0.834 | 0.66800 | 0.66800 | 0.77100 | 28 |
| HP_ABNORMAL_INVOLUNTARY_EYE_MOVEMENTS | 45 | -0.156 | -0.859 | 0.66900 | 0.66900 | 0.77100 | 194 |
| HP_DELAYED_MYELINATION | 10 | -0.258 | -0.831 | 0.67000 | 0.67000 | 0.77100 | 231 |
| GOMF_HYDROLASE_ACTIVITY_ACTING_ON_ESTER_BONDS | 21 | 0.232 | 0.838 | 0.67200 | 0.67200 | 0.77100 | 268 |
| HP_ABNORMALITY_OF_THE_FOREHEAD | 52 | 0.187 | 0.853 | 0.67200 | 0.67200 | 0.77100 | 77 |
| HP_WIDE_MOUTH | 13 | -0.235 | -0.837 | 0.67200 | 0.67200 | 0.77100 | 151 |
| GOBP_SENSORY_SYSTEM_DEVELOPMENT | 17 | 0.243 | 0.826 | 0.67200 | 0.67200 | 0.77100 | 74 |
| GOBP_MICROTUBULE_BASED_PROCESS | 35 | 0.203 | 0.844 | 0.67300 | 0.67300 | 0.77100 | 142 |
| GOMF_PROTEIN_MACROMOLECULE_ADAPTOR_ACTIVITY | 16 | 0.248 | 0.829 | 0.67300 | 0.67300 | 0.77100 | 235 |
| HP_FUNCTIONAL_ABNORMALITY_OF_MALE_INTERNAL_GENITALIA | 10 | 0.287 | 0.829 | 0.67500 | 0.67500 | 0.77100 | 3 |
| HP_TOE_SYNDACTYLY | 10 | -0.257 | -0.827 | 0.67600 | 0.67600 | 0.77100 | 207 |
| HP_ABNORMAL_INTESTINE_MORPHOLOGY | 38 | 0.2 | 0.847 | 0.67700 | 0.67700 | 0.77100 | 77 |
| HP_BOWING_OF_THE_LEGS | 14 | 0.257 | 0.824 | 0.67700 | 0.67700 | 0.77100 | 260 |
| HP_ABNORMAL_HAND_MORPHOLOGY | 19 | 0.236 | 0.83 | 0.67700 | 0.67700 | 0.77100 | 148 |
| HP_ABNORMAL_STOMACH_MORPHOLOGY | 12 | -0.239 | -0.828 | 0.67700 | 0.67700 | 0.77100 | 165 |
| HP_ABNORMAL_SCALP_MORPHOLOGY | 24 | 0.222 | 0.832 | 0.67800 | 0.67800 | 0.77100 | 14 |
| HP_POLYHYDRAMNIOS | 13 | 0.261 | 0.819 | 0.67800 | 0.67800 | 0.77100 | 59 |
| GOBP_TRNA_MODIFICATION | 11 | -0.248 | -0.825 | 0.67900 | 0.67900 | 0.77100 | 540 |
| HP_RETROGNATHIA | 23 | 0.225 | 0.83 | 0.67900 | 0.67900 | 0.77100 | 26 |
| HP_INFERTILITY | 10 | 0.286 | 0.825 | 0.68000 | 0.68000 | 0.77100 | 3 |
| HP_ABNORMALITY_OF_THE_SPLEEN | 27 | 0.214 | 0.83 | 0.68800 | 0.68800 | 0.77900 | 188 |
| GOCC_NUCLEAR_OUTER_MEMBRANE_ENDOPLASMIC_RETICULUM_MEMBRANE_ | 61 | -0.147 | -0.868 | 0.68800 | 0.68800 | 0.77900 | 104 |
| HP_MYOPIA | 19 | -0.2 | -0.821 | 0.69000 | 0.69000 | 0.78000 | 165 |
| HP_ABNORMALITY_OF_THE_NECK | 29 | -0.176 | -0.837 | 0.69100 | 0.69100 | 0.78000 | 215 |
| GOBP_NEGATIVE_REGULATION_OF_MOLECULAR_FUNCTION | 38 | 0.197 | 0.834 | 0.69200 | 0.69200 | 0.78000 | 140 |
| GOBP_REGULATION_OF_DNA_BIOSYNTHETIC_PROCESS | 12 | -0.236 | -0.819 | 0.69200 | 0.69200 | 0.78000 | 196 |

| | | | | | | | |
|---|---|---|---|---|---|---|---|
| HP_ABNORMALITY_OF_THE_ABDOMINAL_WALL | 21 | 0.227 | 0.82 | 0.69300 | 0.69300 | 0.78000 | 162 |
| HP_ABNORMALITY_OF_THE_MAXILLA | 10 | 0.282 | 0.814 | 0.69300 | 0.69300 | 0.78000 | 59 |
| GOBP_RESPIRATORY_SYSTEM_DEVELOPMENT | 15 | 0.249 | 0.814 | 0.69400 | 0.69400 | 0.78000 | 77 |
| HP_REGIONAL_ABNORMALITY_OF_SKIN | 27 | 0.212 | 0.824 | 0.69600 | 0.69600 | 0.78000 | 14 |
| HP_LIMB_UNDERGROWTH | 11 | 0.271 | 0.806 | 0.69600 | 0.69600 | 0.78000 | 134 |
| GOBP_PROTEASOMAL_PROTEIN_CATABOLIC_PROCESS | 27 | 0.212 | 0.823 | 0.69700 | 0.69700 | 0.78000 | 15 |
| HP_ABNORMAL_RENAL_PHYSIOLOGY | 35 | -0.164 | -0.832 | 0.69900 | 0.69900 | 0.78000 | 165 |
| HP_MUSCLE_WEAKNESS | 35 | -0.164 | -0.832 | 0.69900 | 0.69900 | 0.78000 | 144 |
| GOMF_HYDROLASE_ACTIVITY_ACTING_ON_ACID_ANHYDRIDES | 26 | 0.213 | 0.817 | 0.69900 | 0.69900 | 0.78000 | 45 |
| GOMF_PHOSPHORIC_ESTER_HYDROLASE_ACTIVITY | 11 | 0.269 | 0.802 | 0.70000 | 0.70000 | 0.78000 | 149 |
| GOCC_MOTILE_CILIUM | 10 | 0.281 | 0.809 | 0.70000 | 0.70000 | 0.78000 | 169 |
| HP_RESPIRATORY_INSUFFICIENCY | 21 | 0.225 | 0.813 | 0.70100 | 0.70100 | 0.78000 | 210 |
| HP_ABNORMALITY_OF_THE_ADRENAL_GLANDS | 10 | 0.28 | 0.809 | 0.70100 | 0.70100 | 0.78000 | 71 |
| HP_CONGENITAL_MALFORMATION_OF_THE_GREAT_ARTERIES | 27 | 0.211 | 0.819 | 0.70400 | 0.70400 | 0.78200 | 14 |
| HP_ABNORMAL_CARDIAC_SEPTUM_MORPHOLOGY | 30 | 0.206 | 0.82 | 0.70600 | 0.70600 | 0.78400 | 77 |
| HP_APLASIA_HYPOPLASIA_AFFECTING_THE_EYE | 14 | 0.25 | 0.801 | 0.70700 | 0.70700 | 0.78400 | 266 |
| HP_ABNORMAL_CIRCULATING_METABOLITE_CONCENTRATION | 39 | -0.159 | -0.833 | 0.70800 | 0.70800 | 0.78400 | 127 |
| HP_ABNORMAL_NASAL_BRIDGE_MORPHOLOGY | 49 | -0.151 | -0.848 | 0.70900 | 0.70900 | 0.78400 | 215 |
| HP_ABNORMALITY_IRIS_MORPHOLOGY | 14 | -0.218 | -0.803 | 0.71000 | 0.71000 | 0.78400 | 213 |
| HP_ABNORMALITY_OF_THE_KIDNEY | 56 | 0.177 | 0.822 | 0.71100 | 0.71100 | 0.78400 | 210 |
| GOCC_LIPID_DROPLET | 10 | 0.278 | 0.801 | 0.71200 | 0.71200 | 0.78400 | 229 |
| HP_ABNORMAL_MALE_REPRODUCTIVE_SYSTEM_PHYSIOLOGY | 11 | 0.266 | 0.793 | 0.71200 | 0.71200 | 0.78400 | 3 |
| HP_PAIN | 31 | 0.203 | 0.815 | 0.71200 | 0.71200 | 0.78400 | 266 |
| HP_DECREASED_FERTILITY | 11 | 0.266 | 0.792 | 0.71300 | 0.71300 | 0.78400 | 3 |
| HP_ABNORMAL_CEREBRAL_CORTEX_MORPHOLOGY | 31 | -0.166 | -0.813 | 0.71300 | 0.71300 | 0.78400 | 190 |
| GOMF_GUANYL_NUCLEOTIDE_EXCHANGE_FACTOR_ACTIVITY | 10 | -0.247 | -0.794 | 0.71500 | 0.71500 | 0.78500 | 6 |
| HP_APNEA | 16 | 0.238 | 0.793 | 0.71700 | 0.71700 | 0.78500 | 2 |
| HP_CONSTIPATION | 18 | 0.229 | 0.794 | 0.71800 | 0.71800 | 0.78500 | 77 |
| HP_HYPERTELORISM | 41 | 0.189 | 0.814 | 0.71800 | 0.71800 | 0.78500 | 77 |
| HP_ABNORMALITY_OF_THE_LARYNX | 15 | 0.242 | 0.793 | 0.71800 | 0.71800 | 0.78500 | 59 |
| GOBP_PROTEOLYSIS | 57 | 0.176 | 0.817 | 0.71900 | 0.71900 | 0.78500 | 25 |
| GOBP_ENDOCYTOSIS | 21 | -0.187 | -0.796 | 0.72000 | 0.72000 | 0.78500 | 161 |
| HP_ABNORMALITY_OF_SKELETAL_MATURATION | 23 | 0.215 | 0.795 | 0.72000 | 0.72000 | 0.78500 | 218 |
| HP_PECTUS_EXCAVATUM | 14 | 0.246 | 0.788 | 0.72300 | 0.72300 | 0.78800 | 59 |
| HP_HYPOGONADISM | 14 | 0.245 | 0.785 | 0.72700 | 0.72700 | 0.79100 | 94 |
| HP_ABNORMAL_CIRCULATING_HORMONE_CONCENTRATION | 18 | 0.227 | 0.784 | 0.72800 | 0.72800 | 0.79200 | 71 |
| HP_CHRONIC_KIDNEY_DISEASE | 10 | 0.271 | 0.781 | 0.73300 | 0.73300 | 0.79600 | 162 |
| HP_SKELETAL_MUSCLE_ATROPHY | 21 | -0.184 | -0.784 | 0.73300 | 0.73300 | 0.79600 | 527 |
| HP_ABNORMAL_DIAPHYSIS_MORPHOLOGY | 19 | 0.222 | 0.781 | 0.73500 | 0.73500 | 0.79600 | 286 |
| HP_ABNORMAL_FEMALE_REPRODUCTIVE_SYSTEM_PHYSIOLOGY | 15 | -0.208 | -0.785 | 0.73500 | 0.73500 | 0.79600 | 165 |
| HP_ABNORMALITY_OF_REFRACTION | 33 | -0.162 | -0.806 | 0.73600 | 0.73600 | 0.79600 | 183 |
| HP_TALIPES_EQUINOVARUS | 11 | 0.259 | 0.773 | 0.73600 | 0.73600 | 0.79600 | 59 |
| HP_RETINAL_DYSTROPHY | 16 | 0.232 | 0.776 | 0.73700 | 0.73700 | 0.79600 | 3 |

| | | | | | | | |
|---|---|---|---|---|---|---|---|
| HP_ABNORMAL_LOWER_LIMB_BONE_MORPHOLOGY | 43 | -0.15 | -0.812 | 0.73700 | 0.73700 | 0.79600 | 215 |
| GOBP_PROTEIN_POLYUBIQUITINATION | 12 | -0.225 | -0.78 | 0.73900 | 0.73900 | 0.79600 | 382 |
| HP_HIGHLY_ARCHED_EYEBROW | 13 | -0.221 | -0.788 | 0.73900 | 0.73900 | 0.79600 | 559 |
| HP_HYPOPLASIA_OF_PENIS | 19 | -0.191 | -0.784 | 0.74200 | 0.74200 | 0.79700 | 213 |
| HP_ABNORMALITY_OF_TOE | 35 | 0.189 | 0.787 | 0.74200 | 0.74200 | 0.79700 | 162 |
| HP_ABNORMALITY_OF_THE_NAIL | 19 | 0.22 | 0.773 | 0.74300 | 0.74300 | 0.79700 | 71 |
| GOMF_DNA_BINDING_TRANSCRIPTION_ACTIVATOR_ACTIVITY | 19 | 0.22 | 0.772 | 0.74400 | 0.74400 | 0.79700 | 249 |
| HP_ABNORMAL_CIRCULATING_LIPID_CONCENTRATION | 14 | -0.211 | -0.777 | 0.74400 | 0.74400 | 0.79700 | 149 |
| HP_OSTEOPOROSIS | 11 | -0.234 | -0.778 | 0.74500 | 0.74500 | 0.79700 | 183 |
| GOCC_GOLGI_APPARATUS | 68 | 0.168 | 0.808 | 0.74500 | 0.74500 | 0.79700 | 178 |
| GOBP_POSITIVE_REGULATION_OF_ESTABLISHMENT_OF_PROTEIN_LOCALIZATION | 16 | 0.231 | 0.77 | 0.74600 | 0.74600 | 0.79800 | 112 |
| HP_ATTENTION_DEFICIT_HYPERACTIVITY_DISORDER | 21 | 0.215 | 0.776 | 0.74700 | 0.74700 | 0.79800 | 94 |
| GOCC_CENTROSOME | 33 | 0.191 | 0.781 | 0.74800 | 0.74800 | 0.79800 | 235 |
| GOCC_TRANSPORTER_COMPLEX | 10 | 0.266 | 0.767 | 0.74900 | 0.74900 | 0.79800 | 107 |
| GOCC_DENDRITIC_TREE | 18 | 0.221 | 0.765 | 0.74900 | 0.74900 | 0.79800 | 141 |
| HP_ABNORMALITY_OF_THE_OUTER_EAR | 52 | 0.174 | 0.792 | 0.75000 | 0.75000 | 0.79800 | 77 |
| HP_ABNORMAL_FINGER_PHALANX_MORPHOLOGY | 36 | 0.187 | 0.785 | 0.75100 | 0.75100 | 0.79800 | 162 |
| HP_APRAXIA | 12 | -0.221 | -0.766 | 0.75400 | 0.75400 | 0.80100 | 559 |
| HP_GENERALIZED_HYPOTONIA | 45 | -0.146 | -0.8 | 0.75500 | 0.75500 | 0.80100 | 120 |
| HP_ABNORMALITY_OF_LOWER_LIP | 10 | 0.264 | 0.76 | 0.75700 | 0.75700 | 0.80200 | 14 |
| HP_ABNORMAL_THUMB_MORPHOLOGY | 22 | 0.21 | 0.766 | 0.75900 | 0.75900 | 0.80300 | 162 |
| GOBP_REGULATION_OF_SUPRAMOLECULAR_FIBER_ORGANIZATION | 13 | 0.24 | 0.753 | 0.75900 | 0.75900 | 0.80300 | 104 |
| HP_ABNORMAL_MORPHOLOGY_OF_THE_GREAT_VESSELS | 36 | 0.185 | 0.775 | 0.76200 | 0.76200 | 0.80500 | 156 |
| GOCC_VACUOLAR_MEMBRANE | 23 | -0.174 | -0.765 | 0.76300 | 0.76300 | 0.80600 | 184 |
| HP_ABNORMAL_PALATE_MORPHOLOGY | 53 | 0.17 | 0.78 | 0.76500 | 0.76500 | 0.80600 | 77 |
| GOBP_GENERATION_OF_PRECURSOR_METABOLITES_AND_ENERGY | 18 | -0.19 | -0.768 | 0.76500 | 0.76500 | 0.80600 | 70 |
| GOBP_HISTONE_DEACETYLATION | 10 | -0.233 | -0.749 | 0.76700 | 0.76700 | 0.80600 | 315 |
| HP_ABNORMAL_FORM_OF_THE_VERTEBRAL_BODIES | 16 | -0.197 | -0.762 | 0.76800 | 0.76800 | 0.80600 | 99 |
| HP_AORTIC_ANEURYSM | 10 | 0.259 | 0.748 | 0.76900 | 0.76900 | 0.80600 | 205 |
| GOBP_VACUOLE_ORGANIZATION | 10 | -0.232 | -0.747 | 0.76900 | 0.76900 | 0.80600 | 45 |
| GOBP_POSITIVE_REGULATION_OF_PROTEOLYSIS | 10 | 0.259 | 0.748 | 0.76900 | 0.76900 | 0.80600 | 13 |
| GOBP_CELL_CYCLE_G2_M_PHASE_TRANSITION | 13 | -0.215 | -0.764 | 0.77000 | 0.77000 | 0.80600 | 464 |
| HP_ABNORMAL_VASCULAR_MORPHOLOGY | 41 | 0.179 | 0.772 | 0.77000 | 0.77000 | 0.80600 | 162 |
| HP_ABNORMAL_CORTICAL_GYRATION | 17 | -0.193 | -0.765 | 0.77100 | 0.77100 | 0.80600 | 144 |
| GOBP_PEPTIDYL_AMINO_ACID_MODIFICATION | 59 | -0.139 | -0.814 | 0.77200 | 0.77200 | 0.80600 | 248 |
| GOBP_DNA_PACKAGING | 17 | 0.22 | 0.746 | 0.77200 | 0.77200 | 0.80600 | 83 |
| HP_BLINDNESS | 10 | -0.231 | -0.743 | 0.77300 | 0.77300 | 0.80600 | 207 |
| HP_ABNORMALITY_OF_ACID_BASE_HOMEOSTASIS | 14 | 0.233 | 0.747 | 0.77400 | 0.77400 | 0.80600 | 148 |
| HP_ABNORMAL_JAW_MORPHOLOGY | 54 | 0.168 | 0.774 | 0.77400 | 0.77400 | 0.80600 | 71 |
| HP_VASCULAR_DILATATION | 13 | 0.235 | 0.739 | 0.77500 | 0.77500 | 0.80600 | 205 |
| GOBP_LYMPHOCYTE_ACTIVATION | 21 | 0.207 | 0.748 | 0.77700 | 0.77700 | 0.80700 | 218 |
| GOBP_ENERGY_DERIVATION_BY_OXIDATION_OF_ORGANIC_COMPOUNDS | 12 | -0.214 | -0.741 | 0.77800 | 0.77800 | 0.80800 | 70 |
| HP_ABNORMALITY_OF_CRANIAL_SUTURES | 22 | -0.174 | -0.756 | 0.78100 | 0.78100 | 0.81000 | 238 |

| | | | | | | | |
|---|---|---|---|---|---|---|---|
| GOMF_PROTEIN_DOMAIN_SPECIFIC_BINDING | 32 | 0.186 | 0.755 | 0.78300 | 0.78300 | 0.81100 | 133 |
| GOBP_NEGATIVE_REGULATION_OF_ORGANELLE_ORGANIZATION | 18 | -0.187 | -0.754 | 0.78400 | 0.78400 | 0.81100 | 285 |
| HP_LOWER_EXTREMITY_JOINT_DISLOCATION | 17 | -0.191 | -0.758 | 0.78400 | 0.78400 | 0.81100 | 231 |
| GOCC_ORGANELLE_SUBCOMPARTMENT | 77 | -0.129 | -0.81 | 0.78500 | 0.78500 | 0.81100 | 105 |
| GOBP_ESTABLISHMENT_OF_PROTEIN_LOCALIZATION_TO_ORGANELLE | 16 | -0.193 | -0.748 | 0.78600 | 0.78600 | 0.81100 | 355 |
| GOCC_MICROTUBULE_ORGANIZING_CENTER | 40 | 0.175 | 0.754 | 0.78700 | 0.78700 | 0.81100 | 235 |
| GOBP_NEGATIVE_REGULATION_OF_TRANSCRIPTION_BY_RNA_POLYMERASE_II | 51 | 0.167 | 0.761 | 0.78900 | 0.78900 | 0.81300 | 116 |
| HP_ABNORMALITY_OF_GLOBE_SIZE | 11 | 0.245 | 0.729 | 0.79300 | 0.79300 | 0.81500 | 266 |
| GOBP_CILIUM_ORGANIZATION | 24 | 0.195 | 0.732 | 0.79300 | 0.79300 | 0.81500 | 345 |
| HP_ABNORMALITY_OF_THE_THYROID_GLAND | 16 | 0.219 | 0.733 | 0.79400 | 0.79400 | 0.81500 | 218 |
| HP_SHORT_NOSE | 20 | -0.178 | -0.746 | 0.79500 | 0.79500 | 0.81600 | 165 |
| GOBP_REGULATION_OF_CELL_CYCLE_G2_M_PHASE_TRANSITION | 12 | -0.21 | -0.727 | 0.79600 | 0.79600 | 0.81600 | 62 |
| HP_REDUCED_VISUAL_ACUITY | 14 | -0.2 | -0.735 | 0.80100 | 0.80100 | 0.82000 | 207 |
| GOCC_MITOCHONDRIAL_MATRIX | 15 | -0.196 | -0.739 | 0.80200 | 0.80200 | 0.82000 | 101 |
| HP_RENAL_CYST | 16 | 0.217 | 0.725 | 0.80200 | 0.80200 | 0.82000 | 360 |
| HP_ABNORMAL_MYELINATION | 21 | -0.173 | -0.734 | 0.80300 | 0.80300 | 0.82000 | 239 |
| HP_EXTERNAL_GENITAL_HYPOPLASIA | 20 | -0.176 | -0.739 | 0.80500 | 0.80500 | 0.82200 | 285 |
| HP_ABNORMAL_RENAL_CORTEX_MORPHOLOGY | 10 | 0.247 | 0.711 | 0.80600 | 0.80600 | 0.82200 | 178 |
| GOBP_DNA_BIOSYNTHETIC_PROCESS | 13 | -0.207 | -0.736 | 0.80800 | 0.80800 | 0.82300 | 196 |
| HP_OSTEOPENIA | 12 | 0.232 | 0.709 | 0.80900 | 0.80900 | 0.82300 | 255 |
| GOBP_POSITIVE_REGULATION_OF_PROTEIN_LOCALIZATION | 23 | -0.166 | -0.728 | 0.80900 | 0.80900 | 0.82300 | 132 |
| GOBP_SUPRAMOLECULAR_FIBER_ORGANIZATION | 19 | 0.202 | 0.712 | 0.81000 | 0.81000 | 0.82300 | 104 |
| HP_APLASIA_HYPOPLASIA_INVOLVING_BONES_OF_THE_FEET | 17 | 0.209 | 0.709 | 0.81500 | 0.81500 | 0.82700 | 59 |
| HP_ABNORMALITY_OF_THE_MIDFACE | 31 | 0.181 | 0.726 | 0.81800 | 0.81800 | 0.82800 | 71 |
| HP_ABNORMALITY_OF_THE_ANUS | 10 | 0.244 | 0.703 | 0.81800 | 0.81800 | 0.82800 | 14 |
| HP_MOTOR_DELAY | 30 | 0.182 | 0.725 | 0.81900 | 0.81900 | 0.82800 | 99 |
| HP_ABNORMALITY_OF_THE_FEMALE_GENITALIA | 31 | 0.18 | 0.725 | 0.81900 | 0.81900 | 0.82800 | 77 |
| HP_ABNORMAL_RENAL_MORPHOLOGY | 40 | 0.168 | 0.723 | 0.82000 | 0.82000 | 0.82900 | 178 |
| HP_ENLARGED_POSTERIOR_FOSSA | 15 | 0.214 | 0.702 | 0.82700 | 0.82700 | 0.83500 | 87 |
| GOBP_POSITIVE_REGULATION_OF_ORGANELLE_ORGANIZATION | 26 | 0.182 | 0.699 | 0.82800 | 0.82800 | 0.83500 | 20 |
| GOMF_PROTEIN_HETERODIMERIZATION_ACTIVITY | 12 | 0.226 | 0.693 | 0.82800 | 0.82800 | 0.83500 | 103 |
| HP_ABNORMAL_SYSTEMIC_ARTERIAL_MORPHOLOGY | 23 | -0.161 | -0.708 | 0.83100 | 0.83100 | 0.83700 | 300 |
| HP_ABNORMAL_NIPPLE_MORPHOLOGY | 11 | 0.231 | 0.687 | 0.83300 | 0.83300 | 0.83800 | 14 |
| GOBP_VIRAL_LIFE_CYCLE | 17 | -0.181 | -0.718 | 0.83400 | 0.83400 | 0.83800 | 204 |
| HP_THICKENED_SKIN | 13 | -0.201 | -0.713 | 0.83500 | 0.83500 | 0.83800 | 194 |
| HP_LOW_POSTERIOR_HAIRLINE | 11 | 0.23 | 0.684 | 0.83500 | 0.83500 | 0.83800 | 14 |
| HP_LARGE_FACE | 15 | 0.212 | 0.693 | 0.83700 | 0.83700 | 0.83900 | 454 |
| HP_COLOBOMA | 11 | -0.209 | -0.697 | 0.83800 | 0.83800 | 0.83900 | 567 |
| HP_ABNORMALITY_OF_GLOBE_LOCATION | 50 | 0.158 | 0.716 | 0.83800 | 0.83800 | 0.83900 | 77 |
| HP_ABNORMALITY_OF_THE_FRONTAL_HAIRLINE | 14 | -0.19 | -0.7 | 0.84100 | 0.84100 | 0.84000 | 581 |
| HP_SMOOTH_PHILTRUM | 14 | -0.19 | -0.7 | 0.84100 | 0.84100 | 0.84000 | 581 |
| HP_ABNORMALITY_OF_THE_FONTANELLES_OR_CRANIAL_SUTURES | 23 | -0.159 | -0.698 | 0.84100 | 0.84100 | 0.84000 | 215 |
| GOBP_LEUKOCYTE_DIFFERENTIATION | 21 | 0.189 | 0.681 | 0.84200 | 0.84200 | 0.84000 | 218 |

| | | | | | | | |
|---|---|---|---|---|---|---|---|
| HP_APLASIA_HYPOPLASIA_INVOLVING_BONES_OF_THE_LOWER_LIMBS | 20 | 0.192 | 0.683 | 0.84300 | 0.84300 | 0.84000 | 162 |
| HP_ABNORMAL_VERTEBRAL_MORPHOLOGY | 20 | -0.167 | -0.7 | 0.84600 | 0.84600 | 0.84100 | 171 |
| HP_ABNORMAL_DERMATOGLYPHICS | 18 | -0.174 | -0.701 | 0.84600 | 0.84600 | 0.84100 | 381 |
| HP_DOWNSLANTED_PALPEBRAL_FISSURES | 27 | -0.149 | -0.694 | 0.84900 | 0.84900 | 0.84300 | 215 |
| GOBP_NUCLEAR_TRANSPORT | 14 | -0.188 | -0.691 | 0.85000 | 0.85000 | 0.84300 | 273 |
| GOMF_ACYLTRANSFERASE_ACTIVITY | 13 | 0.213 | 0.669 | 0.85000 | 0.85000 | 0.84300 | 564 |
| GOMF_ZINC_ION_BINDING | 33 | 0.168 | 0.688 | 0.85300 | 0.85300 | 0.84400 | 39 |
| HP_ABNORMALITY_OF_THE_AUTONOMIC_NERVOUS_SYSTEM | 11 | -0.204 | -0.679 | 0.85400 | 0.85400 | 0.84400 | 215 |
| GOBP_LYMPHOCYTE_ACTIVATION_INVOLVED_IN_IMMUNE_RESPONSE | 10 | 0.23 | 0.664 | 0.85400 | 0.85400 | 0.84400 | 218 |
| HP_ABNORMALITY_OF_THE_UTERUS | 12 | 0.217 | 0.664 | 0.85500 | 0.85500 | 0.84400 | 77 |
| HP_SYNDACTYLY | 20 | 0.188 | 0.671 | 0.85500 | 0.85500 | 0.84400 | 71 |
| GOBP_REGULATION_OF_CELL_DEVELOPMENT | 15 | 0.205 | 0.67 | 0.86000 | 0.86000 | 0.84800 | 77 |
| HP_ANORECTAL_ANOMALY | 15 | -0.181 | -0.684 | 0.86100 | 0.86100 | 0.84800 | 215 |
| HP_ABNORMALITY_OF_THE_URINARY_SYSTEM_PHYSIOLOGY | 43 | 0.158 | 0.688 | 0.86200 | 0.86200 | 0.84800 | 178 |
| GOCC_LATE_ENDOSOME | 12 | 0.214 | 0.656 | 0.86300 | 0.86300 | 0.84800 | 17 |
| HP_APLASIA_HYPOPLASIA_INVOLVING_BONES_OF_THE_UPPER_LIMBS | 25 | 0.176 | 0.667 | 0.86400 | 0.86400 | 0.84800 | 71 |
| HP_ABNORMAL_FEMORAL_NECK_HEAD_MORPHOLOGY | 12 | 0.213 | 0.65 | 0.86800 | 0.86800 | 0.84800 | 564 |
| GOBP_CELLULAR_PROTEIN_CATABOLIC_PROCESS | 36 | 0.161 | 0.675 | 0.86800 | 0.86800 | 0.84800 | 15 |
| GOCC_INTRACELLULAR_PROTEIN_CONTAINING_COMPLEX | 40 | -0.132 | -0.698 | 0.87000 | 0.87000 | 0.84800 | 434 |
| GOBP_NUCLEAR_EXPORT | 10 | -0.202 | -0.652 | 0.87000 | 0.87000 | 0.84800 | 312 |
| GOBP_NEGATIVE_REGULATION_OF_CELL_CYCLE | 16 | -0.174 | -0.673 | 0.87200 | 0.87200 | 0.84800 | 139 |
| HP_MICROPENIS | 10 | 0.222 | 0.642 | 0.87200 | 0.87200 | 0.84800 | 266 |
| GOBP_POSITIVE_REGULATION_OF_CELL_PROJECTION_ORGANIZATION | 15 | -0.178 | -0.672 | 0.87300 | 0.87300 | 0.84800 | 156 |
| HP_TRIANGULAR_SHAPED_PHALANGES_OF_THE_HAND | 12 | 0.211 | 0.646 | 0.87400 | 0.87400 | 0.84800 | 77 |
| GOBP_PURINE_CONTAINING_COMPOUND_METABOLIC_PROCESS | 13 | 0.205 | 0.643 | 0.87400 | 0.87400 | 0.84800 | 9 |
| GOBP_RIBOSE_PHOSPHATE_METABOLIC_PROCESS | 13 | 0.205 | 0.643 | 0.87400 | 0.87400 | 0.84800 | 9 |
| HP_SPARSE_HAIR | 16 | 0.197 | 0.658 | 0.87500 | 0.87500 | 0.84800 | 8 |
| HP_FEEDING_DIFFICULTIES | 51 | 0.15 | 0.681 | 0.87500 | 0.87500 | 0.84800 | 87 |
| GOBP_REGULATION_OF_AUTOPHAGY | 17 | -0.172 | -0.682 | 0.87500 | 0.87500 | 0.84800 | 247 |
| HP_ABNORMALITY_OF_COAGULATION | 14 | 0.201 | 0.645 | 0.87500 | 0.87500 | 0.84800 | 14 |
| GOBP_MODIFICATION_DEPENDENT_MACROMOLECULE_CATABOLIC_PROCESS | 31 | -0.137 | -0.67 | 0.87600 | 0.87600 | 0.84800 | 406 |
| HP_POSITIONAL_FOOT_DEFORMITY | 14 | 0.2 | 0.642 | 0.87800 | 0.87800 | 0.84800 | 162 |
| GOBP_NEGATIVE_REGULATION_OF_CELLULAR_CATABOLIC_PROCESS | 19 | -0.163 | -0.668 | 0.87800 | 0.87800 | 0.84800 | 90 |
| GOBP_EYE_MORPHOGENESIS | 10 | 0.22 | 0.634 | 0.87900 | 0.87900 | 0.84800 | 74 |
| HP_APLASIA_HYPOPLASIA_INVOLVING_THE_SKELETON | 59 | 0.145 | 0.678 | 0.87900 | 0.87900 | 0.84800 | 14 |
| HP_ABNORMAL_PATTERN_OF_RESPIRATION | 33 | 0.161 | 0.661 | 0.88000 | 0.88000 | 0.84800 | 27 |
| GOBP_CARBOHYDRATE_METABOLIC_PROCESS | 21 | -0.157 | -0.667 | 0.88000 | 0.88000 | 0.84800 | 108 |
| GOBP_ORGANELLE_ASSEMBLY | 45 | 0.153 | 0.673 | 0.88000 | 0.88000 | 0.84800 | 3 |
| GOCC_CONDENSED_CHROMOSOME | 10 | -0.199 | -0.641 | 0.88000 | 0.88000 | 0.84800 | 574 |
| GOBP_BIOLOGICAL_PROCESS_INVOLVED_IN_INTERACTION_WITH_HOST | 10 | 0.218 | 0.629 | 0.88400 | 0.88400 | 0.85000 | 203 |
| HP_ELEVATED_HEPATIC_TRANSAMINASE | 13 | 0.2 | 0.63 | 0.88500 | 0.88500 | 0.85100 | 360 |
| HP_ABNORMALITY_OF_RETINAL_PIGMENTATION | 14 | -0.177 | -0.651 | 0.88600 | 0.88600 | 0.85100 | 84 |
| HP_ABNORMAL_CNS_MYELINATION | 10 | -0.197 | -0.634 | 0.88900 | 0.88900 | 0.85300 | 53 |

| | | | | | | | |
|---|---|---|---|---|---|---|---|
| HP_ABNORMAL_BLEEDING | 19 | 0.179 | 0.629 | 0.89100 | 0.89100 | 0.85300 | 129 |
| GOCC_MICROTUBULE_CYTOSKELETON | 62 | 0.143 | 0.674 | 0.89100 | 0.89100 | 0.85300 | 349 |
| HP_ABNORMALITY_OF_THE_NASAL_DORSUM | 10 | 0.215 | 0.62 | 0.89100 | 0.89100 | 0.85300 | 266 |
| GOCC_GOLGI_MEMBRANE | 31 | 0.161 | 0.646 | 0.89200 | 0.89200 | 0.85300 | 239 |
| GOMF_UBIQUITIN_LIKE_PROTEIN_LIGASE_ACTIVITY | 13 | 0.198 | 0.621 | 0.89200 | 0.89200 | 0.85300 | 25 |
| HP_ABNORMALITY_OF_THE_SCROTUM | 11 | -0.192 | -0.639 | 0.89500 | 0.89500 | 0.85500 | 300 |
| GOCC_SOMATODENDRITIC_COMPARTMENT | 22 | 0.17 | 0.622 | 0.89900 | 0.89900 | 0.85700 | 141 |
| HP_CHOLESTASIS | 13 | 0.195 | 0.612 | 0.89900 | 0.89900 | 0.85700 | 151 |
| HP_ABNORMAL_FINGERTIP_MORPHOLOGY | 10 | 0.212 | 0.611 | 0.90000 | 0.90000 | 0.85700 | 564 |
| HP_ABNORMALITY_OF_THE_SKULL_BASE | 18 | 0.179 | 0.619 | 0.90200 | 0.90200 | 0.85800 | 162 |
| GOBP_NUCLEIC_ACID_PHOSPHODIESTER_BOND_HYDROLYSIS | 10 | -0.193 | -0.62 | 0.90400 | 0.90400 | 0.85900 | 93 |
| GOBP_REGULATION_OF_WNT_SIGNALING_PATHWAY | 12 | 0.199 | 0.61 | 0.90400 | 0.90400 | 0.85900 | 74 |
| HP_ABNORMALITY_OF_THE_BREAST | 13 | 0.193 | 0.606 | 0.90500 | 0.90500 | 0.85900 | 14 |
| GOMF_HISTONE_BINDING | 20 | 0.171 | 0.609 | 0.90900 | 0.90900 | 0.86200 | 46 |
| GOBP_REGULATION_OF_MITOCHONDRION_ORGANIZATION | 11 | 0.201 | 0.598 | 0.91000 | 0.91000 | 0.86200 | 202 |
| GOBP_PROTEIN_MODIFICATION_BY_SMALL_PROTEIN_CONJUGATION | 38 | -0.123 | -0.639 | 0.91100 | 0.91100 | 0.86200 | 437 |
| HP_INTELLECTUAL_DISABILITY_MILD | 22 | -0.146 | -0.632 | 0.91500 | 0.91500 | 0.86500 | 156 |
| GOBP_REGULATION_OF_NERVOUS_SYSTEM_DEVELOPMENT | 14 | -0.165 | -0.608 | 0.92000 | 0.92000 | 0.86800 | 181 |
| GOBP_MITOCHONDRION_ORGANIZATION | 26 | 0.156 | 0.598 | 0.92000 | 0.92000 | 0.86800 | 149 |
| HP_FEEDING_DIFFICULTIES_IN_INFANCY | 28 | 0.154 | 0.603 | 0.92000 | 0.92000 | 0.86800 | 71 |
| HP_ABNORMALITY_OF_THE_SYNOVIA | 16 | -0.159 | -0.614 | 0.92200 | 0.92200 | 0.86900 | 505 |
| HP_ABNORMALITY_OF_THE_MUSCULATURE_OF_THE_LIMBS | 11 | 0.196 | 0.585 | 0.92200 | 0.92200 | 0.86900 | 262 |
| GOBP_CHROMATIN_ORGANIZATION | 47 | 0.138 | 0.615 | 0.92300 | 0.92300 | 0.86900 | 83 |
| HP_ABNORMAL_ENZYME_COENZYME_ACTIVITY | 20 | -0.145 | -0.609 | 0.92500 | 0.92500 | 0.86900 | 72 |
| HP_APLASIA_HYPOPLASIA_OF_THE_EXTREMITIES | 31 | 0.148 | 0.596 | 0.92500 | 0.92500 | 0.86900 | 162 |
| GOCC_MITOCHONDRION | 58 | 0.134 | 0.627 | 0.92600 | 0.92600 | 0.86900 | 228 |
| GOMF_TRANSITION_METAL_ION_BINDING | 38 | 0.14 | 0.595 | 0.92800 | 0.92800 | 0.87100 | 138 |
| GOBP_PROTEIN_CATABOLIC_PROCESS | 41 | 0.138 | 0.597 | 0.93200 | 0.93200 | 0.87300 | 15 |
| GOBP_B_CELL_DIFFERENTIATION | 11 | 0.192 | 0.571 | 0.93200 | 0.93200 | 0.87300 | 218 |
| HP_ABNORMAL_CARDIAC_ATRIUM_MORPHOLOGY | 28 | 0.148 | 0.581 | 0.93300 | 0.93300 | 0.87300 | 77 |
| GOCC_CHROMOSOME_CENTROMERIC_REGION | 10 | 0.198 | 0.57 | 0.93500 | 0.93500 | 0.87400 | 574 |
| GOCC_TRANSFERASE_COMPLEX | 42 | -0.112 | -0.603 | 0.93600 | 0.93600 | 0.87500 | 465 |
| GOBP_NUCLEOBASE_CONTAINING_SMALL_MOLECULE_METABOLIC_PROCESS | 17 | 0.168 | 0.569 | 0.93700 | 0.93700 | 0.87500 | 9 |
| HP_APLASIA_HYPOPLASIA_AFFECTING_BONES_OF_THE_AXIAL_SKELETON | 50 | 0.132 | 0.599 | 0.93800 | 0.93800 | 0.87500 | 77 |
| GOBP_GLYCOSYLATION | 12 | 0.182 | 0.557 | 0.93900 | 0.93900 | 0.87500 | 250 |
| GOCC_NUCLEAR_CHROMOSOME | 16 | 0.166 | 0.556 | 0.94400 | 0.94400 | 0.87800 | 597 |
| GOMF_ISOMERASE_ACTIVITY | 10 | -0.178 | -0.572 | 0.94600 | 0.94600 | 0.87800 | 265 |
| GOCC_POLYMERIC_CYTOSKELETAL_FIBER | 17 | 0.163 | 0.554 | 0.94600 | 0.94600 | 0.87800 | 117 |
| HP_ABNORMALITY_OF_THE_PERIPHERAL_NERVOUS_SYSTEM | 16 | 0.165 | 0.551 | 0.94600 | 0.94600 | 0.87800 | 598 |
| HP_ABNORMAL_EYEBROW_MORPHOLOGY | 32 | -0.117 | -0.578 | 0.94600 | 0.94600 | 0.87800 | 176 |
| GOBP_NEGATIVE_REGULATION_OF_CELLULAR_COMPONENT_ORGANIZATION | 25 | 0.145 | 0.55 | 0.94900 | 0.94900 | 0.87900 | 132 |
| HP_ABNORMALITY_OF_LONG_BONE_MORPHOLOGY | 40 | 0.131 | 0.562 | 0.95000 | 0.95000 | 0.87900 | 162 |
| HP_CRYPTORCHIDISM | 38 | 0.131 | 0.558 | 0.95000 | 0.95000 | 0.87900 | 224 |

| | | | | | | | |
|---|---|---|---|---|---|---|---|
| GOBP_RESPONSE_TO_INSULIN | 10 | -0.175 | -0.563 | 0.95100 | 0.95100 | 0.87900 | 275 |
| HP_BROAD_NASAL_TIP | 11 | -0.169 | -0.562 | 0.95300 | 0.95300 | 0.88000 | 151 |
| HP_CEREBELLAR_MALFORMATION | 18 | 0.155 | 0.535 | 0.95300 | 0.95300 | 0.88000 | 335 |
| GOMF_KINASE_ACTIVATOR_ACTIVITY | 10 | 0.187 | 0.538 | 0.95300 | 0.95300 | 0.88000 | 245 |
| GOBP_NEGATIVE_REGULATION_OF_PROTEIN_MODIFICATION_PROCESS | 24 | 0.139 | 0.519 | 0.96400 | 0.96400 | 0.88800 | 126 |
| HP_INGUINAL_HERNIA | 16 | 0.153 | 0.512 | 0.96600 | 0.96600 | 0.88900 | 162 |
| HP_SOMATIC_SENSORY_DYSFUNCTION | 14 | 0.157 | 0.505 | 0.96800 | 0.96800 | 0.88900 | 293 |
| HP_ABNORMALITY_OF_EPIPHYSIS_MORPHOLOGY | 16 | 0.151 | 0.504 | 0.96800 | 0.96800 | 0.88900 | 71 |
| HP_ABNORMAL_CEREBELLAR_VERMIS_MORPHOLOGY | 19 | -0.132 | -0.542 | 0.97000 | 0.97000 | 0.88900 | 144 |
| HP_APLASIA_HYPOPLASIA_OF_THE_CEREBELLAR_VERMIS | 19 | -0.132 | -0.542 | 0.97000 | 0.97000 | 0.88900 | 144 |
| GOBP_NEGATIVE_REGULATION_OF_NUCLEOBASE_CONTAINING_COMPOUND_M | 80 | -0.093 | -0.589 | 0.97000 | 0.97000 | 0.88900 | 223 |
| GOBP_NEGATIVE_REGULATION_OF_BIOSYNTHETIC_PROCESS | 79 | 0.114 | 0.564 | 0.97100 | 0.97100 | 0.88900 | 90 |
| GOBP_PROTEIN_MODIFICATION_BY_SMALL_PROTEIN_CONJUGATION_OR_REMO | 41 | 0.121 | 0.52 | 0.97100 | 0.97100 | 0.88900 | 450 |
| GOBP_CELL_PROJECTION_ASSEMBLY | 30 | 0.126 | 0.503 | 0.97200 | 0.97200 | 0.89000 | 345 |
| HP_ABNORMALITY_OF_THE_HAIRLINE | 19 | 0.138 | 0.484 | 0.97500 | 0.97500 | 0.89100 | 14 |
| GOBP_B_CELL_ACTIVATION | 12 | 0.16 | 0.49 | 0.97600 | 0.97600 | 0.89200 | 218 |
| HP_ABNORMALITY_OF_THE_NASAL_TIP | 19 | 0.136 | 0.48 | 0.97700 | 0.97700 | 0.89200 | 59 |
| HP_REDUCED_TENDON_REFLEXES | 14 | 0.146 | 0.468 | 0.98200 | 0.98200 | 0.89200 | 611 |
| HP_ABNORMALITY_OF_THE_SPINAL_CORD | 10 | 0.164 | 0.472 | 0.98200 | 0.98200 | 0.89200 | 598 |
| GOMF_UBIQUITIN_LIKE_PROTEIN_TRANSFERASE_ACTIVITY | 21 | 0.127 | 0.46 | 0.98300 | 0.98300 | 0.89200 | 625 |
| HP_ABNORMALITY_OF_MUSCLE_SIZE | 25 | -0.11 | -0.5 | 0.98400 | 0.98400 | 0.89200 | 127 |
| HP_DEVELOPMENTAL_REGRESSION | 12 | -0.146 | -0.504 | 0.98400 | 0.98400 | 0.89200 | 153 |
| GOBP_MACROMOLECULE_CATABOLIC_PROCESS | 61 | 0.103 | 0.485 | 0.98500 | 0.98500 | 0.89200 | 74 |
| HP_HIP_DISLOCATION | 11 | -0.148 | -0.494 | 0.98500 | 0.98500 | 0.89200 | 165 |
| HP_HYPOREFLEXIA | 11 | -0.147 | -0.489 | 0.98600 | 0.98600 | 0.89200 | 214 |
| GOCC_CHROMOSOME_TELOMERIC_REGION | 12 | -0.144 | -0.5 | 0.98600 | 0.98600 | 0.89200 | 196 |
| HP_JOINT_LAXITY | 11 | 0.155 | 0.463 | 0.98700 | 0.98700 | 0.89200 | 604 |
| GOBP_REGULATION_OF_GENE_EXPRESSION_EPIGENETIC | 12 | -0.143 | -0.496 | 0.98700 | 0.98700 | 0.89200 | 315 |
| HP_ABNORMAL_RETINAL_MORPHOLOGY | 34 | 0.11 | 0.452 | 0.98800 | 0.98800 | 0.89200 | 3 |
| GOCC_CATALYTIC_COMPLEX | 74 | 0.097 | 0.476 | 0.98900 | 0.98900 | 0.89200 | 45 |
| GOBP_ORGANONITROGEN_COMPOUND_CATABOLIC_PROCESS | 51 | 0.1 | 0.455 | 0.99000 | 0.99000 | 0.89200 | 15 |
| GOMF_GTPASE_BINDING | 12 | 0.148 | 0.454 | 0.99000 | 0.99000 | 0.89200 | 462 |
| HP_ABNORMAL_HAIR_PATTERN | 23 | 0.116 | 0.429 | 0.99000 | 0.99000 | 0.89200 | 14 |
| GOBP_ENDOMEMBRANE_SYSTEM_ORGANIZATION | 26 | -0.1 | -0.461 | 0.99200 | 0.99200 | 0.89300 | 645 |
| GOBP_ATP_METABOLIC_PROCESS | 10 | -0.145 | -0.467 | 0.99400 | 0.99400 | 0.89300 | 70 |
| GOCC_CENTRIOLE | 10 | -0.144 | -0.463 | 0.99400 | 0.99400 | 0.89300 | 491 |
| HP_CENTRAL_NERVOUS_SYSTEM_CYST | 12 | 0.134 | 0.408 | 0.99700 | 0.99700 | 0.89300 | 335 |
| HP_CEREBELLAR_CYST | 12 | 0.134 | 0.408 | 0.99700 | 0.99700 | 0.89300 | 335 |
| HP_DILATED_FOURTH_VENTRICLE | 12 | 0.134 | 0.408 | 0.99700 | 0.99700 | 0.89300 | 335 |
| GOCC_MICROTUBULE | 14 | -0.113 | -0.416 | 0.99700 | 0.99700 | 0.89300 | 406 |
| GOBP_REGULATION_OF_NEUROGENESIS | 12 | -0.121 | -0.42 | 0.99800 | 0.99800 | 0.89300 | 509 |
| GOBP_PROTEIN_TARGETING | 13 | 0.111 | 0.348 | 0.99900 | 0.99900 | 0.89300 | 437 |
| HP_DEPRESSION | 12 | 0.119 | 0.365 | 0.99900 | 0.99900 | 0.89300 | 176 |

| | | | | | | | |
|---|---|---|---|---|---|---|---|
| HP_CRANIOSYNOSTOSIS | 11 | 0.106 | 0.316 | 1.00000 | 1.00000 | 0.89300 | 134 |

**Supplemental Table S14.** Significantly different proteins between *TP53* mutation compared to WT *TP53*.

| Gene | Protein ID | Mean Difference | Mean expression (*TP53* mutation) | Mean expression (WT *TP53*) | Statistic | p.value | Parameter | conf.low | conf.high | P.adjust |
|---|---|---|---|---|---|---|---|---|---|---|
| ITGA2B | P08514 | -5.767 | 18.526 | 24.293 | -6.359 | 0.000 | 25.314 | -7.634 | -3.900 | 0.003 |
| CD9 | P21926 | -3.184 | 19.334 | 22.518 | -5.785 | 0.000 | 25.862 | -4.315 | -2.052 | 0.003 |
| PPBP | P02775 | -4.586 | 20.362 | 24.949 | -6.082 | 0.000 | 20.796 | -6.156 | -3.017 | 0.003 |
| CYB5R3 | P00387 | -1.400 | 22.965 | 24.365 | -5.680 | 0.000 | 25.962 | -1.906 | -0.893 | 0.003 |
| ILK | Q13418 | -3.850 | 18.936 | 22.786 | -5.739 | 0.000 | 24.538 | -5.233 | -2.467 | 0.003 |
| PFN1 | P07737 | -1.261 | 27.133 | 28.394 | -5.609 | 0.000 | 22.890 | -1.726 | -0.796 | 0.005 |
| PFKP | Q01813 | -2.277 | 19.146 | 21.423 | -5.391 | 0.000 | 25.441 | -3.147 | -1.408 | 0.005 |
| DNAJC11 | Q9NVH1 | 1.415 | 21.652 | 20.238 | 5.188 | 0.000 | 24.270 | 0.852 | 1.977 | 0.008 |
| THBS1 | P07996 | -4.758 | 18.583 | 23.341 | -4.996 | 0.000 | 23.310 | -6.727 | -2.789 | 0.011 |
| MAOB | P27338 | -2.030 | 18.442 | 20.472 | -4.930 | 0.000 | 24.680 | -2.879 | -1.182 | 0.011 |
| RAB32 | Q13637 | -1.787 | 18.658 | 20.445 | -4.893 | 0.000 | 24.766 | -2.540 | -1.035 | 0.011 |
| SLC2A3,SLC2A14 | P11169,Q8TDB8 | -3.322 | 19.050 | 22.372 | -5.014 | 0.000 | 21.670 | -4.697 | -1.947 | 0.011 |
| F13A1 | P00488 | -5.550 | 17.979 | 23.529 | -5.020 | 0.000 | 21.169 | -7.848 | -3.252 | 0.011 |
| CALD1 | Q05682 | -3.402 | 18.766 | 22.168 | -4.899 | 0.000 | 23.357 | -4.837 | -1.967 | 0.011 |
| NEXN | Q0ZGT2 | -2.404 | 18.618 | 21.022 | -4.821 | 0.000 | 23.050 | -3.435 | -1.372 | 0.012 |
| ITGB3,ITGB3 | P05106 | -4.346 | 19.507 | 23.852 | -4.815 | 0.000 | 22.702 | -6.214 | -2.477 | 0.012 |
| RAB27B | O00194 | -3.036 | 19.239 | 22.275 | -4.709 | 0.000 | 25.289 | -4.363 | -1.709 | 0.012 |
| GP5 | P40197 | -2.379 | 19.045 | 21.424 | -4.602 | 0.000 | 24.872 | -3.443 | -1.314 | 0.015 |
| RSU1 | Q15404 | -3.551 | 19.242 | 22.793 | -4.680 | 0.000 | 19.897 | -5.134 | -1.967 | 0.018 |
| SDPR | O95810 | -3.900 | 19.609 | 23.509 | -4.786 | 0.000 | 17.863 | -5.612 | -2.187 | 0.018 |
| CROCC,CROCCP3 | Q5TZA2,Q8IVE0,Q5TZA2,Q8IVE0 | 1.982 | 23.268 | 21.286 | 4.469 | 0.000 | 24.438 | 1.067 | 2.897 | 0.018 |
| TLN1 | Q9Y490 | -1.963 | 27.266 | 29.229 | -4.430 | 0.000 | 25.701 | -2.875 | -1.052 | 0.018 |
| FLNA | P21333 | -3.941 | 23.918 | 27.859 | -4.496 | 0.000 | 23.115 | -5.754 | -2.128 | 0.018 |
| PARVB | Q9HBI1 | -4.976 | 18.421 | 23.397 | -5.352 | 0.000 | 12.049 | -7.000 | -2.951 | 0.018 |
| SNX1 | Q13596,Q13596 | 1.175 | 21.359 | 20.184 | 4.514 | 0.000 | 21.789 | 0.635 | 1.715 | 0.018 |
| SH3BGRL3 | Q9H299 | -1.379 | 23.178 | 24.557 | -4.821 | 0.000 | 15.734 | -1.986 | -0.772 | 0.019 |
| ALOX5 | P09917 | -1.459 | 18.614 | 20.073 | -4.282 | 0.000 | 25.449 | -2.161 | -0.758 | 0.021 |
| MMRN1 | Q13201 | -2.704 | 19.261 | 21.964 | -4.345 | 0.000 | 23.276 | -3.990 | -1.417 | 0.021 |
| SUPT16H | Q9Y5B9 | 1.816 | 22.969 | 21.153 | 4.293 | 0.000 | 23.709 | 0.942 | 2.690 | 0.023 |
| LGALSL | Q3ZCW2 | -2.803 | 19.321 | 22.124 | -4.502 | 0.000 | 18.026 | -4.112 | -1.495 | 0.023 |
| UNC13D | Q70J99 | -2.428 | 18.533 | 20.961 | -4.248 | 0.000 | 24.189 | -3.607 | -1.249 | 0.023 |
| VCL | P18206 | -6.279 | 19.788 | 26.066 | -5.176 | 0.000 | 11.143 | -8.944 | -3.613 | 0.023 |
| WDR61 | Q9GZS3,Q9GZS3 | 1.135 | 21.945 | 20.810 | 4.170 | 0.000 | 25.822 | 0.575 | 1.694 | 0.023 |
| CD36 | P16671 | -3.498 | 18.759 | 22.258 | -4.557 | 0.000 | 16.170 | -5.124 | -1.872 | 0.023 |
| SKAP2 | O75563 | -2.012 | 18.788 | 20.800 | -4.193 | 0.000 | 24.209 | -3.002 | -1.022 | 0.023 |
| IGLL5,IGLC1 | B9A064,P0CG04,B9A064,P0CG04 | -3.773 | 20.236 | 24.008 | -4.173 | 0.000 | 24.462 | -5.637 | -1.909 | 0.023 |

| | | | | | | | | | | |
|---|---|---|---|---|---|---|---|---|---|---|
| SCFD1 | Q8WVM8 | 0.751 | 23.574 | 22.823 | 4.160 | 0.000 | 24.747 | 0.379 | 1.124 | 0.023 |
| GP1BA | P07359 | -2.855 | 19.515 | 22.370 | -4.127 | 0.000 | 25.361 | -4.279 | -1.431 | 0.023 |
| SRRT | Q9BXP5 | 1.292 | 23.069 | 21.777 | 4.138 | 0.000 | 24.691 | 0.649 | 1.935 | 0.023 |
| ITGB1 | P05556 | -2.721 | 19.502 | 22.223 | -4.254 | 0.000 | 20.756 | -4.052 | -1.390 | 0.023 |
| PRPF40A | O75400,O75400 | 0.969 | 20.785 | 19.815 | 4.135 | 0.000 | 23.876 | 0.485 | 1.453 | 0.023 |
| PYGL | P06737 | -1.740 | 19.012 | 20.752 | -4.079 | 0.000 | 25.978 | -2.617 | -0.863 | 0.023 |
| SLC1A4 | P43007 | 1.502 | 22.125 | 20.623 | 4.144 | 0.000 | 22.551 | 0.752 | 2.253 | 0.024 |
| TPM1 | P09493 | -2.243 | 19.052 | 21.295 | -4.099 | 0.000 | 23.007 | -3.375 | -1.111 | 0.025 |
| MALSU1 | Q96EH3,Q96EH3 | -0.947 | 18.387 | 19.334 | -4.218 | 0.000 | 19.538 | -1.417 | -0.478 | 0.025 |
| HDAC2 | Q92769 | 1.627 | 21.636 | 20.009 | 4.409 | 0.000 | 15.768 | 0.844 | 2.411 | 0.025 |
| FERMT3 | Q86UX7 | -1.825 | 24.751 | 26.576 | -4.047 | 0.000 | 23.942 | -2.756 | -0.894 | 0.026 |
| RPL31 | P62899 | 1.872 | 24.112 | 22.240 | 3.988 | 0.000 | 25.541 | 0.906 | 2.838 | 0.026 |
| GANAB | Q14697,Q14697 | 0.707 | 28.140 | 27.433 | 4.147 | 0.001 | 19.436 | 0.351 | 1.063 | 0.027 |
| MYLK | Q15746 | -1.629 | 19.463 | 21.092 | -3.931 | 0.001 | 25.517 | -2.481 | -0.776 | 0.028 |
| EMC1 | Q8N766 | 0.911 | 23.057 | 22.146 | 4.042 | 0.001 | 21.231 | 0.442 | 1.379 | 0.028 |
| ELANE | P08246 | -2.201 | 19.020 | 21.221 | -3.946 | 0.001 | 24.585 | -3.351 | -1.051 | 0.028 |
| ARRB1 | P49407 | -1.774 | 19.221 | 20.995 | -3.913 | 0.001 | 25.944 | -2.706 | -0.842 | 0.028 |
| EDF1 | O60869 | 1.321 | 23.188 | 21.867 | 3.887 | 0.001 | 25.719 | 0.622 | 2.020 | 0.030 |
| ITGA2 | P17301 | -2.304 | 18.627 | 20.932 | -4.073 | 0.001 | 19.057 | -3.488 | -1.121 | 0.030 |
| IGLC3,IGLC2,IGLC6 | P0CG06,P0CG05,P0CF74,P0CG06,P0CG05,P0CF74 | -3.063 | 18.417 | 21.479 | -3.958 | 0.001 | 21.813 | -4.668 | -1.457 | 0.031 |
| CTAGE5 | O15320 | 1.774 | 22.046 | 20.271 | 4.255 | 0.001 | 15.006 | 0.886 | 2.663 | 0.031 |
| SELP | P16109 | -2.255 | 19.274 | 21.529 | -3.865 | 0.001 | 24.937 | -3.457 | -1.053 | 0.031 |
| NDUFB9 | Q9Y6M9,Q9Y6M9 | 1.204 | 23.198 | 21.994 | 3.800 | 0.001 | 25.943 | 0.553 | 1.856 | 0.034 |
| TMX2 | Q9Y320 | 1.383 | 22.238 | 20.855 | 3.857 | 0.001 | 23.232 | 0.642 | 2.125 | 0.034 |
| BUB3 | O43684 | 1.208 | 22.706 | 21.498 | 3.794 | 0.001 | 25.432 | 0.553 | 1.864 | 0.034 |
| UQCRQ | O14949,O14949 | 1.356 | 22.079 | 20.723 | 3.792 | 0.001 | 25.243 | 0.620 | 2.091 | 0.034 |
| ERO1L | Q96HE7 | 1.140 | 25.035 | 23.895 | 4.313 | 0.001 | 12.753 | 0.568 | 1.712 | 0.036 |
| SART3 | Q15020,Q15020 | 1.357 | 22.310 | 20.953 | 3.810 | 0.001 | 22.744 | 0.620 | 2.094 | 0.036 |
| LMNB2 | Q03252 | 0.616 | 25.879 | 25.263 | 3.749 | 0.001 | 25.292 | 0.278 | 0.955 | 0.036 |
| THUMPD1 | Q9NXG2 | 1.242 | 22.175 | 20.933 | 3.982 | 0.001 | 17.294 | 0.585 | 1.899 | 0.036 |
| PACSIN2 | Q9UNF0 | -1.405 | 19.294 | 20.699 | -3.834 | 0.001 | 20.821 | -2.168 | -0.643 | 0.037 |
| CBR4 | Q8N4T8,Q8N4T8 | 1.486 | 21.603 | 20.117 | 3.899 | 0.001 | 18.834 | 0.688 | 2.284 | 0.037 |
| ISOC2 | Q96AB3 | 1.599 | 22.921 | 21.322 | 4.047 | 0.001 | 15.481 | 0.759 | 2.439 | 0.037 |
| ITGB2 | P05107 | -1.627 | 18.890 | 20.517 | -3.701 | 0.001 | 25.772 | -2.530 | -0.723 | 0.037 |
| GP9 | P14770 | -2.344 | 19.027 | 21.371 | -3.699 | 0.001 | 25.816 | -3.647 | -1.041 | 0.037 |
| GP1BB | P13224 | -2.411 | 19.079 | 21.490 | -3.692 | 0.001 | 25.996 | -3.753 | -1.069 | 0.037 |
| SFXN3 | Q9BWM7 | -1.756 | 18.737 | 20.493 | -3.813 | 0.001 | 20.480 | -2.716 | -0.797 | 0.037 |
| GNG7 | O60262 | 1.523 | 22.276 | 20.753 | 3.755 | 0.001 | 22.654 | 0.683 | 2.363 | 0.037 |
| MAPRE2 | Q15555 | -2.144 | 19.212 | 21.356 | -3.711 | 0.001 | 22.513 | -3.340 | -0.947 | 0.040 |

| | | | | | | | | | | |
|---|---|---|---|---|---|---|---|---|---|---|
| LTBP1 | Q14766 | -1.768 | 18.565 | 20.333 | -3.642 | 0.001 | 24.869 | -2.769 | -0.768 | 0.042 |
| ZYX | Q15942 | -3.685 | 19.253 | 22.938 | -4.083 | 0.001 | 13.244 | -5.631 | -1.739 | 0.042 |
| SNCA | P37840 | -2.422 | 19.326 | 21.748 | -3.870 | 0.001 | 16.501 | -3.746 | -1.099 | 0.042 |
| HLA-C | Q29963,Q29963 | -1.797 | 18.774 | 20.571 | -3.735 | 0.001 | 19.660 | -2.802 | -0.792 | 0.043 |
| FDPS | P14324,P14324 | 1.631 | 22.964 | 21.332 | 3.608 | 0.001 | 24.970 | 0.700 | 2.562 | 0.043 |
| PTGS1 | P23219 | -1.799 | 19.033 | 20.833 | -3.722 | 0.001 | 19.762 | -2.808 | -0.790 | 0.043 |
| SYK | P43405 | -1.596 | 18.692 | 20.288 | -4.034 | 0.001 | 13.034 | -2.450 | -0.741 | 0.044 |
| CTTN | Q14247 | -2.121 | 19.218 | 21.338 | -3.600 | 0.001 | 23.976 | -3.336 | -0.905 | 0.044 |
| TREML1 | Q86YW5 | -1.587 | 18.871 | 20.458 | -3.579 | 0.001 | 24.718 | -2.501 | -0.673 | 0.044 |
| GP6 | Q9HCN6 | -1.579 | 18.779 | 20.358 | -3.663 | 0.001 | 20.763 | -2.476 | -0.682 | 0.044 |
| MYL9 | P24844 | -2.079 | 19.396 | 21.475 | -3.559 | 0.001 | 25.578 | -3.281 | -0.877 | 0.044 |
| STT3A | P46977 | 0.666 | 26.256 | 25.590 | 3.683 | 0.002 | 19.520 | 0.288 | 1.043 | 0.045 |
| MT-ND4 | P03905,P03905 | 0.997 | 21.159 | 20.162 | 3.618 | 0.002 | 21.884 | 0.426 | 1.569 | 0.045 |
| ARAP1 | Q96P48 | -1.026 | 18.937 | 19.962 | -3.627 | 0.002 | 20.754 | -1.614 | -0.437 | 0.046 |
| ALDH1A1 | P00352 | -1.010 | 18.676 | 19.686 | -3.958 | 0.002 | 13.098 | -1.561 | -0.459 | 0.046 |
| APEX1 | P27695 | 0.906 | 25.798 | 24.892 | 3.560 | 0.002 | 22.695 | 0.379 | 1.433 | 0.047 |
| INF2 | Q27J81 | -1.770 | 18.999 | 20.769 | -3.524 | 0.002 | 24.174 | -2.807 | -0.734 | 0.047 |
| BZW2 | Q9Y6E2,Q9Y6E2 | 1.077 | 22.536 | 21.459 | 3.608 | 0.002 | 20.249 | 0.455 | 1.699 | 0.047 |
| RPL36 | Q9Y3U8 | 2.296 | 23.573 | 21.277 | 3.640 | 0.002 | 19.060 | 0.976 | 3.616 | 0.047 |
| RAP1B | P61224,A6NIZ1 | -1.466 | 24.906 | 26.372 | -3.542 | 0.002 | 21.949 | -2.324 | -0.608 | 0.049 |
| GIMAP1 | Q8WWP7 | -0.940 | 18.531 | 19.471 | -3.455 | 0.002 | 25.802 | -1.499 | -0.380 | 0.051 |
| CLU | P10909 | -1.966 | 19.212 | 21.178 | -3.522 | 0.002 | 20.552 | -3.129 | -0.804 | 0.054 |
| VKORC1L1 | Q8N0U8,Q8N0U8 | 1.385 | 21.341 | 19.956 | 4.222 | 0.002 | 9.295 | 0.646 | 2.123 | 0.054 |
| ANXA3 | P12429 | -2.144 | 19.226 | 21.370 | -3.528 | 0.002 | 19.943 | -3.411 | -0.876 | 0.054 |
| CAT | P04040 | -1.625 | 23.419 | 25.043 | -4.255 | 0.002 | 8.998 | -2.489 | -0.761 | 0.054 |
| MYL12A,MYL12B | P19105,O14950 | -1.594 | 24.491 | 26.085 | -3.823 | 0.002 | 12.907 | -2.495 | -0.692 | 0.054 |
| RPL19 | P84098 | 1.640 | 23.559 | 21.919 | 3.426 | 0.002 | 24.822 | 0.654 | 2.626 | 0.054 |
| HBD | P02042 | -1.226 | 18.915 | 20.141 | -3.427 | 0.002 | 23.067 | -1.966 | -0.486 | 0.057 |
| EHD3 | Q9NZN3 | -2.291 | 19.771 | 22.062 | -3.385 | 0.002 | 25.132 | -3.684 | -0.897 | 0.057 |
| NRD1 | O43847,O43847 | 0.878 | 20.860 | 19.982 | 3.414 | 0.002 | 23.063 | 0.346 | 1.410 | 0.057 |
| ATP2C1 | P98194,P98194 | -0.796 | 18.941 | 19.737 | -3.620 | 0.002 | 15.577 | -1.263 | -0.329 | 0.057 |
| CAMK1 | Q14012 | -1.465 | 18.724 | 20.189 | -3.556 | 0.002 | 17.179 | -2.333 | -0.596 | 0.057 |
| PGLS | O95336 | 0.904 | 23.409 | 22.505 | 4.085 | 0.002 | 9.462 | 0.407 | 1.401 | 0.059 |
| TIAL1 | Q01085,Q01085 | 0.644 | 21.083 | 20.439 | 3.405 | 0.003 | 21.990 | 0.252 | 1.037 | 0.060 |
| FYB | O15117 | -1.699 | 19.432 | 21.130 | -3.348 | 0.003 | 24.399 | -2.745 | -0.652 | 0.061 |
| NDUFS7 | O75251 | 0.985 | 21.978 | 20.993 | 3.313 | 0.003 | 25.948 | 0.374 | 1.596 | 0.063 |
| CAPZA1 | P52907 | -0.595 | 23.951 | 24.546 | -3.391 | 0.003 | 20.897 | -0.960 | -0.230 | 0.063 |
| FCER1G | P30273 | -1.542 | 18.933 | 20.474 | -3.361 | 0.003 | 22.353 | -2.492 | -0.591 | 0.063 |
| NDUFA7,NDUFA7 | O95182,O95182 | 0.864 | 21.494 | 20.630 | 3.444 | 0.003 | 18.627 | 0.338 | 1.390 | 0.063 |

| | | | | | | | | | | |
|---|---|---|---|---|---|---|---|---|---|---|
| PRKG1 | Q13976 | -1.439 | 19.155 | 20.593 | -3.279 | 0.003 | 25.995 | -2.340 | -0.537 | 0.066 |
| NDUFA9 | Q16795,Q16795 | 0.697 | 23.763 | 23.066 | 3.267 | 0.003 | 25.700 | 0.258 | 1.136 | 0.068 |
| RCC1 | P18754,P18754 | 1.049 | 21.556 | 20.507 | 3.330 | 0.003 | 21.459 | 0.395 | 1.703 | 0.068 |
| FAF2 | Q96CS3,Q96CS3 | 0.882 | 21.660 | 20.778 | 3.719 | 0.003 | 11.288 | 0.362 | 1.402 | 0.069 |
| CLEC1B | Q9P126 | -1.300 | 18.819 | 20.119 | -3.301 | 0.003 | 21.932 | -2.116 | -0.483 | 0.069 |
| GOLGA3 | Q08378 | 1.508 | 22.415 | 20.906 | 3.708 | 0.003 | 11.362 | 0.617 | 2.400 | 0.069 |
| IDE | P14735 | 1.125 | 22.517 | 21.392 | 3.238 | 0.003 | 26.000 | 0.411 | 1.839 | 0.069 |
| SRP9 | P49458,P49458 | 1.061 | 23.875 | 22.814 | 3.236 | 0.003 | 25.986 | 0.387 | 1.735 | 0.069 |
| RNMT | O43148,O43148 | 1.192 | 21.484 | 20.292 | 3.309 | 0.003 | 20.927 | 0.443 | 1.941 | 0.070 |
| F11R | Q9Y624 | -1.972 | 19.387 | 21.359 | -3.587 | 0.003 | 12.734 | -3.162 | -0.781 | 0.071 |
| PSTPIP2 | Q9H939 | -1.763 | 18.646 | 20.409 | -3.609 | 0.003 | 12.240 | -2.825 | -0.701 | 0.071 |
| NCK2 | O43639 | -0.995 | 19.167 | 20.162 | -3.207 | 0.004 | 26.000 | -1.633 | -0.357 | 0.072 |
| DOK2 | O60496 | -1.534 | 18.514 | 20.048 | -3.447 | 0.004 | 15.101 | -2.481 | -0.586 | 0.072 |
| TUBA8 | Q9NY65 | -1.903 | 19.204 | 21.107 | -3.428 | 0.004 | 14.934 | -3.087 | -0.719 | 0.075 |
| DNAJC10 | Q8IXB1 | 1.666 | 22.975 | 21.309 | 3.387 | 0.004 | 15.215 | 0.619 | 2.713 | 0.079 |
| RNPS1 | Q15287,Q15287 | 0.766 | 20.959 | 20.192 | 3.192 | 0.004 | 22.531 | 0.269 | 1.263 | 0.081 |
| PUF60 | Q9UHX1 | 0.885 | 23.318 | 22.433 | 3.185 | 0.004 | 22.145 | 0.309 | 1.460 | 0.083 |
| FAM120A | Q9NZB2,Q9NZB2 | 0.984 | 22.270 | 21.286 | 3.119 | 0.005 | 25.039 | 0.334 | 1.633 | 0.087 |
| PCK2 | Q16822 | 2.316 | 24.896 | 22.580 | 3.628 | 0.005 | 10.138 | 0.896 | 3.736 | 0.087 |
| TBL2 | Q9Y4P3 | 1.282 | 23.921 | 22.639 | 3.673 | 0.005 | 9.583 | 0.500 | 2.064 | 0.088 |
| PCNA | P12004 | 1.286 | 24.426 | 23.140 | 3.202 | 0.005 | 19.349 | 0.446 | 2.125 | 0.088 |
| COPB2 | P35606 | 0.585 | 26.057 | 25.472 | 3.333 | 0.005 | 14.091 | 0.209 | 0.960 | 0.092 |
| HNRNPA3 | P51991 | 0.583 | 26.411 | 25.828 | 3.193 | 0.005 | 18.178 | 0.200 | 0.966 | 0.093 |
| ERP29 | P30040 | 0.591 | 26.885 | 26.294 | 3.158 | 0.005 | 19.321 | 0.200 | 0.982 | 0.095 |
| ANO6 | Q4KMQ2 | -1.204 | 18.885 | 20.089 | -3.102 | 0.005 | 22.035 | -2.009 | -0.399 | 0.096 |
| VIM | P08670,P08670 | 1.450 | 29.414 | 27.963 | 3.283 | 0.005 | 14.506 | 0.506 | 2.395 | 0.096 |
| BPI | P17213 | -1.943 | 18.709 | 20.652 | -3.239 | 0.005 | 15.632 | -3.218 | -0.669 | 0.096 |
| ESAM | Q96AP7 | -1.194 | 18.651 | 19.845 | -3.560 | 0.005 | 9.737 | -1.945 | -0.444 | 0.097 |
| EML4 | Q9HC35 | 1.021 | 23.430 | 22.409 | 3.269 | 0.005 | 14.383 | 0.353 | 1.689 | 0.097 |
| C6orf25 | O95866 | -1.534 | 18.987 | 20.520 | -3.115 | 0.005 | 20.080 | -2.560 | -0.507 | 0.097 |
| TXLNA | P40222,P40222 | 0.957 | 20.353 | 19.396 | 3.497 | 0.006 | 10.237 | 0.349 | 1.565 | 0.098 |
| MYO1G | B0I1T2 | -0.821 | 19.136 | 19.958 | -3.030 | 0.006 | 24.936 | -1.380 | -0.263 | 0.098 |
| ANXA1 | P04083 | -3.222 | 20.776 | 23.997 | -3.499 | 0.006 | 10.099 | -5.270 | -1.173 | 0.098 |
| SRC | P12931 | -2.063 | 19.680 | 21.743 | -3.022 | 0.006 | 25.439 | -3.468 | -0.658 | 0.098 |
| COTL1 | Q14019 | -3.510 | 19.411 | 22.922 | -3.568 | 0.006 | 9.307 | -5.725 | -1.296 | 0.099 |
| MYL6 | P60660 | -1.143 | 24.190 | 25.332 | -3.211 | 0.006 | 14.775 | -1.902 | -0.383 | 0.100 |
| ACTN1 | P12814 | -1.851 | 25.706 | 27.557 | -3.230 | 0.006 | 14.233 | -3.079 | -0.624 | 0.100 |
| CPSF6 | Q16630 | 0.969 | 22.749 | 21.780 | 3.051 | 0.006 | 21.589 | 0.310 | 1.629 | 0.100 |
| ELAVL1 | Q15717 | 0.887 | 23.822 | 22.935 | 3.103 | 0.006 | 18.500 | 0.288 | 1.486 | 0.100 |

| | | | | | | | | | | |
|---|---|---|---|---|---|---|---|---|---|---|
| PSMC4 | P43686,P43686 | 0.648 | 23.988 | 23.340 | 3.173 | 0.006 | 15.594 | 0.214 | 1.082 | 0.101 |
| THRAP3 | Q9Y2W1 | 1.289 | 22.984 | 21.695 | 3.119 | 0.006 | 17.209 | 0.418 | 2.159 | 0.102 |
| NDUFA8 | P51970 | 0.692 | 24.002 | 23.310 | 3.067 | 0.006 | 19.111 | 0.220 | 1.164 | 0.103 |
| RPN1 | P04843 | 0.640 | 28.237 | 27.597 | 3.441 | 0.006 | 9.992 | 0.225 | 1.054 | 0.103 |
| RAB6B | Q9NRW1 | -2.085 | 18.577 | 20.661 | -3.726 | 0.006 | 7.522 | -3.389 | -0.780 | 0.105 |
| SFXN2 | Q96NB2,Q96NB2 | -1.275 | 18.911 | 20.186 | -3.611 | 0.007 | 8.146 | -2.086 | -0.463 | 0.106 |
| MNDA | P41218 | -2.225 | 18.658 | 20.883 | -3.159 | 0.007 | 14.571 | -3.729 | -0.720 | 0.106 |
| NDUFA12 | Q9UI09,Q9UI09 | 0.871 | 21.704 | 20.833 | 2.959 | 0.007 | 24.840 | 0.265 | 1.477 | 0.106 |
| TM9SF1,TM9SF1 | O15321 | 1.086 | 21.645 | 20.559 | 3.506 | 0.007 | 8.944 | 0.385 | 1.788 | 0.106 |
| DIABLO | Q9NR28 | 1.217 | 24.592 | 23.374 | 2.941 | 0.007 | 25.977 | 0.366 | 2.068 | 0.107 |
| PDE5A | O76074 | -1.602 | 18.745 | 20.348 | -3.036 | 0.007 | 17.753 | -2.712 | -0.492 | 0.112 |
| HPSE | Q9Y251 | -1.625 | 19.148 | 20.773 | -2.938 | 0.008 | 21.524 | -2.774 | -0.477 | 0.120 |
| SSRP1 | Q08945 | 1.147 | 22.484 | 21.338 | 2.933 | 0.008 | 21.703 | 0.335 | 1.958 | 0.120 |
| NDUFB11 | Q9NX14,Q9NX14 | 0.864 | 22.267 | 21.403 | 2.909 | 0.008 | 23.324 | 0.250 | 1.477 | 0.120 |
| GPD1L | Q8N335,Q8N335 | -1.604 | 18.934 | 20.538 | -3.215 | 0.008 | 11.296 | -2.699 | -0.509 | 0.122 |
| ALOX12 | P18054 | -1.897 | 19.194 | 21.091 | -2.992 | 0.008 | 17.445 | -3.232 | -0.562 | 0.122 |
| PSPC1 | Q8WXF1,Q8WXF1 | 0.844 | 22.695 | 21.850 | 2.920 | 0.008 | 21.504 | 0.244 | 1.445 | 0.122 |
| IGHM | P01871 | -0.941 | 19.042 | 19.983 | -2.939 | 0.008 | 19.359 | -1.611 | -0.272 | 0.125 |
| XPOT | O43592 | 1.565 | 22.315 | 20.750 | 3.383 | 0.008 | 8.702 | 0.513 | 2.616 | 0.126 |
| NANS | Q9NR45 | 1.056 | 25.909 | 24.853 | 3.217 | 0.009 | 10.577 | 0.330 | 1.782 | 0.126 |
| CD74 | P04233,P04233 | -1.401 | 21.088 | 22.490 | -2.923 | 0.009 | 19.321 | -2.404 | -0.399 | 0.126 |
| GSN | P06396 | -4.856 | 20.699 | 25.556 | -3.481 | 0.009 | 7.795 | -8.088 | -1.625 | 0.126 |
| FLOT1 | O75955,O75955 | 1.224 | 23.534 | 22.310 | 2.852 | 0.009 | 24.696 | 0.339 | 2.108 | 0.126 |
| SARNP | P82979 | 0.882 | 23.598 | 22.715 | 2.827 | 0.009 | 25.998 | 0.241 | 1.524 | 0.129 |
| PRTN3 | P24158 | -2.544 | 19.888 | 22.431 | -3.106 | 0.009 | 11.970 | -4.329 | -0.759 | 0.131 |
| NDUFS8 | O00217 | 0.746 | 22.663 | 21.917 | 2.848 | 0.009 | 22.938 | 0.204 | 1.289 | 0.131 |
| NDUFB5 | O43674,O43674 | 1.047 | 21.843 | 20.796 | 2.813 | 0.009 | 25.983 | 0.282 | 1.812 | 0.132 |
| MAX | P61244,P61244 | -0.488 | 18.731 | 19.218 | -2.809 | 0.009 | 25.997 | -0.845 | -0.131 | 0.132 |
| PF4 | P02776 | -2.977 | 20.267 | 23.244 | -3.098 | 0.010 | 11.347 | -5.085 | -0.870 | 0.138 |
| NUDC | Q9Y266,Q9Y266 | 1.012 | 23.641 | 22.630 | 2.892 | 0.010 | 17.396 | 0.275 | 1.749 | 0.140 |
| FNDC3A | Q9Y2H6 | 0.710 | 20.647 | 19.937 | 2.840 | 0.010 | 19.926 | 0.188 | 1.231 | 0.141 |
| PRDX4 | Q13162 | 0.943 | 28.099 | 27.155 | 2.979 | 0.010 | 13.314 | 0.261 | 1.626 | 0.143 |
| NDUFB4 | O95168 | 0.816 | 22.349 | 21.534 | 2.776 | 0.011 | 23.900 | 0.209 | 1.422 | 0.143 |
| AP3B1 | O00203,O00203 | 1.035 | 22.284 | 21.249 | 2.920 | 0.011 | 15.055 | 0.280 | 1.790 | 0.143 |
| MECR | Q9BV79,Q9BV79 | -1.007 | 19.503 | 20.510 | -2.870 | 0.011 | 17.138 | -1.747 | -0.267 | 0.143 |
| CLC | Q05315,Q05315 | -1.167 | 18.909 | 20.076 | -2.771 | 0.011 | 24.237 | -2.036 | -0.298 | 0.143 |
| CA1 | P00915 | -0.897 | 18.501 | 19.398 | -2.755 | 0.011 | 25.615 | -1.567 | -0.227 | 0.143 |
| CTSA | P10619 | -1.617 | 19.844 | 21.461 | -3.374 | 0.011 | 7.517 | -2.735 | -0.499 | 0.143 |
| CPOX | P36551,P36551 | 1.206 | 21.576 | 20.370 | 3.157 | 0.011 | 9.620 | 0.350 | 2.062 | 0.143 |

| | | | | | | | | | | |
|---|---|---|---|---|---|---|---|---|---|---|
| UBLCP1 | Q8WVY7,Q8WVY7 | 0.789 | 20.262 | 19.473 | 3.232 | 0.011 | 8.668 | 0.234 | 1.344 | 0.144 |
| UGDH | O60701 | 1.317 | 22.027 | 20.710 | 2.871 | 0.011 | 16.292 | 0.346 | 2.289 | 0.144 |
| SEC61A1 | P61619 | 0.564 | 24.704 | 24.139 | 2.760 | 0.011 | 23.814 | 0.142 | 0.986 | 0.144 |
| MPP1 | Q00013 | -1.180 | 19.112 | 20.292 | -2.842 | 0.011 | 17.381 | -2.054 | -0.306 | 0.145 |
| FTL | P02792,P02792 | -1.335 | 19.990 | 21.325 | -2.898 | 0.011 | 14.362 | -2.320 | -0.349 | 0.149 |
| COPS2 | P61201,P61201 | 0.606 | 21.631 | 21.025 | 2.711 | 0.012 | 25.927 | 0.146 | 1.066 | 0.151 |
| EIF1AY | O14602 | 1.164 | 22.679 | 21.515 | 2.865 | 0.012 | 14.889 | 0.297 | 2.030 | 0.151 |
| SNAP23 | O00161 | -2.137 | 19.371 | 21.508 | -3.486 | 0.012 | 6.322 | -3.619 | -0.655 | 0.151 |
| PABPN1 | Q86U42 | 1.309 | 22.975 | 21.666 | 2.767 | 0.012 | 19.724 | 0.321 | 2.298 | 0.151 |
| SRP68 | Q9UHB9 | 1.054 | 23.997 | 22.943 | 3.030 | 0.012 | 10.478 | 0.284 | 1.825 | 0.151 |
| SYVN1 | Q86TM6 | 1.292 | 22.708 | 21.416 | 3.302 | 0.012 | 7.410 | 0.377 | 2.208 | 0.151 |
| KIAA0368 | Q5VYK3 | 1.725 | 22.413 | 20.688 | 3.334 | 0.012 | 7.173 | 0.508 | 2.943 | 0.151 |
| SEC24D | O94855 | 1.305 | 22.143 | 20.837 | 2.899 | 0.012 | 13.430 | 0.336 | 2.275 | 0.151 |
| NDUFB8,NDUFB8 | O95169,O95169 | 0.827 | 22.764 | 21.937 | 2.691 | 0.012 | 26.000 | 0.195 | 1.460 | 0.152 |
| NDUFA11 | Q86Y39,Q86Y39 | 1.155 | 21.818 | 20.663 | 2.721 | 0.012 | 22.579 | 0.276 | 2.034 | 0.152 |
| NDUFS5 | O43920 | 0.751 | 22.829 | 22.078 | 2.830 | 0.013 | 15.267 | 0.186 | 1.316 | 0.154 |
| SNRNP200 | O75643 | 1.057 | 24.229 | 23.171 | 2.758 | 0.013 | 18.650 | 0.254 | 1.861 | 0.155 |
| GUCY1B3 | Q02153 | -0.815 | 18.693 | 19.508 | -2.897 | 0.013 | 12.755 | -1.424 | -0.206 | 0.155 |
| HUWE1 | Q7Z6Z7,Q7Z6Z7 | 0.737 | 21.996 | 21.259 | 2.774 | 0.013 | 16.663 | 0.176 | 1.298 | 0.159 |
| SPARC | P09486 | -1.331 | 19.022 | 20.353 | -2.684 | 0.013 | 23.186 | -2.356 | -0.306 | 0.159 |
| F5 | P12259 | -1.338 | 19.763 | 21.101 | -2.668 | 0.013 | 25.056 | -2.371 | -0.305 | 0.159 |
| CBX5 | P45973 | 1.667 | 22.346 | 20.679 | 3.147 | 0.014 | 8.074 | 0.447 | 2.886 | 0.162 |
| NONO | Q15233 | 1.130 | 24.799 | 23.670 | 2.725 | 0.014 | 18.123 | 0.259 | 2.000 | 0.164 |
| MRE11A | P49959 | 1.128 | 22.062 | 20.935 | 2.935 | 0.014 | 10.766 | 0.280 | 1.975 | 0.164 |
| SNRPA1 | P09661 | 0.835 | 22.886 | 22.050 | 2.695 | 0.014 | 19.610 | 0.188 | 1.483 | 0.167 |
| PRKDC | P78527 | 1.043 | 26.252 | 25.209 | 2.621 | 0.015 | 25.142 | 0.224 | 1.862 | 0.172 |
| RBM3 | P98179 | 1.399 | 23.082 | 21.683 | 2.613 | 0.015 | 25.768 | 0.298 | 2.500 | 0.172 |
| DLD | P09622,P09622 | -0.442 | 23.901 | 24.344 | -2.614 | 0.015 | 25.563 | -0.791 | -0.094 | 0.172 |
| AASDHPPT | Q9NRN7 | 1.565 | 22.461 | 20.896 | 2.955 | 0.015 | 9.707 | 0.380 | 2.751 | 0.172 |
| GOLT1B | Q9Y3E0 | 0.775 | 21.787 | 21.012 | 3.167 | 0.015 | 7.314 | 0.201 | 1.348 | 0.172 |
| KHSRP | Q92945 | 1.121 | 25.139 | 24.019 | 2.970 | 0.015 | 9.440 | 0.273 | 1.968 | 0.172 |
| MAP4 | P27816 | 1.572 | 23.617 | 22.045 | 3.045 | 0.015 | 8.408 | 0.392 | 2.752 | 0.172 |
| WDR1 | O75083 | -1.162 | 24.611 | 25.773 | -2.655 | 0.015 | 20.335 | -2.074 | -0.250 | 0.172 |
| CNST | Q6PJW8 | -1.154 | 19.040 | 20.194 | -2.882 | 0.015 | 10.845 | -2.037 | -0.271 | 0.172 |
| GDI2 | P50395,P50395 | 0.469 | 26.658 | 26.189 | 2.922 | 0.015 | 10.001 | 0.111 | 0.827 | 0.172 |
| TMEM40 | Q8WWA1 | -1.237 | 19.011 | 20.247 | -2.706 | 0.015 | 16.484 | -2.203 | -0.270 | 0.172 |
| CMTM5 | Q96DZ9 | -1.266 | 19.062 | 20.329 | -2.601 | 0.015 | 24.648 | -2.269 | -0.263 | 0.173 |
| STOM | P27105 | -3.582 | 20.854 | 24.437 | -3.152 | 0.016 | 7.112 | -6.261 | -0.904 | 0.176 |
| AP2S1 | P53680,P53680 | 0.657 | 21.185 | 20.528 | 2.874 | 0.016 | 10.310 | 0.150 | 1.164 | 0.178 |

| | | | | | | | | | | |
|---|---|---|---|---|---|---|---|---|---|---|
| TMEM147 | Q9BVK8,Q9BVK8 | 0.698 | 19.931 | 19.233 | 2.778 | 0.016 | 12.530 | 0.153 | 1.243 | 0.178 |
| RANBP3 | Q9H6Z4 | 1.180 | 21.583 | 20.403 | 2.830 | 0.016 | 11.177 | 0.264 | 2.096 | 0.178 |
| TRAM1 | Q15629 | 0.773 | 23.771 | 22.998 | 2.921 | 0.016 | 9.269 | 0.177 | 1.370 | 0.180 |
| CAP1 | Q01518 | -1.333 | 25.690 | 27.023 | -3.071 | 0.017 | 7.527 | -2.345 | -0.321 | 0.180 |
| CAND1 | Q86VP6,Q86VP6 | 0.534 | 24.848 | 24.314 | 2.577 | 0.017 | 23.711 | 0.106 | 0.961 | 0.180 |
| DDX17 | Q92841 | 0.929 | 24.972 | 24.043 | 2.761 | 0.017 | 12.486 | 0.199 | 1.660 | 0.180 |
| NDUFA4 | O00483,O00483 | 1.398 | 23.467 | 22.068 | 2.579 | 0.017 | 22.963 | 0.276 | 2.520 | 0.180 |
| AP3D1 | O14617,O14617 | 1.161 | 21.396 | 20.235 | 3.130 | 0.017 | 6.899 | 0.281 | 2.040 | 0.180 |
| TCEA1 | P23193 | 1.301 | 23.287 | 21.985 | 2.992 | 0.017 | 8.111 | 0.301 | 2.302 | 0.180 |
| LIMS1 | P48059 | -1.739 | 19.293 | 21.032 | -2.566 | 0.017 | 23.728 | -3.139 | -0.340 | 0.180 |
| MARCKS,MARCKS | P29966,P29966 | 1.654 | 23.602 | 21.948 | 2.725 | 0.017 | 13.310 | 0.346 | 2.961 | 0.180 |
| NDUFS3 | O75489 | 0.377 | 24.254 | 23.877 | 2.554 | 0.017 | 25.320 | 0.073 | 0.682 | 0.180 |
| BCAT2 | O15382 | -0.884 | 21.861 | 22.744 | -2.600 | 0.017 | 19.640 | -1.594 | -0.174 | 0.182 |
| RASA3 | Q14644 | -1.331 | 18.808 | 20.139 | -2.769 | 0.017 | 11.620 | -2.382 | -0.280 | 0.183 |
| S100A8 | P05109 | -2.457 | 22.846 | 25.303 | -2.802 | 0.018 | 10.656 | -4.395 | -0.520 | 0.184 |
| PPIL1 | Q9Y3C6 | 0.874 | 21.115 | 20.241 | 2.672 | 0.018 | 14.137 | 0.173 | 1.575 | 0.188 |
| TJP2 | Q9UDY2 | -1.225 | 19.084 | 20.309 | -2.557 | 0.018 | 21.377 | -2.221 | -0.230 | 0.188 |
| ECI2 | O75521,O75521 | 0.780 | 21.624 | 20.845 | 2.523 | 0.018 | 25.371 | 0.144 | 1.416 | 0.188 |
| TCERG1 | O14776 | 0.871 | 21.241 | 20.370 | 2.523 | 0.018 | 25.082 | 0.160 | 1.583 | 0.188 |
| NDUFV1 | P49821,P49821 | 0.736 | 23.488 | 22.753 | 2.512 | 0.019 | 25.831 | 0.133 | 1.338 | 0.190 |
| UFC1 | Q9Y3C8 | 0.863 | 23.150 | 22.287 | 2.510 | 0.019 | 25.394 | 0.155 | 1.570 | 0.191 |
| SFPQ | P23246 | 0.865 | 26.392 | 25.527 | 2.726 | 0.019 | 11.513 | 0.170 | 1.559 | 0.192 |
| TSPAN14 | Q8NG11,Q8NG11 | -1.614 | 19.257 | 20.871 | -2.991 | 0.019 | 7.288 | -2.880 | -0.348 | 0.192 |
| PPIF | P30405 | -2.081 | 20.822 | 22.902 | -3.253 | 0.019 | 5.584 | -3.674 | -0.487 | 0.192 |
| PEPD | P12955,P12955 | 0.587 | 23.201 | 22.615 | 2.494 | 0.019 | 25.935 | 0.103 | 1.070 | 0.192 |
| COPA | P53621,P53621 | 0.663 | 26.479 | 25.815 | 2.604 | 0.019 | 15.764 | 0.123 | 1.204 | 0.192 |
| ENDOD1 | O94919 | -1.597 | 19.173 | 20.770 | -2.624 | 0.020 | 14.571 | -2.897 | -0.296 | 0.192 |
| SEC23A | Q15436,Q15436 | 1.213 | 22.362 | 21.149 | 2.648 | 0.020 | 13.537 | 0.227 | 2.199 | 0.192 |
| S100A9 | P06702 | -2.232 | 23.197 | 25.429 | -2.652 | 0.020 | 13.308 | -4.046 | -0.418 | 0.192 |
| TSPAN33 | Q86UF1 | -0.625 | 19.120 | 19.745 | -2.497 | 0.020 | 24.498 | -1.141 | -0.109 | 0.192 |
| UFL1 | O94874 | 0.640 | 24.640 | 24.000 | 2.616 | 0.020 | 14.777 | 0.118 | 1.162 | 0.192 |
| COPG1 | Q9Y678 | 0.639 | 26.095 | 25.456 | 2.748 | 0.020 | 10.329 | 0.123 | 1.155 | 0.195 |
| FLII | Q13045 | -0.953 | 19.885 | 20.838 | -2.522 | 0.020 | 20.434 | -1.740 | -0.166 | 0.195 |
| RRM1 | P23921,P23921 | -0.978 | 18.593 | 19.571 | -2.963 | 0.020 | 7.275 | -1.752 | -0.204 | 0.195 |
| RPS15 | P62841 | 0.979 | 23.054 | 22.075 | 2.477 | 0.020 | 25.135 | 0.165 | 1.792 | 0.196 |
| SOD2 | P04179 | -1.982 | 20.828 | 22.810 | -2.859 | 0.020 | 8.324 | -3.570 | -0.394 | 0.196 |
| NDUFS6 | O75380,O75380 | 0.887 | 20.982 | 20.094 | 2.894 | 0.020 | 7.848 | 0.178 | 1.596 | 0.196 |
| SLC39A7 | Q92504 | 0.748 | 21.024 | 20.276 | 2.546 | 0.021 | 17.593 | 0.130 | 1.366 | 0.196 |
| PRPF8 | Q6P2Q9 | 1.225 | 27.309 | 26.084 | 2.469 | 0.021 | 24.635 | 0.203 | 2.248 | 0.198 |

| | | | | | | | | | | |
|---|---|---|---|---|---|---|---|---|---|---|
| HP1BP3 | Q5SSJ5 | 0.919 | 24.044 | 23.125 | 2.615 | 0.021 | 13.369 | 0.162 | 1.677 | 0.199 |
| RBM25 | P49756 | 0.646 | 20.205 | 19.559 | 2.573 | 0.021 | 15.174 | 0.111 | 1.181 | 0.199 |
| ADAR | P55265 | 1.012 | 23.754 | 22.742 | 2.610 | 0.021 | 13.187 | 0.176 | 1.848 | 0.201 |
| DYNC1H1 | Q14204,Q14204 | 0.952 | 24.978 | 24.026 | 2.531 | 0.022 | 16.740 | 0.157 | 1.746 | 0.203 |
| CALU | O43852 | 1.116 | 26.636 | 25.521 | 2.732 | 0.022 | 9.580 | 0.200 | 2.031 | 0.205 |
| USP14 | P54578 | 0.610 | 24.337 | 23.727 | 2.741 | 0.022 | 9.244 | 0.109 | 1.111 | 0.207 |
| RPL8 | P62917 | 1.134 | 24.779 | 23.645 | 2.597 | 0.023 | 12.594 | 0.188 | 2.080 | 0.209 |
| NDUFS1 | P28331,P28331 | 0.463 | 24.887 | 24.423 | 2.436 | 0.023 | 23.574 | 0.070 | 0.856 | 0.209 |
| RASGRP2 | Q7LDG7 | -1.043 | 19.329 | 20.372 | -2.480 | 0.023 | 18.586 | -1.925 | -0.162 | 0.209 |
| SSR3 | Q9UNL2 | 0.867 | 23.607 | 22.741 | 2.475 | 0.023 | 18.981 | 0.134 | 1.600 | 0.209 |
| TMCO1 | Q9UM00,Q9UM00 | 0.776 | 21.211 | 20.435 | 2.551 | 0.023 | 14.105 | 0.124 | 1.429 | 0.209 |
| CNDP2 | Q96KP4 | 0.474 | 26.160 | 25.686 | 2.517 | 0.023 | 15.829 | 0.074 | 0.874 | 0.209 |
| PDXK | O00764 | 0.710 | 24.463 | 23.752 | 2.506 | 0.023 | 16.320 | 0.110 | 1.310 | 0.210 |
| MIA3 | Q5JRA6 | 0.964 | 23.136 | 22.172 | 2.524 | 0.023 | 15.073 | 0.150 | 1.777 | 0.210 |
| ARHGAP1 | Q07960 | -0.781 | 22.757 | 23.538 | -2.550 | 0.024 | 13.412 | -1.441 | -0.122 | 0.213 |
| SEC11C | Q9BY50 | 0.520 | 25.885 | 25.365 | 2.403 | 0.024 | 25.976 | 0.075 | 0.965 | 0.213 |
| LONP1 | P36776 | 1.253 | 25.729 | 24.475 | 2.966 | 0.024 | 6.269 | 0.230 | 2.277 | 0.213 |
| MGAT2 | Q10469 | 1.203 | 22.433 | 21.230 | 2.870 | 0.024 | 6.881 | 0.208 | 2.198 | 0.218 |
| ECE1 | P42892,P42892 | -0.865 | 18.527 | 19.392 | -2.577 | 0.025 | 11.626 | -1.599 | -0.131 | 0.219 |
| ARHGDIB | P52566 | -1.166 | 24.726 | 25.892 | -2.797 | 0.025 | 7.455 | -2.139 | -0.192 | 0.219 |
| PITRM1 | Q5JRX3 | 0.945 | 23.261 | 22.317 | 2.389 | 0.025 | 24.308 | 0.129 | 1.760 | 0.219 |
| COPB1 | P53618 | 0.586 | 26.005 | 25.420 | 2.644 | 0.025 | 9.769 | 0.090 | 1.081 | 0.219 |
| CTNNBL1 | Q8WYA6 | 0.496 | 20.222 | 19.726 | 2.467 | 0.025 | 16.256 | 0.070 | 0.922 | 0.219 |
| KHDRBS1 | Q07666 | 1.108 | 24.173 | 23.065 | 2.528 | 0.025 | 12.991 | 0.161 | 2.055 | 0.219 |
| PRKD2 | Q9BZL6,Q9BZL6 | -1.683 | 19.643 | 21.326 | -2.875 | 0.025 | 6.625 | -3.084 | -0.283 | 0.219 |
| ALG2 | Q9H553 | 0.674 | 23.187 | 22.513 | 2.380 | 0.025 | 24.432 | 0.090 | 1.258 | 0.219 |
| NHP2 | Q9NX24,Q9NX24 | 1.492 | 21.194 | 19.702 | 2.936 | 0.026 | 6.107 | 0.254 | 2.730 | 0.219 |
| TMPO | P42167 | 0.990 | 23.732 | 22.743 | 2.714 | 0.026 | 8.328 | 0.155 | 1.825 | 0.219 |
| ADRM1 | Q16186 | 0.668 | 21.269 | 20.601 | 2.492 | 0.026 | 14.308 | 0.094 | 1.242 | 0.219 |
| GIMAP4 | Q9NUV9 | -0.792 | 19.133 | 19.925 | -2.426 | 0.026 | 18.440 | -1.477 | -0.107 | 0.220 |
| SEC63 | Q9UGP8 | 0.921 | 23.382 | 22.461 | 2.457 | 0.026 | 15.716 | 0.125 | 1.717 | 0.221 |
| TPM3 | P06753 | -1.058 | 24.138 | 25.196 | -2.669 | 0.026 | 8.830 | -1.957 | -0.159 | 0.221 |
| HSD17B4 | P51659 | -0.895 | 23.975 | 24.871 | -2.841 | 0.026 | 6.714 | -1.647 | -0.144 | 0.221 |
| ARFGAP2 | Q8N6H7 | 0.682 | 20.919 | 20.236 | 2.453 | 0.026 | 15.676 | 0.092 | 1.273 | 0.222 |
| NCKAP1 | Q9Y2A7 | -1.404 | 18.576 | 19.980 | -3.020 | 0.027 | 5.431 | -2.572 | -0.237 | 0.223 |
| SHMT2 | P34897 | 0.970 | 25.017 | 24.047 | 2.418 | 0.027 | 17.525 | 0.126 | 1.815 | 0.224 |
| CUTA | O60888,O60888 | 0.938 | 23.693 | 22.755 | 2.359 | 0.027 | 24.037 | 0.117 | 1.759 | 0.224 |
| SLIRP | Q9GZT3 | 0.842 | 22.021 | 21.179 | 2.401 | 0.027 | 18.501 | 0.107 | 1.577 | 0.225 |
| H1FX | Q92522 | 1.614 | 25.202 | 23.588 | 2.735 | 0.027 | 7.543 | 0.239 | 2.990 | 0.225 |

| | | | | | | | | | | |
|---|---|---|---|---|---|---|---|---|---|---|
| SULT1A1,SULT1A2 | P50225,P50226,P50225,P50226 | -1.040 | 19.020 | 20.061 | -2.450 | 0.027 | 14.886 | -1.946 | -0.134 | 0.225 |
| RPL10A | P62906 | 0.593 | 25.336 | 24.743 | 2.429 | 0.027 | 16.077 | 0.076 | 1.110 | 0.225 |
| DDX46 | Q7L014,Q7L014 | 1.011 | 22.273 | 21.261 | 2.483 | 0.027 | 13.048 | 0.132 | 1.891 | 0.225 |
| HNRNPL | P14866 | 0.548 | 25.932 | 25.384 | 2.609 | 0.028 | 9.311 | 0.075 | 1.021 | 0.226 |
| STX11 | O75558 | -1.807 | 19.376 | 21.183 | -2.633 | 0.028 | 8.852 | -3.363 | -0.250 | 0.226 |
| XPO1 | O14980,O14980 | 0.766 | 24.212 | 23.446 | 2.470 | 0.028 | 13.370 | 0.098 | 1.433 | 0.226 |
| AK1 | P00568 | -1.216 | 19.064 | 20.280 | -2.728 | 0.028 | 7.422 | -2.258 | -0.174 | 0.226 |
| ATP6V1C1 | P21283,P21283 | -0.947 | 19.525 | 20.472 | -2.775 | 0.028 | 6.864 | -1.757 | -0.137 | 0.227 |
| GALNT2 | Q10471 | 1.035 | 23.434 | 22.399 | 2.592 | 0.028 | 9.272 | 0.136 | 1.934 | 0.228 |
| TSR1 | Q2NL82,Q2NL82 | -0.854 | 18.816 | 19.670 | -2.606 | 0.028 | 8.992 | -1.595 | -0.113 | 0.228 |
| PSMD10 | O75832,O75832 | 0.803 | 21.558 | 20.755 | 2.365 | 0.028 | 19.616 | 0.094 | 1.512 | 0.228 |
| NDUFA5 | Q16718 | 0.405 | 24.223 | 23.818 | 2.353 | 0.029 | 20.555 | 0.046 | 0.763 | 0.229 |
| RPL27 | P61353 | 1.040 | 25.283 | 24.242 | 2.463 | 0.029 | 12.771 | 0.126 | 1.954 | 0.229 |
| HNRNPA2B1 | P22626 | 0.442 | 28.420 | 27.978 | 2.323 | 0.029 | 24.351 | 0.050 | 0.835 | 0.229 |
| RPS17L,RPS17 | P0CW22,P08708 | 0.823 | 24.567 | 23.744 | 2.354 | 0.029 | 19.524 | 0.093 | 1.553 | 0.229 |
| GNAQ | P50148 | -1.389 | 19.328 | 20.717 | -2.406 | 0.029 | 15.358 | -2.617 | -0.161 | 0.229 |
| C1QBP | Q07021,Q07021 | -0.540 | 26.266 | 26.806 | -2.309 | 0.029 | 25.963 | -1.021 | -0.059 | 0.229 |
| TAGLN2 | P37802 | -1.939 | 24.658 | 26.597 | -2.786 | 0.029 | 6.492 | -3.611 | -0.267 | 0.229 |
| IARS | P41252,P41252 | 1.117 | 24.866 | 23.749 | 2.326 | 0.030 | 22.057 | 0.121 | 2.114 | 0.231 |
| PDXDC1 | Q6P996 | 0.887 | 22.022 | 21.134 | 2.318 | 0.030 | 22.728 | 0.095 | 1.680 | 0.231 |
| TNPO1 | Q92973,Q92973 | 0.776 | 23.087 | 22.311 | 2.590 | 0.030 | 8.782 | 0.096 | 1.457 | 0.231 |
| LRBA | P50851 | -1.586 | 19.785 | 21.371 | -2.610 | 0.030 | 8.405 | -2.975 | -0.196 | 0.231 |
| SLC1A5 | Q15758 | 1.508 | 23.141 | 21.634 | 2.517 | 0.030 | 10.299 | 0.178 | 2.837 | 0.231 |
| SRP72 | O76094 | 1.155 | 24.211 | 23.056 | 2.552 | 0.030 | 9.351 | 0.137 | 2.172 | 0.232 |
| EHD1 | Q9H4M9 | -1.785 | 20.134 | 21.920 | -2.626 | 0.030 | 8.019 | -3.352 | -0.218 | 0.232 |
| RPA2 | P15927,P15927 | 0.995 | 22.087 | 21.092 | 2.383 | 0.030 | 15.600 | 0.108 | 1.882 | 0.232 |
| RPL7A | P62424 | 0.895 | 25.877 | 24.982 | 2.546 | 0.030 | 9.431 | 0.105 | 1.685 | 0.232 |
| PARP1 | P09874 | 0.962 | 26.853 | 25.890 | 2.309 | 0.030 | 22.597 | 0.099 | 1.825 | 0.232 |
| PLEK | P08567 | -4.156 | 21.060 | 25.216 | -2.864 | 0.031 | 5.665 | -7.757 | -0.554 | 0.232 |
| CHD4 | Q14839 | 0.960 | 22.045 | 21.085 | 2.335 | 0.031 | 18.831 | 0.099 | 1.821 | 0.233 |
| TTC37 | Q6PGP7 | 0.809 | 21.865 | 21.056 | 2.377 | 0.031 | 15.297 | 0.085 | 1.533 | 0.233 |
| ME2 | P23368,P23368 | -0.815 | 24.191 | 25.005 | -2.470 | 0.031 | 11.089 | -1.540 | -0.089 | 0.233 |
| LYN | P07948 | -1.439 | 19.528 | 20.966 | -2.547 | 0.031 | 9.097 | -2.714 | -0.163 | 0.233 |
| DUSP3 | P51452 | -1.496 | 19.224 | 20.720 | -2.486 | 0.031 | 10.485 | -2.829 | -0.163 | 0.233 |
| LASP1 | Q14847 | -0.916 | 22.668 | 23.584 | -2.610 | 0.031 | 7.956 | -1.726 | -0.106 | 0.233 |
| STK24 | Q9Y6E0 | -1.063 | 19.642 | 20.704 | -2.550 | 0.031 | 8.935 | -2.006 | -0.119 | 0.233 |
| LCP2 | Q13094 | -0.866 | 19.916 | 20.783 | -2.284 | 0.033 | 21.699 | -1.654 | -0.079 | 0.240 |
| CYP20A1 | Q6UW02 | 1.334 | 22.967 | 21.633 | 2.700 | 0.033 | 6.552 | 0.149 | 2.519 | 0.240 |
| CMAS | Q8NFW8 | 0.772 | 20.514 | 19.742 | 2.489 | 0.033 | 9.732 | 0.078 | 1.466 | 0.240 |

| | | | | | | | | | | |
|---|---|---|---|---|---|---|---|---|---|---|
| DDX5 | P17844 | 0.774 | 25.381 | 24.607 | 2.347 | 0.033 | 15.300 | 0.072 | 1.475 | 0.241 |
| SEC23B | Q15437 | 0.905 | 24.542 | 23.637 | 2.283 | 0.033 | 21.233 | 0.081 | 1.728 | 0.241 |
| EMC3 | Q9P0I2 | 0.640 | 21.360 | 20.720 | 2.252 | 0.033 | 25.939 | 0.056 | 1.224 | 0.241 |
| HSD17B11 | Q8NBQ5 | -1.004 | 20.312 | 21.316 | -2.447 | 0.033 | 10.676 | -1.911 | -0.098 | 0.241 |
| CAPZA2 | P47755 | -0.781 | 23.169 | 23.950 | -2.461 | 0.034 | 10.044 | -1.488 | -0.074 | 0.243 |
| HNRNPH1 | P31943 | 0.528 | 25.813 | 25.285 | 2.312 | 0.034 | 17.055 | 0.046 | 1.009 | 0.243 |
| GPX4 | P36969,P36969 | 0.870 | 21.717 | 20.847 | 2.239 | 0.034 | 25.935 | 0.071 | 1.669 | 0.245 |
| KDELR1 | P24390 | 0.818 | 22.726 | 21.908 | 2.258 | 0.034 | 22.440 | 0.067 | 1.569 | 0.245 |
| HNRNPK | P61978 | 0.406 | 27.254 | 26.847 | 2.327 | 0.035 | 14.751 | 0.034 | 0.779 | 0.248 |
| TUBA4A | P68366 | -1.588 | 23.659 | 25.247 | -2.710 | 0.035 | 6.081 | -3.018 | -0.159 | 0.248 |
| ARCN1 | P48444 | 0.532 | 25.451 | 24.919 | 2.412 | 0.035 | 10.903 | 0.046 | 1.017 | 0.248 |
| YWHAH | Q04917 | -1.298 | 22.761 | 24.059 | -2.610 | 0.035 | 7.029 | -2.473 | -0.123 | 0.248 |
| ACTB | P60709 | -0.885 | 30.322 | 31.207 | -2.272 | 0.035 | 18.946 | -1.700 | -0.070 | 0.248 |
| ATP6V1A | P38606 | -0.787 | 22.927 | 23.714 | -2.667 | 0.035 | 6.355 | -1.499 | -0.075 | 0.249 |
| ADD3 | Q9UEY8,Q9UEY8 | -1.143 | 18.665 | 19.808 | -2.487 | 0.035 | 8.732 | -2.187 | -0.099 | 0.250 |
| ADAM10 | O14672 | -1.440 | 19.617 | 21.057 | -2.423 | 0.036 | 10.107 | -2.763 | -0.118 | 0.251 |
| LMAN1 | P49257 | 0.708 | 26.635 | 25.927 | 2.371 | 0.036 | 11.792 | 0.056 | 1.360 | 0.251 |
| TMPO | P42166 | 1.000 | 25.353 | 24.353 | 2.419 | 0.036 | 10.084 | 0.080 | 1.919 | 0.252 |
| UBE2I | P63279 | 0.952 | 22.971 | 22.019 | 2.270 | 0.037 | 16.903 | 0.067 | 1.838 | 0.255 |
| HK1 | P19367 | -1.701 | 21.204 | 22.905 | -2.326 | 0.037 | 13.122 | -3.280 | -0.122 | 0.256 |
| EIF3E | P60228,P60228 | 0.478 | 23.651 | 23.173 | 2.202 | 0.037 | 25.593 | 0.031 | 0.924 | 0.256 |
| SMARCA5,SMARCA1 | O60264,P28370,O60264,P28370 | 1.118 | 21.432 | 20.314 | 2.502 | 0.037 | 7.983 | 0.087 | 2.148 | 0.256 |
| SPCS3 | P61009 | 0.443 | 25.467 | 25.024 | 2.228 | 0.037 | 20.849 | 0.029 | 0.857 | 0.256 |
| UGP2 | Q16851,Q16851 | 0.613 | 24.174 | 23.561 | 2.196 | 0.038 | 24.699 | 0.038 | 1.188 | 0.259 |
| NUP205 | Q92621,Q92621 | 0.452 | 20.576 | 20.123 | 2.189 | 0.038 | 25.890 | 0.028 | 0.877 | 0.259 |
| EIF3L | Q9Y262,Q9Y262 | 0.711 | 23.459 | 22.748 | 2.189 | 0.038 | 25.885 | 0.043 | 1.379 | 0.259 |
| PSMD3 | O43242,O43242 | 0.615 | 23.575 | 22.959 | 2.215 | 0.038 | 20.356 | 0.036 | 1.195 | 0.262 |
| UBASH3B | Q8TF42 | -0.844 | 19.078 | 19.922 | -2.184 | 0.039 | 25.115 | -1.641 | -0.048 | 0.263 |
| UBAP2L | Q14157 | 0.731 | 21.550 | 20.819 | 2.295 | 0.039 | 13.230 | 0.044 | 1.418 | 0.263 |
| XRN2 | Q9H0D6 | 0.690 | 21.487 | 20.797 | 2.237 | 0.039 | 17.295 | 0.040 | 1.340 | 0.263 |
| HDLBP | Q00341 | 1.237 | 25.944 | 24.708 | 2.386 | 0.039 | 9.694 | 0.077 | 2.397 | 0.264 |
| SEC61B | P60468 | 0.903 | 24.601 | 23.698 | 2.172 | 0.039 | 25.686 | 0.048 | 1.758 | 0.264 |
| RAB1B | Q9H0U4 | -0.604 | 22.809 | 23.413 | -2.230 | 0.039 | 17.302 | -1.175 | -0.033 | 0.264 |
| DFFA | O00273,O00273 | -0.850 | 18.514 | 19.364 | -2.584 | 0.039 | 6.380 | -1.643 | -0.057 | 0.264 |
| MRPL45,MRPL45 | Q9BRJ2,Q9BRJ2 | -0.666 | 19.861 | 20.528 | -2.181 | 0.040 | 23.271 | -1.298 | -0.035 | 0.265 |
| APMAP | Q9HDC9 | -0.899 | 23.440 | 24.339 | -2.602 | 0.040 | 6.130 | -1.740 | -0.058 | 0.265 |
| ERLEC1 | Q96DZ1 | 1.174 | 24.310 | 23.136 | 2.492 | 0.040 | 7.382 | 0.071 | 2.277 | 0.265 |
| RTCB | Q9Y3I0 | 0.431 | 24.710 | 24.280 | 2.311 | 0.040 | 11.546 | 0.023 | 0.839 | 0.267 |
| PDCD6 | O75340,O75340 | 0.318 | 22.441 | 22.123 | 2.521 | 0.041 | 6.789 | 0.018 | 0.619 | 0.270 |

| | | | | | | | | | | |
|---|---|---|---|---|---|---|---|---|---|---|
| WARS | P23381 | 1.917 | 27.345 | 25.428 | 2.607 | 0.041 | 5.913 | 0.111 | 3.722 | 0.270 |
| CRKL | P46109 | -1.157 | 21.456 | 22.613 | -2.559 | 0.041 | 6.290 | -2.251 | -0.063 | 0.272 |
| ANO10 | Q9NW15,Q9NW15 | -0.720 | 18.766 | 19.486 | -2.487 | 0.042 | 7.037 | -1.403 | -0.036 | 0.273 |
| YWHAZ | P63104 | -0.812 | 27.278 | 28.091 | -2.393 | 0.042 | 8.580 | -1.586 | -0.039 | 0.273 |
| EIF5 | P55010 | 0.722 | 23.394 | 22.672 | 2.246 | 0.042 | 13.753 | 0.031 | 1.412 | 0.273 |
| PDLIM1 | O00151 | -1.794 | 22.584 | 24.378 | -2.329 | 0.042 | 10.029 | -3.509 | -0.079 | 0.274 |
| PRPF19 | Q9UMS4 | 0.891 | 23.977 | 23.085 | 2.291 | 0.042 | 11.327 | 0.038 | 1.745 | 0.274 |
| TARDBP | Q13148 | 0.687 | 22.513 | 21.826 | 2.149 | 0.042 | 23.335 | 0.026 | 1.348 | 0.274 |
| ALAD | P13716 | -1.296 | 19.777 | 21.073 | -2.516 | 0.042 | 6.546 | -2.532 | -0.061 | 0.274 |
| NARS2 | Q96I59,Q96I59 | -0.756 | 18.674 | 19.430 | -2.577 | 0.043 | 5.883 | -1.478 | -0.035 | 0.275 |
| PHPT1 | Q9NRX4 | 0.894 | 22.822 | 21.929 | 2.178 | 0.043 | 18.159 | 0.032 | 1.755 | 0.275 |
| HSP90AA1 | P07900 | 0.488 | 27.164 | 26.676 | 2.288 | 0.043 | 11.065 | 0.019 | 0.958 | 0.275 |
| PPIB | P23284 | 0.601 | 29.362 | 28.761 | 2.268 | 0.043 | 11.846 | 0.023 | 1.180 | 0.275 |
| ST6GAL1 | P15907 | 1.237 | 22.305 | 21.069 | 2.469 | 0.044 | 6.829 | 0.046 | 2.427 | 0.280 |
| VWA5A | O00534,O00534 | -0.926 | 18.432 | 19.358 | -2.569 | 0.044 | 5.777 | -1.817 | -0.036 | 0.280 |
| TMOD3 | Q9NYL9 | -1.517 | 20.551 | 22.068 | -2.453 | 0.044 | 6.938 | -2.982 | -0.052 | 0.280 |
| FKBP3 | Q00688,Q00688 | 0.762 | 24.481 | 23.719 | 2.179 | 0.044 | 16.252 | 0.022 | 1.502 | 0.280 |
| ILF3 | Q12906 | 0.680 | 25.881 | 25.201 | 2.293 | 0.044 | 10.146 | 0.021 | 1.338 | 0.280 |
| CLIC4 | Q9Y696 | 0.979 | 24.146 | 23.166 | 2.187 | 0.045 | 15.464 | 0.027 | 1.931 | 0.280 |
| BCS1L | Q9Y276,Q9Y276 | -0.949 | 18.676 | 19.625 | -2.498 | 0.045 | 6.348 | -1.866 | -0.031 | 0.280 |
| BZW1 | Q7L1Q6,Q7L1Q6 | 0.503 | 22.239 | 21.736 | 2.116 | 0.045 | 24.709 | 0.013 | 0.993 | 0.280 |
| PRPF4 | O43172,O43172 | 0.744 | 20.912 | 20.168 | 2.305 | 0.045 | 9.637 | 0.021 | 1.467 | 0.281 |
| EFHD2 | Q96C19 | -1.762 | 19.690 | 21.452 | -2.481 | 0.045 | 6.464 | -3.471 | -0.054 | 0.281 |
| AP2A1 | O95782,O95782 | 0.784 | 23.253 | 22.469 | 2.291 | 0.045 | 9.949 | 0.021 | 1.547 | 0.281 |
| TBCB | Q99426,Q99426 | -1.344 | 19.669 | 21.012 | -2.690 | 0.046 | 4.750 | -2.649 | -0.039 | 0.284 |
| RBM14 | Q96PK6 | 1.555 | 22.799 | 21.244 | 2.357 | 0.046 | 8.132 | 0.038 | 3.072 | 0.284 |
| NUDT5 | Q9UKK9 | 0.553 | 23.543 | 22.991 | 2.227 | 0.046 | 11.926 | 0.012 | 1.094 | 0.285 |
| MRPS26 | Q9BYN8,Q9BYN8 | -0.750 | 18.941 | 19.690 | -2.376 | 0.046 | 7.633 | -1.483 | -0.016 | 0.286 |
| M6PR | P20645,P20645 | -0.804 | 21.778 | 22.582 | -2.457 | 0.047 | 6.425 | -1.592 | -0.016 | 0.288 |
| CPPED1 | Q9BRF8 | -0.856 | 19.212 | 20.067 | -2.185 | 0.047 | 13.753 | -1.697 | -0.014 | 0.288 |
| DKC1 | O60832 | 0.872 | 23.800 | 22.927 | 2.101 | 0.047 | 22.695 | 0.013 | 1.732 | 0.288 |
| CSNK2A1,CSNK2A3 | P68400,Q8NEV1 | 1.132 | 22.792 | 21.660 | 2.263 | 0.048 | 9.752 | 0.014 | 2.251 | 0.292 |
| ATP2A3 | Q93084 | -1.740 | 22.740 | 24.480 | -2.392 | 0.049 | 6.905 | -3.465 | -0.015 | 0.296 |
| HBA1 | P69905 | -1.373 | 25.647 | 27.020 | -2.365 | 0.049 | 7.206 | -2.738 | -0.008 | 0.297 |
| LDHB | P07195,P07195 | -0.958 | 26.488 | 27.446 | -2.525 | 0.049 | 5.441 | -1.911 | -0.006 | 0.297 |
| GPX1 | P07203 | -1.119 | 22.630 | 23.749 | -2.183 | 0.049 | 12.334 | -2.233 | -0.006 | 0.297 |
| SF3A1 | Q15459 | 0.791 | 23.624 | 22.834 | 2.079 | 0.050 | 21.945 | 0.002 | 1.579 | 0.297 |
| PRDX2 | P32119,P32119 | 0.614 | 26.588 | 25.974 | 2.183 | 0.050 | 12.078 | 0.002 | 1.226 | 0.297 |
| PSMC5 | P62195,P62195 | 0.486 | 23.800 | 23.314 | 2.146 | 0.050 | 14.272 | 0.001 | 0.972 | 0.297 |

| | | | | | | | | | | |
|---|---|---|---|---|---|---|---|---|---|---|
| NDUFB10 | O96000,O96000 | 0.437 | 23.470 | 23.033 | 2.088 | 0.050 | 20.301 | 0.001 | 0.872 | 0.297 |
| ACBD3 | Q9H3P7 | 0.730 | 24.060 | 23.330 | 2.162 | 0.050 | 13.089 | 0.001 | 1.460 | 0.297 |
| PDS5B | Q9NTI5 | 1.456 | 21.493 | 20.037 | 2.542 | 0.050 | 5.220 | 0.002 | 2.911 | 0.297 |
| P4HA1 | P13674,P13674 | 2.754 | 22.555 | 19.801 | 2.633 | 0.050 | 4.637 | 0.000 | 5.509 | 0.298 |

**Supplemental Table S15.** Gene set enrichment analysis of the Hallmark library from significantly expressed proteins of the comparison between *TP53* mutation to WT *TP53* samples.

| ID | setSize | enrichmentScore | NES | pvalue | p.adjust | qvalue | rank |
|---|---|---|---|---|---|---|---|
| HALLMARK_COAGULATION | 13 | -0.822 | -2.375 | 0.0000 | 0.0000 | 0.0000 | 58 |
| HALLMARK_APICAL_JUNCTION | 16 | -0.679 | -2.074 | 0.0002 | 0.0002 | 0.0010 | 126 |
| HALLMARK_EPITHELIAL_MESENCHYMAL_TRANSITION | 14 | -0.684 | -2.021 | 0.0006 | 0.0006 | 0.0016 | 52 |
| HALLMARK_COMPLEMENT | 14 | -0.665 | -1.963 | 0.0011 | 0.0011 | 0.0024 | 126 |
| HALLMARK_E2F_TARGETS | 11 | 0.591 | 2.131 | 0.0024 | 0.0024 | 0.0040 | 106 |
| HALLMARK_G2M_CHECKPOINT | 10 | 0.589 | 2.033 | 0.0044 | 0.0044 | 0.0062 | 176 |
| HALLMARK_MYC_TARGETS_V1 | 19 | 0.421 | 1.920 | 0.0077 | 0.0077 | 0.0093 | 201 |
| HALLMARK_MYOGENESIS | 10 | -0.568 | -1.526 | 0.0569 | 0.0569 | 0.0599 | 153 |
| HALLMARK_ADIPOGENESIS | 11 | -0.432 | -1.195 | 0.2716 | 0.2716 | 0.2541 | 86 |
| HALLMARK_OXIDATIVE_PHOSPHORYLATION | 21 | 0.225 | 1.068 | 0.3517 | 0.3517 | 0.2961 | 172 |
| HALLMARK_HEME_METABOLISM | 11 | -0.342 | -0.947 | 0.5327 | 0.5327 | 0.4078 | 133 |
| HALLMARK_MTORC1_SIGNALING | 15 | -0.280 | -0.841 | 0.6616 | 0.6616 | 0.4643 | 94 |
| HALLMARK_GLYCOLYSIS | 10 | 0.229 | 0.791 | 0.7306 | 0.7306 | 0.4732 | 1 |
| HALLMARK_FATTY_ACID_METABOLISM | 13 | -0.222 | -0.641 | 0.8710 | 0.8710 | 0.5239 | 163 |
| HALLMARK_PROTEIN_SECRETION | 10 | -0.169 | -0.455 | 0.9816 | 0.9816 | 0.5511 | 248 |

**Supplemental Table S16.** Gene set enrichment analysis of the C5 library from significantly expressed proteins of the comparison between *TP53* mutation to WT *TP53* samples.

| ID | setSize | enrichmentScore | NES | pvalue | p.adjust | qvalue | rank |
|---|---|---|---|---|---|---|---|
| GOBP_BIOLOGICAL_ADHESION | 68 | -0.719295157 | -2.9290204 | 0.00000 | 0.00000 | 0.00000 | 110 |
| GOBP_CELL_ACTIVATION | 59 | -0.72391404 | -2.8922632 | 0.00000 | 0.00000 | 0.00000 | 110 |
| GOCC_SECRETORY_VESICLE | 65 | -0.69389998 | -2.8087195 | 0.00000 | 0.00000 | 0.00000 | 123 |
| GOBP_WOUND_HEALING | 48 | -0.727848871 | -2.8218642 | 0.00000 | 0.00000 | 0.00000 | 116 |
| GOCC_SECRETORY_GRANULE | 64 | -0.690364908 | -2.7873244 | 0.00000 | 0.00000 | 0.00000 | 123 |
| GOBP_RESPONSE_TO_WOUNDING | 51 | -0.717310921 | -2.8145994 | 0.00000 | 0.00000 | 0.00000 | 116 |
| GOMF_RNA_BINDING | 102 | 0.443262352 | 3.29855387 | 0.00000 | 0.00000 | 0.00000 | 176 |
| GOBP_CELL_MIGRATION | 58 | -0.679083241 | -2.7069892 | 0.00000 | 0.00000 | 0.00000 | 142 |
| GOBP_REGULATION_OF_BODY_FLUID_LEVELS | 41 | -0.719849094 | -2.7200284 | 0.00000 | 0.00000 | 0.00000 | 116 |
| GOBP_HEMOSTASIS | 40 | -0.727053068 | -2.7308927 | 0.00000 | 0.00000 | 0.00000 | 116 |
| GOBP_CELL_CELL_ADHESION | 49 | -0.689063232 | -2.687334 | 0.00000 | 0.00000 | 0.00000 | 110 |
| GOBP_LOCOMOTION | 66 | -0.639982671 | -2.5922807 | 0.00000 | 0.00000 | 0.00000 | 142 |
| GOBP_PLATELET_ACTIVATION | 29 | -0.770039881 | -2.7225039 | 0.00000 | 0.00000 | 0.00000 | 107 |
| GOBP_CELL_SUBSTRATE_ADHESION | 25 | -0.760679728 | -2.6026158 | 0.00000 | 0.00000 | 0.00000 | 85 |
| GOBP_REGULATION_OF_CELL_ADHESION | 41 | -0.657404654 | -2.4840753 | 0.00000 | 0.00000 | 0.00000 | 110 |
| GOBP_REGULATION_OF_CELL_ACTIVATION | 36 | -0.679688794 | -2.505056 | 0.00000 | 0.00000 | 0.00000 | 110 |
| GOMF_SIGNALING_RECEPTOR_BINDING | 44 | -0.645092353 | -2.4648532 | 0.00000 | 0.00000 | 0.00000 | 112 |
| GOBP_DEFENSE_RESPONSE | 57 | -0.602755739 | -2.3959145 | 0.00000 | 0.00000 | 0.00000 | 110 |
| GOCC_VESICLE_LUMEN | 39 | -0.662369445 | -2.4762092 | 0.00000 | 0.00000 | 0.00000 | 123 |
| GOBP_MRNA_METABOLIC_PROCESS | 42 | 0.516484686 | 3.16397603 | 0.00000 | 0.00000 | 0.00000 | 174 |
| GOBP_RNA_PROCESSING | 46 | 0.497741185 | 3.1189621 | 0.00000 | 0.00000 | 0.00000 | 159 |
| GOBP_INFLAMMATORY_RESPONSE | 29 | -0.70002292 | -2.4749564 | 0.00000 | 0.00000 | 0.00000 | 106 |
| GOBP_CELL_MATRIX_ADHESION | 17 | -0.799698869 | -2.4892454 | 0.00000 | 0.00000 | 0.00000 | 85 |
| GOCC_PLATELET_ALPHA_GRANULE | 17 | -0.792123255 | -2.4656646 | 0.00000 | 0.00000 | 0.00000 | 65 |
| GOBP_PLATELET_AGGREGATION | 21 | -0.745542845 | -2.4504475 | 0.00000 | 0.00000 | 0.00000 | 107 |
| GOBP_REGULATION_OF_CELLULAR_COMPONENT_MOVEMENT | 39 | -0.64109985 | -2.3966947 | 0.00000 | 0.00000 | 0.00000 | 146 |
| GOCC_RIBONUCLEOPROTEIN_COMPLEX | 50 | 0.459987101 | 2.91388207 | 0.00000 | 0.00000 | 0.00000 | 165 |
| GOBP_BIOLOGICAL_PROCESS_INVOLVED_IN_INTERSPECIES_INTERAC | 61 | -0.580001624 | -2.3250179 | 0.00000 | 0.00000 | 0.00000 | 60 |
| GOBP_DEFENSE_RESPONSE_TO_OTHER_ORGANISM | 41 | -0.620198384 | -2.3434874 | 0.00000 | 0.00000 | 0.00000 | 110 |
| GOBP_HOMOTYPIC_CELL_CELL_ADHESION | 22 | -0.722835085 | -2.398857 | 0.00000 | 0.00000 | 0.00000 | 107 |
| GOBP_VESICLE_MEDIATED_TRANSPORT | 68 | -0.550975436 | -2.2436107 | 0.00000 | 0.00000 | 0.00001 | 72 |
| GOBP_PHAGOCYTOSIS | 21 | -0.730590067 | -2.4013007 | 0.00000 | 0.00000 | 0.00001 | 53 |
| GOCC_NUCLEAR_PROTEIN_CONTAINING_COMPLEX | 46 | 0.462153619 | 2.89596213 | 0.00000 | 0.00000 | 0.00001 | 162 |
| GOCC_CELL_SUBSTRATE_JUNCTION | 49 | -0.593186145 | -2.3134151 | 0.00000 | 0.00000 | 0.00001 | 86 |
| GOBP_ACTIN_FILAMENT_BASED_PROCESS | 43 | -0.609059144 | -2.3215979 | 0.00000 | 0.00000 | 0.00001 | 153 |
| GOBP_MRNA_PROCESSING | 36 | 0.505468146 | 2.94561373 | 0.00000 | 0.00000 | 0.00001 | 174 |
| GOBP_LEUKOCYTE_MIGRATION | 29 | -0.660889507 | -2.3365988 | 0.00000 | 0.00000 | 0.00002 | 137 |
| GOCC_ANCHORING_JUNCTION | 62 | -0.548670955 | -2.2055296 | 0.00000 | 0.00000 | 0.00002 | 139 |
| GOCC_EXTERNAL_SIDE_OF_PLASMA_MEMBRANE | 18 | -0.737611481 | -2.3286128 | 0.00000 | 0.00000 | 0.00002 | 41 |
| HP_SUBCUTANEOUS_HEMORRHAGE | 21 | -0.712013558 | -2.3402435 | 0.00000 | 0.00000 | 0.00002 | 116 |
| GOMF_CYTOSKELETAL_PROTEIN_BINDING | 47 | -0.57935297 | -2.2405912 | 0.00000 | 0.00000 | 0.00002 | 126 |
| GOCC_NUCLEAR_BODY | 41 | 0.456416053 | 2.76840653 | 0.00000 | 0.00000 | 0.00002 | 159 |
| GOBP_IMMUNE_RESPONSE | 54 | -0.566629007 | -2.2366609 | 0.00000 | 0.00000 | 0.00002 | 110 |
| GOBP_INTEGRIN_MEDIATED_SIGNALING_PATHWAY | 15 | -0.776942356 | -2.3357933 | 0.00000 | 0.00000 | 0.00003 | 105 |
| GOMF_INTEGRIN_BINDING | 15 | -0.77443609 | -2.3282585 | 0.00000 | 0.00000 | 0.00003 | 106 |
| GOMF_CALCIUM_ION_BINDING | 21 | -0.702323317 | -2.3083937 | 0.00000 | 0.00000 | 0.00003 | 76 |
| HP_PURPURA | 14 | -0.783517296 | -2.3154564 | 0.00000 | 0.00000 | 0.00004 | 66 |
| GOBP_POSITIVE_REGULATION_OF_CELL_ADHESION | 27 | -0.661211253 | -2.3058225 | 0.00000 | 0.00000 | 0.00004 | 110 |
| HP_PETECHIAE | 11 | -0.808276521 | -2.237784 | 0.00000 | 0.00000 | 0.00004 | 66 |
| GOBP_ANATOMICAL_STRUCTURE_FORMATION_INVOLVED_IN_MORPH | 37 | -0.609112808 | -2.2560171 | 0.00000 | 0.00000 | 0.00004 | 104 |
| GOCC_CATALYTIC_COMPLEX | 72 | 0.34453696 | 2.38971329 | 0.00000 | 0.00000 | 0.00005 | 227 |
| GOBP_POSITIVE_REGULATION_OF_CATALYTIC_ACTIVITY | 42 | -0.583540184 | -2.2141795 | 0.00000 | 0.00000 | 0.00005 | 112 |
| GOBP_CYTOKINE_PRODUCTION | 25 | -0.665762911 | -2.2778641 | 0.00000 | 0.00000 | 0.00005 | 110 |
| GOCC_SECRETORY_GRANULE_MEMBRANE | 19 | -0.709598951 | -2.2705364 | 0.00000 | 0.00000 | 0.00005 | 105 |
| GOBP_REGULATION_OF_PLATELET_ACTIVATION | 16 | -0.746945172 | -2.2893543 | 0.00000 | 0.00000 | 0.00005 | 107 |
| HP_MACULAR_PURPURA | 13 | -0.775381927 | -2.2481405 | 0.00000 | 0.00000 | 0.00005 | 66 |
| GOMF_CELL_ADHESION_MOLECULE_BINDING | 50 | -0.560604886 | -2.1946598 | 0.00000 | 0.00000 | 0.00005 | 108 |
| HP_ABNORMAL_BLEEDING | 36 | -0.605326426 | -2.2309866 | 0.00001 | 0.00001 | 0.00006 | 120 |
| GOCC_INTRINSIC_COMPONENT_OF_PLASMA_MEMBRANE | 26 | -0.64874637 | -2.2404855 | 0.00001 | 0.00001 | 0.00006 | 121 |
| GOCC_CHROMOSOME | 34 | 0.482187273 | 2.73954783 | 0.00001 | 0.00001 | 0.00007 | 118 |
| GOCC_EXTERNAL_ENCAPSULATING_STRUCTURE | 16 | -0.741587427 | -2.272933 | 0.00001 | 0.00001 | 0.00007 | 85 |
| GOCC_CELL_SURFACE | 36 | -0.601887487 | -2.2183121 | 0.00001 | 0.00001 | 0.00007 | 110 |
| GOBP_CIRCULATORY_SYSTEM_DEVELOPMENT | 30 | -0.6290595 | -2.2435214 | 0.00001 | 0.00001 | 0.00007 | 105 |
| GOBP_ENDOCYTOSIS | 32 | -0.621273562 | -2.245375 | 0.00001 | 0.00001 | 0.00007 | 98 |
| GOCC_SUPRAMOLECULAR_POLYMER | 34 | -0.605954698 | -2.211021 | 0.00001 | 0.00001 | 0.00008 | 75 |
| GOBP_CELL_JUNCTION_ORGANIZATION | 27 | -0.643693285 | -2.2447326 | 0.00001 | 0.00001 | 0.00009 | 137 |
| GOCC_CONTRACTILE_FIBER | 13 | -0.764423408 | -2.2163674 | 0.00001 | 0.00001 | 0.00009 | 69 |
| GOBP_REGULATION_OF_TRANSPORT | 55 | -0.535968684 | -2.1213203 | 0.00001 | 0.00001 | 0.00011 | 110 |
| GOBP_REGULATION_OF_CELL_POPULATION_PROLIFERATION | 41 | -0.569097803 | -2.1503982 | 0.00001 | 0.00001 | 0.00011 | 110 |
| GOCC_CELL_CELL_JUNCTION | 29 | -0.62370131 | -2.2051186 | 0.00001 | 0.00001 | 0.00011 | 139 |
| HP_BRUISING_SUSCEPTIBILITY | 16 | -0.732787723 | -2.2459623 | 0.00001 | 0.00001 | 0.00011 | 116 |
| GOBP_NEGATIVE_REGULATION_OF_RESPONSE_TO_STIMULUS | 42 | -0.56790323 | -2.1548468 | 0.00001 | 0.00001 | 0.00012 | 110 |

| | | | | | | | |
|---|---|---|---|---|---|---|---|
| GOBP_RNA_SPLICING | 31 | 0.485639687 | 2.67237213 | 0.00001 | 0.00001 | 0.00012 | 228 |
| GOBP_TAXIS | 28 | -0.635678478 | -2.2343677 | 0.00001 | 0.00001 | 0.00012 | 137 |
| GOBP_DEFENSE_RESPONSE_TO_BACTERIUM | 13 | -0.758831258 | -2.2001535 | 0.00002 | 0.00002 | 0.00013 | 98 |
| HP_GASTROINTESTINAL_HEMORRHAGE | 11 | -0.779776035 | -2.1588779 | 0.00002 | 0.00002 | 0.00014 | 38 |
| HP_ABNORMAL_FEMALE_REPRODUCTIVE_SYSTEM_PHYSIOLOGY | 11 | -0.778428552 | -2.1551473 | 0.00002 | 0.00002 | 0.00014 | 37 |
| GOCC_COLLAGEN_CONTAINING_EXTRACELLULAR_MATRIX | 14 | -0.754905126 | -2.2309015 | 0.00002 | 0.00002 | 0.00014 | 74 |
| GOBP_POSITIVE_REGULATION_OF_INTRACELLULAR_SIGNAL_TRANSDU | 36 | -0.585779486 | -2.1589445 | 0.00002 | 0.00002 | 0.00016 | 110 |
| GOBP_MYELOID_LEUKOCYTE_ACTIVATION | 15 | -0.733893908 | -2.2063728 | 0.00002 | 0.00002 | 0.00019 | 110 |
| GOBP_REGULATION_OF_HYDROLASE_ACTIVITY | 33 | -0.592916438 | -2.1539156 | 0.00002 | 0.00002 | 0.00019 | 146 |
| GOBP_POSITIVE_REGULATION_OF_MOLECULAR_FUNCTION | 56 | -0.528178312 | -2.0960142 | 0.00003 | 0.00003 | 0.00019 | 112 |
| GOBP_MAINTENANCE_OF_LOCATION | 21 | -0.666621214 | -2.1910481 | 0.00003 | 0.00003 | 0.00019 | 81 |
| GOBP_VASCULATURE_DEVELOPMENT | 24 | -0.652743695 | -2.2137618 | 0.00003 | 0.00003 | 0.00020 | 105 |
| GOCC_ACTIN_CYTOSKELETON | 39 | -0.5743945 | -2.1473227 | 0.00003 | 0.00003 | 0.00021 | 83 |
| GOBP_REGULATION_OF_VESICLE_MEDIATED_TRANSPORT | 26 | -0.623596402 | -2.1536286 | 0.00003 | 0.00003 | 0.00021 | 108 |
| GOBP_TUBE_MORPHOGENESIS | 23 | -0.652872625 | -2.1941792 | 0.00003 | 0.00003 | 0.00022 | 92 |
| GOBP_REGULATION_OF_RESPONSE_TO_EXTERNAL_STIMULUS | 43 | -0.552019067 | -2.1041738 | 0.00004 | 0.00004 | 0.00026 | 110 |
| HP_VASCULAR_SKIN_ABNORMALITY | 32 | -0.597672357 | -2.1600767 | 0.00004 | 0.00004 | 0.00027 | 116 |
| GOCC_SIDE_OF_MEMBRANE | 28 | -0.618905894 | -2.1754132 | 0.00004 | 0.00004 | 0.00027 | 119 |
| GOBP_POSITIVE_REGULATION_OF_CELLULAR_COMPONENT_ORGANIZ | 41 | -0.553348434 | -2.0908875 | 0.00004 | 0.00004 | 0.00027 | 150 |
| GOBP_CELL_SUBSTRATE_JUNCTION_ORGANIZATION | 10 | -0.777227723 | -2.0898376 | 0.00004 | 0.00004 | 0.00028 | 101 |
| GOCC_CHROMOSOMAL_REGION | 13 | 0.666591162 | 2.59251756 | 0.00004 | 0.00004 | 0.00028 | 112 |
| GOBP_ACTIN_FILAMENT_ORGANIZATION | 29 | -0.603056171 | -2.1321269 | 0.00004 | 0.00004 | 0.00029 | 153 |
| GOBP_POSITIVE_REGULATION_OF_LOCOMOTION | 24 | -0.642250795 | -2.1781754 | 0.00004 | 0.00004 | 0.00029 | 137 |
| GOBP_POSITIVE_REGULATION_OF_SIGNALING | 45 | -0.548511877 | -2.1027591 | 0.00004 | 0.00004 | 0.00029 | 110 |
| GOBP_CYTOSKELETON_ORGANIZATION | 59 | -0.508797713 | -2.0328061 | 0.00005 | 0.00005 | 0.00029 | 75 |
| GOMF_CHROMATIN_BINDING | 16 | 0.614150581 | 2.60980318 | 0.00005 | 0.00005 | 0.00031 | 149 |
| GOBP_CELL_CHEMOTAXIS | 19 | -0.667967848 | -2.1373275 | 0.00005 | 0.00005 | 0.00034 | 137 |
| HP_EPISTAXIS | 14 | -0.736129207 | -2.1754148 | 0.00005 | 0.00005 | 0.00034 | 65 |
| GOCC_PLASMA_MEMBRANE_REGION | 30 | -0.59366141 | -2.1172752 | 0.00006 | 0.00006 | 0.00035 | 109 |
| GOBP_RESPONSE_TO_GROWTH_FACTOR | 19 | -0.666304472 | -2.1320051 | 0.00006 | 0.00006 | 0.00035 | 101 |
| HP_ABNORMALITY_OF_THE_COAGULATION_CASCADE | 15 | -0.717384776 | -2.1567399 | 0.00006 | 0.00006 | 0.00035 | 37 |
| GOCC_PLATELET_ALPHA_GRANULE_LUMEN | 10 | -0.768989229 | -2.0676856 | 0.00006 | 0.00006 | 0.00035 | 65 |
| GOBP_BLOOD_VESSEL_MORPHOGENESIS | 20 | -0.65606677 | -2.1291178 | 0.00006 | 0.00006 | 0.00036 | 104 |
| GOBP_TUBE_DEVELOPMENT | 27 | -0.613514225 | -2.13949 | 0.00006 | 0.00006 | 0.00037 | 92 |
| GOCC_VESICLE_MEMBRANE | 50 | -0.524198779 | -2.0521369 | 0.00007 | 0.00007 | 0.00038 | 110 |
| GOBP_REGULATION_OF_ANATOMICAL_STRUCTURE_MORPHOGENESIS | 28 | -0.608709721 | -2.1395744 | 0.00007 | 0.00007 | 0.00039 | 84 |
| GOBP_ACTIN_FILAMENT_BUNDLE_ORGANIZATION | 14 | -0.732527554 | -2.1647711 | 0.00007 | 0.00007 | 0.00040 | 72 |
| GOBP_REGULATED_EXOCYTOSIS | 10 | -0.764851485 | -2.0565599 | 0.00007 | 0.00007 | 0.00041 | 106 |
| GOBP_REGULATION_OF_INTRACELLULAR_SIGNAL_TRANSDUCTION | 57 | -0.493721334 | -1.9625099 | 0.00007 | 0.00007 | 0.00042 | 110 |
| GOBP_SUPRAMOLECULAR_FIBER_ORGANIZATION | 37 | -0.557605928 | -2.0652472 | 0.00008 | 0.00008 | 0.00042 | 75 |
| GOCC_PLASMA_MEMBRANE_SIGNALING_RECEPTOR_COMPLEX | 12 | -0.73826698 | -2.0945545 | 0.00008 | 0.00008 | 0.00043 | 38 |
| HP_ABNORMALITY_OF_THE_MENSTRUAL_CYCLE | 10 | -0.763093554 | -2.0518331 | 0.00008 | 0.00008 | 0.00043 | 37 |
| GOBP_RNA_SPLICING_VIA_TRANSESTERIFICATION_REACTIONS | 27 | 0.487831606 | 2.54848629 | 0.00008 | 0.00008 | 0.00043 | 159 |
| GOBP_IMMUNE_EFFECTOR_PROCESS | 22 | -0.634958085 | -2.1072215 | 0.00008 | 0.00008 | 0.00044 | 110 |
| GOBP_CELL_MORPHOGENESIS | 41 | -0.539881354 | -2.0400007 | 0.00008 | 0.00008 | 0.00044 | 122 |
| HP_ABNORMAL_URINE_CYTOLOGY | 13 | -0.725614179 | -2.103844 | 0.00008 | 0.00008 | 0.00044 | 38 |
| GOCC_CHROMOSOME_TELOMERIC_REGION | 10 | 0.706202011 | 2.44684964 | 0.00009 | 0.00009 | 0.00046 | 112 |
| GOMF_ACTIN_BINDING | 31 | -0.581137271 | -2.0904979 | 0.00009 | 0.00009 | 0.00048 | 83 |
| GOBP_MYELOID_LEUKOCYTE_MIGRATION | 18 | -0.669653066 | -2.1140705 | 0.00010 | 0.00010 | 0.00051 | 136 |
| GOBP_POSITIVE_REGULATION_OF_TRANSPORT | 34 | -0.566475547 | -2.0669686 | 0.00010 | 0.00010 | 0.00051 | 103 |
| GOBP_REGULATION_OF_CELL_SUBSTRATE_ADHESION | 13 | -0.721432952 | -2.0917209 | 0.00010 | 0.00010 | 0.00051 | 53 |
| GOBP_POSITIVE_REGULATION_OF_PHOSPHORUS_METABOLIC_PROCE | 24 | -0.626994088 | -2.1264327 | 0.00010 | 0.00010 | 0.00052 | 110 |
| GOCC_NUCLEAR_SPECK | 25 | 0.508483778 | 2.56940789 | 0.00010 | 0.00010 | 0.00052 | 159 |
| HP_GENERALIZED_ABNORMALITY_OF_SKIN | 40 | -0.551559387 | -2.0717188 | 0.00011 | 0.00011 | 0.00054 | 116 |
| GOBP_CHROMOSOME_SEGREGATION | 10 | 0.70049505 | 2.42707615 | 0.00011 | 0.00011 | 0.00056 | 131 |
| GOBP_POSITIVE_REGULATION_OF_CELL_ACTIVATION | 20 | -0.645171108 | -2.0937583 | 0.00011 | 0.00011 | 0.00056 | 110 |
| GOCC_RECEPTOR_COMPLEX | 15 | -0.701460041 | -2.1088639 | 0.00012 | 0.00012 | 0.00057 | 38 |
| HP_ABNORMALITY_OF_REPRODUCTIVE_SYSTEM_PHYSIOLOGY | 18 | -0.66385636 | -2.0957705 | 0.00012 | 0.00012 | 0.00058 | 37 |
| GOBP_REGULATION_OF_CELL_CELL_ADHESION | 25 | -0.602489034 | -2.0613767 | 0.00013 | 0.00013 | 0.00062 | 110 |
| GOBP_CELL_MORPHOGENESIS_INVOLVED_IN_DIFFERENTIATION | 25 | -0.602351032 | -2.0609046 | 0.00013 | 0.00013 | 0.00062 | 65 |
| HP_ABNORMAL_THROMBOCYTE_MORPHOLOGY | 27 | -0.598450688 | -2.0869594 | 0.00013 | 0.00013 | 0.00063 | 66 |
| GOBP_HUMORAL_IMMUNE_RESPONSE | 12 | -0.726324987 | -2.0606736 | 0.00014 | 0.00014 | 0.00065 | 58 |
| HP_ABNORMALITY_OF_COAGULATION | 19 | -0.650006096 | -2.0798544 | 0.00014 | 0.00014 | 0.00067 | 37 |
| GOBP_INNATE_IMMUNE_RESPONSE | 32 | -0.572468367 | -2.0689858 | 0.00016 | 0.00016 | 0.00071 | 110 |
| GOBP_REGULATION_OF_RESPONSE_TO_STRESS | 53 | -0.506085646 | -1.9928088 | 0.00016 | 0.00016 | 0.00071 | 110 |
| GOCC_ENDOCYTIC_VESICLE | 21 | -0.635101194 | -2.0874482 | 0.00016 | 0.00016 | 0.00071 | 120 |
| GOBP_POSITIVE_REGULATION_OF_HYDROLASE_ACTIVITY | 24 | -0.618077374 | -2.0961919 | 0.00016 | 0.00016 | 0.00074 | 112 |
| GOCC_CELL_PROJECTION_MEMBRANE | 12 | -0.722638385 | -2.0502142 | 0.00017 | 0.00017 | 0.00074 | 109 |
| GOBP_CHROMOSOME_ORGANIZATION | 28 | 0.45215947 | 2.39091512 | 0.00017 | 0.00017 | 0.00074 | 131 |
| GOBP_RESPONSE_TO_LIPID | 36 | -0.5468359 | -2.0154143 | 0.00018 | 0.00018 | 0.00080 | 60 |
| GOBP_LEUKOCYTE_CELL_CELL_ADHESION | 21 | -0.632123866 | -2.0776624 | 0.00018 | 0.00018 | 0.00080 | 110 |
| GOBP_POSITIVE_REGULATION_OF_DEVELOPMENTAL_PROCESS | 37 | -0.545978042 | -2.0221801 | 0.00019 | 0.00019 | 0.00082 | 110 |
| GOBP_LEUKOCYTE_CHEMOTAXIS | 17 | -0.671972776 | -2.0916688 | 0.00020 | 0.00020 | 0.00086 | 133 |
| GOBP_POSITIVE_REGULATION_OF_MULTICELLULAR_ORGANISMAL_PRO | 38 | -0.537292119 | -1.9992086 | 0.00020 | 0.00020 | 0.00086 | 110 |
| GOBP_IMPORT_INTO_CELL | 10 | -0.740174831 | -1.9902084 | 0.00022 | 0.00022 | 0.00093 | 33 |

| | | | | | | | |
|---|---|---|---|---|---|---|---|
| GOCC_PHAGOCYTIC_VESICLE | 10 | -0.739361284 | -1.9880209 | 0.00022 | 0.00022 | 0.00093 | 92 |
| GOBP_ION_TRANSPORT | 44 | -0.518909134 | -1.9827159 | 0.00022 | 0.00022 | 0.00093 | 136 |
| GOBP_BIOLOGICAL_PROCESS_INVOLVED_IN_SYMBIOTIC_INTERACTIC | 15 | -0.689712956 | -2.0735475 | 0.00023 | 0.00023 | 0.00095 | 57 |
| GOCC_NUCLEOLUS | 34 | 0.404533974 | 2.29836048 | 0.00023 | 0.00023 | 0.00095 | 153 |
| GOBP_REGULATION_OF_RESPONSE_TO_WOUNDING | 17 | -0.669372195 | -2.0835739 | 0.00023 | 0.00023 | 0.00095 | 107 |
| GOBP_REGULATION_OF_SUPRAMOLECULAR_FIBER_ORGANIZATION | 21 | -0.627465988 | -2.0623529 | 0.00023 | 0.00023 | 0.00096 | 153 |
| GOBP_AMEBOIDAL_TYPE_CELL_MIGRATION | 19 | -0.63942875 | -2.0460096 | 0.00024 | 0.00024 | 0.00097 | 141 |
| GOCC_SPECIFIC_GRANULE | 14 | -0.712546904 | -2.1057242 | 0.00024 | 0.00024 | 0.00099 | 105 |
| GOCC_CELL_LEADING_EDGE | 20 | -0.629036425 | -2.0413968 | 0.00025 | 0.00025 | 0.00099 | 64 |
| GOBP_ACTIN_CYTOSKELETON_REORGANIZATION | 12 | -0.710716866 | -2.0163914 | 0.00025 | 0.00025 | 0.00101 | 57 |
| GOCC_SUPRAMOLECULAR_COMPLEX | 45 | -0.517717856 | -1.9847081 | 0.00026 | 0.00026 | 0.00101 | 75 |
| GOBP_CELL_CELL_SIGNALING | 28 | -0.585187751 | -2.0568962 | 0.00026 | 0.00026 | 0.00103 | 122 |
| HP_PROLONGED_BLEEDING_TIME | 10 | -0.73520245 | -1.9768385 | 0.00027 | 0.00027 | 0.00104 | 37 |
| GOBP_POSITIVE_REGULATION_OF_CYTOKINE_PRODUCTION | 17 | -0.667420437 | -2.0774986 | 0.00027 | 0.00027 | 0.00105 | 110 |
| GOBP_REGULATION_OF_WOUND_HEALING | 15 | -0.685287981 | -2.0602443 | 0.00027 | 0.00027 | 0.00106 | 107 |
| HP_ABNORMALITY_OF_BLOOD_CIRCULATION | 19 | -0.634941673 | -2.0316521 | 0.00028 | 0.00028 | 0.00109 | 38 |
| GOBP_SUBSTRATE_ADHESION_DEPENDENT_CELL_SPREADING | 11 | -0.719422431 | -1.9917837 | 0.00029 | 0.00029 | 0.00112 | 80 |
| GOBP_POSITIVE_REGULATION_OF_CELL_DEATH | 26 | -0.580777825 | -2.005752 | 0.00030 | 0.00030 | 0.00112 | 58 |
| GOCC_INNER_MITOCHONDRIAL_MEMBRANE_PROTEIN_COMPLEX | 23 | 0.474219526 | 2.3419871 | 0.00031 | 0.00031 | 0.00116 | 172 |
| GOBP_CELLULAR_MACROMOLECULE_BIOSYNTHETIC_PROCESS | 52 | 0.343279614 | 2.22972325 | 0.00032 | 0.00032 | 0.00119 | 96 |
| GOCC_CHROMATIN | 22 | 0.486535871 | 2.36641668 | 0.00032 | 0.00032 | 0.00120 | 110 |
| GOBP_MUSCLE_CONTRACTION | 16 | -0.664706187 | -2.0372954 | 0.00034 | 0.00034 | 0.00125 | 107 |
| GOBP_CELL_PROJECTION_ORGANIZATION | 39 | -0.529914999 | -1.9810401 | 0.00035 | 0.00035 | 0.00127 | 150 |
| GOBP_CELL_JUNCTION_ASSEMBLY | 19 | -0.628328656 | -2.0104921 | 0.00035 | 0.00035 | 0.00128 | 137 |
| GOCC_RUFFLE | 11 | -0.715623335 | -1.9812656 | 0.00035 | 0.00035 | 0.00129 | 109 |
| GOMF_LIPID_BINDING | 21 | -0.615376085 | -2.0226158 | 0.00037 | 0.00037 | 0.00134 | 60 |
| GOBP_POSITIVE_REGULATION_OF_PROTEIN_PHOSPHORYLATION | 22 | -0.602411622 | -1.9992103 | 0.00037 | 0.00037 | 0.00135 | 110 |
| GOCC_MEMBRANE_MICRODOMAIN | 16 | -0.662404371 | -2.0302404 | 0.00038 | 0.00038 | 0.00135 | 108 |
| HP_INTRACRANIAL_HEMORRHAGE | 13 | -0.692774404 | -2.0086284 | 0.00041 | 0.00041 | 0.00145 | 120 |
| GOBP_TISSUE_MORPHOGENESIS | 14 | -0.698716725 | -2.0648531 | 0.00042 | 0.00042 | 0.00149 | 83 |
| GOCC_NADH_DEHYDROGENASE_COMPLEX | 20 | 0.500382742 | 2.33644615 | 0.00043 | 0.00043 | 0.00151 | 172 |
| GOBP_REGULATION_OF_SYSTEM_PROCESS | 13 | -0.690731336 | -2.0027047 | 0.00043 | 0.00043 | 0.00151 | 107 |
| GOBP_EXOCYTOSIS | 15 | -0.676172975 | -2.0328411 | 0.00045 | 0.00045 | 0.00155 | 106 |
| GOBP_RESPONSE_TO_OXYGEN_CONTAINING_COMPOUND | 48 | -0.489296866 | -1.897 | 0.00045 | 0.00045 | 0.00155 | 60 |
| GOBP_POSITIVE_REGULATION_OF_EPITHELIAL_CELL_MIGRATION | 11 | -0.708728231 | -1.9621759 | 0.00045 | 0.00045 | 0.00155 | 53 |
| GOBP_REGULATION_OF_EPITHELIAL_CELL_MIGRATION | 11 | -0.708728231 | -1.9621759 | 0.00045 | 0.00045 | 0.00155 | 53 |
| GOBP_RESPONSE_TO_BACTERIUM | 28 | -0.571481223 | -2.0087186 | 0.00049 | 0.00049 | 0.00166 | 106 |
| GOBP_RESPONSE_TO_MOLECULE_OF_BACTERIAL_ORIGIN | 20 | -0.611717629 | -1.9851926 | 0.00050 | 0.00050 | 0.00168 | 60 |
| GOBP_REGULATION_OF_PHOSPHORUS_METABOLIC_PROCESS | 36 | -0.52799362 | -1.9459694 | 0.00050 | 0.00050 | 0.00169 | 112 |
| GOBP_CELL_ADHESION_MEDIATED_BY_INTEGRIN | 10 | -0.716970911 | -1.9278169 | 0.00057 | 0.00057 | 0.00190 | 106 |
| GOCC_PLASMA_MEMBRANE_PROTEIN_COMPLEX | 21 | -0.602229992 | -1.9794073 | 0.00057 | 0.00057 | 0.00190 | 112 |
| GOBP_DEVELOPMENTAL_GROWTH | 10 | -0.715589248 | -1.9241019 | 0.00058 | 0.00058 | 0.00192 | 49 |
| GOBP_HOMEOSTATIC_PROCESS | 57 | -0.4569943 | -1.8165223 | 0.00062 | 0.00062 | 0.00203 | 132 |
| GOBP_CATION_TRANSPORT | 38 | -0.516638973 | -1.9223603 | 0.00063 | 0.00063 | 0.00208 | 131 |
| GOCC_BLOOD_MICROPARTICLE | 13 | -0.675848441 | -1.9595533 | 0.00071 | 0.00071 | 0.00232 | 23 |
| GOMF_OXIDOREDUCTASE_ACTIVITY_ACTING_ON_NAD_P_H_QUINONI | 19 | 0.49533231 | 2.26633254 | 0.00071 | 0.00071 | 0.00232 | 172 |
| GOBP_METAL_ION_TRANSPORT | 24 | -0.585485671 | -1.9856581 | 0.00073 | 0.00073 | 0.00236 | 131 |
| GOBP_GRANULOCYTE_CHEMOTAXIS | 13 | -0.672285757 | -1.9492237 | 0.00077 | 0.00077 | 0.00248 | 110 |
| GOBP_NEGATIVE_REGULATION_OF_CELL_ACTIVATION | 12 | -0.685279021 | -1.9442211 | 0.00079 | 0.00079 | 0.00254 | 110 |
| GOCC_RESPIRASOME | 22 | 0.462598619 | 2.2499905 | 0.00081 | 0.00081 | 0.00257 | 172 |
| GOBP_POSITIVE_REGULATION_OF_IMMUNE_SYSTEM_PROCESS | 40 | -0.509411949 | -1.9134083 | 0.00081 | 0.00081 | 0.00258 | 110 |
| GOBP_GRANULOCYTE_MIGRATION | 15 | -0.659031287 | -1.9813064 | 0.00082 | 0.00082 | 0.00259 | 136 |
| HP_ABNORMALITY_OF_URINE_HOMEOSTASIS | 25 | -0.562309866 | -1.9239063 | 0.00089 | 0.00089 | 0.00278 | 38 |
| GOBP_POSITIVE_REGULATION_OF_KINASE_ACTIVITY | 15 | -0.656247719 | -1.9729379 | 0.00092 | 0.00092 | 0.00286 | 110 |
| GOBP_RECEPTOR_INTERNALIZATION | 11 | -0.686089789 | -1.8994994 | 0.00093 | 0.00093 | 0.00289 | 98 |
| GOCC_ACTIN_FILAMENT | 11 | -0.683485571 | -1.8922894 | 0.00094 | 0.00094 | 0.00291 | 75 |
| GOCC_ACTIN_FILAMENT_BUNDLE | 10 | -0.702816388 | -1.8897577 | 0.00095 | 0.00095 | 0.00291 | 83 |
| GOBP_NEGATIVE_REGULATION_OF_TRANSPORT | 15 | -0.655101702 | -1.9694925 | 0.00096 | 0.00096 | 0.00295 | 54 |
| GOBP_RECEPTOR_METABOLIC_PROCESS | 13 | -0.667555018 | -1.9355074 | 0.00100 | 0.00100 | 0.00304 | 98 |
| GOBP_REGULATION_OF_ACTIN_FILAMENT_BASED_PROCESS | 20 | -0.595837747 | -1.933658 | 0.00101 | 0.00101 | 0.00306 | 153 |
| GOBP_CIRCULATORY_SYSTEM_PROCESS | 20 | -0.595740034 | -1.9333408 | 0.00102 | 0.00102 | 0.00308 | 116 |
| GOBP_CHEMICAL_HOMEOSTASIS | 32 | -0.534227904 | -1.930779 | 0.00105 | 0.00105 | 0.00313 | 119 |
| GOBP_REGULATION_OF_ACTIN_FILAMENT_ORGANIZATION | 18 | -0.617059801 | -1.9480355 | 0.00107 | 0.00107 | 0.00320 | 153 |
| HP_ABNORMAL_CEREBRAL_VASCULAR_MORPHOLOGY | 19 | -0.602159392 | -1.9267571 | 0.00109 | 0.00109 | 0.00321 | 38 |
| GOBP_GROWTH | 19 | -0.602134106 | -1.9266762 | 0.00109 | 0.00109 | 0.00321 | 53 |
| GOBP_POSITIVE_REGULATION_OF_CELL_POPULATION_PROLIFERATION | 22 | -0.576895495 | -1.9145305 | 0.00109 | 0.00109 | 0.00322 | 110 |
| GOBP_POSITIVE_REGULATION_OF_PROTEIN_MODIFICATION_PROCES | 25 | -0.555644939 | -1.9011027 | 0.00114 | 0.00114 | 0.00332 | 110 |
| GOBP_TISSUE_MIGRATION | 14 | -0.67022939 | -1.9806671 | 0.00120 | 0.00120 | 0.00345 | 53 |
| GOBP_REGULATION_OF_INFLAMMATORY_RESPONSE | 14 | -0.67011969 | -1.9803429 | 0.00120 | 0.00120 | 0.00345 | 106 |
| GOBP_CELL_CELL_JUNCTION_ORGANIZATION | 13 | -0.663360135 | -1.9233447 | 0.00120 | 0.00120 | 0.00345 | 136 |
| GOBP_REGULATION_OF_CYTOSKELETON_ORGANIZATION | 28 | -0.552600794 | -1.9423552 | 0.00120 | 0.00120 | 0.00345 | 153 |
| GOBP_REGULATION_OF_IMMUNE_SYSTEM_PROCESS | 51 | -0.47847377 | -1.8774453 | 0.00121 | 0.00121 | 0.00345 | 110 |
| GOBP_NEGATIVE_REGULATION_OF_MULTICELLULAR_ORGANISMAL_PR | 23 | -0.579381667 | -1.9471903 | 0.00122 | 0.00122 | 0.00345 | 122 |
| GOBP_G_PROTEIN_COUPLED_RECEPTOR_SIGNALING_PATHWAY | 12 | -0.66919756 | -1.898596 | 0.00122 | 0.00122 | 0.00345 | 72 |
| GOBP_REGULATION_OF_CELLULAR_COMPONENT_BIOGENESIS | 41 | -0.493726461 | -1.8655994 | 0.00122 | 0.00122 | 0.00345 | 109 |

| | | | | | | | |
|---|---|---|---|---|---|---|---|
| GOBP_REGULATION_OF_CELL_DIFFERENTIATION | 35 | -0.509607249 | -1.8660783 | 0.00125 | 0.00125 | 0.00351 | 137 |
| GOBP_RIBONUCLEOPROTEIN_COMPLEX_BIOGENESIS | 23 | 0.440901379 | 2.17744164 | 0.00126 | 0.00126 | 0.00352 | 173 |
| GOBP_REGULATION_OF_CELL_DEATH | 54 | -0.460535239 | -1.8178758 | 0.00127 | 0.00127 | 0.00352 | 64 |
| GOBP_RESPONSE_TO_ENDOGENOUS_STIMULUS | 45 | -0.487432384 | -1.8686066 | 0.00129 | 0.00129 | 0.00358 | 106 |
| GOBP_POSITIVE_REGULATION_OF_PROTEIN_KINASE_ACTIVITY | 13 | -0.65886767 | -1.9103193 | 0.00134 | 0.00134 | 0.00369 | 103 |
| GOCC_MICROTUBULE_ORGANIZING_CENTER | 16 | 0.513174714 | 2.18071112 | 0.00135 | 0.00135 | 0.00372 | 112 |
| GOBP_POSITIVE_REGULATION_OF_MAPK_CASCADE | 13 | -0.656333504 | -1.9029717 | 0.00136 | 0.00136 | 0.00372 | 110 |
| GOBP_DIVALENT_INORGANIC_CATION_HOMEOSTASIS | 18 | -0.609505785 | -1.9241877 | 0.00138 | 0.00138 | 0.00376 | 45 |
| GOBP_REGULATION_OF_ION_TRANSPORT | 18 | -0.608858749 | -1.9221451 | 0.00140 | 0.00140 | 0.00380 | 136 |
| GOBP_REGULATION_OF_PROTEIN_CONTAINING_COMPLEX_ASSEMBLY | 23 | -0.576782941 | -1.9384565 | 0.00143 | 0.00143 | 0.00387 | 153 |
| GOBP_REGULATION_OF_PROTEIN_PHOSPHORYLATION | 33 | -0.510817247 | -1.85567 | 0.00145 | 0.00145 | 0.00389 | 112 |
| GOBP_ACTOMYOSIN_STRUCTURE_ORGANIZATION | 13 | -0.653366584 | -1.8943695 | 0.00148 | 0.00148 | 0.00396 | 153 |
| GOBP_LEUKOCYTE_MEDIATED_IMMUNITY | 17 | -0.618327675 | -1.9246861 | 0.00150 | 0.00150 | 0.00401 | 136 |
| GOBP_REGULATION_OF_MRNA_METABOLIC_PROCESS | 19 | 0.470886076 | 2.15448178 | 0.00153 | 0.00153 | 0.00406 | 228 |
| GOMF_MRNA_BINDING | 19 | 0.470886076 | 2.15448178 | 0.00153 | 0.00153 | 0.00406 | 228 |
| GOBP_SECRETION | 25 | -0.550820784 | -1.8845972 | 0.00155 | 0.00155 | 0.00407 | 110 |
| GOMF_NADH_DEHYDROGENASE_ACTIVITY | 18 | 0.482595155 | 2.16492023 | 0.00159 | 0.00159 | 0.00415 | 172 |
| GOMF_OXIDOREDUCTION_DRIVEN_ACTIVE_TRANSMEMBRANE_TRANS | 18 | 0.482595155 | 2.16492023 | 0.00159 | 0.00159 | 0.00415 | 172 |
| HP_DECREASED_HEAD_CIRCUMFERENCE | 56 | 0.293995301 | 1.94346412 | 0.00162 | 0.00162 | 0.00421 | 134 |
| GOBP_MAINTENANCE_OF_LOCATION_IN_CELL | 15 | -0.642028115 | -1.9301882 | 0.00166 | 0.00166 | 0.00429 | 119 |
| GOCC_AZUROPHIL_GRANULE | 16 | -0.620781357 | -1.9026677 | 0.00171 | 0.00171 | 0.00440 | 118 |
| GOBP_RESPONSE_TO_CYTOKINE | 28 | -0.542465319 | -1.9067297 | 0.00171 | 0.00171 | 0.00440 | 53 |
| GOCC_TRANSPORTER_COMPLEX | 24 | 0.419955426 | 2.10587871 | 0.00173 | 0.00173 | 0.00442 | 172 |
| GOBP_MUSCLE_SYSTEM_PROCESS | 18 | -0.600863358 | -1.8969039 | 0.00175 | 0.00175 | 0.00444 | 107 |
| GOBP_ION_HOMEOSTASIS | 25 | -0.54693205 | -1.8712921 | 0.00185 | 0.00185 | 0.00469 | 119 |
| GOBP_TRANSMEMBRANE_TRANSPORT | 42 | -0.484041163 | -1.8366413 | 0.00195 | 0.00195 | 0.00491 | 39 |
| GOBP_NCRNA_METABOLIC_PROCESS | 17 | 0.470198691 | 2.05182206 | 0.00197 | 0.00197 | 0.00495 | 126 |
| GOCC_FICOLIN_1_RICH_GRANULE | 17 | -0.609995442 | -1.8987502 | 0.00201 | 0.00201 | 0.00503 | 104 |
| GOBP_REGULATION_OF_ORGANELLE_ORGANIZATION | 50 | -0.461202949 | -1.8055204 | 0.00203 | 0.00203 | 0.00506 | 58 |
| GOBP_RIBONUCLEOPROTEIN_COMPLEX_SUBUNIT_ORGANIZATION | 12 | 0.576641844 | 2.16229919 | 0.00203 | 0.00203 | 0.00506 | 164 |
| GOBP_MEMBRANE_ORGANIZATION | 40 | -0.485620557 | -1.8240452 | 0.00210 | 0.00210 | 0.00520 | 66 |
| GOBP_NEGATIVE_REGULATION_OF_RESPONSE_TO_EXTERNAL_STIMUL | 18 | -0.597287237 | -1.8856142 | 0.00220 | 0.00220 | 0.00543 | 107 |
| GOBP_RECEPTOR_MEDIATED_ENDOCYTOSIS | 18 | -0.59044858 | -1.8640248 | 0.00223 | 0.00223 | 0.00548 | 98 |
| GOBP_METAL_ION_HOMEOSTASIS | 21 | -0.568202142 | -1.8675647 | 0.00229 | 0.00229 | 0.00561 | 119 |
| GOBP_NEUTROPHIL_MIGRATION | 13 | -0.644076661 | -1.8674343 | 0.00235 | 0.00235 | 0.00573 | 136 |
| HP_ABNORMAL_CSF_METABOLITE_LEVEL | 18 | 0.468641403 | 2.10232375 | 0.00237 | 0.00237 | 0.00574 | 172 |
| GOBP_REGULATION_OF_SECRETION | 14 | -0.651741992 | -1.926033 | 0.00238 | 0.00238 | 0.00574 | 110 |
| GOBP_NEGATIVE_REGULATION_OF_SIGNALING | 32 | -0.509650312 | -1.841952 | 0.00238 | 0.00238 | 0.00574 | 119 |
| HP_INCREASED_CSF_LACTATE | 17 | 0.457792452 | 1.99768453 | 0.00243 | 0.00243 | 0.00583 | 172 |
| GOBP_REGULATION_OF_LEUKOCYTE_MIGRATION | 14 | -0.634818654 | -1.876021 | 0.00245 | 0.00245 | 0.00585 | 137 |
| GOBP_POSITIVE_REGULATION_OF_RESPONSE_TO_WOUNDING | 10 | -0.692828547 | -1.8629021 | 0.00247 | 0.00247 | 0.00588 | 28 |
| GOBP_REGULATION_OF_GTPASE_ACTIVITY | 11 | -0.667493797 | -1.8480148 | 0.00256 | 0.00256 | 0.00605 | 146 |
| GOBP_REGULATION_OF_METAL_ION_TRANSPORT | 11 | -0.665477704 | -1.842433 | 0.00256 | 0.00256 | 0.00605 | 131 |
| GOMF_MOLECULAR_TRANSDUCER_ACTIVITY | 18 | -0.588356485 | -1.8574202 | 0.00260 | 0.00260 | 0.00613 | 121 |
| GOCC_LAMELLIPODIUM | 12 | -0.657641443 | -1.8658099 | 0.00265 | 0.00265 | 0.00621 | 49 |
| GOCC_ENDOPLASMIC_RETICULUM_PROTEIN_CONTAINING_COMPLEX | 16 | 0.492462312 | 2.09269477 | 0.00268 | 0.00268 | 0.00623 | 218 |
| GOBP_ADAPTIVE_IMMUNE_RESPONSE | 15 | -0.627655729 | -1.8869792 | 0.00268 | 0.00268 | 0.00623 | 110 |
| HP_FUNCTIONAL_ABNORMALITY_OF_THE_INNER_EAR | 43 | -0.475628341 | -1.8129894 | 0.00268 | 0.00268 | 0.00623 | 42 |
| GOBP_AXON_DEVELOPMENT | 10 | -0.687798766 | -1.8493778 | 0.00274 | 0.00274 | 0.00634 | 49 |
| GOBP_REGULATION_OF_CELL_MORPHOGENESIS | 18 | -0.583907833 | -1.8433759 | 0.00285 | 0.00285 | 0.00656 | 84 |
| GOBP_REACTIVE_OXYGEN_SPECIES_METABOLIC_PROCESS | 17 | -0.597441381 | -1.8596728 | 0.00312 | 0.00312 | 0.00714 | 141 |
| HP_HYPERTRICHOSIS | 17 | 0.44510076 | 1.94230136 | 0.00312 | 0.00312 | 0.00714 | 171 |
| GOBP_LYMPHOCYTE_ACTIVATION | 25 | -0.537868179 | -1.8402807 | 0.00317 | 0.00317 | 0.00722 | 110 |
| GOBP_REGULATION_OF_DNA_BINDING_TRANSCRIPTION_FACTOR_AC | 15 | -0.620605504 | -1.8657835 | 0.00319 | 0.00319 | 0.00722 | 92 |
| GOCC_CENTROSOME | 12 | 0.556988849 | 2.08860413 | 0.00319 | 0.00319 | 0.00722 | 112 |
| GOBP_PHOSPHORYLATION | 47 | -0.458568486 | -1.773469 | 0.00323 | 0.00323 | 0.00728 | 112 |
| GOMF_KINASE_BINDING | 32 | -0.501461341 | -1.8123558 | 0.00329 | 0.00329 | 0.00739 | 105 |
| GOBP_CELL_PROJECTION_ASSEMBLY | 19 | -0.571945236 | -1.8300795 | 0.00331 | 0.00331 | 0.00741 | 150 |
| GOMF_KINASE_ACTIVITY | 18 | -0.580505133 | -1.8326337 | 0.00335 | 0.00335 | 0.00743 | 142 |
| GOBP_MAPK_CASCADE | 18 | -0.5803673 | -1.8321986 | 0.00335 | 0.00335 | 0.00743 | 110 |
| GOBP_RNA_LOCALIZATION | 13 | 0.532818877 | 2.07224814 | 0.00348 | 0.00348 | 0.00771 | 126 |
| GOCC_SPLICEOSOMAL_COMPLEX | 16 | 0.48241206 | 2.04998671 | 0.00352 | 0.00352 | 0.00775 | 222 |
| GOBP_ENZYME_LINKED_RECEPTOR_PROTEIN_SIGNALING_PATHWAY | 24 | -0.543309578 | -1.842619 | 0.00355 | 0.00355 | 0.00779 | 106 |
| GOBP_REGULATION_OF_CYSTEINE_TYPE_ENDOPEPTIDASE_ACTIVITY | 12 | -0.647375099 | -1.836683 | 0.00358 | 0.00358 | 0.00783 | 92 |
| HP_ABNORMAL_RENAL_PHYSIOLOGY | 36 | -0.486634428 | -1.7935362 | 0.00368 | 0.00368 | 0.00804 | 38 |
| GOBP_REGULATION_OF_ANATOMICAL_STRUCTURE_SIZE | 20 | -0.559564007 | -1.8159397 | 0.00378 | 0.00378 | 0.00822 | 153 |
| GOBP_RNA_CATABOLIC_PROCESS | 12 | 0.55011411 | 2.06282514 | 0.00386 | 0.00386 | 0.00837 | 174 |
| GOBP_NEGATIVE_REGULATION_OF_CELL_DEATH | 31 | -0.50448827 | -1.814772 | 0.00387 | 0.00387 | 0.00837 | 63 |
| GOCC_ADHERENS_JUNCTION | 15 | -0.613667218 | -1.8449242 | 0.00396 | 0.00396 | 0.00852 | 132 |
| GOCC_TERTIARY_GRANULE | 10 | -0.679663636 | -1.8275038 | 0.00398 | 0.00398 | 0.00853 | 105 |
| GOBP_NEGATIVE_REGULATION_OF_CELL_POPULATION_PROLIFERATIO | 16 | -0.603170313 | -1.8486906 | 0.00416 | 0.00416 | 0.00889 | 120 |
| HP_ABNORMALITY_OF_THE_MITOCHONDRION | 16 | 0.472361809 | 2.00727865 | 0.00427 | 0.00427 | 0.00909 | 226 |
| GOBP_POSITIVE_REGULATION_OF_PROTEIN_CONTAINING_COMPLEX_ | 13 | -0.629067188 | -1.8239159 | 0.00431 | 0.00431 | 0.00916 | 126 |
| GOBP_REGULATION_OF_PROTEIN_KINASE_ACTIVITY | 20 | -0.553557032 | -1.7964454 | 0.00463 | 0.00463 | 0.00980 | 112 |
| GOBP_NEGATIVE_REGULATION_OF_DEVELOPMENTAL_PROCESS | 16 | -0.599215548 | -1.8365695 | 0.00467 | 0.00467 | 0.00984 | 125 |

| | | | | | | | |
|---|---|---|---|---|---|---|---|
| GOBP_CELLULAR_RESPONSE_TO_ENDOGENOUS_STIMULUS | 30 | -0.4951897 | -1.7660789 | 0.00470 | 0.00470 | 0.00988 | 106 |
| GOBP_REGULATION_OF_MULTICELLULAR_ORGANISMAL_DEVELOPMEN | 27 | -0.510864384 | -1.7815223 | 0.00477 | 0.00477 | 0.00999 | 131 |
| GOBP_POSITIVE_REGULATION_OF_CELL_CELL_ADHESION | 16 | -0.598220797 | -1.8335206 | 0.00479 | 0.00479 | 0.01000 | 110 |
| GOBP_POSITIVE_REGULATION_OF_ORGANELLE_ORGANIZATION | 18 | -0.57407186 | -1.8123241 | 0.00483 | 0.00483 | 0.01006 | 53 |
| GOBP_NEGATIVE_REGULATION_OF_CELL_ADHESION | 14 | -0.614991902 | -1.8174289 | 0.00502 | 0.00502 | 0.01042 | 110 |
| GOBP_INORGANIC_ION_TRANSMEMBRANE_TRANSPORT | 25 | -0.526535022 | -1.801505 | 0.00505 | 0.00505 | 0.01044 | 131 |
| GOBP_CELLULAR_ION_HOMEOSTASIS | 21 | -0.5483324 | -1.8022569 | 0.00507 | 0.00507 | 0.01045 | 45 |
| GOCC_SOMATODENDRITIC_COMPARTMENT | 15 | -0.6029974 | -1.8128466 | 0.00523 | 0.00523 | 0.01075 | 64 |
| GOCC_LEADING_EDGE_MEMBRANE | 10 | -0.66836248 | -1.7971168 | 0.00535 | 0.00535 | 0.01095 | 109 |
| GOCC_VACUOLE | 39 | -0.465531543 | -1.7403482 | 0.00543 | 0.00543 | 0.01108 | 118 |
| GOMF_OXIDOREDUCTASE_ACTIVITY_ACTING_ON_NAD_P_H | 20 | 0.421196145 | 1.96669875 | 0.00547 | 0.00547 | 0.01113 | 172 |
| GOBP_REGULATION_OF_APOPTOTIC_SIGNALING_PATHWAY | 22 | -0.537027017 | -1.7822198 | 0.00564 | 0.00564 | 0.01144 | 58 |
| HP_INTELLECTUAL_DISABILITY_SEVERE | 21 | 0.408631687 | 1.94289541 | 0.00573 | 0.00573 | 0.01153 | 224 |
| HP_LEUKODYSTROPHY | 15 | 0.500619275 | 2.07856104 | 0.00573 | 0.00573 | 0.01153 | 171 |
| GOBP_ACTIN_POLYMERIZATION_OR_DEPOLYMERIZATION | 15 | -0.601430661 | -1.8081364 | 0.00574 | 0.00574 | 0.01153 | 153 |
| GOBP_NEGATIVE_REGULATION_OF_CELL_DIFFERENTIATION | 11 | -0.648677941 | -1.7959214 | 0.00580 | 0.00580 | 0.01160 | 125 |
| GOBP_NEGATIVE_REGULATION_OF_INTRACELLULAR_SIGNAL_TRANSDU | 19 | -0.560363473 | -1.7930208 | 0.00589 | 0.00589 | 0.01175 | 119 |
| HP_DIARRHEA | 15 | -0.598463589 | -1.7992162 | 0.00600 | 0.00600 | 0.01193 | 136 |
| GOBP_REGULATION_OF_IMMUNE_EFFECTOR_PROCESS | 10 | -0.664063885 | -1.7855586 | 0.00604 | 0.00604 | 0.01194 | 134 |
| HP_GENERALIZED_HYPOTONIA | 35 | 0.311943625 | 1.79528385 | 0.00607 | 0.00607 | 0.01194 | 161 |
| GOMF_ACTIN_FILAMENT_BINDING | 18 | -0.566632391 | -1.788838 | 0.00607 | 0.00607 | 0.01194 | 75 |
| GOBP_POSITIVE_REGULATION_OF_GENE_EXPRESSION | 34 | -0.477648814 | -1.7428556 | 0.00608 | 0.00608 | 0.01194 | 98 |
| GOBP_ERK1_AND_ERK2_CASCADE | 11 | -0.644745063 | -1.7850329 | 0.00634 | 0.00634 | 0.01236 | 110 |
| GOBP_REGULATION_OF_PROTEIN_SERINE_THREONINE_KINASE_ACTIV | 11 | -0.644352926 | -1.7839473 | 0.00634 | 0.00634 | 0.01236 | 106 |
| GOBP_DNA_REPAIR | 15 | 0.493374544 | 2.04848106 | 0.00658 | 0.00658 | 0.01280 | 131 |
| GOBP_ENDOTHELIAL_CELL_MIGRATION | 10 | -0.658958157 | -1.7718302 | 0.00686 | 0.00686 | 0.01330 | 47 |
| GOBP_EXPORT_FROM_CELL | 23 | -0.526794739 | -1.7704557 | 0.00690 | 0.00690 | 0.01333 | 106 |
| GOBP_NCRNA_PROCESSING | 15 | 0.491276013 | 2.039768 | 0.00692 | 0.00692 | 0.01334 | 126 |
| GOBP_NEUTROPHIL_CHEMOTAXIS | 11 | -0.639749894 | -1.7712034 | 0.00701 | 0.00701 | 0.01347 | 133 |
| GOCC_NUCLEAR_OUTER_MEMBRANE_ENDOPLASMIC_RETICULUM_M | 47 | 0.279552037 | 1.77476516 | 0.00716 | 0.00716 | 0.01369 | 208 |
| GOBP_VIRAL_LIFE_CYCLE | 13 | -0.615842891 | -1.7855734 | 0.00719 | 0.00719 | 0.01369 | 57 |
| GOBP_MUSCLE_STRUCTURE_DEVELOPMENT | 20 | -0.542396667 | -1.7602269 | 0.00719 | 0.00719 | 0.01369 | 153 |
| GOBP_REGULATION_OF_ENDOCYTOSIS | 13 | -0.615204738 | -1.7837231 | 0.00745 | 0.00745 | 0.01414 | 108 |
| GOBP_POSITIVE_REGULATION_OF_CELL_DIFFERENTIATION | 24 | -0.518293095 | -1.7577764 | 0.00756 | 0.00756 | 0.01431 | 110 |
| GOBP_POSITIVE_REGULATION_OF_SUPRAMOLECULAR_FIBER_ORGANI | 10 | -0.655940594 | -1.7637164 | 0.00768 | 0.00768 | 0.01450 | 150 |
| GOCC_VACUOLAR_LUMEN | 16 | -0.579789036 | -1.7770281 | 0.00782 | 0.00782 | 0.01470 | 118 |
| GOBP_PROTEIN_LOCALIZATION_TO_ENDOPLASMIC_RETICULUM | 11 | 0.544341806 | 1.97234769 | 0.00811 | 0.00811 | 0.01522 | 150 |
| HP_ABNORMAL_CNS_MYELINATION | 25 | 0.369518869 | 1.86720745 | 0.00832 | 0.00832 | 0.01555 | 177 |
| GOCC_ENDOCYTIC_VESICLE_MEMBRANE | 11 | -0.635047803 | -1.7581852 | 0.00836 | 0.00836 | 0.01555 | 110 |
| GOBP_REGULATION_OF_MEMBRANE_POTENTIAL | 13 | -0.608847732 | -1.7652916 | 0.00837 | 0.00837 | 0.01555 | 136 |
| GOBP_POSITIVE_REGULATION_OF_RESPONSE_TO_EXTERNAL_STIMULU | 22 | -0.529440633 | -1.7570431 | 0.00840 | 0.00840 | 0.01557 | 110 |
| HP_ABNORMAL_ACTIVITY_OF_MITOCHONDRIAL_RESPIRATORY_CHAIN | 14 | 0.486560713 | 1.95521431 | 0.00849 | 0.00849 | 0.01563 | 172 |
| GOCC_RIBOSOME | 14 | 0.484460511 | 1.94677477 | 0.00849 | 0.00849 | 0.01563 | 132 |
| GOBP_CELLULAR_RESPONSE_TO_OXYGEN_CONTAINING_COMPOUND | 33 | -0.469653046 | -1.7061309 | 0.00870 | 0.00870 | 0.01598 | 60 |
| GOBP_NEGATIVE_REGULATION_OF_MOLECULAR_FUNCTION | 32 | -0.472627894 | -1.7081474 | 0.00874 | 0.00874 | 0.01601 | 112 |
| GOBP_CHROMATIN_ORGANIZATION | 12 | 0.519352113 | 1.9474734 | 0.00896 | 0.00896 | 0.01637 | 129 |
| GOMF_CADHERIN_BINDING | 37 | -0.461389247 | -1.7088822 | 0.00912 | 0.00912 | 0.01660 | 76 |
| GOBP_REGULATION_OF_ACTIN_FILAMENT_LENGTH | 14 | -0.596351724 | -1.7623433 | 0.00914 | 0.00914 | 0.01660 | 153 |
| GOBP_CATION_TRANSMEMBRANE_TRANSPORT | 30 | -0.477975702 | -1.7046857 | 0.00928 | 0.00928 | 0.01677 | 131 |
| GOBP_CALCIUM_ION_TRANSPORT | 15 | -0.582647395 | -1.7516665 | 0.00932 | 0.00932 | 0.01677 | 131 |
| GOBP_REGULATION_OF_PEPTIDASE_ACTIVITY | 15 | -0.582475144 | -1.7511486 | 0.00932 | 0.00932 | 0.01677 | 106 |
| GOMF_CATALYTIC_ACTIVITY_ACTING_ON_A_NUCLEIC_ACID | 13 | 0.496938583 | 1.9327019 | 0.00977 | 0.00977 | 0.01750 | 173 |
| HP_ABNORMAL_INVOLUNTARY_EYE_MOVEMENTS | 34 | 0.312585421 | 1.77595462 | 0.00978 | 0.00978 | 0.01750 | 177 |
| GOBP_SIGNAL_RELEASE | 10 | -0.645942779 | -1.7368339 | 0.00988 | 0.00988 | 0.01763 | 106 |
| GOBP_POSITIVE_REGULATION_OF_PROTEIN_METABOLIC_PROCESS | 46 | -0.431160584 | -1.6611293 | 0.00995 | 0.00995 | 0.01770 | 110 |
| HP_ABNORMAL_ADIPOSE_TISSUE_MORPHOLOGY | 11 | -0.625419135 | -1.7315274 | 0.00998 | 0.00998 | 0.01770 | 42 |
| HP_FUNCTIONAL_ABNORMALITY_OF_THE_GASTROINTESTINAL_TRACT | 52 | -0.420839967 | -1.6544446 | 0.01024 | 0.01024 | 0.01812 | 52 |
| GOBP_POSITIVE_REGULATION_OF_PEPTIDASE_ACTIVITY | 10 | -0.641110978 | -1.723842 | 0.01043 | 0.01043 | 0.01840 | 106 |
| HP_GLUCOSE_INTOLERANCE | 21 | 0.377436753 | 1.79457482 | 0.01047 | 0.01047 | 0.01841 | 161 |
| GOCC_MEMBRANE_COAT | 13 | 0.485542514 | 1.88838011 | 0.01062 | 0.01062 | 0.01863 | 206 |
| GOBP_REGULATION_OF_PROTEIN_LOCALIZATION | 31 | -0.469466272 | -1.6887891 | 0.01071 | 0.01071 | 0.01870 | 28 |
| GOBP_MUSCLE_CELL_DIFFERENTIATION | 13 | -0.598315813 | -1.7347554 | 0.01072 | 0.01072 | 0.01870 | 153 |
| HP_ABNORMALITY_OF_THE_GINGIVA | 20 | -0.532057992 | -1.726675 | 0.01097 | 0.01097 | 0.01907 | 45 |
| GOBP_REGULATION_OF_CATION_TRANSMEMBRANE_TRANSPORT | 12 | -0.611155416 | -1.7339233 | 0.01099 | 0.01099 | 0.01907 | 131 |
| GOBP_REGULATION_OF_CELLULAR_COMPONENT_SIZE | 16 | -0.568108319 | -1.7412272 | 0.01122 | 0.01122 | 0.01942 | 153 |
| GOBP_HEART_DEVELOPMENT | 10 | -0.638192134 | -1.7159937 | 0.01166 | 0.01166 | 0.02012 | 69 |
| HP_ABNORMALITY_OF_RETINAL_PIGMENTATION | 13 | 0.473815461 | 1.84277107 | 0.01189 | 0.01189 | 0.02041 | 224 |
| HP_POOR_EYE_CONTACT | 13 | 0.473815461 | 1.84277107 | 0.01189 | 0.01189 | 0.02041 | 224 |
| GOCC_PRESYNAPSE | 10 | -0.637509236 | -1.7141575 | 0.01207 | 0.01207 | 0.02066 | 66 |
| HP_REGIONAL_ABNORMALITY_OF_SKIN | 16 | -0.565844839 | -1.7342897 | 0.01210 | 0.01210 | 0.02066 | 42 |
| GOBP_REGULATION_OF_TRANSMEMBRANE_TRANSPORT | 16 | -0.565013722 | -1.7317424 | 0.01236 | 0.01236 | 0.02103 | 136 |
| GOBP_REGULATION_OF_MRNA_PROCESSING | 14 | 0.465 | 1.86857391 | 0.01251 | 0.01251 | 0.02111 | 228 |
| GOBP_REGULATION_OF_RNA_SPLICING | 14 | 0.465 | 1.86857391 | 0.01251 | 0.01251 | 0.02111 | 228 |
| GOBP_ENDOPLASMIC_RETICULUM_TO_GOLGI_VESICLE_MEDIATED_TF | 14 | 0.46 | 1.84848172 | 0.01251 | 0.01251 | 0.02111 | 230 |
| GOBP_MITOCHONDRIAL_RESPIRATORY_CHAIN_COMPLEX_ASSEMBLY | 17 | 0.404660709 | 1.76583173 | 0.01263 | 0.01263 | 0.02119 | 224 |

| | | | | | | | |
|---|---|---|---|---|---|---|---|
| GOBP_NADH_DEHYDROGENASE_COMPLEX_ASSEMBLY | 17 | 0.404660709 | 1.76583173 | 0.01263 | 0.01263 | 0.02119 | 224 |
| HP_ABNORMALITY_OF_THE_URINARY_SYSTEM_PHYSIOLOGY | 43 | -0.43309765 | -1.6508718 | 0.01265 | 0.01265 | 0.02119 | 78 |
| GOBP_CELL_GROWTH | 12 | -0.603552447 | -1.7123527 | 0.01271 | 0.01271 | 0.02121 | 49 |
| GOCC_AXON | 16 | -0.563159664 | -1.7260598 | 0.01273 | 0.01273 | 0.02121 | 49 |
| GOBP_SMALL_GTPASE_MEDIATED_SIGNAL_TRANSDUCTION | 16 | -0.562032679 | -1.7226056 | 0.01311 | 0.01311 | 0.02176 | 146 |
| HP_ABNORMAL_SCALP_MORPHOLOGY | 13 | 0.469369351 | 1.82547918 | 0.01317 | 0.01317 | 0.02176 | 198 |
| HP_LETHARGY | 13 | 0.46882793 | 1.82337347 | 0.01317 | 0.01317 | 0.02176 | 226 |
| GOCC_NEURON_PROJECTION | 33 | -0.459461722 | -1.6691084 | 0.01334 | 0.01334 | 0.02198 | 67 |
| GOBP_REGULATION_OF_COAGULATION | 10 | -0.633467462 | -1.7032899 | 0.01358 | 0.01358 | 0.02226 | 107 |
| GOBP_TRANSMEMBRANE_RECEPTOR_PROTEIN_SERINE_THREONINE_ | 10 | -0.633135801 | -1.7023981 | 0.01358 | 0.01358 | 0.02226 | 74 |
| GOBP_MRNA_CATABOLIC_PROCESS | 10 | 0.53875521 | 1.86667975 | 0.01361 | 0.01361 | 0.02226 | 174 |
| GOBP_NEGATIVE_REGULATION_OF_APOPTOTIC_SIGNALING_PATHWAY | 15 | -0.569876597 | -1.7132725 | 0.01366 | 0.01366 | 0.02228 | 58 |
| GOCC_TRANSCRIPTION_REGULATOR_COMPLEX | 14 | 0.445750045 | 1.79121915 | 0.01385 | 0.01385 | 0.02252 | 131 |
| HP_ABNORMALITY_OF_BONE_MARROW_CELL_MORPHOLOGY | 11 | -0.618237341 | -1.711644 | 0.01402 | 0.01402 | 0.02275 | 96 |
| GOCC_FICOLIN_1_RICH_GRANULE_LUMEN | 13 | -0.589600057 | -1.709485 | 0.01438 | 0.01438 | 0.02322 | 123 |
| GOBP_CELLULAR_COMPONENT_MORPHOGENESIS | 20 | -0.524299452 | -1.7014964 | 0.01438 | 0.01438 | 0.02322 | 53 |
| GOBP_PROGRAMMED_CELL_DEATH | 67 | -0.392775078 | -1.5943661 | 0.01463 | 0.01463 | 0.02351 | 110 |
| GOMF_PHOSPHOLIPID_BINDING | 13 | -0.588953583 | -1.7076106 | 0.01464 | 0.01464 | 0.02351 | 59 |
| GOBP_NERVOUS_SYSTEM_PROCESS | 10 | -0.6290508 | -1.6914142 | 0.01468 | 0.01468 | 0.02352 | 56 |
| GOBP_REGULATION_OF_MRNA_SPLICING_VIA_SPLICEOSOME | 13 | 0.463840399 | 1.80397588 | 0.01487 | 0.01487 | 0.02376 | 228 |
| GOBP_ACTIN_FILAMENT_POLYMERIZATION | 13 | -0.58609806 | -1.6993313 | 0.01516 | 0.01516 | 0.02417 | 153 |
| GOMF_PROTEIN_KINASE_ACTIVITY | 12 | -0.598182806 | -1.6971184 | 0.01536 | 0.01536 | 0.02442 | 107 |
| GOBP_CALCIUM_ION_TRANSMEMBRANE_TRANSPORT | 14 | -0.57260109 | -1.6921552 | 0.01546 | 0.01546 | 0.02451 | 131 |
| GOBP_NON_MEMBRANE_BOUNDED_ORGANELLE_ASSEMBLY | 17 | -0.54335951 | -1.6913306 | 0.01570 | 0.01570 | 0.02484 | 53 |
| GOMF_PROTEIN_CONTAINING_COMPLEX_BINDING | 82 | -0.372300583 | -1.5450456 | 0.01580 | 0.01580 | 0.02493 | 112 |
| GOBP_REGULATION_OF_CHEMOTAXIS | 13 | -0.584731525 | -1.6953691 | 0.01595 | 0.01595 | 0.02510 | 133 |
| GOBP_TISSUE_DEVELOPMENT | 44 | -0.425277251 | -1.6249549 | 0.01614 | 0.01614 | 0.02526 | 141 |
| GOBP_POSITIVE_REGULATION_OF_LEUKOCYTE_CELL_CELL_ADHESION | 12 | -0.595228526 | -1.6887367 | 0.01616 | 0.01616 | 0.02526 | 110 |
| GOBP_REGULATION_OF_DEFENSE_RESPONSE | 23 | -0.499772095 | -1.6796378 | 0.01617 | 0.01617 | 0.02526 | 53 |
| GOBP_REGULATION_OF_ION_TRANSMEMBRANE_TRANSPORT | 14 | -0.570520153 | -1.6860056 | 0.01623 | 0.01623 | 0.02528 | 136 |
| GOBP_POSITIVE_REGULATION_OF_TRANSFERASE_ACTIVITY | 20 | -0.519742641 | -1.6867083 | 0.01634 | 0.01634 | 0.02536 | 110 |
| GOCC_MITOCHONDRIAL_PROTEIN_CONTAINING_COMPLEX | 27 | 0.333094695 | 1.74012354 | 0.01636 | 0.01636 | 0.02536 | 234 |
| GOCC_MICROTUBULE_CYTOSKELETON | 23 | 0.346790023 | 1.712662 | 0.01694 | 0.01694 | 0.02619 | 118 |
| GOBP_TRANSMEMBRANE_RECEPTOR_PROTEIN_TYROSINE_KINASE_SI | 15 | -0.560216627 | -1.6842308 | 0.01698 | 0.01698 | 0.02619 | 106 |
| GOCC_PERINUCLEAR_REGION_OF_CYTOPLASM | 26 | -0.478990168 | -1.654222 | 0.01730 | 0.01730 | 0.02663 | 106 |
| GOBP_RESPONSE_TO_ORGANIC_CYCLIC_COMPOUND | 29 | -0.463438017 | -1.6385019 | 0.01788 | 0.01788 | 0.02745 | 58 |
| GOBP_NEGATIVE_REGULATION_OF_IMMUNE_SYSTEM_PROCESS | 16 | -0.550947196 | -1.6886291 | 0.01816 | 0.01816 | 0.02780 | 110 |
| GOBP_NEUROGENESIS | 37 | -0.437588844 | -1.6207309 | 0.01825 | 0.01825 | 0.02787 | 58 |
| GOBP_REGULATION_OF_PROTEIN_POLYMERIZATION | 14 | -0.567284104 | -1.6764424 | 0.01842 | 0.01842 | 0.02807 | 153 |
| GOBP_ESTABLISHMENT_OF_RNA_LOCALIZATION | 11 | 0.498451166 | 1.80606927 | 0.01855 | 0.01855 | 0.02813 | 153 |
| GOBP_NUCLEOBASE_CONTAINING_COMPOUND_TRANSPORT | 11 | 0.498451166 | 1.80606927 | 0.01855 | 0.01855 | 0.02813 | 153 |
| GOCC_CELL_CORTEX | 19 | -0.520190845 | -1.6644786 | 0.01890 | 0.01890 | 0.02860 | 64 |
| GOBP_RESPONSE_TO_ABIOTIC_STIMULUS | 27 | -0.472411211 | -1.6474256 | 0.01896 | 0.01896 | 0.02861 | 86 |
| GOBP_REGULATION_OF_PROTEIN_MODIFICATION_PROCESS | 41 | -0.424347162 | -1.6034421 | 0.01955 | 0.01955 | 0.02943 | 112 |
| HP_CONGESTIVE_HEART_FAILURE | 11 | -0.604937763 | -1.6748229 | 0.01968 | 0.01968 | 0.02956 | 46 |
| GOBP_ATP_SYNTHESIS_COUPLED_ELECTRON_TRANSPORT | 21 | 0.35373669 | 1.68188962 | 0.01977 | 0.01977 | 0.02962 | 172 |
| GOBP_ORGANIC_ACID_TRANSPORT | 10 | -0.618357318 | -1.6626612 | 0.01990 | 0.01990 | 0.02974 | 33 |
| HP_OPHTHALMOPARESIS | 15 | 0.411918403 | 1.71027682 | 0.02030 | 0.02030 | 0.03026 | 171 |
| GOCC_AZUROPHIL_GRANULE_LUMEN | 13 | -0.574189765 | -1.6648044 | 0.02052 | 0.02052 | 0.03053 | 118 |
| GOCC_ENDOSOME | 20 | -0.511612369 | -1.6603234 | 0.02097 | 0.02097 | 0.03112 | 71 |
| HP_MUSCLE_ABNORMALITY_RELATED_TO_MITOCHONDRIAL_DYSFUN | 11 | 0.482313935 | 1.74759823 | 0.02125 | 0.02125 | 0.03147 | 161 |
| HP_HYPOPLASIA_OF_PENIS | 15 | 0.406015038 | 1.68576617 | 0.02168 | 0.02168 | 0.03196 | 252 |
| GOBP_PROTEIN_POLYMERIZATION | 16 | -0.542561781 | -1.6629281 | 0.02169 | 0.02169 | 0.03196 | 153 |
| GOBP_ANIMAL_ORGAN_MORPHOGENESIS | 10 | -0.613976918 | -1.650883 | 0.02182 | 0.02182 | 0.03208 | 92 |
| GOMF_RIBONUCLEOPROTEIN_COMPLEX_BINDING | 13 | 0.436962281 | 1.69944106 | 0.02209 | 0.02209 | 0.03240 | 170 |
| GOCC_CATALYTIC_STEP_2_SPLICEOSOME | 11 | 0.476426799 | 1.726267 | 0.02280 | 0.02280 | 0.03325 | 222 |
| GOBP_MULTICELLULAR_ORGANISMAL_HOMEOSTASIS | 16 | -0.539986984 | -1.6550365 | 0.02282 | 0.02282 | 0.03325 | 63 |
| GOBP_ION_TRANSMEMBRANE_TRANSPORT | 33 | -0.442837069 | -1.6087152 | 0.02283 | 0.02283 | 0.03325 | 136 |
| GOBP_CELLULAR_RESPONSE_TO_REACTIVE_OXYGEN_SPECIES | 11 | -0.596830228 | -1.6523765 | 0.02305 | 0.02305 | 0.03346 | 86 |
| GOBP_RAS_PROTEIN_SIGNAL_TRANSDUCTION | 13 | -0.569120829 | -1.6501075 | 0.02314 | 0.02314 | 0.03346 | 144 |
| GOCC_VESICLE_COAT | 11 | 0.474755091 | 1.7202098 | 0.02318 | 0.02318 | 0.03346 | 206 |
| GOCC_RIBOSOMAL_SUBUNIT | 11 | 0.474068065 | 1.71772045 | 0.02318 | 0.02318 | 0.03346 | 116 |
| HP_DECREASED_ACTIVITY_OF_MITOCHONDRIAL_COMPLEX_I | 11 | 0.47146402 | 1.70828505 | 0.02396 | 0.02396 | 0.03449 | 224 |
| GOMF_ACTIVE_TRANSMEMBRANE_TRANSPORTER_ACTIVITY | 24 | 0.322978655 | 1.61958586 | 0.02401 | 0.02401 | 0.03449 | 172 |
| GOBP_REGULATION_OF_CELLULAR_LOCALIZATION | 34 | -0.435706435 | -1.5898153 | 0.02454 | 0.02454 | 0.03518 | 106 |
| GOBP_RESPONSE_TO_INORGANIC_SUBSTANCE | 25 | -0.47182011 | -1.6143016 | 0.02468 | 0.02468 | 0.03529 | 56 |
| GOBP_POSITIVE_REGULATION_OF_CELL_PROJECTION_ORGANIZATION | 12 | -0.576429344 | -1.6354011 | 0.02490 | 0.02490 | 0.03552 | 146 |
| GOBP_CELL_CYCLE_PROCESS | 28 | 0.307355524 | 1.62522521 | 0.02502 | 0.02502 | 0.03561 | 118 |
| HP_ABNORMAL_SKIN_MORPHOLOGY_OF_THE_PALM | 12 | -0.57563818 | -1.6331565 | 0.02529 | 0.02529 | 0.03592 | 42 |
| HP_LOWER_LIMB_SPASTICITY | 10 | 0.500873936 | 1.73542866 | 0.02575 | 0.02575 | 0.03649 | 171 |
| GOBP_POSITIVE_REGULATION_OF_IMMUNE_RESPONSE | 27 | -0.459627333 | -1.6028448 | 0.02629 | 0.02629 | 0.03717 | 110 |
| GOBP_ORGANIC_ANION_TRANSPORT | 10 | -0.60434772 | -1.6249917 | 0.02648 | 0.02648 | 0.03730 | 92 |
| HP_VISUAL_IMPAIRMENT | 42 | -0.413954729 | -1.570706 | 0.02650 | 0.02650 | 0.03730 | 38 |
| HP_ABNORMALITY_OF_THE_PALM | 15 | -0.54317964 | -1.633011 | 0.02668 | 0.02668 | 0.03746 | 46 |
| HP_PIGMENTARY_RETINOPATHY | 10 | 0.493948991 | 1.7114351 | 0.02686 | 0.02686 | 0.03763 | 171 |

| | | | | | | | |
|---|---|---|---|---|---|---|---|
| GOCC_TRANSFERASE_COMPLEX | 16 | 0.385024309 | 1.63614217 | 0.02704 | 0.02704 | 0.03780 | 103 |
| GOBP_NEGATIVE_REGULATION_OF_CELL_CELL_ADHESION | 10 | -0.602168797 | -1.6191329 | 0.02744 | 0.02744 | 0.03828 | 110 |
| HP_MOTOR_SEIZURE | 12 | 0.447831699 | 1.67928521 | 0.02771 | 0.02771 | 0.03857 | 66 |
| GOBP_PEPTIDE_BIOSYNTHETIC_PROCESS | 33 | 0.278942113 | 1.57239832 | 0.02860 | 0.02860 | 0.03963 | 132 |
| HP_ABNORMAL_BRAINSTEM_MORPHOLOGY | 15 | 0.393522339 | 1.63389674 | 0.02860 | 0.02860 | 0.03963 | 161 |
| HP_ABNORMAL_ORAL_MUCOSA_MORPHOLOGY | 23 | -0.479307206 | -1.6108593 | 0.02961 | 0.02961 | 0.04094 | 116 |
| GOBP_ORGANONITROGEN_COMPOUND_BIOSYNTHETIC_PROCESS | 53 | 0.237265433 | 1.55025726 | 0.02978 | 0.02978 | 0.04108 | 100 |
| HP_ABNORMAL_INFLAMMATORY_RESPONSE | 41 | -0.412454579 | -1.5585047 | 0.03009 | 0.03009 | 0.04142 | 116 |
| GOBP_SYNAPSE_ORGANIZATION | 10 | -0.596534653 | -1.6039836 | 0.03060 | 0.03060 | 0.04203 | 174 |
| GOBP_PROTEIN_MODIFICATION_BY_SMALL_PROTEIN_CONJUGATION_ | 17 | 0.367027183 | 1.60160903 | 0.03082 | 0.03082 | 0.04213 | 211 |
| HP_POOR_HEAD_CONTROL | 17 | 0.366395647 | 1.59885318 | 0.03082 | 0.03082 | 0.04213 | 238 |
| GOMF_GTPASE_ACTIVITY | 10 | -0.596282794 | -1.6033064 | 0.03087 | 0.03087 | 0.04213 | 135 |
| HP_VENTRICULOMEGALY | 24 | 0.310194832 | 1.55548101 | 0.03115 | 0.03115 | 0.04241 | 263 |
| HP_ABNORMAL_CIRCULATING_CARBOHYDRATE_CONCENTRATION | 13 | 0.418615818 | 1.62808768 | 0.03229 | 0.03229 | 0.04377 | 161 |
| HP_INCREASED_SERUM_PYRUVATE | 13 | 0.418615818 | 1.62808768 | 0.03229 | 0.03229 | 0.04377 | 161 |
| HP_REDUCED_VISUAL_ACUITY | 27 | -0.452024573 | -1.5763319 | 0.03257 | 0.03257 | 0.04406 | 38 |
| HP_PARAPLEGIA_PARAPARESIS | 10 | 0.483663654 | 1.67579845 | 0.03274 | 0.03274 | 0.04420 | 174 |
| GOBP_ADAPTIVE_IMMUNE_RESPONSE_BASED_ON_SOMATIC_RECOMI | 11 | -0.580597868 | -1.6074358 | 0.03343 | 0.03343 | 0.04503 | 110 |
| HP_ABNORMALITY_OF_ACID_BASE_HOMEOSTASIS | 23 | 0.320369841 | 1.58218292 | 0.03513 | 0.03513 | 0.04722 | 235 |
| HP_ABNORMALITY_OF_SKIN_PHYSIOLOGY | 19 | -0.494626938 | -1.5826806 | 0.03523 | 0.03523 | 0.04725 | 96 |
| GOBP_RRNA_METABOLIC_PROCESS | 10 | 0.477822966 | 1.65556163 | 0.03569 | 0.03569 | 0.04776 | 126 |
| GOBP_IMMUNE_SYSTEM_DEVELOPMENT | 29 | -0.440757758 | -1.558315 | 0.03577 | 0.03577 | 0.04777 | 137 |
| GOBP_POSITIVE_REGULATION_OF_CELLULAR_COMPONENT_BIOGENE | 20 | -0.488852076 | -1.5864599 | 0.03669 | 0.03669 | 0.04890 | 150 |
| GOBP_B_CELL_ACTIVATION | 12 | -0.560286826 | -1.5896028 | 0.03682 | 0.03682 | 0.04896 | 110 |
| GOMF_GUANYL_NUCLEOTIDE_BINDING | 18 | -0.501127719 | -1.5820421 | 0.03717 | 0.03717 | 0.04933 | 112 |
| GOBP_REGULATION_OF_LYMPHOCYTE_ACTIVATION | 17 | -0.509394663 | -1.5856072 | 0.03727 | 0.03727 | 0.04935 | 110 |
| GOBP_IMMUNE_RESPONSE_REGULATING_CELL_SURFACE_RECEPTOR | 15 | -0.524761375 | -1.5776384 | 0.03740 | 0.03740 | 0.04942 | 106 |
| HP_ABNORMALITY_OF_THE_OUTER_EAR | 36 | 0.26671142 | 1.5542598 | 0.03824 | 0.03824 | 0.05040 | 86 |
| GOBP_ESTABLISHMENT_OR_MAINTENANCE_OF_CELL_POLARITY | 13 | -0.544836082 | -1.5796964 | 0.03830 | 0.03830 | 0.05040 | 28 |
| HP_ABNORMAL_GLUCOSE_HOMEOSTASIS | 25 | 0.307772887 | 1.55520022 | 0.03885 | 0.03885 | 0.05101 | 161 |
| GOBP_VIRAL_PROCESS | 19 | -0.490778571 | -1.5703668 | 0.03904 | 0.03904 | 0.05115 | 57 |
| HP_ABNORMALITY_OF_THE_LYMPHATIC_SYSTEM | 23 | -0.466809157 | -1.5688557 | 0.03948 | 0.03948 | 0.05162 | 116 |
| GOBP_REGULATION_OF_IMMUNE_RESPONSE | 34 | -0.419432264 | -1.5304338 | 0.04019 | 0.04019 | 0.05232 | 110 |
| HP_COGNITIVE_IMPAIRMENT | 15 | -0.520957697 | -1.5662031 | 0.04021 | 0.04021 | 0.05232 | 33 |
| GOBP_GLIOGENESIS | 13 | -0.541833791 | -1.5709916 | 0.04026 | 0.04026 | 0.05232 | 58 |
| GOBP_NEGATIVE_REGULATION_OF_SUPRAMOLECULAR_FIBER_ORGAN | 11 | -0.570749757 | -1.5801705 | 0.04044 | 0.04044 | 0.05244 | 153 |
| HP_LEUKOENCEPHALOPATHY | 14 | 0.386956312 | 1.55496015 | 0.04064 | 0.04064 | 0.05244 | 161 |
| HP_ABNORMAL_VENTRICULAR_SEPTUM_MORPHOLOGY | 22 | 0.321378207 | 1.56312164 | 0.04067 | 0.04067 | 0.05244 | 171 |
| GOCC_OXIDOREDUCTASE_COMPLEX | 30 | 0.277857624 | 1.51108573 | 0.04069 | 0.04069 | 0.05244 | 172 |
| GOBP_CELLULAR_NITROGEN_COMPOUND_CATABOLIC_PROCESS | 16 | 0.365788152 | 1.55439905 | 0.04104 | 0.04104 | 0.05278 | 174 |
| GOCC_SYNAPSE | 39 | -0.404492498 | -1.5121592 | 0.04167 | 0.04167 | 0.05307 | 150 |
| HP_ABNORMAL_BRAINSTEM_MRI_SIGNAL_INTENSITY | 10 | 0.47029703 | 1.62948576 | 0.04194 | 0.04194 | 0.05307 | 224 |
| HP_ABNORMAL_MITOCHONDRIA_IN_MUSCLE_TISSUE | 10 | 0.47029703 | 1.62948576 | 0.04194 | 0.04194 | 0.05307 | 224 |
| HP_FETAL_DISTRESS | 10 | 0.47029703 | 1.62948576 | 0.04194 | 0.04194 | 0.05307 | 224 |
| HP_FOCAL_T2_HYPERINTENSE_BRAINSTEM_LESION | 10 | 0.47029703 | 1.62948576 | 0.04194 | 0.04194 | 0.05307 | 224 |
| HP_MITOCHONDRIAL_MYOPATHY | 10 | 0.47029703 | 1.62948576 | 0.04194 | 0.04194 | 0.05307 | 224 |
| HP_PAROXYSMAL_INVOLUNTARY_EYE_MOVEMENTS | 10 | 0.47029703 | 1.62948576 | 0.04194 | 0.04194 | 0.05307 | 224 |
| HP_PROXIMAL_TUBULOPATHY | 10 | 0.47029703 | 1.62948576 | 0.04194 | 0.04194 | 0.05307 | 224 |
| GOMF_ELECTRON_TRANSFER_ACTIVITY | 20 | 0.333448945 | 1.55697917 | 0.04225 | 0.04225 | 0.05335 | 172 |
| GOBP_POSITIVE_REGULATION_OF_PROTEOLYSIS | 17 | -0.502767212 | -1.5649778 | 0.04275 | 0.04275 | 0.05388 | 106 |
| GOBP_REGULATION_OF_CELL_CYCLE_PROCESS | 16 | 0.362779373 | 1.54161339 | 0.04297 | 0.04297 | 0.05405 | 131 |
| GOBP_CELLULAR_RESPONSE_TO_MOLECULE_OF_BACTERIAL_ORIGIN | 10 | -0.580265982 | -1.5602398 | 0.04308 | 0.04308 | 0.05408 | 60 |
| GOBP_POSITIVE_REGULATION_OF_TRANSCRIPTION_BY_RNA_POLYMEI | 22 | 0.316379763 | 1.53881017 | 0.04486 | 0.04486 | 0.05609 | 131 |
| GOBP_STRIATED_MUSCLE_CELL_DIFFERENTIATION | 10 | -0.578002615 | -1.554154 | 0.04487 | 0.04487 | 0.05609 | 153 |
| HP_ABNORMAL_SOCIAL_BEHAVIOR | 17 | 0.356758539 | 1.5567994 | 0.04497 | 0.04497 | 0.05611 | 161 |
| GOBP_NUCLEAR_EXPORT | 10 | 0.462277749 | 1.60170054 | 0.04525 | 0.04525 | 0.05635 | 123 |
| GOBP_ORGANELLE_ASSEMBLY | 26 | -0.444989761 | -1.5367995 | 0.04571 | 0.04571 | 0.05681 | 100 |
| GOBP_IMMUNE_RESPONSE_REGULATING_SIGNALING_PATHWAY | 19 | -0.48218246 | -1.5428614 | 0.04677 | 0.04677 | 0.05801 | 106 |
| HP_BLINDNESS | 21 | -0.469579041 | -1.5434106 | 0.04781 | 0.04781 | 0.05918 | 38 |
| GOBP_MALE_GAMETE_GENERATION | 11 | 0.435318735 | 1.57731758 | 0.04791 | 0.04791 | 0.05919 | 173 |
| GOBP_NEGATIVE_REGULATION_OF_CELLULAR_COMPONENT_ORGANI | 30 | -0.423825397 | -1.5115603 | 0.04849 | 0.04849 | 0.05978 | 58 |
| HP_ENCEPHALOPATHY | 14 | 0.38 | 1.52700663 | 0.04913 | 0.04913 | 0.06043 | 262 |
| GOBP_MONONUCLEAR_CELL_MIGRATION | 11 | -0.559856449 | -1.5500114 | 0.04920 | 0.04920 | 0.06043 | 137 |
| GOBP_CIRCADIAN_RHYTHM | 10 | 0.456808373 | 1.58275024 | 0.04967 | 0.04967 | 0.06088 | 174 |
| GOBP_ENDOMEMBRANE_SYSTEM_ORGANIZATION | 23 | -0.456689103 | -1.5348442 | 0.04983 | 0.04983 | 0.06095 | 66 |
| GOBP_REGULATION_OF_INTRACELLULAR_PROTEIN_TRANSPORT | 10 | -0.571450253 | -1.5365358 | 0.05049 | 0.05049 | 0.06165 | 18 |
| HP_HEADACHE | 14 | -0.518640939 | -1.5326918 | 0.05319 | 0.05319 | 0.06482 | 73 |
| GOBP_LYMPHOCYTE_MEDIATED_IMMUNITY | 10 | -0.568399396 | -1.5283325 | 0.05338 | 0.05338 | 0.06491 | 172 |
| HP_PROGRESSIVE_CEREBELLAR_ATAXIA | 10 | 0.448019802 | 1.55229959 | 0.05372 | 0.05372 | 0.06520 | 233 |
| HP_PAIN | 26 | -0.438619791 | -1.5148004 | 0.05402 | 0.05402 | 0.06544 | 126 |
| GOCC_ENDOSOME_MEMBRANE | 13 | -0.525432359 | -1.5234373 | 0.05425 | 0.05425 | 0.06559 | 71 |
| HP_ABNORMALITY_OF_CENTRAL_NERVOUS_SYSTEM_ELECTROPHYSIC | 11 | 0.425338515 | 1.54115563 | 0.05564 | 0.05564 | 0.06715 | 61 |
| GOBP_NEGATIVE_REGULATION_OF_PROTEIN_CONTAINING_COMPLEX | 10 | -0.56519316 | -1.5197115 | 0.05598 | 0.05598 | 0.06743 | 153 |
| GOBP_ANATOMICAL_STRUCTURE_HOMEOSTASIS | 12 | -0.538604852 | -1.5280884 | 0.05628 | 0.05628 | 0.06766 | 57 |
| HP_ABNORMAL_SYSTEMIC_BLOOD_PRESSURE | 14 | -0.514432265 | -1.5202542 | 0.05706 | 0.05706 | 0.06846 | 129 |

| | | | | | | | |
|---|---|---|---|---|---|---|---|
| GOBP_ANION_TRANSPORT | 13 | -0.52294728 | -1.5162321 | 0.05752 | 0.05752 | 0.06888 | 33 |
| GOBP_RESPONSE_TO_TOXIC_SUBSTANCE | 17 | -0.48718516 | -1.5164751 | 0.05771 | 0.05771 | 0.06888 | 147 |
| HP_ABNORMAL_VASCULAR_PHYSIOLOGY | 12 | -0.536762149 | -1.5228604 | 0.05774 | 0.05774 | 0.06888 | 116 |
| HP_ABNORMAL_IMMUNE_SYSTEM_MORPHOLOGY | 29 | -0.422306647 | -1.4930805 | 0.05792 | 0.05792 | 0.06897 | 96 |
| GOBP_RESPONSE_TO_XENOBIOTIC_STIMULUS | 16 | -0.49498136 | -1.5170962 | 0.05813 | 0.05813 | 0.06902 | 145 |
| GOBP_MODIFICATION_DEPENDENT_MACROMOLECULE_CATABOLIC_ | 19 | 0.32907172 | 1.5056275 | 0.05819 | 0.05819 | 0.06902 | 193 |
| HP_ABNORMALITY_OF_THE_KIDNEY | 58 | -0.360703132 | -1.4378495 | 0.05934 | 0.05934 | 0.07026 | 83 |
| HP_ABNORMALITY_OF_THE_HAIRLINE | 10 | 0.440781162 | 1.52721914 | 0.05960 | 0.05960 | 0.07043 | 198 |
| GOMF_PROTEIN_SERINE_THREONINE_KINASE_ACTIVITY | 10 | -0.56070277 | -1.5076376 | 0.05982 | 0.05982 | 0.07056 | 107 |
| GOCC_CORTICAL_CYTOSKELETON | 12 | -0.533576369 | -1.513822 | 0.06012 | 0.06012 | 0.07067 | 64 |
| GOBP_REGULATION_OF_CELL_DEVELOPMENT | 16 | -0.493316954 | -1.5119949 | 0.06014 | 0.06014 | 0.07067 | 137 |
| GOBP_PROCESS_UTILIZING_AUTOPHAGIC_MECHANISM | 17 | -0.485448758 | -1.5110701 | 0.06033 | 0.06033 | 0.07076 | 96 |
| HP_ABNORMAL_LARGE_INTESTINE_MORPHOLOGY | 16 | -0.492530119 | -1.5095833 | 0.06115 | 0.06115 | 0.07159 | 96 |
| HP_DEPRESSION | 12 | -0.53288408 | -1.5118579 | 0.06158 | 0.06158 | 0.07188 | 34 |
| GOCC_POLYMERIC_CYTOSKELETAL_FIBER | 23 | -0.446551103 | -1.5007723 | 0.06172 | 0.06172 | 0.07188 | 115 |
| GOBP_LEUKOCYTE_PROLIFERATION | 11 | -0.546646664 | -1.5134389 | 0.06174 | 0.06174 | 0.07188 | 150 |
| GOBP_SYNAPTIC_SIGNALING | 14 | -0.508758449 | -1.5034869 | 0.06221 | 0.06221 | 0.07229 | 137 |
| HP_ABNORMALITY_OF_THE_SPLEEN | 22 | -0.448143152 | -1.4872429 | 0.06291 | 0.06291 | 0.07296 | 116 |
| GOBP_MITOCHONDRIAL_ELECTRON_TRANSPORT_NADH_TO_UBIQUIN | 19 | 0.323884646 | 1.48189467 | 0.06358 | 0.06358 | 0.07356 | 226 |
| HP_ABNORMAL_LEUKOCYTE_PHYSIOLOGY | 13 | -0.515320099 | -1.4941178 | 0.06366 | 0.06366 | 0.07356 | 96 |
| GOBP_OXIDATIVE_PHOSPHORYLATION | 23 | 0.292422965 | 1.4441641 | 0.06399 | 0.06399 | 0.07381 | 172 |
| GOBP_RIBOSOME_BIOGENESIS | 13 | 0.381546135 | 1.48391565 | 0.06415 | 0.06415 | 0.07385 | 261 |
| GOMF_MOLECULAR_FUNCTION_REGULATOR | 44 | -0.379330064 | -1.4493939 | 0.06433 | 0.06433 | 0.07393 | 147 |
| GOBP_NEGATIVE_REGULATION_OF_LOCOMOTION | 13 | -0.514481994 | -1.4916878 | 0.06497 | 0.06497 | 0.07452 | 142 |
| GOCC_PIGMENT_GRANULE | 15 | -0.497274973 | -1.4950035 | 0.06510 | 0.06510 | 0.07454 | 28 |
| GOCC_POSTSYNAPSE | 18 | -0.47255977 | -1.4918541 | 0.06642 | 0.06642 | 0.07590 | 150 |
| HP_ABNORMAL_INTESTINE_MORPHOLOGY | 27 | -0.422157698 | -1.4721781 | 0.06653 | 0.06653 | 0.07590 | 96 |
| HP_ABNORMALITY_OF_TEMPERATURE_REGULATION | 12 | -0.528378255 | -1.4990743 | 0.06714 | 0.06714 | 0.07632 | 157 |
| HP_FEVER | 12 | -0.528378255 | -1.4990743 | 0.06714 | 0.06714 | 0.07632 | 157 |
| GOCC_NUCLEAR_PERIPHERY | 12 | 0.395593311 | 1.48340101 | 0.06764 | 0.06764 | 0.07655 | 135 |
| GOBP_DNA_METABOLIC_PROCESS | 25 | 0.280667527 | 1.41823474 | 0.06765 | 0.06765 | 0.07655 | 131 |
| GOBP_T_CELL_ACTIVATION | 16 | -0.486444558 | -1.4909313 | 0.06771 | 0.06771 | 0.07655 | 110 |
| GOBP_CELLULAR_RESPONSE_TO_LIPID | 21 | -0.452570633 | -1.4875075 | 0.06846 | 0.06846 | 0.07726 | 60 |
| GOBP_CELLULAR_RESPONSE_TO_BIOTIC_STIMULUS | 12 | -0.527261579 | -1.4959061 | 0.06860 | 0.06860 | 0.07728 | 106 |
| GOBP_CELLULAR_RESPONSE_TO_TOXIC_SUBSTANCE | 12 | -0.526504161 | -1.4937572 | 0.06926 | 0.06926 | 0.07768 | 147 |
| GOBP_DETOXIFICATION | 12 | -0.526504161 | -1.4937572 | 0.06926 | 0.06926 | 0.07768 | 147 |
| HP_OTITIS_MEDIA | 10 | -0.551963419 | -1.4841389 | 0.06943 | 0.06943 | 0.07768 | 96 |
| GOBP_APOPTOTIC_SIGNALING_PATHWAY | 29 | -0.412428326 | -1.4581553 | 0.06946 | 0.06946 | 0.07768 | 58 |
| GOBP_LIPID_METABOLIC_PROCESS | 33 | -0.399729175 | -1.4521151 | 0.07109 | 0.07109 | 0.07916 | 149 |
| GOBP_CELL_PART_MORPHOGENESIS | 14 | -0.501682544 | -1.4825762 | 0.07110 | 0.07110 | 0.07916 | 53 |
| HP_ABNORMAL_MYELINATION | 29 | 0.261915391 | 1.41136774 | 0.07116 | 0.07116 | 0.07916 | 177 |
| HP_ABNORMAL_CARDIAC_ATRIUM_MORPHOLOGY | 16 | 0.343115229 | 1.45805157 | 0.07146 | 0.07146 | 0.07936 | 147 |
| GOMF_PRIMARY_ACTIVE_TRANSMEMBRANE_TRANSPORTER_ACTIVITY | 22 | 0.296121592 | 1.44027834 | 0.07297 | 0.07297 | 0.08089 | 260 |
| GOBP_RESPONSE_TO_METAL_ION | 16 | -0.480554416 | -1.4728783 | 0.07439 | 0.07439 | 0.08232 | 56 |
| HP_DERMATOLOGICAL_MANIFESTATIONS_OF_SYSTEMIC_DISORDERS | 11 | -0.532723402 | -1.4748912 | 0.07495 | 0.07495 | 0.08269 | 116 |
| GOBP_PEPTIDYL_AMINO_ACID_MODIFICATION | 35 | -0.392375199 | -1.4367983 | 0.07499 | 0.07499 | 0.08269 | 110 |
| GOBP_CELLULAR_OXIDANT_DETOXIFICATION | 11 | -0.532002267 | -1.4728947 | 0.07576 | 0.07576 | 0.08336 | 141 |
| GOMF_TRANSFERASE_ACTIVITY_TRANSFERRING_PHOSPHORUS_CON | 24 | -0.431665543 | -1.4639815 | 0.07587 | 0.07587 | 0.08336 | 142 |
| HP_ABNORMALITY_OF_HUMORAL_IMMUNITY | 11 | -0.530825731 | -1.4696373 | 0.07751 | 0.07751 | 0.08502 | 96 |
| GOBP_REGULATION_OF_TRANSFERASE_ACTIVITY | 29 | -0.406561925 | -1.4374144 | 0.07765 | 0.07765 | 0.08502 | 112 |
| HP_RENAL_TUBULAR_DYSFUNCTION | 12 | 0.388276976 | 1.45596612 | 0.07783 | 0.07783 | 0.08503 | 161 |
| GOBP_POSITIVE_REGULATION_OF_DEFENSE_RESPONSE | 14 | -0.496070548 | -1.4659916 | 0.07792 | 0.07792 | 0.08503 | 106 |
| GOBP_CELLULAR_HOMEOSTASIS | 30 | -0.402968794 | -1.4371758 | 0.07932 | 0.07932 | 0.08640 | 45 |
| HP_ABNORMAL_NASAL_BRIDGE_MORPHOLOGY | 34 | 0.243741682 | 1.38481879 | 0.08036 | 0.08036 | 0.08731 | 84 |
| GOBP_AMIDE_BIOSYNTHETIC_PROCESS | 35 | 0.240346021 | 1.38322856 | 0.08044 | 0.08044 | 0.08731 | 96 |
| GOBP_RESPONSE_TO_REACTIVE_OXYGEN_SPECIES | 14 | -0.493047819 | -1.4570588 | 0.08153 | 0.08153 | 0.08835 | 142 |
| HP_ABNORMALLY_LAX_OR_HYPEREXTENSIBLE_SKIN | 10 | -0.542032362 | -1.4574359 | 0.08178 | 0.08178 | 0.08846 | 46 |
| GOBP_TELOMERE_MAINTENANCE | 10 | 0.421686126 | 1.46105863 | 0.08241 | 0.08241 | 0.08884 | 126 |
| GOBP_TELOMERE_ORGANIZATION | 10 | 0.421686126 | 1.46105863 | 0.08241 | 0.08241 | 0.08884 | 126 |
| HP_MICROPENIS | 11 | 0.401985112 | 1.45653778 | 0.08269 | 0.08269 | 0.08899 | 252 |
| HP_INCREASED_SERUM_LACTATE | 20 | 0.305929785 | 1.42848346 | 0.08338 | 0.08338 | 0.08956 | 172 |
| GOBP_MITOTIC_CELL_CYCLE_PROCESS | 19 | 0.310667277 | 1.42142021 | 0.08351 | 0.08351 | 0.08956 | 112 |
| GOBP_ANTIGEN_RECEPTOR_MEDIATED_SIGNALING_PATHWAY | 12 | -0.515539148 | -1.4626481 | 0.08476 | 0.08476 | 0.09074 | 166 |
| GOBP_REGULATION_OF_PROTEOLYSIS | 24 | -0.424415269 | -1.4393924 | 0.08532 | 0.08532 | 0.09119 | 106 |
| GOBP_LEUKOCYTE_DIFFERENTIATION | 17 | -0.463236616 | -1.4419298 | 0.08800 | 0.08800 | 0.09389 | 110 |
| GOBP_VESICLE_ORGANIZATION | 14 | -0.487907216 | -1.4418672 | 0.08836 | 0.08836 | 0.09411 | 70 |
| GOBP_REGULATION_OF_SMALL_GTPASE_MEDIATED_SIGNAL_TRANSDU | 10 | -0.537123238 | -1.4442361 | 0.08919 | 0.08919 | 0.09484 | 146 |
| GOBP_NEGATIVE_REGULATION_OF_CATALYTIC_ACTIVITY | 21 | -0.435633341 | -1.4318381 | 0.09092 | 0.09092 | 0.09652 | 112 |
| HP_ABNORMALITY_OF_DENTAL_MORPHOLOGY | 10 | 0.416520516 | 1.44316082 | 0.09124 | 0.09124 | 0.09670 | 55 |
| HP_INTRAUTERINE_GROWTH_RETARDATION | 23 | 0.280073527 | 1.38317499 | 0.09222 | 0.09222 | 0.09757 | 133 |
| GOCC_GLUTAMATERGIC_SYNAPSE | 15 | -0.474856564 | -1.427605 | 0.09497 | 0.09497 | 0.10031 | 126 |
| GOBP_EPITHELIUM_DEVELOPMENT | 29 | -0.397181096 | -1.4042482 | 0.09519 | 0.09519 | 0.10037 | 86 |
| GOBP_PROTEIN_LOCALIZATION_TO_PLASMA_MEMBRANE | 12 | -0.50689813 | -1.4381325 | 0.09575 | 0.09575 | 0.10079 | 88 |
| GOBP_RESPIRATORY_ELECTRON_TRANSPORT_CHAIN | 22 | 0.279566581 | 1.35975795 | 0.10227 | 0.10227 | 0.10747 | 172 |
| HP_OPTIC_DISC_PALLOR | 13 | 0.358694374 | 1.39504019 | 0.10280 | 0.10280 | 0.10785 | 161 |

| | | | | | | | |
|---|---|---|---|---|---|---|---|
| HP_INCREASED_HEAD_CIRCUMFERENCE | 10 | 0.406459249 | 1.40830053 | 0.10338 | 0.10338 | 0.10819 | 160 |
| GOBP_CARBOHYDRATE_METABOLIC_PROCESS | 18 | -0.44739154 | -1.4123989 | 0.10347 | 0.10347 | 0.10819 | 89 |
| HP_ABNORMAL_URINE_PROTEIN_LEVEL | 10 | -0.526873905 | -1.4166773 | 0.10456 | 0.10456 | 0.10914 | 78 |
| GOBP_NEURON_DEVELOPMENT | 26 | -0.403742838 | -1.3943507 | 0.10733 | 0.10733 | 0.11185 | 174 |
| HP_ABNORMAL_MYELOID_LEUKOCYTE_MORPHOLOGY | 20 | -0.43192078 | -1.4017022 | 0.10923 | 0.10923 | 0.11351 | 66 |
| GOBP_PROTEIN_LOCALIZATION_TO_CELL_PERIPHERY | 13 | -0.485883969 | -1.4087708 | 0.10928 | 0.10928 | 0.11351 | 105 |
| GOBP_RIBOSE_PHOSPHATE_METABOLIC_PROCESS | 14 | -0.476526537 | -1.4082349 | 0.10948 | 0.10948 | 0.11352 | 178 |
| GOBP_AEROBIC_RESPIRATION | 25 | 0.260900922 | 1.31835256 | 0.10985 | 0.10985 | 0.11372 | 172 |
| GOBP_PEPTIDYL_SERINE_MODIFICATION | 13 | -0.485018667 | -1.4062619 | 0.11124 | 0.11124 | 0.11497 | 103 |
| HP_GASTROESOPHAGEAL_REFLUX | 15 | 0.326681501 | 1.35637494 | 0.11255 | 0.11255 | 0.11613 | 122 |
| HP_ABNORMAL_CARDIAC_SEPTUM_MORPHOLOGY | 28 | 0.250306188 | 1.32356146 | 0.11332 | 0.11332 | 0.11673 | 161 |
| HP_ABNORMAL_EYEBROW_MORPHOLOGY | 17 | 0.31383243 | 1.36948128 | 0.11369 | 0.11369 | 0.11693 | 160 |
| GOBP_CELLULAR_AMIDE_METABOLIC_PROCESS | 47 | 0.203314536 | 1.29076346 | 0.11480 | 0.11480 | 0.11787 | 96 |
| GOBP_REGULATION_OF_T_CELL_ACTIVATION | 12 | -0.493811706 | -1.4010047 | 0.11588 | 0.11588 | 0.11878 | 150 |
| HP_DEVELOPMENTAL_REGRESSION | 10 | 0.398514851 | 1.38077477 | 0.11626 | 0.11626 | 0.11898 | 253 |
| GOMF_PEPTIDE_BINDING | 16 | -0.455579641 | -1.3963317 | 0.11751 | 0.11751 | 0.11987 | 110 |
| HP_THICKENED_SKIN | 13 | -0.481804542 | -1.3969429 | 0.11752 | 0.11752 | 0.11987 | 46 |
| GOBP_REGULATION_OF_INTRINSIC_APOPTOTIC_SIGNALING_PATHWA | 16 | -0.454922454 | -1.3943174 | 0.11890 | 0.11890 | 0.12109 | 58 |
| GOBP_PEPTIDE_METABOLIC_PROCESS | 41 | 0.21376953 | 1.29662609 | 0.11929 | 0.11929 | 0.12128 | 132 |
| HP_CONSTITUTIONAL_SYMPTOM | 36 | -0.366982359 | -1.3525475 | 0.12014 | 0.12014 | 0.12196 | 143 |
| HP_WIDE_NASAL_BRIDGE | 22 | 0.272061058 | 1.32325253 | 0.12261 | 0.12261 | 0.12426 | 83 |
| GOBP_RESPONSE_TO_RADIATION | 11 | -0.49785208 | -1.3783469 | 0.12685 | 0.12685 | 0.12835 | 141 |
| HP_MYOPIA | 11 | 0.374689826 | 1.35763707 | 0.12713 | 0.12713 | 0.12842 | 263 |
| HP_ABNORMAL_CONJUGATE_EYE_MOVEMENT | 40 | 0.20909294 | 1.26537602 | 0.12829 | 0.12829 | 0.12938 | 177 |
| HP_THIN_VERMILION_BORDER | 13 | 0.345327937 | 1.34305522 | 0.12914 | 0.12914 | 0.13004 | 122 |
| HP_HYPERKERATOSIS | 10 | -0.512357428 | -1.3776449 | 0.13021 | 0.13021 | 0.13091 | 42 |
| GOBP_ENDOTHELIUM_DEVELOPMENT | 10 | -0.511838762 | -1.3762503 | 0.13090 | 0.13090 | 0.13139 | 141 |
| HP_ABNORMALITY_OF_CIRCULATING_ENZYME_LEVEL | 12 | -0.482592644 | -1.3691748 | 0.13323 | 0.13323 | 0.13351 | 42 |
| GOBP_REGULATION_OF_INTRACELLULAR_TRANSPORT | 17 | -0.436086617 | -1.3574192 | 0.13411 | 0.13411 | 0.13418 | 103 |
| HP_ABNORMALITY_OF_THE_VOICE | 14 | -0.461576726 | -1.3640551 | 0.13679 | 0.13679 | 0.13660 | 34 |
| GOCC_DENDRITIC_TREE | 11 | -0.492117587 | -1.3624704 | 0.13696 | 0.13696 | 0.13660 | 64 |
| HP_ABNORMAL_CIRCULATING_PROTEIN_CONCENTRATION | 23 | -0.39901777 | -1.3410219 | 0.14128 | 0.14128 | 0.14045 | 73 |
| GOBP_POSTTRANSCRIPTIONAL_REGULATION_OF_GENE_EXPRESSION | 28 | 0.243128356 | 1.28560674 | 0.14128 | 0.14128 | 0.14045 | 126 |
| GOBP_REGULATION_OF_CELL_PROJECTION_ORGANIZATION | 18 | -0.426276137 | -1.3457384 | 0.14201 | 0.14201 | 0.14095 | 146 |
| GOBP_POSITIVE_REGULATION_OF_CELL_DEVELOPMENT | 14 | -0.457395161 | -1.3516978 | 0.14400 | 0.14400 | 0.14270 | 106 |
| GOBP_CELLULAR_LIPID_METABOLIC_PROCESS | 20 | -0.412343492 | -1.3381684 | 0.14629 | 0.14629 | 0.14474 | 148 |
| GOCC_INTRACELLULAR_PROTEIN_CONTAINING_COMPLEX | 16 | 0.304125231 | 1.29236546 | 0.14679 | 0.14679 | 0.14499 | 252 |
| HP_ABNORMAL_ELASTICITY_OF_SKIN | 12 | -0.4758483 | -1.3500403 | 0.14700 | 0.14700 | 0.14499 | 46 |
| GOBP_RESPONSE_TO_STEROID_HORMONE | 14 | -0.454231797 | -1.3423494 | 0.14992 | 0.14992 | 0.14764 | 54 |
| HP_ABNORMALITY_OF_DIGESTIVE_SYSTEM_MORPHOLOGY | 31 | -0.368163726 | -1.3243781 | 0.15119 | 0.15119 | 0.14865 | 137 |
| HP_FATIGUE | 12 | -0.471850139 | -1.338697 | 0.15402 | 0.15402 | 0.15120 | 157 |
| GOBP_GLYCOPROTEIN_METABOLIC_PROCESS | 11 | 0.358133951 | 1.29764913 | 0.15533 | 0.15533 | 0.15225 | 186 |
| GOBP_CELLULAR_PROTEIN_CATABOLIC_PROCESS | 24 | 0.252797679 | 1.26766132 | 0.15639 | 0.15639 | 0.15305 | 193 |
| GOBP_CELLULAR_COMPONENT_DISASSEMBLY | 21 | -0.401981064 | -1.3212299 | 0.15745 | 0.15745 | 0.15373 | 105 |
| GOBP_VASCULAR_PROCESS_IN_CIRCULATORY_SYSTEM | 11 | -0.4812228 | -1.3323073 | 0.15786 | 0.15786 | 0.15373 | 141 |
| GOBP_POSITIVE_REGULATION_OF_INTRACELLULAR_TRANSPORT | 10 | -0.495682709 | -1.3328093 | 0.15807 | 0.15807 | 0.15373 | 103 |
| HP_ADULT_ONSET | 10 | -0.495669501 | -1.3327738 | 0.15807 | 0.15807 | 0.15373 | 117 |
| GOBP_NEGATIVE_REGULATION_OF_CYTOSKELETON_ORGANIZATION | 11 | -0.480562084 | -1.330478 | 0.15880 | 0.15880 | 0.15420 | 153 |
| GOMF_PROTEIN_HOMODIMERIZATION_ACTIVITY | 24 | -0.387748525 | -1.3150381 | 0.16107 | 0.16107 | 0.15583 | 105 |
| GOMF_ANTIOXIDANT_ACTIVITY | 10 | -0.494690973 | -1.3301427 | 0.16109 | 0.16109 | 0.15583 | 141 |
| GOBP_REGULATION_OF_NEURON_PROJECTION_DEVELOPMENT | 10 | -0.49448648 | -1.3295929 | 0.16122 | 0.16122 | 0.15583 | 142 |
| HP_HYPOPLASIA_OF_THE_CORPUS_CALLOSUM | 22 | 0.262430465 | 1.27641119 | 0.16148 | 0.16148 | 0.15584 | 122 |
| HP_MYOPATHY | 13 | -0.458205482 | -1.3285198 | 0.16314 | 0.16314 | 0.15719 | 42 |
| GOBP_POSITIVE_REGULATION_OF_CELLULAR_BIOSYNTHETIC_PROCES | 47 | -0.332830399 | -1.2871892 | 0.16435 | 0.16435 | 0.15812 | 72 |
| HP_ABNORMAL_LOCATION_OF_EARS | 27 | 0.239474955 | 1.25104366 | 0.16643 | 0.16643 | 0.15987 | 86 |
| GOBP_ORGANELLE_FISSION | 10 | 0.372658379 | 1.29118723 | 0.16777 | 0.16777 | 0.16091 | 103 |
| GOBP_CELLULAR_RESPONSE_TO_ORGANIC_CYCLIC_COMPOUND | 17 | -0.421477469 | -1.3119449 | 0.16827 | 0.16827 | 0.16114 | 58 |
| GOBP_MONONUCLEAR_CELL_DIFFERENTIATION | 13 | -0.453726448 | -1.3155333 | 0.17176 | 0.17176 | 0.16424 | 110 |
| GOBP_NEURON_DIFFERENTIATION | 29 | -0.3655015 | -1.2922438 | 0.17272 | 0.17272 | 0.16491 | 109 |
| GOBP_ACTIVATION_OF_IMMUNE_RESPONSE | 23 | -0.386842997 | -1.3001049 | 0.17350 | 0.17350 | 0.16540 | 106 |
| GOBP_CELLULAR_RESPONSE_TO_CHEMICAL_STRESS | 22 | -0.388562964 | -1.2895154 | 0.17755 | 0.17755 | 0.16900 | 86 |
| GOBP_RESPONSE_TO_OXYGEN_LEVELS | 13 | -0.450249702 | -1.3054529 | 0.17922 | 0.17922 | 0.17008 | 86 |
| HP_SPLENOMEGALY | 19 | -0.404273498 | -1.2935725 | 0.17923 | 0.17923 | 0.17008 | 116 |
| HP_ABNORMALITY_OF_FLUID_REGULATION | 21 | -0.392167626 | -1.2889751 | 0.17991 | 0.17991 | 0.17047 | 114 |
| GOCC_CLATHRIN_COATED_VESICLE | 10 | -0.483818112 | -1.3009074 | 0.18071 | 0.18071 | 0.17097 | 32 |
| GOBP_REGULATION_OF_PROTEIN_STABILITY | 16 | -0.422678868 | -1.2954923 | 0.18321 | 0.18321 | 0.17307 | 58 |
| GOBP_MYELOID_CELL_DIFFERENTIATION | 15 | -0.430135103 | -1.2931548 | 0.18522 | 0.18522 | 0.17471 | 136 |
| GOBP_PROTEIN_CONTAINING_COMPLEX_DISASSEMBLY | 10 | -0.47988765 | -1.290339 | 0.18702 | 0.18702 | 0.17614 | 10 |
| GOMF_CARBOHYDRATE_BINDING | 13 | -0.445360086 | -1.2912759 | 0.18980 | 0.18980 | 0.17841 | 81 |
| HP_ABNORMALITY_OF_LONG_BONE_MORPHOLOGY | 17 | 0.282939144 | 1.23467119 | 0.18999 | 0.18999 | 0.17841 | 129 |
| GOBP_ESTABLISHMENT_OF_PROTEIN_LOCALIZATION | 82 | -0.296631707 | -1.2310201 | 0.19201 | 0.19201 | 0.18002 | 92 |
| GOMF_STRUCTURAL_MOLECULE_ACTIVITY | 33 | -0.34763181 | -1.2628585 | 0.19575 | 0.19575 | 0.18299 | 74 |
| GOBP_PROTEIN_LOCALIZATION_TO_NUCLEUS | 10 | -0.476196759 | -1.2804148 | 0.19608 | 0.19608 | 0.18299 | 92 |
| GOBP_PROTEIN_MODIFICATION_BY_SMALL_PROTEIN_CONJUGATION | 13 | 0.318988541 | 1.2406156 | 0.19626 | 0.19626 | 0.18299 | 211 |
| GOBP_PURINE_CONTAINING_COMPOUND_METABOLIC_PROCESS | 13 | -0.441513597 | -1.2801234 | 0.19634 | 0.19634 | 0.18299 | 178 |

| | | | | | | | |
|---|---|---|---|---|---|---|---|
| HP_DEVIATION_OF_THE_HAND_OR_OF_FINGERS_OF_THE_HAND | 13 | 0.318259871 | 1.23778164 | 0.19796 | 0.19796 | 0.18423 | 99 |
| GOBP_RESPONSE_TO_HYDROGEN_PEROXIDE | 11 | -0.461821018 | -1.2785918 | 0.19925 | 0.19925 | 0.18515 | 142 |
| HP_ABNORMALITY_OF_NEUTROPHILS | 19 | -0.396126041 | -1.2675027 | 0.20083 | 0.20083 | 0.18635 | 66 |
| GOBP_RESPONSE_TO_EXTRACELLULAR_STIMULUS | 12 | -0.448539248 | -1.2725612 | 0.20183 | 0.20183 | 0.18699 | 142 |
| GOBP_ORGANELLE_LOCALIZATION | 24 | -0.370561012 | -1.2567472 | 0.20338 | 0.20338 | 0.18815 | 66 |
| GOBP_CENTRAL_NERVOUS_SYSTEM_DEVELOPMENT | 28 | -0.357897901 | -1.2579874 | 0.20583 | 0.20583 | 0.19014 | 58 |
| GOBP_NITROGEN_COMPOUND_TRANSPORT | 96 | -0.285960914 | -1.2047062 | 0.20627 | 0.20627 | 0.19026 | 114 |
| HP_LACTIC_ACIDOSIS | 16 | 0.285315261 | 1.2124334 | 0.20811 | 0.20811 | 0.19168 | 172 |
| HP_TALIPES_EQUINOVARUS | 11 | -0.457300732 | -1.2660769 | 0.21030 | 0.21030 | 0.19341 | 178 |
| GOMF_AMIDE_BINDING | 19 | -0.392559018 | -1.2560892 | 0.21090 | 0.21090 | 0.19368 | 86 |
| HP_ABNORMALITY_OF_THE_BLADDER | 14 | -0.426510869 | -1.2604283 | 0.21175 | 0.21175 | 0.19417 | 83 |
| GOBP_CELLULAR_MACROMOLECULE_LOCALIZATION | 90 | -0.287026289 | -1.2015691 | 0.21844 | 0.21844 | 0.20002 | 110 |
| GOBP_EPITHELIAL_CELL_DEVELOPMENT | 12 | -0.441622308 | -1.252937 | 0.21931 | 0.21931 | 0.20052 | 141 |
| HP_EXTERNAL_GENITAL_HYPOPLASIA | 17 | 0.273944577 | 1.19542129 | 0.22031 | 0.22031 | 0.20115 | 252 |
| HP_ABNORMAL_CORTICAL_GYRATION | 14 | 0.29668599 | 1.19221441 | 0.22465 | 0.22465 | 0.20463 | 129 |
| GOMF_PROTEIN_DIMERIZATION_ACTIVITY | 30 | -0.346133133 | -1.2344732 | 0.22478 | 0.22478 | 0.20463 | 89 |
| HP_ABNORMAL_CORPUS_CALLOSUM_MORPHOLOGY | 30 | 0.215806913 | 1.17363253 | 0.22770 | 0.22770 | 0.20698 | 133 |
| HP_ABNORMAL_NERVOUS_SYSTEM_ELECTROPHYSIOLOGY | 14 | 0.294924046 | 1.18513414 | 0.23091 | 0.23091 | 0.20942 | 111 |
| HP_ABNORMALITY_OF_THE_FOREHEAD | 28 | 0.223648893 | 1.18260383 | 0.23105 | 0.23105 | 0.20942 | 84 |
| HP_ABNORMALITY_OF_UPPER_LIP_VERMILLION | 15 | 0.285311784 | 1.18460872 | 0.23155 | 0.23155 | 0.20957 | 96 |
| GOBP_MONOCARBOXYLIC_ACID_METABOLIC_PROCESS | 21 | -0.374059605 | -1.2294577 | 0.23195 | 0.23195 | 0.20963 | 173 |
| GOMF_IDENTICAL_PROTEIN_BINDING | 73 | -0.291117175 | -1.1940061 | 0.23405 | 0.23405 | 0.21123 | 123 |
| GOBP_ORGANOPHOSPHATE_METABOLIC_PROCESS | 22 | -0.366676809 | -1.2168823 | 0.23998 | 0.23998 | 0.21626 | 163 |
| GOBP_PROTEIN_MATURATION | 14 | -0.414759879 | -1.2257016 | 0.24124 | 0.24124 | 0.21657 | 7 |
| HP_ABNORMAL_THUMB_MORPHOLOGY | 13 | 0.302665568 | 1.17713202 | 0.24129 | 0.24129 | 0.21657 | 96 |
| HP_HEMOLYTIC_ANEMIA | 10 | -0.455540566 | -1.2248737 | 0.24136 | 0.24136 | 0.21657 | 166 |
| GOBP_EPITHELIAL_CELL_DIFFERENTIATION | 19 | -0.380051032 | -1.2160668 | 0.24380 | 0.24380 | 0.21845 | 61 |
| GOBP_LOCALIZATION_WITHIN_MEMBRANE | 34 | -0.331371172 | -1.2091145 | 0.24668 | 0.24668 | 0.22071 | 88 |
| HP_INCREASED_BLOOD_PRESSURE | 12 | -0.429956529 | -1.2198397 | 0.24712 | 0.24712 | 0.22079 | 129 |
| GOCC_NUCLEAR_ENVELOPE | 15 | 0.279618331 | 1.16096962 | 0.25092 | 0.25092 | 0.22387 | 103 |
| GOBP_REGULATION_OF_ESTABLISHMENT_OF_PROTEIN_LOCALIZATION | 18 | -0.383216233 | -1.2097998 | 0.25291 | 0.25291 | 0.22532 | 104 |
| HP_ABNORMALITY_OF_THE_URETER | 12 | -0.427019075 | -1.2115058 | 0.25454 | 0.25454 | 0.22624 | 34 |
| GOBP_POSITIVE_REGULATION_OF_PROTEIN_LOCALIZATION | 15 | -0.403291853 | -1.2124534 | 0.25466 | 0.25466 | 0.22624 | 103 |
| HP_OPTIC_ATROPHY | 24 | 0.233274399 | 1.16976126 | 0.25633 | 0.25633 | 0.22740 | 171 |
| HP_GLAUCOMA | 13 | -0.414582501 | -1.2020395 | 0.26301 | 0.26301 | 0.23242 | 34 |
| HP_ABNORMAL_BLOOD_GLUCOSE_CONCENTRATION | 14 | 0.286091616 | 1.14964157 | 0.26306 | 0.26306 | 0.23242 | 161 |
| HP_RESPIRATORY_TRACT_INFECTION | 25 | -0.348774528 | -1.1933092 | 0.26310 | 0.26310 | 0.23242 | 96 |
| GOCC_GOLGI_APPARATUS | 58 | -0.295566539 | -1.1781994 | 0.26465 | 0.26465 | 0.23346 | 116 |
| HP_MACULE | 11 | 0.320619221 | 1.16171966 | 0.26546 | 0.26546 | 0.23384 | 126 |
| GOMF_RIBONUCLEOTIDE_BINDING | 60 | -0.293272382 | -1.1747913 | 0.26609 | 0.26609 | 0.23407 | 112 |
| HP_THIN_UPPER_LIP_VERMILION | 12 | 0.307826042 | 1.15429015 | 0.26691 | 0.26691 | 0.23441 | 122 |
| HP_ABNORMALITY_OF_CARDIOVASCULAR_SYSTEM_ELECTROPHYSIOL | 17 | -0.385198359 | -1.1990179 | 0.26723 | 0.26723 | 0.23441 | 42 |
| GOBP_REGULATION_OF_GROWTH | 10 | -0.4428416 | -1.1907283 | 0.26880 | 0.26880 | 0.23546 | 144 |
| GOMF_RNA_POLYMERASE_II_SPECIFIC_DNA_BINDING_TRANSCRIPTIC | 13 | 0.29473536 | 1.14628972 | 0.26933 | 0.26933 | 0.23559 | 127 |
| HP_IMPAIRED_SOCIAL_INTERACTIONS | 14 | 0.283416659 | 1.13889242 | 0.27200 | 0.27200 | 0.23720 | 224 |
| GOBP_NEGATIVE_REGULATION_OF_PROTEIN_METABOLIC_PROCESS | 33 | -0.325825516 | -1.1836418 | 0.27215 | 0.27215 | 0.23720 | 155 |
| GOBP_CELL_DIVISION | 13 | 0.294145414 | 1.14399529 | 0.27230 | 0.27230 | 0.23720 | 104 |
| HP_ABNORMAL_CIRCULATING_LIPID_CONCENTRATION | 11 | -0.429337055 | -1.1886571 | 0.27865 | 0.27865 | 0.24239 | 78 |
| HP_DELAYED_GROSS_MOTOR_DEVELOPMENT | 13 | 0.292222114 | 1.13651516 | 0.27910 | 0.27910 | 0.24245 | 238 |
| HP_ABNORMALITY_OF_PRENATAL_DEVELOPMENT_OR_BIRTH | 39 | -0.313067185 | -1.1703738 | 0.28025 | 0.28025 | 0.24311 | 52 |
| HP_ABNORMALITY_OF_THE_PINNA | 24 | 0.228370991 | 1.14517298 | 0.28164 | 0.28164 | 0.24397 | 82 |
| GOBP_SMALL_MOLECULE_METABOLIC_PROCESS | 53 | -0.296052152 | -1.1657619 | 0.28311 | 0.28311 | 0.24483 | 165 |
| GOMF_PROTEIN_TYROSINE_KINASE_BINDING | 10 | -0.436449025 | -1.1735397 | 0.28430 | 0.28430 | 0.24483 | 150 |
| HP_ELEVATED_HEPATIC_TRANSAMINASE | 11 | -0.426686971 | -1.1813201 | 0.28444 | 0.28444 | 0.24483 | 190 |
| GOBP_REGULATION_OF_HEMOPOIESIS | 10 | -0.436217317 | -1.1729167 | 0.28458 | 0.28458 | 0.24483 | 110 |
| GOBP_REGULATION_OF_LEUKOCYTE_DIFFERENTIATION | 10 | -0.436217317 | -1.1729167 | 0.28458 | 0.28458 | 0.24483 | 110 |
| GOBP_MICROTUBULE_BASED_PROCESS | 20 | -0.362417992 | -1.1761464 | 0.28624 | 0.28624 | 0.24592 | 62 |
| HP_ABNORMAL_EXTERNAL_GENITALIA | 33 | 0.196950295 | 1.11020996 | 0.28769 | 0.28769 | 0.24668 | 133 |
| GOBP_REGULATION_OF_HORMONE_LEVELS | 12 | -0.41412134 | -1.1749133 | 0.28791 | 0.28791 | 0.24668 | 167 |
| HP_INGUINAL_HERNIA | 11 | -0.423934921 | -1.1737008 | 0.29159 | 0.29159 | 0.24949 | 107 |
| HP_AUTOIMMUNITY | 10 | -0.432875917 | -1.1639322 | 0.29377 | 0.29377 | 0.25101 | 45 |
| GOBP_FATTY_ACID_METABOLIC_PROCESS | 14 | -0.394458716 | -1.1657075 | 0.29418 | 0.29418 | 0.25102 | 163 |
| GOBP_RESPONSE_TO_NITROGEN_COMPOUND | 38 | -0.311368124 | -1.1585687 | 0.29566 | 0.29566 | 0.25194 | 112 |
| GOBP_MICROTUBULE_CYTOSKELETON_ORGANIZATION | 14 | -0.393204419 | -1.1620008 | 0.29817 | 0.29817 | 0.25374 | 62 |
| GOBP_RESPONSE_TO_OXIDATIVE_STRESS | 30 | -0.323995605 | -1.1555204 | 0.30170 | 0.30170 | 0.25639 | 142 |
| GOMF_TRANSITION_METAL_ION_BINDING | 30 | 0.205104149 | 1.11542721 | 0.30282 | 0.30282 | 0.25699 | 113 |
| GOBP_NEGATIVE_REGULATION_OF_BIOSYNTHETIC_PROCESS | 39 | -0.307507811 | -1.1495906 | 0.30352 | 0.30352 | 0.25724 | 53 |
| GOBP_POSITIVE_REGULATION_OF_ESTABLISHMENT_OF_PROTEIN_LOC | 11 | -0.418223499 | -1.1578882 | 0.30615 | 0.30615 | 0.25911 | 103 |
| HP_ABNORMAL_EYELID_MORPHOLOGY | 39 | 0.183143993 | 1.0995706 | 0.31186 | 0.31186 | 0.26359 | 86 |
| HP_FUNCTIONAL_MOTOR_DEFICIT | 17 | -0.369057176 | -1.1487748 | 0.31422 | 0.31422 | 0.26523 | 154 |
| HP_ABNORMALITY_OF_THE_NAIL | 13 | -0.395326468 | -1.1462086 | 0.31529 | 0.31529 | 0.26578 | 11 |
| HP_CONSTIPATION | 14 | -0.385794198 | -1.140102 | 0.31929 | 0.31929 | 0.26866 | 34 |
| GOCC_EXTRINSIC_COMPONENT_OF_MEMBRANE | 10 | -0.423702953 | -1.1392676 | 0.31957 | 0.31957 | 0.26866 | 137 |
| HP_POSITIONAL_FOOT_DEFORMITY | 13 | -0.392404183 | -1.1377357 | 0.32222 | 0.32222 | 0.27021 | 34 |
| HP_SLEEP_DISTURBANCE | 16 | -0.371610094 | -1.1389687 | 0.32228 | 0.32228 | 0.27021 | 33 |

| | | | | | | | |
|---|---|---|---|---|---|---|---|
| HP_LONG_FINGERS | 13 | -0.392112939 | -1.1368913 | 0.32301 | 0.32301 | 0.27046 | 107 |
| HP_HYPERPIGMENTATION_OF_THE_SKIN | 14 | 0.272275581 | 1.09412268 | 0.32827 | 0.32827 | 0.27412 | 133 |
| GOMF_GTPASE_BINDING | 14 | -0.382526 | -1.1304438 | 0.32857 | 0.32857 | 0.27412 | 135 |
| GOBP_NEGATIVE_REGULATION_OF_ORGANELLE_ORGANIZATION | 23 | -0.336010082 | -1.1292652 | 0.32870 | 0.32870 | 0.27412 | 58 |
| GOMF_PHOSPHOPROTEIN_BINDING | 10 | -0.419862833 | -1.1289421 | 0.33013 | 0.33013 | 0.27496 | 171 |
| GOBP_GLAND_DEVELOPMENT | 11 | -0.40626151 | -1.1247704 | 0.33715 | 0.33715 | 0.28043 | 56 |
| GOMF_ATP_HYDROLYSIS_ACTIVITY | 12 | 0.289978859 | 1.08736655 | 0.33945 | 0.33945 | 0.28196 | 108 |
| HP_FEEDING_DIFFICULTIES | 40 | 0.177660742 | 1.07515654 | 0.34595 | 0.34595 | 0.28699 | 161 |
| HP_ABNORMAL_CONJUNCTIVA_MORPHOLOGY | 11 | -0.401036482 | -1.1103045 | 0.35036 | 0.35036 | 0.29024 | 112 |
| HP_ABNORMAL_ERYTHROCYTE_MORPHOLOGY | 36 | -0.301596232 | -1.1115608 | 0.35083 | 0.35083 | 0.29024 | 130 |
| HP_EMOTIONAL_LABILITY | 12 | 0.286069652 | 1.07270775 | 0.35126 | 0.35126 | 0.29024 | 299 |
| GOBP_CELLULAR_RESPIRATION | 26 | 0.210013453 | 1.085539 | 0.35172 | 0.35172 | 0.29024 | 172 |
| HP_ABNORMAL_RESPIRATORY_SYSTEM_MORPHOLOGY | 40 | -0.294681652 | -1.1068573 | 0.35256 | 0.35256 | 0.29046 | 157 |
| GOBP_POSITIVE_REGULATION_OF_CELLULAR_AMIDE_METABOLIC_PRO | 11 | -0.399349377 | -1.1056336 | 0.35293 | 0.35293 | 0.29046 | 58 |
| HP_ABNORMALITY_OF_MOUTH_SIZE | 11 | -0.399140734 | -1.1050559 | 0.35373 | 0.35373 | 0.29075 | 46 |
| HP_ABNORMAL_HAIR_MORPHOLOGY | 43 | 0.174633461 | 1.06796551 | 0.35569 | 0.35569 | 0.29197 | 161 |
| HP_ABNORMALITY_OF_THE_UPPER_RESPIRATORY_TRACT | 15 | -0.36789488 | -1.1060362 | 0.35691 | 0.35691 | 0.29259 | 96 |
| HP_UNUSUAL_INFECTION | 29 | -0.310110074 | -1.0964054 | 0.36137 | 0.36137 | 0.29586 | 96 |
| HP_ABNORMAL_FEAR_ANXIETY_RELATED_BEHAVIOR | 13 | -0.379001421 | -1.0988758 | 0.36196 | 0.36196 | 0.29596 | 34 |
| HP_ABNORMAL_CORNEA_MORPHOLOGY | 21 | -0.334325608 | -1.0988602 | 0.36525 | 0.36525 | 0.29826 | 87 |
| HP_HYPERREFLEXIA | 36 | 0.181480148 | 1.05757488 | 0.36807 | 0.36807 | 0.30017 | 161 |
| GOBP_RESPONSE_TO_HORMONE | 24 | -0.3215462 | -1.0905148 | 0.37154 | 0.37154 | 0.30261 | 54 |
| GOCC_VACUOLAR_MEMBRANE | 18 | -0.3444617 | -1.0874531 | 0.37410 | 0.37410 | 0.30429 | 112 |
| HP_ABDOMINAL_PAIN | 12 | -0.384715829 | -1.0914863 | 0.37465 | 0.37465 | 0.30435 | 123 |
| HP_ABNORMAL_MYOCARDIUM_MORPHOLOGY | 31 | -0.301410174 | -1.0842487 | 0.38179 | 0.38179 | 0.30963 | 42 |
| HP_ABSENT_SPEECH | 11 | 0.286832988 | 1.0392999 | 0.38215 | 0.38215 | 0.30963 | 199 |
| HP_ABNORMAL_URINE_METABOLITE_LEVEL | 16 | -0.353084198 | -1.0821876 | 0.38457 | 0.38457 | 0.31119 | 78 |
| HP_IMPAIRMENT_IN_PERSONALITY_FUNCTIONING | 27 | -0.307928822 | -1.0738311 | 0.38758 | 0.38758 | 0.31322 | 34 |
| GOBP_ORGANIC_HYDROXY_COMPOUND_METABOLIC_PROCESS | 15 | -0.357693381 | -1.0753665 | 0.38869 | 0.38869 | 0.31372 | 152 |
| GOBP_ORGANIC_ACID_METABOLIC_PROCESS | 32 | -0.298196068 | -1.0777249 | 0.39176 | 0.39176 | 0.31578 | 165 |
| GOBP_PROTEOLYSIS | 51 | -0.273491966 | -1.0731334 | 0.39485 | 0.39485 | 0.31787 | 106 |
| HP_LOCALIZED_SKIN_LESION | 32 | -0.296696273 | -1.0723044 | 0.39687 | 0.39687 | 0.31908 | 92 |
| GOBP_CELLULAR_RESPONSE_TO_DNA_DAMAGE_STIMULUS | 24 | 0.208403946 | 1.04504765 | 0.40169 | 0.40169 | 0.32254 | 160 |
| GOBP_ESTABLISHMENT_OF_ORGANELLE_LOCALIZATION | 21 | -0.324693225 | -1.0672005 | 0.40316 | 0.40316 | 0.32331 | 48 |
| HP_PERIPHERAL_NEUROPATHY | 23 | -0.315394301 | -1.0599795 | 0.40564 | 0.40564 | 0.32488 | 81 |
| HP_EPICANTHUS | 24 | 0.207092259 | 1.03847016 | 0.40688 | 0.40688 | 0.32533 | 83 |
| HP_DYSPNEA | 12 | -0.374973522 | -1.0638461 | 0.40723 | 0.40723 | 0.32533 | 126 |
| HP_ABNORMAL_PATTERN_OF_RESPIRATION | 29 | -0.297320613 | -1.0511878 | 0.41410 | 0.41410 | 0.33039 | 6 |
| HP_PUBERTY_AND_GONADAL_DISORDERS | 10 | 0.292138417 | 1.01220157 | 0.41832 | 0.41832 | 0.33325 | 99 |
| GOCC_ORGANELLE_INNER_MEMBRANE | 34 | 0.179841145 | 1.02176778 | 0.41875 | 0.41875 | 0.33325 | 172 |
| HP_JOINT_LAXITY | 10 | 0.291725832 | 1.01077204 | 0.42384 | 0.42384 | 0.33688 | 77 |
| HP_ABNORMAL_ABDOMEN_MORPHOLOGY | 38 | -0.281327772 | -1.0467916 | 0.42753 | 0.42753 | 0.33937 | 116 |
| HP_ABNORMALITY_OF_THE_MIDDLE_EAR | 19 | -0.325157868 | -1.0404226 | 0.42929 | 0.42929 | 0.34034 | 45 |
| GOBP_POSITIVE_REGULATION_OF_TRANSLATION | 10 | -0.385380765 | -1.0362255 | 0.43153 | 0.43153 | 0.34169 | 38 |
| GOBP_PROTEASOME_MEDIATED_UBIQUITIN_DEPENDENT_PROTEIN_C | 14 | 0.250397019 | 1.00620502 | 0.43234 | 0.43234 | 0.34189 | 193 |
| GOBP_INTRINSIC_APOPTOTIC_SIGNALING_PATHWAY | 22 | -0.312402383 | -1.036763 | 0.43433 | 0.43433 | 0.34298 | 79 |
| GOMF_SEQUENCE_SPECIFIC_DNA_BINDING | 12 | 0.267685399 | 1.00377024 | 0.43480 | 0.43480 | 0.34298 | 174 |
| GOBP_NEGATIVE_REGULATION_OF_PHOSPHORUS_METABOLIC_PROC | 14 | -0.349767589 | -1.0336359 | 0.43727 | 0.43727 | 0.34449 | 150 |
| HP_NEOPLASM | 36 | -0.281154019 | -1.0362192 | 0.43859 | 0.43859 | 0.34500 | 137 |
| GOBP_SMALL_MOLECULE_BIOSYNTHETIC_PROCESS | 21 | -0.315261838 | -1.0362014 | 0.43902 | 0.43902 | 0.34500 | 111 |
| HP_ABNORMALITY_OF_FACIAL_MUSCULATURE | 10 | -0.381589362 | -1.0260311 | 0.44374 | 0.44374 | 0.34819 | 33 |
| GOCC_CILIUM | 11 | -0.370562717 | -1.0259352 | 0.44419 | 0.44419 | 0.34819 | 130 |
| GOMF_PROTEIN_DOMAIN_SPECIFIC_BINDING | 26 | -0.29645703 | -1.0238326 | 0.44815 | 0.44815 | 0.35085 | 179 |
| HP_FUNCTIONAL_ABNORMALITY_OF_THE_MIDDLE_EAR | 11 | -0.368789055 | -1.0210247 | 0.44918 | 0.44918 | 0.35122 | 34 |
| GOBP_NEGATIVE_REGULATION_OF_TRANSFERASE_ACTIVITY | 10 | -0.378461277 | -1.0176202 | 0.45307 | 0.45307 | 0.35382 | 112 |
| GOMF_ION_TRANSMEMBRANE_TRANSPORTER_ACTIVITY | 18 | -0.321316196 | -1.0143836 | 0.45750 | 0.45750 | 0.35683 | 183 |
| HP_ABNORMALITY_OF_FACIAL_SOFT_TISSUE | 19 | -0.31700703 | -1.014342 | 0.45826 | 0.45826 | 0.35699 | 117 |
| GOBP_RHYTHMIC_PROCESS | 15 | 0.238676414 | 0.99097961 | 0.45941 | 0.45941 | 0.35744 | 174 |
| HP_ABNORMAL_LEUKOCYTE_COUNT | 18 | -0.320660036 | -1.0123121 | 0.46047 | 0.46047 | 0.35782 | 96 |
| HP_ABNORMALITY_OF_HINDBRAIN_MORPHOLOGY | 30 | 0.183534033 | 0.99812147 | 0.46244 | 0.46244 | 0.35890 | 129 |
| HP_NEOPLASM_BY_HISTOLOGY | 11 | -0.363196064 | -1.00554 | 0.46697 | 0.46697 | 0.36197 | 53 |
| HP_ABNORMAL_FACIAL_SHAPE | 30 | 0.183014609 | 0.99529666 | 0.46870 | 0.46870 | 0.36286 | 134 |
| HP_ABNORMALITY_OF_THE_ABDOMINAL_WALL | 13 | -0.345893474 | -1.0028827 | 0.47124 | 0.47124 | 0.36393 | 107 |
| HP_DISPLACEMENT_OF_THE_URETHRAL_MEATUS | 13 | -0.345872662 | -1.0028223 | 0.47124 | 0.47124 | 0.36393 | 46 |
| HP_ABNORMALITY_OF_THE_CALVARIA | 29 | 0.184980453 | 0.99679306 | 0.47491 | 0.47491 | 0.36631 | 104 |
| HP_APLASIA_HYPOPLASIA_INVOLVING_THE_SKELETON | 31 | 0.179649491 | 0.98857302 | 0.48245 | 0.48245 | 0.37167 | 133 |
| HP_SHORT_DIGIT | 15 | 0.234855943 | 0.97511709 | 0.48386 | 0.48386 | 0.37229 | 129 |
| GOBP_ORGANOPHOSPHATE_BIOSYNTHETIC_PROCESS | 10 | -0.369687611 | -0.9940292 | 0.48491 | 0.48491 | 0.37264 | 78 |
| HP_ABNORMALITY_OF_THE_VASCULATURE_OF_THE_EYE | 16 | -0.323918158 | -0.992795 | 0.48556 | 0.48556 | 0.37269 | 112 |
| GOMF_ADENYL_NUCLEOTIDE_BINDING | 44 | -0.262215264 | -1.0019064 | 0.48741 | 0.48741 | 0.37365 | 181 |
| GOBP_EMBRYO_DEVELOPMENT | 17 | -0.318699017 | -0.9920235 | 0.48835 | 0.48835 | 0.37391 | 38 |
| HP_ABNORMALITY_OF_CRANIAL_SUTURES | 12 | -0.348662407 | -0.9891982 | 0.48974 | 0.48974 | 0.37452 | 34 |
| HP_ABNORMALITY_OF_THE_LOWER_URINARY_TRACT | 24 | -0.290994855 | -0.9869008 | 0.49279 | 0.49279 | 0.37610 | 52 |
| GOBP_REGULATION_OF_PROTEIN_MODIFICATION_BY_SMALL_PROTEI | 10 | 0.275679936 | 0.9551762 | 0.49301 | 0.49301 | 0.37610 | 187 |
| GOBP_ELECTRON_TRANSPORT_CHAIN | 24 | 0.194292577 | 0.97428578 | 0.49708 | 0.49708 | 0.37875 | 172 |

| | | | | | | | |
|---|---|---|---|---|---|---|---|
| GOCC_TRANSPORT_VESICLE_MEMBRANE | 10 | -0.363904553 | -0.9784795 | 0.49890 | 0.49890 | 0.37967 | 68 |
| GOMF_TRANSCRIPTION_FACTOR_BINDING | 25 | 0.192734993 | 0.97390484 | 0.50033 | 0.50033 | 0.38030 | 42 |
| HP_CHOLESTASIS | 11 | -0.35143273 | -0.9729722 | 0.50458 | 0.50458 | 0.38268 | 163 |
| HP_ABNORMALITY_OF_THE_NASAL_ALAE | 14 | 0.238531037 | 0.95852231 | 0.50469 | 0.50469 | 0.38268 | 76 |
| HP_AUTISTIC_BEHAVIOR | 23 | 0.193800814 | 0.95710738 | 0.50753 | 0.50753 | 0.38437 | 161 |
| HP_DELAYED_ABILITY_TO_WALK | 11 | 0.26046404 | 0.94375564 | 0.50966 | 0.50966 | 0.38552 | 238 |
| HP_APLASIA_HYPOPLASIA_AFFECTING_THE_EYE | 11 | -0.349471638 | -0.9675427 | 0.51227 | 0.51227 | 0.38702 | 62 |
| HP_ABNORMALITY_OF_THE_FEMALE_GENITALIA | 11 | 0.258873853 | 0.93799381 | 0.51507 | 0.51507 | 0.38867 | 52 |
| HP_ABNORMAL_CIRCULATING_METABOLITE_CONCENTRATION | 53 | -0.247842028 | -0.9759253 | 0.52050 | 0.52050 | 0.39219 | 81 |
| GOBP_MULTICELLULAR_ORGANISM_REPRODUCTION | 19 | -0.300882854 | -0.9627487 | 0.52099 | 0.52099 | 0.39219 | 53 |
| GOMF_DNA_BINDING_TRANSCRIPTION_FACTOR_BINDING | 19 | 0.207097108 | 0.94754755 | 0.52263 | 0.52263 | 0.39295 | 42 |
| HP_ABNORMAL_RENAL_MORPHOLOGY | 33 | -0.26532062 | -0.9638428 | 0.52509 | 0.52509 | 0.39417 | 83 |
| GOCC_TRANSPORT_VESICLE | 23 | -0.285265038 | -0.9587209 | 0.52551 | 0.52551 | 0.39417 | 68 |
| GOBP_SMALL_MOLECULE_CATABOLIC_PROCESS | 11 | 0.25688445 | 0.93078548 | 0.52782 | 0.52782 | 0.39543 | 15 |
| GOBP_HEAD_DEVELOPMENT | 16 | -0.311901243 | -0.9559637 | 0.53007 | 0.53007 | 0.39665 | 54 |
| GOBP_REGULATION_OF_RESPONSE_TO_BIOTIC_STIMULUS | 14 | -0.322174127 | -0.9520915 | 0.53156 | 0.53156 | 0.39688 | 106 |
| HP_ABNORMALITY_OF_HEPATOBILIARY_SYSTEM_PHYSIOLOGY | 12 | -0.336114184 | -0.9535974 | 0.53251 | 0.53251 | 0.39688 | 66 |
| GOBP_NUCLEOBASE_CONTAINING_SMALL_MOLECULE_METABOLIC_P | 18 | -0.302358879 | -0.9545361 | 0.53333 | 0.53333 | 0.39688 | 178 |
| GOBP_NEGATIVE_REGULATION_OF_GENE_EXPRESSION | 32 | -0.264713727 | -0.9567147 | 0.53355 | 0.53355 | 0.39688 | 72 |
| HP_ABNORMAL_FORM_OF_THE_VERTEBRAL_BODIES | 13 | 0.240396546 | 0.93495429 | 0.53356 | 0.53356 | 0.39688 | 147 |
| HP_AORTIC_ANEURYSM | 10 | -0.350874534 | -0.9434439 | 0.53828 | 0.53828 | 0.39992 | 143 |
| HP_ABNORMALITY_OF_THE_BILIARY_SYSTEM | 15 | -0.315444686 | -0.9483504 | 0.54072 | 0.54072 | 0.40117 | 163 |
| HP_ABNORMALITY_OF_THE_FONTANELLES_OR_CRANIAL_SUTURES | 13 | -0.325008722 | -0.9423295 | 0.54144 | 0.54144 | 0.40117 | 34 |
| HP_ABNORMALITY_OF_THE_ABDOMINAL_ORGANS | 52 | -0.244040013 | -0.9593924 | 0.54188 | 0.54188 | 0.40117 | 96 |
| HP_ABNORMALITY_OF_THE_URETHRA | 14 | -0.316925851 | -0.9365817 | 0.55023 | 0.55023 | 0.40687 | 46 |
| GOMF_HYDROLASE_ACTIVITY_ACTING_ON_ACID_ANHYDRIDES | 23 | -0.278787144 | -0.9369499 | 0.55345 | 0.55345 | 0.40877 | 112 |
| GOMF_OXIDOREDUCTASE_ACTIVITY | 50 | 0.14860831 | 0.94138963 | 0.56049 | 0.56049 | 0.41348 | 35 |
| HP_ABNORMALITY_OF_BONE_MINERAL_DENSITY | 19 | -0.290345545 | -0.929032 | 0.56150 | 0.56150 | 0.41374 | 129 |
| GOCC_GOLGI_ASSOCIATED_VESICLE | 10 | -0.341584158 | -0.9184636 | 0.56737 | 0.56737 | 0.41757 | 277 |
| GOBP_MITOTIC_CELL_CYCLE_PHASE_TRANSITION | 10 | -0.341118067 | -0.9172104 | 0.56888 | 0.56888 | 0.41771 | 28 |
| GOBP_PROTEIN_CONTAINING_COMPLEX_ORGANIZATION | 83 | -0.2278626 | -0.9470425 | 0.56889 | 0.56889 | 0.41771 | 75 |
| GOBP_POSITIVE_REGULATION_OF_RNA_METABOLIC_PROCESS | 36 | 0.158012986 | 0.92082009 | 0.57170 | 0.57170 | 0.41928 | 131 |
| GOBP_LIPID_BIOSYNTHETIC_PROCESS | 15 | -0.306004238 | -0.9199687 | 0.57250 | 0.57250 | 0.41938 | 111 |
| HP_HERNIA | 14 | -0.310336054 | -0.9171075 | 0.57329 | 0.57329 | 0.41946 | 107 |
| GOBP_REGULATION_OF_CELLULAR_RESPONSE_TO_STRESS | 25 | -0.269003305 | -0.9203772 | 0.57532 | 0.57532 | 0.42034 | 150 |
| HP_DOWNSLANTED_PALPEBRAL_FISSURES | 16 | -0.299671839 | -0.9184811 | 0.57584 | 0.57584 | 0.42034 | 46 |
| GOBP_ORGANIC_ACID_BIOSYNTHETIC_PROCESS | 14 | -0.3093884 | -0.914307 | 0.57651 | 0.57651 | 0.42034 | 165 |
| GOMF_ENZYME_REGULATOR_ACTIVITY | 30 | -0.258729068 | -0.9227493 | 0.57726 | 0.57726 | 0.42040 | 147 |
| GOMF_TRANSCRIPTION_COREGULATOR_ACTIVITY | 12 | -0.320610724 | -0.9096122 | 0.58204 | 0.58204 | 0.42302 | 72 |
| GOBP_NUCLEAR_TRANSPORT | 17 | -0.292820603 | -0.911471 | 0.58220 | 0.58220 | 0.42302 | 18 |
| GOBP_CELLULAR_RESPONSE_TO_NITROGEN_COMPOUND | 18 | -0.288972605 | -0.9122761 | 0.58538 | 0.58538 | 0.42483 | 112 |
| GOBP_RESPONSE_TO_PEPTIDE | 13 | -0.310772057 | -0.9010517 | 0.59190 | 0.59190 | 0.42881 | 106 |
| GOBP_NEGATIVE_REGULATION_OF_PHOSPHORYLATION | 12 | -0.317243489 | -0.9000589 | 0.59224 | 0.59224 | 0.42881 | 171 |
| GOBP_REGULATION_OF_CELLULAR_AMIDE_METABOLIC_PROCESS | 22 | -0.273761935 | -0.9085278 | 0.59292 | 0.59292 | 0.42881 | 58 |
| HP_ABNORMAL_CRANIAL_NERVE_MORPHOLOGY | 11 | -0.324321971 | -0.8979137 | 0.59639 | 0.59639 | 0.43083 | 6 |
| GOBP_REPRODUCTION | 28 | -0.255288235 | -0.8973212 | 0.60477 | 0.60477 | 0.43638 | 53 |
| HP_ABNORMAL_HEART_VALVE_MORPHOLOGY | 12 | -0.312493082 | -0.8865814 | 0.60760 | 0.60760 | 0.43727 | 107 |
| HP_ABNORMALITY_OF_THE_OPTIC_NERVE | 33 | 0.160067017 | 0.90229871 | 0.60919 | 0.60919 | 0.43727 | 161 |
| HP_PEDIATRIC_ONSET | 29 | -0.25357621 | -0.8965279 | 0.60932 | 0.60932 | 0.43727 | 66 |
| HP_ABNORMAL_JAW_MORPHOLOGY | 30 | 0.167636017 | 0.91166256 | 0.60955 | 0.60955 | 0.43727 | 66 |
| HP_PATENT_DUCTUS_ARTERIOSUS | 11 | -0.319691685 | -0.8850943 | 0.61000 | 0.61000 | 0.43727 | 107 |
| GOMF_TRANSCRIPTION_REGULATOR_ACTIVITY | 13 | -0.305451416 | -0.8856251 | 0.61020 | 0.61020 | 0.43727 | 72 |
| HP_ORAL_CLEFT | 16 | -0.289213655 | -0.8864272 | 0.61304 | 0.61304 | 0.43881 | 46 |
| GOCC_COATED_VESICLE | 23 | -0.264606025 | -0.8892899 | 0.61589 | 0.61589 | 0.43987 | 116 |
| GOBP_CELL_CYCLE_PHASE_TRANSITION | 11 | -0.317991271 | -0.8803866 | 0.61593 | 0.61593 | 0.43987 | 28 |
| GOBP_ORGANIC_CYCLIC_COMPOUND_CATABOLIC_PROCESS | 19 | 0.192426015 | 0.88042175 | 0.61692 | 0.61692 | 0.44007 | 174 |
| GOBP_CELLULAR_RESPONSE_TO_STRESS | 62 | -0.225190408 | -0.9052131 | 0.61822 | 0.61822 | 0.44050 | 58 |
| GOCC_GOLGI_APPARATUS_SUBCOMPARTMENT | 13 | -0.302160553 | -0.8760836 | 0.62301 | 0.62301 | 0.44341 | 24 |
| HP_BABINSKI_SIGN | 10 | -0.323833598 | -0.8707353 | 0.62541 | 0.62541 | 0.44461 | 33 |
| HP_RECURRENT_RESPIRATORY_INFECTIONS | 19 | -0.273048875 | -0.8736871 | 0.62914 | 0.62914 | 0.44663 | 52 |
| GOCC_ENVELOPE | 56 | 0.136871508 | 0.90479292 | 0.63036 | 0.63036 | 0.44663 | 365 |
| HP_ABNORMAL_INTERNAL_GENITALIA | 12 | -0.305868661 | -0.8677871 | 0.63038 | 0.63038 | 0.44663 | 53 |
| GOBP_CELLULAR_RESPONSE_TO_PEPTIDE | 10 | 0.247919698 | 0.85899249 | 0.63539 | 0.63539 | 0.44967 | 112 |
| HP_JOINT_HYPERMOBILITY | 12 | -0.303040708 | -0.8597639 | 0.63766 | 0.63766 | 0.45067 | 34 |
| HP_SKELETAL_MUSCLE_ATROPHY | 15 | -0.286705447 | -0.861949 | 0.63824 | 0.63824 | 0.45067 | 256 |
| HP_PHENOTYPIC_VARIABILITY | 11 | 0.23573201 | 0.85414253 | 0.64219 | 0.64219 | 0.45295 | 319 |
| HP_ABNORMALITY_OF_PELVIC_GIRDLE_BONE_MORPHOLOGY | 17 | 0.198382817 | 0.86568986 | 0.64325 | 0.64325 | 0.45318 | 129 |
| GOMF_ENZYME_ACTIVATOR_ACTIVITY | 18 | -0.271408164 | -0.8568258 | 0.64461 | 0.64461 | 0.45331 | 147 |
| GOCC_ENDOPLASMIC_RETICULUM_LUMEN | 15 | -0.285022843 | -0.8568904 | 0.64488 | 0.64488 | 0.45331 | 116 |
| GOBP_NEGATIVE_REGULATION_OF_CATABOLIC_PROCESS | 13 | -0.295217782 | -0.8559537 | 0.64680 | 0.64680 | 0.45397 | 33 |
| HP_CRYPTORCHIDISM | 17 | -0.27510705 | -0.8563335 | 0.64726 | 0.64726 | 0.45397 | 46 |
| HP_ABNORMAL_UVEA_MORPHOLOGY | 14 | -0.288313678 | -0.8520268 | 0.64992 | 0.64992 | 0.45491 | 112 |
| GOBP_MITOTIC_CELL_CYCLE | 22 | -0.258723915 | -0.8586214 | 0.65006 | 0.65006 | 0.45491 | 101 |
| HP_MYOCLONUS | 11 | -0.306242266 | -0.8478584 | 0.65098 | 0.65098 | 0.45505 | 33 |
| HP_ABNORMAL_LIP_MORPHOLOGY | 30 | 0.161820038 | 0.88003326 | 0.65493 | 0.65493 | 0.45729 | 84 |

| | | | | | | | |
|---|---|---|---|---|---|---|---|
| HP_HEMATOLOGICAL_NEOPLASM | 12 | -0.297226981 | -0.8432696 | 0.65660 | 0.65660 | 0.45761 | 96 |
| GOCC_MITOCHONDRIAL_MATRIX | 23 | 0.173561046 | 0.85715098 | 0.65684 | 0.65684 | 0.45761 | 275 |
| HP_ABNORMALITY_OF_THE_DENTITION | 27 | -0.244915888 | -0.8540879 | 0.65779 | 0.65779 | 0.45776 | 86 |
| GOBP_PROTEIN_COMPLEX_OLIGOMERIZATION | 13 | -0.290080601 | -0.841059 | 0.66314 | 0.66314 | 0.46043 | 123 |
| HP_PES_CAVUS | 11 | -0.302324338 | -0.8370113 | 0.66325 | 0.66325 | 0.46043 | 81 |
| HP_OPTIC_NEUROPATHY | 11 | 0.232030567 | 0.84073086 | 0.66383 | 0.66383 | 0.46043 | 161 |
| GOBP_GENERATION_OF_PRECURSOR_METABOLITES_AND_ENERGY | 33 | -0.234926943 | -0.8534302 | 0.66625 | 0.66625 | 0.46160 | 86 |
| HP_PES_PLANUS | 17 | -0.269580493 | -0.8391309 | 0.66758 | 0.66758 | 0.46201 | 153 |
| HP_ATROPHY_DEGENERATION_AFFECTING_THE_CENTRAL_NERVOUS_ | 39 | 0.144308823 | 0.86640974 | 0.67076 | 0.67076 | 0.46369 | 174 |
| GOMF_HYDROLASE_ACTIVITY_ACTING_ON_ESTER_BONDS | 13 | -0.287146596 | -0.8325521 | 0.67281 | 0.67281 | 0.46460 | 171 |
| HP_CONGENITAL_ONSET | 13 | -0.286059639 | -0.8294006 | 0.67582 | 0.67582 | 0.46616 | 4 |
| HP_ABNORMALITY_OF_THE_MIDFACE | 16 | -0.27022467 | -0.8282268 | 0.67911 | 0.67911 | 0.46792 | 178 |
| HP_ABNORMALITY_OF_EXTRAPYRAMIDAL_MOTOR_FUNCTION | 10 | -0.308083925 | -0.828387 | 0.68016 | 0.68016 | 0.46813 | 143 |
| HP_NAUSEA_AND_VOMITING | 24 | -0.244653905 | -0.8297368 | 0.68412 | 0.68412 | 0.47033 | 33 |
| HP_APNEA | 15 | -0.273257774 | -0.82152 | 0.68777 | 0.68777 | 0.47233 | 6 |
| HP_ONSET | 50 | -0.215340969 | -0.8430183 | 0.68927 | 0.68927 | 0.47284 | 144 |
| GOBP_INTRACELLULAR_PROTEIN_TRANSPORT | 58 | -0.212730444 | -0.8479947 | 0.69092 | 0.69092 | 0.47345 | 92 |
| GOBP_MULTI_ORGANISM_REPRODUCTIVE_PROCESS | 22 | -0.248341516 | -0.8241656 | 0.69232 | 0.69232 | 0.47389 | 53 |
| GOBP_POSITIVE_REGULATION_OF_DNA_METABOLIC_PROCESS | 10 | -0.30321849 | -0.8153047 | 0.69361 | 0.69361 | 0.47401 | 82 |
| GOBP_ACTIVATION_OF_INNATE_IMMUNE_RESPONSE | 10 | -0.302747562 | -0.8140384 | 0.69553 | 0.69553 | 0.47401 | 106 |
| GOBP_POSITIVE_REGULATION_OF_INNATE_IMMUNE_RESPONSE | 10 | -0.302747562 | -0.8140384 | 0.69553 | 0.69553 | 0.47401 | 106 |
| GOBP_POSITIVE_REGULATION_OF_RESPONSE_TO_BIOTIC_STIMULUS | 10 | -0.302747562 | -0.8140384 | 0.69553 | 0.69553 | 0.47401 | 106 |
| HP_CEREBELLAR_ATROPHY | 15 | 0.197994987 | 0.82207116 | 0.69834 | 0.69834 | 0.47541 | 335 |
| HP_INTELLECTUAL_DISABILITY_MILD | 14 | -0.274060475 | -0.8099057 | 0.70003 | 0.70003 | 0.47604 | 34 |
| HP_ABNORMALITY_OF_THE_LENS | 27 | -0.234339546 | -0.8172054 | 0.70083 | 0.70083 | 0.47607 | 34 |
| HP_ABNORMAL_ESOPHAGUS_PHYSIOLOGY | 31 | -0.227082903 | -0.8168747 | 0.70491 | 0.70491 | 0.47833 | 34 |
| GOBP_INTRACELLULAR_TRANSPORT | 76 | -0.205310627 | -0.8466736 | 0.70701 | 0.70701 | 0.47923 | 92 |
| HP_HYPOREFLEXIA | 13 | -0.275675012 | -0.7992915 | 0.70902 | 0.70902 | 0.48008 | 181 |
| GOBP_REGULATION_OF_PROTEIN_CATABOLIC_PROCESS | 18 | -0.25367748 | -0.8008506 | 0.71214 | 0.71214 | 0.48167 | 87 |
| HP_VASCULAR_DILATATION | 12 | -0.280187861 | -0.7949275 | 0.71368 | 0.71368 | 0.48219 | 143 |
| GOBP_ENERGY_DERIVATION_BY_OXIDATION_OF_ORGANIC_COMPOL | 28 | 0.157725023 | 0.83401359 | 0.71597 | 0.71597 | 0.48266 | 172 |
| HP_APLASIA_HYPOPLASIA_INVOLVING_BONES_OF_THE_LOWER_LIMBS | 10 | -0.294988714 | -0.7931762 | 0.71679 | 0.71679 | 0.48266 | 11 |
| HP_RETROGNATHIA | 14 | 0.2025 | 0.8137338 | 0.71728 | 0.71728 | 0.48266 | 333 |
| HP_REDUCED_CONSCIOUSNESS_CONFUSION | 21 | -0.243307225 | -0.7997013 | 0.71746 | 0.71746 | 0.48266 | 33 |
| HP_APLASIA_HYPOPLASIA_OF_THE_CEREBELLUM | 10 | -0.294197551 | -0.7910489 | 0.71995 | 0.71995 | 0.48330 | 11 |
| HP_CEREBELLAR_HYPOPLASIA | 10 | -0.294197551 | -0.7910489 | 0.71995 | 0.71995 | 0.48330 | 11 |
| HP_ABNORMAL_STERNUM_MORPHOLOGY | 16 | -0.256518413 | -0.7862177 | 0.72299 | 0.72299 | 0.48482 | 164 |
| GOBP_OSSIFICATION | 10 | -0.291182012 | -0.7829406 | 0.72901 | 0.72901 | 0.48833 | 131 |
| HP_ABNORMAL_CELLULAR_PHENOTYPE | 30 | -0.223474786 | -0.797016 | 0.73063 | 0.73063 | 0.48855 | 96 |
| HP_SHORT_NOSE | 11 | -0.281982498 | -0.7806931 | 0.73106 | 0.73106 | 0.48855 | 11 |
| HP_APLASIA_HYPOPLASIA_OF_THE_EXTREMITIES | 19 | 0.174687056 | 0.79925931 | 0.73168 | 0.73168 | 0.48855 | 129 |
| HP_CONGENITAL_MALFORMATION_OF_THE_GREAT_ARTERIES | 13 | -0.267573522 | -0.775802 | 0.73464 | 0.73464 | 0.48940 | 167 |
| HP_PECTUS_EXCAVATUM | 13 | -0.267281736 | -0.774956 | 0.73529 | 0.73529 | 0.48940 | 46 |
| GOBP_ALCOHOL_METABOLIC_PROCESS | 10 | -0.289423619 | -0.7782125 | 0.73642 | 0.73642 | 0.48940 | 147 |
| HP_ABNORMAL_LOWER_LIMB_BONE_MORPHOLOGY | 16 | -0.252503696 | -0.7739128 | 0.73660 | 0.73660 | 0.48940 | 34 |
| GOBP_HOMEOSTASIS_OF_NUMBER_OF_CELLS | 11 | -0.28038887 | -0.776281 | 0.73686 | 0.73686 | 0.48940 | 110 |
| GOCC_ORGANELLE_SUBCOMPARTMENT | 56 | 0.127298304 | 0.84150898 | 0.74464 | 0.74464 | 0.49405 | 208 |
| GOBP_NEGATIVE_REGULATION_OF_TRANSCRIPTION_BY_RNA_POLYMI | 20 | -0.237142371 | -0.7695925 | 0.74631 | 0.74631 | 0.49463 | 45 |
| HP_ABNORMALITY_OF_THE_CURVATURE_OF_THE_VERTEBRAL_COLUM | 38 | 0.138032564 | 0.81674233 | 0.74924 | 0.74924 | 0.49605 | 128 |
| GOBP_ORGANONITROGEN_COMPOUND_CATABOLIC_PROCESS | 37 | -0.211837993 | -0.7846004 | 0.75022 | 0.75022 | 0.49617 | 90 |
| HP_HYPERTROPHIC_CARDIOMYOPATHY | 23 | -0.229545362 | -0.7714578 | 0.75241 | 0.75241 | 0.49680 | 2 |
| HP_ABNORMAL_HAIR_QUANTITY | 31 | -0.216099086 | -0.7773631 | 0.75276 | 0.75276 | 0.49680 | 11 |
| HP_ABNORMAL_5TH_FINGER_MORPHOLOGY | 10 | -0.282956089 | -0.7608224 | 0.75453 | 0.75453 | 0.49745 | 11 |
| GOBP_PROTEIN_CATABOLIC_PROCESS | 30 | -0.215926956 | -0.7700969 | 0.76112 | 0.76112 | 0.50126 | 87 |
| HP_ABNORMAL_PERIPHERAL_NERVOUS_SYSTEM_MORPHOLOGY | 15 | -0.249210566 | -0.7492247 | 0.76398 | 0.76398 | 0.50240 | 81 |
| GOBP_REGULATION_OF_CATABOLIC_PROCESS | 32 | -0.212158758 | -0.7667733 | 0.76445 | 0.76445 | 0.50240 | 58 |
| HP_DEPRESSED_NASAL_BRIDGE | 12 | -0.263895037 | -0.7487028 | 0.76732 | 0.76732 | 0.50375 | 46 |
| GOCC_MEMBRANE_PROTEIN_COMPLEX | 77 | -0.193691333 | -0.7998507 | 0.76845 | 0.76845 | 0.50397 | 51 |
| HP_ABNORMAL_CEREBRAL_CORTEX_MORPHOLOGY | 25 | 0.157572842 | 0.79622777 | 0.77093 | 0.77093 | 0.50483 | 200 |
| HP_SHORT_ATTENTION_SPAN | 11 | -0.267886557 | -0.7416673 | 0.77137 | 0.77137 | 0.50483 | 34 |
| HP_ANORECTAL_ANOMALY | 12 | -0.262087281 | -0.7435739 | 0.77261 | 0.77261 | 0.50511 | 92 |
| HP_ABNORMAL_HAIR_PATTERN | 11 | -0.266948989 | -0.7390716 | 0.77420 | 0.77420 | 0.50562 | 11 |
| GOMF_TRANSPORTER_ACTIVITY | 40 | -0.204825311 | -0.7693468 | 0.77505 | 0.77505 | 0.50565 | 20 |
| HP_ABNORMAL_PYRAMIDAL_SIGN | 13 | -0.254536988 | -0.7380039 | 0.77882 | 0.77882 | 0.50694 | 33 |
| GOBP_NEGATIVE_REGULATION_OF_NUCLEOBASE_CONTAINING_COM | 37 | -0.205841961 | -0.7623924 | 0.77893 | 0.77893 | 0.50694 | 53 |
| GOBP_RESPONSE_TO_PEPTIDE_HORMONE | 11 | -0.26469764 | -0.7328385 | 0.77946 | 0.77946 | 0.50694 | 106 |
| HP_INFANTILE_ONSET | 23 | -0.221334877 | -0.7438639 | 0.78131 | 0.78131 | 0.50762 | 157 |
| HP_ABNORMAL_SYSTEMIC_ARTERIAL_MORPHOLOGY | 20 | -0.226924116 | -0.7364314 | 0.78240 | 0.78240 | 0.50780 | 107 |
| GOBP_HISTONE_MODIFICATION | 13 | 0.195843827 | 0.76167911 | 0.78462 | 0.78462 | 0.50812 | 31 |
| GOBP_REGULATION_OF_INNATE_IMMUNE_RESPONSE | 11 | -0.262896807 | -0.7278527 | 0.78471 | 0.78471 | 0.50812 | 106 |
| HP_ABNORMALITY_OF_THE_NECK | 20 | -0.225926197 | -0.7331929 | 0.78532 | 0.78532 | 0.50812 | 46 |
| HP_ABNORMAL_FINGER_PHALANX_MORPHOLOGY | 15 | -0.242411798 | -0.7287849 | 0.78759 | 0.78759 | 0.50906 | 34 |
| GOMF_ENDOPEPTIDASE_ACTIVITY | 10 | -0.270514204 | -0.7273682 | 0.78869 | 0.78869 | 0.50925 | 45 |
| HP_ABNORMAL_GRANULOCYTE_COUNT | 12 | -0.254805298 | -0.7229141 | 0.79208 | 0.79208 | 0.51038 | 96 |
| HP_ABNORMAL_NEUTROPHIL_COUNT | 12 | -0.254805298 | -0.7229141 | 0.79208 | 0.79208 | 0.51038 | 96 |

| | | | | | | | |
|---|---|---|---|---|---|---|---|
| GOCC_MITOCHONDRIAL_ENVELOPE | 41 | 0.131367292 | 0.79681262 | 0.79487 | 0.79487 | 0.51166 | 365 |
| HP_CLINODACTYLY | 11 | -0.259079426 | -0.717284 | 0.79604 | 0.79604 | 0.51188 | 11 |
| GOBP_ATP_METABOLIC_PROCESS | 29 | 0.143103837 | 0.77113506 | 0.79850 | 0.79850 | 0.51294 | 172 |
| HP_ABNORMALITY_OF_DENTAL_STRUCTURE | 16 | -0.232878914 | -0.7137637 | 0.79977 | 0.79977 | 0.51323 | 46 |
| GOMF_CATION_TRANSMEMBRANE_TRANSPORTER_ACTIVITY | 14 | -0.241816096 | -0.7146168 | 0.80113 | 0.80113 | 0.51357 | 183 |
| HP_ABNORMALITY_OF_THE_SYNOVIA | 11 | -0.255934228 | -0.7085762 | 0.80480 | 0.80480 | 0.51540 | 11 |
| HP_ABNORMAL_TRACHEOBRONCHIAL_MORPHOLOGY | 10 | -0.264049612 | -0.709986 | 0.80571 | 0.80571 | 0.51545 | 157 |
| HP_ABNORMAL_HEART_VALVE_PHYSIOLOGY | 13 | -0.244923741 | -0.7101313 | 0.80758 | 0.80758 | 0.51575 | 107 |
| GOMF_MOLECULAR_ADAPTOR_ACTIVITY | 12 | -0.247442276 | -0.7020243 | 0.81075 | 0.81075 | 0.51575 | 150 |
| GOMF_PROTEIN_MACROMOLECULE_ADAPTOR_ACTIVITY | 12 | -0.247442276 | -0.7020243 | 0.81075 | 0.81075 | 0.51575 | 150 |
| HP_IRREGULAR_HYPERPIGMENTATION | 12 | 0.196808641 | 0.73799564 | 0.81133 | 0.81133 | 0.51575 | 133 |
| HP_ABNORMALITY_IRIS_MORPHOLOGY | 13 | -0.243434653 | -0.7058138 | 0.81163 | 0.81163 | 0.51575 | 112 |
| HP_POSTERIORLY_ROTATED_EARS | 13 | -0.243399678 | -0.7057124 | 0.81190 | 0.81190 | 0.51575 | 46 |
| GOMF_GTPASE_ACTIVATOR_ACTIVITY | 12 | -0.247092036 | -0.7010306 | 0.81195 | 0.81195 | 0.51575 | 147 |
| HP_ABNORMALITY_OF_THE_CURVATURE_OF_THE_CORNEA | 12 | -0.246338795 | -0.6988935 | 0.81300 | 0.81300 | 0.51590 | 11 |
| HP_ABNORMAL_VASCULAR_MORPHOLOGY | 31 | -0.200122711 | -0.7198921 | 0.81554 | 0.81554 | 0.51699 | 112 |
| HP_KYPHOSIS | 13 | -0.242155898 | -0.7021062 | 0.81712 | 0.81712 | 0.51747 | 11 |
| HP_ATROPHY_DEGENERATION_AFFECTING_THE_CEREBRUM | 26 | 0.144993422 | 0.74945682 | 0.81862 | 0.81862 | 0.51754 | 126 |
| GOBP_CELL_CYCLE | 36 | -0.195393706 | -0.7201416 | 0.81889 | 0.81889 | 0.51754 | 103 |
| GOBP_ESTABLISHMENT_OF_PROTEIN_LOCALIZATION_TO_ORGANELLE | 26 | -0.205310766 | -0.7090534 | 0.82088 | 0.82088 | 0.51819 | 18 |
| GOBP_PROTEASOMAL_PROTEIN_CATABOLIC_PROCESS | 17 | 0.167840815 | 0.73241269 | 0.82213 | 0.82213 | 0.51819 | 193 |
| HP_HYPERTELORISM | 26 | 0.144353166 | 0.7461474 | 0.82345 | 0.82345 | 0.51819 | 84 |
| HP_APLASIA_HYPOPLASIA_INVOLVING_BONES_OF_THE_UPPER_LIMBS | 13 | -0.239326289 | -0.6939021 | 0.82431 | 0.82431 | 0.51819 | 46 |
| HP_SLANTING_OF_THE_PALPEBRAL_FISSURE | 23 | -0.208824748 | -0.7018198 | 0.82459 | 0.82459 | 0.51819 | 46 |
| HP_FEEDING_DIFFICULTIES_IN_INFANCY | 17 | -0.221304444 | -0.6888606 | 0.82550 | 0.82550 | 0.51819 | 34 |
| HP_UPPER_MOTOR_NEURON_DYSFUNCTION | 41 | -0.19034592 | -0.7192429 | 0.82570 | 0.82570 | 0.51819 | 33 |
| GOMF_ZINC_ION_BINDING | 17 | -0.220576532 | -0.6865948 | 0.82849 | 0.82849 | 0.51915 | 43 |
| HP_UPSLANTED_PALPEBRAL_FISSURE | 12 | -0.240847985 | -0.6833154 | 0.82890 | 0.82890 | 0.51915 | 46 |
| HP_ABNORMAL_NEPHRON_MORPHOLOGY | 11 | -0.245909584 | -0.6808221 | 0.83230 | 0.83230 | 0.52076 | 236 |
| HP_ABNORMAL_CARDIAC_VENTRICLE_MORPHOLOGY | 27 | -0.199779874 | -0.6966864 | 0.83471 | 0.83471 | 0.52152 | 35 |
| GOBP_REGULATION_OF_RESPONSE_TO_DNA_DAMAGE_STIMULUS | 10 | 0.203213047 | 0.70409283 | 0.83517 | 0.83517 | 0.52152 | 149 |
| GOBP_PROTEIN_TARGETING_TO_MEMBRANE | 10 | -0.254252086 | -0.6836421 | 0.83631 | 0.83631 | 0.52170 | 15 |
| HP_ABNORMALITY_OF_THE_BASAL_GANGLIA | 17 | -0.217871047 | -0.6781734 | 0.83759 | 0.83759 | 0.52179 | 33 |
| HP_NEOPLASM_OF_THE_GASTROINTESTINAL_TRACT | 10 | 0.202573794 | 0.70187794 | 0.83812 | 0.83812 | 0.52179 | 82 |
| HP_ABNORMAL_SHAPE_OF_THE_FRONTAL_REGION | 10 | -0.252685792 | -0.6794306 | 0.84042 | 0.84042 | 0.52189 | 11 |
| GOBP_REGULATION_OF_DNA_METABOLIC_PROCESS | 12 | -0.235485241 | -0.6681007 | 0.84161 | 0.84161 | 0.52189 | 82 |
| GOMF_INORGANIC_MOLECULAR_ENTITY_TRANSMEMBRANE_TRANSP( | 12 | 0.191542289 | 0.7182478 | 0.84189 | 0.84189 | 0.52189 | 337 |
| HP_ABNORMAL_ENZYME_COENZYME_ACTIVITY | 22 | -0.205710952 | -0.6826884 | 0.84202 | 0.84202 | 0.52189 | 197 |
| HP_ABNORMALITY_OF_SKIN_PIGMENTATION | 21 | -0.206119641 | -0.6774733 | 0.84243 | 0.84243 | 0.52189 | 112 |
| GOBP_MITOCHONDRION_ORGANIZATION | 36 | -0.187472449 | -0.6909471 | 0.84390 | 0.84390 | 0.52197 | 73 |
| HP_ABNORMAL_JOINT_MORPHOLOGY | 36 | -0.187326061 | -0.6904075 | 0.84424 | 0.84424 | 0.52197 | 11 |
| HP_PTOSIS | 38 | -0.186942482 | -0.6955937 | 0.84596 | 0.84596 | 0.52252 | 11 |
| HP_ABNORMALITY_OF_FINGER | 29 | 0.135548172 | 0.73042029 | 0.85019 | 0.85019 | 0.52462 | 129 |
| HP_VOMITING | 16 | -0.21277801 | -0.6521553 | 0.85676 | 0.85676 | 0.52787 | 11 |
| HP_INVOLUNTARY_MOVEMENTS | 31 | 0.134328296 | 0.7391801 | 0.85714 | 0.85714 | 0.52787 | 105 |
| HP_ABNORMAL_TESTIS_MORPHOLOGY | 22 | -0.199706513 | -0.6627616 | 0.85930 | 0.85930 | 0.52868 | 46 |
| HP_REGULATION_OF_BINDING | 12 | -0.228312497 | -0.6477507 | 0.86015 | 0.86015 | 0.52868 | 103 |
| GOBP_PROTEIN_LOCALIZATION_TO_ORGANELLE | 41 | -0.17914908 | -0.6769344 | 0.86337 | 0.86337 | 0.53014 | 108 |
| HP_AGENESIS_OF_CORPUS_CALLOSUM | 10 | 0.194525111 | 0.67399086 | 0.86424 | 0.86424 | 0.53015 | 133 |
| HP_ABNORMALITY_OF_THE_CEREBROSPINAL_FLUID | 28 | -0.18954625 | -0.6662425 | 0.86579 | 0.86579 | 0.53058 | 34 |
| HP_RESPIRATORY_INSUFFICIENCY | 22 | -0.196709349 | -0.652815 | 0.86807 | 0.86807 | 0.53146 | 30 |
| HP_ABNORMALITY_OF_THE_AMNIOTIC_FLUID | 12 | -0.224733838 | -0.6375976 | 0.86969 | 0.86969 | 0.53158 | 163 |
| HP_ABNORMAL_REFLEX | 48 | 0.11314346 | 0.72041873 | 0.87060 | 0.87060 | 0.53158 | 177 |
| HP_ABNORMAL_BONE_STRUCTURE | 26 | -0.190137402 | -0.6566512 | 0.87139 | 0.87139 | 0.53158 | 92 |
| GOBP_REGULATION_OF_CELLULAR_PROTEIN_CATABOLIC_PROCESS | 11 | -0.231174492 | -0.6400267 | 0.87166 | 0.87166 | 0.53158 | 275 |
| HP_ABNORMALITY_OF_THE_PHILTRUM | 23 | -0.192390958 | -0.646589 | 0.87442 | 0.87442 | 0.53275 | 46 |
| GOBP_NEGATIVE_REGULATION_OF_PROTEIN_MODIFICATION_PROCES | 17 | -0.203290347 | -0.6327876 | 0.87935 | 0.87935 | 0.53523 | 150 |
| HP_ABNORMAL_AORTIC_MORPHOLOGY | 17 | -0.202887157 | -0.6315326 | 0.88034 | 0.88034 | 0.53531 | 34 |
| GOMF_ATP_DEPENDENT_ACTIVITY | 19 | 0.147424892 | 0.6745246 | 0.88308 | 0.88308 | 0.53601 | 276 |
| GOMF_PEPTIDASE_ACTIVITY | 15 | -0.207857897 | -0.6249023 | 0.88320 | 0.88320 | 0.53601 | 105 |
| GOBP_CARBOHYDRATE_DERIVATIVE_BIOSYNTHETIC_PROCESS | 16 | 0.160706297 | 0.68291363 | 0.88508 | 0.88508 | 0.53629 | 100 |
| GOMF_NUCLEOSIDE_TRIPHOSPHATASE_REGULATOR_ACTIVITY | 15 | -0.20682315 | -0.6217915 | 0.88537 | 0.88537 | 0.53629 | 147 |
| HP_ABNORMALITY_OF_FACIAL_SKELETON | 36 | -0.175249918 | -0.6458998 | 0.88823 | 0.88823 | 0.53750 | 46 |
| HP_ABNORMALITY_OF_TOE | 20 | -0.189543223 | -0.6151201 | 0.89309 | 0.89309 | 0.53992 | 34 |
| HP_ABNORMALITY_OF_LOWER_LIMB_JOINT | 20 | -0.187783667 | -0.6094098 | 0.89784 | 0.89784 | 0.54217 | 34 |
| GOBP_RESPONSE_TO_ENDOPLASMIC_RETICULUM_STRESS | 17 | -0.195906344 | -0.6098032 | 0.89854 | 0.89854 | 0.54217 | 7 |
| GOCC_GOLGI_MEMBRANE | 29 | 0.124675325 | 0.67183044 | 0.90112 | 0.90112 | 0.54320 | 366 |
| HP_ABNORMALITY_OF_MUSCLE_SIZE | 18 | -0.192372827 | -0.6073141 | 0.90496 | 0.90496 | 0.54499 | 312 |
| HP_DYSARTHRIA | 24 | -0.17694318 | -0.6000978 | 0.91078 | 0.91078 | 0.54754 | 33 |
| HP_APLASIA_HYPOPLASIA_AFFECTING_BONES_OF_THE_AXIAL_SKELET( | 24 | -0.176699641 | -0.5992718 | 0.91137 | 0.91137 | 0.54754 | 46 |
| GOBP_EMBRYO_DEVELOPMENT_ENDING_IN_BIRTH_OR_EGG_HATCH | 10 | -0.222349291 | -0.5978607 | 0.91232 | 0.91232 | 0.54754 | 234 |
| HP_ABNORMAL_TENDON_MORPHOLOGY | 14 | -0.19885697 | -0.5876637 | 0.91319 | 0.91319 | 0.54754 | 11 |
| HP_ABNORMALITY_OF_GLOBE_LOCATION | 35 | -0.167379528 | -0.6129098 | 0.91356 | 0.91356 | 0.54754 | 53 |
| HP_ABNORMAL_VERTEBRAL_MORPHOLOGY | 15 | -0.19100875 | -0.5742472 | 0.92047 | 0.92047 | 0.55115 | 11 |
| HP_ABNORMALITY_OF_REFRACTION | 17 | -0.185233492 | -0.5765816 | 0.92347 | 0.92347 | 0.55238 | 11 |

| | | | | | | | |
|---|---|---|---|---|---|---|---|
| HP_ABNORMALITY_OF_THE_PALPEBRAL_FISSURES | 26 | -0.169201007 | -0.5843461 | 0.92529 | 0.92529 | 0.55238 | 46 |
| HP_ABNORMAL_THORAX_MORPHOLOGY | 24 | -0.17082696 | -0.5793548 | 0.92614 | 0.92614 | 0.55238 | 46 |
| HP_WEAKNESS_DUE_TO_UPPER_MOTOR_NEURON_DYSFUNCTION | 15 | 0.155125919 | 0.64407966 | 0.92620 | 0.92620 | 0.55238 | 238 |
| HP_ABNORMALITY_OF_THE_CERVICAL_SPINE | 11 | -0.207109519 | -0.5734008 | 0.92693 | 0.92693 | 0.55238 | 46 |
| HP_GAIT_DISTURBANCE | 34 | -0.16055959 | -0.5858534 | 0.92851 | 0.92851 | 0.55279 | 33 |
| HP_ABNORMALITY_OF_THE_EXTERNAL_NOSE | 21 | -0.171557675 | -0.5638751 | 0.92973 | 0.92973 | 0.55299 | 34 |
| HP_ABNORMAL_PENIS_MORPHOLOGY | 26 | -0.166689968 | -0.5756741 | 0.93207 | 0.93207 | 0.55386 | 46 |
| HP_REDUCED_TENDON_REFLEXES | 17 | -0.179662798 | -0.5592415 | 0.93469 | 0.93469 | 0.55394 | 312 |
| GOBP_GOLGI_VESICLE_TRANSPORT | 23 | -0.167302389 | -0.5622711 | 0.93471 | 0.93471 | 0.55394 | 50 |
| GOBP_REGULATION_OF_CELLULAR_MACROMOLECULE_BIOSYNTHETI( | 27 | -0.16250364 | -0.5666941 | 0.93486 | 0.93486 | 0.55394 | 53 |
| HP_ABNORMALITY_OF_NEURONAL_MIGRATION | 17 | -0.17800654 | -0.554086 | 0.93643 | 0.93643 | 0.55435 | 11 |
| GOBP_POSITIVE_REGULATION_OF_CATABOLIC_PROCESS | 13 | -0.189590912 | -0.5496994 | 0.93869 | 0.93869 | 0.55516 | 58 |
| GOCC_INTRINSIC_COMPONENT_OF_ORGANELLE_MEMBRANE | 16 | -0.175879355 | -0.5390625 | 0.94124 | 0.94124 | 0.55615 | 124 |
| HP_DYSPHAGIA | 21 | -0.164835664 | -0.5417812 | 0.94325 | 0.94325 | 0.55635 | 148 |
| GOBP_CARBOHYDRATE_DERIVATIVE_METABOLIC_PROCESS | 34 | -0.152451325 | -0.5562678 | 0.94416 | 0.94416 | 0.55635 | 151 |
| GOBP_POSITIVE_REGULATION_OF_NUCLEOBASE_CONTAINING_COMF | 42 | -0.148994754 | -0.5653443 | 0.94426 | 0.94426 | 0.55635 | 82 |
| HP_CEREBRAL_CORTICAL_ATROPHY | 10 | -0.203470098 | -0.5470976 | 0.94745 | 0.94745 | 0.55704 | 112 |
| HP_MUSCLE_WEAKNESS | 38 | -0.149047306 | -0.5545897 | 0.94777 | 0.94777 | 0.55704 | 20 |
| HP_ABNORMALITY_OF_UPPER_LIP | 27 | -0.15562922 | -0.5427212 | 0.94870 | 0.94870 | 0.55704 | 46 |
| GOBP_REGULATION_OF_CELL_CYCLE | 22 | -0.162776054 | -0.5402013 | 0.94898 | 0.94898 | 0.55704 | 53 |
| HP_SPASTICITY | 36 | 0.113357098 | 0.66058807 | 0.95029 | 0.95029 | 0.55728 | 305 |
| HP_ABNORMALITY_OF_COORDINATION | 43 | -0.145441064 | -0.554389 | 0.95178 | 0.95178 | 0.55734 | 33 |
| GOBP_CELLULAR_RESPONSE_TO_HORMONE_STIMULUS | 13 | -0.18248704 | -0.5291025 | 0.95216 | 0.95216 | 0.55734 | 53 |
| HP_LONG_PHILTRUM | 14 | -0.177940436 | -0.525851 | 0.95415 | 0.95415 | 0.55798 | 46 |
| HP_ABNORMAL_ANTERIOR_EYE_SEGMENT_MORPHOLOGY | 43 | -0.140661741 | -0.5361712 | 0.95964 | 0.95964 | 0.56059 | 11 |
| HP_ABNORMALITY_OF_THE_CEREBRAL_SUBCORTEX | 48 | -0.138595681 | -0.5373343 | 0.96039 | 0.96039 | 0.56059 | 66 |
| GOBP_ESTABLISHMENT_OF_PROTEIN_LOCALIZATION_TO_MEMBRANE | 17 | -0.161107127 | -0.5014827 | 0.96298 | 0.96298 | 0.56150 | 15 |
| HP_ABNORMAL_MORPHOLOGY_OF_THE_GREAT_VESSELS | 22 | -0.152186772 | -0.5050589 | 0.96483 | 0.96483 | 0.56150 | 34 |
| HP_LANGUAGE_IMPAIRMENT | 29 | 0.108172514 | 0.58290273 | 0.96554 | 0.96554 | 0.56150 | 134 |
| HP_HYPERTONIA | 41 | -0.13509829 | -0.5104836 | 0.96683 | 0.96683 | 0.56150 | 33 |
| GOBP_PROTEIN_TARGETING | 20 | -0.152284264 | -0.4942045 | 0.96696 | 0.96696 | 0.56150 | 355 |
| HP_ABNORMALITY_OF_THE_MUSCULATURE_OF_THE_LIMBS | 17 | -0.157855181 | -0.4913603 | 0.96734 | 0.96734 | 0.56150 | 11 |
| GOCC_ENDOPLASMIC_RETICULUM | 75 | 0.087582715 | 0.60809294 | 0.96830 | 0.96830 | 0.56154 | 208 |
| HP_ABNORMAL_PALATE_MORPHOLOGY | 30 | -0.138908872 | -0.4954142 | 0.96951 | 0.96951 | 0.56172 | 164 |
| GOBP_MACROMOLECULE_CATABOLIC_PROCESS | 44 | -0.133396843 | -0.5097001 | 0.97078 | 0.97078 | 0.56193 | 87 |
| HP_ABNORMAL_LIVER_MORPHOLOGY | 37 | 0.103168319 | 0.60727704 | 0.97239 | 0.97239 | 0.56205 | 351 |
| HP_TREMOR | 10 | 0.156987804 | 0.54393155 | 0.97314 | 0.97314 | 0.56205 | 134 |
| HP_IMMUNODEFICIENCY | 13 | 0.141704937 | 0.55112123 | 0.97366 | 0.97366 | 0.56205 | 126 |
| GOBP_GAMETE_GENERATION | 15 | -0.156379131 | -0.470137 | 0.97715 | 0.97715 | 0.56354 | 90 |
| HP_PROGRESSIVE | 18 | 0.122302795 | 0.54864992 | 0.97879 | 0.97879 | 0.56395 | 54 |
| HP_ABNORMAL_CEREBRAL_VENTRICLE_MORPHOLOGY | 30 | 0.100828873 | 0.54834224 | 0.97966 | 0.97966 | 0.56395 | 147 |
| HP_HEPATOMEGALY | 27 | 0.102716412 | 0.5366019 | 0.98080 | 0.98080 | 0.56408 | 133 |
| HP_HYPERACTIVITY | 12 | 0.140412693 | 0.52652137 | 0.98411 | 0.98411 | 0.56547 | 160 |
| GOBP_DEVELOPMENTAL_PROCESS_INVOLVED_IN_REPRODUCTION | 18 | -0.137422623 | -0.4338383 | 0.98612 | 0.98612 | 0.56548 | 53 |
| HP_ABNORMAL_SIZE_OF_THE_PALPEBRAL_FISSURES | 10 | 0.14464957 | 0.50118202 | 0.98823 | 0.98823 | 0.56548 | 43 |
| HP_RENAL_CYST | 10 | -0.163992352 | -0.4409485 | 0.98834 | 0.98834 | 0.56548 | 83 |
| HP_HIGH_PALATE | 24 | -0.123943388 | -0.4203505 | 0.99031 | 0.99031 | 0.56548 | 11 |
| HP_ABNORMAL_EMOTION_AFFECT_BEHAVIOR | 27 | -0.120660044 | -0.4207742 | 0.99093 | 0.99093 | 0.56548 | 126 |
| GOBP_PROTEIN_FOLDING | 13 | -0.145561627 | -0.422041 | 0.99137 | 0.99137 | 0.56548 | 58 |
| HP_DYSTONIA | 26 | -0.120500902 | -0.4161573 | 0.99287 | 0.99287 | 0.56548 | 33 |
| HP_ATTENTION_DEFICIT_HYPERACTIVITY_DISORDER | 10 | -0.151854497 | -0.4083118 | 0.99300 | 0.99300 | 0.56548 | 178 |
| GOCC_COATED_VESICLE_MEMBRANE | 18 | -0.127028878 | -0.4010256 | 0.99356 | 0.99356 | 0.56548 | 22 |
| HP_ABNORMALITY_OF_THE_THYROID_GLAND | 10 | -0.142537992 | -0.3832612 | 0.99547 | 0.99547 | 0.56548 | 194 |
| HP_ABNORMALITY_OF_THE_LIVER | 42 | -0.097557556 | -0.3701715 | 0.99660 | 0.99660 | 0.56548 | 92 |
| HP_PACE_OF_PROGRESSION | 21 | 0.09578785 | 0.45543647 | 0.99767 | 0.99767 | 0.56548 | 54 |
| HP_MOTOR_DELAY | 29 | -0.097610003 | -0.3451037 | 0.99862 | 0.99862 | 0.56548 | 11 |
| GOCC_MITOCHONDRION | 72 | -0.085585375 | -0.3505747 | 0.99886 | 0.99886 | 0.56548 | 86 |
| HP_ABNORMAL_RETINAL_MORPHOLOGY | 34 | -0.088200397 | -0.3218276 | 0.99910 | 0.99910 | 0.56548 | 46 |
| HP_FAILURE_TO_THRIVE | 43 | -0.0790598 | -0.3013584 | 0.99945 | 0.99945 | 0.56548 | 373 |
| GOBP_NEGATIVE_REGULATION_OF_CELLULAR_CATABOLIC_PROCESS | 12 | -0.110903511 | -0.3146469 | 0.99947 | 0.99947 | 0.56548 | 295 |

**Supplemental Table S17.** Significantly different proteins between monoallelic *TP53* compared to WT *TP53* .

| gene | proteinId | Mean Difference | Mean expression (monoallelic *TP53* ) | Mean expression (WT *TP53* ) | statistic | p.value | parameter | conf.low | conf.high | p.adjust |
|---|---|---|---|---|---|---|---|---|---|---|
| SUGT1 | Q9Y2Z0 | -1.045 | 18.780 | 19.826 | -4.788 | 0.000 | 26.923 | -1.493 | -0.597 | 0.139 |
| IDI1 | Q13907 | -0.943 | 19.016 | 19.958 | -4.545 | 0.000 | 25.171 | -1.370 | -0.516 | 0.153 |
| ADD1 | P35611 | 1.010 | 21.038 | 20.028 | 4.117 | 0.000 | 26.738 | 0.506 | 1.513 | 0.213 |
| ATP1B3 | P54709,P54709 | -1.476 | 19.318 | 20.794 | -4.081 | 0.000 | 25.789 | -2.221 | -0.732 | 0.213 |
| SERPINB6 | P35237 | 1.731 | 23.110 | 21.379 | 3.983 | 0.001 | 24.581 | 0.835 | 2.627 | 0.213 |
| LSM1 | O15116,O15116 | -1.447 | 18.867 | 20.314 | -4.008 | 0.001 | 22.305 | -2.195 | -0.699 | 0.213 |
| TMEM208 | Q9BTX3 | -1.044 | 18.896 | 19.940 | -3.933 | 0.001 | 25.232 | -1.590 | -0.497 | 0.213 |
| ALDH16A1 | Q8IZ83,Q8IZ83 | 1.100 | 22.269 | 21.168 | 3.615 | 0.001 | 24.519 | 0.473 | 1.728 | 0.434 |
| DLD | P09622,P09622 | -0.727 | 23.617 | 24.344 | -3.612 | 0.002 | 19.091 | -1.148 | -0.306 | 0.478 |
| OAS3 | Q9Y6K5,Q9Y6K5 | -0.967 | 18.365 | 19.332 | -3.975 | 0.002 | 11.187 | -1.501 | -0.433 | 0.478 |
| AP1G1 | O43747,O43747 | 0.792 | 22.287 | 21.495 | 3.401 | 0.002 | 23.949 | 0.311 | 1.272 | 0.478 |
| MRPS28 | Q9Y2Q9,Q9Y2Q9 | -0.782 | 19.295 | 20.077 | -3.624 | 0.002 | 15.460 | -1.241 | -0.323 | 0.478 |
| PLEC | Q15149,Q15149 | 2.212 | 26.327 | 24.115 | 3.652 | 0.002 | 14.735 | 0.919 | 3.505 | 0.478 |
| GMFB | P60983 | -1.290 | 18.914 | 20.204 | -3.411 | 0.003 | 16.563 | -2.089 | -0.491 | 0.620 |
| NDUFAF2 | Q8N183,Q8N183 | -1.659 | 19.982 | 21.641 | -3.514 | 0.004 | 12.107 | -2.687 | -0.632 | 0.620 |
| ASNA1 | O43681,O43681 | -0.373 | 22.477 | 22.850 | -3.169 | 0.004 | 23.188 | -0.616 | -0.130 | 0.620 |
| PDHX | O00330 | -1.046 | 20.477 | 21.523 | -3.253 | 0.004 | 18.568 | -1.720 | -0.372 | 0.620 |
| TKT | P29401 | -0.642 | 26.650 | 27.292 | -3.175 | 0.004 | 22.141 | -1.062 | -0.223 | 0.620 |
| UBE2L6 | O14933 | -1.214 | 18.475 | 19.689 | -3.118 | 0.005 | 24.597 | -2.016 | -0.411 | 0.620 |
| NHP2 | Q9NX24,Q9NX24 | -1.111 | 18.591 | 19.702 | -3.305 | 0.005 | 14.190 | -1.830 | -0.391 | 0.655 |
| VDAC1 | P21796,P21796 | -0.732 | 26.678 | 27.410 | -3.152 | 0.005 | 18.055 | -1.221 | -0.244 | 0.655 |
| PSMB4 | P28070 | -0.638 | 23.238 | 23.876 | -3.219 | 0.006 | 15.300 | -1.060 | -0.216 | 0.655 |
| TALDO1 | P37837 | -0.369 | 25.447 | 25.817 | -2.989 | 0.006 | 26.682 | -0.623 | -0.116 | 0.664 |
| PDCD4 | Q53EL6,Q53EL6 | -2.218 | 20.272 | 22.490 | -3.112 | 0.008 | 13.482 | -3.752 | -0.684 | 0.847 |
| LGALS3 | P17931 | -2.123 | 19.946 | 22.069 | -3.537 | 0.008 | 7.634 | -3.519 | -0.727 | 0.847 |
| RRM2 | P31350,P31350 | -1.171 | 18.699 | 19.870 | -2.952 | 0.009 | 17.624 | -2.006 | -0.336 | 0.852 |
| FLOT1 | O75955,O75955 | 1.283 | 23.593 | 22.310 | 2.790 | 0.010 | 26.995 | 0.339 | 2.226 | 0.852 |
| S100A6 | P06703 | 1.889 | 24.763 | 22.874 | 2.838 | 0.010 | 21.993 | 0.509 | 3.269 | 0.852 |
| PSMB6 | P28072,P28072 | -1.697 | 19.815 | 21.512 | -3.172 | 0.010 | 10.285 | -2.885 | -0.509 | 0.852 |
| CLTC | Q00610,Q00610 | 1.068 | 27.490 | 26.422 | 2.715 | 0.011 | 26.705 | 0.260 | 1.875 | 0.981 |
| GAPDH | P04406,P04406 | 0.538 | 29.477 | 28.939 | 2.769 | 0.012 | 19.584 | 0.132 | 0.944 | 0.993 |
| HTATSF1 | O43719 | -0.788 | 18.860 | 19.649 | -2.761 | 0.015 | 14.941 | -1.397 | -0.180 | 0.997 |
| MRPL45,MRPL45 | Q9BRJ2,Q9BRJ2 | -0.803 | 19.724 | 20.528 | -2.616 | 0.015 | 25.354 | -1.435 | -0.171 | 0.997 |
| AP3B1 | O00203,O00203 | 0.728 | 21.977 | 21.249 | 2.600 | 0.015 | 26.595 | 0.153 | 1.303 | 0.997 |
| GIMAP4 | Q9NUV9 | -0.718 | 19.206 | 19.925 | -2.566 | 0.016 | 26.917 | -1.293 | -0.144 | 0.997 |
| KCNAB2 | Q13303 | -0.828 | 19.223 | 20.050 | -2.625 | 0.016 | 19.713 | -1.486 | -0.169 | 0.997 |
| EIF3J | O75822,O75822 | -0.830 | 22.340 | 23.170 | -2.674 | 0.016 | 16.382 | -1.487 | -0.173 | 0.997 |
| CTSD | P07339,P07339 | -0.577 | 24.749 | 25.326 | -2.735 | 0.017 | 13.007 | -1.032 | -0.121 | 0.997 |
| GRHPR | Q9UBQ7,Q9UBQ7 | -0.496 | 22.882 | 23.378 | -2.576 | 0.017 | 22.660 | -0.894 | -0.097 | 0.997 |
| TNFAIP8 | O95379 | -1.274 | 18.987 | 20.262 | -2.673 | 0.017 | 15.154 | -2.289 | -0.259 | 0.997 |
| APMAP | Q9HDC9 | -0.618 | 23.721 | 24.339 | -2.691 | 0.017 | 14.187 | -1.109 | -0.126 | 0.997 |
| PSMC6 | P62333 | -0.748 | 22.353 | 23.102 | -2.668 | 0.019 | 13.057 | -1.354 | -0.143 | 0.997 |
| HLA-B | P10319,P10319 | 1.013 | 24.390 | 23.377 | 2.493 | 0.020 | 24.416 | 0.175 | 1.851 | 0.997 |
| STIP1 | P31948,P31948 | -0.391 | 25.051 | 25.442 | -2.476 | 0.021 | 22.629 | -0.718 | -0.064 | 0.997 |

| | | | | | | | | | | |
|---|---|---|---|---|---|---|---|---|---|---|
| PSMA4 | P25789 | -0.497 | 24.173 | 24.669 | -2.660 | 0.022 | 11.438 | -0.905 | -0.088 | 0.997 |
| TUBG2,TUBG1 | Q9NRH3,P23258 | -0.566 | 18.835 | 19.400 | -2.571 | 0.022 | 14.597 | -1.036 | -0.096 | 0.997 |
| MCTS1 | Q9ULC4,Q9ULC4 | -1.049 | 20.056 | 21.105 | -2.559 | 0.022 | 14.898 | -1.924 | -0.175 | 0.997 |
| AP2M1 | Q96CW1,Q96CW1 | 0.641 | 20.330 | 19.688 | 2.466 | 0.022 | 21.584 | 0.101 | 1.182 | 0.997 |
| BAG1 | Q99933,Q99933 | -0.590 | 19.126 | 19.716 | -2.432 | 0.023 | 24.647 | -1.089 | -0.090 | 0.997 |
| MRPL49 | Q13405 | -1.023 | 19.789 | 20.812 | -2.464 | 0.023 | 20.293 | -1.888 | -0.158 | 0.997 |
| SEC24A | O95486,O95486 | 0.732 | 22.225 | 21.493 | 2.468 | 0.023 | 19.792 | 0.113 | 1.351 | 0.997 |
| C21orf33 | P30042,P30042 | -1.020 | 23.715 | 24.736 | -2.712 | 0.024 | 9.154 | -1.869 | -0.171 | 0.997 |
| PGP | A6NDG6 | -0.718 | 18.806 | 19.524 | -2.577 | 0.024 | 12.282 | -1.324 | -0.113 | 0.997 |
| ACP2 | P11117,P11117 | -1.130 | 20.746 | 21.876 | -2.598 | 0.024 | 11.443 | -2.082 | -0.177 | 0.997 |
| PSMA6 | P60900,P60900 | -0.432 | 24.565 | 24.997 | -2.398 | 0.025 | 22.407 | -0.806 | -0.059 | 0.997 |
| PSMA7 | O14818 | -0.406 | 24.539 | 24.945 | -2.425 | 0.027 | 16.245 | -0.761 | -0.052 | 0.997 |
| SMIM20 | Q8N5G0 | -0.660 | 19.101 | 19.761 | -2.385 | 0.027 | 19.297 | -1.238 | -0.081 | 0.997 |
| MRPS26 | Q9BYN8,Q9BYN8 | -1.021 | 18.669 | 19.690 | -2.708 | 0.028 | 7.715 | -1.896 | -0.146 | 0.997 |
| HSPB1 | P04792 | 1.798 | 24.437 | 22.638 | 2.423 | 0.028 | 15.714 | 0.222 | 3.374 | 0.997 |
| MRPL12 | P52815 | -0.766 | 22.578 | 23.345 | -2.428 | 0.028 | 15.302 | -1.438 | -0.095 | 0.997 |
| MYO1F | O00160 | -1.050 | 18.492 | 19.542 | -2.425 | 0.029 | 14.671 | -1.974 | -0.125 | 0.997 |
| PSMA1,PSMA1 | P25786 | -0.464 | 24.138 | 24.601 | -2.408 | 0.030 | 14.485 | -0.875 | -0.052 | 0.997 |
| KIF13B | Q9NQT8,Q9NQT8 | -0.545 | 19.189 | 19.735 | -2.358 | 0.030 | 17.425 | -1.033 | -0.058 | 0.997 |
| CPT2 | P23786 | -0.658 | 22.988 | 23.646 | -2.285 | 0.030 | 27.000 | -1.250 | -0.067 | 0.997 |
| CECR5 | Q9BXW7,Q9BXW7 | -0.944 | 19.168 | 20.113 | -2.629 | 0.031 | 7.829 | -1.775 | -0.113 | 0.997 |
| ACOT9 | Q9Y305 | 0.647 | 20.742 | 20.095 | 2.289 | 0.031 | 23.474 | 0.063 | 1.232 | 0.997 |
| ADH5 | P11766,P11766 | 0.465 | 23.252 | 22.786 | 2.262 | 0.032 | 26.645 | 0.043 | 0.888 | 0.997 |
| TOMM6 | Q96B49 | -0.792 | 18.944 | 19.736 | -2.296 | 0.032 | 20.967 | -1.509 | -0.075 | 0.997 |
| ORMDL1,ORMDL2,ORMDL3 | Q9P0S3,Q53FV1,Q8N138,Q9P0S3,Q53FV1,Q8N138 | 0.596 | 20.354 | 19.758 | 2.321 | 0.032 | 18.013 | 0.057 | 1.135 | 0.997 |
| APOD | P05090,P05090 | -0.654 | 18.903 | 19.556 | -2.258 | 0.033 | 25.650 | -1.249 | -0.058 | 0.997 |
| PEX19 | P40855,P40855 | -0.671 | 19.052 | 19.723 | -2.299 | 0.033 | 19.103 | -1.283 | -0.060 | 0.997 |
| PSMB1 | P20618 | -0.589 | 23.622 | 24.212 | -2.420 | 0.033 | 11.572 | -1.122 | -0.057 | 0.997 |
| OGDH | Q02218,Q02218 | -0.719 | 23.832 | 24.551 | -2.490 | 0.034 | 9.086 | -1.371 | -0.067 | 0.997 |
| HN1 | Q9UK76 | -0.937 | 19.216 | 20.154 | -2.309 | 0.036 | 15.001 | -1.803 | -0.072 | 0.997 |
| UCHL3 | P15374 | -1.233 | 19.511 | 20.744 | -2.433 | 0.037 | 9.411 | -2.372 | -0.094 | 0.997 |
| CUL4B | Q13620,Q13620 | 0.816 | 21.502 | 20.686 | 2.244 | 0.037 | 19.117 | 0.055 | 1.576 | 0.997 |
| UBA3 | Q8TBC4,Q8TBC4 | 0.765 | 21.246 | 20.480 | 2.227 | 0.038 | 19.870 | 0.048 | 1.482 | 0.997 |
| PGAM1 | P18669,P18669 | -0.481 | 25.886 | 26.367 | -2.196 | 0.038 | 24.671 | -0.932 | -0.030 | 0.997 |
| KPNB1 | Q14974,Q14974 | -0.372 | 25.305 | 25.677 | -2.197 | 0.038 | 24.229 | -0.722 | -0.023 | 0.997 |
| PAFAH1B1 | P43034,P43034 | 0.618 | 22.301 | 21.682 | 2.174 | 0.039 | 26.869 | 0.035 | 1.202 | 0.997 |
| SSBP1 | Q04837,Q04837 | -0.819 | 24.443 | 25.261 | -2.393 | 0.040 | 9.052 | -1.592 | -0.046 | 0.997 |
| ADCK3 | Q8NI60,Q8NI60 | 0.765 | 20.283 | 19.517 | 2.263 | 0.041 | 13.189 | 0.036 | 1.495 | 0.997 |
| GRSF1 | Q12849,Q12849 | -0.532 | 19.329 | 19.861 | -2.130 | 0.043 | 26.761 | -1.045 | -0.019 | 0.997 |
| DR1 | Q01658 | -0.787 | 19.162 | 19.949 | -2.263 | 0.043 | 11.899 | -1.545 | -0.029 | 0.997 |
| DNAJC7 | Q99615,Q99615 | -0.876 | 19.155 | 20.031 | -2.427 | 0.045 | 7.185 | -1.725 | -0.027 | 0.997 |
| GLUD1,GLUD2 | P00367,P49448 | -0.775 | 24.736 | 25.512 | -2.278 | 0.045 | 10.218 | -1.531 | -0.019 | 0.997 |
| MPDU1 | O75352,O75352 | -0.724 | 20.867 | 21.591 | -2.301 | 0.046 | 9.492 | -1.431 | -0.018 | 0.997 |
| HLA-A | P01892,P01892 | 0.906 | 26.371 | 25.464 | 2.270 | 0.046 | 10.119 | 0.018 | 1.794 | 0.997 |
| PFKFB2 | O60825,O60825 | -0.634 | 19.052 | 19.686 | -2.141 | 0.047 | 17.557 | -1.257 | -0.011 | 0.997 |
| APOOL | Q6UXV4,Q6UXV4 | 0.969 | 21.080 | 20.111 | 2.184 | 0.047 | 13.727 | 0.016 | 1.923 | 0.997 |
| SIRT3 | Q9NTG7,Q9NTG7 | -0.655 | 19.472 | 20.127 | -2.158 | 0.047 | 15.628 | -1.300 | -0.010 | 0.997 |

| | | | | | | | | | | |
|---|---|---|---|---|---|---|---|---|---|---|
| TMEM41B | Q5BJD5 | 0.526 | 20.123 | 19.596 | 2.104 | 0.047 | 22.194 | 0.008 | 1.045 | 0.997 |
| MEF2D | Q14814,Q14814 | 0.979 | 20.418 | 19.439 | 2.413 | 0.048 | 6.805 | 0.014 | 1.944 | 0.997 |
| AP1M1 | Q9BXS5,Q9BXS5 | 0.557 | 22.230 | 21.673 | 2.079 | 0.048 | 24.531 | 0.005 | 1.109 | 0.997 |
| CBR1 | P16152,P16152 | 0.625 | 24.321 | 23.695 | 2.203 | 0.048 | 11.692 | 0.005 | 1.246 | 0.997 |
| ALDH5A1 | P51649,P51649 | -1.623 | 20.227 | 21.851 | -2.225 | 0.050 | 10.137 | -3.246 | -0.001 | 0.997 |

**Supplemental Table S18.** Gene set enrichment analysis of the C5 library from significantly expressed proteins of the comparison between monoallelic *TP53* to WT *TP53* samples of the FIMM cohort.

| ID | setSize | enrichmentScore | NES | pvalue | p.adjust | qvalue | rank |
|---|---|---|---|---|---|---|---|
| GOCC_MEMBRANE_PROTEIN_COMPLEX | 13 | 0.588 | 2.146 | 0.00100 | 0.00100 | 0.05900 | 22 |
| GOCC_VESICLE_MEMBRANE | 12 | 0.521 | 1.847 | 0.01000 | 0.01000 | 0.22900 | 23 |
| GOBP_INTRACELLULAR_PROTEIN_TRANSPORT | 11 | 0.538 | 1.844 | 0.01300 | 0.01300 | 0.22900 | 23 |
| GOBP_INTRACELLULAR_TRANSPORT | 15 | 0.45 | 1.724 | 0.01800 | 0.01800 | 0.22900 | 28 |
| GOCC_ORGANELLE_SUBCOMPARTMENT | 12 | 0.491 | 1.74 | 0.02000 | 0.02000 | 0.22900 | 28 |
| GOCC_MITOCHONDRIAL_MATRIX | 15 | -0.5 | -1.696 | 0.02400 | 0.02400 | 0.22900 | 40 |
| GOBP_NITROGEN_COMPOUND_TRANSPORT | 16 | 0.421 | 1.651 | 0.02800 | 0.02800 | 0.22900 | 23 |
| GOBP_CELLULAR_RESPONSE_TO_STRESS | 11 | 0.489 | 1.676 | 0.03000 | 0.03000 | 0.22900 | 5 |
| GOBP_CELLULAR_MACROMOLECULE_LOCALIZATION | 16 | 0.415 | 1.627 | 0.03200 | 0.03200 | 0.22900 | 19 |
| GOCC_RIBONUCLEOPROTEIN_COMPLEX | 13 | -0.504 | -1.632 | 0.03700 | 0.03700 | 0.23900 | 35 |
| GOMF_PROTEIN_DIMERIZATION_ACTIVITY | 11 | 0.462 | 1.583 | 0.04500 | 0.04500 | 0.26700 | 15 |
| GOBP_IMMUNE_RESPONSE | 11 | 0.457 | 1.566 | 0.04900 | 0.04900 | 0.26700 | 17 |
| GOCC_MICROTUBULE_CYTOSKELETON | 10 | 0.469 | 1.548 | 0.05500 | 0.05500 | 0.27800 | 44 |
| GOBP_GENERATION_OF_PRECURSOR_METABOLITES_AND_ENERGY | 10 | 0.444 | 1.467 | 0.07800 | 0.07800 | 0.36200 | 55 |
| GOCC_ORGANELLE_INNER_MEMBRANE | 10 | -0.475 | -1.409 | 0.10800 | 0.10800 | 0.43200 | 45 |
| GOCC_SUPRAMOLECULAR_COMPLEX | 13 | 0.373 | 1.362 | 0.11900 | 0.11900 | 0.43200 | 7 |
| GOBP_TISSUE_DEVELOPMENT | 11 | 0.396 | 1.357 | 0.12000 | 0.12000 | 0.43200 | 21 |
| HP_ABNORMAL_EYELID_MORPHOLOGY | 12 | 0.379 | 1.344 | 0.13000 | 0.13000 | 0.43200 | 21 |
| GOBP_NEGATIVE_REGULATION_OF_RESPONSE_TO_STIMULUS | 11 | -0.447 | -1.37 | 0.13000 | 0.13000 | 0.43200 | 5 |
| GOBP_ESTABLISHMENT_OF_PROTEIN_LOCALIZATION | 16 | 0.341 | 1.337 | 0.13200 | 0.13200 | 0.43200 | 23 |
| GOBP_MACROMOLECULE_CATABOLIC_PROCESS | 14 | -0.404 | -1.341 | 0.14500 | 0.14500 | 0.45100 | 12 |
| GOCC_MITOCHONDRION | 25 | -0.328 | -1.284 | 0.17600 | 0.17600 | 0.50700 | 46 |
| GOMF_MOLECULAR_FUNCTION_REGULATOR | 10 | -0.43 | -1.276 | 0.18200 | 0.18200 | 0.50700 | 10 |
| GOCC_VACUOLE | 10 | 0.383 | 1.265 | 0.18900 | 0.18900 | 0.50700 | 23 |
| GOBP_ORGANONITROGEN_COMPOUND_CATABOLIC_PROCESS | 15 | -0.372 | -1.261 | 0.20300 | 0.20300 | 0.50700 | 12 |
| GOCC_MITOCHONDRIAL_ENVELOPE | 14 | -0.371 | -1.231 | 0.21900 | 0.21900 | 0.50700 | 45 |

| | | | | | | | |
|---|---|---|---|---|---|---|---|
| GOBP_REGULATION_OF_TRANSPORT | 12 | -0.389 | -1.225 | 0.22900 | 0.22900 | 0.50700 | 7 |
| HP_ABNORMAL_HAIR_MORPHOLOGY | 10 | 0.366 | 1.207 | 0.23200 | 0.23200 | 0.50700 | 19 |
| HP_ABNORMALITY_OF_THE_OUTER_EAR | 12 | 0.337 | 1.193 | 0.24500 | 0.24500 | 0.50700 | 21 |
| GOBP_CELLULAR_MACROMOLECULE_BIOSYNTHETIC_PROCESS | 10 | -0.402 | -1.192 | 0.25400 | 0.25400 | 0.50700 | 39 |
| GOBP_DEFENSE_RESPONSE | 12 | -0.378 | -1.193 | 0.25400 | 0.25400 | 0.50700 | 3 |
| HP_ABNORMALITY_OF_THE_LIVER | 11 | -0.388 | -1.189 | 0.25600 | 0.25600 | 0.50700 | 49 |
| HP_ABNORMAL_LIVER_MORPHOLOGY | 11 | -0.388 | -1.189 | 0.25600 | 0.25600 | 0.50700 | 49 |
| GOMF_RNA_BINDING | 23 | -0.307 | -1.173 | 0.26900 | 0.26900 | 0.50800 | 35 |
| HP_ABNORMAL_ANTERIOR_EYE_SEGMENT_MORPHOLOGY | 10 | 0.348 | 1.15 | 0.28200 | 0.28200 | 0.50800 | 17 |
| GOBP_REGULATION_OF_IMMUNE_SYSTEM_PROCESS | 10 | 0.345 | 1.14 | 0.29300 | 0.29300 | 0.50800 | 17 |
| GOBP_CELLULAR_PROTEIN_CATABOLIC_PROCESS | 11 | -0.374 | -1.147 | 0.29500 | 0.29500 | 0.50800 | 12 |
| GOMF_IDENTICAL_PROTEIN_BINDING | 17 | 0.283 | 1.134 | 0.29600 | 0.29600 | 0.50800 | 10 |
| GOBP_BIOLOGICAL_PROCESS_INVOLVED_IN_INTERSPECIES_INTER/ | 10 | -0.383 | -1.134 | 0.31200 | 0.31200 | 0.52200 | 3 |
| HP_HEPATOMEGALY | 10 | -0.369 | -1.093 | 0.35500 | 0.35500 | 0.56300 | 49 |
| GOBP_PROTEIN_CATABOLIC_PROCESS | 12 | -0.344 | -1.084 | 0.35700 | 0.35700 | 0.56300 | 12 |
| GOBP_REGULATION_OF_PROTEIN_LOCALIZATION | 10 | -0.366 | -1.084 | 0.36300 | 0.36300 | 0.56300 | 5 |
| GOBP_REGULATION_OF_RESPONSE_TO_STRESS | 11 | 0.296 | 1.013 | 0.43600 | 0.43600 | 0.63200 | 14 |
| HP_GENERALIZED_HYPOTONIA | 13 | -0.313 | -1.013 | 0.43900 | 0.43900 | 0.63200 | 48 |
| GOBP_CELLULAR_LIPID_METABOLIC_PROCESS | 10 | 0.303 | 1 | 0.44500 | 0.44500 | 0.63200 | 54 |
| GOBP_LIPID_METABOLIC_PROCESS | 10 | 0.303 | 1 | 0.44500 | 0.44500 | 0.63200 | 54 |
| HP_FAILURE_TO_THRIVE | 12 | -0.311 | -0.98 | 0.47900 | 0.47900 | 0.66500 | 49 |
| GOCC_SECRETORY_GRANULE | 11 | 0.273 | 0.934 | 0.52700 | 0.52700 | 0.67800 | 11 |
| GOCC_SECRETORY_VESICLE | 11 | 0.273 | 0.934 | 0.52700 | 0.52700 | 0.67800 | 11 |
| HP_ABNORMALITY_OF_THE_KIDNEY | 10 | 0.278 | 0.917 | 0.53700 | 0.53700 | 0.67800 | 8 |
| HP_ABNORMALITY_OF_THE_URINARY_SYSTEM_PHYSIOLOGY | 11 | -0.304 | -0.933 | 0.54000 | 0.54000 | 0.67800 | 6 |
| HP_HYPERTONIA | 10 | -0.315 | -0.933 | 0.54300 | 0.54300 | 0.67800 | 43 |
| HP_ABNORMALITY_OF_THE_ABDOMINAL_ORGANS | 12 | -0.292 | -0.919 | 0.56400 | 0.56400 | 0.67800 | 49 |
| GOBP_PROTEOLYSIS | 18 | -0.256 | -0.914 | 0.56600 | 0.56600 | 0.67800 | 12 |
| GOCC_ENVELOPE | 18 | 0.224 | 0.913 | 0.57800 | 0.57800 | 0.67800 | 2 |
| GOCC_CATALYTIC_COMPLEX | 21 | -0.242 | -0.9 | 0.58100 | 0.58100 | 0.67800 | 19 |

| | | | | | | | |
|---|---|---|---|---|---|---|---|
| GOCC_ENDOPLASMIC_RETICULUM | 11 | 0.257 | 0.88 | 0.59900 | 0.59900 | 0.68600 | 59 |
| HP_ABNORMALITY_OF_COORDINATION | 15 | 0.231 | 0.884 | 0.62100 | 0.62100 | 0.69500 | 21 |
| GOMF_OXIDOREDUCTASE_ACTIVITY | 12 | -0.276 | -0.87 | 0.62800 | 0.62800 | 0.69500 | 40 |
| GOBP_SMALL_MOLECULE_METABOLIC_PROCESS | 19 | -0.236 | -0.856 | 0.64100 | 0.64100 | 0.69800 | 75 |
| GOBP_PROTEIN_CONTAINING_COMPLEX_ORGANIZATION | 13 | -0.26 | -0.841 | 0.66200 | 0.66200 | 0.70800 | 17 |
| HP_ABNORMAL_ABDOMEN_MORPHOLOGY | 11 | -0.266 | -0.814 | 0.68900 | 0.68900 | 0.72500 | 49 |
| GOBP_CELL_CYCLE | 12 | -0.255 | -0.804 | 0.70800 | 0.70800 | 0.73300 | 19 |
| GOBP_ORGANIC_ACID_METABOLIC_PROCESS | 13 | -0.244 | -0.789 | 0.73100 | 0.73100 | 0.74500 | 73 |
| HP_ABNORMAL_CIRCULATING_METABOLITE_CONCENTRATION | 12 | -0.244 | -0.769 | 0.74700 | 0.74700 | 0.75000 | 6 |
| GOBP_MONOCARBOXYLIC_ACID_METABOLIC_PROCESS | 12 | -0.241 | -0.758 | 0.76000 | 0.76000 | 0.75100 | 73 |
| GOBP_ORGANONITROGEN_COMPOUND_BIOSYNTHETIC_PROCESS | 13 | -0.229 | -0.74 | 0.78800 | 0.78800 | 0.76800 | 39 |
| GOBP_AMIDE_BIOSYNTHETIC_PROCESS | 10 | -0.238 | -0.707 | 0.82200 | 0.82200 | 0.78000 | 38 |
| GOBP_ORGANOPHOSPHATE_METABOLIC_PROCESS | 13 | -0.218 | -0.706 | 0.82400 | 0.82400 | 0.78000 | 75 |
| HP_UPPER_MOTOR_NEURON_DYSFUNCTION | 11 | -0.209 | -0.641 | 0.89100 | 0.89100 | 0.83000 | 18 |
| GOBP_CARBOHYDRATE_DERIVATIVE_METABOLIC_PROCESS | 12 | 0.178 | 0.629 | 0.91000 | 0.91000 | 0.83600 | 55 |
| HP_DECREASED_HEAD_CIRCUMFERENCE | 16 | -0.159 | -0.549 | 0.95200 | 0.95200 | 0.85400 | 45 |
| GOBP_CELLULAR_AMIDE_METABOLIC_PROCESS | 13 | -0.173 | -0.56 | 0.95500 | 0.95500 | 0.85400 | 40 |
| GOCC_INTRACELLULAR_PROTEIN_CONTAINING_COMPLEX | 13 | -0.163 | -0.528 | 0.96900 | 0.96900 | 0.85500 | 19 |

### Supplemental Figures

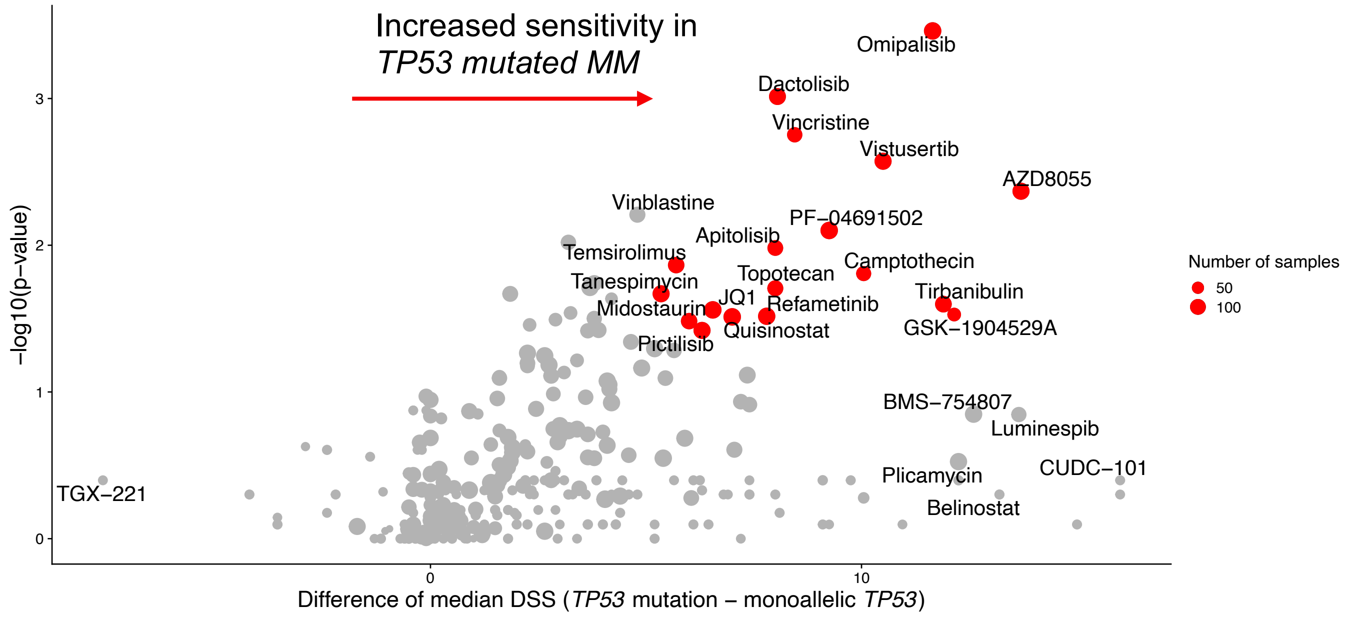

**Supplemental Figure S1. *Ex vivo* drug screening highlighting the difference in response to treatment between patients with *TP53* mutation and monoallelic *TP53*.** Volcano plot illustrating the comparison of drug sensitivity between MM samples with *TP53* mutation and monoallelic *TP53*. Compounds with significant difference are labeled (p-value ≤ 0.05), red dots represent compounds with significant difference below 5 DSS points.

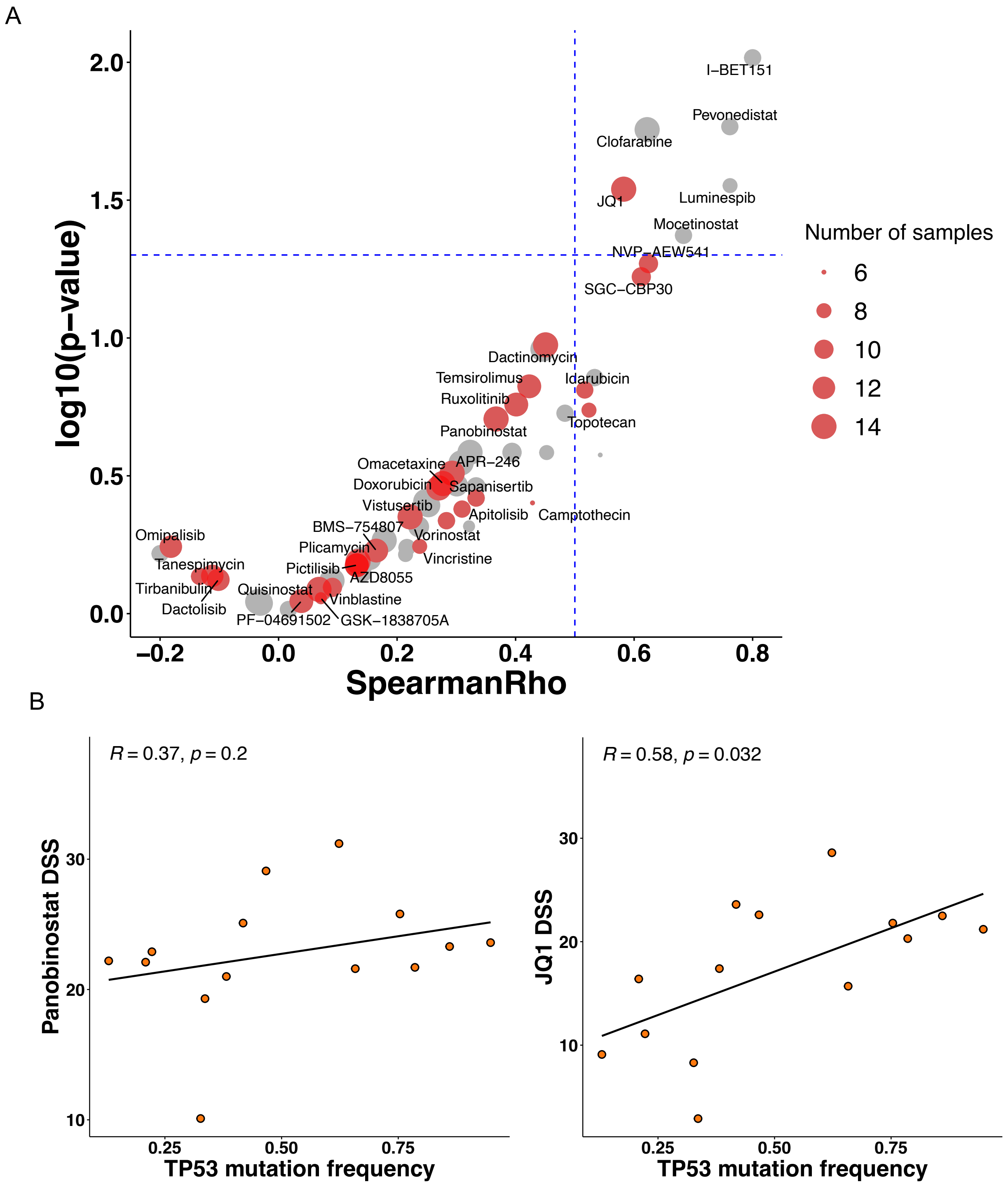

**Supplemental Figure S2. *TP53* mutation frequency weakly correlates with drug sensitivity of MM cells.** (A) Scatter plot summarizing results from the Spearman correlation of Variant Allele Frequency (VAF) for *TP53* mutation to DSS. The x-axis representes the Spearman correlation coefficient (SpearmanRho), indicating the strength and direction of the correlation between *TP53* VAF and DSS. The y-axis represents the negative logarithm (base10) of the p-value (log10(p-value)), indicating the statistical significance of the correlation. Compounds labeled in red show a significant increase in sensitivity in *TP53* mutated MM cells, as presented in **Figure 2B**. The blue lines indicate limits of significance for p-value and spearman Rho at 0.5. (B) Representative correlation between compounds and *TP53* mutation frequency.

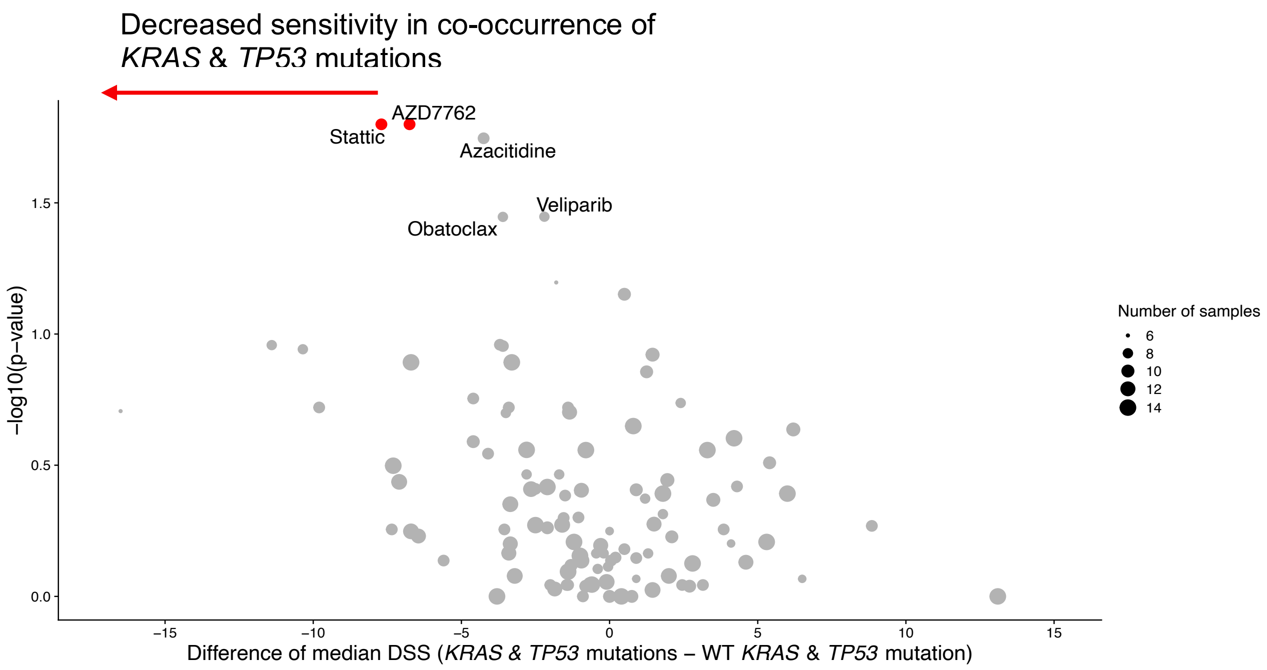

**Supplemental Figure S3.** ***Ex vivo*** **drug screening results highlighting the difference in response between MM samples with** ***KRAS*** **and** ***TP53*** **mutation co-occurrence compared to WT** ***KRAS*** **and** ***TP53*** **mutation.** Volcano plot illustrating the comparison of drug sensitivity between MM samples with *KRAS* and *TP53* mutation co-occurrence compared to WT *KRAS* and *TP53* mutation. Compounds with significant difference are labeled (p-value ≤ 0.05), red dots represent compounds with significant difference below 5 DSS points.

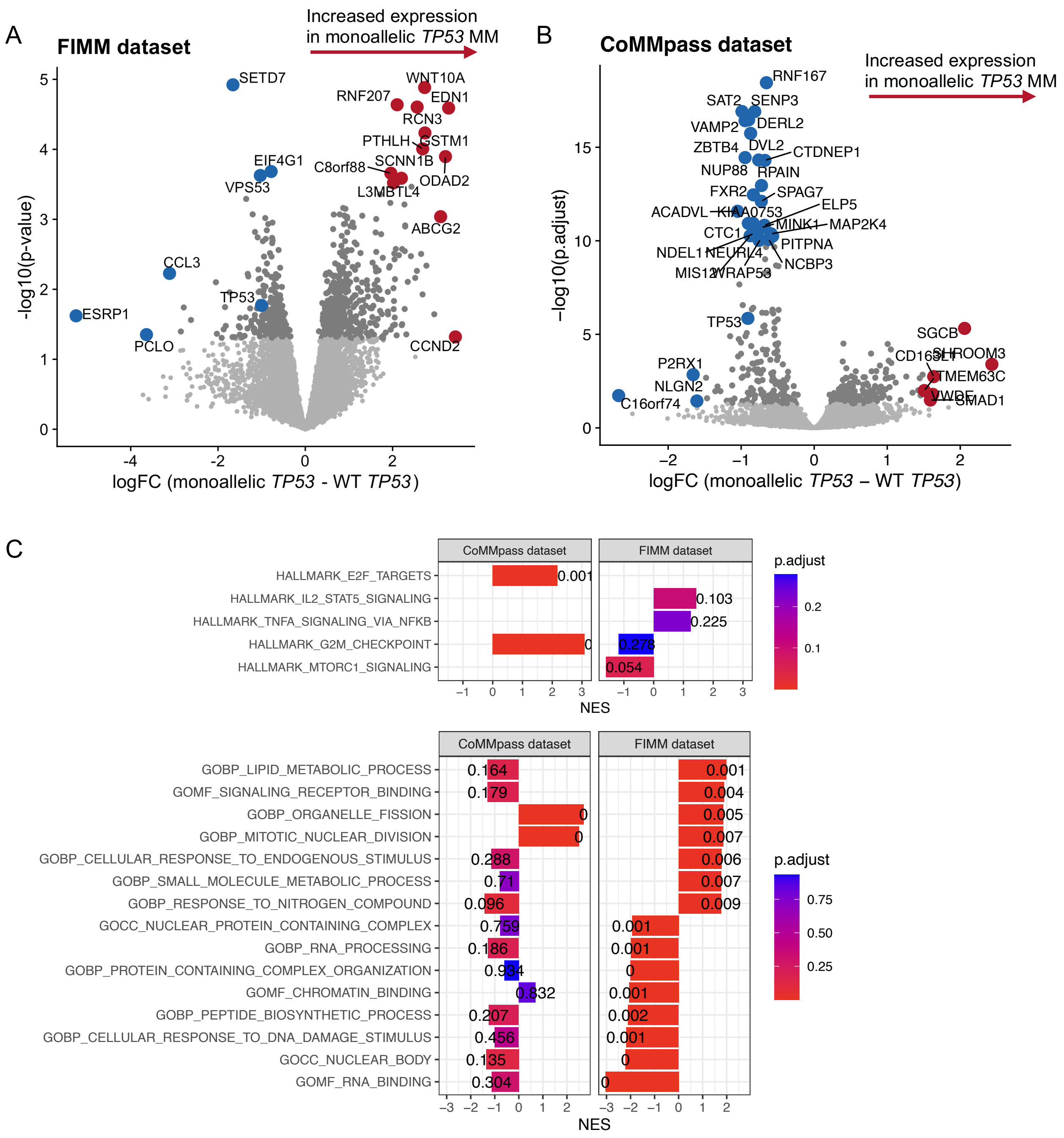

**Supplemental Figure S4. Monoallelic *TP53* leads to reduced expression of multiple genes in the 17p region.** (A) DGE in *TP53* mutated samples compared to WT *TP53* samples from the FIMM dataset. The x-axis shows the log2 fold change in expression and the y-axis shows the negative log10 of the p-value. Red dots represent genes with increased expression in monoallelic *TP53* MM, while decreased expression is represented with blue dots. (B) DGE analysis of the CoMMpass dataset, similarly presented, however the y-axis shows the negative log10 of the adjusted p-value with cut-off for significance for adjusted p-value ≤ 0.05. (C) GSEA results compared between both datasets, ordered by the NES of the FIMM dataset. The gene sets shown have an adjusted p-value $\leq 0.1$ in at least one of the two datasets.

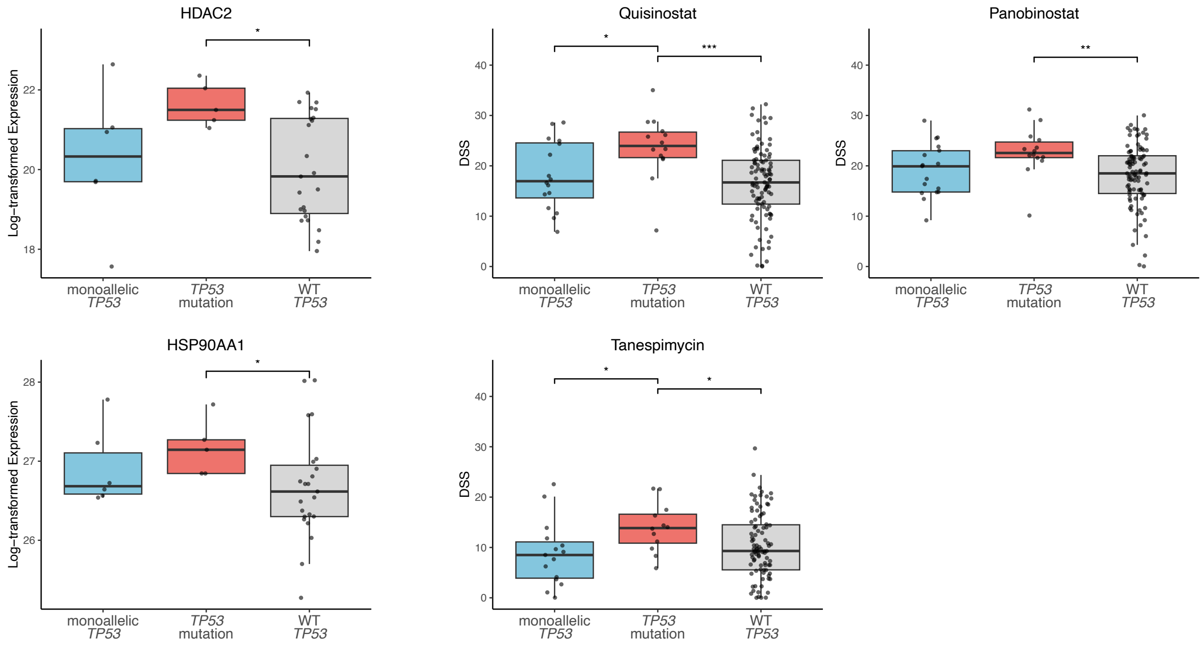

**Supplemental Figure S5. Drug target expression levels are associated with targeted inhibitor's sensitivity observed *ex vivo*.** Box plot showing the distribution of HDAC2 and HSP90AA1 protein expression levels and corresponding selective HDAC inhibitors quizinostat and panobinostat and HSP90 inhibitor tanespimycin. The center line represents the median value. The upper and lower limits of the box represent the upper (75th percentile) and lower (25th percentile) quartiles, respectively. The whiskers extend to the largest and smallest values within 1.5 times the interquartile range from the quartiles. Points beyond the whiskers are outliers and are plotted individually. Statistical significance indicated as * p-value < 0.05, ** p-value < 0.01, *** p-value < 0.001 by Wilcoxon rank-sum test.

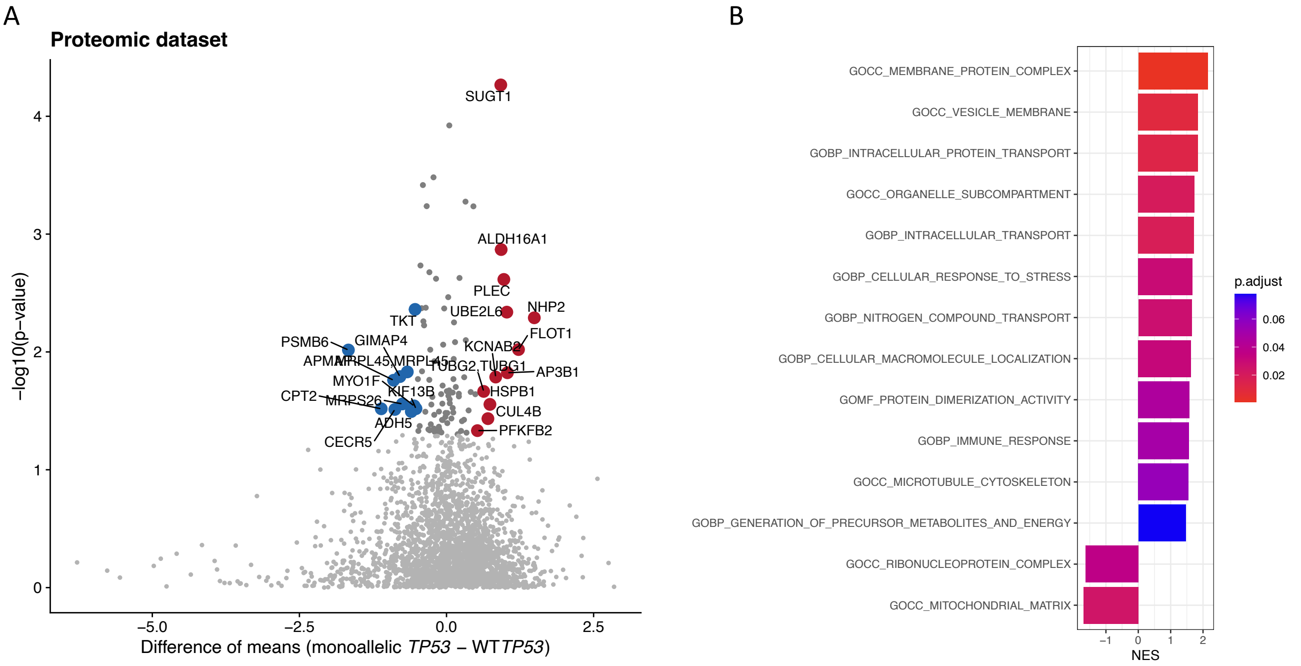

**Supplemental Figure S6. Monoallelic *TP53* MM has minimal impact on the MM proteomic profile.** (A) Significantly increased expression of proteins from samples with monoallelic *TP53* compared to WT *TP53* MM samples is represented in a volcano plot. The x-axis shows difference in mean level of protein expression, with the y-axis representing negative log10 of the p-value. Red dots represent proteins with increased expression in monoallelic *TP53* MM, while decreased expression is represented with blue dots. (B) GSEA results, ordered by the NES.
